# Sex Differences in Comparative Effectiveness and Safety of Second-line Antidiabetic Agents: Real-world Evidence from Large-scale Multinational Study

**DOI:** 10.64898/2026.04.10.26350252

**Authors:** Xiyu Ding, Vidhu Vadini, Chungsoo Kim, Fan Bu, Hsin Yi Chen, Yi Chai, Talita Duarte-Salles, Jason C. Hsu, Rohan Khera, Wallis C.Y. Lau, Kenneth K.C. Man, Paul Nagy, Anna Ostropolets, Andrea Pistillo, Nicole Pratt, Elena Roel, Sarah Seager, Mui Van Zandt, Lu Yuan, George Hripcsak, Nestoras Mathioudakis, Marc A Suchard, Akihiko Nishimura

## Abstract

**Importance:** Women have been under-represented in clinical trials of type 2 diabetes mellitus (T2D), and evidence on sex differences in effectiveness of T2D treatments remains limited.

**Objective:** To assess sex differences in comparative effectiveness and safety of four second-line antidiabetic agents: glucagon-like peptide-1 receptor agonists (GLP-1RA), sodium-glucose cotransporter-2 inhibitors (SGLT2i), dipeptidyl peptidase-4 inhibitors (DPP4i), and sulfonylureas (SU).

**Design:** Retrospective cohort study using an active-comparator new-user design, following each participant till treatment discontinuation or end of data.

**Setting:** Multinational study across ten real-world databases from the Observational Health Data Sciences and Informatics (OHDSI) network in the United States, United Kingdom, Germany, and Spain.

**Participants:** 5.15 million adults with T2D who initiated one of the four second-line therapies following metformin during 1992–2021.

**Exposures:** GLP-1RA, SGLT2i, DPP4i, or SU.

**Main Outcomes and Measures:** Cardiovascular effectiveness as measured through 7 outcomes (major adverse cardiovascular events and glycemic control) and safety through 18 outcomes as highlighted by ADA guideline. Hazard ratios (HRs) are estimated separately for women and men using propensity score-stratified Cox models with empirical calibration. Sex differences were tested using Z-tests on log-HR differences.

**Results:** Drug initiation rates differed by sex with 9.28% of women initiating on GLP-1RA, 11.91% SGLT2i, 27.81% DPP4i, and 50.99% SU; the rates among the men were 5.41%, 12.84%, 24.64%, and 57.10%. No significant sex differences were observed for cardiovascular effectiveness outcomes. Several safety outcomes showed significant sex differences that are consistent across drug comparisons. Focusing on GLP-1RA compared to SGLT2i for brevity, GLP-1RA users experienced the following comparative benefits and risks: higher risk of acute pancreatitis among women (HR 1.39 [1.13–1.70]) while non-differential risk among men (HR 0.91 [0.74–1.12]) with p = 0.005 for the test of difference; non-differential risk of hypotension among women (HR 1.08 [0.98–1.19]) while lower risk among men (HR 0.87 [0.78–0.96]) with p = 0.003. Where no sex differences were found, our findings were consistent with existing evidence.

**Conclusions and Relevance:** This large-scale multinational study on antidiabetic agents identified clinically relevant sex differences, which are biologically plausible but previously lacked clinical evidence. Our findings reinforce the importance of tailoring T2D management according to sex.

## Introduction

The management of type 2 diabetes mellitus (T2D) has been transformed with the emergence of glucagon-like peptide-1 receptor agonists (GLP-1RA) and sodium-glucose cotransporter-2 inhibitors (SGLT2i), which offer additional cardiovascular and renal benefits^1^. Current clinical guidelines recommend these drugs as the preferred second-line therapies following metformin and, in patients with established or high-risk cardiovascular or renal disease, as first-line therapies^2,3^. Beyond their cardio-renal benefits, these agents are considered generally safer than older agents such as dipeptidyl peptidase-4 inhibitors (DPP4i) and sulfonylureas (SU)^1^.

Despite the growing clinical preference for GLP-1RA and SGLT2i, limited evidence exists on their comparative effectiveness and safety in real-world populations, both between these two drug classes and against older agents^4,5^. Most randomized controlled trials (RCTs) have evaluated these drugs against placebo or as add-on therapies, providing no direct comparisons. The dearth of evidence is particularly pronounced among clinically relevant subpopulations, who are often inadequately represented in RCTs.

Particularly important among such understudied populations are women. Most cardiovascular outcome trials of antidiabetic agents enrolled only 20-40% women, limiting detection of sex specific differences in efficacy and safety^6,7^. While the prevalence of T2D is lower than in men, women experience greater cardiovascular morbidity and mortality than men ^8–10^. Additionally, hormonal factors and comorbidities may predispose women to unique benefits and risks from specific drugs^11^. Given these important biological differences, it is critical to study effectiveness and safety profiles of antidiabetic agents separately by men and women.

Real-world data from electronic health records (EHR) and administrative claims present an opportunity to fill these critical evidence gaps on sex differences in T2D treatments. Even with such databases’ larger sample sizes and more balanced representation, observational studies confined to a single database often lack sufficient statistical power to detect differences between the sexes. Single-database studies also leave concerns about the generalizability and robustness of their findings.

To overcome the limitations of existing studies and make the most of available real-world data, we conducted a multi-national, multi-database, federated study to comprehensively characterize effectiveness and safety profiles of four second-line T2DM medications: GLP-1RA, SGLT2i, DPP4i, and SU. We estimated their relative effectiveness and safety separately in women and men and tested for sex differences across a broad set of effectiveness and safety endpoints. We followed a pre-specified, published analysis plan implementing established best practices to ensure the rigor of the resulting evidence^12^.

## Methods

### Data

Conducted as part of the Large-scale Evidence Generation and Evaluation across a Network of Databases (LEGEND) initiative by the Observational Health Data Sciences and Informatics (OHDSI) collaborative, this study included ten databases—six claims and four EHR databases—representing patient experiences from the United States, Germany, Spain and the United Kingdom and covering the periods 1992–2021 (Table 1). All participating databases have been standardized to the Observational Medical Outcomes Partnership (OMOP) common data model version 5.3. All data were de-identified with their use approved in compliance with local regulations. Details of the databases and represented populations are provided in Supplement A, Table 1.

**Table 1.** Population size, median on-treatment time, and median follow-up time for new users of the second-line antihyperglycemic agents across data sources.

| Drug Class | Data Source | Female |  |  | Male |  |  |
| --- | --- | --- | --- | --- | --- | --- | --- |
|  |  | Patients, No. | On-treatment time Median (IQR), days | Total Follow-up time Median (IQR), days | Patients, No. | On-treatment time Median (IQR), days | Total Follow-up time Median (IQR), days |
| GLP-1RA | CCAE | 20,594 | 100 (29–253) | 518 (206–1,087) | 12,998 | 108 (36–276) | 464 (109–943) |
|  | DAG | 390 | 83 (29–286) | 608 (285–1,273) | 401 | 85 (29–299) | 690 (281–1,505) |
|  | ODOD | 11,664 | 90 (29–223) | 373 (141–779) | 8,520 | 94 (29–236) | 362 (141–743) |
|  | MDCR | 1,231 | 89 (29–263) | 572 (171–1,206) | 1,008 | 99 (31–270) | 482 (152–1,074) |
|  | MDCD | 2,034 | 90 (29–226) | 601 (260–1,177) | 727 | 95 (29–203) | 490 (208–954) |
|  | IMRD | 635 | 169 (64–407) | 1,325 (560–2,189) | 577 | 197 (74–408) | 1,387 (583–2,298) |
|  | Open Claims | 170,648 | 97 (31–250) | 650 (265–1,293) | 117,212 | 105 (33–269) | 642 (268–1,244) |
|  | OEHR | 13,640 | 55 (29–97) | 597 (311–1,127) | 9,105 | 54 (29–92) | 582 (306–1,045) |
|  | VA | 303 | 97 (30–266) | 370 (116–751) | 1,584 | 98 (33–253) | 358 (137–755) |
|  | SIDIAP | 636 | 217 (76–616) | 716 (211–1,930) | 543 | 251 (90–582) | 756 (238–1,940) |
|  | **Total (Eligible)** |  | **221,775 (219,811)** |  |  | **152,675 (150,427)** |  |
| SGLT2i | CCAE | 18,887 | 120 (47–293) | 543 (231–1,076) | 24,150 | 154 (59–354) | 513 (215–1,002) |
|  | DAG | 2,338 | 132 (97–318) | 691 (322–1,214) | 4,062 | 154 (97–356) | 642 (310–1,197) |
|  | ODOD | 12,845 | 98 (29–247) | 461 (174–912) | 17,486 | 121 (53–289) | 440 (163–862) |
|  | MDCR | 1,489 | 92 (29–258) | 415 (159–891) | 2,137 | 118 (40–338) | 425 (155–872) |
|  | MDCD | 2,435 | 88 (29–198) | 571 (243–1,105) | 1,507 | 95 (29–251) | 496 (213–966) |
|  | IMRD | 1,759 | 195 (67–431) | 523 (243–922) | 2,386 | 181 (60–454) | 521 (219–886) |
|  | Open Claims | 222,787 | 104 (30–275) | 947 (405–1,768) | 265,602 | 141 (57–349) | 870 (377–1,629) |
|  | OEHR | 16,666 | 56 (29–101) | 765 (378–1,346) | 20,231 | 55 (29–100) | 694 (356–1,244) |
|  | VA | 930 | 90 (46–195) | 265 (116–563) | 14,178 | 111 (57–247) | 288 (125–601) |
|  | SIDIAP | 2,956 | 246 (90–628) | 611 (211–1,262) | 5,168 | 281 (97–644) | 561 (198–1,010) |
|  | **Total (Eligible)** |  | **283,092 (282,162)** |  |  | **356,902** |  |
| DPP4i | CCAE | 45,501 | 119 (48–303) | 718 (291–1,428) | 52,897 | 146 (59–344) | 718 (294–1,413) |
|  | DAG | 10,124 | 196 (97–447) | 1,261 (604–2,238) | 13,128 | 195 (97–441) | 1,282 (610–2,288) |
|  | ODOD | 29,521 | 118 (43–299) | 770 (316–1,495) | 30,865 | 128 (54–321) | 779 (319–1,526) |
|  | MDCR | 8,813 | 158 (60–393) | 817 (339–1,538) | 9,568 | 180 (76–435) | 829 (335–1,557) |
|  | MDCD | 7,773 | 101 (29–247) | 863 (383–1,496) | 4,188 | 112 (30–266) | 804 (365–1,433) |
|  | IMRD | 8,548 | 250 (87–587) | 1,049 (481–1,852) | 12,175 | 280 (99–639) | 1,043 (471–1,824) |
|  | Open Claims | 487,437 | 113 (29–311) | 1,738 (917–2,630) | 470,166 | 126 (46–347) | 1,747 (946–2,625) |
|  | OEHR | 38,897 | 89 (89–174) | 1,210 (613–1,970) | 38,289 | 89 (89–172) | 1,187 (609–1,941) |
|  | VA | 2,334 | 112 (60–284) | 666 (316–1,259) | 26,562 | 163 (89–364) | 742 (344–1,333) |
|  | SIDIAP | 19,644 | 478 (142–1,076) | 1,386 (624–2,457) | 26,891 | 541 (180–1,127) | 1,335 (649–2,413) |
|  | **Total** |  | **658,592** |  |  | **684,724** |  |
| SU | CCAE | 78,632 | 113 (41–298) | 731 (295–1,513) | 97,654 | 146 (59–362) | 720 (293–1,465) |
|  | DAG | 3,301 | 183 (119–419) | 2,273 (1,102–3,670) | 3,726 | 185 (119–418) | 2,229 (1,089–3,669) |
|  | ODOD | 60,389 | 135 (56–356) | 806 (318–1,588) | 70,808 | 160 (62–393) | 767 (307–1,528) |
|  | MDCR | 20,652 | 155 (57–404) | 889 (362–1,783) | 23,623 | 190 (74–487) | 904 (356–1,806) |
|  | MDCD | 16,863 | 90 (29–235) | 938 (414–1,702) | 9,898 | 110 (29–276) | 860 (368–1,550) |
|  | IMRD | 2,855 | 303 (88–750) | 2,224 (1,102–3,546) | 3,793 | 359 (112–863) | 2,223 (1,105–3,416) |
|  | Open Claims | 918,353 | 136 (57–362) | 1,679 (833–2,617) | 965,429 | 173 (73–435) | 1,612 (795–2,529) |
|  | OEHR | 83,370 | 90 (89–195) | 1,325 (661–2,144) | 95,917 | 89 (89–194) | 1,278 (631–2,097) |
|  | VA | 15,658 | 145 (90–329) | 2,269 (1,115–3,778) | 305,887 | 187 (90–448) | 2,554 (1,295–4,026) |
|  | SIDIAP | 7,595 | 576 (160–1,223) | 3,137 (1,987–4,066) | 9,904 | 664 (196–1,379) | 3,054 (1,941–3,929) |
|  | **Total** |  | **1,207,668** |  |  | **1,586,639** |  |
| **Overall (Eligible)** |  |  | **2,371,127 (2,368,233)** |  |  | **2,780,940 (2,778,692)** |  |

We greyed out database-drug pairs with < 1,000 new users; these pairs are excluded from the subsequent analyses.
Abbreviations: SGLT2i = sodium-glucose co-transporter-2 inhibitor; GLP-1 RA = glucagon-like peptide-1 receptor agonist; DPP4i = dipeptidyl peptidase-4 inhibitor; SU = sulfonylurea.

### Study design and exposures

We conducted a retrospective cohort study on patients with T2DM diagnoses who were prescribed one of the four second-line agents following metformin monotherapy. A schematic of the study design is presented in Figure 1, and the full list of ingredient-level agents for the four second-line drug classes is in Supplement A, Table 2. We additionally required that patients within each drug cohort have: (1) ≥1 year of continuous observation prior to the index date, (2) ≥90 days of prior metformin use, and (3) no prior exposure to long-term (>30 days) insulin therapy, comparator drugs, or other glucose-lowering medications. To ensure sufficient statistical power, we limited our analyses to comparisons in which each drug cohort included at least 1,000 patients.

**Table 2.** Baseline Patient Characteristics for New Female/Male Users of GLP-1RA (T) and SGLT2i (C) in the USOC database.

| Characteristic | Female |  |  |  |  |  | Male |  |  |  |  |  |
| --- | --- | --- | --- | --- | --- | --- | --- | --- | --- | --- | --- | --- |
|  | Before stratification |  |  | After stratification |  |  | Before stratification |  |  | After stratification |  |  |
|  | T (%) | C (%) | StdDiff | T (%) | C (%) | StdDiff | T (%) | C (%) | StdDiff | T (%) | C (%) | StdDiff |
| **Age group** |  |  |  |  |  |  |  |  |  |  |  |  |
| 15-19 | 0.2 | 0.1 | 0.02 | 0.2 | 0.2 | 0.01 | 0.2 | 0.1 | 0.03 | 0.1 | 0.1 | 0.02 |
| 20-24 | 0.9 | 0.5 | 0.05 | 0.7 | 0.7 | 0.00 | 0.5 | 0.3 | 0.04 | 0.4 | 0.3 | 0.01 |
| 25-29 | 1.7 | 1.0 | 0.06 | 1.3 | 1.3 | 0.00 | 1.0 | 0.6 | 0.05 | 0.7 | 0.7 | 0.01 |
| 30-34 | 3.1 | 1.9 | 0.08 | 2.5 | 2.4 | 0.00 | 2.3 | 1.5 | 0.06 | 1.7 | 1.7 | 0.00 |
| 35-39 | 5.4 | 3.6 | 0.09 | 4.5 | 4.4 | 0.00 | 4.7 | 3.4 | 0.07 | 3.7 | 3.8 | 0.00 |
| 40-44 | 8.5 | 6.3 | 0.08 | 7.2 | 7.3 | 0.00 | NA | NA | NA | NA | NA | NA |
| 45-49 | 12.0 | 9.8 | 0.07 | 10.7 | 10.8 | 0.00 | 11.8 | 10.0 | 0.06 | 10.5 | 10.6 | 0.00 |
| 50-54 | 15.5 | 14.0 | 0.04 | 14.5 | 14.7 | 0.00 | 15.1 | 13.8 | 0.04 | 14.2 | 14.2 | 0.00 |
| 55-59 | 16.4 | 16.6 | -0.01 | 16.7 | 16.4 | 0.01 | 16.8 | 16.6 | 0.00 | 16.5 | 16.7 | 0.00 |
| 60-64 | 14.5 | 16.4 | -0.05 | 15.4 | 15.5 | 0.00 | 15.5 | 17.1 | -0.04 | 16.7 | 16.6 | 0.00 |
| 65-69 | 10.8 | 12.8 | -0.06 | 11.9 | 12.0 | 0.00 | 11.6 | 13.3 | -0.05 | 12.8 | 12.7 | 0.00 |
| 70-74 | 6.5 | 8.7 | -0.09 | 7.9 | 7.7 | 0.01 | 7.1 | 8.9 | -0.07 | 8.3 | 8.4 | 0.00 |
| 75-79 | 3.0 | 5.2 | -0.11 | 4.2 | 4.2 | 0.00 | 3.6 | 5.4 | -0.09 | 4.8 | 4.9 | 0.00 |
| 80-84 | 1.3 | 2.9 | -0.11 | 2.1 | 2.2 | -0.01 | 1.6 | 2.8 | -0.08 | 2.5 | 2.4 | 0.00 |
| 85-89 | 0.1 | 0.2 | -0.03 | 0.1 | 0.2 | -0.01 | 0.1 | 0.2 | -0.03 | 0.1 | 0.2 | -0.02 |
| **Medical history: General** |  |  |  |  |  |  |  |  |  |  |  |  |
| Acute respiratory disease | 16.8 | 16.3 | 0.01 | 16.5 | 16.7 | -0.01 | 11.3 | 11.0 | 0.01 | 11.1 | 11.2 | 0.00 |
| Attention deficit hyperactivity disorder | 1.2 | 0.6 | 0.06 | 0.9 | 0.9 | 0.00 | 1.0 | 0.6 | 0.05 | 0.7 | 0.7 | 0.00 |
| Chronic liver disease | 0.8 | 1.1 | -0.04 | 1.0 | 1.0 | 0.00 | 0.8 | 1.0 | -0.02 | 0.9 | 0.9 | 0.00 |
| Chronic obstructive lung disease | 4.1 | 4.5 | -0.02 | 4.4 | 4.5 | 0.00 | 3.9 | 4.2 | -0.02 | 4.2 | 4.2 | 0.00 |
| Crohn disease | NA | NA | NA | NA | NA | NA | 0.1 | 0.1 | 0.00 | 0.1 | 0.1 | 0.00 |
| Dementia | 0.3 | 0.5 | -0.03 | 0.4 | 0.4 | 0.00 | NA | NA | NA | NA | NA | NA |
| Depressive disorder | 12.8 | 9.8 | 0.10 | 11.3 | 11.3 | 0.00 | 6.6 | 4.6 | 0.08 | 5.3 | 5.3 | 0.00 |
| Gastroesophageal reflux disease | 10.9 | 10.0 | 0.03 | 10.5 | 10.5 | 0.00 | 7.5 | 6.9 | 0.02 | 7.1 | 7.1 | 0.00 |
| Gastrointestinal hemorrhage | 1.1 | 1.2 | -0.01 | 1.2 | 1.2 | 0.00 | 1.1 | 1.1 | 0.00 | 1.1 | 1.2 | 0.00 |
| Human immunodeficiency virus infection | 0.2 | 0.2 | 0.01 | 0.2 | 0.2 | 0.00 | 0.5 | 0.4 | 0.02 | 0.4 | 0.4 | 0.00 |
| Hyperlipidemia | 37.9 | 42.7 | -0.10 | 40.3 | 40.8 | -0.01 | 42.8 | 46.0 | -0.06 | 44.8 | 45.1 | 0.00 |
| Hypertensive disorder | 44.9 | 48.2 | -0.07 | 46.6 | 47.0 | -0.01 | 49.1 | 49.9 | -0.02 | 49.7 | 49.7 | 0.00 |
| Lesion of liver | 0.5 | 0.6 | -0.01 | 0.5 | 0.6 | 0.00 | 0.6 | 0.6 | 0.00 | 0.6 | 0.6 | -0.01 |
| Obesity | 21.6 | 14.4 | 0.19 | 17.9 | 17.7 | 0.01 | 18.0 | 11.4 | 0.19 | 13.8 | 13.2 | 0.02 |
| Osteoarthritis | 17.2 | 15.5 | 0.04 | 16.4 | 16.6 | 0.00 | 13.3 | 11.8 | 0.04 | 12.5 | 12.2 | 0.01 |
| Pneumonia | 2.2 | 2.1 | 0.01 | 2.2 | 2.2 | 0.00 | 2.0 | 2.0 | 0.00 | 2.0 | 2.0 | 0.00 |
| Psoriasis | 1.4 | 1.0 | 0.03 | 1.2 | 1.2 | 0.00 | 1.3 | 1.0 | 0.02 | 1.2 | 1.1 | 0.01 |
| Renal impairment | 4.0 | 4.8 | -0.04 | 4.6 | 4.6 | 0.00 | 4.7 | 5.4 | -0.03 | 5.3 | 5.2 | 0.00 |
| Rheumatoid arthritis | 1.5 | 1.3 | 0.01 | 1.4 | 1.4 | 0.00 | 0.6 | 0.5 | 0.00 | 0.5 | 0.6 | 0.00 |
| Schizophrenia | NA | NA | NA | NA | NA | NA | 0.3 | 0.3 | 0.01 | 0.3 | 0.3 | 0.00 |
| Ulcerative colitis | 0.2 | 0.2 | 0.00 | 0.2 | 0.2 | 0.00 | 0.2 | 0.2 | 0.00 | 0.2 | 0.2 | 0.00 |
| Urinary tract infectious disease | 6.6 | 6.4 | 0.01 | 6.6 | 6.5 | 0.00 | 1.9 | 1.8 | 0.01 | 1.9 | 1.8 | 0.00 |
| Viral hepatitis C | 0.2 | 0.2 | -0.01 | 0.2 | 0.2 | 0.00 | 0.3 | 0.3 | -0.01 | 0.3 | 0.3 | 0.01 |
| Visual system disorder | 16.9 | 18.4 | -0.04 | 17.9 | 17.8 | 0.00 | 14.5 | 15.6 | -0.03 | 15.2 | 15.2 | 0.00 |
| **Medical history: Cardiovascular disease** |  |  |  |  |  |  |  |  |  |  |  |  |
| Atrial fibrillation | 1.8 | 2.7 | -0.06 | 2.3 | 2.3 | 0.00 | 3.6 | 4.6 | -0.05 | 4.2 | 4.3 | 0.00 |
| Cerebrovascular disease | 1.8 | 2.4 | -0.04 | 2.2 | 2.1 | 0.00 | 1.9 | 2.6 | -0.05 | 2.4 | 2.5 | -0.01 |
| Coronary arteriosclerosis | 3.6 | 5.7 | -0.10 | 4.7 | 4.8 | 0.00 | 8.2 | 12.0 | -0.13 | 10.6 | 10.9 | -0.01 |
| Heart disease | 11.3 | 14.8 | -0.10 | 13.3 | 13.4 | 0.00 | 16.3 | 21.2 | -0.13 | 19.4 | 19.8 | -0.01 |
| Heart failure | 2.3 | 4.0 | -0.10 | 3.1 | 3.4 | -0.02 | 2.9 | 5.2 | -0.11 | 4.1 | 4.6 | -0.03 |
| Ischemic heart disease | 2.0 | 3.2 | -0.08 | 2.6 | 2.7 | -0.01 | 3.5 | 5.5 | -0.10 | 4.7 | 5.0 | -0.01 |
| Peripheral vascular disease | 3.2 | 3.9 | -0.04 | 3.5 | 3.6 | 0.00 | 4.2 | 4.6 | -0.02 | 4.5 | 4.6 | 0.00 |
| Pulmonary embolism | 0.5 | 0.5 | 0.01 | 0.5 | 0.5 | 0.01 | 0.5 | 0.4 | 0.01 | 0.5 | 0.4 | 0.00 |
| Venous thrombosis | NA | NA | NA | NA | NA | NA | 0.7 | 0.8 | -0.01 | 0.7 | 0.8 | 0.00 |
| **Medical history: Neoplasms** |  |  |  |  |  |  |  |  |  |  |  |  |
| Hematologic neoplasm | 0.4 | 0.5 | -0.01 | 0.5 | 0.5 | 0.00 | 0.5 | 0.6 | 0.00 | 0.6 | 0.6 | 0.00 |
| Malignant lymphoma | 0.2 | 0.3 | -0.01 | 0.3 | 0.3 | 0.00 | 0.3 | 0.4 | -0.01 | 0.3 | 0.4 | 0.00 |
| Malignant neoplasm of anorectum | 0.1 | 0.1 | -0.01 | 0.1 | 0.1 | 0.00 | 0.1 | 0.1 | -0.01 | 0.1 | 0.1 | -0.01 |
| Malignant neoplastic disease | 4.4 | 5.0 | -0.03 | 4.8 | 4.7 | 0.00 | 4.4 | 4.9 | -0.03 | 4.8 | 4.7 | 0.00 |
| Malignant tumor of breast | 1.7 | 2.0 | -0.03 | 1.9 | 1.9 | 0.00 | NA | NA | NA | NA | NA | NA |
| Malignant tumor of colon | 0.1 | 0.2 | -0.01 | 0.2 | 0.2 | 0.00 | 0.2 | 0.3 | -0.01 | 0.2 | 0.2 | 0.00 |
| Malignant tumor of lung | 0.1 | 0.2 | -0.01 | 0.1 | 0.1 | 0.00 | 0.1 | 0.1 | -0.01 | 0.1 | 0.1 | 0.00 |
| Malignant tumor of urinary bladder | 0.1 | 0.1 | 0.00 | 0.1 | 0.1 | 0.00 | 0.3 | 0.3 | 0.00 | 0.3 | 0.3 | 0.01 |
| Primary malignant neoplasm of prostate | NA | NA | NA | NA | NA | NA | 1.5 | 1.7 | -0.02 | 1.7 | 1.6 | 0.00 |
| **Medication use** |  |  |  |  |  |  |  |  |  |  |  |  |
| Agents acting on renin-angiotensin system | 57.2 | 64.6 | -0.15 | 61.2 | 61.8 | -0.01 | 69.0 | 71.5 | -0.06 | 70.8 | 70.9 | 0.00 |
| Antibacterials for systemic use | 63.0 | 61.0 | 0.04 | 61.9 | 62.4 | -0.01 | 50.5 | 49.0 | 0.03 | 49.7 | 49.5 | 0.00 |
| Antidepressants | 47.3 | 38.3 | 0.18 | 42.7 | 42.9 | 0.00 | 27.4 | 21.1 | 0.15 | 23.3 | 23.2 | 0.00 |
| Antiepileptics | 26.4 | 21.9 | 0.10 | 24.2 | 24.3 | 0.00 | 17.7 | 14.7 | 0.08 | 16.0 | 15.6 | 0.01 |
| Anti-inflammatory and antirheumatic products | 41.9 | 40.5 | 0.03 | 41.3 | 41.1 | 0.00 | 33.2 | 32.0 | 0.03 | 32.6 | 32.3 | 0.00 |
| Antineoplastic agents | 6.3 | 5.2 | 0.04 | 5.7 | 5.7 | 0.00 | 4.5 | 3.9 | 0.03 | 4.2 | 4.1 | 0.00 |
| Antipsoriatics | 2.1 | 1.5 | 0.04 | 1.7 | 1.7 | 0.00 | 0.6 | 0.5 | 0.01 | 0.6 | 0.5 | 0.00 |
| Antithrombotic agents | NA | NA | NA | NA | NA | NA | 16.8 | 21.3 | -0.11 | 19.7 | 20.0 | -0.01 |
| Beta blocking agents | 27.4 | 31.4 | -0.09 | 29.7 | 30.0 | -0.01 | 30.3 | 35.1 | -0.10 | 33.6 | 33.7 | 0.00 |
| Calcium channel blockers | 20.4 | 23.1 | -0.07 | 22.0 | 22.0 | 0.00 | 25.6 | 25.7 | 0.00 | 25.8 | 25.9 | 0.00 |
| Diuretics | 43.0 | 43.4 | -0.01 | 43.4 | 43.5 | 0.00 | 37.3 | 36.3 | 0.02 | 36.5 | 36.7 | 0.00 |
| Drugs for acid related disorders | 37.5 | 36.3 | 0.02 | 37.1 | 37.2 | 0.00 | NA | NA | NA | NA | NA | NA |
| Drugs for obstructive airway diseases | 46.8 | 43.2 | 0.07 | 45.0 | 45.3 | -0.01 | 34.4 | 31.6 | 0.06 | 32.7 | 32.4 | 0.01 |
| Immunosuppressants | 4.1 | 3.5 | 0.03 | 3.8 | 3.8 | 0.00 | 2.2 | 1.9 | 0.02 | 2.1 | 2.0 | 0.00 |
| Lipid modifying agents | 60.7 | 68.5 | -0.16 | 65.4 | 65.2 | 0.00 | 71.1 | 75.3 | -0.10 | 74.1 | 74.1 | 0.00 |
| Opioids | 26.9 | 25.7 | 0.03 | 26.6 | 26.4 | 0.00 | 21.7 | 20.6 | 0.03 | 21.3 | 21.1 | 0.00 |
| Psycholeptics | 30.3 | 26.4 | 0.09 | 28.4 | 28.4 | 0.00 | 20.0 | 17.4 | 0.07 | 18.3 | 18.3 | 0.00 |
| Psychostimulants | 5.6 | 3.5 | 0.10 | 4.5 | 4.4 | 0.00 | NA | NA | NA | NA | NA | NA |

We also reported the standardized difference of population proportions (StdDiff) before and after propensity score adjustment. Stratification improved patient cohort balance, reducing all StdDiffs to well below the recommended threshold. Abbreviations: SGLT2i = sodium-glucose co-transporter-2 inhibitor; GLP-1RA = glucagon-like peptide-1 receptor agonist.

**Figure 1.**
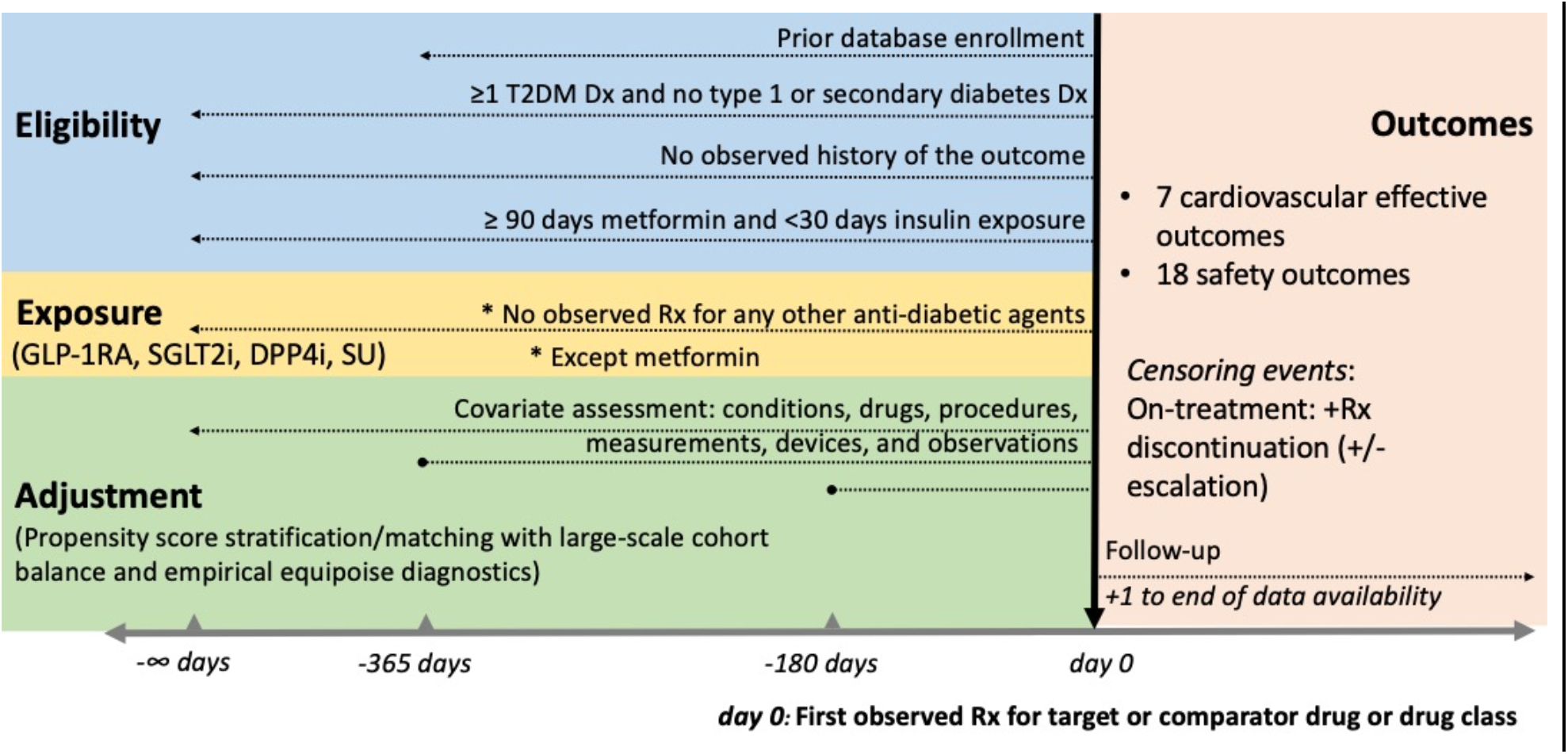
Study design schematic.

We used a new-user, active-comparator design to approximate a hypothetical target trial, comparing one drug class (target) against another (comparator), taking the first prescription of the respective agents as the index date^13,14^.

### Outcomes

We assessed comparative effectiveness through 7 cardiovascular outcomes and safety through 18 adverse outcomes.

For the cardiovascular outcomes, we used 3- and 4-point major adverse cardiovascular events (MACE) as the two primary outcomes, with the former consisting of acute myocardial infarction, stroke, and sudden cardiac death and the latter consisting additionally of heart failure with hospitalization. The secondary outcomes of interest included the four individual components of MACE, and glycemic control, defined as first hemoglobin A1c measurement with value ≤ 7%.

For the safety outcomes, we identified them as highlighted in the American Diabetes Association guidelines^15^ and previous RCTs. We categorized these as 1) “cardiovascular”: hypotension, venous thromboembolism, peripheral edema; 2)”metabolic/endocrine”: abnormal weight gain, abnormal weight loss, diabetic ketoacidosis, hypoglycemia; 3) “gastrointestinal”: acute pancreatitis, diarrhea, nausea, vomiting;, 4) “renal”: acute renal failure, hyperkalemia; 5) “musculoskeletal/peripheral vascular”: bone fracture, joint pain, lower extremity amputation; and 6) others: photosensitivity, genitourinary infection. Outcome phenotype definitions, which have been validated and deployed in previous work, are detailed in the published protocol ^12^.

### Statistical analyses

For each outcome, we restricted our analyses to patients with no outcome history prior to treatment initiation. We employed an on-treatment time-at-risk definition, following patients from the day after treatment initiation to the earliest of treatment discontinuation or escalation with additional T2D agents^12^.

For each target-comparator pair, we calculated propensity scores (PS) with L1-regularized logistic regression that includes thousands of covariates, covering a wide range of baseline clinical information^16^. We stratified patients based on PS deciles and used the stratified Cox proportional hazard model to estimate hazard ratios (HR). To address residual bias, we empirically calibrated the HR estimates and 95% confidence intervals (CIs) using up to 100 pre-selected negative control outcomes, a list of which are provided in Supplement A, Table 3.2^17,18^.

We calculated the pooled HR estimate, 95% CI, and p-value for each comparison by applying random-effects meta-analysis to the estimates from individual databases^19^. We applied Z-test to the difference between the log-HR estimates from the male and female cohorts to quantify significance of sex differences^20^. We highlighted an HR estimate as potentially significant if the p-value was below 0.05.

Analyses were performed on R 4.2.3, using the study package openly available at https://github.com/ohdsi-studies/LegendT2dm. We reported the results in accordance with STROBE guidelines^21^.

### Diagnostics and Sensitivity analyses

To ensure reliability of the reported results, we applied a suite of study diagnostics. Specifically, our meta-analysis only included the individual-database results that met the following criteria: 1) empirical equipoise: ≥25% of patients from both target and comparator cohorts having PS of 0.3–0.7 to ensure sufficient overlap; 2) covariate balance: standardized mean differences <0.15 across all covariates after PS adjustment to minimize residual confounding; and 3) minimum detectable relative risks (MDRR) of 4.0. These thresholds were preselected based on established best practices for observational studies^22^. The full diagnostic results are available in the supplement and at https://data.ohdsi.org/LegendT2dmClassEvidenceExplorer/.

We additionally conducted two sensitivity analyses. First, we assessed to what extent the estimates were influenced by the largest database in our study, the US Open Claims (USOC). We also recalculated the HR estimates under an alternative time-at-risk definition, in which we allow for treatment escalation while remaining on the initially prescribed drug.

## Results

### Patient characteristics

Table 1 presents the patient population size, median on-treatment time, and median follow-up time for each participating database. We identified 5,146,925 T2D patients meeting the inclusion criteria, the largest contributions coming from USOC (3,617,634), VA (366,198), CCAE (351,313), and OptumEHR (316,115). Women represented 46.01% (2,368,233) of the cohort. Among the women, the fractions initiating on GLP-1RA, SGLT2i, DPP4i, and SU were 9.28%, 11.91%, 27.81%, and 50.99% respectively; among the men, the fractions were 5.41%, 12.84%, 24.64%, and 57.10%. Notably, GLP1-RA initiation was more common among women.

Table 2 presents the baseline characteristics by sex before and after PS stratification of GLP-1RA and SGLT2i users in the USOC database. For the other drug pairs and databases, analogous tables are provided in Supplement B and C, Table 157-199.

### Effectiveness for cardiovascular protection and glycemic control

Figure 2 presents the HR estimates and CIs for all effectiveness outcomes and comparisons. Compared with SU, the use of GLP-1RA, SGLT2i, and DPP4i were associated with lower risks of cardiovascular outcomes, though no significant sex differences were observed. For 3-point MACE specifically, HRs relative to SU were 0.73 (95% CI, 0.57–0.92) and 0.67 (0.47–0.96) among women and men for GLP-1RA; 0.76 (0.63–0.91) and 0.77 (0.64– 0.92) for SGLT2i; and 0.81 (0.70–0.94) and 0.86 (0.76–0.97) for DPP4i. Similar trends in HRs were observed for 4-point MACE. The estimates also suggested superior cardiovascular protection of GLP-1RA and SGLT2i over DPP4i for both sexes; however, statistical significance was not consistently achieved across the outcomes.

**Figure 2.**
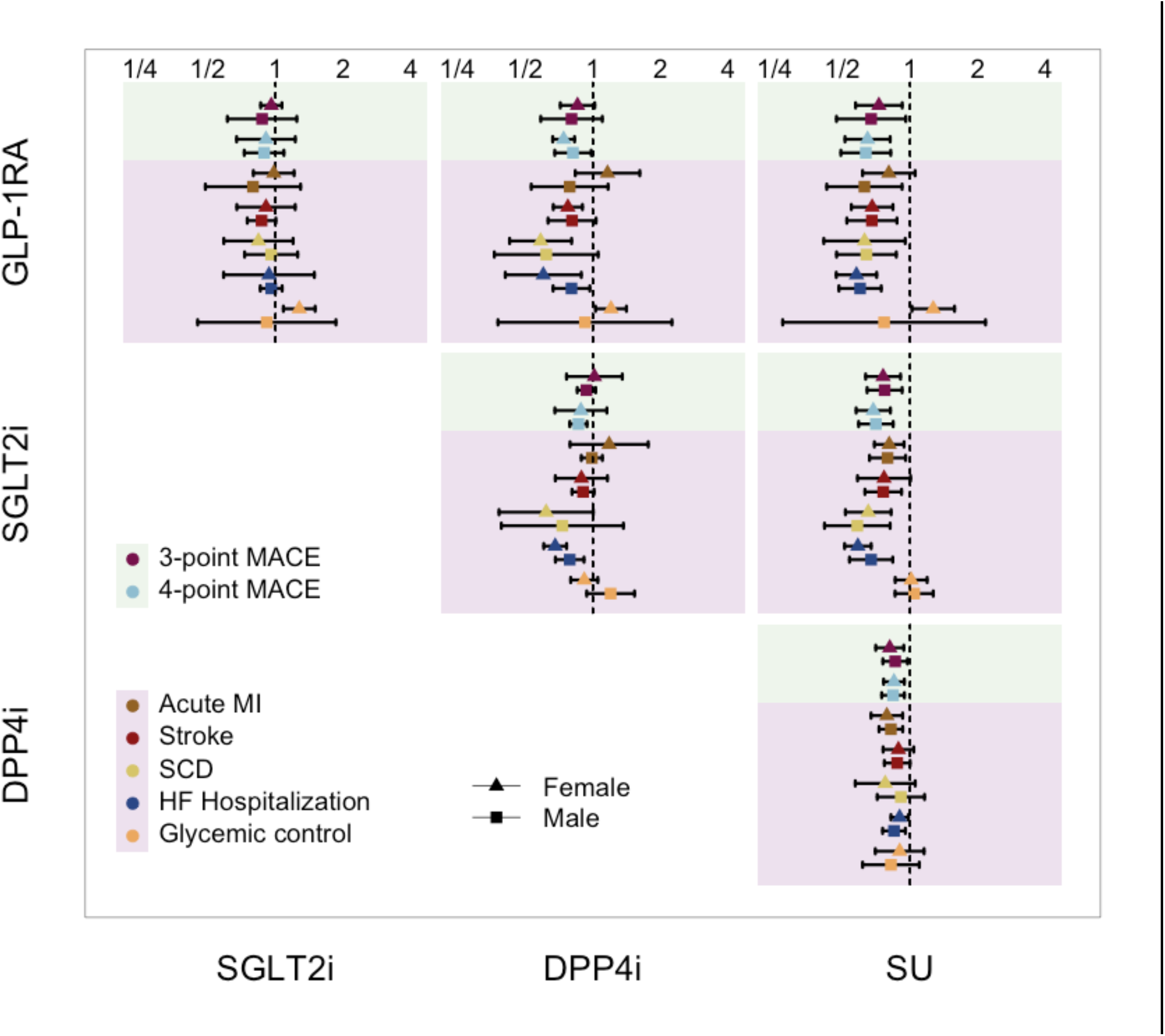
Meta-analyzed Hazard Ratio Estimates for the Cardiovascular Effectiveness Outcomes. Hazard ratios (HRs) compare the agents listed on the vertical axis versus the agents listed on the horizontal axis. For example, the top-left panel shows the HRs of GLP-1RA relative to SGLT2i. The precise values of HRs and the p-values for the test of sex differences are presented in Supplement A, Table 3.

For glycemic control, the female users of GLP-1RA were more likely to achieve HbA1c ≤ 7.0 with HR of 1.28 (1.08–1.50), 1.21 (1.03–1.41), and 1.27 (1.02–1.58) when compared with the users of SGLT2i, DPP4i, and SU; the same comparisons showed no significant differences among men with HR of 0.91 (0.45–1.86), 0.92 (0.38–2.25) and 0.77 (0.27–2.17).

### Cardiovascular side effects

For the safety outcomes, we focus our in-text discussions on the outcomes for which we found either significant or suggestive evidence of sex differences. We refer readers to Figure 3 for the HR estimates and CIs for all other outcomes and comparisons.

**Figure 3.**
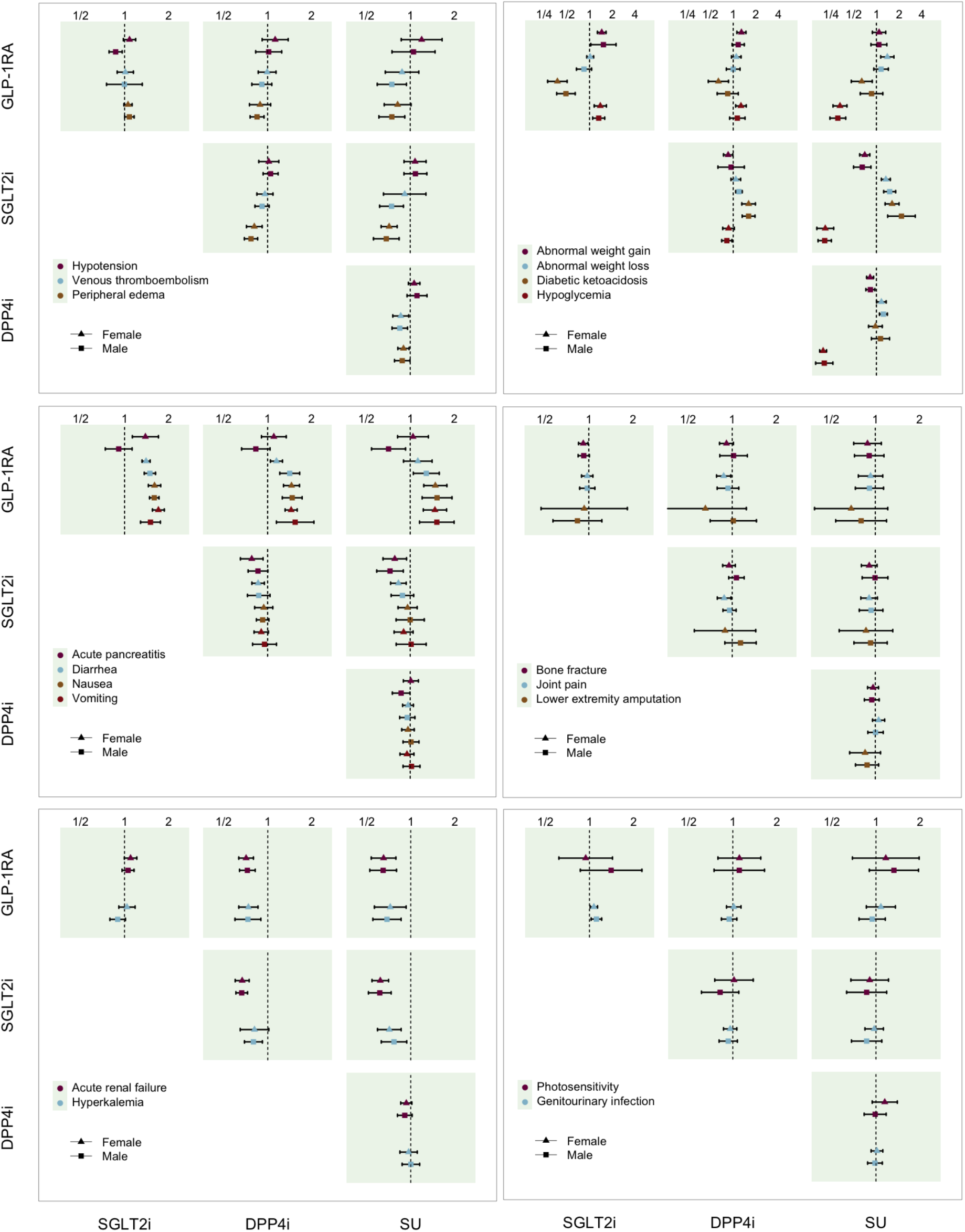
Meta-analyzed Hazard Ratio Estimates for the Safety Outcomes. Hazard ratios (HRs) compare the agents listed on the vertical axis versus the agents listed on the horizontal axis. For example, the top-left panel shows the HRs of GLP-1RA relative to SGLT2i. The precise values of HRs and the p-values for the test of sex differences are presented in Supplement A, Table 3.

Significant sex difference was found in the relative risk of hypotension, where men using GLP-1RA compared to SGLT2i had a lower risk with HR of 0.87 (0.78–0.96) while women with HR of 1.08 (0.98–1.19); the test of sex difference yielded p = 0.0025.

No significant sex differences were found among venous thromboembolism and peripheral edema.

### Metabolic/endocrine side effects

For diabetic ketoacidosis, GLP-1RA appeared to be associated with lower risks, particularly among women—the HR was 0.37 (0.28–0.50) among women vs 0.49 (0.37–0.64) among men when compared to SGLT2i; 0.64 (0.47–0.87) vs 0.85 (0.62–1.17) when compared to DPP4i; 0.64 (0.46–0.88) vs 0.86 (0.61–1.21) when compared to SU. These sex differences were not significant, however.

No significant sex differences were found for hypoglycemia, abnormal weight gain or abnormal weight loss. We note however that, for hypoglycemia, GLP-1RA compared to SGLT2i was associated with a higher risk both in men and women. We also found in agreement with existing evidence that, compared with the other agents, SGLT2i was associated with higher risk of diabetic ketoacidosis and SU with higher risk of hypoglycemia.

### Gastrointestinal (GI) side effects

For acute pancreatitis, GLP-1RA was associated with higher risks among women but neutral to lower risks among men—when compared against SGLT2i, the HR was 1.39 (1.13–1.70) among women vs 0.91 (0.74–1.12) among men; against DPP4i, 1.10 (0.91–1.34) vs 0.83 (0.67–1.04); and against SU, 1.04 (0.82–1.33) vs 0.71 (0.54–0.93). The tests of sex differences yielded significance for the comparisons against SGLT2i and SU with p = 0.005 and p = 0.036 respectively. For diarrhea, GLP-1RA was associated with higher risks, particularly among men—the HR was 1.40 (1.31–1.49) among women vs 1.49 (1.36–1.63) among men when compared against SGLT2i; 1.15 (1.05–1.27) vs 1.42 (1.12–1.66) against DPP4i; 1.12 (0.90–1.41) vs 1.29 (1.05–1.58) against SU; the sex difference was significant for the comparison against DPP4i with p = 0.025.

No significant sex differences were found for the other outcomes. On the other hand, in agreement with existing evidence for nausea and vomiting, GLP-1RA were associated with higher risks compared to the other agents for both sexes.

### Side effects in renal, musculoskeletal and other organ systems

No significant sex differences were observed for the outcomes under these categories. We note however that, for acute renal failure and hyperkalemia, GLP-1RA and SGLT2i were generally associated with lower risks compared to DPP4i and SU for both sexes.

### Sensitivity analyses

Findings from the alternative on-treatment design were largely consistent with the primary analysis. When excluding USOC, the reduced sample sizes generally lowered statistical power and yielded wider confidence intervals; nonetheless, most HR estimates retained the same directions and the key findings remained unchanged. The full sensitivity analyses results are presented in Supplement B and C, Table 1-156.

## Discussion

This multinational study of over 5 million patients provides comprehensive evidence on sex differences in second-line T2D treatments. We found no sex differences in cardiovascular effectiveness, confirming that benefits of GLP-1RA and SGLT2i observed in male-dominated trials extend to women. Our study also rigorously confirmed findings from prior studies: an elevated risk of hypoglycemia with SU, of diabetic ketoacidosis with SGLT2i, and of GI adverse effects with GLP-1RA, especially among women^23–30^. Further, our study identified important sex differences in acute pancreatitis and gastrointestinal symptoms.

Importantly, some findings would have been obscured without sex-stratified analyses. For example, if aggregating the female and male patients, the resulting HR for glycemic control would have been 1.07 (0.72–1.60) in the GLP-1RA vs DPP4i comparison. Similar phenomena were observed in the relative risks of acute pancreatitis and hypotension, where the aggregate analyses would have yielded HRs of 1.14 (0.99–1.31) and 0.97 (0.91–1.04) in the GLP-1RA vs SGLT2i comparison.

We found that women were more likely to achieve HbA1c ≤7.0 with GLP-1RA compared to the other agents, when no such benefit was observed in men. While the wide confidence interval for men warrants us to interpret this finding with caution, the overall pattern aligns with emerging evidence suggesting that GLP-1RA are more effective for glucose control in women than men^31,32^. Notably, a recent analysis of the real-world data from UK Clinical Practice Research Datalink (CPRD) and the HARMONY-7 trial results both identified sex as a major treatment effect modifier, with women experiencing substantially larger HbA1c reductions than men^32^. The evidence among the broader literature remains mixed, however^31^. Our study contributes to this ongoing debate and underscores the need for future work with balanced sex representation.

GLP-1RA, which act by increasing insulin secretion while suppressing glucagon release only when glucose levels are elevated, are generally considered to carry a low risk of hypoglycemia^14,24^. Nevertheless, some studies have reported increased risks of hypoglycemia with certain GLP-1RA compounds, particularly when used concomitantly with other glucose-lowering therapies^31,33^. Our study reinforces this concern, having found that GLP-1RA compared with SGLT2i was associated with a higher risk of hypoglycemia in both sexes; this finding contrasts with another observational study that found a non-differential risk^23^.

Our study contributes much-needed evidence on the controversial association between GLP-1RA use and acute pancreatitis^28,34–36^. Our study found GLP-1RA compared against SGLT2i to be associated with a higher risk of acute pancreatitis in women but not in men. While such sex difference in pancreatitis has been reported by a prior study, its use of a cross-sectional and single-database design makes its finding less reliable than ours^26^.

Findings from RCTs have also been mixed regarding this association^27,37–40^; our finding suggests that this may be due to the lack of statistical power in these RCTs and of limited female representations.

SGLT2i compared to GLP-1RA was associated with a higher risk of hypotension among men but not among women in our study. Aside from the sex difference, this finding is consistent with existing evidence that the stronger blood pressure-lowering effect of SGLT2i is largely attributable to osmotic diuresis and natriuresis leading to plasma volume contraction^41–45^. Our finding of the sex difference, however, raises the possibility that this prior evidence has been driven by the male-dominant compositions of RCTs. For example, in a systematic review of 22 RCTs reporting increased odds ratio of 1.24 (1.08–1.43) of orthostatic hypotension with high-dose SGLT2i versus placebo, 65.5% of the participants were men^42^. The meta-analysis showing stronger blood pressure-lowering effects of SGLT2i over GLP-1RA was also based on male-dominated RCTs, with majorities enrolling 60-70% men^41^. Our sex-stratified analysis of real-world data suggests the effect might differ for women.

Some hypotheses have been proposed to explain why GLP-1RA may affect men and women differently^11,31,46^. Estrogens and progesterone modulate GLP-1 receptor signaling and downstream endocrine and metabolic responsiveness, potentially amplifying insulinotropic and glucose-lowering effects in women^11,47–49^. In addition, women tend to exhibit higher drug levels at equivalent doses due to lower volume of distribution and slower clearance. This has been linked to greater gastrointestinal intolerance, such as nausea and vomiting, in exposure–response analyses of GLP-1RA^11,31,50,51^. Sex differences in gastrointestinal physiology, including slower baseline gastric emptying in women, may also contribute to the higher rates of GLP-1RA associated adverse events observed in our study^11^.

### Study Strengths and Limitations

To our knowledge, this is the largest, most comprehensive, and most rigorous observational study to date evaluating sex differences in comparative effectiveness and safety of the second-line T2DM treatments. We deployed the standardized analytic pipelines to the OHDSI databases covering diverse populations, thereby ensuring the reproducibility and generalizability of our findings. We systematically investigated a wide range of clinically relevant outcomes, providing a comprehensive characterization of benefit-risk profiles across treatment classes.

Alongside the notable strengths highlighted above, our study also has limitations. First, as with all observational studies, we cannot fully exclude the possibility of residual confounding. But the large-scale approach to propensity score adjustment, which incorporates the broad range of clinical information, has been shown as less prone to residual confounding. And the negative control calibration is likely to have detected any remaining bias.

Second, we applied the alpha level of 0.05 without formal adjustment for multiplicity. We did so because our study was designed not so much to confirm specific hypotheses, but rather to provide a comprehensive and systematic comparison of the effectiveness and safety profiles; our approach in particular avoids the issue of potential p-hacking and publication bias from selective reporting^52^. Accordingly, we have emphasized the general trends across comparisons and caution against over-interpretation of individual statistical significances.

Third, we defined the exposures at the drug-class level, ignoring potential heterogeneity within the classes. We focused on the class-level comparisons since 1) ingredient-level comparisons substantially diminishes statistical power due to reduced sample sizes within treatment groups and 2) prior ingredient-level comparisons indicated limited within-class heterogeneity, at least when pooling both sexes^53^.

Fourth, as with all studies based on health databases, we cannot fully ascertain exposures and outcomes from available information in data. The drug exposures inferred from prescription and dispensation records can deviate from patients’ actual usages. And variation in coding and healthcare practices could have affected the degree of outcome ascertainments across the databases. On the other hand, our multi-database approach allowed us to confirm the results’ consistency across databases and enhance confidence in the overall findings.

## Data Availability

The patient-level raw data used in this study cannot be publicly shared due to strict data privacy regulations, ethical considerations, and commercial Data Use Agreements. The underlying databases (e.g., Merative MarketScan, Optum, IQVIA, VA, SIDIAP) are subject to licensing restrictions and institutional governance. Researchers interested in accessing the raw patient-level data must apply directly to the respective data custodians for access or licensing.
However, the comprehensive aggregated summary statistics, baseline characteristics, propensity score diagnostics, and full comparative effectiveness analysis results are publicly available through interactive online repositories. Cohort characteristics analysis results can be accessed at:
https://data.ohdsi.org/LegendT2dmClassCohortExplorer/
and comparative effectiveness analysis results at:
https://data.ohdsi.org/LegendT2dmClassEvidenceExplorer/.

https://data.ohdsi.org/LegendT2dmClassCohortExplorer/

https://data.ohdsi.org/LegendT2dmClassEvidenceExplorer/

