## Supplement A for "Sex Differences in Comparative Effectiveness and Safety of Second-line Antidiabetic Agents: Real-world Evidence from Large-scale Multinational Study"

for

#### Contents


**1 Data sources** **2**

**2 Pharmacological agents included in the drug classes** **3**

**3 Meta-analyzed comparative effectiveness estimates** **4**

**4 Exposure cohort definitions and OMOP concepts** **7**

4.1 Example (GLP1RA) new-user cohort . . . . . . . . . . . . . . . . . . . . . . . . . . . . . . . 7

4.2 Negative control concepts . . . . . . . . . . . . . . . . . . . . . . . . . . . . . . . . . . . . 10

### 1 Data sources

**Supplementary Table 1:** Data sources and the populations they cover.

| Data source | Population | Patients | History | Data capture process and short description |
|---|---|---|---|---|
| | | **Administrative claims** | | |
| IBM MarketScan Commercial Claims and Encounters (CCAE) | Commercially insured, < 65 years | 142M | 2000 – | Adjudicated health insurance claims (e.g. inpatient, outpatient, and outpatient pharmacy) from large employers and health plans who provide private healthcare coverage to employees, their spouses and dependents. |
| IBM MarketScan Medicare Supplemental Database (MDCR) | Commercially insured, 65$+$ years | 10M | 2000 – | Adjudicated health insurance claims of retirees with primary or Medicare supplemental coverage through privately insured fee-for-service, point-of-service or capitated health plans. |
| IBM MarketScan Multi-State Medicaid Database (MDCD) | Medicaid enrollees, racially diverse | 26M | 2006 – | Adjudicated health insurance claims for Medicaid enrollees from multiple states and includes hospital discharge diagnoses, outpatient diagnoses and procedures, and outpatient pharmacy claims. |
| IQVIA Open Claims (Open Claims) | General | 160M | 2010 – | Pre-adjudicated claims at the anonymized patient-level collected from office-based physicians and specialists via office management software and clearinghouse switch sources for the purpose of reimbursement. |
| Optum Clinformatics Data Mart (OptumDOD) | Commercially or Medicare insured | 85M | 2000 – | Inpatient and outpatient healthcare insurance claims. |
| | | **Electronic health records (EHRs)** | | |
| IQVIA Disease Analyzer Germany (DAG) | Germany, general | 37M | 1992 – | Collection from patient management software used by general practitioners and selected specialists to document patients' medical records within their office-based practice during a visit. |
| Optum Electronic Health Records (OptumEHR) | US, general | 93M | 2006 – | Clinical information, prescriptions, lab results, vital signs, body measurements, diagnoses and procedures derived from clinical notes using natural language processing. |
| Information System for Research in Primary Care (SIDIAP) | 80% of all Catalonia (Spain) | 7.7M | 2006 – | Primary care partially linked to inpatient data with pharmacy dispensations and primary care laboratories. Healthcare is universal and taxpayer funded in the region, and PCPs are gatekeeps for all care and responsible for repeat prescriptions. |
| UK IQVIA Medical Research Data (UKIMRD) | United Kingdom, general | 15M | 1996 – | Primary care records from the UK general practices covering a 6% representative sample and includes demographics, diagnoses, prescriptions, consultations, referrals and laboratory investigations. |
| US Department of Veterans Affairs (VA) | Veterans, older, racially diverse | 12M | 2000 – | National VA health care system, the largest integrated provider of medical services in the US, provided at 170 VA medical centers and 1,063 outpatient sites. |

#### 2 Pharmacological agents included in the drug classes

**Supplementary Table 2:** Drug classes and example agents.

| GLP1-RA | SGLT2i | DPP4i | SU |
|---|---|---|---|
| Albiglutide | Canagliflozin | Alogliptin | Chlorpropamide |
| Dulaglutide | Dapagliflozin | Linagliptin | Glimepiride |
| Exenatide | Empagliflozin | Saxagliptin | Glipizide |
| Liraglutide | Ertugliflozin | Sitagliptin | Gliquidone |
| Lixisenatide | | Vildagliptin | Glyburide |
| Semaglutide | | | Tolazamide |
| | | | Tolbutamide |

### 3 Meta-analyzed comparative effectiveness estimates

**Supplementary Table 3:** Meta-analyzed comparative effectiveness estimates from PS-stratification, on-treatment time-at-risk designs across outcomes and drug class comparisons. We report calibrated hazard ratios (HRs), 95% confidence intervals (CIs), and p-values for sex differences. P-values correspond to Z-tests for differences between log-HRs in female and male cohorts.

| Target | Comparator | Female HR (95% CI) | Male HR (95% CI) | Overall HR (95% CI) | P value for sex difference |
|---|---|---|---|---|---|
| **3-point MACE** | | | | | |
| DPP4i | SU | 0.81 (0.70–0.94) | 0.86 (0.76–0.97) | 0.85 (0.78–0.94) | 0.568 |
| GLP-1RA | DPP4i | 0.86 (0.72–1.02) | 0.80 (0.59–1.10) | 0.87 (0.68–1.10) | 0.739 |
| GLP-1RA | SGLT2i | 0.96 (0.86–1.07) | 0.87 (0.61–1.24) | 0.88 (0.81–0.95) | 0.620 |
| GLP-1RA | SU | 0.73 (0.57–0.92) | 0.67 (0.47–0.96) | 0.69 (0.55–0.86) | 0.716 |
| SGLT2i | DPP4i | 1.02 (0.76–1.35) | 0.94 (0.85–1.03) | 0.91 (0.80–1.05) | 0.595 |
| SGLT2i | SU | 0.76 (0.63–0.91) | 0.77 (0.64–0.92) | 0.75 (0.63–0.90) | 0.904 |
| **4-point MACE** | | | | | |
| DPP4i | SU | 0.85 (0.76–0.94) | 0.84 (0.75–0.94) | 0.85 (0.78–0.94) | 0.905 |
| GLP-1RA | DPP4i | 0.74 (0.66–0.83) | 0.82 (0.68–0.98) | 0.78 (0.68–0.89) | 0.390 |
| GLP-1RA | SGLT2i | 0.91 (0.67–1.23) | 0.89 (0.73–1.09) | 0.91 (0.86–0.98) | 0.918 |
| GLP-1RA | SU | 0.65 (0.51–0.82) | 0.64 (0.49–0.82) | 0.62 (0.51–0.76) | 0.918 |
| SGLT2i | DPP4i | 0.89 (0.68–1.16) | 0.86 (0.79–0.94) | 0.87 (0.75–1.02) | 0.861 |
| SGLT2i | SU | 0.69 (0.58–0.82) | 0.70 (0.59–0.84) | 0.71 (0.59–0.86) | 0.832 |
| **Acute myocardial infarction** | | | | | |
| DPP4i | SU | 0.79 (0.67–0.93) | 0.82 (0.73–0.93) | 0.83 (0.75–0.92) | 0.689 |
| GLP-1RA | DPP4i | 1.16 (0.84–1.62) | 0.79 (0.53–1.17) | 0.98 (0.75–1.28) | 0.139 |
| GLP-1RA | SGLT2i | 0.98 (0.80–1.21) | 0.79 (0.49–1.29) | 0.89 (0.72–1.11) | 0.432 |
| GLP-1RA | SU | 0.80 (0.61–1.05) | 0.63 (0.43–0.92) | 0.68 (0.55–0.84) | 0.297 |
| SGLT2i | DPP4i | 1.18 (0.79–1.76) | 0.99 (0.89–1.10) | 1.00 (0.87–1.16) | 0.403 |
| SGLT2i | SU | 0.81 (0.69–0.94) | 0.79 (0.66–0.96) | 0.81 (0.67–0.96) | 0.890 |
| **Stroke** | | | | | |
| DPP4i | SU | 0.89 (0.76–1.04) | 0.88 (0.77–1.00) | 0.90 (0.81–1.00) | 0.908 |
| GLP-1RA | DPP4i | 0.77 (0.67–0.90) | 0.81 (0.63–1.03) | 0.78 (0.67–0.91) | 0.770 |
| GLP-1RA | SGLT2i | 0.91 (0.67–1.22) | 0.87 (0.75–1.00) | 0.93 (0.84–1.02) | 0.789 |
| GLP-1RA | SU | 0.68 (0.55–0.84) | 0.68 (0.52–0.87) | 0.69 (0.55–0.86) | 0.988 |
| SGLT2i | DPP4i | 0.89 (0.68–1.16) | 0.91 (0.81–1.01) | 0.85 (0.73–0.98) | 0.909 |
| SGLT2i | SU | 0.77 (0.58–1.01) | 0.76 (0.63–0.92) | 0.75 (0.60–0.94) | 0.960 |
| **Sudden cardiac death** | | | | | |
| DPP4i | SU | 0.78 (0.57–1.05) | 0.91 (0.71–1.16) | 0.83 (0.66–1.04) | 0.422 |
| GLP-1RA | DPP4i | 0.59 (0.43–0.81) | 0.62 (0.36–1.06) | 0.68 (0.54–0.86) | 0.855 |
| GLP-1RA | SGLT2i | 0.84 (0.59–1.20) | 0.96 (0.73–1.26) | 0.88 (0.71–1.08) | 0.569 |
| GLP-1RA | SU | 0.63 (0.41–0.95) | 0.64 (0.47–0.87) | 0.67 (0.48–0.92) | 0.940 |
| SGLT2i | DPP4i | 0.62 (0.38–1.00) | 0.73 (0.39–1.37) | 0.84 (0.51–1.38) | 0.680 |
| SGLT2i | SU | 0.65 (0.52–0.82) | 0.58 (0.42–0.81) | 0.65 (0.53–0.80) | 0.591 |
| **Hospitalization with heart failure** | | | | | |
| DPP4i | SU | 0.90 (0.82–0.98) | 0.85 (0.75–0.95) | 0.87 (0.79–0.96) | 0.442 |
| GLP-1RA | DPP4i | 0.60 (0.41–0.89) | 0.80 (0.67–0.97) | 0.75 (0.65–0.86) | 0.191 |
| GLP-1RA | SGLT2i | 0.94 (0.59–1.49) | 0.96 (0.86–1.07) | 1.00 (0.89–1.14) | 0.925 |
| GLP-1RA | SU | 0.58 (0.47–0.71) | 0.60 (0.48–0.74) | 0.58 (0.48–0.71) | 0.808 |
| SGLT2i | DPP4i | 0.68 (0.61–0.76) | 0.79 (0.68–0.91) | 0.77 (0.66–0.91) | 0.120 |
| SGLT2i | SU | 0.59 (0.51–0.67) | 0.67 (0.54–0.84) | 0.66 (0.53–0.81) | 0.307 |
| **Glycemic control** | | | | | |
| DPP4i | SU | 0.90 (0.70–1.15) | 0.82 (0.61–1.10) | 0.83 (0.67–1.02) | 0.646 |
| GLP-1RA | DPP4i | 1.21 (1.03–1.41) | 0.92 (0.38–2.25) | 1.07 (0.72–1.60) | 0.564 |
| GLP-1RA | SGLT2i | 1.28 (1.08–1.50) | 0.91 (0.45–1.86) | 1.18 (1.06–1.32) | 0.370 |
| GLP-1RA | SU | 1.27 (1.02–1.58) | 0.77 (0.27–2.17) | 1.24 (1.01–1.53) | 0.353 |
| SGLT2i | DPP4i | 0.92 (0.80–1.05) | 1.20 (0.94–1.53) | 1.08 (0.89–1.29) | 0.060 |
| SGLT2i | SU | 1.01 (0.86–1.19) | 1.04 (0.86–1.27) | 1.03 (0.86–1.24) | 0.815 |
| **Abnormal weight gain** | | | | | |
| DPP4i | SU | 0.82 (0.74–0.91) | 0.83 (0.72–0.95) | 0.82 (0.74–0.91) | 0.926 |
| GLP-1RA | DPP4i | 1.29 (1.12–1.48) | 1.17 (0.97–1.41) | 1.25 (1.07–1.46) | 0.404 |
| GLP-1RA | SGLT2i | 1.45 (1.27–1.65) | 1.52 (1.04–2.24) | 1.51 (1.36–1.68) | 0.804 |
| GLP-1RA | SU | 1.08 (0.88–1.33) | 1.07 (0.84–1.37) | 1.15 (0.85–1.57) | 0.972 |

| | | | | | |
|---|---|---|---|---|---|
| SGLT2i | DPP4i | 0.86 (0.75–0.99) | 0.94 (0.63–1.41) | 0.83 (0.71–0.97) | 0.679 |
| SGLT2i | SU | 0.70 (0.60–0.81) | 0.65 (0.50–0.84) | 0.66 (0.55–0.80) | 0.623 |
| **Abnormal weight loss** | | | | | |
| DPP4i | SU | 1.16 (1.00–1.34) | 1.23 (1.09–1.39) | 1.19 (1.07–1.34) | 0.523 |
| GLP-1RA | DPP4i | 1.10 (0.95–1.28) | 1.00 (0.81–1.24) | 1.08 (0.90–1.29) | 0.471 |
| GLP-1RA | SGLT2i | 1.02 (0.92–1.13) | 0.84 (0.67–1.06) | 0.94 (0.83–1.07) | 0.145 |
| GLP-1RA | SU | 1.39 (1.14–1.70) | 1.15 (0.92–1.43) | 1.25 (1.03–1.53) | 0.212 |
| SGLT2i | DPP4i | 1.09 (0.94–1.26) | 1.20 (1.08–1.32) | 1.19 (1.02–1.38) | 0.294 |
| SGLT2i | SU | 1.33 (1.16–1.52) | 1.49 (1.24–1.79) | 1.42 (1.19–1.70) | 0.318 |
| **Diabetic ketoacidosis** | | | | | |
| DPP4i | SU | 0.97 (0.78–1.19) | 1.13 (0.85–1.49) | 1.02 (0.89–1.16) | 0.383 |
| GLP-1RA | DPP4i | 0.64 (0.47–0.87) | 0.85 (0.62–1.17) | 0.71 (0.54–0.93) | 0.210 |
| GLP-1RA | SGLT2i | 0.37 (0.28–0.50) | 0.49 (0.37–0.64) | 0.42 (0.32–0.54) | 0.212 |
| GLP-1RA | SU | 0.64 (0.46–0.88) | 0.86 (0.61–1.21) | 0.76 (0.58–1.00) | 0.211 |
| SGLT2i | DPP4i | 1.61 (1.31–1.98) | 1.60 (1.32–1.95) | 1.61 (1.34–1.93) | 0.994 |
| SGLT2i | SU | 1.61 (1.30–1.99) | 2.14 (1.41–3.26) | 1.85 (1.37–2.50) | 0.233 |
| **Hypoglycemia** | | | | | |
| DPP4i | SU | 0.20 (0.18–0.22) | 0.20 (0.16–0.26) | 0.21 (0.16–0.29) | 0.805 |
| GLP-1RA | DPP4i | 1.27 (1.08–1.49) | 1.14 (0.90–1.44) | 1.23 (1.05–1.45) | 0.446 |
| GLP-1RA | SGLT2i | 1.38 (1.15–1.67) | 1.32 (1.10–1.58) | 1.34 (1.11–1.62) | 0.713 |
| GLP-1RA | SU | 0.33 (0.27–0.41) | 0.31 (0.24–0.39) | 0.32 (0.26–0.39) | 0.688 |
| SGLT2i | DPP4i | 0.87 (0.73–1.04) | 0.83 (0.71–0.97) | 0.87 (0.72–1.05) | 0.659 |
| SGLT2i | SU | 0.21 (0.16–0.27) | 0.21 (0.17–0.25) | 0.21 (0.17–0.27) | 0.919 |
| **Acute pancreatitis** | | | | | |
| DPP4i | SU | 1.01 (0.90–1.13) | 0.86 (0.75–0.99) | 0.92 (0.83–1.03) | 0.099 |
| GLP-1RA | DPP4i | 1.10 (0.91–1.34) | 0.83 (0.67–1.04) | 0.96 (0.80–1.15) | 0.061 |
| GLP-1RA | SGLT2i | 1.39 (1.13–1.70) | 0.91 (0.74–1.12) | 1.14 (0.99–1.31) | 0.005 |
| GLP-1RA | SU | 1.04 (0.82–1.33) | 0.71 (0.54–0.93) | 0.92 (0.70–1.23) | 0.036 |
| SGLT2i | DPP4i | 0.78 (0.65–0.93) | 0.86 (0.74–1.01) | 0.92 (0.73–1.16) | 0.416 |
| SGLT2i | SU | 0.78 (0.65–0.94) | 0.73 (0.59–0.90) | 0.79 (0.63–0.98) | 0.600 |
| **Diarrhea** | | | | | |
| DPP4i | SU | 0.96 (0.89–1.05) | 0.95 (0.85–1.07) | 0.96 (0.87–1.05) | 0.876 |
| GLP-1RA | DPP4i | 1.15 (1.05–1.27) | 1.42 (1.21–1.66) | 1.25 (1.08–1.45) | 0.025 |
| GLP-1RA | SGLT2i | 1.40 (1.31–1.49) | 1.49 (1.36–1.63) | 1.44 (1.37–1.51) | 0.280 |
| GLP-1RA | SU | 1.12 (0.90–1.41) | 1.29 (1.05–1.58) | 1.21 (0.95–1.53) | 0.389 |
| SGLT2i | DPP4i | 0.86 (0.78–0.95) | 0.87 (0.73–1.04) | 0.89 (0.77–1.01) | 0.925 |
| SGLT2i | SU | 0.83 (0.73–0.94) | 0.88 (0.74–1.05) | 0.86 (0.72–1.02) | 0.567 |
| **Nausea** | | | | | |
| DPP4i | SU | 0.96 (0.87–1.06) | 1.01 (0.89–1.15) | 1.00 (0.90–1.11) | 0.556 |
| GLP-1RA | DPP4i | 1.46 (1.29–1.65) | 1.48 (1.26–1.72) | 1.46 (1.25–1.71) | 0.915 |
| GLP-1RA | SGLT2i | 1.60 (1.45–1.76) | 1.59 (1.48–1.72) | 1.58 (1.45–1.72) | 0.949 |
| GLP-1RA | SU | 1.48 (1.24–1.78) | 1.52 (1.20–1.93) | 1.55 (1.26–1.91) | 0.858 |
| SGLT2i | DPP4i | 0.94 (0.82–1.09) | 0.93 (0.84–1.02) | 0.95 (0.80–1.11) | 0.847 |
| SGLT2i | SU | 0.96 (0.82–1.11) | 1.00 (0.80–1.24) | 0.98 (0.80–1.20) | 0.766 |
| **Vomiting** | | | | | |
| DPP4i | SU | 0.95 (0.85–1.06) | 1.02 (0.89–1.16) | 0.99 (0.89–1.10) | 0.415 |
| GLP-1RA | DPP4i | 1.45 (1.32–1.60) | 1.55 (1.15–2.08) | 1.49 (1.27–1.74) | 0.689 |
| GLP-1RA | SGLT2i | 1.70 (1.55–1.87) | 1.50 (1.28–1.76) | 1.63 (1.55–1.72) | 0.179 |
| GLP-1RA | SU | 1.47 (1.23–1.77) | 1.52 (1.16–1.99) | 1.47 (1.21–1.78) | 0.859 |
| SGLT2i | DPP4i | 0.91 (0.81–1.01) | 0.95 (0.79–1.15) | 0.95 (0.80–1.12) | 0.643 |
| SGLT2i | SU | 0.90 (0.77–1.04) | 1.01 (0.80–1.28) | 0.96 (0.79–1.17) | 0.403 |
| **Acute renal failure** | | | | | |
| DPP4i | SU | 0.93 (0.85–1.02) | 0.91 (0.81–1.02) | 0.92 (0.83–1.01) | 0.745 |
| GLP-1RA | DPP4i | 0.71 (0.63–0.80) | 0.73 (0.64–0.82) | 0.73 (0.64–0.83) | 0.815 |
| GLP-1RA | SGLT2i | 1.10 (1.00–1.21) | 1.06 (0.96–1.16) | 1.12 (1.00–1.25) | 0.565 |
| GLP-1RA | SU | 0.65 (0.54–0.79) | 0.65 (0.52–0.80) | 0.68 (0.54–0.86) | 0.964 |
| SGLT2i | DPP4i | 0.67 (0.60–0.75) | 0.66 (0.61–0.73) | 0.68 (0.59–0.78) | 0.917 |
| SGLT2i | SU | 0.62 (0.54–0.71) | 0.61 (0.51–0.73) | 0.62 (0.52–0.74) | 0.944 |
| **Hyperkalemia** | | | | | |
| DPP4i | SU | 0.97 (0.84–1.11) | 1.00 (0.87–1.15) | 0.98 (0.87–1.10) | 0.716 |
| GLP-1RA | DPP4i | 0.74 (0.63–0.86) | 0.73 (0.60–0.90) | 0.78 (0.64–0.95) | 0.955 |
| GLP-1RA | SGLT2i | 1.04 (0.91–1.18) | 0.90 (0.79–1.01) | 0.96 (0.88–1.05) | 0.107 |
| GLP-1RA | SU | 0.72 (0.56–0.93) | 0.69 (0.55–0.86) | 0.74 (0.57–0.94) | 0.763 |
| SGLT2i | DPP4i | 0.81 (0.65–1.02) | 0.80 (0.69–0.92) | 0.82 (0.69–0.98) | 0.902 |
| SGLT2i | SU | 0.71 (0.59–0.86) | 0.77 (0.63–0.94) | 0.76 (0.62–0.93) | 0.600 |
| **Hypotension** | | | | | |

| | | | | | |
|---|---|---|---|---|---|
| DPP4i | SU | 1.06 (0.97–1.16) | 1.11 (0.95–1.30) | 1.08 (0.97–1.20) | 0.623 |
| GLP-1RA | DPP4i | 1.13 (0.92–1.39) | 1.02 (0.83–1.25) | 1.10 (0.90–1.35) | 0.490 |
| GLP-1RA | SGLT2i | 1.08 (0.98–1.19) | 0.87 (0.78–0.96) | 0.97 (0.91–1.05) | 0.003 |
| GLP-1RA | SU | 1.20 (0.87–1.65) | 1.05 (0.75–1.48) | 1.14 (0.77–1.71) | 0.591 |
| SGLT2i | DPP4i | 1.02 (0.87–1.19) | 1.05 (0.93–1.18) | 1.07 (0.92–1.26) | 0.775 |
| SGLT2i | SU | 1.08 (0.91–1.28) | 1.08 (0.90–1.30) | 1.13 (0.93–1.37) | 0.966 |
| **Venous thromboembolism** | | | | | |
| DPP4i | SU | 0.86 (0.76–0.97) | 0.85 (0.75–0.96) | 0.86 (0.77–0.96) | 0.864 |
| GLP-1RA | DPP4i | 0.99 (0.86–1.14) | 0.91 (0.78–1.07) | 0.95 (0.82–1.11) | 0.436 |
| GLP-1RA | SGLT2i | 1.01 (0.89–1.15) | 1.00 (0.75–1.32) | 0.99 (0.90–1.09) | 0.928 |
| GLP-1RA | SU | 0.88 (0.67–1.14) | 0.75 (0.59–0.94) | 0.83 (0.63–1.09) | 0.368 |
| SGLT2i | DPP4i | 0.96 (0.85–1.09) | 0.92 (0.82–1.03) | 0.94 (0.82–1.09) | 0.601 |
| SGLT2i | SU | 0.92 (0.65–1.28) | 0.74 (0.62–0.90) | 0.82 (0.66–1.02) | 0.289 |
| **Peripheral edema** | | | | | |
| DPP4i | SU | 0.90 (0.82–0.98) | 0.88 (0.78–0.99) | 0.90 (0.81–0.99) | 0.798 |
| GLP-1RA | DPP4i | 0.89 (0.75–1.05) | 0.85 (0.76–0.95) | 0.90 (0.76–1.06) | 0.643 |
| GLP-1RA | SGLT2i | 1.05 (0.99–1.13) | 1.08 (1.00–1.16) | 1.07 (1.02–1.13) | 0.674 |
| GLP-1RA | SU | 0.82 (0.66–1.01) | 0.75 (0.61–0.92) | 0.80 (0.63–1.01) | 0.543 |
| SGLT2i | DPP4i | 0.81 (0.72–0.92) | 0.77 (0.69–0.86) | 0.79 (0.67–0.92) | 0.529 |
| SGLT2i | SU | 0.72 (0.63–0.81) | 0.68 (0.56–0.84) | 0.72 (0.60–0.87) | 0.710 |
| **Bone fracture** | | | | | |
| DPP4i | SU | 0.96 (0.88–1.05) | 0.94 (0.84–1.06) | 0.97 (0.88–1.06) | 0.785 |
| GLP-1RA | DPP4i | 0.91 (0.82–1.02) | 1.02 (0.82–1.27) | 0.97 (0.82–1.15) | 0.366 |
| GLP-1RA | SGLT2i | 0.92 (0.85–0.99) | 0.92 (0.84–1.01) | 0.92 (0.87–0.97) | 0.897 |
| GLP-1RA | SU | 0.88 (0.71–1.10) | 0.91 (0.72–1.14) | 0.88 (0.69–1.13) | 0.868 |
| SGLT2i | DPP4i | 0.95 (0.86–1.05) | 1.07 (0.95–1.21) | 1.03 (0.88–1.19) | 0.145 |
| SGLT2i | SU | 0.91 (0.80–1.03) | 0.99 (0.81–1.22) | 0.97 (0.80–1.16) | 0.450 |
| **Joint pain** | | | | | |
| DPP4i | SU | 1.05 (0.96–1.16) | 1.00 (0.89–1.13) | 1.04 (0.94–1.15) | 0.537 |
| GLP-1RA | DPP4i | 0.88 (0.78–0.98) | 0.94 (0.79–1.11) | 0.90 (0.78–1.03) | 0.514 |
| GLP-1RA | SGLT2i | 0.97 (0.89–1.06) | 0.98 (0.87–1.10) | 0.99 (0.92–1.06) | 0.989 |
| GLP-1RA | SU | 0.93 (0.77–1.12) | 0.91 (0.73–1.14) | 0.93 (0.76–1.13) | 0.910 |
| SGLT2i | DPP4i | 0.88 (0.79–0.98) | 0.96 (0.87–1.07) | 0.92 (0.79–1.08) | 0.265 |
| SGLT2i | SU | 0.91 (0.80–1.03) | 0.93 (0.78–1.13) | 0.94 (0.78–1.15) | 0.787 |
| **Lower extremity amputation** | | | | | |
| DPP4i | SU | 0.85 (0.67–1.08) | 0.88 (0.73–1.05) | 0.88 (0.76–1.03) | 0.841 |
| GLP-1RA | DPP4i | 0.66 (0.35–1.25) | 1.02 (0.71–1.46) | 0.99 (0.68–1.45) | 0.245 |
| GLP-1RA | SGLT2i | 0.93 (0.47–1.83) | 0.84 (0.57–1.23) | 0.85 (0.63–1.15) | 0.789 |
| GLP-1RA | SU | 0.68 (0.38–1.21) | 0.80 (0.54–1.19) | 0.75 (0.53–1.05) | 0.662 |
| SGLT2i | DPP4i | 0.89 (0.55–1.45) | 1.14 (0.89–1.46) | 1.16 (0.91–1.48) | 0.379 |
| SGLT2i | SU | 0.86 (0.57–1.31) | 0.93 (0.71–1.20) | 0.92 (0.72–1.17) | 0.775 |
| **Photosensitivity** | | | | | |
| DPP4i | SU | 1.15 (0.94–1.40) | 0.99 (0.83–1.17) | 1.03 (0.90–1.17) | 0.260 |
| GLP-1RA | DPP4i | 1.11 (0.79–1.55) | 1.11 (0.75–1.65) | 1.09 (0.83–1.44) | 0.996 |
| GLP-1RA | SGLT2i | 0.94 (0.62–1.43) | 1.40 (0.87–2.28) | 1.17 (0.87–1.57) | 0.222 |
| GLP-1RA | SU | 1.17 (0.69–1.97) | 1.33 (0.90–1.95) | 1.23 (0.90–1.68) | 0.700 |
| SGLT2i | DPP4i | 1.02 (0.75–1.38) | 0.82 (0.61–1.10) | 0.91 (0.72–1.16) | 0.310 |
| SGLT2i | SU | 0.91 (0.67–1.22) | 0.86 (0.63–1.18) | 0.88 (0.68–1.14) | 0.828 |
| **Genitourinary infection** | | | | | |
| DPP4i | SU | 1.02 (0.93–1.11) | 0.98 (0.88–1.10) | 1.01 (0.92–1.12) | 0.674 |
| GLP-1RA | DPP4i | 1.02 (0.91–1.14) | 0.94 (0.84–1.07) | 1.01 (0.87–1.18) | 0.390 |
| GLP-1RA | SGLT2i | 1.07 (1.01–1.13) | 1.11 (1.03–1.21) | 1.10 (1.04–1.15) | 0.443 |
| GLP-1RA | SU | 1.08 (0.86–1.36) | 0.94 (0.77–1.16) | 1.08 (0.80–1.45) | 0.383 |
| SGLT2i | DPP4i | 0.96 (0.86–1.07) | 0.93 (0.81–1.07) | 0.98 (0.84–1.15) | 0.736 |
| SGLT2i | SU | 0.97 (0.84–1.12) | 0.86 (0.68–1.09) | 0.93 (0.76–1.12) | 0.403 |

### 4 Exposure cohort definitions and OMOP concepts

#### 4.1 Example (GLP1RA) new-user cohort

##### 4.1.1 Cohort Entry Events

People with continuous observation of 365 days before event may enter the cohort when observing any of the following:

1. drug exposure of ‘GLP-1 receptor agonists’ for the first time in the person’s history.

Limit cohort entry events to the earliest event per person.

Restrict entry events to with all of the following criteria:

1. with the following event criteria: who are >= 18 years old.
2. having at least 1 condition occurrence of ‘Type 2 diabetes mellitus’, starting anytime on or before cohort entry start date; allow events outside observation period.
3. having no condition occurrences of ‘Type 1 diabetes mellitus’, starting anytime on or before cohort entry start date; allow events outside observation period.
4. having no condition occurrences of ‘Secondary diabetes mellitus’, starting anytime on or before cohort entry start date; allow events outside observation period.

##### 4.1.2 Additional Inclusion Criteria

**I. No prior DPP4 inhibitor exposure** Entry events having no drug exposures of ‘DPP4 inhibitors’, starting anytime on or before cohort entry start date; allow events outside observation period.

**II. No prior SGLT-2 inhibitor exposure** Entry events having no drug exposures of ‘SGLT2 inhibitors’, starting anytime on or before cohort entry start date; allow events outside observation period.

**III. No prior SU exposure** Entry events having no drug exposures of ‘Sulfonylureas’, starting anytime on or before cohort entry start date; allow events outside observation period.

**IV. No prior other anti-diabetic exposure** Entry events having no drug exposures of ‘Other anti-diabetics’, starting anytime on or before cohort entry start date; allow events outside observation period.

**V. Prior metformin use** Entry events with any of the following criteria:

1. having at least 1 drug era of ‘Metformin’, starting anytime up to 90 days before cohort entry start date; allow events outside observation period; with era length >= 90 days.
2. having at least 3 drug exposures of ‘Metformin’, starting anytime on or before cohort entry start date; allow events outside observation period.

**VI. No prior insulin use or combo initiation: Proxy for < 30 days drug era anytime before index and no combination use on index** Entry events with all of the following criteria:

1. having no drug eras of ‘Insulin’, starting anytime up to 30 days before cohort entry start date; allow events outside observation period; with era length > 30 days.
2. having no drug eras of ‘Insulin’, starting between 30 days before and 0 days after cohort entry start date; allow events outside observation period.

##### 4.1.3 Cohort Exit

The cohort end date will be based on a continuous exposure to 'GLP-1 receptor agonists': allowing 30 days between exposures, adding 0 days after exposure ends, and using days supply and exposure end date for exposure duration.

##### 4.1.4 Cohort Eras

Remaining events will be combined into cohort eras if they are within 0 days of each other.

##### 4.1.5 Concept: DPP4 inhibitors

| Concept ID | Concept Name | Code | Vocabulary | Excluded | Descendants | Mapped |
|---|---|---|---|---|---|---|
| 43013884 | alogliptin | 1368001 | RxNorm | NO | YES | NO |
| 40239216 | linagliptin | 1100699 | RxNorm | NO | YES | NO |
| 40166035 | saxagliptin | 857974 | RxNorm | NO | YES | NO |
| 1580747 | sitagliptin | 593411 | RxNorm | NO | YES | NO |
| 19122137 | vildagliptin | 596554 | RxNorm | NO | YES | NO |

##### 4.1.6 Concept: GLP-1 receptor agonists

| Concept ID | Concept Name | Code | Vocabulary | Excluded | Descendants | Mapped |
|---|---|---|---|---|---|---|
| 44816332 | albiglutide | 1534763 | RxNorm | NO | YES | NO |
| 45774435 | dulaglutide | 1551291 | RxNorm | NO | YES | NO |
| 1583722 | exenatide | 60548 | RxNorm | NO | YES | NO |
| 40170911 | liraglutide | 475968 | RxNorm | NO | YES | NO |
| 44506754 | lixisenatide | 1440051 | RxNorm | NO | YES | NO |
| 793143 | semaglutide | 1991302 | RxNorm | NO | YES | NO |

##### 4.1.7 Concept: SGLT2 inhibitors

| Concept ID | Concept Name | Code | Vocabulary | Excluded | Descendants | Mapped |
|---|---|---|---|---|---|---|
| 43526465 | canagliflozin | 1373458 | RxNorm | NO | YES | NO |
| 44785829 | dapagliflozin | 1488564 | RxNorm | NO | YES | NO |
| 45774751 | empagliflozin | 1545653 | RxNorm | NO | YES | NO |
| 793293 | ertugliflozin | 1992672 | RxNorm | NO | YES | NO |

##### 4.1.8 Concept: Sulfonylureas

| Concept ID | Concept Name | Code | Vocabulary | Excluded | Descendants | Mapped |
|---|---|---|---|---|---|---|
| 1594973 | chlorpropamide | 2404 | RxNorm | NO | YES | NO |
| 1597756 | glimepiride | 25789 | RxNorm | NO | YES | NO |
| 1560171 | glipizide | 4821 | RxNorm | NO | YES | NO |
| 19097821 | gliquidone | 25793 | RxNorm | NO | YES | NO |
| 1559684 | glyburide | 4815 | RxNorm | NO | YES | NO |
| 1502809 | tolazamide | 10633 | RxNorm | NO | YES | NO |
| 1502855 | tolbutamide | 10635 | RxNorm | NO | YES | NO |

##### 4.1.9 Concept: Other anti-diabetics

| Concept ID | Concept Name | Code | Vocabulary | Excluded | Descendants | Mapped |
|---|---|---|---|---|---|---|
| 1529331 | acarbose | 16681 | RxNorm | NO | YES | NO |
| 1530014 | acetohexamide | 173 | RxNorm | NO | YES | NO |
| 730548 | bromocriptine | 1760 | RxNorm | NO | YES | NO |
| 19033498 | carbutamide | 2068 | RxNorm | NO | YES | NO |
| 19001409 | glibornuride | 102846 | RxNorm | NO | YES | NO |
| 19059796 | gliclazide | 4816 | RxNorm | NO | YES | NO |
| 19001441 | glymidine | 102848 | RxNorm | NO | YES | NO |
| 1510202 | miglitol | 30009 | RxNorm | NO | YES | NO |
| 1502826 | nateglinide | 274332 | RxNorm | NO | YES | NO |
| 1525215 | pioglitazone | 33738 | RxNorm | NO | YES | NO |
| 1516766 | repaglinide | 73044 | RxNorm | NO | YES | NO |
| 1547504 | rosiglitazone | 84108 | RxNorm | NO | YES | NO |
| 1515249 | troglitazone | 72610 | RxNorm | NO | YES | NO |

##### 4.1.10 Concept: Insulin

| Concept ID | Concept Name | Code | Vocabulary | Excluded | Descendants | Mapped |
|---|---|---|---|---|---|---|
| 1596977 | insulin, regular, human | 253182 | RxNorm | NO | YES | NO |
| 1550023 | insulin lispro | 86009 | RxNorm | NO | YES | NO |
| 1567198 | insulin aspart, human | 51428 | RxNorm | NO | YES | NO |
| 1502905 | insulin glargine | 274783 | RxNorm | NO | YES | NO |
| 1513876 | insulin lispro protamine, human | 314684 | RxNorm | NO | YES | NO |
| 1531601 | insulin aspart protamine, human | 352385 | RxNorm | NO | YES | NO |
| 1586346 | insulin, regular, pork | 221109 | RxNorm | NO | YES | NO |
| 1544838 | insulin glulisine, human | 400008 | RxNorm | NO | YES | NO |
| 1516976 | insulin detemir | 139825 | RxNorm | NO | YES | NO |
| 1590165 | insulin, regular, beef-pork | 235275 | RxNorm | NO | YES | NO |
| 1513849 | lente insulin, human | 314683 | RxNorm | NO | YES | NO |
| 1562586 | lente insulin, pork | 93108 | RxNorm | NO | YES | NO |
| 1588986 | insulin human, rDNA origin | 631657 | RxNorm | NO | YES | NO |
| 1513843 | lente insulin, beef-pork | 314682 | RxNorm | NO | YES | NO |
| 1586369 | ultralente insulin, human | 221110 | RxNorm | NO | YES | NO |
| 35605670 | insulin argine | 1740938 | RxNorm | NO | YES | NO |
| 35602717 | insulin degludec | 1670007 | RxNorm | NO | YES | NO |
| 21600713 | INSULINS AND ANALOGUES | A10A | ATC | NO | YES | NO |
| 19078608 | insulin, protamine zinc, beef-pork 100 UNT/ML Injectable Suspension | 311053 | RxNorm | NO | YES | NO |

##### 4.1.11 Concept: Metformin

| Concept ID | Concept Name | Code | Vocabulary | Excluded | Descendants | Mapped |
|---|---|---|---|---|---|---|
| 1503297 | metformin | 6809 | RxNorm | NO | YES | NO |

##### 4.1.12 Concept: Secondary diabetes mellitus

| Concept ID | Concept Name | Code | Vocabulary | Excluded | Descendants | Mapped |
|---|---|---|---|---|---|---|
| 195771 | Secondary diabetes mellitus | 8801005 | SNOMED | NO | YES | NO |

##### 4.1.13 Concept: Type 1 diabetes mellitus

| Concept ID | Concept Name | Code | Vocabulary | Excluded | Descendants | Mapped |
|---|---|---|---|---|---|---|
| 201254 | Type 1 diabetes mellitus | 46635009 | SNOMED | NO | YES | NO |
| 435216 | Disorder due to type 1 diabetes mellitus | 420868002 | SNOMED | NO | YES | NO |
| 200687 | Renal disorder due to type 1 diabetes mellitus | 421893009 | SNOMED | NO | YES | NO |
| 377821 | Disorder of nervous system due to type 1 diabetes mellitus | 421468001 | SNOMED | NO | YES | NO |
| 318712 | Peripheral circulatory disorder due to type 1 diabetes mellitus | 421365002 | SNOMED | NO | YES | NO |

##### 4.1.14 Concept: Type 2 diabetes mellitus

| Concept ID | Concept Name | Code | Vocabulary | Excluded | Descendants | Mapped |
|---|---|---|---|---|---|---|
| 201826 | Type 2 diabetes mellitus | 44054006 | SNOMED | NO | YES | NO |
| 443734 | Ketoacidosis due to type 2 diabetes mellitus | 421750000 | SNOMED | NO | YES | NO |
| 443767 | Disorder of eye due to diabetes mellitus | 25093002 | SNOMED | NO | YES | NO |
| 192279 | Disorder of kidney due to diabetes mellitus | 127013003 | SNOMED | NO | YES | NO |
| 443735 | Coma due to diabetes mellitus | 420662003 | SNOMED | NO | YES | NO |
| 376065 | Disorder of nervous system due to type 2 diabetes mellitus | 421326000 | SNOMED | NO | YES | NO |
| 443729 | Peripheral circulatory disorder due to type 2 diabetes mellitus | 422166005 | SNOMED | NO | YES | NO |
| 443732 | Disorder due to type 2 diabetes mellitus | 422014003 | SNOMED | NO | YES | NO |

---

Note the above specifies an on-treatment time-at-risk; intent-to-treat analyses set the cohort end date to the observational end date for each patient.

#### 4.2 Negative control concepts

Negative outcome controls specified through condition occurrences that map to (a descendent of) the indicated concept ID

| | Concept ID |
|---|---|
| Abnormal posture | 439935 |
| Abnormal pupil | 436409 |
| Abrasion and/or friction burn of multiple sites | 443585 |
| Abrasion and/or friction burn of trunk without infection | 199192 |
| Absence of breast | 4088290 |
| Absent kidney | 4092879 |
| Acquired hallux valgus | 75911 |
| Acquired keratoderma | 137951 |
| Anal and rectal polyp | 73241 |
| Anomaly of jaw size | 45757682 |
| Benign paroxysmal positional vertigo | 81878 |
| Bizarre personal appearance | 4216219 |
| Burn of forearm | 133655 |
| Cachexia | 134765 |
| Calcaneal spur | 73560 |
| Cannabis abuse | 434327 |

*(Continued on Next Page...)*

Negative outcome controls specified through condition occurrences that map to (a descendent of) the indicated concept ID *(continued)*

| | Concept ID |
|---|---|
| Changes in skin texture | 140842 |
| Chondromalacia of patella | 81378 |
| Cocaine abuse | 432303 |
| Colostomy present | 4201390 |
| Complication due to Crohn's disease | 46269889 |
| Complication of gastrostomy | 434675 |
| Contact dermatitis | 134438 |
| Contusion of knee | 78619 |
| Crohn's disease | 201606 |
| Derangement of knee | 76786 |
| Developmental delay | 436077 |
| Deviated nasal septum | 377910 |
| Difficulty sleeping | 4115402 |
| Disproportion of reconstructed breast | 45757370 |
| Effects of hunger | 433111 |
| Endometriosis | 433527 |
| Epidermoid cyst | 4170770 |
| Exhaustion due to excessive exertion | 437448 |
| Feces contents abnormal | 4092896 |
| Feces contents abnormal | 4092896 |
| Foreign body in ear | 374801 |
| Foreign body in orifice | 259995 |
| Foreskin deficient | 4096540 |
| Galactosemia | 439788 |
| Ganglion cyst | 40481632 |
| Ganglion cyst | 40481632 |
| Genetic disorder carrier | 4168318 |
| Hammer toe | 433577 |
| Hereditary thrombophilia | 4231770 |
| High risk sexual behavior | 4012570 |
| Homocystinuria | 4012934 |
| Impacted cerumen | 374375 |
| Impacted cerumen | 374375 |
| Impingement syndrome of shoulder region | 4344500 |
| Inadequate sleep hygiene | 40481897 |
| Ingrowing nail | 139099 |
| Injury of knee | 444132 |
| Jellyfish poisoning | 4265896 |
| Kwashiorkor | 432593 |
| Lagophthalmos | 381021 |
| Late effect of contusion | 434203 |
| Late effect of motor vehicle accident | 438329 |
| Lipid storage disease | 4027782 |
| Lymphangioma | 433997 |
| Macular drusen | 4083487 |
| Malingering | 4051630 |
| Marfan's syndrome | 258540 |
| Mechanical complication of internal orthopedic device, implant AND/OR graft | 432798 |
| Melena | 4103703 |
| Minimal cognitive impairment | 439795 |
| Nicotine dependence | 4209423 |
| Nicotine dependence | 4209423 |
| Noise effects on inner ear | 377572 |
| Non-toxic multinodular goiter | 136368 |
| Nonspecific tuberculin test reaction | 40480893 |
| Nonspecific tuberculin test reaction | 40480893 |

*(Continued on Next Page...)*

Negative outcome controls specified through condition occurrences that map to (a descendent of) the indicated concept ID *(continued)*

| | Concept ID |
|---|---|
| Opioid abuse | 438130 |
| Opioid abuse | 438130 |
| Opioid intoxication | 4299094 |
| Passing flatus | 4091513 |
| Physiological development failure | 437092 |
| Poisoning by tranquilizer | 433951 |
| Postviral fatigue syndrome | 4202045 |
| Presbyopia | 373478 |
| Psychalgia | 439790 |
| Ptotic breast | 81634 |
| Regular astigmatism | 380706 |
| Senile hyperkeratosis | 141932 |
| Social exclusion | 4019836 |
| Somatic dysfunction of lumbar region | 36713918 |
| Splinter of face without major open wound | 443172 |
| Sprain of ankle | 81151 |
| Strain of rotator cuff capsule | 72748 |
| Symbolic dysfunction | 432436 |
| Tear film insufficiency | 378427 |
| Tobacco dependence syndrome | 437264 |
| Tooth loss | 433244 |
| Toxic effect of lead compound | 436876 |
| Toxic effect of tobacco and nicotine | 440612 |
| Tracheostomy present | 4201387 |
| Unsatisfactory tooth restoration | 45757285 |
| Verruca vulgaris | 140641 |
| Wrist joint pain | 4115367 |
| Wristdrop | 440193 |
