## Supplement B and C for "Sex Differences in Comparative Effectiveness and Safety of Second-line Antidiabetic Agents: Real-world Evidence from Large-scale Multinational Study"

### Supplementary Material B

for

### FEMALE SUBJECTS

in

### Sex Differences in Comparative Effectiveness and Safety of Second-line Antidiabetic Agents: Real-world Evidence from Large-scale Multinational Study

### 1 Exposures, outcomes and risk across data sources and outcomes

#### 1.1 GLP1RA (female) vs SGLT2I (female)

##### 1.1.1 3-point MACE

**Supplementary Table 1:** Exposures, outcomes and risk estimates for 3-point MACE in GLP1RA (female) (T) vs SGLT2I (female) (C) new-user cohorts under four different propensity score (PS) adjustment / time-at-risk (TAR) designs across data sources. We report total patient exposure time in 1,000 patient-year (KYr) units, uncalibrated and calibrated hazard ratios (HRs) and their 95% confidence intervals (CIs). Greyed out entries did not pass study diagnostics and are included only for completeness.

| | Patients | | Exposure (KYr) | | Outcomes | | Uncalibrated | | Calibrated | |
|---|---|---|---|---|---|---|---|---|---|---|
| | T | C | T | C | T | C | HR (95% CI) | $p$ | HR (95% CI) | $p$ |
| **PS-matching / on-treatment TAR** | | | | | | | | | | |
| CCAE | 10,652 | 18,456 | 6.9 | 13.4 | 31 | 88 | 0.71 (0.41 - 1.20) | 0.22 | 0.73 (0.43 - 1.25) | 0.25 |
| ODOD | 7,259 | 12,275 | 4.1 | 7.6 | 46 | 85 | 1.05 (0.65 - 1.69) | 0.84 | 1.03 (0.64 - 1.67) | 0.88 |
| MDCR | 698 | 1,406 | 0.4 | 0.9 | 5 | 12 | 0.83 (0.11 - 4.37) | 0.84 | 0.72 (0.12 - 4.51) | 0.73 |
| MDCD | 1,407 | 2,280 | 0.8 | 1.2 | 7 | 20 | 0.92 (0.31 - 2.55) | 0.88 | 0.89 (0.31 - 2.56) | 0.83 |
| Open Claims | 164,256 | 263,893 | 111.1 | 196.2 | 558 | 1,135 | 0.94 (0.81 - 1.08) | 0.38 | 0.92 (0.80 - 1.07) | 0.29 |
| OEHR | 9,548 | 16,375 | 2.8 | 5.1 | 20 | 31 | 1.02 (0.47 - 2.15) | 0.95 | 0.94 (0.44 - 2.00) | 0.86 |
| Meta-analysis | 193,122 | 313,279 | 125.7 | 223.5 | 662 | 1,359 | 0.93 (0.82 - 1.06) | 0.29 | 0.92 (0.80 - 1.05) | 0.22 |
| Meta-analysis (w/o USOC) | 28,866 | 49,386 | 14.6 | 27.3 | 104 | 224 | 0.91 (0.67 - 1.24) | 0.55 | 0.90 (0.66 - 1.23) | 0.53 |
| **PS-stratification / on-treatment TAR** | | | | | | | | | | |
| CCAE | 14,071 | 18,456 | 8.6 | 13.4 | 39 | 88 | 0.71 (0.47 - 1.05) | 0.09 | 0.72 (0.48 - 1.09) | 0.12 |
| ODOD | 9,783 | 12,278 | 5.2 | 7.6 | 59 | 85 | 1.20 (0.83 - 1.72) | 0.32 | 1.18 (0.82 - 1.70) | 0.37 |
| MDCR | 775 | 1,406 | 0.5 | 0.9 | 5 | 12 | 0.61 (0.18 - 1.77) | 0.39 | 0.54 (0.17 - 1.66) | 0.28 |
| MDCD | 1,662 | 2,284 | 0.9 | 1.2 | 8 | 20 | 0.59 (0.24 - 1.34) | 0.23 | 0.60 (0.25 - 1.44) | 0.26 |
| Open Claims | 223,161 | 263,915 | 139.1 | 196.3 | 676 | 1,135 | 0.97 (0.88 - 1.08) | 0.60 | 0.96 (0.86 - 1.06) | 0.43 |
| OEHR | 12,715 | 16,377 | 3.7 | 5.1 | 25 | 31 | 1.26 (0.71 - 2.22) | 0.43 | 1.11 (0.63 - 1.97) | 0.73 |
| Meta-analysis | 261,392 | 313,310 | 157.5 | 223.5 | 807 | 1,359 | 0.97 (0.88 - 1.07) | 0.54 | 0.96 (0.87 - 1.06) | 0.41 |
| Meta-analysis (w/o USOC) | 38,231 | 49,395 | 18.4 | 27.3 | 131 | 224 | 0.94 (0.67 - 1.33) | 0.73 | 0.94 (0.66 - 1.33) | 0.72 |
| **PS-matching / on-treatment TAR without escalation** | | | | | | | | | | |
| CCAE | 10,652 | 18,456 | 6.3 | 11.5 | 25 | 71 | 0.84 (0.45 - 1.53) | 0.57 | 0.86 (0.47 - 1.60) | 0.64 |
| ODOD | 7,259 | 12,275 | 3.7 | 6.7 | 41 | 77 | 1.05 (0.63 - 1.74) | 0.85 | 1.02 (0.61 - 1.71) | 0.90 |
| MDCR | 698 | 1,406 | 0.4 | 0.8 | 5 | 10 | 0.83 (0.11 - 4.37) | 0.84 | 0.74 (0.12 - 4.59) | 0.74 |
| MDCD | 1,407 | 2,280 | 0.7 | 1.0 | 6 | 12 | 1.65 (0.49 - 5.82) | 0.43 | 1.64 (0.47 - 5.65) | 0.44 |
| Open Claims | 164,256 | 263,893 | 99.3 | 165.0 | 483 | 947 | 0.92 (0.79 - 1.08) | 0.31 | 0.90 (0.77 - 1.05) | 0.20 |
| OEHR | 9,548 | 16,375 | 2.4 | 4.5 | 15 | 26 | 1.01 (0.44 - 2.21) | 0.99 | 0.91 (0.41 - 2.05) | 0.82 |
| Meta-analysis | 193,122 | 313,279 | 112.3 | 188.7 | 570 | 1,133 | 0.94 (0.81 - 1.08) | 0.37 | 0.92 (0.79 - 1.06) | 0.25 |
| Meta-analysis (w/o USOC) | 28,866 | 49,386 | 13.1 | 23.7 | 87 | 186 | 1.01 (0.72 - 1.41) | 0.97 | 1.00 (0.71 - 1.41) | 0.92 |
| **PS-stratification / on-treatment TAR without escalation** | | | | | | | | | | |
| CCAE | 14,071 | 18,456 | 7.8 | 11.5 | 31 | 71 | 0.68 (0.43 - 1.05) | 0.09 | 0.70 (0.45 - 1.11) | 0.13 |
| ODOD | 9,783 | 12,278 | 4.8 | 6.7 | 53 | 77 | 1.18 (0.80 - 1.72) | 0.41 | 1.19 (0.81 - 1.74) | 0.39 |
| MDCR | 775 | 1,406 | 0.4 | 0.8 | 5 | 10 | 0.83 (0.24 - 2.51) | 0.75 | 0.75 (0.23 - 2.39) | 0.62 |
| MDCD | 1,662 | 2,284 | 0.8 | 1.0 | 7 | 12 | 0.70 (0.25 - 1.81) | 0.48 | 0.74 (0.28 - 2.01) | 0.56 |
| Open Claims | 223,161 | 263,915 | 125.6 | 165.0 | 596 | 947 | 0.98 (0.87 - 1.09) | 0.69 | 0.96 (0.85 - 1.07) | 0.46 |
| OEHR | 12,715 | 16,377 | 3.2 | 4.5 | 20 | 26 | 1.11 (0.58 - 2.09) | 0.74 | 1.00 (0.53 - 1.89) | 0.92 |
| Meta-analysis | 261,392 | 313,310 | 142.2 | 188.8 | 707 | 1,133 | 0.97 (0.88 - 1.08) | 0.58 | 0.96 (0.86 - 1.07) | 0.45 |
| Meta-analysis (w/o USOC) | 38,231 | 49,395 | 16.6 | 23.7 | 111 | 186 | 0.93 (0.68 - 1.27) | 0.65 | 0.95 (0.69 - 1.29) | 0.72 |

##### 1.1.2 4-point MACE

**Supplementary Table 2:** Exposures, outcomes and risk estimates for 4-point MACE in GLP1RA (female) (T) vs SGLT2I (female) (C) new-user cohorts under four different propensity score (PS) adjustment / time-at-risk (TAR) designs across data sources. We report total patient exposure time in 1,000 patient-year (KYr) units, uncalibrated and calibrated hazard ratios (HRs) and their 95% confidence intervals (CIs). Greyed out entries did not pass study diagnostics and are included only for completeness.

| | Patients | | Exposure (KYr) | | Outcomes | | Uncalibrated | | Calibrated | |
|---|---|---|---|---|---|---|---|---|---|---|
| | T | C | T | C | T | C | HR (95% CI) | $p$ | HR (95% CI) | $p$ |
| **PS-matching / on-treatment TAR** | | | | | | | | | | |
| CCAE | 10,491 | 18,202 | 6.8 | 13.2 | 38 | 121 | 0.63 (0.39 - 0.98) | 0.05 | 0.64 (0.40 - 1.02) | 0.06 |
| ODOD | 7,024 | 11,795 | 4.0 | 7.3 | 83 | 129 | 1.21 (0.81 - 1.77) | 0.35 | 1.19 (0.80 - 1.75) | 0.41 |
| MDCR | 652 | 1,334 | 0.4 | 0.9 | 8 | 20 | 1.15 (0.29 - 4.09) | 0.83 | 1.01 (0.27 - 3.78) | 0.90 |
| MDCD | 1,331 | 2,154 | 0.8 | 1.2 | 11 | 35 | 0.38 (0.13 - 0.97) | 0.07 | 0.37 (0.13 - 1.03) | 0.06 |
| Open Claims | 160,005 | 255,636 | 108.2 | 191.0 | 852 | 1,703 | 0.98 (0.87 - 1.10) | 0.73 | 0.96 (0.86 - 1.08) | 0.56 |
| OEHR | 9,389 | 16,143 | 2.7 | 5.1 | 32 | 59 | 0.90 (0.51 - 1.56) | 0.72 | 0.83 (0.47 - 1.44) | 0.51 |
| Meta-analysis | 188,240 | 303,930 | 122.5 | 217.7 | 1,016 | 2,047 | 0.89 (0.70 - 1.14) | 0.36 | 0.88 (0.68 - 1.13) | 0.31 |
| Meta-analysis (w/o USOC) | 28,235 | 48,294 | 14.3 | 26.7 | 164 | 344 | 0.80 (0.53 - 1.22) | 0.30 | 0.79 (0.52 - 1.21) | 0.29 |
| **PS-stratification / on-treatment TAR** | | | | | | | | | | |
| CCAE | 13,882 | 18,202 | 8.5 | 13.2 | 46 | 121 | 0.59 (0.40 - 0.84) | <0.01 | 0.60 (0.42 - 0.87) | <0.01 |
| ODOD | 9,492 | 11,798 | 5.0 | 7.3 | 99 | 130 | 1.27 (0.95 - 1.68) | 0.10 | 1.25 (0.94 - 1.66) | 0.14 |
| MDCR | 736 | 1,334 | 0.4 | 0.9 | 9 | 20 | 0.68 (0.27 - 1.60) | 0.41 | 0.60 (0.25 - 1.47) | 0.27 |
| MDCD | 1,563 | 2,158 | 0.9 | 1.2 | 17 | 35 | 0.67 (0.35 - 1.22) | 0.21 | 0.69 (0.37 - 1.27) | 0.24 |
| Open Claims | 218,158 | 255,645 | 135.9 | 191.0 | 1,041 | 1,703 | 0.98 (0.91 - 1.07) | 0.71 | 0.97 (0.89 - 1.06) | 0.49 |
| OEHR | 12,521 | 16,145 | 3.6 | 5.1 | 41 | 59 | 1.09 (0.71 - 1.67) | 0.68 | 0.96 (0.63 - 1.48) | 0.85 |
| Meta-analysis | 255,616 | 303,948 | 153.9 | 217.7 | 1,244 | 2,048 | 0.92 (0.70 - 1.21) | 0.54 | 0.90 (0.68 - 1.19) | 0.48 |
| Meta-analysis (w/o USOC) | 37,458 | 48,303 | 18.0 | 26.7 | 203 | 345 | 0.88 (0.59 - 1.32) | 0.54 | 0.88 (0.58 - 1.32) | 0.53 |
| **PS-matching / on-treatment TAR without escalation** | | | | | | | | | | |
| CCAE | 10,491 | 18,202 | 6.1 | 11.4 | 31 | 102 | 0.66 (0.39 - 1.10) | 0.12 | 0.68 (0.41 - 1.15) | 0.16 |
| ODOD | 7,024 | 11,795 | 3.6 | 6.5 | 74 | 112 | 1.29 (0.85 - 1.95) | 0.23 | 1.26 (0.83 - 1.91) | 0.30 |
| MDCR | 652 | 1,334 | 0.4 | 0.8 | 8 | 17 | 1.15 (0.29 - 4.09) | 0.83 | 1.03 (0.27 - 3.84) | 0.89 |
| MDCD | 1,331 | 2,154 | 0.7 | 1.0 | 10 | 24 | 0.51 (0.16 - 1.36) | 0.22 | 0.51 (0.18 - 1.46) | 0.22 |
| Open Claims | 160,005 | 255,636 | 96.8 | 160.6 | 732 | 1,411 | 0.96 (0.85 - 1.09) | 0.57 | 0.94 (0.83 - 1.07) | 0.37 |
| OEHR | 9,389 | 16,143 | 2.4 | 4.4 | 24 | 49 | 0.91 (0.49 - 1.65) | 0.75 | 0.82 (0.45 - 1.52) | 0.54 |
| Meta-analysis | 188,240 | 303,930 | 109.6 | 183.9 | 871 | 1,698 | 0.96 (0.86 - 1.08) | 0.47 | 0.94 (0.83 - 1.06) | 0.31 |
| Meta-analysis (w/o USOC) | 28,235 | 48,294 | 12.8 | 23.2 | 139 | 287 | 0.88 (0.60 - 1.30) | 0.53 | 0.88 (0.59 - 1.30) | 0.51 |
| **PS-stratification / on-treatment TAR without escalation** | | | | | | | | | | |
| CCAE | 13,882 | 18,202 | 7.7 | 11.4 | 37 | 102 | 0.55 (0.36 - 0.81) | <0.01 | 0.57 (0.38 - 0.85) | <0.01 |
| ODOD | 9,492 | 11,798 | 4.6 | 6.5 | 89 | 112 | 1.32 (0.97 - 1.78) | 0.07 | 1.33 (0.98 - 1.80) | 0.07 |
| MDCR | 736 | 1,334 | 0.4 | 0.8 | 9 | 17 | 0.84 (0.33 - 2.00) | 0.71 | 0.76 (0.31 - 1.87) | 0.55 |
| MDCD | 1,563 | 2,158 | 0.8 | 1.0 | 14 | 24 | 0.78 (0.38 - 1.55) | 0.49 | 0.83 (0.41 - 1.68) | 0.62 |
| Open Claims | 218,158 | 255,645 | 122.8 | 160.7 | 905 | 1,411 | 0.98 (0.89 - 1.07) | 0.66 | 0.96 (0.87 - 1.05) | 0.40 |
| OEHR | 12,521 | 16,145 | 3.1 | 4.4 | 33 | 49 | 1.02 (0.63 - 1.64) | 0.93 | 0.92 (0.57 - 1.48) | 0.72 |
| Meta-analysis | 255,616 | 303,948 | 139.0 | 183.9 | 1,078 | 1,698 | 0.92 (0.68 - 1.24) | 0.58 | 0.91 (0.67 - 1.23) | 0.53 |
| Meta-analysis (w/o USOC) | 37,458 | 48,303 | 16.2 | 23.2 | 173 | 287 | 0.89 (0.57 - 1.39) | 0.60 | 0.90 (0.58 - 1.41) | 0.65 |

##### 1.1.3 Acute myocardial infarction

**Supplementary Table 3:** Exposures, outcomes and risk estimates for Acute myocardial infarction in GLP1RA (female) (T) vs SGLT2I (female) (C) new-user cohorts under four different propensity score (PS) adjustment / time-at-risk (TAR) designs across data sources. We report total patient exposure time in 1,000 patient-year (KYr) units, uncalibrated and calibrated hazard ratios (HRs) and their 95% confidence intervals (CIs). Greyed out entries did not pass study diagnostics and are included only for completeness.

| | Patients | | Exposure (KYr) | | Outcomes | | Uncalibrated | | Calibrated | |
|---|---|---|---|---|---|---|---|---|---|---|
| | T | C | T | C | T | C | HR (95% CI) | $p$ | HR (95% CI) | $p$ |
| **PS-matching / on-treatment TAR** | | | | | | | | | | |
| CCAE | 10,787 | 18,670 | 7.0 | 13.5 | 14 | 35 | 0.66 (0.25 - 1.56) | 0.37 | 0.67 (0.27 - 1.68) | 0.40 |
| ODOD | 7,385 | 12,541 | 4.2 | 7.7 | 20 | 34 | 1.28 (0.57 - 2.87) | 0.54 | 1.26 (0.56 - 2.84) | 0.58 |
| MDCR | 723 | 1,439 | 0.4 | 0.9 | <5 | 7 | 0.88 (0.04 - 9.66) | 0.93 | 0.77 (0.05 - 12.0) | 0.85 |
| MDCD | 1,449 | 2,357 | 0.8 | 1.3 | <5 | 8 | 1.26 (0.23 - 6.92) | 0.79 | 1.21 (0.22 - 6.66) | 0.83 |
| Open Claims | 167,713 | 269,863 | 113.4 | 200.4 | 237 | 547 | 0.99 (0.80 - 1.22) | 0.90 | 0.97 (0.78 - 1.20) | 0.79 |
| OEHR | 9,618 | 16,489 | 2.8 | 5.2 | 12 | 17 | 1.24 (0.36 - 3.94) | 0.72 | 1.14 (0.34 - 3.77) | 0.84 |
| Meta-analysis | 195,503 | 317,563 | 127.4 | 226.8 | 283 | 633 | 0.99 (0.81 - 1.21) | 0.92 | 0.97 (0.80 - 1.19) | 0.80 |
| Meta-analysis (w/o USOC) | 27,790 | 47,700 | 14.0 | 26.4 | 46 | 86 | 1.01 (0.59 - 1.74) | 0.97 | 1.00 (0.58 - 1.72) | 0.95 |
| **PS-stratification / on-treatment TAR** | | | | | | | | | | |
| CCAE | 14,254 | 18,670 | 8.7 | 13.5 | 18 | 35 | 0.93 (0.50 - 1.67) | 0.81 | 0.95 (0.52 - 1.74) | 0.86 |
| ODOD | 9,933 | 12,545 | 5.3 | 7.7 | 25 | 34 | 1.29 (0.72 - 2.27) | 0.38 | 1.27 (0.72 - 2.26) | 0.41 |
| MDCR | 805 | 1,439 | 0.5 | 0.9 | <5 | 7 | 0.50 (0.07 - 2.39) | 0.45 | 0.44 (0.08 - 2.61) | 0.37 |
| MDCD | 1,709 | 2,362 | 0.9 | 1.3 | <5 | 8 | 0.63 (0.13 - 2.37) | 0.53 | 0.64 (0.15 - 2.74) | 0.55 |
| Open Claims | 227,503 | 269,891 | 141.8 | 200.4 | 281 | 548 | 0.89 (0.76 - 1.04) | 0.14 | 0.88 (0.75 - 1.02) | 0.10 |
| OEHR | 12,808 | 16,491 | 3.7 | 5.2 | 15 | 17 | 1.68 (0.78 - 3.57) | 0.18 | 1.49 (0.70 - 3.18) | 0.31 |
| Meta-analysis | 264,498 | 317,597 | 159.5 | 226.8 | 339 | 634 | 1.01 (0.78 - 1.31) | 0.93 | 1.00 (0.77 - 1.29) | 0.96 |
| Meta-analysis (w/o USOC) | 36,995 | 47,706 | 17.7 | 26.5 | 58 | 86 | 1.22 (0.85 - 1.75) | 0.29 | 1.21 (0.84 - 1.75) | 0.31 |
| **PS-matching / on-treatment TAR without escalation** | | | | | | | | | | |
| CCAE | 10,787 | 18,670 | 6.3 | 11.7 | 12 | 28 | 0.97 (0.35 - 2.47) | 0.96 | 1.00 (0.38 - 2.65) | 0.96 |
| ODOD | 7,385 | 12,541 | 3.8 | 6.9 | 18 | 29 | 1.45 (0.58 - 3.56) | 0.42 | 1.41 (0.57 - 3.49) | 0.46 |
| MDCR | 723 | 1,439 | 0.4 | 0.8 | <5 | 6 | 0.88 (0.04 - 9.66) | 0.93 | 0.78 (0.05 - 12.2) | 0.86 |
| MDCD | 1,449 | 2,357 | 0.7 | 1.0 | <5 | 6 | 1.26 (0.23 - 6.92) | 0.79 | 1.25 (0.23 - 6.86) | 0.80 |
| Open Claims | 167,713 | 269,863 | 101.3 | 168.5 | 210 | 457 | 0.98 (0.78 - 1.23) | 0.87 | 0.96 (0.76 - 1.21) | 0.72 |
| OEHR | 9,618 | 16,489 | 2.4 | 4.5 | 9 | 16 | 1.07 (0.31 - 3.40) | 0.91 | 0.97 (0.29 - 3.22) | 0.93 |
| Meta-analysis | 195,503 | 317,563 | 113.9 | 191.5 | 249 | 530 | 1.01 (0.81 - 1.25) | 0.96 | 0.98 (0.79 - 1.22) | 0.87 |
| Meta-analysis (w/o USOC) | 27,790 | 47,700 | 12.6 | 23.0 | 39 | 73 | 1.17 (0.66 - 2.09) | 0.59 | 1.16 (0.65 - 2.08) | 0.61 |
| **PS-stratification / on-treatment TAR without escalation** | | | | | | | | | | |
| CCAE | 14,254 | 18,670 | 7.9 | 11.7 | 16 | 28 | 0.99 (0.51 - 1.89) | 0.98 | 1.03 (0.53 - 2.00) | 0.93 |
| ODOD | 9,933 | 12,545 | 4.9 | 6.9 | 23 | 29 | 1.40 (0.76 - 2.58) | 0.28 | 1.42 (0.77 - 2.61) | 0.27 |
| MDCR | 805 | 1,439 | 0.4 | 0.8 | <5 | 6 | 0.73 (0.10 - 3.51) | 0.73 | 0.66 (0.11 - 3.88) | 0.65 |
| MDCD | 1,709 | 2,362 | 0.8 | 1.0 | <5 | 6 | 0.74 (0.14 - 3.03) | 0.69 | 0.78 (0.17 - 3.58) | 0.75 |
| Open Claims | 227,503 | 269,891 | 128.0 | 168.5 | 251 | 458 | 0.93 (0.78 - 1.09) | 0.37 | 0.91 (0.76 - 1.07) | 0.26 |
| OEHR | 12,808 | 16,491 | 3.2 | 4.5 | 12 | 16 | 1.38 (0.59 - 3.13) | 0.44 | 1.24 (0.54 - 2.85) | 0.62 |
| Meta-analysis | 264,498 | 317,597 | 144.0 | 191.5 | 302 | 531 | 1.00 (0.81 - 1.22) | 0.97 | 0.98 (0.80 - 1.21) | 0.87 |
| Meta-analysis (w/o USOC) | 36,995 | 47,706 | 16.0 | 23.0 | 51 | 73 | 1.24 (0.83 - 1.83) | 0.29 | 1.26 (0.84 - 1.87) | 0.26 |

##### 1.1.4 Acute renal failure

**Supplementary Table 4:** Exposures, outcomes and risk estimates for Acute renal failure in GLP1RA (female) (T) vs SGLT2I (female) (C) new-user cohorts under four different propensity score (PS) adjustment / time-at-risk (TAR) designs across data sources. We report total patient exposure time in 1,000 patient-year (KYr) units, uncalibrated and calibrated hazard ratios (HRs) and their 95% confidence intervals (CIs). Greyed out entries did not pass study diagnostics and are included only for completeness.

| | Patients | | Exposure (KYr) | | Outcomes | | Uncalibrated | | Calibrated | |
|---|---|---|---|---|---|---|---|---|---|---|
| | T | C | T | C | T | C | HR (95% CI) | $p$ | HR (95% CI) | $p$ |
| **PS-matching / on-treatment TAR** | | | | | | | | | | |
| CCAE | 10,761 | 18,692 | 7.0 | 13.5 | 39 | 62 | 1.01 (0.57 - 1.76) | 0.97 | 1.03 (0.59 - 1.81) | 0.90 |
| ODOD | 7,251 | 12,443 | 4.1 | 7.6 | 58 | 120 | 0.86 (0.55 - 1.33) | 0.51 | 0.85 (0.54 - 1.32) | 0.48 |
| MDCR | 702 | 1,448 | 0.4 | 0.9 | 8 | 13 | 0.69 (0.21 - 2.08) | 0.53 | 0.60 (0.19 - 1.91) | 0.39 |
| MDCD | 1,404 | 2,308 | 0.8 | 1.2 | 13 | 21 | 1.81 (0.64 - 5.52) | 0.28 | 1.75 (0.59 - 5.12) | 0.32 |
| Open Claims | 166,240 | 269,436 | 112.0 | 200.1 | 766 | 1,385 | 1.09 (0.97 - 1.23) | 0.16 | 1.07 (0.95 - 1.21) | 0.28 |
| OEHR | 9,599 | 16,499 | 2.8 | 5.2 | 35 | 39 | 2.22 (1.23 - 4.06) | <0.01 | 2.03 (1.11 - 3.68) | 0.02 |
| Meta-analysis | 195,255 | 319,378 | 126.7 | 227.7 | 911 | 1,627 | 1.18 (0.88 - 1.58) | 0.27 | 1.16 (0.86 - 1.56) | 0.32 |
| Meta-analysis (w/o USOC) | 29,015 | 49,942 | 14.7 | 27.6 | 145 | 242 | 1.28 (0.79 - 2.06) | 0.31 | 1.27 (0.79 - 2.04) | 0.33 |
| **PS-stratification / on-treatment TAR** | | | | | | | | | | |
| CCAE | 14,196 | 18,692 | 8.7 | 13.5 | 56 | 62 | 1.26 (0.84 - 1.87) | 0.26 | 1.29 (0.86 - 1.92) | 0.22 |
| ODOD | 9,772 | 12,447 | 5.2 | 7.6 | 71 | 120 | 0.99 (0.72 - 1.37) | 0.97 | 0.98 (0.71 - 1.36) | 0.89 |
| MDCR | 783 | 1,449 | 0.5 | 0.9 | 8 | 13 | 1.10 (0.42 - 2.75) | 0.84 | 0.97 (0.38 - 2.48) | 0.90 |
| MDCD | 1,659 | 2,313 | 0.9 | 1.2 | 16 | 21 | 1.06 (0.53 - 2.09) | 0.87 | 1.08 (0.54 - 2.16) | 0.82 |
| Open Claims | 225,203 | 269,448 | 140.0 | 200.1 | 936 | 1,386 | 1.09 (1.00 - 1.19) | 0.06 | 1.07 (0.98 - 1.18) | 0.18 |
| OEHR | 12,747 | 16,501 | 3.7 | 5.2 | 43 | 39 | 1.66 (1.04 - 2.64) | 0.03 | 1.46 (0.92 - 2.33) | 0.12 |
| Meta-analysis | 263,577 | 319,401 | 158.5 | 227.7 | 1,122 | 1,628 | 1.10 (1.02 - 1.20) | 0.02 | 1.09 (0.99 - 1.19) | 0.07 |
| Meta-analysis (w/o USOC) | 38,374 | 49,953 | 18.5 | 27.6 | 186 | 242 | 1.19 (0.95 - 1.49) | 0.13 | 1.19 (0.95 - 1.49) | 0.15 |
| **PS-matching / on-treatment TAR without escalation** | | | | | | | | | | |
| CCAE | 10,761 | 18,692 | 6.3 | 11.7 | 34 | 53 | 1.01 (0.56 - 1.80) | 0.97 | 1.04 (0.58 - 1.88) | 0.89 |
| ODOD | 7,251 | 12,443 | 3.7 | 6.8 | 51 | 103 | 0.90 (0.56 - 1.43) | 0.66 | 0.88 (0.55 - 1.41) | 0.60 |
| MDCR | 702 | 1,448 | 0.4 | 0.8 | 7 | 12 | 0.55 (0.15 - 1.77) | 0.35 | 0.49 (0.14 - 1.71) | 0.27 |
| MDCD | 1,404 | 2,308 | 0.7 | 1.0 | 12 | 14 | 2.14 (0.63 - 8.40) | 0.25 | 2.12 (0.58 - 7.75) | 0.26 |
| Open Claims | 166,240 | 269,436 | 100.1 | 168.4 | 650 | 1,116 | 1.07 (0.94 - 1.22) | 0.30 | 1.05 (0.91 - 1.20) | 0.54 |
| OEHR | 9,599 | 16,499 | 2.4 | 4.5 | 23 | 29 | 1.87 (0.95 - 3.74) | 0.07 | 1.70 (0.86 - 3.37) | 0.13 |
| Meta-analysis | 195,255 | 319,378 | 113.2 | 192.3 | 770 | 1,315 | 1.08 (0.96 - 1.22) | 0.20 | 1.06 (0.93 - 1.20) | 0.41 |
| Meta-analysis (w/o USOC) | 29,015 | 49,942 | 13.2 | 24.0 | 120 | 199 | 1.19 (0.81 - 1.74) | 0.38 | 1.18 (0.80 - 1.73) | 0.41 |
| **PS-stratification / on-treatment TAR without escalation** | | | | | | | | | | |
| CCAE | 14,196 | 18,692 | 7.9 | 11.7 | 50 | 53 | 1.29 (0.84 - 1.96) | 0.25 | 1.33 (0.87 - 2.04) | 0.19 |
| ODOD | 9,772 | 12,447 | 4.8 | 6.8 | 64 | 103 | 1.01 (0.71 - 1.41) | 0.97 | 1.01 (0.72 - 1.43) | 0.90 |
| MDCR | 783 | 1,449 | 0.4 | 0.8 | 7 | 12 | 0.89 (0.32 - 2.29) | 0.82 | 0.80 (0.30 - 2.14) | 0.66 |
| MDCD | 1,659 | 2,313 | 0.8 | 1.0 | 14 | 14 | 1.37 (0.63 - 3.00) | 0.42 | 1.46 (0.67 - 3.20) | 0.35 |
| Open Claims | 225,203 | 269,448 | 126.4 | 168.4 | 809 | 1,117 | 1.10 (1.00 - 1.22) | 0.05 | 1.08 (0.98 - 1.20) | 0.16 |
| OEHR | 12,747 | 16,501 | 3.2 | 4.5 | 29 | 29 | 1.36 (0.78 - 2.36) | 0.28 | 1.21 (0.70 - 2.12) | 0.50 |
| Meta-analysis | 263,577 | 319,401 | 143.0 | 192.4 | 966 | 1,316 | 1.11 (1.02 - 1.22) | 0.02 | 1.10 (1.00 - 1.21) | 0.06 |
| Meta-analysis (w/o USOC) | 38,374 | 49,953 | 16.7 | 24.0 | 157 | 199 | 1.17 (0.93 - 1.47) | 0.18 | 1.19 (0.94 - 1.50) | 0.15 |

##### 1.1.5 Glycemic control

**Supplementary Table 5:** Exposures, outcomes and risk estimates for Glycemic control in GLP1RA (female) (T) vs SGLT2I (female) (C) new-user cohorts under four different propensity score (PS) adjustment / time-at-risk (TAR) designs across data sources. We report total patient exposure time in 1,000 patient-year (KYr) units, uncalibrated and calibrated hazard ratios (HRs) and their 95% confidence intervals (CIs). Greyed out entries did not pass study diagnostics and are included only for completeness.

| | Patients | | Exposure (KYr) | | Outcomes | | Uncalibrated | | Calibrated | |
|---|---|---|---|---|---|---|---|---|---|---|
| | T | C | T | C | T | C | HR (95% CI) | $p$ | HR (95% CI) | $p$ |
| **PS-matching / on-treatment TAR** | | | | | | | | | | |
| CCAE | 10,790 | 18,683 | 7.0 | 13.5 | 10 | 22 | 1.58 (0.55 - 4.40) | 0.39 | 1.62 (0.57 - 4.59) | 0.37 |
| ODOD | 4,647 | 7,822 | 2.5 | 4.4 | 290 | 456 | 1.53 (1.23 - 1.91) | <0.01 | 1.50 (1.21 - 1.88) | <0.01 |
| MDCR | 732 | 1,481 | 0.4 | 0.9 | 0 | 0 | | | | |
| MDCD | 1,485 | 2,430 | 0.8 | 1.3 | 0 | 0 | | | | |
| Open Claims | 170,783 | 276,514 | 115.3 | 205.0 | 0 | 0 | | | | |
| OEHR | 9,714 | 16,664 | 2.8 | 5.2 | 0 | 0 | | | | |
| Meta-analysis | 15,437 | 26,505 | 9.5 | 18.0 | 300 | 478 | 1.53 (1.24 - 1.90) | <0.01 | 1.51 (1.21 - 1.87) | <0.01 |
| Meta-analysis (w/o USOC) | 15,437 | 26,505 | 9.5 | 18.0 | 300 | 478 | 1.53 (1.24 - 1.90) | <0.01 | 1.52 (1.22 - 1.89) | <0.01 |
| **PS-stratification / on-treatment TAR** | | | | | | | | | | |
| CCAE | 14,261 | 18,683 | 8.7 | 13.5 | 11 | 22 | 1.28 (0.57 - 2.72) | 0.54 | 1.31 (0.60 - 2.87) | 0.50 |
| ODOD | 6,078 | 7,839 | 3.1 | 4.4 | 346 | 456 | 1.26 (1.08 - 1.46) | <0.01 | 1.24 (1.06 - 1.45) | 0.01 |
| MDCR | 815 | 1,482 | 0.5 | 0.9 | 0 | 0 | | | | |
| MDCD | 1,747 | 2,435 | 1.0 | 1.3 | 0 | 0 | | | | |
| Open Claims | 230,932 | 276,521 | 143.9 | 205.0 | 0 | 0 | | | | |
| OEHR | 12,899 | 16,666 | 3.7 | 5.2 | 0 | 0 | | | | |
| Meta-analysis | 20,339 | 26,522 | 11.8 | 18.0 | 357 | 478 | 1.26 (1.08 - 1.46) | <0.01 | 1.24 (1.06 - 1.45) | <0.01 |
| Meta-analysis (w/o USOC) | 20,339 | 26,522 | 11.8 | 18.0 | 357 | 478 | 1.26 (1.08 - 1.46) | <0.01 | 1.25 (1.08 - 1.46) | <0.01 |
| **PS-matching / on-treatment TAR without escalation** | | | | | | | | | | |
| CCAE | 10,790 | 18,683 | 6.3 | 11.7 | 10 | 18 | 2.33 (0.75 - 7.46) | 0.15 | 2.40 (0.76 - 7.58) | 0.14 |
| ODOD | 4,647 | 7,822 | 2.3 | 3.9 | 261 | 397 | 1.49 (1.17 - 1.89) | <0.01 | 1.45 (1.14 - 1.85) | <0.01 |
| MDCR | 732 | 1,481 | 0.4 | 0.8 | 0 | 0 | | | | |
| MDCD | 1,485 | 2,430 | 0.7 | 1.1 | 0 | 0 | | | | |
| Open Claims | 170,783 | 276,514 | 103.0 | 172.3 | 0 | 0 | | | | |
| OEHR | 9,714 | 16,664 | 2.4 | 4.6 | 0 | 0 | | | | |
| Meta-analysis | 15,437 | 26,505 | 8.6 | 15.6 | 271 | 415 | 1.51 (1.20 - 1.91) | <0.01 | 1.48 (1.16 - 1.88) | <0.01 |
| Meta-analysis (w/o USOC) | 15,437 | 26,505 | 8.6 | 15.6 | 271 | 415 | 1.51 (1.20 - 1.91) | <0.01 | 1.50 (1.19 - 1.91) | <0.01 |
| **PS-stratification / on-treatment TAR without escalation** | | | | | | | | | | |
| CCAE | 14,261 | 18,683 | 7.9 | 11.7 | 11 | 18 | 1.49 (0.64 - 3.30) | 0.34 | 1.55 (0.68 - 3.51) | 0.29 |
| ODOD | 6,078 | 7,839 | 2.8 | 3.9 | 313 | 397 | 1.29 (1.09 - 1.51) | <0.01 | 1.30 (1.10 - 1.53) | <0.01 |
| MDCR | 815 | 1,482 | 0.4 | 0.8 | 0 | 0 | | | | |
| MDCD | 1,747 | 2,435 | 0.8 | 1.1 | 0 | 0 | | | | |
| Open Claims | 230,932 | 276,521 | 129.8 | 172.3 | 0 | 0 | | | | |
| OEHR | 12,899 | 16,666 | 3.2 | 4.6 | 0 | 0 | | | | |
| Meta-analysis | 20,339 | 26,522 | 10.7 | 15.6 | 324 | 415 | 1.29 (1.10 - 1.52) | <0.01 | 1.28 (1.08 - 1.50) | <0.01 |
| Meta-analysis (w/o USOC) | 20,339 | 26,522 | 10.7 | 15.6 | 324 | 415 | 1.29 (1.10 - 1.52) | <0.01 | 1.31 (1.12 - 1.54) | <0.01 |

##### 1.1.6 Hospitalization with heart failure

**Supplementary Table 6:** Exposures, outcomes and risk estimates for Hospitalization with heart failure in GLP1RA (female) (T) vs SGLT2I (female) (C) new-user cohorts under four different propensity score (PS) adjustment / time-at-risk (TAR) designs across data sources. We report total patient exposure time in 1,000 patient-year (KYr) units, uncalibrated and calibrated hazard ratios (HRs) and their 95% confidence intervals (CIs). Greyed out entries did not pass study diagnostics and are included only for completeness.

| | Patients | | Exposure (KYr) | | Outcomes | | Uncalibrated | | Calibrated | |
|---|---|---|---|---|---|---|---|---|---|---|
| | T | C | T | C | T | C | HR (95% CI) | $p$ | HR (95% CI) | $p$ |
| **PS-matching / on-treatment TAR** | | | | | | | | | | |
| CCAE | 10,691 | 18,562 | 7.0 | 13.5 | 12 | 47 | 0.27 (0.10 - 0.61) | <0.01 | 0.28 (0.11 - 0.68) | <0.01 |
| ODOD | 7,194 | 12,161 | 4.1 | 7.5 | 55 | 82 | 1.35 (0.82 - 2.21) | 0.23 | 1.33 (0.81 - 2.18) | 0.27 |
| MDCR | 684 | 1,394 | 0.4 | 0.9 | 7 | 14 | 1.11 (0.21 - 5.12) | 0.90 | 0.97 (0.20 - 4.73) | 0.91 |
| MDCD | 1,382 | 2,261 | 0.8 | 1.2 | 11 | 20 | 0.43 (0.10 - 1.36) | 0.21 | 0.41 (0.11 - 1.55) | 0.19 |
| Open Claims | 165,236 | 265,014 | 111.7 | 197.8 | 493 | 960 | 1.12 (0.97 - 1.30) | 0.13 | 1.10 (0.95 - 1.29) | 0.22 |
| OEHR | 9,525 | 16,385 | 2.8 | 5.1 | 20 | 36 | 0.96 (0.47 - 1.88) | 0.90 | 0.87 (0.44 - 1.75) | 0.70 |
| Meta-analysis | 194,028 | 314,383 | 126.3 | 225.1 | 591 | 1,145 | 0.81 (0.47 - 1.40) | 0.45 | 0.80 (0.46 - 1.38) | 0.42 |
| Meta-analysis (w/o USOC) | 28,792 | 49,369 | 14.6 | 27.3 | 98 | 185 | 0.69 (0.33 - 1.45) | 0.33 | 0.68 (0.33 - 1.43) | 0.32 |
| **PS-stratification / on-treatment TAR** | | | | | | | | | | |
| CCAE | 14,134 | 18,562 | 8.7 | 13.5 | 12 | 47 | 0.38 (0.18 - 0.73) | <0.01 | 0.39 (0.20 - 0.77) | <0.01 |
| ODOD | 9,699 | 12,165 | 5.2 | 7.5 | 60 | 82 | 1.31 (0.91 - 1.87) | 0.14 | 1.30 (0.90 - 1.86) | 0.17 |
| MDCR | 771 | 1,395 | 0.5 | 0.9 | 8 | 14 | 1.03 (0.37 - 2.66) | 0.95 | 0.90 (0.34 - 2.43) | 0.84 |
| MDCD | 1,619 | 2,266 | 0.9 | 1.2 | 16 | 20 | 1.06 (0.52 - 2.13) | 0.87 | 1.08 (0.53 - 2.19) | 0.83 |
| Open Claims | 224,509 | 265,019 | 139.9 | 197.8 | 587 | 960 | 1.06 (0.95 - 1.19) | 0.27 | 1.05 (0.93 - 1.17) | 0.47 |
| OEHR | 12,672 | 16,387 | 3.6 | 5.1 | 25 | 36 | 1.11 (0.64 - 1.89) | 0.71 | 0.98 (0.57 - 1.68) | 0.90 |
| Meta-analysis | 262,633 | 314,399 | 158.3 | 225.1 | 700 | 1,145 | 0.97 (0.69 - 1.37) | 0.88 | 0.96 (0.68 - 1.36) | 0.82 |
| Meta-analysis (w/o USOC) | 38,124 | 49,380 | 18.4 | 27.4 | 113 | 185 | 0.92 (0.55 - 1.51) | 0.73 | 0.91 (0.55 - 1.51) | 0.72 |
| **PS-matching / on-treatment TAR without escalation** | | | | | | | | | | |
| CCAE | 10,691 | 18,562 | 6.3 | 11.6 | 9 | 42 | 0.32 (0.12 - 0.74) | 0.02 | 0.33 (0.13 - 0.83) | 0.02 |
| ODOD | 7,194 | 12,161 | 3.7 | 6.7 | 49 | 70 | 1.45 (0.85 - 2.47) | 0.17 | 1.41 (0.83 - 2.41) | 0.22 |
| MDCR | 684 | 1,394 | 0.4 | 0.8 | 6 | 13 | 0.78 (0.11 - 4.08) | 0.79 | 0.69 (0.11 - 4.30) | 0.70 |
| MDCD | 1,382 | 2,261 | 0.7 | 1.0 | 10 | 14 | 0.52 (0.12 - 1.71) | 0.34 | 0.52 (0.13 - 1.98) | 0.34 |
| Open Claims | 165,236 | 265,014 | 99.8 | 166.2 | 425 | 794 | 1.10 (0.93 - 1.29) | 0.27 | 1.07 (0.91 - 1.27) | 0.44 |
| OEHR | 9,525 | 16,385 | 2.4 | 4.5 | 15 | 29 | 0.90 (0.40 - 1.93) | 0.79 | 0.82 (0.38 - 1.79) | 0.62 |
| Meta-analysis | 194,028 | 314,383 | 112.9 | 190.0 | 508 | 949 | 0.87 (0.54 - 1.42) | 0.59 | 0.85 (0.53 - 1.39) | 0.53 |
| Meta-analysis (w/o USOC) | 28,792 | 49,369 | 13.1 | 23.8 | 83 | 155 | 0.75 (0.37 - 1.52) | 0.43 | 0.75 (0.37 - 1.52) | 0.42 |
| **PS-stratification / on-treatment TAR without escalation** | | | | | | | | | | |
| CCAE | 14,134 | 18,562 | 7.9 | 11.6 | 9 | 42 | 0.30 (0.13 - 0.61) | <0.01 | 0.31 (0.14 - 0.67) | <0.01 |
| ODOD | 9,699 | 12,165 | 4.7 | 6.7 | 54 | 70 | 1.39 (0.94 - 2.03) | 0.09 | 1.40 (0.95 - 2.06) | 0.09 |
| MDCR | 771 | 1,395 | 0.4 | 0.8 | 7 | 13 | 0.84 (0.28 - 2.25) | 0.74 | 0.76 (0.27 - 2.14) | 0.60 |
| MDCD | 1,619 | 2,266 | 0.8 | 1.0 | 13 | 14 | 1.27 (0.56 - 2.83) | 0.57 | 1.35 (0.60 - 3.03) | 0.48 |
| Open Claims | 224,509 | 265,019 | 126.3 | 166.2 | 507 | 794 | 1.06 (0.94 - 1.20) | 0.34 | 1.04 (0.92 - 1.17) | 0.58 |
| OEHR | 12,672 | 16,387 | 3.2 | 4.5 | 20 | 29 | 1.05 (0.56 - 1.92) | 0.88 | 0.94 (0.51 - 1.74) | 0.84 |
| Meta-analysis | 262,633 | 314,399 | 142.9 | 190.0 | 603 | 949 | 0.95 (0.60 - 1.51) | 0.83 | 0.94 (0.59 - 1.49) | 0.78 |
| Meta-analysis (w/o USOC) | 38,124 | 49,380 | 16.6 | 23.8 | 96 | 155 | 0.90 (0.48 - 1.68) | 0.73 | 0.91 (0.48 - 1.71) | 0.77 |

##### 1.1.7 Measured renal dysfunction

**Supplementary Table 7:** Exposures, outcomes and risk estimates for Measured renal dysfunction in GLP1RA (female) (T) vs SGLT2I (female) (C) new-user cohorts under four different propensity score (PS) adjustment / time-at-risk (TAR) designs across data sources. We report total patient exposure time in 1,000 patient-year (KYr) units, uncalibrated and calibrated hazard ratios (HRs) and their 95% confidence intervals (CIs). Greyed out entries did not pass study diagnostics and are included only for completeness.

| | Patients | | Exposure (KYr) | | Outcomes | | Uncalibrated | | Calibrated | |
|---|---|---|---|---|---|---|---|---|---|---|
| | T | C | T | C | T | C | HR (95% CI) | $p$ | HR (95% CI) | $p$ |
| **PS-matching / on-treatment TAR** | | | | | | | | | | |
| CCAE | 10,862 | 18,849 | 7.1 | 13.7 | <5 | <5 | | | | |
| ODOD | 7,487 | 12,830 | 4.2 | 7.9 | <5 | 10 | 1.29 (0.16 - 8.55) | 0.80 | 1.27 (0.17 - 9.31) | 0.81 |
| MDCR | 736 | 1,488 | 0.4 | 0.9 | 0 | 0 | | | | |
| MDCD | 1,485 | 2,430 | 0.8 | 1.3 | 0 | 0 | | | | |
| Open Claims | 170,783 | 276,514 | 115.3 | 205.0 | 0 | 0 | | | | |
| OEHR | 9,714 | 16,664 | 2.8 | 5.2 | 0 | 0 | | | | |
| **PS-stratification / on-treatment TAR** | | | | | | | | | | |
| CCAE | 14,338 | 18,849 | 8.8 | 13.7 | <5 | <5 | 3.20 (0.36 - 28.1) | 0.29 | 3.27 (0.37 - 28.8) | 0.29 |
| ODOD | 10,060 | 12,834 | 5.4 | 7.9 | <5 | 10 | 0.33 (0.05 - 1.33) | 0.19 | 0.33 (0.06 - 1.70) | 0.18 |
| MDCR | 819 | 1,489 | 0.5 | 0.9 | 0 | 0 | | | | |
| MDCD | 1,747 | 2,435 | 1.0 | 1.3 | 0 | 0 | | | | |
| Open Claims | 230,932 | 276,521 | 143.9 | 205.0 | 0 | 0 | | | | |
| OEHR | 12,899 | 16,666 | 3.7 | 5.2 | 0 | 0 | | | | |
| **PS-matching / on-treatment TAR without escalation** | | | | | | | | | | |
| CCAE | 10,862 | 18,849 | 6.4 | 11.8 | <5 | <5 | | | | |
| ODOD | 7,487 | 12,830 | 3.9 | 7.0 | <5 | 8 | 2.45 (0.27 - 22.1) | 0.42 | 2.39 (0.26 - 21.6) | 0.44 |
| MDCR | 736 | 1,488 | 0.4 | 0.8 | 0 | 0 | | | | |
| MDCD | 1,485 | 2,430 | 0.7 | 1.1 | 0 | 0 | | | | |
| Open Claims | 170,783 | 276,514 | 103.0 | 172.3 | 0 | 0 | | | | |
| OEHR | 9,714 | 16,664 | 2.4 | 4.6 | 0 | 0 | | | | |
| **PS-stratification / on-treatment TAR without escalation** | | | | | | | | | | |
| CCAE | 14,338 | 18,849 | 8.0 | 11.8 | <5 | <5 | 3.22 (0.37 - 28.3) | 0.29 | 3.34 (0.38 - 29.3) | 0.28 |
| ODOD | 10,060 | 12,834 | 4.9 | 7.0 | <5 | 8 | 0.47 (0.07 - 1.97) | 0.38 | 0.47 (0.09 - 2.54) | 0.38 |
| MDCR | 819 | 1,489 | 0.4 | 0.8 | 0 | 0 | | | | |
| MDCD | 1,747 | 2,435 | 0.8 | 1.1 | 0 | 0 | | | | |
| Open Claims | 230,932 | 276,521 | 129.8 | 172.3 | 0 | 0 | | | | |
| OEHR | 12,899 | 16,666 | 3.2 | 4.6 | 0 | 0 | | | | |

##### 1.1.8 Stroke

**Supplementary Table 8:** Exposures, outcomes and risk estimates for Stroke in GLP1RA (female) (T) vs SGLT2I (female) (C) new-user cohorts under four different propensity score (PS) adjustment / time-at-risk (TAR) designs across data sources. We report total patient exposure time in 1,000 patient-year (KYr) units, uncalibrated and calibrated hazard ratios (HRs) and their 95% confidence intervals (CIs). Greyed out entries did not pass study diagnostics and are included only for completeness.

| | Patients | | Exposure (KYr) | | Outcomes | | Uncalibrated | | Calibrated | |
|---|---|---|---|---|---|---|---|---|---|---|
| | T | C | T | C | T | C | HR (95% CI) | $p$ | HR (95% CI) | $p$ |
| **PS-matching / on-treatment TAR** | | | | | | | | | | |
| CCAE | 10,750 | 18,673 | 7.0 | 13.5 | 17 | 51 | 0.92 (0.44 - 1.87) | 0.82 | 0.94 (0.46 - 1.94) | 0.87 |
| ODOD | 7,366 | 12,560 | 4.2 | 7.7 | 26 | 53 | 0.81 (0.43 - 1.47) | 0.50 | 0.80 (0.43 - 1.47) | 0.48 |
| MDCR | 711 | 1,452 | 0.4 | 0.9 | <5 | 5 | 1.00 (0.04 - 25.3) | 1.00 | 0.87 (0.03 - 22.1) | 0.93 |
| MDCD | 1,435 | 2,340 | 0.8 | 1.3 | 6 | 12 | 0.95 (0.29 - 2.84) | 0.93 | 0.92 (0.29 - 2.89) | 0.87 |
| Open Claims | 167,178 | 270,181 | 113.0 | 200.6 | 324 | 602 | 0.99 (0.82 - 1.19) | 0.93 | 0.98 (0.81 - 1.18) | 0.80 |
| OEHR | 9,640 | 16,550 | 2.8 | 5.2 | 7 | 14 | 1.00 (0.32 - 2.87) | 0.99 | 0.91 (0.31 - 2.71) | 0.86 |
| Meta-analysis | 196,369 | 320,304 | 127.7 | 228.3 | 380 | 732 | 0.97 (0.82 - 1.15) | 0.74 | 0.96 (0.80 - 1.14) | 0.61 |
| Meta-analysis (w/o USOC) | 29,191 | 50,123 | 14.8 | 27.7 | 56 | 130 | 0.88 (0.59 - 1.32) | 0.55 | 0.88 (0.59 - 1.31) | 0.53 |
| **PS-stratification / on-treatment TAR** | | | | | | | | | | |
| CCAE | 14,188 | 18,673 | 8.7 | 13.5 | 22 | 51 | 0.64 (0.37 - 1.09) | 0.11 | 0.66 (0.38 - 1.13) | 0.13 |
| ODOD | 9,918 | 12,563 | 5.3 | 7.7 | 33 | 53 | 1.07 (0.66 - 1.70) | 0.79 | 1.05 (0.66 - 1.69) | 0.83 |
| MDCR | 791 | 1,453 | 0.5 | 0.9 | <5 | 5 | 0.69 (0.14 - 3.03) | 0.65 | 0.61 (0.13 - 2.89) | 0.54 |
| MDCD | 1,694 | 2,344 | 0.9 | 1.3 | 7 | 12 | 0.86 (0.31 - 2.23) | 0.77 | 0.88 (0.33 - 2.37) | 0.80 |
| Open Claims | 226,441 | 270,188 | 141.2 | 200.6 | 396 | 602 | 1.05 (0.92 - 1.21) | 0.47 | 1.03 (0.90 - 1.19) | 0.65 |
| OEHR | 12,803 | 16,552 | 3.7 | 5.2 | 9 | 14 | 0.83 (0.33 - 2.00) | 0.69 | 0.74 (0.30 - 1.80) | 0.50 |
| Meta-analysis | 265,044 | 320,320 | 159.8 | 228.3 | 467 | 732 | 0.99 (0.83 - 1.18) | 0.92 | 0.98 (0.82 - 1.17) | 0.80 |
| Meta-analysis (w/o USOC) | 38,603 | 50,132 | 18.6 | 27.7 | 71 | 130 | 0.86 (0.63 - 1.17) | 0.33 | 0.85 (0.62 - 1.17) | 0.32 |
| **PS-matching / on-treatment TAR without escalation** | | | | | | | | | | |
| CCAE | 10,750 | 18,673 | 6.3 | 11.7 | 13 | 41 | 1.01 (0.41 - 2.40) | 0.98 | 1.04 (0.43 - 2.52) | 0.93 |
| ODOD | 7,366 | 12,560 | 3.8 | 6.8 | 23 | 50 | 0.80 (0.42 - 1.48) | 0.49 | 0.78 (0.41 - 1.47) | 0.45 |
| MDCR | 711 | 1,452 | 0.4 | 0.8 | <5 | <5 | 1.00 (0.04 - 25.3) | 1.00 | 0.89 (0.04 - 22.5) | 0.93 |
| MDCD | 1,435 | 2,340 | 0.7 | 1.0 | 5 | 6 | 1.94 (0.50 - 8.01) | 0.35 | 1.92 (0.48 - 7.69) | 0.36 |
| Open Claims | 167,178 | 270,181 | 100.9 | 168.7 | 278 | 505 | 0.94 (0.76 - 1.15) | 0.54 | 0.91 (0.74 - 1.13) | 0.40 |
| OEHR | 9,640 | 16,550 | 2.4 | 4.5 | 5 | 9 | 1.19 (0.33 - 4.10) | 0.79 | 1.08 (0.31 - 3.80) | 0.91 |
| Meta-analysis | 185,294 | 301,414 | 111.0 | 187.2 | 314 | 596 | 0.93 (0.76 - 1.12) | 0.44 | 0.90 (0.74 - 1.10) | 0.33 |
| Meta-analysis (w/o USOC) | 18,116 | 31,233 | 10.1 | 18.5 | 36 | 91 | 0.87 (0.52 - 1.45) | 0.58 | 0.86 (0.51 - 1.44) | 0.57 |
| **PS-stratification / on-treatment TAR without escalation** | | | | | | | | | | |
| CCAE | 14,188 | 18,673 | 7.9 | 11.7 | 16 | 41 | 0.56 (0.29 - 1.02) | 0.07 | 0.58 (0.31 - 1.08) | 0.09 |
| ODOD | 9,918 | 12,563 | 4.8 | 6.8 | 29 | 50 | 0.97 (0.58 - 1.58) | 0.91 | 0.98 (0.59 - 1.61) | 0.92 |
| MDCR | 791 | 1,453 | 0.4 | 0.8 | <5 | <5 | 0.93 (0.17 - 4.52) | 0.93 | 0.84 (0.16 - 4.29) | 0.83 |
| MDCD | 1,694 | 2,344 | 0.8 | 1.0 | 6 | 6 | 1.21 (0.36 - 4.00) | 0.76 | 1.29 (0.39 - 4.27) | 0.69 |
| Open Claims | 226,441 | 270,188 | 127.4 | 168.7 | 348 | 505 | 1.04 (0.90 - 1.21) | 0.59 | 1.02 (0.88 - 1.18) | 0.82 |
| OEHR | 12,803 | 16,552 | 3.2 | 4.5 | 7 | 9 | 0.87 (0.29 - 2.45) | 0.79 | 0.78 (0.27 - 2.24) | 0.64 |
| Meta-analysis | 250,547 | 301,424 | 140.1 | 187.2 | 393 | 596 | 0.92 (0.68 - 1.24) | 0.58 | 0.91 (0.67 - 1.22) | 0.53 |
| Meta-analysis (w/o USOC) | 24,106 | 31,236 | 12.7 | 18.5 | 45 | 91 | 0.76 (0.44 - 1.31) | 0.32 | 0.77 (0.45 - 1.33) | 0.35 |

##### 1.1.9 Sudden cardiac death

**Supplementary Table 9:** Exposures, outcomes and risk estimates for Sudden cardiac death in GLP1RA (female) (T) vs SGLT2I (female) (C) new-user cohorts under four different propensity score (PS) adjustment / time-at-risk (TAR) designs across data sources. We report total patient exposure time in 1,000 patient-year (KYr) units, uncalibrated and calibrated hazard ratios (HRs) and their 95% confidence intervals (CIs). Greyed out entries did not pass study diagnostics and are included only for completeness.

| | Patients | | Exposure (KYr) | | Outcomes | | Uncalibrated | | Calibrated | |
|---|---|---|---|---|---|---|---|---|---|---|
| | T | C | T | C | T | C | HR (95% CI) | $p$ | HR (95% CI) | $p$ |
| **PS-matching / on-treatment TAR** | | | | | | | | | | |
| CCAE | 10,874 | 18,867 | 7.1 | 13.7 | 5 | 8 | 0.99 (0.28 - 3.36) | 0.98 | 1.01 (0.29 - 3.50) | 0.97 |
| ODOD | 7,487 | 12,824 | 4.2 | 7.9 | <5 | 6 | | 0.09 | | 0.10 |
| MDCR | 734 | 1,486 | 0.4 | 0.9 | 0 | 0 | | | | |
| MDCD | 1,485 | 2,428 | 0.8 | 1.3 | 0 | <5 | | | | |
| Open Claims | 170,550 | 276,019 | 115.2 | 204.6 | 56 | 121 | 0.94 (0.58 - 1.50) | 0.80 | 0.93 (0.58 - 1.49) | 0.75 |
| OEHR | 9,707 | 16,655 | 2.8 | 5.2 | <5 | <5 | 0.50 (0.02 - 5.22) | 0.62 | 0.46 (0.03 - 6.85) | 0.57 |
| Meta-analysis | 170,550 | 276,019 | 115.2 | 204.6 | 56 | 121 | 0.94 (0.58 - 1.50) | 0.80 | 0.93 (0.58 - 1.49) | 0.75 |
| **PS-stratification / on-treatment TAR** | | | | | | | | | | |
| CCAE | 14,348 | 18,867 | 8.8 | 13.7 | 5 | 8 | 0.78 (0.23 - 2.40) | 0.68 | 0.80 (0.25 - 2.57) | 0.71 |
| ODOD | 10,066 | 12,828 | 5.4 | 7.9 | 6 | 6 | 1.69 (0.49 - 5.79) | 0.41 | 1.67 (0.48 - 5.73) | 0.42 |
| MDCR | 815 | 1,487 | 0.5 | 0.9 | 0 | 0 | | | | |
| MDCD | 1,746 | 2,433 | 1.0 | 1.3 | 0 | <5 | | | | |
| Open Claims | 230,619 | 276,028 | 143.7 | 204.6 | 66 | 121 | 0.91 (0.65 - 1.25) | 0.56 | 0.89 (0.64 - 1.24) | 0.49 |
| OEHR | 12,892 | 16,657 | 3.7 | 5.2 | <5 | <5 | 0.72 (0.09 - 4.05) | 0.73 | 0.63 (0.10 - 4.18) | 0.64 |
| Meta-analysis | 230,619 | 276,028 | 143.7 | 204.6 | 66 | 121 | 0.91 (0.65 - 1.25) | 0.56 | 0.89 (0.65 - 1.24) | 0.50 |
| **PS-matching / on-treatment TAR without escalation** | | | | | | | | | | |
| CCAE | 10,874 | 18,867 | 6.4 | 11.8 | <5 | 7 | 0.98 (0.24 - 3.84) | 0.98 | 1.01 (0.25 - 4.07) | 0.97 |
| ODOD | 7,487 | 12,824 | 3.9 | 7.0 | <5 | 6 | | 0.09 | | 0.10 |
| MDCR | 734 | 1,486 | 0.4 | 0.8 | 0 | 0 | | | | |
| MDCD | 1,485 | 2,428 | 0.7 | 1.1 | 0 | <5 | | | | |
| Open Claims | 170,550 | 276,019 | 102.9 | 172.0 | 48 | 105 | 0.88 (0.53 - 1.45) | 0.63 | 0.86 (0.52 - 1.42) | 0.56 |
| OEHR | 9,707 | 16,655 | 2.4 | 4.5 | <5 | <5 | 0.50 (0.02 - 5.22) | 0.62 | 0.45 (0.03 - 6.81) | 0.57 |
| Meta-analysis | 170,550 | 276,019 | 102.9 | 172.0 | 48 | 105 | 0.88 (0.53 - 1.45) | 0.63 | 0.86 (0.52 - 1.43) | 0.57 |
| **PS-stratification / on-treatment TAR without escalation** | | | | | | | | | | |
| CCAE | 14,348 | 18,867 | 8.0 | 11.8 | <5 | 7 | 0.69 (0.18 - 2.37) | 0.58 | 0.72 (0.20 - 2.62) | 0.62 |
| ODOD | 10,066 | 12,828 | 4.9 | 7.0 | 6 | 6 | 1.66 (0.48 - 5.71) | 0.43 | 1.67 (0.48 - 5.78) | 0.42 |
| MDCR | 815 | 1,487 | 0.4 | 0.8 | 0 | 0 | | | | |
| MDCD | 1,746 | 2,433 | 0.8 | 1.1 | 0 | <5 | | | | |
| Open Claims | 230,619 | 276,028 | 129.7 | 172.0 | 57 | 105 | 0.85 (0.60 - 1.21) | 0.37 | 0.83 (0.58 - 1.19) | 0.31 |
| OEHR | 12,892 | 16,657 | 3.2 | 4.5 | <5 | <5 | 0.37 (0.02 - 2.87) | 0.44 | 0.33 (0.03 - 4.22) | 0.39 |
| Meta-analysis | 230,619 | 276,028 | 129.7 | 172.0 | 57 | 105 | 0.85 (0.60 - 1.21) | 0.37 | 0.84 (0.59 - 1.20) | 0.34 |

##### 1.1.10 Abnormal weight gain

**Supplementary Table 10:** Exposures, outcomes and risk estimates for Abnormal weight gain in GLP1RA (female) (T) vs SGLT2I (female) (C) new-user cohorts under four different propensity score (PS) adjustment / time-at-risk (TAR) designs across data sources. We report total patient exposure time in 1,000 patient-year (KYr) units, uncalibrated and calibrated hazard ratios (HRs) and their 95% confidence intervals (CIs). Greyed out entries did not pass study diagnostics and are included only for completeness.

| | Patients | | Exposure (KYr) | | Outcomes | | Uncalibrated | | Calibrated | |
|---|---|---|---|---|---|---|---|---|---|---|
| | T | C | T | C | T | C | HR (95% CI) | $p$ | HR (95% CI) | $p$ |
| **PS-matching / on-treatment TAR** | | | | | | | | | | |
| CCAE | 9,579 | 17,049 | 6.2 | 12.4 | 64 | 80 | 1.35 (0.83 - 2.18) | 0.22 | 1.38 (0.85 - 2.24) | 0.20 |
| ODOD | 6,779 | 11,831 | 3.8 | 7.2 | 38 | 54 | 0.95 (0.51 - 1.73) | 0.87 | 0.93 (0.51 - 1.73) | 0.83 |
| MDCR | 701 | 1,454 | 0.4 | 0.9 | <5 | <5 | 1.73 (0.06 - 46.9) | 0.74 | 1.51 (0.06 - 40.9) | 0.81 |
| MDCD | 1,271 | 2,172 | 0.7 | 1.2 | 8 | 5 | 3.00 (0.38 - 60.6) | 0.39 | 2.89 (0.23 - 36.3) | 0.41 |
| Open Claims | 154,383 | 255,338 | 103.9 | 188.9 | 379 | 486 | 1.25 (1.03 - 1.52) | 0.02 | 1.23 (1.01 - 1.50) | 0.04 |
| OEHR | 8,813 | 15,453 | 2.5 | 4.8 | 30 | 29 | 2.19 (1.02 - 4.91) | 0.05 | 2.00 (0.91 - 4.40) | 0.09 |
| Meta-analysis | 179,554 | 299,671 | 116.5 | 213.3 | 511 | 649 | 1.27 (1.07 - 1.51) | <0.01 | 1.25 (1.05 - 1.49) | 0.01 |
| Meta-analysis (w/o USOC) | 25,171 | 44,333 | 12.6 | 24.4 | 132 | 163 | 1.34 (0.89 - 2.01) | 0.16 | 1.33 (0.88 - 2.00) | 0.17 |
| **PS-stratification / on-treatment TAR** | | | | | | | | | | |
| CCAE | 12,191 | 17,049 | 7.5 | 12.4 | 85 | 80 | 1.39 (0.99 - 1.95) | 0.05 | 1.42 (1.01 - 2.00) | 0.04 |
| ODOD | 8,795 | 11,835 | 4.7 | 7.2 | 47 | 54 | 1.09 (0.71 - 1.67) | 0.71 | 1.07 (0.70 - 1.65) | 0.75 |
| MDCR | 769 | 1,455 | 0.4 | 0.9 | <5 | <5 | 2.96 (0.45 - 23.7) | 0.28 | 2.61 (0.36 - 18.8) | 0.34 |
| MDCD | 1,464 | 2,177 | 0.8 | 1.2 | 10 | 5 | 1.55 (0.54 - 5.07) | 0.45 | 1.58 (0.51 - 4.86) | 0.43 |
| Open Claims | 201,930 | 255,345 | 126.3 | 188.9 | 548 | 486 | 1.40 (1.23 - 1.61) | <0.01 | 1.38 (1.20 - 1.58) | <0.01 |
| OEHR | 11,328 | 15,455 | 3.2 | 4.8 | 45 | 29 | 1.63 (0.99 - 2.71) | 0.06 | 1.43 (0.87 - 2.38) | 0.17 |
| Meta-analysis | 234,244 | 299,684 | 141.7 | 213.3 | 725 | 649 | 1.39 (1.24 - 1.56) | <0.01 | 1.37 (1.21 - 1.55) | <0.01 |
| Meta-analysis (w/o USOC) | 32,314 | 44,339 | 15.4 | 24.4 | 177 | 163 | 1.34 (1.06 - 1.69) | 0.02 | 1.33 (1.05 - 1.69) | 0.02 |
| **PS-matching / on-treatment TAR without escalation** | | | | | | | | | | |
| CCAE | 9,579 | 17,049 | 5.6 | 10.7 | 60 | 68 | 1.40 (0.84 - 2.33) | 0.20 | 1.44 (0.86 - 2.40) | 0.17 |
| ODOD | 6,779 | 11,831 | 3.5 | 6.4 | 35 | 48 | 1.05 (0.54 - 2.00) | 0.88 | 1.03 (0.54 - 1.97) | 0.92 |
| MDCR | 701 | 1,454 | 0.4 | 0.8 | <5 | <5 | 1.73 (0.06 - 46.9) | 0.74 | 1.54 (0.06 - 41.7) | 0.80 |
| MDCD | 1,271 | 2,172 | 0.6 | 1.0 | 6 | <5 | 2.00 (0.19 - 43.0) | 0.62 | 1.98 (0.13 - 29.7) | 0.62 |
| Open Claims | 154,383 | 255,338 | 92.8 | 159.0 | 331 | 388 | 1.37 (1.11 - 1.70) | <0.01 | 1.34 (1.08 - 1.66) | <0.01 |
| OEHR | 8,813 | 15,453 | 2.2 | 4.2 | 24 | 24 | 3.29 (1.35 - 8.80) | 0.01 | 2.99 (1.17 - 7.63) | 0.02 |
| Meta-analysis | 179,554 | 299,671 | 104.1 | 180.2 | 450 | 528 | 1.39 (1.16 - 1.67) | <0.01 | 1.36 (1.13 - 1.65) | <0.01 |
| Meta-analysis (w/o USOC) | 25,171 | 44,333 | 11.3 | 21.2 | 119 | 140 | 1.54 (0.89 - 2.65) | 0.12 | 1.53 (0.89 - 2.63) | 0.13 |
| **PS-stratification / on-treatment TAR without escalation** | | | | | | | | | | |
| CCAE | 12,191 | 17,049 | 6.8 | 10.7 | 80 | 68 | 1.50 (1.05 - 2.15) | 0.03 | 1.55 (1.08 - 2.23) | 0.02 |
| ODOD | 8,795 | 11,835 | 4.3 | 6.4 | 44 | 48 | 1.11 (0.70 - 1.74) | 0.66 | 1.12 (0.71 - 1.76) | 0.64 |
| MDCR | 769 | 1,455 | 0.4 | 0.8 | <5 | <5 | 3.07 (0.48 - 24.4) | 0.26 | 2.76 (0.39 - 19.8) | 0.31 |
| MDCD | 1,464 | 2,177 | 0.7 | 1.0 | 8 | <5 | 1.37 (0.42 - 5.18) | 0.63 | 1.45 (0.42 - 5.09) | 0.56 |
| Open Claims | 201,930 | 255,345 | 113.7 | 159.0 | 498 | 388 | 1.49 (1.29 - 1.73) | <0.01 | 1.46 (1.26 - 1.69) | <0.01 |
| OEHR | 11,328 | 15,455 | 2.8 | 4.2 | 38 | 24 | 1.67 (0.97 - 2.94) | 0.07 | 1.50 (0.86 - 2.61) | 0.16 |
| Meta-analysis | 234,244 | 299,684 | 127.6 | 180.2 | 660 | 528 | 1.47 (1.29 - 1.67) | <0.01 | 1.45 (1.27 - 1.65) | <0.01 |
| Meta-analysis (w/o USOC) | 32,314 | 44,339 | 13.9 | 21.2 | 162 | 140 | 1.40 (1.08 - 1.79) | <0.01 | 1.42 (1.10 - 1.83) | <0.01 |

##### 1.1.11 Abnormal weight loss

**Supplementary Table 11:** Exposures, outcomes and risk estimates for Abnormal weight loss in GLP1RA (female) (T) vs SGLT2I (female) (C) new-user cohorts under four different propensity score (PS) adjustment / time-at-risk (TAR) designs across data sources. We report total patient exposure time in 1,000 patient-year (KYr) units, uncalibrated and calibrated hazard ratios (HRs) and their 95% confidence intervals (CIs). Greyed out entries did not pass study diagnostics and are included only for completeness.

| | Patients | | Exposure (KYr) | | Outcomes | | Uncalibrated | | Calibrated | |
|---|---|---|---|---|---|---|---|---|---|---|
| | T | C | T | C | T | C | HR (95% CI) | $p$ | HR (95% CI) | $p$ |
| **PS-matching / on-treatment TAR** | | | | | | | | | | |
| CCAE | 10,615 | 18,444 | 6.9 | 13.3 | 67 | 112 | 1.07 (0.71 - 1.61) | 0.75 | 1.10 (0.73 - 1.65) | 0.67 |
| ODOD | 7,288 | 12,376 | 4.1 | 7.5 | 68 | 114 | 1.07 (0.71 - 1.60) | 0.75 | 1.05 (0.70 - 1.58) | 0.81 |
| MDCR | 719 | 1,437 | 0.4 | 0.9 | 9 | 11 | 2.31 (0.55 - 11.6) | 0.28 | 2.02 (0.44 - 9.26) | 0.37 |
| MDCD | 1,418 | 2,325 | 0.8 | 1.2 | 12 | 24 | 1.15 (0.40 - 3.29) | 0.79 | 1.11 (0.38 - 3.20) | 0.85 |
| Open Claims | 165,133 | 266,944 | 111.3 | 197.5 | 581 | 1,078 | 1.03 (0.89 - 1.19) | 0.66 | 1.02 (0.88 - 1.18) | 0.83 |
| OEHR | 9,470 | 16,230 | 2.7 | 5.1 | 23 | 46 | 0.56 (0.26 - 1.10) | 0.11 | 0.51 (0.25 - 1.04) | 0.07 |
| Meta-analysis | 193,924 | 316,319 | 125.9 | 224.7 | 751 | 1,374 | 1.02 (0.90 - 1.16) | 0.74 | 1.01 (0.88 - 1.15) | 0.89 |
| Meta-analysis (w/o USOC) | 28,791 | 49,375 | 14.5 | 27.2 | 170 | 296 | 0.99 (0.76 - 1.28) | 0.92 | 0.98 (0.75 - 1.27) | 0.86 |
| **PS-stratification / on-treatment TAR** | | | | | | | | | | |
| CCAE | 13,986 | 18,444 | 8.5 | 13.3 | 89 | 112 | 1.09 (0.80 - 1.47) | 0.58 | 1.11 (0.82 - 1.51) | 0.49 |
| ODOD | 9,740 | 12,379 | 5.2 | 7.5 | 81 | 115 | 1.14 (0.83 - 1.55) | 0.41 | 1.12 (0.83 - 1.53) | 0.48 |
| MDCR | 800 | 1,438 | 0.5 | 0.9 | 9 | 11 | 1.51 (0.58 - 3.83) | 0.39 | 1.33 (0.52 - 3.42) | 0.56 |
| MDCD | 1,674 | 2,330 | 0.9 | 1.2 | 15 | 24 | 0.70 (0.35 - 1.36) | 0.31 | 0.72 (0.36 - 1.42) | 0.35 |
| Open Claims | 223,055 | 266,948 | 138.7 | 197.5 | 738 | 1,078 | 1.01 (0.92 - 1.12) | 0.80 | 1.00 (0.90 - 1.11) | 0.90 |
| OEHR | 12,562 | 16,231 | 3.6 | 5.1 | 32 | 46 | 0.95 (0.59 - 1.54) | 0.85 | 0.84 (0.52 - 1.36) | 0.50 |
| Meta-analysis | 261,017 | 316,332 | 157.0 | 224.7 | 955 | 1,375 | 1.02 (0.93 - 1.12) | 0.65 | 1.01 (0.91 - 1.11) | 0.87 |
| Meta-analysis (w/o USOC) | 37,962 | 49,384 | 18.2 | 27.2 | 217 | 297 | 1.05 (0.87 - 1.27) | 0.62 | 1.04 (0.86 - 1.26) | 0.67 |
| **PS-matching / on-treatment TAR without escalation** | | | | | | | | | | |
| CCAE | 10,615 | 18,444 | 6.2 | 11.5 | 56 | 93 | 1.01 (0.65 - 1.56) | 0.96 | 1.04 (0.67 - 1.62) | 0.86 |
| ODOD | 7,288 | 12,376 | 3.7 | 6.7 | 60 | 99 | 0.97 (0.62 - 1.49) | 0.88 | 0.94 (0.61 - 1.47) | 0.80 |
| MDCR | 719 | 1,437 | 0.4 | 0.8 | 9 | 10 | 2.31 (0.55 - 11.6) | 0.28 | 2.06 (0.45 - 9.43) | 0.36 |
| MDCD | 1,418 | 2,325 | 0.7 | 1.0 | 9 | 21 | 0.93 (0.23 - 3.38) | 0.92 | 0.92 (0.24 - 3.52) | 0.89 |
| Open Claims | 165,133 | 266,944 | 99.5 | 166.1 | 517 | 907 | 1.09 (0.93 - 1.27) | 0.29 | 1.06 (0.91 - 1.24) | 0.48 |
| OEHR | 9,470 | 16,230 | 2.4 | 4.4 | 19 | 39 | 0.57 (0.25 - 1.21) | 0.16 | 0.52 (0.23 - 1.15) | 0.11 |
| Meta-analysis | 193,924 | 316,319 | 112.5 | 189.8 | 661 | 1,159 | 1.05 (0.91 - 1.20) | 0.51 | 1.02 (0.89 - 1.18) | 0.77 |
| Meta-analysis (w/o USOC) | 28,791 | 49,375 | 13.0 | 23.6 | 144 | 252 | 0.92 (0.69 - 1.22) | 0.56 | 0.91 (0.69 - 1.21) | 0.54 |
| **PS-stratification / on-treatment TAR without escalation** | | | | | | | | | | |
| CCAE | 13,986 | 18,444 | 7.8 | 11.5 | 74 | 93 | 1.07 (0.76 - 1.48) | 0.70 | 1.11 (0.79 - 1.55) | 0.56 |
| ODOD | 9,740 | 12,379 | 4.7 | 6.7 | 72 | 100 | 1.19 (0.85 - 1.65) | 0.31 | 1.20 (0.86 - 1.67) | 0.30 |
| MDCR | 800 | 1,438 | 0.4 | 0.8 | 9 | 10 | 1.54 (0.58 - 4.00) | 0.38 | 1.38 (0.53 - 3.62) | 0.52 |
| MDCD | 1,674 | 2,330 | 0.8 | 1.0 | 10 | 21 | 0.54 (0.24 - 1.16) | 0.13 | 0.57 (0.26 - 1.27) | 0.18 |
| Open Claims | 223,055 | 266,948 | 125.2 | 166.1 | 666 | 907 | 1.03 (0.92 - 1.14) | 0.64 | 1.00 (0.90 - 1.12) | 0.91 |
| OEHR | 12,562 | 16,231 | 3.1 | 4.4 | 27 | 39 | 0.99 (0.58 - 1.67) | 0.98 | 0.89 (0.53 - 1.51) | 0.67 |
| Meta-analysis | 261,017 | 316,332 | 141.7 | 189.8 | 849 | 1,160 | 1.03 (0.94 - 1.14) | 0.53 | 1.02 (0.92 - 1.13) | 0.76 |
| Meta-analysis (w/o USOC) | 37,962 | 49,384 | 16.4 | 23.6 | 183 | 253 | 1.04 (0.84 - 1.30) | 0.71 | 1.06 (0.85 - 1.32) | 0.61 |

##### 1.1.12 Acute pancreatitis

**Supplementary Table 12:** Exposures, outcomes and risk estimates for Acute pancreatitis in GLP1RA (female) (T) vs SGLT2I (female) (C) new-user cohorts under four different propensity score (PS) adjustment / time-at-risk (TAR) designs across data sources. We report total patient exposure time in 1,000 patient-year (KYr) units, uncalibrated and calibrated hazard ratios (HRs) and their 95% confidence intervals (CIs). Greyed out entries did not pass study diagnostics and are included only for completeness.

| | Patients | | Exposure (KYr) | | Outcomes | | Uncalibrated | | Calibrated | |
|---|---|---|---|---|---|---|---|---|---|---|
| | T | C | T | C | T | C | HR (95% CI) | $p$ | HR (95% CI) | $p$ |
| **PS-matching / on-treatment TAR** | | | | | | | | | | |
| CCAE | 10,791 | 18,710 | 7.1 | 13.6 | 10 | 22 | 0.74 (0.25 - 1.92) | 0.56 | 0.75 (0.27 - 2.08) | 0.59 |
| ODOD | 7,436 | 12,737 | 4.2 | 7.8 | 13 | 17 | 1.07 (0.38 - 2.88) | 0.89 | 1.05 (0.38 - 2.90) | 0.92 |
| MDCR | 730 | 1,472 | 0.4 | 0.9 | <5 | <5 | | | | |
| MDCD | 1,464 | 2,385 | 0.8 | 1.3 | <5 | 9 | 0.31 (0.02 - 1.94) | 0.33 | 0.29 (0.03 - 3.26) | 0.32 |
| Open Claims | 168,780 | 272,682 | 114.1 | 202.2 | 177 | 275 | 1.28 (0.97 - 1.67) | 0.08 | 1.25 (0.96 - 1.65) | 0.11 |
| OEHR | 9,670 | 16,580 | 2.8 | 5.2 | 9 | 9 | 0.98 (0.19 - 4.64) | 0.98 | 0.90 (0.18 - 4.45) | 0.89 |
| Meta-analysis | 196,677 | 320,709 | 128.2 | 228.8 | 209 | 323 | 1.21 (0.94 - 1.56) | 0.13 | 1.19 (0.93 - 1.54) | 0.17 |
| Meta-analysis (w/o USOC) | 27,897 | 48,027 | 14.1 | 26.6 | 32 | 48 | 0.90 (0.47 - 1.74) | 0.76 | 0.90 (0.47 - 1.73) | 0.74 |
| **PS-stratification / on-treatment TAR** | | | | | | | | | | |
| CCAE | 14,283 | 18,712 | 8.8 | 13.6 | 11 | 22 | 1.00 (0.45 - 2.12) | 1.00 | 1.02 (0.47 - 2.23) | 0.95 |
| ODOD | 10,013 | 12,741 | 5.3 | 7.8 | 16 | 17 | 1.41 (0.67 - 2.93) | 0.36 | 1.39 (0.67 - 2.90) | 0.38 |
| MDCR | 816 | 1,473 | 0.5 | 0.9 | <5 | <5 | 3.17 (0.43 - 27.7) | 0.28 | 2.78 (0.35 - 22.4) | 0.34 |
| MDCD | 1,724 | 2,390 | 1.0 | 1.3 | <5 | 9 | 0.37 (0.08 - 1.37) | 0.18 | 0.38 (0.09 - 1.61) | 0.19 |
| Open Claims | 228,642 | 272,688 | 142.6 | 202.2 | 216 | 275 | 1.21 (1.00 - 1.47) | 0.05 | 1.19 (0.98 - 1.45) | 0.09 |
| OEHR | 12,842 | 16,582 | 3.7 | 5.2 | 10 | 9 | 1.48 (0.57 - 3.88) | 0.42 | 1.31 (0.50 - 3.40) | 0.59 |
| Meta-analysis | 265,780 | 320,723 | 160.4 | 228.8 | 253 | 323 | 1.22 (1.02 - 1.46) | 0.03 | 1.20 (1.00 - 1.44) | 0.05 |
| Meta-analysis (w/o USOC) | 37,138 | 48,035 | 17.8 | 26.6 | 37 | 48 | 1.26 (0.79 - 2.01) | 0.33 | 1.25 (0.79 - 2.00) | 0.34 |
| **PS-matching / on-treatment TAR without escalation** | | | | | | | | | | |
| CCAE | 10,791 | 18,710 | 6.4 | 11.7 | 8 | 17 | 0.78 (0.20 - 2.59) | 0.70 | 0.80 (0.22 - 2.87) | 0.73 |
| ODOD | 7,436 | 12,737 | 3.9 | 6.9 | 12 | 13 | 1.08 (0.35 - 3.11) | 0.89 | 1.06 (0.36 - 3.13) | 0.92 |
| MDCR | 730 | 1,472 | 0.4 | 0.8 | <5 | <5 | | | | |
| MDCD | 1,464 | 2,385 | 0.7 | 1.1 | <5 | 8 | | | | |
| Open Claims | 168,780 | 272,682 | 101.9 | 170.1 | 156 | 211 | 1.42 (1.05 - 1.91) | 0.02 | 1.38 (1.03 - 1.86) | 0.03 |
| OEHR | 9,670 | 16,580 | 2.4 | 4.5 | 8 | 7 | 0.81 (0.10 - 5.07) | 0.83 | 0.73 (0.11 - 5.12) | 0.76 |
| Meta-analysis | 187,007 | 304,129 | 112.1 | 188.7 | 176 | 241 | 1.35 (1.02 - 1.79) | 0.03 | 1.32 (1.00 - 1.75) | 0.05 |
| Meta-analysis (w/o USOC) | 18,227 | 31,447 | 10.2 | 18.6 | 20 | 30 | 0.94 (0.41 - 2.16) | 0.89 | 0.94 (0.41 - 2.14) | 0.87 |
| **PS-stratification / on-treatment TAR without escalation** | | | | | | | | | | |
| CCAE | 14,283 | 18,712 | 8.0 | 11.7 | 9 | 17 | 1.21 (0.49 - 2.79) | 0.68 | 1.25 (0.52 - 3.00) | 0.62 |
| ODOD | 10,013 | 12,741 | 4.9 | 6.9 | 14 | 13 | 1.57 (0.70 - 3.54) | 0.27 | 1.59 (0.71 - 3.56) | 0.27 |
| MDCR | 816 | 1,473 | 0.4 | 0.8 | <5 | <5 | 3.16 (0.42 - 27.7) | 0.28 | 2.84 (0.35 - 23.0) | 0.33 |
| MDCD | 1,724 | 2,390 | 0.8 | 1.1 | <5 | 8 | 0.14 (0.01 - 0.83) | 0.10 | 0.15 (0.01 - 1.58) | 0.11 |
| Open Claims | 228,642 | 272,688 | 128.7 | 170.1 | 192 | 211 | 1.41 (1.14 - 1.75) | <0.01 | 1.38 (1.11 - 1.71) | <0.01 |
| OEHR | 12,842 | 16,582 | 3.2 | 4.5 | 9 | 7 | 1.64 (0.58 - 4.84) | 0.36 | 1.47 (0.51 - 4.25) | 0.48 |
| Meta-analysis | 252,938 | 304,141 | 141.5 | 188.7 | 215 | 241 | 1.41 (1.15 - 1.72) | <0.01 | 1.39 (1.13 - 1.70) | <0.01 |
| Meta-analysis (w/o USOC) | 24,296 | 31,453 | 12.9 | 18.6 | 23 | 30 | 1.39 (0.77 - 2.52) | 0.28 | 1.41 (0.78 - 2.56) | 0.26 |

##### 1.1.13 Bone fracture

**Supplementary Table 13:** Exposures, outcomes and risk estimates for Bone fracture in GLP1RA (female) (T) vs SGLT2I (female) (C) new-user cohorts under four different propensity score (PS) adjustment / time-at-risk (TAR) designs across data sources. We report total patient exposure time in 1,000 patient-year (KYr) units, uncalibrated and calibrated hazard ratios (HRs) and their 95% confidence intervals (CIs). Greyed out entries did not pass study diagnostics and are included only for completeness.

| | Patients | | Exposure (KYr) | | Outcomes | | Uncalibrated | | Calibrated | |
|---|---|---|---|---|---|---|---|---|---|---|
| | T | C | T | C | T | C | HR (95% CI) | $p$ | HR (95% CI) | $p$ |
| **PS-matching / on-treatment TAR** | | | | | | | | | | |
| CCAE | 9,687 | 16,973 | 6.2 | 12.0 | 117 | 230 | 0.94 (0.68 - 1.29) | 0.69 | 0.96 (0.69 - 1.33) | 0.80 |
| ODOD | 6,688 | 11,475 | 3.7 | 6.9 | 114 | 222 | 1.07 (0.76 - 1.48) | 0.70 | 1.05 (0.75 - 1.46) | 0.79 |
| MDCR | 619 | 1,279 | 0.4 | 0.8 | 9 | 35 | 0.25 (0.06 - 0.77) | 0.04 | 0.22 (0.06 - 0.80) | 0.02 |
| MDCD | 1,275 | 2,097 | 0.7 | 1.1 | 26 | 33 | 1.21 (0.59 - 2.47) | 0.60 | 1.16 (0.57 - 2.39) | 0.70 |
| Open Claims | 144,944 | 237,925 | 96.8 | 174.9 | 1,400 | 2,816 | 0.89 (0.81 - 0.98) | 0.01 | 0.87 (0.79 - 0.96) | 0.01 |
| OEHR | 8,944 | 15,398 | 2.6 | 4.8 | 34 | 69 | 1.18 (0.67 - 2.05) | 0.56 | 1.08 (0.62 - 1.88) | 0.80 |
| Meta-analysis | 171,538 | 283,868 | 110.0 | 199.8 | 1,691 | 3,370 | 0.92 (0.83 - 1.02) | 0.12 | 0.91 (0.81 - 1.01) | 0.09 |
| Meta-analysis (w/o USOC) | 26,594 | 45,943 | 13.2 | 24.9 | 291 | 554 | 1.04 (0.85 - 1.27) | 0.73 | 1.03 (0.84 - 1.27) | 0.78 |
| **PS-stratification / on-treatment TAR** | | | | | | | | | | |
| CCAE | 12,748 | 16,973 | 7.7 | 12.0 | 150 | 230 | 0.96 (0.77 - 1.21) | 0.75 | 0.99 (0.79 - 1.24) | 0.87 |
| ODOD | 8,968 | 11,478 | 4.7 | 6.9 | 139 | 222 | 0.93 (0.74 - 1.18) | 0.57 | 0.92 (0.73 - 1.17) | 0.54 |
| MDCR | 684 | 1,280 | 0.4 | 0.8 | 10 | 35 | 0.46 (0.21 - 0.93) | 0.04 | 0.41 (0.19 - 0.86) | 0.02 |
| MDCD | 1,490 | 2,102 | 0.8 | 1.1 | 30 | 33 | 1.20 (0.70 - 2.02) | 0.50 | 1.22 (0.72 - 2.07) | 0.47 |
| Open Claims | 194,529 | 237,944 | 120.1 | 175.0 | 1,753 | 2,817 | 0.93 (0.87 - 0.99) | 0.03 | 0.91 (0.85 - 0.98) | 0.02 |
| OEHR | 11,791 | 15,400 | 3.4 | 4.8 | 44 | 69 | 0.92 (0.61 - 1.38) | 0.69 | 0.81 (0.54 - 1.22) | 0.34 |
| Meta-analysis | 229,526 | 283,897 | 136.7 | 199.8 | 2,116 | 3,371 | 0.94 (0.88 - 0.99) | 0.03 | 0.92 (0.86 - 0.99) | 0.03 |
| Meta-analysis (w/o USOC) | 34,997 | 45,953 | 16.6 | 24.9 | 363 | 554 | 0.96 (0.83 - 1.11) | 0.61 | 0.96 (0.83 - 1.11) | 0.58 |
| **PS-matching / on-treatment TAR without escalation** | | | | | | | | | | |
| CCAE | 9,687 | 16,973 | 5.6 | 10.4 | 104 | 196 | 0.99 (0.70 - 1.40) | 0.97 | 1.02 (0.72 - 1.45) | 0.87 |
| ODOD | 6,688 | 11,475 | 3.4 | 6.1 | 98 | 192 | 1.14 (0.79 - 1.63) | 0.47 | 1.11 (0.78 - 1.59) | 0.58 |
| MDCR | 619 | 1,279 | 0.4 | 0.7 | 9 | 30 | 0.28 (0.06 - 0.86) | 0.05 | 0.25 (0.07 - 0.91) | 0.04 |
| MDCD | 1,275 | 2,097 | 0.6 | 0.9 | 24 | 28 | 1.00 (0.44 - 2.21) | 0.99 | 0.99 (0.44 - 2.23) | 0.87 |
| Open Claims | 144,944 | 237,925 | 86.6 | 147.3 | 1,217 | 2,345 | 0.89 (0.81 - 0.99) | 0.03 | 0.87 (0.78 - 0.97) | 0.02 |
| OEHR | 8,944 | 15,398 | 2.2 | 4.2 | 30 | 59 | 1.36 (0.74 - 2.47) | 0.31 | 1.24 (0.68 - 2.25) | 0.50 |
| Meta-analysis | 171,538 | 283,868 | 98.5 | 168.9 | 1,473 | 2,820 | 0.97 (0.83 - 1.12) | 0.67 | 0.95 (0.81 - 1.10) | 0.49 |
| Meta-analysis (w/o USOC) | 26,594 | 45,943 | 11.8 | 21.6 | 256 | 475 | 1.09 (0.88 - 1.36) | 0.43 | 1.09 (0.87 - 1.36) | 0.49 |
| **PS-stratification / on-treatment TAR without escalation** | | | | | | | | | | |
| CCAE | 12,748 | 16,973 | 7.0 | 10.4 | 135 | 196 | 0.99 (0.78 - 1.26) | 0.93 | 1.03 (0.81 - 1.31) | 0.83 |
| ODOD | 8,968 | 11,478 | 4.3 | 6.2 | 122 | 192 | 0.92 (0.71 - 1.18) | 0.50 | 0.92 (0.72 - 1.19) | 0.56 |
| MDCR | 684 | 1,280 | 0.4 | 0.7 | 10 | 30 | 0.50 (0.22 - 1.02) | 0.07 | 0.45 (0.21 - 0.95) | 0.04 |
| MDCD | 1,490 | 2,102 | 0.7 | 0.9 | 27 | 28 | 1.28 (0.73 - 2.25) | 0.39 | 1.36 (0.77 - 2.40) | 0.30 |
| Open Claims | 194,529 | 237,944 | 108.5 | 147.3 | 1,546 | 2,346 | 0.92 (0.86 - 0.99) | 0.02 | 0.90 (0.83 - 0.97) | 0.01 |
| OEHR | 11,791 | 15,400 | 2.9 | 4.2 | 40 | 59 | 0.93 (0.60 - 1.43) | 0.74 | 0.83 (0.54 - 1.29) | 0.42 |
| Meta-analysis | 229,526 | 283,897 | 123.4 | 168.9 | 1,870 | 2,821 | 0.93 (0.87 - 0.99) | 0.02 | 0.92 (0.85 - 0.99) | 0.02 |
| Meta-analysis (w/o USOC) | 34,997 | 45,953 | 14.9 | 21.6 | 324 | 475 | 0.97 (0.83 - 1.14) | 0.72 | 0.99 (0.84 - 1.16) | 0.84 |

###### 1.1.14 Diabetic ketoacidosis

**Supplementary Table 14:** Exposures, outcomes and risk estimates for Diabetic ketoacidosis in GLP1RA (female) (T) vs SGLT2I (female) (C) new-user cohorts under four different propensity score (PS) adjustment / time-at-risk (TAR) designs across data sources. We report total patient exposure time in 1,000 patient-year (KYr) units, uncalibrated and calibrated hazard ratios (HRs) and their 95% confidence intervals (CIs). Greyed out entries did not pass study diagnostics and are included only for completeness.

| | Patients | | Exposure (KYr) | | Outcomes | | Uncalibrated | | Calibrated | |
|---|---|---|---|---|---|---|---|---|---|---|
| | T | C | T | C | T | C | HR (95% CI) | $p$ | HR (95% CI) | $p$ |
| **PS-matching / on-treatment TAR** | | | | | | | | | | |
| CCAE | 10,874 | 18,865 | 7.1 | 13.7 | 5 | 31 | 0.20 (0.03 - 0.73) | 0.05 | 0.21 (0.04 - 1.00) | 0.05 |
| ODOD | 7,491 | 12,829 | 4.2 | 7.9 | <5 | 24 | 0.20 (0.03 - 0.75) | 0.05 | 0.20 (0.04 - 0.98) | 0.05 |
| MDCR | 735 | 1,485 | 0.4 | 0.9 | 0 | <5 | | | | |
| MDCD | 1,478 | 2,422 | 0.8 | 1.3 | <5 | 5 | 0.39 (0.02 - 3.08) | 0.47 | 0.37 (0.03 - 4.75) | 0.45 |
| Open Claims | 170,551 | 276,150 | 115.2 | 204.7 | 61 | 254 | 0.41 (0.29 - 0.58) | <0.01 | 0.41 (0.29 - 0.58) | <0.01 |
| OEHR | 9,710 | 16,655 | 2.8 | 5.2 | 5 | 11 | 1.68 (0.45 - 6.34) | 0.44 | 1.54 (0.41 - 5.78) | 0.53 |
| Meta-analysis | 188,916 | 307,844 | 126.5 | 226.2 | <71 | 309 | 0.38 (0.27 - 0.55) | <0.01 | 0.38 (0.26 - 0.55) | <0.01 |
| Meta-analysis (w/o USOC) | 18,365 | 31,694 | 11.3 | 21.5 | <10 | 55 | 0.20 (0.07 - 0.62) | <0.01 | 0.20 (0.07 - 0.62) | <0.01 |
| **PS-stratification / on-treatment TAR** | | | | | | | | | | |
| CCAE | 14,354 | 18,865 | 8.8 | 13.7 | 5 | 31 | 0.27 (0.09 - 0.65) | <0.01 | 0.28 (0.10 - 0.74) | 0.01 |
| ODOD | 10,062 | 12,833 | 5.4 | 7.9 | 6 | 24 | 0.29 (0.10 - 0.69) | 0.01 | 0.28 (0.11 - 0.74) | 0.01 |
| MDCR | 819 | 1,486 | 0.5 | 0.9 | 0 | <5 | | | | |
| MDCD | 1,739 | 2,427 | 1.0 | 1.3 | <5 | 5 | 0.20 (0.01 - 1.29) | 0.19 | 0.21 (0.02 - 2.30) | 0.20 |
| Open Claims | 230,627 | 276,157 | 143.7 | 204.7 | 78 | 254 | 0.41 (0.31 - 0.53) | <0.01 | 0.40 (0.30 - 0.52) | <0.01 |
| OEHR | 12,891 | 16,657 | 3.7 | 5.2 | 8 | 11 | 1.06 (0.38 - 2.79) | 0.91 | 0.93 (0.35 - 2.52) | 0.89 |
| Meta-analysis | 267,934 | 324,512 | 161.7 | 231.4 | 97 | 320 | 0.41 (0.32 - 0.52) | <0.01 | 0.40 (0.32 - 0.52) | <0.01 |
| Meta-analysis (w/o USOC) | 37,307 | 48,355 | 17.9 | 26.8 | 19 | 66 | 0.43 (0.18 - 1.03) | 0.06 | 0.43 (0.18 - 1.02) | 0.06 |
| **PS-matching / on-treatment TAR without escalation** | | | | | | | | | | |
| CCAE | 10,874 | 18,865 | 6.4 | 11.8 | <5 | 24 | 0.20 (0.03 - 0.73) | 0.05 | 0.21 (0.04 - 1.00) | 0.05 |
| ODOD | 7,491 | 12,829 | 3.9 | 7.0 | <5 | 20 | 0.22 (0.03 - 0.80) | 0.06 | 0.21 (0.04 - 1.03) | 0.06 |
| MDCR | 735 | 1,485 | 0.4 | 0.8 | 0 | <5 | | | | |
| MDCD | 1,478 | 2,422 | 0.7 | 1.1 | <5 | <5 | 0.62 (0.03 - 6.63) | 0.73 | 0.61 (0.04 - 9.37) | 0.73 |
| Open Claims | 170,551 | 276,150 | 102.9 | 172.1 | 52 | 200 | 0.41 (0.28 - 0.59) | <0.01 | 0.40 (0.28 - 0.58) | <0.01 |
| OEHR | 9,710 | 16,655 | 2.4 | 4.6 | <5 | 7 | 1.40 (0.17 - 9.13) | 0.74 | 1.27 (0.17 - 9.26) | 0.82 |
| Meta-analysis | 188,916 | 307,844 | 113.2 | 190.8 | <62 | 244 | 0.38 (0.27 - 0.54) | <0.01 | 0.38 (0.26 - 0.53) | <0.01 |
| Meta-analysis (w/o USOC) | 18,365 | 31,694 | 10.3 | 18.8 | <10 | 44 | 0.21 (0.07 - 0.64) | <0.01 | 0.21 (0.07 - 0.64) | <0.01 |
| **PS-stratification / on-treatment TAR without escalation** | | | | | | | | | | |
| CCAE | 14,354 | 18,865 | 8.0 | 11.8 | <5 | 24 | 0.21 (0.05 - 0.60) | 0.01 | 0.21 (0.06 - 0.75) | 0.02 |
| ODOD | 10,062 | 12,833 | 4.9 | 7.0 | 5 | 20 | 0.27 (0.09 - 0.71) | 0.02 | 0.27 (0.09 - 0.78) | 0.02 |
| MDCR | 819 | 1,486 | 0.4 | 0.8 | 0 | <5 | | | | |
| MDCD | 1,739 | 2,427 | 0.8 | 1.1 | <5 | <5 | 0.43 (0.02 - 3.43) | 0.52 | 0.46 (0.04 - 5.85) | 0.55 |
| Open Claims | 230,627 | 276,157 | 129.7 | 172.1 | 67 | 200 | 0.41 (0.30 - 0.54) | <0.01 | 0.40 (0.30 - 0.53) | <0.01 |
| OEHR | 12,891 | 16,657 | 3.2 | 4.6 | <5 | 7 | 0.78 (0.18 - 2.99) | 0.73 | 0.70 (0.17 - 2.88) | 0.62 |
| Meta-analysis | 255,043 | 307,855 | 142.6 | 190.8 | <77 | 244 | 0.38 (0.28 - 0.51) | <0.01 | 0.37 (0.28 - 0.50) | <0.01 |
| Meta-analysis (w/o USOC) | 24,416 | 31,698 | 12.9 | 18.8 | <10 | 44 | 0.24 (0.11 - 0.54) | <0.01 | 0.24 (0.11 - 0.55) | <0.01 |

##### 1.1.15 Diarrhea

**Supplementary Table 15:** Exposures, outcomes and risk estimates for Diarrhea in GLP1RA (female) (T) vs SGLT2I (female) (C) new-user cohorts under four different propensity score (PS) adjustment / time-at-risk (TAR) designs across data sources. We report total patient exposure time in 1,000 patient-year (KYr) units, uncalibrated and calibrated hazard ratios (HRs) and their 95% confidence intervals (CIs). Greyed out entries did not pass study diagnostics and are included only for completeness.

| | Patients | | Exposure (KYr) | | Outcomes | | Uncalibrated | | Calibrated | |
|---|---|---|---|---|---|---|---|---|---|---|
| | T | C | T | C | T | C | HR (95% CI) | $p$ | HR (95% CI) | $p$ |
| **PS-matching / on-treatment TAR** | | | | | | | | | | |
| CCAE | 9,039 | 15,863 | 5.7 | 11.4 | 285 | 371 | 1.32 (1.06 - 1.65) | 0.01 | 1.35 (1.08 - 1.69) | 0.01 |
| ODOD | 6,071 | 10,571 | 3.4 | 6.4 | 181 | 261 | 1.54 (1.17 - 2.02) | <0.01 | 1.51 (1.15 - 1.99) | <0.01 |
| MDCR | 608 | 1,227 | 0.4 | 0.8 | 10 | 29 | 1.06 (0.26 - 3.87) | 0.93 | 0.93 (0.24 - 3.56) | 0.88 |
| MDCD | 1,098 | 1,789 | 0.6 | 0.9 | 49 | 57 | 1.69 (0.94 - 3.05) | 0.08 | 1.63 (0.90 - 2.93) | 0.12 |
| Open Claims | 139,745 | 230,065 | 93.2 | 169.4 | 2,342 | 3,165 | 1.41 (1.30 - 1.52) | <0.01 | 1.38 (1.28 - 1.50) | <0.01 |
| OEHR | 8,173 | 14,216 | 2.3 | 4.4 | 140 | 177 | 1.81 (1.33 - 2.46) | <0.01 | 1.65 (1.21 - 2.25) | <0.01 |
| Meta-analysis | 164,126 | 272,504 | 105.2 | 192.4 | 2,997 | 4,031 | 1.43 (1.33 - 1.53) | <0.01 | 1.40 (1.30 - 1.52) | <0.01 |
| Meta-analysis (w/o USOC) | 24,381 | 42,439 | 11.9 | 23.0 | 655 | 866 | 1.50 (1.30 - 1.74) | <0.01 | 1.49 (1.28 - 1.73) | <0.01 |
| **PS-stratification / on-treatment TAR** | | | | | | | | | | |
| CCAE | 11,815 | 15,864 | 7.0 | 11.4 | 360 | 371 | 1.40 (1.19 - 1.64) | <0.01 | 1.43 (1.21 - 1.68) | <0.01 |
| ODOD | 8,096 | 10,574 | 4.2 | 6.4 | 244 | 262 | 1.28 (1.05 - 1.55) | 0.01 | 1.26 (1.04 - 1.53) | 0.03 |
| MDCR | 680 | 1,228 | 0.4 | 0.8 | 12 | 29 | 0.68 (0.32 - 1.37) | 0.30 | 0.60 (0.29 - 1.24) | 0.17 |
| MDCD | 1,299 | 1,793 | 0.7 | 0.9 | 64 | 57 | 1.34 (0.92 - 1.96) | 0.13 | 1.37 (0.93 - 2.00) | 0.12 |
| Open Claims | 186,585 | 230,071 | 115.0 | 169.4 | 3,055 | 3,165 | 1.34 (1.27 - 1.42) | <0.01 | 1.32 (1.24 - 1.41) | <0.01 |
| OEHR | 10,772 | 14,218 | 3.0 | 4.4 | 190 | 177 | 1.51 (1.21 - 1.88) | <0.01 | 1.33 (1.06 - 1.66) | 0.02 |
| Meta-analysis | 218,567 | 272,520 | 129.9 | 192.5 | 3,913 | 4,032 | 1.35 (1.29 - 1.42) | <0.01 | 1.33 (1.25 - 1.42) | <0.01 |
| Meta-analysis (w/o USOC) | 31,982 | 42,449 | 14.9 | 23.0 | 858 | 867 | 1.38 (1.24 - 1.53) | <0.01 | 1.37 (1.23 - 1.53) | <0.01 |
| **PS-matching / on-treatment TAR without escalation** | | | | | | | | | | |
| CCAE | 9,039 | 15,863 | 5.2 | 9.9 | 255 | 322 | 1.37 (1.08 - 1.73) | <0.01 | 1.41 (1.11 - 1.78) | <0.01 |
| ODOD | 6,071 | 10,571 | 3.1 | 5.7 | 171 | 233 | 1.57 (1.18 - 2.08) | <0.01 | 1.53 (1.15 - 2.04) | <0.01 |
| MDCR | 608 | 1,227 | 0.3 | 0.7 | 9 | 26 | 1.37 (0.33 - 5.39) | 0.66 | 1.21 (0.30 - 4.94) | 0.79 |
| MDCD | 1,098 | 1,789 | 0.5 | 0.8 | 43 | 47 | 1.62 (0.85 - 3.08) | 0.14 | 1.60 (0.84 - 3.06) | 0.17 |
| Open Claims | 139,745 | 230,065 | 83.6 | 143.4 | 2,096 | 2,586 | 1.48 (1.36 - 1.61) | <0.01 | 1.44 (1.32 - 1.57) | <0.01 |
| OEHR | 8,173 | 14,216 | 2.0 | 3.8 | 126 | 147 | 1.92 (1.38 - 2.69) | <0.01 | 1.75 (1.25 - 2.44) | <0.01 |
| Meta-analysis | 164,126 | 272,504 | 94.4 | 163.6 | 2,691 | 3,335 | 1.49 (1.39 - 1.61) | <0.01 | 1.46 (1.34 - 1.59) | <0.01 |
| Meta-analysis (w/o USOC) | 24,381 | 42,439 | 10.8 | 20.2 | 595 | 749 | 1.55 (1.32 - 1.80) | <0.01 | 1.54 (1.31 - 1.80) | <0.01 |
| **PS-stratification / on-treatment TAR without escalation** | | | | | | | | | | |
| CCAE | 11,815 | 15,864 | 6.4 | 9.9 | 325 | 322 | 1.45 (1.22 - 1.71) | <0.01 | 1.50 (1.27 - 1.78) | <0.01 |
| ODOD | 8,096 | 10,574 | 3.9 | 5.7 | 232 | 234 | 1.34 (1.10 - 1.64) | <0.01 | 1.35 (1.11 - 1.66) | <0.01 |
| MDCR | 680 | 1,228 | 0.4 | 0.7 | 10 | 26 | 0.61 (0.27 - 1.29) | 0.22 | 0.55 (0.25 - 1.20) | 0.14 |
| MDCD | 1,299 | 1,793 | 0.6 | 0.8 | 56 | 47 | 1.41 (0.94 - 2.14) | 0.10 | 1.50 (0.99 - 2.28) | 0.07 |
| Open Claims | 186,585 | 230,071 | 104.0 | 143.4 | 2,764 | 2,586 | 1.41 (1.33 - 1.50) | <0.01 | 1.38 (1.29 - 1.47) | <0.01 |
| OEHR | 10,772 | 14,218 | 2.6 | 3.8 | 171 | 147 | 1.66 (1.31 - 2.11) | <0.01 | 1.49 (1.17 - 1.89) | <0.01 |
| Meta-analysis | 218,567 | 272,520 | 117.5 | 163.6 | 3,548 | 3,336 | 1.42 (1.35 - 1.50) | <0.01 | 1.40 (1.31 - 1.49) | <0.01 |
| Meta-analysis (w/o USOC) | 31,982 | 42,449 | 13.5 | 20.2 | 784 | 750 | 1.46 (1.30 - 1.62) | <0.01 | 1.48 (1.32 - 1.66) | <0.01 |

##### 1.1.16 Genitourinary infection

**Supplementary Table 16:** Exposures, outcomes and risk estimates for Genitourinary infection in GLP1RA (female) (T) vs SGLT2I (female) (C) new-user cohorts under four different propensity score (PS) adjustment / time-at-risk (TAR) designs across data sources. We report total patient exposure time in 1,000 patient-year (KYr) units, uncalibrated and calibrated hazard ratios (HRs) and their 95% confidence intervals (CIs). Greyed out entries did not pass study diagnostics and are included only for completeness.

| | Patients | | Exposure (KYr) | | Outcomes | | Uncalibrated | | Calibrated | |
|---|---|---|---|---|---|---|---|---|---|---|
| | T | C | T | C | T | C | HR (95% CI) | $p$ | HR (95% CI) | $p$ |
| **PS-matching / on-treatment TAR** | | | | | | | | | | |
| CCAE | 7,431 | 12,966 | 4.6 | 9.0 | 340 | 553 | 1.18 (0.98 - 1.42) | 0.09 | 1.21 (1.00 - 1.46) | 0.07 |
| ODOD | 5,164 | 8,864 | 2.8 | 5.2 | 246 | 402 | 1.14 (0.91 - 1.43) | 0.26 | 1.12 (0.89 - 1.41) | 0.38 |
| MDCR | 503 | 1,114 | 0.3 | 0.7 | 24 | 54 | 0.76 (0.31 - 1.72) | 0.54 | 0.67 (0.28 - 1.57) | 0.36 |
| MDCD | 920 | 1,600 | 0.5 | 0.8 | 52 | 79 | 0.92 (0.53 - 1.56) | 0.76 | 0.88 (0.51 - 1.52) | 0.67 |
| Open Claims | 109,958 | 183,236 | 73.2 | 136.5 | 2,971 | 5,340 | 1.07 (1.00 - 1.14) | 0.04 | 1.05 (0.98 - 1.13) | 0.19 |
| OEHR | 7,379 | 12,670 | 2.1 | 3.8 | 159 | 254 | 1.11 (0.83 - 1.46) | 0.48 | 1.01 (0.76 - 1.34) | 0.77 |
| Meta-analysis | 130,852 | 219,336 | 83.1 | 155.4 | 3,768 | 6,628 | 1.08 (1.02 - 1.15) | <0.01 | 1.07 (1.00 - 1.14) | 0.07 |
| Meta-analysis (w/o USOC) | 20,894 | 36,100 | 9.9 | 18.9 | 797 | 1,288 | 1.14 (1.00 - 1.29) | 0.05 | 1.13 (0.99 - 1.28) | 0.09 |
| **PS-stratification / on-treatment TAR** | | | | | | | | | | |
| CCAE | 9,602 | 12,966 | 5.6 | 9.0 | 411 | 553 | 1.17 (1.01 - 1.34) | 0.03 | 1.19 (1.03 - 1.37) | 0.02 |
| ODOD | 6,814 | 8,866 | 3.5 | 5.2 | 298 | 402 | 1.16 (0.98 - 1.37) | 0.08 | 1.15 (0.97 - 1.35) | 0.15 |
| MDCR | 545 | 1,115 | 0.3 | 0.7 | 25 | 54 | 1.03 (0.61 - 1.71) | 0.91 | 0.91 (0.54 - 1.52) | 0.71 |
| MDCD | 1,123 | 1,602 | 0.6 | 0.8 | 64 | 79 | 1.04 (0.73 - 1.48) | 0.81 | 1.07 (0.75 - 1.52) | 0.73 |
| Open Claims | 144,211 | 183,262 | 89.1 | 136.5 | 3,714 | 5,341 | 1.08 (1.03 - 1.13) | <0.01 | 1.06 (1.00 - 1.12) | 0.07 |
| OEHR | 9,671 | 12,672 | 2.6 | 3.8 | 218 | 254 | 1.17 (0.96 - 1.43) | 0.11 | 1.04 (0.85 - 1.27) | 0.72 |
| Meta-analysis | 171,421 | 219,368 | 101.3 | 155.4 | 4,705 | 6,629 | 1.10 (1.05 - 1.15) | <0.01 | 1.08 (1.02 - 1.15) | 0.01 |
| Meta-analysis (w/o USOC) | 27,210 | 36,106 | 12.3 | 18.9 | 991 | 1,288 | 1.16 (1.06 - 1.27) | <0.01 | 1.15 (1.05 - 1.27) | <0.01 |
| **PS-matching / on-treatment TAR without escalation** | | | | | | | | | | |
| CCAE | 7,431 | 12,966 | 4.2 | 7.8 | 309 | 485 | 1.16 (0.95 - 1.43) | 0.15 | 1.20 (0.97 - 1.47) | 0.12 |
| ODOD | 5,164 | 8,864 | 2.5 | 4.7 | 223 | 355 | 1.17 (0.92 - 1.50) | 0.20 | 1.14 (0.89 - 1.47) | 0.34 |
| MDCR | 503 | 1,114 | 0.3 | 0.6 | 19 | 49 | 0.35 (0.10 - 0.94) | 0.06 | 0.31 (0.10 - 0.95) | 0.04 |
| MDCD | 920 | 1,600 | 0.4 | 0.7 | 40 | 59 | 0.95 (0.51 - 1.74) | 0.87 | 0.94 (0.51 - 1.75) | 0.80 |
| Open Claims | 109,958 | 183,236 | 65.5 | 115.3 | 2,616 | 4,528 | 1.05 (0.98 - 1.12) | 0.18 | 1.02 (0.95 - 1.10) | 0.62 |
| OEHR | 7,379 | 12,670 | 1.8 | 3.3 | 135 | 218 | 1.06 (0.78 - 1.44) | 0.69 | 0.97 (0.71 - 1.32) | 0.74 |
| Meta-analysis | 130,852 | 219,336 | 74.4 | 131.8 | 3,323 | 5,645 | 1.06 (1.00 - 1.13) | 0.04 | 1.04 (0.96 - 1.12) | 0.34 |
| Meta-analysis (w/o USOC) | 20,894 | 36,100 | 9.0 | 16.6 | 707 | 1,117 | 1.13 (0.99 - 1.30) | 0.07 | 1.13 (0.98 - 1.30) | 0.13 |
| **PS-stratification / on-treatment TAR without escalation** | | | | | | | | | | |
| CCAE | 9,602 | 12,966 | 5.1 | 7.8 | 378 | 485 | 1.15 (0.99 - 1.33) | 0.06 | 1.19 (1.03 - 1.38) | 0.03 |
| ODOD | 6,814 | 8,866 | 3.2 | 4.7 | 272 | 355 | 1.17 (0.98 - 1.39) | 0.08 | 1.18 (0.99 - 1.40) | 0.09 |
| MDCR | 545 | 1,115 | 0.3 | 0.6 | 19 | 49 | 0.76 (0.42 - 1.31) | 0.34 | 0.68 (0.38 - 1.21) | 0.20 |
| MDCD | 1,123 | 1,602 | 0.5 | 0.7 | 50 | 59 | 1.06 (0.70 - 1.59) | 0.79 | 1.13 (0.75 - 1.70) | 0.60 |
| Open Claims | 144,211 | 183,262 | 80.4 | 115.3 | 3,305 | 4,529 | 1.07 (1.02 - 1.12) | <0.01 | 1.05 (0.99 - 1.11) | 0.16 |
| OEHR | 9,671 | 12,672 | 2.3 | 3.3 | 186 | 218 | 1.14 (0.92 - 1.42) | 0.23 | 1.02 (0.82 - 1.27) | 0.76 |
| Meta-analysis | 171,421 | 219,368 | 91.6 | 131.9 | 4,191 | 5,646 | 1.09 (1.04 - 1.14) | <0.01 | 1.07 (1.01 - 1.13) | 0.02 |
| Meta-analysis (w/o USOC) | 27,210 | 36,106 | 11.2 | 16.6 | 886 | 1,117 | 1.15 (1.04 - 1.26) | <0.01 | 1.17 (1.05 - 1.29) | <0.01 |

##### 1.1.17 Hyperkalemia

**Supplementary Table 17:** Exposures, outcomes and risk estimates for Hyperkalemia in GLP1RA (female) (T) vs SGLT2I (female) (C) new-user cohorts under four different propensity score (PS) adjustment / time-at-risk (TAR) designs across data sources. We report total patient exposure time in 1,000 patient-year (KYr) units, uncalibrated and calibrated hazard ratios (HRs) and their 95% confidence intervals (CIs). Greyed out entries did not pass study diagnostics and are included only for completeness.

| | Patients | | Exposure (KYr) | | Outcomes | | Uncalibrated | | Calibrated | |
|---|---|---|---|---|---|---|---|---|---|---|
| | T | C | T | C | T | C | HR (95% CI) | $p$ | HR (95% CI) | $p$ |
| **PS-matching / on-treatment TAR** | | | | | | | | | | |
| CCAE | 10,686 | 18,560 | 6.9 | 13.4 | 32 | 76 | 1.03 (0.59 - 1.78) | 0.92 | 1.05 (0.60 - 1.83) | 0.86 |
| ODOD | 7,258 | 12,377 | 4.1 | 7.6 | 56 | 109 | 0.80 (0.50 - 1.25) | 0.34 | 0.79 (0.50 - 1.24) | 0.31 |
| MDCR | 717 | 1,450 | 0.4 | 0.9 | <5 | 5 | 0.87 (0.11 - 5.36) | 0.89 | 0.76 (0.11 - 5.24) | 0.78 |
| MDCD | 1,450 | 2,358 | 0.8 | 1.3 | 5 | 7 | 1.27 (0.31 - 4.96) | 0.74 | 1.22 (0.30 - 4.93) | 0.78 |
| Open Claims | 167,890 | 271,508 | 113.2 | 201.2 | 361 | 716 | 0.96 (0.80 - 1.15) | 0.67 | 0.95 (0.79 - 1.14) | 0.56 |
| OEHR | 9,580 | 16,393 | 2.8 | 5.1 | 24 | 32 | 0.77 (0.32 - 1.72) | 0.53 | 0.70 (0.30 - 1.62) | 0.41 |
| Meta-analysis | 195,414 | 318,838 | 127.0 | 227.2 | 473 | 933 | 0.94 (0.80 - 1.10) | 0.43 | 0.92 (0.78 - 1.09) | 0.34 |
| Meta-analysis (w/o USOC) | 27,524 | 47,330 | 13.8 | 26.1 | 112 | 217 | 0.87 (0.63 - 1.20) | 0.39 | 0.86 (0.62 - 1.19) | 0.37 |
| **PS-stratification / on-treatment TAR** | | | | | | | | | | |
| CCAE | 14,113 | 18,560 | 8.6 | 13.4 | 36 | 76 | 0.81 (0.52 - 1.23) | 0.33 | 0.83 (0.54 - 1.27) | 0.38 |
| ODOD | 9,779 | 12,381 | 5.2 | 7.6 | 65 | 109 | 0.94 (0.67 - 1.31) | 0.73 | 0.93 (0.67 - 1.30) | 0.69 |
| MDCR | 798 | 1,451 | 0.5 | 0.9 | 5 | 5 | 1.22 (0.28 - 4.91) | 0.79 | 1.07 (0.26 - 4.46) | 0.92 |
| MDCD | 1,710 | 2,363 | 0.9 | 1.3 | 6 | 7 | 1.21 (0.36 - 3.94) | 0.76 | 1.23 (0.37 - 4.08) | 0.74 |
| Open Claims | 227,232 | 271,518 | 141.3 | 201.2 | 425 | 717 | 1.04 (0.92 - 1.19) | 0.52 | 1.03 (0.90 - 1.17) | 0.72 |
| OEHR | 12,701 | 16,395 | 3.7 | 5.1 | 29 | 32 | 1.24 (0.72 - 2.12) | 0.43 | 1.09 (0.64 - 1.88) | 0.75 |
| Meta-analysis | 263,825 | 318,854 | 158.8 | 227.2 | 555 | 934 | 1.02 (0.91 - 1.14) | 0.72 | 1.01 (0.89 - 1.13) | 0.90 |
| Meta-analysis (w/o USOC) | 36,593 | 47,336 | 17.5 | 26.1 | 130 | 217 | 0.95 (0.75 - 1.20) | 0.66 | 0.94 (0.74 - 1.20) | 0.64 |
| **PS-matching / on-treatment TAR without escalation** | | | | | | | | | | |
| CCAE | 10,686 | 18,560 | 6.3 | 11.5 | 30 | 67 | 0.95 (0.52 - 1.71) | 0.86 | 0.98 (0.54 - 1.78) | 0.92 |
| ODOD | 7,258 | 12,377 | 3.7 | 6.7 | 55 | 97 | 0.81 (0.50 - 1.30) | 0.40 | 0.79 (0.49 - 1.28) | 0.36 |
| MDCR | 717 | 1,450 | 0.4 | 0.8 | <5 | 5 | 0.87 (0.11 - 5.36) | 0.89 | 0.77 (0.11 - 5.33) | 0.79 |
| MDCD | 1,450 | 2,358 | 0.7 | 1.0 | 5 | 7 | 1.27 (0.31 - 4.96) | 0.74 | 1.26 (0.31 - 5.07) | 0.75 |
| Open Claims | 167,890 | 271,508 | 101.1 | 169.2 | 316 | 579 | 0.99 (0.81 - 1.21) | 0.94 | 0.97 (0.79 - 1.18) | 0.75 |
| OEHR | 9,580 | 16,393 | 2.4 | 4.5 | 22 | 26 | 0.77 (0.28 - 1.90) | 0.59 | 0.70 (0.27 - 1.81) | 0.46 |
| Meta-analysis | 195,414 | 318,838 | 113.5 | 191.9 | 423 | 769 | 0.96 (0.80 - 1.13) | 0.60 | 0.93 (0.78 - 1.12) | 0.45 |
| Meta-analysis (w/o USOC) | 27,524 | 47,330 | 12.4 | 22.7 | 107 | 190 | 0.85 (0.60 - 1.20) | 0.36 | 0.84 (0.60 - 1.20) | 0.35 |
| **PS-stratification / on-treatment TAR without escalation** | | | | | | | | | | |
| CCAE | 14,113 | 18,560 | 7.8 | 11.5 | 34 | 67 | 0.80 (0.51 - 1.24) | 0.33 | 0.83 (0.53 - 1.30) | 0.42 |
| ODOD | 9,779 | 12,381 | 4.8 | 6.7 | 63 | 97 | 1.03 (0.73 - 1.46) | 0.85 | 1.04 (0.74 - 1.48) | 0.81 |
| MDCR | 798 | 1,451 | 0.4 | 0.8 | 5 | 5 | 1.19 (0.28 - 4.79) | 0.81 | 1.07 (0.26 - 4.46) | 0.92 |
| MDCD | 1,710 | 2,363 | 0.8 | 1.0 | 6 | 7 | 1.15 (0.35 - 3.78) | 0.81 | 1.23 (0.37 - 4.07) | 0.74 |
| Open Claims | 227,232 | 271,518 | 127.6 | 169.2 | 372 | 580 | 1.07 (0.93 - 1.23) | 0.37 | 1.04 (0.90 - 1.20) | 0.58 |
| OEHR | 12,701 | 16,395 | 3.2 | 4.5 | 27 | 26 | 1.42 (0.79 - 2.54) | 0.24 | 1.27 (0.71 - 2.27) | 0.43 |
| Meta-analysis | 263,825 | 318,854 | 143.3 | 191.9 | 496 | 770 | 1.05 (0.93 - 1.19) | 0.41 | 1.04 (0.91 - 1.18) | 0.57 |
| Meta-analysis (w/o USOC) | 36,593 | 47,336 | 15.8 | 22.7 | 124 | 190 | 1.01 (0.77 - 1.33) | 0.92 | 1.03 (0.78 - 1.36) | 0.83 |

##### 1.1.18 Hypoglycemia

**Supplementary Table 18:** Exposures, outcomes and risk estimates for Hypoglycemia in GLP1RA (female) (T) vs SGLT2I (female) (C) new-user cohorts under four different propensity score (PS) adjustment / time-at-risk (TAR) designs across data sources. We report total patient exposure time in 1,000 patient-year (KYr) units, uncalibrated and calibrated hazard ratios (HRs) and their 95% confidence intervals (CIs). Greyed out entries did not pass study diagnostics and are included only for completeness.

| | Patients | | Exposure (KYr) | | Outcomes | | Uncalibrated | | Calibrated | |
|---|---|---|---|---|---|---|---|---|---|---|
| | T | C | T | C | T | C | HR (95% CI) | $p$ | HR (95% CI) | $p$ |
| **PS-matching / on-treatment TAR** | | | | | | | | | | |
| CCAE | 10,584 | 18,425 | 6.9 | 13.3 | 26 | 50 | 0.94 (0.50 - 1.72) | 0.84 | 0.96 (0.52 - 1.78) | 0.90 |
| ODOD | 7,341 | 12,576 | 4.1 | 7.7 | 32 | 50 | 1.19 (0.62 - 2.27) | 0.59 | 1.17 (0.61 - 2.24) | 0.64 |
| MDCR | 721 | 1,472 | 0.4 | 0.9 | <5 | 0 | | | | |
| MDCD | 1,427 | 2,344 | 0.8 | 1.3 | <5 | 12 | 0.22 (0.01 - 1.28) | 0.20 | 0.21 (0.02 - 2.21) | 0.19 |
| Open Claims | 167,043 | 270,929 | 112.8 | 200.8 | 311 | 416 | 1.57 (1.27 - 1.93) | <0.01 | 1.54 (1.25 - 1.90) | <0.01 |
| OEHR | 9,568 | 16,442 | 2.8 | 5.2 | 17 | 18 | 1.79 (0.68 - 4.83) | 0.24 | 1.63 (0.61 - 4.34) | 0.33 |
| Meta-analysis | 194,536 | 318,372 | 126.6 | 226.9 | 386 | 534 | 1.41 (1.09 - 1.82) | <0.01 | 1.38 (1.07 - 1.79) | 0.01 |
| Meta-analysis (w/o USOC) | 27,493 | 47,443 | 13.9 | 26.1 | 75 | 118 | 1.15 (0.77 - 1.73) | 0.49 | 1.14 (0.76 - 1.72) | 0.52 |
| **PS-stratification / on-treatment TAR** | | | | | | | | | | |
| CCAE | 13,925 | 18,425 | 8.6 | 13.3 | 43 | 50 | 1.08 (0.69 - 1.68) | 0.75 | 1.10 (0.70 - 1.72) | 0.68 |
| ODOD | 9,812 | 12,580 | 5.2 | 7.7 | 45 | 50 | 1.01 (0.64 - 1.57) | 0.98 | 0.99 (0.63 - 1.56) | 0.93 |
| MDCR | 804 | 1,473 | 0.5 | 0.9 | <5 | 0 | | | | |
| MDCD | 1,676 | 2,349 | 0.9 | 1.3 | 8 | 12 | 0.62 (0.23 - 1.62) | 0.34 | 0.63 (0.24 - 1.70) | 0.37 |
| Open Claims | 224,703 | 270,936 | 140.1 | 200.8 | 400 | 416 | 1.28 (1.10 - 1.48) | <0.01 | 1.26 (1.08 - 1.46) | <0.01 |
| OEHR | 12,655 | 16,444 | 3.6 | 5.2 | 30 | 18 | 1.88 (1.01 - 3.57) | 0.05 | 1.66 (0.88 - 3.12) | 0.13 |
| Meta-analysis | 262,771 | 320,734 | 158.5 | 228.2 | 526 | 546 | 1.24 (1.09 - 1.41) | <0.01 | 1.22 (1.06 - 1.40) | <0.01 |
| Meta-analysis (w/o USOC) | 38,068 | 49,798 | 18.4 | 27.4 | 126 | 130 | 1.12 (0.80 - 1.56) | 0.51 | 1.11 (0.79 - 1.56) | 0.54 |
| **PS-matching / on-treatment TAR without escalation** | | | | | | | | | | |
| CCAE | 10,584 | 18,425 | 6.2 | 11.5 | 21 | 37 | 1.13 (0.57 - 2.20) | 0.72 | 1.16 (0.59 - 2.29) | 0.67 |
| ODOD | 7,341 | 12,576 | 3.8 | 6.8 | 26 | 37 | 1.14 (0.54 - 2.44) | 0.73 | 1.11 (0.52 - 2.38) | 0.78 |
| MDCR | 721 | 1,472 | 0.4 | 0.8 | <5 | 0 | | | | |
| MDCD | 1,427 | 2,344 | 0.7 | 1.0 | <5 | 9 | 0.00 (0.00 - 0.68) | 0.20 | 0.00 (0.00 - 25325) | 0.20 |
| Open Claims | 167,043 | 270,929 | 100.8 | 168.9 | 260 | 277 | 1.60 (1.27 - 2.04) | <0.01 | 1.57 (1.23 - 1.99) | <0.01 |
| OEHR | 9,568 | 16,442 | 2.4 | 4.5 | 12 | 9 | 2.23 (0.65 - 8.83) | 0.23 | 2.03 (0.55 - 7.47) | 0.29 |
| Meta-analysis | 194,536 | 318,372 | 113.2 | 191.6 | 319 | 360 | 1.52 (1.23 - 1.88) | <0.01 | 1.49 (1.20 - 1.85) | <0.01 |
| Meta-analysis (w/o USOC) | 27,493 | 47,443 | 12.4 | 22.8 | 59 | 83 | 1.24 (0.77 - 1.98) | 0.37 | 1.23 (0.77 - 1.97) | 0.39 |
| **PS-stratification / on-treatment TAR without escalation** | | | | | | | | | | |
| CCAE | 13,925 | 18,425 | 7.8 | 11.5 | 37 | 37 | 1.19 (0.72 - 1.95) | 0.50 | 1.23 (0.75 - 2.03) | 0.42 |
| ODOD | 9,812 | 12,580 | 4.8 | 6.8 | 39 | 37 | 1.01 (0.61 - 1.67) | 0.98 | 1.02 (0.61 - 1.68) | 0.93 |
| MDCR | 804 | 1,473 | 0.4 | 0.8 | <5 | 0 | | | | |
| MDCD | 1,676 | 2,349 | 0.8 | 1.0 | 6 | 9 | 0.56 (0.18 - 1.66) | 0.31 | 0.59 (0.19 - 1.83) | 0.37 |
| Open Claims | 224,703 | 270,936 | 126.5 | 168.9 | 337 | 277 | 1.47 (1.24 - 1.75) | <0.01 | 1.44 (1.20 - 1.71) | <0.01 |
| OEHR | 12,655 | 16,444 | 3.2 | 4.5 | 23 | 9 | 2.74 (1.23 - 6.56) | 0.02 | 2.45 (1.06 - 5.66) | 0.04 |
| Meta-analysis | 261,095 | 318,385 | 142.2 | 191.7 | 436 | 360 | 1.40 (1.17 - 1.69) | <0.01 | 1.38 (1.15 - 1.67) | <0.01 |
| Meta-analysis (w/o USOC) | 36,392 | 47,449 | 15.7 | 22.8 | 99 | 83 | 1.34 (0.82 - 2.19) | 0.24 | 1.36 (0.84 - 2.23) | 0.22 |

##### 1.1.19 Hypotension

**Supplementary Table 19:** Exposures, outcomes and risk estimates for Hypotension in GLP1RA (female) (T) vs SGLT2I (female) (C) new-user cohorts under four different propensity score (PS) adjustment / time-at-risk (TAR) designs across data sources. We report total patient exposure time in 1,000 patient-year (KYr) units, uncalibrated and calibrated hazard ratios (HRs) and their 95% confidence intervals (CIs). Greyed out entries did not pass study diagnostics and are included only for completeness.

| | Patients | | Exposure (KYr) | | Outcomes | | Uncalibrated | | Calibrated | |
|---|---|---|---|---|---|---|---|---|---|---|
| | T | C | T | C | T | C | HR (95% CI) | $p$ | HR (95% CI) | $p$ |
| **PS-matching / on-treatment TAR** | | | | | | | | | | |
| CCAE | 10,655 | 18,471 | 6.9 | 13.3 | 60 | 110 | 1.20 (0.78 - 1.83) | 0.40 | 1.23 (0.80 - 1.88) | 0.36 |
| ODOD | 7,194 | 12,288 | 4.0 | 7.5 | 70 | 148 | 0.96 (0.62 - 1.45) | 0.83 | 0.94 (0.61 - 1.44) | 0.78 |
| MDCR | 692 | 1,424 | 0.4 | 0.9 | 7 | 25 | 0.53 (0.14 - 1.54) | 0.29 | 0.46 (0.14 - 1.50) | 0.20 |
| MDCD | 1,396 | 2,259 | 0.8 | 1.2 | 22 | 42 | 0.34 (0.14 - 0.76) | 0.01 | 0.33 (0.14 - 0.78) | 0.01 |
| Open Claims | 165,007 | 267,166 | 111.1 | 198.1 | 734 | 1,306 | 1.09 (0.96 - 1.24) | 0.18 | 1.07 (0.94 - 1.22) | 0.30 |
| OEHR | 9,495 | 16,278 | 2.7 | 5.1 | 44 | 60 | 1.45 (0.86 - 2.45) | 0.16 | 1.33 (0.79 - 2.25) | 0.30 |
| Meta-analysis | 193,747 | 316,462 | 125.5 | 225.2 | 930 | 1,666 | 1.08 (0.97 - 1.21) | 0.18 | 1.06 (0.94 - 1.20) | 0.32 |
| Meta-analysis (w/o USOC) | 28,740 | 49,296 | 14.5 | 27.1 | 196 | 360 | 0.97 (0.62 - 1.51) | 0.88 | 0.96 (0.61 - 1.49) | 0.85 |
| **PS-stratification / on-treatment TAR** | | | | | | | | | | |
| CCAE | 14,028 | 18,471 | 8.6 | 13.3 | 81 | 110 | 1.05 (0.77 - 1.44) | 0.75 | 1.08 (0.78 - 1.48) | 0.66 |
| ODOD | 9,654 | 12,292 | 5.1 | 7.5 | 90 | 148 | 0.85 (0.63 - 1.13) | 0.26 | 0.84 (0.62 - 1.12) | 0.25 |
| MDCR | 776 | 1,425 | 0.5 | 0.9 | 7 | 25 | 0.56 (0.20 - 1.32) | 0.23 | 0.49 (0.19 - 1.26) | 0.14 |
| MDCD | 1,644 | 2,263 | 0.9 | 1.2 | 25 | 42 | 0.71 (0.42 - 1.19) | 0.21 | 0.73 (0.43 - 1.23) | 0.25 |
| Open Claims | 222,878 | 267,171 | 138.4 | 198.1 | 937 | 1,308 | 1.11 (1.01 - 1.22) | 0.02 | 1.09 (0.99 - 1.20) | 0.09 |
| OEHR | 12,645 | 16,280 | 3.6 | 5.1 | 55 | 60 | 1.46 (0.99 - 2.17) | 0.06 | 1.29 (0.87 - 1.92) | 0.22 |
| Meta-analysis | 260,849 | 316,477 | 156.6 | 225.2 | 1,188 | 1,668 | 1.04 (0.88 - 1.23) | 0.62 | 1.03 (0.87 - 1.22) | 0.75 |
| Meta-analysis (w/o USOC) | 37,971 | 49,306 | 18.2 | 27.1 | 251 | 360 | 1.00 (0.76 - 1.31) | 0.98 | 0.99 (0.75 - 1.30) | 0.92 |
| **PS-matching / on-treatment TAR without escalation** | | | | | | | | | | |
| CCAE | 10,655 | 18,471 | 6.2 | 11.5 | 55 | 84 | 1.27 (0.80 - 2.01) | 0.31 | 1.31 (0.82 - 2.08) | 0.27 |
| ODOD | 7,194 | 12,288 | 3.7 | 6.6 | 64 | 126 | 0.90 (0.57 - 1.39) | 0.63 | 0.87 (0.56 - 1.37) | 0.57 |
| MDCR | 692 | 1,424 | 0.4 | 0.8 | 7 | 24 | 0.48 (0.13 - 1.41) | 0.22 | 0.43 (0.13 - 1.39) | 0.16 |
| MDCD | 1,396 | 2,259 | 0.7 | 1.0 | 19 | 32 | 0.22 (0.07 - 0.60) | <0.01 | 0.22 (0.07 - 0.67) | <0.01 |
| Open Claims | 165,007 | 267,166 | 99.3 | 166.7 | 621 | 1,065 | 1.05 (0.91 - 1.21) | 0.50 | 1.02 (0.89 - 1.18) | 0.76 |
| OEHR | 9,495 | 16,278 | 2.4 | 4.4 | 34 | 42 | 1.49 (0.81 - 2.73) | 0.20 | 1.36 (0.74 - 2.49) | 0.33 |
| Meta-analysis | 193,747 | 316,462 | 112.3 | 190.3 | 793 | 1,349 | 1.05 (0.92 - 1.18) | 0.48 | 1.02 (0.90 - 1.17) | 0.75 |
| Meta-analysis (w/o USOC) | 28,740 | 49,296 | 13.0 | 23.6 | 172 | 284 | 0.93 (0.54 - 1.59) | 0.78 | 0.92 (0.54 - 1.58) | 0.76 |
| **PS-stratification / on-treatment TAR without escalation** | | | | | | | | | | |
| CCAE | 14,028 | 18,471 | 7.8 | 11.5 | 73 | 84 | 1.20 (0.85 - 1.70) | 0.29 | 1.25 (0.88 - 1.77) | 0.21 |
| ODOD | 9,654 | 12,292 | 4.7 | 6.6 | 82 | 126 | 0.90 (0.66 - 1.23) | 0.53 | 0.91 (0.67 - 1.25) | 0.57 |
| MDCR | 776 | 1,425 | 0.4 | 0.8 | 7 | 24 | 0.59 (0.22 - 1.39) | 0.27 | 0.53 (0.21 - 1.35) | 0.19 |
| MDCD | 1,644 | 2,263 | 0.8 | 1.0 | 20 | 32 | 0.71 (0.39 - 1.27) | 0.26 | 0.76 (0.42 - 1.37) | 0.38 |
| Open Claims | 222,878 | 267,171 | 125.0 | 166.7 | 807 | 1,066 | 1.10 (1.00 - 1.22) | 0.06 | 1.08 (0.97 - 1.19) | 0.19 |
| OEHR | 12,645 | 16,280 | 3.2 | 4.4 | 43 | 42 | 1.63 (1.03 - 2.59) | 0.04 | 1.46 (0.92 - 2.33) | 0.12 |
| Meta-analysis | 260,849 | 316,477 | 141.4 | 190.3 | 1,025 | 1,350 | 1.09 (1.00 - 1.20) | 0.05 | 1.08 (0.98 - 1.19) | 0.13 |
| Meta-analysis (w/o USOC) | 37,971 | 49,306 | 16.4 | 23.6 | 218 | 284 | 1.08 (0.80 - 1.46) | 0.63 | 1.10 (0.81 - 1.49) | 0.56 |

###### 1.1.20 Joint pain

**Supplementary Table 20:** Exposures, outcomes and risk estimates for Joint pain in GLP1RA (female) (T) vs SGLT2I (female) (C) new-user cohorts under four different propensity score (PS) adjustment / time-at-risk (TAR) designs across data sources. We report total patient exposure time in 1,000 patient-year (KYr) units, uncalibrated and calibrated hazard ratios (HRs) and their 95% confidence intervals (CIs). Greyed out entries did not pass study diagnostics and are included only for completeness.

| | Patients | | Exposure (KYr) | | Outcomes | | Uncalibrated | | Calibrated | |
|---|---|---|---|---|---|---|---|---|---|---|
| | T | C | T | C | T | C | HR (95% CI) | $p$ | HR (95% CI) | $p$ |
| **PS-matching / on-treatment TAR** | | | | | | | | | | |
| CCAE | 9,829 | 17,271 | 6.4 | 12.5 | 115 | 175 | 0.97 (0.68 - 1.38) | 0.88 | 1.00 (0.70 - 1.42) | 0.90 |
| ODOD | 6,780 | 11,746 | 3.8 | 7.1 | 78 | 129 | 0.77 (0.50 - 1.16) | 0.22 | 0.76 (0.50 - 1.15) | 0.20 |
| MDCR | 677 | 1,411 | 0.4 | 0.9 | 5 | 14 | 0.72 (0.19 - 2.45) | 0.62 | 0.63 (0.17 - 2.28) | 0.48 |
| MDCD | 1,299 | 2,153 | 0.7 | 1.2 | 22 | 31 | 1.77 (0.69 - 4.62) | 0.24 | 1.70 (0.65 - 4.41) | 0.29 |
| Open Claims | 155,108 | 253,902 | 104.5 | 188.4 | 828 | 1,464 | 0.94 (0.83 - 1.07) | 0.35 | 0.93 (0.81 - 1.05) | 0.26 |
| OEHR | 8,894 | 15,385 | 2.6 | 4.8 | 47 | 80 | 1.04 (0.60 - 1.80) | 0.88 | 0.95 (0.55 - 1.65) | 0.84 |
| Meta-analysis | 181,910 | 300,457 | 118.0 | 214.0 | 1,090 | 1,879 | 0.94 (0.84 - 1.05) | 0.30 | 0.93 (0.83 - 1.04) | 0.22 |
| Meta-analysis (w/o USOC) | 26,802 | 46,555 | 13.4 | 25.6 | 262 | 415 | 0.95 (0.75 - 1.20) | 0.66 | 0.94 (0.74 - 1.19) | 0.63 |
| **PS-stratification / on-treatment TAR** | | | | | | | | | | |
| CCAE | 12,834 | 17,271 | 7.8 | 12.5 | 140 | 175 | 1.18 (0.93 - 1.51) | 0.17 | 1.21 (0.95 - 1.55) | 0.14 |
| ODOD | 8,959 | 11,750 | 4.7 | 7.2 | 97 | 129 | 0.97 (0.72 - 1.29) | 0.82 | 0.96 (0.71 - 1.28) | 0.77 |
| MDCR | 743 | 1,412 | 0.4 | 0.9 | 5 | 14 | 0.55 (0.17 - 1.54) | 0.29 | 0.48 (0.16 - 1.46) | 0.20 |
| MDCD | 1,513 | 2,157 | 0.8 | 1.2 | 26 | 31 | 1.15 (0.66 - 2.00) | 0.62 | 1.18 (0.68 - 2.05) | 0.58 |
| Open Claims | 207,412 | 253,928 | 129.2 | 188.5 | 1,119 | 1,465 | 0.98 (0.90 - 1.06) | 0.58 | 0.96 (0.88 - 1.05) | 0.40 |
| OEHR | 11,723 | 15,387 | 3.3 | 4.8 | 61 | 80 | 0.97 (0.68 - 1.39) | 0.88 | 0.86 (0.60 - 1.23) | 0.43 |
| Meta-analysis | 242,441 | 300,493 | 145.9 | 214.0 | 1,443 | 1,880 | 1.00 (0.92 - 1.07) | 0.92 | 0.98 (0.90 - 1.07) | 0.69 |
| Meta-analysis (w/o USOC) | 35,029 | 46,565 | 16.7 | 25.6 | 324 | 415 | 1.07 (0.91 - 1.26) | 0.40 | 1.07 (0.91 - 1.25) | 0.45 |
| **PS-matching / on-treatment TAR without escalation** | | | | | | | | | | |
| CCAE | 9,829 | 17,271 | 5.7 | 10.8 | 101 | 150 | 0.92 (0.63 - 1.34) | 0.68 | 0.95 (0.65 - 1.39) | 0.80 |
| ODOD | 6,780 | 11,746 | 3.5 | 6.3 | 66 | 113 | 0.71 (0.45 - 1.10) | 0.13 | 0.69 (0.44 - 1.08) | 0.12 |
| MDCR | 677 | 1,411 | 0.4 | 0.8 | 5 | 13 | 0.64 (0.13 - 2.48) | 0.55 | 0.57 (0.13 - 2.46) | 0.45 |
| MDCD | 1,299 | 2,153 | 0.6 | 1.0 | 21 | 28 | 1.62 (0.63 - 4.19) | 0.32 | 1.60 (0.62 - 4.12) | 0.34 |
| Open Claims | 155,108 | 253,902 | 93.3 | 158.6 | 735 | 1,232 | 0.93 (0.81 - 1.06) | 0.28 | 0.90 (0.78 - 1.04) | 0.17 |
| OEHR | 8,894 | 15,385 | 2.2 | 4.2 | 40 | 72 | 0.93 (0.51 - 1.66) | 0.80 | 0.84 (0.47 - 1.52) | 0.57 |
| Meta-analysis | 181,910 | 300,457 | 105.4 | 180.8 | 963 | 1,595 | 0.92 (0.81 - 1.03) | 0.16 | 0.89 (0.79 - 1.02) | 0.10 |
| Meta-analysis (w/o USOC) | 26,802 | 46,555 | 12.1 | 22.2 | 228 | 363 | 0.88 (0.69 - 1.14) | 0.34 | 0.88 (0.68 - 1.13) | 0.33 |
| **PS-stratification / on-treatment TAR without escalation** | | | | | | | | | | |
| CCAE | 12,834 | 17,271 | 7.1 | 10.8 | 124 | 150 | 1.16 (0.90 - 1.51) | 0.25 | 1.21 (0.93 - 1.57) | 0.17 |
| ODOD | 8,959 | 11,750 | 4.4 | 6.3 | 84 | 113 | 0.91 (0.67 - 1.25) | 0.57 | 0.92 (0.67 - 1.26) | 0.62 |
| MDCR | 743 | 1,412 | 0.4 | 0.8 | 5 | 13 | 0.61 (0.18 - 1.75) | 0.38 | 0.54 (0.18 - 1.68) | 0.29 |
| MDCD | 1,513 | 2,157 | 0.7 | 1.0 | 25 | 28 | 1.20 (0.67 - 2.14) | 0.53 | 1.28 (0.72 - 2.28) | 0.42 |
| Open Claims | 207,412 | 253,928 | 116.4 | 158.6 | 1,006 | 1,233 | 0.97 (0.89 - 1.07) | 0.58 | 0.95 (0.87 - 1.05) | 0.35 |
| OEHR | 11,723 | 15,387 | 2.9 | 4.2 | 53 | 72 | 0.91 (0.61 - 1.33) | 0.63 | 0.81 (0.55 - 1.20) | 0.31 |
| Meta-analysis | 242,441 | 300,493 | 131.5 | 180.9 | 1,292 | 1,596 | 0.99 (0.91 - 1.07) | 0.77 | 0.97 (0.89 - 1.06) | 0.57 |
| Meta-analysis (w/o USOC) | 35,029 | 46,565 | 15.1 | 22.2 | 286 | 363 | 1.04 (0.87 - 1.23) | 0.68 | 1.05 (0.89 - 1.25) | 0.57 |

##### 1.1.21 Lower extremity amputation

**Supplementary Table 21:** Exposures, outcomes and risk estimates for Lower extremity amputation in GLP1RA (female) (T) vs SGLT2I (female) (C) new-user cohorts under four different propensity score (PS) adjustment / time-at-risk (TAR) designs across data sources. We report total patient exposure time in 1,000 patient-year (KYr) units, uncalibrated and calibrated hazard ratios (HRs) and their 95% confidence intervals (CIs). Greyed out entries did not pass study diagnostics and are included only for completeness.

| | Patients | | Exposure (KYr) | | Outcomes | | Uncalibrated | | Calibrated | |
|---|---|---|---|---|---|---|---|---|---|---|
| | T | C | T | C | T | C | HR (95% CI) | $p$ | HR (95% CI) | $p$ |
| **PS-matching / on-treatment TAR** | | | | | | | | | | |
| CCAE | 10,881 | 18,874 | 7.1 | 13.7 | <5 | 5 | 1.00 (0.04 - 25.3) | 1.00 | 1.02 (0.04 - 25.8) | 0.99 |
| ODOD | 7,484 | 12,827 | 4.2 | 7.9 | 0 | 5 | | | | |
| MDCR | 734 | 1,485 | 0.4 | 0.9 | 0 | <5 | | | | |
| MDCD | 1,483 | 2,426 | 0.8 | 1.3 | 0 | 0 | | | | |
| Open Claims | 170,652 | 276,297 | 115.2 | 204.7 | 21 | 39 | 1.31 (0.58 - 2.89) | 0.51 | 1.29 (0.58 - 2.87) | 0.53 |
| OEHR | 9,708 | 16,652 | 2.8 | 5.2 | 0 | <5 | | | | |
| Meta-analysis | 170,652 | 276,297 | 115.2 | 204.7 | 21 | 39 | 1.31 (0.58 - 2.89) | 0.51 | 1.29 (0.58 - 2.87) | 0.53 |
| **PS-stratification / on-treatment TAR** | | | | | | | | | | |
| CCAE | 14,357 | 18,874 | 8.8 | 13.7 | <5 | 5 | 1.02 (0.14 - 5.23) | 0.98 | 1.04 (0.17 - 6.42) | 0.96 |
| ODOD | 10,063 | 12,831 | 5.4 | 7.9 | 0 | 5 | | | | |
| MDCR | 818 | 1,486 | 0.5 | 0.9 | 0 | <5 | | | | |
| MDCD | 1,745 | 2,431 | 1.0 | 1.3 | <5 | 0 | | | | |
| Open Claims | 230,754 | 276,304 | 143.8 | 204.7 | 22 | 39 | 1.04 (0.59 - 1.81) | 0.89 | 1.02 (0.58 - 1.80) | 0.94 |
| OEHR | 12,890 | 16,654 | 3.7 | 5.2 | 0 | <5 | | | | |
| Meta-analysis | 230,754 | 276,304 | 143.8 | 204.7 | 22 | 39 | 1.04 (0.59 - 1.81) | 0.89 | 1.03 (0.58 - 1.81) | 0.93 |
| **PS-matching / on-treatment TAR without escalation** | | | | | | | | | | |
| CCAE | 10,881 | 18,874 | 6.4 | 11.8 | <5 | <5 | 1.00 (0.04 - 25.3) | 1.00 | 1.03 (0.04 - 26.0) | 0.98 |
| ODOD | 7,484 | 12,827 | 3.9 | 7.0 | 0 | <5 | | | | |
| MDCR | 734 | 1,485 | 0.4 | 0.8 | 0 | <5 | | | | |
| MDCD | 1,483 | 2,426 | 0.7 | 1.1 | 0 | 0 | | | | |
| Open Claims | 170,652 | 276,297 | 103.0 | 172.1 | 15 | 27 | 1.13 (0.45 - 2.76) | 0.79 | 1.10 (0.44 - 2.73) | 0.83 |
| OEHR | 9,708 | 16,652 | 2.4 | 4.5 | 0 | <5 | | | | |
| Meta-analysis | 170,652 | 276,297 | 103.0 | 172.1 | 15 | 27 | 1.13 (0.45 - 2.76) | 0.79 | 1.10 (0.44 - 2.74) | 0.83 |
| **PS-stratification / on-treatment TAR without escalation** | | | | | | | | | | |
| CCAE | 14,357 | 18,874 | 8.0 | 11.8 | <5 | <5 | 1.06 (0.14 - 5.84) | 0.95 | 1.10 (0.17 - 7.09) | 0.92 |
| ODOD | 10,063 | 12,831 | 4.9 | 7.0 | 0 | <5 | | | | |
| MDCR | 818 | 1,486 | 0.4 | 0.8 | 0 | <5 | | | | |
| MDCD | 1,745 | 2,431 | 0.8 | 1.1 | <5 | 0 | | | | |
| Open Claims | 230,754 | 276,304 | 129.7 | 172.1 | 16 | 27 | 0.94 (0.47 - 1.83) | 0.86 | 0.92 (0.47 - 1.81) | 0.81 |
| OEHR | 12,890 | 16,654 | 3.2 | 4.6 | 0 | <5 | | | | |
| Meta-analysis | 230,754 | 276,304 | 129.7 | 172.1 | 16 | 27 | 0.94 (0.47 - 1.83) | 0.86 | 0.93 (0.47 - 1.83) | 0.83 |

##### 1.1.22 Nausea

**Supplementary Table 22:** Exposures, outcomes and risk estimates for Nausea in GLP1RA (female) (T) vs SGLT2I (female) (C) new-user cohorts under four different propensity score (PS) adjustment / time-at-risk (TAR) designs across data sources. We report total patient exposure time in 1,000 patient-year (KYr) units, uncalibrated and calibrated hazard ratios (HRs) and their 95% confidence intervals (CIs). Greyed out entries did not pass study diagnostics and are included only for completeness.

| | Patients | | Exposure (KYr) | | Outcomes | | Uncalibrated | | Calibrated | |
|---|---|---|---|---|---|---|---|---|---|---|
| | T | C | T | C | T | C | HR (95% CI) | $p$ | HR (95% CI) | $p$ |
| **PS-matching / on-treatment TAR** | | | | | | | | | | |
| CCAE | 8,590 | 15,256 | 5.4 | 10.9 | 385 | 515 | 1.46 (1.22 - 1.76) | <0.01 | 1.50 (1.24 - 1.80) | <0.01 |
| ODOD | 5,818 | 10,199 | 3.2 | 6.2 | 292 | 321 | 1.79 (1.44 - 2.23) | <0.01 | 1.76 (1.41 - 2.20) | <0.01 |
| MDCR | 643 | 1,258 | 0.4 | 0.8 | 23 | 32 | 3.16 (1.26 - 8.58) | 0.02 | 2.76 (1.06 - 7.20) | 0.04 |
| MDCD | 909 | 1,538 | 0.5 | 0.8 | 87 | 79 | 1.69 (1.06 - 2.71) | 0.03 | 1.62 (1.01 - 2.61) | 0.06 |
| Open Claims | 132,962 | 221,448 | 89.5 | 165.5 | 3,475 | 3,960 | 1.73 (1.63 - 1.85) | <0.01 | 1.71 (1.59 - 1.83) | <0.01 |
| OEHR | 7,954 | 14,002 | 2.2 | 4.3 | 203 | 230 | 1.80 (1.38 - 2.34) | <0.01 | 1.64 (1.26 - 2.14) | <0.01 |
| Meta-analysis | 156,233 | 262,443 | 100.9 | 187.7 | 4,442 | 5,105 | 1.71 (1.61 - 1.82) | <0.01 | 1.68 (1.57 - 1.81) | <0.01 |
| Meta-analysis (w/o USOC) | 23,271 | 40,995 | 11.3 | 22.2 | 967 | 1,145 | 1.64 (1.45 - 1.85) | <0.01 | 1.62 (1.43 - 1.84) | <0.01 |
| **PS-stratification / on-treatment TAR** | | | | | | | | | | |
| CCAE | 11,176 | 15,258 | 6.6 | 10.9 | 504 | 515 | 1.36 (1.19 - 1.56) | <0.01 | 1.39 (1.21 - 1.60) | <0.01 |
| ODOD | 7,686 | 10,200 | 4.0 | 6.2 | 406 | 321 | 1.67 (1.43 - 1.97) | <0.01 | 1.65 (1.41 - 1.95) | <0.01 |
| MDCR | 721 | 1,258 | 0.4 | 0.8 | 25 | 32 | 1.20 (0.66 - 2.13) | 0.55 | 1.05 (0.59 - 1.88) | 0.82 |
| MDCD | 1,056 | 1,543 | 0.6 | 0.8 | 93 | 80 | 1.67 (1.22 - 2.30) | <0.01 | 1.71 (1.24 - 2.35) | <0.01 |
| Open Claims | 175,582 | 221,452 | 109.4 | 165.5 | 4,646 | 3,960 | 1.55 (1.48 - 1.62) | <0.01 | 1.52 (1.44 - 1.61) | <0.01 |
| OEHR | 10,399 | 14,004 | 2.9 | 4.3 | 273 | 230 | 1.58 (1.31 - 1.91) | <0.01 | 1.40 (1.15 - 1.69) | <0.01 |
| Meta-analysis | 205,899 | 262,457 | 123.4 | 187.7 | 5,922 | 5,106 | 1.54 (1.46 - 1.62) | <0.01 | 1.52 (1.42 - 1.62) | <0.01 |
| Meta-analysis (w/o USOC) | 30,317 | 41,005 | 14.1 | 22.2 | 1,276 | 1,146 | 1.53 (1.37 - 1.71) | <0.01 | 1.53 (1.36 - 1.71) | <0.01 |
| **PS-matching / on-treatment TAR without escalation** | | | | | | | | | | |
| CCAE | 8,590 | 15,256 | 4.9 | 9.5 | 353 | 450 | 1.49 (1.23 - 1.82) | <0.01 | 1.54 (1.26 - 1.87) | <0.01 |
| ODOD | 5,818 | 10,199 | 2.9 | 5.5 | 272 | 277 | 2.04 (1.61 - 2.60) | <0.01 | 1.99 (1.57 - 2.53) | <0.01 |
| MDCR | 643 | 1,258 | 0.4 | 0.7 | 20 | 29 | 3.27 (1.21 - 9.72) | 0.03 | 2.91 (1.02 - 8.25) | 0.05 |
| MDCD | 909 | 1,538 | 0.4 | 0.7 | 80 | 64 | 1.62 (0.98 - 2.66) | 0.06 | 1.60 (0.97 - 2.64) | 0.08 |
| Open Claims | 132,962 | 221,448 | 80.3 | 140.3 | 3,121 | 3,228 | 1.80 (1.68 - 1.93) | <0.01 | 1.75 (1.62 - 1.89) | <0.01 |
| OEHR | 7,954 | 14,002 | 1.9 | 3.8 | 178 | 190 | 1.84 (1.39 - 2.44) | <0.01 | 1.67 (1.26 - 2.22) | <0.01 |
| Meta-analysis | 156,233 | 262,443 | 90.5 | 159.8 | 4,004 | 4,209 | 1.77 (1.62 - 1.93) | <0.01 | 1.73 (1.56 - 1.91) | <0.01 |
| Meta-analysis (w/o USOC) | 23,271 | 40,995 | 10.2 | 19.5 | 883 | 981 | 1.73 (1.47 - 2.04) | <0.01 | 1.72 (1.46 - 2.03) | <0.01 |
| **PS-stratification / on-treatment TAR without escalation** | | | | | | | | | | |
| CCAE | 11,176 | 15,258 | 6.1 | 9.5 | 465 | 450 | 1.39 (1.21 - 1.60) | <0.01 | 1.44 (1.25 - 1.67) | <0.01 |
| ODOD | 7,686 | 10,200 | 3.7 | 5.5 | 382 | 277 | 1.80 (1.52 - 2.14) | <0.01 | 1.82 (1.53 - 2.16) | <0.01 |
| MDCR | 721 | 1,258 | 0.4 | 0.7 | 22 | 29 | 1.19 (0.64 - 2.20) | 0.58 | 1.07 (0.58 - 1.99) | 0.81 |
| MDCD | 1,056 | 1,543 | 0.5 | 0.7 | 86 | 65 | 1.82 (1.29 - 2.58) | <0.01 | 1.94 (1.37 - 2.75) | <0.01 |
| Open Claims | 175,582 | 221,452 | 98.9 | 140.3 | 4,231 | 3,228 | 1.64 (1.56 - 1.72) | <0.01 | 1.60 (1.51 - 1.69) | <0.01 |
| OEHR | 10,399 | 14,004 | 2.5 | 3.8 | 239 | 190 | 1.68 (1.37 - 2.07) | <0.01 | 1.51 (1.22 - 1.86) | <0.01 |
| Meta-analysis | 205,899 | 262,457 | 111.6 | 159.8 | 5,403 | 4,210 | 1.62 (1.48 - 1.78) | <0.01 | 1.60 (1.45 - 1.76) | <0.01 |
| Meta-analysis (w/o USOC) | 30,317 | 41,005 | 12.7 | 19.5 | 1,172 | 982 | 1.63 (1.41 - 1.88) | <0.01 | 1.65 (1.43 - 1.92) | <0.01 |

##### 1.1.23 Peripheral edema

**Supplementary Table 23:** Exposures, outcomes and risk estimates for Peripheral edema in GLP1RA (female) (T) vs SGLT2I (female) (C) new-user cohorts under four different propensity score (PS) adjustment / time-at-risk (TAR) designs across data sources. We report total patient exposure time in 1,000 patient-year (KYr) units, uncalibrated and calibrated hazard ratios (HRs) and their 95% confidence intervals (CIs). Greyed out entries did not pass study diagnostics and are included only for completeness.

| | Patients | | Exposure (KYr) | | Outcomes | | Uncalibrated | | Calibrated | |
|---|---|---|---|---|---|---|---|---|---|---|
| | T | C | T | C | T | C | HR (95% CI) | $p$ | HR (95% CI) | $p$ |
| **PS-matching / on-treatment TAR** | | | | | | | | | | |
| CCAE | 8,076 | 14,268 | 5.1 | 10.1 | 258 | 434 | 1.13 (0.90 - 1.40) | 0.29 | 1.15 (0.92 - 1.44) | 0.24 |
| ODOD | 5,356 | 9,262 | 3.0 | 5.5 | 169 | 324 | 0.91 (0.70 - 1.17) | 0.45 | 0.89 (0.69 - 1.16) | 0.41 |
| MDCR | 511 | 1,086 | 0.3 | 0.7 | 23 | 30 | 0.98 (0.39 - 2.37) | 0.97 | 0.86 (0.35 - 2.11) | 0.74 |
| MDCD | 937 | 1,533 | 0.5 | 0.8 | 44 | 69 | 0.74 (0.42 - 1.28) | 0.29 | 0.71 (0.41 - 1.25) | 0.26 |
| Open Claims | 121,526 | 201,851 | 81.0 | 149.5 | 2,047 | 3,611 | 1.06 (0.98 - 1.14) | 0.17 | 1.04 (0.96 - 1.13) | 0.41 |
| OEHR | 7,661 | 13,267 | 2.2 | 4.1 | 97 | 157 | 1.20 (0.84 - 1.70) | 0.31 | 1.10 (0.77 - 1.56) | 0.62 |
| Meta-analysis | 143,556 | 240,181 | 91.8 | 170.0 | 2,615 | 4,595 | 1.05 (0.98 - 1.13) | 0.16 | 1.03 (0.96 - 1.12) | 0.42 |
| Meta-analysis (w/o USOC) | 22,030 | 38,330 | 10.8 | 20.4 | 568 | 984 | 1.03 (0.87 - 1.21) | 0.75 | 1.02 (0.86 - 1.21) | 0.80 |
| **PS-stratification / on-treatment TAR** | | | | | | | | | | |
| CCAE | 10,423 | 14,270 | 6.2 | 10.1 | 318 | 434 | 1.05 (0.90 - 1.23) | 0.53 | 1.08 (0.92 - 1.26) | 0.40 |
| ODOD | 7,141 | 9,264 | 3.7 | 5.5 | 219 | 325 | 0.94 (0.78 - 1.13) | 0.51 | 0.93 (0.77 - 1.12) | 0.48 |
| MDCR | 569 | 1,087 | 0.3 | 0.7 | 25 | 30 | 1.54 (0.86 - 2.75) | 0.14 | 1.35 (0.76 - 2.42) | 0.32 |
| MDCD | 1,097 | 1,536 | 0.6 | 0.8 | 48 | 69 | 0.94 (0.64 - 1.39) | 0.77 | 0.96 (0.65 - 1.43) | 0.82 |
| Open Claims | 160,550 | 201,879 | 99.1 | 149.5 | 2,527 | 3,614 | 1.09 (1.03 - 1.15) | <0.01 | 1.07 (1.01 - 1.14) | 0.06 |
| OEHR | 9,971 | 13,269 | 2.8 | 4.1 | 129 | 157 | 1.09 (0.84 - 1.40) | 0.52 | 0.96 (0.74 - 1.24) | 0.75 |
| Meta-analysis | 189,182 | 240,218 | 112.4 | 170.0 | 3,241 | 4,599 | 1.07 (1.02 - 1.13) | <0.01 | 1.06 (0.99 - 1.12) | 0.09 |
| Meta-analysis (w/o USOC) | 28,632 | 38,339 | 13.3 | 20.4 | 714 | 985 | 1.01 (0.91 - 1.13) | 0.81 | 1.01 (0.90 - 1.12) | 0.82 |
| **PS-matching / on-treatment TAR without escalation** | | | | | | | | | | |
| CCAE | 8,076 | 14,268 | 4.6 | 8.7 | 232 | 372 | 1.14 (0.90 - 1.44) | 0.28 | 1.17 (0.93 - 1.49) | 0.22 |
| ODOD | 5,356 | 9,262 | 2.7 | 4.9 | 158 | 287 | 0.94 (0.72 - 1.23) | 0.66 | 0.92 (0.70 - 1.21) | 0.57 |
| MDCR | 511 | 1,086 | 0.3 | 0.6 | 22 | 28 | 0.82 (0.32 - 2.03) | 0.68 | 0.73 (0.29 - 1.85) | 0.52 |
| MDCD | 937 | 1,533 | 0.5 | 0.6 | 37 | 60 | 0.59 (0.31 - 1.07) | 0.10 | 0.58 (0.31 - 1.09) | 0.10 |
| Open Claims | 121,526 | 201,851 | 72.4 | 126.4 | 1,762 | 2,984 | 1.05 (0.97 - 1.15) | 0.23 | 1.03 (0.94 - 1.12) | 0.59 |
| OEHR | 7,661 | 13,267 | 1.9 | 3.6 | 89 | 131 | 1.15 (0.78 - 1.68) | 0.49 | 1.04 (0.71 - 1.54) | 0.79 |
| Meta-analysis | 143,556 | 240,181 | 82.1 | 144.3 | 2,278 | 3,834 | 1.05 (0.97 - 1.13) | 0.22 | 1.02 (0.94 - 1.12) | 0.62 |
| Meta-analysis (w/o USOC) | 22,030 | 38,330 | 9.7 | 17.8 | 516 | 850 | 1.01 (0.82 - 1.24) | 0.94 | 1.00 (0.81 - 1.23) | 0.87 |
| **PS-stratification / on-treatment TAR without escalation** | | | | | | | | | | |
| CCAE | 10,423 | 14,270 | 5.6 | 8.7 | 283 | 372 | 1.07 (0.90 - 1.26) | 0.46 | 1.11 (0.93 - 1.32) | 0.27 |
| ODOD | 7,141 | 9,264 | 3.4 | 4.9 | 204 | 288 | 0.97 (0.79 - 1.18) | 0.74 | 0.97 (0.80 - 1.19) | 0.79 |
| MDCR | 569 | 1,087 | 0.3 | 0.6 | 24 | 28 | 1.59 (0.87 - 2.90) | 0.13 | 1.43 (0.78 - 2.61) | 0.26 |
| MDCD | 1,097 | 1,536 | 0.5 | 0.6 | 40 | 60 | 0.82 (0.53 - 1.24) | 0.35 | 0.87 (0.57 - 1.33) | 0.54 |
| Open Claims | 160,550 | 201,879 | 89.4 | 126.4 | 2,212 | 2,985 | 1.08 (1.02 - 1.15) | 0.01 | 1.06 (0.99 - 1.13) | 0.15 |
| OEHR | 9,971 | 13,269 | 2.4 | 3.6 | 119 | 131 | 1.20 (0.91 - 1.57) | 0.19 | 1.07 (0.82 - 1.41) | 0.63 |
| Meta-analysis | 189,182 | 240,218 | 101.4 | 144.3 | 2,858 | 3,836 | 1.07 (1.01 - 1.13) | 0.01 | 1.05 (0.99 - 1.13) | 0.12 |
| Meta-analysis (w/o USOC) | 28,632 | 38,339 | 12.0 | 17.8 | 646 | 851 | 1.03 (0.92 - 1.16) | 0.55 | 1.05 (0.94 - 1.18) | 0.42 |

##### 1.1.24 Photosensitivity

**Supplementary Table 24:** Exposures, outcomes and risk estimates for Photosensitivity in GLP1RA (female) (T) vs SGLT2I (female) (C) new-user cohorts under four different propensity score (PS) adjustment / time-at-risk (TAR) designs across data sources. We report total patient exposure time in 1,000 patient-year (KYr) units, uncalibrated and calibrated hazard ratios (HRs) and their 95% confidence intervals (CIs). Greyed out entries did not pass study diagnostics and are included only for completeness.

| | Patients | | Exposure (KYr) | | Outcomes | | Uncalibrated | | Calibrated | |
|---|---|---|---|---|---|---|---|---|---|---|
| | T | C | T | C | T | C | HR (95% CI) | $p$ | HR (95% CI) | $p$ |
| **PS-matching / on-treatment TAR** | | | | | | | | | | |
| CCAE | 10,789 | 18,681 | 7.0 | 13.5 | <5 | 11 | 0.42 (0.06 - 1.78) | 0.31 | 0.42 (0.08 - 2.30) | 0.32 |
| ODOD | 7,449 | 12,733 | 4.2 | 7.8 | 7 | <5 | 4.88 (1.16 - 24.9) | 0.04 | 4.79 (1.04 - 22.2) | 0.05 |
| MDCR | 722 | 1,472 | 0.4 | 0.9 | <5 | <5 | 2.00 (0.08 - 50.5) | 0.67 | 1.75 (0.07 - 44.1) | 0.74 |
| MDCD | 1,473 | 2,411 | 0.8 | 1.3 | <5 | <5 | 1.41 (0.05 - 36.7) | 0.83 | 1.36 (0.05 - 35.4) | 0.85 |
| Open Claims | 168,804 | 273,492 | 113.8 | 202.5 | 42 | 72 | 1.46 (0.85 - 2.50) | 0.17 | 1.44 (0.84 - 2.47) | 0.19 |
| OEHR | 9,639 | 16,533 | 2.8 | 5.2 | 6 | <5 | | 0.20 | | 0.23 |
| Meta-analysis | 168,804 | 273,492 | 113.8 | 202.5 | 42 | 72 | 1.46 (0.85 - 2.50) | 0.17 | 1.44 (0.84 - 2.47) | 0.19 |
| **PS-stratification / on-treatment TAR** | | | | | | | | | | |
| CCAE | 14,215 | 18,681 | 8.7 | 13.5 | 7 | 11 | 0.65 (0.21 - 1.90) | 0.44 | 0.66 (0.22 - 1.99) | 0.46 |
| ODOD | 10,003 | 12,737 | 5.3 | 7.8 | 7 | <5 | 6.44 (1.63 - 31.6) | 0.01 | 6.36 (1.44 - 28.0) | 0.01 |
| MDCR | 797 | 1,473 | 0.5 | 0.9 | <5 | <5 | 2.35 (0.09 - 60.4) | 0.61 | 2.07 (0.08 - 53.1) | 0.66 |
| MDCD | 1,727 | 2,416 | 1.0 | 1.3 | <5 | <5 | 1.45 (0.05 - 38.0) | 0.83 | 1.48 (0.06 - 38.8) | 0.82 |
| Open Claims | 228,019 | 273,499 | 141.9 | 202.5 | 50 | 72 | 1.07 (0.72 - 1.58) | 0.75 | 1.05 (0.71 - 1.56) | 0.81 |
| OEHR | 12,790 | 16,535 | 3.7 | 5.2 | 8 | <5 | 3.46 (0.85 - 17.7) | 0.11 | 3.05 (0.67 - 13.9) | 0.15 |
| Meta-analysis | 242,234 | 292,180 | 150.6 | 216.0 | 57 | 83 | 1.01 (0.69 - 1.46) | 0.97 | 0.99 (0.68 - 1.44) | 0.97 |
| Meta-analysis (w/o USOC) | 14,215 | 18,681 | 8.7 | 13.5 | 7 | 11 | 0.65 (0.21 - 1.90) | 0.44 | 0.65 (0.22 - 1.93) | 0.44 |
| **PS-matching / on-treatment TAR without escalation** | | | | | | | | | | |
| CCAE | 10,789 | 18,681 | 6.3 | 11.7 | <5 | 10 | 0.21 (0.01 - 1.22) | 0.19 | 0.22 (0.02 - 2.26) | 0.20 |
| ODOD | 7,449 | 12,733 | 3.8 | 6.9 | 7 | <5 | 4.72 (1.13 - 24.1) | 0.05 | 4.60 (1.00 - 21.2) | 0.05 |
| MDCR | 722 | 1,472 | 0.4 | 0.8 | <5 | <5 | 2.00 (0.08 - 50.5) | 0.67 | 1.78 (0.07 - 44.9) | 0.73 |
| MDCD | 1,473 | 2,411 | 0.7 | 1.1 | <5 | <5 | 1.41 (0.05 - 36.7) | 0.83 | 1.40 (0.05 - 36.4) | 0.84 |
| Open Claims | 168,804 | 273,492 | 101.7 | 170.2 | 31 | 59 | 1.11 (0.58 - 2.07) | 0.76 | 1.08 (0.57 - 2.05) | 0.82 |
| OEHR | 9,639 | 16,533 | 2.4 | 4.5 | <5 | <5 | 4.00 (0.32 - 98.5) | 0.34 | 3.64 (0.21 - 63.6) | 0.38 |
| Meta-analysis | 168,804 | 273,492 | 101.7 | 170.2 | 31 | 59 | 1.11 (0.58 - 2.07) | 0.76 | 1.08 (0.57 - 2.05) | 0.81 |
| **PS-stratification / on-treatment TAR without escalation** | | | | | | | | | | |
| CCAE | 14,215 | 18,681 | 7.9 | 11.7 | 7 | 10 | 0.78 (0.25 - 2.36) | 0.67 | 0.81 (0.26 - 2.50) | 0.72 |
| ODOD | 10,003 | 12,737 | 4.9 | 6.9 | 7 | <5 | 6.44 (1.63 - 31.6) | 0.01 | 6.50 (1.47 - 28.6) | 0.01 |
| MDCR | 797 | 1,473 | 0.4 | 0.8 | <5 | <5 | 2.23 (0.09 - 57.2) | 0.63 | 2.01 (0.08 - 51.4) | 0.68 |
| MDCD | 1,727 | 2,416 | 0.8 | 1.1 | <5 | <5 | 1.40 (0.05 - 36.8) | 0.84 | 1.49 (0.06 - 39.2) | 0.81 |
| Open Claims | 228,019 | 273,499 | 128.0 | 170.2 | 38 | 59 | 0.99 (0.62 - 1.54) | 0.95 | 0.96 (0.61 - 1.52) | 0.88 |
| OEHR | 12,790 | 16,535 | 3.2 | 4.5 | 5 | <5 | 1.81 (0.36 - 10.8) | 0.50 | 1.62 (0.30 - 8.87) | 0.58 |
| Meta-analysis | 242,234 | 292,180 | 136.0 | 181.9 | 45 | 69 | 0.96 (0.63 - 1.45) | 0.83 | 0.94 (0.62 - 1.43) | 0.78 |
| Meta-analysis (w/o USOC) | 14,215 | 18,681 | 7.9 | 11.7 | 7 | 10 | 0.78 (0.25 - 2.36) | 0.67 | 0.80 (0.26 - 2.45) | 0.69 |

##### 1.1.25 Venous thromboembolism

**Supplementary Table 25:** Exposures, outcomes and risk estimates for Venous thromboembolism in GLP1RA (female) (T) vs SGLT2I (female) (C) new-user cohorts under four different propensity score (PS) adjustment / time-at-risk (TAR) designs across data sources. We report total patient exposure time in 1,000 patient-year (KYr) units, uncalibrated and calibrated hazard ratios (HRs) and their 95% confidence intervals (CIs). Greyed out entries did not pass study diagnostics and are included only for completeness.

| | Patients | | Exposure (KYr) | | Outcomes | | Uncalibrated | | Calibrated | |
|---|---|---|---|---|---|---|---|---|---|---|
| | T | C | T | C | T | C | HR (95% CI) | $p$ | HR (95% CI) | $p$ |
| **PS-matching / on-treatment TAR** | | | | | | | | | | |
| CCAE | 10,610 | 18,464 | 6.9 | 13.4 | 31 | 59 | 1.08 (0.58 - 1.99) | 0.82 | 1.10 (0.59 - 2.05) | 0.76 |
| ODOD | 7,217 | 12,413 | 4.1 | 7.6 | 32 | 60 | 0.77 (0.42 - 1.39) | 0.40 | 0.76 (0.42 - 1.39) | 0.38 |
| MDCR | 704 | 1,425 | 0.4 | 0.9 | 5 | 5 | 1.73 (0.37 - 9.06) | 0.50 | 1.51 (0.31 - 7.47) | 0.62 |
| MDCD | 1,412 | 2,319 | 0.8 | 1.2 | 5 | 10 | 0.56 (0.08 - 2.71) | 0.52 | 0.54 (0.09 - 3.17) | 0.50 |
| Open Claims | 164,239 | 267,171 | 110.9 | 198.1 | 406 | 730 | 0.96 (0.81 - 1.15) | 0.68 | 0.95 (0.80 - 1.13) | 0.56 |
| OEHR | 9,470 | 16,263 | 2.8 | 5.1 | 17 | 38 | 0.67 (0.30 - 1.41) | 0.31 | 0.61 (0.28 - 1.33) | 0.22 |
| Meta-analysis | 191,536 | 314,311 | 124.6 | 224.1 | 486 | 887 | 0.94 (0.81 - 1.10) | 0.46 | 0.93 (0.79 - 1.09) | 0.36 |
| Meta-analysis (w/o USOC) | 27,297 | 47,140 | 13.8 | 26.0 | 80 | 157 | 0.84 (0.58 - 1.23) | 0.38 | 0.84 (0.57 - 1.22) | 0.37 |
| **PS-stratification / on-treatment TAR** | | | | | | | | | | |
| CCAE | 13,935 | 18,464 | 8.5 | 13.4 | 37 | 59 | 0.94 (0.59 - 1.48) | 0.79 | 0.96 (0.61 - 1.52) | 0.86 |
| ODOD | 9,685 | 12,418 | 5.2 | 7.6 | 43 | 60 | 1.05 (0.68 - 1.62) | 0.81 | 1.04 (0.67 - 1.61) | 0.86 |
| MDCR | 778 | 1,426 | 0.4 | 0.9 | 5 | 5 | 1.96 (0.53 - 7.19) | 0.31 | 1.72 (0.47 - 6.32) | 0.41 |
| MDCD | 1,661 | 2,324 | 0.9 | 1.2 | 6 | 10 | 0.86 (0.28 - 2.41) | 0.79 | 0.88 (0.30 - 2.57) | 0.82 |
| Open Claims | 221,626 | 267,177 | 138.1 | 198.1 | 506 | 730 | 1.01 (0.89 - 1.14) | 0.92 | 0.99 (0.87 - 1.12) | 0.87 |
| OEHR | 12,519 | 16,265 | 3.6 | 5.1 | 25 | 38 | 0.92 (0.53 - 1.56) | 0.75 | 0.81 (0.47 - 1.39) | 0.45 |
| Meta-analysis | 257,765 | 314,324 | 155.4 | 224.1 | 611 | 887 | 1.00 (0.90 - 1.12) | 0.98 | 0.99 (0.88 - 1.11) | 0.83 |
| Meta-analysis (w/o USOC) | 36,139 | 47,147 | 17.3 | 26.0 | 105 | 157 | 0.98 (0.74 - 1.28) | 0.86 | 0.97 (0.74 - 1.28) | 0.84 |
| **PS-matching / on-treatment TAR without escalation** | | | | | | | | | | |
| CCAE | 10,610 | 18,464 | 6.2 | 11.5 | 30 | 49 | 0.98 (0.50 - 1.89) | 0.95 | 1.01 (0.52 - 1.96) | 0.94 |
| ODOD | 7,217 | 12,413 | 3.7 | 6.7 | 28 | 53 | 0.72 (0.37 - 1.34) | 0.31 | 0.70 (0.36 - 1.34) | 0.29 |
| MDCR | 704 | 1,425 | 0.4 | 0.8 | <5 | <5 | 1.34 (0.24 - 7.50) | 0.74 | 1.19 (0.21 - 6.65) | 0.84 |
| MDCD | 1,412 | 2,319 | 0.7 | 1.0 | <5 | 7 | 0.40 (0.02 - 2.87) | 0.47 | 0.40 (0.03 - 4.76) | 0.47 |
| Open Claims | 164,239 | 267,171 | 99.1 | 166.7 | 352 | 597 | 0.96 (0.80 - 1.16) | 0.68 | 0.94 (0.78 - 1.13) | 0.51 |
| OEHR | 9,470 | 16,263 | 2.4 | 4.4 | 15 | 32 | 0.80 (0.33 - 1.82) | 0.61 | 0.73 (0.31 - 1.70) | 0.47 |
| Meta-analysis | 191,536 | 314,311 | 111.5 | 189.3 | 425 | 731 | 0.94 (0.79 - 1.11) | 0.45 | 0.91 (0.77 - 1.09) | 0.33 |
| Meta-analysis (w/o USOC) | 27,297 | 47,140 | 12.4 | 22.7 | 73 | 134 | 0.83 (0.55 - 1.24) | 0.36 | 0.82 (0.55 - 1.24) | 0.35 |
| **PS-stratification / on-treatment TAR without escalation** | | | | | | | | | | |
| CCAE | 13,935 | 18,464 | 7.7 | 11.5 | 35 | 49 | 1.06 (0.65 - 1.71) | 0.81 | 1.10 (0.68 - 1.78) | 0.70 |
| ODOD | 9,685 | 12,418 | 4.7 | 6.7 | 39 | 53 | 1.07 (0.67 - 1.69) | 0.78 | 1.08 (0.68 - 1.72) | 0.76 |
| MDCR | 778 | 1,426 | 0.4 | 0.8 | <5 | <5 | 1.59 (0.31 - 7.38) | 0.57 | 1.43 (0.29 - 7.01) | 0.66 |
| MDCD | 1,661 | 2,324 | 0.8 | 1.0 | <5 | 7 | 0.39 (0.06 - 1.67) | 0.27 | 0.41 (0.08 - 2.25) | 0.31 |
| Open Claims | 221,626 | 267,177 | 124.6 | 166.7 | 444 | 597 | 1.02 (0.89 - 1.17) | 0.73 | 1.00 (0.87 - 1.15) | 0.92 |
| OEHR | 12,519 | 16,265 | 3.1 | 4.4 | 22 | 32 | 0.93 (0.51 - 1.66) | 0.80 | 0.83 (0.46 - 1.50) | 0.55 |
| Meta-analysis | 257,765 | 314,324 | 140.2 | 189.3 | 540 | 731 | 1.02 (0.91 - 1.16) | 0.70 | 1.01 (0.89 - 1.15) | 0.87 |
| Meta-analysis (w/o USOC) | 36,139 | 47,147 | 15.6 | 22.7 | 96 | 134 | 1.03 (0.77 - 1.38) | 0.84 | 1.05 (0.78 - 1.40) | 0.77 |

##### 1.1.26 Vomiting

**Supplementary Table 26:** Exposures, outcomes and risk estimates for Vomiting in GLP1RA (female) (T) vs SGLT2I (female) (C) new-user cohorts under four different propensity score (PS) adjustment / time-at-risk (TAR) designs across data sources. We report total patient exposure time in 1,000 patient-year (KYr) units, uncalibrated and calibrated hazard ratios (HRs) and their 95% confidence intervals (CIs). Greyed out entries did not pass study diagnostics and are included only for completeness.

| | Patients | | Exposure (KYr) | | Outcomes | | Uncalibrated | | Calibrated | |
|---|---|---|---|---|---|---|---|---|---|---|
| | T | C | T | C | T | C | HR (95% CI) | $p$ | HR (95% CI) | $p$ |
| **PS-matching / on-treatment TAR** | | | | | | | | | | |
| CCAE | 9,342 | 16,473 | 6.0 | 11.9 | 303 | 353 | 1.64 (1.32 - 2.04) | <0.01 | 1.68 (1.35 - 2.09) | <0.01 |
| ODOD | 6,403 | 11,061 | 3.6 | 6.7 | 215 | 209 | 1.88 (1.45 - 2.46) | <0.01 | 1.85 (1.42 - 2.42) | <0.01 |
| MDCR | 663 | 1,345 | 0.4 | 0.8 | 17 | 21 | 3.02 (1.07 - 9.81) | 0.05 | 2.64 (0.87 - 7.99) | 0.09 |
| MDCD | 1,035 | 1,722 | 0.6 | 0.9 | 63 | 70 | 1.52 (0.95 - 2.43) | 0.08 | 1.46 (0.91 - 2.34) | 0.14 |
| Open Claims | 143,217 | 235,916 | 96.6 | 176.1 | 2,492 | 2,889 | 1.66 (1.54 - 1.78) | <0.01 | 1.63 (1.50 - 1.76) | <0.01 |
| OEHR | 8,617 | 15,026 | 2.5 | 4.6 | 126 | 137 | 2.07 (1.49 - 2.90) | <0.01 | 1.89 (1.35 - 2.65) | <0.01 |
| Meta-analysis | 168,614 | 280,198 | 109.2 | 200.2 | 3,199 | 3,658 | 1.68 (1.57 - 1.80) | <0.01 | 1.65 (1.53 - 1.78) | <0.01 |
| Meta-analysis (w/o USOC) | 25,397 | 44,282 | 12.6 | 24.2 | 707 | 769 | 1.77 (1.53 - 2.04) | <0.01 | 1.76 (1.52 - 2.03) | <0.01 |
| **PS-stratification / on-treatment TAR** | | | | | | | | | | |
| CCAE | 12,234 | 16,475 | 7.4 | 11.9 | 393 | 353 | 1.57 (1.34 - 1.83) | <0.01 | 1.60 (1.36 - 1.88) | <0.01 |
| ODOD | 8,551 | 11,062 | 4.5 | 6.7 | 299 | 210 | 1.94 (1.60 - 2.35) | <0.01 | 1.91 (1.58 - 2.33) | <0.01 |
| MDCR | 743 | 1,346 | 0.4 | 0.8 | 19 | 21 | 1.44 (0.73 - 2.84) | 0.29 | 1.27 (0.64 - 2.50) | 0.50 |
| MDCD | 1,209 | 1,727 | 0.6 | 0.9 | 72 | 71 | 1.55 (1.09 - 2.19) | 0.01 | 1.58 (1.11 - 2.24) | 0.01 |
| Open Claims | 191,085 | 235,920 | 119.1 | 176.1 | 3,332 | 2,889 | 1.55 (1.47 - 1.63) | <0.01 | 1.52 (1.43 - 1.62) | <0.01 |
| OEHR | 11,301 | 15,028 | 3.2 | 4.6 | 169 | 137 | 1.68 (1.32 - 2.15) | <0.01 | 1.48 (1.16 - 1.90) | <0.01 |
| Meta-analysis | 224,380 | 280,212 | 134.8 | 200.2 | 4,265 | 3,660 | 1.62 (1.49 - 1.76) | <0.01 | 1.59 (1.45 - 1.75) | <0.01 |
| Meta-analysis (w/o USOC) | 33,295 | 44,292 | 15.7 | 24.2 | 933 | 771 | 1.69 (1.52 - 1.88) | <0.01 | 1.68 (1.50 - 1.87) | <0.01 |
| **PS-matching / on-treatment TAR without escalation** | | | | | | | | | | |
| CCAE | 9,342 | 16,473 | 5.4 | 10.3 | 279 | 301 | 1.79 (1.42 - 2.25) | <0.01 | 1.84 (1.46 - 2.32) | <0.01 |
| ODOD | 6,403 | 11,061 | 3.3 | 6.0 | 198 | 178 | 2.00 (1.51 - 2.65) | <0.01 | 1.95 (1.46 - 2.58) | <0.01 |
| MDCR | 663 | 1,345 | 0.4 | 0.7 | 16 | 20 | 3.20 (1.01 - 12.1) | 0.07 | 2.84 (0.82 - 9.82) | 0.11 |
| MDCD | 1,035 | 1,722 | 0.5 | 0.8 | 58 | 55 | 1.39 (0.83 - 2.33) | 0.21 | 1.38 (0.82 - 2.31) | 0.25 |
| Open Claims | 143,217 | 235,916 | 86.6 | 148.9 | 2,232 | 2,319 | 1.74 (1.60 - 1.89) | <0.01 | 1.70 (1.56 - 1.85) | <0.01 |
| OEHR | 8,617 | 15,026 | 2.1 | 4.1 | 109 | 112 | 1.95 (1.36 - 2.82) | <0.01 | 1.77 (1.23 - 2.56) | <0.01 |
| Meta-analysis | 168,614 | 280,198 | 97.9 | 170.0 | 2,876 | 2,965 | 1.76 (1.64 - 1.89) | <0.01 | 1.72 (1.58 - 1.87) | <0.01 |
| Meta-analysis (w/o USOC) | 25,397 | 44,282 | 11.3 | 21.1 | 644 | 646 | 1.83 (1.57 - 2.14) | <0.01 | 1.82 (1.56 - 2.13) | <0.01 |
| **PS-stratification / on-treatment TAR without escalation** | | | | | | | | | | |
| CCAE | 12,234 | 16,475 | 6.7 | 10.3 | 363 | 301 | 1.67 (1.41 - 1.98) | <0.01 | 1.73 (1.46 - 2.06) | <0.01 |
| ODOD | 8,551 | 11,062 | 4.1 | 6.0 | 276 | 179 | 2.07 (1.69 - 2.55) | <0.01 | 2.09 (1.70 - 2.57) | <0.01 |
| MDCR | 743 | 1,346 | 0.4 | 0.7 | 17 | 20 | 1.31 (0.65 - 2.62) | 0.45 | 1.18 (0.58 - 2.37) | 0.66 |
| MDCD | 1,209 | 1,727 | 0.6 | 0.8 | 66 | 56 | 1.74 (1.19 - 2.55) | <0.01 | 1.85 (1.26 - 2.72) | <0.01 |
| Open Claims | 191,085 | 235,920 | 107.6 | 148.9 | 3,019 | 2,319 | 1.65 (1.56 - 1.75) | <0.01 | 1.61 (1.51 - 1.72) | <0.01 |
| OEHR | 11,301 | 15,028 | 2.8 | 4.1 | 146 | 112 | 1.80 (1.38 - 2.35) | <0.01 | 1.61 (1.23 - 2.11) | <0.01 |
| Meta-analysis | 224,380 | 280,212 | 121.9 | 170.0 | 3,870 | 2,967 | 1.73 (1.58 - 1.88) | <0.01 | 1.70 (1.55 - 1.87) | <0.01 |
| Meta-analysis (w/o USOC) | 33,295 | 44,292 | 14.2 | 21.1 | 851 | 648 | 1.81 (1.62 - 2.02) | <0.01 | 1.84 (1.64 - 2.06) | <0.01 |

#### 1.2 SGLT2I (female) vs DPP4I (female)

##### 1.2.1 3-point MACE

**Supplementary Table 27:** Exposures, outcomes and risk estimates for 3-point MACE in SGLT2I (female) (T) vs DPP4I (female) (C) new-user cohorts under four different propensity score (PS) adjustment / time-at-risk (TAR) designs across data sources. We report total patient exposure time in 1,000 patient-year (KYr) units, uncalibrated and calibrated hazard ratios (HRs) and their 95% confidence intervals (CIs). Greyed out entries did not pass study diagnostics and are included only for completeness.

| | Patients | | Exposure (KYr) | | Outcomes | | Uncalibrated | | Calibrated | |
|---|---|---|---|---|---|---|---|---|---|---|
| | T | C | T | C | T | C | HR (95% CI) | $p$ | HR (95% CI) | $p$ |
| **PS-matching / on-treatment TAR** | | | | | | | | | | |
| CCAE | 16,056 | 25,285 | 12.1 | 19.0 | 79 | 105 | 1.26 (0.85 - 1.87) | 0.26 | 1.28 (0.86 - 1.91) | 0.23 |
| DAG | 2,939 | 8,792 | 2.4 | 9.3 | 0 | 0 | | | | |
| ODOD | 10,337 | 19,536 | 6.7 | 14.8 | 78 | 306 | 0.82 (0.59 - 1.12) | 0.21 | 0.85 (0.61 - 1.17) | 0.32 |
| MDCR | 1,234 | 4,463 | 0.8 | 3.8 | 10 | 99 | 0.78 (0.32 - 1.70) | 0.55 | 0.76 (0.33 - 1.76) | 0.54 |
| MDCD | 2,197 | 5,957 | 1.2 | 4.2 | 20 | 86 | 0.84 (0.43 - 1.55) | 0.58 | 0.82 (0.43 - 1.55) | 0.54 |
| Open Claims | 222,562 | 424,612 | 175.7 | 370.6 | 1,044 | 3,895 | 0.82 (0.74 - 0.90) | <0.01 | 0.85 (0.74 - 0.98) | 0.03 |
| OEHR | 14,837 | 33,015 | 4.7 | 17.4 | 29 | 142 | 1.81 (1.05 - 3.05) | 0.03 | 1.83 (1.07 - 3.12) | 0.03 |
| SIDIAP | 2,719 | 14,071 | 3.8 | 30.4 | 13 | 94 | 1.83 (0.85 - 3.67) | 0.11 | 1.94 (0.93 - 4.04) | 0.08 |
| Meta-analysis | 268,708 | 522,476 | 204.2 | 456.4 | 1,263 | 4,628 | 1.07 (0.80 - 1.43) | 0.64 | 1.11 (0.83 - 1.50) | 0.48 |
| Meta-analysis (w/o USOC) | 46,146 | 97,864 | 28.5 | 85.7 | 219 | 733 | 1.18 (0.84 - 1.66) | 0.34 | 1.21 (0.86 - 1.70) | 0.28 |
| **PS-stratification / on-treatment TAR** | | | | | | | | | | |
| CCAE | 18,456 | 25,317 | 13.4 | 19.0 | 88 | 105 | 1.33 (0.99 - 1.79) | 0.06 | 1.34 (0.99 - 1.80) | 0.06 |
| DAG | 3,243 | 8,833 | 2.5 | 9.3 | 0 | 0 | | | | |
| ODOD | 12,278 | 19,723 | 7.6 | 15.0 | 85 | 317 | 0.77 (0.59 - 1.00) | 0.05 | 0.79 (0.61 - 1.03) | 0.09 |
| MDCR | 1,395 | 4,506 | 0.9 | 3.8 | 12 | 102 | 0.64 (0.32 - 1.18) | 0.18 | 0.64 (0.33 - 1.25) | 0.20 |
| MDCD | 2,284 | 5,975 | 1.2 | 4.2 | 20 | 89 | 0.84 (0.49 - 1.37) | 0.51 | 0.80 (0.48 - 1.34) | 0.41 |
| Open Claims | 263,915 | 424,925 | 196.3 | 370.9 | 1,135 | 3,909 | 0.79 (0.74 - 0.85) | <0.01 | 0.83 (0.74 - 0.94) | <0.01 |
| OEHR | 16,377 | 34,132 | 5.1 | 17.9 | 31 | 151 | 1.10 (0.71 - 1.66) | 0.67 | 1.16 (0.76 - 1.78) | 0.50 |
| SIDIAP | 2,858 | 14,547 | 3.9 | 31.9 | 13 | 99 | 1.52 (0.78 - 2.76) | 0.19 | 1.52 (0.81 - 2.87) | 0.21 |
| Meta-analysis | 313,884 | 518,644 | 226.2 | 454.7 | 1,352 | 4,581 | 1.00 (0.77 - 1.29) | 0.97 | 1.02 (0.78 - 1.35) | 0.86 |
| Meta-analysis (w/o USOC) | 49,969 | 93,719 | 30.0 | 83.8 | 217 | 672 | 1.10 (0.79 - 1.51) | 0.58 | 1.12 (0.81 - 1.55) | 0.49 |
| **PS-matching / on-treatment TAR without escalation** | | | | | | | | | | |
| CCAE | 16,056 | 25,285 | 10.4 | 15.5 | 63 | 85 | 1.09 (0.70 - 1.69) | 0.69 | 1.08 (0.69 - 1.67) | 0.75 |
| DAG | 2,939 | 8,792 | 2.2 | 8.3 | 0 | 0 | | | | |
| ODOD | 10,337 | 19,536 | 6.0 | 12.5 | 70 | 270 | 0.83 (0.59 - 1.17) | 0.29 | 0.86 (0.61 - 1.21) | 0.40 |
| MDCR | 1,234 | 4,463 | 0.7 | 3.3 | 9 | 78 | 1.07 (0.42 - 2.46) | 0.88 | 1.00 (0.41 - 2.41) | 0.90 |
| MDCD | 2,197 | 5,957 | 1.0 | 3.2 | 12 | 66 | 0.69 (0.29 - 1.49) | 0.37 | 0.67 (0.30 - 1.51) | 0.34 |
| Open Claims | 222,562 | 424,612 | 146.8 | 294.7 | 870 | 3,161 | 0.81 (0.73 - 0.90) | <0.01 | 0.84 (0.75 - 0.94) | 0.01 |
| OEHR | 14,837 | 33,015 | 4.1 | 14.0 | 24 | 94 | 1.77 (0.97 - 3.18) | 0.06 | 1.81 (1.00 - 3.29) | 0.05 |
| SIDIAP | 2,719 | 14,071 | 3.3 | 24.1 | 12 | 77 | 1.65 (0.73 - 3.40) | 0.20 | 1.89 (0.88 - 4.07) | 0.11 |
| Meta-analysis | 268,708 | 522,476 | 171.5 | 363.9 | 1,051 | 3,753 | 0.99 (0.76 - 1.28) | 0.94 | 1.02 (0.78 - 1.34) | 0.88 |
| Meta-analysis (w/o USOC) | 46,146 | 97,864 | 24.7 | 69.2 | 181 | 592 | 1.10 (0.79 - 1.52) | 0.58 | 1.11 (0.80 - 1.54) | 0.53 |
| **PS-stratification / on-treatment TAR without escalation** | | | | | | | | | | |
| CCAE | 18,456 | 25,317 | 11.5 | 15.5 | 71 | 85 | 1.28 (0.92 - 1.79) | 0.14 | 1.24 (0.89 - 1.74) | 0.21 |
| DAG | 3,243 | 8,833 | 2.3 | 8.4 | 0 | 0 | | | | |
| ODOD | 12,278 | 19,723 | 6.7 | 12.6 | 77 | 281 | 0.75 (0.57 - 0.99) | 0.04 | 0.76 (0.57 - 1.00) | 0.05 |
| MDCR | 1,395 | 4,506 | 0.8 | 3.3 | 10 | 80 | 0.75 (0.35 - 1.47) | 0.43 | 0.74 (0.36 - 1.52) | 0.41 |
| MDCD | 2,284 | 5,975 | 1.0 | 3.2 | 12 | 68 | 0.60 (0.30 - 1.09) | 0.12 | 0.57 (0.30 - 1.07) | 0.09 |
| Open Claims | 263,915 | 424,925 | 165.0 | 294.9 | 947 | 3,175 | 0.79 (0.73 - 0.85) | <0.01 | 0.82 (0.74 - 0.91) | <0.01 |
| OEHR | 16,377 | 34,132 | 4.5 | 14.4 | 26 | 99 | 1.30 (0.79 - 2.07) | 0.29 | 1.36 (0.84 - 2.20) | 0.23 |
| SIDIAP | 2,858 | 14,547 | 3.4 | 25.3 | 12 | 79 | 1.42 (0.70 - 2.66) | 0.30 | 1.49 (0.76 - 2.90) | 0.25 |
| Meta-analysis | 313,884 | 518,644 | 191.2 | 362.7 | 1,133 | 3,719 | 0.99 (0.76 - 1.30) | 0.96 | 1.02 (0.76 - 1.35) | 0.91 |
| Meta-analysis (w/o USOC) | 49,969 | 93,719 | 26.1 | 67.8 | 186 | 544 | 1.10 (0.78 - 1.55) | 0.58 | 1.10 (0.79 - 1.55) | 0.57 |

##### 1.2.2 4-point MACE

**Supplementary Table 28:** Exposures, outcomes and risk estimates for 4-point MACE in SGLT2I (female) (T) vs DPP4I (female) (C) new-user cohorts under four different propensity score (PS) adjustment / time-at-risk (TAR) designs across data sources. We report total patient exposure time in 1,000 patient-year (KYr) units, uncalibrated and calibrated hazard ratios (HRs) and their 95% confidence intervals (CIs). Greyed out entries did not pass study diagnostics and are included only for completeness.

| | Patients | | Exposure (KYr) | | Outcomes | | Uncalibrated | | Calibrated | |
|---|---|---|---|---|---|---|---|---|---|---|
| | T | C | T | C | T | C | HR (95% CI) | $p$ | HR (95% CI) | $p$ |
| **PS-matching / on-treatment TAR** | | | | | | | | | | |
| CCAE | 15,870 | 24,934 | 11.9 | 18.7 | 110 | 170 | 1.23 (0.89 - 1.70) | 0.21 | 1.25 (0.90 - 1.74) | 0.20 |
| DAG | 2,939 | 8,792 | 2.4 | 9.3 | 0 | 0 | | | | |
| ODOD | 9,977 | 18,473 | 6.5 | 13.9 | 117 | 500 | 0.69 (0.53 - 0.89) | <0.01 | 0.71 (0.55 - 0.92) | 0.01 |
| MDCR | 1,170 | 4,153 | 0.8 | 3.5 | 18 | 147 | 0.76 (0.36 - 1.46) | 0.44 | 0.74 (0.37 - 1.50) | 0.42 |
| MDCD | 2,075 | 5,549 | 1.1 | 3.8 | 35 | 147 | 0.98 (0.61 - 1.55) | 0.93 | 0.96 (0.60 - 1.53) | 0.83 |
| Open Claims | 217,181 | 410,711 | 171.9 | 357.2 | 1,536 | 6,409 | 0.74 (0.68 - 0.80) | <0.01 | 0.77 (0.68 - 0.88) | <0.01 |
| OEHR | 14,644 | 32,499 | 4.6 | 17.0 | 57 | 284 | 1.11 (0.75 - 1.61) | 0.61 | 1.12 (0.77 - 1.64) | 0.57 |
| SIDIAP | 2,701 | 14,001 | 3.7 | 30.3 | 13 | 118 | 1.40 (0.66 - 2.73) | 0.36 | 1.48 (0.73 - 3.02) | 0.29 |
| Meta-analysis | 262,448 | 506,167 | 199.9 | 440.9 | 1,868 | 7,628 | 0.92 (0.73 - 1.16) | 0.50 | 0.96 (0.75 - 1.22) | 0.73 |
| Meta-analysis (w/o USOC) | 45,267 | 95,456 | 28.0 | 83.7 | 332 | 1,219 | 1.00 (0.76 - 1.32) | 0.99 | 1.03 (0.77 - 1.36) | 0.86 |
| **PS-stratification / on-treatment TAR** | | | | | | | | | | |
| CCAE | 18,202 | 24,965 | 13.2 | 18.7 | 121 | 170 | 1.10 (0.86 - 1.40) | 0.46 | 1.10 (0.86 - 1.41) | 0.46 |
| DAG | 3,243 | 8,833 | 2.5 | 9.3 | 0 | 0 | | | | |
| ODOD | 11,798 | 18,772 | 7.3 | 14.1 | 130 | 528 | 0.66 (0.54 - 0.81) | <0.01 | 0.68 (0.55 - 0.83) | <0.01 |
| MDCR | 1,323 | 4,178 | 0.9 | 3.5 | 20 | 148 | 0.63 (0.37 - 1.03) | 0.08 | 0.64 (0.38 - 1.07) | 0.09 |
| MDCD | 2,158 | 5,565 | 1.2 | 3.8 | 35 | 149 | 0.85 (0.57 - 1.24) | 0.42 | 0.82 (0.55 - 1.20) | 0.31 |
| Open Claims | 255,645 | 410,993 | 191.0 | 357.5 | 1,703 | 6,427 | 0.74 (0.70 - 0.78) | <0.01 | 0.77 (0.69 - 0.87) | <0.01 |
| OEHR | 16,145 | 33,583 | 5.1 | 17.5 | 59 | 306 | 1.01 (0.74 - 1.37) | 0.92 | 1.07 (0.79 - 1.46) | 0.66 |
| SIDIAP | 2,834 | 14,477 | 3.9 | 31.8 | 14 | 127 | 1.31 (0.69 - 2.30) | 0.38 | 1.31 (0.71 - 2.39) | 0.40 |
| Meta-analysis | 304,624 | 502,790 | 220.5 | 439.6 | 2,027 | 7,558 | 0.88 (0.70 - 1.11) | 0.27 | 0.90 (0.70 - 1.16) | 0.42 |
| Meta-analysis (w/o USOC) | 48,979 | 91,797 | 29.4 | 82.1 | 324 | 1,131 | 0.95 (0.69 - 1.29) | 0.73 | 0.97 (0.71 - 1.33) | 0.85 |
| **PS-matching / on-treatment TAR without escalation** | | | | | | | | | | |
| CCAE | 15,870 | 24,934 | 10.3 | 15.2 | 92 | 143 | 1.08 (0.76 - 1.54) | 0.66 | 1.07 (0.75 - 1.52) | 0.73 |
| DAG | 2,939 | 8,792 | 2.2 | 8.3 | 0 | 0 | | | | |
| ODOD | 9,977 | 18,473 | 5.8 | 11.7 | 100 | 427 | 0.65 (0.49 - 0.86) | <0.01 | 0.67 (0.51 - 0.89) | <0.01 |
| MDCR | 1,170 | 4,153 | 0.7 | 3.1 | 16 | 122 | 0.95 (0.44 - 1.89) | 0.89 | 0.89 (0.43 - 1.83) | 0.75 |
| MDCD | 2,075 | 5,549 | 0.9 | 3.0 | 24 | 112 | 0.93 (0.53 - 1.59) | 0.80 | 0.90 (0.52 - 1.57) | 0.73 |
| Open Claims | 217,181 | 410,711 | 143.7 | 284.2 | 1,264 | 5,166 | 0.73 (0.67 - 0.79) | <0.01 | 0.76 (0.69 - 0.83) | <0.01 |
| OEHR | 14,644 | 32,499 | 4.0 | 13.7 | 47 | 194 | 1.22 (0.79 - 1.85) | 0.36 | 1.25 (0.81 - 1.91) | 0.32 |
| SIDIAP | 2,701 | 14,001 | 3.3 | 24.0 | 12 | 95 | 1.41 (0.64 - 2.85) | 0.36 | 1.62 (0.77 - 3.42) | 0.22 |
| Meta-analysis | 262,448 | 506,167 | 168.0 | 351.8 | 1,539 | 6,137 | 0.89 (0.70 - 1.13) | 0.35 | 0.92 (0.72 - 1.18) | 0.51 |
| Meta-analysis (w/o USOC) | 45,267 | 95,456 | 24.3 | 67.6 | 275 | 971 | 0.97 (0.72 - 1.31) | 0.84 | 0.98 (0.73 - 1.33) | 0.91 |
| **PS-stratification / on-treatment TAR without escalation** | | | | | | | | | | |
| CCAE | 18,202 | 24,965 | 11.4 | 15.3 | 102 | 143 | 1.04 (0.80 - 1.36) | 0.76 | 1.01 (0.77 - 1.32) | 0.91 |
| DAG | 3,243 | 8,833 | 2.3 | 8.4 | 0 | 0 | | | | |
| ODOD | 11,798 | 18,772 | 6.5 | 11.9 | 112 | 453 | 0.62 (0.50 - 0.78) | <0.01 | 0.63 (0.50 - 0.79) | <0.01 |
| MDCR | 1,323 | 4,178 | 0.8 | 3.1 | 17 | 122 | 0.72 (0.40 - 1.22) | 0.25 | 0.71 (0.40 - 1.24) | 0.23 |
| MDCD | 2,158 | 5,565 | 1.0 | 3.0 | 24 | 113 | 0.68 (0.42 - 1.05) | 0.10 | 0.64 (0.41 - 1.02) | 0.07 |
| Open Claims | 255,645 | 410,993 | 160.7 | 284.4 | 1,411 | 5,184 | 0.73 (0.68 - 0.77) | <0.01 | 0.76 (0.69 - 0.84) | <0.01 |
| OEHR | 16,145 | 33,583 | 4.4 | 14.1 | 49 | 209 | 1.11 (0.78 - 1.55) | 0.56 | 1.16 (0.82 - 1.63) | 0.43 |
| SIDIAP | 2,834 | 14,477 | 3.4 | 25.2 | 13 | 99 | 1.26 (0.64 - 2.28) | 0.48 | 1.31 (0.70 - 2.48) | 0.42 |
| Meta-analysis | 304,624 | 502,790 | 186.3 | 350.9 | 1,687 | 6,088 | 0.86 (0.67 - 1.11) | 0.26 | 0.89 (0.68 - 1.16) | 0.37 |
| Meta-analysis (w/o USOC) | 48,979 | 91,797 | 25.7 | 66.5 | 276 | 904 | 0.93 (0.66 - 1.32) | 0.70 | 0.94 (0.66 - 1.32) | 0.71 |

##### 1.2.3 Acute myocardial infarction

**Supplementary Table 29:** Exposures, outcomes and risk estimates for Acute myocardial infarction in SGLT2I (female) (T) vs DPP4I (female) (C) new-user cohorts under four different propensity score (PS) adjustment / time-at-risk (TAR) designs across data sources. We report total patient exposure time in 1,000 patient-year (KYr) units, uncalibrated and calibrated hazard ratios (HRs) and their 95% confidence intervals (CIs). Greyed out entries did not pass study diagnostics and are included only for completeness.

| | Patients | | Exposure (KYr) | | Outcomes | | Uncalibrated | | Calibrated | |
|---|---|---|---|---|---|---|---|---|---|---|
| | T | C | T | C | T | C | HR (95% CI) | $p$ | HR (95% CI) | $p$ |
| **PS-matching / on-treatment TAR** | | | | | | | | | | |
| CCAE | 16,247 | 25,644 | 12.2 | 19.2 | 34 | 42 | 1.22 (0.64 - 2.33) | 0.54 | 1.25 (0.65 - 2.38) | 0.51 |
| DAG | 2,939 | 8,792 | 2.4 | 9.3 | 0 | 0 | | | | |
| ODOD | 10,559 | 20,205 | 6.9 | 15.4 | 31 | 106 | 0.83 (0.46 - 1.45) | 0.53 | 0.86 (0.49 - 1.53) | 0.62 |
| MDCR | 1,262 | 4,678 | 0.9 | 4.0 | 7 | 45 | 1.14 (0.36 - 3.01) | 0.81 | 1.12 (0.39 - 3.23) | 0.84 |
| MDCD | 2,273 | 6,190 | 1.2 | 4.3 | 8 | 28 | 1.49 (0.53 - 3.89) | 0.43 | 1.45 (0.54 - 3.93) | 0.47 |
| Open Claims | 227,528 | 436,789 | 179.4 | 382.1 | 488 | 1,651 | 0.84 (0.73 - 0.97) | 0.02 | 0.88 (0.73 - 1.05) | 0.16 |
| OEHR | 14,941 | 33,414 | 4.7 | 17.6 | 15 | 65 | 1.46 (0.66 - 3.11) | 0.34 | 1.48 (0.68 - 3.22) | 0.33 |
| SIDIAP | 2,735 | 14,188 | 3.8 | 30.7 | <5 | 28 | 0.76 (0.11 - 2.92) | 0.74 | 0.80 (0.16 - 4.08) | 0.79 |
| Meta-analysis | 274,283 | 536,430 | 208.3 | 469.4 | <581 | 1,920 | 0.93 (0.75 - 1.14) | 0.47 | 0.96 (0.77 - 1.20) | 0.73 |
| Meta-analysis (w/o USOC) | 46,755 | 99,641 | 28.9 | 87.3 | <93 | 269 | 1.11 (0.78 - 1.56) | 0.57 | 1.13 (0.80 - 1.60) | 0.48 |
| **PS-stratification / on-treatment TAR** | | | | | | | | | | |
| CCAE | 18,670 | 25,678 | 13.5 | 19.3 | 35 | 42 | 1.40 (0.87 - 2.24) | 0.16 | 1.41 (0.88 - 2.25) | 0.16 |
| DAG | 3,243 | 8,833 | 2.5 | 9.3 | 0 | 0 | | | | |
| ODOD | 12,545 | 20,404 | 7.7 | 15.6 | 34 | 108 | 0.96 (0.62 - 1.46) | 0.86 | 0.99 (0.64 - 1.51) | 0.93 |
| MDCR | 1,428 | 4,740 | 0.9 | 4.0 | 7 | 47 | 1.12 (0.43 - 2.52) | 0.80 | 1.14 (0.47 - 2.74) | 0.78 |
| MDCD | 2,362 | 6,208 | 1.3 | 4.4 | 8 | 28 | 1.09 (0.45 - 2.37) | 0.83 | 1.05 (0.46 - 2.39) | 0.91 |
| Open Claims | 269,891 | 437,223 | 200.4 | 382.5 | 548 | 1,660 | 0.88 (0.79 - 0.98) | 0.02 | 0.93 (0.80 - 1.07) | 0.32 |
| OEHR | 16,491 | 34,445 | 5.2 | 18.2 | 17 | 67 | 1.29 (0.69 - 2.28) | 0.41 | 1.36 (0.75 - 2.47) | 0.31 |
| SIDIAP | 2,880 | 14,673 | 3.9 | 32.2 | <5 | 28 | 1.20 (0.33 - 3.40) | 0.76 | 1.20 (0.37 - 3.86) | 0.77 |
| Meta-analysis | 320,477 | 532,423 | 230.7 | 467.7 | <639 | 1,905 | 1.02 (0.81 - 1.27) | 0.87 | 1.05 (0.82 - 1.33) | 0.70 |
| Meta-analysis (w/o USOC) | 50,586 | 95,200 | 30.4 | 85.2 | <91 | 245 | 1.17 (0.89 - 1.54) | 0.25 | 1.20 (0.91 - 1.58) | 0.20 |
| **PS-matching / on-treatment TAR without escalation** | | | | | | | | | | |
| CCAE | 16,247 | 25,644 | 10.5 | 15.7 | 27 | 29 | 1.27 (0.63 - 2.57) | 0.50 | 1.25 (0.62 - 2.54) | 0.54 |
| DAG | 2,939 | 8,792 | 2.2 | 8.3 | 0 | 0 | | | | |
| ODOD | 10,559 | 20,205 | 6.1 | 12.9 | 26 | 96 | 0.69 (0.36 - 1.25) | 0.24 | 0.71 (0.38 - 1.33) | 0.29 |
| MDCR | 1,262 | 4,678 | 0.8 | 3.5 | 6 | 39 | 1.15 (0.36 - 3.16) | 0.80 | 1.08 (0.36 - 3.21) | 0.88 |
| MDCD | 2,273 | 6,190 | 1.0 | 3.4 | 6 | 19 | 1.66 (0.53 - 4.93) | 0.37 | 1.61 (0.53 - 4.91) | 0.40 |
| Open Claims | 227,528 | 436,789 | 149.9 | 303.7 | 406 | 1,333 | 0.85 (0.73 - 1.00) | 0.05 | 0.89 (0.75 - 1.05) | 0.20 |
| OEHR | 14,941 | 33,414 | 4.1 | 14.2 | 14 | 40 | 1.97 (0.79 - 4.78) | 0.14 | 2.01 (0.82 - 4.96) | 0.13 |
| SIDIAP | 2,735 | 14,188 | 3.3 | 24.3 | <5 | 21 | 0.36 (0.02 - 1.99) | 0.39 | 0.42 (0.04 - 4.24) | 0.46 |
| Meta-analysis | 271,548 | 522,242 | 171.6 | 349.9 | 479 | 1,517 | 0.99 (0.72 - 1.36) | 0.97 | 1.03 (0.74 - 1.42) | 0.88 |
| Meta-analysis (w/o USOC) | 44,020 | 85,453 | 21.7 | 46.2 | 73 | 184 | 1.18 (0.72 - 1.92) | 0.51 | 1.20 (0.73 - 1.95) | 0.47 |
| **PS-stratification / on-treatment TAR without escalation** | | | | | | | | | | |
| CCAE | 18,670 | 25,678 | 11.7 | 15.7 | 28 | 29 | 1.66 (0.96 - 2.88) | 0.07 | 1.61 (0.93 - 2.80) | 0.09 |
| DAG | 3,243 | 8,833 | 2.3 | 8.4 | 0 | 0 | | | | |
| ODOD | 12,545 | 20,404 | 6.9 | 13.1 | 29 | 98 | 0.85 (0.53 - 1.33) | 0.49 | 0.86 (0.54 - 1.35) | 0.51 |
| MDCR | 1,428 | 4,740 | 0.8 | 3.5 | 6 | 41 | 1.13 (0.40 - 2.68) | 0.81 | 1.11 (0.43 - 2.86) | 0.83 |
| MDCD | 2,362 | 6,208 | 1.0 | 3.4 | 6 | 19 | 1.18 (0.42 - 2.88) | 0.74 | 1.11 (0.42 - 2.93) | 0.83 |
| Open Claims | 269,891 | 437,223 | 168.5 | 304.1 | 458 | 1,342 | 0.89 (0.79 - 0.99) | 0.04 | 0.93 (0.81 - 1.06) | 0.30 |
| OEHR | 16,491 | 34,445 | 4.5 | 14.6 | 16 | 42 | 1.92 (0.98 - 3.62) | 0.05 | 2.00 (1.04 - 3.87) | 0.04 |
| SIDIAP | 2,880 | 14,673 | 3.4 | 25.5 | <5 | 21 | 0.85 (0.19 - 2.77) | 0.82 | 0.89 (0.23 - 3.43) | 0.87 |
| Meta-analysis | 317,597 | 517,750 | 191.5 | 347.5 | 531 | 1,511 | 1.15 (0.78 - 1.71) | 0.47 | 1.18 (0.79 - 1.76) | 0.41 |
| Meta-analysis (w/o USOC) | 47,706 | 80,527 | 23.0 | 43.4 | 73 | 169 | 1.35 (0.80 - 2.26) | 0.26 | 1.35 (0.80 - 2.27) | 0.26 |

##### 1.2.4 Acute renal failure

**Supplementary Table 30:** Exposures, outcomes and risk estimates for Acute renal failure in SGLT2I (female) (T) vs DPP4I (female) (C) new-user cohorts under four different propensity score (PS) adjustment / time-at-risk (TAR) designs across data sources. We report total patient exposure time in 1,000 patient-year (KYr) units, uncalibrated and calibrated hazard ratios (HRs) and their 95% confidence intervals (CIs). Greyed out entries did not pass study diagnostics and are included only for completeness.

| | Patients | | Exposure (KYr) | | Outcomes | | Uncalibrated | | Calibrated | |
|---|---|---|---|---|---|---|---|---|---|---|
| | T | C | T | C | T | C | HR (95% CI) | $p$ | HR (95% CI) | $p$ |
| **PS-matching / on-treatment TAR** | | | | | | | | | | |
| CCAE | 16,249 | 25,527 | 12.2 | 19.2 | 57 | 138 | 0.69 (0.45 - 1.05) | 0.09 | 0.70 (0.46 - 1.08) | 0.12 |
| DAG | 2,939 | 8,792 | 2.4 | 9.3 | 0 | 0 | | | | |
| ODOD | 10,409 | 19,326 | 6.8 | 14.5 | 109 | 481 | 0.72 (0.54 - 0.96) | 0.03 | 0.75 (0.56 - 1.00) | 0.06 |
| MDCR | 1,266 | 4,529 | 0.9 | 3.8 | 12 | 135 | 0.54 (0.24 - 1.08) | 0.10 | 0.53 (0.25 - 1.12) | 0.10 |
| MDCD | 2,226 | 5,994 | 1.2 | 4.1 | 19 | 141 | 0.36 (0.18 - 0.67) | <0.01 | 0.35 (0.18 - 0.69) | <0.01 |
| Open Claims | 226,364 | 430,258 | 178.7 | 373.9 | 1,226 | 5,473 | 0.72 (0.66 - 0.78) | <0.01 | 0.75 (0.66 - 0.85) | <0.01 |
| OEHR | 14,927 | 33,215 | 4.7 | 17.4 | 36 | 241 | 0.89 (0.55 - 1.39) | 0.62 | 0.90 (0.57 - 1.43) | 0.67 |
| SIDIAP | 2,778 | 14,248 | 3.9 | 30.8 | <5 | 12 | 0.57 (0.03 - 4.06) | 0.66 | 0.60 (0.05 - 7.24) | 0.69 |
| Meta-analysis | 270,175 | 514,320 | 203.7 | 429.2 | 1,447 | 6,474 | 0.71 (0.66 - 0.77) | <0.01 | 0.74 (0.67 - 0.82) | <0.01 |
| Meta-analysis (w/o USOC) | 43,811 | 84,062 | 24.9 | 55.2 | 221 | 1,001 | 0.68 (0.52 - 0.90) | <0.01 | 0.70 (0.53 - 0.92) | 0.01 |
| **PS-stratification / on-treatment TAR** | | | | | | | | | | |
| CCAE | 18,692 | 25,555 | 13.5 | 19.2 | 62 | 139 | 0.68 (0.50 - 0.92) | 0.02 | 0.68 (0.50 - 0.93) | 0.02 |
| DAG | 3,243 | 8,833 | 2.5 | 9.3 | 0 | 0 | | | | |
| ODOD | 12,447 | 19,568 | 7.6 | 14.7 | 120 | 495 | 0.68 (0.54 - 0.83) | <0.01 | 0.69 (0.56 - 0.86) | <0.01 |
| MDCR | 1,438 | 4,575 | 0.9 | 3.8 | 13 | 144 | 0.52 (0.27 - 0.93) | 0.04 | 0.53 (0.28 - 0.98) | 0.05 |
| MDCD | 2,313 | 6,013 | 1.2 | 4.1 | 21 | 141 | 0.52 (0.31 - 0.81) | <0.01 | 0.49 (0.31 - 0.80) | <0.01 |
| Open Claims | 269,448 | 430,532 | 200.1 | 374.2 | 1,386 | 5,492 | 0.68 (0.63 - 0.72) | <0.01 | 0.71 (0.63 - 0.80) | <0.01 |
| OEHR | 16,501 | 34,223 | 5.2 | 17.9 | 39 | 253 | 0.80 (0.55 - 1.14) | 0.23 | 0.85 (0.59 - 1.22) | 0.39 |
| SIDIAP | 2,956 | 14,730 | 4.0 | 32.3 | <5 | 13 | 0.32 (0.02 - 1.92) | 0.35 | 0.32 (0.03 - 3.44) | 0.35 |
| Meta-analysis | 317,088 | 509,878 | 226.5 | 426.0 | 1,607 | 6,379 | 0.68 (0.64 - 0.72) | <0.01 | 0.70 (0.63 - 0.78) | <0.01 |
| Meta-analysis (w/o USOC) | 47,640 | 79,346 | 26.4 | 51.8 | 221 | 887 | 0.70 (0.60 - 0.82) | <0.01 | 0.72 (0.61 - 0.84) | <0.01 |
| **PS-matching / on-treatment TAR without escalation** | | | | | | | | | | |
| CCAE | 16,249 | 25,527 | 10.5 | 15.6 | 48 | 113 | 0.67 (0.42 - 1.06) | 0.09 | 0.66 (0.41 - 1.06) | 0.10 |
| DAG | 2,939 | 8,792 | 2.2 | 8.3 | 0 | 0 | | | | |
| ODOD | 10,409 | 19,326 | 6.0 | 12.3 | 95 | 401 | 0.71 (0.52 - 0.96) | 0.03 | 0.73 (0.54 - 1.00) | 0.06 |
| MDCR | 1,266 | 4,529 | 0.8 | 3.3 | 11 | 114 | 0.60 (0.26 - 1.22) | 0.19 | 0.56 (0.26 - 1.20) | 0.14 |
| MDCD | 2,226 | 5,994 | 1.0 | 3.2 | 13 | 109 | 0.41 (0.19 - 0.82) | 0.02 | 0.40 (0.19 - 0.84) | 0.02 |
| Open Claims | 226,364 | 430,258 | 149.4 | 297.6 | 976 | 4,352 | 0.69 (0.63 - 0.76) | <0.01 | 0.72 (0.65 - 0.80) | <0.01 |
| OEHR | 14,927 | 33,215 | 4.1 | 14.0 | 26 | 165 | 0.87 (0.49 - 1.47) | 0.61 | 0.89 (0.51 - 1.54) | 0.68 |
| SIDIAP | 2,778 | 14,248 | 3.4 | 24.4 | <5 | 12 | 0.55 (0.03 - 3.91) | 0.64 | 0.64 (0.05 - 7.57) | 0.72 |
| Meta-analysis | 270,175 | 514,320 | 171.0 | 342.7 | 1,158 | 5,140 | 0.69 (0.63 - 0.75) | <0.01 | 0.71 (0.64 - 0.79) | <0.01 |
| Meta-analysis (w/o USOC) | 43,811 | 84,062 | 21.6 | 45.1 | 182 | 788 | 0.69 (0.55 - 0.86) | <0.01 | 0.70 (0.56 - 0.88) | <0.01 |
| **PS-stratification / on-treatment TAR without escalation** | | | | | | | | | | |
| CCAE | 18,692 | 25,555 | 11.7 | 15.6 | 53 | 113 | 0.69 (0.49 - 0.96) | 0.03 | 0.67 (0.48 - 0.94) | 0.02 |
| DAG | 3,243 | 8,833 | 2.3 | 8.4 | 0 | 0 | | | | |
| ODOD | 12,447 | 19,568 | 6.8 | 12.4 | 103 | 413 | 0.66 (0.52 - 0.84) | <0.01 | 0.67 (0.53 - 0.84) | <0.01 |
| MDCR | 1,438 | 4,575 | 0.8 | 3.4 | 12 | 120 | 0.54 (0.27 - 0.98) | 0.06 | 0.53 (0.28 - 1.01) | 0.06 |
| MDCD | 2,313 | 6,013 | 1.0 | 3.2 | 14 | 109 | 0.42 (0.23 - 0.72) | <0.01 | 0.40 (0.22 - 0.71) | <0.01 |
| Open Claims | 269,448 | 430,532 | 168.4 | 297.8 | 1,117 | 4,368 | 0.65 (0.60 - 0.70) | <0.01 | 0.68 (0.61 - 0.75) | <0.01 |
| OEHR | 16,501 | 34,223 | 4.5 | 14.5 | 29 | 173 | 0.74 (0.47 - 1.11) | 0.16 | 0.77 (0.50 - 1.18) | 0.25 |
| SIDIAP | 2,956 | 14,730 | 3.5 | 25.6 | <5 | 13 | 0.31 (0.02 - 1.87) | 0.34 | 0.33 (0.03 - 3.51) | 0.36 |
| Meta-analysis | 317,088 | 509,878 | 191.3 | 340.3 | 1,302 | 5,067 | 0.65 (0.61 - 0.70) | <0.01 | 0.67 (0.60 - 0.75) | <0.01 |
| Meta-analysis (w/o USOC) | 47,640 | 79,346 | 23.0 | 42.5 | 185 | 699 | 0.68 (0.57 - 0.81) | <0.01 | 0.68 (0.57 - 0.82) | <0.01 |

##### 1.2.5 Glycemic control

**Supplementary Table 31:** Exposures, outcomes and risk estimates for Glycemic control in SGLT2I (female) (T) vs DPP4I (female) (C) new-user cohorts under four different propensity score (PS) adjustment / time-at-risk (TAR) designs across data sources. We report total patient exposure time in 1,000 patient-year (KYr) units, uncalibrated and calibrated hazard ratios (HRs) and their 95% confidence intervals (CIs). Greyed out entries did not pass study diagnostics and are included only for completeness.

| | Patients | | Exposure (KYr) | | Outcomes | | Uncalibrated | | Calibrated | |
|---|---|---|---|---|---|---|---|---|---|---|
| | T | C | T | C | T | C | HR (95% CI) | $p$ | HR (95% CI) | $p$ |
| **PS-matching / on-treatment TAR** | | | | | | | | | | |
| CCAE | 16,210 | 25,462 | 12.2 | 19.2 | 22 | 41 | 1.04 (0.47 - 2.21) | 0.92 | 1.06 (0.49 - 2.30) | 0.88 |
| DAG | 1,012 | 3,111 | 0.7 | 2.7 | 95 | 380 | 0.84 (0.63 - 1.12) | 0.24 | 0.74 (0.55 - 0.99) | 0.08 |
| ODOD | 6,625 | 12,653 | 3.9 | 8.5 | 403 | 977 | 0.87 (0.73 - 1.03) | 0.11 | 0.90 (0.76 - 1.07) | 0.28 |
| MDCR | 1,294 | 4,793 | 0.9 | 4.1 | 0 | 6 | | | | |
| MDCD | 2,343 | 6,366 | 1.3 | 4.5 | 0 | 0 | | | | |
| Open Claims | 231,830 | 446,127 | 182.9 | 391.2 | 0 | 0 | | | | |
| OEHR | 15,066 | 33,703 | 4.8 | 17.8 | 0 | 0 | | | | |
| SIDIAP | 186 | 964 | 0.2 | 1.2 | 52 | 367 | 0.76 (0.53 - 1.07) | 0.13 | 0.80 (0.57 - 1.15) | 0.26 |
| Meta-analysis | 24,033 | 42,190 | 17.0 | 31.7 | 572 | 1,765 | 0.85 (0.74 - 0.97) | 0.02 | 0.88 (0.76 - 1.03) | 0.11 |
| Meta-analysis (w/o USOC) | 24,033 | 42,190 | 17.0 | 31.7 | 572 | 1,765 | 0.85 (0.74 - 0.97) | 0.02 | 0.87 (0.76 - 1.00) | 0.06 |
| **PS-stratification / on-treatment TAR** | | | | | | | | | | |
| CCAE | 18,683 | 25,497 | 13.5 | 19.2 | 22 | 42 | 1.05 (0.61 - 1.79) | 0.85 | 1.06 (0.61 - 1.82) | 0.84 |
| DAG | 1,073 | 3,152 | 0.7 | 2.8 | 100 | 385 | 0.92 (0.72 - 1.17) | 0.50 | 0.81 (0.63 - 1.03) | 0.12 |
| ODOD | 7,839 | 12,767 | 4.4 | 8.6 | 456 | 984 | 0.95 (0.84 - 1.07) | 0.41 | 0.97 (0.86 - 1.10) | 0.71 |
| MDCR | 1,471 | 4,858 | 0.9 | 4.1 | 0 | 6 | | | | |
| MDCD | 2,435 | 6,385 | 1.3 | 4.5 | 0 | 0 | | | | |
| Open Claims | 276,521 | 446,563 | 205.0 | 391.6 | 0 | 0 | | | | |
| OEHR | 16,666 | 34,746 | 5.2 | 18.4 | 0 | 0 | | | | |
| SIDIAP | 193 | 1,065 | 0.2 | 1.4 | 55 | 418 | 0.78 (0.57 - 1.05) | 0.11 | 0.78 (0.57 - 1.06) | 0.15 |
| Meta-analysis | 27,788 | 42,481 | 18.9 | 32.0 | 633 | 1,829 | 0.93 (0.84 - 1.03) | 0.15 | 0.96 (0.84 - 1.09) | 0.51 |
| Meta-analysis (w/o USOC) | 27,788 | 42,481 | 18.9 | 32.0 | 633 | 1,829 | 0.93 (0.84 - 1.03) | 0.15 | 0.95 (0.86 - 1.05) | 0.38 |
| **PS-matching / on-treatment TAR without escalation** | | | | | | | | | | |
| CCAE | 16,210 | 25,462 | 10.5 | 15.6 | 18 | 36 | 1.09 (0.45 - 2.54) | 0.84 | 1.08 (0.45 - 2.56) | 0.87 |
| DAG | 1,012 | 3,111 | 0.6 | 2.5 | 78 | 334 | 0.75 (0.54 - 1.02) | 0.08 | 0.70 (0.50 - 0.96) | 0.05 |
| ODOD | 6,625 | 12,653 | 3.5 | 7.2 | 349 | 820 | 0.88 (0.73 - 1.06) | 0.18 | 0.91 (0.75 - 1.10) | 0.36 |
| MDCR | 1,294 | 4,793 | 0.8 | 3.6 | 0 | <5 | | | | |
| MDCD | 2,343 | 6,366 | 1.1 | 3.5 | 0 | 0 | | | | |
| Open Claims | 231,830 | 446,127 | 152.7 | 310.8 | 0 | 0 | | | | |
| OEHR | 15,066 | 33,703 | 4.1 | 14.3 | 0 | 0 | | | | |
| SIDIAP | 186 | 964 | 0.2 | 1.0 | 45 | 294 | 0.78 (0.52 - 1.13) | 0.21 | 0.89 (0.61 - 1.32) | 0.60 |
| Meta-analysis | 24,033 | 42,190 | 14.8 | 26.2 | 490 | 1,484 | 0.84 (0.73 - 0.97) | 0.02 | 0.87 (0.74 - 1.02) | 0.09 |
| Meta-analysis (w/o USOC) | 24,033 | 42,190 | 14.8 | 26.2 | 490 | 1,484 | 0.84 (0.73 - 0.97) | 0.02 | 0.85 (0.73 - 0.99) | 0.05 |
| **PS-stratification / on-treatment TAR without escalation** | | | | | | | | | | |
| CCAE | 18,683 | 25,497 | 11.7 | 15.6 | 18 | 37 | 0.92 (0.50 - 1.63) | 0.77 | 0.89 (0.49 - 1.61) | 0.70 |
| DAG | 1,073 | 3,152 | 0.7 | 2.5 | 82 | 339 | 0.84 (0.64 - 1.08) | 0.19 | 0.73 (0.56 - 0.95) | 0.04 |
| ODOD | 7,839 | 12,767 | 3.9 | 7.3 | 397 | 827 | 0.94 (0.82 - 1.06) | 0.31 | 0.94 (0.82 - 1.07) | 0.38 |
| MDCR | 1,471 | 4,858 | 0.8 | 3.6 | 0 | <5 | | | | |
| MDCD | 2,435 | 6,385 | 1.1 | 3.5 | 0 | 0 | | | | |
| Open Claims | 276,521 | 446,563 | 172.3 | 311.2 | 0 | 0 | | | | |
| OEHR | 16,666 | 34,746 | 4.6 | 14.8 | 0 | 0 | | | | |
| SIDIAP | 193 | 1,065 | 0.2 | 1.1 | 48 | 338 | 0.74 (0.53 - 1.02) | 0.08 | 0.78 (0.56 - 1.08) | 0.18 |
| Meta-analysis | 27,788 | 42,481 | 16.4 | 26.5 | 545 | 1,541 | 0.90 (0.80 - 1.00) | 0.04 | 0.92 (0.80 - 1.05) | 0.22 |
| Meta-analysis (w/o USOC) | 27,788 | 42,481 | 16.4 | 26.5 | 545 | 1,541 | 0.90 (0.80 - 1.00) | 0.04 | 0.90 (0.80 - 1.00) | 0.07 |

##### 1.2.6 Hospitalization with heart failure

**Supplementary Table 32:** Exposures, outcomes and risk estimates for Hospitalization with heart failure in SGLT2I (female) (T) vs DPP4I (female) (C) new-user cohorts under four different propensity score (PS) adjustment / time-at-risk (TAR) designs across data sources. We report total patient exposure time in 1,000 patient-year (KYr) units, uncalibrated and calibrated hazard ratios (HRs) and their 95% confidence intervals (CIs). Greyed out entries did not pass study diagnostics and are included only for completeness.

| | Patients | | Exposure (KYr) | | Outcomes | | Uncalibrated | | Calibrated | |
|---|---|---|---|---|---|---|---|---|---|---|
| | T | C | T | C | T | C | HR (95% CI) | $p$ | HR (95% CI) | $p$ |
| **PS-matching / on-treatment TAR** | | | | | | | | | | |
| CCAE | 16,166 | 25,458 | 12.2 | 19.1 | 45 | 96 | 0.91 (0.57 - 1.45) | 0.71 | 0.93 (0.58 - 1.49) | 0.78 |
| DAG | 2,939 | 8,792 | 2.4 | 9.3 | 0 | 0 | | | | |
| ODOD | 10,265 | 19,245 | 6.7 | 14.5 | 68 | 360 | 0.69 (0.49 - 0.95) | 0.02 | 0.71 (0.51 - 0.99) | 0.05 |
| MDCR | 1,218 | 4,401 | 0.8 | 3.7 | 13 | 102 | 0.93 (0.41 - 1.93) | 0.85 | 0.91 (0.42 - 1.99) | 0.82 |
| MDCD | 2,184 | 5,838 | 1.2 | 4.0 | 20 | 96 | 0.88 (0.46 - 1.60) | 0.69 | 0.86 (0.46 - 1.60) | 0.64 |
| Open Claims | 224,538 | 427,200 | 177.7 | 372.7 | 839 | 4,237 | 0.70 (0.63 - 0.78) | <0.01 | 0.73 (0.64 - 0.85) | <0.01 |
| OEHR | 14,834 | 32,983 | 4.7 | 17.3 | 36 | 211 | 0.77 (0.48 - 1.22) | 0.29 | 0.78 (0.49 - 1.26) | 0.33 |
| SIDIAP | 2,760 | 14,201 | 3.9 | 30.8 | <5 | 29 | 0.65 (0.03 - 4.19) | 0.73 | 0.68 (0.06 - 7.84) | 0.76 |
| Meta-analysis | 270,747 | 524,925 | 206.3 | 458.5 | <1,013 | 5,029 | 0.72 (0.65 - 0.78) | <0.01 | 0.74 (0.66 - 0.83) | <0.01 |
| Meta-analysis (w/o USOC) | 46,209 | 97,725 | 28.6 | 85.8 | <174 | 792 | 0.77 (0.62 - 0.96) | 0.02 | 0.79 (0.64 - 0.98) | 0.04 |
| **PS-stratification / on-treatment TAR** | | | | | | | | | | |
| CCAE | 18,562 | 25,492 | 13.5 | 19.2 | 47 | 96 | 0.72 (0.49 - 1.02) | 0.07 | 0.72 (0.50 - 1.03) | 0.08 |
| DAG | 3,243 | 8,833 | 2.5 | 9.3 | 0 | 0 | | | | |
| ODOD | 12,165 | 19,513 | 7.5 | 14.7 | 82 | 378 | 0.63 (0.49 - 0.82) | <0.01 | 0.65 (0.50 - 0.84) | <0.01 |
| MDCR | 1,384 | 4,436 | 0.9 | 3.8 | 14 | 103 | 0.69 (0.36 - 1.23) | 0.24 | 0.70 (0.38 - 1.29) | 0.26 |
| MDCD | 2,266 | 5,856 | 1.2 | 4.1 | 20 | 99 | 0.79 (0.47 - 1.28) | 0.36 | 0.76 (0.46 - 1.25) | 0.28 |
| Open Claims | 265,019 | 427,498 | 197.8 | 373.0 | 960 | 4,257 | 0.67 (0.62 - 0.73) | <0.01 | 0.71 (0.62 - 0.80) | <0.01 |
| OEHR | 16,387 | 34,086 | 5.1 | 17.9 | 36 | 225 | 0.85 (0.57 - 1.23) | 0.41 | 0.90 (0.61 - 1.32) | 0.60 |
| SIDIAP | 2,929 | 14,683 | 4.0 | 32.3 | <5 | 33 | 0.84 (0.18 - 2.77) | 0.81 | 0.84 (0.22 - 3.28) | 0.81 |
| Meta-analysis | 315,062 | 521,272 | 227.9 | 457.0 | <1,130 | 4,989 | 0.68 (0.63 - 0.73) | <0.01 | 0.70 (0.62 - 0.78) | <0.01 |
| Meta-analysis (w/o USOC) | 50,043 | 93,774 | 30.1 | 84.0 | <170 | 732 | 0.70 (0.58 - 0.84) | <0.01 | 0.72 (0.60 - 0.87) | <0.01 |
| **PS-matching / on-treatment TAR without escalation** | | | | | | | | | | |
| CCAE | 16,166 | 25,458 | 10.5 | 15.6 | 40 | 84 | 0.81 (0.48 - 1.32) | 0.41 | 0.80 (0.48 - 1.32) | 0.39 |
| DAG | 2,939 | 8,792 | 2.2 | 8.3 | 0 | 0 | | | | |
| ODOD | 10,265 | 19,245 | 5.9 | 12.2 | 57 | 300 | 0.62 (0.43 - 0.88) | <0.01 | 0.64 (0.45 - 0.92) | 0.02 |
| MDCR | 1,218 | 4,401 | 0.7 | 3.3 | 12 | 84 | 1.18 (0.48 - 2.62) | 0.70 | 1.10 (0.47 - 2.58) | 0.82 |
| MDCD | 2,184 | 5,838 | 1.0 | 3.1 | 14 | 73 | 1.00 (0.47 - 1.99) | 0.99 | 0.96 (0.47 - 1.98) | 0.89 |
| Open Claims | 224,538 | 427,200 | 148.4 | 296.4 | 683 | 3,393 | 0.68 (0.60 - 0.76) | <0.01 | 0.70 (0.62 - 0.79) | <0.01 |
| OEHR | 14,834 | 32,983 | 4.1 | 13.9 | 29 | 148 | 0.91 (0.53 - 1.51) | 0.72 | 0.93 (0.55 - 1.57) | 0.79 |
| SIDIAP | 2,760 | 14,201 | 3.4 | 24.3 | <5 | 23 | 0.63 (0.03 - 4.12) | 0.71 | 0.73 (0.06 - 8.32) | 0.80 |
| Meta-analysis | 267,987 | 510,724 | 169.9 | 341.3 | 823 | 3,998 | 0.69 (0.62 - 0.76) | <0.01 | 0.71 (0.63 - 0.80) | <0.01 |
| Meta-analysis (w/o USOC) | 43,449 | 83,524 | 21.5 | 44.9 | 140 | 605 | 0.75 (0.59 - 0.96) | 0.02 | 0.76 (0.60 - 0.97) | 0.03 |
| **PS-stratification / on-treatment TAR without escalation** | | | | | | | | | | |
| CCAE | 18,562 | 25,492 | 11.6 | 15.6 | 42 | 84 | 0.68 (0.46 - 1.00) | 0.05 | 0.66 (0.45 - 0.97) | 0.04 |
| DAG | 3,243 | 8,833 | 2.3 | 8.4 | 0 | 0 | | | | |
| ODOD | 12,165 | 19,513 | 6.7 | 12.4 | 70 | 317 | 0.59 (0.44 - 0.78) | <0.01 | 0.59 (0.45 - 0.79) | <0.01 |
| MDCR | 1,384 | 4,436 | 0.8 | 3.3 | 13 | 85 | 0.85 (0.43 - 1.58) | 0.64 | 0.84 (0.44 - 1.61) | 0.61 |
| MDCD | 2,266 | 5,856 | 1.0 | 3.2 | 14 | 73 | 0.66 (0.35 - 1.15) | 0.17 | 0.62 (0.34 - 1.13) | 0.13 |
| Open Claims | 265,019 | 427,498 | 166.2 | 296.7 | 794 | 3,410 | 0.66 (0.61 - 0.72) | <0.01 | 0.69 (0.62 - 0.78) | <0.01 |
| OEHR | 16,387 | 34,086 | 4.5 | 14.4 | 29 | 159 | 0.87 (0.55 - 1.32) | 0.52 | 0.91 (0.59 - 1.40) | 0.67 |
| SIDIAP | 2,929 | 14,683 | 3.5 | 25.5 | <5 | 25 | 0.91 (0.19 - 3.14) | 0.90 | 0.96 (0.24 - 3.85) | 0.94 |
| Meta-analysis | 315,062 | 521,272 | 192.5 | 364.6 | <940 | 3,995 | 0.66 (0.62 - 0.72) | <0.01 | 0.68 (0.61 - 0.76) | <0.01 |
| Meta-analysis (w/o USOC) | 50,043 | 93,774 | 26.3 | 68.0 | <146 | 585 | 0.67 (0.55 - 0.82) | <0.01 | 0.67 (0.55 - 0.83) | <0.01 |

##### 1.2.7 Measured renal dysfunction

**Supplementary Table 33:** Exposures, outcomes and risk estimates for Measured renal dysfunction in SGLT2I (female) (T) vs DPP4I (female) (C) new-user cohorts under four different propensity score (PS) adjustment / time-at-risk (TAR) designs across data sources. We report total patient exposure time in 1,000 patient-year (KYr) units, uncalibrated and calibrated hazard ratios (HRs) and their 95% confidence intervals (CIs). Greyed out entries did not pass study diagnostics and are included only for completeness.

| | Patients | | Exposure (KYr) | | Outcomes | | Uncalibrated | | Calibrated | |
|---|---|---|---|---|---|---|---|---|---|---|
| | T | C | T | C | T | C | HR (95% CI) | $p$ | HR (95% CI) | $p$ |
| **PS-matching / on-treatment TAR** | | | | | | | | | | |
| CCAE | 16,380 | 25,894 | 12.3 | 19.5 | <5 | 7 | | | | |
| DAG | 2,939 | 8,792 | 2.4 | 9.3 | 0 | 0 | | | | |
| ODOD | 10,719 | 20,598 | 7.0 | 15.7 | 10 | 41 | 1.14 (0.38 - 3.17) | 0.80 | 1.19 (0.41 - 3.44) | 0.75 |
| MDCR | 1,303 | 4,842 | 0.9 | 4.1 | 0 | 0 | | | | |
| MDCD | 2,343 | 6,366 | 1.3 | 4.5 | 0 | 0 | | | | |
| Open Claims | 231,830 | 446,127 | 182.9 | 391.2 | 0 | 0 | | | | |
| OEHR | 15,066 | 33,703 | 4.8 | 17.8 | 0 | 0 | | | | |
| SIDIAP | 2,772 | 14,209 | 3.9 | 30.7 | 0 | 95 | | | | |
| Meta-analysis | 10,719 | 20,598 | 7.0 | 15.7 | 10 | 41 | 1.14 (0.38 - 3.17) | 0.80 | 1.19 (0.41 - 3.45) | 0.75 |
| Meta-analysis (w/o USOC) | 10,719 | 20,598 | 7.0 | 15.7 | 10 | 41 | 1.14 (0.38 - 3.17) | 0.80 | 1.17 (0.40 - 3.40) | 0.77 |
| **PS-stratification / on-treatment TAR** | | | | | | | | | | |
| CCAE | 18,849 | 25,930 | 13.7 | 19.5 | <5 | 7 | 0.57 (0.08 - 2.47) | 0.51 | 0.57 (0.10 - 3.11) | 0.52 |
| DAG | 3,243 | 8,833 | 2.5 | 9.3 | 0 | 0 | | | | |
| ODOD | 12,834 | 20,806 | 7.9 | 15.9 | 10 | 46 | 0.90 (0.41 - 1.83) | 0.79 | 0.93 (0.44 - 1.96) | 0.84 |
| MDCR | 1,478 | 4,909 | 0.9 | 4.2 | 0 | 0 | | | | |
| MDCD | 2,435 | 6,385 | 1.3 | 4.5 | 0 | 0 | | | | |
| Open Claims | 276,521 | 446,563 | 205.0 | 391.6 | 0 | 0 | | | | |
| OEHR | 16,666 | 34,746 | 5.2 | 18.4 | 0 | 0 | | | | |
| SIDIAP | 2,949 | 14,687 | 4.0 | 32.2 | 0 | 107 | | | | |
| Meta-analysis | 12,834 | 20,806 | 7.9 | 15.9 | 10 | 46 | 0.90 (0.41 - 1.83) | 0.79 | 0.93 (0.44 - 1.97) | 0.85 |
| Meta-analysis (w/o USOC) | 12,834 | 20,806 | 7.9 | 15.9 | 10 | 46 | 0.90 (0.41 - 1.83) | 0.79 | 0.93 (0.44 - 1.96) | 0.84 |
| **PS-matching / on-treatment TAR without escalation** | | | | | | | | | | |
| CCAE | 16,380 | 25,894 | 10.6 | 15.9 | <5 | 6 | | | | |
| DAG | 2,939 | 8,792 | 2.2 | 8.3 | 0 | 0 | | | | |
| ODOD | 10,719 | 20,598 | 6.2 | 13.2 | 8 | 33 | 1.38 (0.40 - 4.36) | 0.60 | 1.42 (0.43 - 4.72) | 0.56 |
| MDCR | 1,303 | 4,842 | 0.8 | 3.6 | 0 | 0 | | | | |
| MDCD | 2,343 | 6,366 | 1.1 | 3.5 | 0 | 0 | | | | |
| Open Claims | 231,830 | 446,127 | 152.7 | 310.8 | 0 | 0 | | | | |
| OEHR | 15,066 | 33,703 | 4.1 | 14.3 | 0 | 0 | | | | |
| SIDIAP | 2,772 | 14,209 | 3.4 | 24.3 | 0 | 80 | | | | |
| Meta-analysis | 10,719 | 20,598 | 6.2 | 13.2 | 8 | 33 | 1.38 (0.40 - 4.36) | 0.60 | 1.42 (0.43 - 4.72) | 0.57 |
| Meta-analysis (w/o USOC) | 10,719 | 20,598 | 6.2 | 13.2 | 8 | 33 | 1.38 (0.40 - 4.36) | 0.60 | 1.40 (0.42 - 4.64) | 0.58 |
| **PS-stratification / on-treatment TAR without escalation** | | | | | | | | | | |
| CCAE | 18,849 | 25,930 | 11.8 | 15.9 | <5 | 6 | 0.59 (0.08 - 2.74) | 0.56 | 0.58 (0.10 - 3.29) | 0.54 |
| DAG | 3,243 | 8,833 | 2.3 | 8.4 | 0 | 0 | | | | |
| ODOD | 12,834 | 20,806 | 7.0 | 13.3 | 8 | 38 | 0.90 (0.37 - 1.96) | 0.80 | 0.90 (0.39 - 2.08) | 0.81 |
| MDCR | 1,478 | 4,909 | 0.8 | 3.6 | 0 | 0 | | | | |
| MDCD | 2,435 | 6,385 | 1.1 | 3.5 | 0 | 0 | | | | |
| Open Claims | 276,521 | 446,563 | 172.3 | 311.2 | 0 | 0 | | | | |
| OEHR | 16,666 | 34,746 | 4.6 | 14.8 | 0 | 0 | | | | |
| SIDIAP | 2,949 | 14,687 | 3.5 | 25.5 | 0 | 89 | | | | |
| Meta-analysis | 12,834 | 20,806 | 7.0 | 13.3 | 8 | 38 | 0.90 (0.37 - 1.96) | 0.80 | 0.92 (0.40 - 2.13) | 0.84 |
| Meta-analysis (w/o USOC) | 12,834 | 20,806 | 7.0 | 13.3 | 8 | 38 | 0.90 (0.37 - 1.96) | 0.80 | 0.90 (0.39 - 2.07) | 0.81 |

##### 1.2.8 Stroke

**Supplementary Table 34:** Exposures, outcomes and risk estimates for Stroke in SGLT2I (female) (T) vs DPP4I (female) (C) new-user cohorts under four different propensity score (PS) adjustment / time-at-risk (TAR) designs across data sources. We report total patient exposure time in 1,000 patient-year (KYr) units, uncalibrated and calibrated hazard ratios (HRs) and their 95% confidence intervals (CIs). Greyed out entries did not pass study diagnostics and are included only for completeness.

| | Patients | | Exposure (KYr) | | Outcomes | | Uncalibrated | | Calibrated | |
|---|---|---|---|---|---|---|---|---|---|---|
| | T | C | T | C | T | C | HR (95% CI) | $p$ | HR (95% CI) | $p$ |
| **PS-matching / on-treatment TAR** | | | | | | | | | | |
| CCAE | 16,223 | 25,555 | 12.2 | 19.2 | 44 | 69 | 1.14 (0.69 - 1.86) | 0.60 | 1.16 (0.71 - 1.90) | 0.56 |
| DAG | 2,939 | 8,792 | 2.4 | 9.3 | 0 | 0 | | | | |
| ODOD | 10,508 | 19,921 | 6.8 | 15.2 | 50 | 198 | 0.87 (0.58 - 1.27) | 0.47 | 0.90 (0.60 - 1.33) | 0.60 |
| MDCR | 1,273 | 4,612 | 0.9 | 3.9 | <5 | 63 | 0.48 (0.11 - 1.52) | 0.28 | 0.47 (0.13 - 1.78) | 0.27 |
| MDCD | 2,255 | 6,115 | 1.2 | 4.3 | 12 | 61 | 0.75 (0.31 - 1.70) | 0.52 | 0.74 (0.31 - 1.74) | 0.49 |
| Open Claims | 226,598 | 433,211 | 179.1 | 379.0 | 562 | 2,320 | 0.77 (0.68 - 0.88) | <0.01 | 0.81 (0.69 - 0.95) | 0.01 |
| OEHR | 14,961 | 33,293 | 4.7 | 17.6 | 13 | 74 | 1.40 (0.65 - 2.90) | 0.38 | 1.42 (0.67 - 3.01) | 0.36 |
| SIDIAP | 2,762 | 14,143 | 3.8 | 30.6 | 10 | 72 | 2.09 (0.91 - 4.44) | 0.07 | 2.21 (1.00 - 4.90) | 0.05 |
| Meta-analysis | 273,307 | 532,238 | 207.9 | 465.8 | 691 | 2,794 | 0.97 (0.75 - 1.27) | 0.85 | 1.01 (0.77 - 1.33) | 0.93 |
| Meta-analysis (w/o USOC) | 46,709 | 99,027 | 28.8 | 86.8 | 129 | 474 | 1.10 (0.81 - 1.48) | 0.55 | 1.12 (0.83 - 1.52) | 0.46 |
| **PS-stratification / on-treatment TAR** | | | | | | | | | | |
| CCAE | 18,673 | 25,589 | 13.5 | 19.3 | 51 | 69 | 1.17 (0.80 - 1.72) | 0.41 | 1.18 (0.80 - 1.73) | 0.40 |
| DAG | 3,243 | 8,833 | 2.5 | 9.3 | 0 | 0 | | | | |
| ODOD | 12,563 | 20,120 | 7.7 | 15.3 | 53 | 206 | 0.77 (0.55 - 1.06) | 0.12 | 0.79 (0.56 - 1.09) | 0.16 |
| MDCR | 1,442 | 4,659 | 0.9 | 3.9 | 5 | 64 | 0.38 (0.13 - 0.93) | 0.06 | 0.39 (0.14 - 1.05) | 0.06 |
| MDCD | 2,344 | 6,133 | 1.3 | 4.3 | 12 | 62 | 0.67 (0.34 - 1.23) | 0.23 | 0.64 (0.33 - 1.23) | 0.18 |
| Open Claims | 270,188 | 433,555 | 200.6 | 379.4 | 602 | 2,328 | 0.73 (0.66 - 0.80) | <0.01 | 0.76 (0.66 - 0.88) | <0.01 |
| OEHR | 16,552 | 34,421 | 5.2 | 18.1 | 14 | 79 | 0.98 (0.51 - 1.78) | 0.95 | 1.04 (0.55 - 1.94) | 0.91 |
| SIDIAP | 2,932 | 14,618 | 4.0 | 32.1 | 10 | 77 | 1.77 (0.82 - 3.47) | 0.12 | 1.77 (0.86 - 3.65) | 0.13 |
| Meta-analysis | 320,908 | 528,303 | 231.0 | 464.1 | 730 | 2,759 | 0.93 (0.70 - 1.22) | 0.58 | 0.95 (0.71 - 1.27) | 0.74 |
| Meta-analysis (w/o USOC) | 50,720 | 94,748 | 30.4 | 84.8 | 128 | 431 | 1.04 (0.75 - 1.44) | 0.82 | 1.06 (0.77 - 1.47) | 0.71 |
| **PS-matching / on-treatment TAR without escalation** | | | | | | | | | | |
| CCAE | 16,223 | 25,555 | 10.5 | 15.7 | 35 | 60 | 0.87 (0.49 - 1.52) | 0.64 | 0.86 (0.48 - 1.52) | 0.61 |
| DAG | 2,939 | 8,792 | 2.2 | 8.3 | 0 | 0 | | | | |
| ODOD | 10,508 | 19,921 | 6.0 | 12.7 | 47 | 177 | 0.91 (0.59 - 1.39) | 0.68 | 0.94 (0.62 - 1.45) | 0.80 |
| MDCR | 1,273 | 4,612 | 0.8 | 3.4 | <5 | 48 | 0.77 (0.17 - 2.70) | 0.72 | 0.72 (0.18 - 2.92) | 0.65 |
| MDCD | 2,255 | 6,115 | 1.0 | 3.3 | 6 | 52 | 0.34 (0.08 - 1.04) | 0.10 | 0.33 (0.09 - 1.20) | 0.10 |
| Open Claims | 226,598 | 433,211 | 149.5 | 301.2 | 470 | 1,875 | 0.76 (0.66 - 0.87) | <0.01 | 0.79 (0.68 - 0.91) | <0.01 |
| OEHR | 14,961 | 33,293 | 4.1 | 14.1 | 9 | 53 | 0.99 (0.39 - 2.27) | 0.98 | 1.01 (0.42 - 2.43) | 0.94 |
| SIDIAP | 2,762 | 14,143 | 3.4 | 24.2 | 10 | 61 | 2.18 (0.94 - 4.70) | 0.06 | 2.49 (1.11 - 5.59) | 0.03 |
| Meta-analysis | 273,307 | 532,238 | 174.5 | 371.3 | 577 | 2,278 | 0.86 (0.68 - 1.09) | 0.21 | 0.89 (0.69 - 1.13) | 0.34 |
| Meta-analysis (w/o USOC) | 46,709 | 99,027 | 25.0 | 70.0 | 107 | 403 | 0.98 (0.65 - 1.47) | 0.91 | 0.99 (0.66 - 1.49) | 0.94 |
| **PS-stratification / on-treatment TAR without escalation** | | | | | | | | | | |
| CCAE | 18,673 | 25,589 | 11.7 | 15.7 | 41 | 60 | 1.04 (0.68 - 1.57) | 0.87 | 1.01 (0.66 - 1.53) | 0.94 |
| DAG | 3,243 | 8,833 | 2.3 | 8.4 | 0 | 0 | | | | |
| ODOD | 12,563 | 20,120 | 6.8 | 12.9 | 50 | 185 | 0.79 (0.55 - 1.10) | 0.17 | 0.79 (0.56 - 1.11) | 0.18 |
| MDCR | 1,442 | 4,659 | 0.8 | 3.4 | <5 | 48 | 0.47 (0.13 - 1.25) | 0.19 | 0.46 (0.15 - 1.41) | 0.18 |
| MDCD | 2,344 | 6,133 | 1.0 | 3.3 | 6 | 52 | 0.35 (0.13 - 0.77) | 0.02 | 0.33 (0.14 - 0.80) | 0.01 |
| Open Claims | 270,188 | 433,555 | 168.7 | 301.5 | 505 | 1,883 | 0.72 (0.65 - 0.80) | <0.01 | 0.75 (0.66 - 0.86) | <0.01 |
| OEHR | 16,552 | 34,421 | 4.5 | 14.6 | 9 | 56 | 0.81 (0.35 - 1.67) | 0.60 | 0.85 (0.39 - 1.84) | 0.68 |
| SIDIAP | 2,932 | 14,618 | 3.5 | 25.4 | 10 | 63 | 1.87 (0.85 - 3.73) | 0.10 | 1.95 (0.93 - 4.09) | 0.08 |
| Meta-analysis | 320,908 | 528,303 | 195.2 | 370.1 | 615 | 2,247 | 0.87 (0.68 - 1.12) | 0.28 | 0.89 (0.68 - 1.16) | 0.39 |
| Meta-analysis (w/o USOC) | 50,720 | 94,748 | 26.5 | 68.6 | 110 | 364 | 0.98 (0.71 - 1.36) | 0.92 | 0.99 (0.71 - 1.36) | 0.92 |

##### 1.2.9 Sudden cardiac death

**Supplementary Table 35:** Exposures, outcomes and risk estimates for Sudden cardiac death in SGLT2I (female) (T) vs DPP4I (female) (C) new-user cohorts under four different propensity score (PS) adjustment / time-at-risk (TAR) designs across data sources. We report total patient exposure time in 1,000 patient-year (KYr) units, uncalibrated and calibrated hazard ratios (HRs) and their 95% confidence intervals (CIs). Greyed out entries did not pass study diagnostics and are included only for completeness.

| | Patients | | Exposure (KYr) | | Outcomes | | Uncalibrated | | Calibrated | |
|---|---|---|---|---|---|---|---|---|---|---|
| | T | C | T | C | T | C | HR (95% CI) | $p$ | HR (95% CI) | $p$ |
| **PS-matching / on-treatment TAR** | | | | | | | | | | |
| CCAE | 16,402 | 25,916 | 12.3 | 19.5 | 7 | 11 | 1.08 (0.34 - 3.32) | 0.89 | 1.10 (0.35 - 3.44) | 0.86 |
| DAG | 2,939 | 8,792 | 2.4 | 9.3 | 0 | 0 | | | | |
| ODOD | 10,719 | 20,603 | 7.0 | 15.7 | 5 | 54 | 0.71 (0.23 - 1.91) | 0.54 | 0.74 (0.25 - 2.16) | 0.58 |
| MDCR | 1,303 | 4,830 | 0.9 | 4.1 | 0 | 5 | | | | |
| MDCD | 2,341 | 6,354 | 1.3 | 4.4 | <5 | 14 | 0.60 (0.09 - 2.43) | 0.54 | 0.59 (0.11 - 3.05) | 0.53 |
| Open Claims | 231,530 | 445,456 | 182.6 | 390.5 | 102 | 475 | 0.78 (0.58 - 1.04) | 0.09 | 0.81 (0.59 - 1.10) | 0.19 |
| OEHR | 15,059 | 33,690 | 4.8 | 17.8 | <5 | 17 | 3.72 (0.58 - 21.0) | 0.15 | 3.77 (0.63 - 22.6) | 0.15 |
| SIDIAP | 2,778 | 14,273 | 3.9 | 30.9 | 0 | 0 | | | | |
| Meta-analysis | 273,710 | 525,665 | 206.7 | 443.5 | <119 | 557 | 0.82 (0.62 - 1.07) | 0.14 | 0.85 (0.64 - 1.12) | 0.25 |
| Meta-analysis (w/o USOC) | 42,180 | 80,209 | 24.1 | 53.1 | <17 | 82 | 1.13 (0.51 - 2.51) | 0.76 | 1.16 (0.52 - 2.57) | 0.72 |
| **PS-stratification / on-treatment TAR** | | | | | | | | | | |
| CCAE | 18,867 | 25,952 | 13.7 | 19.5 | 8 | 11 | 1.07 (0.40 - 2.78) | 0.89 | 1.08 (0.41 - 2.85) | 0.88 |
| DAG | 3,243 | 8,833 | 2.5 | 9.3 | 0 | 0 | | | | |
| ODOD | 12,828 | 20,812 | 7.9 | 15.9 | 6 | 56 | 0.36 (0.13 - 0.82) | 0.03 | 0.37 (0.15 - 0.92) | 0.03 |
| MDCR | 1,476 | 4,897 | 0.9 | 4.2 | 0 | 5 | | | | |
| MDCD | 2,433 | 6,373 | 1.3 | 4.5 | <5 | 17 | 1.04 (0.24 - 3.23) | 0.95 | 0.99 (0.27 - 3.67) | 0.96 |
| Open Claims | 276,028 | 445,883 | 204.6 | 390.9 | 121 | 476 | 0.67 (0.53 - 0.83) | <0.01 | 0.70 (0.55 - 0.89) | <0.01 |
| OEHR | 16,657 | 34,732 | 5.2 | 18.4 | <5 | 19 | 1.47 (0.39 - 4.41) | 0.53 | 1.56 (0.46 - 5.24) | 0.48 |
| SIDIAP | 2,955 | 14,757 | 4.0 | 32.4 | 0 | 0 | | | | |
| Meta-analysis | 324,380 | 527,379 | 231.4 | 444.7 | <140 | 562 | 0.68 (0.55 - 0.83) | <0.01 | 0.70 (0.56 - 0.87) | <0.01 |
| Meta-analysis (w/o USOC) | 48,352 | 81,496 | 26.8 | 53.8 | <19 | 86 | 0.78 (0.33 - 1.84) | 0.57 | 0.80 (0.34 - 1.88) | 0.61 |
| **PS-matching / on-treatment TAR without escalation** | | | | | | | | | | |
| CCAE | 16,402 | 25,916 | 10.6 | 15.9 | 6 | 7 | 0.81 (0.21 - 2.88) | 0.76 | 0.80 (0.21 - 2.99) | 0.74 |
| DAG | 2,939 | 8,792 | 2.2 | 8.3 | 0 | 0 | | | | |
| ODOD | 10,719 | 20,603 | 6.2 | 13.2 | 5 | 42 | 0.75 (0.23 - 2.09) | 0.61 | 0.78 (0.26 - 2.33) | 0.66 |
| MDCR | 1,303 | 4,830 | 0.8 | 3.6 | 0 | 5 | | | | |
| MDCD | 2,341 | 6,354 | 1.1 | 3.5 | <5 | 8 | 0.58 (0.03 - 3.65) | 0.65 | 0.56 (0.05 - 6.19) | 0.64 |
| Open Claims | 231,530 | 445,456 | 152.4 | 310.3 | 88 | 387 | 0.80 (0.58 - 1.09) | 0.16 | 0.83 (0.60 - 1.14) | 0.27 |
| OEHR | 15,059 | 33,690 | 4.1 | 14.3 | <5 | 10 | | 0.12 | | 0.11 |
| SIDIAP | 2,778 | 14,273 | 3.4 | 24.4 | 0 | 0 | | | | |
| Meta-analysis | 242,249 | 466,059 | 158.6 | 323.5 | 93 | 429 | 0.80 (0.59 - 1.08) | 0.14 | 0.82 (0.60 - 1.12) | 0.21 |
| Meta-analysis (w/o USOC) | 10,719 | 20,603 | 6.2 | 13.2 | 5 | 42 | 0.75 (0.23 - 2.09) | 0.61 | 0.77 (0.26 - 2.29) | 0.63 |
| **PS-stratification / on-treatment TAR without escalation** | | | | | | | | | | |
| CCAE | 18,867 | 25,952 | 11.8 | 15.9 | 7 | 7 | 1.33 (0.43 - 4.10) | 0.62 | 1.29 (0.42 - 3.97) | 0.66 |
| DAG | 3,243 | 8,833 | 2.3 | 8.4 | 0 | 0 | | | | |
| ODOD | 12,828 | 20,812 | 7.0 | 13.4 | 6 | 44 | 0.38 (0.14 - 0.88) | 0.04 | 0.38 (0.15 - 0.96) | 0.04 |
| MDCR | 1,476 | 4,897 | 0.8 | 3.6 | 0 | 5 | | | | |
| MDCD | 2,433 | 6,373 | 1.1 | 3.5 | <5 | 10 | 1.52 (0.33 - 5.20) | 0.55 | 1.44 (0.36 - 5.70) | 0.60 |
| Open Claims | 276,028 | 445,883 | 172.0 | 310.7 | 105 | 388 | 0.69 (0.54 - 0.87) | <0.01 | 0.72 (0.56 - 0.92) | 0.01 |
| OEHR | 16,657 | 34,732 | 4.5 | 14.7 | <5 | 10 | 2.14 (0.52 - 7.32) | 0.26 | 2.23 (0.60 - 8.37) | 0.23 |
| SIDIAP | 2,955 | 14,757 | 3.5 | 25.6 | 0 | 0 | | | | |
| Meta-analysis | 288,856 | 466,695 | 179.0 | 324.0 | 111 | 432 | 0.61 (0.38 - 0.97) | 0.04 | 0.62 (0.38 - 1.00) | 0.05 |
| Meta-analysis (w/o USOC) | 12,828 | 20,812 | 7.0 | 13.4 | 6 | 44 | 0.38 (0.14 - 0.88) | 0.04 | 0.38 (0.15 - 0.96) | 0.04 |

##### 1.2.10 Abnormal weight gain

**Supplementary Table 36:** Exposures, outcomes and risk estimates for Abnormal weight gain in SGLT2I (female) (T) vs DPP4I (female) (C) new-user cohorts under four different propensity score (PS) adjustment / time-at-risk (TAR) designs across data sources. We report total patient exposure time in 1,000 patient-year (KYr) units, uncalibrated and calibrated hazard ratios (HRs) and their 95% confidence intervals (CIs). Greyed out entries did not pass study diagnostics and are included only for completeness.

| | Patients | | Exposure (KYr) | | Outcomes | | Uncalibrated | | Calibrated | |
|---|---|---|---|---|---|---|---|---|---|---|
| | T | C | T | C | T | C | HR (95% CI) | $p$ | HR (95% CI) | $p$ |
| **PS-matching / on-treatment TAR** | | | | | | | | | | |
| CCAE | 14,880 | 23,938 | 11.2 | 18.0 | 74 | 144 | 0.85 (0.57 - 1.26) | 0.42 | 0.87 (0.58 - 1.29) | 0.50 |
| DAG | 2,894 | 8,701 | 2.3 | 9.2 | <5 | 13 | 1.04 (0.14 - 4.91) | 0.96 | 0.91 (0.16 - 5.33) | 0.91 |
| ODOD | 9,959 | 19,506 | 6.4 | 14.8 | 45 | 114 | 0.77 (0.46 - 1.25) | 0.30 | 0.79 (0.48 - 1.31) | 0.37 |
| MDCR | 1,274 | 4,750 | 0.9 | 4.0 | <5 | 14 | 1.46 (0.20 - 6.86) | 0.67 | 1.43 (0.25 - 8.30) | 0.69 |
| MDCD | 2,097 | 5,787 | 1.1 | 4.0 | 5 | 41 | 0.39 (0.09 - 1.20) | 0.15 | 0.38 (0.10 - 1.39) | 0.15 |
| Open Claims | 215,276 | 422,234 | 169.3 | 370.3 | 427 | 1,112 | 0.80 (0.69 - 0.94) | <0.01 | 0.84 (0.69 - 1.01) | 0.07 |
| OEHR | 14,026 | 31,802 | 4.4 | 16.7 | 25 | 145 | 0.58 (0.30 - 1.07) | 0.10 | 0.59 (0.31 - 1.12) | 0.11 |
| SIDIAP | 2,750 | 14,120 | 3.8 | 30.5 | <5 | 28 | 0.91 (0.13 - 3.70) | 0.91 | 0.96 (0.18 - 5.09) | 0.95 |
| Meta-analysis | 258,988 | 517,387 | 196.3 | 454.4 | <581 | 1,584 | 0.79 (0.69 - 0.90) | <0.01 | 0.82 (0.70 - 0.95) | 0.01 |
| Meta-analysis (w/o USOC) | 43,712 | 95,153 | 27.0 | 84.1 | <154 | 472 | 0.75 (0.57 - 0.98) | 0.03 | 0.76 (0.58 - 1.00) | 0.06 |
| **PS-stratification / on-treatment TAR** | | | | | | | | | | |
| CCAE | 17,049 | 23,974 | 12.4 | 18.0 | 80 | 145 | 0.78 (0.59 - 1.04) | 0.09 | 0.78 (0.59 - 1.05) | 0.11 |
| DAG | 3,193 | 8,742 | 2.5 | 9.2 | <5 | 13 | 1.89 (0.46 - 6.25) | 0.34 | 1.65 (0.45 - 6.08) | 0.45 |
| ODOD | 11,835 | 19,719 | 7.2 | 15.0 | 54 | 114 | 0.82 (0.57 - 1.16) | 0.28 | 0.84 (0.59 - 1.20) | 0.35 |
| MDCR | 1,444 | 4,816 | 0.9 | 4.1 | <5 | 14 | 1.13 (0.17 - 4.23) | 0.88 | 1.15 (0.23 - 5.65) | 0.87 |
| MDCD | 2,177 | 5,803 | 1.2 | 4.0 | 5 | 41 | 0.37 (0.13 - 0.89) | 0.05 | 0.36 (0.13 - 0.95) | 0.04 |
| Open Claims | 255,345 | 422,689 | 188.9 | 370.8 | 486 | 1,116 | 0.86 (0.77 - 0.97) | 0.01 | 0.90 (0.78 - 1.05) | 0.21 |
| OEHR | 15,455 | 32,773 | 4.8 | 17.2 | 29 | 150 | 0.71 (0.46 - 1.08) | 0.12 | 0.76 (0.49 - 1.16) | 0.21 |
| SIDIAP | 2,925 | 14,600 | 4.0 | 32.0 | <5 | 29 | 0.81 (0.12 - 3.01) | 0.80 | 0.81 (0.16 - 4.02) | 0.80 |
| Meta-analysis | 302,609 | 513,755 | 217.3 | 453.0 | <654 | 1,554 | 0.84 (0.76 - 0.93) | <0.01 | 0.86 (0.76 - 0.99) | 0.03 |
| Meta-analysis (w/o USOC) | 47,264 | 91,066 | 28.3 | 82.2 | <168 | 438 | 0.78 (0.64 - 0.95) | 0.01 | 0.80 (0.65 - 0.97) | 0.03 |
| **PS-matching / on-treatment TAR without escalation** | | | | | | | | | | |
| CCAE | 14,880 | 23,938 | 9.6 | 14.7 | 63 | 115 | 0.83 (0.54 - 1.26) | 0.39 | 0.82 (0.54 - 1.25) | 0.38 |
| DAG | 2,894 | 8,701 | 2.1 | 8.3 | <5 | 10 | 1.79 (0.23 - 10.0) | 0.55 | 1.66 (0.25 - 11.1) | 0.60 |
| ODOD | 9,959 | 19,506 | 5.7 | 12.5 | 39 | 92 | 0.70 (0.40 - 1.20) | 0.21 | 0.72 (0.42 - 1.26) | 0.26 |
| MDCR | 1,274 | 4,750 | 0.8 | 3.5 | <5 | 13 | 1.66 (0.23 - 8.56) | 0.58 | 1.56 (0.25 - 9.59) | 0.63 |
| MDCD | 2,097 | 5,787 | 0.9 | 3.1 | <5 | 33 | 0.31 (0.05 - 1.15) | 0.15 | 0.30 (0.06 - 1.46) | 0.14 |
| Open Claims | 215,276 | 422,234 | 141.5 | 294.8 | 339 | 846 | 0.81 (0.67 - 0.97) | 0.02 | 0.84 (0.70 - 1.01) | 0.09 |
| OEHR | 14,026 | 31,802 | 3.8 | 13.4 | 20 | 98 | 0.69 (0.33 - 1.36) | 0.30 | 0.71 (0.35 - 1.43) | 0.34 |
| SIDIAP | 2,750 | 14,120 | 3.3 | 24.2 | <5 | 23 | 1.15 (0.16 - 5.20) | 0.87 | 1.32 (0.23 - 7.52) | 0.76 |
| Meta-analysis | 256,238 | 503,267 | 161.6 | 338.5 | <466 | 1,184 | 0.79 (0.68 - 0.92) | <0.01 | 0.81 (0.69 - 0.96) | 0.02 |
| Meta-analysis (w/o USOC) | 40,962 | 81,033 | 20.1 | 43.7 | <127 | 338 | 0.74 (0.55 - 1.00) | 0.05 | 0.75 (0.56 - 1.01) | 0.06 |
| **PS-stratification / on-treatment TAR without escalation** | | | | | | | | | | |
| CCAE | 17,049 | 23,974 | 10.7 | 14.7 | 68 | 115 | 0.78 (0.57 - 1.06) | 0.12 | 0.76 (0.55 - 1.04) | 0.09 |
| DAG | 3,193 | 8,742 | 2.3 | 8.3 | <5 | 10 | 3.03 (0.62 - 11.3) | 0.14 | 2.63 (0.61 - 11.3) | 0.20 |
| ODOD | 11,835 | 19,719 | 6.4 | 12.7 | 48 | 92 | 0.81 (0.55 - 1.19) | 0.29 | 0.82 (0.56 - 1.20) | 0.30 |
| MDCR | 1,444 | 4,816 | 0.8 | 3.6 | <5 | 13 | 1.38 (0.21 - 5.30) | 0.69 | 1.36 (0.27 - 6.81) | 0.71 |
| MDCD | 2,177 | 5,803 | 1.0 | 3.1 | <5 | 33 | 0.37 (0.11 - 0.97) | 0.08 | 0.35 (0.12 - 1.05) | 0.06 |
| Open Claims | 255,345 | 422,689 | 159.0 | 295.2 | 388 | 850 | 0.86 (0.76 - 0.98) | 0.02 | 0.90 (0.78 - 1.05) | 0.19 |
| OEHR | 15,455 | 32,773 | 4.2 | 13.8 | 24 | 101 | 0.75 (0.46 - 1.20) | 0.25 | 0.79 (0.49 - 1.28) | 0.35 |
| SIDIAP | 2,925 | 14,600 | 3.5 | 25.4 | <5 | 24 | 0.89 (0.13 - 3.41) | 0.89 | 0.93 (0.18 - 4.73) | 0.93 |
| Meta-analysis | 299,684 | 499,155 | 180.2 | 336.4 | 528 | 1,158 | 0.84 (0.75 - 0.94) | <0.01 | 0.86 (0.75 - 0.99) | 0.04 |
| Meta-analysis (w/o USOC) | 44,339 | 76,466 | 21.2 | 41.2 | 140 | 308 | 0.78 (0.63 - 0.97) | 0.03 | 0.79 (0.63 - 0.98) | 0.03 |

##### 1.2.11 Abnormal weight loss

**Supplementary Table 37:** Exposures, outcomes and risk estimates for Abnormal weight loss in SGLT2I (female) (T) vs DPP4I (female) (C) new-user cohorts under four different propensity score (PS) adjustment / time-at-risk (TAR) designs across data sources. We report total patient exposure time in 1,000 patient-year (KYr) units, uncalibrated and calibrated hazard ratios (HRs) and their 95% confidence intervals (CIs). Greyed out entries did not pass study diagnostics and are included only for completeness.

| | Patients | | Exposure (KYr) | | Outcomes | | Uncalibrated | | Calibrated | |
|---|---|---|---|---|---|---|---|---|---|---|
| | T | C | T | C | T | C | HR (95% CI) | $p$ | HR (95% CI) | $p$ |
| **PS-matching / on-treatment TAR** | | | | | | | | | | |
| CCAE | 16,030 | 25,270 | 12.0 | 18.9 | 104 | 154 | 1.06 (0.74 - 1.51) | 0.76 | 1.08 (0.75 - 1.54) | 0.69 |
| DAG | 2,860 | 8,535 | 2.3 | 8.9 | 21 | 70 | 1.02 (0.52 - 1.91) | 0.96 | 0.89 (0.47 - 1.71) | 0.73 |
| ODOD | 10,339 | 19,502 | 6.7 | 14.8 | 99 | 223 | 1.29 (0.93 - 1.80) | 0.12 | 1.34 (0.96 - 1.87) | 0.09 |
| MDCR | 1,247 | 4,569 | 0.9 | 3.9 | 9 | 66 | 0.96 (0.40 - 2.11) | 0.93 | 0.95 (0.41 - 2.19) | 0.88 |
| MDCD | 2,245 | 6,043 | 1.2 | 4.2 | 24 | 76 | 1.41 (0.73 - 2.64) | 0.30 | 1.37 (0.72 - 2.60) | 0.35 |
| Open Claims | 223,805 | 427,915 | 176.2 | 373.0 | 956 | 2,614 | 0.98 (0.88 - 1.09) | 0.76 | 1.03 (0.89 - 1.19) | 0.74 |
| OEHR | 14,682 | 32,695 | 4.6 | 17.1 | 42 | 195 | 1.13 (0.73 - 1.73) | 0.59 | 1.14 (0.74 - 1.77) | 0.56 |
| SIDIAP | 2,749 | 14,055 | 3.8 | 30.3 | 17 | 131 | 0.91 (0.48 - 1.63) | 0.77 | 0.97 (0.52 - 1.78) | 0.86 |
| Meta-analysis | 272,710 | 534,015 | 206.8 | 467.3 | 1,263 | 3,463 | 1.04 (0.93 - 1.17) | 0.49 | 1.08 (0.95 - 1.24) | 0.26 |
| Meta-analysis (w/o USOC) | 48,905 | 106,100 | 30.6 | 94.3 | 307 | 849 | 1.15 (0.95 - 1.38) | 0.14 | 1.17 (0.98 - 1.41) | 0.10 |
| **PS-stratification / on-treatment TAR** | | | | | | | | | | |
| CCAE | 18,444 | 25,301 | 13.3 | 18.9 | 112 | 154 | 1.13 (0.87 - 1.45) | 0.36 | 1.13 (0.88 - 1.46) | 0.36 |
| DAG | 3,163 | 8,568 | 2.5 | 9.0 | 21 | 70 | 1.10 (0.63 - 1.85) | 0.73 | 0.96 (0.56 - 1.65) | 0.82 |
| ODOD | 12,379 | 19,749 | 7.5 | 15.0 | 115 | 229 | 1.27 (0.99 - 1.62) | 0.06 | 1.30 (1.01 - 1.67) | 0.04 |
| MDCR | 1,427 | 4,653 | 0.9 | 3.9 | 11 | 68 | 0.93 (0.44 - 1.80) | 0.84 | 0.94 (0.47 - 1.90) | 0.87 |
| MDCD | 2,330 | 6,060 | 1.2 | 4.2 | 24 | 76 | 1.09 (0.66 - 1.75) | 0.74 | 1.04 (0.64 - 1.70) | 0.87 |
| Open Claims | 266,948 | 428,313 | 197.5 | 373.4 | 1,078 | 2,622 | 1.00 (0.93 - 1.08) | 0.97 | 1.05 (0.92 - 1.19) | 0.47 |
| OEHR | 16,231 | 33,687 | 5.1 | 17.6 | 46 | 206 | 0.97 (0.68 - 1.36) | 0.86 | 1.03 (0.73 - 1.45) | 0.86 |
| SIDIAP | 2,925 | 14,525 | 4.0 | 31.8 | 17 | 140 | 0.93 (0.53 - 1.56) | 0.80 | 0.93 (0.54 - 1.61) | 0.80 |
| Meta-analysis | 320,090 | 530,143 | 229.9 | 465.7 | 1,389 | 3,421 | 1.05 (0.95 - 1.16) | 0.35 | 1.08 (0.94 - 1.24) | 0.27 |
| Meta-analysis (w/o USOC) | 53,142 | 101,830 | 32.4 | 92.3 | 311 | 799 | 1.13 (0.97 - 1.30) | 0.11 | 1.15 (0.99 - 1.34) | 0.07 |
| **PS-matching / on-treatment TAR without escalation** | | | | | | | | | | |
| CCAE | 16,030 | 25,270 | 10.3 | 15.4 | 86 | 128 | 1.03 (0.70 - 1.51) | 0.87 | 1.02 (0.69 - 1.50) | 0.90 |
| DAG | 2,860 | 8,535 | 2.1 | 8.0 | 19 | 61 | 1.00 (0.48 - 2.00) | 0.99 | 0.94 (0.46 - 1.92) | 0.80 |
| ODOD | 10,339 | 19,502 | 5.9 | 12.4 | 89 | 184 | 1.42 (1.00 - 2.02) | 0.05 | 1.47 (1.03 - 2.09) | 0.04 |
| MDCR | 1,247 | 4,569 | 0.8 | 3.3 | 8 | 60 | 1.20 (0.44 - 2.93) | 0.70 | 1.12 (0.44 - 2.88) | 0.81 |
| MDCD | 2,245 | 6,043 | 1.0 | 3.3 | 21 | 53 | 1.89 (0.90 - 3.95) | 0.09 | 1.84 (0.88 - 3.85) | 0.11 |
| Open Claims | 223,805 | 427,915 | 147.2 | 296.4 | 810 | 2,130 | 0.99 (0.88 - 1.11) | 0.84 | 1.03 (0.91 - 1.16) | 0.72 |
| OEHR | 14,682 | 32,695 | 4.0 | 13.8 | 36 | 155 | 1.25 (0.77 - 1.99) | 0.36 | 1.28 (0.79 - 2.05) | 0.33 |
| SIDIAP | 2,749 | 14,055 | 3.4 | 24.0 | 15 | 106 | 0.95 (0.47 - 1.79) | 0.89 | 1.09 (0.56 - 2.13) | 0.79 |
| Meta-analysis | 272,710 | 534,015 | 173.9 | 373.3 | 1,076 | 2,817 | 1.11 (0.94 - 1.31) | 0.21 | 1.15 (0.96 - 1.37) | 0.14 |
| Meta-analysis (w/o USOC) | 48,905 | 106,100 | 26.7 | 76.9 | 266 | 687 | 1.22 (1.00 - 1.49) | 0.05 | 1.24 (1.02 - 1.52) | 0.04 |
| **PS-stratification / on-treatment TAR without escalation** | | | | | | | | | | |
| CCAE | 18,444 | 25,301 | 11.5 | 15.4 | 93 | 128 | 1.12 (0.85 - 1.48) | 0.43 | 1.09 (0.82 - 1.44) | 0.57 |
| DAG | 3,163 | 8,568 | 2.2 | 8.1 | 19 | 61 | 1.20 (0.66 - 2.09) | 0.54 | 1.04 (0.59 - 1.85) | 0.82 |
| ODOD | 12,379 | 19,749 | 6.7 | 12.6 | 100 | 190 | 1.31 (1.00 - 1.71) | 0.05 | 1.31 (1.00 - 1.72) | 0.05 |
| MDCR | 1,427 | 4,653 | 0.8 | 3.4 | 10 | 62 | 0.96 (0.43 - 1.92) | 0.91 | 0.94 (0.45 - 1.98) | 0.87 |
| MDCD | 2,330 | 6,060 | 1.0 | 3.3 | 21 | 53 | 1.24 (0.71 - 2.10) | 0.44 | 1.17 (0.68 - 2.02) | 0.57 |
| Open Claims | 266,948 | 428,313 | 166.1 | 296.7 | 907 | 2,138 | 0.99 (0.91 - 1.08) | 0.88 | 1.04 (0.93 - 1.16) | 0.53 |
| OEHR | 16,231 | 33,687 | 4.4 | 14.2 | 39 | 161 | 1.01 (0.69 - 1.46) | 0.96 | 1.06 (0.72 - 1.54) | 0.78 |
| SIDIAP | 2,925 | 14,525 | 3.5 | 25.2 | 15 | 113 | 0.95 (0.51 - 1.65) | 0.87 | 0.99 (0.55 - 1.78) | 0.87 |
| Meta-analysis | 320,090 | 530,143 | 194.5 | 372.2 | 1,173 | 2,791 | 1.06 (0.94 - 1.20) | 0.32 | 1.09 (0.94 - 1.26) | 0.27 |
| Meta-analysis (w/o USOC) | 53,142 | 101,830 | 28.3 | 75.5 | 266 | 653 | 1.15 (0.98 - 1.35) | 0.08 | 1.16 (0.98 - 1.36) | 0.09 |

##### 1.2.12 Acute pancreatitis

**Supplementary Table 38:** Exposures, outcomes and risk estimates for Acute pancreatitis in SGLT2I (female) (T) vs DPP4I (female) (C) new-user cohorts under four different propensity score (PS) adjustment / time-at-risk (TAR) designs across data sources. We report total patient exposure time in 1,000 patient-year (KYr) units, uncalibrated and calibrated hazard ratios (HRs) and their 95% confidence intervals (CIs). Greyed out entries did not pass study diagnostics and are included only for completeness.

| | Patients | | Exposure (KYr) | | Outcomes | | Uncalibrated | | Calibrated | |
|---|---|---|---|---|---|---|---|---|---|---|
| | T | C | T | C | T | C | HR (95% CI) | $p$ | HR (95% CI) | $p$ |
| **PS-matching / on-treatment TAR** | | | | | | | | | | |
| CCAE | 16,286 | 25,742 | 12.3 | 19.3 | 21 | 45 | 0.92 (0.45 - 1.84) | 0.82 | 0.94 (0.47 - 1.90) | 0.87 |
| DAG | 2,939 | 8,792 | 2.4 | 9.3 | 0 | 0 | | | | |
| ODOD | 10,645 | 20,480 | 6.9 | 15.6 | 16 | 49 | 0.81 (0.39 - 1.61) | 0.57 | 0.84 (0.41 - 1.71) | 0.64 |
| MDCR | 1,287 | 4,814 | 0.9 | 4.1 | <5 | 6 | 2.56 (0.33 - 14.9) | 0.33 | 2.52 (0.37 - 17.0) | 0.34 |
| MDCD | 2,301 | 6,249 | 1.2 | 4.4 | 9 | 21 | 0.64 (0.17 - 1.96) | 0.47 | 0.62 (0.19 - 2.10) | 0.45 |
| Open Claims | 228,911 | 441,231 | 180.6 | 386.7 | 242 | 831 | 0.81 (0.66 - 0.99) | 0.05 | 0.85 (0.68 - 1.06) | 0.17 |
| OEHR | 14,999 | 33,550 | 4.7 | 17.8 | 9 | 34 | 1.20 (0.43 - 3.16) | 0.72 | 1.22 (0.45 - 3.32) | 0.70 |
| SIDIAP | 2,770 | 14,242 | 3.9 | 30.8 | <5 | 12 | 1.18 (0.06 - 6.68) | 0.89 | 1.25 (0.12 - 12.9) | 0.85 |
| Meta-analysis | 273,142 | 527,252 | 205.7 | 443.8 | 297 | 980 | 0.83 (0.69 - 0.99) | 0.04 | 0.86 (0.71 - 1.04) | 0.13 |
| Meta-analysis (w/o USOC) | 44,231 | 86,021 | 25.2 | 57.1 | 55 | 149 | 0.89 (0.58 - 1.35) | 0.57 | 0.91 (0.60 - 1.38) | 0.65 |
| **PS-stratification / on-treatment TAR** | | | | | | | | | | |
| CCAE | 18,712 | 25,778 | 13.6 | 19.4 | 22 | 45 | 0.74 (0.43 - 1.24) | 0.26 | 0.74 (0.44 - 1.26) | 0.27 |
| DAG | 3,243 | 8,833 | 2.5 | 9.3 | 0 | 0 | | | | |
| ODOD | 12,741 | 20,685 | 7.8 | 15.8 | 17 | 52 | 0.93 (0.50 - 1.67) | 0.81 | 0.95 (0.52 - 1.74) | 0.88 |
| MDCR | 1,462 | 4,881 | 0.9 | 4.1 | <5 | 6 | 3.57 (0.46 - 18.8) | 0.18 | 3.62 (0.56 - 23.1) | 0.18 |
| MDCD | 2,390 | 6,268 | 1.3 | 4.4 | 9 | 21 | 1.24 (0.51 - 2.79) | 0.62 | 1.18 (0.51 - 2.76) | 0.70 |
| Open Claims | 272,688 | 441,635 | 202.2 | 387.1 | 275 | 835 | 0.79 (0.68 - 0.91) | <0.01 | 0.83 (0.69 - 0.99) | 0.04 |
| OEHR | 16,582 | 34,586 | 5.2 | 18.3 | 9 | 36 | 1.05 (0.46 - 2.20) | 0.90 | 1.11 (0.51 - 2.45) | 0.79 |
| SIDIAP | 2,947 | 14,724 | 4.0 | 32.3 | <5 | 13 | 0.92 (0.05 - 4.92) | 0.94 | 0.92 (0.09 - 9.17) | 0.94 |
| Meta-analysis | 320,723 | 522,684 | 228.8 | 440.5 | 323 | 968 | 0.80 (0.70 - 0.91) | <0.01 | 0.82 (0.70 - 0.97) | 0.02 |
| Meta-analysis (w/o USOC) | 48,035 | 81,049 | 26.6 | 53.5 | 48 | 133 | 0.86 (0.60 - 1.23) | 0.40 | 0.88 (0.62 - 1.26) | 0.49 |
| **PS-matching / on-treatment TAR without escalation** | | | | | | | | | | |
| CCAE | 16,286 | 25,742 | 10.5 | 15.8 | 16 | 37 | 0.74 (0.34 - 1.54) | 0.43 | 0.73 (0.34 - 1.55) | 0.42 |
| DAG | 2,939 | 8,792 | 2.2 | 8.3 | 0 | 0 | | | | |
| ODOD | 10,645 | 20,480 | 6.1 | 13.1 | 12 | 39 | 0.79 (0.35 - 1.67) | 0.55 | 0.81 (0.37 - 1.78) | 0.61 |
| MDCR | 1,287 | 4,814 | 0.8 | 3.6 | <5 | 5 | 3.00 (0.36 - 20.5) | 0.29 | 2.81 (0.37 - 21.1) | 0.32 |
| MDCD | 2,301 | 6,249 | 1.0 | 3.4 | 8 | 14 | 0.75 (0.19 - 2.50) | 0.66 | 0.73 (0.20 - 2.61) | 0.63 |
| Open Claims | 228,911 | 441,231 | 150.9 | 307.3 | 189 | 643 | 0.83 (0.66 - 1.03) | 0.10 | 0.86 (0.69 - 1.08) | 0.22 |
| OEHR | 14,999 | 33,550 | 4.1 | 14.2 | 7 | 26 | 1.44 (0.41 - 4.54) | 0.55 | 1.47 (0.44 - 4.87) | 0.53 |
| SIDIAP | 2,770 | 14,242 | 3.4 | 24.4 | <5 | 8 | 1.94 (0.10 - 12.7) | 0.59 | 2.23 (0.20 - 25.1) | 0.52 |
| Meta-analysis | 273,142 | 527,252 | 172.7 | 353.9 | 232 | 759 | 0.83 (0.68 - 1.01) | 0.07 | 0.86 (0.69 - 1.06) | 0.15 |
| Meta-analysis (w/o USOC) | 44,231 | 86,021 | 21.8 | 46.6 | 43 | 116 | 0.84 (0.53 - 1.33) | 0.45 | 0.85 (0.53 - 1.35) | 0.49 |
| **PS-stratification / on-treatment TAR without escalation** | | | | | | | | | | |
| CCAE | 18,712 | 25,778 | 11.7 | 15.8 | 17 | 37 | 0.67 (0.36 - 1.19) | 0.18 | 0.65 (0.36 - 1.18) | 0.16 |
| DAG | 3,243 | 8,833 | 2.3 | 8.4 | 0 | 0 | | | | |
| ODOD | 12,741 | 20,685 | 6.9 | 13.3 | 13 | 42 | 0.88 (0.43 - 1.68) | 0.71 | 0.88 (0.45 - 1.74) | 0.72 |
| MDCR | 1,462 | 4,881 | 0.8 | 3.6 | <5 | 5 | 4.19 (0.51 - 25.4) | 0.15 | 4.12 (0.59 - 29.0) | 0.16 |
| MDCD | 2,390 | 6,268 | 1.1 | 3.4 | 8 | 14 | 1.33 (0.51 - 3.30) | 0.55 | 1.26 (0.49 - 3.21) | 0.63 |
| Open Claims | 272,688 | 441,635 | 170.1 | 307.7 | 211 | 646 | 0.76 (0.64 - 0.89) | <0.01 | 0.79 (0.66 - 0.95) | 0.02 |
| OEHR | 16,582 | 34,586 | 4.5 | 14.7 | 7 | 27 | 0.97 (0.37 - 2.25) | 0.95 | 1.02 (0.41 - 2.51) | 0.95 |
| SIDIAP | 2,947 | 14,724 | 3.5 | 25.6 | <5 | 9 | 1.32 (0.07 - 7.32) | 0.82 | 1.38 (0.14 - 14.0) | 0.79 |
| Meta-analysis | 320,723 | 522,684 | 193.2 | 351.4 | 248 | 752 | 0.76 (0.65 - 0.89) | <0.01 | 0.78 (0.65 - 0.93) | <0.01 |
| Meta-analysis (w/o USOC) | 48,035 | 81,049 | 23.2 | 43.8 | 37 | 106 | 0.79 (0.53 - 1.18) | 0.25 | 0.79 (0.53 - 1.19) | 0.26 |

##### 1.2.13 Bone fracture

**Supplementary Table 39:** Exposures, outcomes and risk estimates for Bone fracture in SGLT2I (female) (T) vs DPP4I (female) (C) new-user cohorts under four different propensity score (PS) adjustment / time-at-risk (TAR) designs across data sources. We report total patient exposure time in 1,000 patient-year (KYr) units, uncalibrated and calibrated hazard ratios (HRs) and their 95% confidence intervals (CIs). Greyed out entries did not pass study diagnostics and are included only for completeness.

| | Patients | | Exposure (KYr) | | Outcomes | | Uncalibrated | | Calibrated | |
|---|---|---|---|---|---|---|---|---|---|---|
| | T | C | T | C | T | C | HR (95% CI) | $p$ | HR (95% CI) | $p$ |
| **PS-matching / on-treatment TAR** | | | | | | | | | | |
| CCAE | 14,745 | 23,331 | 10.8 | 17.1 | 203 | 386 | 0.89 (0.69 - 1.15) | 0.39 | 0.91 (0.71 - 1.18) | 0.52 |
| DAG | 2,574 | 7,485 | 2.1 | 7.8 | 30 | 182 | 0.62 (0.35 - 1.03) | 0.08 | 0.54 (0.32 - 0.93) | 0.04 |
| ODOD | 9,580 | 18,098 | 6.1 | 13.5 | 206 | 484 | 1.18 (0.92 - 1.49) | 0.18 | 1.22 (0.96 - 1.55) | 0.12 |
| MDCR | 1,130 | 3,987 | 0.7 | 3.3 | 34 | 148 | 1.45 (0.82 - 2.50) | 0.20 | 1.42 (0.81 - 2.49) | 0.23 |
| MDCD | 2,026 | 5,487 | 1.1 | 3.7 | 33 | 151 | 1.17 (0.69 - 1.94) | 0.55 | 1.14 (0.68 - 1.92) | 0.63 |
| Open Claims | 200,013 | 381,565 | 156.2 | 327.8 | 2,577 | 7,138 | 0.93 (0.87 - 1.00) | 0.04 | 0.97 (0.86 - 1.10) | 0.69 |
| OEHR | 13,943 | 30,967 | 4.4 | 16.1 | 66 | 348 | 0.84 (0.58 - 1.19) | 0.34 | 0.85 (0.59 - 1.22) | 0.40 |
| SIDIAP | 2,357 | 11,917 | 3.3 | 25.2 | 50 | 559 | 0.83 (0.58 - 1.16) | 0.29 | 0.88 (0.62 - 1.25) | 0.49 |
| Meta-analysis | 246,566 | 483,639 | 185.2 | 416.9 | 3,177 | 9,325 | 0.94 (0.88 - 0.99) | 0.03 | 0.97 (0.89 - 1.07) | 0.55 |
| Meta-analysis (w/o USOC) | 46,553 | 102,074 | 29.0 | 89.1 | 600 | 2,187 | 0.93 (0.80 - 1.09) | 0.37 | 0.95 (0.81 - 1.12) | 0.57 |
| **PS-stratification / on-treatment TAR** | | | | | | | | | | |
| CCAE | 16,973 | 23,365 | 12.0 | 17.1 | 230 | 388 | 0.89 (0.75 - 1.05) | 0.16 | 0.89 (0.75 - 1.06) | 0.21 |
| DAG | 2,825 | 7,508 | 2.2 | 7.8 | 31 | 186 | 0.77 (0.50 - 1.15) | 0.22 | 0.68 (0.45 - 1.03) | 0.08 |
| ODOD | 11,478 | 18,317 | 6.9 | 13.7 | 222 | 500 | 1.11 (0.94 - 1.32) | 0.22 | 1.14 (0.96 - 1.36) | 0.15 |
| MDCR | 1,272 | 4,110 | 0.8 | 3.4 | 35 | 159 | 1.10 (0.72 - 1.63) | 0.66 | 1.11 (0.74 - 1.68) | 0.62 |
| MDCD | 2,102 | 5,506 | 1.1 | 3.8 | 33 | 154 | 0.74 (0.49 - 1.08) | 0.13 | 0.70 (0.47 - 1.04) | 0.09 |
| Open Claims | 237,944 | 382,107 | 175.0 | 328.3 | 2,817 | 7,167 | 0.93 (0.89 - 0.97) | <0.01 | 0.98 (0.87 - 1.09) | 0.67 |
| OEHR | 15,400 | 31,977 | 4.8 | 16.6 | 69 | 368 | 0.83 (0.62 - 1.08) | 0.18 | 0.87 (0.66 - 1.16) | 0.37 |
| SIDIAP | 2,515 | 12,279 | 3.4 | 26.3 | 55 | 605 | 0.85 (0.63 - 1.13) | 0.29 | 0.85 (0.63 - 1.15) | 0.35 |
| Meta-analysis | 287,135 | 475,553 | 204.3 | 409.8 | 3,424 | 9,214 | 0.93 (0.89 - 0.97) | <0.01 | 0.96 (0.87 - 1.06) | 0.39 |
| Meta-analysis (w/o USOC) | 49,191 | 93,446 | 29.4 | 81.5 | 607 | 2,047 | 0.92 (0.80 - 1.05) | 0.23 | 0.94 (0.82 - 1.08) | 0.42 |
| **PS-matching / on-treatment TAR without escalation** | | | | | | | | | | |
| CCAE | 14,745 | 23,331 | 9.3 | 14.1 | 172 | 302 | 0.89 (0.68 - 1.17) | 0.40 | 0.88 (0.66 - 1.15) | 0.40 |
| DAG | 2,574 | 7,485 | 1.9 | 7.0 | 27 | 167 | 0.60 (0.31 - 1.06) | 0.09 | 0.56 (0.30 - 1.02) | 0.07 |
| ODOD | 9,580 | 18,098 | 5.4 | 11.4 | 177 | 399 | 1.18 (0.91 - 1.52) | 0.20 | 1.22 (0.94 - 1.58) | 0.15 |
| MDCR | 1,130 | 3,987 | 0.7 | 2.9 | 29 | 123 | 1.47 (0.79 - 2.68) | 0.22 | 1.37 (0.74 - 2.54) | 0.32 |
| MDCD | 2,026 | 5,487 | 0.9 | 2.9 | 28 | 117 | 1.08 (0.61 - 1.87) | 0.78 | 1.05 (0.60 - 1.84) | 0.85 |
| Open Claims | 200,013 | 381,565 | 130.6 | 261.2 | 2,137 | 5,665 | 0.93 (0.86 - 1.00) | 0.04 | 0.96 (0.88 - 1.05) | 0.52 |
| OEHR | 13,943 | 30,967 | 3.8 | 13.0 | 56 | 256 | 0.80 (0.54 - 1.18) | 0.27 | 0.82 (0.56 - 1.22) | 0.35 |
| SIDIAP | 2,357 | 11,917 | 2.9 | 20.0 | 41 | 457 | 0.78 (0.53 - 1.13) | 0.21 | 0.90 (0.61 - 1.32) | 0.62 |
| Meta-analysis | 246,566 | 483,639 | 156.0 | 334.1 | 2,649 | 7,425 | 0.93 (0.87 - 0.99) | 0.03 | 0.96 (0.87 - 1.05) | 0.37 |
| Meta-analysis (w/o USOC) | 46,553 | 102,074 | 25.3 | 72.9 | 512 | 1,760 | 0.92 (0.79 - 1.08) | 0.33 | 0.94 (0.80 - 1.10) | 0.46 |
| **PS-stratification / on-treatment TAR without escalation** | | | | | | | | | | |
| CCAE | 16,973 | 23,365 | 10.4 | 14.1 | 196 | 304 | 0.91 (0.75 - 1.10) | 0.33 | 0.88 (0.73 - 1.07) | 0.24 |
| DAG | 2,825 | 7,508 | 2.0 | 7.0 | 28 | 171 | 0.71 (0.45 - 1.08) | 0.12 | 0.62 (0.40 - 0.95) | 0.04 |
| ODOD | 11,478 | 18,317 | 6.2 | 11.5 | 192 | 413 | 1.12 (0.93 - 1.35) | 0.23 | 1.13 (0.93 - 1.36) | 0.24 |
| MDCR | 1,272 | 4,110 | 0.7 | 3.0 | 30 | 134 | 1.09 (0.69 - 1.67) | 0.72 | 1.07 (0.68 - 1.67) | 0.78 |
| MDCD | 2,102 | 5,506 | 0.9 | 2.9 | 28 | 118 | 0.79 (0.51 - 1.20) | 0.29 | 0.75 (0.49 - 1.15) | 0.20 |
| Open Claims | 237,944 | 382,107 | 147.3 | 261.6 | 2,346 | 5,689 | 0.93 (0.88 - 0.98) | <0.01 | 0.97 (0.89 - 1.06) | 0.55 |
| OEHR | 15,400 | 31,977 | 4.2 | 13.4 | 59 | 272 | 0.85 (0.63 - 1.15) | 0.31 | 0.89 (0.66 - 1.21) | 0.49 |
| SIDIAP | 2,515 | 12,279 | 3.0 | 20.8 | 46 | 497 | 0.79 (0.57 - 1.08) | 0.15 | 0.83 (0.60 - 1.14) | 0.30 |
| Meta-analysis | 287,135 | 475,553 | 173.0 | 328.4 | 2,867 | 7,346 | 0.93 (0.89 - 0.98) | <0.01 | 0.95 (0.86 - 1.05) | 0.33 |
| Meta-analysis (w/o USOC) | 49,191 | 93,446 | 25.7 | 66.9 | 521 | 1,657 | 0.91 (0.78 - 1.06) | 0.24 | 0.92 (0.78 - 1.07) | 0.28 |

##### 1.2.14 Diabetic ketoacidosis

**Supplementary Table 40:** Exposures, outcomes and risk estimates for Diabetic ketoacidosis in SGLT2I (female) (T) vs DPP4I (female) (C) new-user cohorts under four different propensity score (PS) adjustment / time-at-risk (TAR) designs across data sources. We report total patient exposure time in 1,000 patient-year (KYr) units, uncalibrated and calibrated hazard ratios (HRs) and their 95% confidence intervals (CIs). Greyed out entries did not pass study diagnostics and are included only for completeness.

| | Patients | | Exposure (KYr) | | Outcomes | | Uncalibrated | | Calibrated | |
|---|---|---|---|---|---|---|---|---|---|---|
| | T | C | T | C | T | C | HR (95% CI) | $p$ | HR (95% CI) | $p$ |
| **PS-matching / on-treatment TAR** | | | | | | | | | | |
| CCAE | 16,394 | 25,898 | 12.3 | 19.5 | 25 | 19 | 1.59 (0.73 - 3.58) | 0.26 | 1.62 (0.73 - 3.59) | 0.24 |
| DAG | 2,939 | 8,792 | 2.4 | 9.3 | 0 | 0 | | | | |
| ODOD | 10,722 | 20,613 | 7.0 | 15.7 | 21 | 28 | 1.86 (0.93 - 3.79) | 0.08 | 1.93 (0.96 - 3.90) | 0.07 |
| MDCR | 1,300 | 4,828 | 0.9 | 4.1 | <5 | 5 | | | | |
| MDCD | 2,335 | 6,347 | 1.3 | 4.4 | <5 | 16 | 1.04 (0.22 - 3.97) | 0.95 | 1.02 (0.24 - 4.37) | 0.95 |
| Open Claims | 231,509 | 445,431 | 182.6 | 390.4 | 212 | 359 | 1.57 (1.24 - 1.98) | <0.01 | 1.63 (1.27 - 2.11) | <0.01 |
| OEHR | 15,056 | 33,689 | 4.8 | 17.8 | 8 | 22 | 1.69 (0.59 - 4.97) | 0.33 | 1.72 (0.59 - 4.98) | 0.32 |
| SIDIAP | 2,778 | 14,274 | 3.9 | 30.9 | 0 | 0 | | | | |
| Meta-analysis | 276,016 | 531,978 | 207.9 | 447.9 | <271 | 444 | 1.58 (1.29 - 1.95) | <0.01 | 1.64 (1.32 - 2.05) | <0.01 |
| Meta-analysis (w/o USOC) | 44,507 | 86,547 | 25.3 | 57.5 | <59 | 85 | 1.65 (1.05 - 2.58) | 0.03 | 1.69 (1.08 - 2.65) | 0.02 |
| **PS-stratification / on-treatment TAR** | | | | | | | | | | |
| CCAE | 18,865 | 25,934 | 13.7 | 19.5 | 31 | 19 | 2.02 (1.12 - 3.72) | 0.02 | 2.03 (1.11 - 3.69) | 0.02 |
| DAG | 3,243 | 8,833 | 2.5 | 9.3 | 0 | 0 | | | | |
| ODOD | 12,833 | 20,819 | 7.9 | 15.9 | 24 | 28 | 1.76 (0.94 - 3.27) | 0.08 | 1.80 (0.97 - 3.36) | 0.06 |
| MDCR | 1,475 | 4,895 | 0.9 | 4.2 | <5 | 5 | 0.92 (0.05 - 6.83) | 0.95 | 0.94 (0.08 - 11.5) | 0.96 |
| MDCD | 2,427 | 6,366 | 1.3 | 4.5 | 5 | 16 | 0.88 (0.27 - 2.39) | 0.81 | 0.84 (0.28 - 2.48) | 0.75 |
| Open Claims | 276,157 | 445,863 | 204.7 | 390.8 | 254 | 359 | 1.42 (1.19 - 1.69) | <0.01 | 1.49 (1.21 - 1.82) | <0.01 |
| OEHR | 16,657 | 34,731 | 5.2 | 18.4 | 11 | 24 | 1.55 (0.68 - 3.40) | 0.28 | 1.65 (0.74 - 3.68) | 0.23 |
| SIDIAP | 2,956 | 14,758 | 4.0 | 32.4 | 0 | 0 | | | | |
| Meta-analysis | 324,512 | 527,347 | 231.4 | 444.6 | 320 | 430 | 1.48 (1.26 - 1.74) | <0.01 | 1.52 (1.27 - 1.83) | <0.01 |
| Meta-analysis (w/o USOC) | 48,355 | 81,484 | 26.8 | 53.8 | 66 | 71 | 1.81 (1.24 - 2.64) | <0.01 | 1.85 (1.27 - 2.71) | <0.01 |
| **PS-matching / on-treatment TAR without escalation** | | | | | | | | | | |
| CCAE | 16,394 | 25,898 | 10.6 | 15.9 | 18 | 13 | 1.77 (0.72 - 4.57) | 0.23 | 1.74 (0.69 - 4.38) | 0.24 |
| DAG | 2,939 | 8,792 | 2.2 | 8.3 | 0 | 0 | | | | |
| ODOD | 10,722 | 20,613 | 6.2 | 13.2 | 19 | 22 | 1.79 (0.85 - 3.86) | 0.13 | 1.85 (0.87 - 3.96) | 0.11 |
| MDCR | 1,300 | 4,828 | 0.8 | 3.6 | <5 | <5 | | | | |
| MDCD | 2,335 | 6,347 | 1.1 | 3.5 | <5 | 11 | 0.82 (0.12 - 3.74) | 0.82 | 0.79 (0.14 - 4.49) | 0.79 |
| Open Claims | 231,509 | 445,431 | 152.5 | 310.4 | 164 | 250 | 1.56 (1.20 - 2.04) | <0.01 | 1.63 (1.24 - 2.13) | <0.01 |
| OEHR | 15,056 | 33,689 | 4.1 | 14.3 | <5 | 13 | 1.00 (0.24 - 3.84) | 1.00 | 1.02 (0.26 - 4.06) | 0.96 |
| SIDIAP | 2,778 | 14,274 | 3.4 | 24.4 | 0 | 0 | | | | |
| Meta-analysis | 258,625 | 491,942 | 169.2 | 339.5 | 201 | 285 | 1.60 (1.26 - 2.04) | <0.01 | 1.65 (1.28 - 2.12) | <0.01 |
| Meta-analysis (w/o USOC) | 27,116 | 46,511 | 16.7 | 29.1 | 37 | 35 | 1.78 (0.99 - 3.20) | 0.05 | 1.81 (1.01 - 3.25) | 0.05 |
| **PS-stratification / on-treatment TAR without escalation** | | | | | | | | | | |
| CCAE | 18,865 | 25,934 | 11.8 | 15.9 | 24 | 13 | 2.04 (1.02 - 4.25) | 0.05 | 1.98 (0.97 - 4.04) | 0.06 |
| DAG | 3,243 | 8,833 | 2.3 | 8.4 | 0 | 0 | | | | |
| ODOD | 12,833 | 20,819 | 7.0 | 13.4 | 20 | 22 | 1.82 (0.91 - 3.63) | 0.09 | 1.82 (0.91 - 3.64) | 0.09 |
| MDCR | 1,475 | 4,895 | 0.8 | 3.6 | <5 | <5 | 1.12 (0.05 - 9.94) | 0.93 | 1.10 (0.08 - 15.2) | 0.94 |
| MDCD | 2,427 | 6,366 | 1.1 | 3.5 | <5 | 11 | 0.75 (0.16 - 2.57) | 0.68 | 0.71 (0.18 - 2.82) | 0.63 |
| Open Claims | 276,157 | 445,863 | 172.1 | 310.7 | 200 | 250 | 1.53 (1.25 - 1.88) | <0.01 | 1.60 (1.29 - 1.99) | <0.01 |
| OEHR | 16,657 | 34,731 | 4.6 | 14.8 | 7 | 14 | 1.20 (0.42 - 3.17) | 0.72 | 1.26 (0.46 - 3.44) | 0.66 |
| SIDIAP | 2,956 | 14,758 | 3.5 | 25.6 | 0 | 0 | | | | |
| Meta-analysis | 324,512 | 527,347 | 195.4 | 354.8 | 251 | 299 | 1.57 (1.30 - 1.89) | <0.01 | 1.61 (1.31 - 1.98) | <0.01 |
| Meta-analysis (w/o USOC) | 48,355 | 81,484 | 23.3 | 44.0 | 51 | 49 | 1.75 (1.12 - 2.73) | 0.01 | 1.76 (1.13 - 2.74) | 0.01 |

##### 1.2.15 Diarrhea

**Supplementary Table 41:** Exposures, outcomes and risk estimates for Diarrhea in SGLT2I (female) (T) vs DPP4I (female) (C) new-user cohorts under four different propensity score (PS) adjustment / time-at-risk (TAR) designs across data sources. We report total patient exposure time in 1,000 patient-year (KYr) units, uncalibrated and calibrated hazard ratios (HRs) and their 95% confidence intervals (CIs). Greyed out entries did not pass study diagnostics and are included only for completeness.

| | Patients | | Exposure (KYr) | | Outcomes | | Uncalibrated | | Calibrated | |
|---|---|---|---|---|---|---|---|---|---|---|
| | T | C | T | C | T | C | HR (95% CI) | $p$ | HR (95% CI) | $p$ |
| **PS-matching / on-treatment TAR** | | | | | | | | | | |
| CCAE | 13,703 | 21,595 | 10.2 | 15.7 | 329 | 589 | 0.82 (0.68 - 1.00) | 0.05 | 0.84 (0.69 - 1.02) | 0.12 |
| DAG | 2,883 | 8,641 | 2.3 | 9.1 | 7 | 22 | 1.58 (0.47 - 4.62) | 0.43 | 1.39 (0.44 - 4.34) | 0.58 |
| ODOD | 8,810 | 16,500 | 5.6 | 12.1 | 238 | 625 | 0.86 (0.69 - 1.06) | 0.15 | 0.89 (0.71 - 1.10) | 0.30 |
| MDCR | 1,060 | 3,818 | 0.7 | 3.1 | 27 | 131 | 0.69 (0.38 - 1.21) | 0.22 | 0.68 (0.38 - 1.22) | 0.22 |
| MDCD | 1,724 | 4,705 | 0.9 | 3.1 | 55 | 216 | 0.74 (0.49 - 1.11) | 0.16 | 0.73 (0.48 - 1.10) | 0.15 |
| Open Claims | 193,021 | 370,847 | 151.0 | 317.9 | 2,852 | 8,032 | 0.83 (0.78 - 0.89) | <0.01 | 0.87 (0.77 - 0.98) | 0.03 |
| OEHR | 12,832 | 28,602 | 4.0 | 14.4 | 159 | 695 | 0.80 (0.62 - 1.01) | 0.07 | 0.81 (0.63 - 1.03) | 0.11 |
| SIDIAP | 2,601 | 13,232 | 3.6 | 28.3 | 53 | 432 | 0.83 (0.57 - 1.19) | 0.33 | 0.88 (0.61 - 1.28) | 0.53 |
| Meta-analysis | 236,667 | 468,127 | 178.8 | 405.3 | 3,713 | 10,717 | 0.83 (0.79 - 0.88) | <0.01 | 0.87 (0.79 - 0.94) | <0.01 |
| Meta-analysis (w/o USOC) | 43,646 | 97,280 | 27.7 | 87.4 | 861 | 2,685 | 0.83 (0.74 - 0.93) | <0.01 | 0.85 (0.76 - 0.95) | 0.01 |
| **PS-stratification / on-treatment TAR** | | | | | | | | | | |
| CCAE | 15,864 | 21,619 | 11.4 | 15.7 | 371 | 589 | 0.85 (0.74 - 0.97) | 0.02 | 0.85 (0.74 - 0.98) | 0.04 |
| DAG | 3,170 | 8,679 | 2.5 | 9.1 | 7 | 23 | 1.52 (0.56 - 3.73) | 0.39 | 1.33 (0.51 - 3.44) | 0.56 |
| ODOD | 10,574 | 16,684 | 6.4 | 12.2 | 262 | 634 | 0.87 (0.74 - 1.01) | 0.07 | 0.89 (0.76 - 1.04) | 0.17 |
| MDCR | 1,217 | 3,941 | 0.8 | 3.2 | 29 | 139 | 0.86 (0.55 - 1.31) | 0.50 | 0.87 (0.56 - 1.35) | 0.55 |
| MDCD | 1,793 | 4,717 | 0.9 | 3.1 | 57 | 217 | 0.80 (0.58 - 1.08) | 0.16 | 0.77 (0.56 - 1.04) | 0.10 |
| Open Claims | 230,071 | 371,526 | 169.4 | 318.5 | 3,165 | 8,067 | 0.86 (0.82 - 0.90) | <0.01 | 0.90 (0.80 - 1.00) | 0.07 |
| OEHR | 14,218 | 29,436 | 4.4 | 14.8 | 177 | 720 | 0.87 (0.72 - 1.03) | 0.11 | 0.92 (0.77 - 1.10) | 0.39 |
| SIDIAP | 2,780 | 13,682 | 3.8 | 29.7 | 58 | 451 | 0.82 (0.60 - 1.09) | 0.19 | 0.82 (0.61 - 1.10) | 0.24 |
| Meta-analysis | 276,677 | 461,626 | 197.7 | 400.1 | 4,040 | 10,484 | 0.86 (0.82 - 0.89) | <0.01 | 0.88 (0.80 - 0.97) | 0.01 |
| Meta-analysis (w/o USOC) | 46,606 | 90,100 | 28.3 | 81.5 | 875 | 2,417 | 0.86 (0.79 - 0.93) | <0.01 | 0.88 (0.80 - 0.96) | <0.01 |
| **PS-matching / on-treatment TAR without escalation** | | | | | | | | | | |
| CCAE | 13,703 | 21,595 | 8.9 | 13.0 | 282 | 479 | 0.86 (0.70 - 1.06) | 0.16 | 0.85 (0.69 - 1.05) | 0.18 |
| DAG | 2,883 | 8,641 | 2.1 | 8.2 | 7 | 21 | 1.45 (0.43 - 4.29) | 0.53 | 1.35 (0.42 - 4.27) | 0.61 |
| ODOD | 8,810 | 16,500 | 5.0 | 10.3 | 213 | 522 | 0.83 (0.66 - 1.04) | 0.11 | 0.86 (0.68 - 1.08) | 0.21 |
| MDCR | 1,060 | 3,818 | 0.6 | 2.7 | 24 | 114 | 0.81 (0.43 - 1.45) | 0.50 | 0.76 (0.41 - 1.40) | 0.38 |
| MDCD | 1,724 | 4,705 | 0.8 | 2.5 | 45 | 177 | 0.73 (0.46 - 1.13) | 0.17 | 0.71 (0.45 - 1.11) | 0.15 |
| Open Claims | 193,021 | 370,847 | 127.1 | 254.9 | 2,324 | 6,448 | 0.83 (0.78 - 0.89) | <0.01 | 0.87 (0.80 - 0.94) | 0.01 |
| OEHR | 12,832 | 28,602 | 3.5 | 11.7 | 131 | 549 | 0.73 (0.56 - 0.95) | 0.02 | 0.75 (0.57 - 0.98) | 0.05 |
| SIDIAP | 2,601 | 13,232 | 3.2 | 22.5 | 46 | 340 | 0.76 (0.50 - 1.15) | 0.21 | 0.88 (0.58 - 1.34) | 0.57 |
| Meta-analysis | 236,667 | 468,127 | 151.4 | 326.9 | 3,067 | 8,624 | 0.83 (0.78 - 0.88) | <0.01 | 0.85 (0.78 - 0.93) | <0.01 |
| Meta-analysis (w/o USOC) | 43,646 | 97,280 | 24.3 | 72.0 | 743 | 2,176 | 0.81 (0.72 - 0.92) | <0.01 | 0.82 (0.73 - 0.93) | <0.01 |
| **PS-stratification / on-treatment TAR without escalation** | | | | | | | | | | |
| CCAE | 15,864 | 21,619 | 9.9 | 13.0 | 322 | 479 | 0.87 (0.75 - 1.01) | 0.06 | 0.84 (0.73 - 0.98) | 0.04 |
| DAG | 3,170 | 8,679 | 2.2 | 8.2 | 7 | 22 | 1.51 (0.55 - 3.71) | 0.40 | 1.31 (0.51 - 3.40) | 0.58 |
| ODOD | 10,574 | 16,684 | 5.7 | 10.4 | 234 | 530 | 0.90 (0.76 - 1.06) | 0.19 | 0.90 (0.76 - 1.06) | 0.23 |
| MDCR | 1,217 | 3,941 | 0.7 | 2.8 | 26 | 121 | 0.89 (0.55 - 1.39) | 0.61 | 0.87 (0.55 - 1.39) | 0.57 |
| MDCD | 1,793 | 4,717 | 0.8 | 2.5 | 47 | 178 | 0.77 (0.54 - 1.07) | 0.12 | 0.73 (0.52 - 1.02) | 0.07 |
| Open Claims | 230,071 | 371,526 | 143.4 | 255.4 | 2,586 | 6,473 | 0.84 (0.80 - 0.88) | <0.01 | 0.87 (0.80 - 0.95) | <0.01 |
| OEHR | 14,218 | 29,436 | 3.8 | 12.0 | 147 | 568 | 0.84 (0.69 - 1.01) | 0.07 | 0.87 (0.72 - 1.07) | 0.23 |
| SIDIAP | 2,780 | 13,682 | 3.3 | 23.6 | 50 | 357 | 0.80 (0.58 - 1.10) | 0.18 | 0.84 (0.61 - 1.16) | 0.35 |
| Meta-analysis | 276,677 | 461,626 | 168.3 | 322.7 | 3,346 | 8,429 | 0.84 (0.81 - 0.88) | <0.01 | 0.86 (0.78 - 0.95) | <0.01 |
| Meta-analysis (w/o USOC) | 46,606 | 90,100 | 24.9 | 67.3 | 760 | 1,956 | 0.87 (0.79 - 0.95) | <0.01 | 0.87 (0.79 - 0.96) | <0.01 |

##### 1.2.16 Genitourinary infection

**Supplementary Table 42:** Exposures, outcomes and risk estimates for Genitourinary infection in SGLT2I (female) (T) vs DPP4I (female) (C) new-user cohorts under four different propensity score (PS) adjustment / time-at-risk (TAR) designs across data sources. We report total patient exposure time in 1,000 patient-year (KYr) units, uncalibrated and calibrated hazard ratios (HRs) and their 95% confidence intervals (CIs). Greyed out entries did not pass study diagnostics and are included only for completeness.

| | Patients | | Exposure (KYr) | | Outcomes | | Uncalibrated | | Calibrated | |
|---|---|---|---|---|---|---|---|---|---|---|
| | T | C | T | C | T | C | HR (95% CI) | $p$ | HR (95% CI) | $p$ |
| **PS-matching / on-treatment TAR** | | | | | | | | | | |
| CCAE | 11,146 | 17,561 | 8.0 | 12.5 | 498 | 857 | 0.96 (0.82 - 1.13) | 0.63 | 0.98 (0.83 - 1.15) | 0.80 |
| DAG | 1,996 | 5,825 | 1.6 | 5.8 | 83 | 302 | 1.37 (1.00 - 1.88) | 0.05 | 1.20 (0.87 - 1.66) | 0.33 |
| ODOD | 7,390 | 13,219 | 4.6 | 9.5 | 365 | 945 | 0.91 (0.76 - 1.08) | 0.27 | 0.94 (0.79 - 1.12) | 0.53 |
| MDCR | 963 | 3,293 | 0.6 | 2.6 | 51 | 246 | 0.95 (0.63 - 1.43) | 0.83 | 0.94 (0.62 - 1.42) | 0.75 |
| MDCD | 1,545 | 4,091 | 0.8 | 2.7 | 78 | 265 | 0.84 (0.58 - 1.19) | 0.34 | 0.82 (0.57 - 1.17) | 0.31 |
| Open Claims | 153,187 | 292,252 | 121.4 | 246.4 | 4,837 | 13,138 | 0.91 (0.87 - 0.95) | <0.01 | 0.95 (0.85 - 1.06) | 0.39 |
| OEHR | 11,514 | 25,196 | 3.5 | 12.5 | 227 | 934 | 0.95 (0.77 - 1.15) | 0.58 | 0.96 (0.78 - 1.17) | 0.69 |
| SIDIAP | 1,802 | 9,133 | 2.6 | 18.9 | 146 | 1,140 | 1.11 (0.89 - 1.37) | 0.34 | 1.18 (0.95 - 1.46) | 0.21 |
| Meta-analysis | 189,714 | 371,177 | 143.5 | 312.7 | 6,258 | 17,692 | 0.95 (0.89 - 1.01) | 0.11 | 0.98 (0.89 - 1.08) | 0.73 |
| Meta-analysis (w/o USOC) | 36,527 | 78,925 | 22.2 | 66.4 | 1,421 | 4,554 | 0.98 (0.89 - 1.09) | 0.77 | 1.01 (0.91 - 1.12) | 0.81 |
| **PS-stratification / on-treatment TAR** | | | | | | | | | | |
| CCAE | 12,966 | 17,577 | 9.0 | 12.5 | 553 | 857 | 0.91 (0.81 - 1.01) | 0.09 | 0.91 (0.81 - 1.02) | 0.16 |
| DAG | 2,201 | 5,902 | 1.7 | 5.9 | 89 | 311 | 1.17 (0.90 - 1.51) | 0.23 | 1.03 (0.79 - 1.33) | 0.67 |
| ODOD | 8,866 | 13,327 | 5.2 | 9.5 | 402 | 959 | 0.87 (0.76 - 0.98) | 0.03 | 0.89 (0.78 - 1.01) | 0.09 |
| MDCR | 1,106 | 3,317 | 0.7 | 2.6 | 54 | 249 | 1.03 (0.74 - 1.42) | 0.84 | 1.05 (0.76 - 1.45) | 0.76 |
| MDCD | 1,602 | 4,111 | 0.8 | 2.7 | 79 | 269 | 0.94 (0.72 - 1.21) | 0.63 | 0.90 (0.69 - 1.17) | 0.44 |
| Open Claims | 183,262 | 292,863 | 136.5 | 246.8 | 5,341 | 13,216 | 0.89 (0.86 - 0.92) | <0.01 | 0.94 (0.84 - 1.04) | 0.26 |
| OEHR | 12,672 | 26,236 | 3.8 | 12.9 | 254 | 987 | 0.99 (0.85 - 1.14) | 0.87 | 1.05 (0.90 - 1.22) | 0.61 |
| SIDIAP | 1,919 | 9,437 | 2.7 | 19.8 | 154 | 1,212 | 1.04 (0.87 - 1.25) | 0.64 | 1.04 (0.86 - 1.26) | 0.65 |
| Meta-analysis | 221,886 | 365,342 | 158.9 | 307.5 | 6,793 | 17,542 | 0.92 (0.87 - 0.97) | <0.01 | 0.95 (0.85 - 1.06) | 0.33 |
| Meta-analysis (w/o USOC) | 38,624 | 72,479 | 22.4 | 60.6 | 1,452 | 4,326 | 0.96 (0.88 - 1.04) | 0.31 | 0.98 (0.89 - 1.07) | 0.68 |
| **PS-matching / on-treatment TAR without escalation** | | | | | | | | | | |
| CCAE | 11,146 | 17,561 | 7.0 | 10.3 | 435 | 706 | 0.99 (0.83 - 1.17) | 0.90 | 0.97 (0.81 - 1.16) | 0.79 |
| DAG | 1,996 | 5,825 | 1.4 | 5.2 | 75 | 279 | 1.40 (1.00 - 1.94) | 0.04 | 1.31 (0.94 - 1.82) | 0.15 |
| ODOD | 7,390 | 13,219 | 4.1 | 8.1 | 321 | 805 | 0.95 (0.79 - 1.14) | 0.59 | 0.98 (0.82 - 1.18) | 0.80 |
| MDCR | 963 | 3,293 | 0.6 | 2.3 | 46 | 202 | 0.98 (0.63 - 1.49) | 0.92 | 0.92 (0.60 - 1.41) | 0.70 |
| MDCD | 1,545 | 4,091 | 0.7 | 2.1 | 58 | 192 | 0.68 (0.44 - 1.04) | 0.08 | 0.66 (0.43 - 1.02) | 0.07 |
| Open Claims | 153,187 | 292,252 | 101.8 | 197.4 | 4,088 | 10,580 | 0.93 (0.88 - 0.98) | <0.01 | 0.97 (0.90 - 1.03) | 0.50 |
| OEHR | 11,514 | 25,196 | 3.0 | 10.1 | 196 | 727 | 1.01 (0.81 - 1.26) | 0.92 | 1.03 (0.83 - 1.29) | 0.74 |
| SIDIAP | 1,802 | 9,133 | 2.2 | 15.2 | 134 | 905 | 1.19 (0.94 - 1.49) | 0.15 | 1.36 (1.07 - 1.72) | 0.03 |
| Meta-analysis | 189,714 | 371,177 | 121.3 | 252.2 | 5,329 | 14,286 | 0.98 (0.90 - 1.07) | 0.70 | 1.01 (0.91 - 1.13) | 0.79 |
| Meta-analysis (w/o USOC) | 36,527 | 78,925 | 19.5 | 54.8 | 1,241 | 3,706 | 1.02 (0.90 - 1.16) | 0.79 | 1.03 (0.91 - 1.18) | 0.67 |
| **PS-stratification / on-treatment TAR without escalation** | | | | | | | | | | |
| CCAE | 12,966 | 17,577 | 7.8 | 10.3 | 485 | 706 | 0.93 (0.82 - 1.04) | 0.21 | 0.90 (0.79 - 1.02) | 0.13 |
| DAG | 2,201 | 5,902 | 1.5 | 5.3 | 81 | 288 | 1.13 (0.86 - 1.48) | 0.37 | 0.98 (0.75 - 1.29) | 0.68 |
| ODOD | 8,866 | 13,327 | 4.7 | 8.2 | 355 | 819 | 0.88 (0.77 - 1.00) | 0.06 | 0.88 (0.77 - 1.01) | 0.09 |
| MDCR | 1,106 | 3,317 | 0.6 | 2.3 | 49 | 205 | 1.12 (0.78 - 1.58) | 0.53 | 1.10 (0.77 - 1.56) | 0.61 |
| MDCD | 1,602 | 4,111 | 0.7 | 2.2 | 59 | 195 | 0.88 (0.64 - 1.18) | 0.40 | 0.83 (0.61 - 1.13) | 0.26 |
| Open Claims | 183,262 | 292,863 | 115.3 | 197.8 | 4,529 | 10,651 | 0.91 (0.87 - 0.94) | <0.01 | 0.95 (0.87 - 1.03) | 0.27 |
| OEHR | 12,672 | 26,236 | 3.3 | 10.5 | 218 | 770 | 1.01 (0.86 - 1.19) | 0.91 | 1.06 (0.89 - 1.25) | 0.58 |
| SIDIAP | 1,919 | 9,437 | 2.3 | 15.9 | 141 | 968 | 1.10 (0.90 - 1.33) | 0.35 | 1.15 (0.94 - 1.40) | 0.26 |
| Meta-analysis | 221,886 | 365,342 | 135.0 | 247.9 | 5,809 | 14,202 | 0.94 (0.89 - 0.99) | 0.03 | 0.96 (0.86 - 1.07) | 0.44 |
| Meta-analysis (w/o USOC) | 38,624 | 72,479 | 19.7 | 50.2 | 1,280 | 3,551 | 0.97 (0.89 - 1.06) | 0.51 | 0.97 (0.89 - 1.07) | 0.60 |

##### 1.2.17 Hyperkalemia

**Supplementary Table 43:** Exposures, outcomes and risk estimates for Hyperkalemia in SGLT2I (female) (T) vs DPP4I (female) (C) new-user cohorts under four different propensity score (PS) adjustment / time-at-risk (TAR) designs across data sources. We report total patient exposure time in 1,000 patient-year (KYr) units, uncalibrated and calibrated hazard ratios (HRs) and their 95% confidence intervals (CIs). Greyed out entries did not pass study diagnostics and are included only for completeness.

| | Patients | | Exposure (KYr) | | Outcomes | | Uncalibrated | | Calibrated | |
|---|---|---|---|---|---|---|---|---|---|---|
| | T | C | T | C | T | C | HR (95% CI) | $p$ | HR (95% CI) | $p$ |
| **PS-matching / on-treatment TAR** | | | | | | | | | | |
| CCAE | 16,149 | 25,490 | 12.1 | 19.0 | 74 | 127 | 0.85 (0.54 - 1.32) | 0.47 | 0.86 (0.55 - 1.36) | 0.54 |
| DAG | 2,759 | 8,171 | 2.2 | 8.5 | 25 | 145 | 0.75 (0.43 - 1.26) | 0.29 | 0.66 (0.38 - 1.13) | 0.15 |
| ODOD | 10,330 | 19,477 | 6.7 | 14.6 | 98 | 361 | 0.80 (0.58 - 1.09) | 0.17 | 0.83 (0.60 - 1.14) | 0.27 |
| MDCR | 1,274 | 4,650 | 0.9 | 3.9 | 5 | 66 | 0.51 (0.17 - 1.30) | 0.20 | 0.50 (0.18 - 1.40) | 0.20 |
| MDCD | 2,274 | 6,148 | 1.2 | 4.2 | 7 | 69 | 0.39 (0.13 - 0.96) | 0.07 | 0.38 (0.14 - 1.04) | 0.06 |
| Open Claims | 227,815 | 435,705 | 179.5 | 378.9 | 641 | 2,940 | 0.66 (0.59 - 0.75) | <0.01 | 0.69 (0.59 - 0.81) | <0.01 |
| OEHR | 14,821 | 33,041 | 4.7 | 17.3 | 30 | 225 | 0.88 (0.54 - 1.37) | 0.57 | 0.89 (0.56 - 1.41) | 0.63 |
| SIDIAP | 2,677 | 13,494 | 3.7 | 28.9 | 35 | 233 | 0.94 (0.58 - 1.49) | 0.81 | 1.00 (0.62 - 1.61) | 0.84 |
| Meta-analysis | 276,825 | 541,526 | 210.1 | 471.4 | 910 | 4,100 | 0.73 (0.64 - 0.85) | <0.01 | 0.76 (0.65 - 0.90) | <0.01 |
| Meta-analysis (w/o USOC) | 49,010 | 105,821 | 30.6 | 92.5 | 269 | 1,160 | 0.81 (0.68 - 0.98) | 0.03 | 0.83 (0.69 - 1.01) | 0.07 |
| **PS-stratification / on-treatment TAR** | | | | | | | | | | |
| CCAE | 18,560 | 25,525 | 13.4 | 19.1 | 76 | 127 | 0.96 (0.71 - 1.29) | 0.78 | 0.96 (0.72 - 1.30) | 0.81 |
| DAG | 3,033 | 8,195 | 2.4 | 8.5 | 26 | 145 | 0.68 (0.43 - 1.05) | 0.09 | 0.60 (0.38 - 0.93) | 0.03 |
| ODOD | 12,381 | 19,703 | 7.6 | 14.7 | 109 | 371 | 0.79 (0.62 - 0.99) | 0.04 | 0.81 (0.64 - 1.02) | 0.08 |
| MDCR | 1,440 | 4,709 | 0.9 | 4.0 | 5 | 71 | 0.38 (0.13 - 0.89) | 0.05 | 0.38 (0.14 - 1.01) | 0.05 |
| MDCD | 2,363 | 6,166 | 1.3 | 4.2 | 7 | 70 | 0.44 (0.18 - 0.92) | 0.05 | 0.42 (0.19 - 0.95) | 0.04 |
| Open Claims | 271,518 | 436,158 | 201.2 | 379.3 | 717 | 2,958 | 0.66 (0.60 - 0.72) | <0.01 | 0.69 (0.61 - 0.79) | <0.01 |
| OEHR | 16,395 | 34,043 | 5.1 | 17.8 | 32 | 237 | 0.64 (0.43 - 0.94) | 0.03 | 0.68 (0.46 - 1.01) | 0.06 |
| SIDIAP | 2,857 | 13,932 | 3.9 | 30.2 | 40 | 254 | 1.31 (0.90 - 1.87) | 0.15 | 1.31 (0.90 - 1.90) | 0.19 |
| Meta-analysis | 324,744 | 537,556 | 233.5 | 469.7 | 1,000 | 4,092 | 0.80 (0.65 - 0.99) | 0.04 | 0.82 (0.66 - 1.03) | 0.10 |
| Meta-analysis (w/o USOC) | 53,226 | 101,398 | 32.3 | 90.3 | 283 | 1,134 | 0.85 (0.68 - 1.07) | 0.18 | 0.88 (0.70 - 1.10) | 0.26 |
| **PS-matching / on-treatment TAR without escalation** | | | | | | | | | | |
| CCAE | 16,149 | 25,490 | 10.4 | 15.5 | 65 | 102 | 0.81 (0.49 - 1.30) | 0.39 | 0.80 (0.49 - 1.30) | 0.38 |
| DAG | 2,759 | 8,171 | 2.0 | 7.6 | 22 | 128 | 0.74 (0.41 - 1.29) | 0.31 | 0.69 (0.39 - 1.23) | 0.23 |
| ODOD | 10,330 | 19,477 | 5.9 | 12.3 | 86 | 299 | 0.80 (0.57 - 1.11) | 0.19 | 0.82 (0.59 - 1.15) | 0.27 |
| MDCR | 1,274 | 4,650 | 0.8 | 3.4 | 5 | 57 | 0.50 (0.16 - 1.30) | 0.20 | 0.47 (0.17 - 1.33) | 0.16 |
| MDCD | 2,274 | 6,148 | 1.0 | 3.3 | 7 | 55 | 0.48 (0.16 - 1.23) | 0.16 | 0.47 (0.17 - 1.31) | 0.15 |
| Open Claims | 227,815 | 435,705 | 150.0 | 301.3 | 511 | 2,333 | 0.67 (0.58 - 0.76) | <0.01 | 0.69 (0.60 - 0.80) | <0.01 |
| OEHR | 14,821 | 33,041 | 4.1 | 13.9 | 25 | 167 | 0.87 (0.51 - 1.43) | 0.59 | 0.89 (0.53 - 1.49) | 0.66 |
| SIDIAP | 2,677 | 13,494 | 3.3 | 22.9 | 31 | 195 | 0.93 (0.56 - 1.51) | 0.79 | 1.07 (0.65 - 1.77) | 0.77 |
| Meta-analysis | 276,825 | 541,526 | 176.7 | 376.8 | 747 | 3,279 | 0.72 (0.63 - 0.83) | <0.01 | 0.75 (0.64 - 0.87) | <0.01 |
| Meta-analysis (w/o USOC) | 49,010 | 105,821 | 26.7 | 75.5 | 236 | 946 | 0.81 (0.66 - 0.99) | 0.04 | 0.82 (0.67 - 1.00) | 0.06 |
| **PS-stratification / on-treatment TAR without escalation** | | | | | | | | | | |
| CCAE | 18,560 | 25,525 | 11.5 | 15.6 | 67 | 102 | 1.02 (0.74 - 1.41) | 0.91 | 0.99 (0.72 - 1.37) | 0.92 |
| DAG | 3,033 | 8,195 | 2.2 | 7.7 | 23 | 128 | 0.65 (0.40 - 1.02) | 0.07 | 0.56 (0.35 - 0.91) | 0.02 |
| ODOD | 12,381 | 19,703 | 6.7 | 12.4 | 97 | 308 | 0.81 (0.63 - 1.03) | 0.09 | 0.81 (0.63 - 1.04) | 0.11 |
| MDCR | 1,440 | 4,709 | 0.8 | 3.5 | 5 | 61 | 0.43 (0.15 - 1.04) | 0.10 | 0.43 (0.16 - 1.14) | 0.09 |
| MDCD | 2,363 | 6,166 | 1.0 | 3.3 | 7 | 56 | 0.52 (0.21 - 1.09) | 0.12 | 0.49 (0.21 - 1.11) | 0.09 |
| Open Claims | 271,518 | 436,158 | 169.2 | 301.7 | 580 | 2,348 | 0.65 (0.59 - 0.71) | <0.01 | 0.68 (0.60 - 0.77) | <0.01 |
| OEHR | 16,395 | 34,043 | 4.5 | 14.3 | 26 | 174 | 0.64 (0.41 - 0.98) | 0.05 | 0.67 (0.43 - 1.04) | 0.09 |
| SIDIAP | 2,857 | 13,932 | 3.4 | 24.0 | 35 | 212 | 1.21 (0.80 - 1.77) | 0.36 | 1.26 (0.84 - 1.88) | 0.29 |
| Meta-analysis | 324,744 | 537,556 | 197.5 | 375.6 | 828 | 3,272 | 0.79 (0.65 - 0.98) | 0.03 | 0.81 (0.65 - 1.02) | 0.07 |
| Meta-analysis (w/o USOC) | 53,226 | 101,398 | 28.3 | 73.9 | 248 | 924 | 0.86 (0.69 - 1.06) | 0.16 | 0.86 (0.69 - 1.07) | 0.18 |

##### 1.2.18 Hypoglycemia

**Supplementary Table 44:** Exposures, outcomes and risk estimates for Hypoglycemia in SGLT2I (female) (T) vs DPP4I (female) (C) new-user cohorts under four different propensity score (PS) adjustment / time-at-risk (TAR) designs across data sources. We report total patient exposure time in 1,000 patient-year (KYr) units, uncalibrated and calibrated hazard ratios (HRs) and their 95% confidence intervals (CIs). Greyed out entries did not pass study diagnostics and are included only for completeness.

| | Patients | | Exposure (KYr) | | Outcomes | | Uncalibrated | | Calibrated | |
|---|---|---|---|---|---|---|---|---|---|---|
| | T | C | T | C | T | C | HR (95% CI) | $p$ | HR (95% CI) | $p$ |
| **PS-matching / on-treatment TAR** | | | | | | | | | | |
| CCAE | 16,052 | 25,420 | 12.0 | 19.1 | 46 | 73 | 1.19 (0.72 - 1.96) | 0.50 | 1.21 (0.73 - 2.01) | 0.47 |
| DAG | 2,924 | 8,714 | 2.4 | 9.2 | <5 | 26 | 0.38 (0.06 - 1.45) | 0.24 | 0.34 (0.07 - 1.67) | 0.19 |
| ODOD | 10,519 | 20,248 | 6.8 | 15.5 | 46 | 135 | 0.84 (0.51 - 1.33) | 0.46 | 0.87 (0.53 - 1.40) | 0.56 |
| MDCR | 1,288 | 4,765 | 0.9 | 4.0 | 0 | 25 | | | | |
| MDCD | 2,263 | 6,160 | 1.2 | 4.3 | 12 | 47 | 0.84 (0.34 - 1.91) | 0.69 | 0.82 (0.34 - 1.94) | 0.65 |
| Open Claims | 227,219 | 437,467 | 179.1 | 383.0 | 367 | 1,254 | 0.82 (0.69 - 0.97) | 0.02 | 0.85 (0.70 - 1.04) | 0.13 |
| OEHR | 14,853 | 33,249 | 4.7 | 17.5 | 17 | 131 | 0.56 (0.28 - 1.05) | 0.09 | 0.57 (0.29 - 1.11) | 0.10 |
| SIDIAP | 2,773 | 14,240 | 3.9 | 30.8 | 0 | 28 | | | | |
| Meta-analysis | 273,830 | 531,258 | 206.3 | 448.5 | <493 | 1,666 | 0.83 (0.71 - 0.96) | 0.01 | 0.86 (0.73 - 1.01) | 0.07 |
| Meta-analysis (w/o USOC) | 46,611 | 93,791 | 27.2 | 65.5 | <126 | 412 | 0.85 (0.63 - 1.13) | 0.26 | 0.87 (0.65 - 1.16) | 0.35 |
| **PS-stratification / on-treatment TAR** | | | | | | | | | | |
| CCAE | 18,425 | 25,454 | 13.3 | 19.1 | 50 | 73 | 0.95 (0.65 - 1.38) | 0.81 | 0.96 (0.66 - 1.40) | 0.83 |
| DAG | 3,226 | 8,750 | 2.5 | 9.2 | <5 | 26 | 0.47 (0.11 - 1.42) | 0.25 | 0.41 (0.11 - 1.50) | 0.18 |
| ODOD | 12,580 | 20,448 | 7.7 | 15.6 | 50 | 137 | 0.93 (0.65 - 1.31) | 0.67 | 0.95 (0.67 - 1.35) | 0.78 |
| MDCR | 1,462 | 4,831 | 0.9 | 4.1 | 0 | 25 | | | | |
| MDCD | 2,349 | 6,175 | 1.3 | 4.3 | 12 | 47 | 0.86 (0.43 - 1.61) | 0.66 | 0.83 (0.43 - 1.60) | 0.57 |
| Open Claims | 270,936 | 438,007 | 200.8 | 383.5 | 416 | 1,258 | 0.77 (0.68 - 0.87) | <0.01 | 0.81 (0.69 - 0.95) | 0.01 |
| OEHR | 16,444 | 34,279 | 5.2 | 18.0 | 18 | 133 | 0.57 (0.33 - 0.94) | 0.04 | 0.61 (0.36 - 1.02) | 0.07 |
| SIDIAP | 2,950 | 14,724 | 4.0 | 32.3 | <5 | 29 | 0.19 (0.01 - 0.98) | 0.16 | 0.19 (0.02 - 1.87) | 0.16 |
| Meta-analysis | 324,561 | 541,662 | 233.4 | 477.8 | <544 | 1,656 | 0.78 (0.70 - 0.87) | <0.01 | 0.80 (0.70 - 0.93) | <0.01 |
| Meta-analysis (w/o USOC) | 53,625 | 103,655 | 32.7 | 94.3 | <128 | 398 | 0.80 (0.60 - 1.06) | 0.12 | 0.82 (0.61 - 1.08) | 0.17 |
| **PS-matching / on-treatment TAR without escalation** | | | | | | | | | | |
| CCAE | 16,052 | 25,420 | 10.3 | 15.6 | 34 | 41 | 1.12 (0.60 - 2.08) | 0.72 | 1.10 (0.59 - 2.05) | 0.76 |
| DAG | 2,924 | 8,714 | 2.2 | 8.3 | <5 | 23 | 0.50 (0.07 - 1.92) | 0.40 | 0.46 (0.09 - 2.34) | 0.36 |
| ODOD | 10,519 | 20,248 | 6.0 | 13.0 | 33 | 76 | 1.05 (0.57 - 1.86) | 0.88 | 1.08 (0.60 - 1.95) | 0.80 |
| MDCR | 1,288 | 4,765 | 0.8 | 3.5 | 0 | 16 | | | | |
| MDCD | 2,263 | 6,160 | 1.0 | 3.3 | 9 | 25 | 0.82 (0.28 - 2.14) | 0.71 | 0.80 (0.29 - 2.20) | 0.67 |
| Open Claims | 227,219 | 437,467 | 149.6 | 304.8 | 238 | 682 | 0.89 (0.72 - 1.09) | 0.27 | 0.93 (0.75 - 1.14) | 0.50 |
| OEHR | 14,853 | 33,249 | 4.1 | 14.1 | 9 | 63 | 0.41 (0.15 - 0.94) | 0.06 | 0.42 (0.17 - 1.05) | 0.06 |
| SIDIAP | 2,773 | 14,240 | 3.4 | 24.4 | 0 | 12 | | | | |
| Meta-analysis | 273,830 | 531,258 | 173.3 | 359.0 | <328 | 910 | 0.89 (0.74 - 1.06) | 0.19 | 0.91 (0.76 - 1.11) | 0.36 |
| Meta-analysis (w/o USOC) | 46,611 | 93,791 | 23.6 | 54.3 | <90 | 228 | 0.87 (0.61 - 1.25) | 0.45 | 0.88 (0.61 - 1.27) | 0.50 |
| **PS-stratification / on-treatment TAR without escalation** | | | | | | | | | | |
| CCAE | 18,425 | 25,454 | 11.5 | 15.6 | 37 | 41 | 1.12 (0.70 - 1.78) | 0.64 | 1.08 (0.68 - 1.73) | 0.74 |
| DAG | 3,226 | 8,750 | 2.3 | 8.3 | <5 | 23 | 0.34 (0.05 - 1.26) | 0.19 | 0.30 (0.06 - 1.46) | 0.14 |
| ODOD | 12,580 | 20,448 | 6.8 | 13.2 | 37 | 77 | 1.07 (0.69 - 1.64) | 0.76 | 1.07 (0.69 - 1.66) | 0.75 |
| MDCR | 1,462 | 4,831 | 0.8 | 3.6 | 0 | 16 | | | | |
| MDCD | 2,349 | 6,175 | 1.0 | 3.4 | 9 | 25 | 1.16 (0.50 - 2.49) | 0.71 | 1.10 (0.49 - 2.46) | 0.81 |
| Open Claims | 270,936 | 438,007 | 168.9 | 305.2 | 277 | 684 | 0.83 (0.71 - 0.96) | 0.01 | 0.87 (0.73 - 1.02) | 0.11 |
| OEHR | 16,444 | 34,279 | 4.5 | 14.5 | 9 | 63 | 0.45 (0.20 - 0.88) | 0.03 | 0.47 (0.22 - 0.98) | 0.05 |
| SIDIAP | 2,950 | 14,724 | 3.5 | 25.6 | <5 | 12 | 0.41 (0.02 - 2.36) | 0.45 | 0.43 (0.04 - 4.48) | 0.48 |
| Meta-analysis | 321,611 | 526,938 | 194.0 | 356.8 | <365 | 888 | 0.85 (0.73 - 0.99) | 0.04 | 0.87 (0.73 - 1.04) | 0.13 |
| Meta-analysis (w/o USOC) | 50,675 | 88,931 | 25.1 | 51.6 | <88 | 204 | 0.82 (0.51 - 1.31) | 0.40 | 0.82 (0.51 - 1.31) | 0.41 |

##### 1.2.19 Hypotension

**Supplementary Table 45:** Exposures, outcomes and risk estimates for Hypotension in SGLT2I (female) (T) vs DPP4I (female) (C) new-user cohorts under four different propensity score (PS) adjustment / time-at-risk (TAR) designs across data sources. We report total patient exposure time in 1,000 patient-year (KYr) units, uncalibrated and calibrated hazard ratios (HRs) and their 95% confidence intervals (CIs). Greyed out entries did not pass study diagnostics and are included only for completeness.

| | Patients | | Exposure (KYr) | | Outcomes | | Uncalibrated | | Calibrated | |
|---|---|---|---|---|---|---|---|---|---|---|
| | T | C | T | C | T | C | HR (95% CI) | $p$ | HR (95% CI) | $p$ |
| **PS-matching / on-treatment TAR** | | | | | | | | | | |
| CCAE | 16,059 | 25,318 | 12.0 | 18.9 | 99 | 150 | 1.17 (0.82 - 1.65) | 0.38 | 1.19 (0.84 - 1.69) | 0.35 |
| DAG | 2,827 | 8,489 | 2.3 | 8.9 | 6 | 54 | 0.36 (0.12 - 0.88) | 0.05 | 0.32 (0.12 - 0.86) | 0.03 |
| ODOD | 10,269 | 19,416 | 6.7 | 14.6 | 130 | 365 | 1.23 (0.93 - 1.61) | 0.14 | 1.27 (0.97 - 1.68) | 0.09 |
| MDCR | 1,244 | 4,557 | 0.8 | 3.9 | 25 | 93 | 2.32 (1.19 - 4.47) | 0.01 | 2.28 (1.17 - 4.44) | 0.02 |
| MDCD | 2,178 | 5,890 | 1.2 | 4.0 | 39 | 120 | 1.56 (0.94 - 2.54) | 0.08 | 1.52 (0.92 - 2.50) | 0.11 |
| Open Claims | 224,435 | 428,978 | 176.9 | 373.7 | 1,160 | 3,602 | 0.94 (0.85 - 1.03) | 0.17 | 0.98 (0.85 - 1.12) | 0.75 |
| OEHR | 14,728 | 32,933 | 4.6 | 17.2 | 57 | 256 | 1.11 (0.74 - 1.65) | 0.60 | 1.13 (0.76 - 1.68) | 0.56 |
| SIDIAP | 2,758 | 14,150 | 3.8 | 30.6 | 8 | 59 | 1.43 (0.52 - 3.51) | 0.46 | 1.52 (0.59 - 3.93) | 0.40 |
| Meta-analysis | 274,695 | 540,426 | 208.9 | 474.2 | <1,504 | 4,630 | 1.08 (0.91 - 1.27) | 0.38 | 1.12 (0.93 - 1.34) | 0.23 |
| Meta-analysis (w/o USOC) | 50,260 | 111,448 | 32.0 | 100.5 | <344 | 1,028 | 1.18 (0.97 - 1.43) | 0.11 | 1.20 (0.99 - 1.47) | 0.08 |
| **PS-stratification / on-treatment TAR** | | | | | | | | | | |
| CCAE | 18,471 | 25,349 | 13.3 | 19.0 | 110 | 150 | 1.20 (0.92 - 1.55) | 0.17 | 1.20 (0.93 - 1.56) | 0.18 |
| DAG | 3,116 | 8,520 | 2.4 | 9.0 | 7 | 55 | 0.48 (0.19 - 1.05) | 0.09 | 0.42 (0.18 - 0.99) | 0.05 |
| ODOD | 12,292 | 19,645 | 7.5 | 14.8 | 148 | 379 | 1.13 (0.91 - 1.38) | 0.27 | 1.15 (0.93 - 1.42) | 0.20 |
| MDCR | 1,415 | 4,606 | 0.9 | 3.9 | 25 | 98 | 1.37 (0.83 - 2.20) | 0.21 | 1.39 (0.85 - 2.27) | 0.20 |
| MDCD | 2,263 | 5,907 | 1.2 | 4.0 | 42 | 120 | 1.26 (0.85 - 1.81) | 0.23 | 1.20 (0.82 - 1.75) | 0.35 |
| Open Claims | 267,171 | 429,492 | 198.1 | 374.2 | 1,308 | 3,619 | 0.93 (0.87 - 1.00) | 0.04 | 0.98 (0.86 - 1.10) | 0.70 |
| OEHR | 16,280 | 33,949 | 5.1 | 17.7 | 60 | 269 | 1.10 (0.80 - 1.48) | 0.55 | 1.16 (0.85 - 1.58) | 0.36 |
| SIDIAP | 2,935 | 14,629 | 4.0 | 32.1 | 8 | 64 | 1.34 (0.55 - 2.84) | 0.49 | 1.34 (0.59 - 3.03) | 0.50 |
| Meta-analysis | 320,265 | 531,584 | 230.4 | 466.7 | 1,641 | 4,536 | 1.03 (0.90 - 1.18) | 0.67 | 1.06 (0.90 - 1.24) | 0.49 |
| Meta-analysis (w/o USOC) | 53,094 | 102,092 | 32.3 | 92.5 | 333 | 917 | 1.12 (0.96 - 1.30) | 0.14 | 1.14 (0.98 - 1.33) | 0.09 |
| **PS-matching / on-treatment TAR without escalation** | | | | | | | | | | |
| CCAE | 16,059 | 25,318 | 10.3 | 15.5 | 76 | 115 | 1.17 (0.79 - 1.75) | 0.43 | 1.16 (0.77 - 1.73) | 0.50 |
| DAG | 2,827 | 8,489 | 2.1 | 8.0 | 6 | 43 | 0.52 (0.17 - 1.32) | 0.21 | 0.48 (0.17 - 1.35) | 0.17 |
| ODOD | 10,269 | 19,416 | 5.9 | 12.4 | 110 | 308 | 1.28 (0.95 - 1.71) | 0.11 | 1.32 (0.98 - 1.77) | 0.08 |
| MDCR | 1,244 | 4,557 | 0.8 | 3.4 | 24 | 87 | 2.59 (1.28 - 5.14) | <0.01 | 2.42 (1.21 - 4.85) | 0.01 |
| MDCD | 2,178 | 5,890 | 1.0 | 3.2 | 29 | 80 | 1.14 (0.61 - 2.04) | 0.67 | 1.10 (0.60 - 2.01) | 0.75 |
| Open Claims | 224,435 | 428,978 | 147.9 | 297.3 | 936 | 2,869 | 0.95 (0.86 - 1.05) | 0.33 | 0.99 (0.88 - 1.10) | 0.85 |
| OEHR | 14,728 | 32,933 | 4.0 | 13.8 | 41 | 176 | 1.10 (0.68 - 1.73) | 0.70 | 1.12 (0.70 - 1.79) | 0.64 |
| SIDIAP | 2,758 | 14,150 | 3.4 | 24.2 | 7 | 47 | 1.46 (0.45 - 4.04) | 0.50 | 1.68 (0.56 - 5.05) | 0.36 |
| Meta-analysis | 274,695 | 540,426 | 175.9 | 379.4 | <1,210 | 3,659 | 1.05 (0.90 - 1.23) | 0.52 | 1.08 (0.92 - 1.28) | 0.35 |
| Meta-analysis (w/o USOC) | 50,260 | 111,448 | 27.9 | 82.1 | <274 | 790 | 1.17 (0.97 - 1.42) | 0.11 | 1.19 (0.98 - 1.44) | 0.09 |
| **PS-stratification / on-treatment TAR without escalation** | | | | | | | | | | |
| CCAE | 18,471 | 25,349 | 11.5 | 15.5 | 84 | 115 | 1.13 (0.84 - 1.52) | 0.41 | 1.10 (0.82 - 1.48) | 0.54 |
| DAG | 3,116 | 8,520 | 2.2 | 8.1 | 7 | 43 | 0.59 (0.23 - 1.33) | 0.24 | 0.52 (0.21 - 1.24) | 0.15 |
| ODOD | 12,292 | 19,645 | 6.6 | 12.5 | 126 | 320 | 1.13 (0.89 - 1.41) | 0.31 | 1.13 (0.90 - 1.42) | 0.31 |
| MDCR | 1,415 | 4,606 | 0.8 | 3.4 | 24 | 91 | 1.47 (0.87 - 2.40) | 0.14 | 1.44 (0.87 - 2.40) | 0.17 |
| MDCD | 2,263 | 5,907 | 1.0 | 3.2 | 32 | 80 | 1.29 (0.82 - 1.98) | 0.26 | 1.22 (0.78 - 1.89) | 0.39 |
| Open Claims | 267,171 | 429,492 | 166.7 | 297.7 | 1,066 | 2,886 | 0.91 (0.85 - 0.99) | 0.02 | 0.96 (0.86 - 1.06) | 0.44 |
| OEHR | 16,280 | 33,949 | 4.4 | 14.3 | 42 | 186 | 1.02 (0.70 - 1.45) | 0.93 | 1.06 (0.73 - 1.54) | 0.76 |
| SIDIAP | 2,935 | 14,629 | 3.5 | 25.4 | 7 | 51 | 1.43 (0.55 - 3.21) | 0.43 | 1.49 (0.62 - 3.61) | 0.38 |
| Meta-analysis | 320,265 | 531,584 | 195.0 | 373.5 | 1,332 | 3,601 | 1.00 (0.87 - 1.14) | 0.96 | 1.02 (0.87 - 1.19) | 0.80 |
| Meta-analysis (w/o USOC) | 53,094 | 102,092 | 28.3 | 75.8 | 266 | 715 | 1.09 (0.93 - 1.28) | 0.27 | 1.09 (0.93 - 1.28) | 0.28 |

##### 1.2.20 Joint pain

**Supplementary Table 46:** Exposures, outcomes and risk estimates for Joint pain in SGLT2I (female) (T) vs DPP4I (female) (C) new-user cohorts under four different propensity score (PS) adjustment / time-at-risk (TAR) designs across data sources. We report total patient exposure time in 1,000 patient-year (KYr) units, uncalibrated and calibrated hazard ratios (HRs) and their 95% confidence intervals (CIs). Greyed out entries did not pass study diagnostics and are included only for completeness.

| | Patients | | Exposure (KYr) | | Outcomes | | Uncalibrated | | Calibrated | |
|---|---|---|---|---|---|---|---|---|---|---|
| | T | C | T | C | T | C | HR (95% CI) | $p$ | HR (95% CI) | $p$ |
| **PS-matching / on-treatment TAR** | | | | | | | | | | |
| CCAE | 15,034 | 23,820 | 11.3 | 17.7 | 152 | 297 | 0.72 (0.53 - 0.96) | 0.03 | 0.73 (0.54 - 0.98) | 0.05 |
| DAG | 2,422 | 7,333 | 1.9 | 7.5 | 41 | 178 | 0.64 (0.38 - 1.02) | 0.07 | 0.56 (0.34 - 0.91) | 0.03 |
| ODOD | 9,854 | 18,883 | 6.4 | 14.2 | 112 | 286 | 0.97 (0.69 - 1.34) | 0.85 | 1.00 (0.72 - 1.40) | 0.90 |
| MDCR | 1,242 | 4,550 | 0.8 | 3.9 | 11 | 61 | 0.65 (0.21 - 1.70) | 0.42 | 0.63 (0.22 - 1.83) | 0.41 |
| MDCD | 2,068 | 5,622 | 1.1 | 3.8 | 30 | 122 | 0.69 (0.39 - 1.18) | 0.19 | 0.67 (0.39 - 1.17) | 0.18 |
| Open Claims | 213,048 | 411,581 | 168.2 | 358.5 | 1,309 | 3,498 | 0.83 (0.75 - 0.91) | <0.01 | 0.86 (0.75 - 0.99) | 0.05 |
| OEHR | 13,960 | 31,427 | 4.3 | 16.3 | 75 | 277 | 1.25 (0.85 - 1.82) | 0.25 | 1.27 (0.87 - 1.86) | 0.24 |
| SIDIAP | 1,982 | 10,479 | 2.8 | 22.0 | 58 | 606 | 0.78 (0.56 - 1.07) | 0.14 | 0.83 (0.60 - 1.15) | 0.30 |
| Meta-analysis | 259,656 | 513,802 | 197.3 | 445.5 | 1,784 | 5,325 | 0.83 (0.77 - 0.90) | <0.01 | 0.86 (0.78 - 0.96) | <0.01 |
| Meta-analysis (w/o USOC) | 46,608 | 102,221 | 29.1 | 87.0 | 475 | 1,827 | 0.83 (0.69 - 0.99) | 0.04 | 0.85 (0.70 - 1.02) | 0.08 |
| **PS-stratification / on-treatment TAR** | | | | | | | | | | |
| CCAE | 17,271 | 23,853 | 12.5 | 17.7 | 175 | 297 | 0.80 (0.66 - 0.97) | 0.03 | 0.80 (0.66 - 0.98) | 0.04 |
| DAG | 2,670 | 7,357 | 2.0 | 7.6 | 46 | 179 | 0.86 (0.60 - 1.22) | 0.41 | 0.75 (0.52 - 1.08) | 0.15 |
| ODOD | 11,750 | 19,065 | 7.2 | 14.4 | 129 | 288 | 0.93 (0.74 - 1.16) | 0.51 | 0.95 (0.76 - 1.19) | 0.67 |
| MDCR | 1,402 | 4,613 | 0.9 | 3.9 | 14 | 61 | 0.74 (0.37 - 1.39) | 0.38 | 0.75 (0.39 - 1.46) | 0.41 |
| MDCD | 2,157 | 5,641 | 1.2 | 3.8 | 31 | 123 | 0.78 (0.51 - 1.16) | 0.24 | 0.75 (0.49 - 1.13) | 0.17 |
| Open Claims | 253,928 | 411,935 | 188.5 | 358.8 | 1,465 | 3,504 | 0.84 (0.79 - 0.90) | <0.01 | 0.88 (0.78 - 1.00) | 0.05 |
| OEHR | 15,387 | 32,362 | 4.8 | 16.8 | 80 | 288 | 1.06 (0.81 - 1.38) | 0.67 | 1.12 (0.86 - 1.47) | 0.43 |
| SIDIAP | 2,113 | 10,863 | 2.9 | 23.1 | 59 | 654 | 0.79 (0.59 - 1.04) | 0.10 | 0.79 (0.59 - 1.05) | 0.15 |
| Meta-analysis | 303,119 | 505,435 | 217.8 | 438.3 | 1,954 | 5,210 | 0.85 (0.80 - 0.90) | <0.01 | 0.87 (0.79 - 0.97) | 0.01 |
| Meta-analysis (w/o USOC) | 49,191 | 93,500 | 29.3 | 79.5 | 489 | 1,706 | 0.88 (0.78 - 0.98) | 0.02 | 0.90 (0.80 - 1.01) | 0.08 |
| **PS-matching / on-treatment TAR without escalation** | | | | | | | | | | |
| CCAE | 15,034 | 23,820 | 9.7 | 14.4 | 129 | 241 | 0.79 (0.57 - 1.07) | 0.13 | 0.77 (0.56 - 1.06) | 0.14 |
| DAG | 2,422 | 7,333 | 1.7 | 6.8 | 39 | 167 | 0.65 (0.39 - 1.06) | 0.10 | 0.61 (0.37 - 1.01) | 0.07 |
| ODOD | 9,854 | 18,883 | 5.6 | 12.0 | 97 | 238 | 1.06 (0.74 - 1.50) | 0.75 | 1.09 (0.77 - 1.56) | 0.63 |
| MDCR | 1,242 | 4,550 | 0.7 | 3.4 | 10 | 52 | 0.74 (0.23 - 2.02) | 0.59 | 0.70 (0.24 - 2.05) | 0.51 |
| MDCD | 2,068 | 5,622 | 0.9 | 3.0 | 27 | 97 | 0.74 (0.40 - 1.33) | 0.33 | 0.72 (0.39 - 1.32) | 0.30 |
| Open Claims | 213,048 | 411,581 | 140.6 | 285.4 | 1,098 | 2,775 | 0.84 (0.76 - 0.94) | <0.01 | 0.88 (0.78 - 0.98) | 0.06 |
| OEHR | 13,960 | 31,427 | 3.8 | 13.1 | 68 | 215 | 1.51 (0.99 - 2.29) | 0.05 | 1.55 (1.01 - 2.35) | 0.05 |
| SIDIAP | 1,982 | 10,479 | 2.4 | 17.5 | 44 | 465 | 0.64 (0.42 - 0.92) | 0.02 | 0.73 (0.49 - 1.08) | 0.14 |
| Meta-analysis | 259,656 | 513,802 | 166.0 | 356.6 | 1,509 | 4,247 | 0.86 (0.71 - 1.04) | 0.11 | 0.89 (0.73 - 1.08) | 0.24 |
| Meta-analysis (w/o USOC) | 46,608 | 102,221 | 25.4 | 71.2 | 411 | 1,472 | 0.86 (0.68 - 1.10) | 0.24 | 0.88 (0.69 - 1.12) | 0.30 |
| **PS-stratification / on-treatment TAR without escalation** | | | | | | | | | | |
| CCAE | 17,271 | 23,853 | 10.8 | 14.5 | 150 | 241 | 0.80 (0.64 - 0.99) | 0.04 | 0.77 (0.63 - 0.96) | 0.03 |
| DAG | 2,670 | 7,357 | 1.8 | 6.8 | 44 | 167 | 0.89 (0.61 - 1.28) | 0.54 | 0.78 (0.54 - 1.12) | 0.21 |
| ODOD | 11,750 | 19,065 | 6.3 | 12.1 | 113 | 240 | 0.95 (0.74 - 1.21) | 0.66 | 0.95 (0.74 - 1.21) | 0.68 |
| MDCR | 1,402 | 4,613 | 0.8 | 3.4 | 13 | 52 | 0.85 (0.41 - 1.64) | 0.64 | 0.83 (0.41 - 1.67) | 0.61 |
| MDCD | 2,157 | 5,641 | 1.0 | 3.0 | 28 | 97 | 0.86 (0.54 - 1.31) | 0.49 | 0.81 (0.52 - 1.26) | 0.36 |
| Open Claims | 253,928 | 411,935 | 158.6 | 285.6 | 1,233 | 2,781 | 0.86 (0.80 - 0.92) | <0.01 | 0.90 (0.81 - 0.99) | 0.05 |
| OEHR | 15,387 | 32,362 | 4.2 | 13.5 | 72 | 225 | 1.11 (0.83 - 1.48) | 0.46 | 1.16 (0.87 - 1.56) | 0.33 |
| SIDIAP | 2,113 | 10,863 | 2.5 | 18.4 | 45 | 505 | 0.68 (0.48 - 0.93) | 0.02 | 0.71 (0.51 - 0.99) | 0.06 |
| Meta-analysis | 303,119 | 505,435 | 184.3 | 350.9 | 1,657 | 4,159 | 0.86 (0.81 - 0.92) | <0.01 | 0.88 (0.79 - 0.98) | 0.02 |
| Meta-analysis (w/o USOC) | 49,191 | 93,500 | 25.7 | 65.3 | 424 | 1,378 | 0.88 (0.75 - 1.02) | 0.09 | 0.88 (0.75 - 1.03) | 0.12 |

##### 1.2.21 Lower extremity amputation

**Supplementary Table 47:** Exposures, outcomes and risk estimates for Lower extremity amputation in SGLT2I (female) (T) vs DPP4I (female) (C) new-user cohorts under four different propensity score (PS) adjustment / time-at-risk (TAR) designs across data sources. We report total patient exposure time in 1,000 patient-year (KYr) units, uncalibrated and calibrated hazard ratios (HRs) and their 95% confidence intervals (CIs). Greyed out entries did not pass study diagnostics and are included only for completeness.

| | Patients | | Exposure (KYr) | | Outcomes | | Uncalibrated | | Calibrated | |
|---|---|---|---|---|---|---|---|---|---|---|
| | T | C | T | C | T | C | HR (95% CI) | $p$ | HR (95% CI) | $p$ |
| **PS-matching / on-treatment TAR** | | | | | | | | | | |
| CCAE | 16,405 | 25,915 | 12.3 | 19.5 | 5 | <5 | 2.00 (0.43 - 10.3) | 0.39 | 2.04 (0.42 - 9.97) | 0.38 |
| DAG | 2,939 | 8,792 | 2.4 | 9.3 | 0 | 0 | | | | |
| ODOD | 10,719 | 20,618 | 7.0 | 15.8 | 5 | 6 | 2.44 (0.41 - 15.3) | 0.33 | 2.53 (0.41 - 15.6) | 0.32 |
| MDCR | 1,302 | 4,829 | 0.9 | 4.1 | <5 | <5 | | 0.29 | | 0.29 |
| MDCD | 2,339 | 6,352 | 1.3 | 4.4 | 0 | <5 | | | | |
| Open Claims | 231,643 | 445,651 | 182.7 | 390.7 | 35 | 108 | 1.03 (0.55 - 1.87) | 0.93 | 1.07 (0.58 - 2.00) | 0.83 |
| OEHR | 15,057 | 33,693 | 4.8 | 17.8 | <5 | 7 | 1.08 (0.05 - 8.32) | 0.95 | 1.09 (0.09 - 13.7) | 0.95 |
| SIDIAP | 2,778 | 14,267 | 3.9 | 30.9 | 0 | <5 | | | | |
| Meta-analysis | 231,643 | 445,651 | 182.7 | 390.7 | 35 | 108 | 1.03 (0.55 - 1.87) | 0.93 | 1.07 (0.58 - 1.98) | 0.84 |
| **PS-stratification / on-treatment TAR** | | | | | | | | | | |
| CCAE | 18,874 | 25,951 | 13.7 | 19.5 | 5 | <5 | 2.04 (0.51 - 8.68) | 0.33 | 2.04 (0.50 - 8.43) | 0.32 |
| DAG | 3,243 | 8,833 | 2.5 | 9.3 | 0 | 0 | | | | |
| ODOD | 12,831 | 20,824 | 7.9 | 15.9 | 5 | 6 | 2.72 (0.67 - 10.8) | 0.16 | 2.78 (0.69 - 11.2) | 0.15 |
| MDCR | 1,475 | 4,896 | 0.9 | 4.2 | <5 | <5 | 3.13 (0.13 - 43.3) | 0.44 | 3.17 (0.17 - 57.4) | 0.44 |
| MDCD | 2,431 | 6,371 | 1.3 | 4.5 | 0 | 5 | | | | |
| Open Claims | 276,304 | 446,087 | 204.7 | 391.1 | 39 | 108 | 1.05 (0.70 - 1.56) | 0.79 | 1.11 (0.73 - 1.67) | 0.63 |
| OEHR | 16,654 | 34,735 | 5.2 | 18.4 | <5 | 7 | 2.01 (0.28 - 9.25) | 0.43 | 2.13 (0.37 - 12.2) | 0.40 |
| SIDIAP | 2,956 | 14,749 | 4.0 | 32.4 | 0 | <5 | | | | |
| Meta-analysis | 276,304 | 446,087 | 204.7 | 391.1 | 39 | 108 | 1.05 (0.70 - 1.56) | 0.79 | 1.08 (0.72 - 1.64) | 0.70 |
| **PS-matching / on-treatment TAR without escalation** | | | | | | | | | | |
| CCAE | 16,405 | 25,915 | 10.6 | 15.9 | <5 | <5 | 1.41 (0.26 - 7.80) | 0.69 | 1.39 (0.25 - 7.68) | 0.70 |
| DAG | 2,939 | 8,792 | 2.2 | 8.3 | 0 | 0 | | | | |
| ODOD | 10,719 | 20,618 | 6.2 | 13.2 | <5 | 6 | 2.15 (0.35 - 13.8) | 0.41 | 2.22 (0.35 - 13.8) | 0.39 |
| MDCR | 1,302 | 4,829 | 0.8 | 3.6 | <5 | <5 | | 0.29 | | 0.30 |
| MDCD | 2,339 | 6,352 | 1.1 | 3.5 | 0 | <5 | | | | |
| Open Claims | 231,643 | 445,651 | 152.5 | 310.5 | 25 | 86 | 1.00 (0.47 - 1.99) | 0.99 | 1.04 (0.50 - 2.13) | 0.92 |
| OEHR | 15,057 | 33,693 | 4.1 | 14.3 | <5 | <5 | | | | |
| SIDIAP | 2,778 | 14,267 | 3.4 | 24.4 | 0 | <5 | | | | |
| Meta-analysis | 231,643 | 445,651 | 152.5 | 310.5 | 25 | 86 | 1.00 (0.47 - 1.99) | 0.99 | 1.03 (0.50 - 2.12) | 0.94 |
| **PS-stratification / on-treatment TAR without escalation** | | | | | | | | | | |
| CCAE | 18,874 | 25,951 | 11.8 | 15.9 | <5 | <5 | 1.97 (0.42 - 10.3) | 0.40 | 1.92 (0.39 - 9.44) | 0.42 |
| DAG | 3,243 | 8,833 | 2.3 | 8.4 | 0 | 0 | | | | |
| ODOD | 12,831 | 20,824 | 7.0 | 13.4 | <5 | 6 | 1.83 (0.33 - 8.43) | 0.47 | 1.84 (0.36 - 9.28) | 0.46 |
| MDCR | 1,475 | 4,896 | 0.8 | 3.6 | <5 | <5 | 2.99 (0.13 - 41.5) | 0.46 | 2.94 (0.16 - 53.3) | 0.47 |
| MDCD | 2,431 | 6,371 | 1.1 | 3.5 | 0 | <5 | | | | |
| Open Claims | 276,304 | 446,087 | 172.1 | 310.9 | 27 | 86 | 0.87 (0.54 - 1.39) | 0.58 | 0.91 (0.57 - 1.48) | 0.71 |
| OEHR | 16,654 | 34,735 | 4.6 | 14.8 | <5 | <5 | 1.58 (0.08 - 12.3) | 0.72 | 1.65 (0.13 - 20.9) | 0.70 |
| SIDIAP | 2,956 | 14,749 | 3.5 | 25.6 | 0 | <5 | | | | |
| Meta-analysis | 276,304 | 446,087 | 172.1 | 310.9 | 27 | 86 | 0.87 (0.54 - 1.39) | 0.58 | 0.89 (0.55 - 1.45) | 0.65 |

##### 1.2.22 Nausea

**Supplementary Table 48:** Exposures, outcomes and risk estimates for Nausea in SGLT2I (female) (T) vs DPP4I (female) (C) new-user cohorts under four different propensity score (PS) adjustment / time-at-risk (TAR) designs across data sources. We report total patient exposure time in 1,000 patient-year (KYr) units, uncalibrated and calibrated hazard ratios (HRs) and their 95% confidence intervals (CIs). Greyed out entries did not pass study diagnostics and are included only for completeness.

| | Patients | | Exposure (KYr) | | Outcomes | | Uncalibrated | | Calibrated | |
|---|---|---|---|---|---|---|---|---|---|---|
| | T | C | T | C | T | C | HR (95% CI) | $p$ | HR (95% CI) | $p$ |
| **PS-matching / on-treatment TAR** | | | | | | | | | | |
| CCAE | 13,325 | 21,231 | 9.9 | 15.6 | 461 | 771 | 0.91 (0.77 - 1.08) | 0.29 | 0.93 (0.79 - 1.10) | 0.49 |
| DAG | 2,604 | 7,941 | 2.1 | 8.3 | 27 | 114 | 0.94 (0.53 - 1.59) | 0.82 | 0.82 (0.48 - 1.42) | 0.52 |
| ODOD | 8,575 | 16,310 | 5.5 | 12.1 | 276 | 718 | 0.87 (0.72 - 1.05) | 0.15 | 0.90 (0.74 - 1.10) | 0.32 |
| MDCR | 1,099 | 4,068 | 0.7 | 3.4 | 30 | 178 | 0.76 (0.46 - 1.23) | 0.28 | 0.75 (0.46 - 1.23) | 0.28 |
| MDCD | 1,504 | 4,121 | 0.8 | 2.7 | 79 | 324 | 0.82 (0.58 - 1.13) | 0.23 | 0.80 (0.57 - 1.11) | 0.21 |
| Open Claims | 186,937 | 363,943 | 148.2 | 316.2 | 3,541 | 9,321 | 0.88 (0.84 - 0.93) | <0.01 | 0.92 (0.82 - 1.03) | 0.20 |
| OEHR | 12,664 | 28,430 | 3.9 | 14.5 | 211 | 742 | 1.18 (0.96 - 1.46) | 0.12 | 1.20 (0.97 - 1.49) | 0.13 |
| SIDIAP | 2,595 | 13,378 | 3.6 | 28.9 | 18 | 287 | 0.31 (0.15 - 0.60) | <0.01 | 0.33 (0.16 - 0.67) | <0.01 |
| Meta-analysis | 229,573 | 460,318 | 175.3 | 404.2 | 4,623 | 12,303 | 0.91 (0.81 - 1.03) | 0.14 | 0.95 (0.83 - 1.09) | 0.47 |
| Meta-analysis (w/o USOC) | 42,636 | 96,375 | 27.1 | 88.1 | 1,082 | 2,982 | 0.91 (0.73 - 1.13) | 0.40 | 0.93 (0.75 - 1.16) | 0.55 |
| **PS-stratification / on-treatment TAR** | | | | | | | | | | |
| CCAE | 15,258 | 21,256 | 10.9 | 15.7 | 515 | 774 | 0.93 (0.83 - 1.05) | 0.24 | 0.94 (0.83 - 1.06) | 0.34 |
| DAG | 2,873 | 7,967 | 2.2 | 8.3 | 27 | 114 | 1.06 (0.66 - 1.65) | 0.80 | 0.93 (0.59 - 1.47) | 0.74 |
| ODOD | 10,200 | 16,497 | 6.2 | 12.2 | 321 | 730 | 0.88 (0.76 - 1.01) | 0.07 | 0.90 (0.78 - 1.04) | 0.17 |
| MDCR | 1,249 | 4,115 | 0.8 | 3.4 | 32 | 179 | 0.72 (0.47 - 1.07) | 0.11 | 0.73 (0.48 - 1.10) | 0.14 |
| MDCD | 1,543 | 4,133 | 0.8 | 2.7 | 80 | 325 | 0.78 (0.60 - 1.00) | 0.05 | 0.74 (0.57 - 0.96) | 0.03 |
| Open Claims | 221,452 | 364,477 | 165.5 | 316.7 | 3,960 | 9,346 | 0.88 (0.84 - 0.91) | <0.01 | 0.92 (0.82 - 1.03) | 0.15 |
| OEHR | 14,004 | 29,486 | 4.3 | 15.0 | 230 | 773 | 1.12 (0.95 - 1.31) | 0.16 | 1.19 (1.01 - 1.40) | 0.06 |
| SIDIAP | 2,767 | 13,824 | 3.7 | 30.3 | 21 | 300 | 0.60 (0.36 - 0.94) | 0.03 | 0.60 (0.37 - 0.97) | 0.04 |
| Meta-analysis | 266,554 | 453,507 | 192.8 | 398.2 | 5,074 | 12,037 | 0.92 (0.84 - 1.02) | 0.10 | 0.95 (0.83 - 1.08) | 0.44 |
| Meta-analysis (w/o USOC) | 45,102 | 89,030 | 27.4 | 81.5 | 1,114 | 2,691 | 0.94 (0.82 - 1.08) | 0.39 | 0.96 (0.84 - 1.11) | 0.62 |
| **PS-matching / on-treatment TAR without escalation** | | | | | | | | | | |
| CCAE | 13,325 | 21,231 | 8.6 | 12.9 | 400 | 637 | 0.87 (0.72 - 1.03) | 0.12 | 0.85 (0.71 - 1.03) | 0.15 |
| DAG | 2,604 | 7,941 | 1.9 | 7.4 | 23 | 100 | 0.89 (0.48 - 1.58) | 0.70 | 0.83 (0.45 - 1.51) | 0.56 |
| ODOD | 8,575 | 16,310 | 4.9 | 10.3 | 244 | 607 | 0.83 (0.67 - 1.02) | 0.07 | 0.85 (0.69 - 1.05) | 0.16 |
| MDCR | 1,099 | 4,068 | 0.7 | 2.9 | 27 | 149 | 0.73 (0.42 - 1.23) | 0.26 | 0.69 (0.40 - 1.18) | 0.18 |
| MDCD | 1,504 | 4,121 | 0.7 | 2.2 | 64 | 251 | 0.90 (0.62 - 1.29) | 0.56 | 0.87 (0.60 - 1.26) | 0.49 |
| Open Claims | 186,937 | 363,943 | 124.9 | 253.3 | 2,881 | 7,391 | 0.87 (0.82 - 0.92) | <0.01 | 0.90 (0.84 - 0.97) | 0.07 |
| OEHR | 12,664 | 28,430 | 3.4 | 11.7 | 174 | 565 | 1.16 (0.91 - 1.47) | 0.22 | 1.19 (0.93 - 1.51) | 0.20 |
| SIDIAP | 2,595 | 13,378 | 3.2 | 22.9 | 14 | 220 | 0.33 (0.15 - 0.67) | <0.01 | 0.38 (0.18 - 0.82) | 0.02 |
| Meta-analysis | 229,573 | 460,318 | 148.7 | 325.6 | 3,809 | 9,796 | 0.88 (0.83 - 0.93) | <0.01 | 0.91 (0.83 - 0.99) | 0.02 |
| Meta-analysis (w/o USOC) | 42,636 | 96,375 | 23.8 | 72.3 | 928 | 2,405 | 0.90 (0.73 - 1.11) | 0.32 | 0.91 (0.74 - 1.13) | 0.41 |
| **PS-stratification / on-treatment TAR without escalation** | | | | | | | | | | |
| CCAE | 15,258 | 21,256 | 9.5 | 12.9 | 450 | 640 | 0.94 (0.83 - 1.07) | 0.36 | 0.92 (0.81 - 1.04) | 0.23 |
| DAG | 2,873 | 7,967 | 2.0 | 7.5 | 23 | 100 | 1.12 (0.67 - 1.80) | 0.66 | 0.97 (0.59 - 1.59) | 0.80 |
| ODOD | 10,200 | 16,497 | 5.5 | 10.4 | 277 | 618 | 0.87 (0.74 - 1.01) | 0.08 | 0.87 (0.75 - 1.02) | 0.10 |
| MDCR | 1,249 | 4,115 | 0.7 | 3.0 | 29 | 150 | 0.80 (0.51 - 1.21) | 0.30 | 0.78 (0.51 - 1.21) | 0.28 |
| MDCD | 1,543 | 4,133 | 0.7 | 2.2 | 65 | 252 | 0.76 (0.57 - 1.00) | 0.05 | 0.72 (0.54 - 0.95) | 0.03 |
| Open Claims | 221,452 | 364,477 | 140.3 | 253.7 | 3,228 | 7,416 | 0.86 (0.83 - 0.90) | <0.01 | 0.90 (0.83 - 0.98) | 0.04 |
| OEHR | 14,004 | 29,486 | 3.8 | 12.2 | 190 | 589 | 1.14 (0.95 - 1.36) | 0.15 | 1.19 (1.00 - 1.43) | 0.09 |
| SIDIAP | 2,767 | 13,824 | 3.3 | 24.1 | 16 | 229 | 0.53 (0.30 - 0.87) | 0.02 | 0.55 (0.32 - 0.95) | 0.04 |
| Meta-analysis | 266,554 | 453,507 | 164.4 | 320.7 | 4,184 | 9,592 | 0.92 (0.82 - 1.03) | 0.15 | 0.94 (0.82 - 1.09) | 0.41 |
| Meta-analysis (w/o USOC) | 45,102 | 89,030 | 24.1 | 67.0 | 956 | 2,176 | 0.94 (0.80 - 1.10) | 0.46 | 0.94 (0.80 - 1.11) | 0.51 |

##### 1.2.23 Peripheral edema

**Supplementary Table 49:** Exposures, outcomes and risk estimates for Peripheral edema in SGLT2I (female) (T) vs DPP4I (female) (C) new-user cohorts under four different propensity score (PS) adjustment / time-at-risk (TAR) designs across data sources. We report total patient exposure time in 1,000 patient-year (KYr) units, uncalibrated and calibrated hazard ratios (HRs) and their 95% confidence intervals (CIs). Greyed out entries did not pass study diagnostics and are included only for completeness.

| | Patients | | Exposure (KYr) | | Outcomes | | Uncalibrated | | Calibrated | |
|---|---|---|---|---|---|---|---|---|---|---|
| | T | C | T | C | T | C | HR (95% CI) | $p$ | HR (95% CI) | $p$ |
| **PS-matching / on-treatment TAR** | | | | | | | | | | |
| CCAE | 12,430 | 19,749 | 9.1 | 14.4 | 392 | 705 | 0.80 (0.67 - 0.96) | 0.02 | 0.82 (0.68 - 0.98) | 0.05 |
| DAG | 2,349 | 7,030 | 1.9 | 7.2 | 31 | 264 | 0.56 (0.35 - 0.86) | 0.01 | 0.49 (0.31 - 0.77) | <0.01 |
| ODOD | 7,836 | 14,517 | 4.9 | 10.3 | 290 | 992 | 0.74 (0.62 - 0.88) | <0.01 | 0.77 (0.64 - 0.92) | <0.01 |
| MDCR | 947 | 3,271 | 0.6 | 2.6 | 28 | 253 | 0.58 (0.33 - 0.96) | 0.05 | 0.57 (0.34 - 0.97) | 0.05 |
| MDCD | 1,489 | 4,025 | 0.8 | 2.5 | 68 | 292 | 0.80 (0.56 - 1.13) | 0.21 | 0.78 (0.55 - 1.11) | 0.20 |
| Open Claims | 170,651 | 326,752 | 134.0 | 276.5 | 3,312 | 11,135 | 0.77 (0.73 - 0.81) | <0.01 | 0.80 (0.71 - 0.90) | <0.01 |
| OEHR | 12,122 | 27,295 | 3.8 | 13.6 | 150 | 721 | 1.03 (0.81 - 1.30) | 0.83 | 1.04 (0.82 - 1.32) | 0.74 |
| SIDIAP | 2,316 | 12,341 | 3.3 | 26.1 | 45 | 587 | 0.75 (0.51 - 1.08) | 0.14 | 0.80 (0.55 - 1.16) | 0.27 |
| Meta-analysis | 210,322 | 415,605 | 158.9 | 355.3 | 4,299 | 14,785 | 0.77 (0.74 - 0.81) | <0.01 | 0.80 (0.74 - 0.88) | <0.01 |
| Meta-analysis (w/o USOC) | 39,671 | 88,853 | 24.9 | 78.8 | 987 | 3,650 | 0.80 (0.71 - 0.90) | <0.01 | 0.81 (0.72 - 0.92) | <0.01 |
| **PS-stratification / on-treatment TAR** | | | | | | | | | | |
| CCAE | 14,270 | 19,774 | 10.1 | 14.4 | 434 | 706 | 0.90 (0.80 - 1.02) | 0.12 | 0.91 (0.80 - 1.03) | 0.19 |
| DAG | 2,574 | 7,105 | 2.0 | 7.3 | 33 | 272 | 0.52 (0.35 - 0.75) | <0.01 | 0.46 (0.31 - 0.67) | <0.01 |
| ODOD | 9,264 | 14,641 | 5.5 | 10.4 | 325 | 1,009 | 0.75 (0.66 - 0.86) | <0.01 | 0.77 (0.67 - 0.89) | <0.01 |
| MDCR | 1,079 | 3,319 | 0.7 | 2.6 | 30 | 262 | 0.57 (0.38 - 0.84) | <0.01 | 0.58 (0.39 - 0.87) | 0.01 |
| MDCD | 1,536 | 4,052 | 0.8 | 2.6 | 69 | 299 | 0.82 (0.62 - 1.07) | 0.15 | 0.78 (0.59 - 1.03) | 0.10 |
| Open Claims | 201,879 | 327,260 | 149.5 | 276.9 | 3,614 | 11,193 | 0.77 (0.74 - 0.80) | <0.01 | 0.80 (0.72 - 0.90) | <0.01 |
| OEHR | 13,269 | 28,054 | 4.1 | 14.0 | 157 | 756 | 0.89 (0.74 - 1.07) | 0.22 | 0.94 (0.78 - 1.14) | 0.58 |
| SIDIAP | 2,465 | 12,766 | 3.4 | 27.4 | 48 | 623 | 0.72 (0.52 - 0.97) | 0.04 | 0.72 (0.52 - 0.98) | 0.06 |
| Meta-analysis | 243,721 | 409,600 | 174.6 | 350.4 | 4,611 | 14,559 | 0.79 (0.72 - 0.87) | <0.01 | 0.81 (0.71 - 0.93) | <0.01 |
| Meta-analysis (w/o USOC) | 41,842 | 82,340 | 25.1 | 73.4 | 997 | 3,366 | 0.79 (0.68 - 0.91) | <0.01 | 0.81 (0.70 - 0.93) | <0.01 |
| **PS-matching / on-treatment TAR without escalation** | | | | | | | | | | |
| CCAE | 12,430 | 19,749 | 7.9 | 11.8 | 334 | 587 | 0.78 (0.64 - 0.95) | 0.01 | 0.77 (0.63 - 0.94) | 0.02 |
| DAG | 2,349 | 7,030 | 1.7 | 6.5 | 29 | 241 | 0.57 (0.35 - 0.89) | 0.02 | 0.53 (0.33 - 0.85) | 0.01 |
| ODOD | 7,836 | 14,517 | 4.4 | 8.7 | 254 | 823 | 0.70 (0.58 - 0.86) | <0.01 | 0.73 (0.60 - 0.89) | <0.01 |
| MDCR | 947 | 3,271 | 0.6 | 2.3 | 26 | 223 | 0.62 (0.35 - 1.05) | 0.09 | 0.58 (0.34 - 1.01) | 0.06 |
| MDCD | 1,489 | 4,025 | 0.6 | 2.0 | 60 | 221 | 0.88 (0.59 - 1.29) | 0.52 | 0.85 (0.58 - 1.26) | 0.46 |
| Open Claims | 170,651 | 326,752 | 112.6 | 221.6 | 2,729 | 8,869 | 0.77 (0.73 - 0.82) | <0.01 | 0.80 (0.75 - 0.87) | <0.01 |
| OEHR | 12,122 | 27,295 | 3.3 | 11.1 | 126 | 555 | 1.01 (0.77 - 1.31) | 0.95 | 1.03 (0.79 - 1.34) | 0.78 |
| SIDIAP | 2,316 | 12,341 | 2.9 | 20.8 | 39 | 463 | 0.81 (0.53 - 1.21) | 0.33 | 0.93 (0.61 - 1.41) | 0.73 |
| Meta-analysis | 210,322 | 415,605 | 134.4 | 286.2 | 3,576 | 11,832 | 0.78 (0.74 - 0.82) | <0.01 | 0.80 (0.74 - 0.87) | <0.01 |
| Meta-analysis (w/o USOC) | 39,671 | 88,853 | 21.8 | 64.7 | 847 | 2,963 | 0.78 (0.68 - 0.90) | <0.01 | 0.79 (0.69 - 0.91) | <0.01 |
| **PS-stratification / on-treatment TAR without escalation** | | | | | | | | | | |
| CCAE | 14,270 | 19,774 | 8.7 | 11.8 | 372 | 588 | 0.90 (0.78 - 1.03) | 0.12 | 0.87 (0.76 - 1.00) | 0.08 |
| DAG | 2,574 | 7,105 | 1.8 | 6.6 | 31 | 248 | 0.53 (0.35 - 0.77) | <0.01 | 0.46 (0.31 - 0.68) | <0.01 |
| ODOD | 9,264 | 14,641 | 4.9 | 8.8 | 288 | 839 | 0.77 (0.67 - 0.89) | <0.01 | 0.77 (0.67 - 0.90) | <0.01 |
| MDCR | 1,079 | 3,319 | 0.6 | 2.3 | 28 | 229 | 0.60 (0.39 - 0.90) | 0.02 | 0.59 (0.39 - 0.90) | 0.02 |
| MDCD | 1,536 | 4,052 | 0.6 | 2.0 | 60 | 228 | 0.90 (0.66 - 1.21) | 0.50 | 0.85 (0.63 - 1.15) | 0.32 |
| Open Claims | 201,879 | 327,260 | 126.4 | 221.9 | 2,985 | 8,914 | 0.76 (0.73 - 0.80) | <0.01 | 0.80 (0.73 - 0.87) | <0.01 |
| OEHR | 13,269 | 28,054 | 3.6 | 11.4 | 131 | 580 | 0.90 (0.73 - 1.11) | 0.34 | 0.95 (0.77 - 1.16) | 0.63 |
| SIDIAP | 2,465 | 12,766 | 3.0 | 21.8 | 41 | 493 | 0.73 (0.51 - 1.01) | 0.06 | 0.76 (0.54 - 1.07) | 0.15 |
| Meta-analysis | 243,721 | 409,600 | 148.5 | 282.3 | 3,848 | 11,662 | 0.79 (0.73 - 0.87) | <0.01 | 0.81 (0.72 - 0.92) | <0.01 |
| Meta-analysis (w/o USOC) | 41,842 | 82,340 | 22.0 | 60.4 | 863 | 2,748 | 0.80 (0.70 - 0.92) | <0.01 | 0.80 (0.70 - 0.92) | <0.01 |

##### 1.2.24 Photosensitivity

**Supplementary Table 50:** Exposures, outcomes and risk estimates for Photosensitivity in SGLT2I (female) (T) vs DPP4I (female) (C) new-user cohorts under four different propensity score (PS) adjustment / time-at-risk (TAR) designs across data sources. We report total patient exposure time in 1,000 patient-year (KYr) units, uncalibrated and calibrated hazard ratios (HRs) and their 95% confidence intervals (CIs). Greyed out entries did not pass study diagnostics and are included only for completeness.

| | Patients | | Exposure (KYr) | | Outcomes | | Uncalibrated | | Calibrated | |
|---|---|---|---|---|---|---|---|---|---|---|
| | T | C | T | C | T | C | HR (95% CI) | $p$ | HR (95% CI) | $p$ |
| **PS-matching / on-treatment TAR** | | | | | | | | | | |
| CCAE | 16,226 | 25,629 | 12.2 | 19.2 | 9 | 14 | 1.14 (0.29 - 3.94) | 0.84 | 1.17 (0.32 - 4.30) | 0.82 |
| DAG | 2,924 | 8,758 | 2.4 | 9.2 | <5 | <5 | | | | |
| ODOD | 10,639 | 20,438 | 6.9 | 15.6 | <5 | 17 | 0.44 (0.07 - 1.87) | 0.34 | 0.46 (0.09 - 2.47) | 0.37 |
| MDCR | 1,291 | 4,759 | 0.9 | 4.0 | <5 | 7 | 0.74 (0.03 - 6.84) | 0.82 | 0.72 (0.05 - 10.2) | 0.81 |
| MDCD | 2,325 | 6,314 | 1.3 | 4.4 | <5 | 6 | | | | |
| Open Claims | 229,224 | 441,027 | 180.6 | 386.2 | 67 | 195 | 1.16 (0.79 - 1.69) | 0.44 | 1.21 (0.82 - 1.79) | 0.34 |
| OEHR | 14,958 | 33,449 | 4.7 | 17.7 | <5 | 30 | 0.27 (0.01 - 1.57) | 0.28 | 0.28 (0.03 - 2.89) | 0.28 |
| SIDIAP | 2,767 | 14,239 | 3.9 | 30.8 | 0 | 7 | | | | |
| Meta-analysis | 271,047 | 520,543 | 204.4 | 438.7 | <86 | 256 | 1.08 (0.76 - 1.53) | 0.68 | 1.12 (0.78 - 1.60) | 0.54 |
| Meta-analysis (w/o USOC) | 41,823 | 79,516 | 23.8 | 52.5 | <19 | 61 | 0.67 (0.26 - 1.73) | 0.41 | 0.69 (0.27 - 1.77) | 0.44 |
| **PS-stratification / on-treatment TAR** | | | | | | | | | | |
| CCAE | 18,681 | 25,665 | 13.5 | 19.3 | 11 | 14 | 1.53 (0.63 - 3.61) | 0.34 | 1.53 (0.64 - 3.67) | 0.34 |
| DAG | 3,228 | 8,796 | 2.5 | 9.3 | <5 | <5 | | 0.42 | | 0.46 |
| ODOD | 12,737 | 20,640 | 7.8 | 15.8 | <5 | 17 | 0.37 (0.08 - 1.25) | 0.16 | 0.38 (0.10 - 1.50) | 0.17 |
| MDCR | 1,462 | 4,825 | 0.9 | 4.1 | <5 | 7 | 1.01 (0.05 - 7.18) | 1.00 | 1.02 (0.08 - 12.3) | 0.97 |
| MDCD | 2,416 | 6,333 | 1.3 | 4.4 | <5 | 6 | 0.27 (0.01 - 1.68) | 0.28 | 0.26 (0.02 - 2.84) | 0.27 |
| Open Claims | 273,499 | 441,578 | 202.5 | 386.7 | 72 | 195 | 1.06 (0.79 - 1.42) | 0.68 | 1.12 (0.82 - 1.52) | 0.49 |
| OEHR | 16,535 | 34,484 | 5.2 | 18.2 | <5 | 30 | 0.65 (0.15 - 1.95) | 0.51 | 0.69 (0.19 - 2.48) | 0.57 |
| SIDIAP | 2,944 | 14,723 | 4.0 | 32.3 | 0 | 8 | | | | |
| Meta-analysis | 321,452 | 522,367 | 229.0 | 439.9 | <93 | 256 | 1.03 (0.79 - 1.35) | 0.80 | 1.06 (0.80 - 1.41) | 0.66 |
| Meta-analysis (w/o USOC) | 47,953 | 80,789 | 26.5 | 53.2 | <21 | 61 | 0.82 (0.35 - 1.92) | 0.65 | 0.84 (0.36 - 1.96) | 0.69 |
| **PS-matching / on-treatment TAR without escalation** | | | | | | | | | | |
| CCAE | 16,226 | 25,629 | 10.4 | 15.7 | 9 | 13 | 1.94 (0.47 - 7.53) | 0.35 | 1.91 (0.47 - 7.67) | 0.36 |
| DAG | 2,924 | 8,758 | 2.2 | 8.3 | <5 | <5 | | | | |
| ODOD | 10,639 | 20,438 | 6.1 | 13.1 | <5 | 16 | 0.40 (0.06 - 1.70) | 0.29 | 0.42 (0.08 - 2.23) | 0.31 |
| MDCR | 1,291 | 4,759 | 0.8 | 3.5 | <5 | 6 | 0.73 (0.03 - 6.83) | 0.82 | 0.69 (0.05 - 9.72) | 0.78 |
| MDCD | 2,325 | 6,314 | 1.0 | 3.4 | <5 | <5 | | | | |
| Open Claims | 229,224 | 441,027 | 150.8 | 306.9 | 55 | 168 | 1.09 (0.72 - 1.63) | 0.69 | 1.13 (0.75 - 1.71) | 0.57 |
| OEHR | 14,958 | 33,449 | 4.1 | 14.2 | <5 | 26 | 0.26 (0.01 - 1.50) | 0.26 | 0.27 (0.03 - 2.79) | 0.27 |
| SIDIAP | 2,767 | 14,239 | 3.4 | 24.4 | 0 | 6 | | | | |
| Meta-analysis | 271,047 | 520,543 | 171.4 | 349.8 | <74 | 223 | 1.04 (0.71 - 1.52) | 0.84 | 1.07 (0.73 - 1.58) | 0.72 |
| Meta-analysis (w/o USOC) | 41,823 | 79,516 | 20.7 | 43.0 | <19 | 55 | 0.73 (0.21 - 2.52) | 0.61 | 0.74 (0.21 - 2.56) | 0.63 |
| **PS-stratification / on-treatment TAR without escalation** | | | | | | | | | | |
| CCAE | 18,681 | 25,665 | 11.7 | 15.7 | 10 | 13 | 1.46 (0.58 - 3.56) | 0.41 | 1.42 (0.57 - 3.51) | 0.45 |
| DAG | 3,228 | 8,796 | 2.3 | 8.3 | <5 | <5 | | 0.41 | | 0.46 |
| ODOD | 12,737 | 20,640 | 6.9 | 13.2 | <5 | 16 | 0.38 (0.08 - 1.30) | 0.17 | 0.38 (0.10 - 1.52) | 0.17 |
| MDCR | 1,462 | 4,825 | 0.8 | 3.6 | <5 | 6 | 1.05 (0.05 - 7.66) | 0.97 | 1.03 (0.08 - 12.6) | 0.97 |
| MDCD | 2,416 | 6,333 | 1.1 | 3.4 | <5 | <5 | 0.34 (0.02 - 2.41) | 0.39 | 0.32 (0.03 - 3.83) | 0.37 |
| Open Claims | 273,499 | 441,578 | 170.2 | 307.3 | 59 | 168 | 1.02 (0.74 - 1.40) | 0.89 | 1.07 (0.77 - 1.49) | 0.69 |
| OEHR | 16,535 | 34,484 | 4.5 | 14.6 | <5 | 26 | 0.68 (0.15 - 2.09) | 0.56 | 0.71 (0.19 - 2.61) | 0.61 |
| SIDIAP | 2,944 | 14,723 | 3.5 | 25.6 | 0 | 7 | | | | |
| Meta-analysis | 321,452 | 522,367 | 193.3 | 350.9 | <79 | 223 | 1.00 (0.75 - 1.33) | 0.98 | 1.02 (0.75 - 1.38) | 0.90 |
| Meta-analysis (w/o USOC) | 47,953 | 80,789 | 23.1 | 43.6 | <20 | 55 | 0.83 (0.38 - 1.84) | 0.65 | 0.84 (0.38 - 1.84) | 0.66 |

##### 1.2.25 Venous thromboembolism

**Supplementary Table 51:** Exposures, outcomes and risk estimates for Venous thromboembolism in SGLT2I (female) (T) vs DPP4I (female) (C) new-user cohorts under four different propensity score (PS) adjustment / time-at-risk (TAR) designs across data sources. We report total patient exposure time in 1,000 patient-year (KYr) units, uncalibrated and calibrated hazard ratios (HRs) and their 95% confidence intervals (CIs). Greyed out entries did not pass study diagnostics and are included only for completeness.

| | Patients | | Exposure (KYr) | | Outcomes | | Uncalibrated | | Calibrated | |
|---|---|---|---|---|---|---|---|---|---|---|
| | T | C | T | C | T | C | HR (95% CI) | $p$ | HR (95% CI) | $p$ |
| **PS-matching / on-treatment TAR** | | | | | | | | | | |
| CCAE | 16,050 | 25,231 | 12.1 | 18.9 | 55 | 101 | 1.08 (0.67 - 1.72) | 0.76 | 1.10 (0.68 - 1.76) | 0.71 |
| DAG | 2,690 | 8,055 | 2.1 | 8.4 | 13 | 94 | 0.77 (0.35 - 1.55) | 0.48 | 0.67 (0.32 - 1.42) | 0.32 |
| ODOD | 10,381 | 19,876 | 6.7 | 15.1 | 53 | 165 | 0.92 (0.59 - 1.42) | 0.71 | 0.95 (0.61 - 1.49) | 0.84 |
| MDCR | 1,249 | 4,593 | 0.9 | 3.9 | 5 | 49 | 0.99 (0.30 - 2.77) | 0.99 | 0.97 (0.32 - 2.94) | 0.92 |
| MDCD | 2,233 | 6,071 | 1.2 | 4.2 | 10 | 42 | 1.59 (0.64 - 3.72) | 0.30 | 1.55 (0.65 - 3.74) | 0.33 |
| Open Claims | 224,268 | 429,979 | 176.8 | 375.9 | 655 | 1,999 | 0.94 (0.83 - 1.07) | 0.35 | 0.98 (0.84 - 1.15) | 0.84 |
| OEHR | 14,678 | 32,822 | 4.6 | 17.2 | 36 | 118 | 2.13 (1.25 - 3.62) | <0.01 | 2.16 (1.27 - 3.68) | <0.01 |
| SIDIAP | 2,655 | 13,693 | 3.7 | 29.6 | 13 | 150 | 0.66 (0.30 - 1.33) | 0.28 | 0.70 (0.33 - 1.49) | 0.37 |
| Meta-analysis | 274,352 | 540,752 | 208.7 | 475.2 | <840 | 2,695 | 1.04 (0.82 - 1.33) | 0.74 | 1.08 (0.84 - 1.40) | 0.54 |
| Meta-analysis (w/o USOC) | 50,084 | 110,773 | 31.8 | 99.3 | <185 | 696 | 1.08 (0.78 - 1.50) | 0.62 | 1.11 (0.80 - 1.54) | 0.53 |
| **PS-stratification / on-treatment TAR** | | | | | | | | | | |
| CCAE | 18,464 | 25,266 | 13.4 | 18.9 | 59 | 101 | 1.01 (0.71 - 1.41) | 0.97 | 1.01 (0.72 - 1.42) | 0.93 |
| DAG | 2,972 | 8,077 | 2.3 | 8.4 | 15 | 94 | 0.82 (0.44 - 1.43) | 0.51 | 0.72 (0.40 - 1.29) | 0.28 |
| ODOD | 12,418 | 20,057 | 7.6 | 15.3 | 60 | 170 | 0.91 (0.66 - 1.25) | 0.58 | 0.93 (0.68 - 1.29) | 0.69 |
| MDCR | 1,415 | 4,651 | 0.9 | 3.9 | 5 | 49 | 0.47 (0.16 - 1.13) | 0.13 | 0.48 (0.18 - 1.27) | 0.14 |
| MDCD | 2,324 | 6,091 | 1.2 | 4.2 | 10 | 42 | 0.98 (0.45 - 1.96) | 0.97 | 0.94 (0.45 - 1.96) | 0.87 |
| Open Claims | 267,177 | 430,441 | 198.1 | 376.3 | 730 | 2,014 | 0.94 (0.86 - 1.04) | 0.23 | 0.99 (0.86 - 1.14) | 0.89 |
| OEHR | 16,265 | 33,807 | 5.1 | 17.7 | 38 | 124 | 1.40 (0.93 - 2.07) | 0.10 | 1.48 (0.99 - 2.21) | 0.06 |
| SIDIAP | 2,822 | 14,164 | 3.8 | 31.0 | 15 | 161 | 0.86 (0.47 - 1.47) | 0.60 | 0.86 (0.48 - 1.52) | 0.61 |
| Meta-analysis | 320,118 | 531,812 | 230.2 | 467.7 | 917 | 2,664 | 0.96 (0.88 - 1.04) | 0.30 | 0.98 (0.87 - 1.11) | 0.80 |
| Meta-analysis (w/o USOC) | 52,941 | 101,371 | 32.2 | 91.4 | 187 | 650 | 1.01 (0.84 - 1.21) | 0.94 | 1.03 (0.86 - 1.24) | 0.75 |
| **PS-matching / on-treatment TAR without escalation** | | | | | | | | | | |
| CCAE | 16,050 | 25,231 | 10.4 | 15.4 | 46 | 80 | 1.20 (0.71 - 2.01) | 0.50 | 1.18 (0.70 - 1.99) | 0.55 |
| DAG | 2,690 | 8,055 | 2.0 | 7.6 | 11 | 86 | 0.62 (0.26 - 1.34) | 0.26 | 0.58 (0.25 - 1.32) | 0.20 |
| ODOD | 10,381 | 19,876 | 6.0 | 12.7 | 46 | 147 | 0.73 (0.44 - 1.18) | 0.22 | 0.76 (0.46 - 1.24) | 0.28 |
| MDCR | 1,249 | 4,593 | 0.8 | 3.4 | <5 | 43 | 0.82 (0.22 - 2.51) | 0.75 | 0.76 (0.23 - 2.59) | 0.67 |
| MDCD | 2,233 | 6,071 | 1.0 | 3.3 | 7 | 30 | 1.48 (0.49 - 3.96) | 0.46 | 1.43 (0.50 - 4.05) | 0.50 |
| Open Claims | 224,268 | 429,979 | 147.8 | 298.9 | 528 | 1,611 | 0.90 (0.78 - 1.03) | 0.12 | 0.93 (0.81 - 1.08) | 0.40 |
| OEHR | 14,678 | 32,822 | 4.0 | 13.9 | 31 | 82 | 2.28 (1.24 - 4.18) | <0.01 | 2.33 (1.27 - 4.29) | <0.01 |
| SIDIAP | 2,655 | 13,693 | 3.2 | 23.4 | 11 | 116 | 0.76 (0.32 - 1.59) | 0.50 | 0.87 (0.39 - 1.94) | 0.74 |
| Meta-analysis | 274,352 | 540,752 | 175.5 | 380.0 | <685 | 2,172 | 1.01 (0.75 - 1.35) | 0.96 | 1.04 (0.77 - 1.40) | 0.81 |
| Meta-analysis (w/o USOC) | 50,084 | 110,773 | 27.8 | 81.0 | <157 | 561 | 1.05 (0.72 - 1.53) | 0.80 | 1.07 (0.73 - 1.55) | 0.75 |
| **PS-stratification / on-treatment TAR without escalation** | | | | | | | | | | |
| CCAE | 18,464 | 25,266 | 11.5 | 15.4 | 49 | 80 | 1.01 (0.68 - 1.46) | 0.98 | 0.98 (0.67 - 1.43) | 0.90 |
| DAG | 2,972 | 8,077 | 2.1 | 7.6 | 13 | 86 | 0.79 (0.40 - 1.44) | 0.47 | 0.69 (0.36 - 1.30) | 0.26 |
| ODOD | 12,418 | 20,057 | 6.7 | 12.8 | 53 | 152 | 0.86 (0.61 - 1.20) | 0.39 | 0.86 (0.61 - 1.22) | 0.41 |
| MDCR | 1,415 | 4,651 | 0.8 | 3.4 | <5 | 43 | 0.40 (0.12 - 1.06) | 0.11 | 0.40 (0.13 - 1.19) | 0.10 |
| MDCD | 2,324 | 6,091 | 1.0 | 3.3 | 7 | 30 | 1.05 (0.41 - 2.37) | 0.92 | 0.99 (0.41 - 2.39) | 0.94 |
| Open Claims | 267,177 | 430,441 | 166.7 | 299.3 | 597 | 1,625 | 0.92 (0.83 - 1.02) | 0.12 | 0.96 (0.85 - 1.09) | 0.59 |
| OEHR | 16,265 | 33,807 | 4.4 | 14.3 | 32 | 86 | 1.53 (0.97 - 2.38) | 0.06 | 1.60 (1.02 - 2.52) | 0.05 |
| SIDIAP | 2,822 | 14,164 | 3.4 | 24.6 | 13 | 124 | 0.91 (0.47 - 1.63) | 0.77 | 0.95 (0.51 - 1.78) | 0.86 |
| Meta-analysis | 320,118 | 531,812 | 194.8 | 374.0 | 757 | 2,153 | 0.94 (0.86 - 1.03) | 0.17 | 0.96 (0.85 - 1.09) | 0.53 |
| Meta-analysis (w/o USOC) | 52,941 | 101,371 | 28.1 | 74.7 | 160 | 528 | 1.00 (0.80 - 1.25) | 0.97 | 1.01 (0.81 - 1.26) | 0.92 |

##### 1.2.26 Vomiting

**Supplementary Table 52:** Exposures, outcomes and risk estimates for Vomiting in SGLT2I (female) (T) vs DPP4I (female) (C) new-user cohorts under four different propensity score (PS) adjustment / time-at-risk (TAR) designs across data sources. We report total patient exposure time in 1,000 patient-year (KYr) units, uncalibrated and calibrated hazard ratios (HRs) and their 95% confidence intervals (CIs). Greyed out entries did not pass study diagnostics and are included only for completeness.

| | Patients | | Exposure (KYr) | | Outcomes | | Uncalibrated | | Calibrated | |
|---|---|---|---|---|---|---|---|---|---|---|
| | T | C | T | C | T | C | HR (95% CI) | $p$ | HR (95% CI) | $p$ |
| **PS-matching / on-treatment TAR** | | | | | | | | | | |
| CCAE | 14,340 | 22,669 | 10.7 | 16.9 | 318 | 531 | 0.90 (0.74 - 1.10) | 0.31 | 0.92 (0.75 - 1.12) | 0.47 |
| DAG | 2,601 | 7,938 | 2.1 | 8.3 | 27 | 115 | 0.92 (0.52 - 1.55) | 0.75 | 0.80 (0.46 - 1.39) | 0.47 |
| ODOD | 9,254 | 17,588 | 6.0 | 13.2 | 179 | 512 | 0.85 (0.67 - 1.07) | 0.18 | 0.88 (0.70 - 1.12) | 0.32 |
| MDCR | 1,171 | 4,258 | 0.8 | 3.5 | 20 | 130 | 0.76 (0.40 - 1.36) | 0.38 | 0.75 (0.41 - 1.37) | 0.37 |
| MDCD | 1,673 | 4,598 | 0.9 | 3.1 | 70 | 243 | 1.06 (0.74 - 1.50) | 0.75 | 1.03 (0.72 - 1.47) | 0.81 |
| Open Claims | 198,473 | 383,325 | 157.4 | 334.5 | 2,551 | 7,135 | 0.84 (0.79 - 0.90) | <0.01 | 0.88 (0.78 - 0.99) | 0.05 |
| OEHR | 13,565 | 30,253 | 4.2 | 15.6 | 125 | 502 | 1.06 (0.81 - 1.39) | 0.65 | 1.08 (0.82 - 1.41) | 0.61 |
| SIDIAP | 2,588 | 13,345 | 3.6 | 28.8 | 23 | 309 | 0.54 (0.30 - 0.93) | 0.04 | 0.58 (0.33 - 1.02) | 0.06 |
| Meta-analysis | 242,494 | 479,716 | 184.9 | 420.4 | 3,293 | 9,347 | 0.86 (0.81 - 0.92) | <0.01 | 0.90 (0.82 - 0.99) | 0.03 |
| Meta-analysis (w/o USOC) | 44,021 | 96,391 | 27.5 | 85.9 | 742 | 2,212 | 0.92 (0.81 - 1.04) | 0.17 | 0.94 (0.82 - 1.07) | 0.36 |
| **PS-stratification / on-treatment TAR** | | | | | | | | | | |
| CCAE | 16,475 | 22,697 | 11.9 | 16.9 | 353 | 533 | 0.93 (0.81 - 1.07) | 0.31 | 0.93 (0.81 - 1.08) | 0.40 |
| DAG | 2,869 | 7,964 | 2.2 | 8.3 | 27 | 115 | 1.06 (0.66 - 1.65) | 0.80 | 0.93 (0.59 - 1.47) | 0.73 |
| ODOD | 11,062 | 17,785 | 6.7 | 13.3 | 210 | 522 | 0.86 (0.72 - 1.02) | 0.09 | 0.88 (0.74 - 1.05) | 0.19 |
| MDCR | 1,336 | 4,310 | 0.8 | 3.6 | 21 | 132 | 0.69 (0.41 - 1.11) | 0.15 | 0.70 (0.43 - 1.15) | 0.17 |
| MDCD | 1,727 | 4,612 | 0.9 | 3.1 | 71 | 245 | 0.95 (0.72 - 1.25) | 0.72 | 0.91 (0.69 - 1.20) | 0.52 |
| Open Claims | 235,920 | 383,765 | 176.1 | 334.9 | 2,889 | 7,163 | 0.87 (0.83 - 0.91) | <0.01 | 0.92 (0.82 - 1.02) | 0.14 |
| OEHR | 15,028 | 31,401 | 4.6 | 16.2 | 137 | 529 | 1.03 (0.84 - 1.26) | 0.75 | 1.09 (0.89 - 1.35) | 0.43 |
| SIDIAP | 2,759 | 13,789 | 3.7 | 30.2 | 26 | 323 | 0.69 (0.44 - 1.04) | 0.09 | 0.69 (0.45 - 1.07) | 0.11 |
| Meta-analysis | 284,113 | 477,401 | 205.3 | 419.8 | 3,642 | 9,185 | 0.88 (0.85 - 0.92) | <0.01 | 0.91 (0.82 - 1.00) | 0.06 |
| Meta-analysis (w/o USOC) | 48,193 | 93,636 | 29.2 | 84.9 | 753 | 2,022 | 0.92 (0.84 - 1.01) | 0.09 | 0.95 (0.86 - 1.04) | 0.28 |
| **PS-matching / on-treatment TAR without escalation** | | | | | | | | | | |
| CCAE | 14,340 | 22,669 | 9.3 | 13.8 | 269 | 431 | 0.83 (0.67 - 1.03) | 0.10 | 0.82 (0.66 - 1.02) | 0.12 |
| DAG | 2,601 | 7,938 | 1.9 | 7.4 | 23 | 101 | 0.87 (0.46 - 1.53) | 0.63 | 0.81 (0.44 - 1.47) | 0.50 |
| ODOD | 9,254 | 17,588 | 5.3 | 11.2 | 158 | 427 | 0.86 (0.66 - 1.10) | 0.22 | 0.88 (0.69 - 1.14) | 0.36 |
| MDCR | 1,171 | 4,258 | 0.7 | 3.1 | 19 | 107 | 0.72 (0.36 - 1.34) | 0.32 | 0.67 (0.35 - 1.29) | 0.25 |
| MDCD | 1,673 | 4,598 | 0.8 | 2.5 | 56 | 181 | 1.15 (0.77 - 1.69) | 0.49 | 1.11 (0.75 - 1.65) | 0.62 |
| Open Claims | 198,473 | 383,325 | 132.2 | 267.3 | 2,040 | 5,628 | 0.81 (0.76 - 0.87) | <0.01 | 0.85 (0.78 - 0.92) | <0.01 |
| OEHR | 13,565 | 30,253 | 3.7 | 12.6 | 102 | 385 | 1.11 (0.82 - 1.50) | 0.48 | 1.14 (0.84 - 1.54) | 0.43 |
| SIDIAP | 2,588 | 13,345 | 3.2 | 22.9 | 18 | 231 | 0.58 (0.30 - 1.04) | 0.09 | 0.67 (0.36 - 1.24) | 0.22 |
| Meta-analysis | 242,494 | 479,716 | 156.2 | 337.8 | 2,666 | 7,384 | 0.86 (0.78 - 0.95) | <0.01 | 0.89 (0.79 - 1.00) | 0.04 |
| Meta-analysis (w/o USOC) | 44,021 | 96,391 | 24.1 | 70.4 | 626 | 1,756 | 0.91 (0.78 - 1.05) | 0.20 | 0.92 (0.79 - 1.07) | 0.31 |
| **PS-stratification / on-treatment TAR without escalation** | | | | | | | | | | |
| CCAE | 16,475 | 22,697 | 10.3 | 13.9 | 301 | 433 | 0.94 (0.80 - 1.09) | 0.42 | 0.91 (0.78 - 1.07) | 0.29 |
| DAG | 2,869 | 7,964 | 2.0 | 7.5 | 23 | 101 | 1.12 (0.67 - 1.79) | 0.66 | 0.97 (0.59 - 1.59) | 0.80 |
| ODOD | 11,062 | 17,785 | 6.0 | 11.3 | 179 | 434 | 0.88 (0.72 - 1.06) | 0.17 | 0.88 (0.72 - 1.06) | 0.20 |
| MDCR | 1,336 | 4,310 | 0.7 | 3.1 | 20 | 109 | 0.78 (0.46 - 1.28) | 0.35 | 0.77 (0.46 - 1.29) | 0.33 |
| MDCD | 1,727 | 4,612 | 0.8 | 2.5 | 56 | 183 | 0.93 (0.68 - 1.27) | 0.68 | 0.88 (0.64 - 1.21) | 0.47 |
| Open Claims | 235,920 | 383,765 | 148.9 | 267.7 | 2,319 | 5,654 | 0.85 (0.81 - 0.90) | <0.01 | 0.89 (0.82 - 0.98) | 0.03 |
| OEHR | 15,028 | 31,401 | 4.1 | 13.1 | 112 | 407 | 1.04 (0.83 - 1.30) | 0.73 | 1.09 (0.86 - 1.37) | 0.51 |
| SIDIAP | 2,759 | 13,789 | 3.3 | 24.0 | 20 | 241 | 0.63 (0.37 - 1.00) | 0.06 | 0.66 (0.40 - 1.07) | 0.11 |
| Meta-analysis | 284,113 | 477,401 | 174.5 | 337.4 | 2,954 | 7,270 | 0.88 (0.82 - 0.95) | <0.01 | 0.91 (0.81 - 1.01) | 0.08 |
| Meta-analysis (w/o USOC) | 48,193 | 93,636 | 25.7 | 69.8 | 635 | 1,616 | 0.93 (0.83 - 1.04) | 0.19 | 0.93 (0.83 - 1.04) | 0.25 |

#### 1.3 SGLT2I (female) vs SU (female)

##### 1.3.1 3-point MACE

**Supplementary Table 53:** Exposures, outcomes and risk estimates for 3-point MACE in SGLT2I (female) (T) vs SU (female) (C) new-user cohorts under four different propensity score (PS) adjustment / time-at-risk (TAR) designs across data sources. We report total patient exposure time in 1,000 patient-year (KYr) units, uncalibrated and calibrated hazard ratios (HRs) and their 95% confidence intervals (CIs). Greyed out entries did not pass study diagnostics and are included only for completeness.

| | Patients | | Exposure (KYr) | | Outcomes | | Uncalibrated | | Calibrated | |
|---|---|---|---|---|---|---|---|---|---|---|
| | T | C | T | C | T | C | HR (95% CI) | $p$ | HR (95% CI) | $p$ |
| **PS-matching / on-treatment TAR** | | | | | | | | | | |
| CCAE | 15,697 | 34,271 | 11.6 | 24.4 | 75 | 178 | 1.00 (0.67 - 1.46) | 0.98 | 0.95 (0.64 - 1.41) | 0.81 |
| DAG | 3,180 | 682 | 2.5 | 0.7 | 0 | 0 | | | | |
| ODOD | 11,353 | 38,676 | 7.1 | 31.9 | 78 | 748 | 0.82 (0.60 - 1.10) | 0.20 | 0.84 (0.62 - 1.15) | 0.30 |
| MDCR | 1,260 | 6,287 | 0.8 | 5.0 | 10 | 147 | 0.47 (0.20 - 0.97) | 0.06 | 0.48 (0.22 - 1.07) | 0.08 |
| MDCD | 2,240 | 10,840 | 1.2 | 6.7 | 20 | 127 | 1.22 (0.66 - 2.14) | 0.52 | 1.11 (0.62 - 2.01) | 0.73 |
| Open Claims | 250,635 | 792,791 | 188.5 | 747.0 | 1,095 | 8,730 | 0.74 (0.68 - 0.81) | <0.01 | 0.76 (0.63 - 0.92) | <0.01 |
| OEHR | 15,228 | 62,852 | 4.8 | 35.7 | 31 | 432 | 1.18 (0.75 - 1.80) | 0.47 | 1.23 (0.78 - 1.92) | 0.43 |
| SIDIAP | 2,768 | 954 | 3.8 | 1.7 | 13 | <5 | 1.69 (0.48 - 7.88) | 0.46 | 1.98 (0.49 - 8.04) | 0.34 |
| Meta-analysis | 296,413 | 945,717 | 214.1 | 850.8 | 1,309 | 10,362 | 0.85 (0.71 - 1.03) | 0.10 | 0.85 (0.67 - 1.09) | 0.19 |
| Meta-analysis (w/o USOC) | 45,778 | 152,926 | 25.5 | 103.9 | 214 | 1,632 | 0.93 (0.73 - 1.19) | 0.57 | 0.92 (0.72 - 1.17) | 0.50 |
| **PS-stratification / on-treatment TAR** | | | | | | | | | | |
| CCAE | 18,456 | 34,871 | 13.4 | 24.6 | 88 | 182 | 1.02 (0.77 - 1.35) | 0.88 | 0.99 (0.75 - 1.32) | 0.92 |
| DAG | 3,231 | 1,228 | 2.5 | 1.2 | 0 | 0 | | | | |
| ODOD | 12,278 | 39,450 | 7.6 | 32.7 | 85 | 781 | 0.76 (0.59 - 0.97) | 0.03 | 0.80 (0.62 - 1.02) | 0.08 |
| MDCR | 1,406 | 6,710 | 0.9 | 5.4 | 12 | 159 | 0.49 (0.25 - 0.90) | 0.03 | 0.48 (0.25 - 0.92) | 0.03 |
| MDCD | 2,284 | 11,111 | 1.2 | 6.9 | 20 | 131 | 1.06 (0.63 - 1.72) | 0.81 | 0.98 (0.59 - 1.62) | 0.90 |
| Open Claims | 263,915 | 801,770 | 196.3 | 755.2 | 1,135 | 9,013 | 0.71 (0.66 - 0.75) | <0.01 | 0.72 (0.61 - 0.86) | <0.01 |
| OEHR | 16,377 | 69,021 | 5.1 | 39.4 | 31 | 512 | 0.82 (0.55 - 1.20) | 0.33 | 0.86 (0.58 - 1.27) | 0.47 |
| SIDIAP | 2,852 | 1,992 | 3.9 | 4.3 | 13 | 11 | 1.04 (0.38 - 2.85) | 0.95 | 0.94 (0.34 - 2.59) | 0.88 |
| Meta-analysis | 311,026 | 945,112 | 222.3 | 851.9 | 1,339 | 10,488 | 0.79 (0.67 - 0.93) | <0.01 | 0.80 (0.64 - 0.99) | 0.04 |
| Meta-analysis (w/o USOC) | 47,111 | 143,342 | 26.1 | 96.7 | 204 | 1,475 | 0.86 (0.71 - 1.03) | 0.11 | 0.87 (0.72 - 1.05) | 0.16 |
| **PS-matching / on-treatment TAR without escalation** | | | | | | | | | | |
| CCAE | 15,697 | 34,271 | 10.0 | 21.0 | 59 | 154 | 1.02 (0.67 - 1.53) | 0.94 | 0.96 (0.63 - 1.45) | 0.84 |
| DAG | 3,180 | 682 | 2.3 | 0.6 | 0 | 0 | | | | |
| ODOD | 11,353 | 38,676 | 6.3 | 28.5 | 70 | 660 | 0.84 (0.61 - 1.14) | 0.27 | 0.85 (0.62 - 1.16) | 0.33 |
| MDCR | 1,260 | 6,287 | 0.7 | 4.6 | 10 | 135 | 0.52 (0.22 - 1.09) | 0.11 | 0.53 (0.24 - 1.18) | 0.12 |
| MDCD | 2,240 | 10,840 | 1.0 | 5.6 | 12 | 107 | 0.85 (0.39 - 1.70) | 0.66 | 0.81 (0.39 - 1.69) | 0.58 |
| Open Claims | 250,635 | 792,791 | 158.3 | 629.5 | 911 | 7,430 | 0.74 (0.67 - 0.81) | <0.01 | 0.76 (0.63 - 0.91) | <0.01 |
| OEHR | 15,228 | 62,852 | 4.2 | 30.4 | 26 | 327 | 1.18 (0.72 - 1.86) | 0.50 | 1.22 (0.75 - 1.96) | 0.45 |
| SIDIAP | 2,768 | 954 | 3.4 | 1.4 | 12 | <5 | 1.39 (0.35 - 6.83) | 0.66 | 1.79 (0.40 - 7.94) | 0.45 |
| Meta-analysis | 296,413 | 945,717 | 180.5 | 719.6 | 1,088 | 8,813 | 0.82 (0.69 - 0.97) | 0.02 | 0.82 (0.66 - 1.03) | 0.08 |
| Meta-analysis (w/o USOC) | 45,778 | 152,926 | 22.2 | 90.1 | 177 | 1,383 | 0.91 (0.74 - 1.11) | 0.35 | 0.89 (0.72 - 1.10) | 0.29 |
| **PS-stratification / on-treatment TAR without escalation** | | | | | | | | | | |
| CCAE | 18,456 | 34,871 | 11.5 | 21.2 | 71 | 157 | 0.92 (0.67 - 1.25) | 0.61 | 0.88 (0.65 - 1.20) | 0.44 |
| DAG | 3,231 | 1,228 | 2.3 | 1.2 | 0 | 0 | | | | |
| ODOD | 12,278 | 39,450 | 6.7 | 29.2 | 77 | 691 | 0.77 (0.59 - 0.99) | 0.05 | 0.80 (0.62 - 1.04) | 0.10 |
| MDCR | 1,406 | 6,710 | 0.8 | 4.9 | 10 | 146 | 0.47 (0.22 - 0.90) | 0.04 | 0.44 (0.22 - 0.89) | 0.02 |
| MDCD | 2,284 | 11,111 | 1.0 | 5.7 | 12 | 110 | 0.80 (0.40 - 1.43) | 0.48 | 0.72 (0.38 - 1.35) | 0.31 |
| Open Claims | 263,915 | 801,770 | 165.0 | 636.6 | 947 | 7,674 | 0.70 (0.65 - 0.75) | <0.01 | 0.71 (0.61 - 0.83) | <0.01 |
| OEHR | 16,377 | 69,021 | 4.5 | 33.5 | 26 | 381 | 0.86 (0.55 - 1.29) | 0.48 | 0.86 (0.56 - 1.33) | 0.54 |
| SIDIAP | 2,852 | 1,992 | 3.4 | 3.5 | 12 | 11 | 0.88 (0.31 - 2.50) | 0.81 | 0.89 (0.31 - 2.56) | 0.83 |
| Meta-analysis | 311,026 | 945,112 | 187.8 | 720.6 | 1,121 | 8,903 | 0.76 (0.66 - 0.87) | <0.01 | 0.76 (0.63 - 0.91) | <0.01 |
| Meta-analysis (w/o USOC) | 47,111 | 143,342 | 22.7 | 83.9 | 174 | 1,229 | 0.84 (0.70 - 1.00) | 0.05 | 0.83 (0.69 - 0.99) | 0.05 |

##### 1.3.2 4-point MACE

**Supplementary Table 54:** Exposures, outcomes and risk estimates for 4-point MACE in SGLT2I (female) (T) vs SU (female) (C) new-user cohorts under four different propensity score (PS) adjustment / time-at-risk (TAR) designs across data sources. We report total patient exposure time in 1,000 patient-year (KYr) units, uncalibrated and calibrated hazard ratios (HRs) and their 95% confidence intervals (CIs). Greyed out entries did not pass study diagnostics and are included only for completeness.

| | Patients | | Exposure (KYr) | | Outcomes | | Uncalibrated | | Calibrated | |
|---|---|---|---|---|---|---|---|---|---|---|
| | T | C | T | C | T | C | HR (95% CI) | $p$ | HR (95% CI) | $p$ |
| **PS-matching / on-treatment TAR** | | | | | | | | | | |
| CCAE | 15,500 | 33,636 | 11.4 | 23.9 | 105 | 289 | 1.00 (0.73 - 1.36) | 0.98 | 0.95 (0.69 - 1.30) | 0.76 |
| DAG | 3,180 | 682 | 2.5 | 0.7 | 0 | 0 | | | | |
| ODOD | 10,944 | 36,544 | 6.9 | 29.9 | 119 | 1,184 | 0.72 (0.57 - 0.92) | <0.01 | 0.75 (0.59 - 0.95) | 0.03 |
| MDCR | 1,196 | 5,866 | 0.8 | 4.7 | 18 | 225 | 0.47 (0.22 - 0.90) | 0.04 | 0.49 (0.24 - 0.98) | 0.05 |
| MDCD | 2,122 | 10,183 | 1.1 | 6.3 | 35 | 206 | 1.10 (0.68 - 1.72) | 0.69 | 1.01 (0.63 - 1.60) | 0.88 |
| Open Claims | 243,852 | 766,558 | 184.1 | 718.2 | 1,628 | 14,529 | 0.67 (0.63 - 0.72) | <0.01 | 0.69 (0.57 - 0.83) | <0.01 |
| OEHR | 15,021 | 61,694 | 4.7 | 34.7 | 59 | 805 | 1.00 (0.72 - 1.37) | 0.99 | 1.04 (0.74 - 1.47) | 0.83 |
| SIDIAP | 2,745 | 952 | 3.8 | 1.7 | 14 | <5 | 1.58 (0.45 - 7.40) | 0.52 | 1.85 (0.45 - 7.56) | 0.39 |
| Meta-analysis | 288,635 | 914,481 | 209.1 | 817.7 | 1,964 | 17,238 | 0.80 (0.66 - 0.98) | 0.03 | 0.80 (0.63 - 1.03) | 0.08 |
| Meta-analysis (w/o USOC) | 44,783 | 147,923 | 25.0 | 99.5 | 336 | 2,709 | 0.87 (0.69 - 1.08) | 0.21 | 0.85 (0.68 - 1.07) | 0.18 |
| **PS-stratification / on-treatment TAR** | | | | | | | | | | |
| CCAE | 18,202 | 34,311 | 13.2 | 24.2 | 121 | 294 | 0.88 (0.69 - 1.10) | 0.27 | 0.85 (0.68 - 1.08) | 0.21 |
| DAG | 3,231 | 1,228 | 2.5 | 1.2 | 0 | 0 | | | | |
| ODOD | 11,798 | 37,406 | 7.3 | 30.8 | 130 | 1,261 | 0.66 (0.54 - 0.80) | <0.01 | 0.69 (0.57 - 0.84) | <0.01 |
| MDCR | 1,334 | 6,198 | 0.9 | 4.9 | 20 | 245 | 0.59 (0.35 - 0.96) | 0.04 | 0.58 (0.35 - 0.97) | 0.05 |
| MDCD | 2,158 | 10,389 | 1.2 | 6.4 | 35 | 208 | 1.09 (0.73 - 1.58) | 0.67 | 1.00 (0.68 - 1.47) | 0.89 |
| Open Claims | 255,645 | 774,956 | 191.0 | 725.9 | 1,703 | 15,047 | 0.64 (0.61 - 0.68) | <0.01 | 0.66 (0.56 - 0.78) | <0.01 |
| OEHR | 16,145 | 67,681 | 5.1 | 38.2 | 59 | 943 | 0.79 (0.59 - 1.04) | 0.10 | 0.82 (0.62 - 1.09) | 0.21 |
| SIDIAP | 2,828 | 1,990 | 3.9 | 4.3 | 14 | 12 | 0.87 (0.33 - 2.32) | 0.79 | 0.79 (0.30 - 2.11) | 0.65 |
| Meta-analysis | 301,790 | 914,354 | 216.6 | 819.0 | 2,013 | 17,545 | 0.71 (0.61 - 0.82) | <0.01 | 0.72 (0.59 - 0.88) | <0.01 |
| Meta-analysis (w/o USOC) | 46,145 | 139,398 | 25.5 | 93.1 | 310 | 2,498 | 0.76 (0.63 - 0.91) | <0.01 | 0.77 (0.64 - 0.92) | <0.01 |
| **PS-matching / on-treatment TAR without escalation** | | | | | | | | | | |
| CCAE | 15,500 | 33,636 | 9.9 | 20.6 | 86 | 250 | 0.91 (0.64 - 1.27) | 0.58 | 0.86 (0.61 - 1.20) | 0.38 |
| DAG | 3,180 | 682 | 2.3 | 0.6 | 0 | 0 | | | | |
| ODOD | 10,944 | 36,544 | 6.1 | 26.8 | 101 | 1,043 | 0.70 (0.53 - 0.90) | <0.01 | 0.71 (0.54 - 0.91) | 0.01 |
| MDCR | 1,196 | 5,866 | 0.7 | 4.3 | 17 | 209 | 0.54 (0.25 - 1.04) | 0.08 | 0.55 (0.27 - 1.11) | 0.10 |
| MDCD | 2,122 | 10,183 | 0.9 | 5.2 | 24 | 167 | 0.92 (0.53 - 1.54) | 0.77 | 0.88 (0.51 - 1.50) | 0.65 |
| Open Claims | 243,852 | 766,558 | 154.6 | 605.6 | 1,344 | 12,307 | 0.66 (0.61 - 0.71) | <0.01 | 0.67 (0.57 - 0.80) | <0.01 |
| OEHR | 15,021 | 61,694 | 4.1 | 29.6 | 49 | 626 | 1.00 (0.69 - 1.41) | 1.00 | 1.03 (0.72 - 1.48) | 0.84 |
| SIDIAP | 2,745 | 952 | 3.3 | 1.4 | 13 | <5 | 1.20 (0.30 - 5.86) | 0.81 | 1.54 (0.35 - 6.77) | 0.58 |
| Meta-analysis | 288,635 | 914,481 | 176.3 | 692.2 | 1,621 | 14,602 | 0.76 (0.64 - 0.90) | <0.01 | 0.76 (0.61 - 0.95) | 0.01 |
| Meta-analysis (w/o USOC) | 44,783 | 147,923 | 21.7 | 86.5 | 277 | 2,295 | 0.81 (0.68 - 0.98) | 0.03 | 0.80 (0.66 - 0.96) | 0.02 |
| **PS-stratification / on-treatment TAR without escalation** | | | | | | | | | | |
| CCAE | 18,202 | 34,311 | 11.4 | 20.9 | 102 | 253 | 0.83 (0.64 - 1.06) | 0.14 | 0.79 (0.61 - 1.02) | 0.08 |
| DAG | 3,231 | 1,228 | 2.3 | 1.2 | 0 | 0 | | | | |
| ODOD | 11,798 | 37,406 | 6.5 | 27.5 | 112 | 1,109 | 0.64 (0.51 - 0.78) | <0.01 | 0.66 (0.53 - 0.82) | <0.01 |
| MDCR | 1,334 | 6,198 | 0.8 | 4.5 | 17 | 228 | 0.58 (0.32 - 0.97) | 0.05 | 0.53 (0.31 - 0.93) | 0.03 |
| MDCD | 2,158 | 10,389 | 1.0 | 5.3 | 24 | 169 | 0.92 (0.57 - 1.42) | 0.72 | 0.83 (0.52 - 1.31) | 0.43 |
| Open Claims | 255,645 | 774,956 | 160.7 | 612.4 | 1,411 | 12,760 | 0.63 (0.60 - 0.67) | <0.01 | 0.65 (0.56 - 0.75) | <0.01 |
| OEHR | 16,145 | 67,681 | 4.4 | 32.6 | 49 | 722 | 0.80 (0.58 - 1.07) | 0.15 | 0.80 (0.58 - 1.10) | 0.22 |
| SIDIAP | 2,828 | 1,990 | 3.4 | 3.5 | 13 | 12 | 0.77 (0.28 - 2.11) | 0.61 | 0.78 (0.28 - 2.15) | 0.64 |
| Meta-analysis | 301,790 | 914,354 | 182.9 | 693.4 | 1,674 | 14,844 | 0.69 (0.60 - 0.78) | <0.01 | 0.69 (0.58 - 0.82) | <0.01 |
| Meta-analysis (w/o USOC) | 46,145 | 139,398 | 22.3 | 81.0 | 263 | 2,084 | 0.73 (0.62 - 0.87) | <0.01 | 0.73 (0.61 - 0.86) | <0.01 |

##### 1.3.3 Acute myocardial infarction

**Supplementary Table 55:** Exposures, outcomes and risk estimates for Acute myocardial infarction in SGLT2I (female) (T) vs SU (female) (C) new-user cohorts under four different propensity score (PS) adjustment / time-at-risk (TAR) designs across data sources. We report total patient exposure time in 1,000 patient-year (KYr) units, uncalibrated and calibrated hazard ratios (HRs) and their 95% confidence intervals (CIs). Greyed out entries did not pass study diagnostics and are included only for completeness.

| | Patients | | Exposure (KYr) | | Outcomes | | Uncalibrated | | Calibrated | |
|---|---|---|---|---|---|---|---|---|---|---|
| | T | C | T | C | T | C | HR (95% CI) | $p$ | HR (95% CI) | $p$ |
| **PS-matching / on-treatment TAR** | | | | | | | | | | |
| CCAE | 15,900 | 34,793 | 11.7 | 24.9 | 31 | 63 | 1.36 (0.70 - 2.59) | 0.36 | 1.30 (0.67 - 2.50) | 0.44 |
| DAG | 3,180 | 682 | 2.5 | 0.7 | 0 | 0 | | | | |
| ODOD | 11,601 | 40,002 | 7.3 | 33.3 | 32 | 330 | 0.47 (0.28 - 0.77) | <0.01 | 0.49 (0.29 - 0.82) | <0.01 |
| MDCR | 1,290 | 6,575 | 0.9 | 5.3 | 6 | 58 | 0.66 (0.18 - 1.87) | 0.48 | 0.68 (0.21 - 2.18) | 0.52 |
| MDCD | 2,318 | 11,314 | 1.3 | 7.1 | 8 | 48 | 1.54 (0.56 - 3.82) | 0.38 | 1.41 (0.54 - 3.66) | 0.49 |
| Open Claims | 256,294 | 815,039 | 192.5 | 770.0 | 527 | 3,768 | 0.82 (0.73 - 0.93) | <0.01 | 0.84 (0.68 - 1.04) | 0.11 |
| OEHR | 15,344 | 63,602 | 4.8 | 36.3 | 17 | 213 | 1.19 (0.65 - 2.08) | 0.55 | 1.24 (0.69 - 2.25) | 0.51 |
| SIDIAP | 2,787 | 963 | 3.9 | 1.7 | <5 | <5 | 1.00 (0.09 - 22.8) | 1.00 | 1.17 (0.07 - 19.1) | 0.91 |
| Meta-analysis | 302,747 | 971,325 | 218.4 | 876.9 | 621 | 4,480 | 0.89 (0.63 - 1.26) | 0.50 | 0.89 (0.61 - 1.30) | 0.54 |
| Meta-analysis (w/o USOC) | 46,453 | 156,286 | 25.9 | 106.9 | 94 | 712 | 0.94 (0.57 - 1.55) | 0.79 | 0.92 (0.56 - 1.52) | 0.74 |
| **PS-stratification / on-treatment TAR** | | | | | | | | | | |
| CCAE | 18,670 | 35,410 | 13.5 | 25.1 | 35 | 66 | 1.23 (0.77 - 1.92) | 0.38 | 1.19 (0.75 - 1.89) | 0.46 |
| DAG | 3,231 | 1,228 | 2.5 | 1.2 | 0 | 0 | | | | |
| ODOD | 12,545 | 40,829 | 7.7 | 34.1 | 34 | 346 | 0.63 (0.42 - 0.92) | 0.02 | 0.66 (0.45 - 0.98) | 0.04 |
| MDCR | 1,439 | 7,041 | 0.9 | 5.7 | 7 | 65 | 0.79 (0.30 - 1.78) | 0.60 | 0.78 (0.32 - 1.88) | 0.58 |
| MDCD | 2,362 | 11,609 | 1.3 | 7.3 | 8 | 53 | 1.20 (0.50 - 2.52) | 0.66 | 1.10 (0.49 - 2.46) | 0.82 |
| Open Claims | 269,891 | 824,655 | 200.4 | 779.0 | 548 | 3,897 | 0.80 (0.73 - 0.88) | <0.01 | 0.82 (0.68 - 0.99) | 0.04 |
| OEHR | 16,491 | 69,757 | 5.2 | 40.0 | 17 | 248 | 0.86 (0.49 - 1.42) | 0.57 | 0.90 (0.53 - 1.52) | 0.69 |
| SIDIAP | 2,874 | 2,005 | 3.9 | 4.3 | <5 | 5 | 0.44 (0.06 - 2.69) | 0.39 | 0.40 (0.06 - 2.66) | 0.34 |
| Meta-analysis | 317,597 | 970,651 | 226.8 | 878.2 | 634 | 4,557 | 0.80 (0.73 - 0.88) | <0.01 | 0.81 (0.69 - 0.96) | 0.01 |
| Meta-analysis (w/o USOC) | 47,706 | 145,996 | 26.5 | 99.2 | 86 | 660 | 0.86 (0.58 - 1.29) | 0.46 | 0.87 (0.58 - 1.30) | 0.50 |
| **PS-matching / on-treatment TAR without escalation** | | | | | | | | | | |
| CCAE | 15,900 | 34,793 | 10.1 | 21.4 | 24 | 56 | 1.51 (0.75 - 2.96) | 0.24 | 1.42 (0.71 - 2.82) | 0.32 |
| DAG | 3,180 | 682 | 2.3 | 0.6 | 0 | 0 | | | | |
| ODOD | 11,601 | 40,002 | 6.5 | 29.7 | 27 | 288 | 0.46 (0.26 - 0.77) | <0.01 | 0.47 (0.27 - 0.81) | <0.01 |
| MDCR | 1,290 | 6,575 | 0.8 | 4.8 | 6 | 54 | 0.69 (0.19 - 2.03) | 0.54 | 0.70 (0.21 - 2.31) | 0.56 |
| MDCD | 2,318 | 11,314 | 1.0 | 5.9 | 6 | 39 | 1.62 (0.53 - 4.45) | 0.37 | 1.54 (0.53 - 4.46) | 0.43 |
| Open Claims | 256,294 | 815,039 | 161.6 | 648.6 | 439 | 3,217 | 0.84 (0.73 - 0.96) | <0.01 | 0.86 (0.70 - 1.06) | 0.16 |
| OEHR | 15,344 | 63,602 | 4.2 | 30.8 | 16 | 157 | 1.49 (0.78 - 2.67) | 0.21 | 1.54 (0.83 - 2.85) | 0.19 |
| SIDIAP | 2,787 | 963 | 3.4 | 1.4 | <5 | <5 | | | | |
| Meta-analysis | 302,747 | 971,325 | 184.1 | 741.2 | 518 | 3,811 | 0.95 (0.63 - 1.45) | 0.83 | 0.96 (0.62 - 1.48) | 0.84 |
| Meta-analysis (w/o USOC) | 46,453 | 156,286 | 22.5 | 92.6 | 79 | 594 | 1.01 (0.57 - 1.82) | 0.96 | 0.99 (0.55 - 1.78) | 0.96 |
| **PS-stratification / on-treatment TAR without escalation** | | | | | | | | | | |
| CCAE | 18,670 | 35,410 | 11.7 | 21.6 | 28 | 58 | 1.10 (0.66 - 1.80) | 0.71 | 1.05 (0.63 - 1.73) | 0.85 |
| DAG | 3,231 | 1,228 | 2.3 | 1.2 | 0 | 0 | | | | |
| ODOD | 12,545 | 40,829 | 6.9 | 30.5 | 29 | 303 | 0.62 (0.40 - 0.92) | 0.02 | 0.64 (0.42 - 0.97) | 0.04 |
| MDCR | 1,439 | 7,041 | 0.8 | 5.2 | 6 | 61 | 0.80 (0.29 - 1.88) | 0.64 | 0.75 (0.29 - 1.90) | 0.54 |
| MDCD | 2,362 | 11,609 | 1.0 | 6.0 | 6 | 42 | 1.24 (0.45 - 2.92) | 0.65 | 1.12 (0.44 - 2.86) | 0.81 |
| Open Claims | 269,891 | 824,655 | 168.5 | 656.4 | 458 | 3,333 | 0.80 (0.72 - 0.89) | <0.01 | 0.82 (0.69 - 0.97) | 0.02 |
| OEHR | 16,491 | 69,757 | 4.5 | 33.9 | 16 | 184 | 1.05 (0.58 - 1.77) | 0.87 | 1.06 (0.60 - 1.85) | 0.86 |
| SIDIAP | 2,874 | 2,005 | 3.4 | 3.6 | <5 | 5 | 0.17 (0.01 - 1.42) | 0.13 | 0.17 (0.02 - 1.68) | 0.13 |
| Meta-analysis | 317,597 | 970,651 | 191.5 | 742.4 | 531 | 3,878 | 0.81 (0.73 - 0.89) | <0.01 | 0.81 (0.69 - 0.94) | <0.01 |
| Meta-analysis (w/o USOC) | 47,706 | 145,996 | 23.0 | 86.0 | 73 | 545 | 0.87 (0.59 - 1.28) | 0.47 | 0.86 (0.58 - 1.27) | 0.44 |

##### 1.3.4 Acute renal failure

**Supplementary Table 56:** Exposures, outcomes and risk estimates for Acute renal failure in SGLT2I (female) (T) vs SU (female) (C) new-user cohorts under four different propensity score (PS) adjustment / time-at-risk (TAR) designs across data sources. We report total patient exposure time in 1,000 patient-year (KYr) units, uncalibrated and calibrated hazard ratios (HRs) and their 95% confidence intervals (CIs). Greyed out entries did not pass study diagnostics and are included only for completeness.

| | Patients | | Exposure (KYr) | | Outcomes | | Uncalibrated | | Calibrated | |
|---|---|---|---|---|---|---|---|---|---|---|
| | T | C | T | C | T | C | HR (95% CI) | $p$ | HR (95% CI) | $p$ |
| **PS-matching / on-treatment TAR** | | | | | | | | | | |
| CCAE | 15,896 | 34,558 | 11.7 | 24.6 | 52 | 224 | 0.54 (0.35 - 0.82) | <0.01 | 0.52 (0.34 - 0.79) | <0.01 |
| DAG | 3,180 | 682 | 2.5 | 0.7 | 0 | 0 | | | | |
| ODOD | 11,479 | 38,470 | 7.2 | 31.6 | 112 | 1,096 | 0.73 (0.57 - 0.93) | 0.01 | 0.75 (0.59 - 0.97) | 0.04 |
| MDCR | 1,294 | 6,359 | 0.9 | 5.1 | 12 | 190 | 0.46 (0.21 - 0.90) | 0.04 | 0.47 (0.23 - 0.98) | 0.05 |
| MDCD | 2,271 | 10,872 | 1.2 | 6.7 | 20 | 206 | 0.60 (0.31 - 1.08) | 0.11 | 0.55 (0.29 - 1.03) | 0.06 |
| Open Claims | 255,466 | 803,475 | 192.0 | 754.8 | 1,320 | 11,815 | 0.67 (0.62 - 0.72) | <0.01 | 0.68 (0.57 - 0.82) | <0.01 |
| OEHR | 15,348 | 63,238 | 4.8 | 35.8 | 38 | 574 | 0.91 (0.59 - 1.36) | 0.64 | 0.94 (0.62 - 1.45) | 0.82 |
| SIDIAP | 2,849 | 965 | 4.0 | 1.7 | <5 | <5 | | | | |
| Meta-analysis | 301,754 | 956,972 | 217.8 | 858.7 | 1,554 | 14,105 | 0.67 (0.63 - 0.72) | <0.01 | 0.67 (0.57 - 0.79) | <0.01 |
| Meta-analysis (w/o USOC) | 46,288 | 153,497 | 25.8 | 103.9 | 234 | 2,290 | 0.68 (0.56 - 0.83) | <0.01 | 0.67 (0.55 - 0.82) | <0.01 |
| **PS-stratification / on-treatment TAR** | | | | | | | | | | |
| CCAE | 18,692 | 35,143 | 13.5 | 24.9 | 62 | 226 | 0.61 (0.45 - 0.83) | <0.01 | 0.60 (0.44 - 0.81) | <0.01 |
| DAG | 3,231 | 1,228 | 2.5 | 1.2 | 0 | 0 | | | | |
| ODOD | 12,447 | 39,271 | 7.6 | 32.4 | 120 | 1,156 | 0.68 (0.55 - 0.83) | <0.01 | 0.71 (0.58 - 0.87) | <0.01 |
| MDCR | 1,449 | 6,718 | 0.9 | 5.4 | 13 | 214 | 0.53 (0.28 - 0.93) | 0.04 | 0.52 (0.28 - 0.95) | 0.04 |
| MDCD | 2,313 | 11,156 | 1.2 | 6.9 | 21 | 214 | 0.62 (0.37 - 0.97) | 0.05 | 0.57 (0.35 - 0.92) | 0.02 |
| Open Claims | 269,448 | 812,179 | 200.1 | 762.8 | 1,386 | 12,217 | 0.63 (0.59 - 0.67) | <0.01 | 0.65 (0.55 - 0.76) | <0.01 |
| OEHR | 16,501 | 69,263 | 5.2 | 39.4 | 39 | 669 | 0.77 (0.54 - 1.08) | 0.15 | 0.81 (0.57 - 1.14) | 0.25 |
| SIDIAP | 2,950 | 2,012 | 4.0 | 4.3 | <5 | <5 | 0.87 (0.03 - 22.9) | 0.93 | 0.79 (0.03 - 20.8) | 0.89 |
| Meta-analysis | 317,088 | 955,856 | 226.5 | 859.5 | 1,607 | 14,268 | 0.64 (0.60 - 0.67) | <0.01 | 0.64 (0.55 - 0.74) | <0.01 |
| Meta-analysis (w/o USOC) | 47,640 | 143,677 | 26.4 | 96.7 | 221 | 2,051 | 0.68 (0.58 - 0.79) | <0.01 | 0.68 (0.58 - 0.80) | <0.01 |
| **PS-matching / on-treatment TAR without escalation** | | | | | | | | | | |
| CCAE | 15,896 | 34,558 | 10.1 | 21.2 | 44 | 185 | 0.52 (0.32 - 0.82) | <0.01 | 0.50 (0.31 - 0.79) | <0.01 |
| DAG | 3,180 | 682 | 2.3 | 0.6 | 0 | 0 | | | | |
| ODOD | 11,479 | 38,470 | 6.4 | 28.3 | 98 | 961 | 0.76 (0.58 - 0.98) | 0.04 | 0.76 (0.59 - 1.00) | 0.06 |
| MDCR | 1,294 | 6,359 | 0.8 | 4.6 | 11 | 170 | 0.58 (0.26 - 1.16) | 0.16 | 0.59 (0.28 - 1.25) | 0.18 |
| MDCD | 2,271 | 10,872 | 1.0 | 5.6 | 13 | 160 | 0.51 (0.22 - 1.06) | 0.10 | 0.49 (0.22 - 1.07) | 0.08 |
| Open Claims | 255,466 | 803,475 | 161.3 | 636.5 | 1,059 | 9,892 | 0.65 (0.59 - 0.70) | <0.01 | 0.66 (0.55 - 0.79) | <0.01 |
| OEHR | 15,348 | 63,238 | 4.2 | 30.6 | 28 | 405 | 0.82 (0.50 - 1.30) | 0.41 | 0.85 (0.52 - 1.37) | 0.52 |
| SIDIAP | 2,849 | 965 | 3.5 | 1.4 | <5 | <5 | | | | |
| Meta-analysis | 301,754 | 956,972 | 183.7 | 726.8 | 1,253 | 11,773 | 0.65 (0.60 - 0.70) | <0.01 | 0.65 (0.56 - 0.77) | <0.01 |
| Meta-analysis (w/o USOC) | 46,288 | 153,497 | 22.4 | 90.3 | 194 | 1,881 | 0.69 (0.57 - 0.84) | <0.01 | 0.68 (0.55 - 0.82) | <0.01 |
| **PS-stratification / on-treatment TAR without escalation** | | | | | | | | | | |
| CCAE | 18,692 | 35,143 | 11.7 | 21.4 | 53 | 187 | 0.63 (0.45 - 0.88) | <0.01 | 0.61 (0.43 - 0.85) | <0.01 |
| DAG | 3,231 | 1,228 | 2.3 | 1.2 | 0 | 0 | | | | |
| ODOD | 12,447 | 39,271 | 6.8 | 29.0 | 103 | 1,018 | 0.67 (0.54 - 0.83) | <0.01 | 0.70 (0.56 - 0.87) | <0.01 |
| MDCR | 1,449 | 6,718 | 0.8 | 4.9 | 12 | 192 | 0.56 (0.28 - 1.00) | 0.07 | 0.52 (0.28 - 0.98) | 0.05 |
| MDCD | 2,313 | 11,156 | 1.0 | 5.7 | 14 | 166 | 0.53 (0.28 - 0.91) | 0.03 | 0.48 (0.27 - 0.85) | 0.01 |
| Open Claims | 269,448 | 812,179 | 168.4 | 643.5 | 1,117 | 10,227 | 0.61 (0.57 - 0.65) | <0.01 | 0.62 (0.54 - 0.72) | <0.01 |
| OEHR | 16,501 | 69,263 | 4.5 | 33.6 | 29 | 473 | 0.72 (0.47 - 1.06) | 0.11 | 0.72 (0.48 - 1.09) | 0.15 |
| SIDIAP | 2,950 | 2,012 | 3.5 | 3.6 | <5 | <5 | 0.94 (0.04 - 24.4) | 0.97 | 0.95 (0.04 - 24.8) | 0.96 |
| Meta-analysis | 317,088 | 955,856 | 191.3 | 727.5 | 1,302 | 11,905 | 0.62 (0.58 - 0.66) | <0.01 | 0.62 (0.54 - 0.71) | <0.01 |
| Meta-analysis (w/o USOC) | 47,640 | 143,677 | 23.0 | 84.0 | 185 | 1,678 | 0.67 (0.57 - 0.79) | <0.01 | 0.66 (0.56 - 0.79) | <0.01 |

##### 1.3.5 Glycemic control

**Supplementary Table 57:** Exposures, outcomes and risk estimates for Glycemic control in SGLT2I (female) (T) vs SU (female) (C) new-user cohorts under four different propensity score (PS) adjustment / time-at-risk (TAR) designs across data sources. We report total patient exposure time in 1,000 patient-year (KYr) units, uncalibrated and calibrated hazard ratios (HRs) and their 95% confidence intervals (CIs). Greyed out entries did not pass study diagnostics and are included only for completeness.

| | Patients | | Exposure (KYr) | | Outcomes | | Uncalibrated | | Calibrated | |
|---|---|---|---|---|---|---|---|---|---|---|
| | T | C | T | C | T | C | HR (95% CI) | $p$ | HR (95% CI) | $p$ |
| **PS-matching / on-treatment TAR** | | | | | | | | | | |
| CCAE | 15,937 | 34,825 | 11.8 | 24.9 | 18 | 39 | 0.86 (0.38 - 1.86) | 0.71 | 0.82 (0.37 - 1.83) | 0.63 |
| DAG | 1,045 | 274 | 0.7 | 0.2 | 97 | 29 | 0.71 (0.41 - 1.27) | 0.24 | 0.78 (0.44 - 1.38) | 0.35 |
| ODOD | 7,412 | 26,878 | 4.3 | 19.9 | 430 | 2,143 | 0.93 (0.81 - 1.08) | 0.36 | 0.96 (0.83 - 1.12) | 0.67 |
| MDCR | 1,327 | 6,736 | 0.9 | 5.5 | 0 | 9 | | | | |
| MDCD | 2,388 | 11,679 | 1.3 | 7.3 | 0 | 0 | | | | |
| Open Claims | 261,894 | 832,892 | 196.5 | 789.1 | 0 | 0 | | | | |
| OEHR | 15,494 | 64,240 | 4.9 | 36.8 | 0 | 0 | | | | |
| SIDIAP | 95 | 222 | 0.1 | 0.3 | 32 | 75 | 0.80 (0.45 - 1.39) | 0.45 | 0.94 (0.53 - 1.66) | 0.74 |
| Meta-analysis | 23,444 | 61,925 | 16.2 | 45.1 | 480 | 2,257 | 0.92 (0.80 - 1.06) | 0.26 | 0.92 (0.75 - 1.13) | 0.43 |
| Meta-analysis (w/o USOC) | 23,444 | 61,925 | 16.2 | 45.1 | 480 | 2,257 | 0.92 (0.80 - 1.06) | 0.26 | 0.91 (0.79 - 1.05) | 0.21 |
| **PS-stratification / on-treatment TAR** | | | | | | | | | | |
| CCAE | 18,683 | 35,323 | 13.5 | 25.1 | 22 | 47 | 1.18 (0.67 - 2.04) | 0.56 | 1.15 (0.66 - 2.01) | 0.62 |
| DAG | 1,071 | 535 | 0.7 | 0.5 | 100 | 62 | 0.92 (0.59 - 1.44) | 0.70 | 0.59 (0.38 - 0.93) | 0.04 |
| ODOD | 7,839 | 27,419 | 4.4 | 20.4 | 456 | 2,208 | 1.05 (0.94 - 1.17) | 0.42 | 1.10 (0.98 - 1.23) | 0.15 |
| MDCR | 1,482 | 7,185 | 0.9 | 5.9 | 0 | 10 | | | | |
| MDCD | 2,435 | 11,946 | 1.3 | 7.5 | 0 | 0 | | | | |
| Open Claims | 276,521 | 842,971 | 205.0 | 798.5 | 0 | 0 | | | | |
| OEHR | 16,666 | 70,488 | 5.2 | 40.6 | 0 | 0 | | | | |
| SIDIAP | 193 | 251 | 0.2 | 0.3 | 55 | 87 | 0.95 (0.61 - 1.46) | 0.80 | 0.86 (0.55 - 1.34) | 0.52 |
| Meta-analysis | 26,522 | 62,742 | 18.0 | 45.5 | 478 | 2,255 | 1.05 (0.94 - 1.17) | 0.37 | 1.06 (0.89 - 1.27) | 0.52 |
| Meta-analysis (w/o USOC) | 26,522 | 62,742 | 18.0 | 45.5 | 478 | 2,255 | 1.05 (0.94 - 1.17) | 0.37 | 1.06 (0.95 - 1.19) | 0.36 |
| **PS-matching / on-treatment TAR without escalation** | | | | | | | | | | |
| CCAE | 15,937 | 34,825 | 10.1 | 21.4 | 16 | 36 | 0.80 (0.33 - 1.78) | 0.60 | 0.75 (0.33 - 1.74) | 0.51 |
| DAG | 1,045 | 274 | 0.7 | 0.2 | 79 | 29 | 0.72 (0.40 - 1.35) | 0.29 | 0.78 (0.43 - 1.44) | 0.37 |
| ODOD | 7,412 | 26,878 | 3.8 | 17.8 | 373 | 1,953 | 0.89 (0.76 - 1.04) | 0.15 | 0.90 (0.77 - 1.06) | 0.26 |
| MDCR | 1,327 | 6,736 | 0.8 | 5.0 | 0 | 9 | | | | |
| MDCD | 2,388 | 11,679 | 1.1 | 6.1 | 0 | 0 | | | | |
| Open Claims | 261,894 | 832,892 | 164.9 | 664.4 | 0 | 0 | | | | |
| OEHR | 15,494 | 64,240 | 4.2 | 31.2 | 0 | 0 | | | | |
| SIDIAP | 95 | 222 | 0.1 | 0.2 | 26 | 63 | 0.70 (0.37 - 1.28) | 0.27 | 0.90 (0.48 - 1.69) | 0.70 |
| Meta-analysis | 23,444 | 61,925 | 14.0 | 39.4 | 415 | 2,052 | 0.88 (0.75 - 1.02) | 0.09 | 0.88 (0.71 - 1.08) | 0.22 |
| Meta-analysis (w/o USOC) | 23,444 | 61,925 | 14.0 | 39.4 | 415 | 2,052 | 0.88 (0.75 - 1.02) | 0.09 | 0.86 (0.73 - 1.00) | 0.07 |
| **PS-stratification / on-treatment TAR without escalation** | | | | | | | | | | |
| CCAE | 18,683 | 35,323 | 11.7 | 21.6 | 18 | 43 | 1.10 (0.59 - 1.98) | 0.75 | 1.05 (0.58 - 1.92) | 0.87 |
| DAG | 1,071 | 535 | 0.7 | 0.5 | 82 | 62 | 0.73 (0.45 - 1.17) | 0.19 | 0.47 (0.29 - 0.75) | <0.01 |
| ODOD | 7,839 | 27,419 | 3.9 | 18.2 | 397 | 2,016 | 1.01 (0.89 - 1.13) | 0.90 | 1.04 (0.92 - 1.18) | 0.53 |
| MDCR | 1,482 | 7,185 | 0.8 | 5.3 | 0 | 10 | | | | |
| MDCD | 2,435 | 11,946 | 1.1 | 6.2 | 0 | 0 | | | | |
| Open Claims | 276,521 | 842,971 | 172.3 | 672.6 | 0 | 0 | | | | |
| OEHR | 16,666 | 70,488 | 4.6 | 34.4 | 0 | 0 | | | | |
| SIDIAP | 193 | 251 | 0.2 | 0.3 | 48 | 75 | 0.90 (0.55 - 1.45) | 0.66 | 0.91 (0.56 - 1.48) | 0.70 |
| Meta-analysis | 26,522 | 62,742 | 15.6 | 39.7 | 415 | 2,059 | 1.01 (0.90 - 1.13) | 0.86 | 1.01 (0.86 - 1.19) | 0.88 |
| Meta-analysis (w/o USOC) | 26,522 | 62,742 | 15.6 | 39.7 | 415 | 2,059 | 1.01 (0.90 - 1.13) | 0.86 | 1.00 (0.89 - 1.13) | 0.87 |

##### 1.3.6 Hospitalization with heart failure

**Supplementary Table 58:** Exposures, outcomes and risk estimates for Hospitalization with heart failure in SGLT2I (female) (T) vs SU (female) (C) new-user cohorts under four different propensity score (PS) adjustment / time-at-risk (TAR) designs across data sources. We report total patient exposure time in 1,000 patient-year (KYr) units, uncalibrated and calibrated hazard ratios (HRs) and their 95% confidence intervals (CIs). Greyed out entries did not pass study diagnostics and are included only for completeness.

| | Patients | | Exposure (KYr) | | Outcomes | | Uncalibrated | | Calibrated | |
|---|---|---|---|---|---|---|---|---|---|---|
| | T | C | T | C | T | C | HR (95% CI) | $p$ | HR (95% CI) | $p$ |
| **PS-matching / on-treatment TAR** | | | | | | | | | | |
| CCAE | 15,826 | 34,412 | 11.7 | 24.6 | 42 | 148 | 0.93 (0.58 - 1.44) | 0.74 | 0.88 (0.56 - 1.39) | 0.60 |
| DAG | 3,180 | 682 | 2.5 | 0.7 | 0 | 0 | | | | |
| ODOD | 11,278 | 38,129 | 7.1 | 31.5 | 75 | 836 | 0.64 (0.48 - 0.86) | <0.01 | 0.67 (0.49 - 0.90) | <0.01 |
| MDCR | 1,252 | 6,184 | 0.8 | 4.9 | 14 | 152 | 0.56 (0.24 - 1.16) | 0.15 | 0.58 (0.26 - 1.28) | 0.18 |
| MDCD | 2,226 | 10,744 | 1.2 | 6.7 | 20 | 146 | 0.77 (0.40 - 1.39) | 0.42 | 0.71 (0.38 - 1.32) | 0.28 |
| Open Claims | 252,612 | 797,448 | 190.5 | 750.2 | 903 | 9,400 | 0.62 (0.57 - 0.68) | <0.01 | 0.64 (0.53 - 0.77) | <0.01 |
| OEHR | 15,240 | 62,764 | 4.8 | 35.5 | 35 | 552 | 0.80 (0.52 - 1.20) | 0.31 | 0.84 (0.54 - 1.29) | 0.48 |
| SIDIAP | 2,824 | 963 | 3.9 | 1.7 | <5 | <5 | | | | |
| Meta-analysis | 298,434 | 949,681 | 216.1 | 853.4 | 1,089 | 11,234 | 0.66 (0.58 - 0.75) | <0.01 | 0.66 (0.54 - 0.81) | <0.01 |
| Meta-analysis (w/o USOC) | 45,822 | 152,233 | 25.6 | 103.2 | 186 | 1,834 | 0.73 (0.60 - 0.89) | <0.01 | 0.72 (0.59 - 0.87) | <0.01 |
| **PS-stratification / on-treatment TAR** | | | | | | | | | | |
| CCAE | 18,562 | 35,113 | 13.5 | 24.9 | 47 | 153 | 0.72 (0.50 - 1.02) | 0.07 | 0.70 (0.49 - 1.00) | 0.05 |
| DAG | 3,231 | 1,228 | 2.5 | 1.2 | 0 | 0 | | | | |
| ODOD | 12,165 | 39,032 | 7.5 | 32.4 | 82 | 895 | 0.60 (0.47 - 0.76) | <0.01 | 0.63 (0.49 - 0.80) | <0.01 |
| MDCR | 1,395 | 6,557 | 0.9 | 5.2 | 14 | 169 | 0.69 (0.36 - 1.23) | 0.23 | 0.68 (0.37 - 1.25) | 0.23 |
| MDCD | 2,266 | 10,936 | 1.2 | 6.8 | 20 | 147 | 0.84 (0.49 - 1.35) | 0.48 | 0.77 (0.47 - 1.27) | 0.31 |
| Open Claims | 265,019 | 806,578 | 197.8 | 758.7 | 960 | 9,828 | 0.59 (0.55 - 0.63) | <0.01 | 0.60 (0.51 - 0.72) | <0.01 |
| OEHR | 16,387 | 68,880 | 5.1 | 39.1 | 36 | 646 | 0.74 (0.51 - 1.05) | 0.11 | 0.77 (0.54 - 1.11) | 0.19 |
| SIDIAP | 2,923 | 2,011 | 4.0 | 4.3 | <5 | <5 | 0.95 (0.06 - 23.8) | 0.98 | 0.87 (0.04 - 16.9) | 0.92 |
| Meta-analysis | 312,133 | 949,603 | 223.9 | 855.1 | 1,125 | 11,522 | 0.60 (0.56 - 0.64) | <0.01 | 0.60 (0.52 - 0.71) | <0.01 |
| Meta-analysis (w/o USOC) | 47,114 | 143,025 | 26.1 | 96.3 | 165 | 1,694 | 0.66 (0.55 - 0.79) | <0.01 | 0.67 (0.56 - 0.80) | <0.01 |
| **PS-matching / on-treatment TAR without escalation** | | | | | | | | | | |
| CCAE | 15,826 | 34,412 | 10.1 | 21.1 | 37 | 130 | 0.77 (0.47 - 1.24) | 0.30 | 0.73 (0.45 - 1.18) | 0.21 |
| DAG | 3,180 | 682 | 2.3 | 0.6 | 0 | 0 | | | | |
| ODOD | 11,278 | 38,129 | 6.3 | 28.2 | 63 | 735 | 0.60 (0.43 - 0.81) | <0.01 | 0.60 (0.44 - 0.83) | <0.01 |
| MDCR | 1,252 | 6,184 | 0.7 | 4.5 | 13 | 141 | 0.59 (0.25 - 1.24) | 0.20 | 0.60 (0.27 - 1.34) | 0.22 |
| MDCD | 2,226 | 10,744 | 1.0 | 5.5 | 14 | 111 | 0.80 (0.39 - 1.52) | 0.52 | 0.76 (0.39 - 1.50) | 0.44 |
| Open Claims | 252,612 | 797,448 | 159.9 | 632.3 | 742 | 7,921 | 0.61 (0.55 - 0.67) | <0.01 | 0.62 (0.52 - 0.75) | <0.01 |
| OEHR | 15,240 | 62,764 | 4.2 | 30.3 | 28 | 421 | 0.74 (0.45 - 1.17) | 0.22 | 0.77 (0.48 - 1.24) | 0.30 |
| SIDIAP | 2,824 | 963 | 3.5 | 1.4 | <5 | <5 | | | | |
| Meta-analysis | 298,434 | 949,681 | 182.1 | 721.9 | 897 | 9,459 | 0.62 (0.57 - 0.68) | <0.01 | 0.62 (0.52 - 0.73) | <0.01 |
| Meta-analysis (w/o USOC) | 45,822 | 152,233 | 22.2 | 89.6 | 155 | 1,538 | 0.67 (0.55 - 0.83) | <0.01 | 0.66 (0.53 - 0.82) | <0.01 |
| **PS-stratification / on-treatment TAR without escalation** | | | | | | | | | | |
| CCAE | 18,562 | 35,113 | 11.6 | 21.4 | 42 | 134 | 0.70 (0.48 - 1.02) | 0.07 | 0.67 (0.46 - 0.98) | 0.04 |
| DAG | 3,231 | 1,228 | 2.3 | 1.2 | 0 | 0 | | | | |
| ODOD | 12,165 | 39,032 | 6.7 | 29.0 | 70 | 785 | 0.58 (0.44 - 0.75) | <0.01 | 0.60 (0.46 - 0.78) | <0.01 |
| MDCR | 1,395 | 6,557 | 0.8 | 4.8 | 13 | 158 | 0.70 (0.36 - 1.28) | 0.28 | 0.65 (0.34 - 1.24) | 0.20 |
| MDCD | 2,266 | 10,936 | 1.0 | 5.6 | 14 | 112 | 0.71 (0.38 - 1.23) | 0.25 | 0.64 (0.35 - 1.16) | 0.14 |
| Open Claims | 265,019 | 806,578 | 166.2 | 639.7 | 794 | 8,291 | 0.58 (0.53 - 0.62) | <0.01 | 0.59 (0.50 - 0.69) | <0.01 |
| OEHR | 16,387 | 68,880 | 4.5 | 33.3 | 29 | 487 | 0.73 (0.48 - 1.06) | 0.12 | 0.73 (0.49 - 1.10) | 0.16 |
| SIDIAP | 2,923 | 2,011 | 3.5 | 3.6 | <5 | <5 | 1.04 (0.06 - 25.8) | 0.98 | 1.05 (0.05 - 21.1) | 0.96 |
| Meta-analysis | 312,133 | 949,603 | 189.0 | 723.4 | 935 | 9,697 | 0.59 (0.54 - 0.63) | <0.01 | 0.59 (0.51 - 0.67) | <0.01 |
| Meta-analysis (w/o USOC) | 47,114 | 143,025 | 22.8 | 83.7 | 141 | 1,406 | 0.64 (0.53 - 0.77) | <0.01 | 0.63 (0.52 - 0.77) | <0.01 |

##### 1.3.7 Measured renal dysfunction

**Supplementary Table 59:** Exposures, outcomes and risk estimates for Measured renal dysfunction in SGLT2I (female) (T) vs SU (female) (C) new-user cohorts under four different propensity score (PS) adjustment / time-at-risk (TAR) designs across data sources. We report total patient exposure time in 1,000 patient-year (KYr) units, uncalibrated and calibrated hazard ratios (HRs) and their 95% confidence intervals (CIs). Greyed out entries did not pass study diagnostics and are included only for completeness.

| | Patients | | Exposure (KYr) | | Outcomes | | Uncalibrated | | Calibrated | |
|---|---|---|---|---|---|---|---|---|---|---|
| | T | C | T | C | T | C | HR (95% CI) | $p$ | HR (95% CI) | $p$ |
| **PS-matching / on-treatment TAR** | | | | | | | | | | |
| CCAE | 16,054 | 35,135 | 11.8 | 25.1 | <5 | 13 | 0.38 (0.02 - 2.95) | 0.45 | 0.36 (0.03 - 4.56) | 0.43 |
| DAG | 3,180 | 682 | 2.5 | 0.7 | 0 | 0 | | | | |
| ODOD | 11,834 | 40,860 | 7.4 | 34.2 | 10 | 56 | 1.38 (0.41 - 4.04) | 0.58 | 1.42 (0.45 - 4.48) | 0.55 |
| MDCR | 1,333 | 6,800 | 0.9 | 5.5 | 0 | <5 | | | | |
| MDCD | 2,388 | 11,679 | 1.3 | 7.3 | 0 | 0 | | | | |
| Open Claims | 261,894 | 832,892 | 196.5 | 789.1 | 0 | 0 | | | | |
| OEHR | 15,494 | 64,240 | 4.9 | 36.8 | 0 | 0 | | | | |
| SIDIAP | 2,843 | 962 | 4.0 | 1.7 | 0 | <5 | | | | |
| Meta-analysis | 11,834 | 40,860 | 7.4 | 34.2 | 10 | 56 | 1.38 (0.41 - 4.04) | 0.58 | 1.38 (0.43 - 4.38) | 0.59 |
| Meta-analysis (w/o USOC) | 11,834 | 40,860 | 7.4 | 34.2 | 10 | 56 | 1.38 (0.41 - 4.04) | 0.58 | 1.36 (0.43 - 4.27) | 0.60 |
| **PS-stratification / on-treatment TAR** | | | | | | | | | | |
| CCAE | 18,849 | 35,766 | 13.7 | 25.3 | <5 | 13 | 0.24 (0.04 - 1.05) | 0.10 | 0.24 (0.04 - 1.30) | 0.10 |
| DAG | 3,231 | 1,228 | 2.5 | 1.2 | 0 | 0 | | | | |
| ODOD | 12,834 | 41,754 | 7.9 | 35.0 | 10 | 64 | 1.39 (0.63 - 2.84) | 0.39 | 1.46 (0.68 - 3.11) | 0.33 |
| MDCR | 1,489 | 7,293 | 0.9 | 5.9 | 0 | <5 | | | | |
| MDCD | 2,435 | 11,946 | 1.3 | 7.5 | 0 | 0 | | | | |
| Open Claims | 276,521 | 842,971 | 205.0 | 798.5 | 0 | 0 | | | | |
| OEHR | 16,666 | 70,488 | 5.2 | 40.6 | 0 | 0 | | | | |
| SIDIAP | 2,943 | 2,006 | 4.0 | 4.3 | 0 | 5 | | | | |
| Meta-analysis | 12,834 | 41,754 | 7.9 | 35.0 | 10 | 64 | 1.39 (0.63 - 2.84) | 0.39 | 1.40 (0.65 - 3.03) | 0.39 |
| Meta-analysis (w/o USOC) | 12,834 | 41,754 | 7.9 | 35.0 | 10 | 64 | 1.39 (0.63 - 2.84) | 0.39 | 1.41 (0.66 - 3.00) | 0.38 |
| **PS-matching / on-treatment TAR without escalation** | | | | | | | | | | |
| CCAE | 16,054 | 35,135 | 10.2 | 21.6 | <5 | 12 | 0.49 (0.02 - 4.78) | 0.60 | 0.46 (0.03 - 6.66) | 0.57 |
| DAG | 3,180 | 682 | 2.3 | 0.6 | 0 | 0 | | | | |
| ODOD | 11,834 | 40,860 | 6.5 | 30.5 | 8 | 47 | 1.36 (0.35 - 4.45) | 0.64 | 1.38 (0.38 - 4.95) | 0.62 |
| MDCR | 1,333 | 6,800 | 0.8 | 5.0 | 0 | <5 | | | | |
| MDCD | 2,388 | 11,679 | 1.1 | 6.1 | 0 | 0 | | | | |
| Open Claims | 261,894 | 832,892 | 164.9 | 664.4 | 0 | 0 | | | | |
| OEHR | 15,494 | 64,240 | 4.2 | 31.2 | 0 | 0 | | | | |
| SIDIAP | 2,843 | 962 | 3.5 | 1.4 | 0 | <5 | | | | |
| Meta-analysis | 11,834 | 40,860 | 6.5 | 30.5 | 8 | 47 | 1.36 (0.35 - 4.45) | 0.64 | 1.36 (0.38 - 4.94) | 0.64 |
| Meta-analysis (w/o USOC) | 11,834 | 40,860 | 6.5 | 30.5 | 8 | 47 | 1.36 (0.35 - 4.45) | 0.64 | 1.33 (0.37 - 4.79) | 0.66 |
| **PS-stratification / on-treatment TAR without escalation** | | | | | | | | | | |
| CCAE | 18,849 | 35,766 | 11.8 | 21.8 | <5 | 12 | 0.24 (0.03 - 1.06) | 0.11 | 0.23 (0.04 - 1.28) | 0.09 |
| DAG | 3,231 | 1,228 | 2.3 | 1.2 | 0 | 0 | | | | |
| ODOD | 12,834 | 41,754 | 7.0 | 31.3 | 8 | 55 | 1.33 (0.55 - 2.93) | 0.50 | 1.38 (0.60 - 3.21) | 0.45 |
| MDCR | 1,489 | 7,293 | 0.8 | 5.4 | 0 | <5 | | | | |
| MDCD | 2,435 | 11,946 | 1.1 | 6.2 | 0 | 0 | | | | |
| Open Claims | 276,521 | 842,971 | 172.3 | 672.6 | 0 | 0 | | | | |
| OEHR | 16,666 | 70,488 | 4.6 | 34.4 | 0 | 0 | | | | |
| SIDIAP | 2,943 | 2,006 | 3.5 | 3.6 | 0 | 5 | | | | |
| Meta-analysis | 12,834 | 41,754 | 7.0 | 31.3 | 8 | 55 | 1.33 (0.55 - 2.93) | 0.50 | 1.34 (0.57 - 3.12) | 0.50 |
| Meta-analysis (w/o USOC) | 12,834 | 41,754 | 7.0 | 31.3 | 8 | 55 | 1.33 (0.55 - 2.93) | 0.50 | 1.32 (0.57 - 3.06) | 0.52 |

##### 1.3.8 Stroke

**Supplementary Table 60:** Exposures, outcomes and risk estimates for Stroke in SGLT2I (female) (T) vs SU (female) (C) new-user cohorts under four different propensity score (PS) adjustment / time-at-risk (TAR) designs across data sources. We report total patient exposure time in 1,000 patient-year (KYr) units, uncalibrated and calibrated hazard ratios (HRs) and their 95% confidence intervals (CIs). Greyed out entries did not pass study diagnostics and are included only for completeness.

| | Patients | | Exposure (KYr) | | Outcomes | | Uncalibrated | | Calibrated | |
|---|---|---|---|---|---|---|---|---|---|---|
| | T | C | T | C | T | C | HR (95% CI) | $p$ | HR (95% CI) | $p$ |
| **PS-matching / on-treatment TAR** | | | | | | | | | | |
| CCAE | 15,885 | 34,645 | 11.7 | 24.7 | 43 | 108 | 0.83 (0.49 - 1.36) | 0.47 | 0.79 (0.48 - 1.32) | 0.38 |
| DAG | 3,180 | 682 | 2.5 | 0.7 | 0 | 0 | | | | |
| ODOD | 11,589 | 39,553 | 7.2 | 32.9 | 48 | 409 | 1.13 (0.76 - 1.64) | 0.52 | 1.17 (0.80 - 1.72) | 0.44 |
| MDCR | 1,300 | 6,486 | 0.9 | 5.2 | <5 | 93 | 0.31 (0.09 - 0.81) | 0.04 | 0.32 (0.10 - 0.96) | 0.04 |
| MDCD | 2,298 | 11,202 | 1.2 | 7.0 | 12 | 87 | 0.92 (0.41 - 1.89) | 0.83 | 0.84 (0.40 - 1.80) | 0.66 |
| Open Claims | 255,962 | 809,471 | 192.4 | 764.9 | 577 | 5,068 | 0.68 (0.61 - 0.76) | <0.01 | 0.70 (0.57 - 0.85) | <0.01 |
| OEHR | 15,375 | 63,445 | 4.8 | 36.2 | 14 | 209 | 1.04 (0.50 - 1.98) | 0.92 | 1.08 (0.54 - 2.17) | 0.84 |
| SIDIAP | 2,830 | 956 | 3.9 | 1.7 | 10 | <5 | 2.34 (0.54 - 16.1) | 0.33 | 2.74 (0.50 - 15.0) | 0.25 |
| Meta-analysis | 302,409 | 964,802 | 218.3 | 870.9 | <699 | 5,974 | 0.81 (0.63 - 1.04) | 0.10 | 0.81 (0.60 - 1.08) | 0.16 |
| Meta-analysis (w/o USOC) | 46,447 | 155,331 | 25.9 | 106.0 | <122 | 906 | 0.91 (0.67 - 1.24) | 0.56 | 0.90 (0.66 - 1.22) | 0.50 |
| **PS-stratification / on-treatment TAR** | | | | | | | | | | |
| CCAE | 18,673 | 35,257 | 13.5 | 24.9 | 51 | 109 | 0.96 (0.66 - 1.38) | 0.82 | 0.93 (0.65 - 1.35) | 0.72 |
| DAG | 3,231 | 1,228 | 2.5 | 1.2 | 0 | 0 | | | | |
| ODOD | 12,563 | 40,405 | 7.7 | 33.7 | 53 | 429 | 0.95 (0.69 - 1.29) | 0.74 | 0.99 (0.72 - 1.36) | 0.93 |
| MDCR | 1,453 | 6,939 | 0.9 | 5.6 | 5 | 102 | 0.32 (0.11 - 0.76) | 0.02 | 0.31 (0.12 - 0.83) | 0.02 |
| MDCD | 2,344 | 11,432 | 1.3 | 7.2 | 12 | 87 | 0.82 (0.41 - 1.49) | 0.54 | 0.75 (0.39 - 1.43) | 0.39 |
| Open Claims | 270,188 | 818,848 | 200.6 | 773.4 | 602 | 5,250 | 0.65 (0.60 - 0.71) | <0.01 | 0.67 (0.56 - 0.80) | <0.01 |
| OEHR | 16,552 | 69,698 | 5.2 | 40.0 | 14 | 251 | 0.79 (0.42 - 1.36) | 0.42 | 0.82 (0.46 - 1.47) | 0.52 |
| SIDIAP | 2,926 | 1,998 | 4.0 | 4.3 | 10 | 6 | 1.87 (0.57 - 6.46) | 0.31 | 1.70 (0.50 - 5.70) | 0.40 |
| Meta-analysis | 317,976 | 964,208 | 227.1 | 872.1 | 720 | 6,039 | 0.80 (0.63 - 1.01) | 0.06 | 0.80 (0.61 - 1.06) | 0.12 |
| Meta-analysis (w/o USOC) | 47,788 | 145,360 | 26.4 | 98.6 | 118 | 789 | 0.93 (0.74 - 1.16) | 0.50 | 0.94 (0.75 - 1.17) | 0.58 |
| **PS-matching / on-treatment TAR without escalation** | | | | | | | | | | |
| CCAE | 15,885 | 34,645 | 10.1 | 21.2 | 34 | 92 | 0.80 (0.46 - 1.35) | 0.42 | 0.76 (0.44 - 1.30) | 0.31 |
| DAG | 3,180 | 682 | 2.3 | 0.6 | 0 | 0 | | | | |
| ODOD | 11,589 | 39,553 | 6.4 | 29.4 | 45 | 361 | 1.20 (0.80 - 1.78) | 0.36 | 1.22 (0.82 - 1.82) | 0.34 |
| MDCR | 1,300 | 6,486 | 0.8 | 4.8 | <5 | 85 | 0.38 (0.11 - 1.02) | 0.09 | 0.39 (0.13 - 1.19) | 0.10 |
| MDCD | 2,298 | 11,202 | 1.0 | 5.8 | 6 | 75 | 0.53 (0.17 - 1.34) | 0.22 | 0.50 (0.18 - 1.41) | 0.19 |
| Open Claims | 255,962 | 809,471 | 161.5 | 644.2 | 482 | 4,307 | 0.68 (0.60 - 0.76) | <0.01 | 0.69 (0.57 - 0.85) | <0.01 |
| OEHR | 15,375 | 63,445 | 4.2 | 30.7 | 9 | 167 | 0.85 (0.36 - 1.76) | 0.69 | 0.88 (0.40 - 1.94) | 0.75 |
| SIDIAP | 2,830 | 956 | 3.4 | 1.4 | 10 | <5 | 2.42 (0.56 - 16.6) | 0.31 | 3.11 (0.57 - 16.9) | 0.19 |
| Meta-analysis | 302,409 | 964,802 | 184.0 | 736.1 | <581 | 5,087 | 0.77 (0.58 - 1.02) | 0.07 | 0.78 (0.57 - 1.06) | 0.11 |
| Meta-analysis (w/o USOC) | 46,447 | 155,331 | 22.5 | 91.9 | <99 | 780 | 0.85 (0.59 - 1.21) | 0.36 | 0.83 (0.58 - 1.19) | 0.31 |
| **PS-stratification / on-treatment TAR without escalation** | | | | | | | | | | |
| CCAE | 18,673 | 35,257 | 11.7 | 21.4 | 41 | 93 | 0.88 (0.58 - 1.31) | 0.53 | 0.84 (0.56 - 1.26) | 0.40 |
| DAG | 3,231 | 1,228 | 2.3 | 1.2 | 0 | 0 | | | | |
| ODOD | 12,563 | 40,405 | 6.8 | 30.1 | 50 | 380 | 0.99 (0.71 - 1.36) | 0.94 | 1.02 (0.74 - 1.42) | 0.89 |
| MDCR | 1,453 | 6,939 | 0.8 | 5.1 | <5 | 93 | 0.29 (0.08 - 0.74) | 0.02 | 0.27 (0.09 - 0.79) | 0.02 |
| MDCD | 2,344 | 11,432 | 1.0 | 5.9 | 6 | 75 | 0.49 (0.18 - 1.07) | 0.11 | 0.44 (0.18 - 1.06) | 0.07 |
| Open Claims | 270,188 | 818,848 | 168.7 | 651.7 | 505 | 4,463 | 0.65 (0.59 - 0.72) | <0.01 | 0.66 (0.56 - 0.78) | <0.01 |
| OEHR | 16,552 | 69,698 | 4.5 | 33.9 | 9 | 194 | 0.61 (0.28 - 1.17) | 0.17 | 0.61 (0.30 - 1.26) | 0.19 |
| SIDIAP | 2,926 | 1,998 | 3.5 | 3.5 | 10 | 6 | 1.87 (0.57 - 6.46) | 0.31 | 1.90 (0.56 - 6.38) | 0.31 |
| Meta-analysis | 317,976 | 964,208 | 191.7 | 737.2 | 605 | 5,130 | 0.77 (0.60 - 0.98) | 0.03 | 0.77 (0.58 - 1.01) | 0.05 |
| Meta-analysis (w/o USOC) | 47,788 | 145,360 | 23.0 | 85.4 | 100 | 667 | 0.90 (0.71 - 1.14) | 0.38 | 0.89 (0.70 - 1.13) | 0.35 |

##### 1.3.9 Sudden cardiac death

**Supplementary Table 61:** Exposures, outcomes and risk estimates for Sudden cardiac death in SGLT2I (female) (T) vs SU (female) (C) new-user cohorts under four different propensity score (PS) adjustment / time-at-risk (TAR) designs across data sources. We report total patient exposure time in 1,000 patient-year (KYr) units, uncalibrated and calibrated hazard ratios (HRs) and their 95% confidence intervals (CIs). Greyed out entries did not pass study diagnostics and are included only for completeness.

| | Patients | | Exposure (KYr) | | Outcomes | | Uncalibrated | | Calibrated | |
|---|---|---|---|---|---|---|---|---|---|---|
| | T | C | T | C | T | C | HR (95% CI) | $p$ | HR (95% CI) | $p$ |
| **PS-matching / on-treatment TAR** | | | | | | | | | | |
| CCAE | 16,073 | 35,171 | 11.9 | 25.1 | 7 | 20 | 0.92 (0.24 - 3.14) | 0.90 | 0.88 (0.24 - 3.22) | 0.85 |
| DAG | 3,180 | 682 | 2.5 | 0.7 | 0 | 0 | | | | |
| ODOD | 11,832 | 40,927 | 7.4 | 34.3 | 6 | 118 | 0.77 (0.28 - 1.81) | 0.59 | 0.80 (0.31 - 2.03) | 0.63 |
| MDCR | 1,332 | 6,788 | 0.9 | 5.5 | 0 | 17 | | | | |
| MDCD | 2,386 | 11,636 | 1.3 | 7.3 | <5 | 20 | 1.33 (0.07 - 8.10) | 0.81 | 1.22 (0.11 - 13.2) | 0.87 |
| Open Claims | 261,446 | 831,503 | 196.2 | 787.7 | 115 | 1,013 | 0.63 (0.48 - 0.80) | <0.01 | 0.64 (0.47 - 0.87) | <0.01 |
| OEHR | 15,487 | 64,209 | 4.9 | 36.8 | <5 | 52 | 1.02 (0.23 - 3.28) | 0.98 | 1.06 (0.28 - 4.04) | 0.93 |
| SIDIAP | 2,849 | 965 | 4.0 | 1.7 | 0 | 0 | | | | |
| Meta-analysis | 304,838 | 971,810 | 220.3 | 883.9 | <133 | 1,203 | 0.65 (0.51 - 0.83) | <0.01 | 0.65 (0.49 - 0.86) | <0.01 |
| Meta-analysis (w/o USOC) | 43,392 | 140,307 | 24.1 | 96.2 | <18 | 190 | 0.87 (0.45 - 1.67) | 0.67 | 0.85 (0.44 - 1.65) | 0.63 |
| **PS-stratification / on-treatment TAR** | | | | | | | | | | |
| CCAE | 18,867 | 35,802 | 13.7 | 25.4 | 8 | 20 | 0.89 (0.35 - 2.12) | 0.80 | 0.87 (0.35 - 2.15) | 0.76 |
| DAG | 3,231 | 1,228 | 2.5 | 1.2 | 0 | 0 | | | | |
| ODOD | 12,828 | 41,824 | 7.9 | 35.1 | 6 | 127 | 0.41 (0.15 - 0.89) | 0.04 | 0.43 (0.18 - 1.02) | 0.06 |
| MDCR | 1,487 | 7,275 | 0.9 | 5.9 | 0 | 17 | | | | |
| MDCD | 2,433 | 11,899 | 1.3 | 7.5 | <5 | 20 | 1.28 (0.28 - 4.18) | 0.73 | 1.17 (0.30 - 4.55) | 0.82 |
| Open Claims | 276,028 | 841,576 | 204.6 | 797.0 | 121 | 1,054 | 0.63 (0.51 - 0.76) | <0.01 | 0.64 (0.50 - 0.83) | <0.01 |
| OEHR | 16,657 | 70,454 | 5.2 | 40.6 | <5 | 69 | 0.93 (0.27 - 2.46) | 0.90 | 0.97 (0.32 - 2.94) | 0.95 |
| SIDIAP | 2,949 | 2,013 | 4.0 | 4.3 | 0 | 0 | | | | |
| Meta-analysis | 324,380 | 989,656 | 231.4 | 898.1 | <140 | 1,270 | 0.63 (0.52 - 0.76) | <0.01 | 0.64 (0.50 - 0.80) | <0.01 |
| Meta-analysis (w/o USOC) | 48,352 | 148,080 | 26.8 | 101.1 | <19 | 216 | 0.66 (0.38 - 1.15) | 0.14 | 0.67 (0.39 - 1.16) | 0.15 |
| **PS-matching / on-treatment TAR without escalation** | | | | | | | | | | |
| CCAE | 16,073 | 35,171 | 10.2 | 21.6 | 6 | 17 | 0.82 (0.17 - 3.29) | 0.80 | 0.78 (0.18 - 3.44) | 0.74 |
| DAG | 3,180 | 682 | 2.3 | 0.6 | 0 | 0 | | | | |
| ODOD | 11,832 | 40,927 | 6.5 | 30.6 | 6 | 99 | 0.80 (0.29 - 1.90) | 0.64 | 0.81 (0.31 - 2.08) | 0.66 |
| MDCR | 1,332 | 6,788 | 0.8 | 5.0 | 0 | 17 | | | | |
| MDCD | 2,386 | 11,636 | 1.1 | 6.0 | <5 | 18 | 4.84 (0.22 - 47.3) | 0.25 | 4.61 (0.31 - 68.1) | 0.27 |
| Open Claims | 261,446 | 831,503 | 164.7 | 663.2 | 99 | 849 | 0.63 (0.47 - 0.82) | <0.01 | 0.64 (0.47 - 0.89) | <0.01 |
| OEHR | 15,487 | 64,209 | 4.2 | 31.2 | <5 | 34 | 1.32 (0.28 - 4.66) | 0.69 | 1.37 (0.34 - 5.55) | 0.66 |
| SIDIAP | 2,849 | 965 | 3.5 | 1.4 | 0 | 0 | | | | |
| Meta-analysis | 304,838 | 971,810 | 185.6 | 746.6 | <116 | 999 | 0.66 (0.51 - 0.86) | <0.01 | 0.66 (0.49 - 0.89) | <0.01 |
| Meta-analysis (w/o USOC) | 43,392 | 140,307 | 21.0 | 83.4 | <17 | 150 | 0.91 (0.45 - 1.82) | 0.79 | 0.89 (0.44 - 1.78) | 0.74 |
| **PS-stratification / on-treatment TAR without escalation** | | | | | | | | | | |
| CCAE | 18,867 | 35,802 | 11.8 | 21.8 | 7 | 17 | 0.80 (0.29 - 2.05) | 0.66 | 0.77 (0.29 - 2.05) | 0.60 |
| DAG | 3,231 | 1,228 | 2.3 | 1.2 | 0 | 0 | | | | |
| ODOD | 12,828 | 41,824 | 7.0 | 31.4 | 6 | 108 | 0.46 (0.17 - 1.02) | 0.09 | 0.48 (0.20 - 1.16) | 0.10 |
| MDCR | 1,487 | 7,275 | 0.8 | 5.4 | 0 | 17 | | | | |
| MDCD | 2,433 | 11,899 | 1.1 | 6.2 | <5 | 18 | 1.40 (0.30 - 4.68) | 0.63 | 1.27 (0.32 - 4.97) | 0.74 |
| Open Claims | 276,028 | 841,576 | 172.0 | 671.4 | 105 | 887 | 0.64 (0.52 - 0.80) | <0.01 | 0.66 (0.51 - 0.85) | <0.01 |
| OEHR | 16,657 | 70,454 | 4.5 | 34.4 | <5 | 45 | 1.17 (0.33 - 3.24) | 0.79 | 1.18 (0.37 - 3.71) | 0.78 |
| SIDIAP | 2,949 | 2,013 | 3.5 | 3.6 | 0 | 0 | | | | |
| Meta-analysis | 324,380 | 989,656 | 195.3 | 758.9 | <123 | 1,057 | 0.65 (0.53 - 0.80) | <0.01 | 0.65 (0.52 - 0.82) | <0.01 |
| Meta-analysis (w/o USOC) | 48,352 | 148,080 | 23.3 | 87.5 | <18 | 170 | 0.70 (0.40 - 1.24) | 0.22 | 0.69 (0.39 - 1.23) | 0.21 |

##### 1.3.10 Abnormal weight gain

**Supplementary Table 62:** Exposures, outcomes and risk estimates for Abnormal weight gain in SGLT2I (female) (T) vs SU (female) (C) new-user cohorts under four different propensity score (PS) adjustment / time-at-risk (TAR) designs across data sources. We report total patient exposure time in 1,000 patient-year (KYr) units, uncalibrated and calibrated hazard ratios (HRs) and their 95% confidence intervals (CIs). Greyed out entries did not pass study diagnostics and are included only for completeness.

| | Patients | | Exposure (KYr) | | Outcomes | | Uncalibrated | | Calibrated | |
|---|---|---|---|---|---|---|---|---|---|---|
| | T | C | T | C | T | C | HR (95% CI) | $p$ | HR (95% CI) | $p$ |
| **PS-matching / on-treatment TAR** | | | | | | | | | | |
| CCAE | 14,730 | 33,074 | 10.9 | 23.6 | 68 | 216 | 0.62 (0.43 - 0.90) | 0.01 | 0.60 (0.41 - 0.87) | <0.01 |
| DAG | 3,132 | 671 | 2.5 | 0.6 | <5 | <5 | 0.07 (0.00 - 0.57) | 0.04 | 0.08 (0.01 - 1.01) | 0.05 |
| ODOD | 11,025 | 39,200 | 6.8 | 32.7 | 49 | 283 | 0.61 (0.40 - 0.91) | 0.02 | 0.63 (0.42 - 0.95) | 0.03 |
| MDCR | 1,302 | 6,684 | 0.9 | 5.4 | <5 | 27 | 0.64 (0.09 - 2.92) | 0.61 | 0.66 (0.12 - 3.77) | 0.64 |
| MDCD | 2,135 | 10,801 | 1.2 | 6.8 | 5 | 84 | 0.08 (0.00 - 0.38) | 0.03 | 0.07 (0.01 - 0.67) | 0.02 |
| Open Claims | 242,662 | 794,057 | 181.8 | 752.7 | 448 | 2,374 | 0.78 (0.68 - 0.90) | <0.01 | 0.80 (0.64 - 1.00) | 0.05 |
| OEHR | 14,423 | 61,417 | 4.5 | 34.9 | 27 | 328 | 0.82 (0.47 - 1.36) | 0.46 | 0.85 (0.50 - 1.46) | 0.60 |
| SIDIAP | 2,818 | 956 | 3.9 | 1.7 | <5 | <5 | 0.64 (0.06 - 14.0) | 0.75 | 0.75 (0.05 - 11.5) | 0.84 |
| Meta-analysis | 286,277 | 945,233 | 205.9 | 856.0 | <602 | 3,312 | 0.73 (0.62 - 0.85) | <0.01 | 0.73 (0.59 - 0.90) | <0.01 |
| Meta-analysis (w/o USOC) | 43,615 | 151,176 | 24.1 | 103.4 | <154 | 938 | 0.64 (0.50 - 0.83) | <0.01 | 0.63 (0.49 - 0.82) | <0.01 |
| **PS-stratification / on-treatment TAR** | | | | | | | | | | |
| CCAE | 17,049 | 33,731 | 12.4 | 23.8 | 80 | 218 | 0.58 (0.44 - 0.77) | <0.01 | 0.57 (0.43 - 0.75) | <0.01 |
| DAG | 3,181 | 1,213 | 2.5 | 1.2 | <5 | <5 | 0.30 (0.06 - 1.57) | 0.14 | 0.19 (0.04 - 0.98) | 0.05 |
| ODOD | 11,835 | 40,059 | 7.2 | 33.5 | 54 | 299 | 0.68 (0.49 - 0.93) | 0.02 | 0.71 (0.51 - 0.98) | 0.04 |
| MDCR | 1,455 | 7,166 | 0.9 | 5.8 | <5 | 29 | 0.47 (0.07 - 1.81) | 0.36 | 0.46 (0.09 - 2.34) | 0.35 |
| MDCD | 2,177 | 11,027 | 1.2 | 7.0 | 5 | 86 | 0.29 (0.10 - 0.66) | <0.01 | 0.26 (0.10 - 0.68) | <0.01 |
| Open Claims | 255,345 | 804,381 | 188.9 | 762.3 | 486 | 2,422 | 0.75 (0.68 - 0.84) | <0.01 | 0.77 (0.64 - 0.93) | <0.01 |
| OEHR | 15,455 | 67,433 | 4.8 | 38.5 | 29 | 369 | 0.68 (0.45 - 1.01) | 0.07 | 0.71 (0.47 - 1.07) | 0.12 |
| SIDIAP | 2,919 | 1,994 | 4.0 | 4.3 | <5 | 6 | 0.29 (0.03 - 2.30) | 0.26 | 0.27 (0.03 - 2.26) | 0.23 |
| Meta-analysis | 299,684 | 945,604 | 213.3 | 858.1 | 649 | 3,308 | 0.70 (0.61 - 0.80) | <0.01 | 0.71 (0.58 - 0.86) | <0.01 |
| Meta-analysis (w/o USOC) | 44,339 | 141,223 | 24.4 | 95.9 | 163 | 886 | 0.63 (0.53 - 0.76) | <0.01 | 0.64 (0.53 - 0.78) | <0.01 |
| **PS-matching / on-treatment TAR without escalation** | | | | | | | | | | |
| CCAE | 14,730 | 33,074 | 9.3 | 20.3 | 56 | 176 | 0.66 (0.44 - 0.97) | 0.04 | 0.62 (0.42 - 0.92) | 0.02 |
| DAG | 3,132 | 671 | 2.2 | 0.6 | <5 | <5 | 0.09 (0.00 - 0.68) | 0.06 | 0.09 (0.01 - 1.18) | 0.07 |
| ODOD | 11,025 | 39,200 | 6.0 | 29.2 | 44 | 242 | 0.60 (0.38 - 0.92) | 0.02 | 0.61 (0.39 - 0.94) | 0.03 |
| MDCR | 1,302 | 6,684 | 0.8 | 4.9 | <5 | 21 | 1.26 (0.16 - 7.21) | 0.81 | 1.29 (0.19 - 8.65) | 0.80 |
| MDCD | 2,135 | 10,801 | 1.0 | 5.6 | <5 | 70 | 0.08 (0.00 - 0.39) | 0.03 | 0.08 (0.01 - 0.72) | 0.02 |
| Open Claims | 242,662 | 794,057 | 152.7 | 635.2 | 357 | 1,960 | 0.76 (0.65 - 0.88) | <0.01 | 0.77 (0.62 - 0.96) | 0.02 |
| OEHR | 14,423 | 61,417 | 3.9 | 29.7 | 22 | 259 | 0.80 (0.44 - 1.38) | 0.43 | 0.82 (0.46 - 1.46) | 0.52 |
| SIDIAP | 2,818 | 956 | 3.4 | 1.4 | <5 | <5 | 0.64 (0.06 - 14.0) | 0.75 | 0.82 (0.05 - 12.6) | 0.89 |
| Meta-analysis | 284,975 | 938,549 | 172.9 | 720.0 | <484 | 2,707 | 0.73 (0.64 - 0.83) | <0.01 | 0.73 (0.60 - 0.88) | <0.01 |
| Meta-analysis (w/o USOC) | 42,313 | 144,492 | 20.2 | 84.9 | <127 | 747 | 0.64 (0.47 - 0.88) | <0.01 | 0.63 (0.45 - 0.86) | <0.01 |
| **PS-stratification / on-treatment TAR without escalation** | | | | | | | | | | |
| CCAE | 17,049 | 33,731 | 10.7 | 20.6 | 68 | 178 | 0.59 (0.43 - 0.79) | <0.01 | 0.56 (0.41 - 0.76) | <0.01 |
| DAG | 3,181 | 1,213 | 2.3 | 1.1 | <5 | <5 | 0.21 (0.04 - 1.21) | 0.08 | 0.13 (0.02 - 0.78) | 0.03 |
| ODOD | 11,835 | 40,059 | 6.4 | 30.0 | 48 | 257 | 0.69 (0.49 - 0.97) | 0.04 | 0.72 (0.51 - 1.01) | 0.06 |
| MDCR | 1,455 | 7,166 | 0.8 | 5.3 | <5 | 23 | 0.52 (0.08 - 2.09) | 0.44 | 0.49 (0.09 - 2.53) | 0.39 |
| MDCD | 2,177 | 11,027 | 1.0 | 5.8 | <5 | 72 | 0.27 (0.08 - 0.67) | 0.02 | 0.24 (0.08 - 0.70) | <0.01 |
| Open Claims | 255,345 | 804,381 | 159.0 | 643.5 | 388 | 2,004 | 0.72 (0.64 - 0.81) | <0.01 | 0.73 (0.61 - 0.88) | <0.01 |
| OEHR | 15,455 | 67,433 | 4.2 | 32.7 | 24 | 290 | 0.66 (0.41 - 1.02) | 0.07 | 0.67 (0.42 - 1.05) | 0.10 |
| SIDIAP | 2,919 | 1,994 | 3.5 | 3.5 | <5 | 6 | 0.30 (0.03 - 2.32) | 0.27 | 0.30 (0.04 - 2.56) | 0.28 |
| Meta-analysis | 299,684 | 945,604 | 180.2 | 726.8 | 528 | 2,729 | 0.70 (0.63 - 0.77) | <0.01 | 0.70 (0.60 - 0.81) | <0.01 |
| Meta-analysis (w/o USOC) | 44,339 | 141,223 | 21.2 | 83.3 | 140 | 725 | 0.64 (0.52 - 0.78) | <0.01 | 0.63 (0.51 - 0.78) | <0.01 |

##### 1.3.11 Abnormal weight loss

**Supplementary Table 63:** Exposures, outcomes and risk estimates for Abnormal weight loss in SGLT2I (female) (T) vs SU (female) (C) new-user cohorts under four different propensity score (PS) adjustment / time-at-risk (TAR) designs across data sources. We report total patient exposure time in 1,000 patient-year (KYr) units, uncalibrated and calibrated hazard ratios (HRs) and their 95% confidence intervals (CIs). Greyed out entries did not pass study diagnostics and are included only for completeness.

| | Patients | | Exposure (KYr) | | Outcomes | | Uncalibrated | | Calibrated | |
|---|---|---|---|---|---|---|---|---|---|---|
| | T | C | T | C | T | C | HR (95% CI) | $p$ | HR (95% CI) | $p$ |
| **PS-matching / on-treatment TAR** | | | | | | | | | | |
| CCAE | 15,708 | 34,218 | 11.5 | 24.4 | 93 | 147 | 1.08 (0.73 - 1.59) | 0.69 | 1.03 (0.70 - 1.53) | 0.87 |
| DAG | 3,100 | 659 | 2.4 | 0.6 | 21 | <5 | 1.87 (0.45 - 12.8) | 0.46 | 2.06 (0.39 - 10.9) | 0.41 |
| ODOD | 11,398 | 39,009 | 7.1 | 32.4 | 107 | 469 | 1.26 (0.94 - 1.68) | 0.12 | 1.30 (0.97 - 1.74) | 0.09 |
| MDCR | 1,287 | 6,450 | 0.9 | 5.2 | 10 | 70 | 1.45 (0.50 - 3.67) | 0.46 | 1.51 (0.56 - 4.08) | 0.43 |
| MDCD | 2,284 | 11,065 | 1.2 | 6.8 | 24 | 111 | 1.35 (0.73 - 2.41) | 0.33 | 1.23 (0.68 - 2.24) | 0.50 |
| Open Claims | 252,941 | 802,749 | 189.5 | 758.2 | 1,026 | 3,898 | 1.26 (1.14 - 1.39) | <0.01 | 1.29 (1.06 - 1.57) | 0.01 |
| OEHR | 15,065 | 62,231 | 4.7 | 35.3 | 43 | 380 | 1.12 (0.75 - 1.65) | 0.58 | 1.17 (0.78 - 1.75) | 0.53 |
| SIDIAP | 2,818 | 953 | 3.9 | 1.7 | 17 | 6 | 1.58 (0.50 - 5.93) | 0.47 | 1.85 (0.54 - 6.35) | 0.34 |
| Meta-analysis | 301,501 | 956,675 | 218.8 | 864.0 | 1,320 | 5,081 | 1.25 (1.15 - 1.36) | <0.01 | 1.25 (1.05 - 1.48) | 0.01 |
| Meta-analysis (w/o USOC) | 48,560 | 153,926 | 29.3 | 105.8 | 294 | 1,183 | 1.21 (1.00 - 1.45) | 0.05 | 1.19 (0.98 - 1.43) | 0.09 |
| **PS-stratification / on-treatment TAR** | | | | | | | | | | |
| CCAE | 18,444 | 34,899 | 13.3 | 24.7 | 112 | 149 | 1.53 (1.16 - 2.02) | <0.01 | 1.49 (1.13 - 1.97) | <0.01 |
| DAG | 3,151 | 1,186 | 2.5 | 1.2 | 21 | 8 | 1.79 (0.53 - 6.34) | 0.36 | 1.16 (0.33 - 4.04) | 0.80 |
| ODOD | 12,379 | 39,757 | 7.5 | 33.1 | 115 | 489 | 1.30 (1.03 - 1.63) | 0.03 | 1.36 (1.08 - 1.72) | 0.01 |
| MDCR | 1,438 | 6,921 | 0.9 | 5.6 | 11 | 79 | 1.63 (0.76 - 3.22) | 0.18 | 1.61 (0.78 - 3.31) | 0.21 |
| MDCD | 2,330 | 11,349 | 1.2 | 7.0 | 24 | 114 | 1.25 (0.76 - 2.00) | 0.36 | 1.15 (0.71 - 1.87) | 0.57 |
| Open Claims | 266,948 | 812,384 | 197.5 | 767.2 | 1,078 | 4,035 | 1.29 (1.20 - 1.39) | <0.01 | 1.32 (1.11 - 1.57) | <0.01 |
| OEHR | 16,231 | 68,316 | 5.1 | 39.0 | 46 | 440 | 1.03 (0.73 - 1.42) | 0.86 | 1.07 (0.77 - 1.50) | 0.70 |
| SIDIAP | 2,919 | 1,995 | 4.0 | 4.3 | 17 | 12 | 1.77 (0.68 - 4.70) | 0.25 | 1.61 (0.61 - 4.22) | 0.34 |
| Meta-analysis | 314,002 | 955,356 | 223.5 | 863.9 | 1,351 | 5,113 | 1.29 (1.21 - 1.38) | <0.01 | 1.30 (1.11 - 1.52) | <0.01 |
| Meta-analysis (w/o USOC) | 47,054 | 142,972 | 25.9 | 96.8 | 273 | 1,078 | 1.29 (1.06 - 1.58) | 0.01 | 1.31 (1.06 - 1.61) | 0.01 |
| **PS-matching / on-treatment TAR without escalation** | | | | | | | | | | |
| CCAE | 15,708 | 34,218 | 9.9 | 21.0 | 76 | 119 | 1.10 (0.72 - 1.65) | 0.65 | 1.04 (0.69 - 1.57) | 0.86 |
| DAG | 3,100 | 659 | 2.2 | 0.6 | 19 | <5 | 1.92 (0.47 - 13.0) | 0.44 | 2.08 (0.39 - 11.0) | 0.40 |
| ODOD | 11,398 | 39,009 | 6.3 | 28.9 | 94 | 415 | 1.31 (0.96 - 1.77) | 0.09 | 1.32 (0.97 - 1.80) | 0.09 |
| MDCR | 1,287 | 6,450 | 0.8 | 4.7 | 9 | 60 | 2.33 (0.77 - 6.17) | 0.11 | 2.37 (0.84 - 6.70) | 0.11 |
| MDCD | 2,284 | 11,065 | 1.0 | 5.7 | 21 | 97 | 1.41 (0.74 - 2.64) | 0.29 | 1.35 (0.71 - 2.56) | 0.37 |
| Open Claims | 252,941 | 802,749 | 159.1 | 638.7 | 864 | 3,279 | 1.27 (1.14 - 1.41) | <0.01 | 1.30 (1.07 - 1.57) | <0.01 |
| OEHR | 15,065 | 62,231 | 4.1 | 30.0 | 37 | 298 | 1.22 (0.79 - 1.86) | 0.36 | 1.26 (0.82 - 1.95) | 0.32 |
| SIDIAP | 2,818 | 953 | 3.4 | 1.4 | 15 | <5 | 2.47 (0.59 - 16.8) | 0.29 | 3.17 (0.59 - 17.0) | 0.18 |
| Meta-analysis | 298,683 | 955,722 | 181.2 | 729.0 | 1,101 | 4,268 | 1.27 (1.16 - 1.39) | <0.01 | 1.27 (1.07 - 1.51) | <0.01 |
| Meta-analysis (w/o USOC) | 45,742 | 152,973 | 22.1 | 90.3 | 237 | 989 | 1.27 (1.04 - 1.55) | 0.02 | 1.25 (1.02 - 1.53) | 0.04 |
| **PS-stratification / on-treatment TAR without escalation** | | | | | | | | | | |
| CCAE | 18,444 | 34,899 | 11.5 | 21.2 | 93 | 121 | 1.69 (1.25 - 2.29) | <0.01 | 1.61 (1.19 - 2.19) | <0.01 |
| DAG | 3,151 | 1,186 | 2.2 | 1.1 | 19 | 7 | 2.53 (0.70 - 9.21) | 0.16 | 1.63 (0.45 - 5.89) | 0.47 |
| ODOD | 12,379 | 39,757 | 6.7 | 29.6 | 100 | 435 | 1.31 (1.02 - 1.67) | 0.03 | 1.36 (1.06 - 1.74) | 0.02 |
| MDCR | 1,438 | 6,921 | 0.8 | 5.1 | 10 | 68 | 1.97 (0.88 - 4.00) | 0.08 | 1.83 (0.86 - 3.90) | 0.12 |
| MDCD | 2,330 | 11,349 | 1.0 | 5.8 | 21 | 100 | 1.33 (0.77 - 2.20) | 0.29 | 1.20 (0.71 - 2.03) | 0.50 |
| Open Claims | 266,948 | 812,384 | 166.1 | 646.5 | 907 | 3,398 | 1.32 (1.21 - 1.43) | <0.01 | 1.35 (1.15 - 1.57) | <0.01 |
| OEHR | 16,231 | 68,316 | 4.4 | 33.0 | 39 | 347 | 1.10 (0.76 - 1.57) | 0.60 | 1.11 (0.77 - 1.60) | 0.61 |
| SIDIAP | 2,919 | 1,995 | 3.5 | 3.5 | 15 | 9 | 2.06 (0.70 - 6.24) | 0.19 | 2.10 (0.70 - 6.25) | 0.19 |
| Meta-analysis | 314,002 | 955,356 | 188.7 | 730.4 | 1,139 | 4,301 | 1.33 (1.23 - 1.43) | <0.01 | 1.33 (1.16 - 1.52) | <0.01 |
| Meta-analysis (w/o USOC) | 47,054 | 142,972 | 22.6 | 83.8 | 232 | 903 | 1.36 (1.09 - 1.70) | <0.01 | 1.35 (1.07 - 1.69) | 0.01 |

##### 1.3.12 Acute pancreatitis

**Supplementary Table 64:** Exposures, outcomes and risk estimates for Acute pancreatitis in SGLT2I (female) (T) vs SU (female) (C) new-user cohorts under four different propensity score (PS) adjustment / time-at-risk (TAR) designs across data sources. We report total patient exposure time in 1,000 patient-year (KYr) units, uncalibrated and calibrated hazard ratios (HRs) and their 95% confidence intervals (CIs). Greyed out entries did not pass study diagnostics and are included only for completeness.

| | Patients | | Exposure (KYr) | | Outcomes | | Uncalibrated | | Calibrated | |
|---|---|---|---|---|---|---|---|---|---|---|
| | T | C | T | C | T | C | HR (95% CI) | $p$ | HR (95% CI) | $p$ |
| **PS-matching / on-treatment TAR** | | | | | | | | | | |
| CCAE | 15,936 | 34,825 | 11.8 | 24.9 | 21 | 58 | 0.80 (0.41 - 1.51) | 0.52 | 0.77 (0.40 - 1.48) | 0.44 |
| DAG | 3,180 | 682 | 2.5 | 0.7 | 0 | 0 | | | | |
| ODOD | 11,759 | 40,592 | 7.3 | 34.0 | 16 | 101 | 1.22 (0.57 - 2.42) | 0.60 | 1.25 (0.61 - 2.59) | 0.54 |
| MDCR | 1,317 | 6,709 | 0.9 | 5.4 | <5 | 22 | 0.86 (0.12 - 4.40) | 0.87 | 0.89 (0.15 - 5.49) | 0.90 |
| MDCD | 2,344 | 11,389 | 1.3 | 7.1 | 9 | 36 | 1.08 (0.36 - 2.84) | 0.89 | 0.99 (0.35 - 2.76) | 0.94 |
| Open Claims | 258,324 | 821,575 | 193.9 | 777.8 | 260 | 1,636 | 0.87 (0.73 - 1.03) | 0.11 | 0.89 (0.70 - 1.13) | 0.35 |
| OEHR | 15,419 | 63,891 | 4.8 | 36.6 | 9 | 76 | 1.20 (0.48 - 2.73) | 0.68 | 1.25 (0.52 - 3.00) | 0.63 |
| SIDIAP | 2,840 | 960 | 4.0 | 1.7 | <5 | 0 | | | | |
| Meta-analysis | 303,782 | 972,272 | 219.1 | 880.4 | 315 | 1,907 | 0.89 (0.76 - 1.04) | 0.15 | 0.89 (0.72 - 1.11) | 0.30 |
| Meta-analysis (w/o USOC) | 45,458 | 150,697 | 25.2 | 102.5 | 55 | 271 | 1.03 (0.69 - 1.52) | 0.89 | 1.01 (0.68 - 1.50) | 0.94 |
| **PS-stratification / on-treatment TAR** | | | | | | | | | | |
| CCAE | 18,712 | 35,440 | 13.6 | 25.1 | 22 | 59 | 0.74 (0.43 - 1.24) | 0.27 | 0.72 (0.42 - 1.23) | 0.24 |
| DAG | 3,231 | 1,228 | 2.5 | 1.2 | 0 | 0 | | | | |
| ODOD | 12,741 | 41,471 | 7.8 | 34.8 | 17 | 104 | 0.97 (0.53 - 1.65) | 0.90 | 1.01 (0.58 - 1.78) | 0.95 |
| MDCR | 1,473 | 7,194 | 0.9 | 5.8 | <5 | 22 | 0.50 (0.07 - 1.97) | 0.40 | 0.49 (0.09 - 2.53) | 0.40 |
| MDCD | 2,390 | 11,698 | 1.3 | 7.3 | 9 | 39 | 1.56 (0.66 - 3.35) | 0.28 | 1.44 (0.64 - 3.22) | 0.38 |
| Open Claims | 272,688 | 831,302 | 202.2 | 786.9 | 275 | 1,680 | 0.81 (0.70 - 0.93) | <0.01 | 0.83 (0.67 - 1.02) | 0.08 |
| OEHR | 16,582 | 70,088 | 5.2 | 40.3 | 9 | 83 | 1.15 (0.51 - 2.30) | 0.71 | 1.20 (0.57 - 2.54) | 0.64 |
| SIDIAP | 2,941 | 2,005 | 4.0 | 4.3 | <5 | <5 | 1.02 (0.05 - 11.0) | 0.99 | 0.93 (0.06 - 14.2) | 0.95 |
| Meta-analysis | 320,723 | 978,301 | 228.8 | 887.2 | 323 | 1,926 | 0.82 (0.72 - 0.93) | <0.01 | 0.83 (0.68 - 1.00) | 0.05 |
| Meta-analysis (w/o USOC) | 48,035 | 146,999 | 26.6 | 100.3 | 48 | 246 | 0.90 (0.64 - 1.27) | 0.54 | 0.91 (0.64 - 1.28) | 0.59 |
| **PS-matching / on-treatment TAR without escalation** | | | | | | | | | | |
| CCAE | 15,936 | 34,825 | 10.1 | 21.4 | 16 | 43 | 0.93 (0.44 - 1.88) | 0.85 | 0.88 (0.42 - 1.82) | 0.73 |
| DAG | 3,180 | 682 | 2.3 | 0.6 | 0 | 0 | | | | |
| ODOD | 11,759 | 40,592 | 6.5 | 30.3 | 12 | 84 | 1.41 (0.63 - 2.90) | 0.38 | 1.43 (0.67 - 3.05) | 0.36 |
| MDCR | 1,317 | 6,709 | 0.8 | 4.9 | <5 | 21 | 0.86 (0.12 - 4.40) | 0.87 | 0.88 (0.14 - 5.39) | 0.89 |
| MDCD | 2,344 | 11,389 | 1.0 | 5.9 | 8 | 33 | 1.01 (0.31 - 2.87) | 0.98 | 0.97 (0.32 - 2.94) | 0.93 |
| Open Claims | 258,324 | 821,575 | 162.8 | 655.3 | 201 | 1,343 | 0.82 (0.68 - 0.99) | 0.04 | 0.84 (0.66 - 1.08) | 0.18 |
| OEHR | 15,419 | 63,891 | 4.2 | 31.0 | 7 | 62 | 1.08 (0.37 - 2.70) | 0.88 | 1.11 (0.41 - 2.99) | 0.84 |
| SIDIAP | 2,840 | 960 | 3.5 | 1.4 | <5 | 0 | | | | |
| Meta-analysis | 303,782 | 972,272 | 184.7 | 743.9 | 244 | 1,565 | 0.87 (0.72 - 1.04) | 0.12 | 0.87 (0.69 - 1.09) | 0.23 |
| Meta-analysis (w/o USOC) | 45,458 | 150,697 | 21.9 | 88.6 | 43 | 222 | 1.10 (0.72 - 1.70) | 0.65 | 1.08 (0.70 - 1.66) | 0.73 |
| **PS-stratification / on-treatment TAR without escalation** | | | | | | | | | | |
| CCAE | 18,712 | 35,440 | 11.7 | 21.6 | 17 | 44 | 0.78 (0.42 - 1.41) | 0.42 | 0.74 (0.40 - 1.37) | 0.35 |
| DAG | 3,231 | 1,228 | 2.3 | 1.2 | 0 | 0 | | | | |
| ODOD | 12,741 | 41,471 | 6.9 | 31.1 | 13 | 86 | 1.03 (0.52 - 1.87) | 0.94 | 1.06 (0.56 - 2.01) | 0.85 |
| MDCR | 1,473 | 7,194 | 0.8 | 5.3 | <5 | 21 | 0.51 (0.08 - 2.04) | 0.43 | 0.48 (0.09 - 2.48) | 0.38 |
| MDCD | 2,390 | 11,698 | 1.1 | 6.1 | 8 | 34 | 1.46 (0.59 - 3.30) | 0.39 | 1.32 (0.56 - 3.13) | 0.53 |
| Open Claims | 272,688 | 831,302 | 170.1 | 663.2 | 211 | 1,380 | 0.76 (0.65 - 0.89) | <0.01 | 0.78 (0.63 - 0.96) | 0.02 |
| OEHR | 16,582 | 70,088 | 4.5 | 34.2 | 7 | 68 | 0.96 (0.38 - 2.06) | 0.92 | 0.96 (0.41 - 2.25) | 0.93 |
| SIDIAP | 2,941 | 2,005 | 3.5 | 3.6 | <5 | <5 | 1.02 (0.05 - 10.9) | 0.99 | 1.03 (0.07 - 15.9) | 0.96 |
| Meta-analysis | 320,723 | 978,301 | 193.2 | 750.1 | 248 | 1,578 | 0.78 (0.68 - 0.90) | <0.01 | 0.78 (0.65 - 0.94) | <0.01 |
| Meta-analysis (w/o USOC) | 48,035 | 146,999 | 23.2 | 86.9 | 37 | 198 | 0.90 (0.61 - 1.33) | 0.61 | 0.89 (0.60 - 1.32) | 0.57 |

##### 1.3.13 Bone fracture

**Supplementary Table 65:** Exposures, outcomes and risk estimates for Bone fracture in SGLT2I (female) (T) vs SU (female) (C) new-user cohorts under four different propensity score (PS) adjustment / time-at-risk (TAR) designs across data sources. We report total patient exposure time in 1,000 patient-year (KYr) units, uncalibrated and calibrated hazard ratios (HRs) and their 95% confidence intervals (CIs). Greyed out entries did not pass study diagnostics and are included only for completeness.

| | Patients | | Exposure (KYr) | | Outcomes | | Uncalibrated | | Calibrated | |
|---|---|---|---|---|---|---|---|---|---|---|
| | T | C | T | C | T | C | HR (95% CI) | $p$ | HR (95% CI) | $p$ |
| **PS-matching / on-treatment TAR** | | | | | | | | | | |
| CCAE | 14,510 | 31,898 | 10.5 | 22.4 | 188 | 453 | 1.06 (0.82 - 1.37) | 0.64 | 1.02 (0.78 - 1.32) | 0.87 |
| DAG | 2,771 | 603 | 2.2 | 0.6 | 31 | 17 | 0.29 (0.11 - 0.76) | 0.01 | 0.32 (0.12 - 0.84) | 0.02 |
| ODOD | 10,623 | 36,345 | 6.5 | 29.4 | 217 | 1,069 | 1.10 (0.89 - 1.35) | 0.36 | 1.14 (0.92 - 1.40) | 0.26 |
| MDCR | 1,155 | 5,643 | 0.7 | 4.4 | 34 | 210 | 0.92 (0.56 - 1.47) | 0.74 | 0.96 (0.59 - 1.55) | 0.81 |
| MDCD | 2,052 | 10,040 | 1.1 | 6.1 | 32 | 222 | 0.88 (0.53 - 1.42) | 0.62 | 0.81 (0.49 - 1.32) | 0.41 |
| Open Claims | 225,774 | 719,920 | 168.0 | 667.2 | 2,702 | 14,367 | 0.90 (0.85 - 0.95) | <0.01 | 0.92 (0.77 - 1.10) | 0.38 |
| OEHR | 14,291 | 58,744 | 4.5 | 32.8 | 66 | 804 | 0.74 (0.54 - 1.01) | 0.07 | 0.77 (0.56 - 1.08) | 0.21 |
| SIDIAP | 2,422 | 853 | 3.4 | 1.5 | 52 | 23 | 1.09 (0.54 - 2.27) | 0.82 | 1.27 (0.62 - 2.61) | 0.53 |
| Meta-analysis | 270,827 | 863,443 | 194.7 | 763.9 | 3,291 | 17,148 | 0.94 (0.85 - 1.05) | 0.26 | 0.94 (0.78 - 1.13) | 0.50 |
| Meta-analysis (w/o USOC) | 45,053 | 143,523 | 26.7 | 96.7 | 589 | 2,781 | 0.99 (0.87 - 1.13) | 0.91 | 0.98 (0.85 - 1.12) | 0.74 |
| **PS-stratification / on-treatment TAR** | | | | | | | | | | |
| CCAE | 16,973 | 32,516 | 12.0 | 22.6 | 230 | 458 | 0.93 (0.78 - 1.11) | 0.42 | 0.91 (0.76 - 1.08) | 0.31 |
| DAG | 2,815 | 1,043 | 2.2 | 1.0 | 31 | 30 | 0.68 (0.34 - 1.38) | 0.28 | 0.44 (0.22 - 0.89) | 0.03 |
| ODOD | 11,478 | 37,125 | 6.9 | 30.2 | 222 | 1,114 | 1.06 (0.90 - 1.25) | 0.45 | 1.12 (0.95 - 1.31) | 0.22 |
| MDCR | 1,280 | 6,081 | 0.8 | 4.7 | 35 | 236 | 0.95 (0.63 - 1.40) | 0.82 | 0.94 (0.63 - 1.40) | 0.76 |
| MDCD | 2,102 | 10,397 | 1.1 | 6.4 | 33 | 227 | 0.90 (0.60 - 1.31) | 0.60 | 0.83 (0.56 - 1.22) | 0.35 |
| Open Claims | 237,944 | 728,813 | 175.0 | 675.5 | 2,817 | 14,731 | 0.91 (0.87 - 0.95) | <0.01 | 0.93 (0.79 - 1.09) | 0.38 |
| OEHR | 15,400 | 64,626 | 4.8 | 36.2 | 69 | 932 | 0.73 (0.55 - 0.94) | 0.02 | 0.76 (0.58 - 0.99) | 0.06 |
| SIDIAP | 2,510 | 1,740 | 3.4 | 3.6 | 55 | 62 | 1.12 (0.68 - 1.82) | 0.65 | 1.02 (0.62 - 1.66) | 0.80 |
| Meta-analysis | 281,795 | 863,080 | 198.7 | 764.5 | 3,338 | 17,235 | 0.92 (0.83 - 1.02) | 0.11 | 0.93 (0.78 - 1.10) | 0.40 |
| Meta-analysis (w/o USOC) | 43,851 | 134,267 | 23.8 | 89.0 | 521 | 2,504 | 0.92 (0.76 - 1.11) | 0.38 | 0.93 (0.76 - 1.13) | 0.47 |
| **PS-matching / on-treatment TAR without escalation** | | | | | | | | | | |
| CCAE | 14,510 | 31,898 | 9.0 | 19.3 | 159 | 393 | 1.02 (0.78 - 1.34) | 0.87 | 0.96 (0.74 - 1.27) | 0.80 |
| DAG | 2,771 | 603 | 2.0 | 0.5 | 28 | 17 | 0.31 (0.12 - 0.83) | 0.02 | 0.34 (0.13 - 0.90) | 0.03 |
| ODOD | 10,623 | 36,345 | 5.8 | 26.3 | 188 | 951 | 1.09 (0.87 - 1.35) | 0.47 | 1.10 (0.88 - 1.37) | 0.44 |
| MDCR | 1,155 | 5,643 | 0.6 | 4.0 | 29 | 193 | 0.91 (0.53 - 1.50) | 0.72 | 0.93 (0.55 - 1.56) | 0.76 |
| MDCD | 2,052 | 10,040 | 0.9 | 5.1 | 27 | 187 | 0.70 (0.38 - 1.22) | 0.23 | 0.67 (0.38 - 1.19) | 0.18 |
| Open Claims | 225,774 | 719,920 | 141.2 | 562.8 | 2,253 | 12,118 | 0.90 (0.84 - 0.96) | <0.01 | 0.92 (0.77 - 1.09) | 0.35 |
| OEHR | 14,291 | 58,744 | 3.9 | 28.0 | 56 | 678 | 0.71 (0.50 - 0.99) | 0.05 | 0.73 (0.52 - 1.04) | 0.10 |
| SIDIAP | 2,422 | 853 | 3.0 | 1.2 | 43 | 19 | 1.04 (0.45 - 2.54) | 0.92 | 1.34 (0.57 - 3.17) | 0.52 |
| Meta-analysis | 270,827 | 863,443 | 164.5 | 646.8 | 2,755 | 14,539 | 0.92 (0.84 - 1.00) | 0.05 | 0.92 (0.78 - 1.09) | 0.32 |
| Meta-analysis (w/o USOC) | 45,053 | 143,523 | 23.2 | 83.9 | 502 | 2,421 | 0.95 (0.81 - 1.11) | 0.50 | 0.93 (0.79 - 1.09) | 0.39 |
| **PS-stratification / on-treatment TAR without escalation** | | | | | | | | | | |
| CCAE | 16,973 | 32,516 | 10.4 | 19.5 | 196 | 398 | 0.90 (0.74 - 1.08) | 0.26 | 0.86 (0.71 - 1.04) | 0.13 |
| DAG | 2,815 | 1,043 | 2.0 | 1.0 | 28 | 28 | 0.63 (0.30 - 1.30) | 0.20 | 0.40 (0.19 - 0.83) | 0.02 |
| ODOD | 11,478 | 37,125 | 6.2 | 27.0 | 192 | 992 | 1.04 (0.87 - 1.23) | 0.69 | 1.07 (0.90 - 1.28) | 0.46 |
| MDCR | 1,280 | 6,081 | 0.7 | 4.3 | 30 | 218 | 0.88 (0.56 - 1.33) | 0.55 | 0.82 (0.53 - 1.26) | 0.38 |
| MDCD | 2,102 | 10,397 | 0.9 | 5.3 | 28 | 192 | 0.89 (0.57 - 1.34) | 0.60 | 0.81 (0.53 - 1.23) | 0.33 |
| Open Claims | 237,944 | 728,813 | 147.3 | 570.0 | 2,346 | 12,427 | 0.90 (0.86 - 0.95) | <0.01 | 0.92 (0.80 - 1.06) | 0.28 |
| OEHR | 15,400 | 64,626 | 4.2 | 30.8 | 59 | 786 | 0.73 (0.54 - 0.96) | 0.03 | 0.73 (0.55 - 0.98) | 0.06 |
| SIDIAP | 2,510 | 1,740 | 3.0 | 3.0 | 46 | 54 | 0.98 (0.57 - 1.67) | 0.95 | 1.00 (0.58 - 1.71) | 0.80 |
| Meta-analysis | 281,795 | 863,080 | 168.0 | 647.3 | 2,793 | 14,603 | 0.91 (0.87 - 0.95) | <0.01 | 0.91 (0.80 - 1.03) | 0.12 |
| Meta-analysis (w/o USOC) | 43,851 | 134,267 | 20.7 | 77.4 | 447 | 2,176 | 0.90 (0.75 - 1.08) | 0.26 | 0.89 (0.74 - 1.07) | 0.24 |

##### 1.3.14 Diabetic ketoacidosis

**Supplementary Table 66:** Exposures, outcomes and risk estimates for Diabetic ketoacidosis in SGLT2I (female) (T) vs SU (female) (C) new-user cohorts under four different propensity score (PS) adjustment / time-at-risk (TAR) designs across data sources. We report total patient exposure time in 1,000 patient-year (KYr) units, uncalibrated and calibrated hazard ratios (HRs) and their 95% confidence intervals (CIs). Greyed out entries did not pass study diagnostics and are included only for completeness.

| | Patients | | Exposure (KYr) | | Outcomes | | Uncalibrated | | Calibrated | |
|---|---|---|---|---|---|---|---|---|---|---|
| | T | C | T | C | T | C | HR (95% CI) | $p$ | HR (95% CI) | $p$ |
| **PS-matching / on-treatment TAR** | | | | | | | | | | |
| CCAE | 16,071 | 35,137 | 11.8 | 25.1 | 26 | 33 | 1.89 (0.89 - 3.99) | 0.10 | 1.81 (0.85 - 3.82) | 0.12 |
| DAG | 3,180 | 682 | 2.5 | 0.7 | 0 | 0 | | | | |
| ODOD | 11,833 | 40,906 | 7.4 | 34.3 | 22 | 63 | 2.67 (1.34 - 5.27) | <0.01 | 2.75 (1.39 - 5.46) | <0.01 |
| MDCR | 1,330 | 6,791 | 0.9 | 5.5 | <5 | 6 | 1.41 (0.06 - 17.2) | 0.81 | 1.46 (0.08 - 25.3) | 0.79 |
| MDCD | 2,380 | 11,611 | 1.3 | 7.3 | 5 | 25 | 1.23 (0.38 - 3.39) | 0.71 | 1.13 (0.38 - 3.37) | 0.83 |
| Open Claims | 261,540 | 831,033 | 196.2 | 787.6 | 242 | 707 | 1.67 (1.37 - 2.04) | <0.01 | 1.71 (1.32 - 2.22) | <0.01 |
| OEHR | 15,483 | 64,194 | 4.9 | 36.8 | 11 | 58 | 1.71 (0.70 - 3.84) | 0.22 | 1.78 (0.76 - 4.18) | 0.21 |
| SIDIAP | 2,849 | 965 | 4.0 | 1.7 | 0 | 0 | | | | |
| Meta-analysis | 307,307 | 982,881 | 221.6 | 891.0 | 306 | 886 | 1.73 (1.44 - 2.06) | <0.01 | 1.72 (1.36 - 2.18) | <0.01 |
| Meta-analysis (w/o USOC) | 45,767 | 151,848 | 25.4 | 103.5 | 64 | 179 | 1.96 (1.31 - 2.94) | <0.01 | 1.93 (1.29 - 2.89) | <0.01 |
| **PS-stratification / on-treatment TAR** | | | | | | | | | | |
| CCAE | 18,865 | 35,766 | 13.7 | 25.4 | 31 | 33 | 1.91 (1.12 - 3.27) | 0.02 | 1.86 (1.09 - 3.18) | 0.02 |
| DAG | 3,231 | 1,228 | 2.5 | 1.2 | 0 | 0 | | | | |
| ODOD | 12,833 | 41,815 | 7.9 | 35.1 | 24 | 67 | 1.92 (1.10 - 3.29) | 0.02 | 2.02 (1.16 - 3.49) | 0.01 |
| MDCR | 1,486 | 7,279 | 0.9 | 5.9 | <5 | 7 | 1.55 (0.07 - 13.7) | 0.75 | 1.52 (0.11 - 21.2) | 0.76 |
| MDCD | 2,427 | 11,887 | 1.3 | 7.5 | 5 | 25 | 1.23 (0.39 - 3.23) | 0.70 | 1.13 (0.39 - 3.25) | 0.82 |
| Open Claims | 276,157 | 841,119 | 204.7 | 797.0 | 254 | 720 | 1.48 (1.27 - 1.74) | <0.01 | 1.52 (1.22 - 1.90) | <0.01 |
| OEHR | 16,657 | 70,431 | 5.2 | 40.6 | 11 | 64 | 1.74 (0.80 - 3.45) | 0.14 | 1.81 (0.87 - 3.75) | 0.12 |
| SIDIAP | 2,950 | 2,013 | 4.0 | 4.3 | 0 | 0 | | | | |
| Meta-analysis | 324,512 | 989,131 | 231.4 | 898.0 | 320 | 884 | 1.55 (1.34 - 1.79) | <0.01 | 1.56 (1.28 - 1.91) | <0.01 |
| Meta-analysis (w/o USOC) | 48,355 | 148,012 | 26.8 | 101.1 | 66 | 164 | 1.88 (1.34 - 2.63) | <0.01 | 1.90 (1.35 - 2.67) | <0.01 |
| **PS-matching / on-treatment TAR without escalation** | | | | | | | | | | |
| CCAE | 16,071 | 35,137 | 10.2 | 21.6 | 20 | 25 | 1.61 (0.66 - 3.85) | 0.29 | 1.52 (0.63 - 3.67) | 0.35 |
| DAG | 3,180 | 682 | 2.3 | 0.6 | 0 | 0 | | | | |
| ODOD | 11,833 | 40,906 | 6.5 | 30.6 | 19 | 51 | 2.40 (1.17 - 4.88) | 0.02 | 2.43 (1.19 - 4.97) | 0.02 |
| MDCR | 1,330 | 6,791 | 0.8 | 5.0 | <5 | <5 | 1.41 (0.06 - 17.2) | 0.81 | 1.44 (0.08 - 24.9) | 0.81 |
| MDCD | 2,380 | 11,611 | 1.1 | 6.0 | <5 | 18 | 1.04 (0.23 - 3.45) | 0.95 | 0.99 (0.26 - 3.85) | 0.95 |
| Open Claims | 261,540 | 831,033 | 164.7 | 663.3 | 190 | 525 | 1.66 (1.34 - 2.06) | <0.01 | 1.70 (1.30 - 2.23) | <0.01 |
| OEHR | 15,483 | 64,194 | 4.2 | 31.2 | 7 | 36 | 1.43 (0.43 - 4.04) | 0.53 | 1.48 (0.48 - 4.53) | 0.50 |
| SIDIAP | 2,849 | 965 | 3.5 | 1.4 | 0 | 0 | | | | |
| Meta-analysis | 304,927 | 971,270 | 185.7 | 746.7 | 236 | 637 | 1.70 (1.39 - 2.07) | <0.01 | 1.70 (1.33 - 2.17) | <0.01 |
| Meta-analysis (w/o USOC) | 43,387 | 140,237 | 21.0 | 83.4 | 46 | 112 | 1.91 (1.16 - 3.14) | 0.01 | 1.87 (1.13 - 3.08) | 0.02 |
| **PS-stratification / on-treatment TAR without escalation** | | | | | | | | | | |
| CCAE | 18,865 | 35,766 | 11.8 | 21.8 | 24 | 25 | 2.07 (1.11 - 3.84) | 0.02 | 1.97 (1.06 - 3.67) | 0.03 |
| DAG | 3,231 | 1,228 | 2.3 | 1.2 | 0 | 0 | | | | |
| ODOD | 12,833 | 41,815 | 7.0 | 31.4 | 20 | 54 | 2.06 (1.12 - 3.70) | 0.02 | 2.13 (1.17 - 3.88) | 0.01 |
| MDCR | 1,486 | 7,279 | 0.8 | 5.4 | <5 | 5 | 1.91 (0.08 - 21.6) | 0.65 | 1.78 (0.11 - 29.0) | 0.69 |
| MDCD | 2,427 | 11,887 | 1.1 | 6.2 | <5 | 18 | 1.05 (0.23 - 3.42) | 0.94 | 0.95 (0.25 - 3.65) | 0.94 |
| Open Claims | 276,157 | 841,119 | 172.1 | 671.5 | 200 | 536 | 1.52 (1.26 - 1.81) | <0.01 | 1.55 (1.24 - 1.94) | <0.01 |
| OEHR | 16,657 | 70,431 | 4.6 | 34.4 | 7 | 41 | 1.74 (0.64 - 4.15) | 0.24 | 1.76 (0.69 - 4.47) | 0.24 |
| SIDIAP | 2,950 | 2,013 | 3.5 | 3.6 | 0 | 0 | | | | |
| Meta-analysis | 324,512 | 989,131 | 195.4 | 759.1 | 251 | 656 | 1.61 (1.34 - 1.92) | <0.01 | 1.61 (1.30 - 1.99) | <0.01 |
| Meta-analysis (w/o USOC) | 48,355 | 148,012 | 23.3 | 87.5 | 51 | 120 | 2.00 (1.35 - 2.96) | <0.01 | 1.98 (1.34 - 2.93) | <0.01 |

##### 1.3.15 Diarrhea

**Supplementary Table 67:** Exposures, outcomes and risk estimates for Diarrhea in SGLT2I (female) (T) vs SU (female) (C) new-user cohorts under four different propensity score (PS) adjustment / time-at-risk (TAR) designs across data sources. We report total patient exposure time in 1,000 patient-year (KYr) units, uncalibrated and calibrated hazard ratios (HRs) and their 95% confidence intervals (CIs). Greyed out entries did not pass study diagnostics and are included only for completeness.

| | Patients | | Exposure (KYr) | | Outcomes | | Uncalibrated | | Calibrated | |
|---|---|---|---|---|---|---|---|---|---|---|
| | T | C | T | C | T | C | HR (95% CI) | $p$ | HR (95% CI) | $p$ |
| **PS-matching / on-treatment TAR** | | | | | | | | | | |
| CCAE | 13,530 | 29,669 | 9.9 | 20.6 | 312 | 781 | 0.74 (0.61 - 0.89) | <0.01 | 0.71 (0.59 - 0.86) | <0.01 |
| DAG | 3,105 | 653 | 2.4 | 0.6 | 7 | <5 | 3.30 (0.47 - 65.5) | 0.34 | 3.63 (0.31 - 43.1) | 0.31 |
| ODOD | 9,768 | 32,870 | 6.0 | 26.4 | 244 | 1,368 | 0.79 (0.65 - 0.95) | 0.01 | 0.81 (0.67 - 0.98) | 0.05 |
| MDCR | 1,100 | 5,279 | 0.7 | 4.1 | 25 | 202 | 0.47 (0.25 - 0.84) | 0.02 | 0.49 (0.26 - 0.91) | 0.03 |
| MDCD | 1,761 | 8,860 | 0.9 | 5.2 | 57 | 412 | 0.71 (0.48 - 1.02) | 0.07 | 0.65 (0.45 - 0.95) | 0.03 |
| Open Claims | 218,164 | 701,722 | 162.6 | 650.6 | 3,008 | 15,651 | 0.83 (0.79 - 0.88) | <0.01 | 0.85 (0.71 - 1.02) | 0.09 |
| OEHR | 13,181 | 54,532 | 4.0 | 29.4 | 162 | 1,524 | 0.86 (0.69 - 1.06) | 0.16 | 0.89 (0.71 - 1.13) | 0.49 |
| SIDIAP | 2,679 | 892 | 3.7 | 1.6 | 56 | 20 | 0.66 (0.32 - 1.40) | 0.28 | 0.77 (0.37 - 1.63) | 0.52 |
| Meta-analysis | 260,183 | 833,824 | 187.9 | 738.0 | 3,864 | 19,958 | 0.81 (0.76 - 0.86) | <0.01 | 0.81 (0.69 - 0.95) | 0.01 |
| Meta-analysis (w/o USOC) | 42,019 | 132,102 | 25.3 | 87.4 | 856 | 4,307 | 0.77 (0.69 - 0.85) | <0.01 | 0.75 (0.68 - 0.84) | <0.01 |
| **PS-stratification / on-treatment TAR** | | | | | | | | | | |
| CCAE | 15,864 | 30,271 | 11.4 | 20.9 | 371 | 798 | 0.83 (0.72 - 0.95) | <0.01 | 0.81 (0.70 - 0.93) | <0.01 |
| DAG | 3,158 | 1,189 | 2.5 | 1.2 | 7 | 5 | 3.80 (0.82 - 16.7) | 0.08 | 2.46 (0.54 - 11.1) | 0.25 |
| ODOD | 10,574 | 33,671 | 6.4 | 27.1 | 262 | 1,413 | 0.81 (0.70 - 0.93) | <0.01 | 0.85 (0.73 - 0.98) | 0.04 |
| MDCR | 1,228 | 5,815 | 0.8 | 4.5 | 29 | 223 | 0.60 (0.38 - 0.92) | 0.02 | 0.59 (0.38 - 0.92) | 0.02 |
| MDCD | 1,793 | 9,021 | 0.9 | 5.4 | 57 | 420 | 0.74 (0.55 - 0.98) | 0.04 | 0.68 (0.51 - 0.91) | 0.01 |
| Open Claims | 230,071 | 711,007 | 169.4 | 659.1 | 3,165 | 16,015 | 0.85 (0.81 - 0.88) | <0.01 | 0.87 (0.74 - 1.02) | 0.09 |
| OEHR | 14,218 | 59,849 | 4.4 | 32.3 | 177 | 1,724 | 0.89 (0.75 - 1.05) | 0.18 | 0.93 (0.78 - 1.11) | 0.50 |
| SIDIAP | 2,774 | 1,870 | 3.8 | 4.0 | 58 | 50 | 0.82 (0.49 - 1.39) | 0.47 | 0.75 (0.44 - 1.27) | 0.30 |
| Meta-analysis | 270,727 | 834,798 | 191.5 | 739.4 | 3,975 | 19,950 | 0.84 (0.81 - 0.88) | <0.01 | 0.85 (0.74 - 0.99) | 0.03 |
| Meta-analysis (w/o USOC) | 40,656 | 123,791 | 22.1 | 80.3 | 810 | 3,935 | 0.84 (0.77 - 0.91) | <0.01 | 0.85 (0.77 - 0.93) | <0.01 |
| **PS-matching / on-treatment TAR without escalation** | | | | | | | | | | |
| CCAE | 13,530 | 29,669 | 8.6 | 17.9 | 273 | 680 | 0.75 (0.61 - 0.91) | <0.01 | 0.71 (0.58 - 0.86) | <0.01 |
| DAG | 3,105 | 653 | 2.2 | 0.6 | 7 | <5 | 3.30 (0.47 - 65.5) | 0.34 | 3.58 (0.30 - 42.4) | 0.32 |
| ODOD | 9,768 | 32,870 | 5.3 | 23.7 | 219 | 1,247 | 0.79 (0.65 - 0.96) | 0.02 | 0.80 (0.66 - 0.98) | 0.04 |
| MDCR | 1,100 | 5,279 | 0.7 | 3.8 | 22 | 182 | 0.53 (0.27 - 0.98) | 0.06 | 0.54 (0.28 - 1.04) | 0.07 |
| MDCD | 1,761 | 8,860 | 0.8 | 4.4 | 47 | 345 | 0.74 (0.49 - 1.09) | 0.14 | 0.71 (0.47 - 1.06) | 0.11 |
| Open Claims | 218,164 | 701,722 | 137.5 | 552.1 | 2,460 | 13,269 | 0.82 (0.77 - 0.86) | <0.01 | 0.84 (0.70 - 0.99) | 0.04 |
| OEHR | 13,181 | 54,532 | 3.5 | 25.4 | 135 | 1,207 | 0.86 (0.68 - 1.09) | 0.22 | 0.89 (0.70 - 1.13) | 0.41 |
| SIDIAP | 2,679 | 892 | 3.3 | 1.3 | 49 | 18 | 0.62 (0.28 - 1.40) | 0.24 | 0.79 (0.35 - 1.78) | 0.59 |
| Meta-analysis | 260,183 | 833,824 | 159.6 | 628.7 | 3,205 | 16,948 | 0.81 (0.77 - 0.85) | <0.01 | 0.81 (0.69 - 0.94) | <0.01 |
| Meta-analysis (w/o USOC) | 42,019 | 132,102 | 22.1 | 76.6 | 745 | 3,679 | 0.78 (0.69 - 0.87) | <0.01 | 0.76 (0.67 - 0.86) | <0.01 |
| **PS-stratification / on-treatment TAR without escalation** | | | | | | | | | | |
| CCAE | 15,864 | 30,271 | 9.9 | 18.1 | 322 | 696 | 0.83 (0.72 - 0.96) | 0.01 | 0.79 (0.68 - 0.92) | <0.01 |
| DAG | 3,158 | 1,189 | 2.2 | 1.1 | 7 | <5 | 5.86 (1.18 - 28.2) | 0.03 | 3.76 (0.77 - 18.4) | 0.11 |
| ODOD | 10,574 | 33,671 | 5.7 | 24.4 | 234 | 1,292 | 0.79 (0.68 - 0.92) | <0.01 | 0.82 (0.70 - 0.96) | 0.02 |
| MDCR | 1,228 | 5,815 | 0.7 | 4.1 | 26 | 200 | 0.63 (0.39 - 0.97) | 0.05 | 0.58 (0.37 - 0.92) | 0.03 |
| MDCD | 1,793 | 9,021 | 0.8 | 4.5 | 47 | 353 | 0.72 (0.52 - 0.99) | 0.05 | 0.65 (0.47 - 0.90) | 0.01 |
| Open Claims | 230,071 | 711,007 | 143.4 | 559.6 | 2,586 | 13,584 | 0.83 (0.79 - 0.87) | <0.01 | 0.84 (0.73 - 0.97) | 0.02 |
| OEHR | 14,218 | 59,849 | 3.8 | 27.9 | 147 | 1,372 | 0.90 (0.75 - 1.08) | 0.28 | 0.91 (0.75 - 1.11) | 0.45 |
| SIDIAP | 2,774 | 1,870 | 3.3 | 3.3 | 50 | 37 | 0.80 (0.44 - 1.45) | 0.46 | 0.81 (0.45 - 1.47) | 0.51 |
| Meta-analysis | 270,727 | 834,798 | 162.8 | 630.1 | 3,289 | 16,944 | 0.83 (0.80 - 0.86) | <0.01 | 0.83 (0.73 - 0.94) | <0.01 |
| Meta-analysis (w/o USOC) | 40,656 | 123,791 | 19.4 | 70.5 | 703 | 3,360 | 0.83 (0.76 - 0.91) | <0.01 | 0.82 (0.75 - 0.91) | <0.01 |

##### 1.3.16 Genitourinary infection

**Supplementary Table 68:** Exposures, outcomes and risk estimates for Genitourinary infection in SGLT2I (female) (T) vs SU (female) (C) new-user cohorts under four different propensity score (PS) adjustment / time-at-risk (TAR) designs across data sources. We report total patient exposure time in 1,000 patient-year (KYr) units, uncalibrated and calibrated hazard ratios (HRs) and their 95% confidence intervals (CIs). Greyed out entries did not pass study diagnostics and are included only for completeness.

| | Patients | | Exposure (KYr) | | Outcomes | | Uncalibrated | | Calibrated | |
|---|---|---|---|---|---|---|---|---|---|---|
| | T | C | T | C | T | C | HR (95% CI) | $p$ | HR (95% CI) | $p$ |
| **PS-matching / on-treatment TAR** | | | | | | | | | | |
| CCAE | 11,136 | 24,608 | 7.9 | 16.9 | 472 | 1,046 | 1.12 (0.96 - 1.31) | 0.14 | 1.07 (0.92 - 1.26) | 0.43 |
| DAG | 2,162 | 481 | 1.7 | 0.4 | 87 | 19 | 1.69 (0.86 - 3.67) | 0.15 | 1.86 (0.90 - 3.87) | 0.11 |
| ODOD | 8,195 | 27,060 | 4.9 | 20.9 | 378 | 2,001 | 0.83 (0.72 - 0.97) | 0.02 | 0.86 (0.74 - 1.00) | 0.09 |
| MDCR | 996 | 4,606 | 0.6 | 3.5 | 51 | 301 | 1.12 (0.73 - 1.70) | 0.59 | 1.16 (0.76 - 1.78) | 0.50 |
| MDCD | 1,575 | 7,460 | 0.8 | 4.5 | 78 | 447 | 0.90 (0.65 - 1.23) | 0.51 | 0.82 (0.60 - 1.14) | 0.26 |
| Open Claims | 174,087 | 563,118 | 131.3 | 518.4 | 5,119 | 25,863 | 0.94 (0.90 - 0.98) | <0.01 | 0.96 (0.81 - 1.15) | 0.66 |
| OEHR | 11,752 | 48,208 | 3.5 | 25.6 | 237 | 1,906 | 1.17 (0.98 - 1.40) | 0.09 | 1.22 (0.99 - 1.49) | 0.19 |
| SIDIAP | 1,816 | 677 | 2.6 | 1.1 | 148 | 61 | 0.99 (0.66 - 1.48) | 0.94 | 1.15 (0.77 - 1.73) | 0.50 |
| Meta-analysis | 209,557 | 675,737 | 151.6 | 590.9 | 6,483 | 31,625 | 0.99 (0.89 - 1.11) | 0.89 | 0.99 (0.82 - 1.19) | 0.90 |
| Meta-analysis (w/o USOC) | 35,470 | 112,619 | 20.4 | 72.6 | 1,364 | 5,762 | 1.01 (0.88 - 1.17) | 0.86 | 1.00 (0.86 - 1.15) | 0.87 |
| **PS-stratification / on-treatment TAR** | | | | | | | | | | |
| CCAE | 12,966 | 24,966 | 9.0 | 17.0 | 553 | 1,057 | 1.01 (0.90 - 1.13) | 0.84 | 0.98 (0.88 - 1.11) | 0.79 |
| DAG | 2,193 | 813 | 1.7 | 0.8 | 89 | 35 | 1.36 (0.83 - 2.28) | 0.23 | 0.88 (0.53 - 1.47) | 0.60 |
| ODOD | 8,866 | 27,519 | 5.2 | 21.3 | 402 | 2,071 | 0.90 (0.80 - 1.01) | 0.08 | 0.94 (0.84 - 1.06) | 0.40 |
| MDCR | 1,115 | 4,982 | 0.7 | 3.8 | 54 | 341 | 1.07 (0.76 - 1.49) | 0.68 | 1.05 (0.75 - 1.48) | 0.76 |
| MDCD | 1,602 | 7,632 | 0.8 | 4.6 | 79 | 458 | 0.98 (0.76 - 1.26) | 0.88 | 0.90 (0.70 - 1.17) | 0.46 |
| Open Claims | 183,262 | 571,242 | 136.5 | 525.7 | 5,341 | 26,626 | 0.91 (0.88 - 0.94) | <0.01 | 0.94 (0.80 - 1.10) | 0.42 |
| OEHR | 12,672 | 53,535 | 3.8 | 28.5 | 254 | 2,210 | 1.04 (0.90 - 1.20) | 0.58 | 1.08 (0.93 - 1.26) | 0.39 |
| SIDIAP | 1,915 | 1,459 | 2.7 | 2.9 | 154 | 169 | 0.89 (0.66 - 1.20) | 0.45 | 0.81 (0.60 - 1.09) | 0.20 |
| Meta-analysis | 217,766 | 677,262 | 154.6 | 592.5 | 6,550 | 31,964 | 0.95 (0.89 - 1.01) | 0.10 | 0.96 (0.82 - 1.12) | 0.56 |
| Meta-analysis (w/o USOC) | 34,504 | 106,020 | 18.1 | 66.9 | 1,209 | 5,338 | 0.98 (0.90 - 1.07) | 0.61 | 0.99 (0.90 - 1.09) | 0.81 |
| **PS-matching / on-treatment TAR without escalation** | | | | | | | | | | |
| CCAE | 11,136 | 24,608 | 6.8 | 14.7 | 412 | 886 | 1.12 (0.95 - 1.32) | 0.19 | 1.06 (0.89 - 1.25) | 0.56 |
| DAG | 2,162 | 481 | 1.5 | 0.4 | 79 | 16 | 1.90 (0.91 - 4.49) | 0.12 | 2.06 (0.92 - 4.59) | 0.09 |
| ODOD | 8,195 | 27,060 | 4.4 | 18.9 | 333 | 1,800 | 0.86 (0.74 - 1.01) | 0.07 | 0.88 (0.75 - 1.03) | 0.14 |
| MDCR | 996 | 4,606 | 0.6 | 3.2 | 46 | 271 | 1.22 (0.77 - 1.87) | 0.39 | 1.24 (0.79 - 1.93) | 0.37 |
| MDCD | 1,575 | 7,460 | 0.7 | 3.8 | 58 | 373 | 0.73 (0.50 - 1.05) | 0.10 | 0.70 (0.48 - 1.01) | 0.07 |
| Open Claims | 174,087 | 563,118 | 110.7 | 440.7 | 4,339 | 22,098 | 0.94 (0.90 - 0.99) | 0.01 | 0.97 (0.82 - 1.14) | 0.71 |
| OEHR | 11,752 | 48,208 | 3.1 | 22.1 | 203 | 1,556 | 1.21 (0.99 - 1.46) | 0.06 | 1.25 (1.02 - 1.52) | 0.06 |
| SIDIAP | 1,816 | 677 | 2.3 | 1.0 | 135 | 55 | 1.00 (0.66 - 1.54) | 1.00 | 1.28 (0.84 - 1.97) | 0.30 |
| Meta-analysis | 209,557 | 675,737 | 128.6 | 504.4 | 5,526 | 27,039 | 1.00 (0.88 - 1.12) | 0.94 | 1.00 (0.83 - 1.20) | 0.91 |
| Meta-analysis (w/o USOC) | 35,470 | 112,619 | 17.9 | 63.7 | 1,187 | 4,941 | 1.01 (0.87 - 1.19) | 0.87 | 0.99 (0.84 - 1.17) | 0.86 |
| **PS-stratification / on-treatment TAR without escalation** | | | | | | | | | | |
| CCAE | 12,966 | 24,966 | 7.8 | 14.8 | 485 | 896 | 1.04 (0.92 - 1.18) | 0.51 | 0.99 (0.88 - 1.13) | 0.84 |
| DAG | 2,193 | 813 | 1.5 | 0.7 | 81 | 31 | 1.42 (0.84 - 2.46) | 0.20 | 0.91 (0.53 - 1.57) | 0.64 |
| ODOD | 8,866 | 27,519 | 4.7 | 19.3 | 355 | 1,869 | 0.90 (0.79 - 1.01) | 0.08 | 0.93 (0.82 - 1.05) | 0.28 |
| MDCR | 1,115 | 4,982 | 0.6 | 3.5 | 49 | 309 | 1.13 (0.79 - 1.59) | 0.50 | 1.05 (0.74 - 1.50) | 0.77 |
| MDCD | 1,602 | 7,632 | 0.7 | 3.9 | 59 | 382 | 0.85 (0.63 - 1.13) | 0.29 | 0.77 (0.57 - 1.03) | 0.09 |
| Open Claims | 183,262 | 571,242 | 115.3 | 447.2 | 4,529 | 22,753 | 0.92 (0.89 - 0.95) | <0.01 | 0.94 (0.82 - 1.08) | 0.39 |
| OEHR | 12,672 | 53,535 | 3.3 | 24.6 | 218 | 1,810 | 1.10 (0.94 - 1.28) | 0.23 | 1.11 (0.94 - 1.31) | 0.35 |
| SIDIAP | 1,915 | 1,459 | 2.3 | 2.4 | 141 | 142 | 0.94 (0.68 - 1.28) | 0.67 | 0.95 (0.69 - 1.30) | 0.64 |
| Meta-analysis | 217,766 | 677,262 | 131.2 | 505.9 | 5,587 | 27,328 | 0.97 (0.89 - 1.06) | 0.51 | 0.97 (0.84 - 1.12) | 0.70 |
| Meta-analysis (w/o USOC) | 34,504 | 106,020 | 15.9 | 58.7 | 1,058 | 4,575 | 1.00 (0.89 - 1.13) | 0.96 | 0.99 (0.88 - 1.12) | 0.86 |

##### 1.3.17 Hyperkalemia

**Supplementary Table 69:** Exposures, outcomes and risk estimates for Hyperkalemia in SGLT2I (female) (T) vs SU (female) (C) new-user cohorts under four different propensity score (PS) adjustment / time-at-risk (TAR) designs across data sources. We report total patient exposure time in 1,000 patient-year (KYr) units, uncalibrated and calibrated hazard ratios (HRs) and their 95% confidence intervals (CIs). Greyed out entries did not pass study diagnostics and are included only for completeness.

| | Patients | | Exposure (KYr) | | Outcomes | | Uncalibrated | | Calibrated | |
|---|---|---|---|---|---|---|---|---|---|---|
| | T | C | T | C | T | C | HR (95% CI) | $p$ | HR (95% CI) | $p$ |
| **PS-matching / on-treatment TAR** | | | | | | | | | | |
| CCAE | 15,821 | 34,604 | 11.6 | 24.6 | 64 | 200 | 0.55 (0.34 - 0.84) | <0.01 | 0.52 (0.33 - 0.82) | <0.01 |
| DAG | 2,974 | 640 | 2.3 | 0.6 | 25 | 9 | 0.63 (0.21 - 2.16) | 0.43 | 0.69 (0.21 - 2.22) | 0.53 |
| ODOD | 11,422 | 38,860 | 7.1 | 31.8 | 105 | 758 | 0.86 (0.65 - 1.13) | 0.29 | 0.89 (0.68 - 1.17) | 0.43 |
| MDCR | 1,299 | 6,566 | 0.9 | 5.3 | 5 | 89 | 0.50 (0.17 - 1.22) | 0.17 | 0.52 (0.19 - 1.40) | 0.20 |
| MDCD | 2,321 | 11,258 | 1.3 | 6.9 | 7 | 121 | 0.37 (0.14 - 0.83) | 0.03 | 0.34 (0.14 - 0.83) | 0.02 |
| Open Claims | 257,321 | 815,085 | 193.0 | 765.4 | 685 | 5,838 | 0.69 (0.62 - 0.76) | <0.01 | 0.70 (0.57 - 0.86) | <0.01 |
| OEHR | 15,234 | 63,024 | 4.8 | 35.7 | 31 | 465 | 0.99 (0.63 - 1.52) | 0.97 | 1.03 (0.66 - 1.63) | 0.88 |
| SIDIAP | 2,756 | 929 | 3.8 | 1.6 | 37 | 16 | 0.87 (0.35 - 2.19) | 0.76 | 1.01 (0.40 - 2.54) | 0.84 |
| Meta-analysis | 306,174 | 970,326 | 222.4 | 871.3 | 934 | 7,487 | 0.72 (0.61 - 0.85) | <0.01 | 0.72 (0.58 - 0.90) | <0.01 |
| Meta-analysis (w/o USOC) | 48,853 | 155,241 | 29.4 | 105.9 | 249 | 1,649 | 0.73 (0.56 - 0.96) | 0.02 | 0.72 (0.55 - 0.94) | 0.02 |
| **PS-stratification / on-treatment TAR** | | | | | | | | | | |
| CCAE | 18,560 | 35,219 | 13.4 | 24.8 | 76 | 203 | 0.77 (0.57 - 1.02) | 0.08 | 0.75 (0.56 - 1.00) | 0.06 |
| DAG | 3,022 | 1,171 | 2.4 | 1.2 | 26 | 12 | 0.59 (0.26 - 1.48) | 0.24 | 0.38 (0.16 - 0.92) | 0.04 |
| ODOD | 12,381 | 39,679 | 7.6 | 32.6 | 109 | 790 | 0.81 (0.65 - 1.00) | 0.06 | 0.85 (0.68 - 1.06) | 0.15 |
| MDCR | 1,451 | 7,008 | 0.9 | 5.6 | 5 | 98 | 0.36 (0.12 - 0.87) | 0.04 | 0.35 (0.13 - 0.95) | 0.04 |
| MDCD | 2,363 | 11,556 | 1.3 | 7.2 | 7 | 128 | 0.31 (0.13 - 0.64) | <0.01 | 0.29 (0.13 - 0.64) | <0.01 |
| Open Claims | 271,518 | 824,713 | 201.2 | 774.2 | 717 | 6,075 | 0.66 (0.61 - 0.72) | <0.01 | 0.68 (0.57 - 0.81) | <0.01 |
| OEHR | 16,395 | 69,036 | 5.1 | 39.2 | 32 | 550 | 0.73 (0.49 - 1.06) | 0.11 | 0.76 (0.52 - 1.12) | 0.19 |
| SIDIAP | 2,851 | 1,941 | 3.9 | 4.1 | 40 | 39 | 0.66 (0.35 - 1.22) | 0.18 | 0.60 (0.32 - 1.11) | 0.11 |
| Meta-analysis | 318,854 | 968,647 | 227.2 | 870.9 | 934 | 7,618 | 0.71 (0.63 - 0.81) | <0.01 | 0.72 (0.60 - 0.87) | <0.01 |
| Meta-analysis (w/o USOC) | 47,336 | 143,934 | 26.1 | 96.6 | 217 | 1,543 | 0.78 (0.67 - 0.92) | <0.01 | 0.79 (0.67 - 0.93) | <0.01 |
| **PS-matching / on-treatment TAR without escalation** | | | | | | | | | | |
| CCAE | 15,821 | 34,604 | 10.0 | 21.2 | 55 | 176 | 0.49 (0.29 - 0.80) | <0.01 | 0.46 (0.28 - 0.77) | <0.01 |
| DAG | 2,974 | 640 | 2.1 | 0.6 | 22 | 8 | 0.65 (0.20 - 2.60) | 0.51 | 0.70 (0.19 - 2.57) | 0.58 |
| ODOD | 11,422 | 38,860 | 6.3 | 28.5 | 93 | 655 | 0.89 (0.66 - 1.19) | 0.44 | 0.90 (0.67 - 1.21) | 0.52 |
| MDCR | 1,299 | 6,566 | 0.8 | 4.8 | 5 | 81 | 0.67 (0.22 - 1.67) | 0.44 | 0.69 (0.25 - 1.88) | 0.47 |
| MDCD | 2,321 | 11,258 | 1.0 | 5.8 | 7 | 106 | 0.42 (0.16 - 0.96) | 0.06 | 0.40 (0.16 - 1.00) | 0.05 |
| Open Claims | 257,321 | 815,085 | 162.1 | 645.3 | 551 | 4,869 | 0.66 (0.59 - 0.75) | <0.01 | 0.68 (0.56 - 0.83) | <0.01 |
| OEHR | 15,234 | 63,024 | 4.1 | 30.3 | 26 | 352 | 0.99 (0.59 - 1.60) | 0.98 | 1.03 (0.62 - 1.69) | 0.89 |
| SIDIAP | 2,756 | 929 | 3.4 | 1.3 | 32 | 14 | 0.73 (0.28 - 1.91) | 0.52 | 0.94 (0.36 - 2.45) | 0.81 |
| Meta-analysis | 306,174 | 970,326 | 187.7 | 737.2 | 769 | 6,253 | 0.71 (0.58 - 0.87) | <0.01 | 0.71 (0.56 - 0.91) | <0.01 |
| Meta-analysis (w/o USOC) | 48,853 | 155,241 | 25.6 | 91.9 | 218 | 1,384 | 0.74 (0.56 - 0.97) | 0.03 | 0.72 (0.55 - 0.95) | 0.02 |
| **PS-stratification / on-treatment TAR without escalation** | | | | | | | | | | |
| CCAE | 18,560 | 35,219 | 11.5 | 21.4 | 67 | 179 | 0.75 (0.55 - 1.02) | 0.07 | 0.72 (0.53 - 0.98) | 0.04 |
| DAG | 3,022 | 1,171 | 2.2 | 1.1 | 23 | 10 | 0.60 (0.25 - 1.58) | 0.27 | 0.38 (0.15 - 0.97) | 0.05 |
| ODOD | 12,381 | 39,679 | 6.7 | 29.2 | 97 | 685 | 0.83 (0.65 - 1.04) | 0.11 | 0.86 (0.67 - 1.08) | 0.21 |
| MDCR | 1,451 | 7,008 | 0.8 | 5.1 | 5 | 90 | 0.38 (0.13 - 0.92) | 0.06 | 0.35 (0.13 - 0.96) | 0.04 |
| MDCD | 2,363 | 11,556 | 1.0 | 5.9 | 7 | 112 | 0.37 (0.15 - 0.78) | 0.02 | 0.34 (0.15 - 0.76) | <0.01 |
| Open Claims | 271,518 | 824,713 | 169.2 | 652.9 | 580 | 5,079 | 0.65 (0.59 - 0.71) | <0.01 | 0.66 (0.56 - 0.78) | <0.01 |
| OEHR | 16,395 | 69,036 | 4.5 | 33.3 | 26 | 410 | 0.74 (0.47 - 1.12) | 0.17 | 0.75 (0.49 - 1.15) | 0.22 |
| SIDIAP | 2,851 | 1,941 | 3.4 | 3.4 | 35 | 33 | 0.64 (0.32 - 1.26) | 0.20 | 0.65 (0.33 - 1.28) | 0.23 |
| Meta-analysis | 318,854 | 968,647 | 191.9 | 736.8 | 770 | 6,353 | 0.71 (0.62 - 0.82) | <0.01 | 0.71 (0.59 - 0.86) | <0.01 |
| Meta-analysis (w/o USOC) | 47,336 | 143,934 | 22.7 | 83.9 | 190 | 1,274 | 0.79 (0.67 - 0.94) | <0.01 | 0.78 (0.66 - 0.93) | <0.01 |

##### 1.3.18 Hypoglycemia

**Supplementary Table 70:** Exposures, outcomes and risk estimates for Hypoglycemia in SGLT2I (female) (T) vs SU (female) (C) new-user cohorts under four different propensity score (PS) adjustment / time-at-risk (TAR) designs across data sources. We report total patient exposure time in 1,000 patient-year (KYr) units, uncalibrated and calibrated hazard ratios (HRs) and their 95% confidence intervals (CIs). Greyed out entries did not pass study diagnostics and are included only for completeness.

| | Patients | | Exposure (KYr) | | Outcomes | | Uncalibrated | | Calibrated | |
|---|---|---|---|---|---|---|---|---|---|---|
| | T | C | T | C | T | C | HR (95% CI) | $p$ | HR (95% CI) | $p$ |
| **PS-matching / on-treatment TAR** | | | | | | | | | | |
| CCAE | 15,761 | 34,654 | 11.6 | 24.6 | 42 | 323 | 0.25 (0.15 - 0.39) | <0.01 | 0.24 (0.15 - 0.38) | <0.01 |
| DAG | 3,163 | 678 | 2.5 | 0.7 | <5 | 5 | 0.12 (0.01 - 0.91) | 0.10 | 0.14 (0.01 - 1.66) | 0.12 |
| ODOD | 11,617 | 40,283 | 7.2 | 33.4 | 47 | 920 | 0.34 (0.24 - 0.48) | <0.01 | 0.35 (0.25 - 0.50) | <0.01 |
| MDCR | 1,320 | 6,722 | 0.9 | 5.4 | 0 | 143 | | | | |
| MDCD | 2,312 | 11,360 | 1.2 | 7.0 | 12 | 243 | 0.38 (0.19 - 0.70) | <0.01 | 0.35 (0.18 - 0.67) | <0.01 |
| Open Claims | 256,842 | 818,296 | 192.7 | 771.9 | 398 | 8,212 | 0.21 (0.19 - 0.24) | <0.01 | 0.22 (0.18 - 0.27) | <0.01 |
| OEHR | 15,300 | 63,521 | 4.8 | 36.0 | 17 | 703 | 0.20 (0.10 - 0.33) | <0.01 | 0.20 (0.11 - 0.37) | <0.01 |
| SIDIAP | 2,843 | 962 | 4.0 | 1.7 | <5 | 7 | 0.03 (0.00 - 0.26) | 0.01 | 0.03 (0.00 - 0.52) | 0.02 |
| Meta-analysis | 301,832 | 968,114 | 217.6 | 872.9 | 516 | 10,401 | 0.26 (0.20 - 0.33) | <0.01 | 0.26 (0.19 - 0.34) | <0.01 |
| Meta-analysis (w/o USOC) | 44,990 | 149,818 | 24.9 | 101.0 | 118 | 2,189 | 0.29 (0.22 - 0.38) | <0.01 | 0.28 (0.22 - 0.37) | <0.01 |
| **PS-stratification / on-treatment TAR** | | | | | | | | | | |
| CCAE | 18,425 | 35,271 | 13.3 | 24.9 | 50 | 326 | 0.28 (0.20 - 0.38) | <0.01 | 0.28 (0.20 - 0.38) | <0.01 |
| DAG | 3,214 | 1,220 | 2.5 | 1.2 | <5 | 11 | 0.23 (0.04 - 1.00) | 0.06 | 0.15 (0.03 - 0.71) | 0.02 |
| ODOD | 12,580 | 41,160 | 7.7 | 34.2 | 50 | 959 | 0.31 (0.23 - 0.41) | <0.01 | 0.33 (0.24 - 0.44) | <0.01 |
| MDCR | 1,473 | 7,208 | 0.9 | 5.8 | 0 | 162 | | | | |
| MDCD | 2,349 | 11,604 | 1.3 | 7.2 | 12 | 248 | 0.32 (0.17 - 0.56) | <0.01 | 0.29 (0.16 - 0.54) | <0.01 |
| Open Claims | 270,936 | 828,559 | 200.8 | 781.5 | 416 | 8,484 | 0.24 (0.21 - 0.26) | <0.01 | 0.24 (0.20 - 0.29) | <0.01 |
| OEHR | 16,444 | 69,682 | 5.2 | 39.7 | 18 | 803 | 0.20 (0.12 - 0.31) | <0.01 | 0.21 (0.13 - 0.34) | <0.01 |
| SIDIAP | 2,944 | 2,010 | 4.0 | 4.3 | <5 | 20 | 0.02 (0.00 - 0.16) | <0.01 | 0.02 (0.00 - 0.25) | <0.01 |
| Meta-analysis | 318,385 | 974,672 | 226.9 | 880.3 | 534 | 10,572 | 0.25 (0.22 - 0.30) | <0.01 | 0.26 (0.21 - 0.31) | <0.01 |
| Meta-analysis (w/o USOC) | 47,449 | 146,113 | 26.1 | 98.7 | 118 | 2,088 | 0.28 (0.22 - 0.34) | <0.01 | 0.28 (0.22 - 0.35) | <0.01 |
| **PS-matching / on-treatment TAR without escalation** | | | | | | | | | | |
| CCAE | 15,761 | 34,654 | 10.0 | 21.2 | 30 | 264 | 0.22 (0.13 - 0.37) | <0.01 | 0.21 (0.12 - 0.36) | <0.01 |
| DAG | 3,163 | 678 | 2.3 | 0.6 | <5 | 5 | | | | |
| ODOD | 11,617 | 40,283 | 6.4 | 29.8 | 34 | 807 | 0.31 (0.20 - 0.45) | <0.01 | 0.31 (0.21 - 0.47) | <0.01 |
| MDCR | 1,320 | 6,722 | 0.8 | 4.9 | 0 | 128 | | | | |
| MDCD | 2,312 | 11,360 | 1.0 | 5.8 | 9 | 197 | 0.37 (0.16 - 0.75) | 0.01 | 0.35 (0.16 - 0.76) | <0.01 |
| Open Claims | 256,842 | 818,296 | 161.8 | 650.9 | 260 | 6,809 | 0.17 (0.14 - 0.20) | <0.01 | 0.17 (0.14 - 0.22) | <0.01 |
| OEHR | 15,300 | 63,521 | 4.2 | 30.6 | 8 | 522 | 0.13 (0.06 - 0.26) | <0.01 | 0.14 (0.06 - 0.30) | <0.01 |
| SIDIAP | 2,843 | 962 | 3.5 | 1.4 | <5 | 5 | 0.03 (0.00 - 0.41) | 0.03 | 0.04 (0.00 - 0.85) | 0.04 |
| Meta-analysis | 301,832 | 968,114 | 183.4 | 738.4 | 341 | 8,599 | 0.22 (0.16 - 0.31) | <0.01 | 0.22 (0.15 - 0.32) | <0.01 |
| Meta-analysis (w/o USOC) | 44,990 | 149,818 | 21.6 | 87.5 | 81 | 1,790 | 0.25 (0.17 - 0.36) | <0.01 | 0.25 (0.17 - 0.36) | <0.01 |
| **PS-stratification / on-treatment TAR without escalation** | | | | | | | | | | |
| CCAE | 18,425 | 35,271 | 11.5 | 21.4 | 37 | 264 | 0.25 (0.17 - 0.36) | <0.01 | 0.24 (0.17 - 0.35) | <0.01 |
| DAG | 3,214 | 1,220 | 2.3 | 1.1 | <5 | 11 | 0.16 (0.02 - 0.82) | 0.05 | 0.10 (0.02 - 0.65) | 0.02 |
| ODOD | 12,580 | 41,160 | 6.8 | 30.6 | 37 | 844 | 0.26 (0.18 - 0.36) | <0.01 | 0.27 (0.19 - 0.38) | <0.01 |
| MDCR | 1,473 | 7,208 | 0.8 | 5.3 | 0 | 145 | | | | |
| MDCD | 2,349 | 11,604 | 1.0 | 6.0 | 9 | 202 | 0.29 (0.14 - 0.55) | <0.01 | 0.26 (0.13 - 0.53) | <0.01 |
| Open Claims | 270,936 | 828,559 | 168.9 | 659.3 | 277 | 7,050 | 0.19 (0.17 - 0.21) | <0.01 | 0.19 (0.16 - 0.23) | <0.01 |
| OEHR | 16,444 | 69,682 | 4.5 | 33.7 | 9 | 590 | 0.12 (0.06 - 0.23) | <0.01 | 0.12 (0.06 - 0.24) | <0.01 |
| SIDIAP | 2,944 | 2,010 | 3.5 | 3.6 | <5 | 15 | 0.03 (0.00 - 0.24) | <0.01 | 0.03 (0.00 - 0.41) | <0.01 |
| Meta-analysis | 318,385 | 974,672 | 191.7 | 745.0 | 360 | 8,748 | 0.21 (0.17 - 0.26) | <0.01 | 0.21 (0.16 - 0.27) | <0.01 |
| Meta-analysis (w/o USOC) | 47,449 | 146,113 | 22.8 | 85.7 | 83 | 1,698 | 0.22 (0.16 - 0.32) | <0.01 | 0.22 (0.15 - 0.32) | <0.01 |

##### 1.3.19 Hypotension

**Supplementary Table 71:** Exposures, outcomes and risk estimates for Hypotension in SGLT2I (female) (T) vs SU (female) (C) new-user cohorts under four different propensity score (PS) adjustment / time-at-risk (TAR) designs across data sources. We report total patient exposure time in 1,000 patient-year (KYr) units, uncalibrated and calibrated hazard ratios (HRs) and their 95% confidence intervals (CIs). Greyed out entries did not pass study diagnostics and are included only for completeness.

| | Patients | | Exposure (KYr) | | Outcomes | | Uncalibrated | | Calibrated | |
|---|---|---|---|---|---|---|---|---|---|---|
| | T | C | T | C | T | C | HR (95% CI) | $p$ | HR (95% CI) | $p$ |
| **PS-matching / on-treatment TAR** | | | | | | | | | | |
| CCAE | 15,744 | 34,322 | 11.6 | 24.5 | 90 | 187 | 1.38 (0.96 - 1.97) | 0.08 | 1.31 (0.92 - 1.89) | 0.15 |
| DAG | 3,055 | 659 | 2.4 | 0.6 | 7 | <5 | 0.82 (0.09 - 18.8) | 0.89 | 0.91 (0.06 - 12.9) | 0.86 |
| ODOD | 11,340 | 38,641 | 7.0 | 31.9 | 141 | 724 | 1.27 (0.99 - 1.61) | 0.06 | 1.31 (1.02 - 1.67) | 0.04 |
| MDCR | 1,279 | 6,447 | 0.8 | 5.2 | 23 | 116 | 1.70 (0.90 - 3.08) | 0.09 | 1.77 (0.96 - 3.26) | 0.08 |
| MDCD | 2,228 | 10,689 | 1.2 | 6.6 | 42 | 177 | 1.26 (0.75 - 2.05) | 0.36 | 1.16 (0.70 - 1.91) | 0.58 |
| Open Claims | 253,429 | 803,417 | 190.2 | 756.9 | 1,208 | 6,608 | 1.04 (0.96 - 1.13) | 0.31 | 1.07 (0.88 - 1.29) | 0.50 |
| OEHR | 15,130 | 62,599 | 4.7 | 35.4 | 56 | 519 | 1.43 (0.97 - 2.06) | 0.06 | 1.49 (1.01 - 2.19) | 0.08 |
| SIDIAP | 2,828 | 957 | 3.9 | 1.7 | 8 | 5 | 0.28 (0.03 - 2.07) | 0.24 | 0.33 (0.04 - 2.77) | 0.31 |
| Meta-analysis | 299,150 | 956,115 | 215.5 | 860.5 | 1,560 | 8,331 | 1.23 (1.05 - 1.44) | 0.01 | 1.22 (0.98 - 1.52) | 0.07 |
| Meta-analysis (w/o USOC) | 45,721 | 152,698 | 25.4 | 103.5 | 352 | 1,723 | 1.34 (1.14 - 1.58) | <0.01 | 1.32 (1.12 - 1.56) | <0.01 |
| **PS-stratification / on-treatment TAR** | | | | | | | | | | |
| CCAE | 18,471 | 35,027 | 13.3 | 24.7 | 110 | 194 | 1.17 (0.90 - 1.52) | 0.23 | 1.14 (0.88 - 1.49) | 0.34 |
| DAG | 3,105 | 1,187 | 2.4 | 1.2 | 7 | <5 | 1.06 (0.17 - 6.57) | 0.95 | 0.69 (0.11 - 4.28) | 0.69 |
| ODOD | 12,292 | 39,562 | 7.5 | 32.8 | 148 | 771 | 1.23 (1.01 - 1.50) | 0.04 | 1.29 (1.06 - 1.58) | 0.02 |
| MDCR | 1,425 | 6,870 | 0.9 | 5.5 | 25 | 131 | 1.71 (1.03 - 2.74) | 0.03 | 1.68 (1.03 - 2.74) | 0.05 |
| MDCD | 2,263 | 11,080 | 1.2 | 6.9 | 42 | 188 | 1.48 (1.00 - 2.13) | 0.04 | 1.36 (0.93 - 1.98) | 0.12 |
| Open Claims | 267,171 | 813,252 | 198.1 | 766.1 | 1,308 | 6,875 | 1.00 (0.93 - 1.06) | 0.94 | 1.02 (0.86 - 1.21) | 0.80 |
| OEHR | 16,280 | 68,740 | 5.1 | 39.0 | 60 | 601 | 1.28 (0.95 - 1.70) | 0.10 | 1.33 (0.99 - 1.79) | 0.07 |
| SIDIAP | 2,929 | 2,001 | 4.0 | 4.3 | 8 | 8 | 0.47 (0.10 - 1.96) | 0.32 | 0.43 (0.10 - 1.89) | 0.26 |
| Meta-analysis | 314,214 | 956,581 | 224.0 | 862.6 | 1,626 | 8,441 | 1.12 (0.98 - 1.29) | 0.10 | 1.13 (0.93 - 1.38) | 0.22 |
| Meta-analysis (w/o USOC) | 47,043 | 143,329 | 25.9 | 96.5 | 318 | 1,566 | 1.23 (1.07 - 1.41) | <0.01 | 1.24 (1.07 - 1.43) | <0.01 |
| **PS-matching / on-treatment TAR without escalation** | | | | | | | | | | |
| CCAE | 15,744 | 34,322 | 10.0 | 21.1 | 68 | 151 | 1.26 (0.84 - 1.87) | 0.26 | 1.19 (0.80 - 1.78) | 0.41 |
| DAG | 3,055 | 659 | 2.2 | 0.6 | 7 | <5 | 0.83 (0.09 - 18.9) | 0.89 | 0.90 (0.06 - 12.7) | 0.85 |
| ODOD | 11,340 | 38,641 | 6.2 | 28.5 | 119 | 638 | 1.23 (0.95 - 1.58) | 0.12 | 1.24 (0.96 - 1.61) | 0.12 |
| MDCR | 1,279 | 6,447 | 0.7 | 4.7 | 23 | 107 | 1.77 (0.89 - 3.32) | 0.09 | 1.80 (0.93 - 3.48) | 0.09 |
| MDCD | 2,228 | 10,689 | 1.0 | 5.5 | 32 | 136 | 1.23 (0.69 - 2.12) | 0.46 | 1.18 (0.67 - 2.06) | 0.58 |
| Open Claims | 253,429 | 803,417 | 159.8 | 638.0 | 975 | 5,490 | 1.05 (0.96 - 1.15) | 0.28 | 1.08 (0.90 - 1.30) | 0.44 |
| OEHR | 15,130 | 62,599 | 4.1 | 30.1 | 39 | 370 | 1.07 (0.66 - 1.66) | 0.79 | 1.10 (0.69 - 1.75) | 0.70 |
| SIDIAP | 2,828 | 957 | 3.5 | 1.4 | 7 | <5 | 0.29 (0.03 - 2.08) | 0.25 | 0.37 (0.04 - 3.04) | 0.36 |
| Meta-analysis | 299,150 | 956,115 | 181.8 | 727.9 | 1,256 | 6,892 | 1.12 (1.00 - 1.26) | 0.05 | 1.13 (0.93 - 1.35) | 0.21 |
| Meta-analysis (w/o USOC) | 45,721 | 152,698 | 22.0 | 89.9 | 281 | 1,402 | 1.24 (1.04 - 1.48) | 0.02 | 1.22 (1.01 - 1.46) | 0.05 |
| **PS-stratification / on-treatment TAR without escalation** | | | | | | | | | | |
| CCAE | 18,471 | 35,027 | 11.5 | 21.3 | 84 | 157 | 1.10 (0.82 - 1.48) | 0.51 | 1.05 (0.78 - 1.42) | 0.74 |
| DAG | 3,105 | 1,187 | 2.2 | 1.1 | 7 | <5 | 0.95 (0.15 - 6.02) | 0.95 | 0.61 (0.10 - 3.85) | 0.60 |
| ODOD | 12,292 | 39,562 | 6.6 | 29.3 | 126 | 680 | 1.20 (0.97 - 1.48) | 0.09 | 1.24 (1.00 - 1.54) | 0.05 |
| MDCR | 1,425 | 6,870 | 0.8 | 5.0 | 24 | 120 | 2.07 (1.23 - 3.35) | <0.01 | 1.92 (1.17 - 3.17) | 0.01 |
| MDCD | 2,263 | 11,080 | 1.0 | 5.7 | 32 | 144 | 1.49 (0.96 - 2.27) | 0.07 | 1.35 (0.87 - 2.08) | 0.19 |
| Open Claims | 267,171 | 813,252 | 166.7 | 646.0 | 1,066 | 5,725 | 0.99 (0.92 - 1.06) | 0.71 | 1.01 (0.86 - 1.17) | 0.90 |
| OEHR | 16,280 | 68,740 | 4.4 | 33.2 | 42 | 427 | 1.24 (0.86 - 1.74) | 0.23 | 1.25 (0.88 - 1.78) | 0.26 |
| SIDIAP | 2,929 | 2,001 | 3.5 | 3.6 | 7 | 7 | 0.54 (0.11 - 2.40) | 0.43 | 0.55 (0.12 - 2.54) | 0.44 |
| Meta-analysis | 314,214 | 956,581 | 189.3 | 729.8 | 1,318 | 6,989 | 1.08 (0.95 - 1.23) | 0.27 | 1.08 (0.91 - 1.28) | 0.40 |
| Meta-analysis (w/o USOC) | 47,043 | 143,329 | 22.6 | 83.8 | 252 | 1,264 | 1.18 (1.01 - 1.38) | 0.04 | 1.17 (1.00 - 1.37) | 0.07 |

##### 1.3.20 Joint pain

**Supplementary Table 72:** Exposures, outcomes and risk estimates for Joint pain in SGLT2I (female) (T) vs SU (female) (C) new-user cohorts under four different propensity score (PS) adjustment / time-at-risk (TAR) designs across data sources. We report total patient exposure time in 1,000 patient-year (KYr) units, uncalibrated and calibrated hazard ratios (HRs) and their 95% confidence intervals (CIs). Greyed out entries did not pass study diagnostics and are included only for completeness.

| | Patients | | Exposure (KYr) | | Outcomes | | Uncalibrated | | Calibrated | |
|---|---|---|---|---|---|---|---|---|---|---|
| | T | C | T | C | T | C | HR (95% CI) | $p$ | HR (95% CI) | $p$ |
| **PS-matching / on-treatment TAR** | | | | | | | | | | |
| CCAE | 14,820 | 32,693 | 10.9 | 23.2 | 140 | 354 | 0.82 (0.61 - 1.10) | 0.18 | 0.78 (0.58 - 1.05) | 0.12 |
| DAG | 2,618 | 573 | 2.0 | 0.5 | 45 | 16 | 0.91 (0.42 - 2.10) | 0.82 | 1.00 (0.45 - 2.25) | 0.61 |
| ODOD | 10,889 | 37,908 | 6.8 | 31.5 | 119 | 552 | 1.06 (0.80 - 1.39) | 0.67 | 1.09 (0.83 - 1.44) | 0.54 |
| MDCR | 1,265 | 6,408 | 0.8 | 5.2 | 12 | 55 | 1.32 (0.48 - 3.33) | 0.58 | 1.36 (0.51 - 3.61) | 0.54 |
| MDCD | 2,114 | 10,446 | 1.1 | 6.4 | 31 | 155 | 0.91 (0.50 - 1.59) | 0.76 | 0.84 (0.47 - 1.49) | 0.55 |
| Open Claims | 240,761 | 775,933 | 180.9 | 732.9 | 1,373 | 6,030 | 0.89 (0.81 - 0.96) | <0.01 | 0.91 (0.75 - 1.09) | 0.32 |
| OEHR | 14,340 | 60,148 | 4.4 | 33.8 | 76 | 588 | 0.98 (0.69 - 1.37) | 0.91 | 1.02 (0.72 - 1.46) | 0.87 |
| SIDIAP | 2,038 | 728 | 2.8 | 1.2 | 57 | 40 | 0.55 (0.30 - 1.01) | 0.06 | 0.65 (0.35 - 1.19) | 0.18 |
| Meta-analysis | 286,227 | 924,264 | 207.8 | 834.1 | 1,808 | 7,774 | 0.89 (0.83 - 0.96) | <0.01 | 0.89 (0.75 - 1.05) | 0.18 |
| Meta-analysis (w/o USOC) | 45,466 | 148,331 | 26.9 | 101.2 | 435 | 1,744 | 0.92 (0.79 - 1.08) | 0.31 | 0.91 (0.77 - 1.06) | 0.26 |
| **PS-stratification / on-treatment TAR** | | | | | | | | | | |
| CCAE | 17,271 | 33,274 | 12.5 | 23.4 | 175 | 355 | 0.82 (0.67 - 1.00) | 0.05 | 0.80 (0.65 - 0.98) | 0.04 |
| DAG | 2,660 | 1,030 | 2.0 | 1.0 | 46 | 31 | 0.91 (0.49 - 1.70) | 0.76 | 0.59 (0.32 - 1.10) | 0.12 |
| ODOD | 11,750 | 38,689 | 7.2 | 32.2 | 129 | 565 | 0.93 (0.75 - 1.15) | 0.52 | 0.98 (0.79 - 1.21) | 0.84 |
| MDCR | 1,412 | 6,861 | 0.9 | 5.5 | 14 | 60 | 1.09 (0.54 - 2.10) | 0.80 | 1.07 (0.54 - 2.12) | 0.84 |
| MDCD | 2,157 | 10,726 | 1.2 | 6.6 | 31 | 160 | 1.21 (0.79 - 1.80) | 0.36 | 1.11 (0.74 - 1.68) | 0.62 |
| Open Claims | 253,928 | 785,790 | 188.5 | 742.0 | 1,465 | 6,137 | 0.90 (0.85 - 0.96) | <0.01 | 0.93 (0.78 - 1.09) | 0.38 |
| OEHR | 15,387 | 66,105 | 4.8 | 37.3 | 80 | 643 | 0.92 (0.71 - 1.18) | 0.54 | 0.96 (0.74 - 1.25) | 0.79 |
| SIDIAP | 2,108 | 1,550 | 2.9 | 3.2 | 59 | 96 | 0.67 (0.44 - 1.02) | 0.07 | 0.61 (0.40 - 0.94) | 0.03 |
| Meta-analysis | 298,336 | 923,858 | 212.9 | 835.0 | 1,849 | 7,700 | 0.90 (0.85 - 0.95) | <0.01 | 0.91 (0.78 - 1.05) | 0.20 |
| Meta-analysis (w/o USOC) | 44,408 | 138,068 | 24.4 | 93.0 | 384 | 1,563 | 0.88 (0.78 - 1.00) | 0.05 | 0.89 (0.78 - 1.02) | 0.12 |
| **PS-matching / on-treatment TAR without escalation** | | | | | | | | | | |
| CCAE | 14,820 | 32,693 | 9.4 | 20.0 | 118 | 313 | 0.82 (0.60 - 1.12) | 0.22 | 0.77 (0.56 - 1.06) | 0.12 |
| DAG | 2,618 | 573 | 1.8 | 0.5 | 43 | 15 | 0.97 (0.45 - 2.24) | 0.94 | 1.05 (0.47 - 2.36) | 0.58 |
| ODOD | 10,889 | 37,908 | 6.0 | 28.2 | 103 | 494 | 1.14 (0.85 - 1.53) | 0.37 | 1.16 (0.86 - 1.56) | 0.35 |
| MDCR | 1,265 | 6,408 | 0.7 | 4.7 | 11 | 48 | 1.21 (0.40 - 3.26) | 0.72 | 1.23 (0.43 - 3.51) | 0.70 |
| MDCD | 2,114 | 10,446 | 0.9 | 5.3 | 28 | 127 | 0.85 (0.43 - 1.58) | 0.62 | 0.81 (0.42 - 1.55) | 0.53 |
| Open Claims | 240,761 | 775,933 | 152.0 | 618.5 | 1,151 | 5,053 | 0.88 (0.80 - 0.96) | <0.01 | 0.90 (0.75 - 1.08) | 0.27 |
| OEHR | 14,340 | 60,148 | 3.9 | 28.8 | 69 | 473 | 1.01 (0.70 - 1.43) | 0.97 | 1.04 (0.72 - 1.50) | 0.83 |
| SIDIAP | 2,038 | 728 | 2.5 | 1.0 | 43 | 35 | 0.49 (0.24 - 0.98) | 0.05 | 0.63 (0.31 - 1.27) | 0.21 |
| Meta-analysis | 286,227 | 924,264 | 175.4 | 706.5 | 1,523 | 6,543 | 0.89 (0.82 - 0.97) | <0.01 | 0.89 (0.76 - 1.05) | 0.18 |
| Meta-analysis (w/o USOC) | 45,466 | 148,331 | 23.4 | 88.0 | 372 | 1,490 | 0.93 (0.76 - 1.14) | 0.47 | 0.91 (0.74 - 1.12) | 0.38 |
| **PS-stratification / on-treatment TAR without escalation** | | | | | | | | | | |
| CCAE | 17,271 | 33,274 | 10.8 | 20.2 | 150 | 314 | 0.79 (0.64 - 0.98) | 0.03 | 0.75 (0.61 - 0.94) | 0.01 |
| DAG | 2,660 | 1,030 | 1.8 | 0.9 | 44 | 29 | 0.89 (0.48 - 1.70) | 0.73 | 0.57 (0.30 - 1.09) | 0.11 |
| ODOD | 11,750 | 38,689 | 6.3 | 28.9 | 113 | 507 | 0.90 (0.72 - 1.12) | 0.36 | 0.93 (0.74 - 1.17) | 0.55 |
| MDCR | 1,412 | 6,861 | 0.8 | 5.0 | 13 | 53 | 1.13 (0.53 - 2.26) | 0.75 | 1.05 (0.51 - 2.16) | 0.88 |
| MDCD | 2,157 | 10,726 | 1.0 | 5.5 | 28 | 131 | 1.29 (0.82 - 1.97) | 0.26 | 1.16 (0.75 - 1.81) | 0.51 |
| Open Claims | 253,928 | 785,790 | 158.6 | 626.4 | 1,233 | 5,146 | 0.91 (0.85 - 0.98) | <0.01 | 0.93 (0.80 - 1.08) | 0.36 |
| OEHR | 15,387 | 66,105 | 4.2 | 31.8 | 72 | 518 | 1.01 (0.77 - 1.32) | 0.93 | 1.02 (0.77 - 1.35) | 0.85 |
| SIDIAP | 2,108 | 1,550 | 2.5 | 2.7 | 45 | 81 | 0.57 (0.35 - 0.91) | 0.02 | 0.58 (0.36 - 0.93) | 0.03 |
| Meta-analysis | 298,336 | 923,858 | 179.9 | 707.2 | 1,568 | 6,485 | 0.91 (0.85 - 0.96) | <0.01 | 0.91 (0.80 - 1.03) | 0.14 |
| Meta-analysis (w/o USOC) | 44,408 | 138,068 | 21.3 | 80.8 | 335 | 1,339 | 0.88 (0.77 - 1.01) | 0.07 | 0.87 (0.76 - 1.00) | 0.07 |

##### 1.3.21 Lower extremity amputation

**Supplementary Table 73:** Exposures, outcomes and risk estimates for Lower extremity amputation in SGLT2I (female) (T) vs SU (female) (C) new-user cohorts under four different propensity score (PS) adjustment / time-at-risk (TAR) designs across data sources. We report total patient exposure time in 1,000 patient-year (KYr) units, uncalibrated and calibrated hazard ratios (HRs) and their 95% confidence intervals (CIs). Greyed out entries did not pass study diagnostics and are included only for completeness.

| | Patients | | Exposure (KYr) | | Outcomes | | Uncalibrated | | Calibrated | |
|---|---|---|---|---|---|---|---|---|---|---|
| | T | C | T | C | T | C | HR (95% CI) | $p$ | HR (95% CI) | $p$ |
| **PS-matching / on-treatment TAR** | | | | | | | | | | |
| CCAE | 16,075 | 35,099 | 11.9 | 25.1 | <5 | 5 | 2.00 (0.22 - 18.3) | 0.54 | 1.91 (0.21 - 17.5) | 0.57 |
| DAG | 3,180 | 682 | 2.5 | 0.7 | 0 | 0 | | | | |
| ODOD | 11,831 | 40,935 | 7.4 | 34.3 | 5 | 22 | 1.86 (0.45 - 6.48) | 0.36 | 1.91 (0.50 - 7.28) | 0.34 |
| MDCR | 1,330 | 6,790 | 0.9 | 5.5 | <5 | <5 | | 0.34 | | 0.34 |
| MDCD | 2,384 | 11,649 | 1.3 | 7.3 | 0 | 7 | | | | |
| Open Claims | 261,706 | 831,984 | 196.3 | 788.1 | 39 | 313 | 0.88 (0.54 - 1.37) | 0.58 | 0.90 (0.55 - 1.47) | 0.67 |
| OEHR | 15,481 | 64,204 | 4.9 | 36.8 | <5 | 21 | 0.48 (0.02 - 3.09) | 0.56 | 0.50 (0.04 - 5.66) | 0.58 |
| SIDIAP | 2,849 | 964 | 4.0 | 1.7 | 0 | 0 | | | | |
| Meta-analysis | 273,537 | 872,919 | 203.7 | 822.3 | 44 | 335 | 0.97 (0.59 - 1.61) | 0.91 | 0.97 (0.57 - 1.64) | 0.91 |
| Meta-analysis (w/o USOC) | 11,831 | 40,935 | 7.4 | 34.3 | 5 | 22 | 1.86 (0.45 - 6.48) | 0.36 | 1.82 (0.48 - 6.94) | 0.38 |
| **PS-stratification / on-treatment TAR** | | | | | | | | | | |
| CCAE | 18,874 | 35,794 | 13.7 | 25.4 | 5 | 5 | 3.28 (0.85 - 12.3) | 0.08 | 3.19 (0.84 - 12.2) | 0.09 |
| DAG | 3,231 | 1,228 | 2.5 | 1.2 | 0 | 0 | | | | |
| ODOD | 12,831 | 41,835 | 7.9 | 35.2 | 5 | 24 | 1.77 (0.52 - 5.08) | 0.33 | 1.86 (0.59 - 5.83) | 0.29 |
| MDCR | 1,486 | 7,280 | 0.9 | 5.9 | <5 | <5 | | 0.10 | | 0.10 |
| MDCD | 2,431 | 11,911 | 1.3 | 7.5 | 0 | 7 | | | | |
| Open Claims | 276,304 | 842,001 | 204.7 | 797.4 | 39 | 323 | 0.86 (0.59 - 1.21) | 0.40 | 0.88 (0.60 - 1.30) | 0.52 |
| OEHR | 16,654 | 70,443 | 5.2 | 40.6 | <5 | 24 | 1.14 (0.17 - 4.27) | 0.87 | 1.19 (0.24 - 5.90) | 0.83 |
| SIDIAP | 2,950 | 2,011 | 4.0 | 4.3 | 0 | 0 | | | | |
| Meta-analysis | 305,789 | 954,279 | 217.8 | 873.1 | <49 | 371 | 0.95 (0.64 - 1.39) | 0.78 | 0.95 (0.63 - 1.44) | 0.82 |
| Meta-analysis (w/o USOC) | 29,485 | 112,278 | 13.1 | 75.7 | <10 | 48 | 1.53 (0.60 - 3.87) | 0.37 | 1.55 (0.61 - 3.92) | 0.36 |
| **PS-matching / on-treatment TAR without escalation** | | | | | | | | | | |
| CCAE | 16,075 | 35,099 | 10.2 | 21.6 | <5 | 5 | 1.09 (0.05 - 12.4) | 0.95 | 1.02 (0.06 - 16.6) | 0.98 |
| DAG | 3,180 | 682 | 2.3 | 0.6 | 0 | 0 | | | | |
| ODOD | 11,831 | 40,935 | 6.5 | 30.6 | <5 | 18 | 1.75 (0.34 - 7.12) | 0.47 | 1.78 (0.39 - 8.13) | 0.46 |
| MDCR | 1,330 | 6,790 | 0.8 | 5.0 | <5 | <5 | | | | |
| MDCD | 2,384 | 11,649 | 1.1 | 6.0 | 0 | <5 | | | | |
| Open Claims | 261,706 | 831,984 | 164.8 | 663.6 | 27 | 239 | 0.81 (0.47 - 1.35) | 0.44 | 0.83 (0.48 - 1.45) | 0.51 |
| OEHR | 15,481 | 64,204 | 4.2 | 31.2 | <5 | 13 | 0.00 (0.00 - 2.27) | 0.23 | 0.00 (0.00 - 252124) | 0.24 |
| SIDIAP | 2,849 | 964 | 3.5 | 1.4 | 0 | 0 | | | | |
| Meta-analysis | 261,706 | 831,984 | 164.8 | 663.6 | 27 | 239 | 0.81 (0.47 - 1.35) | 0.44 | 0.81 (0.47 - 1.41) | 0.46 |
| **PS-stratification / on-treatment TAR without escalation** | | | | | | | | | | |
| CCAE | 18,874 | 35,794 | 11.8 | 21.8 | <5 | 5 | 2.45 (0.56 - 9.87) | 0.22 | 2.34 (0.56 - 9.87) | 0.25 |
| DAG | 3,231 | 1,228 | 2.3 | 1.2 | 0 | 0 | | | | |
| ODOD | 12,831 | 41,835 | 7.0 | 31.4 | <5 | 20 | 1.08 (0.22 - 3.83) | 0.92 | 1.11 (0.27 - 4.66) | 0.88 |
| MDCR | 1,486 | 7,280 | 0.8 | 5.4 | <5 | <5 | | 0.09 | | 0.09 |
| MDCD | 2,431 | 11,911 | 1.1 | 6.2 | 0 | <5 | | | | |
| Open Claims | 276,304 | 842,001 | 172.1 | 671.7 | 27 | 248 | 0.85 (0.54 - 1.27) | 0.44 | 0.86 (0.55 - 1.34) | 0.51 |
| OEHR | 16,654 | 70,443 | 4.6 | 34.3 | <5 | 16 | 0.91 (0.05 - 5.53) | 0.94 | 0.92 (0.08 - 9.96) | 0.94 |
| SIDIAP | 2,950 | 2,011 | 3.5 | 3.6 | 0 | 0 | | | | |
| Meta-analysis | 289,135 | 883,836 | 179.1 | 703.1 | <32 | 268 | 0.86 (0.58 - 1.29) | 0.47 | 0.86 (0.57 - 1.31) | 0.49 |
| Meta-analysis (w/o USOC) | 12,831 | 41,835 | 7.0 | 31.4 | <5 | 20 | 1.08 (0.22 - 3.83) | 0.92 | 1.06 (0.25 - 4.46) | 0.93 |

##### 1.3.22 Nausea

**Supplementary Table 74:** Exposures, outcomes and risk estimates for Nausea in SGLT2I (female) (T) vs SU (female) (C) new-user cohorts under four different propensity score (PS) adjustment / time-at-risk (TAR) designs across data sources. We report total patient exposure time in 1,000 patient-year (KYr) units, uncalibrated and calibrated hazard ratios (HRs) and their 95% confidence intervals (CIs). Greyed out entries did not pass study diagnostics and are included only for completeness.

| | Patients | | Exposure (KYr) | | Outcomes | | Uncalibrated | | Calibrated | |
|---|---|---|---|---|---|---|---|---|---|---|
| | T | C | T | C | T | C | HR (95% CI) | $p$ | HR (95% CI) | $p$ |
| **PS-matching / on-treatment TAR** | | | | | | | | | | |
| CCAE | 13,066 | 28,698 | 9.5 | 20.3 | 435 | 1,000 | 0.96 (0.82 - 1.13) | 0.63 | 0.92 (0.78 - 1.08) | 0.35 |
| DAG | 2,817 | 604 | 2.2 | 0.6 | 27 | 6 | 2.88 (0.74 - 19.1) | 0.20 | 3.17 (0.62 - 16.1) | 0.17 |
| ODOD | 9,424 | 32,387 | 5.8 | 26.5 | 296 | 1,541 | 0.99 (0.84 - 1.18) | 0.95 | 1.03 (0.87 - 1.22) | 0.78 |
| MDCR | 1,133 | 5,621 | 0.7 | 4.4 | 31 | 212 | 0.86 (0.49 - 1.44) | 0.58 | 0.89 (0.52 - 1.53) | 0.68 |
| MDCD | 1,514 | 7,446 | 0.8 | 4.6 | 78 | 483 | 0.70 (0.50 - 0.97) | 0.04 | 0.64 (0.46 - 0.90) | 0.01 |
| Open Claims | 210,078 | 681,648 | 158.9 | 643.8 | 3,770 | 18,261 | 0.92 (0.88 - 0.96) | <0.01 | 0.94 (0.79 - 1.12) | 0.50 |
| OEHR | 13,014 | 54,321 | 3.9 | 29.7 | 215 | 1,686 | 1.08 (0.90 - 1.30) | 0.40 | 1.13 (0.92 - 1.39) | 0.44 |
| SIDIAP | 2,664 | 897 | 3.7 | 1.6 | 20 | 12 | 0.34 (0.10 - 1.15) | 0.09 | 0.40 (0.12 - 1.39) | 0.16 |
| Meta-analysis | 250,893 | 811,018 | 183.4 | 730.9 | 4,845 | 23,195 | 0.93 (0.89 - 0.98) | <0.01 | 0.93 (0.79 - 1.09) | 0.37 |
| Meta-analysis (w/o USOC) | 40,815 | 129,370 | 24.5 | 87.0 | 1,075 | 4,934 | 0.95 (0.83 - 1.08) | 0.41 | 0.93 (0.81 - 1.07) | 0.33 |
| **PS-stratification / on-treatment TAR** | | | | | | | | | | |
| CCAE | 15,258 | 29,286 | 10.9 | 20.5 | 515 | 1,013 | 0.98 (0.87 - 1.10) | 0.73 | 0.95 (0.85 - 1.07) | 0.50 |
| DAG | 2,864 | 1,091 | 2.2 | 1.1 | 27 | 15 | 1.11 (0.48 - 2.67) | 0.81 | 0.72 (0.31 - 1.70) | 0.48 |
| ODOD | 10,200 | 33,096 | 6.2 | 27.2 | 321 | 1,590 | 0.91 (0.80 - 1.04) | 0.17 | 0.96 (0.84 - 1.09) | 0.56 |
| MDCR | 1,258 | 5,994 | 0.8 | 4.7 | 32 | 233 | 0.80 (0.52 - 1.19) | 0.28 | 0.78 (0.52 - 1.19) | 0.28 |
| MDCD | 1,543 | 7,617 | 0.8 | 4.7 | 80 | 494 | 0.89 (0.69 - 1.14) | 0.37 | 0.82 (0.64 - 1.06) | 0.15 |
| Open Claims | 221,452 | 690,768 | 165.5 | 652.4 | 3,960 | 18,680 | 0.89 (0.86 - 0.92) | <0.01 | 0.91 (0.78 - 1.07) | 0.26 |
| OEHR | 14,004 | 59,772 | 4.3 | 32.8 | 230 | 1,880 | 1.08 (0.93 - 1.25) | 0.32 | 1.13 (0.96 - 1.32) | 0.21 |
| SIDIAP | 2,761 | 1,897 | 3.7 | 4.1 | 21 | 41 | 0.55 (0.27 - 1.08) | 0.10 | 0.50 (0.25 - 1.01) | 0.06 |
| Meta-analysis | 260,914 | 812,922 | 186.8 | 732.8 | 5,026 | 23,163 | 0.95 (0.87 - 1.03) | 0.18 | 0.95 (0.81 - 1.12) | 0.56 |
| Meta-analysis (w/o USOC) | 39,462 | 122,154 | 21.4 | 80.5 | 1,066 | 4,483 | 0.98 (0.90 - 1.07) | 0.70 | 0.99 (0.90 - 1.10) | 0.84 |
| **PS-matching / on-treatment TAR without escalation** | | | | | | | | | | |
| CCAE | 13,066 | 28,698 | 8.2 | 17.6 | 383 | 845 | 0.99 (0.84 - 1.18) | 0.93 | 0.94 (0.79 - 1.11) | 0.49 |
| DAG | 2,817 | 604 | 2.0 | 0.5 | 23 | 6 | 2.80 (0.71 - 18.6) | 0.22 | 3.03 (0.59 - 15.5) | 0.19 |
| ODOD | 9,424 | 32,387 | 5.2 | 23.8 | 257 | 1,362 | 1.00 (0.83 - 1.19) | 0.98 | 1.01 (0.84 - 1.21) | 0.84 |
| MDCR | 1,133 | 5,621 | 0.6 | 4.0 | 28 | 192 | 1.05 (0.59 - 1.78) | 0.86 | 1.07 (0.62 - 1.86) | 0.79 |
| MDCD | 1,514 | 7,446 | 0.7 | 3.9 | 63 | 411 | 0.77 (0.53 - 1.09) | 0.15 | 0.73 (0.51 - 1.05) | 0.10 |
| Open Claims | 210,078 | 681,648 | 134.4 | 546.6 | 3,072 | 15,114 | 0.91 (0.87 - 0.96) | <0.01 | 0.94 (0.79 - 1.11) | 0.46 |
| OEHR | 13,014 | 54,321 | 3.5 | 25.6 | 179 | 1,333 | 1.15 (0.93 - 1.41) | 0.18 | 1.19 (0.96 - 1.47) | 0.16 |
| SIDIAP | 2,664 | 897 | 3.2 | 1.3 | 16 | 10 | 0.41 (0.11 - 1.46) | 0.17 | 0.52 (0.14 - 1.91) | 0.33 |
| Meta-analysis | 250,893 | 811,018 | 156.0 | 622.9 | 3,998 | 19,267 | 0.96 (0.88 - 1.04) | 0.32 | 0.96 (0.81 - 1.13) | 0.64 |
| Meta-analysis (w/o USOC) | 40,815 | 129,370 | 21.5 | 76.3 | 926 | 4,153 | 1.00 (0.89 - 1.12) | 0.98 | 0.98 (0.87 - 1.11) | 0.75 |
| **PS-stratification / on-treatment TAR without escalation** | | | | | | | | | | |
| CCAE | 15,258 | 29,286 | 9.5 | 17.8 | 450 | 858 | 1.02 (0.90 - 1.16) | 0.77 | 0.97 (0.85 - 1.11) | 0.71 |
| DAG | 2,864 | 1,091 | 2.0 | 1.0 | 23 | 14 | 1.18 (0.51 - 2.88) | 0.70 | 0.76 (0.32 - 1.82) | 0.55 |
| ODOD | 10,200 | 33,096 | 5.5 | 24.4 | 277 | 1,408 | 0.90 (0.78 - 1.04) | 0.16 | 0.93 (0.81 - 1.08) | 0.39 |
| MDCR | 1,258 | 5,994 | 0.7 | 4.3 | 29 | 210 | 0.84 (0.54 - 1.28) | 0.44 | 0.79 (0.51 - 1.21) | 0.30 |
| MDCD | 1,543 | 7,617 | 0.7 | 4.0 | 65 | 420 | 0.85 (0.64 - 1.12) | 0.26 | 0.77 (0.58 - 1.02) | 0.08 |
| Open Claims | 221,452 | 690,768 | 140.3 | 554.1 | 3,228 | 15,476 | 0.89 (0.85 - 0.93) | <0.01 | 0.91 (0.79 - 1.04) | 0.18 |
| OEHR | 14,004 | 59,772 | 3.8 | 28.3 | 190 | 1,496 | 1.10 (0.93 - 1.29) | 0.27 | 1.11 (0.93 - 1.32) | 0.37 |
| SIDIAP | 2,761 | 1,897 | 3.3 | 3.4 | 16 | 34 | 0.45 (0.19 - 0.99) | 0.06 | 0.45 (0.20 - 1.04) | 0.07 |
| Meta-analysis | 260,914 | 812,922 | 159.1 | 624.7 | 4,145 | 19,238 | 0.96 (0.87 - 1.05) | 0.37 | 0.96 (0.82 - 1.11) | 0.57 |
| Meta-analysis (w/o USOC) | 39,462 | 122,154 | 18.8 | 70.6 | 917 | 3,762 | 1.00 (0.90 - 1.11) | 0.98 | 0.99 (0.88 - 1.11) | 0.82 |

##### 1.3.23 Peripheral edema

**Supplementary Table 75:** Exposures, outcomes and risk estimates for Peripheral edema in SGLT2I (female) (T) vs SU (female) (C) new-user cohorts under four different propensity score (PS) adjustment / time-at-risk (TAR) designs across data sources. We report total patient exposure time in 1,000 patient-year (KYr) units, uncalibrated and calibrated hazard ratios (HRs) and their 95% confidence intervals (CIs). Greyed out entries did not pass study diagnostics and are included only for completeness.

| | Patients | | Exposure (KYr) | | Outcomes | | Uncalibrated | | Calibrated | |
|---|---|---|---|---|---|---|---|---|---|---|
| | T | C | T | C | T | C | HR (95% CI) | $p$ | HR (95% CI) | $p$ |
| **PS-matching / on-treatment TAR** | | | | | | | | | | |
| CCAE | 12,325 | 27,200 | 8.9 | 18.7 | 369 | 1,065 | 0.79 (0.66 - 0.93) | <0.01 | 0.75 (0.63 - 0.90) | <0.01 |
| DAG | 2,530 | 557 | 2.0 | 0.5 | 33 | 20 | 0.51 (0.24 - 1.10) | 0.08 | 0.56 (0.26 - 1.20) | 0.15 |
| ODOD | 8,641 | 29,325 | 5.3 | 22.7 | 310 | 2,178 | 0.77 (0.66 - 0.90) | <0.01 | 0.80 (0.68 - 0.93) | 0.01 |
| MDCR | 983 | 4,713 | 0.6 | 3.6 | 28 | 377 | 0.55 (0.32 - 0.90) | 0.02 | 0.57 (0.34 - 0.95) | 0.04 |
| MDCD | 1,517 | 7,521 | 0.8 | 4.3 | 69 | 454 | 0.75 (0.51 - 1.07) | 0.12 | 0.69 (0.47 - 0.99) | 0.05 |
| Open Claims | 192,709 | 626,018 | 144.3 | 569.4 | 3,477 | 24,047 | 0.71 (0.67 - 0.74) | <0.01 | 0.72 (0.60 - 0.86) | <0.01 |
| OEHR | 12,363 | 51,739 | 3.8 | 27.4 | 149 | 1,830 | 0.83 (0.67 - 1.03) | 0.10 | 0.87 (0.69 - 1.10) | 0.39 |
| SIDIAP | 2,386 | 847 | 3.4 | 1.5 | 47 | 35 | 0.54 (0.30 - 0.96) | 0.04 | 0.63 (0.35 - 1.13) | 0.15 |
| Meta-analysis | 230,924 | 747,363 | 167.0 | 647.5 | 4,449 | 29,986 | 0.73 (0.68 - 0.78) | <0.01 | 0.73 (0.62 - 0.86) | <0.01 |
| Meta-analysis (w/o USOC) | 38,215 | 121,345 | 22.7 | 78.2 | 972 | 5,939 | 0.77 (0.70 - 0.85) | <0.01 | 0.76 (0.69 - 0.84) | <0.01 |
| **PS-stratification / on-treatment TAR** | | | | | | | | | | |
| CCAE | 14,270 | 27,767 | 10.1 | 18.9 | 434 | 1,084 | 0.76 (0.67 - 0.86) | <0.01 | 0.74 (0.65 - 0.84) | <0.01 |
| DAG | 2,565 | 990 | 2.0 | 1.0 | 33 | 47 | 0.32 (0.17 - 0.60) | <0.01 | 0.21 (0.11 - 0.39) | <0.01 |
| ODOD | 9,264 | 29,843 | 5.5 | 23.2 | 325 | 2,258 | 0.73 (0.64 - 0.82) | <0.01 | 0.76 (0.67 - 0.87) | <0.01 |
| MDCR | 1,087 | 4,955 | 0.7 | 3.7 | 30 | 397 | 0.52 (0.34 - 0.76) | <0.01 | 0.51 (0.34 - 0.76) | <0.01 |
| MDCD | 1,536 | 7,651 | 0.8 | 4.4 | 69 | 460 | 0.89 (0.67 - 1.15) | 0.38 | 0.81 (0.62 - 1.07) | 0.16 |
| Open Claims | 201,879 | 633,848 | 149.5 | 576.2 | 3,614 | 24,744 | 0.70 (0.68 - 0.73) | <0.01 | 0.72 (0.61 - 0.84) | <0.01 |
| OEHR | 13,269 | 56,725 | 4.1 | 30.0 | 157 | 2,118 | 0.74 (0.62 - 0.88) | <0.01 | 0.77 (0.65 - 0.93) | 0.01 |
| SIDIAP | 2,460 | 1,790 | 3.4 | 3.7 | 48 | 84 | 0.64 (0.40 - 1.02) | 0.07 | 0.58 (0.37 - 0.94) | 0.03 |
| Meta-analysis | 238,682 | 748,183 | 169.2 | 648.3 | 4,530 | 30,204 | 0.71 (0.68 - 0.74) | <0.01 | 0.72 (0.62 - 0.83) | <0.01 |
| Meta-analysis (w/o USOC) | 36,803 | 114,335 | 19.7 | 72.1 | 916 | 5,460 | 0.74 (0.69 - 0.81) | <0.01 | 0.75 (0.69 - 0.82) | <0.01 |
| **PS-matching / on-treatment TAR without escalation** | | | | | | | | | | |
| CCAE | 12,325 | 27,200 | 7.7 | 16.2 | 313 | 924 | 0.80 (0.66 - 0.95) | 0.01 | 0.75 (0.62 - 0.90) | <0.01 |
| DAG | 2,530 | 557 | 1.8 | 0.5 | 31 | 17 | 0.47 (0.21 - 1.10) | 0.07 | 0.51 (0.22 - 1.17) | 0.13 |
| ODOD | 8,641 | 29,325 | 4.7 | 20.5 | 273 | 1,955 | 0.73 (0.61 - 0.86) | <0.01 | 0.74 (0.62 - 0.88) | <0.01 |
| MDCR | 983 | 4,713 | 0.6 | 3.3 | 26 | 353 | 0.55 (0.32 - 0.90) | 0.03 | 0.56 (0.33 - 0.95) | 0.04 |
| MDCD | 1,517 | 7,521 | 0.6 | 3.7 | 60 | 393 | 0.86 (0.58 - 1.25) | 0.45 | 0.82 (0.56 - 1.21) | 0.34 |
| Open Claims | 192,709 | 626,018 | 121.8 | 483.6 | 2,870 | 20,251 | 0.70 (0.66 - 0.74) | <0.01 | 0.71 (0.60 - 0.85) | <0.01 |
| OEHR | 12,363 | 51,739 | 3.3 | 23.7 | 125 | 1,501 | 0.87 (0.68 - 1.09) | 0.23 | 0.90 (0.70 - 1.14) | 0.43 |
| SIDIAP | 2,386 | 847 | 3.0 | 1.2 | 40 | 30 | 0.60 (0.30 - 1.16) | 0.13 | 0.77 (0.39 - 1.50) | 0.45 |
| Meta-analysis | 230,924 | 747,363 | 141.7 | 552.1 | 3,707 | 25,407 | 0.73 (0.68 - 0.79) | <0.01 | 0.73 (0.62 - 0.87) | <0.01 |
| Meta-analysis (w/o USOC) | 38,215 | 121,345 | 19.8 | 68.5 | 837 | 5,156 | 0.77 (0.70 - 0.85) | <0.01 | 0.75 (0.68 - 0.84) | <0.01 |
| **PS-stratification / on-treatment TAR without escalation** | | | | | | | | | | |
| CCAE | 14,270 | 27,767 | 8.7 | 16.4 | 372 | 940 | 0.77 (0.67 - 0.87) | <0.01 | 0.73 (0.64 - 0.84) | <0.01 |
| DAG | 2,565 | 990 | 1.8 | 0.9 | 31 | 44 | 0.40 (0.21 - 0.76) | <0.01 | 0.26 (0.14 - 0.49) | <0.01 |
| ODOD | 9,264 | 29,843 | 4.9 | 20.8 | 288 | 2,029 | 0.73 (0.64 - 0.83) | <0.01 | 0.76 (0.66 - 0.87) | <0.01 |
| MDCR | 1,087 | 4,955 | 0.6 | 3.5 | 28 | 373 | 0.53 (0.34 - 0.78) | <0.01 | 0.49 (0.32 - 0.74) | <0.01 |
| MDCD | 1,536 | 7,651 | 0.6 | 3.7 | 60 | 396 | 0.93 (0.69 - 1.23) | 0.60 | 0.83 (0.62 - 1.12) | 0.25 |
| Open Claims | 201,879 | 633,848 | 126.4 | 489.6 | 2,985 | 20,855 | 0.70 (0.67 - 0.73) | <0.01 | 0.71 (0.62 - 0.82) | <0.01 |
| OEHR | 13,269 | 56,725 | 3.6 | 25.9 | 131 | 1,731 | 0.76 (0.62 - 0.92) | <0.01 | 0.77 (0.63 - 0.94) | 0.03 |
| SIDIAP | 2,460 | 1,790 | 3.0 | 3.1 | 41 | 72 | 0.54 (0.33 - 0.90) | 0.02 | 0.55 (0.33 - 0.92) | 0.03 |
| Meta-analysis | 238,682 | 748,183 | 143.6 | 552.9 | 3,776 | 25,555 | 0.72 (0.68 - 0.75) | <0.01 | 0.72 (0.63 - 0.81) | <0.01 |
| Meta-analysis (w/o USOC) | 36,803 | 114,335 | 17.2 | 63.2 | 791 | 4,700 | 0.75 (0.69 - 0.82) | <0.01 | 0.74 (0.68 - 0.81) | <0.01 |

##### 1.3.24 Photosensitivity

**Supplementary Table 76:** Exposures, outcomes and risk estimates for Photosensitivity in SGLT2I (female) (T) vs SU (female) (C) new-user cohorts under four different propensity score (PS) adjustment / time-at-risk (TAR) designs across data sources. We report total patient exposure time in 1,000 patient-year (KYr) units, uncalibrated and calibrated hazard ratios (HRs) and their 95% confidence intervals (CIs). Greyed out entries did not pass study diagnostics and are included only for completeness.

| | Patients | | Exposure (KYr) | | Outcomes | | Uncalibrated | | Calibrated | |
|---|---|---|---|---|---|---|---|---|---|---|
| | T | C | T | C | T | C | HR (95% CI) | $p$ | HR (95% CI) | $p$ |
| **PS-matching / on-treatment TAR** | | | | | | | | | | |
| CCAE | 15,922 | 34,859 | 11.7 | 24.8 | 9 | 31 | 1.02 (0.37 - 2.67) | 0.97 | 0.97 (0.36 - 2.63) | 0.96 |
| DAG | 3,165 | 680 | 2.5 | 0.7 | <5 | 0 | | | | |
| ODOD | 11,746 | 40,624 | 7.3 | 33.9 | <5 | 31 | 0.48 (0.07 - 1.89) | 0.37 | 0.49 (0.10 - 2.53) | 0.40 |
| MDCR | 1,320 | 6,692 | 0.9 | 5.4 | <5 | 8 | 0.97 (0.04 - 9.26) | 0.98 | 1.00 (0.07 - 14.7) | 0.97 |
| MDCD | 2,373 | 11,605 | 1.3 | 7.3 | <5 | 8 | 1.09 (0.05 - 8.01) | 0.95 | 1.00 (0.08 - 12.2) | 0.98 |
| Open Claims | 259,076 | 825,095 | 194.3 | 780.5 | 65 | 362 | 0.84 (0.58 - 1.19) | 0.34 | 0.86 (0.57 - 1.28) | 0.45 |
| OEHR | 15,371 | 63,759 | 4.8 | 36.5 | <5 | 52 | 0.48 (0.11 - 1.48) | 0.27 | 0.50 (0.13 - 1.84) | 0.31 |
| SIDIAP | 2,837 | 962 | 4.0 | 1.7 | 0 | <5 | | | | |
| Meta-analysis | 302,115 | 964,337 | 218.1 | 875.7 | <84 | 476 | 0.81 (0.59 - 1.12) | 0.20 | 0.81 (0.57 - 1.15) | 0.24 |
| Meta-analysis (w/o USOC) | 43,039 | 139,242 | 23.8 | 95.3 | <19 | 114 | 0.71 (0.35 - 1.44) | 0.34 | 0.69 (0.34 - 1.41) | 0.31 |
| **PS-stratification / on-treatment TAR** | | | | | | | | | | |
| CCAE | 18,681 | 35,476 | 13.5 | 25.1 | 11 | 31 | 0.83 (0.37 - 1.75) | 0.63 | 0.81 (0.37 - 1.76) | 0.59 |
| DAG | 3,216 | 1,225 | 2.5 | 1.2 | <5 | <5 | 0.39 (0.01 - 7.40) | 0.56 | 0.25 (0.01 - 6.19) | 0.40 |
| ODOD | 12,737 | 41,516 | 7.8 | 34.8 | <5 | 33 | 0.44 (0.10 - 1.42) | 0.23 | 0.46 (0.12 - 1.77) | 0.26 |
| MDCR | 1,473 | 7,176 | 0.9 | 5.8 | <5 | 8 | 0.67 (0.03 - 5.36) | 0.76 | 0.66 (0.05 - 8.70) | 0.75 |
| MDCD | 2,416 | 11,868 | 1.3 | 7.5 | <5 | 8 | 0.56 (0.03 - 3.46) | 0.63 | 0.51 (0.05 - 5.63) | 0.58 |
| Open Claims | 273,499 | 835,013 | 202.5 | 789.7 | 72 | 375 | 0.97 (0.74 - 1.28) | 0.85 | 1.00 (0.73 - 1.37) | 0.95 |
| OEHR | 16,535 | 69,939 | 5.2 | 40.2 | <5 | 61 | 0.61 (0.14 - 1.79) | 0.44 | 0.64 (0.18 - 2.26) | 0.49 |
| SIDIAP | 2,938 | 2,004 | 4.0 | 4.3 | 0 | <5 | | | | |
| Meta-analysis | 321,452 | 981,944 | 229.0 | 889.8 | <93 | 500 | 0.92 (0.71 - 1.18) | 0.49 | 0.92 (0.69 - 1.23) | 0.58 |
| Meta-analysis (w/o USOC) | 47,953 | 146,931 | 26.5 | 100.1 | <21 | 125 | 0.68 (0.38 - 1.24) | 0.21 | 0.69 (0.38 - 1.25) | 0.23 |
| **PS-matching / on-treatment TAR without escalation** | | | | | | | | | | |
| CCAE | 15,922 | 34,859 | 10.1 | 21.4 | 8 | 28 | 1.34 (0.47 - 3.69) | 0.58 | 1.27 (0.45 - 3.57) | 0.65 |
| DAG | 3,165 | 680 | 2.3 | 0.6 | <5 | 0 | | | | |
| ODOD | 11,746 | 40,624 | 6.5 | 30.3 | <5 | 31 | 0.44 (0.07 - 1.78) | 0.33 | 0.45 (0.09 - 2.32) | 0.34 |
| MDCR | 1,320 | 6,692 | 0.8 | 4.9 | <5 | 6 | 0.97 (0.04 - 9.18) | 0.98 | 0.98 (0.07 - 14.3) | 0.97 |
| MDCD | 2,373 | 11,605 | 1.1 | 6.0 | <5 | 6 | 1.09 (0.05 - 8.01) | 0.95 | 1.04 (0.08 - 12.7) | 0.97 |
| Open Claims | 259,076 | 825,095 | 163.0 | 657.2 | 53 | 312 | 0.89 (0.60 - 1.30) | 0.55 | 0.91 (0.60 - 1.38) | 0.66 |
| OEHR | 15,371 | 63,759 | 4.2 | 30.9 | <5 | 45 | 0.49 (0.11 - 1.52) | 0.28 | 0.50 (0.14 - 1.87) | 0.31 |
| SIDIAP | 2,837 | 962 | 3.5 | 1.4 | 0 | <5 | | | | |
| Meta-analysis | 302,115 | 964,337 | 183.8 | 739.7 | <71 | 416 | 0.87 (0.61 - 1.22) | 0.41 | 0.87 (0.60 - 1.26) | 0.45 |
| Meta-analysis (w/o USOC) | 43,039 | 139,242 | 20.8 | 82.6 | <18 | 104 | 0.79 (0.38 - 1.64) | 0.53 | 0.77 (0.37 - 1.61) | 0.49 |
| **PS-stratification / on-treatment TAR without escalation** | | | | | | | | | | |
| CCAE | 18,681 | 35,476 | 11.7 | 21.6 | 10 | 28 | 0.85 (0.36 - 1.87) | 0.70 | 0.81 (0.36 - 1.85) | 0.62 |
| DAG | 3,216 | 1,225 | 2.3 | 1.1 | <5 | <5 | 0.98 (0.02 - 54.9) | 0.99 | 0.63 (0.01 - 35.3) | 0.82 |
| ODOD | 12,737 | 41,516 | 6.9 | 31.1 | <5 | 32 | 0.46 (0.10 - 1.48) | 0.25 | 0.47 (0.12 - 1.81) | 0.27 |
| MDCR | 1,473 | 7,176 | 0.8 | 5.3 | <5 | 6 | 0.73 (0.03 - 6.17) | 0.81 | 0.68 (0.05 - 9.26) | 0.77 |
| MDCD | 2,416 | 11,868 | 1.1 | 6.2 | <5 | 6 | 0.78 (0.04 - 5.17) | 0.84 | 0.70 (0.06 - 8.12) | 0.78 |
| Open Claims | 273,499 | 835,013 | 170.2 | 665.2 | 59 | 324 | 0.96 (0.70 - 1.29) | 0.80 | 0.98 (0.70 - 1.37) | 0.90 |
| OEHR | 16,535 | 69,939 | 4.5 | 34.0 | <5 | 51 | 0.67 (0.16 - 2.00) | 0.54 | 0.68 (0.19 - 2.44) | 0.56 |
| SIDIAP | 2,938 | 2,004 | 3.5 | 3.6 | 0 | <5 | | | | |
| Meta-analysis | 321,452 | 981,944 | 193.3 | 751.9 | <79 | 435 | 0.91 (0.69 - 1.19) | 0.47 | 0.91 (0.67 - 1.22) | 0.51 |
| Meta-analysis (w/o USOC) | 47,953 | 146,931 | 23.1 | 86.7 | <20 | 111 | 0.71 (0.38 - 1.31) | 0.27 | 0.70 (0.38 - 1.29) | 0.26 |

##### 1.3.25 Venous thromboembolism

**Supplementary Table 77:** Exposures, outcomes and risk estimates for Venous thromboembolism in SGLT2I (female) (T) vs SU (female) (C) new-user cohorts under four different propensity score (PS) adjustment / time-at-risk (TAR) designs across data sources. We report total patient exposure time in 1,000 patient-year (KYr) units, uncalibrated and calibrated hazard ratios (HRs) and their 95% confidence intervals (CIs). Greyed out entries did not pass study diagnostics and are included only for completeness.

| | Patients | | Exposure (KYr) | | Outcomes | | Uncalibrated | | Calibrated | |
|---|---|---|---|---|---|---|---|---|---|---|
| | T | C | T | C | T | C | HR (95% CI) | $p$ | HR (95% CI) | $p$ |
| **PS-matching / on-treatment TAR** | | | | | | | | | | |
| CCAE | 15,702 | 34,115 | 11.6 | 24.3 | 50 | 153 | 0.83 (0.53 - 1.27) | 0.40 | 0.79 (0.51 - 1.23) | 0.30 |
| DAG | 2,914 | 625 | 2.3 | 0.6 | 15 | 7 | 0.63 (0.17 - 2.62) | 0.51 | 0.70 (0.18 - 2.71) | 0.59 |
| ODOD | 11,476 | 39,372 | 7.1 | 32.8 | 58 | 373 | 1.18 (0.82 - 1.68) | 0.37 | 1.22 (0.85 - 1.75) | 0.30 |
| MDCR | 1,277 | 6,479 | 0.9 | 5.2 | 5 | 81 | 0.72 (0.23 - 1.84) | 0.53 | 0.74 (0.26 - 2.10) | 0.58 |
| MDCD | 2,277 | 11,099 | 1.2 | 6.9 | 10 | 76 | 2.09 (0.89 - 4.58) | 0.08 | 1.91 (0.84 - 4.35) | 0.12 |
| Open Claims | 253,292 | 804,409 | 190.1 | 758.7 | 696 | 4,734 | 0.81 (0.73 - 0.90) | <0.01 | 0.83 (0.68 - 1.02) | 0.08 |
| OEHR | 15,125 | 62,308 | 4.7 | 35.3 | 38 | 328 | 1.44 (0.93 - 2.18) | 0.10 | 1.50 (0.97 - 2.32) | 0.11 |
| SIDIAP | 2,721 | 929 | 3.8 | 1.6 | 14 | 5 | 0.50 (0.11 - 2.30) | 0.37 | 0.59 (0.13 - 2.64) | 0.50 |
| Meta-analysis | 299,149 | 957,782 | 215.6 | 863.2 | 857 | 5,745 | 1.04 (0.80 - 1.37) | 0.76 | 1.04 (0.77 - 1.42) | 0.79 |
| Meta-analysis (w/o USOC) | 45,857 | 153,373 | 25.5 | 104.5 | 161 | 1,011 | 1.16 (0.87 - 1.55) | 0.30 | 1.14 (0.86 - 1.53) | 0.38 |
| **PS-stratification / on-treatment TAR** | | | | | | | | | | |
| CCAE | 18,464 | 34,895 | 13.4 | 24.6 | 59 | 155 | 0.80 (0.57 - 1.10) | 0.18 | 0.78 (0.56 - 1.08) | 0.15 |
| DAG | 2,961 | 1,106 | 2.3 | 1.1 | 15 | 13 | 0.43 (0.16 - 1.23) | 0.11 | 0.28 (0.10 - 0.78) | 0.02 |
| ODOD | 12,418 | 40,278 | 7.6 | 33.7 | 60 | 393 | 0.94 (0.69 - 1.27) | 0.71 | 0.99 (0.73 - 1.34) | 0.92 |
| MDCR | 1,426 | 6,917 | 0.9 | 5.6 | 5 | 87 | 0.47 (0.15 - 1.15) | 0.14 | 0.46 (0.17 - 1.25) | 0.13 |
| MDCD | 2,324 | 11,379 | 1.2 | 7.1 | 10 | 84 | 0.88 (0.41 - 1.72) | 0.72 | 0.81 (0.39 - 1.66) | 0.56 |
| Open Claims | 267,177 | 813,929 | 198.1 | 767.6 | 730 | 4,915 | 0.77 (0.70 - 0.83) | <0.01 | 0.78 (0.66 - 0.94) | <0.01 |
| OEHR | 16,265 | 68,607 | 5.1 | 39.0 | 38 | 385 | 1.47 (1.01 - 2.09) | 0.04 | 1.53 (1.06 - 2.21) | 0.03 |
| SIDIAP | 2,816 | 1,941 | 3.8 | 4.2 | 15 | 15 | 0.77 (0.30 - 1.95) | 0.58 | 0.70 (0.27 - 1.79) | 0.46 |
| Meta-analysis | 314,324 | 957,709 | 224.1 | 864.9 | 887 | 5,848 | 0.93 (0.71 - 1.23) | 0.62 | 0.94 (0.69 - 1.28) | 0.69 |
| Meta-analysis (w/o USOC) | 47,147 | 143,780 | 26.0 | 97.3 | 157 | 933 | 1.03 (0.73 - 1.44) | 0.89 | 1.04 (0.74 - 1.45) | 0.84 |
| **PS-matching / on-treatment TAR without escalation** | | | | | | | | | | |
| CCAE | 15,702 | 34,115 | 9.9 | 20.9 | 41 | 137 | 0.73 (0.45 - 1.17) | 0.21 | 0.69 (0.43 - 1.12) | 0.14 |
| DAG | 2,914 | 625 | 2.1 | 0.6 | 13 | 7 | 0.43 (0.10 - 1.94) | 0.26 | 0.46 (0.11 - 2.01) | 0.31 |
| ODOD | 11,476 | 39,372 | 6.3 | 29.3 | 51 | 336 | 1.09 (0.74 - 1.59) | 0.66 | 1.10 (0.75 - 1.62) | 0.63 |
| MDCR | 1,277 | 6,479 | 0.8 | 4.7 | <5 | 75 | 0.61 (0.17 - 1.68) | 0.40 | 0.62 (0.20 - 1.94) | 0.42 |
| MDCD | 2,277 | 11,099 | 1.0 | 5.7 | 7 | 57 | 2.23 (0.80 - 5.56) | 0.11 | 2.13 (0.80 - 5.62) | 0.13 |
| Open Claims | 253,292 | 804,409 | 159.7 | 639.2 | 569 | 4,046 | 0.76 (0.67 - 0.85) | <0.01 | 0.77 (0.64 - 0.94) | 0.01 |
| OEHR | 15,125 | 62,308 | 4.1 | 30.0 | 32 | 255 | 1.46 (0.89 - 2.31) | 0.12 | 1.50 (0.93 - 2.42) | 0.11 |
| SIDIAP | 2,721 | 929 | 3.3 | 1.3 | 13 | <5 | 0.40 (0.08 - 2.34) | 0.29 | 0.51 (0.09 - 2.81) | 0.44 |
| Meta-analysis | 299,149 | 957,782 | 181.8 | 729.9 | <705 | 4,906 | 0.98 (0.72 - 1.32) | 0.88 | 0.98 (0.70 - 1.37) | 0.90 |
| Meta-analysis (w/o USOC) | 45,857 | 153,373 | 22.1 | 90.7 | <136 | 860 | 1.09 (0.77 - 1.55) | 0.62 | 1.07 (0.75 - 1.52) | 0.72 |
| **PS-stratification / on-treatment TAR without escalation** | | | | | | | | | | |
| CCAE | 18,464 | 34,895 | 11.5 | 21.2 | 49 | 139 | 0.76 (0.53 - 1.07) | 0.13 | 0.72 (0.50 - 1.03) | 0.08 |
| DAG | 2,961 | 1,106 | 2.1 | 1.0 | 13 | 13 | 0.32 (0.11 - 0.97) | 0.04 | 0.20 (0.07 - 0.61) | <0.01 |
| ODOD | 12,418 | 40,278 | 6.7 | 30.1 | 53 | 355 | 0.91 (0.65 - 1.24) | 0.56 | 0.94 (0.68 - 1.30) | 0.71 |
| MDCR | 1,426 | 6,917 | 0.8 | 5.1 | <5 | 80 | 0.38 (0.11 - 1.02) | 0.09 | 0.36 (0.12 - 1.09) | 0.07 |
| MDCD | 2,324 | 11,379 | 1.0 | 5.9 | 7 | 64 | 0.78 (0.31 - 1.72) | 0.58 | 0.71 (0.30 - 1.67) | 0.43 |
| Open Claims | 267,177 | 813,929 | 166.7 | 646.9 | 597 | 4,205 | 0.74 (0.67 - 0.81) | <0.01 | 0.75 (0.64 - 0.89) | <0.01 |
| OEHR | 16,265 | 68,607 | 4.4 | 33.1 | 32 | 296 | 1.55 (1.02 - 2.28) | 0.03 | 1.56 (1.04 - 2.34) | 0.04 |
| SIDIAP | 2,816 | 1,941 | 3.4 | 3.4 | 13 | 12 | 0.81 (0.29 - 2.19) | 0.68 | 0.82 (0.30 - 2.24) | 0.71 |
| Meta-analysis | 314,324 | 957,709 | 189.3 | 731.4 | 731 | 4,995 | 0.91 (0.67 - 1.25) | 0.58 | 0.92 (0.65 - 1.28) | 0.60 |
| Meta-analysis (w/o USOC) | 47,147 | 143,780 | 22.7 | 84.4 | 134 | 790 | 1.01 (0.68 - 1.50) | 0.96 | 1.00 (0.67 - 1.49) | 0.96 |

##### 1.3.26 Vomiting

**Supplementary Table 78:** Exposures, outcomes and risk estimates for Vomiting in SGLT2I (female) (T) vs SU (female) (C) new-user cohorts under four different propensity score (PS) adjustment / time-at-risk (TAR) designs across data sources. We report total patient exposure time in 1,000 patient-year (KYr) units, uncalibrated and calibrated hazard ratios (HRs) and their 95% confidence intervals (CIs). Greyed out entries did not pass study diagnostics and are included only for completeness.

| | Patients | | Exposure (KYr) | | Outcomes | | Uncalibrated | | Calibrated | |
|---|---|---|---|---|---|---|---|---|---|---|
| | T | C | T | C | T | C | HR (95% CI) | $p$ | HR (95% CI) | $p$ |
| **PS-matching / on-treatment TAR** | | | | | | | | | | |
| CCAE | 14,041 | 30,666 | 10.3 | 21.7 | 300 | 694 | 0.91 (0.76 - 1.10) | 0.34 | 0.87 (0.72 - 1.06) | 0.19 |
| DAG | 2,813 | 604 | 2.2 | 0.6 | 27 | 6 | 3.42 (0.90 - 22.5) | 0.13 | 3.77 (0.75 - 18.9) | 0.11 |
| ODOD | 10,217 | 34,872 | 6.3 | 28.8 | 197 | 1,190 | 0.83 (0.67 - 1.01) | 0.06 | 0.85 (0.70 - 1.04) | 0.15 |
| MDCR | 1,211 | 5,956 | 0.8 | 4.7 | 20 | 179 | 0.72 (0.37 - 1.31) | 0.31 | 0.75 (0.40 - 1.41) | 0.38 |
| MDCD | 1,706 | 8,339 | 0.9 | 5.1 | 70 | 352 | 1.14 (0.81 - 1.59) | 0.44 | 1.05 (0.75 - 1.47) | 0.78 |
| Open Claims | 223,660 | 716,261 | 168.9 | 678.9 | 2,751 | 13,887 | 0.95 (0.89 - 1.00) | 0.05 | 0.97 (0.81 - 1.16) | 0.71 |
| OEHR | 13,955 | 57,490 | 4.3 | 32.0 | 129 | 1,172 | 1.07 (0.84 - 1.35) | 0.56 | 1.12 (0.87 - 1.44) | 0.53 |
| SIDIAP | 2,657 | 896 | 3.7 | 1.6 | 25 | 13 | 0.33 (0.11 - 0.99) | 0.06 | 0.39 (0.13 - 1.20) | 0.11 |
| Meta-analysis | 267,447 | 854,480 | 195.2 | 772.9 | 3,492 | 17,487 | 0.94 (0.90 - 0.99) | 0.02 | 0.94 (0.80 - 1.10) | 0.43 |
| Meta-analysis (w/o USOC) | 43,787 | 138,219 | 26.3 | 94.0 | 741 | 3,600 | 0.92 (0.79 - 1.08) | 0.32 | 0.91 (0.78 - 1.06) | 0.26 |
| **PS-stratification / on-treatment TAR** | | | | | | | | | | |
| CCAE | 16,475 | 31,156 | 11.9 | 21.9 | 353 | 699 | 0.98 (0.85 - 1.13) | 0.80 | 0.96 (0.83 - 1.10) | 0.58 |
| DAG | 2,860 | 1,091 | 2.2 | 1.1 | 27 | 15 | 1.11 (0.48 - 2.67) | 0.80 | 0.72 (0.31 - 1.70) | 0.48 |
| ODOD | 11,062 | 35,657 | 6.7 | 29.5 | 210 | 1,236 | 0.81 (0.69 - 0.95) | <0.01 | 0.85 (0.72 - 1.00) | 0.06 |
| MDCR | 1,346 | 6,340 | 0.8 | 5.0 | 21 | 196 | 0.70 (0.41 - 1.12) | 0.16 | 0.69 (0.42 - 1.13) | 0.16 |
| MDCD | 1,727 | 8,546 | 0.9 | 5.3 | 71 | 361 | 1.16 (0.88 - 1.51) | 0.29 | 1.07 (0.81 - 1.40) | 0.66 |
| Open Claims | 235,920 | 725,274 | 176.1 | 687.4 | 2,889 | 14,218 | 0.89 (0.85 - 0.93) | <0.01 | 0.92 (0.78 - 1.08) | 0.29 |
| OEHR | 15,028 | 63,337 | 4.6 | 35.4 | 137 | 1,315 | 0.96 (0.79 - 1.16) | 0.68 | 1.00 (0.82 - 1.22) | 0.85 |
| SIDIAP | 2,753 | 1,889 | 3.7 | 4.0 | 26 | 41 | 0.67 (0.35 - 1.25) | 0.22 | 0.61 (0.32 - 1.15) | 0.14 |
| Meta-analysis | 278,485 | 855,424 | 199.3 | 774.2 | 3,589 | 17,468 | 0.90 (0.86 - 0.93) | <0.01 | 0.90 (0.78 - 1.05) | 0.17 |
| Meta-analysis (w/o USOC) | 42,565 | 130,150 | 23.2 | 86.8 | 700 | 3,250 | 0.91 (0.81 - 1.04) | 0.16 | 0.92 (0.81 - 1.05) | 0.27 |
| **PS-matching / on-treatment TAR without escalation** | | | | | | | | | | |
| CCAE | 14,041 | 30,666 | 8.9 | 18.8 | 261 | 586 | 0.93 (0.76 - 1.15) | 0.52 | 0.88 (0.72 - 1.08) | 0.26 |
| DAG | 2,813 | 604 | 2.0 | 0.5 | 23 | 6 | 3.34 (0.87 - 22.0) | 0.14 | 3.62 (0.72 - 18.2) | 0.13 |
| ODOD | 10,217 | 34,872 | 5.6 | 25.8 | 168 | 1,057 | 0.81 (0.65 - 1.00) | 0.06 | 0.82 (0.66 - 1.02) | 0.09 |
| MDCR | 1,211 | 5,956 | 0.7 | 4.3 | 19 | 156 | 0.81 (0.41 - 1.49) | 0.52 | 0.83 (0.43 - 1.57) | 0.57 |
| MDCD | 1,706 | 8,339 | 0.8 | 4.3 | 56 | 298 | 1.13 (0.77 - 1.63) | 0.53 | 1.08 (0.74 - 1.57) | 0.71 |
| Open Claims | 223,660 | 716,261 | 142.6 | 575.1 | 2,209 | 11,482 | 0.94 (0.88 - 1.00) | 0.04 | 0.96 (0.81 - 1.14) | 0.66 |
| OEHR | 13,955 | 57,490 | 3.8 | 27.5 | 106 | 931 | 1.09 (0.83 - 1.41) | 0.54 | 1.12 (0.86 - 1.47) | 0.46 |
| SIDIAP | 2,657 | 896 | 3.2 | 1.3 | 20 | 10 | 0.32 (0.09 - 1.10) | 0.08 | 0.41 (0.12 - 1.45) | 0.17 |
| Meta-analysis | 267,447 | 854,480 | 165.6 | 657.0 | 2,839 | 14,520 | 0.94 (0.89 - 0.99) | 0.02 | 0.94 (0.81 - 1.09) | 0.41 |
| Meta-analysis (w/o USOC) | 43,787 | 138,219 | 23.0 | 82.0 | 630 | 3,038 | 0.93 (0.80 - 1.08) | 0.35 | 0.91 (0.78 - 1.07) | 0.27 |
| **PS-stratification / on-treatment TAR without escalation** | | | | | | | | | | |
| CCAE | 16,475 | 31,156 | 10.3 | 19.0 | 301 | 591 | 1.02 (0.87 - 1.19) | 0.80 | 0.97 (0.83 - 1.14) | 0.76 |
| DAG | 2,860 | 1,091 | 2.0 | 1.0 | 23 | 14 | 1.18 (0.51 - 2.88) | 0.70 | 0.76 (0.32 - 1.82) | 0.56 |
| ODOD | 11,062 | 35,657 | 6.0 | 26.4 | 179 | 1,101 | 0.78 (0.66 - 0.93) | <0.01 | 0.81 (0.68 - 0.97) | 0.02 |
| MDCR | 1,346 | 6,340 | 0.7 | 4.6 | 20 | 170 | 0.79 (0.46 - 1.29) | 0.37 | 0.73 (0.44 - 1.23) | 0.26 |
| MDCD | 1,727 | 8,546 | 0.8 | 4.4 | 56 | 306 | 1.06 (0.77 - 1.43) | 0.70 | 0.96 (0.70 - 1.30) | 0.78 |
| Open Claims | 235,920 | 725,274 | 148.9 | 582.5 | 2,319 | 11,766 | 0.88 (0.84 - 0.92) | <0.01 | 0.90 (0.78 - 1.03) | 0.14 |
| OEHR | 15,028 | 63,337 | 4.1 | 30.4 | 112 | 1,048 | 0.95 (0.77 - 1.17) | 0.66 | 0.96 (0.77 - 1.20) | 0.76 |
| SIDIAP | 2,753 | 1,889 | 3.3 | 3.3 | 20 | 33 | 0.58 (0.27 - 1.20) | 0.16 | 0.59 (0.28 - 1.25) | 0.18 |
| Meta-analysis | 278,485 | 855,424 | 169.2 | 658.3 | 2,911 | 14,506 | 0.90 (0.82 - 0.99) | 0.03 | 0.90 (0.77 - 1.04) | 0.16 |
| Meta-analysis (w/o USOC) | 42,565 | 130,150 | 20.3 | 75.8 | 592 | 2,740 | 0.91 (0.77 - 1.08) | 0.29 | 0.90 (0.76 - 1.07) | 0.26 |

#### 1.4 GLP1RA (female) vs DPP4I (female)

##### 1.4.1 3-point MACE

**Supplementary Table 79:** Exposures, outcomes and risk estimates for 3-point MACE in GLP1RA (female) (T) vs DPP4I (female) (C) new-user cohorts under four different propensity score (PS) adjustment / time-at-risk (TAR) designs across data sources. We report total patient exposure time in 1,000 patient-year (KYr) units, uncalibrated and calibrated hazard ratios (HRs) and their 95% confidence intervals (CIs). Greyed out entries did not pass study diagnostics and are included only for completeness.

| | Patients | | Exposure (KYr) | | Outcomes | | Uncalibrated | | Calibrated | |
|---|---|---|---|---|---|---|---|---|---|---|
| | T | C | T | C | T | C | HR (95% CI) | $p$ | HR (95% CI) | $p$ |
| **PS-matching / on-treatment TAR** | | | | | | | | | | |
| CCAE | 14,149 | 44,506 | 8.9 | 35.1 | 45 | 203 | 1.14 (0.71 - 1.78) | 0.57 | 1.24 (0.78 - 1.96) | 0.39 |
| ODOD | 7,703 | 27,560 | 4.5 | 21.7 | 52 | 366 | 1.16 (0.79 - 1.67) | 0.45 | 1.13 (0.77 - 1.64) | 0.55 |
| MDCR | 941 | 7,585 | 0.6 | 7.2 | 6 | 157 | 0.59 (0.17 - 1.54) | 0.35 | 0.49 (0.16 - 1.46) | 0.20 |
| MDCD | 1,771 | 7,241 | 1.0 | 5.1 | 9 | 101 | 0.41 (0.14 - 1.00) | 0.08 | 0.42 (0.16 - 1.13) | 0.09 |
| Open Claims | 139,429 | 427,400 | 101.8 | 373.9 | 532 | 3,909 | 0.80 (0.70 - 0.90) | <0.01 | 0.80 (0.68 - 0.95) | 0.02 |
| OEHR | 10,417 | 38,070 | 3.2 | 19.7 | 22 | 155 | 1.20 (0.67 - 2.07) | 0.54 | 1.15 (0.65 - 2.03) | 0.63 |
| Meta-analysis | 173,469 | 544,777 | 119.3 | 455.6 | 660 | 4,734 | 0.94 (0.74 - 1.19) | 0.60 | 0.94 (0.74 - 1.21) | 0.66 |
| Meta-analysis (w/o USOC) | 34,040 | 117,377 | 17.6 | 81.6 | 128 | 825 | 1.06 (0.79 - 1.43) | 0.69 | 1.08 (0.80 - 1.46) | 0.62 |
| **PS-stratification / on-treatment TAR** | | | | | | | | | | |
| CCAE | 19,084 | 44,510 | 11.3 | 35.1 | 54 | 203 | 1.06 (0.75 - 1.48) | 0.72 | 1.11 (0.79 - 1.56) | 0.56 |
| ODOD | 11,356 | 28,169 | 6.0 | 22.3 | 66 | 400 | 0.89 (0.65 - 1.20) | 0.46 | 0.89 (0.65 - 1.21) | 0.47 |
| MDCR | 1,075 | 8,124 | 0.7 | 7.6 | 6 | 182 | 0.43 (0.16 - 0.94) | 0.06 | 0.35 (0.15 - 0.86) | 0.02 |
| MDCD | 1,943 | 7,282 | 1.0 | 5.2 | 9 | 102 | 0.51 (0.23 - 1.00) | 0.07 | 0.52 (0.25 - 1.08) | 0.08 |
| Open Claims | 223,507 | 428,126 | 139.3 | 374.7 | 676 | 3,943 | 0.78 (0.71 - 0.86) | <0.01 | 0.81 (0.69 - 0.97) | 0.03 |
| OEHR | 13,452 | 38,260 | 4.0 | 19.8 | 25 | 161 | 1.14 (0.69 - 1.80) | 0.60 | 1.07 (0.66 - 1.72) | 0.80 |
| Meta-analysis | 269,342 | 546,347 | 161.6 | 457.0 | 830 | 4,809 | 0.86 (0.73 - 1.01) | 0.07 | 0.88 (0.72 - 1.08) | 0.22 |
| Meta-analysis (w/o USOC) | 45,835 | 118,221 | 22.3 | 82.4 | 154 | 866 | 0.94 (0.74 - 1.19) | 0.61 | 0.96 (0.75 - 1.22) | 0.73 |
| **PS-matching / on-treatment TAR without escalation** | | | | | | | | | | |
| CCAE | 14,149 | 44,506 | 8.0 | 28.9 | 36 | 158 | 1.18 (0.71 - 1.92) | 0.52 | 1.26 (0.76 - 2.08) | 0.38 |
| ODOD | 7,703 | 27,560 | 4.0 | 18.2 | 45 | 322 | 1.00 (0.66 - 1.49) | 1.00 | 0.96 (0.64 - 1.44) | 0.85 |
| MDCR | 941 | 7,585 | 0.6 | 6.2 | 6 | 124 | 0.71 (0.21 - 1.88) | 0.55 | 0.57 (0.19 - 1.73) | 0.33 |
| MDCD | 1,771 | 7,241 | 0.8 | 4.0 | 8 | 77 | 0.43 (0.12 - 1.14) | 0.14 | 0.45 (0.15 - 1.37) | 0.16 |
| Open Claims | 139,429 | 427,400 | 90.2 | 297.0 | 455 | 3,167 | 0.74 (0.65 - 0.86) | <0.01 | 0.75 (0.65 - 0.87) | <0.01 |
| OEHR | 10,417 | 38,070 | 2.8 | 15.8 | 17 | 99 | 1.16 (0.58 - 2.19) | 0.67 | 1.14 (0.58 - 2.22) | 0.71 |
| Meta-analysis | 173,469 | 544,777 | 105.8 | 364.0 | 561 | 3,823 | 0.88 (0.69 - 1.12) | 0.30 | 0.89 (0.69 - 1.15) | 0.37 |
| Meta-analysis (w/o USOC) | 34,040 | 117,377 | 15.6 | 67.0 | 106 | 656 | 1.02 (0.78 - 1.35) | 0.87 | 1.05 (0.79 - 1.38) | 0.75 |
| **PS-stratification / on-treatment TAR without escalation** | | | | | | | | | | |
| CCAE | 19,084 | 44,510 | 10.2 | 28.9 | 43 | 158 | 1.06 (0.71 - 1.54) | 0.76 | 1.10 (0.75 - 1.62) | 0.62 |
| ODOD | 11,356 | 28,169 | 5.5 | 18.7 | 59 | 353 | 0.83 (0.60 - 1.15) | 0.28 | 0.83 (0.60 - 1.16) | 0.29 |
| MDCR | 1,075 | 8,124 | 0.6 | 6.6 | 6 | 147 | 0.57 (0.21 - 1.28) | 0.22 | 0.46 (0.19 - 1.14) | 0.10 |
| MDCD | 1,943 | 7,282 | 0.9 | 4.0 | 8 | 78 | 0.55 (0.23 - 1.14) | 0.14 | 0.57 (0.26 - 1.26) | 0.17 |
| Open Claims | 223,507 | 428,126 | 125.7 | 297.6 | 596 | 3,196 | 0.78 (0.70 - 0.86) | <0.01 | 0.80 (0.67 - 0.96) | 0.02 |
| OEHR | 13,452 | 38,260 | 3.4 | 15.9 | 20 | 104 | 1.22 (0.69 - 2.08) | 0.47 | 1.15 (0.66 - 2.00) | 0.65 |
| Meta-analysis | 269,342 | 546,347 | 145.7 | 365.2 | 726 | 3,889 | 0.84 (0.72 - 0.98) | 0.03 | 0.86 (0.72 - 1.02) | 0.08 |
| Meta-analysis (w/o USOC) | 45,835 | 118,221 | 20.0 | 67.6 | 130 | 693 | 0.93 (0.73 - 1.19) | 0.55 | 0.95 (0.74 - 1.21) | 0.68 |

##### 1.4.2 4-point MACE

**Supplementary Table 80:** Exposures, outcomes and risk estimates for 4-point MACE in GLP1RA (female) (T) vs DPP4I (female) (C) new-user cohorts under four different propensity score (PS) adjustment / time-at-risk (TAR) designs across data sources. We report total patient exposure time in 1,000 patient-year (KYr) units, uncalibrated and calibrated hazard ratios (HRs) and their 95% confidence intervals (CIs). Greyed out entries did not pass study diagnostics and are included only for completeness.

| | Patients | | Exposure (KYr) | | Outcomes | | Uncalibrated | | Calibrated | |
|---|---|---|---|---|---|---|---|---|---|---|
| | T | C | T | C | T | C | HR (95% CI) | $p$ | HR (95% CI) | $p$ |
| **PS-matching / on-treatment TAR** | | | | | | | | | | |
| CCAE | 13,948 | 43,925 | 8.8 | 34.7 | 55 | 329 | 0.74 (0.50 - 1.07) | 0.12 | 0.80 (0.55 - 1.18) | 0.29 |
| ODOD | 7,470 | 26,516 | 4.3 | 20.7 | 92 | 635 | 0.81 (0.59 - 1.09) | 0.18 | 0.79 (0.58 - 1.07) | 0.15 |
| MDCR | 900 | 7,079 | 0.6 | 6.7 | 11 | 248 | 0.42 (0.14 - 0.96) | 0.07 | 0.34 (0.13 - 0.89) | 0.03 |
| MDCD | 1,655 | 6,752 | 0.9 | 4.7 | 17 | 180 | 0.40 (0.19 - 0.77) | 0.01 | 0.41 (0.21 - 0.83) | 0.01 |
| Open Claims | 135,919 | 413,784 | 99.2 | 360.7 | 820 | 6,457 | 0.74 (0.67 - 0.82) | <0.01 | 0.75 (0.65 - 0.87) | <0.01 |
| OEHR | 10,256 | 37,505 | 3.2 | 19.2 | 37 | 316 | 1.02 (0.66 - 1.56) | 0.91 | 0.99 (0.64 - 1.52) | 0.88 |
| Meta-analysis | 169,248 | 528,482 | 116.4 | 440.1 | 1,021 | 7,917 | 0.75 (0.69 - 0.82) | <0.01 | 0.76 (0.68 - 0.85) | <0.01 |
| Meta-analysis (w/o USOC) | 33,329 | 114,698 | 17.2 | 79.4 | 201 | 1,460 | 0.77 (0.58 - 1.01) | 0.06 | 0.78 (0.59 - 1.03) | 0.08 |
| **PS-stratification / on-treatment TAR** | | | | | | | | | | |
| CCAE | 18,823 | 43,928 | 11.1 | 34.7 | 65 | 329 | 0.71 (0.52 - 0.94) | 0.02 | 0.74 (0.55 - 0.99) | 0.05 |
| ODOD | 11,041 | 27,017 | 5.8 | 21.2 | 109 | 680 | 0.78 (0.61 - 0.99) | 0.04 | 0.78 (0.61 - 0.99) | 0.06 |
| MDCR | 1,026 | 7,581 | 0.6 | 7.1 | 11 | 286 | 0.49 (0.23 - 0.91) | 0.04 | 0.41 (0.21 - 0.80) | 0.01 |
| MDCD | 1,819 | 6,786 | 1.0 | 4.8 | 18 | 181 | 0.49 (0.28 - 0.80) | <0.01 | 0.49 (0.29 - 0.83) | <0.01 |
| Open Claims | 218,502 | 414,108 | 136.1 | 361.1 | 1,041 | 6,491 | 0.73 (0.68 - 0.79) | <0.01 | 0.76 (0.65 - 0.90) | <0.01 |
| OEHR | 13,257 | 37,672 | 3.9 | 19.3 | 41 | 325 | 0.92 (0.63 - 1.30) | 0.64 | 0.86 (0.60 - 1.23) | 0.43 |
| Meta-analysis | 263,442 | 529,511 | 157.9 | 441.0 | 1,274 | 8,006 | 0.74 (0.69 - 0.79) | <0.01 | 0.76 (0.67 - 0.86) | <0.01 |
| Meta-analysis (w/o USOC) | 44,940 | 115,403 | 21.8 | 80.0 | 233 | 1,515 | 0.74 (0.61 - 0.90) | <0.01 | 0.76 (0.62 - 0.92) | <0.01 |
| **PS-matching / on-treatment TAR without escalation** | | | | | | | | | | |
| CCAE | 13,948 | 43,925 | 7.9 | 28.6 | 44 | 264 | 0.74 (0.48 - 1.11) | 0.16 | 0.79 (0.52 - 1.21) | 0.30 |
| ODOD | 7,470 | 26,516 | 3.9 | 17.4 | 80 | 537 | 0.78 (0.56 - 1.08) | 0.14 | 0.75 (0.54 - 1.04) | 0.10 |
| MDCR | 900 | 7,079 | 0.6 | 5.8 | 11 | 205 | 0.46 (0.16 - 1.07) | 0.11 | 0.37 (0.14 - 0.96) | 0.04 |
| MDCD | 1,655 | 6,752 | 0.8 | 3.7 | 14 | 137 | 0.47 (0.21 - 0.94) | 0.05 | 0.49 (0.23 - 1.04) | 0.07 |
| Open Claims | 135,919 | 413,784 | 88.0 | 286.7 | 695 | 5,190 | 0.72 (0.64 - 0.81) | <0.01 | 0.73 (0.65 - 0.82) | <0.01 |
| OEHR | 10,256 | 37,505 | 2.7 | 15.5 | 29 | 215 | 1.01 (0.60 - 1.64) | 0.97 | 1.00 (0.60 - 1.64) | 0.89 |
| Meta-analysis | 169,248 | 528,482 | 103.2 | 351.9 | 862 | 6,343 | 0.73 (0.66 - 0.81) | <0.01 | 0.74 (0.65 - 0.84) | <0.01 |
| Meta-analysis (w/o USOC) | 33,329 | 114,698 | 15.3 | 65.2 | 167 | 1,153 | 0.77 (0.62 - 0.96) | 0.02 | 0.79 (0.63 - 0.99) | 0.05 |
| **PS-stratification / on-treatment TAR without escalation** | | | | | | | | | | |
| CCAE | 18,823 | 43,928 | 10.1 | 28.6 | 51 | 264 | 0.65 (0.46 - 0.90) | 0.01 | 0.68 (0.48 - 0.95) | 0.03 |
| ODOD | 11,041 | 27,017 | 5.3 | 17.8 | 97 | 574 | 0.74 (0.57 - 0.95) | 0.02 | 0.74 (0.57 - 0.95) | 0.03 |
| MDCR | 1,026 | 7,581 | 0.6 | 6.2 | 11 | 240 | 0.59 (0.28 - 1.11) | 0.13 | 0.48 (0.24 - 0.95) | 0.04 |
| MDCD | 1,819 | 6,786 | 0.8 | 3.7 | 15 | 138 | 0.50 (0.27 - 0.86) | 0.02 | 0.52 (0.29 - 0.92) | 0.03 |
| Open Claims | 218,502 | 414,108 | 123.0 | 287.0 | 905 | 5,220 | 0.73 (0.67 - 0.79) | <0.01 | 0.75 (0.63 - 0.89) | <0.01 |
| OEHR | 13,257 | 37,672 | 3.4 | 15.6 | 33 | 222 | 0.97 (0.63 - 1.44) | 0.87 | 0.90 (0.60 - 1.37) | 0.65 |
| Meta-analysis | 263,442 | 529,511 | 142.5 | 352.7 | 1,101 | 6,418 | 0.73 (0.67 - 0.78) | <0.01 | 0.74 (0.66 - 0.83) | <0.01 |
| Meta-analysis (w/o USOC) | 44,940 | 115,403 | 19.6 | 65.7 | 196 | 1,198 | 0.72 (0.58 - 0.89) | <0.01 | 0.74 (0.60 - 0.91) | <0.01 |

##### 1.4.3 Acute myocardial infarction

**Supplementary Table 81:** Exposures, outcomes and risk estimates for Acute myocardial infarction in GLP1RA (female) (T) vs DPP4I (female) (C) new-user cohorts under four different propensity score (PS) adjustment / time-at-risk (TAR) designs across data sources. We report total patient exposure time in 1,000 patient-year (KYr) units, uncalibrated and calibrated hazard ratios (HRs) and their 95% confidence intervals (CIs). Greyed out entries did not pass study diagnostics and are included only for completeness.

| | Patients | | Exposure (KYr) | | Outcomes | | Uncalibrated | | Calibrated | |
|---|---|---|---|---|---|---|---|---|---|---|
| | T | C | T | C | T | C | HR (95% CI) | $p$ | HR (95% CI) | $p$ |
| **PS-matching / on-treatment TAR** | | | | | | | | | | |
| CCAE | 14,309 | 45,022 | 9.0 | 35.5 | 22 | 85 | 1.67 (0.82 - 3.33) | 0.15 | 1.81 (0.90 - 3.66) | 0.10 |
| ODOD | 7,825 | 28,313 | 4.6 | 22.4 | 24 | 126 | 1.83 (1.01 - 3.22) | 0.04 | 1.78 (0.99 - 3.19) | 0.06 |
| MDCR | 972 | 8,213 | 0.6 | 7.8 | <5 | 67 | 0.70 (0.04 - 3.72) | 0.76 | 0.58 (0.06 - 5.75) | 0.64 |
| MDCD | 1,822 | 7,496 | 1.0 | 5.3 | <5 | 32 | 0.75 (0.16 - 2.44) | 0.67 | 0.76 (0.20 - 2.94) | 0.70 |
| Open Claims | 142,472 | 439,605 | 103.9 | 385.4 | 226 | 1,658 | 0.84 (0.70 - 1.01) | 0.07 | 0.85 (0.68 - 1.05) | 0.16 |
| OEHR | 10,487 | 38,386 | 3.2 | 19.9 | 12 | 69 | 1.33 (0.58 - 2.85) | 0.48 | 1.28 (0.58 - 2.84) | 0.54 |
| Meta-analysis | 176,915 | 558,822 | 121.7 | 468.6 | <289 | 1,970 | 1.20 (0.81 - 1.79) | 0.37 | 1.21 (0.81 - 1.81) | 0.36 |
| Meta-analysis (w/o USOC) | 34,443 | 119,217 | 17.8 | 83.2 | <63 | 312 | 1.55 (1.07 - 2.25) | 0.02 | 1.57 (1.08 - 2.29) | 0.02 |
| **PS-stratification / on-treatment TAR** | | | | | | | | | | |
| CCAE | 19,305 | 45,027 | 11.4 | 35.5 | 25 | 85 | 1.15 (0.68 - 1.88) | 0.59 | 1.20 (0.72 - 1.99) | 0.49 |
| ODOD | 11,516 | 28,975 | 6.1 | 23.0 | 30 | 147 | 1.47 (0.91 - 2.32) | 0.11 | 1.46 (0.91 - 2.35) | 0.12 |
| MDCR | 1,111 | 8,520 | 0.7 | 8.0 | <5 | 74 | 0.35 (0.05 - 1.25) | 0.19 | 0.29 (0.06 - 1.39) | 0.12 |
| MDCD | 1,994 | 7,556 | 1.1 | 5.4 | <5 | 33 | 0.78 (0.22 - 2.16) | 0.67 | 0.79 (0.25 - 2.49) | 0.68 |
| Open Claims | 227,850 | 440,479 | 142.0 | 386.3 | 281 | 1,670 | 0.83 (0.71 - 0.95) | 0.01 | 0.86 (0.70 - 1.05) | 0.16 |
| OEHR | 13,546 | 38,583 | 4.0 | 20.0 | 15 | 69 | 1.58 (0.80 - 2.94) | 0.17 | 1.48 (0.77 - 2.84) | 0.25 |
| Meta-analysis | 274,211 | 560,620 | 164.6 | 470.3 | <356 | 2,004 | 1.09 (0.80 - 1.49) | 0.58 | 1.12 (0.81 - 1.56) | 0.50 |
| Meta-analysis (w/o USOC) | 46,361 | 120,141 | 22.6 | 84.0 | <75 | 334 | 1.32 (0.98 - 1.77) | 0.07 | 1.34 (1.00 - 1.80) | 0.06 |
| **PS-matching / on-treatment TAR without escalation** | | | | | | | | | | |
| CCAE | 14,309 | 45,022 | 8.1 | 29.2 | 19 | 58 | 1.71 (0.81 - 3.49) | 0.15 | 1.82 (0.88 - 3.79) | 0.11 |
| ODOD | 7,825 | 28,313 | 4.1 | 18.8 | 20 | 113 | 1.39 (0.72 - 2.57) | 0.30 | 1.34 (0.71 - 2.52) | 0.37 |
| MDCR | 972 | 8,213 | 0.6 | 6.8 | <5 | 58 | 0.69 (0.04 - 3.67) | 0.75 | 0.55 (0.06 - 5.51) | 0.62 |
| MDCD | 1,822 | 7,496 | 0.9 | 4.2 | <5 | 23 | 1.17 (0.25 - 4.03) | 0.82 | 1.23 (0.31 - 4.92) | 0.77 |
| Open Claims | 142,472 | 439,605 | 92.0 | 306.1 | 196 | 1,336 | 0.84 (0.68 - 1.03) | 0.10 | 0.85 (0.69 - 1.05) | 0.16 |
| OEHR | 10,487 | 38,386 | 2.8 | 16.0 | 9 | 42 | 1.51 (0.54 - 3.92) | 0.41 | 1.49 (0.55 - 4.03) | 0.44 |
| Meta-analysis | 176,915 | 558,822 | 107.9 | 374.3 | <249 | 1,572 | 1.15 (0.79 - 1.68) | 0.46 | 1.17 (0.80 - 1.71) | 0.43 |
| Meta-analysis (w/o USOC) | 34,443 | 119,217 | 15.8 | 68.2 | <53 | 236 | 1.48 (0.98 - 2.24) | 0.06 | 1.52 (1.00 - 2.30) | 0.05 |
| **PS-stratification / on-treatment TAR without escalation** | | | | | | | | | | |
| CCAE | 19,305 | 45,027 | 10.3 | 29.2 | 22 | 58 | 1.36 (0.76 - 2.36) | 0.28 | 1.42 (0.81 - 2.50) | 0.23 |
| ODOD | 11,516 | 28,975 | 5.6 | 19.3 | 26 | 132 | 1.24 (0.73 - 2.01) | 0.41 | 1.23 (0.74 - 2.04) | 0.43 |
| MDCR | 1,111 | 8,520 | 0.6 | 7.0 | <5 | 65 | 0.45 (0.07 - 1.67) | 0.32 | 0.36 (0.07 - 1.81) | 0.22 |
| MDCD | 1,994 | 7,556 | 0.9 | 4.2 | <5 | 23 | 1.34 (0.36 - 3.87) | 0.63 | 1.40 (0.43 - 4.57) | 0.58 |
| Open Claims | 227,850 | 440,479 | 128.1 | 306.8 | 251 | 1,346 | 0.85 (0.72 - 1.00) | 0.05 | 0.87 (0.70 - 1.09) | 0.25 |
| OEHR | 13,546 | 38,583 | 3.5 | 16.1 | 12 | 42 | 1.83 (0.82 - 3.83) | 0.13 | 1.71 (0.79 - 3.71) | 0.18 |
| Meta-analysis | 274,211 | 560,620 | 148.4 | 375.7 | <316 | 1,601 | 1.14 (0.83 - 1.57) | 0.42 | 1.16 (0.84 - 1.62) | 0.37 |
| Meta-analysis (w/o USOC) | 46,361 | 120,141 | 20.3 | 68.9 | <65 | 255 | 1.38 (0.99 - 1.91) | 0.05 | 1.41 (1.02 - 1.95) | 0.04 |

##### 1.4.4 Acute renal failure

**Supplementary Table 82:** Exposures, outcomes and risk estimates for Acute renal failure in GLP1RA (female) (T) vs DPP4I (female) (C) new-user cohorts under four different propensity score (PS) adjustment / time-at-risk (TAR) designs across data sources. We report total patient exposure time in 1,000 patient-year (KYr) units, uncalibrated and calibrated hazard ratios (HRs) and their 95% confidence intervals (CIs). Greyed out entries did not pass study diagnostics and are included only for completeness.

| | Patients | | Exposure (KYr) | | Outcomes | | Uncalibrated | | Calibrated | |
|---|---|---|---|---|---|---|---|---|---|---|
| | T | C | T | C | T | C | HR (95% CI) | $p$ | HR (95% CI) | $p$ |
| **PS-matching / on-treatment TAR** | | | | | | | | | | |
| CCAE | 14,295 | 44,953 | 9.0 | 35.5 | 41 | 225 | 0.48 (0.29 - 0.77) | <0.01 | 0.52 (0.32 - 0.85) | 0.01 |
| ODOD | 7,660 | 27,597 | 4.5 | 21.6 | 61 | 584 | 0.66 (0.46 - 0.92) | 0.02 | 0.64 (0.45 - 0.91) | 0.02 |
| MDCR | 950 | 8,006 | 0.6 | 7.5 | 10 | 211 | 0.61 (0.23 - 1.39) | 0.29 | 0.50 (0.20 - 1.25) | 0.14 |
| MDCD | 1,767 | 7,296 | 1.0 | 5.1 | 16 | 171 | 0.73 (0.38 - 1.28) | 0.30 | 0.74 (0.41 - 1.36) | 0.34 |
| Open Claims | 140,798 | 433,196 | 102.4 | 377.3 | 735 | 5,505 | 0.72 (0.65 - 0.80) | <0.01 | 0.73 (0.63 - 0.85) | <0.01 |
| OEHR | 10,454 | 38,171 | 3.2 | 19.7 | 38 | 259 | 1.27 (0.83 - 1.91) | 0.26 | 1.22 (0.80 - 1.86) | 0.36 |
| Meta-analysis | 174,974 | 551,213 | 120.0 | 459.2 | 891 | 6,744 | 0.74 (0.56 - 0.97) | 0.03 | 0.74 (0.56 - 0.99) | 0.04 |
| Meta-analysis (w/o USOC) | 34,176 | 118,017 | 17.6 | 81.9 | 156 | 1,239 | 0.74 (0.49 - 1.12) | 0.15 | 0.75 (0.50 - 1.14) | 0.18 |
| **PS-stratification / on-treatment TAR** | | | | | | | | | | |
| CCAE | 19,257 | 44,957 | 11.4 | 35.5 | 59 | 225 | 0.64 (0.46 - 0.89) | <0.01 | 0.67 (0.48 - 0.93) | 0.02 |
| ODOD | 11,351 | 28,091 | 6.0 | 22.1 | 79 | 613 | 0.62 (0.47 - 0.82) | <0.01 | 0.62 (0.47 - 0.82) | <0.01 |
| MDCR | 1,089 | 8,325 | 0.7 | 7.8 | 10 | 238 | 0.64 (0.30 - 1.20) | 0.20 | 0.53 (0.26 - 1.06) | 0.07 |
| MDCD | 1,939 | 7,347 | 1.0 | 5.1 | 16 | 172 | 0.50 (0.28 - 0.84) | 0.01 | 0.51 (0.29 - 0.88) | 0.02 |
| Open Claims | 225,549 | 433,780 | 140.2 | 377.9 | 937 | 5,535 | 0.72 (0.67 - 0.78) | <0.01 | 0.75 (0.64 - 0.89) | <0.01 |
| OEHR | 13,485 | 38,353 | 4.0 | 19.8 | 45 | 267 | 1.19 (0.83 - 1.67) | 0.34 | 1.11 (0.78 - 1.59) | 0.57 |
| Meta-analysis | 271,581 | 552,528 | 162.6 | 460.4 | 1,136 | 6,812 | 0.72 (0.57 - 0.91) | <0.01 | 0.74 (0.57 - 0.96) | 0.02 |
| Meta-analysis (w/o USOC) | 46,032 | 118,748 | 22.4 | 82.4 | 199 | 1,277 | 0.71 (0.51 - 1.01) | 0.05 | 0.73 (0.51 - 1.02) | 0.07 |
| **PS-matching / on-treatment TAR without escalation** | | | | | | | | | | |
| CCAE | 14,295 | 44,953 | 8.1 | 29.2 | 36 | 177 | 0.47 (0.27 - 0.77) | <0.01 | 0.50 (0.30 - 0.85) | 0.01 |
| ODOD | 7,660 | 27,597 | 4.0 | 18.2 | 52 | 476 | 0.66 (0.44 - 0.97) | 0.04 | 0.64 (0.43 - 0.95) | 0.03 |
| MDCR | 950 | 8,006 | 0.6 | 6.6 | 9 | 177 | 0.54 (0.16 - 1.38) | 0.26 | 0.43 (0.15 - 1.27) | 0.13 |
| MDCD | 1,767 | 7,296 | 0.8 | 4.0 | 14 | 130 | 0.80 (0.40 - 1.48) | 0.50 | 0.84 (0.43 - 1.62) | 0.60 |
| Open Claims | 140,798 | 433,196 | 90.7 | 300.0 | 626 | 4,363 | 0.71 (0.63 - 0.80) | <0.01 | 0.72 (0.64 - 0.81) | <0.01 |
| OEHR | 10,454 | 38,171 | 2.8 | 15.9 | 23 | 178 | 1.07 (0.62 - 1.78) | 0.79 | 1.06 (0.63 - 1.79) | 0.83 |
| Meta-analysis | 174,974 | 551,213 | 106.4 | 367.3 | 751 | 5,324 | 0.71 (0.64 - 0.79) | <0.01 | 0.72 (0.63 - 0.82) | <0.01 |
| Meta-analysis (w/o USOC) | 34,176 | 118,017 | 15.7 | 67.2 | 125 | 961 | 0.71 (0.51 - 0.99) | 0.04 | 0.72 (0.52 - 1.01) | 0.06 |
| **PS-stratification / on-treatment TAR without escalation** | | | | | | | | | | |
| CCAE | 19,257 | 44,957 | 10.3 | 29.2 | 53 | 177 | 0.65 (0.46 - 0.93) | 0.02 | 0.68 (0.48 - 0.97) | 0.04 |
| ODOD | 11,351 | 28,091 | 5.5 | 18.6 | 69 | 501 | 0.61 (0.45 - 0.82) | <0.01 | 0.61 (0.45 - 0.83) | <0.01 |
| MDCR | 1,089 | 8,325 | 0.6 | 6.8 | 9 | 198 | 0.60 (0.27 - 1.18) | 0.18 | 0.49 (0.24 - 1.02) | 0.06 |
| MDCD | 1,939 | 7,347 | 0.9 | 4.0 | 14 | 131 | 0.56 (0.30 - 0.98) | 0.06 | 0.59 (0.32 - 1.07) | 0.09 |
| Open Claims | 225,549 | 433,780 | 126.5 | 300.5 | 810 | 4,388 | 0.70 (0.64 - 0.77) | <0.01 | 0.72 (0.61 - 0.86) | <0.01 |
| OEHR | 13,485 | 38,353 | 3.5 | 16.0 | 30 | 184 | 0.95 (0.61 - 1.45) | 0.83 | 0.89 (0.58 - 1.38) | 0.63 |
| Meta-analysis | 271,581 | 552,528 | 146.6 | 368.2 | 976 | 5,381 | 0.70 (0.64 - 0.76) | <0.01 | 0.71 (0.63 - 0.80) | <0.01 |
| Meta-analysis (w/o USOC) | 46,032 | 118,748 | 20.1 | 67.7 | 166 | 993 | 0.68 (0.55 - 0.83) | <0.01 | 0.69 (0.56 - 0.85) | <0.01 |

##### 1.4.5 Glycemic control

**Supplementary Table 83:** Exposures, outcomes and risk estimates for Glycemic control in GLP1RA (female) (T) vs DPP4I (female) (C) new-user cohorts under four different propensity score (PS) adjustment / time-at-risk (TAR) designs across data sources. We report total patient exposure time in 1,000 patient-year (KYr) units, uncalibrated and calibrated hazard ratios (HRs) and their 95% confidence intervals (CIs). Greyed out entries did not pass study diagnostics and are included only for completeness.

| | Patients | | Exposure (KYr) | | Outcomes | | Uncalibrated | | Calibrated | |
|---|---|---|---|---|---|---|---|---|---|---|
| | T | C | T | C | T | C | HR (95% CI) | $p$ | HR (95% CI) | $p$ |
| **PS-matching / on-treatment TAR** | | | | | | | | | | |
| CCAE | 14,306 | 44,884 | 9.0 | 35.5 | 18 | 111 | 1.37 (0.69 - 2.59) | 0.35 | 1.48 (0.77 - 2.87) | 0.25 |
| ODOD | 5,103 | 18,673 | 2.8 | 13.2 | 316 | 1,420 | 1.22 (1.02 - 1.44) | 0.03 | 1.19 (1.00 - 1.41) | 0.08 |
| MDCR | 981 | 8,392 | 0.6 | 8.0 | 0 | 15 | | | | |
| MDCD | 1,861 | 7,692 | 1.0 | 5.5 | 0 | 0 | | | | |
| Open Claims | 144,854 | 448,754 | 105.6 | 394.3 | 0 | 0 | | | | |
| OEHR | 10,563 | 38,693 | 3.3 | 20.1 | 0 | 0 | | | | |
| Meta-analysis | 19,409 | 63,557 | 11.8 | 48.7 | 334 | 1,531 | 1.23 (1.04 - 1.45) | 0.02 | 1.24 (1.03 - 1.48) | 0.02 |
| Meta-analysis (w/o USOC) | 19,409 | 63,557 | 11.8 | 48.7 | 334 | 1,531 | 1.23 (1.04 - 1.45) | 0.02 | 1.25 (1.05 - 1.48) | 0.01 |
| **PS-stratification / on-treatment TAR** | | | | | | | | | | |
| CCAE | 19,314 | 44,889 | 11.4 | 35.5 | 19 | 111 | 0.91 (0.51 - 1.53) | 0.74 | 0.95 (0.55 - 1.64) | 0.85 |
| ODOD | 7,236 | 19,175 | 3.6 | 13.6 | 400 | 1,468 | 1.15 (1.01 - 1.31) | 0.04 | 1.14 (1.00 - 1.31) | 0.10 |
| MDCR | 1,124 | 8,751 | 0.7 | 8.3 | 0 | 15 | | | | |
| MDCD | 2,034 | 7,771 | 1.1 | 5.6 | 0 | 0 | | | | |
| Open Claims | 231,281 | 449,866 | 144.1 | 395.4 | 0 | 0 | | | | |
| OEHR | 13,638 | 38,897 | 4.0 | 20.2 | 0 | 0 | | | | |
| Meta-analysis | 26,550 | 64,064 | 15.0 | 49.1 | 419 | 1,579 | 1.13 (1.00 - 1.29) | 0.05 | 1.16 (0.99 - 1.37) | 0.07 |
| Meta-analysis (w/o USOC) | 26,550 | 64,064 | 15.0 | 49.1 | 419 | 1,579 | 1.13 (1.00 - 1.29) | 0.05 | 1.15 (1.01 - 1.32) | 0.05 |
| **PS-matching / on-treatment TAR without escalation** | | | | | | | | | | |
| CCAE | 14,306 | 44,884 | 8.1 | 29.2 | 18 | 97 | 1.53 (0.76 - 3.00) | 0.22 | 1.64 (0.82 - 3.26) | 0.17 |
| ODOD | 5,103 | 18,673 | 2.5 | 11.1 | 288 | 1,187 | 1.24 (1.03 - 1.49) | 0.02 | 1.19 (0.99 - 1.43) | 0.10 |
| MDCR | 981 | 8,392 | 0.6 | 6.9 | 0 | 12 | | | | |
| MDCD | 1,861 | 7,692 | 0.9 | 4.3 | 0 | 0 | | | | |
| Open Claims | 144,854 | 448,754 | 93.5 | 313.0 | 0 | 0 | | | | |
| OEHR | 10,563 | 38,693 | 2.8 | 16.2 | 0 | 0 | | | | |
| Meta-analysis | 19,409 | 63,557 | 10.6 | 40.2 | 306 | 1,284 | 1.26 (1.05 - 1.50) | 0.01 | 1.27 (1.05 - 1.54) | 0.01 |
| Meta-analysis (w/o USOC) | 19,409 | 63,557 | 10.6 | 40.2 | 306 | 1,284 | 1.26 (1.05 - 1.50) | 0.01 | 1.29 (1.07 - 1.54) | <0.01 |
| **PS-stratification / on-treatment TAR without escalation** | | | | | | | | | | |
| CCAE | 19,314 | 44,889 | 10.3 | 29.2 | 19 | 97 | 0.98 (0.55 - 1.67) | 0.96 | 1.02 (0.59 - 1.79) | 0.93 |
| ODOD | 7,236 | 19,175 | 3.3 | 11.4 | 365 | 1,230 | 1.19 (1.04 - 1.37) | 0.01 | 1.19 (1.03 - 1.38) | 0.04 |
| MDCR | 1,124 | 8,751 | 0.6 | 7.2 | 0 | 12 | | | | |
| MDCD | 2,034 | 7,771 | 0.9 | 4.3 | 0 | 0 | | | | |
| Open Claims | 231,281 | 449,866 | 130.0 | 313.9 | 0 | 0 | | | | |
| OEHR | 13,638 | 38,897 | 3.5 | 16.3 | 0 | 0 | | | | |
| Meta-analysis | 26,550 | 64,064 | 13.6 | 40.6 | 384 | 1,327 | 1.18 (1.03 - 1.35) | 0.02 | 1.21 (1.03 - 1.41) | 0.02 |
| Meta-analysis (w/o USOC) | 26,550 | 64,064 | 13.6 | 40.6 | 384 | 1,327 | 1.18 (1.03 - 1.35) | 0.02 | 1.21 (1.05 - 1.39) | 0.01 |

##### 1.4.6 Hospitalization with heart failure

**Supplementary Table 84:** Exposures, outcomes and risk estimates for Hospitalization with heart failure in GLP1RA (female) (T) vs DPP4I (female) (C) new-user cohorts under four different propensity score (PS) adjustment / time-at-risk (TAR) designs across data sources. We report total patient exposure time in 1,000 patient-year (KYr) units, uncalibrated and calibrated hazard ratios (HRs) and their 95% confidence intervals (CIs). Greyed out entries did not pass study diagnostics and are included only for completeness.

| | Patients | | Exposure (KYr) | | Outcomes | | Uncalibrated | | Calibrated | |
|---|---|---|---|---|---|---|---|---|---|---|
| | T | C | T | C | T | C | HR (95% CI) | $p$ | HR (95% CI) | $p$ |
| **PS-matching / on-treatment TAR** | | | | | | | | | | |
| CCAE | 14,194 | 44,751 | 9.0 | 35.4 | 15 | 177 | 0.22 (0.09 - 0.46) | <0.01 | 0.24 (0.11 - 0.54) | <0.01 |
| ODOD | 7,628 | 27,438 | 4.4 | 21.5 | 59 | 451 | 0.74 (0.49 - 1.10) | 0.15 | 0.72 (0.49 - 1.08) | 0.13 |
| MDCR | 932 | 7,765 | 0.6 | 7.4 | 9 | 187 | 0.58 (0.17 - 1.51) | 0.33 | 0.48 (0.16 - 1.43) | 0.19 |
| MDCD | 1,715 | 7,075 | 0.9 | 5.0 | 15 | 124 | 0.58 (0.27 - 1.14) | 0.14 | 0.59 (0.29 - 1.22) | 0.16 |
| Open Claims | 140,316 | 430,165 | 102.3 | 376.1 | 479 | 4,266 | 0.73 (0.64 - 0.83) | <0.01 | 0.74 (0.63 - 0.87) | <0.01 |
| OEHR | 10,378 | 38,019 | 3.2 | 19.6 | 21 | 234 | 0.81 (0.45 - 1.37) | 0.45 | 0.78 (0.45 - 1.35) | 0.38 |
| Meta-analysis | 174,231 | 547,448 | 119.8 | 457.6 | 589 | 5,252 | 0.65 (0.49 - 0.86) | <0.01 | 0.65 (0.49 - 0.87) | <0.01 |
| Meta-analysis (w/o USOC) | 33,915 | 117,283 | 17.5 | 81.5 | 110 | 986 | 0.57 (0.35 - 0.93) | 0.02 | 0.58 (0.36 - 0.94) | 0.03 |
| **PS-stratification / on-treatment TAR** | | | | | | | | | | |
| CCAE | 19,133 | 44,755 | 11.3 | 35.4 | 18 | 177 | 0.31 (0.18 - 0.51) | <0.01 | 0.33 (0.19 - 0.55) | <0.01 |
| ODOD | 11,259 | 27,920 | 6.0 | 22.0 | 66 | 482 | 0.76 (0.56 - 1.01) | 0.07 | 0.75 (0.56 - 1.02) | 0.08 |
| MDCR | 1,067 | 8,032 | 0.7 | 7.6 | 9 | 203 | 0.66 (0.29 - 1.33) | 0.29 | 0.55 (0.26 - 1.19) | 0.13 |
| MDCD | 1,878 | 7,124 | 1.0 | 5.0 | 16 | 125 | 0.66 (0.36 - 1.13) | 0.15 | 0.67 (0.38 - 1.18) | 0.17 |
| Open Claims | 224,856 | 430,696 | 140.1 | 376.6 | 587 | 4,304 | 0.71 (0.64 - 0.79) | <0.01 | 0.74 (0.62 - 0.88) | <0.01 |
| OEHR | 13,410 | 38,193 | 4.0 | 19.7 | 25 | 241 | 0.76 (0.47 - 1.18) | 0.24 | 0.71 (0.45 - 1.13) | 0.16 |
| Meta-analysis | 270,536 | 548,688 | 162.3 | 458.7 | 712 | 5,329 | 0.64 (0.49 - 0.84) | <0.01 | 0.66 (0.49 - 0.88) | <0.01 |
| Meta-analysis (w/o USOC) | 45,680 | 117,992 | 22.2 | 82.1 | 125 | 1,025 | 0.60 (0.41 - 0.89) | 0.01 | 0.61 (0.42 - 0.91) | 0.01 |
| **PS-matching / on-treatment TAR without escalation** | | | | | | | | | | |
| CCAE | 14,194 | 44,751 | 8.0 | 29.1 | 10 | 148 | 0.17 (0.06 - 0.40) | <0.01 | 0.18 (0.07 - 0.48) | <0.01 |
| ODOD | 7,628 | 27,438 | 4.0 | 18.1 | 51 | 370 | 0.78 (0.50 - 1.19) | 0.26 | 0.75 (0.48 - 1.16) | 0.21 |
| MDCR | 932 | 7,765 | 0.6 | 6.4 | 8 | 159 | 0.58 (0.14 - 1.67) | 0.39 | 0.46 (0.13 - 1.62) | 0.23 |
| MDCD | 1,715 | 7,075 | 0.8 | 3.9 | 12 | 93 | 0.75 (0.33 - 1.56) | 0.47 | 0.78 (0.36 - 1.72) | 0.55 |
| Open Claims | 140,316 | 430,165 | 90.7 | 298.9 | 409 | 3,403 | 0.75 (0.65 - 0.86) | <0.01 | 0.76 (0.65 - 0.88) | <0.01 |
| OEHR | 10,378 | 38,019 | 2.8 | 15.8 | 16 | 166 | 0.79 (0.41 - 1.44) | 0.47 | 0.78 (0.42 - 1.47) | 0.46 |
| Meta-analysis | 174,231 | 547,448 | 106.3 | 365.8 | 498 | 4,180 | 0.73 (0.64 - 0.83) | <0.01 | 0.74 (0.64 - 0.86) | <0.01 |
| Meta-analysis (w/o USOC) | 33,915 | 117,283 | 15.6 | 66.9 | 89 | 777 | 0.58 (0.33 - 1.03) | 0.06 | 0.60 (0.34 - 1.06) | 0.08 |
| **PS-stratification / on-treatment TAR without escalation** | | | | | | | | | | |
| CCAE | 19,133 | 44,755 | 10.2 | 29.1 | 12 | 148 | 0.22 (0.11 - 0.40) | <0.01 | 0.23 (0.12 - 0.44) | <0.01 |
| ODOD | 11,259 | 27,920 | 5.4 | 18.5 | 58 | 394 | 0.70 (0.50 - 0.96) | 0.03 | 0.70 (0.50 - 0.97) | 0.04 |
| MDCR | 1,067 | 8,032 | 0.6 | 6.6 | 8 | 175 | 0.64 (0.26 - 1.35) | 0.29 | 0.52 (0.23 - 1.19) | 0.13 |
| MDCD | 1,878 | 7,124 | 0.9 | 3.9 | 13 | 93 | 0.64 (0.33 - 1.16) | 0.16 | 0.66 (0.35 - 1.25) | 0.22 |
| Open Claims | 224,856 | 430,696 | 126.4 | 299.3 | 507 | 3,435 | 0.71 (0.63 - 0.79) | <0.01 | 0.73 (0.60 - 0.88) | <0.01 |
| OEHR | 13,410 | 38,193 | 3.4 | 15.9 | 20 | 171 | 0.78 (0.45 - 1.28) | 0.34 | 0.73 (0.43 - 1.23) | 0.25 |
| Meta-analysis | 270,536 | 548,688 | 146.4 | 366.7 | 610 | 4,241 | 0.59 (0.40 - 0.86) | <0.01 | 0.60 (0.41 - 0.89) | 0.01 |
| Meta-analysis (w/o USOC) | 45,680 | 117,992 | 20.0 | 67.4 | 103 | 806 | 0.55 (0.33 - 0.89) | 0.02 | 0.56 (0.34 - 0.92) | 0.02 |

##### 1.4.7 Measured renal dysfunction

**Supplementary Table 85:** Exposures, outcomes and risk estimates for Measured renal dysfunction in GLP1RA (female) (T) vs DPP4I (female) (C) new-user cohorts under four different propensity score (PS) adjustment / time-at-risk (TAR) designs across data sources. We report total patient exposure time in 1,000 patient-year (KYr) units, uncalibrated and calibrated hazard ratios (HRs) and their 95% confidence intervals (CIs). Greyed out entries did not pass study diagnostics and are included only for completeness.

| | Patients | | Exposure (KYr) | | Outcomes | | Uncalibrated | | Calibrated | |
|---|---|---|---|---|---|---|---|---|---|---|
| | T | C | T | C | T | C | HR (95% CI) | $p$ | HR (95% CI) | $p$ |
| **PS-matching / on-treatment TAR** | | | | | | | | | | |
| CCAE | 14,416 | 45,460 | 9.1 | 36.0 | <5 | 7 | 1.69 (0.20 - 14.3) | 0.63 | 1.83 (0.22 - 15.5) | 0.58 |
| ODOD | 7,909 | 28,772 | 4.6 | 22.8 | <5 | 46 | 0.42 (0.09 - 1.40) | 0.21 | 0.41 (0.11 - 1.59) | 0.20 |
| MDCR | 986 | 8,446 | 0.6 | 8.0 | 0 | 0 | | | | |
| MDCD | 1,861 | 7,692 | 1.0 | 5.5 | 0 | 0 | | | | |
| Open Claims | 144,854 | 448,754 | 105.6 | 394.3 | 0 | 0 | | | | |
| OEHR | 10,563 | 38,693 | 3.3 | 20.1 | 0 | 0 | | | | |
| Meta-analysis | 7,909 | 28,772 | 4.6 | 22.8 | <5 | 46 | 0.42 (0.09 - 1.40) | 0.21 | 0.43 (0.11 - 1.65) | 0.22 |
| Meta-analysis (w/o USOC) | 7,909 | 28,772 | 4.6 | 22.8 | <5 | 46 | 0.42 (0.09 - 1.40) | 0.21 | 0.43 (0.11 - 1.66) | 0.22 |
| **PS-stratification / on-treatment TAR** | | | | | | | | | | |
| CCAE | 19,418 | 45,465 | 11.5 | 36.0 | <5 | 7 | 2.49 (0.34 - 12.1) | 0.32 | 2.59 (0.43 - 15.5) | 0.30 |
| ODOD | 11,650 | 29,464 | 6.2 | 23.4 | <5 | 55 | 0.42 (0.10 - 1.28) | 0.19 | 0.42 (0.11 - 1.53) | 0.19 |
| MDCR | 1,128 | 8,813 | 0.7 | 8.3 | 0 | 0 | | | | |
| MDCD | 2,034 | 7,771 | 1.1 | 5.6 | 0 | 0 | | | | |
| Open Claims | 231,281 | 449,866 | 144.1 | 395.4 | 0 | 0 | | | | |
| OEHR | 13,638 | 38,897 | 4.0 | 20.2 | 0 | 0 | | | | |
| Meta-analysis | 11,650 | 29,464 | 6.2 | 23.4 | <5 | 55 | 0.42 (0.10 - 1.28) | 0.19 | 0.43 (0.12 - 1.59) | 0.21 |
| Meta-analysis (w/o USOC) | 11,650 | 29,464 | 6.2 | 23.4 | <5 | 55 | 0.42 (0.10 - 1.28) | 0.19 | 0.43 (0.12 - 1.57) | 0.20 |
| **PS-matching / on-treatment TAR without escalation** | | | | | | | | | | |
| CCAE | 14,416 | 45,460 | 8.2 | 29.6 | <5 | 6 | 3.24 (0.30 - 70.6) | 0.40 | 3.46 (0.23 - 52.9) | 0.37 |
| ODOD | 7,909 | 28,772 | 4.1 | 19.1 | <5 | 37 | 0.52 (0.08 - 2.29) | 0.46 | 0.50 (0.09 - 2.78) | 0.43 |
| MDCR | 986 | 8,446 | 0.6 | 7.0 | 0 | 0 | | | | |
| MDCD | 1,861 | 7,692 | 0.9 | 4.3 | 0 | 0 | | | | |
| Open Claims | 144,854 | 448,754 | 93.5 | 313.0 | 0 | 0 | | | | |
| OEHR | 10,563 | 38,693 | 2.8 | 16.2 | 0 | 0 | | | | |
| Meta-analysis | 7,909 | 28,772 | 4.1 | 19.1 | <5 | 37 | 0.52 (0.08 - 2.29) | 0.46 | 0.53 (0.10 - 2.93) | 0.47 |
| Meta-analysis (w/o USOC) | 7,909 | 28,772 | 4.1 | 19.1 | <5 | 37 | 0.52 (0.08 - 2.29) | 0.46 | 0.54 (0.10 - 2.96) | 0.48 |
| **PS-stratification / on-treatment TAR without escalation** | | | | | | | | | | |
| CCAE | 19,418 | 45,465 | 10.4 | 29.6 | <5 | 6 | 2.47 (0.33 - 12.4) | 0.33 | 2.57 (0.42 - 15.7) | 0.31 |
| ODOD | 11,650 | 29,464 | 5.6 | 19.6 | <5 | 46 | 0.62 (0.13 - 1.98) | 0.48 | 0.61 (0.16 - 2.36) | 0.48 |
| MDCR | 1,128 | 8,813 | 0.6 | 7.2 | 0 | 0 | | | | |
| MDCD | 2,034 | 7,771 | 0.9 | 4.3 | 0 | 0 | | | | |
| Open Claims | 231,281 | 449,866 | 130.0 | 313.9 | 0 | 0 | | | | |
| OEHR | 13,638 | 38,897 | 3.5 | 16.3 | 0 | 0 | | | | |
| Meta-analysis | 11,650 | 29,464 | 5.6 | 19.6 | <5 | 46 | 0.62 (0.13 - 1.98) | 0.48 | 0.63 (0.16 - 2.42) | 0.50 |
| Meta-analysis (w/o USOC) | 11,650 | 29,464 | 5.6 | 19.6 | <5 | 46 | 0.62 (0.13 - 1.98) | 0.48 | 0.63 (0.16 - 2.42) | 0.50 |

##### 1.4.8 Stroke

**Supplementary Table 86:** Exposures, outcomes and risk estimates for Stroke in GLP1RA (female) (T) vs DPP4I (female) (C) new-user cohorts under four different propensity score (PS) adjustment / time-at-risk (TAR) designs across data sources. We report total patient exposure time in 1,000 patient-year (KYr) units, uncalibrated and calibrated hazard ratios (HRs) and their 95% confidence intervals (CIs). Greyed out entries did not pass study diagnostics and are included only for completeness.

| | Patients | | Exposure (KYr) | | Outcomes | | Uncalibrated | | Calibrated | |
|---|---|---|---|---|---|---|---|---|---|---|
| | T | C | T | C | T | C | HR (95% CI) | $p$ | HR (95% CI) | $p$ |
| **PS-matching / on-treatment TAR** | | | | | | | | | | |
| CCAE | 14,268 | 44,964 | 9.0 | 35.6 | 22 | 123 | 0.74 (0.39 - 1.32) | 0.33 | 0.80 (0.43 - 1.47) | 0.48 |
| ODOD | 7,799 | 28,030 | 4.5 | 22.1 | 27 | 234 | 0.88 (0.52 - 1.44) | 0.62 | 0.86 (0.51 - 1.43) | 0.56 |
| MDCR | 957 | 7,797 | 0.6 | 7.4 | <5 | 108 | 0.62 (0.14 - 1.89) | 0.47 | 0.51 (0.14 - 1.87) | 0.31 |
| MDCD | 1,805 | 7,419 | 1.0 | 5.3 | 8 | 69 | 0.50 (0.14 - 1.34) | 0.22 | 0.51 (0.17 - 1.56) | 0.24 |
| Open Claims | 141,653 | 435,868 | 103.4 | 382.2 | 309 | 2,323 | 0.79 (0.67 - 0.93) | <0.01 | 0.79 (0.65 - 0.97) | 0.03 |
| OEHR | 10,486 | 38,365 | 3.2 | 19.9 | 9 | 80 | 1.23 (0.48 - 2.87) | 0.65 | 1.18 (0.48 - 2.89) | 0.71 |
| Meta-analysis | 176,011 | 554,646 | 121.1 | 465.0 | 375 | 2,829 | 0.79 (0.68 - 0.92) | <0.01 | 0.80 (0.68 - 0.94) | <0.01 |
| Meta-analysis (w/o USOC) | 34,358 | 118,778 | 17.7 | 82.8 | 66 | 506 | 0.83 (0.59 - 1.17) | 0.28 | 0.84 (0.60 - 1.19) | 0.33 |
| **PS-stratification / on-treatment TAR** | | | | | | | | | | |
| CCAE | 19,232 | 44,968 | 11.4 | 35.6 | 29 | 123 | 0.92 (0.57 - 1.44) | 0.73 | 0.96 (0.60 - 1.52) | 0.86 |
| ODOD | 11,498 | 28,658 | 6.1 | 22.7 | 35 | 254 | 0.72 (0.47 - 1.08) | 0.12 | 0.71 (0.47 - 1.09) | 0.13 |
| MDCR | 1,094 | 8,390 | 0.7 | 7.9 | <5 | 120 | 0.44 (0.13 - 1.13) | 0.14 | 0.36 (0.12 - 1.08) | 0.07 |
| MDCD | 1,977 | 7,475 | 1.1 | 5.3 | 8 | 69 | 0.65 (0.27 - 1.35) | 0.29 | 0.66 (0.30 - 1.46) | 0.31 |
| Open Claims | 226,789 | 436,800 | 141.4 | 383.1 | 396 | 2,350 | 0.78 (0.69 - 0.88) | <0.01 | 0.81 (0.67 - 0.98) | 0.04 |
| OEHR | 13,541 | 38,563 | 4.0 | 20.0 | 9 | 86 | 0.83 (0.37 - 1.68) | 0.64 | 0.78 (0.36 - 1.67) | 0.53 |
| Meta-analysis | 273,037 | 556,464 | 163.9 | 466.7 | 477 | 2,882 | 0.78 (0.70 - 0.88) | <0.01 | 0.80 (0.69 - 0.94) | <0.01 |
| Meta-analysis (w/o USOC) | 46,248 | 119,664 | 22.5 | 83.6 | 81 | 532 | 0.79 (0.60 - 1.03) | 0.08 | 0.80 (0.61 - 1.05) | 0.12 |
| **PS-matching / on-treatment TAR without escalation** | | | | | | | | | | |
| CCAE | 14,268 | 44,964 | 8.1 | 29.3 | 16 | 103 | 0.67 (0.33 - 1.29) | 0.26 | 0.72 (0.36 - 1.43) | 0.35 |
| ODOD | 7,799 | 28,030 | 4.1 | 18.5 | 23 | 207 | 0.74 (0.42 - 1.26) | 0.29 | 0.71 (0.41 - 1.24) | 0.24 |
| MDCR | 957 | 7,797 | 0.6 | 6.4 | <5 | 86 | 0.82 (0.19 - 2.56) | 0.77 | 0.66 (0.18 - 2.45) | 0.54 |
| MDCD | 1,805 | 7,419 | 0.9 | 4.1 | 7 | 57 | 0.49 (0.11 - 1.49) | 0.28 | 0.51 (0.14 - 1.87) | 0.31 |
| Open Claims | 141,653 | 435,868 | 91.6 | 303.4 | 263 | 1,877 | 0.75 (0.62 - 0.90) | <0.01 | 0.76 (0.63 - 0.91) | <0.01 |
| OEHR | 10,486 | 38,365 | 2.8 | 16.0 | 7 | 55 | 1.17 (0.39 - 3.14) | 0.77 | 1.15 (0.41 - 3.28) | 0.79 |
| Meta-analysis | 176,011 | 554,646 | 107.3 | 371.3 | 316 | 2,299 | 0.75 (0.63 - 0.88) | <0.01 | 0.76 (0.63 - 0.91) | <0.01 |
| Meta-analysis (w/o USOC) | 34,358 | 118,778 | 15.8 | 67.9 | 53 | 422 | 0.74 (0.50 - 1.08) | 0.12 | 0.76 (0.52 - 1.11) | 0.16 |
| **PS-stratification / on-treatment TAR without escalation** | | | | | | | | | | |
| CCAE | 19,232 | 44,968 | 10.3 | 29.3 | 21 | 103 | 0.78 (0.45 - 1.31) | 0.37 | 0.82 (0.48 - 1.39) | 0.46 |
| ODOD | 11,498 | 28,658 | 5.5 | 19.1 | 31 | 227 | 0.68 (0.43 - 1.05) | 0.09 | 0.68 (0.44 - 1.06) | 0.10 |
| MDCR | 1,094 | 8,390 | 0.6 | 6.9 | <5 | 96 | 0.56 (0.16 - 1.47) | 0.30 | 0.46 (0.15 - 1.37) | 0.17 |
| MDCD | 1,977 | 7,475 | 0.9 | 4.2 | 7 | 57 | 0.59 (0.23 - 1.31) | 0.24 | 0.62 (0.26 - 1.46) | 0.28 |
| Open Claims | 226,789 | 436,800 | 127.5 | 304.2 | 348 | 1,900 | 0.77 (0.67 - 0.88) | <0.01 | 0.79 (0.64 - 0.97) | 0.03 |
| OEHR | 13,541 | 38,563 | 3.5 | 16.1 | 7 | 60 | 0.83 (0.32 - 1.88) | 0.69 | 0.78 (0.32 - 1.90) | 0.59 |
| Meta-analysis | 273,037 | 556,464 | 147.7 | 372.8 | 414 | 2,347 | 0.76 (0.67 - 0.86) | <0.01 | 0.77 (0.67 - 0.90) | <0.01 |
| Meta-analysis (w/o USOC) | 46,248 | 119,664 | 20.2 | 68.6 | 66 | 447 | 0.72 (0.53 - 0.97) | 0.03 | 0.73 (0.54 - 0.99) | 0.05 |

##### 1.4.9 Sudden cardiac death

**Supplementary Table 87:** Exposures, outcomes and risk estimates for Sudden cardiac death in GLP1RA (female) (T) vs DPP4I (female) (C) new-user cohorts under four different propensity score (PS) adjustment / time-at-risk (TAR) designs across data sources. We report total patient exposure time in 1,000 patient-year (KYr) units, uncalibrated and calibrated hazard ratios (HRs) and their 95% confidence intervals (CIs). Greyed out entries did not pass study diagnostics and are included only for completeness.

| | Patients | | Exposure (KYr) | | Outcomes | | Uncalibrated | | Calibrated | |
|---|---|---|---|---|---|---|---|---|---|---|
| | T | C | T | C | T | C | HR (95% CI) | $p$ | HR (95% CI) | $p$ |
| **PS-matching / on-treatment TAR** | | | | | | | | | | |
| CCAE | 14,422 | 45,475 | 9.1 | 36.0 | 6 | 16 | 2.22 (0.50 - 9.69) | 0.29 | 2.41 (0.54 - 10.6) | 0.25 |
| ODOD | 7,916 | 28,775 | 4.6 | 22.8 | 7 | 61 | 1.05 (0.39 - 2.50) | 0.92 | 1.02 (0.40 - 2.58) | 0.95 |
| MDCR | 983 | 8,427 | 0.6 | 8.0 | 0 | 11 | | | | |
| MDCD | 1,860 | 7,678 | 1.0 | 5.5 | 0 | 18 | | | | |
| Open Claims | 144,684 | 448,094 | 105.4 | 393.7 | 54 | 470 | 0.53 (0.36 - 0.78) | <0.01 | 0.54 (0.36 - 0.81) | <0.01 |
| OEHR | 10,558 | 38,677 | 3.3 | 20.1 | <5 | 19 | 0.70 (0.03 - 4.71) | 0.78 | 0.67 (0.06 - 7.87) | 0.75 |
| Meta-analysis | 152,600 | 476,869 | 110.1 | 416.5 | 61 | 531 | 0.65 (0.36 - 1.19) | 0.16 | 0.66 (0.36 - 1.21) | 0.18 |
| Meta-analysis (w/o USOC) | 7,916 | 28,775 | 4.6 | 22.8 | 7 | 61 | 1.05 (0.39 - 2.50) | 0.92 | 1.06 (0.42 - 2.69) | 0.90 |
| **PS-stratification / on-treatment TAR** | | | | | | | | | | |
| CCAE | 19,424 | 45,480 | 11.5 | 36.0 | 6 | 16 | 1.63 (0.55 - 4.35) | 0.36 | 1.70 (0.60 - 4.79) | 0.32 |
| ODOD | 11,656 | 29,466 | 6.2 | 23.5 | 8 | 66 | 0.80 (0.32 - 1.77) | 0.60 | 0.80 (0.34 - 1.87) | 0.60 |
| MDCR | 1,124 | 8,792 | 0.7 | 8.3 | 0 | 14 | | | | |
| MDCD | 2,033 | 7,756 | 1.1 | 5.5 | 0 | 20 | | | | |
| Open Claims | 230,968 | 449,183 | 143.9 | 394.7 | 66 | 479 | 0.58 (0.43 - 0.77) | <0.01 | 0.60 (0.43 - 0.84) | <0.01 |
| OEHR | 13,631 | 38,881 | 4.0 | 20.2 | <5 | 20 | 0.69 (0.10 - 2.70) | 0.66 | 0.64 (0.13 - 3.32) | 0.60 |
| Meta-analysis | 275,679 | 563,010 | 165.6 | 474.4 | <85 | 581 | 0.74 (0.46 - 1.17) | 0.20 | 0.76 (0.47 - 1.22) | 0.25 |
| Meta-analysis (w/o USOC) | 44,711 | 113,827 | 21.7 | 79.7 | <19 | 102 | 1.00 (0.54 - 1.85) | 1.00 | 1.02 (0.55 - 1.88) | 0.95 |
| **PS-matching / on-treatment TAR without escalation** | | | | | | | | | | |
| CCAE | 14,422 | 45,475 | 8.2 | 29.6 | 5 | 11 | 1.64 (0.30 - 7.88) | 0.55 | 1.76 (0.35 - 8.95) | 0.50 |
| ODOD | 7,916 | 28,775 | 4.1 | 19.1 | 7 | 51 | 1.13 (0.39 - 2.91) | 0.82 | 1.08 (0.39 - 2.98) | 0.88 |
| MDCR | 983 | 8,427 | 0.6 | 7.0 | 0 | 7 | | | | |
| MDCD | 1,860 | 7,678 | 0.9 | 4.3 | 0 | 11 | | | | |
| Open Claims | 144,684 | 448,094 | 93.4 | 312.5 | 47 | 381 | 0.52 (0.33 - 0.79) | <0.01 | 0.52 (0.34 - 0.82) | <0.01 |
| OEHR | 10,558 | 38,677 | 2.8 | 16.2 | <5 | 10 | 0.66 (0.03 - 4.56) | 0.74 | 0.65 (0.05 - 7.69) | 0.73 |
| Meta-analysis | 152,600 | 476,869 | 97.5 | 331.6 | 54 | 432 | 0.67 (0.33 - 1.35) | 0.26 | 0.67 (0.33 - 1.38) | 0.28 |
| Meta-analysis (w/o USOC) | 7,916 | 28,775 | 4.1 | 19.1 | 7 | 51 | 1.13 (0.39 - 2.91) | 0.82 | 1.15 (0.42 - 3.17) | 0.78 |
| **PS-stratification / on-treatment TAR without escalation** | | | | | | | | | | |
| CCAE | 19,424 | 45,480 | 10.4 | 29.6 | 5 | 11 | 2.09 (0.59 - 6.59) | 0.23 | 2.17 (0.65 - 7.24) | 0.21 |
| ODOD | 11,656 | 29,466 | 5.6 | 19.7 | 8 | 53 | 0.81 (0.32 - 1.85) | 0.64 | 0.81 (0.34 - 1.94) | 0.64 |
| MDCR | 1,124 | 8,792 | 0.6 | 7.2 | 0 | 10 | | | | |
| MDCD | 2,033 | 7,756 | 0.9 | 4.3 | 0 | 12 | | | | |
| Open Claims | 230,968 | 449,183 | 129.8 | 313.4 | 57 | 389 | 0.55 (0.39 - 0.75) | <0.01 | 0.56 (0.39 - 0.81) | <0.01 |
| OEHR | 13,631 | 38,881 | 3.5 | 16.3 | <5 | 11 | 0.57 (0.03 - 3.36) | 0.65 | 0.54 (0.05 - 5.70) | 0.61 |
| Meta-analysis | 242,624 | 478,649 | 135.4 | 333.1 | 65 | 442 | 0.57 (0.42 - 0.78) | <0.01 | 0.59 (0.43 - 0.81) | <0.01 |
| Meta-analysis (w/o USOC) | 11,656 | 29,466 | 5.6 | 19.7 | 8 | 53 | 0.81 (0.32 - 1.85) | 0.64 | 0.83 (0.35 - 1.99) | 0.68 |

##### 1.4.10 Abnormal weight gain

**Supplementary Table 88:** Exposures, outcomes and risk estimates for Abnormal weight gain in GLP1RA (female) (T) vs DPP4I (female) (C) new-user cohorts under four different propensity score (PS) adjustment / time-at-risk (TAR) designs across data sources. We report total patient exposure time in 1,000 patient-year (KYr) units, uncalibrated and calibrated hazard ratios (HRs) and their 95% confidence intervals (CIs). Greyed out entries did not pass study diagnostics and are included only for completeness.

| | Patients | | Exposure (KYr) | | Outcomes | | Uncalibrated | | Calibrated | |
|---|---|---|---|---|---|---|---|---|---|---|
| | T | C | T | C | T | C | HR (95% CI) | $p$ | HR (95% CI) | $p$ |
| **PS-matching / on-treatment TAR** | | | | | | | | | | |
| CCAE | 12,839 | 42,506 | 8.0 | 33.6 | 85 | 255 | 1.47 (1.03 - 2.07) | 0.03 | 1.59 (1.12 - 2.26) | 0.01 |
| ODOD | 7,201 | 27,139 | 4.2 | 21.4 | 44 | 172 | 1.39 (0.84 - 2.26) | 0.20 | 1.35 (0.82 - 2.22) | 0.24 |
| MDCR | 950 | 8,313 | 0.6 | 7.9 | <5 | 26 | 1.80 (0.23 - 9.85) | 0.54 | 1.48 (0.22 - 9.79) | 0.68 |
| MDCD | 1,607 | 6,993 | 0.9 | 4.9 | 11 | 49 | 0.57 (0.18 - 1.48) | 0.29 | 0.58 (0.21 - 1.66) | 0.31 |
| Open Claims | 132,024 | 424,771 | 95.7 | 373.4 | 375 | 1,127 | 1.17 (0.98 - 1.39) | 0.07 | 1.18 (0.97 - 1.45) | 0.13 |
| OEHR | 9,609 | 36,545 | 2.9 | 18.8 | 36 | 166 | 1.31 (0.79 - 2.15) | 0.29 | 1.26 (0.76 - 2.09) | 0.38 |
| Meta-analysis | 163,280 | 537,954 | 111.7 | 452.2 | 551 | 1,769 | 1.22 (1.06 - 1.41) | <0.01 | 1.23 (1.06 - 1.44) | <0.01 |
| Meta-analysis (w/o USOC) | 31,256 | 113,183 | 16.0 | 78.8 | 176 | 642 | 1.34 (1.06 - 1.71) | 0.02 | 1.36 (1.07 - 1.74) | 0.01 |
| **PS-stratification / on-treatment TAR** | | | | | | | | | | |
| CCAE | 16,752 | 42,511 | 9.9 | 33.6 | 110 | 255 | 1.24 (0.95 - 1.61) | 0.11 | 1.29 (0.99 - 1.68) | 0.07 |
| ODOD | 10,178 | 27,829 | 5.4 | 22.1 | 61 | 179 | 1.13 (0.79 - 1.60) | 0.51 | 1.12 (0.79 - 1.60) | 0.54 |
| MDCR | 1,069 | 8,672 | 0.6 | 8.2 | <5 | 26 | 2.44 (0.62 - 7.51) | 0.16 | 2.03 (0.58 - 7.08) | 0.27 |
| MDCD | 1,722 | 7,067 | 0.9 | 5.0 | 13 | 50 | 1.09 (0.55 - 2.06) | 0.79 | 1.11 (0.57 - 2.15) | 0.77 |
| Open Claims | 202,244 | 425,856 | 126.4 | 374.5 | 549 | 1,130 | 1.22 (1.07 - 1.38) | <0.01 | 1.27 (1.05 - 1.53) | 0.02 |
| OEHR | 12,038 | 36,753 | 3.5 | 18.9 | 53 | 168 | 1.23 (0.86 - 1.73) | 0.25 | 1.15 (0.81 - 1.63) | 0.46 |
| Meta-analysis | 242,934 | 540,016 | 146.1 | 454.1 | 786 | 1,782 | 1.21 (1.09 - 1.34) | <0.01 | 1.24 (1.07 - 1.44) | <0.01 |
| Meta-analysis (w/o USOC) | 40,690 | 114,160 | 19.7 | 79.6 | 237 | 652 | 1.20 (1.01 - 1.43) | 0.04 | 1.22 (1.02 - 1.46) | 0.04 |
| **PS-matching / on-treatment TAR without escalation** | | | | | | | | | | |
| CCAE | 12,839 | 42,506 | 7.2 | 27.7 | 78 | 202 | 1.37 (0.94 - 1.97) | 0.09 | 1.46 (1.01 - 2.12) | 0.05 |
| ODOD | 7,201 | 27,139 | 3.8 | 18.0 | 40 | 139 | 1.46 (0.85 - 2.48) | 0.16 | 1.41 (0.82 - 2.41) | 0.22 |
| MDCR | 950 | 8,313 | 0.6 | 6.9 | <5 | 20 | 2.82 (0.33 - 16.0) | 0.29 | 2.26 (0.33 - 15.7) | 0.41 |
| MDCD | 1,607 | 6,993 | 0.8 | 3.8 | 9 | 35 | 0.65 (0.18 - 1.92) | 0.48 | 0.68 (0.21 - 2.24) | 0.53 |
| Open Claims | 132,024 | 424,771 | 84.7 | 296.9 | 331 | 857 | 1.09 (0.90 - 1.32) | 0.38 | 1.10 (0.91 - 1.33) | 0.38 |
| OEHR | 9,609 | 36,545 | 2.5 | 15.2 | 30 | 113 | 1.69 (0.96 - 2.95) | 0.07 | 1.67 (0.95 - 2.93) | 0.08 |
| Meta-analysis | 163,280 | 537,954 | 99.0 | 361.6 | 488 | 1,346 | 1.23 (1.01 - 1.49) | 0.04 | 1.24 (1.01 - 1.53) | 0.04 |
| Meta-analysis (w/o USOC) | 31,256 | 113,183 | 14.3 | 64.7 | 157 | 489 | 1.40 (1.08 - 1.82) | 0.01 | 1.44 (1.11 - 1.87) | <0.01 |
| **PS-stratification / on-treatment TAR without escalation** | | | | | | | | | | |
| CCAE | 16,752 | 42,511 | 8.9 | 27.7 | 102 | 202 | 1.27 (0.96 - 1.68) | 0.09 | 1.32 (1.00 - 1.76) | 0.06 |
| ODOD | 10,178 | 27,829 | 4.9 | 18.6 | 56 | 145 | 1.14 (0.77 - 1.65) | 0.51 | 1.13 (0.78 - 1.66) | 0.53 |
| MDCR | 1,069 | 8,672 | 0.6 | 7.1 | <5 | 20 | 2.86 (0.69 - 9.40) | 0.11 | 2.33 (0.63 - 8.59) | 0.21 |
| MDCD | 1,722 | 7,067 | 0.8 | 3.9 | 11 | 36 | 1.10 (0.51 - 2.23) | 0.80 | 1.15 (0.55 - 2.40) | 0.72 |
| Open Claims | 202,244 | 425,856 | 113.9 | 297.8 | 499 | 860 | 1.26 (1.10 - 1.45) | <0.01 | 1.30 (1.06 - 1.60) | 0.02 |
| OEHR | 12,038 | 36,753 | 3.1 | 15.3 | 46 | 115 | 1.44 (0.96 - 2.12) | 0.07 | 1.35 (0.91 - 2.01) | 0.17 |
| Meta-analysis | 242,934 | 540,016 | 131.6 | 363.2 | 714 | 1,358 | 1.26 (1.13 - 1.41) | <0.01 | 1.29 (1.12 - 1.48) | <0.01 |
| Meta-analysis (w/o USOC) | 40,690 | 114,160 | 17.7 | 65.4 | 215 | 498 | 1.26 (1.04 - 1.52) | 0.02 | 1.29 (1.06 - 1.56) | 0.01 |

##### 1.4.11 Abnormal weight loss

**Supplementary Table 89:** Exposures, outcomes and risk estimates for Abnormal weight loss in GLP1RA (female) (T) vs DPP4I (female) (C) new-user cohorts under four different propensity score (PS) adjustment / time-at-risk (TAR) designs across data sources. We report total patient exposure time in 1,000 patient-year (KYr) units, uncalibrated and calibrated hazard ratios (HRs) and their 95% confidence intervals (CIs). Greyed out entries did not pass study diagnostics and are included only for completeness.

| | Patients | | Exposure (KYr) | | Outcomes | | Uncalibrated | | Calibrated | |
|---|---|---|---|---|---|---|---|---|---|---|
| | T | C | T | C | T | C | HR (95% CI) | $p$ | HR (95% CI) | $p$ |
| **PS-matching / on-treatment TAR** | | | | | | | | | | |
| CCAE | 14,105 | 44,496 | 8.9 | 35.0 | 81 | 243 | 1.34 (0.91 - 1.94) | 0.13 | 1.45 (0.99 - 2.12) | 0.07 |
| ODOD | 7,697 | 27,623 | 4.5 | 21.7 | 73 | 313 | 1.49 (1.03 - 2.14) | 0.03 | 1.45 (1.00 - 2.10) | 0.05 |
| MDCR | 964 | 8,114 | 0.6 | 7.7 | 11 | 109 | 1.49 (0.66 - 3.10) | 0.31 | 1.23 (0.57 - 2.66) | 0.61 |
| MDCD | 1,790 | 7,318 | 1.0 | 5.2 | 20 | 84 | 0.95 (0.46 - 1.86) | 0.89 | 0.97 (0.49 - 1.95) | 0.89 |
| Open Claims | 139,999 | 430,739 | 101.8 | 376.4 | 539 | 2,632 | 1.02 (0.89 - 1.17) | 0.74 | 1.03 (0.87 - 1.22) | 0.73 |
| OEHR | 10,300 | 37,586 | 3.2 | 19.3 | 27 | 220 | 0.90 (0.51 - 1.50) | 0.69 | 0.86 (0.50 - 1.48) | 0.60 |
| Meta-analysis | 173,891 | 547,762 | 119.3 | 457.6 | 740 | 3,492 | 1.13 (0.94 - 1.36) | 0.21 | 1.14 (0.93 - 1.39) | 0.21 |
| Meta-analysis (w/o USOC) | 33,892 | 117,023 | 17.5 | 81.3 | 201 | 860 | 1.25 (1.00 - 1.57) | 0.05 | 1.27 (1.01 - 1.60) | 0.04 |
| **PS-stratification / on-treatment TAR** | | | | | | | | | | |
| CCAE | 18,985 | 44,501 | 11.2 | 35.0 | 106 | 243 | 1.45 (1.10 - 1.89) | <0.01 | 1.51 (1.15 - 1.98) | <0.01 |
| ODOD | 11,296 | 28,127 | 6.0 | 22.2 | 88 | 328 | 1.20 (0.90 - 1.59) | 0.21 | 1.20 (0.90 - 1.59) | 0.25 |
| MDCR | 1,107 | 8,411 | 0.7 | 7.9 | 12 | 120 | 1.22 (0.60 - 2.32) | 0.56 | 1.02 (0.52 - 2.01) | 0.90 |
| MDCD | 1,955 | 7,383 | 1.0 | 5.2 | 20 | 88 | 1.26 (0.72 - 2.13) | 0.40 | 1.28 (0.74 - 2.20) | 0.38 |
| Open Claims | 223,395 | 431,515 | 138.9 | 377.1 | 739 | 2,654 | 1.03 (0.94 - 1.14) | 0.51 | 1.08 (0.90 - 1.28) | 0.43 |
| OEHR | 13,291 | 37,764 | 3.9 | 19.4 | 34 | 223 | 0.87 (0.57 - 1.29) | 0.50 | 0.82 (0.54 - 1.22) | 0.34 |
| Meta-analysis | 268,922 | 549,290 | 161.0 | 459.0 | 987 | 3,536 | 1.13 (0.96 - 1.33) | 0.13 | 1.16 (0.96 - 1.41) | 0.13 |
| Meta-analysis (w/o USOC) | 45,527 | 117,775 | 22.1 | 81.9 | 248 | 882 | 1.21 (0.98 - 1.49) | 0.07 | 1.23 (1.00 - 1.52) | 0.06 |
| **PS-matching / on-treatment TAR without escalation** | | | | | | | | | | |
| CCAE | 14,105 | 44,496 | 8.0 | 28.8 | 69 | 208 | 1.29 (0.85 - 1.93) | 0.22 | 1.38 (0.92 - 2.08) | 0.14 |
| ODOD | 7,697 | 27,623 | 4.0 | 18.2 | 65 | 263 | 1.53 (1.02 - 2.26) | 0.04 | 1.47 (0.98 - 2.18) | 0.07 |
| MDCR | 964 | 8,114 | 0.6 | 6.7 | 10 | 98 | 1.71 (0.75 - 3.55) | 0.18 | 1.36 (0.63 - 2.96) | 0.45 |
| MDCD | 1,790 | 7,318 | 0.8 | 4.0 | 13 | 59 | 0.88 (0.37 - 1.94) | 0.76 | 0.92 (0.40 - 2.11) | 0.84 |
| Open Claims | 139,999 | 430,739 | 90.2 | 298.7 | 475 | 2,139 | 1.07 (0.93 - 1.24) | 0.35 | 1.08 (0.93 - 1.26) | 0.36 |
| OEHR | 10,300 | 37,586 | 2.7 | 15.6 | 21 | 172 | 0.97 (0.52 - 1.72) | 0.92 | 0.96 (0.53 - 1.73) | 0.87 |
| Meta-analysis | 173,891 | 547,762 | 105.8 | 365.5 | 643 | 2,841 | 1.14 (0.97 - 1.33) | 0.10 | 1.15 (0.97 - 1.37) | 0.11 |
| Meta-analysis (w/o USOC) | 33,892 | 117,023 | 15.5 | 66.7 | 168 | 702 | 1.27 (0.99 - 1.62) | 0.06 | 1.30 (1.01 - 1.66) | 0.05 |
| **PS-stratification / on-treatment TAR without escalation** | | | | | | | | | | |
| CCAE | 18,985 | 44,501 | 10.1 | 28.8 | 88 | 208 | 1.31 (0.97 - 1.76) | 0.08 | 1.36 (1.01 - 1.84) | 0.05 |
| ODOD | 11,296 | 28,127 | 5.4 | 18.6 | 79 | 275 | 1.23 (0.90 - 1.67) | 0.19 | 1.23 (0.90 - 1.67) | 0.22 |
| MDCR | 1,107 | 8,411 | 0.6 | 6.8 | 11 | 109 | 1.33 (0.63 - 2.57) | 0.42 | 1.09 (0.54 - 2.19) | 0.82 |
| MDCD | 1,955 | 7,383 | 0.9 | 4.1 | 13 | 63 | 1.00 (0.50 - 1.88) | 1.00 | 1.04 (0.53 - 2.02) | 0.89 |
| Open Claims | 223,395 | 431,515 | 125.4 | 299.3 | 666 | 2,158 | 1.03 (0.92 - 1.15) | 0.60 | 1.06 (0.88 - 1.27) | 0.57 |
| OEHR | 13,291 | 37,764 | 3.4 | 15.7 | 28 | 175 | 0.88 (0.55 - 1.36) | 0.57 | 0.82 (0.52 - 1.29) | 0.42 |
| Meta-analysis | 268,922 | 549,290 | 145.2 | 366.6 | 874 | 2,879 | 1.08 (0.96 - 1.22) | 0.22 | 1.10 (0.95 - 1.28) | 0.19 |
| Meta-analysis (w/o USOC) | 45,527 | 117,775 | 19.8 | 67.2 | 208 | 721 | 1.17 (0.97 - 1.41) | 0.10 | 1.20 (0.99 - 1.45) | 0.07 |

##### 1.4.12 Acute pancreatitis

**Supplementary Table 90:** Exposures, outcomes and risk estimates for Acute pancreatitis in GLP1RA (female) (T) vs DPP4I (female) (C) new-user cohorts under four different propensity score (PS) adjustment / time-at-risk (TAR) designs across data sources. We report total patient exposure time in 1,000 patient-year (KYr) units, uncalibrated and calibrated hazard ratios (HRs) and their 95% confidence intervals (CIs). Greyed out entries did not pass study diagnostics and are included only for completeness.

| | Patients | | Exposure (KYr) | | Outcomes | | Uncalibrated | | Calibrated | |
|---|---|---|---|---|---|---|---|---|---|---|
| | T | C | T | C | T | C | HR (95% CI) | $p$ | HR (95% CI) | $p$ |
| **PS-matching / on-treatment TAR** | | | | | | | | | | |
| CCAE | 14,343 | 45,207 | 9.1 | 35.7 | 15 | 76 | 0.98 (0.40 - 2.23) | 0.97 | 1.07 (0.45 - 2.54) | 0.89 |
| ODOD | 7,876 | 28,636 | 4.6 | 22.7 | 13 | 62 | 1.21 (0.50 - 2.71) | 0.66 | 1.18 (0.51 - 2.74) | 0.70 |
| MDCR | 982 | 8,389 | 0.6 | 8.0 | <5 | 16 | 3.77 (0.64 - 19.2) | 0.13 | 3.11 (0.57 - 17.0) | 0.19 |
| MDCD | 1,840 | 7,561 | 1.0 | 5.4 | 5 | 25 | 0.99 (0.25 - 3.30) | 0.99 | 1.02 (0.28 - 3.66) | 0.94 |
| Open Claims | 143,294 | 443,745 | 104.5 | 389.7 | 169 | 838 | 0.95 (0.76 - 1.19) | 0.68 | 0.96 (0.75 - 1.24) | 0.78 |
| OEHR | 10,522 | 38,526 | 3.2 | 20.0 | 9 | 39 | 1.03 (0.32 - 2.87) | 0.96 | 0.99 (0.33 - 2.98) | 0.95 |
| Meta-analysis | 177,875 | 563,675 | 122.5 | 473.6 | 211 | 1,040 | 0.97 (0.79 - 1.20) | 0.79 | 0.98 (0.79 - 1.22) | 0.86 |
| Meta-analysis (w/o USOC) | 34,581 | 119,930 | 17.9 | 83.8 | 42 | 202 | 1.07 (0.65 - 1.74) | 0.80 | 1.08 (0.66 - 1.77) | 0.75 |
| **PS-stratification / on-treatment TAR** | | | | | | | | | | |
| CCAE | 19,337 | 45,212 | 11.4 | 35.7 | 16 | 77 | 0.80 (0.43 - 1.42) | 0.46 | 0.83 (0.46 - 1.52) | 0.56 |
| ODOD | 11,600 | 29,302 | 6.2 | 23.3 | 16 | 67 | 1.24 (0.63 - 2.32) | 0.52 | 1.23 (0.64 - 2.37) | 0.53 |
| MDCR | 1,124 | 8,746 | 0.7 | 8.3 | <5 | 18 | 3.05 (0.72 - 10.6) | 0.10 | 2.53 (0.66 - 9.71) | 0.18 |
| MDCD | 2,010 | 7,636 | 1.1 | 5.5 | 5 | 25 | 0.96 (0.30 - 2.58) | 0.94 | 0.97 (0.33 - 2.86) | 0.94 |
| Open Claims | 228,988 | 444,912 | 142.8 | 390.9 | 217 | 842 | 1.01 (0.84 - 1.21) | 0.90 | 1.05 (0.84 - 1.32) | 0.68 |
| OEHR | 13,579 | 38,728 | 4.0 | 20.1 | 10 | 39 | 1.47 (0.66 - 3.02) | 0.32 | 1.38 (0.64 - 2.95) | 0.41 |
| Meta-analysis | 275,514 | 565,790 | 165.5 | 475.5 | 264 | 1,050 | 1.02 (0.87 - 1.20) | 0.79 | 1.05 (0.87 - 1.27) | 0.62 |
| Meta-analysis (w/o USOC) | 46,526 | 120,878 | 22.7 | 84.6 | 47 | 208 | 1.07 (0.75 - 1.53) | 0.72 | 1.09 (0.76 - 1.56) | 0.65 |
| **PS-matching / on-treatment TAR without escalation** | | | | | | | | | | |
| CCAE | 14,343 | 45,207 | 8.1 | 29.4 | 12 | 59 | 1.03 (0.39 - 2.46) | 0.95 | 1.10 (0.44 - 2.78) | 0.84 |
| ODOD | 7,876 | 28,636 | 4.1 | 19.0 | 11 | 52 | 1.78 (0.66 - 4.42) | 0.23 | 1.71 (0.66 - 4.41) | 0.27 |
| MDCR | 982 | 8,389 | 0.6 | 6.9 | <5 | 12 | 4.41 (0.70 - 27.1) | 0.11 | 3.53 (0.57 - 22.0) | 0.18 |
| MDCD | 1,840 | 7,561 | 0.9 | 4.2 | <5 | 16 | 0.29 (0.02 - 1.76) | 0.31 | 0.30 (0.03 - 3.29) | 0.33 |
| Open Claims | 143,294 | 443,745 | 92.6 | 309.4 | 149 | 649 | 0.97 (0.75 - 1.24) | 0.82 | 0.98 (0.76 - 1.26) | 0.89 |
| OEHR | 10,522 | 38,526 | 2.8 | 16.1 | 8 | 29 | 1.21 (0.37 - 3.46) | 0.74 | 1.19 (0.39 - 3.67) | 0.76 |
| Meta-analysis | 176,035 | 556,114 | 107.6 | 373.9 | 180 | 789 | 1.02 (0.81 - 1.28) | 0.87 | 1.03 (0.81 - 1.32) | 0.80 |
| Meta-analysis (w/o USOC) | 32,741 | 112,369 | 15.1 | 64.5 | 31 | 140 | 1.31 (0.74 - 2.31) | 0.36 | 1.34 (0.76 - 2.37) | 0.32 |
| **PS-stratification / on-treatment TAR without escalation** | | | | | | | | | | |
| CCAE | 19,337 | 45,212 | 10.4 | 29.4 | 13 | 60 | 0.78 (0.39 - 1.49) | 0.48 | 0.82 (0.41 - 1.60) | 0.56 |
| ODOD | 11,600 | 29,302 | 5.6 | 19.6 | 14 | 56 | 1.41 (0.68 - 2.75) | 0.33 | 1.41 (0.70 - 2.83) | 0.34 |
| MDCR | 1,124 | 8,746 | 0.6 | 7.2 | <5 | 14 | 3.55 (0.81 - 13.1) | 0.07 | 2.89 (0.72 - 11.7) | 0.14 |
| MDCD | 2,010 | 7,636 | 0.9 | 4.2 | <5 | 16 | 0.42 (0.06 - 1.74) | 0.31 | 0.44 (0.08 - 2.34) | 0.33 |
| Open Claims | 228,988 | 444,912 | 128.8 | 310.4 | 193 | 653 | 1.07 (0.88 - 1.29) | 0.51 | 1.10 (0.86 - 1.40) | 0.47 |
| OEHR | 13,579 | 38,728 | 3.5 | 16.2 | 9 | 29 | 1.56 (0.65 - 3.41) | 0.29 | 1.46 (0.64 - 3.33) | 0.38 |
| Meta-analysis | 273,504 | 558,154 | 148.2 | 375.5 | 229 | 798 | 1.08 (0.91 - 1.29) | 0.38 | 1.10 (0.91 - 1.34) | 0.32 |
| Meta-analysis (w/o USOC) | 44,516 | 113,242 | 19.4 | 65.1 | 36 | 145 | 1.16 (0.75 - 1.77) | 0.50 | 1.18 (0.77 - 1.81) | 0.45 |

##### 1.4.13 Bone fracture

**Supplementary Table 91:** Exposures, outcomes and risk estimates for Bone fracture in GLP1RA (female) (T) vs DPP4I (female) (C) new-user cohorts under four different propensity score (PS) adjustment / time-at-risk (TAR) designs across data sources. We report total patient exposure time in 1,000 patient-year (KYr) units, uncalibrated and calibrated hazard ratios (HRs) and their 95% confidence intervals (CIs). Greyed out entries did not pass study diagnostics and are included only for completeness.

| | Patients | | Exposure (KYr) | | Outcomes | | Uncalibrated | | Calibrated | |
|---|---|---|---|---|---|---|---|---|---|---|
| | T | C | T | C | T | C | HR (95% CI) | $p$ | HR (95% CI) | $p$ |
| **PS-matching / on-treatment TAR** | | | | | | | | | | |
| CCAE | 12,951 | 41,233 | 8.0 | 31.8 | 163 | 694 | 0.98 (0.76 - 1.24) | 0.84 | 1.06 (0.82 - 1.36) | 0.70 |
| ODOD | 7,110 | 25,541 | 4.1 | 19.7 | 123 | 672 | 1.07 (0.81 - 1.41) | 0.62 | 1.05 (0.79 - 1.39) | 0.76 |
| MDCR | 835 | 7,107 | 0.5 | 6.6 | 18 | 301 | 0.86 (0.47 - 1.48) | 0.60 | 0.70 (0.40 - 1.25) | 0.25 |
| MDCD | 1,600 | 6,648 | 0.9 | 4.6 | 32 | 182 | 0.59 (0.34 - 0.98) | 0.05 | 0.61 (0.36 - 1.02) | 0.07 |
| Open Claims | 122,716 | 384,052 | 88.4 | 330.6 | 1,321 | 7,197 | 0.92 (0.84 - 1.00) | 0.04 | 0.93 (0.81 - 1.06) | 0.34 |
| OEHR | 9,754 | 35,795 | 3.0 | 18.3 | 36 | 409 | 0.76 (0.49 - 1.15) | 0.21 | 0.73 (0.48 - 1.12) | 0.17 |
| Meta-analysis | 154,131 | 493,269 | 104.3 | 405.0 | 1,675 | 9,154 | 0.92 (0.85 - 0.99) | 0.03 | 0.93 (0.84 - 1.03) | 0.14 |
| Meta-analysis (w/o USOC) | 31,415 | 109,217 | 15.9 | 74.4 | 354 | 1,957 | 0.90 (0.72 - 1.11) | 0.33 | 0.91 (0.73 - 1.14) | 0.42 |
| **PS-stratification / on-treatment TAR** | | | | | | | | | | |
| CCAE | 17,366 | 41,238 | 10.1 | 31.8 | 203 | 694 | 0.95 (0.79 - 1.13) | 0.56 | 0.99 (0.82 - 1.18) | 0.87 |
| ODOD | 10,452 | 26,228 | 5.4 | 20.3 | 161 | 709 | 1.02 (0.83 - 1.25) | 0.86 | 1.01 (0.82 - 1.25) | 0.85 |
| MDCR | 957 | 7,400 | 0.6 | 6.8 | 18 | 311 | 0.78 (0.45 - 1.27) | 0.35 | 0.65 (0.39 - 1.09) | 0.11 |
| MDCD | 1,744 | 6,705 | 0.9 | 4.7 | 33 | 185 | 0.85 (0.56 - 1.26) | 0.44 | 0.86 (0.58 - 1.29) | 0.49 |
| Open Claims | 194,843 | 385,020 | 120.3 | 331.6 | 1,758 | 7,243 | 0.88 (0.83 - 0.94) | <0.01 | 0.92 (0.78 - 1.07) | 0.29 |
| OEHR | 12,509 | 35,951 | 3.7 | 18.4 | 48 | 416 | 0.69 (0.49 - 0.94) | 0.02 | 0.64 (0.46 - 0.89) | 0.01 |
| Meta-analysis | 236,914 | 495,142 | 140.4 | 406.7 | 2,203 | 9,247 | 0.89 (0.84 - 0.94) | <0.01 | 0.91 (0.81 - 1.03) | 0.14 |
| Meta-analysis (w/o USOC) | 42,071 | 110,122 | 20.1 | 75.1 | 445 | 2,004 | 0.91 (0.78 - 1.06) | 0.20 | 0.92 (0.79 - 1.08) | 0.34 |
| **PS-matching / on-treatment TAR without escalation** | | | | | | | | | | |
| CCAE | 12,951 | 41,233 | 7.2 | 26.3 | 145 | 558 | 0.97 (0.74 - 1.26) | 0.83 | 1.04 (0.80 - 1.36) | 0.79 |
| ODOD | 7,110 | 25,541 | 3.7 | 16.6 | 106 | 551 | 1.08 (0.80 - 1.46) | 0.60 | 1.04 (0.77 - 1.41) | 0.80 |
| MDCR | 835 | 7,107 | 0.5 | 5.8 | 17 | 250 | 0.92 (0.49 - 1.62) | 0.78 | 0.73 (0.40 - 1.33) | 0.32 |
| MDCD | 1,600 | 6,648 | 0.7 | 3.6 | 29 | 136 | 0.77 (0.44 - 1.30) | 0.35 | 0.81 (0.47 - 1.39) | 0.46 |
| Open Claims | 122,716 | 384,052 | 78.4 | 263.2 | 1,134 | 5,706 | 0.91 (0.83 - 1.00) | 0.05 | 0.92 (0.83 - 1.01) | 0.18 |
| OEHR | 9,754 | 35,795 | 2.6 | 14.8 | 31 | 306 | 0.81 (0.49 - 1.29) | 0.39 | 0.80 (0.49 - 1.29) | 0.38 |
| Meta-analysis | 154,131 | 493,269 | 92.5 | 324.4 | 1,445 | 7,257 | 0.92 (0.85 - 1.00) | 0.05 | 0.93 (0.83 - 1.04) | 0.22 |
| Meta-analysis (w/o USOC) | 31,415 | 109,217 | 14.2 | 61.3 | 311 | 1,551 | 0.96 (0.81 - 1.14) | 0.65 | 0.98 (0.82 - 1.17) | 0.83 |
| **PS-stratification / on-treatment TAR without escalation** | | | | | | | | | | |
| CCAE | 17,366 | 41,238 | 9.1 | 26.3 | 182 | 558 | 0.97 (0.79 - 1.17) | 0.73 | 1.01 (0.83 - 1.22) | 0.88 |
| ODOD | 10,452 | 26,228 | 5.0 | 17.1 | 140 | 582 | 1.02 (0.81 - 1.27) | 0.87 | 1.02 (0.81 - 1.27) | 0.86 |
| MDCR | 957 | 7,400 | 0.5 | 6.0 | 17 | 260 | 0.84 (0.48 - 1.40) | 0.53 | 0.69 (0.40 - 1.18) | 0.18 |
| MDCD | 1,744 | 6,705 | 0.8 | 3.6 | 30 | 138 | 0.98 (0.62 - 1.48) | 0.91 | 1.02 (0.66 - 1.57) | 0.86 |
| Open Claims | 194,843 | 385,020 | 108.6 | 263.9 | 1,548 | 5,747 | 0.87 (0.82 - 0.93) | <0.01 | 0.90 (0.76 - 1.06) | 0.23 |
| OEHR | 12,509 | 35,951 | 3.2 | 14.8 | 43 | 310 | 0.77 (0.54 - 1.09) | 0.15 | 0.72 (0.51 - 1.03) | 0.09 |
| Meta-analysis | 236,914 | 495,142 | 126.7 | 325.8 | 1,943 | 7,335 | 0.90 (0.83 - 0.96) | <0.01 | 0.91 (0.82 - 1.02) | 0.11 |
| Meta-analysis (w/o USOC) | 42,071 | 110,122 | 18.1 | 61.9 | 395 | 1,588 | 0.95 (0.84 - 1.09) | 0.48 | 0.98 (0.86 - 1.11) | 0.74 |

##### 1.4.14 Diabetic ketoacidosis

**Supplementary Table 92:** Exposures, outcomes and risk estimates for Diabetic ketoacidosis in GLP1RA (female) (T) vs DPP4I (female) (C) new-user cohorts under four different propensity score (PS) adjustment / time-at-risk (TAR) designs across data sources. We report total patient exposure time in 1,000 patient-year (KYr) units, uncalibrated and calibrated hazard ratios (HRs) and their 95% confidence intervals (CIs). Greyed out entries did not pass study diagnostics and are included only for completeness.

| | Patients | | Exposure (KYr) | | Outcomes | | Uncalibrated | | Calibrated | |
|---|---|---|---|---|---|---|---|---|---|---|
| | T | C | T | C | T | C | HR (95% CI) | $p$ | HR (95% CI) | $p$ |
| **PS-matching / on-treatment TAR** | | | | | | | | | | |
| CCAE | 14,417 | 45,448 | 9.1 | 36.0 | 6 | 29 | 0.62 (0.17 - 1.89) | 0.44 | 0.67 (0.20 - 2.27) | 0.52 |
| ODOD | 7,919 | 28,782 | 4.6 | 22.8 | 5 | 36 | 0.42 (0.09 - 1.33) | 0.20 | 0.41 (0.11 - 1.53) | 0.19 |
| MDCR | 986 | 8,431 | 0.6 | 8.0 | 0 | 10 | | | | |
| MDCD | 1,854 | 7,669 | 1.0 | 5.5 | <5 | 17 | 0.30 (0.02 - 1.68) | 0.31 | 0.31 (0.03 - 3.15) | 0.32 |
| Open Claims | 144,634 | 448,054 | 105.4 | 393.6 | 59 | 362 | 0.60 (0.41 - 0.86) | <0.01 | 0.61 (0.42 - 0.89) | 0.01 |
| OEHR | 10,556 | 38,675 | 3.3 | 20.1 | 5 | 24 | 1.24 (0.36 - 3.77) | 0.72 | 1.19 (0.37 - 3.85) | 0.77 |
| Meta-analysis | 177,526 | 560,959 | 122.4 | 472.4 | 75 | 451 | 0.62 (0.45 - 0.86) | <0.01 | 0.63 (0.45 - 0.88) | <0.01 |
| Meta-analysis (w/o USOC) | 32,892 | 112,905 | 17.0 | 78.9 | 16 | 89 | 0.71 (0.35 - 1.45) | 0.35 | 0.73 (0.36 - 1.48) | 0.38 |
| **PS-stratification / on-treatment TAR** | | | | | | | | | | |
| CCAE | 19,428 | 45,453 | 11.5 | 36.0 | 7 | 29 | 0.55 (0.20 - 1.34) | 0.22 | 0.58 (0.22 - 1.48) | 0.25 |
| ODOD | 11,652 | 29,474 | 6.2 | 23.5 | 7 | 37 | 0.46 (0.17 - 1.09) | 0.10 | 0.46 (0.18 - 1.16) | 0.10 |
| MDCR | 1,128 | 8,794 | 0.7 | 8.3 | 0 | 10 | | | | |
| MDCD | 2,026 | 7,748 | 1.1 | 5.5 | <5 | 17 | 0.25 (0.01 - 1.38) | 0.24 | 0.26 (0.03 - 2.59) | 0.25 |
| Open Claims | 230,974 | 449,158 | 143.9 | 394.7 | 78 | 362 | 0.60 (0.45 - 0.79) | <0.01 | 0.62 (0.45 - 0.86) | <0.01 |
| OEHR | 13,629 | 38,879 | 4.0 | 20.2 | 8 | 25 | 1.08 (0.40 - 2.67) | 0.87 | 1.01 (0.39 - 2.63) | 0.96 |
| Meta-analysis | 275,683 | 562,964 | 165.7 | 474.3 | 100 | 453 | 0.61 (0.47 - 0.78) | <0.01 | 0.62 (0.47 - 0.82) | <0.01 |
| Meta-analysis (w/o USOC) | 44,709 | 113,806 | 21.7 | 79.6 | 22 | 91 | 0.65 (0.38 - 1.11) | 0.11 | 0.66 (0.38 - 1.13) | 0.13 |
| **PS-matching / on-treatment TAR without escalation** | | | | | | | | | | |
| CCAE | 14,417 | 45,448 | 8.2 | 29.6 | 5 | 18 | 0.81 (0.21 - 2.72) | 0.74 | 0.86 (0.24 - 3.13) | 0.82 |
| ODOD | 7,919 | 28,782 | 4.1 | 19.1 | <5 | 29 | 0.91 (0.19 - 3.26) | 0.89 | 0.87 (0.21 - 3.59) | 0.85 |
| MDCR | 986 | 8,431 | 0.6 | 7.0 | 0 | 8 | | | | |
| MDCD | 1,854 | 7,669 | 0.9 | 4.3 | <5 | 12 | 0.31 (0.02 - 1.83) | 0.34 | 0.33 (0.03 - 3.46) | 0.36 |
| Open Claims | 144,634 | 448,054 | 93.4 | 312.6 | 48 | 252 | 0.63 (0.41 - 0.95) | 0.03 | 0.64 (0.42 - 0.97) | 0.04 |
| OEHR | 10,556 | 38,675 | 2.8 | 16.2 | <5 | 14 | 0.55 (0.03 - 3.65) | 0.63 | 0.54 (0.05 - 6.24) | 0.62 |
| Meta-analysis | 166,970 | 522,284 | 105.7 | 361.3 | <58 | 299 | 0.66 (0.45 - 0.97) | 0.04 | 0.67 (0.46 - 0.99) | 0.05 |
| Meta-analysis (w/o USOC) | 22,336 | 74,230 | 12.3 | 48.7 | <10 | 47 | 0.85 (0.33 - 2.21) | 0.74 | 0.87 (0.34 - 2.26) | 0.78 |
| **PS-stratification / on-treatment TAR without escalation** | | | | | | | | | | |
| CCAE | 19,428 | 45,453 | 10.4 | 29.6 | 5 | 18 | 0.56 (0.17 - 1.64) | 0.32 | 0.58 (0.19 - 1.83) | 0.36 |
| ODOD | 11,652 | 29,474 | 5.6 | 19.7 | 6 | 30 | 0.42 (0.14 - 1.07) | 0.09 | 0.42 (0.15 - 1.15) | 0.09 |
| MDCR | 1,128 | 8,794 | 0.6 | 7.2 | 0 | 8 | | | | |
| MDCD | 2,026 | 7,748 | 0.9 | 4.3 | <5 | 12 | 0.28 (0.01 - 1.59) | 0.29 | 0.29 (0.03 - 3.02) | 0.30 |
| Open Claims | 230,974 | 449,158 | 129.9 | 313.5 | 67 | 252 | 0.66 (0.47 - 0.90) | 0.01 | 0.68 (0.47 - 0.97) | 0.03 |
| OEHR | 13,629 | 38,879 | 3.5 | 16.3 | <5 | 14 | 0.53 (0.13 - 1.84) | 0.35 | 0.49 (0.13 - 1.87) | 0.30 |
| Meta-analysis | 262,054 | 524,085 | 145.9 | 362.7 | 78 | 300 | 0.63 (0.47 - 0.84) | <0.01 | 0.64 (0.47 - 0.87) | <0.01 |
| Meta-analysis (w/o USOC) | 31,080 | 74,927 | 16.0 | 49.2 | 11 | 48 | 0.48 (0.22 - 1.01) | 0.05 | 0.49 (0.23 - 1.04) | 0.06 |

##### 1.4.15 Diarrhea

**Supplementary Table 93:** Exposures, outcomes and risk estimates for Diarrhea in GLP1RA (female) (T) vs DPP4I (female) (C) new-user cohorts under four different propensity score (PS) adjustment / time-at-risk (TAR) designs across data sources. We report total patient exposure time in 1,000 patient-year (KYr) units, uncalibrated and calibrated hazard ratios (HRs) and their 95% confidence intervals (CIs). Greyed out entries did not pass study diagnostics and are included only for completeness.

| | Patients | | Exposure (KYr) | | Outcomes | | Uncalibrated | | Calibrated | |
|---|---|---|---|---|---|---|---|---|---|---|
| | T | C | T | C | T | C | HR (95% CI) | $p$ | HR (95% CI) | $p$ |
| **PS-matching / on-treatment TAR** | | | | | | | | | | |
| CCAE | 12,135 | 38,768 | 7.4 | 29.7 | 335 | 1,050 | 1.21 (1.02 - 1.44) | 0.03 | 1.31 (1.10 - 1.57) | 0.01 |
| ODOD | 6,507 | 23,550 | 3.7 | 18.0 | 185 | 867 | 1.06 (0.85 - 1.33) | 0.59 | 1.04 (0.83 - 1.30) | 0.76 |
| MDCR | 840 | 7,035 | 0.5 | 6.4 | 24 | 275 | 1.12 (0.64 - 1.87) | 0.67 | 0.92 (0.54 - 1.58) | 0.77 |
| MDCD | 1,404 | 5,749 | 0.7 | 3.9 | 64 | 272 | 1.20 (0.83 - 1.72) | 0.33 | 1.23 (0.85 - 1.77) | 0.30 |
| Open Claims | 118,300 | 373,356 | 85.1 | 320.7 | 2,145 | 8,088 | 1.16 (1.08 - 1.24) | <0.01 | 1.17 (1.03 - 1.33) | 0.04 |
| OEHR | 8,981 | 33,026 | 2.7 | 16.2 | 168 | 800 | 1.40 (1.12 - 1.73) | <0.01 | 1.34 (1.08 - 1.67) | 0.01 |
| Meta-analysis | 147,327 | 474,449 | 99.6 | 388.6 | 2,897 | 11,077 | 1.17 (1.11 - 1.24) | <0.01 | 1.18 (1.08 - 1.30) | <0.01 |
| Meta-analysis (w/o USOC) | 29,027 | 101,093 | 14.5 | 67.8 | 752 | 2,989 | 1.22 (1.09 - 1.36) | <0.01 | 1.24 (1.10 - 1.39) | <0.01 |
| **PS-stratification / on-treatment TAR** | | | | | | | | | | |
| CCAE | 16,274 | 38,768 | 9.3 | 29.7 | 457 | 1,050 | 1.17 (1.02 - 1.33) | 0.02 | 1.21 (1.06 - 1.39) | <0.01 |
| ODOD | 9,491 | 24,049 | 4.9 | 18.4 | 264 | 897 | 1.00 (0.84 - 1.18) | 0.98 | 0.99 (0.84 - 1.18) | 0.85 |
| MDCR | 960 | 7,235 | 0.6 | 6.6 | 25 | 285 | 0.97 (0.60 - 1.49) | 0.88 | 0.80 (0.51 - 1.26) | 0.36 |
| MDCD | 1,534 | 5,785 | 0.8 | 3.9 | 73 | 273 | 1.06 (0.79 - 1.41) | 0.69 | 1.07 (0.81 - 1.43) | 0.64 |
| Open Claims | 186,885 | 374,449 | 115.2 | 321.8 | 3,059 | 8,141 | 1.11 (1.06 - 1.17) | <0.01 | 1.16 (0.99 - 1.34) | 0.08 |
| OEHR | 11,445 | 33,203 | 3.3 | 16.4 | 199 | 804 | 1.22 (1.02 - 1.45) | 0.02 | 1.14 (0.96 - 1.37) | 0.19 |
| Meta-analysis | 225,629 | 476,254 | 133.4 | 390.2 | 4,052 | 11,165 | 1.11 (1.07 - 1.16) | <0.01 | 1.14 (1.02 - 1.29) | 0.02 |
| Meta-analysis (w/o USOC) | 38,744 | 101,805 | 18.2 | 68.4 | 993 | 3,024 | 1.12 (1.03 - 1.23) | 0.01 | 1.14 (1.04 - 1.26) | 0.01 |
| **PS-matching / on-treatment TAR without escalation** | | | | | | | | | | |
| CCAE | 12,135 | 38,768 | 6.7 | 24.8 | 303 | 865 | 1.27 (1.06 - 1.53) | 0.01 | 1.36 (1.13 - 1.64) | <0.01 |
| ODOD | 6,507 | 23,550 | 3.3 | 15.2 | 174 | 723 | 1.13 (0.89 - 1.44) | 0.30 | 1.09 (0.86 - 1.38) | 0.52 |
| MDCR | 840 | 7,035 | 0.5 | 5.7 | 21 | 244 | 1.31 (0.73 - 2.20) | 0.34 | 1.04 (0.60 - 1.82) | 0.82 |
| MDCD | 1,404 | 5,749 | 0.7 | 3.1 | 55 | 224 | 0.99 (0.65 - 1.47) | 0.95 | 1.03 (0.69 - 1.55) | 0.80 |
| Open Claims | 118,300 | 373,356 | 75.7 | 256.9 | 1,904 | 6,481 | 1.17 (1.08 - 1.26) | <0.01 | 1.18 (1.09 - 1.28) | <0.01 |
| OEHR | 8,981 | 33,026 | 2.3 | 13.2 | 152 | 638 | 1.49 (1.18 - 1.87) | <0.01 | 1.47 (1.16 - 1.86) | <0.01 |
| Meta-analysis | 147,327 | 474,449 | 88.6 | 313.3 | 2,588 | 8,931 | 1.21 (1.11 - 1.31) | <0.01 | 1.22 (1.09 - 1.36) | <0.01 |
| Meta-analysis (w/o USOC) | 29,027 | 101,093 | 13.0 | 56.4 | 684 | 2,450 | 1.25 (1.08 - 1.45) | <0.01 | 1.28 (1.11 - 1.49) | <0.01 |
| **PS-stratification / on-treatment TAR without escalation** | | | | | | | | | | |
| CCAE | 16,274 | 38,768 | 8.5 | 24.8 | 409 | 865 | 1.16 (1.01 - 1.34) | 0.03 | 1.21 (1.05 - 1.39) | 0.01 |
| ODOD | 9,491 | 24,049 | 4.5 | 15.6 | 249 | 752 | 1.06 (0.89 - 1.26) | 0.53 | 1.06 (0.88 - 1.26) | 0.62 |
| MDCR | 960 | 7,235 | 0.5 | 5.8 | 22 | 252 | 0.97 (0.58 - 1.53) | 0.89 | 0.79 (0.49 - 1.27) | 0.35 |
| MDCD | 1,534 | 5,785 | 0.7 | 3.1 | 64 | 225 | 1.01 (0.73 - 1.36) | 0.97 | 1.05 (0.77 - 1.43) | 0.76 |
| Open Claims | 186,885 | 374,449 | 104.1 | 257.8 | 2,767 | 6,522 | 1.12 (1.06 - 1.18) | <0.01 | 1.15 (0.98 - 1.35) | 0.11 |
| OEHR | 11,445 | 33,203 | 2.9 | 13.3 | 180 | 641 | 1.32 (1.09 - 1.58) | <0.01 | 1.23 (1.01 - 1.49) | 0.07 |
| Meta-analysis | 225,629 | 476,254 | 120.6 | 314.7 | 3,669 | 9,005 | 1.13 (1.08 - 1.18) | <0.01 | 1.15 (1.05 - 1.27) | <0.01 |
| Meta-analysis (w/o USOC) | 38,744 | 101,805 | 16.5 | 56.9 | 902 | 2,483 | 1.15 (1.04 - 1.27) | <0.01 | 1.18 (1.06 - 1.31) | <0.01 |

##### 1.4.16 Genitourinary infection

**Supplementary Table 94:** Exposures, outcomes and risk estimates for Genitourinary infection in GLP1RA (female) (T) vs DPP4I (female) (C) new-user cohorts under four different propensity score (PS) adjustment / time-at-risk (TAR) designs across data sources. We report total patient exposure time in 1,000 patient-year (KYr) units, uncalibrated and calibrated hazard ratios (HRs) and their 95% confidence intervals (CIs). Greyed out entries did not pass study diagnostics and are included only for completeness.

| | Patients | | Exposure (KYr) | | Outcomes | | Uncalibrated | | Calibrated | |
|---|---|---|---|---|---|---|---|---|---|---|
| | T | C | T | C | T | C | HR (95% CI) | $p$ | HR (95% CI) | $p$ |
| **PS-matching / on-treatment TAR** | | | | | | | | | | |
| CCAE | 9,865 | 31,463 | 5.8 | 23.4 | 425 | 1,750 | 1.00 (0.85 - 1.16) | 0.95 | 1.08 (0.92 - 1.26) | 0.46 |
| ODOD | 5,459 | 18,987 | 3.0 | 13.9 | 271 | 1,382 | 1.08 (0.90 - 1.29) | 0.42 | 1.05 (0.87 - 1.27) | 0.63 |
| MDCR | 672 | 5,842 | 0.4 | 5.2 | 32 | 489 | 0.84 (0.51 - 1.31) | 0.45 | 0.69 (0.43 - 1.10) | 0.13 |
| MDCD | 1,192 | 4,948 | 0.6 | 3.3 | 73 | 344 | 1.05 (0.73 - 1.48) | 0.79 | 1.07 (0.75 - 1.52) | 0.71 |
| Open Claims | 92,305 | 294,588 | 66.2 | 248.8 | 2,742 | 13,292 | 0.96 (0.90 - 1.02) | 0.17 | 0.97 (0.86 - 1.10) | 0.67 |
| OEHR | 8,089 | 29,595 | 2.4 | 14.3 | 192 | 1,107 | 1.18 (0.96 - 1.44) | 0.11 | 1.13 (0.92 - 1.39) | 0.28 |
| Meta-analysis | 116,910 | 379,581 | 78.0 | 303.7 | 3,703 | 17,875 | 1.01 (0.94 - 1.10) | 0.74 | 1.02 (0.92 - 1.14) | 0.70 |
| Meta-analysis (w/o USOC) | 24,605 | 84,993 | 11.8 | 54.9 | 961 | 4,583 | 1.06 (0.96 - 1.17) | 0.22 | 1.08 (0.97 - 1.20) | 0.19 |
| **PS-stratification / on-treatment TAR** | | | | | | | | | | |
| CCAE | 13,187 | 31,471 | 7.3 | 23.4 | 555 | 1,751 | 0.94 (0.84 - 1.05) | 0.30 | 0.98 (0.88 - 1.10) | 0.77 |
| ODOD | 7,974 | 19,378 | 4.0 | 14.3 | 361 | 1,439 | 1.00 (0.87 - 1.15) | 0.99 | 1.00 (0.86 - 1.15) | 0.83 |
| MDCR | 772 | 6,012 | 0.4 | 5.3 | 35 | 505 | 0.91 (0.61 - 1.32) | 0.64 | 0.76 (0.52 - 1.11) | 0.17 |
| MDCD | 1,310 | 4,991 | 0.7 | 3.3 | 78 | 346 | 1.03 (0.78 - 1.35) | 0.82 | 1.05 (0.80 - 1.38) | 0.74 |
| Open Claims | 144,467 | 295,210 | 89.2 | 249.4 | 3,719 | 13,368 | 0.96 (0.91 - 1.00) | 0.04 | 0.99 (0.86 - 1.15) | 0.89 |
| OEHR | 10,291 | 29,709 | 2.9 | 14.4 | 243 | 1,118 | 1.19 (1.02 - 1.39) | 0.03 | 1.12 (0.95 - 1.31) | 0.25 |
| Meta-analysis | 177,229 | 380,759 | 104.1 | 304.7 | 4,956 | 18,022 | 1.00 (0.92 - 1.08) | 1.00 | 1.03 (0.90 - 1.17) | 0.70 |
| Meta-analysis (w/o USOC) | 32,762 | 85,549 | 14.9 | 55.3 | 1,237 | 4,654 | 1.03 (0.92 - 1.15) | 0.61 | 1.05 (0.93 - 1.17) | 0.47 |
| **PS-matching / on-treatment TAR without escalation** | | | | | | | | | | |
| CCAE | 9,865 | 31,463 | 5.3 | 19.5 | 379 | 1,467 | 1.01 (0.86 - 1.19) | 0.87 | 1.08 (0.92 - 1.28) | 0.41 |
| ODOD | 5,459 | 18,987 | 2.7 | 11.9 | 240 | 1,169 | 1.04 (0.85 - 1.26) | 0.71 | 1.00 (0.82 - 1.22) | 0.81 |
| MDCR | 672 | 5,842 | 0.4 | 4.5 | 25 | 418 | 0.73 (0.42 - 1.20) | 0.24 | 0.59 (0.35 - 0.99) | 0.05 |
| MDCD | 1,192 | 4,948 | 0.6 | 2.6 | 59 | 254 | 1.12 (0.75 - 1.64) | 0.58 | 1.17 (0.79 - 1.74) | 0.46 |
| Open Claims | 92,305 | 294,588 | 58.7 | 199.1 | 2,385 | 10,685 | 0.94 (0.88 - 1.00) | 0.07 | 0.95 (0.88 - 1.02) | 0.34 |
| OEHR | 8,089 | 29,595 | 2.1 | 11.7 | 160 | 863 | 1.12 (0.89 - 1.41) | 0.32 | 1.11 (0.88 - 1.40) | 0.44 |
| Meta-analysis | 116,910 | 379,581 | 69.3 | 244.7 | 3,223 | 14,438 | 0.99 (0.92 - 1.07) | 0.82 | 1.00 (0.90 - 1.12) | 0.86 |
| Meta-analysis (w/o USOC) | 24,605 | 84,993 | 10.6 | 45.6 | 838 | 3,753 | 1.05 (0.95 - 1.17) | 0.35 | 1.08 (0.96 - 1.20) | 0.26 |
| **PS-stratification / on-treatment TAR without escalation** | | | | | | | | | | |
| CCAE | 13,187 | 31,471 | 6.7 | 19.5 | 505 | 1,468 | 0.94 (0.84 - 1.06) | 0.31 | 0.98 (0.87 - 1.10) | 0.74 |
| ODOD | 7,974 | 19,378 | 3.7 | 12.2 | 324 | 1,220 | 1.01 (0.87 - 1.17) | 0.89 | 1.01 (0.87 - 1.17) | 0.82 |
| MDCR | 772 | 6,012 | 0.4 | 4.6 | 27 | 431 | 0.77 (0.49 - 1.17) | 0.24 | 0.63 (0.41 - 0.97) | 0.04 |
| MDCD | 1,310 | 4,991 | 0.6 | 2.6 | 63 | 255 | 1.04 (0.75 - 1.40) | 0.83 | 1.08 (0.79 - 1.47) | 0.65 |
| Open Claims | 144,467 | 295,210 | 80.5 | 199.6 | 3,307 | 10,757 | 0.95 (0.91 - 1.00) | 0.04 | 0.98 (0.83 - 1.15) | 0.79 |
| OEHR | 10,291 | 29,709 | 2.5 | 11.7 | 206 | 874 | 1.19 (1.00 - 1.42) | 0.04 | 1.12 (0.93 - 1.34) | 0.33 |
| Meta-analysis | 177,229 | 380,759 | 94.1 | 245.6 | 4,405 | 14,574 | 1.00 (0.92 - 1.08) | 0.90 | 1.02 (0.91 - 1.14) | 0.78 |
| Meta-analysis (w/o USOC) | 32,762 | 85,549 | 13.5 | 46.0 | 1,098 | 3,817 | 1.03 (0.92 - 1.15) | 0.62 | 1.05 (0.94 - 1.18) | 0.44 |

##### 1.4.17 Hyperkalemia

**Supplementary Table 95:** Exposures, outcomes and risk estimates for Hyperkalemia in GLP1RA (female) (T) vs DPP4I (female) (C) new-user cohorts under four different propensity score (PS) adjustment / time-at-risk (TAR) designs across data sources. We report total patient exposure time in 1,000 patient-year (KYr) units, uncalibrated and calibrated hazard ratios (HRs) and their 95% confidence intervals (CIs). Greyed out entries did not pass study diagnostics and are included only for completeness.

| | Patients | | Exposure (KYr) | | Outcomes | | Uncalibrated | | Calibrated | |
|---|---|---|---|---|---|---|---|---|---|---|
| | T | C | T | C | T | C | HR (95% CI) | $p$ | HR (95% CI) | $p$ |
| **PS-matching / on-treatment TAR** | | | | | | | | | | |
| CCAE | 14,235 | 44,801 | 8.9 | 35.3 | 37 | 205 | 1.19 (0.73 - 1.91) | 0.47 | 1.29 (0.80 - 2.10) | 0.32 |
| ODOD | 7,683 | 27,663 | 4.5 | 21.5 | 51 | 463 | 0.68 (0.45 - 1.00) | 0.06 | 0.66 (0.44 - 0.99) | 0.05 |
| MDCR | 966 | 8,221 | 0.6 | 7.8 | 5 | 104 | 0.49 (0.12 - 1.45) | 0.27 | 0.41 (0.11 - 1.44) | 0.16 |
| MDCD | 1,807 | 7,452 | 1.0 | 5.2 | 8 | 92 | 0.55 (0.18 - 1.36) | 0.25 | 0.56 (0.21 - 1.54) | 0.27 |
| Open Claims | 142,284 | 438,407 | 103.5 | 382.1 | 341 | 2,955 | 0.67 (0.58 - 0.79) | <0.01 | 0.68 (0.57 - 0.82) | <0.01 |
| OEHR | 10,414 | 37,959 | 3.2 | 19.5 | 25 | 255 | 0.67 (0.38 - 1.11) | 0.13 | 0.64 (0.38 - 1.09) | 0.11 |
| Meta-analysis | 176,423 | 556,282 | 121.1 | 463.6 | 462 | 3,970 | 0.72 (0.60 - 0.87) | <0.01 | 0.73 (0.59 - 0.88) | <0.01 |
| Meta-analysis (w/o USOC) | 34,139 | 117,875 | 17.6 | 81.4 | 121 | 1,015 | 0.78 (0.57 - 1.07) | 0.13 | 0.79 (0.57 - 1.09) | 0.16 |
| **PS-stratification / on-treatment TAR** | | | | | | | | | | |
| CCAE | 19,147 | 44,818 | 11.3 | 35.3 | 47 | 206 | 0.95 (0.65 - 1.34) | 0.76 | 0.99 (0.69 - 1.42) | 0.92 |
| ODOD | 11,337 | 28,155 | 6.0 | 22.0 | 67 | 480 | 0.64 (0.47 - 0.86) | <0.01 | 0.64 (0.47 - 0.87) | <0.01 |
| MDCR | 1,105 | 8,538 | 0.7 | 8.0 | 6 | 119 | 0.58 (0.21 - 1.31) | 0.24 | 0.48 (0.19 - 1.19) | 0.12 |
| MDCD | 1,985 | 7,515 | 1.1 | 5.3 | 8 | 94 | 0.52 (0.22 - 1.05) | 0.10 | 0.53 (0.24 - 1.14) | 0.11 |
| Open Claims | 227,579 | 439,399 | 141.5 | 383.1 | 427 | 2,989 | 0.68 (0.60 - 0.76) | <0.01 | 0.70 (0.58 - 0.85) | <0.01 |
| OEHR | 13,435 | 38,147 | 4.0 | 19.6 | 30 | 261 | 0.73 (0.47 - 1.09) | 0.14 | 0.68 (0.45 - 1.04) | 0.09 |
| Meta-analysis | 273,483 | 558,034 | 163.8 | 465.1 | 579 | 4,030 | 0.69 (0.62 - 0.76) | <0.01 | 0.71 (0.61 - 0.82) | <0.01 |
| Meta-analysis (w/o USOC) | 45,904 | 118,635 | 22.3 | 82.1 | 152 | 1,041 | 0.73 (0.59 - 0.90) | <0.01 | 0.74 (0.60 - 0.92) | <0.01 |
| **PS-matching / on-treatment TAR without escalation** | | | | | | | | | | |
| CCAE | 14,235 | 44,801 | 8.0 | 29.0 | 33 | 166 | 1.33 (0.79 - 2.20) | 0.28 | 1.42 (0.85 - 2.38) | 0.19 |
| ODOD | 7,683 | 27,663 | 4.0 | 18.1 | 50 | 379 | 0.74 (0.48 - 1.11) | 0.16 | 0.71 (0.46 - 1.09) | 0.12 |
| MDCR | 966 | 8,221 | 0.6 | 6.8 | 5 | 91 | 0.56 (0.13 - 1.70) | 0.38 | 0.45 (0.12 - 1.63) | 0.23 |
| MDCD | 1,807 | 7,452 | 0.8 | 4.1 | 8 | 71 | 0.64 (0.21 - 1.63) | 0.40 | 0.67 (0.24 - 1.88) | 0.45 |
| Open Claims | 142,284 | 438,407 | 91.7 | 303.6 | 294 | 2,339 | 0.71 (0.60 - 0.83) | <0.01 | 0.71 (0.60 - 0.85) | <0.01 |
| OEHR | 10,414 | 37,959 | 2.8 | 15.7 | 23 | 190 | 0.89 (0.48 - 1.56) | 0.70 | 0.88 (0.48 - 1.58) | 0.67 |
| Meta-analysis | 176,423 | 556,282 | 107.4 | 370.4 | 408 | 3,145 | 0.81 (0.64 - 1.03) | 0.09 | 0.82 (0.64 - 1.06) | 0.13 |
| Meta-analysis (w/o USOC) | 34,139 | 117,875 | 15.6 | 66.9 | 114 | 806 | 0.90 (0.67 - 1.23) | 0.52 | 0.93 (0.68 - 1.26) | 0.63 |
| **PS-stratification / on-treatment TAR without escalation** | | | | | | | | | | |
| CCAE | 19,147 | 44,818 | 10.2 | 29.0 | 43 | 167 | 0.96 (0.65 - 1.40) | 0.84 | 1.00 (0.68 - 1.47) | 0.94 |
| ODOD | 11,337 | 28,155 | 5.4 | 18.4 | 64 | 395 | 0.73 (0.53 - 0.99) | 0.05 | 0.73 (0.53 - 1.00) | 0.06 |
| MDCR | 1,105 | 8,538 | 0.6 | 7.0 | 6 | 104 | 0.60 (0.22 - 1.38) | 0.28 | 0.49 (0.19 - 1.23) | 0.13 |
| MDCD | 1,985 | 7,515 | 0.9 | 4.1 | 8 | 73 | 0.63 (0.27 - 1.33) | 0.26 | 0.66 (0.30 - 1.47) | 0.32 |
| Open Claims | 227,579 | 439,399 | 127.7 | 304.3 | 374 | 2,370 | 0.68 (0.60 - 0.78) | <0.01 | 0.70 (0.58 - 0.86) | <0.01 |
| OEHR | 13,435 | 38,147 | 3.4 | 15.8 | 28 | 193 | 0.82 (0.52 - 1.26) | 0.39 | 0.77 (0.49 - 1.20) | 0.27 |
| Meta-analysis | 273,483 | 558,034 | 147.7 | 371.7 | 517 | 3,198 | 0.72 (0.64 - 0.82) | <0.01 | 0.74 (0.63 - 0.86) | <0.01 |
| Meta-analysis (w/o USOC) | 45,904 | 118,635 | 20.0 | 67.4 | 143 | 828 | 0.80 (0.65 - 0.99) | 0.04 | 0.82 (0.66 - 1.01) | 0.07 |

##### 1.4.18 Hypoglycemia

**Supplementary Table 96:** Exposures, outcomes and risk estimates for Hypoglycemia in GLP1RA (female) (T) vs DPP4I (female) (C) new-user cohorts under four different propensity score (PS) adjustment / time-at-risk (TAR) designs across data sources. We report total patient exposure time in 1,000 patient-year (KYr) units, uncalibrated and calibrated hazard ratios (HRs) and their 95% confidence intervals (CIs). Greyed out entries did not pass study diagnostics and are included only for completeness.

| | Patients | | Exposure (KYr) | | Outcomes | | Uncalibrated | | Calibrated | |
|---|---|---|---|---|---|---|---|---|---|---|
| | T | C | T | C | T | C | HR (95% CI) | $p$ | HR (95% CI) | $p$ |
| **PS-matching / on-treatment TAR** | | | | | | | | | | |
| CCAE | 14,052 | 44,595 | 8.9 | 35.3 | 41 | 120 | 1.33 (0.79 - 2.21) | 0.28 | 1.44 (0.86 - 2.41) | 0.18 |
| ODOD | 7,741 | 28,286 | 4.5 | 22.4 | 32 | 175 | 1.15 (0.67 - 1.89) | 0.61 | 1.12 (0.66 - 1.88) | 0.68 |
| MDCR | 972 | 8,349 | 0.6 | 7.9 | <5 | 42 | 0.59 (0.09 - 2.51) | 0.54 | 0.49 (0.09 - 2.65) | 0.41 |
| MDCD | 1,787 | 7,432 | 1.0 | 5.3 | 9 | 58 | 1.29 (0.48 - 3.13) | 0.60 | 1.31 (0.51 - 3.37) | 0.57 |
| Open Claims | 141,638 | 440,239 | 103.2 | 386.3 | 284 | 1,258 | 0.94 (0.78 - 1.13) | 0.52 | 0.95 (0.77 - 1.18) | 0.66 |
| OEHR | 10,394 | 38,200 | 3.2 | 19.7 | 27 | 137 | 0.99 (0.53 - 1.79) | 0.98 | 0.95 (0.52 - 1.75) | 0.88 |
| Meta-analysis | 175,612 | 558,752 | 120.7 | 468.9 | 393 | 1,748 | 1.00 (0.85 - 1.17) | 0.98 | 1.01 (0.85 - 1.20) | 0.90 |
| Meta-analysis (w/o USOC) | 33,974 | 118,513 | 17.6 | 82.6 | 109 | 490 | 1.18 (0.87 - 1.58) | 0.28 | 1.20 (0.89 - 1.61) | 0.25 |
| **PS-stratification / on-treatment TAR** | | | | | | | | | | |
| CCAE | 18,798 | 44,599 | 11.1 | 35.3 | 60 | 120 | 1.22 (0.84 - 1.74) | 0.29 | 1.27 (0.88 - 1.83) | 0.21 |
| ODOD | 11,351 | 28,947 | 6.0 | 23.0 | 56 | 182 | 1.04 (0.71 - 1.51) | 0.82 | 1.04 (0.71 - 1.52) | 0.85 |
| MDCR | 1,110 | 8,701 | 0.7 | 8.2 | <5 | 45 | 0.51 (0.08 - 1.88) | 0.40 | 0.42 (0.09 - 2.08) | 0.29 |
| MDCD | 1,953 | 7,505 | 1.1 | 5.3 | 9 | 60 | 0.94 (0.42 - 1.92) | 0.88 | 0.95 (0.44 - 2.05) | 0.90 |
| Open Claims | 225,048 | 441,240 | 140.3 | 387.3 | 401 | 1,269 | 0.96 (0.84 - 1.10) | 0.57 | 1.00 (0.82 - 1.22) | 0.92 |
| OEHR | 13,387 | 38,401 | 3.9 | 19.8 | 31 | 139 | 1.02 (0.65 - 1.57) | 0.92 | 0.96 (0.62 - 1.49) | 0.86 |
| Meta-analysis | 270,537 | 560,692 | 162.5 | 470.7 | 557 | 1,770 | 1.00 (0.89 - 1.12) | 0.95 | 1.02 (0.87 - 1.20) | 0.78 |
| Meta-analysis (w/o USOC) | 45,489 | 119,452 | 22.1 | 83.4 | 156 | 501 | 1.09 (0.88 - 1.35) | 0.45 | 1.11 (0.89 - 1.38) | 0.38 |
| **PS-matching / on-treatment TAR without escalation** | | | | | | | | | | |
| CCAE | 14,052 | 44,595 | 8.0 | 29.0 | 34 | 70 | 1.61 (0.89 - 2.88) | 0.11 | 1.72 (0.95 - 3.10) | 0.08 |
| ODOD | 7,741 | 28,286 | 4.1 | 18.8 | 23 | 96 | 1.32 (0.70 - 2.41) | 0.38 | 1.27 (0.68 - 2.36) | 0.46 |
| MDCR | 972 | 8,349 | 0.6 | 6.9 | <5 | 26 | 0.71 (0.10 - 3.35) | 0.70 | 0.57 (0.10 - 3.31) | 0.53 |
| MDCD | 1,787 | 7,432 | 0.8 | 4.1 | 7 | 32 | 1.26 (0.42 - 3.45) | 0.66 | 1.32 (0.46 - 3.79) | 0.60 |
| Open Claims | 141,638 | 440,239 | 91.4 | 307.1 | 233 | 685 | 1.20 (0.96 - 1.49) | 0.10 | 1.21 (0.97 - 1.51) | 0.11 |
| OEHR | 10,394 | 38,200 | 2.8 | 15.9 | 20 | 66 | 1.19 (0.55 - 2.43) | 0.65 | 1.17 (0.56 - 2.46) | 0.68 |
| Meta-analysis | 175,612 | 558,752 | 107.1 | 374.9 | 317 | 949 | 1.25 (1.04 - 1.50) | 0.02 | 1.26 (1.03 - 1.54) | 0.02 |
| Meta-analysis (w/o USOC) | 33,974 | 118,513 | 15.6 | 67.8 | 84 | 264 | 1.37 (0.97 - 1.95) | 0.07 | 1.41 (0.99 - 2.00) | 0.06 |
| **PS-stratification / on-treatment TAR without escalation** | | | | | | | | | | |
| CCAE | 18,798 | 44,599 | 10.1 | 29.0 | 52 | 70 | 1.42 (0.93 - 2.17) | 0.10 | 1.48 (0.97 - 2.26) | 0.07 |
| ODOD | 11,351 | 28,947 | 5.5 | 19.3 | 46 | 101 | 1.21 (0.77 - 1.89) | 0.39 | 1.21 (0.77 - 1.90) | 0.42 |
| MDCR | 1,110 | 8,701 | 0.6 | 7.1 | <5 | 28 | 0.60 (0.09 - 2.37) | 0.54 | 0.49 (0.10 - 2.53) | 0.40 |
| MDCD | 1,953 | 7,505 | 0.9 | 4.2 | 7 | 33 | 0.97 (0.37 - 2.25) | 0.94 | 1.01 (0.41 - 2.49) | 0.93 |
| Open Claims | 225,048 | 441,240 | 126.6 | 307.9 | 337 | 690 | 1.23 (1.05 - 1.45) | 0.01 | 1.27 (1.01 - 1.58) | 0.04 |
| OEHR | 13,387 | 38,401 | 3.4 | 16.0 | 24 | 66 | 1.28 (0.73 - 2.17) | 0.37 | 1.20 (0.69 - 2.06) | 0.54 |
| Meta-analysis | 270,537 | 560,692 | 146.5 | 376.4 | 466 | 960 | 1.25 (1.09 - 1.43) | <0.01 | 1.27 (1.08 - 1.49) | <0.01 |
| Meta-analysis (w/o USOC) | 45,489 | 119,452 | 19.9 | 68.5 | 129 | 270 | 1.28 (0.99 - 1.65) | 0.06 | 1.31 (1.01 - 1.69) | 0.05 |

##### 1.4.19 Hypotension

**Supplementary Table 97:** Exposures, outcomes and risk estimates for Hypotension in GLP1RA (female) (T) vs DPP4I (female) (C) new-user cohorts under four different propensity score (PS) adjustment / time-at-risk (TAR) designs across data sources. We report total patient exposure time in 1,000 patient-year (KYr) units, uncalibrated and calibrated hazard ratios (HRs) and their 95% confidence intervals (CIs). Greyed out entries did not pass study diagnostics and are included only for completeness.

| | Patients | | Exposure (KYr) | | Outcomes | | Uncalibrated | | Calibrated | |
|---|---|---|---|---|---|---|---|---|---|---|
| | T | C | T | C | T | C | HR (95% CI) | $p$ | HR (95% CI) | $p$ |
| **PS-matching / on-treatment TAR** | | | | | | | | | | |
| CCAE | 14,152 | 44,621 | 8.9 | 35.2 | 72 | 257 | 1.11 (0.74 - 1.62) | 0.62 | 1.20 (0.81 - 1.77) | 0.39 |
| ODOD | 7,602 | 27,545 | 4.4 | 21.6 | 74 | 461 | 0.85 (0.60 - 1.19) | 0.35 | 0.83 (0.58 - 1.17) | 0.29 |
| MDCR | 934 | 8,043 | 0.6 | 7.6 | 9 | 155 | 1.06 (0.45 - 2.22) | 0.89 | 0.87 (0.39 - 1.94) | 0.74 |
| MDCD | 1,753 | 7,149 | 0.9 | 5.0 | 28 | 151 | 0.93 (0.53 - 1.58) | 0.80 | 0.95 (0.55 - 1.65) | 0.84 |
| Open Claims | 139,697 | 431,616 | 101.4 | 376.8 | 697 | 3,615 | 0.97 (0.87 - 1.09) | 0.66 | 0.99 (0.84 - 1.15) | 0.86 |
| OEHR | 10,378 | 37,870 | 3.2 | 19.4 | 52 | 281 | 1.66 (1.12 - 2.43) | <0.01 | 1.60 (1.09 - 2.36) | 0.02 |
| Meta-analysis | 173,582 | 548,801 | 118.8 | 457.9 | 923 | 4,765 | 1.06 (0.85 - 1.31) | 0.61 | 1.07 (0.85 - 1.33) | 0.58 |
| Meta-analysis (w/o USOC) | 33,885 | 117,185 | 17.4 | 81.1 | 226 | 1,150 | 1.10 (0.81 - 1.51) | 0.53 | 1.12 (0.82 - 1.54) | 0.47 |
| **PS-stratification / on-treatment TAR** | | | | | | | | | | |
| CCAE | 19,037 | 44,624 | 11.2 | 35.2 | 98 | 257 | 1.09 (0.83 - 1.43) | 0.54 | 1.14 (0.86 - 1.50) | 0.38 |
| ODOD | 11,208 | 28,041 | 5.9 | 22.0 | 103 | 486 | 1.02 (0.78 - 1.32) | 0.90 | 1.01 (0.78 - 1.32) | 0.89 |
| MDCR | 1,073 | 8,338 | 0.6 | 7.8 | 9 | 172 | 0.73 (0.33 - 1.42) | 0.39 | 0.61 (0.29 - 1.25) | 0.18 |
| MDCD | 1,922 | 7,199 | 1.0 | 5.0 | 28 | 153 | 0.96 (0.60 - 1.47) | 0.84 | 0.97 (0.62 - 1.51) | 0.86 |
| Open Claims | 223,218 | 432,712 | 138.6 | 377.9 | 939 | 3,663 | 0.99 (0.90 - 1.08) | 0.80 | 1.03 (0.87 - 1.22) | 0.76 |
| OEHR | 13,380 | 38,051 | 3.9 | 19.5 | 60 | 287 | 1.52 (1.11 - 2.06) | <0.01 | 1.43 (1.04 - 1.95) | 0.03 |
| Meta-analysis | 268,765 | 550,627 | 160.7 | 459.6 | 1,228 | 4,846 | 1.07 (0.93 - 1.24) | 0.33 | 1.10 (0.92 - 1.32) | 0.28 |
| Meta-analysis (w/o USOC) | 45,547 | 117,915 | 22.0 | 81.7 | 289 | 1,183 | 1.14 (0.94 - 1.38) | 0.20 | 1.16 (0.95 - 1.41) | 0.16 |
| **PS-matching / on-treatment TAR without escalation** | | | | | | | | | | |
| CCAE | 14,152 | 44,621 | 8.0 | 29.0 | 62 | 202 | 1.03 (0.66 - 1.57) | 0.90 | 1.10 (0.71 - 1.70) | 0.68 |
| ODOD | 7,602 | 27,545 | 4.0 | 18.1 | 65 | 386 | 0.88 (0.60 - 1.26) | 0.49 | 0.84 (0.58 - 1.22) | 0.39 |
| MDCR | 934 | 8,043 | 0.6 | 6.6 | 9 | 136 | 1.00 (0.42 - 2.12) | 0.99 | 0.80 (0.35 - 1.80) | 0.59 |
| MDCD | 1,753 | 7,149 | 0.8 | 3.9 | 23 | 103 | 1.28 (0.68 - 2.30) | 0.43 | 1.34 (0.73 - 2.47) | 0.37 |
| Open Claims | 139,697 | 431,616 | 89.9 | 299.5 | 584 | 2,874 | 0.94 (0.83 - 1.07) | 0.37 | 0.95 (0.84 - 1.09) | 0.54 |
| OEHR | 10,378 | 37,870 | 2.7 | 15.7 | 41 | 194 | 1.77 (1.12 - 2.74) | 0.01 | 1.74 (1.11 - 2.73) | 0.02 |
| Meta-analysis | 173,582 | 548,801 | 105.4 | 366.2 | 775 | 3,759 | 1.08 (0.85 - 1.36) | 0.52 | 1.09 (0.85 - 1.40) | 0.48 |
| Meta-analysis (w/o USOC) | 33,885 | 117,185 | 15.5 | 66.7 | 191 | 885 | 1.17 (0.85 - 1.61) | 0.34 | 1.20 (0.87 - 1.65) | 0.29 |
| **PS-stratification / on-treatment TAR without escalation** | | | | | | | | | | |
| CCAE | 19,037 | 44,624 | 10.1 | 29.0 | 87 | 202 | 1.10 (0.81 - 1.48) | 0.53 | 1.15 (0.85 - 1.55) | 0.39 |
| ODOD | 11,208 | 28,041 | 5.4 | 18.5 | 93 | 410 | 1.10 (0.82 - 1.45) | 0.52 | 1.09 (0.82 - 1.46) | 0.57 |
| MDCR | 1,073 | 8,338 | 0.6 | 6.8 | 9 | 151 | 0.79 (0.36 - 1.55) | 0.53 | 0.64 (0.31 - 1.34) | 0.25 |
| MDCD | 1,922 | 7,199 | 0.9 | 4.0 | 23 | 104 | 0.96 (0.57 - 1.56) | 0.87 | 1.00 (0.60 - 1.66) | 0.89 |
| Open Claims | 223,218 | 432,712 | 125.1 | 300.4 | 809 | 2,916 | 0.96 (0.87 - 1.05) | 0.36 | 0.98 (0.82 - 1.18) | 0.85 |
| OEHR | 13,380 | 38,051 | 3.4 | 15.8 | 47 | 200 | 1.67 (1.16 - 2.36) | <0.01 | 1.56 (1.09 - 2.24) | 0.02 |
| Meta-analysis | 268,765 | 550,627 | 144.9 | 367.6 | 1,059 | 3,832 | 1.11 (0.92 - 1.33) | 0.30 | 1.13 (0.92 - 1.39) | 0.25 |
| Meta-analysis (w/o USOC) | 45,547 | 117,915 | 19.8 | 67.2 | 250 | 916 | 1.19 (0.96 - 1.48) | 0.11 | 1.22 (0.98 - 1.52) | 0.09 |

##### 1.4.20 Joint pain

**Supplementary Table 98:** Exposures, outcomes and risk estimates for Joint pain in GLP1RA (female) (T) vs DPP4I (female) (C) new-user cohorts under four different propensity score (PS) adjustment / time-at-risk (TAR) designs across data sources. We report total patient exposure time in 1,000 patient-year (KYr) units, uncalibrated and calibrated hazard ratios (HRs) and their 95% confidence intervals (CIs). Greyed out entries did not pass study diagnostics and are included only for completeness.

| | Patients | | Exposure (KYr) | | Outcomes | | Uncalibrated | | Calibrated | |
|---|---|---|---|---|---|---|---|---|---|---|
| | T | C | T | C | T | C | HR (95% CI) | $p$ | HR (95% CI) | $p$ |
| **PS-matching / on-treatment TAR** | | | | | | | | | | |
| CCAE | 13,190 | 42,276 | 8.2 | 33.2 | 128 | 487 | 1.01 (0.75 - 1.36) | 0.94 | 1.10 (0.81 - 1.48) | 0.58 |
| ODOD | 7,217 | 26,640 | 4.2 | 20.9 | 83 | 396 | 0.84 (0.59 - 1.18) | 0.32 | 0.82 (0.58 - 1.16) | 0.27 |
| MDCR | 918 | 8,004 | 0.6 | 7.6 | 5 | 97 | 0.38 (0.11 - 1.01) | 0.09 | 0.32 (0.10 - 0.95) | 0.04 |
| MDCD | 1,623 | 6,807 | 0.9 | 4.7 | 24 | 145 | 0.74 (0.40 - 1.30) | 0.31 | 0.75 (0.42 - 1.36) | 0.36 |
| Open Claims | 131,575 | 413,816 | 95.6 | 361.2 | 780 | 3,517 | 0.79 (0.71 - 0.88) | <0.01 | 0.80 (0.68 - 0.93) | 0.01 |
| OEHR | 9,737 | 36,169 | 3.0 | 18.4 | 51 | 310 | 0.96 (0.61 - 1.46) | 0.85 | 0.92 (0.60 - 1.42) | 0.72 |
| Meta-analysis | 163,342 | 525,708 | 111.8 | 438.3 | 1,066 | 4,855 | 0.83 (0.74 - 0.94) | <0.01 | 0.84 (0.73 - 0.96) | 0.01 |
| Meta-analysis (w/o USOC) | 31,767 | 111,892 | 16.2 | 77.2 | 286 | 1,338 | 0.92 (0.76 - 1.11) | 0.36 | 0.93 (0.77 - 1.13) | 0.48 |
| **PS-stratification / on-treatment TAR** | | | | | | | | | | |
| CCAE | 17,578 | 42,281 | 10.3 | 33.2 | 163 | 487 | 0.96 (0.78 - 1.18) | 0.70 | 1.00 (0.81 - 1.23) | 0.89 |
| ODOD | 10,448 | 27,243 | 5.5 | 21.4 | 109 | 412 | 0.86 (0.67 - 1.11) | 0.25 | 0.86 (0.67 - 1.11) | 0.27 |
| MDCR | 1,038 | 8,335 | 0.6 | 7.8 | 5 | 100 | 0.55 (0.19 - 1.27) | 0.22 | 0.45 (0.17 - 1.19) | 0.11 |
| MDCD | 1,769 | 6,865 | 0.9 | 4.7 | 28 | 148 | 0.74 (0.47 - 1.13) | 0.18 | 0.75 (0.48 - 1.17) | 0.21 |
| Open Claims | 207,745 | 415,056 | 129.4 | 362.4 | 1,119 | 3,534 | 0.83 (0.76 - 0.90) | <0.01 | 0.86 (0.73 - 1.01) | 0.09 |
| OEHR | 12,431 | 36,357 | 3.6 | 18.5 | 66 | 310 | 1.02 (0.75 - 1.38) | 0.88 | 0.96 (0.71 - 1.30) | 0.80 |
| Meta-analysis | 249,971 | 527,802 | 149.7 | 440.3 | 1,485 | 4,891 | 0.86 (0.79 - 0.94) | <0.01 | 0.88 (0.77 - 1.02) | 0.08 |
| Meta-analysis (w/o USOC) | 42,226 | 112,746 | 20.3 | 77.9 | 366 | 1,357 | 0.92 (0.80 - 1.05) | 0.23 | 0.94 (0.82 - 1.08) | 0.39 |
| **PS-matching / on-treatment TAR without escalation** | | | | | | | | | | |
| CCAE | 13,190 | 42,276 | 7.4 | 27.4 | 112 | 402 | 1.03 (0.75 - 1.41) | 0.84 | 1.11 (0.81 - 1.52) | 0.56 |
| ODOD | 7,217 | 26,640 | 3.7 | 17.6 | 70 | 330 | 0.82 (0.56 - 1.17) | 0.28 | 0.78 (0.54 - 1.14) | 0.22 |
| MDCR | 918 | 8,004 | 0.6 | 6.6 | 5 | 84 | 0.39 (0.09 - 1.16) | 0.15 | 0.31 (0.09 - 1.12) | 0.08 |
| MDCD | 1,623 | 6,807 | 0.7 | 3.7 | 22 | 113 | 0.86 (0.44 - 1.59) | 0.64 | 0.90 (0.47 - 1.72) | 0.76 |
| Open Claims | 131,575 | 413,816 | 84.6 | 287.2 | 689 | 2,792 | 0.80 (0.71 - 0.91) | <0.01 | 0.81 (0.71 - 0.92) | <0.01 |
| OEHR | 9,737 | 36,169 | 2.6 | 14.9 | 42 | 241 | 0.85 (0.51 - 1.37) | 0.51 | 0.83 (0.51 - 1.37) | 0.49 |
| Meta-analysis | 163,342 | 525,708 | 99.0 | 350.6 | 935 | 3,878 | 0.83 (0.75 - 0.93) | <0.01 | 0.84 (0.74 - 0.96) | 0.01 |
| Meta-analysis (w/o USOC) | 31,767 | 111,892 | 14.4 | 63.4 | 246 | 1,086 | 0.91 (0.74 - 1.12) | 0.38 | 0.93 (0.76 - 1.15) | 0.54 |
| **PS-stratification / on-treatment TAR without escalation** | | | | | | | | | | |
| CCAE | 17,578 | 42,281 | 9.3 | 27.4 | 143 | 402 | 0.92 (0.73 - 1.15) | 0.46 | 0.96 (0.76 - 1.20) | 0.72 |
| ODOD | 10,448 | 27,243 | 5.0 | 18.0 | 94 | 345 | 0.87 (0.66 - 1.14) | 0.30 | 0.86 (0.65 - 1.14) | 0.33 |
| MDCR | 1,038 | 8,335 | 0.6 | 6.8 | 5 | 87 | 0.64 (0.22 - 1.50) | 0.36 | 0.52 (0.20 - 1.37) | 0.19 |
| MDCD | 1,769 | 6,865 | 0.8 | 3.7 | 26 | 115 | 0.81 (0.50 - 1.28) | 0.38 | 0.84 (0.52 - 1.35) | 0.49 |
| Open Claims | 207,745 | 415,056 | 116.6 | 288.2 | 1,006 | 2,807 | 0.84 (0.77 - 0.92) | <0.01 | 0.86 (0.72 - 1.03) | 0.12 |
| OEHR | 12,431 | 36,357 | 3.2 | 14.9 | 57 | 241 | 1.00 (0.71 - 1.38) | 1.00 | 0.94 (0.67 - 1.31) | 0.72 |
| Meta-analysis | 249,971 | 527,802 | 134.8 | 352.2 | 1,326 | 3,910 | 0.86 (0.80 - 0.92) | <0.01 | 0.88 (0.78 - 0.98) | 0.02 |
| Meta-analysis (w/o USOC) | 42,226 | 112,746 | 18.3 | 64.0 | 320 | 1,103 | 0.91 (0.78 - 1.05) | 0.19 | 0.93 (0.80 - 1.08) | 0.36 |

##### 1.4.21 Lower extremity amputation

**Supplementary Table 99:** Exposures, outcomes and risk estimates for Lower extremity amputation in GLP1RA (female) (T) vs DPP4I (female) (C) new-user cohorts under four different propensity score (PS) adjustment / time-at-risk (TAR) designs across data sources. We report total patient exposure time in 1,000 patient-year (KYr) units, uncalibrated and calibrated hazard ratios (HRs) and their 95% confidence intervals (CIs). Greyed out entries did not pass study diagnostics and are included only for completeness.

| | Patients | | Exposure (KYr) | | Outcomes | | Uncalibrated | | Calibrated | |
|---|---|---|---|---|---|---|---|---|---|---|
| | T | C | T | C | T | C | HR (95% CI) | $p$ | HR (95% CI) | $p$ |
| **PS-matching / on-treatment TAR** | | | | | | | | | | |
| CCAE | 14,428 | 45,476 | 9.1 | 36.0 | <5 | 10 | 0.79 (0.11 - 3.62) | 0.79 | 0.86 (0.15 - 4.90) | 0.86 |
| ODOD | 7,911 | 28,787 | 4.6 | 22.8 | <5 | 10 | 2.26 (0.11 - 17.2) | 0.53 | 2.21 (0.18 - 27.7) | 0.54 |
| MDCR | 985 | 8,426 | 0.6 | 8.0 | 0 | 5 | | | | |
| MDCD | 1,858 | 7,679 | 1.0 | 5.5 | <5 | 5 | 1.42 (0.06 - 16.2) | 0.80 | 1.45 (0.09 - 23.2) | 0.79 |
| Open Claims | 144,729 | 448,280 | 105.5 | 393.9 | 19 | 107 | 0.88 (0.42 - 1.70) | 0.71 | 0.88 (0.44 - 1.79) | 0.74 |
| OEHR | 10,560 | 38,681 | 3.3 | 20.1 | 0 | 7 | | | | |
| Meta-analysis | 144,729 | 448,280 | 105.5 | 393.9 | 19 | 107 | 0.88 (0.42 - 1.70) | 0.71 | 0.88 (0.44 - 1.77) | 0.73 |
| **PS-stratification / on-treatment TAR** | | | | | | | | | | |
| CCAE | 19,436 | 45,481 | 11.5 | 36.0 | <5 | 10 | 1.21 (0.24 - 4.46) | 0.80 | 1.26 (0.29 - 5.38) | 0.76 |
| ODOD | 11,652 | 29,479 | 6.2 | 23.5 | <5 | 10 | 1.55 (0.08 - 9.19) | 0.72 | 1.54 (0.14 - 16.4) | 0.72 |
| MDCR | 1,127 | 8,789 | 0.7 | 8.3 | 0 | 6 | | | | |
| MDCD | 2,031 | 7,756 | 1.1 | 5.5 | <5 | 5 | 1.64 (0.07 - 13.6) | 0.71 | 1.67 (0.12 - 22.7) | 0.70 |
| Open Claims | 231,103 | 449,385 | 144.0 | 395.0 | 22 | 110 | 0.90 (0.51 - 1.51) | 0.69 | 0.93 (0.53 - 1.63) | 0.80 |
| OEHR | 13,629 | 38,885 | 4.0 | 20.2 | 0 | 7 | | | | |
| Meta-analysis | 231,103 | 449,385 | 144.0 | 395.0 | 22 | 110 | 0.90 (0.51 - 1.51) | 0.69 | 0.92 (0.53 - 1.59) | 0.77 |
| **PS-matching / on-treatment TAR without escalation** | | | | | | | | | | |
| CCAE | 14,428 | 45,476 | 8.2 | 29.6 | <5 | 9 | 0.90 (0.12 - 4.38) | 0.91 | 0.96 (0.16 - 5.74) | 0.97 |
| ODOD | 7,911 | 28,787 | 4.1 | 19.1 | <5 | 8 | 2.03 (0.10 - 15.1) | 0.58 | 1.95 (0.16 - 24.2) | 0.60 |
| MDCR | 985 | 8,426 | 0.6 | 7.0 | 0 | 5 | | | | |
| MDCD | 1,858 | 7,679 | 0.9 | 4.3 | <5 | <5 | 2.00 (0.08 - 50.5) | 0.67 | 2.09 (0.08 - 52.9) | 0.65 |
| Open Claims | 144,729 | 448,280 | 93.4 | 312.7 | 13 | 85 | 0.57 (0.22 - 1.29) | 0.21 | 0.58 (0.24 - 1.39) | 0.22 |
| OEHR | 10,560 | 38,681 | 2.8 | 16.2 | 0 | <5 | | | | |
| Meta-analysis | 144,729 | 448,280 | 93.4 | 312.7 | 13 | 85 | 0.57 (0.22 - 1.29) | 0.21 | 0.58 (0.24 - 1.40) | 0.23 |
| **PS-stratification / on-treatment TAR without escalation** | | | | | | | | | | |
| CCAE | 19,436 | 45,481 | 10.4 | 29.6 | <5 | 9 | 1.24 (0.24 - 4.80) | 0.78 | 1.29 (0.29 - 5.73) | 0.74 |
| ODOD | 11,652 | 29,479 | 5.6 | 19.7 | <5 | 8 | 2.23 (0.11 - 14.1) | 0.52 | 2.22 (0.20 - 24.8) | 0.52 |
| MDCR | 1,127 | 8,789 | 0.6 | 7.2 | 0 | 6 | | | | |
| MDCD | 2,031 | 7,756 | 0.9 | 4.3 | <5 | <5 | 2.15 (0.09 - 24.1) | 0.59 | 2.24 (0.14 - 37.1) | 0.57 |
| Open Claims | 231,103 | 449,385 | 129.9 | 313.6 | 16 | 87 | 0.64 (0.33 - 1.18) | 0.18 | 0.66 (0.34 - 1.27) | 0.22 |
| OEHR | 13,629 | 38,885 | 3.5 | 16.3 | 0 | <5 | | | | |
| Meta-analysis | 231,103 | 449,385 | 129.9 | 313.6 | 16 | 87 | 0.64 (0.33 - 1.18) | 0.18 | 0.66 (0.35 - 1.25) | 0.20 |

##### 1.4.22 Nausea

**Supplementary Table 100:** Exposures, outcomes and risk estimates for Nausea in GLP1RA (female) (T) vs DPP4I (female) (C) new-user cohorts under four different propensity score (PS) adjustment / time-at-risk (TAR) designs across data sources. We report total patient exposure time in 1,000 patient-year (KYr) units, uncalibrated and calibrated hazard ratios (HRs) and their 95% confidence intervals (CIs). Greyed out entries did not pass study diagnostics and are included only for completeness.

| | Patients | | Exposure (KYr) | | Outcomes | | Uncalibrated | | Calibrated | |
|---|---|---|---|---|---|---|---|---|---|---|
| | T | C | T | C | T | C | HR (95% CI) | $p$ | HR (95% CI) | $p$ |
| **PS-matching / on-treatment TAR** | | | | | | | | | | |
| CCAE | 11,842 | 38,596 | 7.2 | 30.0 | 441 | 1,285 | 1.28 (1.10 - 1.49) | <0.01 | 1.38 (1.18 - 1.62) | <0.01 |
| ODOD | 6,280 | 23,313 | 3.5 | 18.0 | 306 | 972 | 1.60 (1.34 - 1.92) | <0.01 | 1.56 (1.30 - 1.88) | <0.01 |
| MDCR | 882 | 7,237 | 0.5 | 6.8 | 29 | 306 | 0.95 (0.55 - 1.54) | 0.83 | 0.78 (0.47 - 1.30) | 0.35 |
| MDCD | 1,189 | 5,111 | 0.6 | 3.4 | 98 | 403 | 1.15 (0.85 - 1.54) | 0.37 | 1.17 (0.87 - 1.58) | 0.32 |
| Open Claims | 112,939 | 366,334 | 81.8 | 319.0 | 3,158 | 9,386 | 1.40 (1.32 - 1.48) | <0.01 | 1.41 (1.25 - 1.60) | <0.01 |
| OEHR | 8,753 | 33,160 | 2.6 | 16.5 | 226 | 833 | 1.94 (1.59 - 2.37) | <0.01 | 1.87 (1.53 - 2.29) | <0.01 |
| Meta-analysis | 141,003 | 466,514 | 95.8 | 386.9 | 4,229 | 12,879 | 1.46 (1.24 - 1.71) | <0.01 | 1.47 (1.24 - 1.75) | <0.01 |
| Meta-analysis (w/o USOC) | 28,064 | 100,180 | 14.0 | 68.0 | 1,071 | 3,493 | 1.48 (1.19 - 1.84) | <0.01 | 1.50 (1.21 - 1.87) | <0.01 |
| **PS-stratification / on-treatment TAR** | | | | | | | | | | |
| CCAE | 15,622 | 38,599 | 8.9 | 30.0 | 606 | 1,285 | 1.25 (1.11 - 1.40) | <0.01 | 1.30 (1.16 - 1.47) | <0.01 |
| ODOD | 9,065 | 23,886 | 4.7 | 18.5 | 450 | 1,003 | 1.45 (1.26 - 1.67) | <0.01 | 1.45 (1.25 - 1.67) | <0.01 |
| MDCR | 1,005 | 7,552 | 0.6 | 7.0 | 33 | 317 | 0.93 (0.61 - 1.37) | 0.72 | 0.77 (0.52 - 1.16) | 0.23 |
| MDCD | 1,275 | 5,142 | 0.7 | 3.4 | 106 | 404 | 1.18 (0.93 - 1.48) | 0.17 | 1.19 (0.94 - 1.51) | 0.17 |
| Open Claims | 175,888 | 367,383 | 109.5 | 320.0 | 4,651 | 9,435 | 1.35 (1.29 - 1.41) | <0.01 | 1.40 (1.21 - 1.62) | <0.01 |
| OEHR | 11,092 | 33,348 | 3.2 | 16.7 | 290 | 842 | 1.57 (1.34 - 1.83) | <0.01 | 1.47 (1.26 - 1.72) | <0.01 |
| Meta-analysis | 212,942 | 468,358 | 127.0 | 388.6 | 6,103 | 12,969 | 1.36 (1.26 - 1.46) | <0.01 | 1.40 (1.22 - 1.59) | <0.01 |
| Meta-analysis (w/o USOC) | 37,054 | 100,975 | 17.4 | 68.6 | 1,452 | 3,534 | 1.37 (1.21 - 1.54) | <0.01 | 1.39 (1.22 - 1.58) | <0.01 |
| **PS-matching / on-treatment TAR without escalation** | | | | | | | | | | |
| CCAE | 11,842 | 38,596 | 6.5 | 24.9 | 403 | 1,058 | 1.29 (1.10 - 1.51) | <0.01 | 1.38 (1.17 - 1.63) | <0.01 |
| ODOD | 6,280 | 23,313 | 3.2 | 15.2 | 280 | 821 | 1.70 (1.40 - 2.05) | <0.01 | 1.63 (1.34 - 1.98) | <0.01 |
| MDCR | 882 | 7,237 | 0.5 | 5.9 | 27 | 261 | 1.03 (0.58 - 1.74) | 0.92 | 0.82 (0.47 - 1.43) | 0.51 |
| MDCD | 1,189 | 5,111 | 0.6 | 2.8 | 89 | 316 | 1.16 (0.83 - 1.60) | 0.39 | 1.21 (0.87 - 1.69) | 0.30 |
| Open Claims | 112,939 | 366,334 | 72.7 | 255.3 | 2,808 | 7,440 | 1.44 (1.36 - 1.54) | <0.01 | 1.46 (1.36 - 1.56) | <0.01 |
| OEHR | 8,753 | 33,160 | 2.3 | 13.5 | 196 | 630 | 2.13 (1.71 - 2.66) | <0.01 | 2.10 (1.68 - 2.63) | <0.01 |
| Meta-analysis | 141,003 | 466,514 | 85.2 | 311.6 | 3,776 | 10,265 | 1.52 (1.25 - 1.84) | <0.01 | 1.54 (1.25 - 1.89) | <0.01 |
| Meta-analysis (w/o USOC) | 28,064 | 100,180 | 12.5 | 56.3 | 968 | 2,825 | 1.54 (1.19 - 1.99) | <0.01 | 1.58 (1.22 - 2.04) | <0.01 |
| **PS-stratification / on-treatment TAR without escalation** | | | | | | | | | | |
| CCAE | 15,622 | 38,599 | 8.1 | 24.9 | 557 | 1,058 | 1.29 (1.14 - 1.45) | <0.01 | 1.34 (1.18 - 1.52) | <0.01 |
| ODOD | 9,065 | 23,886 | 4.3 | 15.7 | 420 | 846 | 1.54 (1.33 - 1.78) | <0.01 | 1.53 (1.32 - 1.79) | <0.01 |
| MDCR | 1,005 | 7,552 | 0.5 | 6.1 | 30 | 271 | 1.00 (0.64 - 1.50) | 0.99 | 0.81 (0.53 - 1.25) | 0.36 |
| MDCD | 1,275 | 5,142 | 0.6 | 2.8 | 97 | 316 | 1.24 (0.96 - 1.59) | 0.10 | 1.29 (1.00 - 1.66) | 0.07 |
| Open Claims | 175,888 | 367,383 | 99.0 | 256.1 | 4,235 | 7,482 | 1.41 (1.34 - 1.48) | <0.01 | 1.45 (1.24 - 1.70) | <0.01 |
| OEHR | 11,092 | 33,348 | 2.8 | 13.6 | 253 | 638 | 1.71 (1.44 - 2.02) | <0.01 | 1.60 (1.34 - 1.91) | <0.01 |
| Meta-analysis | 212,942 | 468,358 | 114.8 | 313.0 | 5,562 | 10,340 | 1.43 (1.30 - 1.57) | <0.01 | 1.46 (1.29 - 1.65) | <0.01 |
| Meta-analysis (w/o USOC) | 37,054 | 100,975 | 15.7 | 56.9 | 1,327 | 2,858 | 1.44 (1.25 - 1.66) | <0.01 | 1.47 (1.27 - 1.71) | <0.01 |

##### 1.4.23 Peripheral edema

**Supplementary Table 101:** Exposures, outcomes and risk estimates for Peripheral edema in GLP1RA (female) (T) vs DPP4I (female) (C) new-user cohorts under four different propensity score (PS) adjustment / time-at-risk (TAR) designs across data sources. We report total patient exposure time in 1,000 patient-year (KYr) units, uncalibrated and calibrated hazard ratios (HRs) and their 95% confidence intervals (CIs). Greyed out entries did not pass study diagnostics and are included only for completeness.

| | Patients | | Exposure (KYr) | | Outcomes | | Uncalibrated | | Calibrated | |
|---|---|---|---|---|---|---|---|---|---|---|
| | T | C | T | C | T | C | HR (95% CI) | $p$ | HR (95% CI) | $p$ |
| **PS-matching / on-treatment TAR** | | | | | | | | | | |
| CCAE | 10,948 | 35,719 | 6.7 | 27.2 | 331 | 1,396 | 0.91 (0.77 - 1.08) | 0.28 | 0.99 (0.83 - 1.18) | 0.84 |
| ODOD | 5,713 | 20,985 | 3.2 | 15.4 | 189 | 1,413 | 0.73 (0.59 - 0.90) | <0.01 | 0.71 (0.58 - 0.88) | <0.01 |
| MDCR | 710 | 6,031 | 0.4 | 5.4 | 29 | 487 | 0.54 (0.29 - 0.94) | 0.04 | 0.45 (0.25 - 0.80) | <0.01 |
| MDCD | 1,168 | 4,889 | 0.6 | 3.1 | 56 | 369 | 0.76 (0.52 - 1.09) | 0.15 | 0.77 (0.53 - 1.13) | 0.20 |
| Open Claims | 102,964 | 329,300 | 73.9 | 279.2 | 1,937 | 11,266 | 0.86 (0.81 - 0.92) | <0.01 | 0.87 (0.77 - 0.99) | 0.07 |
| OEHR | 8,443 | 31,545 | 2.5 | 15.4 | 126 | 849 | 1.12 (0.88 - 1.43) | 0.35 | 1.08 (0.85 - 1.38) | 0.57 |
| Meta-analysis | 129,236 | 422,438 | 87.0 | 340.2 | 2,639 | 15,293 | 0.87 (0.78 - 0.98) | 0.02 | 0.88 (0.77 - 1.01) | 0.07 |
| Meta-analysis (w/o USOC) | 26,272 | 93,138 | 13.1 | 61.0 | 702 | 4,027 | 0.88 (0.73 - 1.06) | 0.17 | 0.89 (0.74 - 1.08) | 0.25 |
| **PS-stratification / on-treatment TAR** | | | | | | | | | | |
| CCAE | 14,476 | 35,723 | 8.3 | 27.2 | 411 | 1,396 | 0.91 (0.80 - 1.04) | 0.16 | 0.95 (0.84 - 1.08) | 0.52 |
| ODOD | 8,358 | 21,408 | 4.3 | 15.7 | 261 | 1,486 | 0.74 (0.63 - 0.86) | <0.01 | 0.73 (0.62 - 0.86) | <0.01 |
| MDCR | 804 | 6,251 | 0.5 | 5.6 | 32 | 516 | 0.90 (0.60 - 1.31) | 0.60 | 0.75 (0.51 - 1.11) | 0.16 |
| MDCD | 1,281 | 4,930 | 0.7 | 3.1 | 60 | 377 | 0.75 (0.55 - 1.00) | 0.05 | 0.76 (0.56 - 1.02) | 0.08 |
| Open Claims | 160,813 | 329,848 | 99.2 | 279.7 | 2,532 | 11,328 | 0.84 (0.80 - 0.89) | <0.01 | 0.88 (0.75 - 1.02) | 0.11 |
| OEHR | 10,620 | 31,683 | 3.0 | 15.5 | 147 | 858 | 1.02 (0.84 - 1.24) | 0.83 | 0.96 (0.79 - 1.16) | 0.69 |
| Meta-analysis | 195,548 | 423,592 | 115.5 | 341.2 | 3,411 | 15,445 | 0.85 (0.77 - 0.94) | <0.01 | 0.88 (0.76 - 1.01) | 0.07 |
| Meta-analysis (w/o USOC) | 34,735 | 93,744 | 16.3 | 61.4 | 879 | 4,117 | 0.86 (0.74 - 1.00) | 0.04 | 0.87 (0.75 - 1.02) | 0.10 |
| **PS-matching / on-treatment TAR without escalation** | | | | | | | | | | |
| CCAE | 10,948 | 35,719 | 6.0 | 22.6 | 299 | 1,160 | 0.97 (0.81 - 1.16) | 0.73 | 1.04 (0.86 - 1.25) | 0.74 |
| ODOD | 5,713 | 20,985 | 2.9 | 13.0 | 175 | 1,167 | 0.77 (0.62 - 0.96) | 0.02 | 0.74 (0.59 - 0.93) | 0.01 |
| MDCR | 710 | 6,031 | 0.4 | 4.8 | 28 | 425 | 0.55 (0.28 - 0.99) | 0.06 | 0.44 (0.23 - 0.83) | 0.01 |
| MDCD | 1,168 | 4,889 | 0.5 | 2.5 | 45 | 289 | 0.71 (0.46 - 1.06) | 0.11 | 0.74 (0.49 - 1.13) | 0.19 |
| Open Claims | 102,964 | 329,300 | 65.5 | 223.6 | 1,655 | 8,960 | 0.85 (0.79 - 0.92) | <0.01 | 0.86 (0.79 - 0.93) | <0.01 |
| OEHR | 8,443 | 31,545 | 2.2 | 12.6 | 115 | 649 | 1.12 (0.85 - 1.46) | 0.41 | 1.10 (0.84 - 1.45) | 0.52 |
| Meta-analysis | 129,236 | 422,438 | 77.2 | 274.2 | 2,289 | 12,225 | 0.88 (0.79 - 0.98) | 0.02 | 0.89 (0.78 - 1.02) | 0.09 |
| Meta-analysis (w/o USOC) | 26,272 | 93,138 | 11.7 | 50.6 | 634 | 3,265 | 0.90 (0.75 - 1.08) | 0.26 | 0.92 (0.76 - 1.11) | 0.41 |
| **PS-stratification / on-treatment TAR without escalation** | | | | | | | | | | |
| CCAE | 14,476 | 35,723 | 7.6 | 22.6 | 368 | 1,160 | 0.93 (0.81 - 1.06) | 0.29 | 0.97 (0.84 - 1.11) | 0.67 |
| ODOD | 8,358 | 21,408 | 3.9 | 13.3 | 239 | 1,231 | 0.76 (0.64 - 0.90) | <0.01 | 0.76 (0.64 - 0.91) | <0.01 |
| MDCR | 804 | 6,251 | 0.4 | 4.9 | 31 | 449 | 0.95 (0.63 - 1.40) | 0.81 | 0.78 (0.52 - 1.16) | 0.24 |
| MDCD | 1,281 | 4,930 | 0.6 | 2.5 | 49 | 295 | 0.69 (0.49 - 0.95) | 0.03 | 0.72 (0.51 - 1.00) | 0.06 |
| Open Claims | 160,813 | 329,848 | 89.5 | 224.0 | 2,215 | 9,012 | 0.84 (0.79 - 0.89) | <0.01 | 0.86 (0.73 - 1.01) | 0.09 |
| OEHR | 10,620 | 31,683 | 2.6 | 12.6 | 135 | 658 | 1.15 (0.93 - 1.41) | 0.18 | 1.08 (0.87 - 1.33) | 0.57 |
| Meta-analysis | 195,548 | 423,592 | 104.2 | 274.9 | 3,006 | 12,356 | 0.87 (0.75 - 1.01) | 0.06 | 0.89 (0.75 - 1.05) | 0.17 |
| Meta-analysis (w/o USOC) | 34,735 | 93,744 | 14.7 | 51.0 | 791 | 3,344 | 0.88 (0.73 - 1.07) | 0.20 | 0.90 (0.74 - 1.10) | 0.32 |

##### 1.4.24 Photosensitivity

**Supplementary Table 102:** Exposures, outcomes and risk estimates for Photosensitivity in GLP1RA (female) (T) vs DPP4I (female) (C) new-user cohorts under four different propensity score (PS) adjustment / time-at-risk (TAR) designs across data sources. We report total patient exposure time in 1,000 patient-year (KYr) units, uncalibrated and calibrated hazard ratios (HRs) and their 95% confidence intervals (CIs). Greyed out entries did not pass study diagnostics and are included only for completeness.

| | Patients | | Exposure (KYr) | | Outcomes | | Uncalibrated | | Calibrated | |
|---|---|---|---|---|---|---|---|---|---|---|
| | T | C | T | C | T | C | HR (95% CI) | $p$ | HR (95% CI) | $p$ |
| **PS-matching / on-treatment TAR** | | | | | | | | | | |
| CCAE | 14,289 | 45,011 | 9.0 | 35.5 | 13 | 50 | 0.58 (0.16 - 1.68) | 0.36 | 0.62 (0.19 - 2.03) | 0.44 |
| ODOD | 7,863 | 28,515 | 4.6 | 22.5 | 10 | 45 | 2.25 (0.91 - 5.33) | 0.07 | 2.20 (0.91 - 5.31) | 0.08 |
| MDCR | 961 | 8,205 | 0.6 | 7.8 | <5 | 25 | 1.20 (0.06 - 6.78) | 0.88 | 0.99 (0.10 - 10.2) | 0.96 |
| MDCD | 1,850 | 7,629 | 1.0 | 5.4 | <5 | 11 | 1.27 (0.17 - 7.06) | 0.80 | 1.30 (0.20 - 8.45) | 0.78 |
| Open Claims | 143,120 | 443,736 | 104.1 | 389.4 | 39 | 202 | 1.22 (0.74 - 1.94) | 0.42 | 1.23 (0.75 - 2.02) | 0.42 |
| OEHR | 10,498 | 38,415 | 3.2 | 19.9 | 9 | 37 | 1.46 (0.44 - 4.18) | 0.51 | 1.41 (0.46 - 4.33) | 0.55 |
| Meta-analysis | 175,770 | 555,677 | 121.0 | 467.3 | 71 | 334 | 1.29 (0.88 - 1.88) | 0.19 | 1.30 (0.89 - 1.90) | 0.18 |
| Meta-analysis (w/o USOC) | 32,650 | 111,941 | 16.8 | 77.9 | 32 | 132 | 1.33 (0.61 - 2.91) | 0.47 | 1.35 (0.62 - 2.96) | 0.45 |
| **PS-stratification / on-treatment TAR** | | | | | | | | | | |
| CCAE | 19,239 | 45,015 | 11.4 | 35.5 | 15 | 50 | 0.98 (0.48 - 1.89) | 0.94 | 1.02 (0.51 - 2.03) | 0.95 |
| ODOD | 11,575 | 29,190 | 6.1 | 23.2 | 11 | 45 | 1.45 (0.63 - 3.11) | 0.36 | 1.45 (0.65 - 3.22) | 0.37 |
| MDCR | 1,102 | 8,668 | 0.7 | 8.2 | <5 | 25 | 1.04 (0.06 - 5.54) | 0.97 | 0.87 (0.09 - 8.63) | 0.91 |
| MDCD | 2,013 | 7,706 | 1.1 | 5.5 | <5 | 11 | 0.76 (0.11 - 3.09) | 0.75 | 0.77 (0.15 - 4.05) | 0.76 |
| Open Claims | 228,366 | 444,856 | 142.1 | 390.5 | 52 | 204 | 1.34 (0.91 - 1.93) | 0.14 | 1.39 (0.93 - 2.08) | 0.12 |
| OEHR | 13,527 | 38,618 | 4.0 | 20.0 | 9 | 37 | 1.82 (0.79 - 3.79) | 0.13 | 1.71 (0.78 - 3.73) | 0.19 |
| Meta-analysis | 272,707 | 557,679 | 163.6 | 469.2 | 87 | 336 | 1.33 (1.00 - 1.77) | 0.05 | 1.37 (1.01 - 1.86) | 0.04 |
| Meta-analysis (w/o USOC) | 44,341 | 112,823 | 21.5 | 78.7 | 35 | 132 | 1.33 (0.86 - 2.05) | 0.20 | 1.35 (0.87 - 2.09) | 0.18 |
| **PS-matching / on-treatment TAR without escalation** | | | | | | | | | | |
| CCAE | 14,289 | 45,011 | 8.1 | 29.2 | 11 | 45 | 0.60 (0.16 - 1.78) | 0.40 | 0.64 (0.20 - 2.12) | 0.47 |
| ODOD | 7,863 | 28,515 | 4.1 | 18.9 | 10 | 37 | 2.52 (0.97 - 6.24) | 0.05 | 2.42 (0.96 - 6.14) | 0.06 |
| MDCR | 961 | 8,205 | 0.6 | 6.8 | <5 | 23 | 1.28 (0.07 - 7.37) | 0.84 | 1.02 (0.10 - 10.7) | 0.96 |
| MDCD | 1,850 | 7,629 | 0.9 | 4.2 | <5 | 9 | 1.13 (0.15 - 6.23) | 0.90 | 1.18 (0.18 - 7.63) | 0.86 |
| Open Claims | 143,120 | 443,736 | 92.3 | 309.1 | 27 | 175 | 0.85 (0.47 - 1.47) | 0.59 | 0.86 (0.49 - 1.53) | 0.62 |
| OEHR | 10,498 | 38,415 | 2.8 | 16.0 | 6 | 31 | 1.25 (0.26 - 4.36) | 0.76 | 1.23 (0.30 - 4.99) | 0.78 |
| Meta-analysis | 175,770 | 555,677 | 107.2 | 373.2 | 54 | 288 | 1.12 (0.60 - 2.07) | 0.72 | 1.13 (0.61 - 2.10) | 0.69 |
| Meta-analysis (w/o USOC) | 32,650 | 111,941 | 15.0 | 64.1 | 27 | 113 | 1.32 (0.55 - 3.20) | 0.53 | 1.35 (0.56 - 3.28) | 0.50 |
| **PS-stratification / on-treatment TAR without escalation** | | | | | | | | | | |
| CCAE | 19,239 | 45,015 | 10.3 | 29.2 | 13 | 45 | 0.80 (0.37 - 1.65) | 0.57 | 0.84 (0.40 - 1.76) | 0.64 |
| ODOD | 11,575 | 29,190 | 5.6 | 19.4 | 11 | 37 | 1.67 (0.70 - 3.73) | 0.23 | 1.67 (0.72 - 3.85) | 0.23 |
| MDCR | 1,102 | 8,668 | 0.6 | 7.1 | <5 | 23 | 1.12 (0.06 - 6.03) | 0.92 | 0.91 (0.09 - 9.15) | 0.94 |
| MDCD | 2,013 | 7,706 | 0.9 | 4.3 | <5 | 9 | 0.85 (0.12 - 3.67) | 0.85 | 0.89 (0.16 - 4.84) | 0.89 |
| Open Claims | 228,366 | 444,856 | 128.2 | 310.0 | 39 | 176 | 1.01 (0.64 - 1.56) | 0.96 | 1.04 (0.65 - 1.66) | 0.87 |
| OEHR | 13,527 | 38,618 | 3.5 | 16.1 | 6 | 31 | 1.41 (0.50 - 3.36) | 0.48 | 1.32 (0.51 - 3.40) | 0.58 |
| Meta-analysis | 272,707 | 557,679 | 147.5 | 374.8 | 69 | 289 | 1.09 (0.78 - 1.51) | 0.62 | 1.11 (0.79 - 1.55) | 0.55 |
| Meta-analysis (w/o USOC) | 44,341 | 112,823 | 19.3 | 64.7 | 30 | 113 | 1.18 (0.73 - 1.91) | 0.49 | 1.21 (0.75 - 1.95) | 0.44 |

##### 1.4.25 Venous thromboembolism

**Supplementary Table 103:** Exposures, outcomes and risk estimates for Venous thromboembolism in GLP1RA (female) (T) vs DPP4I (female) (C) new-user cohorts under four different propensity score (PS) adjustment / time-at-risk (TAR) designs across data sources. We report total patient exposure time in 1,000 patient-year (KYr) units, uncalibrated and calibrated hazard ratios (HRs) and their 95% confidence intervals (CIs). Greyed out entries did not pass study diagnostics and are included only for completeness.

| | Patients | | Exposure (KYr) | | Outcomes | | Uncalibrated | | Calibrated | |
|---|---|---|---|---|---|---|---|---|---|---|
| | T | C | T | C | T | C | HR (95% CI) | $p$ | HR (95% CI) | $p$ |
| **PS-matching / on-treatment TAR** | | | | | | | | | | |
| CCAE | 14,066 | 44,438 | 8.8 | 35.0 | 40 | 185 | 0.93 (0.56 - 1.50) | 0.77 | 1.01 (0.61 - 1.65) | 0.93 |
| ODOD | 7,637 | 27,903 | 4.5 | 21.9 | 40 | 230 | 0.90 (0.56 - 1.41) | 0.66 | 0.88 (0.55 - 1.39) | 0.59 |
| MDCR | 947 | 8,008 | 0.6 | 7.6 | 6 | 103 | 0.73 (0.24 - 1.88) | 0.55 | 0.60 (0.21 - 1.70) | 0.34 |
| MDCD | 1,771 | 7,342 | 1.0 | 5.2 | 10 | 61 | 1.71 (0.68 - 4.06) | 0.24 | 1.75 (0.71 - 4.27) | 0.23 |
| Open Claims | 139,255 | 432,685 | 101.4 | 379.1 | 397 | 2,005 | 0.97 (0.84 - 1.13) | 0.72 | 0.98 (0.82 - 1.18) | 0.87 |
| OEHR | 10,290 | 37,693 | 3.2 | 19.4 | 25 | 135 | 1.67 (0.92 - 2.92) | 0.08 | 1.61 (0.90 - 2.85) | 0.11 |
| Meta-analysis | 173,019 | 550,061 | 118.9 | 460.6 | 512 | 2,616 | 1.00 (0.88 - 1.14) | 0.96 | 1.01 (0.87 - 1.17) | 0.87 |
| Meta-analysis (w/o USOC) | 33,764 | 117,376 | 17.4 | 81.5 | 115 | 611 | 1.14 (0.82 - 1.59) | 0.43 | 1.16 (0.83 - 1.62) | 0.39 |
| **PS-stratification / on-treatment TAR** | | | | | | | | | | |
| CCAE | 18,912 | 44,442 | 11.1 | 35.0 | 50 | 185 | 0.88 (0.61 - 1.25) | 0.48 | 0.92 (0.64 - 1.31) | 0.64 |
| ODOD | 11,250 | 28,540 | 6.0 | 22.6 | 52 | 240 | 1.02 (0.70 - 1.44) | 0.93 | 1.01 (0.71 - 1.45) | 0.91 |
| MDCR | 1,082 | 8,418 | 0.6 | 7.9 | 6 | 108 | 0.77 (0.28 - 1.75) | 0.57 | 0.64 (0.26 - 1.59) | 0.34 |
| MDCD | 1,938 | 7,411 | 1.0 | 5.2 | 10 | 61 | 1.06 (0.48 - 2.12) | 0.88 | 1.07 (0.51 - 2.25) | 0.85 |
| Open Claims | 221,964 | 433,661 | 138.3 | 380.1 | 508 | 2,031 | 0.97 (0.86 - 1.09) | 0.59 | 1.01 (0.84 - 1.21) | 0.91 |
| OEHR | 13,249 | 37,874 | 3.9 | 19.5 | 30 | 139 | 1.34 (0.84 - 2.06) | 0.21 | 1.25 (0.80 - 1.97) | 0.35 |
| Meta-analysis | 267,313 | 551,928 | 160.3 | 462.4 | 650 | 2,656 | 0.98 (0.89 - 1.09) | 0.73 | 1.01 (0.87 - 1.17) | 0.89 |
| Meta-analysis (w/o USOC) | 45,349 | 118,267 | 22.0 | 82.3 | 142 | 625 | 1.03 (0.83 - 1.27) | 0.79 | 1.05 (0.85 - 1.30) | 0.68 |
| **PS-matching / on-treatment TAR without escalation** | | | | | | | | | | |
| CCAE | 14,066 | 44,438 | 7.9 | 28.8 | 35 | 147 | 0.84 (0.48 - 1.42) | 0.53 | 0.90 (0.52 - 1.54) | 0.71 |
| ODOD | 7,637 | 27,903 | 4.0 | 18.4 | 34 | 203 | 0.92 (0.55 - 1.51) | 0.75 | 0.89 (0.53 - 1.47) | 0.65 |
| MDCR | 947 | 8,008 | 0.6 | 6.6 | <5 | 87 | 0.48 (0.11 - 1.49) | 0.27 | 0.39 (0.10 - 1.42) | 0.16 |
| MDCD | 1,771 | 7,342 | 0.8 | 4.0 | <5 | 44 | 0.76 (0.20 - 2.29) | 0.65 | 0.79 (0.24 - 2.66) | 0.71 |
| Open Claims | 139,255 | 432,685 | 89.9 | 301.2 | 343 | 1,613 | 0.98 (0.83 - 1.15) | 0.81 | 0.99 (0.84 - 1.17) | 0.90 |
| OEHR | 10,290 | 37,693 | 2.7 | 15.7 | 20 | 91 | 1.94 (1.00 - 3.64) | 0.04 | 1.92 (1.01 - 3.65) | 0.05 |
| Meta-analysis | 173,019 | 550,061 | 105.4 | 368.1 | <437 | 2,098 | 0.99 (0.86 - 1.15) | 0.94 | 1.01 (0.85 - 1.19) | 0.90 |
| Meta-analysis (w/o USOC) | 33,764 | 117,376 | 15.5 | 66.9 | <94 | 485 | 1.06 (0.71 - 1.59) | 0.77 | 1.09 (0.72 - 1.63) | 0.69 |
| **PS-stratification / on-treatment TAR without escalation** | | | | | | | | | | |
| CCAE | 18,912 | 44,442 | 10.0 | 28.8 | 45 | 147 | 0.92 (0.62 - 1.34) | 0.66 | 0.95 (0.65 - 1.41) | 0.82 |
| ODOD | 11,250 | 28,540 | 5.4 | 18.9 | 46 | 211 | 0.93 (0.63 - 1.36) | 0.73 | 0.93 (0.63 - 1.37) | 0.73 |
| MDCR | 1,082 | 8,418 | 0.6 | 6.9 | <5 | 89 | 0.50 (0.15 - 1.33) | 0.23 | 0.41 (0.14 - 1.24) | 0.12 |
| MDCD | 1,938 | 7,411 | 0.9 | 4.1 | <5 | 44 | 0.54 (0.15 - 1.49) | 0.29 | 0.56 (0.18 - 1.76) | 0.33 |
| Open Claims | 221,964 | 433,661 | 124.8 | 302.0 | 445 | 1,637 | 0.97 (0.85 - 1.10) | 0.60 | 0.99 (0.81 - 1.21) | 0.91 |
| OEHR | 13,249 | 37,874 | 3.4 | 15.7 | 25 | 95 | 1.48 (0.87 - 2.44) | 0.13 | 1.39 (0.83 - 2.33) | 0.23 |
| Meta-analysis | 267,313 | 551,928 | 144.5 | 369.5 | <566 | 2,134 | 0.97 (0.87 - 1.09) | 0.65 | 0.99 (0.86 - 1.14) | 0.90 |
| Meta-analysis (w/o USOC) | 45,349 | 118,267 | 19.8 | 67.6 | <121 | 497 | 1.00 (0.76 - 1.31) | 0.99 | 1.02 (0.78 - 1.35) | 0.87 |

##### 1.4.26 Vomiting

**Supplementary Table 104:** Exposures, outcomes and risk estimates for Vomiting in GLP1RA (female) (T) vs DPP4I (female) (C) new-user cohorts under four different propensity score (PS) adjustment / time-at-risk (TAR) designs across data sources. We report total patient exposure time in 1,000 patient-year (KYr) units, uncalibrated and calibrated hazard ratios (HRs) and their 95% confidence intervals (CIs). Greyed out entries did not pass study diagnostics and are included only for completeness.

| | Patients | | Exposure (KYr) | | Outcomes | | Uncalibrated | | Calibrated | |
|---|---|---|---|---|---|---|---|---|---|---|
| | T | C | T | C | T | C | HR (95% CI) | $p$ | HR (95% CI) | $p$ |
| **PS-matching / on-treatment TAR** | | | | | | | | | | |
| CCAE | 12,634 | 40,547 | 7.8 | 31.7 | 348 | 924 | 1.43 (1.20 - 1.70) | <0.01 | 1.55 (1.29 - 1.85) | <0.01 |
| ODOD | 6,827 | 25,080 | 3.9 | 19.5 | 218 | 708 | 1.60 (1.28 - 1.99) | <0.01 | 1.56 (1.25 - 1.95) | <0.01 |
| MDCR | 907 | 7,519 | 0.6 | 7.0 | 22 | 230 | 1.26 (0.67 - 2.23) | 0.45 | 1.04 (0.57 - 1.89) | 0.85 |
| MDCD | 1,350 | 5,634 | 0.7 | 3.9 | 80 | 297 | 1.31 (0.94 - 1.82) | 0.11 | 1.34 (0.96 - 1.87) | 0.10 |
| Open Claims | 121,312 | 385,568 | 88.1 | 337.2 | 2,315 | 7,187 | 1.40 (1.31 - 1.50) | <0.01 | 1.42 (1.25 - 1.61) | <0.01 |
| OEHR | 9,413 | 35,152 | 2.9 | 17.8 | 136 | 573 | 1.62 (1.26 - 2.07) | <0.01 | 1.56 (1.22 - 2.00) | <0.01 |
| Meta-analysis | 151,536 | 491,981 | 103.4 | 410.1 | 3,097 | 9,689 | 1.43 (1.35 - 1.51) | <0.01 | 1.44 (1.31 - 1.57) | <0.01 |
| Meta-analysis (w/o USOC) | 30,224 | 106,413 | 15.3 | 72.9 | 782 | 2,502 | 1.50 (1.34 - 1.67) | <0.01 | 1.52 (1.35 - 1.71) | <0.01 |
| **PS-stratification / on-treatment TAR** | | | | | | | | | | |
| CCAE | 16,862 | 40,551 | 9.8 | 31.7 | 469 | 925 | 1.35 (1.19 - 1.54) | <0.01 | 1.41 (1.23 - 1.61) | <0.01 |
| ODOD | 9,989 | 25,529 | 5.2 | 19.9 | 323 | 732 | 1.45 (1.23 - 1.71) | <0.01 | 1.45 (1.22 - 1.71) | <0.01 |
| MDCR | 1,032 | 7,844 | 0.6 | 7.3 | 24 | 245 | 0.92 (0.56 - 1.45) | 0.73 | 0.77 (0.48 - 1.23) | 0.28 |
| MDCD | 1,441 | 5,668 | 0.7 | 3.9 | 87 | 300 | 1.21 (0.93 - 1.57) | 0.15 | 1.23 (0.94 - 1.60) | 0.15 |
| Open Claims | 191,403 | 386,745 | 119.3 | 338.3 | 3,339 | 7,240 | 1.33 (1.27 - 1.40) | <0.01 | 1.39 (1.19 - 1.62) | <0.01 |
| OEHR | 12,003 | 35,368 | 3.5 | 17.9 | 178 | 580 | 1.54 (1.26 - 1.86) | <0.01 | 1.44 (1.18 - 1.75) | <0.01 |
| Meta-analysis | 231,698 | 493,861 | 138.5 | 411.7 | 4,396 | 9,777 | 1.35 (1.29 - 1.41) | <0.01 | 1.39 (1.24 - 1.56) | <0.01 |
| Meta-analysis (w/o USOC) | 40,295 | 107,116 | 19.3 | 73.5 | 1,057 | 2,537 | 1.40 (1.28 - 1.52) | <0.01 | 1.42 (1.30 - 1.56) | <0.01 |
| **PS-matching / on-treatment TAR without escalation** | | | | | | | | | | |
| CCAE | 12,634 | 40,547 | 7.1 | 26.3 | 321 | 760 | 1.52 (1.27 - 1.83) | <0.01 | 1.63 (1.35 - 1.97) | <0.01 |
| ODOD | 6,827 | 25,080 | 3.5 | 16.5 | 196 | 590 | 1.84 (1.46 - 2.33) | <0.01 | 1.77 (1.40 - 2.24) | <0.01 |
| MDCR | 907 | 7,519 | 0.5 | 6.1 | 21 | 191 | 1.34 (0.70 - 2.43) | 0.36 | 1.07 (0.57 - 2.00) | 0.82 |
| MDCD | 1,350 | 5,634 | 0.6 | 3.1 | 71 | 224 | 1.46 (1.00 - 2.10) | 0.04 | 1.53 (1.06 - 2.22) | 0.03 |
| Open Claims | 121,312 | 385,568 | 78.2 | 269.2 | 2,052 | 5,665 | 1.45 (1.34 - 1.56) | <0.01 | 1.46 (1.35 - 1.58) | <0.01 |
| OEHR | 9,413 | 35,152 | 2.5 | 14.4 | 116 | 437 | 1.70 (1.29 - 2.23) | <0.01 | 1.68 (1.27 - 2.21) | <0.01 |
| Meta-analysis | 151,536 | 491,981 | 91.9 | 329.5 | 2,756 | 7,676 | 1.54 (1.40 - 1.70) | <0.01 | 1.56 (1.38 - 1.76) | <0.01 |
| Meta-analysis (w/o USOC) | 30,224 | 106,413 | 13.7 | 60.3 | 704 | 2,011 | 1.63 (1.45 - 1.84) | <0.01 | 1.67 (1.47 - 1.89) | <0.01 |
| **PS-stratification / on-treatment TAR without escalation** | | | | | | | | | | |
| CCAE | 16,862 | 40,551 | 8.9 | 26.3 | 430 | 761 | 1.44 (1.25 - 1.66) | <0.01 | 1.50 (1.30 - 1.73) | <0.01 |
| ODOD | 9,989 | 25,529 | 4.7 | 16.9 | 294 | 609 | 1.53 (1.28 - 1.82) | <0.01 | 1.52 (1.27 - 1.82) | <0.01 |
| MDCR | 1,032 | 7,844 | 0.6 | 6.3 | 22 | 204 | 1.00 (0.60 - 1.61) | 1.00 | 0.81 (0.50 - 1.34) | 0.44 |
| MDCD | 1,441 | 5,668 | 0.6 | 3.1 | 78 | 227 | 1.29 (0.96 - 1.71) | 0.08 | 1.34 (1.01 - 1.79) | 0.06 |
| Open Claims | 191,403 | 386,745 | 107.8 | 270.1 | 3,025 | 5,707 | 1.40 (1.32 - 1.48) | <0.01 | 1.44 (1.23 - 1.69) | <0.01 |
| OEHR | 12,003 | 35,368 | 3.0 | 14.5 | 154 | 442 | 1.64 (1.32 - 2.03) | <0.01 | 1.53 (1.23 - 1.91) | <0.01 |
| Meta-analysis | 231,698 | 493,861 | 125.1 | 330.9 | 3,981 | 7,746 | 1.42 (1.36 - 1.49) | <0.01 | 1.45 (1.32 - 1.60) | <0.01 |
| Meta-analysis (w/o USOC) | 40,295 | 107,116 | 17.4 | 60.8 | 956 | 2,039 | 1.48 (1.35 - 1.62) | <0.01 | 1.51 (1.37 - 1.67) | <0.01 |

#### 1.5 GLP1RA (female) vs SU (female)

##### 1.5.1 3-point MACE

**Supplementary Table 105:** Exposures, outcomes and risk estimates for 3-point MACE in GLP1RA (female) (T) vs SU (female) (C) new-user cohorts under four different propensity score (PS) adjustment / time-at-risk (TAR) designs across data sources. We report total patient exposure time in 1,000 patient-year (KYr) units, uncalibrated and calibrated hazard ratios (HRs) and their 95% confidence intervals (CIs). Greyed out entries did not pass study diagnostics and are included only for completeness.

| | Patients | | Exposure (KYr) | | Outcomes | | Uncalibrated | | Calibrated | |
|---|---|---|---|---|---|---|---|---|---|---|
| | T | C | T | C | T | C | HR (95% CI) | $p$ | HR (95% CI) | $p$ |
| **PS-matching / on-treatment TAR** | | | | | | | | | | |
| CCAE | 15,035 | 68,011 | 9.3 | 50.2 | 45 | 363 | 0.71 (0.45 - 1.07) | 0.12 | 0.73 (0.46 - 1.17) | 0.24 |
| ODOD | 8,688 | 53,222 | 4.9 | 44.8 | 58 | 889 | 1.02 (0.73 - 1.41) | 0.89 | 1.05 (0.75 - 1.49) | 0.80 |
| MDCR | 1,051 | 12,167 | 0.7 | 11.0 | 8 | 300 | 0.37 (0.15 - 0.76) | 0.02 | 0.29 (0.13 - 0.66) | <0.01 |
| MDCD | 1,801 | 14,292 | 1.0 | 9.1 | 9 | 178 | 0.66 (0.26 - 1.44) | 0.34 | 0.63 (0.27 - 1.47) | 0.29 |
| Open Claims | 171,822 | 796,792 | 115.8 | 754.6 | 584 | 8,693 | 0.68 (0.61 - 0.76) | <0.01 | 0.69 (0.57 - 0.83) | <0.01 |
| OEHR | 10,976 | 76,984 | 3.3 | 43.2 | 21 | 483 | 0.83 (0.48 - 1.37) | 0.49 | 0.79 (0.47 - 1.34) | 0.40 |
| Meta-analysis | 209,373 | 1,021,468 | 135.0 | 912.9 | 725 | 10,906 | 0.74 (0.60 - 0.90) | <0.01 | 0.73 (0.56 - 0.96) | 0.03 |
| Meta-analysis (w/o USOC) | 37,551 | 224,676 | 19.2 | 158.3 | 141 | 2,213 | 0.77 (0.57 - 1.03) | 0.08 | 0.75 (0.54 - 1.04) | 0.11 |
| **PS-stratification / on-treatment TAR** | | | | | | | | | | |
| CCAE | 20,219 | 70,165 | 12.0 | 51.5 | 56 | 372 | 0.79 (0.57 - 1.09) | 0.17 | 0.78 (0.55 - 1.12) | 0.21 |
| ODOD | 11,357 | 57,249 | 6.0 | 48.9 | 66 | 1,113 | 0.88 (0.66 - 1.17) | 0.41 | 0.91 (0.68 - 1.21) | 0.53 |
| MDCR | 1,175 | 16,035 | 0.7 | 14.3 | 8 | 454 | 0.48 (0.21 - 0.95) | 0.06 | 0.40 (0.19 - 0.85) | 0.02 |
| MDCD | 1,943 | 15,757 | 1.0 | 10.2 | 9 | 208 | 0.73 (0.33 - 1.42) | 0.39 | 0.70 (0.34 - 1.45) | 0.35 |
| Open Claims | 223,507 | 810,783 | 139.3 | 767.8 | 676 | 9,194 | 0.67 (0.61 - 0.73) | <0.01 | 0.69 (0.57 - 0.83) | <0.01 |
| OEHR | 13,453 | 81,779 | 4.0 | 46.2 | 25 | 586 | 0.84 (0.53 - 1.28) | 0.45 | 0.79 (0.48 - 1.28) | 0.38 |
| Meta-analysis | 268,536 | 1,019,976 | 161.3 | 914.4 | 823 | 11,265 | 0.75 (0.64 - 0.89) | <0.01 | 0.76 (0.59 - 0.98) | 0.04 |
| Meta-analysis (w/o USOC) | 45,029 | 209,193 | 22.0 | 146.6 | 147 | 2,071 | 0.84 (0.69 - 1.02) | 0.09 | 0.83 (0.62 - 1.12) | 0.25 |
| **PS-matching / on-treatment TAR without escalation** | | | | | | | | | | |
| CCAE | 15,035 | 68,011 | 8.4 | 43.1 | 37 | 303 | 0.60 (0.36 - 0.96) | 0.04 | 0.62 (0.38 - 1.02) | 0.08 |
| ODOD | 8,688 | 53,222 | 4.4 | 39.8 | 51 | 784 | 1.04 (0.72 - 1.46) | 0.83 | 1.07 (0.75 - 1.53) | 0.73 |
| MDCR | 1,051 | 12,167 | 0.6 | 10.1 | 8 | 267 | 0.45 (0.18 - 0.95) | 0.06 | 0.35 (0.15 - 0.81) | 0.02 |
| MDCD | 1,801 | 14,292 | 0.8 | 7.5 | 8 | 144 | 0.50 (0.17 - 1.22) | 0.17 | 0.48 (0.18 - 1.30) | 0.15 |
| Open Claims | 171,822 | 796,792 | 103.4 | 634.7 | 507 | 7,373 | 0.63 (0.56 - 0.71) | <0.01 | 0.64 (0.53 - 0.76) | <0.01 |
| OEHR | 10,976 | 76,984 | 2.8 | 36.7 | 16 | 354 | 0.81 (0.44 - 1.39) | 0.46 | 0.75 (0.42 - 1.34) | 0.34 |
| Meta-analysis | 209,373 | 1,021,468 | 120.5 | 771.9 | 627 | 9,225 | 0.70 (0.55 - 0.88) | <0.01 | 0.69 (0.51 - 0.94) | 0.02 |
| Meta-analysis (w/o USOC) | 37,551 | 224,676 | 17.1 | 137.2 | 120 | 1,852 | 0.73 (0.53 - 1.01) | 0.06 | 0.72 (0.49 - 1.04) | 0.09 |
| **PS-stratification / on-treatment TAR without escalation** | | | | | | | | | | |
| CCAE | 20,219 | 70,165 | 10.9 | 44.3 | 45 | 310 | 0.71 (0.49 - 1.01) | 0.07 | 0.71 (0.49 - 1.02) | 0.07 |
| ODOD | 11,357 | 57,249 | 5.5 | 43.6 | 59 | 989 | 0.87 (0.64 - 1.18) | 0.38 | 0.90 (0.66 - 1.22) | 0.50 |
| MDCR | 1,175 | 16,035 | 0.7 | 13.1 | 8 | 407 | 0.52 (0.23 - 1.04) | 0.09 | 0.42 (0.20 - 0.90) | 0.03 |
| MDCD | 1,943 | 15,757 | 0.9 | 8.5 | 8 | 170 | 0.76 (0.32 - 1.55) | 0.49 | 0.75 (0.34 - 1.63) | 0.47 |
| Open Claims | 223,507 | 810,783 | 125.7 | 646.3 | 596 | 7,802 | 0.66 (0.60 - 0.73) | <0.01 | 0.67 (0.56 - 0.80) | <0.01 |
| OEHR | 13,453 | 81,779 | 3.4 | 39.3 | 20 | 426 | 0.85 (0.50 - 1.37) | 0.53 | 0.79 (0.46 - 1.37) | 0.44 |
| Meta-analysis | 268,536 | 1,019,976 | 145.5 | 773.4 | 720 | 9,527 | 0.72 (0.62 - 0.85) | <0.01 | 0.73 (0.57 - 0.92) | <0.01 |
| Meta-analysis (w/o USOC) | 45,029 | 209,193 | 19.8 | 127.1 | 124 | 1,725 | 0.81 (0.65 - 1.00) | 0.05 | 0.81 (0.58 - 1.11) | 0.20 |

##### 1.5.2 4-point MACE

**Supplementary Table 106:** Exposures, outcomes and risk estimates for 4-point MACE in GLP1RA (female) (T) vs SU (female) (C) new-user cohorts under four different propensity score (PS) adjustment / time-at-risk (TAR) designs across data sources. We report total patient exposure time in 1,000 patient-year (KYr) units, uncalibrated and calibrated hazard ratios (HRs) and their 95% confidence intervals (CIs). Greyed out entries did not pass study diagnostics and are included only for completeness.

| | Patients | | Exposure (KYr) | | Outcomes | | Uncalibrated | | Calibrated | |
|---|---|---|---|---|---|---|---|---|---|---|
| | T | C | T | C | T | C | HR (95% CI) | $p$ | HR (95% CI) | $p$ |
| **PS-matching / on-treatment TAR** | | | | | | | | | | |
| CCAE | 14,823 | 67,020 | 9.2 | 49.4 | 57 | 584 | 0.68 (0.47 - 0.96) | 0.03 | 0.71 (0.48 - 1.05) | 0.13 |
| ODOD | 8,427 | 51,180 | 4.7 | 42.6 | 100 | 1,511 | 0.82 (0.62 - 1.05) | 0.13 | 0.84 (0.64 - 1.11) | 0.31 |
| MDCR | 1,011 | 11,143 | 0.7 | 10.0 | 13 | 445 | 0.33 (0.15 - 0.64) | <0.01 | 0.26 (0.13 - 0.54) | <0.01 |
| MDCD | 1,689 | 13,397 | 0.9 | 8.4 | 17 | 308 | 0.65 (0.32 - 1.21) | 0.21 | 0.62 (0.32 - 1.21) | 0.17 |
| Open Claims | 167,622 | 771,419 | 113.0 | 726.5 | 900 | 14,557 | 0.64 (0.59 - 0.70) | <0.01 | 0.65 (0.55 - 0.78) | <0.01 |
| OEHR | 10,817 | 75,708 | 3.3 | 42.0 | 36 | 916 | 0.77 (0.51 - 1.12) | 0.18 | 0.73 (0.49 - 1.08) | 0.13 |
| Meta-analysis | 204,389 | 989,867 | 131.8 | 878.9 | 1,123 | 18,321 | 0.67 (0.60 - 0.75) | <0.01 | 0.67 (0.54 - 0.83) | <0.01 |
| Meta-analysis (w/o USOC) | 36,767 | 218,448 | 18.8 | 152.4 | 223 | 3,764 | 0.70 (0.56 - 0.87) | <0.01 | 0.68 (0.52 - 0.89) | <0.01 |
| **PS-stratification / on-treatment TAR** | | | | | | | | | | |
| CCAE | 19,944 | 69,057 | 11.8 | 50.6 | 70 | 599 | 0.64 (0.48 - 0.85) | <0.01 | 0.64 (0.46 - 0.87) | <0.01 |
| ODOD | 11,042 | 54,541 | 5.8 | 46.2 | 109 | 1,805 | 0.79 (0.63 - 0.98) | 0.04 | 0.81 (0.65 - 1.01) | 0.08 |
| MDCR | 1,125 | 14,786 | 0.7 | 13.1 | 13 | 731 | 0.42 (0.22 - 0.72) | <0.01 | 0.35 (0.19 - 0.63) | <0.01 |
| MDCD | 1,819 | 14,676 | 1.0 | 9.4 | 18 | 355 | 0.74 (0.43 - 1.21) | 0.25 | 0.71 (0.42 - 1.20) | 0.22 |
| Open Claims | 218,502 | 783,706 | 136.1 | 738.0 | 1,041 | 15,361 | 0.61 (0.57 - 0.65) | <0.01 | 0.62 (0.52 - 0.75) | <0.01 |
| OEHR | 13,258 | 80,292 | 3.9 | 44.8 | 41 | 1,088 | 0.69 (0.49 - 0.96) | 0.04 | 0.65 (0.44 - 0.96) | 0.06 |
| Meta-analysis | 262,746 | 987,596 | 157.7 | 879.5 | 1,261 | 18,853 | 0.66 (0.58 - 0.76) | <0.01 | 0.67 (0.53 - 0.85) | <0.01 |
| Meta-analysis (w/o USOC) | 44,244 | 203,890 | 21.6 | 141.5 | 220 | 3,492 | 0.72 (0.62 - 0.84) | <0.01 | 0.71 (0.54 - 0.94) | 0.02 |
| **PS-matching / on-treatment TAR without escalation** | | | | | | | | | | |
| CCAE | 14,823 | 67,020 | 8.3 | 42.4 | 47 | 503 | 0.65 (0.44 - 0.95) | 0.03 | 0.67 (0.45 - 1.00) | 0.08 |
| ODOD | 8,427 | 51,180 | 4.3 | 37.9 | 89 | 1,325 | 0.88 (0.66 - 1.16) | 0.38 | 0.91 (0.69 - 1.21) | 0.55 |
| MDCR | 1,011 | 11,143 | 0.6 | 9.1 | 13 | 394 | 0.40 (0.18 - 0.78) | 0.01 | 0.31 (0.15 - 0.65) | <0.01 |
| MDCD | 1,689 | 13,397 | 0.8 | 7.0 | 15 | 248 | 0.68 (0.33 - 1.27) | 0.26 | 0.65 (0.33 - 1.27) | 0.22 |
| Open Claims | 167,622 | 771,419 | 100.9 | 611.6 | 772 | 12,285 | 0.61 (0.55 - 0.67) | <0.01 | 0.61 (0.52 - 0.72) | <0.01 |
| OEHR | 10,817 | 75,708 | 2.8 | 35.9 | 28 | 689 | 0.79 (0.51 - 1.20) | 0.29 | 0.74 (0.48 - 1.14) | 0.19 |
| Meta-analysis | 204,389 | 989,867 | 117.7 | 743.9 | 964 | 15,444 | 0.68 (0.56 - 0.81) | <0.01 | 0.68 (0.52 - 0.87) | <0.01 |
| Meta-analysis (w/o USOC) | 36,767 | 218,448 | 16.7 | 132.4 | 192 | 3,159 | 0.73 (0.59 - 0.91) | <0.01 | 0.72 (0.54 - 0.96) | 0.04 |
| **PS-stratification / on-treatment TAR without escalation** | | | | | | | | | | |
| CCAE | 19,944 | 69,057 | 10.7 | 43.5 | 56 | 513 | 0.57 (0.42 - 0.78) | <0.01 | 0.57 (0.42 - 0.78) | <0.01 |
| ODOD | 11,042 | 54,541 | 5.3 | 41.1 | 97 | 1,590 | 0.78 (0.61 - 0.98) | 0.04 | 0.80 (0.63 - 1.02) | 0.08 |
| MDCR | 1,125 | 14,786 | 0.7 | 12.0 | 13 | 659 | 0.47 (0.25 - 0.81) | 0.01 | 0.38 (0.21 - 0.69) | <0.01 |
| MDCD | 1,819 | 14,676 | 0.8 | 7.8 | 15 | 289 | 0.77 (0.42 - 1.32) | 0.37 | 0.76 (0.43 - 1.35) | 0.36 |
| Open Claims | 218,502 | 783,706 | 123.0 | 621.7 | 905 | 12,981 | 0.60 (0.55 - 0.64) | <0.01 | 0.60 (0.51 - 0.72) | <0.01 |
| OEHR | 13,258 | 80,292 | 3.4 | 38.3 | 33 | 827 | 0.71 (0.48 - 1.02) | 0.08 | 0.66 (0.43 - 1.02) | 0.09 |
| Meta-analysis | 262,746 | 987,596 | 142.4 | 744.7 | 1,091 | 15,911 | 0.64 (0.55 - 0.74) | <0.01 | 0.65 (0.51 - 0.82) | <0.01 |
| Meta-analysis (w/o USOC) | 44,244 | 203,890 | 19.4 | 123.0 | 186 | 2,930 | 0.69 (0.58 - 0.84) | <0.01 | 0.69 (0.51 - 0.94) | 0.02 |

##### 1.5.3 Acute myocardial infarction

**Supplementary Table 107:** Exposures, outcomes and risk estimates for Acute myocardial infarction in GLP1RA (female) (T) vs SU (female) (C) new-user cohorts under four different propensity score (PS) adjustment / time-at-risk (TAR) designs across data sources. We report total patient exposure time in 1,000 patient-year (KYr) units, uncalibrated and calibrated hazard ratios (HRs) and their 95% confidence intervals (CIs). Greyed out entries did not pass study diagnostics and are included only for completeness.

| | Patients | | Exposure (KYr) | | Outcomes | | Uncalibrated | | Calibrated | |
|---|---|---|---|---|---|---|---|---|---|---|
| | T | C | T | C | T | C | HR (95% CI) | $p$ | HR (95% CI) | $p$ |
| **PS-matching / on-treatment TAR** | | | | | | | | | | |
| CCAE | 15,204 | 68,858 | 9.4 | 51.0 | 22 | 160 | 1.10 (0.58 - 2.00) | 0.75 | 1.15 (0.61 - 2.19) | 0.68 |
| ODOD | 8,837 | 54,488 | 5.0 | 46.2 | 28 | 385 | 1.19 (0.71 - 1.92) | 0.50 | 1.22 (0.73 - 2.04) | 0.47 |
| MDCR | 1,084 | 13,225 | 0.7 | 12.1 | <5 | 124 | 0.54 (0.15 - 1.43) | 0.27 | 0.42 (0.14 - 1.30) | 0.14 |
| MDCD | 1,847 | 14,776 | 1.0 | 9.5 | <5 | 67 | 1.31 (0.34 - 4.00) | 0.67 | 1.24 (0.36 - 4.25) | 0.73 |
| Open Claims | 175,427 | 818,268 | 118.2 | 777.2 | 251 | 3,750 | 0.70 (0.59 - 0.82) | <0.01 | 0.71 (0.57 - 0.89) | <0.01 |
| OEHR | 11,063 | 77,588 | 3.4 | 43.7 | 12 | 234 | 0.86 (0.41 - 1.62) | 0.66 | 0.82 (0.41 - 1.62) | 0.56 |
| Meta-analysis | 213,462 | 1,047,203 | 137.7 | 939.5 | <323 | 4,720 | 0.86 (0.65 - 1.13) | 0.28 | 0.86 (0.62 - 1.20) | 0.37 |
| Meta-analysis (w/o USOC) | 38,035 | 228,935 | 19.5 | 162.3 | <72 | 970 | 1.03 (0.75 - 1.41) | 0.86 | 1.01 (0.71 - 1.43) | 0.92 |
| **PS-stratification / on-treatment TAR** | | | | | | | | | | |
| CCAE | 20,444 | 71,108 | 12.1 | 52.3 | 25 | 164 | 0.93 (0.56 - 1.49) | 0.77 | 0.91 (0.55 - 1.53) | 0.74 |
| ODOD | 11,517 | 59,011 | 6.1 | 50.9 | 30 | 481 | 0.98 (0.63 - 1.48) | 0.93 | 1.01 (0.66 - 1.54) | 0.94 |
| MDCR | 1,213 | 16,841 | 0.8 | 15.1 | <5 | 183 | 0.57 (0.17 - 1.45) | 0.30 | 0.47 (0.16 - 1.40) | 0.18 |
| MDCD | 1,994 | 16,407 | 1.1 | 10.7 | <5 | 83 | 0.94 (0.27 - 2.50) | 0.91 | 0.91 (0.30 - 2.77) | 0.87 |
| Open Claims | 227,850 | 833,873 | 142.0 | 792.0 | 281 | 3,979 | 0.71 (0.61 - 0.81) | <0.01 | 0.72 (0.58 - 0.89) | <0.01 |
| OEHR | 13,547 | 82,582 | 4.0 | 46.8 | 15 | 277 | 1.04 (0.56 - 1.79) | 0.90 | 0.97 (0.52 - 1.79) | 0.91 |
| Meta-analysis | 273,358 | 1,046,574 | 164.3 | 942.0 | 351 | 4,901 | 0.83 (0.66 - 1.04) | 0.10 | 0.84 (0.62 - 1.13) | 0.25 |
| Meta-analysis (w/o USOC) | 45,508 | 212,701 | 22.3 | 150.0 | 70 | 922 | 0.98 (0.74 - 1.29) | 0.87 | 0.97 (0.67 - 1.39) | 0.86 |
| **PS-matching / on-treatment TAR without escalation** | | | | | | | | | | |
| CCAE | 15,204 | 68,858 | 8.5 | 43.7 | 19 | 136 | 1.07 (0.54 - 2.00) | 0.84 | 1.11 (0.57 - 2.14) | 0.78 |
| ODOD | 8,837 | 54,488 | 4.5 | 41.0 | 24 | 335 | 1.18 (0.67 - 1.97) | 0.54 | 1.22 (0.71 - 2.09) | 0.49 |
| MDCR | 1,084 | 13,225 | 0.6 | 11.0 | <5 | 109 | 0.68 (0.19 - 1.89) | 0.51 | 0.54 (0.17 - 1.69) | 0.30 |
| MDCD | 1,847 | 14,776 | 0.9 | 7.8 | <5 | 51 | 1.53 (0.39 - 5.01) | 0.51 | 1.47 (0.41 - 5.28) | 0.56 |
| Open Claims | 175,427 | 818,268 | 105.5 | 653.4 | 223 | 3,192 | 0.70 (0.59 - 0.84) | <0.01 | 0.71 (0.56 - 0.89) | <0.01 |
| OEHR | 11,063 | 77,588 | 2.9 | 37.0 | 9 | 166 | 0.82 (0.36 - 1.67) | 0.62 | 0.76 (0.36 - 1.64) | 0.50 |
| Meta-analysis | 213,462 | 1,047,203 | 122.8 | 794.0 | <285 | 3,989 | 0.86 (0.65 - 1.14) | 0.29 | 0.86 (0.62 - 1.20) | 0.37 |
| Meta-analysis (w/o USOC) | 38,035 | 228,935 | 17.4 | 140.6 | <62 | 797 | 1.04 (0.75 - 1.46) | 0.81 | 1.02 (0.69 - 1.50) | 0.91 |
| **PS-stratification / on-treatment TAR without escalation** | | | | | | | | | | |
| CCAE | 20,444 | 71,108 | 11.0 | 44.9 | 22 | 139 | 0.88 (0.51 - 1.47) | 0.64 | 0.88 (0.52 - 1.49) | 0.64 |
| ODOD | 11,517 | 59,011 | 5.6 | 45.3 | 26 | 423 | 0.94 (0.58 - 1.45) | 0.78 | 0.96 (0.61 - 1.52) | 0.87 |
| MDCR | 1,213 | 16,841 | 0.7 | 13.8 | <5 | 163 | 0.68 (0.20 - 1.77) | 0.49 | 0.55 (0.19 - 1.65) | 0.29 |
| MDCD | 1,994 | 16,407 | 0.9 | 8.9 | <5 | 66 | 1.27 (0.35 - 3.50) | 0.69 | 1.24 (0.39 - 3.94) | 0.71 |
| Open Claims | 227,850 | 833,873 | 128.1 | 666.3 | 251 | 3,391 | 0.72 (0.62 - 0.83) | <0.01 | 0.73 (0.59 - 0.90) | <0.01 |
| OEHR | 13,547 | 82,582 | 3.5 | 39.8 | 12 | 201 | 1.11 (0.55 - 2.04) | 0.77 | 1.03 (0.52 - 2.06) | 0.92 |
| Meta-analysis | 273,358 | 1,046,574 | 148.2 | 796.3 | 311 | 4,154 | 0.80 (0.65 - 0.98) | 0.03 | 0.80 (0.61 - 1.05) | 0.11 |
| Meta-analysis (w/o USOC) | 45,508 | 212,701 | 20.1 | 130.0 | 60 | 763 | 0.95 (0.70 - 1.29) | 0.75 | 0.95 (0.64 - 1.40) | 0.79 |

##### 1.5.4 Acute renal failure

**Supplementary Table 108:** Exposures, outcomes and risk estimates for Acute renal failure in GLP1RA (female) (T) vs SU (female) (C) new-user cohorts under four different propensity score (PS) adjustment / time-at-risk (TAR) designs across data sources. We report total patient exposure time in 1,000 patient-year (KYr) units, uncalibrated and calibrated hazard ratios (HRs) and their 95% confidence intervals (CIs). Greyed out entries did not pass study diagnostics and are included only for completeness.

| | Patients | | Exposure (KYr) | | Outcomes | | Uncalibrated | | Calibrated | |
|---|---|---|---|---|---|---|---|---|---|---|
| | T | C | T | C | T | C | HR (95% CI) | $p$ | HR (95% CI) | $p$ |
| **PS-matching / on-treatment TAR** | | | | | | | | | | |
| CCAE | 15,192 | 68,720 | 9.4 | 50.8 | 44 | 421 | 0.66 (0.43 - 0.99) | 0.05 | 0.69 (0.44 - 1.08) | 0.14 |
| ODOD | 8,692 | 53,334 | 4.9 | 44.7 | 65 | 1,305 | 0.62 (0.45 - 0.84) | <0.01 | 0.64 (0.46 - 0.89) | 0.02 |
| MDCR | 1,061 | 12,944 | 0.7 | 11.7 | 10 | 339 | 0.57 (0.25 - 1.17) | 0.16 | 0.45 (0.21 - 0.99) | 0.05 |
| MDCD | 1,797 | 14,412 | 1.0 | 9.1 | 16 | 268 | 0.69 (0.33 - 1.32) | 0.29 | 0.65 (0.33 - 1.31) | 0.24 |
| Open Claims | 173,476 | 805,988 | 116.6 | 761.0 | 802 | 11,625 | 0.73 (0.67 - 0.80) | <0.01 | 0.74 (0.62 - 0.89) | <0.01 |
| OEHR | 11,015 | 77,132 | 3.3 | 43.1 | 41 | 629 | 1.25 (0.84 - 1.82) | 0.25 | 1.19 (0.81 - 1.75) | 0.38 |
| Meta-analysis | 211,233 | 1,032,530 | 135.9 | 920.4 | 978 | 14,587 | 0.75 (0.60 - 0.93) | <0.01 | 0.75 (0.56 - 0.99) | 0.05 |
| Meta-analysis (w/o USOC) | 37,757 | 226,542 | 19.3 | 159.4 | 176 | 2,962 | 0.75 (0.55 - 1.03) | 0.08 | 0.74 (0.52 - 1.05) | 0.10 |
| **PS-stratification / on-treatment TAR** | | | | | | | | | | |
| CCAE | 20,403 | 70,859 | 12.1 | 52.0 | 60 | 434 | 0.61 (0.44 - 0.83) | <0.01 | 0.60 (0.42 - 0.85) | <0.01 |
| ODOD | 11,352 | 57,109 | 6.0 | 48.7 | 79 | 1,557 | 0.61 (0.47 - 0.78) | <0.01 | 0.62 (0.48 - 0.81) | <0.01 |
| MDCR | 1,192 | 16,436 | 0.7 | 14.6 | 11 | 489 | 0.65 (0.32 - 1.19) | 0.20 | 0.54 (0.28 - 1.04) | 0.07 |
| MDCD | 1,939 | 15,879 | 1.0 | 10.2 | 16 | 323 | 0.52 (0.29 - 0.87) | 0.02 | 0.50 (0.29 - 0.86) | 0.02 |
| Open Claims | 225,549 | 821,391 | 140.2 | 775.7 | 937 | 12,428 | 0.65 (0.60 - 0.70) | <0.01 | 0.66 (0.55 - 0.80) | <0.01 |
| OEHR | 13,486 | 82,060 | 4.0 | 46.2 | 45 | 763 | 1.09 (0.77 - 1.50) | 0.61 | 1.02 (0.69 - 1.50) | 0.89 |
| Meta-analysis | 270,790 | 1,031,419 | 162.3 | 922.6 | 1,121 | 15,182 | 0.70 (0.55 - 0.89) | <0.01 | 0.71 (0.52 - 0.97) | 0.03 |
| Meta-analysis (w/o USOC) | 45,241 | 210,028 | 22.1 | 146.9 | 184 | 2,754 | 0.73 (0.51 - 1.05) | 0.09 | 0.72 (0.47 - 1.11) | 0.15 |
| **PS-matching / on-treatment TAR without escalation** | | | | | | | | | | |
| CCAE | 15,192 | 68,720 | 8.5 | 43.6 | 40 | 340 | 0.62 (0.39 - 0.95) | 0.04 | 0.64 (0.41 - 1.01) | 0.08 |
| ODOD | 8,692 | 53,334 | 4.4 | 39.7 | 55 | 1,128 | 0.61 (0.43 - 0.86) | <0.01 | 0.63 (0.45 - 0.90) | 0.01 |
| MDCR | 1,061 | 12,944 | 0.6 | 10.7 | 9 | 301 | 0.56 (0.24 - 1.15) | 0.15 | 0.44 (0.20 - 0.97) | 0.05 |
| MDCD | 1,797 | 14,412 | 0.8 | 7.6 | 14 | 208 | 0.70 (0.32 - 1.39) | 0.34 | 0.67 (0.32 - 1.39) | 0.29 |
| Open Claims | 173,476 | 805,988 | 104.1 | 640.4 | 684 | 9,683 | 0.73 (0.66 - 0.80) | <0.01 | 0.73 (0.61 - 0.87) | <0.01 |
| OEHR | 11,015 | 77,132 | 2.9 | 36.7 | 26 | 430 | 1.22 (0.75 - 1.91) | 0.41 | 1.13 (0.71 - 1.81) | 0.61 |
| Meta-analysis | 211,233 | 1,032,530 | 121.3 | 778.8 | 828 | 12,090 | 0.72 (0.66 - 0.79) | <0.01 | 0.72 (0.59 - 0.89) | <0.01 |
| Meta-analysis (w/o USOC) | 37,757 | 226,542 | 17.2 | 138.4 | 144 | 2,407 | 0.72 (0.54 - 0.97) | 0.03 | 0.71 (0.50 - 1.00) | 0.06 |
| **PS-stratification / on-treatment TAR without escalation** | | | | | | | | | | |
| CCAE | 20,403 | 70,859 | 11.0 | 44.8 | 54 | 351 | 0.68 (0.48 - 0.95) | 0.03 | 0.68 (0.48 - 0.96) | 0.03 |
| ODOD | 11,352 | 57,109 | 5.5 | 43.4 | 69 | 1,359 | 0.61 (0.46 - 0.79) | <0.01 | 0.62 (0.47 - 0.82) | <0.01 |
| MDCR | 1,192 | 16,436 | 0.7 | 13.4 | 10 | 430 | 0.68 (0.32 - 1.27) | 0.26 | 0.55 (0.28 - 1.09) | 0.09 |
| MDCD | 1,939 | 15,879 | 0.9 | 8.5 | 14 | 256 | 0.54 (0.29 - 0.95) | 0.04 | 0.53 (0.29 - 0.97) | 0.04 |
| Open Claims | 225,549 | 821,391 | 126.5 | 653.4 | 810 | 10,377 | 0.64 (0.59 - 0.69) | <0.01 | 0.65 (0.54 - 0.77) | <0.01 |
| OEHR | 13,486 | 82,060 | 3.5 | 39.4 | 30 | 527 | 0.92 (0.60 - 1.36) | 0.70 | 0.86 (0.54 - 1.36) | 0.56 |
| Meta-analysis | 270,790 | 1,031,419 | 146.4 | 781.0 | 963 | 12,614 | 0.65 (0.60 - 0.70) | <0.01 | 0.65 (0.54 - 0.79) | <0.01 |
| Meta-analysis (w/o USOC) | 45,241 | 210,028 | 19.9 | 127.6 | 153 | 2,237 | 0.70 (0.56 - 0.88) | <0.01 | 0.69 (0.50 - 0.97) | 0.04 |

##### 1.5.5 Glycemic control

**Supplementary Table 109:** Exposures, outcomes and risk estimates for Glycemic control in GLP1RA (female) (T) vs SU (female) (C) new-user cohorts under four different propensity score (PS) adjustment / time-at-risk (TAR) designs across data sources. We report total patient exposure time in 1,000 patient-year (KYr) units, uncalibrated and calibrated hazard ratios (HRs) and their 95% confidence intervals (CIs). Greyed out entries did not pass study diagnostics and are included only for completeness.

| | Patients | | Exposure (KYr) | | Outcomes | | Uncalibrated | | Calibrated | |
|---|---|---|---|---|---|---|---|---|---|---|
| | T | C | T | C | T | C | HR (95% CI) | $p$ | HR (95% CI) | $p$ |
| **PS-matching / on-treatment TAR** | | | | | | | | | | |
| CCAE | 15,234 | 68,978 | 9.5 | 51.0 | 20 | 285 | 0.99 (0.53 - 1.76) | 0.98 | 1.04 (0.56 - 1.93) | 0.91 |
| ODOD | 5,847 | 38,821 | 3.1 | 29.5 | 336 | 2,844 | 1.34 (1.14 - 1.56) | <0.01 | 1.38 (1.15 - 1.65) | 0.01 |
| MDCR | 1,094 | 13,448 | 0.7 | 12.3 | 0 | 41 | | | | |
| MDCD | 1,887 | 15,102 | 1.0 | 9.7 | 0 | 0 | | | | |
| Open Claims | 178,376 | 835,401 | 120.1 | 795.9 | 0 | 0 | | | | |
| OEHR | 11,145 | 78,232 | 3.4 | 44.2 | 0 | 0 | | | | |
| Meta-analysis | 21,081 | 107,799 | 12.5 | 80.5 | 356 | 3,129 | 1.31 (1.13 - 1.52) | <0.01 | 1.31 (1.04 - 1.66) | 0.02 |
| Meta-analysis (w/o USOC) | 21,081 | 107,799 | 12.5 | 80.5 | 356 | 3,129 | 1.31 (1.13 - 1.52) | <0.01 | 1.28 (1.04 - 1.59) | 0.04 |
| **PS-stratification / on-treatment TAR** | | | | | | | | | | |
| CCAE | 20,459 | 70,856 | 12.1 | 51.9 | 24 | 393 | 0.96 (0.58 - 1.51) | 0.86 | 0.94 (0.57 - 1.56) | 0.83 |
| ODOD | 7,237 | 42,368 | 3.6 | 32.7 | 400 | 3,230 | 1.25 (1.10 - 1.41) | <0.01 | 1.28 (1.13 - 1.46) | <0.01 |
| MDCR | 1,227 | 17,202 | 0.8 | 15.5 | 0 | 101 | | | | |
| MDCD | 2,034 | 16,863 | 1.1 | 11.0 | 0 | 0 | | | | |
| Open Claims | 231,281 | 852,348 | 144.1 | 811.8 | 0 | 0 | | | | |
| OEHR | 13,639 | 83,370 | 4.0 | 47.5 | 0 | 0 | | | | |
| Meta-analysis | 27,696 | 113,224 | 15.8 | 84.6 | 424 | 3,623 | 1.22 (1.03 - 1.43) | 0.02 | 1.23 (0.95 - 1.59) | 0.11 |
| Meta-analysis (w/o USOC) | 27,696 | 113,224 | 15.8 | 84.6 | 424 | 3,623 | 1.22 (1.03 - 1.43) | 0.02 | 1.20 (0.91 - 1.59) | 0.21 |
| **PS-matching / on-treatment TAR without escalation** | | | | | | | | | | |
| CCAE | 15,234 | 68,978 | 8.5 | 43.6 | 20 | 254 | 1.13 (0.60 - 2.03) | 0.69 | 1.17 (0.63 - 2.16) | 0.64 |
| ODOD | 5,847 | 38,821 | 2.8 | 26.2 | 305 | 2,559 | 1.29 (1.09 - 1.51) | <0.01 | 1.33 (1.12 - 1.57) | <0.01 |
| MDCR | 1,094 | 13,448 | 0.7 | 11.2 | 0 | 37 | | | | |
| MDCD | 1,887 | 15,102 | 0.9 | 8.1 | 0 | 0 | | | | |
| Open Claims | 178,376 | 835,401 | 107.1 | 668.8 | 0 | 0 | | | | |
| OEHR | 11,145 | 78,232 | 2.9 | 37.4 | 0 | 0 | | | | |
| Meta-analysis | 21,081 | 107,799 | 11.3 | 69.9 | 325 | 2,813 | 1.28 (1.09 - 1.49) | <0.01 | 1.27 (1.00 - 1.62) | 0.05 |
| Meta-analysis (w/o USOC) | 21,081 | 107,799 | 11.3 | 69.9 | 325 | 2,813 | 1.28 (1.09 - 1.49) | <0.01 | 1.25 (0.97 - 1.60) | 0.10 |
| **PS-stratification / on-treatment TAR without escalation** | | | | | | | | | | |
| CCAE | 20,459 | 70,856 | 11.0 | 44.5 | 24 | 358 | 1.06 (0.64 - 1.68) | 0.81 | 1.06 (0.65 - 1.71) | 0.83 |
| ODOD | 7,237 | 42,368 | 3.3 | 29.2 | 365 | 2,923 | 1.27 (1.12 - 1.45) | <0.01 | 1.31 (1.15 - 1.50) | <0.01 |
| MDCR | 1,227 | 17,202 | 0.7 | 14.1 | 0 | 92 | | | | |
| MDCD | 2,034 | 16,863 | 0.9 | 9.2 | 0 | 0 | | | | |
| Open Claims | 231,281 | 852,348 | 130.0 | 682.7 | 0 | 0 | | | | |
| OEHR | 13,639 | 83,370 | 3.5 | 40.3 | 0 | 0 | | | | |
| Meta-analysis | 27,696 | 113,224 | 14.3 | 73.7 | 389 | 3,281 | 1.26 (1.11 - 1.43) | <0.01 | 1.27 (1.02 - 1.58) | 0.03 |
| Meta-analysis (w/o USOC) | 27,696 | 113,224 | 14.3 | 73.7 | 389 | 3,281 | 1.26 (1.11 - 1.43) | <0.01 | 1.25 (0.95 - 1.65) | 0.12 |

##### 1.5.6 Hospitalization with heart failure

**Supplementary Table 110:** Exposures, outcomes and risk estimates for Hospitalization with heart failure in GLP1RA (female) (T) vs SU (female) (C) new-user cohorts under four different propensity score (PS) adjustment / time-at-risk (TAR) designs across data sources. We report total patient exposure time in 1,000 patient-year (KYr) units, uncalibrated and calibrated hazard ratios (HRs) and their 95% confidence intervals (CIs). Greyed out entries did not pass study diagnostics and are included only for completeness.

| | Patients | | Exposure (KYr) | | Outcomes | | Uncalibrated | | Calibrated | |
|---|---|---|---|---|---|---|---|---|---|---|
| | T | C | T | C | T | C | HR (95% CI) | $p$ | HR (95% CI) | $p$ |
| **PS-matching / on-treatment TAR** | | | | | | | | | | |
| CCAE | 15,081 | 68,367 | 9.4 | 50.6 | 17 | 321 | 0.43 (0.22 - 0.77) | <0.01 | 0.45 (0.23 - 0.86) | 0.02 |
| ODOD | 8,619 | 52,820 | 4.8 | 44.4 | 63 | 1,049 | 0.78 (0.56 - 1.07) | 0.13 | 0.80 (0.57 - 1.12) | 0.26 |
| MDCR | 1,050 | 12,178 | 0.7 | 11.0 | 10 | 333 | 0.44 (0.19 - 0.90) | 0.04 | 0.35 (0.16 - 0.77) | <0.01 |
| MDCD | 1,745 | 13,943 | 0.9 | 8.8 | 15 | 215 | 0.79 (0.38 - 1.49) | 0.50 | 0.75 (0.38 - 1.49) | 0.43 |
| Open Claims | 172,903 | 801,531 | 116.4 | 758.1 | 512 | 9,369 | 0.65 (0.58 - 0.72) | <0.01 | 0.66 (0.54 - 0.79) | <0.01 |
| OEHR | 10,956 | 76,773 | 3.3 | 42.8 | 23 | 618 | 0.74 (0.44 - 1.18) | 0.23 | 0.71 (0.43 - 1.15) | 0.17 |
| Meta-analysis | 210,354 | 1,025,612 | 135.6 | 915.7 | 640 | 11,905 | 0.66 (0.59 - 0.72) | <0.01 | 0.65 (0.53 - 0.81) | <0.01 |
| Meta-analysis (w/o USOC) | 37,451 | 224,081 | 19.2 | 157.5 | 128 | 2,536 | 0.68 (0.54 - 0.86) | <0.01 | 0.66 (0.50 - 0.88) | <0.01 |
| **PS-stratification / on-treatment TAR** | | | | | | | | | | |
| CCAE | 20,267 | 70,548 | 12.1 | 51.8 | 21 | 330 | 0.41 (0.25 - 0.65) | <0.01 | 0.41 (0.25 - 0.67) | <0.01 |
| ODOD | 11,260 | 56,641 | 6.0 | 48.4 | 66 | 1,270 | 0.71 (0.53 - 0.93) | 0.02 | 0.73 (0.55 - 0.97) | 0.03 |
| MDCR | 1,169 | 15,734 | 0.7 | 14.0 | 10 | 530 | 0.45 (0.22 - 0.83) | 0.02 | 0.37 (0.19 - 0.73) | <0.01 |
| MDCD | 1,878 | 15,407 | 1.0 | 9.9 | 16 | 250 | 0.79 (0.44 - 1.35) | 0.42 | 0.77 (0.44 - 1.34) | 0.36 |
| Open Claims | 224,856 | 815,621 | 140.1 | 771.4 | 587 | 10,030 | 0.58 (0.53 - 0.64) | <0.01 | 0.59 (0.49 - 0.72) | <0.01 |
| OEHR | 13,411 | 81,589 | 4.0 | 45.8 | 25 | 751 | 0.65 (0.41 - 0.98) | 0.05 | 0.61 (0.38 - 0.98) | 0.06 |
| Meta-analysis | 269,794 | 1,024,399 | 162.0 | 917.5 | 699 | 12,381 | 0.59 (0.54 - 0.64) | <0.01 | 0.60 (0.48 - 0.74) | <0.01 |
| Meta-analysis (w/o USOC) | 44,938 | 208,778 | 22.0 | 146.1 | 112 | 2,351 | 0.60 (0.45 - 0.82) | <0.01 | 0.60 (0.41 - 0.87) | <0.01 |
| **PS-matching / on-treatment TAR without escalation** | | | | | | | | | | |
| CCAE | 15,081 | 68,367 | 8.4 | 43.4 | 13 | 281 | 0.38 (0.18 - 0.72) | <0.01 | 0.40 (0.20 - 0.79) | 0.01 |
| ODOD | 8,619 | 52,820 | 4.4 | 39.4 | 56 | 913 | 0.83 (0.58 - 1.16) | 0.29 | 0.85 (0.60 - 1.21) | 0.41 |
| MDCR | 1,050 | 12,178 | 0.6 | 10.0 | 9 | 293 | 0.46 (0.18 - 0.98) | 0.07 | 0.36 (0.16 - 0.83) | 0.02 |
| MDCD | 1,745 | 13,943 | 0.8 | 7.3 | 13 | 172 | 0.80 (0.37 - 1.55) | 0.54 | 0.76 (0.37 - 1.56) | 0.47 |
| Open Claims | 172,903 | 801,531 | 104.0 | 637.8 | 438 | 7,864 | 0.64 (0.57 - 0.72) | <0.01 | 0.65 (0.54 - 0.78) | <0.01 |
| OEHR | 10,956 | 76,773 | 2.9 | 36.5 | 18 | 449 | 0.85 (0.49 - 1.40) | 0.54 | 0.79 (0.46 - 1.34) | 0.40 |
| Meta-analysis | 210,354 | 1,025,612 | 121.1 | 774.4 | 547 | 9,972 | 0.66 (0.58 - 0.76) | <0.01 | 0.66 (0.53 - 0.83) | <0.01 |
| Meta-analysis (w/o USOC) | 37,451 | 224,081 | 17.1 | 136.6 | 109 | 2,108 | 0.69 (0.51 - 0.93) | 0.02 | 0.68 (0.47 - 0.96) | 0.04 |
| **PS-stratification / on-treatment TAR without escalation** | | | | | | | | | | |
| CCAE | 20,267 | 70,548 | 10.9 | 44.6 | 15 | 287 | 0.34 (0.19 - 0.58) | <0.01 | 0.34 (0.20 - 0.60) | <0.01 |
| ODOD | 11,260 | 56,641 | 5.4 | 43.1 | 58 | 1,110 | 0.69 (0.51 - 0.93) | 0.02 | 0.71 (0.52 - 0.96) | 0.03 |
| MDCR | 1,169 | 15,734 | 0.7 | 12.8 | 9 | 475 | 0.46 (0.21 - 0.88) | 0.03 | 0.37 (0.18 - 0.76) | <0.01 |
| MDCD | 1,878 | 15,407 | 0.9 | 8.2 | 13 | 201 | 0.78 (0.40 - 1.40) | 0.43 | 0.77 (0.41 - 1.43) | 0.42 |
| Open Claims | 224,856 | 815,621 | 126.4 | 649.4 | 507 | 8,427 | 0.57 (0.51 - 0.63) | <0.01 | 0.57 (0.47 - 0.69) | <0.01 |
| OEHR | 13,411 | 81,589 | 3.4 | 39.1 | 20 | 561 | 0.67 (0.40 - 1.07) | 0.11 | 0.63 (0.37 - 1.07) | 0.11 |
| Meta-analysis | 269,794 | 1,024,399 | 146.2 | 776.2 | 600 | 10,385 | 0.57 (0.52 - 0.64) | <0.01 | 0.58 (0.47 - 0.71) | <0.01 |
| Meta-analysis (w/o USOC) | 44,938 | 208,778 | 19.8 | 126.8 | 93 | 1,958 | 0.57 (0.38 - 0.85) | <0.01 | 0.57 (0.36 - 0.90) | 0.02 |

##### 1.5.7 Measured renal dysfunction

**Supplementary Table 111:** Exposures, outcomes and risk estimates for Measured renal dysfunction in GLP1RA (female) (T) vs SU (female) (C) new-user cohorts under four different propensity score (PS) adjustment / time-at-risk (TAR) designs across data sources. We report total patient exposure time in 1,000 patient-year (KYr) units, uncalibrated and calibrated hazard ratios (HRs) and their 95% confidence intervals (CIs). Greyed out entries did not pass study diagnostics and are included only for completeness.

| | Patients | | Exposure (KYr) | | Outcomes | | Uncalibrated | | Calibrated | |
|---|---|---|---|---|---|---|---|---|---|---|
| | T | C | T | C | T | C | HR (95% CI) | $p$ | HR (95% CI) | $p$ |
| **PS-matching / on-treatment TAR** | | | | | | | | | | |
| CCAE | 15,319 | 69,514 | 9.5 | 51.6 | <5 | 17 | 1.02 (0.13 - 5.68) | 0.98 | 1.06 (0.16 - 7.00) | 0.95 |
| ODOD | 8,942 | 55,439 | 5.0 | 47.1 | <5 | 55 | 0.73 (0.16 - 2.40) | 0.64 | 0.75 (0.19 - 2.91) | 0.68 |
| MDCR | 1,098 | 13,508 | 0.7 | 12.4 | 0 | <5 | | | | |
| MDCD | 1,887 | 15,102 | 1.0 | 9.7 | 0 | 0 | | | | |
| Open Claims | 178,376 | 835,401 | 120.1 | 795.9 | 0 | 0 | | | | |
| OEHR | 11,145 | 78,232 | 3.4 | 44.2 | 0 | 0 | | | | |
| Meta-analysis | 8,942 | 55,439 | 5.0 | 47.1 | <5 | 55 | 0.73 (0.16 - 2.40) | 0.64 | 0.72 (0.18 - 2.85) | 0.64 |
| Meta-analysis (w/o USOC) | 8,942 | 55,439 | 5.0 | 47.1 | <5 | 55 | 0.73 (0.16 - 2.40) | 0.64 | 0.71 (0.18 - 2.78) | 0.63 |
| **PS-stratification / on-treatment TAR** | | | | | | | | | | |
| CCAE | 20,567 | 71,860 | 12.2 | 52.9 | <5 | 17 | 1.07 (0.15 - 4.61) | 0.93 | 1.06 (0.19 - 5.85) | 0.95 |
| ODOD | 11,651 | 60,230 | 6.2 | 52.2 | <5 | 82 | 0.61 (0.14 - 1.88) | 0.46 | 0.62 (0.17 - 2.32) | 0.48 |
| MDCR | 1,231 | 17,494 | 0.8 | 15.8 | 0 | <5 | | | | |
| MDCD | 2,034 | 16,863 | 1.1 | 11.0 | 0 | 0 | | | | |
| Open Claims | 231,281 | 852,348 | 144.1 | 811.8 | 0 | 0 | | | | |
| OEHR | 13,639 | 83,370 | 4.0 | 47.5 | 0 | 0 | | | | |
| Meta-analysis | 11,651 | 60,230 | 6.2 | 52.2 | <5 | 82 | 0.61 (0.14 - 1.88) | 0.46 | 0.62 (0.16 - 2.32) | 0.48 |
| Meta-analysis (w/o USOC) | 11,651 | 60,230 | 6.2 | 52.2 | <5 | 82 | 0.61 (0.14 - 1.88) | 0.46 | 0.60 (0.16 - 2.28) | 0.46 |
| **PS-matching / on-treatment TAR without escalation** | | | | | | | | | | |
| CCAE | 15,319 | 69,514 | 8.6 | 44.2 | <5 | 14 | 1.01 (0.13 - 5.54) | 0.99 | 1.04 (0.16 - 6.75) | 0.96 |
| ODOD | 8,942 | 55,439 | 4.5 | 41.8 | <5 | 44 | 0.83 (0.18 - 2.92) | 0.80 | 0.86 (0.21 - 3.49) | 0.83 |
| MDCR | 1,098 | 13,508 | 0.7 | 11.3 | 0 | <5 | | | | |
| MDCD | 1,887 | 15,102 | 0.9 | 8.1 | 0 | 0 | | | | |
| Open Claims | 178,376 | 835,401 | 107.1 | 668.8 | 0 | 0 | | | | |
| OEHR | 11,145 | 78,232 | 2.9 | 37.4 | 0 | 0 | | | | |
| Meta-analysis | 8,942 | 55,439 | 4.5 | 41.8 | <5 | 44 | 0.83 (0.18 - 2.92) | 0.80 | 0.83 (0.20 - 3.41) | 0.80 |
| Meta-analysis (w/o USOC) | 8,942 | 55,439 | 4.5 | 41.8 | <5 | 44 | 0.83 (0.18 - 2.92) | 0.80 | 0.81 (0.20 - 3.35) | 0.78 |
| **PS-stratification / on-treatment TAR without escalation** | | | | | | | | | | |
| CCAE | 20,567 | 71,860 | 11.1 | 45.4 | <5 | 14 | 1.12 (0.16 - 4.92) | 0.90 | 1.11 (0.20 - 6.22) | 0.91 |
| ODOD | 11,651 | 60,230 | 5.6 | 46.4 | <5 | 70 | 0.68 (0.15 - 2.17) | 0.57 | 0.70 (0.18 - 2.67) | 0.60 |
| MDCR | 1,231 | 17,494 | 0.7 | 14.4 | 0 | <5 | | | | |
| MDCD | 2,034 | 16,863 | 0.9 | 9.2 | 0 | 0 | | | | |
| Open Claims | 231,281 | 852,348 | 130.0 | 682.7 | 0 | 0 | | | | |
| OEHR | 13,639 | 83,370 | 3.5 | 40.3 | 0 | 0 | | | | |
| Meta-analysis | 11,651 | 60,230 | 5.6 | 46.4 | <5 | 70 | 0.68 (0.15 - 2.17) | 0.57 | 0.69 (0.18 - 2.65) | 0.58 |
| Meta-analysis (w/o USOC) | 11,651 | 60,230 | 5.6 | 46.4 | <5 | 70 | 0.68 (0.15 - 2.17) | 0.57 | 0.68 (0.17 - 2.64) | 0.57 |

##### 1.5.8 Stroke

**Supplementary Table 112:** Exposures, outcomes and risk estimates for Stroke in GLP1RA (female) (T) vs SU (female) (C) new-user cohorts under four different propensity score (PS) adjustment / time-at-risk (TAR) designs across data sources. We report total patient exposure time in 1,000 patient-year (KYr) units, uncalibrated and calibrated hazard ratios (HRs) and their 95% confidence intervals (CIs). Greyed out entries did not pass study diagnostics and are included only for completeness.

| | Patients | | Exposure (KYr) | | Outcomes | | Uncalibrated | | Calibrated | |
|---|---|---|---|---|---|---|---|---|---|---|
| | T | C | T | C | T | C | HR (95% CI) | $p$ | HR (95% CI) | $p$ |
| **PS-matching / on-treatment TAR** | | | | | | | | | | |
| CCAE | 15,170 | 68,693 | 9.4 | 50.9 | 24 | 188 | 0.59 (0.29 - 1.09) | 0.11 | 0.61 (0.31 - 1.21) | 0.17 |
| ODOD | 8,800 | 54,222 | 4.9 | 45.8 | 28 | 491 | 0.88 (0.55 - 1.37) | 0.58 | 0.91 (0.57 - 1.45) | 0.70 |
| MDCR | 1,067 | 12,540 | 0.7 | 11.4 | <5 | 197 | 0.27 (0.06 - 0.77) | 0.04 | 0.21 (0.06 - 0.74) | 0.02 |
| MDCD | 1,835 | 14,600 | 1.0 | 9.3 | 8 | 111 | 0.87 (0.35 - 1.95) | 0.75 | 0.83 (0.35 - 1.95) | 0.67 |
| Open Claims | 174,625 | 812,865 | 117.7 | 772.1 | 338 | 5,028 | 0.69 (0.59 - 0.80) | <0.01 | 0.70 (0.57 - 0.86) | <0.01 |
| OEHR | 11,054 | 77,589 | 3.3 | 43.7 | 8 | 248 | 0.72 (0.27 - 1.59) | 0.47 | 0.69 (0.28 - 1.67) | 0.41 |
| Meta-analysis | 212,551 | 1,040,509 | 137.1 | 933.2 | <411 | 6,263 | 0.70 (0.61 - 0.79) | <0.01 | 0.70 (0.55 - 0.87) | <0.01 |
| Meta-analysis (w/o USOC) | 37,926 | 227,644 | 19.4 | 161.1 | <73 | 1,235 | 0.73 (0.53 - 0.99) | 0.04 | 0.71 (0.50 - 1.01) | 0.07 |
| **PS-stratification / on-treatment TAR** | | | | | | | | | | |
| CCAE | 20,377 | 70,931 | 12.1 | 52.2 | 32 | 192 | 0.81 (0.52 - 1.24) | 0.35 | 0.80 (0.50 - 1.26) | 0.36 |
| ODOD | 11,499 | 58,487 | 6.1 | 50.3 | 35 | 633 | 0.90 (0.59 - 1.32) | 0.59 | 0.92 (0.62 - 1.37) | 0.68 |
| MDCR | 1,195 | 16,620 | 0.8 | 14.9 | <5 | 303 | 0.38 (0.11 - 0.96) | 0.08 | 0.32 (0.11 - 0.93) | 0.04 |
| MDCD | 1,977 | 16,180 | 1.1 | 10.5 | 8 | 129 | 1.03 (0.44 - 2.13) | 0.94 | 0.99 (0.45 - 2.20) | 0.93 |
| Open Claims | 226,789 | 828,014 | 141.4 | 786.3 | 396 | 5,350 | 0.68 (0.60 - 0.76) | <0.01 | 0.70 (0.57 - 0.85) | <0.01 |
| OEHR | 13,542 | 82,512 | 4.0 | 46.8 | 9 | 295 | 0.61 (0.28 - 1.20) | 0.19 | 0.57 (0.27 - 1.22) | 0.16 |
| Meta-analysis | 272,207 | 1,039,944 | 163.5 | 935.6 | 472 | 6,470 | 0.71 (0.62 - 0.81) | <0.01 | 0.72 (0.57 - 0.91) | <0.01 |
| Meta-analysis (w/o USOC) | 45,418 | 211,930 | 22.2 | 149.3 | 76 | 1,120 | 0.82 (0.62 - 1.07) | 0.15 | 0.81 (0.57 - 1.15) | 0.25 |
| **PS-matching / on-treatment TAR without escalation** | | | | | | | | | | |
| CCAE | 15,170 | 68,693 | 8.5 | 43.6 | 19 | 158 | 0.50 (0.22 - 1.01) | 0.07 | 0.52 (0.24 - 1.11) | 0.10 |
| ODOD | 8,800 | 54,222 | 4.5 | 40.7 | 24 | 436 | 0.89 (0.53 - 1.43) | 0.64 | 0.92 (0.56 - 1.51) | 0.74 |
| MDCR | 1,067 | 12,540 | 0.6 | 10.4 | <5 | 175 | 0.29 (0.07 - 0.86) | 0.06 | 0.23 (0.07 - 0.82) | 0.02 |
| MDCD | 1,835 | 14,600 | 0.9 | 7.7 | 7 | 94 | 0.75 (0.26 - 1.84) | 0.56 | 0.72 (0.27 - 1.90) | 0.51 |
| Open Claims | 174,625 | 812,865 | 105.0 | 649.1 | 291 | 4,259 | 0.63 (0.53 - 0.73) | <0.01 | 0.63 (0.51 - 0.78) | <0.01 |
| OEHR | 11,054 | 77,589 | 2.9 | 37.0 | 6 | 190 | 0.67 (0.23 - 1.61) | 0.43 | 0.63 (0.24 - 1.67) | 0.36 |
| Meta-analysis | 212,551 | 1,040,509 | 122.2 | 788.6 | <352 | 5,312 | 0.64 (0.55 - 0.74) | <0.01 | 0.64 (0.50 - 0.80) | <0.01 |
| Meta-analysis (w/o USOC) | 37,926 | 227,644 | 17.3 | 139.5 | <61 | 1,053 | 0.69 (0.49 - 0.97) | 0.03 | 0.68 (0.46 - 1.00) | 0.06 |
| **PS-stratification / on-treatment TAR without escalation** | | | | | | | | | | |
| CCAE | 20,377 | 70,931 | 11.0 | 44.8 | 24 | 161 | 0.68 (0.41 - 1.10) | 0.13 | 0.68 (0.41 - 1.12) | 0.14 |
| ODOD | 11,499 | 58,487 | 5.5 | 44.8 | 31 | 564 | 0.87 (0.55 - 1.31) | 0.51 | 0.89 (0.58 - 1.36) | 0.60 |
| MDCR | 1,195 | 16,620 | 0.7 | 13.6 | <5 | 272 | 0.45 (0.13 - 1.14) | 0.14 | 0.36 (0.12 - 1.06) | 0.07 |
| MDCD | 1,977 | 16,180 | 0.9 | 8.8 | 7 | 107 | 1.06 (0.42 - 2.33) | 0.89 | 1.04 (0.44 - 2.48) | 0.91 |
| Open Claims | 226,789 | 828,014 | 127.5 | 661.6 | 348 | 4,533 | 0.66 (0.58 - 0.75) | <0.01 | 0.67 (0.55 - 0.82) | <0.01 |
| OEHR | 13,542 | 82,512 | 3.5 | 39.7 | 7 | 223 | 0.56 (0.22 - 1.20) | 0.18 | 0.52 (0.22 - 1.24) | 0.15 |
| Meta-analysis | 272,207 | 1,039,944 | 147.5 | 790.9 | 410 | 5,481 | 0.67 (0.60 - 0.76) | <0.01 | 0.68 (0.55 - 0.84) | <0.01 |
| Meta-analysis (w/o USOC) | 45,418 | 211,930 | 20.0 | 129.3 | 62 | 948 | 0.75 (0.55 - 1.01) | 0.06 | 0.74 (0.50 - 1.10) | 0.15 |

##### 1.5.9 Sudden cardiac death

**Supplementary Table 113:** Exposures, outcomes and risk estimates for Sudden cardiac death in GLP1RA (female) (T) vs SU (female) (C) new-user cohorts under four different propensity score (PS) adjustment / time-at-risk (TAR) designs across data sources. We report total patient exposure time in 1,000 patient-year (KYr) units, uncalibrated and calibrated hazard ratios (HRs) and their 95% confidence intervals (CIs). Greyed out entries did not pass study diagnostics and are included only for completeness.

| | Patients | | Exposure (KYr) | | Outcomes | | Uncalibrated | | Calibrated | |
|---|---|---|---|---|---|---|---|---|---|---|
| | T | C | T | C | T | C | HR (95% CI) | $p$ | HR (95% CI) | $p$ |
| **PS-matching / on-treatment TAR** | | | | | | | | | | |
| CCAE | 15,327 | 69,542 | 9.5 | 51.6 | 6 | 52 | 1.01 (0.34 - 2.63) | 0.98 | 1.06 (0.38 - 2.96) | 0.92 |
| ODOD | 8,947 | 55,512 | 5.0 | 47.2 | 8 | 122 | 1.32 (0.50 - 3.03) | 0.55 | 1.35 (0.55 - 3.35) | 0.52 |
| MDCR | 1,095 | 13,395 | 0.7 | 12.3 | 0 | 32 | | | | |
| MDCD | 1,886 | 15,066 | 1.0 | 9.7 | 0 | 29 | | | | |
| Open Claims | 178,120 | 834,180 | 119.9 | 794.5 | 58 | 1,004 | 0.55 (0.38 - 0.78) | <0.01 | 0.56 (0.38 - 0.82) | <0.01 |
| OEHR | 11,139 | 78,206 | 3.4 | 44.2 | <5 | 55 | 0.51 (0.03 - 2.94) | 0.57 | 0.48 (0.05 - 5.12) | 0.55 |
| Meta-analysis | 213,533 | 1,037,440 | 137.9 | 937.5 | <77 | 1,233 | 0.76 (0.44 - 1.30) | 0.32 | 0.76 (0.43 - 1.34) | 0.34 |
| Meta-analysis (w/o USOC) | 35,413 | 203,260 | 17.9 | 143.0 | <19 | 229 | 1.10 (0.58 - 2.10) | 0.77 | 1.08 (0.55 - 2.10) | 0.83 |
| **PS-stratification / on-treatment TAR** | | | | | | | | | | |
| CCAE | 20,573 | 71,879 | 12.2 | 52.9 | 6 | 53 | 0.86 (0.31 - 2.04) | 0.76 | 0.85 (0.33 - 2.20) | 0.74 |
| ODOD | 11,657 | 60,285 | 6.2 | 52.3 | 8 | 163 | 0.89 (0.37 - 1.91) | 0.79 | 0.92 (0.40 - 2.10) | 0.84 |
| MDCR | 1,227 | 17,461 | 0.8 | 15.8 | 0 | 54 | | | | |
| MDCD | 2,033 | 16,806 | 1.1 | 11.0 | 0 | 35 | | | | |
| Open Claims | 230,968 | 850,946 | 143.9 | 810.3 | 66 | 1,077 | 0.57 (0.43 - 0.74) | <0.01 | 0.58 (0.42 - 0.80) | <0.01 |
| OEHR | 13,632 | 83,333 | 4.0 | 47.5 | <5 | 81 | 0.48 (0.07 - 1.78) | 0.37 | 0.44 (0.09 - 2.24) | 0.33 |
| Meta-analysis | 276,830 | 1,066,443 | 166.4 | 963.0 | <85 | 1,374 | 0.61 (0.47 - 0.78) | <0.01 | 0.61 (0.45 - 0.84) | <0.01 |
| Meta-analysis (w/o USOC) | 45,862 | 215,497 | 22.5 | 152.7 | <19 | 297 | 0.81 (0.46 - 1.45) | 0.48 | 0.80 (0.43 - 1.50) | 0.50 |
| **PS-matching / on-treatment TAR without escalation** | | | | | | | | | | |
| CCAE | 15,327 | 69,542 | 8.6 | 44.2 | 5 | 38 | 0.81 (0.25 - 2.25) | 0.70 | 0.83 (0.28 - 2.51) | 0.75 |
| ODOD | 8,947 | 55,512 | 4.5 | 41.9 | 8 | 104 | 1.45 (0.54 - 3.44) | 0.43 | 1.50 (0.59 - 3.78) | 0.40 |
| MDCR | 1,095 | 13,395 | 0.6 | 11.2 | 0 | 29 | | | | |
| MDCD | 1,886 | 15,066 | 0.9 | 8.0 | 0 | 25 | | | | |
| Open Claims | 178,120 | 834,180 | 107.0 | 667.7 | 49 | 841 | 0.59 (0.40 - 0.85) | <0.01 | 0.59 (0.39 - 0.89) | 0.01 |
| OEHR | 11,139 | 78,206 | 2.9 | 37.4 | <5 | 36 | 0.64 (0.03 - 4.02) | 0.71 | 0.59 (0.05 - 6.64) | 0.67 |
| Meta-analysis | 213,533 | 1,037,440 | 123.0 | 791.2 | <67 | 1,019 | 0.76 (0.46 - 1.28) | 0.31 | 0.76 (0.44 - 1.32) | 0.33 |
| Meta-analysis (w/o USOC) | 35,413 | 203,260 | 16.0 | 123.5 | <18 | 178 | 1.09 (0.55 - 2.14) | 0.81 | 1.06 (0.52 - 2.15) | 0.87 |
| **PS-stratification / on-treatment TAR without escalation** | | | | | | | | | | |
| CCAE | 20,573 | 71,879 | 11.1 | 45.4 | 5 | 39 | 0.79 (0.25 - 2.05) | 0.67 | 0.79 (0.28 - 2.24) | 0.66 |
| ODOD | 11,657 | 60,285 | 5.6 | 46.5 | 8 | 141 | 1.03 (0.41 - 2.25) | 0.95 | 1.06 (0.45 - 2.47) | 0.90 |
| MDCR | 1,227 | 17,461 | 0.7 | 14.4 | 0 | 50 | | | | |
| MDCD | 2,033 | 16,806 | 0.9 | 9.1 | 0 | 31 | | | | |
| Open Claims | 230,968 | 850,946 | 129.8 | 681.5 | 57 | 904 | 0.54 (0.40 - 0.73) | <0.01 | 0.55 (0.39 - 0.77) | <0.01 |
| OEHR | 13,632 | 83,333 | 3.5 | 40.3 | <5 | 49 | 0.31 (0.02 - 1.80) | 0.33 | 0.29 (0.03 - 3.10) | 0.31 |
| Meta-analysis | 276,830 | 1,066,443 | 150.0 | 813.7 | <75 | 1,133 | 0.62 (0.43 - 0.91) | 0.01 | 0.63 (0.41 - 0.95) | 0.03 |
| Meta-analysis (w/o USOC) | 45,862 | 215,497 | 20.2 | 132.2 | <18 | 229 | 0.86 (0.46 - 1.62) | 0.64 | 0.85 (0.43 - 1.68) | 0.65 |

##### 1.5.10 Abnormal weight gain

**Supplementary Table 114:** Exposures, outcomes and risk estimates for Abnormal weight gain in GLP1RA (female) (T) vs SU (female) (C) new-user cohorts under four different propensity score (PS) adjustment / time-at-risk (TAR) designs across data sources. We report total patient exposure time in 1,000 patient-year (KYr) units, uncalibrated and calibrated hazard ratios (HRs) and their 95% confidence intervals (CIs). Greyed out entries did not pass study diagnostics and are included only for completeness.

| | Patients | | Exposure (KYr) | | Outcomes | | Uncalibrated | | Calibrated | |
|---|---|---|---|---|---|---|---|---|---|---|
| | T | C | T | C | T | C | HR (95% CI) | $p$ | HR (95% CI) | $p$ |
| **PS-matching / on-treatment TAR** | | | | | | | | | | |
| CCAE | 13,857 | 66,170 | 8.5 | 49.0 | 83 | 394 | 1.14 (0.83 - 1.55) | 0.41 | 1.19 (0.83 - 1.70) | 0.40 |
| ODOD | 8,189 | 52,870 | 4.6 | 44.7 | 50 | 409 | 0.85 (0.56 - 1.26) | 0.43 | 0.88 (0.58 - 1.33) | 0.57 |
| MDCR | 1,061 | 13,346 | 0.7 | 12.2 | <5 | 35 | 0.86 (0.13 - 3.48) | 0.86 | 0.68 (0.13 - 3.58) | 0.65 |
| MDCD | 1,632 | 13,899 | 0.9 | 8.9 | 13 | 118 | 1.23 (0.54 - 2.55) | 0.60 | 1.17 (0.54 - 2.56) | 0.70 |
| Open Claims | 161,505 | 795,705 | 108.4 | 758.2 | 413 | 2,381 | 1.00 (0.86 - 1.16) | 0.97 | 1.01 (0.82 - 1.25) | 0.91 |
| OEHR | 10,162 | 74,811 | 3.1 | 41.9 | 34 | 397 | 0.96 (0.60 - 1.49) | 0.86 | 0.92 (0.58 - 1.44) | 0.71 |
| Meta-analysis | 195,345 | 1,003,455 | 125.4 | 902.7 | 593 | 3,699 | 1.01 (0.89 - 1.14) | 0.93 | 1.00 (0.81 - 1.25) | 0.91 |
| Meta-analysis (w/o USOC) | 33,840 | 207,750 | 17.0 | 144.5 | 180 | 1,318 | 1.02 (0.83 - 1.26) | 0.84 | 1.00 (0.77 - 1.30) | 0.90 |
| **PS-stratification / on-treatment TAR** | | | | | | | | | | |
| CCAE | 17,829 | 68,489 | 10.6 | 50.4 | 116 | 399 | 0.97 (0.75 - 1.23) | 0.78 | 0.95 (0.72 - 1.27) | 0.76 |
| ODOD | 10,179 | 57,785 | 5.4 | 49.9 | 61 | 452 | 0.79 (0.57 - 1.08) | 0.15 | 0.81 (0.59 - 1.12) | 0.21 |
| MDCR | 1,171 | 17,288 | 0.7 | 15.6 | <5 | 55 | 1.02 (0.28 - 2.90) | 0.97 | 0.85 (0.27 - 2.73) | 0.79 |
| MDCD | 1,722 | 15,658 | 0.9 | 10.2 | 13 | 130 | 0.81 (0.42 - 1.46) | 0.51 | 0.78 (0.42 - 1.46) | 0.45 |
| Open Claims | 202,244 | 813,475 | 126.4 | 775.1 | 549 | 2,456 | 1.07 (0.96 - 1.20) | 0.20 | 1.10 (0.90 - 1.34) | 0.37 |
| OEHR | 12,039 | 80,074 | 3.5 | 45.2 | 53 | 432 | 1.33 (0.95 - 1.83) | 0.09 | 1.24 (0.85 - 1.82) | 0.33 |
| Meta-analysis | 242,291 | 1,019,823 | 145.9 | 920.6 | 779 | 3,739 | 1.03 (0.89 - 1.20) | 0.67 | 1.04 (0.82 - 1.33) | 0.73 |
| Meta-analysis (w/o USOC) | 40,047 | 206,348 | 19.5 | 145.4 | 230 | 1,283 | 1.00 (0.76 - 1.32) | 0.99 | 0.99 (0.69 - 1.41) | 0.92 |
| **PS-matching / on-treatment TAR without escalation** | | | | | | | | | | |
| CCAE | 13,857 | 66,170 | 7.7 | 42.1 | 75 | 325 | 1.22 (0.87 - 1.68) | 0.24 | 1.26 (0.90 - 1.77) | 0.24 |
| ODOD | 8,189 | 52,870 | 4.1 | 39.8 | 45 | 346 | 0.84 (0.53 - 1.30) | 0.45 | 0.87 (0.56 - 1.36) | 0.56 |
| MDCR | 1,061 | 13,346 | 0.6 | 11.1 | <5 | 30 | 1.18 (0.18 - 4.64) | 0.84 | 0.93 (0.18 - 4.76) | 0.92 |
| MDCD | 1,632 | 13,899 | 0.8 | 7.4 | 11 | 99 | 1.30 (0.53 - 2.88) | 0.54 | 1.25 (0.53 - 2.91) | 0.62 |
| Open Claims | 161,505 | 795,705 | 96.7 | 638.6 | 369 | 1,959 | 1.00 (0.85 - 1.17) | 1.00 | 1.01 (0.81 - 1.24) | 0.92 |
| OEHR | 10,162 | 74,811 | 2.6 | 35.6 | 29 | 310 | 1.07 (0.65 - 1.70) | 0.79 | 0.99 (0.61 - 1.61) | 0.90 |
| Meta-analysis | 195,345 | 1,003,455 | 111.9 | 763.5 | 529 | 3,039 | 1.03 (0.90 - 1.17) | 0.69 | 1.02 (0.82 - 1.28) | 0.83 |
| Meta-analysis (w/o USOC) | 33,840 | 207,750 | 15.2 | 124.9 | 160 | 1,080 | 1.08 (0.87 - 1.35) | 0.49 | 1.06 (0.79 - 1.42) | 0.72 |
| **PS-stratification / on-treatment TAR without escalation** | | | | | | | | | | |
| CCAE | 17,829 | 68,489 | 9.6 | 43.4 | 107 | 330 | 1.01 (0.77 - 1.31) | 0.96 | 1.00 (0.77 - 1.31) | 0.91 |
| ODOD | 10,179 | 57,785 | 4.9 | 44.5 | 56 | 386 | 0.88 (0.62 - 1.22) | 0.45 | 0.90 (0.64 - 1.27) | 0.56 |
| MDCR | 1,171 | 17,288 | 0.7 | 14.2 | <5 | 48 | 1.02 (0.28 - 2.93) | 0.98 | 0.82 (0.25 - 2.67) | 0.75 |
| MDCD | 1,722 | 15,658 | 0.8 | 8.5 | 11 | 110 | 0.86 (0.42 - 1.64) | 0.67 | 0.85 (0.43 - 1.69) | 0.65 |
| Open Claims | 202,244 | 813,475 | 113.9 | 653.4 | 499 | 2,028 | 1.08 (0.96 - 1.22) | 0.20 | 1.09 (0.90 - 1.33) | 0.39 |
| OEHR | 12,039 | 80,074 | 3.1 | 38.5 | 46 | 337 | 1.41 (0.98 - 1.99) | 0.06 | 1.31 (0.87 - 1.99) | 0.25 |
| Meta-analysis | 242,291 | 1,019,823 | 131.4 | 779.7 | 708 | 3,081 | 1.07 (0.97 - 1.19) | 0.16 | 1.08 (0.88 - 1.33) | 0.45 |
| Meta-analysis (w/o USOC) | 40,047 | 206,348 | 17.5 | 126.3 | 209 | 1,053 | 1.06 (0.83 - 1.37) | 0.63 | 1.06 (0.75 - 1.50) | 0.76 |

##### 1.5.11 Abnormal weight loss

**Supplementary Table 115:** Exposures, outcomes and risk estimates for Abnormal weight loss in GLP1RA (female) (T) vs SU (female) (C) new-user cohorts under four different propensity score (PS) adjustment / time-at-risk (TAR) designs across data sources. We report total patient exposure time in 1,000 patient-year (KYr) units, uncalibrated and calibrated hazard ratios (HRs) and their 95% confidence intervals (CIs). Greyed out entries did not pass study diagnostics and are included only for completeness.

| | Patients | | Exposure (KYr) | | Outcomes | | Uncalibrated | | Calibrated | |
|---|---|---|---|---|---|---|---|---|---|---|
| | T | C | T | C | T | C | HR (95% CI) | $p$ | HR (95% CI) | $p$ |
| **PS-matching / on-treatment TAR** | | | | | | | | | | |
| CCAE | 15,023 | 68,111 | 9.3 | 50.4 | 77 | 255 | 1.74 (1.19 - 2.53) | <0.01 | 1.82 (1.20 - 2.76) | 0.01 |
| ODOD | 8,688 | 53,392 | 4.9 | 45.0 | 74 | 599 | 1.36 (0.97 - 1.88) | 0.07 | 1.40 (0.99 - 1.98) | 0.09 |
| MDCR | 1,076 | 13,152 | 0.7 | 12.0 | 12 | 127 | 2.08 (0.88 - 4.42) | 0.07 | 1.65 (0.74 - 3.71) | 0.24 |
| MDCD | 1,813 | 14,392 | 1.0 | 9.1 | 19 | 141 | 1.02 (0.50 - 1.94) | 0.95 | 0.97 (0.49 - 1.91) | 0.89 |
| Open Claims | 172,652 | 806,459 | 115.9 | 765.8 | 591 | 3,941 | 1.29 (1.14 - 1.46) | <0.01 | 1.31 (1.08 - 1.59) | <0.01 |
| OEHR | 10,878 | 76,256 | 3.3 | 42.7 | 27 | 458 | 0.92 (0.55 - 1.47) | 0.73 | 0.87 (0.53 - 1.43) | 0.60 |
| Meta-analysis | 210,130 | 1,031,762 | 135.0 | 925.0 | 800 | 5,521 | 1.31 (1.18 - 1.46) | <0.01 | 1.31 (1.06 - 1.62) | 0.01 |
| Meta-analysis (w/o USOC) | 37,478 | 225,303 | 19.1 | 159.3 | 209 | 1,580 | 1.35 (1.04 - 1.76) | 0.02 | 1.32 (0.98 - 1.79) | 0.09 |
| **PS-stratification / on-treatment TAR** | | | | | | | | | | |
| CCAE | 20,123 | 70,373 | 11.9 | 51.8 | 107 | 264 | 1.70 (1.28 - 2.24) | <0.01 | 1.68 (1.23 - 2.29) | <0.01 |
| ODOD | 11,297 | 57,469 | 6.0 | 49.3 | 88 | 704 | 1.39 (1.05 - 1.81) | 0.02 | 1.42 (1.08 - 1.87) | 0.02 |
| MDCR | 1,208 | 16,711 | 0.8 | 15.0 | 12 | 183 | 2.33 (1.14 - 4.35) | 0.01 | 1.94 (0.99 - 3.80) | 0.06 |
| MDCD | 1,955 | 16,069 | 1.0 | 10.4 | 20 | 167 | 1.27 (0.73 - 2.11) | 0.38 | 1.22 (0.72 - 2.09) | 0.47 |
| Open Claims | 223,395 | 821,471 | 138.9 | 779.9 | 739 | 4,151 | 1.31 (1.19 - 1.44) | <0.01 | 1.34 (1.10 - 1.62) | <0.01 |
| OEHR | 13,292 | 80,965 | 3.9 | 45.7 | 34 | 527 | 1.00 (0.67 - 1.45) | 0.99 | 0.93 (0.60 - 1.44) | 0.77 |
| Meta-analysis | 268,107 | 1,030,278 | 160.7 | 926.6 | 968 | 5,646 | 1.35 (1.18 - 1.53) | <0.01 | 1.36 (1.08 - 1.72) | 0.01 |
| Meta-analysis (w/o USOC) | 44,712 | 208,807 | 21.8 | 146.7 | 229 | 1,495 | 1.37 (1.04 - 1.80) | 0.03 | 1.35 (0.95 - 1.93) | 0.11 |
| **PS-matching / on-treatment TAR without escalation** | | | | | | | | | | |
| CCAE | 15,023 | 68,111 | 8.4 | 43.2 | 63 | 210 | 1.62 (1.06 - 2.45) | 0.02 | 1.68 (1.10 - 2.57) | 0.03 |
| ODOD | 8,688 | 53,392 | 4.4 | 40.0 | 67 | 533 | 1.40 (0.97 - 1.98) | 0.07 | 1.44 (1.00 - 2.07) | 0.06 |
| MDCR | 1,076 | 13,152 | 0.6 | 10.9 | 11 | 114 | 2.80 (1.17 - 6.00) | 0.01 | 2.20 (0.97 - 4.99) | 0.07 |
| MDCD | 1,813 | 14,392 | 0.8 | 7.6 | 13 | 123 | 0.63 (0.23 - 1.46) | 0.33 | 0.60 (0.24 - 1.52) | 0.29 |
| Open Claims | 172,652 | 806,459 | 103.5 | 643.9 | 528 | 3,309 | 1.33 (1.16 - 1.52) | <0.01 | 1.34 (1.10 - 1.62) | <0.01 |
| OEHR | 10,878 | 76,256 | 2.8 | 36.2 | 21 | 355 | 0.95 (0.54 - 1.59) | 0.85 | 0.88 (0.51 - 1.53) | 0.67 |
| Meta-analysis | 210,130 | 1,031,762 | 120.5 | 781.9 | 703 | 4,644 | 1.34 (1.19 - 1.50) | <0.01 | 1.34 (1.08 - 1.66) | <0.01 |
| Meta-analysis (w/o USOC) | 37,478 | 225,303 | 17.0 | 137.9 | 175 | 1,335 | 1.35 (0.95 - 1.91) | 0.10 | 1.32 (0.88 - 1.96) | 0.19 |
| **PS-stratification / on-treatment TAR without escalation** | | | | | | | | | | |
| CCAE | 20,123 | 70,373 | 10.8 | 44.5 | 89 | 219 | 1.77 (1.29 - 2.40) | <0.01 | 1.76 (1.28 - 2.40) | <0.01 |
| ODOD | 11,297 | 57,469 | 5.4 | 43.9 | 79 | 628 | 1.46 (1.08 - 1.94) | 0.01 | 1.50 (1.12 - 2.01) | <0.01 |
| MDCR | 1,208 | 16,711 | 0.7 | 13.7 | 11 | 167 | 2.66 (1.26 - 5.08) | <0.01 | 2.15 (1.07 - 4.32) | 0.04 |
| MDCD | 1,955 | 16,069 | 0.9 | 8.6 | 13 | 146 | 0.92 (0.46 - 1.69) | 0.80 | 0.90 (0.47 - 1.73) | 0.77 |
| Open Claims | 223,395 | 821,471 | 125.4 | 656.3 | 666 | 3,496 | 1.36 (1.22 - 1.50) | <0.01 | 1.37 (1.13 - 1.65) | <0.01 |
| OEHR | 13,292 | 80,965 | 3.4 | 38.8 | 28 | 413 | 1.01 (0.64 - 1.53) | 0.97 | 0.94 (0.58 - 1.52) | 0.81 |
| Meta-analysis | 268,107 | 1,030,278 | 145.0 | 783.4 | 862 | 4,756 | 1.38 (1.26 - 1.52) | <0.01 | 1.39 (1.14 - 1.70) | <0.01 |
| Meta-analysis (w/o USOC) | 44,712 | 208,807 | 19.6 | 127.1 | 196 | 1,260 | 1.42 (1.07 - 1.89) | 0.02 | 1.41 (0.97 - 2.06) | 0.08 |

##### 1.5.12 Acute pancreatitis

**Supplementary Table 116:** Exposures, outcomes and risk estimates for Acute pancreatitis in GLP1RA (female) (T) vs SU (female) (C) new-user cohorts under four different propensity score (PS) adjustment / time-at-risk (TAR) designs across data sources. We report total patient exposure time in 1,000 patient-year (KYr) units, uncalibrated and calibrated hazard ratios (HRs) and their 95% confidence intervals (CIs). Greyed out entries did not pass study diagnostics and are included only for completeness.

| | Patients | | Exposure (KYr) | | Outcomes | | Uncalibrated | | Calibrated | |
|---|---|---|---|---|---|---|---|---|---|---|
| | T | C | T | C | T | C | HR (95% CI) | $p$ | HR (95% CI) | $p$ |
| **PS-matching / on-treatment TAR** | | | | | | | | | | |
| CCAE | 15,250 | 68,961 | 9.5 | 51.1 | 15 | 119 | 0.56 (0.25 - 1.13) | 0.13 | 0.58 (0.27 - 1.27) | 0.19 |
| ODOD | 8,897 | 55,154 | 5.0 | 46.9 | 12 | 130 | 1.24 (0.53 - 2.65) | 0.61 | 1.27 (0.56 - 2.87) | 0.58 |
| MDCR | 1,093 | 13,410 | 0.7 | 12.2 | <5 | 44 | 1.71 (0.36 - 5.96) | 0.45 | 1.35 (0.33 - 5.49) | 0.68 |
| MDCD | 1,866 | 14,860 | 1.0 | 9.6 | 5 | 46 | 1.06 (0.28 - 3.14) | 0.93 | 1.00 (0.30 - 3.36) | 0.95 |
| Open Claims | 176,408 | 823,569 | 118.9 | 783.8 | 186 | 1,639 | 1.07 (0.87 - 1.31) | 0.50 | 1.08 (0.84 - 1.39) | 0.54 |
| OEHR | 11,103 | 77,881 | 3.4 | 44.0 | 9 | 94 | 1.22 (0.49 - 2.73) | 0.65 | 1.16 (0.49 - 2.74) | 0.73 |
| Meta-analysis | 213,524 | 1,040,425 | 137.8 | 935.3 | 227 | 2,028 | 1.05 (0.87 - 1.25) | 0.63 | 1.04 (0.81 - 1.35) | 0.74 |
| Meta-analysis (w/o USOC) | 37,116 | 216,856 | 18.9 | 151.5 | 41 | 389 | 0.93 (0.60 - 1.44) | 0.75 | 0.91 (0.58 - 1.45) | 0.70 |
| **PS-stratification / on-treatment TAR** | | | | | | | | | | |
| CCAE | 20,479 | 71,257 | 12.2 | 52.4 | 18 | 124 | 0.62 (0.34 - 1.08) | 0.11 | 0.61 (0.34 - 1.11) | 0.12 |
| ODOD | 11,601 | 59,795 | 6.2 | 51.8 | 16 | 155 | 1.20 (0.62 - 2.17) | 0.57 | 1.23 (0.66 - 2.29) | 0.53 |
| MDCR | 1,226 | 17,293 | 0.8 | 15.6 | <5 | 63 | 1.48 (0.39 - 4.53) | 0.53 | 1.24 (0.36 - 4.22) | 0.74 |
| MDCD | 2,010 | 16,533 | 1.1 | 10.8 | 5 | 59 | 1.30 (0.42 - 3.26) | 0.62 | 1.25 (0.45 - 3.48) | 0.67 |
| Open Claims | 228,988 | 840,593 | 142.8 | 800.1 | 217 | 1,714 | 0.97 (0.82 - 1.15) | 0.75 | 0.99 (0.79 - 1.26) | 0.92 |
| OEHR | 13,580 | 82,939 | 4.0 | 47.2 | 10 | 105 | 1.51 (0.70 - 2.94) | 0.26 | 1.41 (0.67 - 2.97) | 0.39 |
| Meta-analysis | 274,648 | 1,054,584 | 165.1 | 951.5 | 261 | 2,098 | 0.97 (0.84 - 1.13) | 0.73 | 0.99 (0.77 - 1.26) | 0.90 |
| Meta-analysis (w/o USOC) | 45,660 | 213,991 | 22.4 | 151.4 | 44 | 384 | 1.01 (0.59 - 1.71) | 0.97 | 1.00 (0.56 - 1.78) | 0.95 |
| **PS-matching / on-treatment TAR without escalation** | | | | | | | | | | |
| CCAE | 15,250 | 68,961 | 8.5 | 43.8 | 12 | 91 | 0.51 (0.20 - 1.11) | 0.12 | 0.53 (0.22 - 1.24) | 0.15 |
| ODOD | 8,897 | 55,154 | 4.5 | 41.6 | 11 | 107 | 1.45 (0.58 - 3.29) | 0.40 | 1.50 (0.63 - 3.58) | 0.37 |
| MDCR | 1,093 | 13,410 | 0.6 | 11.2 | <5 | 37 | 1.80 (0.38 - 6.50) | 0.42 | 1.42 (0.34 - 5.88) | 0.64 |
| MDCD | 1,866 | 14,860 | 0.9 | 7.9 | <5 | 40 | 0.75 (0.11 - 2.91) | 0.73 | 0.72 (0.14 - 3.67) | 0.69 |
| Open Claims | 176,408 | 823,569 | 106.1 | 659.0 | 164 | 1,337 | 1.12 (0.90 - 1.38) | 0.32 | 1.12 (0.87 - 1.45) | 0.39 |
| OEHR | 11,103 | 77,881 | 2.9 | 37.2 | 8 | 79 | 1.01 (0.35 - 2.51) | 0.98 | 0.94 (0.35 - 2.52) | 0.91 |
| Meta-analysis | 213,524 | 1,040,425 | 122.9 | 789.6 | <200 | 1,654 | 1.07 (0.88 - 1.31) | 0.48 | 1.07 (0.82 - 1.40) | 0.62 |
| Meta-analysis (w/o USOC) | 37,116 | 216,856 | 16.8 | 130.6 | <36 | 317 | 0.88 (0.54 - 1.44) | 0.61 | 0.86 (0.51 - 1.46) | 0.59 |
| **PS-stratification / on-treatment TAR without escalation** | | | | | | | | | | |
| CCAE | 20,479 | 71,257 | 11.0 | 45.1 | 15 | 96 | 0.64 (0.33 - 1.17) | 0.17 | 0.64 (0.34 - 1.20) | 0.17 |
| ODOD | 11,601 | 59,795 | 5.6 | 46.1 | 14 | 131 | 1.38 (0.67 - 2.66) | 0.36 | 1.42 (0.71 - 2.83) | 0.32 |
| MDCR | 1,226 | 17,293 | 0.7 | 14.2 | <5 | 54 | 1.54 (0.40 - 4.80) | 0.50 | 1.24 (0.36 - 4.32) | 0.74 |
| MDCD | 2,010 | 16,533 | 0.9 | 9.0 | <5 | 50 | 0.62 (0.10 - 2.22) | 0.55 | 0.61 (0.13 - 2.94) | 0.54 |
| Open Claims | 228,988 | 840,593 | 128.8 | 673.2 | 193 | 1,402 | 1.03 (0.86 - 1.23) | 0.72 | 1.04 (0.82 - 1.32) | 0.74 |
| OEHR | 13,580 | 82,939 | 3.5 | 40.0 | 9 | 88 | 1.52 (0.66 - 3.09) | 0.29 | 1.41 (0.64 - 3.14) | 0.41 |
| Meta-analysis | 274,648 | 1,054,584 | 148.9 | 804.4 | 231 | 1,717 | 1.04 (0.88 - 1.22) | 0.68 | 1.04 (0.82 - 1.33) | 0.74 |
| Meta-analysis (w/o USOC) | 45,660 | 213,991 | 20.1 | 131.2 | 38 | 315 | 1.07 (0.62 - 1.87) | 0.80 | 1.07 (0.58 - 1.96) | 0.83 |

##### 1.5.13 Bone fracture

**Supplementary Table 117:** Exposures, outcomes and risk estimates for Bone fracture in GLP1RA (female) (T) vs SU (female) (C) new-user cohorts under four different propensity score (PS) adjustment / time-at-risk (TAR) designs across data sources. We report total patient exposure time in 1,000 patient-year (KYr) units, uncalibrated and calibrated hazard ratios (HRs) and their 95% confidence intervals (CIs). Greyed out entries did not pass study diagnostics and are included only for completeness.

| | Patients | | Exposure (KYr) | | Outcomes | | Uncalibrated | | Calibrated | |
|---|---|---|---|---|---|---|---|---|---|---|
| | T | C | T | C | T | C | HR (95% CI) | $p$ | HR (95% CI) | $p$ |
| **PS-matching / on-treatment TAR** | | | | | | | | | | |
| CCAE | 13,821 | 63,626 | 8.4 | 46.3 | 169 | 968 | 0.96 (0.76 - 1.21) | 0.73 | 1.00 (0.75 - 1.34) | 0.89 |
| ODOD | 8,076 | 50,040 | 4.4 | 41.2 | 133 | 1,432 | 1.13 (0.88 - 1.44) | 0.33 | 1.16 (0.90 - 1.51) | 0.36 |
| MDCR | 941 | 11,539 | 0.6 | 10.2 | 22 | 479 | 1.12 (0.66 - 1.82) | 0.66 | 0.89 (0.53 - 1.49) | 0.66 |
| MDCD | 1,622 | 13,355 | 0.9 | 8.4 | 30 | 296 | 0.82 (0.47 - 1.36) | 0.46 | 0.78 (0.45 - 1.33) | 0.38 |
| Open Claims | 151,263 | 722,142 | 100.8 | 673.0 | 1,461 | 14,383 | 0.86 (0.79 - 0.92) | <0.01 | 0.86 (0.73 - 1.02) | 0.11 |
| OEHR | 10,253 | 72,315 | 3.1 | 39.8 | 42 | 982 | 0.65 (0.43 - 0.93) | 0.03 | 0.62 (0.42 - 0.90) | 0.02 |
| Meta-analysis | 185,976 | 933,017 | 118.2 | 818.8 | 1,857 | 18,540 | 0.91 (0.79 - 1.04) | 0.17 | 0.91 (0.72 - 1.14) | 0.41 |
| Meta-analysis (w/o USOC) | 34,713 | 210,875 | 17.4 | 145.8 | 396 | 4,157 | 0.94 (0.77 - 1.14) | 0.54 | 0.92 (0.72 - 1.18) | 0.55 |
| **PS-stratification / on-treatment TAR** | | | | | | | | | | |
| CCAE | 18,418 | 65,775 | 10.7 | 47.5 | 219 | 987 | 0.98 (0.82 - 1.16) | 0.81 | 0.96 (0.77 - 1.20) | 0.78 |
| ODOD | 10,453 | 53,994 | 5.4 | 45.3 | 161 | 1,660 | 1.01 (0.83 - 1.22) | 0.91 | 1.04 (0.85 - 1.26) | 0.74 |
| MDCR | 1,049 | 14,758 | 0.6 | 12.9 | 23 | 626 | 0.73 (0.45 - 1.12) | 0.17 | 0.61 (0.38 - 0.96) | 0.04 |
| MDCD | 1,744 | 14,781 | 0.9 | 9.4 | 33 | 331 | 1.05 (0.69 - 1.54) | 0.82 | 1.01 (0.68 - 1.51) | 0.87 |
| Open Claims | 194,843 | 737,097 | 120.3 | 686.7 | 1,758 | 15,021 | 0.84 (0.79 - 0.89) | <0.01 | 0.86 (0.72 - 1.03) | 0.10 |
| OEHR | 12,510 | 76,889 | 3.7 | 42.6 | 48 | 1,112 | 0.66 (0.48 - 0.89) | <0.01 | 0.62 (0.43 - 0.89) | 0.03 |
| Meta-analysis | 236,224 | 933,755 | 140.1 | 822.2 | 2,186 | 18,780 | 0.88 (0.77 - 1.01) | 0.07 | 0.89 (0.70 - 1.13) | 0.35 |
| Meta-analysis (w/o USOC) | 41,381 | 196,658 | 19.8 | 135.4 | 428 | 3,759 | 0.90 (0.73 - 1.11) | 0.33 | 0.89 (0.65 - 1.21) | 0.48 |
| **PS-matching / on-treatment TAR without escalation** | | | | | | | | | | |
| CCAE | 13,821 | 63,626 | 7.6 | 39.8 | 150 | 815 | 1.04 (0.81 - 1.32) | 0.78 | 1.07 (0.83 - 1.39) | 0.67 |
| ODOD | 8,076 | 50,040 | 4.0 | 36.7 | 114 | 1,268 | 1.04 (0.80 - 1.35) | 0.76 | 1.08 (0.82 - 1.41) | 0.64 |
| MDCR | 941 | 11,539 | 0.6 | 9.3 | 21 | 442 | 1.08 (0.61 - 1.81) | 0.78 | 0.85 (0.49 - 1.46) | 0.58 |
| MDCD | 1,622 | 13,355 | 0.7 | 7.0 | 27 | 247 | 0.80 (0.43 - 1.39) | 0.45 | 0.76 (0.42 - 1.37) | 0.38 |
| Open Claims | 151,263 | 722,142 | 90.0 | 566.6 | 1,266 | 12,096 | 0.84 (0.77 - 0.91) | <0.01 | 0.84 (0.72 - 0.99) | 0.05 |
| OEHR | 10,253 | 72,315 | 2.6 | 33.9 | 37 | 814 | 0.69 (0.45 - 1.03) | 0.08 | 0.65 (0.43 - 0.98) | 0.05 |
| Meta-analysis | 185,976 | 933,017 | 105.5 | 693.4 | 1,615 | 15,682 | 0.90 (0.79 - 1.02) | 0.10 | 0.90 (0.72 - 1.12) | 0.33 |
| Meta-analysis (w/o USOC) | 34,713 | 210,875 | 15.5 | 126.7 | 349 | 3,586 | 0.97 (0.83 - 1.13) | 0.68 | 0.95 (0.74 - 1.21) | 0.69 |
| **PS-stratification / on-treatment TAR without escalation** | | | | | | | | | | |
| CCAE | 18,418 | 65,775 | 9.7 | 41.0 | 196 | 832 | 0.98 (0.82 - 1.18) | 0.87 | 0.98 (0.81 - 1.18) | 0.83 |
| ODOD | 10,453 | 53,994 | 5.0 | 40.5 | 140 | 1,479 | 0.99 (0.80 - 1.21) | 0.92 | 1.02 (0.82 - 1.25) | 0.86 |
| MDCR | 1,049 | 14,758 | 0.6 | 11.8 | 22 | 574 | 0.71 (0.43 - 1.10) | 0.15 | 0.57 (0.36 - 0.92) | 0.03 |
| MDCD | 1,744 | 14,781 | 0.8 | 7.9 | 30 | 277 | 1.08 (0.69 - 1.62) | 0.74 | 1.06 (0.69 - 1.62) | 0.80 |
| Open Claims | 194,843 | 737,097 | 108.6 | 578.6 | 1,548 | 12,654 | 0.83 (0.78 - 0.88) | <0.01 | 0.84 (0.71 - 0.99) | 0.05 |
| OEHR | 12,510 | 76,889 | 3.2 | 36.3 | 43 | 927 | 0.68 (0.48 - 0.94) | 0.03 | 0.64 (0.43 - 0.95) | 0.05 |
| Meta-analysis | 236,224 | 933,755 | 126.5 | 696.4 | 1,927 | 15,892 | 0.88 (0.78 - 0.99) | 0.04 | 0.88 (0.71 - 1.10) | 0.26 |
| Meta-analysis (w/o USOC) | 41,381 | 196,658 | 17.9 | 117.8 | 379 | 3,238 | 0.91 (0.76 - 1.10) | 0.33 | 0.91 (0.67 - 1.23) | 0.55 |

##### 1.5.14 Diabetic ketoacidosis

**Supplementary Table 118:** Exposures, outcomes and risk estimates for Diabetic ketoacidosis in GLP1RA (female) (T) vs SU (female) (C) new-user cohorts under four different propensity score (PS) adjustment / time-at-risk (TAR) designs across data sources. We report total patient exposure time in 1,000 patient-year (KYr) units, uncalibrated and calibrated hazard ratios (HRs) and their 95% confidence intervals (CIs). Greyed out entries did not pass study diagnostics and are included only for completeness.

| | Patients | | Exposure (KYr) | | Outcomes | | Uncalibrated | | Calibrated | |
|---|---|---|---|---|---|---|---|---|---|---|
| | T | C | T | C | T | C | HR (95% CI) | $p$ | HR (95% CI) | $p$ |
| **PS-matching / on-treatment TAR** | | | | | | | | | | |
| CCAE | 15,323 | 69,477 | 9.5 | 51.5 | 5 | 59 | 0.52 (0.12 - 1.70) | 0.34 | 0.55 (0.14 - 2.11) | 0.39 |
| ODOD | 8,942 | 55,476 | 5.0 | 47.2 | 6 | 83 | 0.47 (0.13 - 1.30) | 0.19 | 0.48 (0.15 - 1.52) | 0.22 |
| MDCR | 1,098 | 13,499 | 0.7 | 12.4 | 0 | 11 | | | | |
| MDCD | 1,881 | 15,037 | 1.0 | 9.7 | <5 | 35 | 0.38 (0.02 - 2.22) | 0.42 | 0.36 (0.03 - 3.82) | 0.40 |
| Open Claims | 178,113 | 833,753 | 119.9 | 794.5 | 67 | 709 | 0.63 (0.46 - 0.86) | <0.01 | 0.64 (0.45 - 0.90) | 0.01 |
| OEHR | 11,136 | 78,175 | 3.4 | 44.2 | 5 | 60 | 0.92 (0.25 - 2.74) | 0.89 | 0.87 (0.26 - 2.92) | 0.82 |
| Meta-analysis | 213,514 | 1,036,881 | 137.9 | 937.4 | 83 | 911 | 0.63 (0.47 - 0.83) | <0.01 | 0.63 (0.45 - 0.88) | <0.01 |
| Meta-analysis (w/o USOC) | 35,401 | 203,128 | 17.9 | 142.9 | 16 | 202 | 0.61 (0.30 - 1.23) | 0.17 | 0.59 (0.29 - 1.22) | 0.16 |
| **PS-stratification / on-treatment TAR** | | | | | | | | | | |
| CCAE | 20,575 | 71,808 | 12.2 | 52.9 | 7 | 64 | 0.56 (0.22 - 1.27) | 0.20 | 0.55 (0.23 - 1.36) | 0.20 |
| ODOD | 11,653 | 60,263 | 6.2 | 52.3 | 7 | 98 | 0.60 (0.23 - 1.36) | 0.26 | 0.61 (0.25 - 1.50) | 0.29 |
| MDCR | 1,231 | 17,473 | 0.8 | 15.8 | 0 | 19 | | | | |
| MDCD | 2,026 | 16,782 | 1.1 | 11.0 | <5 | 39 | 0.34 (0.02 - 1.85) | 0.36 | 0.33 (0.03 - 3.31) | 0.34 |
| Open Claims | 230,974 | 850,481 | 143.9 | 810.3 | 78 | 732 | 0.59 (0.45 - 0.76) | <0.01 | 0.60 (0.44 - 0.82) | <0.01 |
| OEHR | 13,630 | 83,305 | 4.0 | 47.5 | 8 | 68 | 1.24 (0.49 - 2.79) | 0.62 | 1.16 (0.48 - 2.83) | 0.75 |
| Meta-analysis | 276,832 | 1,065,857 | 166.4 | 962.9 | 100 | 962 | 0.62 (0.49 - 0.79) | <0.01 | 0.63 (0.46 - 0.85) | <0.01 |
| Meta-analysis (w/o USOC) | 45,858 | 215,376 | 22.5 | 152.6 | 22 | 230 | 0.75 (0.45 - 1.25) | 0.28 | 0.75 (0.43 - 1.30) | 0.31 |
| **PS-matching / on-treatment TAR without escalation** | | | | | | | | | | |
| CCAE | 15,323 | 69,477 | 8.6 | 44.1 | <5 | 45 | 0.64 (0.14 - 2.23) | 0.53 | 0.66 (0.16 - 2.68) | 0.57 |
| ODOD | 8,942 | 55,476 | 4.5 | 41.9 | 5 | 64 | 0.48 (0.13 - 1.34) | 0.21 | 0.49 (0.16 - 1.56) | 0.23 |
| MDCR | 1,098 | 13,499 | 0.7 | 11.3 | 0 | 9 | | | | |
| MDCD | 1,881 | 15,037 | 0.9 | 8.0 | <5 | 23 | 0.57 (0.03 - 3.34) | 0.64 | 0.55 (0.05 - 5.83) | 0.62 |
| Open Claims | 178,113 | 833,753 | 107.0 | 667.9 | 59 | 525 | 0.70 (0.50 - 0.98) | 0.04 | 0.71 (0.49 - 1.02) | 0.07 |
| OEHR | 11,136 | 78,175 | 2.9 | 37.4 | <5 | 38 | 0.34 (0.02 - 1.94) | 0.36 | 0.31 (0.03 - 3.29) | 0.33 |
| Meta-analysis | 213,514 | 1,036,881 | 123.0 | 791.3 | <74 | 672 | 0.67 (0.49 - 0.92) | 0.01 | 0.67 (0.47 - 0.96) | 0.03 |
| Meta-analysis (w/o USOC) | 35,401 | 203,128 | 16.0 | 123.5 | <15 | 147 | 0.51 (0.22 - 1.16) | 0.11 | 0.50 (0.21 - 1.16) | 0.11 |
| **PS-stratification / on-treatment TAR without escalation** | | | | | | | | | | |
| CCAE | 20,575 | 71,808 | 11.1 | 45.4 | 5 | 50 | 0.53 (0.17 - 1.36) | 0.23 | 0.52 (0.18 - 1.49) | 0.23 |
| ODOD | 11,653 | 60,263 | 5.6 | 46.5 | 6 | 78 | 0.58 (0.20 - 1.44) | 0.28 | 0.60 (0.23 - 1.60) | 0.31 |
| MDCR | 1,231 | 17,473 | 0.7 | 14.4 | 0 | 16 | | | | |
| MDCD | 2,026 | 16,782 | 0.9 | 9.1 | <5 | 26 | 0.50 (0.03 - 2.81) | 0.56 | 0.49 (0.05 - 5.10) | 0.56 |
| Open Claims | 230,974 | 850,481 | 129.9 | 681.6 | 67 | 542 | 0.64 (0.47 - 0.85) | <0.01 | 0.64 (0.46 - 0.90) | 0.01 |
| OEHR | 13,630 | 83,305 | 3.5 | 40.2 | <5 | 44 | 0.80 (0.21 - 2.49) | 0.73 | 0.75 (0.21 - 2.66) | 0.66 |
| Meta-analysis | 276,832 | 1,065,857 | 150.1 | 813.8 | <83 | 714 | 0.63 (0.49 - 0.82) | <0.01 | 0.64 (0.46 - 0.88) | <0.01 |
| Meta-analysis (w/o USOC) | 45,858 | 215,376 | 20.2 | 132.2 | <16 | 172 | 0.61 (0.33 - 1.14) | 0.12 | 0.61 (0.31 - 1.18) | 0.15 |

##### 1.5.15 Diarrhea

**Supplementary Table 119:** Exposures, outcomes and risk estimates for Diarrhea in GLP1RA (female) (T) vs SU (female) (C) new-user cohorts under four different propensity score (PS) adjustment / time-at-risk (TAR) designs across data sources. We report total patient exposure time in 1,000 patient-year (KYr) units, uncalibrated and calibrated hazard ratios (HRs) and their 95% confidence intervals (CIs). Greyed out entries did not pass study diagnostics and are included only for completeness.

| | Patients | | Exposure (KYr) | | Outcomes | | Uncalibrated | | Calibrated | |
|---|---|---|---|---|---|---|---|---|---|---|
| | T | C | T | C | T | C | HR (95% CI) | $p$ | HR (95% CI) | $p$ |
| **PS-matching / on-treatment TAR** | | | | | | | | | | |
| CCAE | 12,969 | 59,928 | 7.8 | 43.4 | 354 | 1,537 | 1.06 (0.90 - 1.25) | 0.48 | 1.11 (0.87 - 1.40) | 0.51 |
| ODOD | 7,324 | 45,848 | 4.0 | 37.2 | 210 | 1,882 | 0.92 (0.76 - 1.11) | 0.38 | 0.94 (0.76 - 1.17) | 0.68 |
| MDCR | 949 | 11,402 | 0.6 | 10.1 | 27 | 440 | 1.09 (0.64 - 1.79) | 0.73 | 0.87 (0.52 - 1.46) | 0.61 |
| MDCD | 1,430 | 11,794 | 0.7 | 7.2 | 65 | 533 | 1.00 (0.70 - 1.41) | 0.98 | 0.96 (0.67 - 1.36) | 0.78 |
| Open Claims | 145,604 | 703,313 | 96.9 | 655.7 | 2,408 | 15,718 | 1.11 (1.05 - 1.18) | <0.01 | 1.13 (0.96 - 1.32) | 0.17 |
| OEHR | 9,403 | 67,296 | 2.8 | 35.7 | 162 | 1,892 | 1.28 (1.04 - 1.55) | 0.02 | 1.22 (0.99 - 1.49) | 0.08 |
| Meta-analysis | 177,679 | 899,581 | 112.8 | 789.2 | 3,226 | 22,002 | 1.09 (1.01 - 1.18) | 0.04 | 1.09 (0.89 - 1.33) | 0.41 |
| Meta-analysis (w/o USOC) | 32,075 | 196,268 | 15.9 | 133.6 | 818 | 6,284 | 1.06 (0.94 - 1.21) | 0.33 | 1.04 (0.85 - 1.27) | 0.71 |
| **PS-stratification / on-treatment TAR** | | | | | | | | | | |
| CCAE | 17,309 | 62,314 | 10.0 | 44.8 | 474 | 1,588 | 1.09 (0.96 - 1.24) | 0.16 | 1.08 (0.89 - 1.30) | 0.52 |
| ODOD | 9,492 | 49,358 | 4.9 | 40.7 | 264 | 2,104 | 0.92 (0.78 - 1.07) | 0.28 | 0.94 (0.80 - 1.10) | 0.51 |
| MDCR | 1,049 | 14,631 | 0.6 | 12.8 | 29 | 580 | 0.84 (0.55 - 1.25) | 0.42 | 0.70 (0.46 - 1.07) | 0.11 |
| MDCD | 1,534 | 13,053 | 0.8 | 8.1 | 73 | 591 | 1.08 (0.82 - 1.42) | 0.56 | 1.05 (0.79 - 1.38) | 0.75 |
| Open Claims | 186,885 | 719,371 | 115.2 | 670.5 | 3,059 | 16,351 | 1.10 (1.05 - 1.15) | <0.01 | 1.12 (0.95 - 1.34) | 0.20 |
| OEHR | 11,446 | 71,651 | 3.3 | 38.2 | 199 | 2,051 | 1.21 (1.02 - 1.43) | 0.02 | 1.13 (0.87 - 1.46) | 0.46 |
| Meta-analysis | 225,132 | 902,694 | 133.3 | 794.2 | 3,996 | 22,094 | 1.08 (0.99 - 1.18) | 0.07 | 1.09 (0.88 - 1.35) | 0.41 |
| Meta-analysis (w/o USOC) | 38,247 | 183,323 | 18.1 | 123.7 | 937 | 5,743 | 1.07 (0.92 - 1.24) | 0.39 | 1.06 (0.81 - 1.38) | 0.70 |
| **PS-matching / on-treatment TAR without escalation** | | | | | | | | | | |
| CCAE | 12,969 | 59,928 | 7.0 | 37.5 | 318 | 1,344 | 1.08 (0.90 - 1.28) | 0.40 | 1.11 (0.92 - 1.35) | 0.42 |
| ODOD | 7,324 | 45,848 | 3.6 | 33.4 | 199 | 1,698 | 0.95 (0.77 - 1.15) | 0.59 | 0.98 (0.80 - 1.20) | 0.80 |
| MDCR | 949 | 11,402 | 0.6 | 9.3 | 23 | 394 | 1.11 (0.62 - 1.86) | 0.71 | 0.87 (0.50 - 1.51) | 0.64 |
| MDCD | 1,430 | 11,794 | 0.7 | 6.0 | 56 | 449 | 1.01 (0.68 - 1.46) | 0.98 | 0.96 (0.65 - 1.42) | 0.79 |
| Open Claims | 145,604 | 703,313 | 86.7 | 555.3 | 2,143 | 13,288 | 1.13 (1.06 - 1.21) | <0.01 | 1.14 (0.98 - 1.33) | 0.11 |
| OEHR | 9,403 | 67,296 | 2.4 | 30.8 | 145 | 1,518 | 1.36 (1.10 - 1.69) | <0.01 | 1.27 (1.02 - 1.58) | 0.05 |
| Meta-analysis | 177,679 | 899,581 | 100.9 | 672.3 | 2,884 | 18,691 | 1.11 (1.02 - 1.22) | 0.02 | 1.11 (0.91 - 1.36) | 0.31 |
| Meta-analysis (w/o USOC) | 32,075 | 196,268 | 14.2 | 117.0 | 741 | 5,403 | 1.10 (0.95 - 1.26) | 0.21 | 1.07 (0.84 - 1.36) | 0.60 |
| **PS-stratification / on-treatment TAR without escalation** | | | | | | | | | | |
| CCAE | 17,309 | 62,314 | 9.1 | 38.8 | 426 | 1,391 | 1.09 (0.95 - 1.24) | 0.22 | 1.08 (0.94 - 1.24) | 0.34 |
| ODOD | 9,492 | 49,358 | 4.5 | 36.6 | 249 | 1,909 | 0.93 (0.79 - 1.10) | 0.41 | 0.96 (0.81 - 1.13) | 0.67 |
| MDCR | 1,049 | 14,631 | 0.6 | 11.8 | 25 | 520 | 0.87 (0.54 - 1.32) | 0.53 | 0.70 (0.45 - 1.10) | 0.14 |
| MDCD | 1,534 | 13,053 | 0.7 | 6.8 | 64 | 500 | 1.10 (0.81 - 1.47) | 0.53 | 1.08 (0.80 - 1.46) | 0.65 |
| Open Claims | 186,885 | 719,371 | 104.1 | 568.4 | 2,767 | 13,842 | 1.12 (1.07 - 1.18) | <0.01 | 1.13 (0.96 - 1.34) | 0.14 |
| OEHR | 11,446 | 71,651 | 2.9 | 33.1 | 180 | 1,645 | 1.38 (1.15 - 1.64) | <0.01 | 1.29 (0.97 - 1.70) | 0.15 |
| Meta-analysis | 225,132 | 902,694 | 120.6 | 676.9 | 3,622 | 18,787 | 1.12 (0.97 - 1.28) | 0.11 | 1.12 (0.90 - 1.41) | 0.31 |
| Meta-analysis (w/o USOC) | 38,247 | 183,323 | 16.4 | 108.5 | 855 | 4,945 | 1.12 (0.91 - 1.37) | 0.29 | 1.11 (0.81 - 1.53) | 0.54 |

##### 1.5.16 Genitourinary infection

**Supplementary Table 120:** Exposures, outcomes and risk estimates for Genitourinary infection in GLP1RA (female) (T) vs SU (female) (C) new-user cohorts under four different propensity score (PS) adjustment / time-at-risk (TAR) designs across data sources. We report total patient exposure time in 1,000 patient-year (KYr) units, uncalibrated and calibrated hazard ratios (HRs) and their 95% confidence intervals (CIs). Greyed out entries did not pass study diagnostics and are included only for completeness.

| | Patients | | Exposure (KYr) | | Outcomes | | Uncalibrated | | Calibrated | |
|---|---|---|---|---|---|---|---|---|---|---|
| | T | C | T | C | T | C | HR (95% CI) | $p$ | HR (95% CI) | $p$ |
| **PS-matching / on-treatment TAR** | | | | | | | | | | |
| CCAE | 10,631 | 49,762 | 6.2 | 35.3 | 451 | 2,355 | 1.05 (0.90 - 1.21) | 0.55 | 1.09 (0.87 - 1.36) | 0.56 |
| ODOD | 6,206 | 37,882 | 3.3 | 29.5 | 302 | 2,889 | 1.01 (0.86 - 1.18) | 0.95 | 1.03 (0.86 - 1.25) | 0.79 |
| MDCR | 773 | 9,213 | 0.5 | 8.0 | 39 | 711 | 1.02 (0.66 - 1.54) | 0.92 | 0.81 (0.53 - 1.24) | 0.36 |
| MDCD | 1,206 | 9,894 | 0.6 | 6.0 | 70 | 615 | 1.07 (0.76 - 1.48) | 0.71 | 1.01 (0.72 - 1.42) | 0.82 |
| Open Claims | 113,858 | 564,876 | 75.7 | 522.7 | 3,094 | 25,854 | 0.97 (0.92 - 1.02) | 0.25 | 0.98 (0.84 - 1.15) | 0.82 |
| OEHR | 8,488 | 60,875 | 2.4 | 31.8 | 208 | 2,409 | 1.30 (1.08 - 1.56) | <0.01 | 1.24 (1.03 - 1.49) | 0.04 |
| Meta-analysis | 141,162 | 732,502 | 88.8 | 633.3 | 4,164 | 34,833 | 1.05 (0.95 - 1.16) | 0.34 | 1.05 (0.85 - 1.29) | 0.66 |
| Meta-analysis (w/o USOC) | 27,304 | 167,626 | 13.1 | 110.6 | 1,070 | 8,979 | 1.09 (0.98 - 1.20) | 0.10 | 1.07 (0.89 - 1.28) | 0.55 |
| **PS-stratification / on-treatment TAR** | | | | | | | | | | |
| CCAE | 14,042 | 51,428 | 7.9 | 36.3 | 598 | 2,428 | 1.07 (0.96 - 1.19) | 0.22 | 1.05 (0.88 - 1.26) | 0.63 |
| ODOD | 7,975 | 40,393 | 4.0 | 32.0 | 361 | 3,269 | 0.97 (0.85 - 1.11) | 0.69 | 1.00 (0.87 - 1.14) | 0.83 |
| MDCR | 849 | 12,248 | 0.5 | 10.5 | 41 | 1,003 | 0.89 (0.62 - 1.24) | 0.51 | 0.74 (0.52 - 1.05) | 0.12 |
| MDCD | 1,310 | 10,858 | 0.7 | 6.6 | 78 | 707 | 1.10 (0.84 - 1.43) | 0.46 | 1.06 (0.82 - 1.39) | 0.67 |
| Open Claims | 144,467 | 578,203 | 89.2 | 534.8 | 3,719 | 27,149 | 0.98 (0.94 - 1.02) | 0.36 | 1.00 (0.84 - 1.19) | 0.90 |
| OEHR | 10,291 | 64,267 | 2.9 | 33.7 | 243 | 2,686 | 1.33 (1.14 - 1.54) | <0.01 | 1.24 (0.97 - 1.59) | 0.20 |
| Meta-analysis | 176,775 | 734,291 | 104.0 | 636.8 | 4,921 | 35,532 | 1.07 (0.94 - 1.22) | 0.32 | 1.08 (0.85 - 1.37) | 0.51 |
| Meta-analysis (w/o USOC) | 32,308 | 156,088 | 14.8 | 102.0 | 1,202 | 8,383 | 1.11 (0.94 - 1.31) | 0.21 | 1.10 (0.83 - 1.45) | 0.53 |
| **PS-matching / on-treatment TAR without escalation** | | | | | | | | | | |
| CCAE | 10,631 | 49,762 | 5.6 | 30.7 | 405 | 2,027 | 1.03 (0.88 - 1.20) | 0.69 | 1.07 (0.90 - 1.27) | 0.60 |
| ODOD | 6,206 | 37,882 | 3.0 | 26.6 | 269 | 2,603 | 0.98 (0.82 - 1.16) | 0.79 | 1.01 (0.85 - 1.20) | 0.83 |
| MDCR | 773 | 9,213 | 0.4 | 7.3 | 30 | 638 | 0.89 (0.54 - 1.40) | 0.63 | 0.70 (0.43 - 1.13) | 0.17 |
| MDCD | 1,206 | 9,894 | 0.6 | 5.1 | 55 | 512 | 1.00 (0.68 - 1.45) | 0.98 | 0.96 (0.66 - 1.40) | 0.79 |
| Open Claims | 113,858 | 564,876 | 67.7 | 443.5 | 2,711 | 22,003 | 0.96 (0.91 - 1.02) | 0.16 | 0.97 (0.83 - 1.12) | 0.67 |
| OEHR | 8,488 | 60,875 | 2.1 | 27.5 | 174 | 1,969 | 1.33 (1.08 - 1.63) | <0.01 | 1.24 (1.01 - 1.52) | 0.07 |
| Meta-analysis | 141,162 | 732,502 | 79.4 | 540.7 | 3,644 | 29,752 | 1.03 (0.93 - 1.14) | 0.60 | 1.03 (0.83 - 1.27) | 0.81 |
| Meta-analysis (w/o USOC) | 27,304 | 167,626 | 11.8 | 97.2 | 933 | 7,749 | 1.07 (0.94 - 1.21) | 0.34 | 1.04 (0.83 - 1.32) | 0.75 |
| **PS-stratification / on-treatment TAR without escalation** | | | | | | | | | | |
| CCAE | 14,042 | 51,428 | 7.2 | 31.6 | 547 | 2,092 | 1.08 (0.96 - 1.21) | 0.20 | 1.07 (0.95 - 1.21) | 0.35 |
| ODOD | 7,975 | 40,393 | 3.7 | 28.9 | 324 | 2,959 | 0.96 (0.84 - 1.11) | 0.61 | 0.99 (0.86 - 1.14) | 0.82 |
| MDCR | 849 | 12,248 | 0.5 | 9.6 | 32 | 901 | 0.74 (0.49 - 1.07) | 0.13 | 0.60 (0.40 - 0.89) | 0.01 |
| MDCD | 1,310 | 10,858 | 0.6 | 5.6 | 63 | 595 | 1.05 (0.78 - 1.40) | 0.74 | 1.03 (0.77 - 1.39) | 0.79 |
| Open Claims | 144,467 | 578,203 | 80.5 | 454.3 | 3,307 | 23,155 | 0.98 (0.94 - 1.02) | 0.29 | 0.99 (0.84 - 1.16) | 0.86 |
| OEHR | 10,291 | 64,267 | 2.5 | 29.2 | 206 | 2,200 | 1.35 (1.15 - 1.59) | <0.01 | 1.26 (0.96 - 1.66) | 0.18 |
| Meta-analysis | 176,775 | 734,291 | 94.0 | 543.9 | 4,384 | 30,406 | 1.07 (0.93 - 1.24) | 0.34 | 1.08 (0.86 - 1.36) | 0.51 |
| Meta-analysis (w/o USOC) | 32,308 | 156,088 | 13.4 | 89.6 | 1,077 | 7,251 | 1.11 (0.93 - 1.33) | 0.23 | 1.11 (0.82 - 1.50) | 0.52 |

##### 1.5.17 Hyperkalemia

**Supplementary Table 121:** Exposures, outcomes and risk estimates for Hyperkalemia in GLP1RA (female) (T) vs SU (female) (C) new-user cohorts under four different propensity score (PS) adjustment / time-at-risk (TAR) designs across data sources. We report total patient exposure time in 1,000 patient-year (KYr) units, uncalibrated and calibrated hazard ratios (HRs) and their 95% confidence intervals (CIs). Greyed out entries did not pass study diagnostics and are included only for completeness.

| | Patients | | Exposure (KYr) | | Outcomes | | Uncalibrated | | Calibrated | |
|---|---|---|---|---|---|---|---|---|---|---|
| | T | C | T | C | T | C | HR (95% CI) | $p$ | HR (95% CI) | $p$ |
| **PS-matching / on-treatment TAR** | | | | | | | | | | |
| CCAE | 15,116 | 68,606 | 9.4 | 50.6 | 41 | 379 | 0.63 (0.39 - 0.99) | 0.05 | 0.66 (0.40 - 1.08) | 0.13 |
| ODOD | 8,696 | 53,210 | 4.9 | 44.3 | 60 | 937 | 0.71 (0.49 - 1.01) | 0.07 | 0.73 (0.50 - 1.07) | 0.15 |
| MDCR | 1,084 | 13,259 | 0.7 | 12.0 | 5 | 155 | 0.71 (0.20 - 1.90) | 0.54 | 0.56 (0.18 - 1.72) | 0.32 |
| MDCD | 1,842 | 14,702 | 1.0 | 9.3 | 8 | 148 | 0.45 (0.15 - 1.11) | 0.12 | 0.43 (0.16 - 1.18) | 0.10 |
| Open Claims | 175,343 | 817,461 | 117.8 | 771.8 | 368 | 5,803 | 0.66 (0.57 - 0.76) | <0.01 | 0.67 (0.55 - 0.82) | <0.01 |
| OEHR | 10,986 | 76,869 | 3.3 | 42.9 | 26 | 530 | 0.98 (0.59 - 1.56) | 0.93 | 0.93 (0.57 - 1.52) | 0.78 |
| Meta-analysis | 213,067 | 1,044,107 | 137.0 | 930.9 | 508 | 7,952 | 0.68 (0.60 - 0.77) | <0.01 | 0.68 (0.54 - 0.84) | <0.01 |
| Meta-analysis (w/o USOC) | 37,724 | 226,646 | 19.3 | 159.1 | 140 | 2,149 | 0.72 (0.57 - 0.92) | <0.01 | 0.71 (0.54 - 0.94) | 0.02 |
| **PS-stratification / on-treatment TAR** | | | | | | | | | | |
| CCAE | 20,285 | 70,883 | 12.0 | 51.9 | 49 | 390 | 0.61 (0.43 - 0.85) | <0.01 | 0.60 (0.41 - 0.87) | 0.01 |
| ODOD | 11,338 | 57,592 | 6.0 | 48.8 | 67 | 1,125 | 0.78 (0.58 - 1.02) | 0.07 | 0.80 (0.60 - 1.05) | 0.12 |
| MDCR | 1,208 | 16,970 | 0.7 | 15.2 | 6 | 220 | 0.58 (0.22 - 1.29) | 0.23 | 0.48 (0.20 - 1.18) | 0.11 |
| MDCD | 1,985 | 16,325 | 1.1 | 10.5 | 8 | 179 | 0.42 (0.18 - 0.87) | 0.03 | 0.41 (0.19 - 0.89) | 0.03 |
| Open Claims | 227,579 | 833,943 | 141.5 | 787.1 | 427 | 6,214 | 0.66 (0.59 - 0.74) | <0.01 | 0.68 (0.55 - 0.83) | <0.01 |
| OEHR | 13,436 | 81,722 | 4.0 | 45.9 | 30 | 642 | 0.88 (0.58 - 1.29) | 0.53 | 0.82 (0.52 - 1.29) | 0.43 |
| Meta-analysis | 272,638 | 1,044,140 | 163.5 | 933.6 | 573 | 8,371 | 0.68 (0.62 - 0.75) | <0.01 | 0.69 (0.55 - 0.86) | <0.01 |
| Meta-analysis (w/o USOC) | 45,059 | 210,197 | 22.0 | 146.5 | 146 | 2,157 | 0.74 (0.61 - 0.90) | <0.01 | 0.73 (0.55 - 0.99) | 0.05 |
| **PS-matching / on-treatment TAR without escalation** | | | | | | | | | | |
| CCAE | 15,116 | 68,606 | 8.4 | 43.4 | 36 | 335 | 0.59 (0.36 - 0.95) | 0.04 | 0.61 (0.37 - 1.01) | 0.07 |
| ODOD | 8,696 | 53,210 | 4.4 | 39.4 | 57 | 810 | 0.79 (0.53 - 1.13) | 0.21 | 0.81 (0.56 - 1.18) | 0.30 |
| MDCR | 1,084 | 13,259 | 0.6 | 11.0 | 5 | 140 | 0.79 (0.22 - 2.15) | 0.68 | 0.62 (0.20 - 1.93) | 0.42 |
| MDCD | 1,842 | 14,702 | 0.9 | 7.7 | 8 | 128 | 0.53 (0.17 - 1.33) | 0.22 | 0.51 (0.18 - 1.41) | 0.20 |
| Open Claims | 175,343 | 817,461 | 105.1 | 649.4 | 318 | 4,817 | 0.64 (0.55 - 0.75) | <0.01 | 0.65 (0.52 - 0.80) | <0.01 |
| OEHR | 10,986 | 76,869 | 2.9 | 36.5 | 24 | 389 | 1.10 (0.64 - 1.81) | 0.72 | 1.02 (0.61 - 1.73) | 0.89 |
| Meta-analysis | 213,067 | 1,044,107 | 122.3 | 787.4 | 448 | 6,619 | 0.70 (0.59 - 0.83) | <0.01 | 0.70 (0.54 - 0.89) | <0.01 |
| Meta-analysis (w/o USOC) | 37,724 | 226,646 | 17.2 | 138.0 | 130 | 1,802 | 0.77 (0.60 - 0.99) | 0.04 | 0.76 (0.55 - 1.03) | 0.10 |
| **PS-stratification / on-treatment TAR without escalation** | | | | | | | | | | |
| CCAE | 20,285 | 70,883 | 10.9 | 44.6 | 44 | 345 | 0.60 (0.41 - 0.86) | <0.01 | 0.60 (0.41 - 0.87) | <0.01 |
| ODOD | 11,338 | 57,592 | 5.4 | 43.5 | 64 | 982 | 0.82 (0.61 - 1.09) | 0.19 | 0.85 (0.63 - 1.13) | 0.28 |
| MDCR | 1,208 | 16,970 | 0.7 | 13.9 | 6 | 192 | 0.63 (0.23 - 1.41) | 0.31 | 0.51 (0.21 - 1.25) | 0.15 |
| MDCD | 1,985 | 16,325 | 0.9 | 8.8 | 8 | 156 | 0.46 (0.20 - 0.96) | 0.06 | 0.46 (0.21 - 1.01) | 0.06 |
| Open Claims | 227,579 | 833,943 | 127.7 | 662.8 | 374 | 5,176 | 0.66 (0.58 - 0.74) | <0.01 | 0.66 (0.54 - 0.81) | <0.01 |
| OEHR | 13,436 | 81,722 | 3.4 | 39.1 | 28 | 474 | 0.99 (0.63 - 1.49) | 0.95 | 0.92 (0.57 - 1.48) | 0.75 |
| Meta-analysis | 272,638 | 1,044,140 | 147.5 | 790.0 | 510 | 6,977 | 0.72 (0.60 - 0.86) | <0.01 | 0.72 (0.56 - 0.93) | 0.01 |
| Meta-analysis (w/o USOC) | 45,059 | 210,197 | 19.8 | 127.2 | 136 | 1,801 | 0.78 (0.60 - 1.01) | 0.06 | 0.78 (0.54 - 1.11) | 0.18 |

##### 1.5.18 Hypoglycemia

**Supplementary Table 122:** Exposures, outcomes and risk estimates for Hypoglycemia in GLP1RA (female) (T) vs SU (female) (C) new-user cohorts under four different propensity score (PS) adjustment / time-at-risk (TAR) designs across data sources. We report total patient exposure time in 1,000 patient-year (KYr) units, uncalibrated and calibrated hazard ratios (HRs) and their 95% confidence intervals (CIs). Greyed out entries did not pass study diagnostics and are included only for completeness.

| | Patients | | Exposure (KYr) | | Outcomes | | Uncalibrated | | Calibrated | |
|---|---|---|---|---|---|---|---|---|---|---|
| | T | C | T | C | T | C | HR (95% CI) | $p$ | HR (95% CI) | $p$ |
| **PS-matching / on-treatment TAR** | | | | | | | | | | |
| CCAE | 14,974 | 68,551 | 9.3 | 50.7 | 46 | 535 | 0.37 (0.25 - 0.54) | <0.01 | 0.39 (0.26 - 0.59) | <0.01 |
| ODOD | 8,791 | 54,693 | 4.9 | 46.0 | 37 | 1,105 | 0.31 (0.20 - 0.46) | <0.01 | 0.32 (0.21 - 0.49) | <0.01 |
| MDCR | 1,083 | 13,413 | 0.7 | 12.2 | <5 | 238 | 0.09 (0.01 - 0.42) | 0.03 | 0.07 (0.01 - 0.65) | 0.02 |
| MDCD | 1,823 | 14,730 | 1.0 | 9.3 | 9 | 288 | 0.21 (0.06 - 0.51) | <0.01 | 0.20 (0.07 - 0.56) | <0.01 |
| Open Claims | 174,559 | 821,394 | 117.4 | 779.2 | 326 | 8,103 | 0.31 (0.27 - 0.36) | <0.01 | 0.32 (0.26 - 0.39) | <0.01 |
| OEHR | 10,983 | 77,456 | 3.3 | 43.3 | 26 | 828 | 0.34 (0.20 - 0.53) | <0.01 | 0.32 (0.20 - 0.52) | <0.01 |
| Meta-analysis | 212,213 | 1,050,237 | 136.7 | 940.7 | <449 | 11,097 | 0.32 (0.28 - 0.36) | <0.01 | 0.32 (0.26 - 0.40) | <0.01 |
| Meta-analysis (w/o USOC) | 37,654 | 228,843 | 19.3 | 161.5 | <123 | 2,994 | 0.33 (0.26 - 0.41) | <0.01 | 0.32 (0.24 - 0.42) | <0.01 |
| **PS-stratification / on-treatment TAR** | | | | | | | | | | |
| CCAE | 19,900 | 70,862 | 11.8 | 52.0 | 60 | 560 | 0.36 (0.27 - 0.49) | <0.01 | 0.36 (0.26 - 0.50) | <0.01 |
| ODOD | 11,352 | 59,401 | 6.0 | 50.9 | 56 | 1,322 | 0.42 (0.30 - 0.56) | <0.01 | 0.43 (0.31 - 0.58) | <0.01 |
| MDCR | 1,212 | 17,322 | 0.8 | 15.5 | <5 | 352 | 0.13 (0.02 - 0.41) | <0.01 | 0.11 (0.02 - 0.47) | <0.01 |
| MDCD | 1,953 | 16,411 | 1.1 | 10.6 | 9 | 341 | 0.30 (0.14 - 0.57) | <0.01 | 0.29 (0.14 - 0.58) | <0.01 |
| Open Claims | 225,048 | 837,735 | 140.3 | 794.4 | 401 | 8,599 | 0.32 (0.29 - 0.36) | <0.01 | 0.33 (0.27 - 0.40) | <0.01 |
| OEHR | 13,388 | 82,491 | 3.9 | 46.4 | 31 | 934 | 0.38 (0.26 - 0.56) | <0.01 | 0.36 (0.23 - 0.55) | <0.01 |
| Meta-analysis | 269,688 | 1,050,489 | 162.1 | 943.8 | 548 | 11,415 | 0.35 (0.31 - 0.40) | <0.01 | 0.35 (0.28 - 0.45) | <0.01 |
| Meta-analysis (w/o USOC) | 44,640 | 212,754 | 21.8 | 149.4 | 147 | 2,816 | 0.39 (0.32 - 0.47) | <0.01 | 0.38 (0.29 - 0.51) | <0.01 |
| **PS-matching / on-treatment TAR without escalation** | | | | | | | | | | |
| CCAE | 14,974 | 68,551 | 8.4 | 43.5 | 40 | 444 | 0.39 (0.25 - 0.57) | <0.01 | 0.40 (0.26 - 0.61) | <0.01 |
| ODOD | 8,791 | 54,693 | 4.5 | 40.9 | 28 | 962 | 0.26 (0.15 - 0.40) | <0.01 | 0.26 (0.16 - 0.43) | <0.01 |
| MDCR | 1,083 | 13,413 | 0.6 | 11.1 | <5 | 212 | 0.09 (0.01 - 0.43) | 0.03 | 0.07 (0.01 - 0.66) | 0.02 |
| MDCD | 1,823 | 14,730 | 0.9 | 7.8 | 7 | 231 | 0.20 (0.05 - 0.55) | <0.01 | 0.19 (0.06 - 0.65) | <0.01 |
| Open Claims | 174,559 | 821,394 | 104.8 | 655.8 | 268 | 6,678 | 0.29 (0.25 - 0.34) | <0.01 | 0.29 (0.24 - 0.36) | <0.01 |
| OEHR | 10,983 | 77,456 | 2.9 | 36.8 | 19 | 610 | 0.31 (0.17 - 0.53) | <0.01 | 0.29 (0.16 - 0.51) | <0.01 |
| Meta-analysis | 212,213 | 1,050,237 | 122.0 | 795.9 | <367 | 9,137 | 0.29 (0.26 - 0.34) | <0.01 | 0.29 (0.23 - 0.37) | <0.01 |
| Meta-analysis (w/o USOC) | 37,654 | 228,843 | 17.2 | 140.1 | <99 | 2,459 | 0.31 (0.24 - 0.40) | <0.01 | 0.30 (0.22 - 0.42) | <0.01 |
| **PS-stratification / on-treatment TAR without escalation** | | | | | | | | | | |
| CCAE | 19,900 | 70,862 | 10.7 | 44.7 | 52 | 468 | 0.36 (0.25 - 0.49) | <0.01 | 0.35 (0.26 - 0.49) | <0.01 |
| ODOD | 11,352 | 59,401 | 5.5 | 45.4 | 46 | 1,161 | 0.38 (0.27 - 0.52) | <0.01 | 0.39 (0.28 - 0.54) | <0.01 |
| MDCR | 1,212 | 17,322 | 0.7 | 14.2 | <5 | 315 | 0.14 (0.02 - 0.44) | <0.01 | 0.11 (0.02 - 0.49) | <0.01 |
| MDCD | 1,953 | 16,411 | 0.9 | 8.8 | 7 | 276 | 0.27 (0.11 - 0.56) | <0.01 | 0.27 (0.12 - 0.60) | <0.01 |
| Open Claims | 225,048 | 837,735 | 126.6 | 669.1 | 337 | 7,127 | 0.31 (0.27 - 0.35) | <0.01 | 0.31 (0.26 - 0.38) | <0.01 |
| OEHR | 13,388 | 82,491 | 3.4 | 39.5 | 24 | 687 | 0.36 (0.23 - 0.55) | <0.01 | 0.34 (0.21 - 0.55) | <0.01 |
| Meta-analysis | 269,688 | 1,050,489 | 146.2 | 798.8 | 459 | 9,443 | 0.33 (0.29 - 0.37) | <0.01 | 0.33 (0.27 - 0.41) | <0.01 |
| Meta-analysis (w/o USOC) | 44,640 | 212,754 | 19.6 | 129.6 | 122 | 2,316 | 0.37 (0.30 - 0.45) | <0.01 | 0.36 (0.26 - 0.50) | <0.01 |

##### 1.5.19 Hypotension

**Supplementary Table 123:** Exposures, outcomes and risk estimates for Hypotension in GLP1RA (female) (T) vs SU (female) (C) new-user cohorts under four different propensity score (PS) adjustment / time-at-risk (TAR) designs across data sources. We report total patient exposure time in 1,000 patient-year (KYr) units, uncalibrated and calibrated hazard ratios (HRs) and their 95% confidence intervals (CIs). Greyed out entries did not pass study diagnostics and are included only for completeness.

| | Patients | | Exposure (KYr) | | Outcomes | | Uncalibrated | | Calibrated | |
|---|---|---|---|---|---|---|---|---|---|---|
| | T | C | T | C | T | C | HR (95% CI) | $p$ | HR (95% CI) | $p$ |
| **PS-matching / on-treatment TAR** | | | | | | | | | | |
| CCAE | 15,039 | 68,260 | 9.3 | 50.5 | 78 | 356 | 1.37 (0.96 - 1.94) | 0.08 | 1.43 (0.97 - 2.11) | 0.11 |
| ODOD | 8,607 | 53,283 | 4.8 | 44.7 | 81 | 915 | 0.86 (0.62 - 1.18) | 0.37 | 0.89 (0.64 - 1.24) | 0.54 |
| MDCR | 1,052 | 12,843 | 0.7 | 11.6 | 10 | 216 | 1.00 (0.46 - 1.97) | 1.00 | 0.79 (0.38 - 1.64) | 0.54 |
| MDCD | 1,780 | 14,216 | 0.9 | 9.0 | 28 | 235 | 1.29 (0.75 - 2.15) | 0.34 | 1.23 (0.72 - 2.08) | 0.47 |
| Open Claims | 172,226 | 805,415 | 115.6 | 762.8 | 761 | 6,533 | 1.07 (0.96 - 1.18) | 0.21 | 1.08 (0.90 - 1.30) | 0.42 |
| OEHR | 10,930 | 76,582 | 3.3 | 42.7 | 56 | 612 | 1.79 (1.24 - 2.55) | <0.01 | 1.71 (1.19 - 2.45) | <0.01 |
| Meta-analysis | 209,634 | 1,030,599 | 134.6 | 921.4 | 1,014 | 8,867 | 1.19 (0.95 - 1.48) | 0.12 | 1.19 (0.89 - 1.58) | 0.24 |
| Meta-analysis (w/o USOC) | 37,408 | 225,184 | 19.0 | 158.6 | 253 | 2,334 | 1.24 (0.93 - 1.66) | 0.15 | 1.21 (0.87 - 1.69) | 0.27 |
| **PS-stratification / on-treatment TAR** | | | | | | | | | | |
| CCAE | 20,175 | 70,624 | 11.9 | 51.9 | 104 | 375 | 1.10 (0.83 - 1.43) | 0.51 | 1.08 (0.80 - 1.47) | 0.65 |
| ODOD | 11,209 | 57,318 | 5.9 | 49.0 | 103 | 1,103 | 1.04 (0.81 - 1.32) | 0.78 | 1.06 (0.83 - 1.36) | 0.66 |
| MDCR | 1,176 | 16,652 | 0.7 | 14.9 | 10 | 323 | 0.94 (0.44 - 1.78) | 0.86 | 0.78 (0.39 - 1.57) | 0.50 |
| MDCD | 1,922 | 15,691 | 1.0 | 10.1 | 28 | 287 | 1.34 (0.85 - 2.05) | 0.20 | 1.29 (0.83 - 2.02) | 0.28 |
| Open Claims | 223,218 | 822,404 | 138.6 | 778.9 | 939 | 6,995 | 1.04 (0.96 - 1.13) | 0.29 | 1.07 (0.89 - 1.29) | 0.50 |
| OEHR | 13,381 | 81,467 | 3.9 | 45.7 | 60 | 717 | 1.65 (1.21 - 2.20) | <0.01 | 1.54 (1.08 - 2.20) | 0.04 |
| Meta-analysis | 267,983 | 1,031,813 | 160.4 | 925.5 | 1,206 | 9,190 | 1.15 (0.95 - 1.39) | 0.14 | 1.17 (0.89 - 1.53) | 0.27 |
| Meta-analysis (w/o USOC) | 44,765 | 209,409 | 21.8 | 146.6 | 267 | 2,195 | 1.22 (0.93 - 1.60) | 0.16 | 1.21 (0.85 - 1.72) | 0.31 |
| **PS-matching / on-treatment TAR without escalation** | | | | | | | | | | |
| CCAE | 15,039 | 68,260 | 8.4 | 43.3 | 67 | 293 | 1.34 (0.92 - 1.95) | 0.13 | 1.39 (0.95 - 2.04) | 0.13 |
| ODOD | 8,607 | 53,283 | 4.4 | 39.8 | 71 | 802 | 0.92 (0.64 - 1.28) | 0.61 | 0.94 (0.67 - 1.33) | 0.76 |
| MDCR | 1,052 | 12,843 | 0.6 | 10.6 | 10 | 192 | 1.18 (0.54 - 2.36) | 0.65 | 0.93 (0.44 - 1.95) | 0.84 |
| MDCD | 1,780 | 14,216 | 0.8 | 7.5 | 23 | 178 | 1.26 (0.68 - 2.22) | 0.44 | 1.21 (0.67 - 2.18) | 0.55 |
| Open Claims | 172,226 | 805,415 | 103.2 | 641.6 | 646 | 5,405 | 1.04 (0.93 - 1.17) | 0.45 | 1.05 (0.88 - 1.26) | 0.61 |
| OEHR | 10,930 | 76,582 | 2.8 | 36.3 | 44 | 423 | 2.21 (1.44 - 3.32) | <0.01 | 2.05 (1.35 - 3.12) | <0.01 |
| Meta-analysis | 209,634 | 1,030,599 | 120.2 | 779.2 | 861 | 7,293 | 1.24 (0.96 - 1.62) | 0.10 | 1.24 (0.90 - 1.71) | 0.19 |
| Meta-analysis (w/o USOC) | 37,408 | 225,184 | 17.0 | 137.6 | 215 | 1,888 | 1.33 (0.95 - 1.85) | 0.10 | 1.30 (0.88 - 1.91) | 0.20 |
| **PS-stratification / on-treatment TAR without escalation** | | | | | | | | | | |
| CCAE | 20,175 | 70,624 | 10.8 | 44.6 | 93 | 311 | 1.08 (0.80 - 1.45) | 0.60 | 1.08 (0.80 - 1.45) | 0.65 |
| ODOD | 11,209 | 57,318 | 5.4 | 43.6 | 93 | 972 | 1.02 (0.78 - 1.32) | 0.87 | 1.05 (0.81 - 1.36) | 0.73 |
| MDCR | 1,176 | 16,652 | 0.7 | 13.6 | 10 | 285 | 1.11 (0.52 - 2.12) | 0.78 | 0.89 (0.44 - 1.81) | 0.76 |
| MDCD | 1,922 | 15,691 | 0.9 | 8.4 | 23 | 226 | 1.34 (0.80 - 2.16) | 0.26 | 1.31 (0.80 - 2.17) | 0.31 |
| Open Claims | 223,218 | 822,404 | 125.1 | 655.8 | 809 | 5,814 | 1.03 (0.94 - 1.13) | 0.48 | 1.04 (0.87 - 1.25) | 0.66 |
| OEHR | 13,381 | 81,467 | 3.4 | 39.0 | 47 | 501 | 1.92 (1.35 - 2.66) | <0.01 | 1.79 (1.19 - 2.67) | 0.01 |
| Meta-analysis | 267,983 | 1,031,813 | 144.7 | 783.0 | 1,042 | 7,598 | 1.19 (0.91 - 1.55) | 0.21 | 1.20 (0.87 - 1.65) | 0.28 |
| Meta-analysis (w/o USOC) | 44,765 | 209,409 | 19.6 | 127.2 | 233 | 1,784 | 1.27 (0.88 - 1.83) | 0.21 | 1.26 (0.81 - 1.96) | 0.31 |

##### 1.5.20 Joint pain

**Supplementary Table 124:** Exposures, outcomes and risk estimates for Joint pain in GLP1RA (female) (T) vs SU (female) (C) new-user cohorts under four different propensity score (PS) adjustment / time-at-risk (TAR) designs across data sources. We report total patient exposure time in 1,000 patient-year (KYr) units, uncalibrated and calibrated hazard ratios (HRs) and their 95% confidence intervals (CIs). Greyed out entries did not pass study diagnostics and are included only for completeness.

| | Patients | | Exposure (KYr) | | Outcomes | | Uncalibrated | | Calibrated | |
|---|---|---|---|---|---|---|---|---|---|---|
| | T | C | T | C | T | C | HR (95% CI) | $p$ | HR (95% CI) | $p$ |
| **PS-matching / on-treatment TAR** | | | | | | | | | | |
| CCAE | 14,119 | 65,351 | 8.7 | 48.1 | 126 | 624 | 0.98 (0.73 - 1.29) | 0.87 | 1.02 (0.73 - 1.42) | 0.89 |
| ODOD | 8,139 | 51,604 | 4.5 | 43.4 | 76 | 737 | 0.75 (0.53 - 1.03) | 0.09 | 0.77 (0.54 - 1.09) | 0.19 |
| MDCR | 1,016 | 12,901 | 0.7 | 11.8 | <5 | 102 | 0.27 (0.01 - 1.32) | 0.25 | 0.21 (0.02 - 2.00) | 0.18 |
| MDCD | 1,652 | 13,655 | 0.9 | 8.6 | 26 | 207 | 1.25 (0.71 - 2.11) | 0.41 | 1.19 (0.69 - 2.05) | 0.55 |
| Open Claims | 161,845 | 777,235 | 108.7 | 738.2 | 878 | 6,030 | 0.89 (0.80 - 0.98) | 0.02 | 0.90 (0.75 - 1.08) | 0.27 |
| OEHR | 10,237 | 73,432 | 3.1 | 40.6 | 50 | 700 | 0.73 (0.49 - 1.07) | 0.11 | 0.70 (0.47 - 1.03) | 0.08 |
| Meta-analysis | 197,008 | 994,178 | 126.5 | 890.8 | <1,161 | 8,400 | 0.88 (0.81 - 0.96) | <0.01 | 0.88 (0.72 - 1.08) | 0.23 |
| Meta-analysis (w/o USOC) | 35,163 | 216,943 | 17.8 | 152.6 | <283 | 2,370 | 0.87 (0.70 - 1.07) | 0.18 | 0.85 (0.65 - 1.10) | 0.25 |
| **PS-stratification / on-treatment TAR** | | | | | | | | | | |
| CCAE | 18,670 | 67,542 | 11.0 | 49.4 | 172 | 634 | 1.03 (0.84 - 1.25) | 0.80 | 1.01 (0.79 - 1.29) | 0.89 |
| ODOD | 10,449 | 56,251 | 5.5 | 48.3 | 109 | 815 | 0.90 (0.70 - 1.14) | 0.38 | 0.92 (0.72 - 1.17) | 0.52 |
| MDCR | 1,137 | 16,702 | 0.7 | 15.0 | 5 | 144 | 0.61 (0.21 - 1.47) | 0.33 | 0.51 (0.19 - 1.36) | 0.19 |
| MDCD | 1,769 | 15,272 | 0.9 | 9.8 | 28 | 230 | 1.26 (0.80 - 1.94) | 0.30 | 1.22 (0.78 - 1.90) | 0.40 |
| Open Claims | 207,745 | 794,699 | 129.4 | 754.5 | 1,119 | 6,218 | 0.92 (0.85 - 0.99) | 0.03 | 0.94 (0.78 - 1.13) | 0.51 |
| OEHR | 12,432 | 78,546 | 3.6 | 43.8 | 66 | 742 | 0.90 (0.67 - 1.18) | 0.46 | 0.84 (0.59 - 1.18) | 0.40 |
| Meta-analysis | 249,296 | 997,038 | 149.4 | 896.1 | 1,466 | 8,409 | 0.93 (0.87 - 0.99) | 0.03 | 0.94 (0.76 - 1.15) | 0.54 |
| Meta-analysis (w/o USOC) | 41,551 | 202,339 | 20.1 | 141.6 | 347 | 2,191 | 0.95 (0.83 - 1.09) | 0.49 | 0.94 (0.72 - 1.23) | 0.68 |
| **PS-matching / on-treatment TAR without escalation** | | | | | | | | | | |
| CCAE | 14,119 | 65,351 | 7.8 | 41.3 | 111 | 536 | 0.98 (0.73 - 1.32) | 0.92 | 1.02 (0.75 - 1.39) | 0.88 |
| ODOD | 8,139 | 51,604 | 4.1 | 38.6 | 63 | 660 | 0.72 (0.50 - 1.02) | 0.07 | 0.74 (0.52 - 1.07) | 0.13 |
| MDCR | 1,016 | 12,901 | 0.6 | 10.7 | <5 | 92 | 0.27 (0.02 - 1.36) | 0.26 | 0.21 (0.02 - 2.04) | 0.18 |
| MDCD | 1,652 | 13,655 | 0.8 | 7.2 | 24 | 174 | 1.53 (0.85 - 2.67) | 0.15 | 1.47 (0.83 - 2.62) | 0.21 |
| Open Claims | 161,845 | 777,235 | 96.9 | 621.6 | 776 | 5,043 | 0.91 (0.81 - 1.01) | 0.08 | 0.91 (0.77 - 1.09) | 0.34 |
| OEHR | 10,237 | 73,432 | 2.6 | 34.6 | 41 | 564 | 0.75 (0.48 - 1.14) | 0.20 | 0.70 (0.46 - 1.08) | 0.12 |
| Meta-analysis | 197,008 | 994,178 | 112.8 | 754.2 | <1,020 | 7,069 | 0.90 (0.82 - 0.99) | 0.03 | 0.90 (0.73 - 1.11) | 0.32 |
| Meta-analysis (w/o USOC) | 35,163 | 216,943 | 15.9 | 132.5 | <244 | 2,026 | 0.89 (0.68 - 1.18) | 0.43 | 0.88 (0.62 - 1.23) | 0.46 |
| **PS-stratification / on-treatment TAR without escalation** | | | | | | | | | | |
| CCAE | 18,670 | 67,542 | 10.0 | 42.5 | 150 | 545 | 0.98 (0.79 - 1.22) | 0.88 | 0.98 (0.78 - 1.22) | 0.84 |
| ODOD | 10,449 | 56,251 | 5.0 | 43.1 | 94 | 734 | 0.82 (0.62 - 1.06) | 0.13 | 0.84 (0.64 - 1.09) | 0.21 |
| MDCR | 1,137 | 16,702 | 0.7 | 13.7 | 5 | 130 | 0.63 (0.21 - 1.53) | 0.37 | 0.51 (0.19 - 1.38) | 0.19 |
| MDCD | 1,769 | 15,272 | 0.8 | 8.2 | 26 | 193 | 1.31 (0.81 - 2.07) | 0.26 | 1.29 (0.80 - 2.07) | 0.31 |
| Open Claims | 207,745 | 794,699 | 116.6 | 636.0 | 1,006 | 5,207 | 0.92 (0.85 - 1.00) | 0.06 | 0.93 (0.78 - 1.11) | 0.46 |
| OEHR | 12,432 | 78,546 | 3.2 | 37.4 | 57 | 602 | 0.90 (0.66 - 1.21) | 0.51 | 0.84 (0.58 - 1.22) | 0.43 |
| Meta-analysis | 249,296 | 997,038 | 134.7 | 758.9 | 1,307 | 7,088 | 0.92 (0.86 - 0.99) | 0.02 | 0.93 (0.77 - 1.12) | 0.43 |
| Meta-analysis (w/o USOC) | 41,551 | 202,339 | 18.1 | 123.0 | 301 | 1,881 | 0.91 (0.78 - 1.05) | 0.20 | 0.90 (0.68 - 1.20) | 0.50 |

##### 1.5.21 Lower extremity amputation

**Supplementary Table 125:** Exposures, outcomes and risk estimates for Lower extremity amputation in GLP1RA (female) (T) vs SU (female) (C) new-user cohorts under four different propensity score (PS) adjustment / time-at-risk (TAR) designs across data sources. We report total patient exposure time in 1,000 patient-year (KYr) units, uncalibrated and calibrated hazard ratios (HRs) and their 95% confidence intervals (CIs). Greyed out entries did not pass study diagnostics and are included only for completeness.

| | Patients | | Exposure (KYr) | | Outcomes | | Uncalibrated | | Calibrated | |
|---|---|---|---|---|---|---|---|---|---|---|
| | T | C | T | C | T | C | HR (95% CI) | $p$ | HR (95% CI) | $p$ |
| **PS-matching / on-treatment TAR** | | | | | | | | | | |
| CCAE | 15,332 | 69,524 | 9.5 | 51.6 | <5 | 16 | 0.67 (0.03 - 4.47) | 0.75 | 0.70 (0.06 - 8.14) | 0.77 |
| ODOD | 8,941 | 55,511 | 5.0 | 47.2 | <5 | 27 | 0.00 (0.00 - 1.12) | 0.22 | 0.00 (0.00 - 131900) | 0.22 |
| MDCR | 1,097 | 13,489 | 0.7 | 12.3 | 0 | 10 | | | | |
| MDCD | 1,884 | 15,079 | 1.0 | 9.7 | <5 | 7 | 0.87 (0.04 - 12.4) | 0.93 | 0.83 (0.05 - 15.3) | 0.90 |
| Open Claims | 178,234 | 834,411 | 120.0 | 794.8 | 21 | 310 | 0.91 (0.48 - 1.60) | 0.75 | 0.92 (0.49 - 1.71) | 0.78 |
| OEHR | 11,141 | 78,091 | 3.4 | 44.1 | 0 | 24 | | | | |
| Meta-analysis | 178,234 | 834,411 | 120.0 | 794.8 | 21 | 310 | 0.91 (0.48 - 1.60) | 0.75 | 0.90 (0.48 - 1.70) | 0.76 |
| **PS-stratification / on-treatment TAR** | | | | | | | | | | |
| CCAE | 20,584 | 71,863 | 12.2 | 52.9 | <5 | 16 | 1.03 (0.23 - 3.45) | 0.96 | 1.02 (0.26 - 4.01) | 0.97 |
| ODOD | 11,653 | 60,303 | 6.2 | 52.3 | <5 | 39 | 0.64 (0.03 - 3.46) | 0.70 | 0.65 (0.07 - 6.59) | 0.72 |
| MDCR | 1,230 | 17,455 | 0.8 | 15.7 | 0 | 12 | | | | |
| MDCD | 2,031 | 16,825 | 1.1 | 11.0 | <5 | 8 | 1.51 (0.07 - 12.7) | 0.76 | 1.46 (0.10 - 20.3) | 0.78 |
| Open Claims | 231,103 | 851,361 | 144.0 | 810.7 | 22 | 331 | 0.73 (0.44 - 1.15) | 0.20 | 0.75 (0.45 - 1.24) | 0.26 |
| OEHR | 13,630 | 83,320 | 4.0 | 47.4 | 0 | 28 | | | | |
| Meta-analysis | 242,756 | 911,664 | 150.2 | 863.0 | <27 | 370 | 0.73 (0.46 - 1.16) | 0.18 | 0.74 (0.44 - 1.22) | 0.24 |
| Meta-analysis (w/o USOC) | 11,653 | 60,303 | 6.2 | 52.3 | <5 | 39 | 0.64 (0.03 - 3.46) | 0.70 | 0.63 (0.06 - 6.43) | 0.70 |
| **PS-matching / on-treatment TAR without escalation** | | | | | | | | | | |
| CCAE | 15,332 | 69,524 | 8.6 | 44.2 | <5 | 13 | 0.79 (0.04 - 5.78) | 0.86 | 0.82 (0.07 - 10.1) | 0.88 |
| ODOD | 8,941 | 55,511 | 4.5 | 41.9 | <5 | 20 | | | | |
| MDCR | 1,097 | 13,489 | 0.7 | 11.2 | 0 | 6 | | | | |
| MDCD | 1,884 | 15,079 | 0.9 | 8.0 | <5 | <5 | 3.00 (0.09 - 99.1) | 0.54 | 2.87 (0.09 - 95.0) | 0.55 |
| Open Claims | 178,234 | 834,411 | 107.1 | 667.9 | 15 | 233 | 0.70 (0.31 - 1.40) | 0.35 | 0.71 (0.33 - 1.51) | 0.37 |
| OEHR | 11,141 | 78,091 | 2.9 | 37.4 | 0 | 13 | | | | |
| Meta-analysis | 178,234 | 834,411 | 107.1 | 667.9 | 15 | 233 | 0.70 (0.31 - 1.40) | 0.35 | 0.70 (0.32 - 1.51) | 0.37 |
| **PS-stratification / on-treatment TAR without escalation** | | | | | | | | | | |
| CCAE | 20,584 | 71,863 | 11.1 | 45.4 | <5 | 13 | 1.34 (0.28 - 4.77) | 0.69 | 1.33 (0.32 - 5.48) | 0.69 |
| ODOD | 11,653 | 60,303 | 5.6 | 46.5 | <5 | 32 | 0.68 (0.04 - 3.80) | 0.75 | 0.70 (0.07 - 7.19) | 0.76 |
| MDCR | 1,230 | 17,455 | 0.7 | 14.4 | 0 | 8 | | | | |
| MDCD | 2,031 | 16,825 | 0.9 | 9.1 | <5 | 5 | 2.68 (0.09 - 38.0) | 0.52 | 2.64 (0.13 - 53.4) | 0.53 |
| Open Claims | 231,103 | 851,361 | 129.9 | 681.8 | 16 | 252 | 0.68 (0.37 - 1.15) | 0.18 | 0.69 (0.38 - 1.23) | 0.21 |
| OEHR | 13,630 | 83,320 | 3.5 | 40.2 | 0 | 16 | | | | |
| Meta-analysis | 242,756 | 911,664 | 135.5 | 728.3 | <21 | 284 | 0.68 (0.39 - 1.17) | 0.16 | 0.68 (0.38 - 1.21) | 0.19 |
| Meta-analysis (w/o USOC) | 11,653 | 60,303 | 5.6 | 46.5 | <5 | 32 | 0.68 (0.04 - 3.80) | 0.75 | 0.68 (0.07 - 7.05) | 0.75 |

##### 1.5.22 Nausea

**Supplementary Table 126:** Exposures, outcomes and risk estimates for Nausea in GLP1RA (female) (T) vs SU (female) (C) new-user cohorts under four different propensity score (PS) adjustment / time-at-risk (TAR) designs across data sources. We report total patient exposure time in 1,000 patient-year (KYr) units, uncalibrated and calibrated hazard ratios (HRs) and their 95% confidence intervals (CIs). Greyed out entries did not pass study diagnostics and are included only for completeness.

| | Patients | | Exposure (KYr) | | Outcomes | | Uncalibrated | | Calibrated | |
|---|---|---|---|---|---|---|---|---|---|---|
| | T | C | T | C | T | C | HR (95% CI) | $p$ | HR (95% CI) | $p$ |
| **PS-matching / on-treatment TAR** | | | | | | | | | | |
| CCAE | 12,637 | 59,138 | 7.6 | 43.5 | 449 | 1,793 | 1.32 (1.14 - 1.54) | <0.01 | 1.38 (1.10 - 1.73) | 0.03 |
| ODOD | 7,071 | 45,139 | 3.8 | 37.3 | 346 | 2,104 | 1.49 (1.28 - 1.74) | <0.01 | 1.54 (1.28 - 1.85) | <0.01 |
| MDCR | 990 | 11,463 | 0.6 | 10.3 | 36 | 433 | 1.07 (0.65 - 1.69) | 0.78 | 0.85 (0.53 - 1.37) | 0.52 |
| MDCD | 1,197 | 10,014 | 0.6 | 6.3 | 96 | 628 | 1.36 (1.01 - 1.81) | 0.04 | 1.29 (0.96 - 1.73) | 0.12 |
| Open Claims | 138,061 | 683,664 | 92.6 | 649.2 | 3,666 | 18,327 | 1.47 (1.40 - 1.55) | <0.01 | 1.49 (1.27 - 1.74) | <0.01 |
| OEHR | 9,169 | 67,499 | 2.7 | 36.3 | 225 | 2,024 | 1.47 (1.23 - 1.75) | <0.01 | 1.40 (1.17 - 1.68) | <0.01 |
| Meta-analysis | 169,125 | 876,917 | 108.0 | 782.8 | 4,818 | 25,309 | 1.45 (1.39 - 1.52) | <0.01 | 1.45 (1.20 - 1.75) | <0.01 |
| Meta-analysis (w/o USOC) | 31,064 | 193,253 | 15.4 | 133.6 | 1,152 | 6,982 | 1.40 (1.28 - 1.53) | <0.01 | 1.37 (1.15 - 1.64) | <0.01 |
| **PS-stratification / on-treatment TAR** | | | | | | | | | | |
| CCAE | 16,677 | 61,481 | 9.6 | 44.9 | 634 | 1,853 | 1.36 (1.22 - 1.52) | <0.01 | 1.34 (1.12 - 1.61) | <0.01 |
| ODOD | 9,066 | 48,730 | 4.7 | 41.0 | 450 | 2,384 | 1.43 (1.26 - 1.62) | <0.01 | 1.47 (1.29 - 1.67) | <0.01 |
| MDCR | 1,099 | 14,996 | 0.7 | 13.2 | 37 | 606 | 1.12 (0.76 - 1.62) | 0.55 | 0.94 (0.64 - 1.37) | 0.74 |
| MDCD | 1,275 | 11,261 | 0.7 | 7.2 | 106 | 727 | 1.33 (1.05 - 1.67) | 0.02 | 1.28 (1.02 - 1.62) | 0.06 |
| Open Claims | 175,888 | 699,022 | 109.5 | 663.8 | 4,651 | 19,013 | 1.38 (1.33 - 1.44) | <0.01 | 1.41 (1.19 - 1.68) | <0.01 |
| OEHR | 11,092 | 71,754 | 3.2 | 38.8 | 290 | 2,206 | 1.45 (1.26 - 1.67) | <0.01 | 1.35 (1.06 - 1.72) | 0.07 |
| Meta-analysis | 212,723 | 880,987 | 127.0 | 788.6 | 6,025 | 25,456 | 1.39 (1.34 - 1.44) | <0.01 | 1.41 (1.15 - 1.72) | <0.01 |
| Meta-analysis (w/o USOC) | 36,835 | 181,965 | 17.4 | 124.8 | 1,374 | 6,443 | 1.41 (1.31 - 1.51) | <0.01 | 1.39 (1.10 - 1.77) | <0.01 |
| **PS-matching / on-treatment TAR without escalation** | | | | | | | | | | |
| CCAE | 12,637 | 59,138 | 6.9 | 37.6 | 410 | 1,506 | 1.34 (1.14 - 1.57) | <0.01 | 1.38 (1.16 - 1.65) | 0.01 |
| ODOD | 7,071 | 45,139 | 3.5 | 33.4 | 322 | 1,866 | 1.58 (1.34 - 1.86) | <0.01 | 1.63 (1.38 - 1.93) | <0.01 |
| MDCR | 990 | 11,463 | 0.6 | 9.4 | 31 | 386 | 1.22 (0.72 - 1.97) | 0.44 | 0.96 (0.58 - 1.59) | 0.82 |
| MDCD | 1,197 | 10,014 | 0.5 | 5.3 | 88 | 528 | 1.35 (0.98 - 1.85) | 0.06 | 1.30 (0.94 - 1.78) | 0.14 |
| Open Claims | 138,061 | 683,664 | 82.9 | 550.0 | 3,298 | 15,134 | 1.55 (1.47 - 1.64) | <0.01 | 1.56 (1.35 - 1.81) | <0.01 |
| OEHR | 9,169 | 67,499 | 2.4 | 31.3 | 196 | 1,608 | 1.51 (1.24 - 1.83) | <0.01 | 1.41 (1.15 - 1.72) | <0.01 |
| Meta-analysis | 169,125 | 876,917 | 96.7 | 667.0 | 4,345 | 21,028 | 1.51 (1.41 - 1.61) | <0.01 | 1.50 (1.24 - 1.82) | <0.01 |
| Meta-analysis (w/o USOC) | 31,064 | 193,253 | 13.8 | 117.0 | 1,047 | 5,894 | 1.45 (1.32 - 1.59) | <0.01 | 1.42 (1.14 - 1.75) | <0.01 |
| **PS-stratification / on-treatment TAR without escalation** | | | | | | | | | | |
| CCAE | 16,677 | 61,481 | 8.7 | 38.9 | 584 | 1,562 | 1.44 (1.28 - 1.62) | <0.01 | 1.43 (1.27 - 1.62) | <0.01 |
| ODOD | 9,066 | 48,730 | 4.3 | 36.8 | 420 | 2,124 | 1.49 (1.30 - 1.70) | <0.01 | 1.53 (1.33 - 1.75) | <0.01 |
| MDCR | 1,099 | 14,996 | 0.6 | 12.1 | 32 | 539 | 1.09 (0.71 - 1.61) | 0.68 | 0.88 (0.59 - 1.32) | 0.56 |
| MDCD | 1,275 | 11,261 | 0.6 | 6.1 | 97 | 621 | 1.42 (1.10 - 1.81) | <0.01 | 1.39 (1.08 - 1.79) | 0.02 |
| Open Claims | 175,888 | 699,022 | 99.0 | 563.0 | 4,235 | 15,739 | 1.47 (1.41 - 1.53) | <0.01 | 1.48 (1.26 - 1.74) | <0.01 |
| OEHR | 11,092 | 71,754 | 2.8 | 33.6 | 253 | 1,758 | 1.55 (1.33 - 1.80) | <0.01 | 1.44 (1.11 - 1.88) | 0.03 |
| Meta-analysis | 212,723 | 880,987 | 114.8 | 672.2 | 5,492 | 21,183 | 1.47 (1.42 - 1.53) | <0.01 | 1.48 (1.24 - 1.78) | <0.01 |
| Meta-analysis (w/o USOC) | 36,835 | 181,965 | 15.8 | 109.3 | 1,257 | 5,444 | 1.48 (1.37 - 1.60) | <0.01 | 1.48 (1.14 - 1.90) | <0.01 |

##### 1.5.23 Peripheral edema

**Supplementary Table 127:** Exposures, outcomes and risk estimates for Peripheral edema in GLP1RA (female) (T) vs SU (female) (C) new-user cohorts under four different propensity score (PS) adjustment / time-at-risk (TAR) designs across data sources. We report total patient exposure time in 1,000 patient-year (KYr) units, uncalibrated and calibrated hazard ratios (HRs) and their 95% confidence intervals (CIs). Greyed out entries did not pass study diagnostics and are included only for completeness.

| | Patients | | Exposure (KYr) | | Outcomes | | Uncalibrated | | Calibrated | |
|---|---|---|---|---|---|---|---|---|---|---|
| | T | C | T | C | T | C | HR (95% CI) | $p$ | HR (95% CI) | $p$ |
| **PS-matching / on-treatment TAR** | | | | | | | | | | |
| CCAE | 11,776 | 55,745 | 7.1 | 39.7 | 350 | 2,266 | 0.93 (0.79 - 1.08) | 0.34 | 0.96 (0.76 - 1.22) | 0.81 |
| ODOD | 6,523 | 41,013 | 3.5 | 32.0 | 208 | 3,009 | 0.69 (0.57 - 0.83) | <0.01 | 0.71 (0.57 - 0.88) | 0.02 |
| MDCR | 816 | 9,763 | 0.5 | 8.4 | 27 | 795 | 0.46 (0.27 - 0.76) | <0.01 | 0.37 (0.22 - 0.62) | <0.01 |
| MDCD | 1,192 | 9,639 | 0.6 | 5.6 | 57 | 597 | 0.94 (0.66 - 1.30) | 0.70 | 0.89 (0.63 - 1.26) | 0.54 |
| Open Claims | 126,791 | 627,042 | 84.2 | 573.5 | 2,132 | 23,997 | 0.77 (0.73 - 0.82) | <0.01 | 0.78 (0.66 - 0.92) | <0.01 |
| OEHR | 8,807 | 64,056 | 2.6 | 33.2 | 126 | 2,285 | 0.83 (0.66 - 1.04) | 0.11 | 0.79 (0.63 - 0.99) | 0.06 |
| Meta-analysis | 155,905 | 807,258 | 98.6 | 692.4 | 2,900 | 32,949 | 0.79 (0.71 - 0.88) | <0.01 | 0.79 (0.64 - 0.98) | 0.03 |
| Meta-analysis (w/o USOC) | 29,114 | 180,216 | 14.4 | 118.9 | 768 | 8,952 | 0.79 (0.66 - 0.95) | 0.01 | 0.77 (0.61 - 0.98) | 0.05 |
| **PS-stratification / on-treatment TAR** | | | | | | | | | | |
| CCAE | 15,438 | 57,760 | 8.9 | 40.8 | 445 | 2,347 | 0.85 (0.75 - 0.96) | <0.01 | 0.84 (0.70 - 1.01) | 0.12 |
| ODOD | 8,359 | 44,279 | 4.3 | 35.1 | 261 | 3,509 | 0.70 (0.60 - 0.81) | <0.01 | 0.72 (0.62 - 0.83) | <0.01 |
| MDCR | 890 | 12,728 | 0.5 | 10.7 | 33 | 1,107 | 0.58 (0.39 - 0.83) | <0.01 | 0.48 (0.33 - 0.71) | <0.01 |
| MDCD | 1,281 | 11,097 | 0.7 | 6.6 | 60 | 726 | 0.96 (0.71 - 1.27) | 0.76 | 0.92 (0.69 - 1.23) | 0.61 |
| Open Claims | 160,813 | 641,277 | 99.2 | 586.1 | 2,532 | 25,278 | 0.77 (0.73 - 0.81) | <0.01 | 0.79 (0.66 - 0.94) | <0.01 |
| OEHR | 10,621 | 68,077 | 3.0 | 35.5 | 147 | 2,577 | 0.87 (0.72 - 1.04) | 0.13 | 0.81 (0.62 - 1.06) | 0.23 |
| Meta-analysis | 195,231 | 811,393 | 115.5 | 697.5 | 3,385 | 33,711 | 0.79 (0.73 - 0.85) | <0.01 | 0.80 (0.64 - 0.98) | 0.03 |
| Meta-analysis (w/o USOC) | 34,418 | 170,116 | 16.2 | 111.4 | 853 | 8,433 | 0.80 (0.70 - 0.92) | <0.01 | 0.79 (0.61 - 1.03) | 0.10 |
| **PS-matching / on-treatment TAR without escalation** | | | | | | | | | | |
| CCAE | 11,776 | 55,745 | 6.4 | 34.4 | 312 | 1,949 | 0.93 (0.79 - 1.10) | 0.43 | 0.97 (0.80 - 1.16) | 0.78 |
| ODOD | 6,523 | 41,013 | 3.2 | 28.7 | 189 | 2,667 | 0.69 (0.56 - 0.84) | <0.01 | 0.71 (0.58 - 0.87) | <0.01 |
| MDCR | 816 | 9,763 | 0.5 | 7.7 | 26 | 725 | 0.49 (0.28 - 0.81) | <0.01 | 0.39 (0.23 - 0.66) | <0.01 |
| MDCD | 1,192 | 9,639 | 0.6 | 4.8 | 46 | 508 | 0.85 (0.57 - 1.23) | 0.39 | 0.81 (0.55 - 1.19) | 0.31 |
| Open Claims | 126,791 | 627,042 | 75.2 | 486.2 | 1,837 | 20,118 | 0.77 (0.72 - 0.82) | <0.01 | 0.77 (0.66 - 0.90) | <0.01 |
| OEHR | 8,807 | 64,056 | 2.2 | 28.8 | 115 | 1,862 | 0.91 (0.72 - 1.15) | 0.46 | 0.85 (0.67 - 1.08) | 0.24 |
| Meta-analysis | 155,905 | 807,258 | 88.0 | 590.6 | 2,525 | 27,829 | 0.80 (0.71 - 0.90) | <0.01 | 0.79 (0.64 - 0.99) | 0.04 |
| Meta-analysis (w/o USOC) | 29,114 | 180,216 | 12.9 | 104.5 | 688 | 7,711 | 0.80 (0.67 - 0.96) | 0.02 | 0.78 (0.60 - 1.02) | 0.09 |
| **PS-stratification / on-treatment TAR without escalation** | | | | | | | | | | |
| CCAE | 15,438 | 57,760 | 8.1 | 35.5 | 398 | 2,024 | 0.85 (0.75 - 0.97) | 0.01 | 0.85 (0.74 - 0.96) | 0.03 |
| ODOD | 8,359 | 44,279 | 3.9 | 31.6 | 239 | 3,128 | 0.73 (0.62 - 0.85) | <0.01 | 0.75 (0.64 - 0.88) | <0.01 |
| MDCR | 890 | 12,728 | 0.5 | 9.9 | 32 | 1,016 | 0.62 (0.42 - 0.90) | 0.02 | 0.50 (0.34 - 0.74) | <0.01 |
| MDCD | 1,281 | 11,097 | 0.6 | 5.6 | 49 | 621 | 0.85 (0.61 - 1.16) | 0.32 | 0.84 (0.61 - 1.16) | 0.32 |
| Open Claims | 160,813 | 641,277 | 89.5 | 497.3 | 2,215 | 21,268 | 0.76 (0.72 - 0.80) | <0.01 | 0.77 (0.65 - 0.91) | <0.01 |
| OEHR | 10,621 | 68,077 | 2.6 | 30.8 | 135 | 2,106 | 0.99 (0.81 - 1.20) | 0.91 | 0.92 (0.69 - 1.23) | 0.66 |
| Meta-analysis | 195,231 | 811,393 | 104.2 | 595.2 | 2,987 | 28,526 | 0.81 (0.73 - 0.91) | <0.01 | 0.82 (0.66 - 1.01) | 0.06 |
| Meta-analysis (w/o USOC) | 34,418 | 170,116 | 14.7 | 97.9 | 772 | 7,258 | 0.84 (0.72 - 0.99) | 0.03 | 0.84 (0.63 - 1.12) | 0.25 |

##### 1.5.24 Photosensitivity

**Supplementary Table 128:** Exposures, outcomes and risk estimates for Photosensitivity in GLP1RA (female) (T) vs SU (female) (C) new-user cohorts under four different propensity score (PS) adjustment / time-at-risk (TAR) designs across data sources. We report total patient exposure time in 1,000 patient-year (KYr) units, uncalibrated and calibrated hazard ratios (HRs) and their 95% confidence intervals (CIs). Greyed out entries did not pass study diagnostics and are included only for completeness.

| | Patients | | Exposure (KYr) | | Outcomes | | Uncalibrated | | Calibrated | |
|---|---|---|---|---|---|---|---|---|---|---|
| | T | C | T | C | T | C | HR (95% CI) | $p$ | HR (95% CI) | $p$ |
| **PS-matching / on-treatment TAR** | | | | | | | | | | |
| CCAE | 15,194 | 69,000 | 9.5 | 51.0 | 14 | 85 | 0.66 (0.27 - 1.46) | 0.34 | 0.69 (0.29 - 1.63) | 0.41 |
| ODOD | 8,890 | 55,046 | 5.0 | 46.7 | 10 | 71 | 2.25 (0.91 - 5.09) | 0.06 | 2.32 (0.98 - 5.50) | 0.06 |
| MDCR | 1,072 | 13,148 | 0.7 | 12.0 | <5 | 27 | 1.78 (0.22 - 10.3) | 0.56 | 1.41 (0.21 - 9.66) | 0.73 |
| MDCD | 1,874 | 15,015 | 1.0 | 9.7 | <5 | 10 | 1.15 (0.15 - 5.87) | 0.88 | 1.10 (0.18 - 6.83) | 0.92 |
| Open Claims | 176,301 | 827,290 | 118.5 | 786.8 | 43 | 373 | 1.21 (0.78 - 1.82) | 0.38 | 1.22 (0.78 - 1.92) | 0.38 |
| OEHR | 11,073 | 77,729 | 3.3 | 43.9 | 9 | 58 | 2.37 (0.90 - 5.78) | 0.07 | 2.26 (0.89 - 5.73) | 0.09 |
| Meta-analysis | 211,458 | 1,029,065 | 136.3 | 928.3 | 76 | 587 | 1.38 (0.83 - 2.28) | 0.22 | 1.37 (0.80 - 2.35) | 0.25 |
| Meta-analysis (w/o USOC) | 35,157 | 201,775 | 17.8 | 141.6 | 33 | 214 | 1.50 (0.65 - 3.46) | 0.34 | 1.47 (0.63 - 3.43) | 0.37 |
| **PS-stratification / on-treatment TAR** | | | | | | | | | | |
| CCAE | 20,378 | 71,317 | 12.1 | 52.4 | 18 | 87 | 0.75 (0.39 - 1.37) | 0.37 | 0.74 (0.39 - 1.40) | 0.37 |
| ODOD | 11,576 | 59,797 | 6.1 | 51.7 | 11 | 81 | 1.97 (0.88 - 4.01) | 0.08 | 2.02 (0.94 - 4.30) | 0.07 |
| MDCR | 1,204 | 17,247 | 0.8 | 15.5 | <5 | 35 | 1.42 (0.20 - 5.79) | 0.68 | 1.19 (0.22 - 6.31) | 0.84 |
| MDCD | 2,013 | 16,755 | 1.1 | 11.0 | <5 | 10 | 1.32 (0.18 - 5.84) | 0.75 | 1.27 (0.23 - 7.14) | 0.79 |
| Open Claims | 228,366 | 844,310 | 142.1 | 802.8 | 52 | 387 | 1.16 (0.81 - 1.62) | 0.40 | 1.18 (0.81 - 1.73) | 0.39 |
| OEHR | 13,528 | 82,778 | 4.0 | 47.1 | 9 | 67 | 2.64 (1.15 - 5.49) | 0.01 | 2.47 (1.10 - 5.53) | 0.03 |
| Meta-analysis | 273,848 | 1,058,202 | 164.3 | 953.9 | 90 | 622 | 1.38 (0.83 - 2.28) | 0.22 | 1.39 (0.81 - 2.40) | 0.23 |
| Meta-analysis (w/o USOC) | 45,482 | 213,892 | 22.2 | 151.1 | 38 | 235 | 1.53 (0.70 - 3.35) | 0.29 | 1.51 (0.67 - 3.42) | 0.32 |
| **PS-matching / on-treatment TAR without escalation** | | | | | | | | | | |
| CCAE | 15,194 | 69,000 | 8.5 | 43.7 | 12 | 74 | 0.69 (0.26 - 1.62) | 0.43 | 0.72 (0.29 - 1.78) | 0.48 |
| ODOD | 8,890 | 55,046 | 4.5 | 41.4 | 10 | 67 | 2.41 (0.97 - 5.53) | 0.05 | 2.48 (1.04 - 5.95) | 0.04 |
| MDCR | 1,072 | 13,148 | 0.6 | 10.9 | <5 | 24 | 1.77 (0.22 - 10.3) | 0.56 | 1.39 (0.20 - 9.51) | 0.74 |
| MDCD | 1,874 | 15,015 | 0.9 | 8.0 | <5 | 8 | 1.11 (0.15 - 5.57) | 0.91 | 1.06 (0.17 - 6.54) | 0.94 |
| Open Claims | 176,301 | 827,290 | 105.8 | 661.2 | 30 | 322 | 0.92 (0.54 - 1.48) | 0.73 | 0.92 (0.55 - 1.55) | 0.76 |
| OEHR | 11,073 | 77,729 | 2.9 | 37.1 | 6 | 50 | 1.55 (0.46 - 4.53) | 0.45 | 1.44 (0.46 - 4.54) | 0.53 |
| Meta-analysis | 211,458 | 1,029,065 | 121.6 | 783.5 | 58 | 513 | 1.17 (0.69 - 1.98) | 0.56 | 1.17 (0.67 - 2.03) | 0.59 |
| Meta-analysis (w/o USOC) | 35,157 | 201,775 | 15.9 | 122.3 | 28 | 191 | 1.37 (0.63 - 2.97) | 0.43 | 1.34 (0.60 - 2.97) | 0.48 |
| **PS-stratification / on-treatment TAR without escalation** | | | | | | | | | | |
| CCAE | 20,378 | 71,317 | 11.0 | 45.0 | 16 | 76 | 0.69 (0.34 - 1.32) | 0.28 | 0.69 (0.35 - 1.35) | 0.28 |
| ODOD | 11,576 | 59,797 | 5.6 | 46.0 | 11 | 76 | 2.02 (0.90 - 4.16) | 0.07 | 2.08 (0.96 - 4.48) | 0.06 |
| MDCR | 1,204 | 17,247 | 0.7 | 14.2 | <5 | 31 | 1.55 (0.22 - 6.46) | 0.61 | 1.26 (0.23 - 6.82) | 0.79 |
| MDCD | 2,013 | 16,755 | 0.9 | 9.1 | <5 | 8 | 1.69 (0.23 - 8.00) | 0.56 | 1.66 (0.28 - 9.85) | 0.58 |
| Open Claims | 228,366 | 844,310 | 128.2 | 675.2 | 39 | 334 | 0.92 (0.61 - 1.35) | 0.69 | 0.93 (0.61 - 1.43) | 0.74 |
| OEHR | 13,528 | 82,778 | 3.5 | 39.9 | 6 | 57 | 1.93 (0.70 - 4.56) | 0.17 | 1.80 (0.69 - 4.73) | 0.24 |
| Meta-analysis | 273,848 | 1,058,202 | 148.2 | 806.1 | 72 | 543 | 1.16 (0.71 - 1.89) | 0.56 | 1.17 (0.69 - 1.97) | 0.57 |
| Meta-analysis (w/o USOC) | 45,482 | 213,892 | 20.0 | 130.9 | 33 | 209 | 1.34 (0.64 - 2.81) | 0.44 | 1.33 (0.61 - 2.91) | 0.48 |

##### 1.5.25 Venous thromboembolism

**Supplementary Table 129:** Exposures, outcomes and risk estimates for Venous thromboembolism in GLP1RA (female) (T) vs SU (female) (C) new-user cohorts under four different propensity score (PS) adjustment / time-at-risk (TAR) designs across data sources. We report total patient exposure time in 1,000 patient-year (KYr) units, uncalibrated and calibrated hazard ratios (HRs) and their 95% confidence intervals (CIs). Greyed out entries did not pass study diagnostics and are included only for completeness.

| | Patients | | Exposure (KYr) | | Outcomes | | Uncalibrated | | Calibrated | |
|---|---|---|---|---|---|---|---|---|---|---|
| | T | C | T | C | T | C | HR (95% CI) | $p$ | HR (95% CI) | $p$ |
| **PS-matching / on-treatment TAR** | | | | | | | | | | |
| CCAE | 14,954 | 68,098 | 9.3 | 50.3 | 43 | 322 | 0.86 (0.56 - 1.27) | 0.46 | 0.89 (0.57 - 1.39) | 0.65 |
| ODOD | 8,642 | 53,731 | 4.9 | 45.3 | 43 | 508 | 1.02 (0.66 - 1.52) | 0.94 | 1.05 (0.68 - 1.61) | 0.86 |
| MDCR | 1,060 | 12,877 | 0.7 | 11.7 | 6 | 180 | 0.63 (0.21 - 1.57) | 0.38 | 0.50 (0.18 - 1.38) | 0.19 |
| MDCD | 1,803 | 14,367 | 1.0 | 9.1 | 10 | 106 | 1.67 (0.71 - 3.57) | 0.22 | 1.59 (0.71 - 3.57) | 0.27 |
| Open Claims | 171,467 | 806,584 | 115.4 | 764.8 | 420 | 4,709 | 0.76 (0.66 - 0.87) | <0.01 | 0.77 (0.63 - 0.94) | 0.01 |
| OEHR | 10,853 | 76,373 | 3.3 | 42.7 | 26 | 403 | 1.62 (0.96 - 2.61) | 0.06 | 1.54 (0.94 - 2.54) | 0.09 |
| Meta-analysis | 208,779 | 1,032,030 | 134.4 | 923.9 | 548 | 6,228 | 0.98 (0.74 - 1.30) | 0.91 | 0.98 (0.70 - 1.37) | 0.91 |
| Meta-analysis (w/o USOC) | 37,312 | 225,446 | 19.1 | 159.1 | 128 | 1,519 | 1.10 (0.81 - 1.49) | 0.55 | 1.07 (0.76 - 1.51) | 0.69 |
| **PS-stratification / on-treatment TAR** | | | | | | | | | | |
| CCAE | 20,049 | 70,354 | 11.8 | 51.6 | 54 | 332 | 0.85 (0.60 - 1.19) | 0.36 | 0.84 (0.58 - 1.22) | 0.39 |
| ODOD | 11,251 | 58,278 | 6.0 | 50.2 | 52 | 622 | 0.98 (0.69 - 1.37) | 0.91 | 1.01 (0.71 - 1.41) | 0.93 |
| MDCR | 1,185 | 16,713 | 0.7 | 15.0 | 7 | 248 | 0.65 (0.26 - 1.36) | 0.30 | 0.54 (0.24 - 1.23) | 0.15 |
| MDCD | 1,938 | 16,115 | 1.0 | 10.5 | 10 | 131 | 0.76 (0.35 - 1.49) | 0.45 | 0.73 (0.35 - 1.51) | 0.40 |
| Open Claims | 221,964 | 823,043 | 138.3 | 780.4 | 508 | 4,999 | 0.78 (0.70 - 0.86) | <0.01 | 0.79 (0.65 - 0.97) | 0.02 |
| OEHR | 13,250 | 81,268 | 3.9 | 45.7 | 30 | 472 | 1.26 (0.82 - 1.86) | 0.27 | 1.17 (0.74 - 1.85) | 0.53 |
| Meta-analysis | 266,514 | 1,032,943 | 160.0 | 927.9 | 644 | 6,425 | 0.90 (0.73 - 1.09) | 0.28 | 0.91 (0.69 - 1.20) | 0.49 |
| Meta-analysis (w/o USOC) | 44,550 | 209,900 | 21.7 | 147.5 | 136 | 1,426 | 0.99 (0.81 - 1.22) | 0.95 | 0.98 (0.72 - 1.34) | 0.90 |
| **PS-matching / on-treatment TAR without escalation** | | | | | | | | | | |
| CCAE | 14,954 | 68,098 | 8.3 | 43.2 | 38 | 279 | 0.85 (0.54 - 1.29) | 0.46 | 0.88 (0.57 - 1.37) | 0.60 |
| ODOD | 8,642 | 53,731 | 4.4 | 40.2 | 38 | 458 | 1.03 (0.65 - 1.58) | 0.90 | 1.06 (0.68 - 1.66) | 0.81 |
| MDCR | 1,060 | 12,877 | 0.6 | 10.7 | <5 | 162 | 0.42 (0.10 - 1.24) | 0.18 | 0.33 (0.09 - 1.17) | 0.09 |
| MDCD | 1,803 | 14,367 | 0.8 | 7.6 | <5 | 89 | 0.92 (0.26 - 2.53) | 0.89 | 0.88 (0.28 - 2.76) | 0.83 |
| Open Claims | 171,467 | 806,584 | 103.0 | 643.1 | 365 | 4,011 | 0.75 (0.65 - 0.87) | <0.01 | 0.76 (0.62 - 0.93) | <0.01 |
| OEHR | 10,853 | 76,373 | 2.8 | 36.2 | 21 | 317 | 1.73 (0.98 - 2.90) | 0.05 | 1.61 (0.93 - 2.77) | 0.10 |
| Meta-analysis | 208,779 | 1,032,030 | 119.9 | 781.0 | <472 | 5,316 | 0.92 (0.69 - 1.23) | 0.59 | 0.92 (0.65 - 1.30) | 0.64 |
| Meta-analysis (w/o USOC) | 37,312 | 225,446 | 17.0 | 137.8 | <107 | 1,305 | 1.03 (0.72 - 1.45) | 0.88 | 1.00 (0.67 - 1.50) | 0.92 |
| **PS-stratification / on-treatment TAR without escalation** | | | | | | | | | | |
| CCAE | 20,049 | 70,354 | 10.7 | 44.4 | 49 | 286 | 0.82 (0.57 - 1.18) | 0.30 | 0.82 (0.57 - 1.18) | 0.31 |
| ODOD | 11,251 | 58,278 | 5.4 | 44.6 | 46 | 564 | 0.94 (0.65 - 1.34) | 0.75 | 0.97 (0.67 - 1.40) | 0.86 |
| MDCR | 1,185 | 16,713 | 0.7 | 13.7 | 5 | 225 | 0.49 (0.17 - 1.16) | 0.15 | 0.40 (0.15 - 1.05) | 0.07 |
| MDCD | 1,938 | 16,115 | 0.9 | 8.7 | <5 | 108 | 0.32 (0.09 - 0.84) | 0.04 | 0.32 (0.11 - 0.95) | 0.04 |
| Open Claims | 221,964 | 823,043 | 124.8 | 656.7 | 445 | 4,270 | 0.77 (0.69 - 0.86) | <0.01 | 0.78 (0.64 - 0.95) | 0.01 |
| OEHR | 13,250 | 81,268 | 3.4 | 38.9 | 25 | 366 | 1.28 (0.80 - 1.97) | 0.28 | 1.19 (0.72 - 1.97) | 0.52 |
| Meta-analysis | 266,514 | 1,032,943 | 144.3 | 784.6 | 565 | 5,486 | 0.87 (0.72 - 1.06) | 0.17 | 0.88 (0.67 - 1.14) | 0.33 |
| Meta-analysis (w/o USOC) | 44,550 | 209,900 | 19.6 | 127.9 | 120 | 1,216 | 0.97 (0.76 - 1.23) | 0.80 | 0.96 (0.69 - 1.36) | 0.84 |

##### 1.5.26 Vomiting

**Supplementary Table 130:** Exposures, outcomes and risk estimates for Vomiting in GLP1RA (female) (T) vs SU (female) (C) new-user cohorts under four different propensity score (PS) adjustment / time-at-risk (TAR) designs across data sources. We report total patient exposure time in 1,000 patient-year (KYr) units, uncalibrated and calibrated hazard ratios (HRs) and their 95% confidence intervals (CIs). Greyed out entries did not pass study diagnostics and are included only for completeness.

| | Patients | | Exposure (KYr) | | Outcomes | | Uncalibrated | | Calibrated | |
|---|---|---|---|---|---|---|---|---|---|---|
| | T | C | T | C | T | C | HR (95% CI) | $p$ | HR (95% CI) | $p$ |
| **PS-matching / on-treatment TAR** | | | | | | | | | | |
| CCAE | 13,485 | 61,930 | 8.2 | 45.6 | 348 | 1,327 | 1.43 (1.20 - 1.70) | <0.01 | 1.49 (1.17 - 1.91) | 0.01 |
| ODOD | 7,691 | 48,204 | 4.3 | 40.2 | 248 | 1,639 | 1.46 (1.21 - 1.76) | <0.01 | 1.51 (1.22 - 1.86) | <0.01 |
| MDCR | 1,024 | 12,009 | 0.6 | 10.8 | 28 | 382 | 1.23 (0.73 - 1.98) | 0.42 | 0.97 (0.59 - 1.61) | 0.83 |
| MDCD | 1,353 | 11,126 | 0.7 | 7.0 | 82 | 488 | 1.25 (0.90 - 1.72) | 0.17 | 1.19 (0.86 - 1.66) | 0.33 |
| Open Claims | 148,913 | 718,518 | 100.1 | 684.7 | 2,639 | 13,928 | 1.44 (1.36 - 1.53) | <0.01 | 1.46 (1.24 - 1.72) | <0.01 |
| OEHR | 9,884 | 71,116 | 3.0 | 39.0 | 139 | 1,408 | 1.54 (1.24 - 1.92) | <0.01 | 1.47 (1.18 - 1.84) | <0.01 |
| Meta-analysis | 182,350 | 922,903 | 116.9 | 827.3 | 3,484 | 19,172 | 1.44 (1.37 - 1.52) | <0.01 | 1.44 (1.19 - 1.74) | <0.01 |
| Meta-analysis (w/o USOC) | 33,437 | 204,385 | 16.8 | 142.6 | 845 | 5,244 | 1.44 (1.30 - 1.59) | <0.01 | 1.41 (1.17 - 1.69) | <0.01 |
| **PS-stratification / on-treatment TAR** | | | | | | | | | | |
| CCAE | 17,941 | 64,184 | 10.5 | 47.0 | 495 | 1,369 | 1.45 (1.27 - 1.65) | <0.01 | 1.43 (1.18 - 1.73) | <0.01 |
| ODOD | 9,990 | 52,035 | 5.2 | 44.2 | 323 | 1,857 | 1.39 (1.20 - 1.62) | <0.01 | 1.43 (1.23 - 1.66) | <0.01 |
| MDCR | 1,132 | 15,584 | 0.7 | 13.8 | 28 | 515 | 0.99 (0.63 - 1.49) | 0.97 | 0.83 (0.54 - 1.27) | 0.41 |
| MDCD | 1,441 | 12,420 | 0.7 | 7.9 | 87 | 565 | 1.47 (1.13 - 1.90) | <0.01 | 1.42 (1.09 - 1.85) | 0.02 |
| Open Claims | 191,403 | 733,774 | 119.3 | 699.2 | 3,339 | 14,509 | 1.37 (1.31 - 1.44) | <0.01 | 1.40 (1.18 - 1.67) | <0.01 |
| OEHR | 12,004 | 75,585 | 3.5 | 41.7 | 178 | 1,545 | 1.35 (1.13 - 1.61) | <0.01 | 1.26 (0.96 - 1.64) | 0.18 |
| Meta-analysis | 231,338 | 925,578 | 138.5 | 832.1 | 4,335 | 19,280 | 1.38 (1.33 - 1.44) | <0.01 | 1.40 (1.14 - 1.71) | <0.01 |
| Meta-analysis (w/o USOC) | 39,935 | 191,804 | 19.2 | 132.9 | 996 | 4,771 | 1.41 (1.29 - 1.53) | <0.01 | 1.39 (1.09 - 1.77) | 0.01 |
| **PS-matching / on-treatment TAR without escalation** | | | | | | | | | | |
| CCAE | 13,485 | 61,930 | 7.4 | 39.3 | 318 | 1,108 | 1.55 (1.29 - 1.86) | <0.01 | 1.61 (1.32 - 1.96) | <0.01 |
| ODOD | 7,691 | 48,204 | 3.9 | 35.9 | 226 | 1,463 | 1.51 (1.24 - 1.83) | <0.01 | 1.56 (1.28 - 1.91) | <0.01 |
| MDCR | 1,024 | 12,009 | 0.6 | 9.9 | 25 | 334 | 1.28 (0.74 - 2.13) | 0.35 | 1.01 (0.59 - 1.71) | 0.84 |
| MDCD | 1,353 | 11,126 | 0.6 | 5.9 | 73 | 414 | 1.32 (0.92 - 1.87) | 0.13 | 1.26 (0.88 - 1.81) | 0.23 |
| Open Claims | 148,913 | 718,518 | 89.5 | 578.7 | 2,366 | 11,482 | 1.51 (1.41 - 1.60) | <0.01 | 1.52 (1.30 - 1.77) | <0.01 |
| OEHR | 9,884 | 71,116 | 2.6 | 33.4 | 120 | 1,114 | 1.64 (1.29 - 2.08) | <0.01 | 1.53 (1.20 - 1.95) | <0.01 |
| Meta-analysis | 182,350 | 922,903 | 104.6 | 703.2 | 3,128 | 15,915 | 1.51 (1.43 - 1.60) | <0.01 | 1.51 (1.25 - 1.82) | <0.01 |
| Meta-analysis (w/o USOC) | 33,437 | 204,385 | 15.0 | 124.4 | 762 | 4,433 | 1.52 (1.36 - 1.70) | <0.01 | 1.49 (1.19 - 1.86) | <0.01 |
| **PS-stratification / on-treatment TAR without escalation** | | | | | | | | | | |
| CCAE | 17,941 | 64,184 | 9.5 | 40.6 | 456 | 1,148 | 1.59 (1.38 - 1.82) | <0.01 | 1.58 (1.37 - 1.82) | <0.01 |
| ODOD | 9,990 | 52,035 | 4.7 | 39.5 | 294 | 1,667 | 1.39 (1.19 - 1.63) | <0.01 | 1.43 (1.22 - 1.68) | <0.01 |
| MDCR | 1,132 | 15,584 | 0.6 | 12.6 | 25 | 449 | 1.01 (0.63 - 1.56) | 0.97 | 0.82 (0.52 - 1.29) | 0.40 |
| MDCD | 1,441 | 12,420 | 0.6 | 6.7 | 78 | 487 | 1.51 (1.13 - 1.98) | <0.01 | 1.48 (1.11 - 1.97) | 0.01 |
| Open Claims | 191,403 | 733,774 | 107.8 | 591.6 | 3,025 | 11,994 | 1.46 (1.39 - 1.53) | <0.01 | 1.47 (1.25 - 1.73) | <0.01 |
| OEHR | 12,004 | 75,585 | 3.0 | 35.9 | 154 | 1,228 | 1.44 (1.18 - 1.74) | <0.01 | 1.34 (1.00 - 1.79) | 0.11 |
| Meta-analysis | 231,338 | 925,578 | 125.1 | 707.5 | 3,929 | 16,037 | 1.46 (1.40 - 1.53) | <0.01 | 1.47 (1.23 - 1.77) | <0.01 |
| Meta-analysis (w/o USOC) | 39,935 | 191,804 | 17.3 | 116.0 | 904 | 4,043 | 1.49 (1.36 - 1.63) | <0.01 | 1.48 (1.14 - 1.92) | <0.01 |

#### 1.6 DPP4I (female) vs SU (female)

##### 1.6.1 3-point MACE

**Supplementary Table 131:** Exposures, outcomes and risk estimates for 3-point MACE in DPP4I (female) (T) vs SU (female) (C) new-user cohorts under four different propensity score (PS) adjustment / time-at-risk (TAR) designs across data sources. We report total patient exposure time in 1,000 patient-year (KYr) units, uncalibrated and calibrated hazard ratios (HRs) and their 95% confidence intervals (CIs). Greyed out entries did not pass study diagnostics and are included only for completeness.

| | Patients | | Exposure (KYr) | | Outcomes | | Uncalibrated | | Calibrated | |
|---|---|---|---|---|---|---|---|---|---|---|
| | T | C | T | C | T | C | HR (95% CI) | $p$ | HR (95% CI) | $p$ |
| **PS-matching / on-treatment TAR** | | | | | | | | | | |
| CCAE | 38,239 | 64,329 | 29.9 | 46.8 | 175 | 336 | 0.86 (0.65 - 1.11) | 0.25 | 0.82 (0.63 - 1.08) | 0.17 |
| DAG | 10,485 | 2,040 | 11.4 | 2.1 | 0 | 0 | | | | |
| ODOD | 26,182 | 57,090 | 20.6 | 48.9 | 376 | 1,113 | 0.86 (0.73 - 1.02) | 0.08 | 0.89 (0.75 - 1.06) | 0.25 |
| MDCR | 7,532 | 14,216 | 7.1 | 12.6 | 169 | 392 | 0.72 (0.55 - 0.94) | 0.02 | 0.73 (0.56 - 0.96) | 0.03 |
| MDCD | 6,936 | 15,653 | 4.9 | 10.2 | 96 | 208 | 1.48 (1.03 - 2.13) | 0.03 | 1.43 (0.99 - 2.06) | 0.06 |
| Open Claims | 418,312 | 810,705 | 363.9 | 767.8 | 3,791 | 9,194 | 0.92 (0.87 - 0.97) | <0.01 | 0.91 (0.82 - 1.01) | 0.08 |
| OEHR | 35,813 | 81,690 | 18.3 | 46.1 | 153 | 586 | 0.73 (0.56 - 0.93) | 0.01 | 0.74 (0.57 - 0.95) | 0.02 |
| VA | 1,770 | 9,474 | 1.3 | 7.3 | <10 | 31 | 1.12 (0.29 - 3.56) | 0.86 | 1.01 (0.29 - 3.55) | 0.94 |
| SIDIAP | 19,287 | 5,403 | 53.0 | 14.1 | 144 | 44 | 0.67 (0.43 - 1.06) | 0.09 | 0.70 (0.45 - 1.10) | 0.14 |
| Meta-analysis | 554,071 | 1,058,560 | 499.0 | 953.8 | <4,914 | 11,904 | 0.86 (0.76 - 0.99) | 0.03 | 0.85 (0.73 - 1.00) | 0.04 |
| Meta-analysis (w/o USOC) | 135,759 | 247,855 | 135.1 | 186.0 | <1,123 | 2,710 | 0.85 (0.72 - 1.01) | 0.06 | 0.85 (0.71 - 1.02) | 0.09 |
| **PS-stratification / on-treatment TAR** | | | | | | | | | | |
| CCAE | 44,510 | 66,329 | 35.1 | 48.0 | 203 | 341 | 0.88 (0.73 - 1.06) | 0.19 | 0.87 (0.72 - 1.05) | 0.16 |
| DAG | 10,516 | 2,367 | 11.4 | 2.5 | 0 | 0 | | | | |
| ODOD | 28,169 | 57,247 | 22.3 | 48.9 | 400 | 1,113 | 0.89 (0.79 - 1.00) | 0.06 | 0.92 (0.81 - 1.04) | 0.23 |
| MDCR | 8,124 | 14,511 | 7.6 | 12.8 | 182 | 401 | 0.76 (0.63 - 0.92) | <0.01 | 0.77 (0.64 - 0.93) | 0.01 |
| MDCD | 7,282 | 15,757 | 5.2 | 10.2 | 102 | 208 | 1.09 (0.84 - 1.40) | 0.52 | 1.05 (0.81 - 1.35) | 0.75 |
| Open Claims | 428,138 | 810,802 | 374.7 | 767.8 | 3,943 | 9,195 | 0.89 (0.85 - 0.92) | <0.01 | 0.87 (0.79 - 0.96) | <0.01 |
| OEHR | 38,260 | 81,775 | 19.8 | 46.2 | 161 | 586 | 0.73 (0.61 - 0.88) | <0.01 | 0.74 (0.62 - 0.89) | <0.01 |
| VA | 2,150 | 12,928 | 1.6 | 10.4 | <10 | 50 | 1.26 (0.46 - 3.28) | 0.65 | 1.09 (0.41 - 2.92) | 0.86 |
| SIDIAP | 19,389 | 7,031 | 53.1 | 19.3 | 144 | 55 | 0.68 (0.48 - 0.99) | 0.04 | 0.66 (0.46 - 0.95) | 0.03 |
| Meta-analysis | 566,590 | 1,037,695 | 512.6 | 943.0 | 5,033 | 11,691 | 0.85 (0.79 - 0.91) | <0.01 | 0.84 (0.74 - 0.94) | <0.01 |
| Meta-analysis (w/o USOC) | 138,452 | 226,893 | 137.9 | 175.2 | 1,090 | 2,496 | 0.82 (0.74 - 0.90) | <0.01 | 0.82 (0.73 - 0.92) | <0.01 |
| **PS-matching / on-treatment TAR without escalation** | | | | | | | | | | |
| CCAE | 38,239 | 64,329 | 24.6 | 40.3 | 135 | 282 | 0.86 (0.64 - 1.15) | 0.33 | 0.85 (0.63 - 1.14) | 0.28 |
| DAG | 10,485 | 2,040 | 10.3 | 2.0 | 0 | 0 | | | | |
| ODOD | 26,182 | 57,090 | 17.3 | 43.5 | 331 | 989 | 0.91 (0.76 - 1.08) | 0.28 | 0.95 (0.79 - 1.14) | 0.61 |
| MDCR | 7,532 | 14,216 | 6.1 | 11.6 | 136 | 352 | 0.70 (0.52 - 0.94) | 0.02 | 0.70 (0.52 - 0.95) | 0.02 |
| MDCD | 6,936 | 15,653 | 3.8 | 8.5 | 72 | 170 | 1.35 (0.90 - 2.02) | 0.14 | 1.30 (0.87 - 1.95) | 0.21 |
| Open Claims | 418,312 | 810,705 | 288.9 | 646.3 | 3,075 | 7,802 | 0.90 (0.85 - 0.96) | <0.01 | 0.90 (0.81 - 0.98) | 0.04 |
| OEHR | 35,813 | 81,690 | 14.7 | 39.3 | 98 | 426 | 0.64 (0.48 - 0.86) | <0.01 | 0.65 (0.48 - 0.88) | <0.01 |
| VA | 1,770 | 9,474 | 1.1 | 6.3 | <10 | 24 | 0.88 (0.18 - 3.29) | 0.87 | 0.80 (0.19 - 3.39) | 0.76 |
| SIDIAP | 19,287 | 5,403 | 39.4 | 11.5 | 114 | 39 | 0.64 (0.38 - 1.12) | 0.11 | 0.69 (0.40 - 1.19) | 0.19 |
| Meta-analysis | 554,071 | 1,058,560 | 396.0 | 807.2 | <3,971 | 10,084 | 0.85 (0.74 - 0.97) | 0.02 | 0.84 (0.72 - 0.99) | 0.04 |
| Meta-analysis (w/o USOC) | 135,759 | 247,855 | 107.1 | 160.9 | <896 | 2,282 | 0.83 (0.69 - 0.99) | 0.04 | 0.83 (0.69 - 1.00) | 0.07 |
| **PS-stratification / on-treatment TAR without escalation** | | | | | | | | | | |
| CCAE | 44,510 | 66,329 | 28.9 | 41.4 | 158 | 286 | 0.86 (0.70 - 1.05) | 0.15 | 0.85 (0.69 - 1.05) | 0.14 |
| DAG | 10,516 | 2,367 | 10.3 | 2.3 | 0 | 0 | | | | |
| ODOD | 28,169 | 57,247 | 18.7 | 43.6 | 353 | 989 | 0.93 (0.82 - 1.06) | 0.28 | 0.98 (0.85 - 1.12) | 0.76 |
| MDCR | 8,124 | 14,511 | 6.6 | 11.8 | 147 | 359 | 0.73 (0.59 - 0.89) | <0.01 | 0.73 (0.60 - 0.90) | <0.01 |
| MDCD | 7,282 | 15,757 | 4.0 | 8.5 | 78 | 170 | 1.04 (0.78 - 1.39) | 0.77 | 1.00 (0.75 - 1.33) | 0.90 |
| Open Claims | 428,138 | 810,802 | 297.6 | 646.3 | 3,196 | 7,803 | 0.90 (0.86 - 0.94) | <0.01 | 0.88 (0.80 - 0.98) | 0.02 |
| OEHR | 38,260 | 81,775 | 15.9 | 39.3 | 104 | 426 | 0.66 (0.52 - 0.82) | <0.01 | 0.66 (0.53 - 0.83) | <0.01 |
| VA | 2,150 | 12,928 | 1.3 | 9.0 | <10 | 41 | 1.07 (0.33 - 3.25) | 0.90 | 0.95 (0.30 - 2.97) | 0.92 |
| SIDIAP | 19,389 | 7,031 | 39.5 | 15.6 | 114 | 48 | 0.66 (0.45 - 0.99) | 0.04 | 0.65 (0.44 - 0.97) | 0.04 |
| Meta-analysis | 566,590 | 1,037,695 | 407.3 | 798.0 | 4,072 | 9,911 | 0.82 (0.72 - 0.92) | <0.01 | 0.81 (0.70 - 0.94) | <0.01 |
| Meta-analysis (w/o USOC) | 138,452 | 226,893 | 109.7 | 151.7 | 876 | 2,108 | 0.78 (0.67 - 0.91) | <0.01 | 0.79 (0.67 - 0.93) | <0.01 |

##### 1.6.2 4-point MACE

**Supplementary Table 132:** Exposures, outcomes and risk estimates for 4-point MACE in DPP4I (female) (T) vs SU (female) (C) new-user cohorts under four different propensity score (PS) adjustment / time-at-risk (TAR) designs across data sources. We report total patient exposure time in 1,000 patient-year (KYr) units, uncalibrated and calibrated hazard ratios (HRs) and their 95% confidence intervals (CIs). Greyed out entries did not pass study diagnostics and are included only for completeness.

| | Patients | | Exposure (KYr) | | Outcomes | | Uncalibrated | | Calibrated | |
|---|---|---|---|---|---|---|---|---|---|---|
| | T | C | T | C | T | C | HR (95% CI) | $p$ | HR (95% CI) | $p$ |
| **PS-matching / on-treatment TAR** | | | | | | | | | | |
| CCAE | 37,689 | 63,317 | 29.5 | 46.0 | 285 | 546 | 0.94 (0.77 - 1.16) | 0.59 | 0.91 (0.74 - 1.12) | 0.40 |
| DAG | 10,485 | 2,040 | 11.4 | 2.1 | 0 | 0 | | | | |
| ODOD | 25,033 | 54,385 | 19.6 | 46.1 | 642 | 1,803 | 0.88 (0.78 - 1.00) | 0.06 | 0.91 (0.80 - 1.04) | 0.28 |
| MDCR | 7,009 | 13,031 | 6.6 | 11.4 | 258 | 622 | 0.73 (0.59 - 0.90) | <0.01 | 0.74 (0.59 - 0.91) | <0.01 |
| MDCD | 6,470 | 14,576 | 4.5 | 9.4 | 170 | 355 | 1.13 (0.87 - 1.48) | 0.36 | 1.09 (0.83 - 1.43) | 0.54 |
| Open Claims | 404,455 | 783,629 | 350.4 | 738.0 | 6,256 | 15,360 | 0.88 (0.84 - 0.92) | <0.01 | 0.87 (0.79 - 0.96) | <0.01 |
| OEHR | 35,226 | 80,207 | 17.8 | 44.7 | 310 | 1,088 | 0.84 (0.71 - 1.00) | 0.05 | 0.85 (0.72 - 1.02) | 0.09 |
| VA | 1,738 | 9,206 | 1.3 | 7.1 | 12 | 58 | 1.25 (0.46 - 3.06) | 0.65 | 1.13 (0.44 - 2.91) | 0.81 |
| SIDIAP | 19,149 | 5,388 | 52.8 | 14.0 | 195 | 48 | 0.82 (0.54 - 1.26) | 0.35 | 0.85 (0.56 - 1.30) | 0.47 |
| Meta-analysis | 536,769 | 1,023,739 | 482.5 | 916.7 | 8,128 | 19,880 | 0.88 (0.85 - 0.91) | <0.01 | 0.87 (0.80 - 0.94) | <0.01 |
| Meta-analysis (w/o USOC) | 132,314 | 240,110 | 132.1 | 178.7 | 1,872 | 4,520 | 0.88 (0.80 - 0.97) | 0.01 | 0.88 (0.79 - 0.98) | 0.04 |
| **PS-stratification / on-treatment TAR** | | | | | | | | | | |
| CCAE | 43,928 | 65,293 | 34.7 | 47.2 | 329 | 558 | 0.89 (0.77 - 1.03) | 0.12 | 0.88 (0.76 - 1.02) | 0.10 |
| DAG | 10,516 | 2,367 | 11.4 | 2.5 | 0 | 0 | | | | |
| ODOD | 27,017 | 54,539 | 21.2 | 46.2 | 680 | 1,805 | 0.91 (0.83 - 1.00) | 0.06 | 0.94 (0.86 - 1.04) | 0.32 |
| MDCR | 7,581 | 13,379 | 7.1 | 11.7 | 286 | 641 | 0.77 (0.66 - 0.89) | <0.01 | 0.78 (0.67 - 0.90) | <0.01 |
| MDCD | 6,786 | 14,676 | 4.8 | 9.4 | 181 | 355 | 1.05 (0.87 - 1.27) | 0.61 | 1.01 (0.83 - 1.23) | 0.86 |
| Open Claims | 414,120 | 783,725 | 361.1 | 738.0 | 6,491 | 15,362 | 0.88 (0.85 - 0.90) | <0.01 | 0.86 (0.78 - 0.95) | <0.01 |
| OEHR | 37,672 | 80,288 | 19.3 | 44.8 | 325 | 1,088 | 0.82 (0.72 - 0.93) | <0.01 | 0.83 (0.72 - 0.94) | <0.01 |
| VA | 2,111 | 12,720 | 1.5 | 10.2 | 15 | 101 | 1.01 (0.49 - 2.00) | 0.99 | 0.88 (0.43 - 1.78) | 0.72 |
| SIDIAP | 19,309 | 7,015 | 52.9 | 19.2 | 195 | 65 | 0.86 (0.63 - 1.20) | 0.38 | 0.84 (0.60 - 1.16) | 0.31 |
| Meta-analysis | 549,627 | 1,004,239 | 496.3 | 907.0 | 8,306 | 19,519 | 0.87 (0.85 - 0.90) | <0.01 | 0.86 (0.78 - 0.95) | <0.01 |
| Meta-analysis (w/o USOC) | 135,507 | 220,514 | 135.2 | 169.0 | 1,815 | 4,157 | 0.86 (0.80 - 0.92) | <0.01 | 0.86 (0.78 - 0.95) | <0.01 |
| **PS-matching / on-treatment TAR without escalation** | | | | | | | | | | |
| CCAE | 37,689 | 63,317 | 24.2 | 39.7 | 227 | 471 | 0.93 (0.74 - 1.16) | 0.54 | 0.91 (0.73 - 1.15) | 0.46 |
| DAG | 10,485 | 2,040 | 10.3 | 2.0 | 0 | 0 | | | | |
| ODOD | 25,033 | 54,385 | 16.4 | 41.1 | 543 | 1,588 | 0.89 (0.78 - 1.02) | 0.11 | 0.94 (0.81 - 1.08) | 0.43 |
| MDCR | 7,009 | 13,031 | 5.8 | 10.5 | 216 | 566 | 0.68 (0.54 - 0.85) | <0.01 | 0.68 (0.54 - 0.86) | <0.01 |
| MDCD | 6,470 | 14,576 | 3.5 | 7.8 | 129 | 289 | 1.03 (0.76 - 1.39) | 0.84 | 0.99 (0.74 - 1.34) | 0.86 |
| Open Claims | 404,455 | 783,629 | 278.5 | 621.7 | 5,035 | 12,980 | 0.88 (0.84 - 0.92) | <0.01 | 0.87 (0.80 - 0.95) | <0.01 |
| OEHR | 35,226 | 80,207 | 14.4 | 38.3 | 210 | 827 | 0.76 (0.62 - 0.93) | <0.01 | 0.77 (0.63 - 0.94) | 0.01 |
| VA | 1,738 | 9,206 | 1.0 | 6.1 | <10 | 48 | 0.80 (0.25 - 2.14) | 0.68 | 0.72 (0.25 - 2.10) | 0.55 |
| SIDIAP | 19,149 | 5,388 | 39.2 | 11.4 | 145 | 43 | 0.71 (0.43 - 1.18) | 0.18 | 0.76 (0.46 - 1.26) | 0.30 |
| Meta-analysis | 536,769 | 1,023,739 | 383.1 | 776.6 | <6,515 | 16,812 | 0.87 (0.83 - 0.91) | <0.01 | 0.86 (0.79 - 0.95) | <0.01 |
| Meta-analysis (w/o USOC) | 132,314 | 240,110 | 104.6 | 154.9 | <1,480 | 3,832 | 0.84 (0.75 - 0.93) | <0.01 | 0.84 (0.75 - 0.95) | <0.01 |
| **PS-stratification / on-treatment TAR without escalation** | | | | | | | | | | |
| CCAE | 43,928 | 65,293 | 28.6 | 40.7 | 264 | 480 | 0.87 (0.74 - 1.02) | 0.08 | 0.86 (0.73 - 1.01) | 0.08 |
| DAG | 10,516 | 2,367 | 10.3 | 2.3 | 0 | 0 | | | | |
| ODOD | 27,017 | 54,539 | 17.8 | 41.1 | 574 | 1,590 | 0.93 (0.84 - 1.02) | 0.13 | 0.97 (0.87 - 1.08) | 0.65 |
| MDCR | 7,581 | 13,379 | 6.2 | 10.8 | 240 | 583 | 0.75 (0.64 - 0.87) | <0.01 | 0.75 (0.64 - 0.88) | <0.01 |
| MDCD | 6,786 | 14,676 | 3.7 | 7.8 | 138 | 289 | 1.02 (0.82 - 1.26) | 0.86 | 0.98 (0.78 - 1.22) | 0.82 |
| Open Claims | 414,120 | 783,725 | 287.0 | 621.7 | 5,220 | 12,982 | 0.88 (0.85 - 0.91) | <0.01 | 0.87 (0.78 - 0.96) | <0.01 |
| OEHR | 37,672 | 80,288 | 15.6 | 38.3 | 222 | 827 | 0.77 (0.66 - 0.90) | <0.01 | 0.78 (0.66 - 0.91) | <0.01 |
| VA | 2,111 | 12,720 | 1.2 | 8.9 | <10 | 83 | 0.69 (0.27 - 1.61) | 0.42 | 0.61 (0.25 - 1.49) | 0.28 |
| SIDIAP | 19,309 | 7,015 | 39.4 | 15.5 | 145 | 58 | 0.77 (0.54 - 1.11) | 0.16 | 0.76 (0.53 - 1.10) | 0.17 |
| Meta-analysis | 549,627 | 1,004,239 | 394.6 | 768.3 | 6,665 | 16,520 | 0.85 (0.80 - 0.91) | <0.01 | 0.85 (0.76 - 0.94) | <0.01 |
| Meta-analysis (w/o USOC) | 135,507 | 220,514 | 107.6 | 146.5 | 1,445 | 3,538 | 0.83 (0.76 - 0.91) | <0.01 | 0.84 (0.75 - 0.94) | <0.01 |

##### 1.6.3 Acute myocardial infarction

**Supplementary Table 133:** Exposures, outcomes and risk estimates for Acute myocardial infarction in DPP4I (female) (T) vs SU (female) (C) new-user cohorts under four different propensity score (PS) adjustment / time-at-risk (TAR) designs across data sources. We report total patient exposure time in 1,000 patient-year (KYr) units, uncalibrated and calibrated hazard ratios (HRs) and their 95% confidence intervals (CIs). Greyed out entries did not pass study diagnostics and are included only for completeness.

| | Patients | | Exposure (KYr) | | Outcomes | | Uncalibrated | | Calibrated | |
|---|---|---|---|---|---|---|---|---|---|---|
| | T | C | T | C | T | C | HR (95% CI) | $p$ | HR (95% CI) | $p$ |
| **PS-matching / on-treatment TAR** | | | | | | | | | | |
| CCAE | 38,717 | 65,227 | 30.3 | 47.6 | 75 | 145 | 0.84 (0.54 - 1.28) | 0.43 | 0.81 (0.53 - 1.25) | 0.35 |
| DAG | 10,485 | 2,040 | 11.4 | 2.1 | 0 | 0 | | | | |
| ODOD | 26,968 | 58,852 | 21.3 | 50.9 | 141 | 481 | 0.74 (0.56 - 0.98) | 0.04 | 0.77 (0.58 - 1.02) | 0.08 |
| MDCR | 7,901 | 14,944 | 7.4 | 13.3 | 71 | 160 | 0.94 (0.63 - 1.39) | 0.77 | 0.95 (0.64 - 1.41) | 0.81 |
| MDCD | 7,213 | 16,301 | 5.1 | 10.7 | 31 | 83 | 0.88 (0.47 - 1.61) | 0.69 | 0.85 (0.46 - 1.58) | 0.62 |
| Open Claims | 430,562 | 833,795 | 375.3 | 792.0 | 1,609 | 3,979 | 0.93 (0.85 - 1.01) | 0.09 | 0.92 (0.82 - 1.04) | 0.21 |
| OEHR | 36,117 | 82,493 | 18.5 | 46.7 | 64 | 277 | 0.56 (0.38 - 0.83) | <0.01 | 0.57 (0.39 - 0.85) | <0.01 |
| VA | 1,796 | 9,581 | 1.4 | 7.4 | <10 | 16 | 0.93 (0.12 - 4.74) | 0.94 | 0.84 (0.14 - 5.22) | 0.85 |
| SIDIAP | 19,424 | 5,426 | 53.5 | 14.2 | 45 | 22 | 0.48 (0.23 - 0.98) | 0.05 | 0.50 (0.24 - 1.03) | 0.06 |
| Meta-analysis | 566,902 | 1,077,038 | 511.4 | 975.3 | 2,036 | 5,147 | 0.80 (0.68 - 0.95) | 0.01 | 0.79 (0.66 - 0.95) | 0.01 |
| Meta-analysis (w/o USOC) | 136,340 | 243,243 | 136.1 | 183.4 | 427 | 1,168 | 0.74 (0.63 - 0.89) | <0.01 | 0.75 (0.62 - 0.89) | <0.01 |
| **PS-stratification / on-treatment TAR** | | | | | | | | | | |
| CCAE | 45,027 | 67,241 | 35.5 | 48.8 | 85 | 146 | 0.91 (0.68 - 1.20) | 0.51 | 0.90 (0.68 - 1.19) | 0.46 |
| DAG | 10,516 | 2,367 | 11.4 | 2.5 | 0 | 0 | | | | |
| ODOD | 28,975 | 59,009 | 23.0 | 50.9 | 147 | 481 | 0.78 (0.64 - 0.94) | 0.01 | 0.80 (0.66 - 0.98) | 0.04 |
| MDCR | 8,520 | 15,256 | 8.0 | 13.6 | 74 | 163 | 0.81 (0.61 - 1.08) | 0.16 | 0.82 (0.61 - 1.10) | 0.21 |
| MDCD | 7,556 | 16,407 | 5.4 | 10.7 | 33 | 83 | 0.85 (0.55 - 1.30) | 0.47 | 0.82 (0.53 - 1.26) | 0.38 |
| Open Claims | 440,491 | 833,892 | 386.3 | 792.0 | 1,670 | 3,979 | 0.89 (0.84 - 0.94) | <0.01 | 0.87 (0.78 - 0.97) | 0.02 |
| OEHR | 38,583 | 82,578 | 20.0 | 46.8 | 69 | 277 | 0.66 (0.50 - 0.87) | <0.01 | 0.67 (0.51 - 0.88) | <0.01 |
| VA | 2,188 | 13,082 | 1.6 | 10.5 | <10 | 29 | 0.49 (0.06 - 2.55) | 0.45 | 0.43 (0.07 - 2.69) | 0.36 |
| SIDIAP | 19,530 | 7,058 | 53.5 | 19.3 | 45 | 26 | 0.53 (0.30 - 0.93) | 0.03 | 0.51 (0.29 - 0.90) | 0.02 |
| Meta-analysis | 581,126 | 1,065,034 | 526.5 | 971.4 | 2,090 | 5,072 | 0.81 (0.73 - 0.91) | <0.01 | 0.80 (0.69 - 0.93) | <0.01 |
| Meta-analysis (w/o USOC) | 140,635 | 231,142 | 140.1 | 179.4 | 420 | 1,093 | 0.77 (0.67 - 0.87) | <0.01 | 0.77 (0.66 - 0.89) | <0.01 |
| **PS-matching / on-treatment TAR without escalation** | | | | | | | | | | |
| CCAE | 38,717 | 65,227 | 24.9 | 41.0 | 52 | 123 | 0.78 (0.48 - 1.26) | 0.33 | 0.77 (0.47 - 1.25) | 0.29 |
| DAG | 10,485 | 2,040 | 10.3 | 2.0 | 0 | 0 | | | | |
| ODOD | 26,968 | 58,852 | 17.9 | 45.2 | 126 | 423 | 0.80 (0.59 - 1.06) | 0.13 | 0.84 (0.62 - 1.12) | 0.26 |
| MDCR | 7,901 | 14,944 | 6.4 | 12.2 | 63 | 143 | 1.07 (0.68 - 1.65) | 0.76 | 1.08 (0.69 - 1.67) | 0.75 |
| MDCD | 7,213 | 16,301 | 4.0 | 8.9 | 21 | 66 | 0.96 (0.46 - 1.89) | 0.90 | 0.92 (0.46 - 1.86) | 0.82 |
| Open Claims | 430,562 | 833,795 | 298.0 | 666.3 | 1,301 | 3,391 | 0.92 (0.84 - 1.01) | 0.08 | 0.92 (0.81 - 1.03) | 0.17 |
| OEHR | 36,117 | 82,493 | 14.9 | 39.7 | 39 | 201 | 0.50 (0.30 - 0.79) | <0.01 | 0.50 (0.31 - 0.82) | <0.01 |
| VA | 1,796 | 9,581 | 1.1 | 6.4 | <10 | 14 | 0.99 (0.13 - 5.28) | 0.99 | 0.89 (0.14 - 5.70) | 0.90 |
| SIDIAP | 19,424 | 5,426 | 39.8 | 11.5 | 34 | 19 | 0.46 (0.18 - 1.15) | 0.10 | 0.49 (0.19 - 1.24) | 0.14 |
| Meta-analysis | 566,902 | 1,077,038 | 405.8 | 824.9 | 1,636 | 4,366 | 0.83 (0.70 - 0.98) | 0.03 | 0.82 (0.68 - 1.00) | 0.05 |
| Meta-analysis (w/o USOC) | 136,340 | 243,243 | 107.8 | 158.6 | 335 | 975 | 0.77 (0.61 - 0.97) | 0.03 | 0.77 (0.61 - 0.98) | 0.04 |
| **PS-stratification / on-treatment TAR without escalation** | | | | | | | | | | |
| CCAE | 45,027 | 67,241 | 29.2 | 42.1 | 58 | 124 | 0.78 (0.56 - 1.08) | 0.13 | 0.77 (0.55 - 1.08) | 0.13 |
| DAG | 10,516 | 2,367 | 10.3 | 2.3 | 0 | 0 | | | | |
| ODOD | 28,975 | 59,009 | 19.3 | 45.3 | 132 | 423 | 0.83 (0.67 - 1.01) | 0.07 | 0.87 (0.70 - 1.07) | 0.21 |
| MDCR | 8,520 | 15,256 | 7.0 | 12.5 | 65 | 145 | 0.84 (0.61 - 1.14) | 0.27 | 0.85 (0.62 - 1.16) | 0.32 |
| MDCD | 7,556 | 16,407 | 4.2 | 8.9 | 23 | 66 | 0.83 (0.49 - 1.36) | 0.48 | 0.80 (0.48 - 1.32) | 0.39 |
| Open Claims | 440,491 | 833,892 | 306.8 | 666.4 | 1,346 | 3,391 | 0.89 (0.83 - 0.95) | <0.01 | 0.88 (0.78 - 0.98) | 0.03 |
| OEHR | 38,583 | 82,578 | 16.1 | 39.8 | 42 | 201 | 0.57 (0.40 - 0.80) | <0.01 | 0.57 (0.40 - 0.81) | <0.01 |
| VA | 2,188 | 13,082 | 1.3 | 9.2 | <10 | 26 | 0.53 (0.07 - 2.89) | 0.51 | 0.47 (0.07 - 3.03) | 0.43 |
| SIDIAP | 19,530 | 7,058 | 39.8 | 15.7 | 34 | 22 | 0.49 (0.26 - 0.94) | 0.03 | 0.48 (0.25 - 0.92) | 0.03 |
| Meta-analysis | 581,126 | 1,065,034 | 418.3 | 821.6 | 1,677 | 4,306 | 0.79 (0.69 - 0.91) | <0.01 | 0.79 (0.67 - 0.93) | <0.01 |
| Meta-analysis (w/o USOC) | 140,635 | 231,142 | 111.5 | 155.3 | 331 | 915 | 0.74 (0.63 - 0.88) | <0.01 | 0.75 (0.62 - 0.90) | <0.01 |

##### 1.6.4 Acute renal failure

**Supplementary Table 134:** Exposures, outcomes and risk estimates for Acute renal failure in DPP4I (female) (T) vs SU (female) (C) new-user cohorts under four different propensity score (PS) adjustment / time-at-risk (TAR) designs across data sources. We report total patient exposure time in 1,000 patient-year (KYr) units, uncalibrated and calibrated hazard ratios (HRs) and their 95% confidence intervals (CIs). Greyed out entries did not pass study diagnostics and are included only for completeness.

| | Patients | | Exposure (KYr) | | Outcomes | | Uncalibrated | | Calibrated | |
|---|---|---|---|---|---|---|---|---|---|---|
| | T | C | T | C | T | C | HR (95% CI) | $p$ | HR (95% CI) | $p$ |
| **PS-matching / on-treatment TAR** | | | | | | | | | | |
| CCAE | 38,636 | 64,976 | 30.2 | 47.3 | 194 | 406 | 0.85 (0.65 - 1.09) | 0.20 | 0.81 (0.63 - 1.06) | 0.13 |
| DAG | 10,485 | 2,040 | 11.4 | 2.1 | 0 | 0 | | | | |
| ODOD | 26,125 | 56,950 | 20.4 | 48.6 | 577 | 1,557 | 0.93 (0.82 - 1.07) | 0.32 | 0.97 (0.84 - 1.11) | 0.70 |
| MDCR | 7,704 | 14,462 | 7.2 | 12.7 | 218 | 458 | 0.85 (0.67 - 1.06) | 0.16 | 0.85 (0.68 - 1.08) | 0.21 |
| MDCD | 7,000 | 15,775 | 4.8 | 10.2 | 157 | 322 | 1.30 (0.97 - 1.74) | 0.07 | 1.26 (0.94 - 1.68) | 0.14 |
| Open Claims | 423,871 | 821,287 | 366.9 | 775.7 | 5,336 | 12,429 | 0.95 (0.91 - 1.00) | 0.04 | 0.94 (0.86 - 1.04) | 0.27 |
| OEHR | 35,910 | 81,969 | 18.3 | 46.1 | 259 | 763 | 1.03 (0.85 - 1.25) | 0.72 | 1.05 (0.87 - 1.27) | 0.63 |
| VA | 1,766 | 9,451 | 1.3 | 7.2 | 16 | 78 | 1.84 (0.79 - 3.97) | 0.14 | 1.66 (0.74 - 3.72) | 0.23 |
| SIDIAP | 19,487 | 5,438 | 53.7 | 14.2 | 16 | <5 | 0.45 (0.08 - 3.63) | 0.41 | 0.47 (0.07 - 3.10) | 0.43 |
| Meta-analysis | 541,012 | 1,064,870 | 449.2 | 947.9 | 6,757 | 16,013 | 0.95 (0.92 - 0.99) | 0.03 | 0.94 (0.87 - 1.03) | 0.18 |
| Meta-analysis (w/o USOC) | 117,141 | 243,583 | 82.3 | 172.2 | 1,421 | 3,584 | 0.98 (0.86 - 1.13) | 0.82 | 0.98 (0.85 - 1.14) | 0.85 |
| **PS-stratification / on-treatment TAR** | | | | | | | | | | |
| CCAE | 44,957 | 66,985 | 35.5 | 48.5 | 225 | 413 | 0.84 (0.71 - 1.00) | 0.05 | 0.83 (0.70 - 0.99) | 0.04 |
| DAG | 10,516 | 2,367 | 11.4 | 2.5 | 0 | 0 | | | | |
| ODOD | 28,091 | 57,107 | 22.1 | 48.7 | 613 | 1,557 | 0.96 (0.87 - 1.06) | 0.42 | 0.99 (0.89 - 1.10) | 0.84 |
| MDCR | 8,325 | 14,834 | 7.8 | 13.1 | 238 | 466 | 0.85 (0.72 - 1.00) | 0.06 | 0.86 (0.73 - 1.02) | 0.12 |
| MDCD | 7,347 | 15,879 | 5.1 | 10.2 | 172 | 323 | 1.10 (0.90 - 1.34) | 0.37 | 1.05 (0.86 - 1.29) | 0.63 |
| Open Claims | 433,792 | 821,409 | 377.9 | 775.7 | 5,536 | 12,429 | 0.94 (0.91 - 0.97) | <0.01 | 0.92 (0.84 - 1.01) | 0.10 |
| OEHR | 38,353 | 82,056 | 19.8 | 46.2 | 267 | 763 | 0.97 (0.84 - 1.13) | 0.73 | 0.99 (0.85 - 1.15) | 0.84 |
| VA | 2,163 | 12,941 | 1.6 | 10.3 | 17 | 116 | 1.64 (0.82 - 3.13) | 0.15 | 1.43 (0.73 - 2.78) | 0.31 |
| SIDIAP | 19,596 | 7,072 | 53.8 | 19.4 | 16 | <5 | 0.91 (0.23 - 5.19) | 0.91 | 0.88 (0.19 - 4.22) | 0.88 |
| Meta-analysis | 573,114 | 1,049,463 | 516.8 | 951.5 | 6,895 | <15,633 | 0.94 (0.91 - 0.96) | <0.01 | 0.92 (0.84 - 1.02) | 0.12 |
| Meta-analysis (w/o USOC) | 139,322 | 228,054 | 138.9 | 175.8 | 1,359 | <3,204 | 0.93 (0.87 - 0.99) | 0.02 | 0.93 (0.84 - 1.02) | 0.15 |
| **PS-matching / on-treatment TAR without escalation** | | | | | | | | | | |
| CCAE | 38,636 | 64,976 | 24.8 | 40.8 | 153 | 334 | 0.82 (0.62 - 1.09) | 0.18 | 0.81 (0.61 - 1.07) | 0.15 |
| DAG | 10,485 | 2,040 | 10.3 | 2.0 | 0 | 0 | | | | |
| ODOD | 26,125 | 56,950 | 17.2 | 43.4 | 471 | 1,359 | 0.93 (0.80 - 1.07) | 0.33 | 0.97 (0.84 - 1.13) | 0.77 |
| MDCR | 7,704 | 14,462 | 6.3 | 11.7 | 180 | 405 | 0.83 (0.64 - 1.06) | 0.14 | 0.83 (0.65 - 1.07) | 0.17 |
| MDCD | 7,000 | 15,775 | 3.8 | 8.5 | 117 | 255 | 1.11 (0.80 - 1.54) | 0.54 | 1.07 (0.77 - 1.49) | 0.70 |
| Open Claims | 423,871 | 821,287 | 291.6 | 653.4 | 4,240 | 10,378 | 0.94 (0.89 - 0.99) | 0.02 | 0.93 (0.85 - 1.02) | 0.18 |
| OEHR | 35,910 | 81,969 | 14.7 | 39.4 | 177 | 527 | 1.06 (0.84 - 1.32) | 0.62 | 1.07 (0.85 - 1.34) | 0.58 |
| VA | 1,766 | 9,451 | 1.1 | 6.3 | 13 | 59 | 2.05 (0.78 - 4.93) | 0.13 | 1.85 (0.74 - 4.64) | 0.20 |
| SIDIAP | 19,487 | 5,438 | 39.9 | 11.6 | 15 | <5 | 0.94 (0.12 - 20.7) | 0.97 | 1.01 (0.08 - 13.2) | 0.98 |
| Meta-analysis | 541,012 | 1,064,870 | 359.5 | 803.3 | 5,351 | 13,317 | 0.94 (0.90 - 0.99) | <0.01 | 0.94 (0.85 - 1.03) | 0.18 |
| Meta-analysis (w/o USOC) | 117,141 | 243,583 | 67.9 | 150.0 | 1,111 | 2,939 | 0.95 (0.84 - 1.07) | 0.41 | 0.96 (0.84 - 1.09) | 0.56 |
| **PS-stratification / on-treatment TAR without escalation** | | | | | | | | | | |
| CCAE | 44,957 | 66,985 | 29.2 | 41.9 | 177 | 339 | 0.85 (0.70 - 1.03) | 0.10 | 0.84 (0.69 - 1.03) | 0.10 |
| DAG | 10,516 | 2,367 | 10.3 | 2.3 | 0 | 0 | | | | |
| ODOD | 28,091 | 57,107 | 18.6 | 43.4 | 501 | 1,359 | 0.96 (0.86 - 1.06) | 0.41 | 1.00 (0.89 - 1.12) | 0.88 |
| MDCR | 8,325 | 14,834 | 6.8 | 12.0 | 198 | 413 | 0.85 (0.71 - 1.02) | 0.08 | 0.86 (0.71 - 1.03) | 0.14 |
| MDCD | 7,347 | 15,879 | 4.0 | 8.5 | 131 | 256 | 1.13 (0.90 - 1.42) | 0.28 | 1.08 (0.86 - 1.36) | 0.53 |
| Open Claims | 433,792 | 821,409 | 300.5 | 653.4 | 4,388 | 10,378 | 0.94 (0.90 - 0.97) | <0.01 | 0.92 (0.83 - 1.02) | 0.13 |
| OEHR | 38,353 | 82,056 | 16.0 | 39.4 | 184 | 527 | 1.00 (0.83 - 1.19) | 0.97 | 1.00 (0.84 - 1.20) | 0.89 |
| VA | 2,163 | 12,941 | 1.3 | 9.0 | 14 | 92 | 2.27 (1.03 - 4.81) | 0.04 | 2.00 (0.92 - 4.33) | 0.08 |
| SIDIAP | 19,596 | 7,072 | 40.0 | 15.7 | 15 | <5 | 0.86 (0.20 - 5.27) | 0.86 | 0.85 (0.17 - 4.34) | 0.85 |
| Meta-analysis | 553,518 | 1,042,391 | 371.0 | 790.1 | 5,448 | 13,016 | 0.94 (0.91 - 0.97) | <0.01 | 0.93 (0.85 - 1.02) | 0.11 |
| Meta-analysis (w/o USOC) | 119,726 | 220,982 | 70.5 | 136.7 | 1,060 | 2,638 | 0.93 (0.86 - 1.00) | 0.05 | 0.94 (0.85 - 1.03) | 0.23 |

##### 1.6.5 Glycemic control

**Supplementary Table 135:** Exposures, outcomes and risk estimates for Glycemic control in DPP4I (female) (T) vs SU (female) (C) new-user cohorts under four different propensity score (PS) adjustment / time-at-risk (TAR) designs across data sources. We report total patient exposure time in 1,000 patient-year (KYr) units, uncalibrated and calibrated hazard ratios (HRs) and their 95% confidence intervals (CIs). Greyed out entries did not pass study diagnostics and are included only for completeness.

| | Patients | | Exposure (KYr) | | Outcomes | | Uncalibrated | | Calibrated | |
|---|---|---|---|---|---|---|---|---|---|---|
| | T | C | T | C | T | C | HR (95% CI) | $p$ | HR (95% CI) | $p$ |
| **PS-matching / on-treatment TAR** | | | | | | | | | | |
| CCAE | 38,694 | 65,531 | 30.3 | 47.7 | 81 | 255 | 0.88 (0.59 - 1.32) | 0.55 | 0.85 (0.57 - 1.28) | 0.44 |
| DAG | 3,753 | 846 | 3.3 | 0.8 | 465 | 95 | 0.91 (0.70 - 1.20) | 0.50 | 0.95 (0.73 - 1.25) | 0.61 |
| ODOD | 18,261 | 42,267 | 13.0 | 32.7 | 1,376 | 3,227 | 0.95 (0.86 - 1.03) | 0.22 | 0.98 (0.89 - 1.08) | 0.74 |
| MDCR | 8,123 | 15,281 | 7.7 | 13.6 | 12 | 44 | 0.90 (0.36 - 2.10) | 0.81 | 0.91 (0.37 - 2.21) | 0.83 |
| MDCD | 7,427 | 16,757 | 5.3 | 11.0 | 0 | 0 | | | | |
| Open Claims | 439,873 | 852,269 | 384.3 | 811.8 | 0 | 0 | | | | |
| OEHR | 36,419 | 83,277 | 18.7 | 47.4 | 0 | 0 | | | | |
| VA | 1,766 | 9,359 | 1.3 | 7.2 | <10 | <10 | 0.55 (0.03 - 4.08) | 0.64 | 0.50 (0.04 - 6.11) | 0.59 |
| SIDIAP | 2,096 | 1,017 | 3.4 | 1.7 | 881 | 348 | 1.09 (0.92 - 1.30) | 0.31 | 1.14 (0.96 - 1.36) | 0.20 |
| Meta-analysis | 70,927 | 124,942 | 57.6 | 96.4 | 2,815 | 3,969 | 0.97 (0.89 - 1.05) | 0.46 | 0.96 (0.86 - 1.07) | 0.45 |
| Meta-analysis (w/o USOC) | 70,927 | 124,942 | 57.6 | 96.4 | 2,815 | 3,969 | 0.97 (0.90 - 1.04) | 0.37 | 0.97 (0.88 - 1.06) | 0.54 |
| **PS-stratification / on-treatment TAR** | | | | | | | | | | |
| CCAE | 44,889 | 66,951 | 35.5 | 48.4 | 111 | 371 | 0.70 (0.55 - 0.88) | <0.01 | 0.69 (0.55 - 0.88) | <0.01 |
| DAG | 3,759 | 1,004 | 3.3 | 0.9 | 465 | 124 | 0.89 (0.72 - 1.12) | 0.32 | 0.88 (0.70 - 1.11) | 0.35 |
| ODOD | 19,175 | 42,367 | 13.6 | 32.7 | 1,468 | 3,230 | 1.01 (0.94 - 1.08) | 0.81 | 1.04 (0.97 - 1.11) | 0.41 |
| MDCR | 8,751 | 15,548 | 8.3 | 13.9 | 15 | 98 | 0.62 (0.33 - 1.08) | 0.11 | 0.63 (0.35 - 1.14) | 0.13 |
| MDCD | 7,771 | 16,863 | 5.6 | 11.0 | 0 | 0 | | | | |
| Open Claims | 449,878 | 852,367 | 395.5 | 811.8 | 0 | 0 | | | | |
| OEHR | 38,897 | 83,366 | 20.2 | 47.5 | 0 | 0 | | | | |
| VA | 2,158 | 12,709 | 1.6 | 10.1 | <10 | 16 | 0.63 (0.03 - 5.55) | 0.73 | 0.55 (0.04 - 7.77) | 0.66 |
| SIDIAP | 2,111 | 1,580 | 3.4 | 2.8 | 885 | 545 | 1.17 (1.04 - 1.33) | 0.01 | 1.14 (1.00 - 1.30) | 0.09 |
| Meta-analysis | 78,685 | 127,450 | 64.1 | 98.6 | 2,944 | 4,368 | 0.91 (0.74 - 1.12) | 0.37 | 0.90 (0.72 - 1.13) | 0.36 |
| Meta-analysis (w/o USOC) | 78,685 | 127,450 | 64.1 | 98.6 | 2,944 | 4,368 | 0.93 (0.79 - 1.09) | 0.37 | 0.93 (0.78 - 1.11) | 0.43 |
| **PS-matching / on-treatment TAR without escalation** | | | | | | | | | | |
| CCAE | 38,694 | 65,531 | 24.9 | 41.0 | 72 | 232 | 0.86 (0.55 - 1.31) | 0.48 | 0.84 (0.54 - 1.30) | 0.44 |
| DAG | 3,753 | 846 | 3.0 | 0.7 | 415 | 94 | 0.90 (0.69 - 1.18) | 0.43 | 0.97 (0.73 - 1.28) | 0.62 |
| ODOD | 18,261 | 42,267 | 10.9 | 29.1 | 1,157 | 2,920 | 0.92 (0.83 - 1.01) | 0.09 | 0.96 (0.87 - 1.07) | 0.57 |
| MDCR | 8,123 | 15,281 | 6.7 | 12.5 | 9 | 42 | 0.67 (0.24 - 1.69) | 0.43 | 0.68 (0.25 - 1.81) | 0.44 |
| MDCD | 7,427 | 16,757 | 4.1 | 9.1 | 0 | 0 | | | | |
| Open Claims | 439,873 | 852,269 | 305.0 | 682.7 | 0 | 0 | | | | |
| OEHR | 36,419 | 83,277 | 15.0 | 40.2 | 0 | 0 | | | | |
| VA | 1,766 | 9,359 | 1.1 | 6.2 | <10 | <10 | 0.99 (0.05 - 8.28) | 0.99 | 0.89 (0.07 - 11.8) | 0.93 |
| SIDIAP | 2,096 | 1,017 | 2.3 | 1.3 | 684 | 287 | 1.16 (0.96 - 1.41) | 0.13 | 1.25 (1.02 - 1.52) | 0.05 |
| Meta-analysis | 70,927 | 124,942 | 47.7 | 84.7 | 2,337 | 3,575 | 0.96 (0.84 - 1.10) | 0.59 | 0.96 (0.82 - 1.12) | 0.60 |
| Meta-analysis (w/o USOC) | 70,927 | 124,942 | 47.7 | 84.7 | 2,337 | 3,575 | 0.96 (0.85 - 1.09) | 0.54 | 0.97 (0.85 - 1.11) | 0.69 |
| **PS-stratification / on-treatment TAR without escalation** | | | | | | | | | | |
| CCAE | 44,889 | 66,951 | 29.2 | 41.6 | 97 | 344 | 0.70 (0.54 - 0.89) | <0.01 | 0.69 (0.54 - 0.89) | <0.01 |
| DAG | 3,759 | 1,004 | 3.0 | 0.8 | 415 | 120 | 0.83 (0.66 - 1.05) | 0.12 | 0.83 (0.66 - 1.05) | 0.19 |
| ODOD | 19,175 | 42,367 | 11.4 | 29.2 | 1,230 | 2,923 | 0.99 (0.92 - 1.06) | 0.77 | 1.04 (0.96 - 1.12) | 0.52 |
| MDCR | 8,751 | 15,548 | 7.2 | 12.7 | 12 | 90 | 0.61 (0.30 - 1.11) | 0.13 | 0.61 (0.32 - 1.17) | 0.14 |
| MDCD | 7,771 | 16,863 | 4.3 | 9.2 | 0 | 0 | | | | |
| Open Claims | 449,878 | 852,367 | 314.0 | 682.7 | 0 | 0 | | | | |
| OEHR | 38,897 | 83,366 | 16.3 | 40.3 | 0 | 0 | | | | |
| VA | 2,158 | 12,709 | 1.3 | 8.8 | <10 | 15 | 1.76 (0.08 - 15.0) | 0.67 | 1.54 (0.12 - 20.7) | 0.74 |
| SIDIAP | 2,111 | 1,580 | 2.3 | 2.2 | 688 | 456 | 1.26 (1.10 - 1.46) | <0.01 | 1.25 (1.08 - 1.45) | <0.01 |
| Meta-analysis | 78,685 | 127,450 | 53.1 | 86.5 | 2,442 | 3,933 | 0.90 (0.71 - 1.14) | 0.40 | 0.90 (0.70 - 1.15) | 0.40 |
| Meta-analysis (w/o USOC) | 78,685 | 127,450 | 53.1 | 86.5 | 2,442 | 3,933 | 0.92 (0.75 - 1.12) | 0.38 | 0.93 (0.75 - 1.14) | 0.48 |

##### 1.6.6 Hospitalization with heart failure

**Supplementary Table 136:** Exposures, outcomes and risk estimates for Hospitalization with heart failure in DPP4I (female) (T) vs SU (female) (C) new-user cohorts under four different propensity score (PS) adjustment / time-at-risk (TAR) designs across data sources. We report total patient exposure time in 1,000 patient-year (KYr) units, uncalibrated and calibrated hazard ratios (HRs) and their 95% confidence intervals (CIs). Greyed out entries did not pass study diagnostics and are included only for completeness.

| | Patients | | Exposure (KYr) | | Outcomes | | Uncalibrated | | Calibrated | |
|---|---|---|---|---|---|---|---|---|---|---|
| | T | C | T | C | T | C | HR (95% CI) | $p$ | HR (95% CI) | $p$ |
| **PS-matching / on-treatment TAR** | | | | | | | | | | |
| CCAE | 38,432 | 64,723 | 30.1 | 47.2 | 155 | 302 | 1.07 (0.82 - 1.41) | 0.61 | 1.04 (0.79 - 1.36) | 0.81 |
| DAG | 10,485 | 2,040 | 11.4 | 2.1 | 0 | 0 | | | | |
| ODOD | 25,936 | 56,484 | 20.3 | 48.4 | 452 | 1,268 | 0.89 (0.76 - 1.03) | 0.12 | 0.92 (0.78 - 1.07) | 0.35 |
| MDCR | 7,431 | 13,869 | 7.0 | 12.2 | 186 | 451 | 0.84 (0.65 - 1.07) | 0.16 | 0.85 (0.66 - 1.08) | 0.21 |
| MDCD | 6,798 | 15,306 | 4.8 | 9.9 | 114 | 250 | 0.88 (0.63 - 1.21) | 0.43 | 0.85 (0.61 - 1.17) | 0.34 |
| Open Claims | 420,971 | 815,544 | 365.9 | 771.4 | 4,146 | 10,029 | 0.88 (0.84 - 0.93) | <0.01 | 0.88 (0.79 - 0.97) | 0.01 |
| OEHR | 35,723 | 81,503 | 18.1 | 45.7 | 233 | 751 | 0.96 (0.78 - 1.17) | 0.67 | 0.97 (0.79 - 1.19) | 0.78 |
| VA | 1,778 | 9,366 | 1.3 | 7.2 | <10 | 39 | 2.45 (0.71 - 7.62) | 0.14 | 2.21 (0.67 - 7.25) | 0.19 |
| SIDIAP | 19,369 | 5,425 | 53.5 | 14.2 | 58 | <5 | 2.60 (0.95 - 9.17) | 0.10 | 2.72 (0.87 - 8.46) | 0.09 |
| Meta-analysis | 556,438 | 1,062,220 | 501.1 | 956.2 | <5,354 | <13,095 | 0.89 (0.85 - 0.94) | <0.01 | 0.88 (0.81 - 0.96) | <0.01 |
| Meta-analysis (w/o USOC) | 135,467 | 246,676 | 135.2 | 184.8 | <1,208 | <3,066 | 0.94 (0.83 - 1.06) | 0.32 | 0.94 (0.82 - 1.07) | 0.41 |
| **PS-stratification / on-treatment TAR** | | | | | | | | | | |
| CCAE | 44,755 | 66,729 | 35.4 | 48.4 | 177 | 311 | 0.95 (0.78 - 1.15) | 0.60 | 0.94 (0.77 - 1.14) | 0.53 |
| DAG | 10,516 | 2,367 | 11.4 | 2.5 | 0 | 0 | | | | |
| ODOD | 27,920 | 56,639 | 22.0 | 48.4 | 482 | 1,270 | 0.93 (0.84 - 1.04) | 0.22 | 0.96 (0.86 - 1.08) | 0.58 |
| MDCR | 8,032 | 14,238 | 7.6 | 12.6 | 203 | 466 | 0.78 (0.66 - 0.93) | <0.01 | 0.79 (0.66 - 0.94) | 0.02 |
| MDCD | 7,124 | 15,407 | 5.0 | 9.9 | 125 | 250 | 0.95 (0.75 - 1.19) | 0.66 | 0.91 (0.72 - 1.15) | 0.48 |
| Open Claims | 430,708 | 815,640 | 376.7 | 771.4 | 4,304 | 10,030 | 0.90 (0.87 - 0.94) | <0.01 | 0.89 (0.80 - 0.98) | 0.02 |
| OEHR | 38,193 | 81,585 | 19.7 | 45.8 | 241 | 751 | 0.92 (0.79 - 1.07) | 0.26 | 0.93 (0.79 - 1.08) | 0.37 |
| VA | 2,167 | 12,966 | 1.6 | 10.4 | <10 | 71 | 0.75 (0.29 - 1.76) | 0.53 | 0.65 (0.26 - 1.61) | 0.36 |
| SIDIAP | 19,541 | 7,057 | 53.6 | 19.4 | 58 | 10 | 2.35 (1.15 - 5.26) | 0.03 | 2.28 (1.06 - 4.88) | 0.04 |
| Meta-analysis | 569,149 | 1,041,888 | 514.9 | 945.9 | 5,465 | 12,838 | 0.90 (0.87 - 0.93) | <0.01 | 0.89 (0.81 - 0.99) | 0.02 |
| Meta-analysis (w/o USOC) | 138,441 | 226,248 | 138.3 | 174.5 | 1,161 | 2,808 | 0.92 (0.81 - 1.04) | 0.18 | 0.92 (0.80 - 1.06) | 0.27 |
| **PS-matching / on-treatment TAR without escalation** | | | | | | | | | | |
| CCAE | 38,432 | 64,723 | 24.7 | 40.6 | 131 | 263 | 1.11 (0.82 - 1.48) | 0.50 | 1.09 (0.81 - 1.46) | 0.59 |
| DAG | 10,485 | 2,040 | 10.3 | 2.0 | 0 | 0 | | | | |
| ODOD | 25,936 | 56,484 | 17.1 | 43.1 | 369 | 1,108 | 0.92 (0.78 - 1.09) | 0.34 | 0.97 (0.82 - 1.14) | 0.73 |
| MDCR | 7,431 | 13,869 | 6.1 | 11.2 | 159 | 410 | 0.83 (0.63 - 1.07) | 0.15 | 0.83 (0.64 - 1.08) | 0.18 |
| MDCD | 6,798 | 15,306 | 3.7 | 8.2 | 86 | 201 | 0.83 (0.57 - 1.19) | 0.32 | 0.80 (0.55 - 1.15) | 0.24 |
| Open Claims | 420,971 | 815,544 | 290.7 | 649.4 | 3,313 | 8,426 | 0.89 (0.84 - 0.95) | <0.01 | 0.89 (0.81 - 0.98) | 0.03 |
| OEHR | 35,723 | 81,503 | 14.6 | 39.1 | 164 | 561 | 0.91 (0.72 - 1.14) | 0.42 | 0.92 (0.73 - 1.16) | 0.50 |
| VA | 1,778 | 9,366 | 1.1 | 6.2 | <10 | 35 | 2.00 (0.51 - 6.59) | 0.29 | 1.80 (0.50 - 6.45) | 0.37 |
| SIDIAP | 19,369 | 5,425 | 39.8 | 11.5 | 37 | <5 | 1.76 (0.53 - 8.01) | 0.41 | 1.89 (0.49 - 7.33) | 0.36 |
| Meta-analysis | 556,438 | 1,062,220 | 397.8 | 809.4 | <4,269 | <11,009 | 0.90 (0.86 - 0.95) | <0.01 | 0.90 (0.81 - 0.99) | 0.03 |
| Meta-analysis (w/o USOC) | 135,467 | 246,676 | 107.1 | 160.0 | <956 | <2,583 | 0.92 (0.83 - 1.03) | 0.15 | 0.93 (0.83 - 1.05) | 0.29 |
| **PS-stratification / on-treatment TAR without escalation** | | | | | | | | | | |
| CCAE | 44,755 | 66,729 | 29.1 | 41.7 | 148 | 270 | 0.95 (0.77 - 1.18) | 0.65 | 0.95 (0.76 - 1.17) | 0.62 |
| DAG | 10,516 | 2,367 | 10.3 | 2.3 | 0 | 0 | | | | |
| ODOD | 27,920 | 56,639 | 18.5 | 43.1 | 394 | 1,110 | 0.92 (0.82 - 1.04) | 0.19 | 0.97 (0.85 - 1.10) | 0.65 |
| MDCR | 8,032 | 14,238 | 6.6 | 11.5 | 175 | 424 | 0.77 (0.64 - 0.93) | <0.01 | 0.78 (0.64 - 0.94) | 0.02 |
| MDCD | 7,124 | 15,407 | 3.9 | 8.2 | 93 | 201 | 0.92 (0.70 - 1.19) | 0.53 | 0.88 (0.67 - 1.15) | 0.37 |
| Open Claims | 430,708 | 815,640 | 299.3 | 649.4 | 3,435 | 8,427 | 0.90 (0.87 - 0.94) | <0.01 | 0.89 (0.80 - 0.99) | 0.03 |
| OEHR | 38,193 | 81,585 | 15.9 | 39.1 | 171 | 561 | 0.92 (0.77 - 1.10) | 0.38 | 0.93 (0.77 - 1.11) | 0.44 |
| VA | 2,167 | 12,966 | 1.3 | 9.1 | <10 | 61 | 0.44 (0.12 - 1.29) | 0.18 | 0.39 (0.12 - 1.28) | 0.12 |
| SIDIAP | 19,541 | 7,057 | 39.9 | 15.7 | 37 | 10 | 1.63 (0.74 - 3.87) | 0.25 | 1.61 (0.70 - 3.71) | 0.27 |
| Meta-analysis | 569,149 | 1,041,888 | 409.2 | 800.6 | 4,360 | 10,802 | 0.90 (0.87 - 0.94) | <0.01 | 0.90 (0.82 - 0.98) | 0.02 |
| Meta-analysis (w/o USOC) | 138,441 | 226,248 | 109.9 | 151.2 | 925 | 2,375 | 0.90 (0.82 - 0.99) | 0.03 | 0.91 (0.81 - 1.02) | 0.13 |

##### 1.6.7 Measured renal dysfunction

**Supplementary Table 137:** Exposures, outcomes and risk estimates for Measured renal dysfunction in DPP4I (female) (T) vs SU (female) (C) new-user cohorts under four different propensity score (PS) adjustment / time-at-risk (TAR) designs across data sources. We report total patient exposure time in 1,000 patient-year (KYr) units, uncalibrated and calibrated hazard ratios (HRs) and their 95% confidence intervals (CIs). Greyed out entries did not pass study diagnostics and are included only for completeness.

| | Patients | | Exposure (KYr) | | Outcomes | | Uncalibrated | | Calibrated | |
|---|---|---|---|---|---|---|---|---|---|---|
| | T | C | T | C | T | C | HR (95% CI) | $p$ | HR (95% CI) | $p$ |
| **PS-matching / on-treatment TAR** | | | | | | | | | | |
| CCAE | 39,095 | 66,019 | 30.6 | 48.2 | 5 | 16 | | | | |
| DAG | 10,485 | 2,040 | 11.4 | 2.1 | 0 | 0 | | | | |
| ODOD | 27,459 | 60,070 | 21.7 | 52.2 | 52 | 82 | 1.55 (0.92 - 2.58) | 0.09 | 1.60 (0.96 - 2.69) | 0.08 |
| MDCR | 8,176 | 15,519 | 7.7 | 13.9 | 0 | <5 | | | | |
| MDCD | 7,427 | 16,757 | 5.3 | 11.0 | 0 | 0 | | | | |
| Open Claims | 439,873 | 852,269 | 384.3 | 811.8 | 0 | 0 | | | | |
| OEHR | 36,419 | 83,277 | 18.7 | 47.4 | 0 | 0 | | | | |
| VA | 1,805 | 9,455 | 1.4 | 7.3 | <10 | 20 | 0.93 (0.13 - 4.40) | 0.93 | 0.83 (0.14 - 4.89) | 0.84 |
| SIDIAP | 19,439 | 5,430 | 53.4 | 14.2 | 141 | 16 | 1.35 (0.74 - 2.65) | 0.36 | 1.41 (0.74 - 2.67) | 0.31 |
| Meta-analysis | 46,898 | 65,500 | 75.1 | 66.3 | 193 | 98 | 1.47 (0.98 - 2.19) | 0.06 | 1.45 (0.96 - 2.18) | 0.07 |
| Meta-analysis (w/o USOC) | 46,898 | 65,500 | 75.1 | 66.3 | 193 | 98 | 1.47 (0.98 - 2.19) | 0.06 | 1.47 (0.98 - 2.20) | 0.06 |
| **PS-stratification / on-treatment TAR** | | | | | | | | | | |
| CCAE | 45,465 | 67,954 | 36.0 | 49.3 | 7 | 16 | 0.46 (0.17 - 1.15) | 0.12 | 0.46 (0.18 - 1.19) | 0.11 |
| DAG | 10,516 | 2,367 | 11.4 | 2.5 | 0 | 0 | | | | |
| ODOD | 29,464 | 60,228 | 23.4 | 52.2 | 55 | 82 | 1.52 (1.06 - 2.18) | 0.02 | 1.57 (1.09 - 2.26) | 0.02 |
| MDCR | 8,813 | 15,840 | 8.3 | 14.2 | 0 | <5 | | | | |
| MDCD | 7,771 | 16,863 | 5.6 | 11.0 | 0 | 0 | | | | |
| Open Claims | 449,878 | 852,367 | 395.5 | 811.8 | 0 | 0 | | | | |
| OEHR | 38,897 | 83,366 | 20.2 | 47.5 | 0 | 0 | | | | |
| VA | 2,193 | 13,010 | 1.6 | 10.4 | <10 | 32 | 0.94 (0.19 - 3.62) | 0.93 | 0.82 (0.19 - 3.58) | 0.79 |
| SIDIAP | 19,548 | 7,062 | 53.5 | 19.4 | 143 | 19 | 1.35 (0.82 - 2.34) | 0.27 | 1.30 (0.77 - 2.21) | 0.33 |
| Meta-analysis | 94,477 | 135,244 | 112.9 | 120.9 | 205 | 117 | 1.13 (0.63 - 2.02) | 0.68 | 1.12 (0.62 - 2.01) | 0.72 |
| Meta-analysis (w/o USOC) | 94,477 | 135,244 | 112.9 | 120.9 | 205 | 117 | 1.15 (0.68 - 1.95) | 0.60 | 1.15 (0.68 - 1.97) | 0.60 |
| **PS-matching / on-treatment TAR without escalation** | | | | | | | | | | |
| CCAE | 39,095 | 66,019 | 25.2 | 41.5 | 5 | 13 | | | | |
| DAG | 10,485 | 2,040 | 10.3 | 2.0 | 0 | 0 | | | | |
| ODOD | 27,459 | 60,070 | 18.2 | 46.4 | 44 | 70 | 1.56 (0.91 - 2.66) | 0.10 | 1.64 (0.96 - 2.80) | 0.07 |
| MDCR | 8,176 | 15,519 | 6.7 | 12.7 | 0 | <5 | | | | |
| MDCD | 7,427 | 16,757 | 4.1 | 9.1 | 0 | 0 | | | | |
| Open Claims | 439,873 | 852,269 | 305.0 | 682.7 | 0 | 0 | | | | |
| OEHR | 36,419 | 83,277 | 15.0 | 40.2 | 0 | 0 | | | | |
| VA | 1,805 | 9,455 | 1.1 | 6.3 | <10 | 19 | 0.95 (0.13 - 4.64) | 0.95 | 0.85 (0.14 - 5.10) | 0.86 |
| SIDIAP | 19,439 | 5,430 | 39.7 | 11.6 | 108 | 14 | 1.57 (0.80 - 3.38) | 0.22 | 1.68 (0.82 - 3.46) | 0.16 |
| Meta-analysis | 46,898 | 65,500 | 57.9 | 57.9 | 152 | 84 | 1.56 (1.02 - 2.40) | 0.04 | 1.56 (1.00 - 2.41) | 0.05 |
| Meta-analysis (w/o USOC) | 46,898 | 65,500 | 57.9 | 57.9 | 152 | 84 | 1.56 (1.02 - 2.40) | 0.04 | 1.58 (1.03 - 2.43) | 0.04 |
| **PS-stratification / on-treatment TAR without escalation** | | | | | | | | | | |
| CCAE | 45,465 | 67,954 | 29.6 | 42.5 | 6 | 13 | 0.52 (0.17 - 1.44) | 0.23 | 0.52 (0.18 - 1.50) | 0.23 |
| DAG | 10,516 | 2,367 | 10.3 | 2.3 | 0 | 0 | | | | |
| ODOD | 29,464 | 60,228 | 19.6 | 46.4 | 46 | 70 | 1.71 (1.15 - 2.53) | <0.01 | 1.79 (1.21 - 2.67) | <0.01 |
| MDCR | 8,813 | 15,840 | 7.2 | 13.0 | 0 | <5 | | | | |
| MDCD | 7,771 | 16,863 | 4.3 | 9.2 | 0 | 0 | | | | |
| Open Claims | 449,878 | 852,367 | 314.0 | 682.7 | 0 | 0 | | | | |
| OEHR | 38,897 | 83,366 | 16.3 | 40.3 | 0 | 0 | | | | |
| VA | 2,193 | 13,010 | 1.3 | 9.1 | <10 | 29 | 0.76 (0.10 - 3.71) | 0.77 | 0.67 (0.11 - 4.04) | 0.66 |
| SIDIAP | 19,548 | 7,062 | 39.8 | 15.7 | 110 | 17 | 1.20 (0.69 - 2.20) | 0.54 | 1.19 (0.67 - 2.12) | 0.57 |
| Meta-analysis | 94,477 | 135,244 | 89.0 | 104.6 | 162 | 100 | 1.21 (0.70 - 2.09) | 0.49 | 1.21 (0.69 - 2.10) | 0.50 |
| Meta-analysis (w/o USOC) | 94,477 | 135,244 | 89.0 | 104.6 | 162 | 100 | 1.22 (0.71 - 2.09) | 0.48 | 1.23 (0.71 - 2.12) | 0.46 |

##### 1.6.8 Stroke

**Supplementary Table 138:** Exposures, outcomes and risk estimates for Stroke in DPP4I (female) (T) vs SU (female) (C) new-user cohorts under four different propensity score (PS) adjustment / time-at-risk (TAR) designs across data sources. We report total patient exposure time in 1,000 patient-year (KYr) units, uncalibrated and calibrated hazard ratios (HRs) and their 95% confidence intervals (CIs). Greyed out entries did not pass study diagnostics and are included only for completeness.

| | Patients | | Exposure (KYr) | | Outcomes | | Uncalibrated | | Calibrated | |
|---|---|---|---|---|---|---|---|---|---|---|
| | T | C | T | C | T | C | HR (95% CI) | $p$ | HR (95% CI) | $p$ |
| **PS-matching / on-treatment TAR** | | | | | | | | | | |
| CCAE | 38,640 | 65,140 | 30.3 | 47.5 | 108 | 178 | 1.08 (0.77 - 1.51) | 0.65 | 1.04 (0.75 - 1.46) | 0.81 |
| DAG | 10,485 | 2,040 | 11.4 | 2.1 | 0 | 0 | | | | |
| ODOD | 26,684 | 58,327 | 21.1 | 50.3 | 244 | 633 | 0.97 (0.79 - 1.20) | 0.80 | 1.01 (0.81 - 1.25) | 0.90 |
| MDCR | 7,786 | 14,724 | 7.3 | 13.1 | 111 | 264 | 0.68 (0.48 - 0.95) | 0.03 | 0.69 (0.49 - 0.97) | 0.04 |
| MDCD | 7,135 | 16,076 | 5.0 | 10.5 | 64 | 129 | 1.59 (0.99 - 2.53) | 0.05 | 1.53 (0.96 - 2.45) | 0.08 |
| Open Claims | 426,881 | 827,934 | 372.1 | 786.3 | 2,257 | 5,349 | 0.90 (0.84 - 0.97) | <0.01 | 0.90 (0.80 - 1.00) | 0.06 |
| OEHR | 36,097 | 82,419 | 18.5 | 46.7 | 82 | 295 | 0.80 (0.57 - 1.11) | 0.20 | 0.82 (0.59 - 1.14) | 0.24 |
| VA | 1,793 | 9,562 | 1.4 | 7.4 | <10 | 16 | 2.37 (0.66 - 7.62) | 0.17 | 2.14 (0.63 - 7.24) | 0.23 |
| SIDIAP | 19,363 | 5,414 | 53.3 | 14.1 | 112 | 23 | 0.90 (0.50 - 1.67) | 0.72 | 0.93 (0.51 - 1.71) | 0.83 |
| Meta-analysis | 562,586 | 1,070,034 | 507.5 | 968.5 | 2,978 | 6,871 | 0.91 (0.86 - 0.97) | <0.01 | 0.90 (0.82 - 0.99) | 0.03 |
| Meta-analysis (w/o USOC) | 135,705 | 242,100 | 135.4 | 182.2 | 721 | 1,522 | 0.95 (0.77 - 1.16) | 0.60 | 0.95 (0.77 - 1.17) | 0.64 |
| **PS-stratification / on-treatment TAR** | | | | | | | | | | |
| CCAE | 44,968 | 67,059 | 35.6 | 48.6 | 123 | 181 | 0.97 (0.76 - 1.23) | 0.80 | 0.96 (0.75 - 1.22) | 0.73 |
| DAG | 10,516 | 2,367 | 11.4 | 2.5 | 0 | 0 | | | | |
| ODOD | 28,658 | 58,485 | 22.7 | 50.3 | 254 | 633 | 1.00 (0.85 - 1.16) | 0.98 | 1.03 (0.88 - 1.20) | 0.75 |
| MDCR | 8,390 | 15,031 | 7.9 | 13.3 | 120 | 268 | 0.75 (0.59 - 0.93) | 0.01 | 0.76 (0.60 - 0.95) | 0.02 |
| MDCD | 7,475 | 16,180 | 5.3 | 10.5 | 69 | 129 | 1.11 (0.81 - 1.52) | 0.51 | 1.07 (0.78 - 1.47) | 0.69 |
| Open Claims | 436,812 | 828,032 | 383.1 | 786.3 | 2,350 | 5,350 | 0.90 (0.85 - 0.94) | <0.01 | 0.88 (0.79 - 0.98) | 0.02 |
| OEHR | 38,563 | 82,508 | 20.0 | 46.8 | 86 | 295 | 0.77 (0.59 - 0.99) | 0.04 | 0.78 (0.60 - 1.01) | 0.06 |
| VA | 2,180 | 13,066 | 1.6 | 10.5 | 10 | 25 | 2.88 (0.98 - 8.36) | 0.05 | 2.50 (0.86 - 7.30) | 0.10 |
| SIDIAP | 19,471 | 7,043 | 53.3 | 19.3 | 112 | 30 | 0.93 (0.60 - 1.50) | 0.76 | 0.90 (0.57 - 1.43) | 0.67 |
| Meta-analysis | 576,862 | 1,058,158 | 522.7 | 964.7 | 3,045 | 6,757 | 0.90 (0.86 - 0.94) | <0.01 | 0.89 (0.80 - 0.98) | 0.02 |
| Meta-analysis (w/o USOC) | 140,050 | 230,126 | 139.5 | 178.4 | 695 | 1,407 | 0.88 (0.77 - 1.01) | 0.07 | 0.89 (0.76 - 1.03) | 0.12 |
| **PS-matching / on-treatment TAR without escalation** | | | | | | | | | | |
| CCAE | 38,640 | 65,140 | 24.9 | 40.9 | 89 | 151 | 1.11 (0.78 - 1.59) | 0.55 | 1.09 (0.76 - 1.57) | 0.63 |
| DAG | 10,485 | 2,040 | 10.3 | 2.0 | 0 | 0 | | | | |
| ODOD | 26,684 | 58,327 | 17.7 | 44.7 | 217 | 564 | 1.00 (0.79 - 1.25) | 0.98 | 1.04 (0.83 - 1.31) | 0.73 |
| MDCR | 7,786 | 14,724 | 6.4 | 12.0 | 89 | 238 | 0.60 (0.41 - 0.87) | <0.01 | 0.61 (0.42 - 0.88) | 0.01 |
| MDCD | 7,135 | 16,076 | 3.9 | 8.7 | 52 | 107 | 1.49 (0.90 - 2.47) | 0.12 | 1.44 (0.87 - 2.38) | 0.17 |
| Open Claims | 426,881 | 827,934 | 295.3 | 661.6 | 1,827 | 4,532 | 0.90 (0.83 - 0.97) | <0.01 | 0.89 (0.80 - 1.00) | 0.06 |
| OEHR | 36,097 | 82,419 | 14.8 | 39.7 | 57 | 223 | 0.80 (0.53 - 1.18) | 0.27 | 0.81 (0.54 - 1.20) | 0.30 |
| VA | 1,793 | 9,562 | 1.1 | 6.4 | <10 | 11 | 2.27 (0.53 - 8.99) | 0.26 | 2.04 (0.49 - 8.46) | 0.33 |
| SIDIAP | 19,363 | 5,414 | 39.6 | 11.5 | 90 | 20 | 0.75 (0.38 - 1.55) | 0.43 | 0.81 (0.40 - 1.62) | 0.56 |
| Meta-analysis | 562,586 | 1,070,034 | 402.7 | 819.1 | 2,421 | 5,835 | 0.92 (0.79 - 1.06) | 0.24 | 0.91 (0.77 - 1.08) | 0.28 |
| Meta-analysis (w/o USOC) | 135,705 | 242,100 | 107.3 | 157.5 | 594 | 1,303 | 0.92 (0.73 - 1.16) | 0.50 | 0.93 (0.73 - 1.18) | 0.57 |
| **PS-stratification / on-treatment TAR without escalation** | | | | | | | | | | |
| CCAE | 44,968 | 67,059 | 29.3 | 41.9 | 103 | 153 | 1.00 (0.77 - 1.31) | 0.98 | 1.00 (0.76 - 1.30) | 0.94 |
| DAG | 10,516 | 2,367 | 10.3 | 2.3 | 0 | 0 | | | | |
| ODOD | 28,658 | 58,485 | 19.1 | 44.8 | 227 | 564 | 1.06 (0.90 - 1.25) | 0.48 | 1.11 (0.94 - 1.31) | 0.26 |
| MDCR | 8,390 | 15,031 | 6.9 | 12.2 | 96 | 242 | 0.71 (0.55 - 0.90) | <0.01 | 0.71 (0.55 - 0.91) | 0.01 |
| MDCD | 7,475 | 16,180 | 4.2 | 8.8 | 57 | 107 | 1.10 (0.78 - 1.54) | 0.59 | 1.05 (0.74 - 1.49) | 0.79 |
| Open Claims | 436,812 | 828,032 | 304.2 | 661.6 | 1,900 | 4,533 | 0.90 (0.86 - 0.96) | <0.01 | 0.89 (0.80 - 0.99) | 0.04 |
| OEHR | 38,563 | 82,508 | 16.1 | 39.7 | 60 | 223 | 0.72 (0.53 - 0.96) | 0.03 | 0.72 (0.53 - 0.98) | 0.04 |
| VA | 2,180 | 13,066 | 1.3 | 9.2 | <10 | 19 | 2.93 (0.85 - 10.2) | 0.09 | 2.58 (0.74 - 8.94) | 0.14 |
| SIDIAP | 19,471 | 7,043 | 39.7 | 15.6 | 90 | 26 | 0.94 (0.58 - 1.59) | 0.82 | 0.93 (0.56 - 1.56) | 0.80 |
| Meta-analysis | 576,862 | 1,058,158 | 415.2 | 815.9 | 2,476 | 5,741 | 0.89 (0.78 - 1.02) | 0.09 | 0.89 (0.76 - 1.04) | 0.14 |
| Meta-analysis (w/o USOC) | 140,050 | 230,126 | 111.0 | 154.3 | 576 | 1,208 | 0.88 (0.72 - 1.07) | 0.19 | 0.89 (0.73 - 1.09) | 0.27 |

##### 1.6.9 Sudden cardiac death

**Supplementary Table 139:** Exposures, outcomes and risk estimates for Sudden cardiac death in DPP4I (female) (T) vs SU (female) (C) new-user cohorts under four different propensity score (PS) adjustment / time-at-risk (TAR) designs across data sources. We report total patient exposure time in 1,000 patient-year (KYr) units, uncalibrated and calibrated hazard ratios (HRs) and their 95% confidence intervals (CIs). Greyed out entries did not pass study diagnostics and are included only for completeness.

| | Patients | | Exposure (KYr) | | Outcomes | | Uncalibrated | | Calibrated | |
|---|---|---|---|---|---|---|---|---|---|---|
| | T | C | T | C | T | C | HR (95% CI) | $p$ | HR (95% CI) | $p$ |
| **PS-matching / on-treatment TAR** | | | | | | | | | | |
| CCAE | 39,112 | 66,040 | 30.7 | 48.2 | 11 | 48 | 0.73 (0.29 - 1.69) | 0.48 | 0.70 (0.29 - 1.69) | 0.43 |
| DAG | 10,485 | 2,040 | 11.4 | 2.1 | 0 | 0 | | | | |
| ODOD | 27,462 | 60,125 | 21.8 | 52.2 | 61 | 163 | 0.79 (0.50 - 1.22) | 0.30 | 0.82 (0.52 - 1.28) | 0.39 |
| MDCR | 8,154 | 15,491 | 7.7 | 13.9 | 14 | 44 | 0.55 (0.23 - 1.23) | 0.17 | 0.56 (0.24 - 1.30) | 0.18 |
| MDCD | 7,412 | 16,700 | 5.3 | 11.0 | 19 | 35 | 1.76 (0.77 - 3.96) | 0.18 | 1.70 (0.75 - 3.84) | 0.21 |
| Open Claims | 439,226 | 850,867 | 383.6 | 810.3 | 462 | 1,077 | 0.98 (0.83 - 1.14) | 0.77 | 0.97 (0.81 - 1.16) | 0.74 |
| OEHR | 36,404 | 83,240 | 18.7 | 47.4 | 20 | 81 | 0.83 (0.41 - 1.57) | 0.58 | 0.84 (0.43 - 1.64) | 0.61 |
| VA | 1,820 | 9,662 | 1.4 | 7.5 | 0 | <10 | | | | |
| SIDIAP | 19,513 | 5,440 | 53.7 | 14.2 | 0 | 0 | | | | |
| Meta-analysis | 557,770 | 1,092,463 | 467.6 | 983.0 | 587 | 1,448 | 0.94 (0.82 - 1.08) | 0.42 | 0.93 (0.80 - 1.09) | 0.39 |
| Meta-analysis (w/o USOC) | 118,544 | 241,596 | 84.0 | 172.7 | 125 | 371 | 0.84 (0.62 - 1.14) | 0.27 | 0.84 (0.62 - 1.15) | 0.29 |
| **PS-stratification / on-treatment TAR** | | | | | | | | | | |
| CCAE | 45,480 | 67,974 | 36.0 | 49.4 | 16 | 49 | 0.62 (0.33 - 1.10) | 0.11 | 0.61 (0.33 - 1.11) | 0.11 |
| DAG | 10,516 | 2,367 | 11.4 | 2.5 | 0 | 0 | | | | |
| ODOD | 29,466 | 60,283 | 23.5 | 52.3 | 66 | 163 | 1.03 (0.76 - 1.39) | 0.84 | 1.06 (0.78 - 1.44) | 0.70 |
| MDCR | 8,792 | 15,812 | 8.3 | 14.1 | 14 | 47 | 0.59 (0.31 - 1.09) | 0.11 | 0.60 (0.32 - 1.13) | 0.12 |
| MDCD | 7,756 | 16,806 | 5.5 | 11.0 | 20 | 35 | 1.48 (0.80 - 2.68) | 0.20 | 1.42 (0.78 - 2.60) | 0.26 |
| Open Claims | 449,195 | 850,965 | 394.8 | 810.4 | 479 | 1,077 | 0.93 (0.84 - 1.04) | 0.23 | 0.92 (0.80 - 1.06) | 0.25 |
| OEHR | 38,881 | 83,329 | 20.2 | 47.5 | 20 | 81 | 0.70 (0.41 - 1.16) | 0.18 | 0.71 (0.42 - 1.20) | 0.20 |
| VA | 2,220 | 13,214 | 1.6 | 10.7 | 0 | <10 | | | | |
| SIDIAP | 19,626 | 7,073 | 53.8 | 19.4 | 0 | 0 | | | | |
| Meta-analysis | 571,814 | 1,078,363 | 482.7 | 973.6 | 595 | 1,417 | 0.91 (0.83 - 1.01) | 0.08 | 0.90 (0.79 - 1.04) | 0.14 |
| Meta-analysis (w/o USOC) | 122,619 | 227,398 | 88.0 | 163.2 | 116 | 340 | 0.78 (0.59 - 1.05) | 0.11 | 0.79 (0.58 - 1.06) | 0.12 |
| **PS-matching / on-treatment TAR without escalation** | | | | | | | | | | |
| CCAE | 39,112 | 66,040 | 25.2 | 41.5 | 8 | 35 | 0.59 (0.19 - 1.62) | 0.34 | 0.58 (0.20 - 1.72) | 0.33 |
| DAG | 10,485 | 2,040 | 10.3 | 2.0 | 0 | 0 | | | | |
| ODOD | 27,462 | 60,125 | 18.3 | 46.5 | 49 | 141 | 0.84 (0.51 - 1.34) | 0.48 | 0.88 (0.54 - 1.43) | 0.61 |
| MDCR | 8,154 | 15,491 | 6.7 | 12.7 | 10 | 41 | 0.49 (0.19 - 1.12) | 0.11 | 0.49 (0.20 - 1.20) | 0.12 |
| MDCD | 7,412 | 16,700 | 4.1 | 9.1 | 11 | 31 | 1.15 (0.42 - 2.94) | 0.78 | 1.10 (0.41 - 2.94) | 0.84 |
| Open Claims | 439,226 | 850,867 | 304.5 | 681.5 | 376 | 904 | 1.02 (0.86 - 1.21) | 0.80 | 1.02 (0.84 - 1.22) | 0.87 |
| OEHR | 36,404 | 83,240 | 15.0 | 40.2 | 11 | 49 | 0.65 (0.27 - 1.43) | 0.32 | 0.66 (0.29 - 1.53) | 0.33 |
| VA | 1,820 | 9,662 | 1.1 | 6.5 | 0 | <10 | | | | |
| SIDIAP | 19,513 | 5,440 | 40.0 | 11.6 | 0 | 0 | | | | |
| Meta-analysis | 557,770 | 1,092,463 | 373.7 | 831.4 | 465 | 1,201 | 0.90 (0.71 - 1.13) | 0.35 | 0.89 (0.70 - 1.14) | 0.36 |
| Meta-analysis (w/o USOC) | 118,544 | 241,596 | 69.2 | 149.9 | 89 | 297 | 0.75 (0.54 - 1.05) | 0.09 | 0.76 (0.54 - 1.06) | 0.11 |
| **PS-stratification / on-treatment TAR without escalation** | | | | | | | | | | |
| CCAE | 45,480 | 67,974 | 29.6 | 42.6 | 11 | 36 | 0.54 (0.25 - 1.07) | 0.09 | 0.54 (0.26 - 1.10) | 0.09 |
| DAG | 10,516 | 2,367 | 10.3 | 2.3 | 0 | 0 | | | | |
| ODOD | 29,466 | 60,283 | 19.7 | 46.5 | 53 | 141 | 1.00 (0.71 - 1.38) | 0.98 | 1.04 (0.74 - 1.46) | 0.81 |
| MDCR | 8,792 | 15,812 | 7.2 | 12.9 | 10 | 43 | 0.46 (0.21 - 0.91) | 0.04 | 0.46 (0.22 - 0.96) | 0.04 |
| MDCD | 7,756 | 16,806 | 4.3 | 9.1 | 12 | 31 | 1.02 (0.48 - 2.06) | 0.95 | 0.98 (0.47 - 2.03) | 0.95 |
| Open Claims | 449,195 | 850,965 | 313.4 | 681.5 | 389 | 904 | 0.95 (0.84 - 1.08) | 0.45 | 0.94 (0.80 - 1.10) | 0.43 |
| OEHR | 38,881 | 83,329 | 16.3 | 40.2 | 11 | 49 | 0.58 (0.28 - 1.12) | 0.13 | 0.59 (0.29 - 1.18) | 0.13 |
| VA | 2,220 | 13,214 | 1.3 | 9.3 | 0 | <10 | | | | |
| SIDIAP | 19,626 | 7,073 | 40.0 | 15.7 | 0 | 0 | | | | |
| Meta-analysis | 571,814 | 1,078,363 | 386.1 | 823.7 | 474 | 1,173 | 0.78 (0.58 - 1.05) | 0.10 | 0.78 (0.57 - 1.05) | 0.10 |
| Meta-analysis (w/o USOC) | 122,619 | 227,398 | 72.7 | 142.2 | 85 | 269 | 0.68 (0.45 - 1.02) | 0.06 | 0.68 (0.45 - 1.03) | 0.07 |

##### 1.6.10 Abnormal weight gain

**Supplementary Table 140:** Exposures, outcomes and risk estimates for Abnormal weight gain in DPP4I (female) (T) vs SU (female) (C) new-user cohorts under four different propensity score (PS) adjustment / time-at-risk (TAR) designs across data sources. We report total patient exposure time in 1,000 patient-year (KYr) units, uncalibrated and calibrated hazard ratios (HRs) and their 95% confidence intervals (CIs). Greyed out entries did not pass study diagnostics and are included only for completeness.

| | Patients | | Exposure (KYr) | | Outcomes | | Uncalibrated | | Calibrated | |
|---|---|---|---|---|---|---|---|---|---|---|
| | T | C | T | C | T | C | HR (95% CI) | $p$ | HR (95% CI) | $p$ |
| **PS-matching / on-treatment TAR** | | | | | | | | | | |
| CCAE | 36,796 | 62,846 | 28.8 | 45.8 | 205 | 377 | 1.01 (0.79 - 1.28) | 0.95 | 0.97 (0.76 - 1.24) | 0.82 |
| DAG | 10,380 | 2,020 | 11.2 | 2.1 | 15 | <5 | 0.56 (0.14 - 2.79) | 0.45 | 0.59 (0.13 - 2.63) | 0.50 |
| ODOD | 26,061 | 57,645 | 20.6 | 49.8 | 157 | 452 | 0.71 (0.55 - 0.92) | 0.01 | 0.74 (0.57 - 0.96) | 0.03 |
| MDCR | 8,073 | 15,327 | 7.6 | 13.7 | 24 | 54 | 0.65 (0.35 - 1.17) | 0.16 | 0.65 (0.36 - 1.20) | 0.18 |
| MDCD | 6,776 | 15,565 | 4.8 | 10.2 | 44 | 130 | 0.63 (0.37 - 1.04) | 0.08 | 0.61 (0.36 - 1.02) | 0.06 |
| Open Claims | 416,823 | 813,408 | 364.2 | 775.1 | 1,087 | 2,456 | 0.85 (0.77 - 0.94) | <0.01 | 0.84 (0.74 - 0.96) | 0.01 |
| OEHR | 34,545 | 79,985 | 17.6 | 45.1 | 153 | 432 | 0.83 (0.64 - 1.08) | 0.17 | 0.84 (0.65 - 1.10) | 0.22 |
| VA | 1,733 | 9,121 | 1.3 | 7.0 | <10 | 22 | 0.00 (0.00 - 2.08) | 0.22 | 0.00 (0.00 - 79831) | 0.21 |
| SIDIAP | 19,342 | 5,409 | 53.2 | 14.1 | 63 | 18 | 0.72 (0.36 - 1.52) | 0.38 | 0.75 (0.37 - 1.55) | 0.45 |
| Meta-analysis | 550,149 | 1,059,306 | 498.0 | 960.9 | <1,743 | 3,941 | 0.84 (0.77 - 0.91) | <0.01 | 0.83 (0.74 - 0.92) | <0.01 |
| Meta-analysis (w/o USOC) | 133,326 | 245,898 | 133.8 | 185.8 | <656 | 1,485 | 0.81 (0.69 - 0.94) | <0.01 | 0.81 (0.69 - 0.95) | 0.02 |
| **PS-stratification / on-treatment TAR** | | | | | | | | | | |
| CCAE | 42,511 | 64,683 | 33.6 | 46.9 | 255 | 380 | 0.81 (0.69 - 0.96) | 0.02 | 0.80 (0.68 - 0.95) | 0.01 |
| DAG | 10,411 | 2,349 | 11.3 | 2.5 | 15 | 5 | 0.75 (0.26 - 2.57) | 0.63 | 0.74 (0.23 - 2.35) | 0.62 |
| ODOD | 27,829 | 57,783 | 22.1 | 49.9 | 179 | 452 | 0.79 (0.65 - 0.94) | <0.01 | 0.81 (0.67 - 0.97) | 0.04 |
| MDCR | 8,672 | 15,644 | 8.2 | 14.0 | 26 | 55 | 0.72 (0.44 - 1.16) | 0.19 | 0.73 (0.45 - 1.19) | 0.22 |
| MDCD | 7,067 | 15,658 | 5.0 | 10.2 | 50 | 130 | 0.74 (0.52 - 1.03) | 0.08 | 0.71 (0.50 - 1.00) | 0.06 |
| Open Claims | 425,868 | 813,494 | 374.5 | 775.2 | 1,130 | 2,456 | 0.87 (0.81 - 0.94) | <0.01 | 0.86 (0.76 - 0.96) | 0.01 |
| OEHR | 36,753 | 80,070 | 18.9 | 45.2 | 168 | 432 | 0.89 (0.74 - 1.08) | 0.24 | 0.90 (0.75 - 1.09) | 0.32 |
| VA | 2,099 | 12,636 | 1.5 | 10.1 | <10 | 38 | 0.29 (0.06 - 1.09) | 0.09 | 0.25 (0.06 - 1.07) | 0.06 |
| SIDIAP | 19,454 | 7,041 | 53.3 | 19.3 | 63 | 24 | 1.09 (0.65 - 1.87) | 0.76 | 1.05 (0.62 - 1.79) | 0.85 |
| Meta-analysis | 561,087 | 1,038,715 | 510.6 | 950.3 | 1,821 | 3,799 | 0.86 (0.81 - 0.91) | <0.01 | 0.85 (0.76 - 0.95) | <0.01 |
| Meta-analysis (w/o USOC) | 135,219 | 225,221 | 136.1 | 175.2 | 691 | 1,343 | 0.83 (0.75 - 0.92) | <0.01 | 0.83 (0.74 - 0.94) | <0.01 |
| **PS-matching / on-treatment TAR without escalation** | | | | | | | | | | |
| CCAE | 36,796 | 62,846 | 23.7 | 39.5 | 163 | 310 | 1.04 (0.80 - 1.36) | 0.75 | 1.02 (0.78 - 1.34) | 0.85 |
| DAG | 10,380 | 2,020 | 10.1 | 2.0 | 12 | <5 | 0.41 (0.09 - 2.19) | 0.28 | 0.45 (0.09 - 2.20) | 0.33 |
| ODOD | 26,061 | 57,645 | 17.3 | 44.5 | 124 | 386 | 0.68 (0.51 - 0.89) | <0.01 | 0.71 (0.53 - 0.94) | 0.02 |
| MDCR | 8,073 | 15,327 | 6.6 | 12.5 | 18 | 47 | 0.63 (0.30 - 1.27) | 0.22 | 0.64 (0.31 - 1.32) | 0.22 |
| MDCD | 6,776 | 15,565 | 3.7 | 8.5 | 30 | 110 | 0.65 (0.36 - 1.14) | 0.15 | 0.63 (0.36 - 1.12) | 0.12 |
| Open Claims | 416,823 | 813,408 | 289.7 | 653.4 | 829 | 2,028 | 0.82 (0.73 - 0.92) | <0.01 | 0.81 (0.71 - 0.93) | <0.01 |
| OEHR | 34,545 | 79,985 | 14.2 | 38.4 | 106 | 337 | 0.70 (0.51 - 0.95) | 0.02 | 0.71 (0.52 - 0.96) | 0.03 |
| VA | 1,733 | 9,121 | 1.0 | 6.1 | <10 | 17 | | | | |
| SIDIAP | 19,342 | 5,409 | 39.6 | 11.5 | 49 | 18 | 0.69 (0.33 - 1.55) | 0.36 | 0.74 (0.34 - 1.62) | 0.46 |
| Meta-analysis | 548,416 | 1,050,185 | 394.7 | 808.3 | 1,319 | 3,236 | 0.79 (0.69 - 0.91) | <0.01 | 0.79 (0.67 - 0.93) | <0.01 |
| Meta-analysis (w/o USOC) | 131,593 | 236,777 | 105.0 | 154.9 | 490 | 1,208 | 0.77 (0.63 - 0.93) | <0.01 | 0.77 (0.64 - 0.94) | 0.01 |
| **PS-stratification / on-treatment TAR without escalation** | | | | | | | | | | |
| CCAE | 42,511 | 64,683 | 27.7 | 40.5 | 202 | 313 | 0.81 (0.67 - 0.98) | 0.03 | 0.81 (0.67 - 0.98) | 0.03 |
| DAG | 10,411 | 2,349 | 10.2 | 2.3 | 12 | 5 | 0.60 (0.19 - 2.15) | 0.41 | 0.60 (0.18 - 2.03) | 0.42 |
| ODOD | 27,829 | 57,783 | 18.6 | 44.5 | 145 | 386 | 0.79 (0.64 - 0.96) | 0.02 | 0.82 (0.67 - 1.01) | 0.08 |
| MDCR | 8,672 | 15,644 | 7.1 | 12.8 | 20 | 48 | 0.67 (0.38 - 1.15) | 0.16 | 0.68 (0.39 - 1.18) | 0.17 |
| MDCD | 7,067 | 15,658 | 3.9 | 8.5 | 36 | 110 | 0.64 (0.42 - 0.94) | 0.03 | 0.61 (0.41 - 0.91) | 0.02 |
| Open Claims | 425,868 | 813,494 | 297.8 | 653.4 | 860 | 2,028 | 0.84 (0.77 - 0.91) | <0.01 | 0.83 (0.73 - 0.94) | <0.01 |
| OEHR | 36,753 | 80,070 | 15.3 | 38.4 | 115 | 337 | 0.81 (0.65 - 1.01) | 0.07 | 0.82 (0.65 - 1.02) | 0.09 |
| VA | 2,099 | 12,636 | 1.2 | 8.8 | <10 | 30 | 0.65 (0.12 - 3.14) | 0.61 | 0.57 (0.11 - 2.96) | 0.51 |
| SIDIAP | 19,454 | 7,041 | 39.7 | 15.6 | 49 | 22 | 0.91 (0.52 - 1.64) | 0.75 | 0.90 (0.51 - 1.60) | 0.73 |
| Meta-analysis | 561,087 | 1,038,715 | 406.1 | 805.3 | 1,391 | 3,134 | 0.83 (0.77 - 0.88) | <0.01 | 0.82 (0.74 - 0.91) | <0.01 |
| Meta-analysis (w/o USOC) | 135,219 | 225,221 | 108.2 | 151.9 | 531 | 1,106 | 0.80 (0.72 - 0.90) | <0.01 | 0.81 (0.71 - 0.92) | <0.01 |

##### 1.6.11 Abnormal weight loss

**Supplementary Table 141:** Exposures, outcomes and risk estimates for Abnormal weight loss in DPP4I (female) (T) vs SU (female) (C) new-user cohorts under four different propensity score (PS) adjustment / time-at-risk (TAR) designs across data sources. We report total patient exposure time in 1,000 patient-year (KYr) units, uncalibrated and calibrated hazard ratios (HRs) and their 95% confidence intervals (CIs). Greyed out entries did not pass study diagnostics and are included only for completeness.

| | Patients | | Exposure (KYr) | | Outcomes | | Uncalibrated | | Calibrated | |
|---|---|---|---|---|---|---|---|---|---|---|
| | T | C | T | C | T | C | HR (95% CI) | $p$ | HR (95% CI) | $p$ |
| **PS-matching / on-treatment TAR** | | | | | | | | | | |
| CCAE | 38,258 | 64,600 | 29.8 | 47.1 | 209 | 251 | 1.27 (0.97 - 1.64) | 0.08 | 1.22 (0.94 - 1.59) | 0.15 |
| DAG | 10,197 | 1,981 | 11.0 | 2.1 | 79 | 15 | 1.65 (0.75 - 4.18) | 0.25 | 1.73 (0.73 - 4.09) | 0.23 |
| ODOD | 26,218 | 57,313 | 20.6 | 49.3 | 291 | 704 | 0.97 (0.80 - 1.17) | 0.72 | 1.00 (0.82 - 1.22) | 0.90 |
| MDCR | 7,809 | 14,790 | 7.3 | 13.1 | 105 | 167 | 1.18 (0.85 - 1.65) | 0.32 | 1.20 (0.86 - 1.67) | 0.31 |
| MDCD | 7,062 | 15,969 | 4.9 | 10.4 | 82 | 167 | 1.03 (0.67 - 1.55) | 0.91 | 0.99 (0.65 - 1.51) | 0.91 |
| Open Claims | 422,034 | 821,394 | 366.6 | 779.9 | 2,548 | 4,151 | 1.18 (1.10 - 1.27) | <0.01 | 1.17 (1.05 - 1.31) | <0.01 |
| OEHR | 35,345 | 80,877 | 18.0 | 45.6 | 204 | 525 | 0.92 (0.72 - 1.16) | 0.47 | 0.93 (0.74 - 1.18) | 0.56 |
| VA | 1,758 | 9,356 | 1.3 | 7.2 | <10 | 37 | 0.33 (0.05 - 1.24) | 0.17 | 0.29 (0.06 - 1.47) | 0.14 |
| SIDIAP | 19,270 | 5,415 | 53.0 | 14.1 | 184 | 34 | 1.69 (1.00 - 2.98) | 0.06 | 1.76 (1.02 - 3.04) | 0.05 |
| Meta-analysis | 567,951 | 1,071,695 | 512.5 | 968.7 | <3,712 | 6,051 | 1.11 (1.00 - 1.24) | 0.06 | 1.10 (0.96 - 1.25) | 0.16 |
| Meta-analysis (w/o USOC) | 145,917 | 250,301 | 145.9 | 188.8 | <1,164 | 1,900 | 1.09 (0.93 - 1.27) | 0.29 | 1.09 (0.93 - 1.28) | 0.33 |
| **PS-stratification / on-treatment TAR** | | | | | | | | | | |
| CCAE | 44,501 | 66,506 | 35.0 | 48.2 | 243 | 255 | 1.30 (1.08 - 1.57) | <0.01 | 1.28 (1.06 - 1.55) | 0.01 |
| DAG | 10,227 | 2,299 | 11.0 | 2.4 | 79 | 17 | 0.85 (0.49 - 1.58) | 0.60 | 0.84 (0.47 - 1.52) | 0.58 |
| ODOD | 28,127 | 57,467 | 22.2 | 49.3 | 328 | 704 | 1.01 (0.88 - 1.16) | 0.84 | 1.05 (0.91 - 1.20) | 0.57 |
| MDCR | 8,411 | 15,103 | 7.9 | 13.4 | 120 | 167 | 1.19 (0.93 - 1.52) | 0.17 | 1.21 (0.94 - 1.55) | 0.17 |
| MDCD | 7,383 | 16,069 | 5.2 | 10.4 | 88 | 167 | 0.95 (0.72 - 1.25) | 0.73 | 0.92 (0.70 - 1.21) | 0.56 |
| Open Claims | 431,527 | 821,490 | 377.1 | 779.9 | 2,654 | 4,151 | 1.26 (1.20 - 1.32) | <0.01 | 1.24 (1.11 - 1.37) | <0.01 |
| OEHR | 37,764 | 80,961 | 19.4 | 45.7 | 223 | 527 | 1.02 (0.87 - 1.21) | 0.79 | 1.03 (0.87 - 1.22) | 0.71 |
| VA | 2,135 | 12,725 | 1.6 | 10.2 | 12 | 55 | 1.09 (0.47 - 2.40) | 0.84 | 0.95 (0.42 - 2.13) | 0.89 |
| SIDIAP | 19,387 | 7,046 | 53.0 | 19.3 | 186 | 44 | 0.95 (0.66 - 1.39) | 0.78 | 0.92 (0.63 - 1.34) | 0.67 |
| Meta-analysis | 579,944 | 1,050,872 | 525.7 | 958.2 | 3,833 | 5,865 | 1.13 (1.02 - 1.25) | 0.02 | 1.12 (0.97 - 1.28) | 0.12 |
| Meta-analysis (w/o USOC) | 148,417 | 229,382 | 148.6 | 178.3 | 1,179 | 1,714 | 1.08 (0.98 - 1.20) | 0.12 | 1.09 (0.96 - 1.23) | 0.20 |
| **PS-matching / on-treatment TAR without escalation** | | | | | | | | | | |
| CCAE | 38,258 | 64,600 | 24.6 | 40.6 | 178 | 209 | 1.40 (1.05 - 1.86) | 0.02 | 1.37 (1.03 - 1.83) | 0.03 |
| DAG | 10,197 | 1,981 | 9.9 | 1.9 | 69 | 13 | 1.55 (0.66 - 4.25) | 0.36 | 1.66 (0.65 - 4.23) | 0.30 |
| ODOD | 26,218 | 57,313 | 17.3 | 43.8 | 241 | 628 | 0.96 (0.78 - 1.18) | 0.71 | 1.01 (0.82 - 1.24) | 0.90 |
| MDCR | 7,809 | 14,790 | 6.3 | 12.0 | 98 | 153 | 1.28 (0.90 - 1.83) | 0.17 | 1.29 (0.90 - 1.85) | 0.17 |
| MDCD | 7,062 | 15,969 | 3.9 | 8.6 | 58 | 146 | 1.18 (0.71 - 1.92) | 0.52 | 1.13 (0.69 - 1.86) | 0.63 |
| Open Claims | 422,034 | 821,394 | 290.9 | 656.3 | 2,071 | 3,496 | 1.17 (1.09 - 1.27) | <0.01 | 1.17 (1.05 - 1.30) | <0.01 |
| OEHR | 35,345 | 80,877 | 14.5 | 38.7 | 160 | 412 | 0.92 (0.71 - 1.19) | 0.52 | 0.93 (0.72 - 1.20) | 0.59 |
| VA | 1,758 | 9,356 | 1.1 | 6.3 | <10 | 32 | 0.48 (0.07 - 1.89) | 0.37 | 0.43 (0.08 - 2.21) | 0.31 |
| SIDIAP | 19,270 | 5,415 | 39.4 | 11.5 | 143 | 28 | 1.35 (0.76 - 2.51) | 0.32 | 1.45 (0.80 - 2.62) | 0.23 |
| Meta-analysis | 567,951 | 1,071,695 | 407.8 | 819.7 | <3,028 | 5,117 | 1.13 (1.01 - 1.27) | 0.04 | 1.13 (0.97 - 1.30) | 0.11 |
| Meta-analysis (w/o USOC) | 145,917 | 250,301 | 116.9 | 163.4 | <957 | 1,621 | 1.11 (0.95 - 1.29) | 0.17 | 1.12 (0.96 - 1.31) | 0.17 |
| **PS-stratification / on-treatment TAR without escalation** | | | | | | | | | | |
| CCAE | 44,501 | 66,506 | 28.8 | 41.6 | 208 | 213 | 1.44 (1.17 - 1.76) | <0.01 | 1.43 (1.16 - 1.76) | <0.01 |
| DAG | 10,227 | 2,299 | 9.9 | 2.2 | 69 | 15 | 0.79 (0.44 - 1.52) | 0.46 | 0.79 (0.42 - 1.47) | 0.48 |
| ODOD | 28,127 | 57,467 | 18.6 | 43.9 | 275 | 628 | 1.00 (0.86 - 1.17) | 0.96 | 1.05 (0.90 - 1.23) | 0.57 |
| MDCR | 8,411 | 15,103 | 6.8 | 12.3 | 109 | 153 | 1.27 (0.98 - 1.65) | 0.07 | 1.28 (0.99 - 1.66) | 0.08 |
| MDCD | 7,383 | 16,069 | 4.1 | 8.6 | 63 | 146 | 0.88 (0.64 - 1.19) | 0.41 | 0.84 (0.61 - 1.15) | 0.29 |
| Open Claims | 431,527 | 821,490 | 299.4 | 656.3 | 2,158 | 3,496 | 1.27 (1.21 - 1.35) | <0.01 | 1.25 (1.12 - 1.40) | <0.01 |
| OEHR | 37,764 | 80,961 | 15.7 | 38.8 | 175 | 413 | 1.07 (0.88 - 1.29) | 0.49 | 1.07 (0.89 - 1.30) | 0.48 |
| VA | 2,135 | 12,725 | 1.2 | 8.9 | <10 | 44 | 0.99 (0.37 - 2.46) | 0.99 | 0.87 (0.34 - 2.25) | 0.78 |
| SIDIAP | 19,387 | 7,046 | 39.5 | 15.6 | 145 | 36 | 0.98 (0.65 - 1.51) | 0.91 | 0.97 (0.63 - 1.48) | 0.87 |
| Meta-analysis | 579,944 | 1,050,872 | 418.7 | 810.6 | 3,139 | 4,954 | 1.16 (1.03 - 1.31) | 0.01 | 1.16 (1.00 - 1.34) | 0.05 |
| Meta-analysis (w/o USOC) | 148,417 | 229,382 | 119.4 | 154.4 | 981 | 1,458 | 1.13 (0.97 - 1.31) | 0.11 | 1.14 (0.97 - 1.34) | 0.13 |

##### 1.6.12 Acute pancreatitis

**Supplementary Table 142:** Exposures, outcomes and risk estimates for Acute pancreatitis in DPP4I (female) (T) vs SU (female) (C) new-user cohorts under four different propensity score (PS) adjustment / time-at-risk (TAR) designs across data sources. We report total patient exposure time in 1,000 patient-year (KYr) units, uncalibrated and calibrated hazard ratios (HRs) and their 95% confidence intervals (CIs). Greyed out entries did not pass study diagnostics and are included only for completeness.

| | Patients | | Exposure (KYr) | | Outcomes | | Uncalibrated | | Calibrated | |
|---|---|---|---|---|---|---|---|---|---|---|
| | T | C | T | C | T | C | HR (95% CI) | $p$ | HR (95% CI) | $p$ |
| **PS-matching / on-treatment TAR** | | | | | | | | | | |
| CCAE | 38,847 | 65,460 | 30.4 | 47.7 | 71 | 112 | 0.97 (0.62 - 1.50) | 0.90 | 0.94 (0.60 - 1.46) | 0.77 |
| DAG | 10,485 | 2,040 | 11.4 | 2.1 | 0 | 0 | | | | |
| ODOD | 27,307 | 59,635 | 21.6 | 51.8 | 62 | 155 | 1.02 (0.66 - 1.54) | 0.93 | 1.05 (0.69 - 1.61) | 0.81 |
| MDCR | 8,109 | 15,336 | 7.6 | 13.7 | 18 | 53 | 0.50 (0.21 - 1.09) | 0.10 | 0.51 (0.22 - 1.17) | 0.11 |
| MDCD | 7,291 | 16,429 | 5.2 | 10.8 | 24 | 58 | 0.98 (0.50 - 1.87) | 0.96 | 0.95 (0.49 - 1.84) | 0.88 |
| Open Claims | 434,881 | 840,518 | 379.6 | 800.1 | 817 | 1,714 | 0.95 (0.84 - 1.06) | 0.35 | 0.94 (0.81 - 1.08) | 0.40 |
| OEHR | 36,246 | 82,846 | 18.6 | 47.1 | 36 | 104 | 1.10 (0.67 - 1.78) | 0.70 | 1.12 (0.68 - 1.83) | 0.66 |
| VA | 1,802 | 9,564 | 1.4 | 7.4 | <10 | 11 | 0.74 (0.04 - 5.21) | 0.81 | 0.67 (0.06 - 7.91) | 0.75 |
| SIDIAP | 19,480 | 5,431 | 53.6 | 14.2 | 18 | 6 | 0.97 (0.28 - 3.87) | 0.97 | 1.02 (0.27 - 3.76) | 0.96 |
| Meta-analysis | 572,161 | 1,085,655 | 516.8 | 985.4 | 1,046 | 2,202 | 0.95 (0.86 - 1.05) | 0.33 | 0.94 (0.83 - 1.07) | 0.33 |
| Meta-analysis (w/o USOC) | 137,280 | 245,137 | 137.1 | 185.3 | 229 | 488 | 0.96 (0.77 - 1.21) | 0.75 | 0.97 (0.76 - 1.22) | 0.77 |
| **PS-stratification / on-treatment TAR** | | | | | | | | | | |
| CCAE | 45,212 | 67,373 | 35.7 | 48.9 | 77 | 115 | 0.97 (0.71 - 1.31) | 0.84 | 0.96 (0.70 - 1.30) | 0.78 |
| DAG | 10,516 | 2,367 | 11.4 | 2.5 | 0 | 0 | | | | |
| ODOD | 29,302 | 59,793 | 23.3 | 51.8 | 67 | 155 | 1.02 (0.75 - 1.38) | 0.88 | 1.05 (0.78 - 1.43) | 0.74 |
| MDCR | 8,746 | 15,653 | 8.3 | 14.0 | 18 | 54 | 0.60 (0.33 - 1.03) | 0.07 | 0.61 (0.34 - 1.07) | 0.09 |
| MDCD | 7,636 | 16,533 | 5.5 | 10.8 | 25 | 59 | 0.99 (0.59 - 1.63) | 0.97 | 0.95 (0.57 - 1.58) | 0.85 |
| Open Claims | 444,924 | 840,612 | 390.9 | 800.1 | 842 | 1,714 | 1.03 (0.95 - 1.12) | 0.48 | 1.01 (0.90 - 1.15) | 0.84 |
| OEHR | 38,728 | 82,935 | 20.1 | 47.2 | 39 | 105 | 1.02 (0.69 - 1.49) | 0.92 | 1.03 (0.70 - 1.52) | 0.87 |
| VA | 2,200 | 13,062 | 1.6 | 10.5 | <10 | 19 | 1.17 (0.14 - 6.11) | 0.87 | 1.01 (0.15 - 6.65) | 0.97 |
| SIDIAP | 19,591 | 7,065 | 53.7 | 19.4 | 18 | 6 | 0.68 (0.26 - 2.06) | 0.47 | 0.66 (0.23 - 1.87) | 0.43 |
| Meta-analysis | 586,503 | 1,073,431 | 532.1 | 981.3 | 1,061 | 2,149 | 1.01 (0.94 - 1.09) | 0.73 | 1.00 (0.89 - 1.13) | 0.92 |
| Meta-analysis (w/o USOC) | 141,579 | 232,819 | 141.2 | 181.2 | 219 | 435 | 0.94 (0.79 - 1.12) | 0.51 | 0.94 (0.78 - 1.14) | 0.56 |
| **PS-matching / on-treatment TAR without escalation** | | | | | | | | | | |
| CCAE | 38,847 | 65,460 | 25.0 | 41.1 | 55 | 86 | 0.96 (0.59 - 1.57) | 0.88 | 0.95 (0.58 - 1.55) | 0.83 |
| DAG | 10,485 | 2,040 | 10.3 | 2.0 | 0 | 0 | | | | |
| ODOD | 27,307 | 59,635 | 18.2 | 46.1 | 51 | 131 | 0.82 (0.50 - 1.32) | 0.43 | 0.86 (0.53 - 1.40) | 0.56 |
| MDCR | 8,109 | 15,336 | 6.7 | 12.5 | 14 | 46 | 0.49 (0.19 - 1.10) | 0.11 | 0.49 (0.20 - 1.18) | 0.11 |
| MDCD | 7,291 | 16,429 | 4.0 | 9.0 | 16 | 49 | 0.87 (0.40 - 1.81) | 0.72 | 0.84 (0.39 - 1.79) | 0.65 |
| Open Claims | 434,881 | 840,518 | 301.4 | 673.2 | 635 | 1,402 | 0.94 (0.82 - 1.07) | 0.33 | 0.93 (0.80 - 1.08) | 0.38 |
| OEHR | 36,246 | 82,846 | 14.9 | 40.0 | 27 | 87 | 1.14 (0.65 - 1.94) | 0.63 | 1.16 (0.67 - 2.00) | 0.60 |
| VA | 1,802 | 9,564 | 1.1 | 6.4 | <10 | <10 | 1.15 (0.05 - 13.2) | 0.92 | 1.03 (0.06 - 17.1) | 0.97 |
| SIDIAP | 19,480 | 5,431 | 39.9 | 11.6 | 13 | <5 | 0.84 (0.18 - 4.43) | 0.83 | 0.90 (0.18 - 4.42) | 0.90 |
| Meta-analysis | 552,681 | 1,080,224 | 370.1 | 821.9 | 798 | 1,801 | 0.93 (0.83 - 1.04) | 0.21 | 0.92 (0.80 - 1.07) | 0.28 |
| Meta-analysis (w/o USOC) | 117,800 | 239,706 | 68.8 | 148.7 | 163 | 399 | 0.89 (0.69 - 1.16) | 0.39 | 0.90 (0.69 - 1.17) | 0.44 |
| **PS-stratification / on-treatment TAR without escalation** | | | | | | | | | | |
| CCAE | 45,212 | 67,373 | 29.4 | 42.2 | 60 | 89 | 1.04 (0.74 - 1.47) | 0.81 | 1.04 (0.73 - 1.47) | 0.84 |
| DAG | 10,516 | 2,367 | 10.3 | 2.3 | 0 | 0 | | | | |
| ODOD | 29,302 | 59,793 | 19.6 | 46.1 | 56 | 131 | 1.08 (0.77 - 1.50) | 0.64 | 1.13 (0.81 - 1.58) | 0.47 |
| MDCR | 8,746 | 15,653 | 7.2 | 12.8 | 14 | 47 | 0.54 (0.28 - 0.99) | 0.06 | 0.55 (0.29 - 1.03) | 0.07 |
| MDCD | 7,636 | 16,533 | 4.2 | 9.0 | 16 | 50 | 0.80 (0.43 - 1.43) | 0.47 | 0.76 (0.42 - 1.40) | 0.39 |
| Open Claims | 444,924 | 840,612 | 310.4 | 673.3 | 653 | 1,402 | 1.02 (0.93 - 1.12) | 0.65 | 1.01 (0.88 - 1.15) | 0.92 |
| OEHR | 38,728 | 82,935 | 16.2 | 40.0 | 29 | 88 | 0.94 (0.59 - 1.45) | 0.78 | 0.94 (0.60 - 1.48) | 0.80 |
| VA | 2,200 | 13,062 | 1.3 | 9.2 | <10 | 16 | 2.68 (0.29 - 14.8) | 0.33 | 2.36 (0.33 - 16.9) | 0.40 |
| SIDIAP | 19,591 | 7,065 | 40.0 | 15.7 | 13 | <5 | 0.62 (0.19 - 2.56) | 0.48 | 0.62 (0.17 - 2.27) | 0.47 |
| Meta-analysis | 566,912 | 1,066,366 | 382.7 | 814.4 | 812 | 1,757 | 1.01 (0.93 - 1.10) | 0.78 | 1.01 (0.90 - 1.13) | 0.89 |
| Meta-analysis (w/o USOC) | 121,988 | 225,754 | 72.3 | 141.1 | 159 | 355 | 0.95 (0.76 - 1.21) | 0.70 | 0.97 (0.76 - 1.23) | 0.78 |

##### 1.6.13 Bone fracture

**Supplementary Table 143:** Exposures, outcomes and risk estimates for Bone fracture in DPP4I (female) (T) vs SU (female) (C) new-user cohorts under four different propensity score (PS) adjustment / time-at-risk (TAR) designs across data sources. We report total patient exposure time in 1,000 patient-year (KYr) units, uncalibrated and calibrated hazard ratios (HRs) and their 95% confidence intervals (CIs). Greyed out entries did not pass study diagnostics and are included only for completeness.

| | Patients | | Exposure (KYr) | | Outcomes | | Uncalibrated | | Calibrated | |
|---|---|---|---|---|---|---|---|---|---|---|
| | T | C | T | C | T | C | HR (95% CI) | $p$ | HR (95% CI) | $p$ |
| **PS-matching / on-treatment TAR** | | | | | | | | | | |
| CCAE | 35,539 | 60,320 | 27.2 | 43.2 | 573 | 892 | 0.98 (0.84 - 1.15) | 0.80 | 0.94 (0.80 - 1.11) | 0.52 |
| DAG | 9,003 | 1,776 | 9.5 | 1.8 | 236 | 55 | 0.86 (0.57 - 1.30) | 0.46 | 0.90 (0.59 - 1.36) | 0.62 |
| ODOD | 24,473 | 53,839 | 18.9 | 45.3 | 656 | 1,658 | 0.95 (0.83 - 1.08) | 0.43 | 0.98 (0.86 - 1.13) | 0.82 |
| MDCR | 6,884 | 13,051 | 6.3 | 11.3 | 286 | 551 | 0.85 (0.69 - 1.05) | 0.13 | 0.86 (0.70 - 1.06) | 0.19 |
| MDCD | 6,435 | 14,654 | 4.4 | 9.4 | 176 | 329 | 1.11 (0.85 - 1.44) | 0.45 | 1.07 (0.82 - 1.40) | 0.64 |
| Open Claims | 376,410 | 737,017 | 322.0 | 686.7 | 6,970 | 15,022 | 0.98 (0.94 - 1.02) | 0.27 | 0.97 (0.88 - 1.07) | 0.54 |
| OEHR | 33,656 | 76,804 | 17.0 | 42.5 | 380 | 1,111 | 0.92 (0.78 - 1.08) | 0.31 | 0.93 (0.79 - 1.10) | 0.43 |
| VA | 1,570 | 8,364 | 1.2 | 6.3 | 24 | 99 | 1.64 (0.82 - 3.15) | 0.15 | 1.48 (0.75 - 2.90) | 0.27 |
| SIDIAP | 16,703 | 4,925 | 44.6 | 12.4 | 1,013 | 252 | 1.26 (1.04 - 1.53) | 0.02 | 1.31 (1.08 - 1.60) | 0.01 |
| Meta-analysis | 510,673 | 970,750 | 451.2 | 858.8 | 10,314 | 19,969 | 0.98 (0.94 - 1.02) | 0.25 | 0.97 (0.89 - 1.05) | 0.43 |
| Meta-analysis (w/o USOC) | 134,263 | 233,733 | 129.1 | 172.1 | 3,344 | 4,947 | 1.00 (0.90 - 1.10) | 0.93 | 1.00 (0.89 - 1.12) | 0.89 |
| **PS-stratification / on-treatment TAR** | | | | | | | | | | |
| CCAE | 41,238 | 62,146 | 31.8 | 44.3 | 694 | 903 | 1.02 (0.92 - 1.13) | 0.70 | 1.01 (0.90 - 1.12) | 0.86 |
| DAG | 9,043 | 2,056 | 9.6 | 2.1 | 237 | 64 | 0.97 (0.71 - 1.34) | 0.86 | 0.96 (0.70 - 1.31) | 0.71 |
| ODOD | 26,228 | 53,992 | 20.3 | 45.3 | 709 | 1,659 | 0.97 (0.89 - 1.07) | 0.57 | 1.00 (0.91 - 1.11) | 0.87 |
| MDCR | 7,400 | 13,337 | 6.8 | 11.5 | 311 | 559 | 0.94 (0.81 - 1.08) | 0.37 | 0.95 (0.82 - 1.10) | 0.54 |
| MDCD | 6,705 | 14,781 | 4.7 | 9.4 | 185 | 331 | 1.08 (0.89 - 1.31) | 0.41 | 1.04 (0.86 - 1.26) | 0.71 |
| Open Claims | 385,029 | 737,112 | 331.6 | 686.7 | 7,245 | 15,022 | 0.97 (0.95 - 1.00) | 0.07 | 0.96 (0.87 - 1.05) | 0.38 |
| OEHR | 35,951 | 76,885 | 18.4 | 42.6 | 416 | 1,112 | 0.95 (0.84 - 1.07) | 0.37 | 0.96 (0.85 - 1.08) | 0.53 |
| VA | 1,908 | 11,555 | 1.4 | 9.0 | 29 | 146 | 1.40 (0.84 - 2.27) | 0.18 | 1.22 (0.74 - 2.00) | 0.45 |
| SIDIAP | 16,780 | 6,472 | 44.7 | 17.1 | 1,013 | 350 | 1.11 (0.96 - 1.27) | 0.16 | 1.07 (0.93 - 1.24) | 0.42 |
| Meta-analysis | 521,669 | 952,000 | 463.1 | 849.6 | 10,625 | 19,669 | 0.98 (0.95 - 1.00) | 0.08 | 0.97 (0.88 - 1.07) | 0.49 |
| Meta-analysis (w/o USOC) | 136,640 | 214,888 | 131.5 | 162.8 | 3,380 | 4,647 | 0.99 (0.94 - 1.04) | 0.74 | 0.99 (0.91 - 1.08) | 0.86 |
| **PS-matching / on-treatment TAR without escalation** | | | | | | | | | | |
| CCAE | 35,539 | 60,320 | 22.4 | 37.3 | 460 | 759 | 0.98 (0.83 - 1.16) | 0.82 | 0.96 (0.81 - 1.14) | 0.68 |
| DAG | 9,003 | 1,776 | 8.6 | 1.7 | 217 | 52 | 0.81 (0.54 - 1.24) | 0.33 | 0.87 (0.57 - 1.33) | 0.57 |
| ODOD | 24,473 | 53,839 | 15.9 | 40.4 | 533 | 1,478 | 0.94 (0.82 - 1.09) | 0.42 | 0.99 (0.85 - 1.14) | 0.85 |
| MDCR | 6,884 | 13,051 | 5.6 | 10.3 | 236 | 509 | 0.80 (0.64 - 1.00) | 0.05 | 0.80 (0.64 - 1.01) | 0.07 |
| MDCD | 6,435 | 14,654 | 3.5 | 7.8 | 131 | 276 | 0.99 (0.73 - 1.35) | 0.96 | 0.96 (0.70 - 1.30) | 0.78 |
| Open Claims | 376,410 | 737,017 | 256.3 | 578.6 | 5,528 | 12,655 | 0.97 (0.92 - 1.02) | 0.18 | 0.96 (0.88 - 1.05) | 0.45 |
| OEHR | 33,656 | 76,804 | 13.7 | 36.3 | 283 | 926 | 0.87 (0.73 - 1.05) | 0.15 | 0.88 (0.73 - 1.06) | 0.21 |
| VA | 1,570 | 8,364 | 0.9 | 5.5 | 18 | 84 | 1.49 (0.68 - 3.11) | 0.30 | 1.34 (0.63 - 2.89) | 0.45 |
| SIDIAP | 16,703 | 4,925 | 33.1 | 10.2 | 783 | 214 | 1.26 (1.02 - 1.58) | 0.04 | 1.36 (1.08 - 1.70) | 0.01 |
| Meta-analysis | 510,673 | 970,750 | 360.1 | 728.1 | 8,189 | 16,953 | 0.97 (0.93 - 1.00) | 0.08 | 0.96 (0.87 - 1.05) | 0.39 |
| Meta-analysis (w/o USOC) | 134,263 | 233,733 | 103.8 | 149.5 | 2,661 | 4,298 | 0.96 (0.86 - 1.07) | 0.44 | 0.97 (0.86 - 1.09) | 0.62 |
| **PS-stratification / on-treatment TAR without escalation** | | | | | | | | | | |
| CCAE | 41,238 | 62,146 | 26.3 | 38.3 | 558 | 768 | 1.00 (0.89 - 1.13) | 0.95 | 1.00 (0.88 - 1.12) | 0.87 |
| DAG | 9,043 | 2,056 | 8.6 | 1.9 | 218 | 60 | 0.98 (0.71 - 1.36) | 0.90 | 0.98 (0.70 - 1.36) | 0.73 |
| ODOD | 26,228 | 53,992 | 17.1 | 40.5 | 582 | 1,478 | 0.95 (0.86 - 1.05) | 0.32 | 0.99 (0.89 - 1.11) | 0.87 |
| MDCR | 7,400 | 13,337 | 6.0 | 10.6 | 260 | 515 | 0.89 (0.76 - 1.04) | 0.13 | 0.89 (0.76 - 1.05) | 0.23 |
| MDCD | 6,705 | 14,781 | 3.6 | 7.9 | 138 | 277 | 1.01 (0.82 - 1.26) | 0.89 | 0.97 (0.78 - 1.21) | 0.80 |
| Open Claims | 385,029 | 737,112 | 264.0 | 578.6 | 5,749 | 12,655 | 0.97 (0.94 - 1.00) | 0.06 | 0.95 (0.86 - 1.05) | 0.37 |
| OEHR | 35,951 | 76,885 | 14.8 | 36.3 | 310 | 927 | 0.89 (0.77 - 1.01) | 0.08 | 0.89 (0.77 - 1.02) | 0.13 |
| VA | 1,908 | 11,555 | 1.1 | 7.9 | 22 | 124 | 1.34 (0.75 - 2.31) | 0.32 | 1.17 (0.67 - 2.07) | 0.59 |
| SIDIAP | 16,780 | 6,472 | 33.2 | 13.9 | 783 | 289 | 1.14 (0.97 - 1.34) | 0.11 | 1.13 (0.96 - 1.33) | 0.22 |
| Meta-analysis | 521,669 | 952,000 | 370.0 | 720.2 | 8,460 | 16,692 | 0.97 (0.94 - 1.00) | 0.02 | 0.96 (0.88 - 1.05) | 0.40 |
| Meta-analysis (w/o USOC) | 136,640 | 214,888 | 106.1 | 141.5 | 2,711 | 4,037 | 0.97 (0.90 - 1.04) | 0.35 | 0.98 (0.89 - 1.07) | 0.66 |

##### 1.6.14 Diabetic ketoacidosis

**Supplementary Table 144:** Exposures, outcomes and risk estimates for Diabetic ketoacidosis in DPP4I (female) (T) vs SU (female) (C) new-user cohorts under four different propensity score (PS) adjustment / time-at-risk (TAR) designs across data sources. We report total patient exposure time in 1,000 patient-year (KYr) units, uncalibrated and calibrated hazard ratios (HRs) and their 95% confidence intervals (CIs). Greyed out entries did not pass study diagnostics and are included only for completeness.

| | Patients | | Exposure (KYr) | | Outcomes | | Uncalibrated | | Calibrated | |
|---|---|---|---|---|---|---|---|---|---|---|
| | T | C | T | C | T | C | HR (95% CI) | $p$ | HR (95% CI) | $p$ |
| **PS-matching / on-treatment TAR** | | | | | | | | | | |
| CCAE | 39,091 | 65,975 | 30.7 | 48.2 | 26 | 60 | 0.99 (0.52 - 1.83) | 0.98 | 0.96 (0.51 - 1.79) | 0.89 |
| DAG | 10,485 | 2,040 | 11.4 | 2.1 | 0 | 0 | | | | |
| ODOD | 27,468 | 60,103 | 21.8 | 52.3 | 36 | 98 | 1.22 (0.73 - 2.00) | 0.44 | 1.26 (0.76 - 2.08) | 0.38 |
| MDCR | 8,158 | 15,498 | 7.7 | 13.9 | 10 | 19 | 0.88 (0.30 - 2.37) | 0.81 | 0.89 (0.31 - 2.52) | 0.83 |
| MDCD | 7,403 | 16,677 | 5.2 | 11.0 | 17 | 39 | 1.30 (0.61 - 2.70) | 0.49 | 1.25 (0.59 - 2.64) | 0.56 |
| Open Claims | 439,162 | 850,402 | 383.5 | 810.3 | 354 | 732 | 1.08 (0.90 - 1.30) | 0.38 | 1.08 (0.88 - 1.31) | 0.48 |
| OEHR | 36,401 | 83,212 | 18.7 | 47.4 | 24 | 68 | 0.68 (0.36 - 1.22) | 0.21 | 0.69 (0.37 - 1.27) | 0.23 |
| VA | 1,820 | 9,661 | 1.4 | 7.5 | <10 | <10 | 0.00 (0.00 - 1.45) | 0.21 | 0.00 (0.00 - 43597) | 0.21 |
| SIDIAP | 19,514 | 5,440 | 53.7 | 14.2 | 0 | 0 | | | | |
| Meta-analysis | 557,683 | 1,091,867 | 467.5 | 982.9 | 467 | 1,016 | 1.06 (0.91 - 1.24) | 0.45 | 1.05 (0.88 - 1.24) | 0.58 |
| Meta-analysis (w/o USOC) | 118,521 | 241,465 | 84.0 | 172.6 | 113 | 284 | 1.00 (0.75 - 1.34) | 0.97 | 1.01 (0.75 - 1.35) | 0.95 |
| **PS-stratification / on-treatment TAR** | | | | | | | | | | |
| CCAE | 45,453 | 67,906 | 36.0 | 49.3 | 29 | 62 | 0.70 (0.43 - 1.10) | 0.13 | 0.69 (0.43 - 1.10) | 0.12 |
| DAG | 10,516 | 2,367 | 11.4 | 2.5 | 0 | 0 | | | | |
| ODOD | 29,474 | 60,261 | 23.5 | 52.3 | 37 | 98 | 0.89 (0.59 - 1.31) | 0.57 | 0.92 (0.62 - 1.37) | 0.68 |
| MDCR | 8,794 | 15,819 | 8.3 | 14.2 | 10 | 19 | 1.11 (0.48 - 2.41) | 0.80 | 1.12 (0.50 - 2.51) | 0.78 |
| MDCD | 7,748 | 16,782 | 5.5 | 11.0 | 17 | 39 | 1.18 (0.62 - 2.15) | 0.61 | 1.13 (0.61 - 2.10) | 0.70 |
| Open Claims | 449,170 | 850,500 | 394.7 | 810.3 | 362 | 732 | 1.09 (0.96 - 1.24) | 0.18 | 1.07 (0.92 - 1.26) | 0.37 |
| OEHR | 38,879 | 83,301 | 20.2 | 47.4 | 25 | 68 | 0.82 (0.50 - 1.31) | 0.41 | 0.83 (0.51 - 1.34) | 0.44 |
| VA | 2,219 | 13,213 | 1.6 | 10.6 | <10 | 12 | 0.37 (0.02 - 4.14) | 0.49 | 0.33 (0.02 - 5.24) | 0.43 |
| SIDIAP | 19,627 | 7,073 | 53.8 | 19.4 | 0 | 0 | | | | |
| Meta-analysis | 571,770 | 1,077,787 | 482.6 | 973.5 | 463 | 979 | 0.95 (0.78 - 1.16) | 0.63 | 0.94 (0.76 - 1.17) | 0.59 |
| Meta-analysis (w/o USOC) | 122,600 | 227,287 | 87.9 | 163.2 | 101 | 247 | 0.83 (0.65 - 1.06) | 0.14 | 0.83 (0.65 - 1.08) | 0.17 |
| **PS-matching / on-treatment TAR without escalation** | | | | | | | | | | |
| CCAE | 39,091 | 65,975 | 25.2 | 41.5 | 16 | 46 | 0.87 (0.41 - 1.77) | 0.71 | 0.86 (0.41 - 1.77) | 0.68 |
| DAG | 10,485 | 2,040 | 10.3 | 2.0 | 0 | 0 | | | | |
| ODOD | 27,468 | 60,103 | 18.2 | 46.5 | 29 | 78 | 1.22 (0.70 - 2.07) | 0.48 | 1.27 (0.74 - 2.19) | 0.39 |
| MDCR | 8,158 | 15,498 | 6.7 | 12.7 | 8 | 16 | 0.75 (0.23 - 2.12) | 0.61 | 0.76 (0.25 - 2.29) | 0.62 |
| MDCD | 7,403 | 16,677 | 4.1 | 9.1 | 12 | 26 | 1.17 (0.47 - 2.75) | 0.73 | 1.12 (0.47 - 2.71) | 0.80 |
| Open Claims | 439,162 | 850,402 | 304.5 | 681.6 | 248 | 542 | 1.02 (0.83 - 1.25) | 0.85 | 1.01 (0.82 - 1.26) | 0.90 |
| OEHR | 36,401 | 83,212 | 15.0 | 40.2 | 13 | 44 | 0.62 (0.27 - 1.32) | 0.24 | 0.63 (0.29 - 1.39) | 0.26 |
| VA | 1,820 | 9,661 | 1.1 | 6.5 | <10 | <10 | 0.00 (0.00 - 4.36) | 0.22 | 0.00 (0.00 - 99802) | 0.22 |
| SIDIAP | 19,514 | 5,440 | 40.0 | 11.6 | 0 | 0 | | | | |
| Meta-analysis | 557,683 | 1,091,867 | 373.7 | 831.6 | 326 | 752 | 1.00 (0.84 - 1.19) | 0.98 | 1.00 (0.82 - 1.21) | 0.95 |
| Meta-analysis (w/o USOC) | 118,521 | 241,465 | 69.2 | 149.9 | 78 | 210 | 0.96 (0.69 - 1.34) | 0.81 | 0.97 (0.69 - 1.35) | 0.85 |
| **PS-stratification / on-treatment TAR without escalation** | | | | | | | | | | |
| CCAE | 45,453 | 67,906 | 29.6 | 42.5 | 18 | 48 | 0.66 (0.36 - 1.15) | 0.15 | 0.65 (0.37 - 1.16) | 0.15 |
| DAG | 10,516 | 2,367 | 10.3 | 2.3 | 0 | 0 | | | | |
| ODOD | 29,474 | 60,261 | 19.7 | 46.5 | 30 | 78 | 0.96 (0.61 - 1.48) | 0.86 | 1.01 (0.64 - 1.57) | 0.96 |
| MDCR | 8,794 | 15,819 | 7.2 | 13.0 | 8 | 16 | 1.15 (0.45 - 2.70) | 0.76 | 1.15 (0.47 - 2.83) | 0.76 |
| MDCD | 7,748 | 16,782 | 4.3 | 9.1 | 12 | 26 | 1.24 (0.58 - 2.54) | 0.57 | 1.18 (0.56 - 2.49) | 0.66 |
| Open Claims | 449,170 | 850,500 | 313.5 | 681.7 | 252 | 542 | 1.07 (0.91 - 1.24) | 0.41 | 1.05 (0.88 - 1.26) | 0.61 |
| OEHR | 38,879 | 83,301 | 16.3 | 40.2 | 14 | 44 | 0.72 (0.37 - 1.33) | 0.31 | 0.72 (0.38 - 1.37) | 0.32 |
| VA | 2,219 | 13,213 | 1.3 | 9.3 | <10 | <10 | 0.40 (0.02 - 5.05) | 0.53 | 0.35 (0.02 - 6.09) | 0.47 |
| SIDIAP | 19,627 | 7,073 | 40.0 | 15.7 | 0 | 0 | | | | |
| Meta-analysis | 571,770 | 1,077,787 | 386.2 | 823.9 | 322 | 728 | 0.97 (0.80 - 1.18) | 0.76 | 0.97 (0.78 - 1.19) | 0.75 |
| Meta-analysis (w/o USOC) | 122,600 | 227,287 | 72.7 | 142.2 | 70 | 186 | 0.84 (0.63 - 1.12) | 0.23 | 0.85 (0.63 - 1.14) | 0.28 |

##### 1.6.15 Diarrhea

**Supplementary Table 145:** Exposures, outcomes and risk estimates for Diarrhea in DPP4I (female) (T) vs SU (female) (C) new-user cohorts under four different propensity score (PS) adjustment / time-at-risk (TAR) designs across data sources. We report total patient exposure time in 1,000 patient-year (KYr) units, uncalibrated and calibrated hazard ratios (HRs) and their 95% confidence intervals (CIs). Greyed out entries did not pass study diagnostics and are included only for completeness.

| | Patients | | Exposure (KYr) | | Outcomes | | Uncalibrated | | Calibrated | |
|---|---|---|---|---|---|---|---|---|---|---|
| | T | C | T | C | T | C | HR (95% CI) | $p$ | HR (95% CI) | $p$ |
| **PS-matching / on-treatment TAR** | | | | | | | | | | |
| CCAE | 33,476 | 56,954 | 25.5 | 40.5 | 871 | 1,460 | 1.02 (0.91 - 1.15) | 0.70 | 0.99 (0.87 - 1.12) | 0.81 |
| DAG | 10,305 | 1,988 | 11.1 | 2.1 | 29 | 7 | 1.39 (0.44 - 6.23) | 0.62 | 1.46 (0.39 - 5.50) | 0.59 |
| ODOD | 22,398 | 49,215 | 17.1 | 40.6 | 830 | 2,101 | 0.96 (0.86 - 1.08) | 0.51 | 1.00 (0.89 - 1.12) | 0.86 |
| MDCR | 6,671 | 12,831 | 6.1 | 11.1 | 251 | 514 | 0.94 (0.76 - 1.15) | 0.55 | 0.95 (0.77 - 1.17) | 0.64 |
| MDCD | 5,573 | 12,971 | 3.7 | 8.1 | 256 | 591 | 0.88 (0.71 - 1.08) | 0.23 | 0.85 (0.68 - 1.05) | 0.15 |
| Open Claims | 366,241 | 719,300 | 312.9 | 670.5 | 7,824 | 16,350 | 0.98 (0.94 - 1.02) | 0.25 | 0.97 (0.88 - 1.07) | 0.55 |
| OEHR | 31,061 | 71,577 | 15.1 | 38.2 | 744 | 2,051 | 0.95 (0.85 - 1.07) | 0.41 | 0.97 (0.86 - 1.09) | 0.61 |
| VA | 1,386 | 7,321 | 1.0 | 5.3 | 37 | 187 | 0.93 (0.56 - 1.51) | 0.78 | 0.84 (0.51 - 1.38) | 0.51 |
| SIDIAP | 18,349 | 5,245 | 50.2 | 13.6 | 621 | 121 | 0.94 (0.73 - 1.23) | 0.66 | 0.98 (0.75 - 1.28) | 0.82 |
| Meta-analysis | 495,460 | 937,402 | 442.6 | 830.0 | 11,463 | 23,382 | 0.97 (0.94 - 1.01) | 0.11 | 0.96 (0.89 - 1.04) | 0.35 |
| Meta-analysis (w/o USOC) | 129,219 | 218,102 | 129.7 | 159.5 | 3,639 | 7,032 | 0.96 (0.91 - 1.02) | 0.23 | 0.97 (0.89 - 1.04) | 0.47 |
| **PS-stratification / on-treatment TAR** | | | | | | | | | | |
| CCAE | 38,768 | 58,736 | 29.7 | 41.6 | 1,050 | 1,493 | 0.98 (0.90 - 1.07) | 0.64 | 0.97 (0.89 - 1.06) | 0.51 |
| DAG | 10,336 | 2,309 | 11.1 | 2.4 | 29 | 8 | 0.82 (0.36 - 2.11) | 0.67 | 0.81 (0.33 - 1.97) | 0.65 |
| ODOD | 24,049 | 49,356 | 18.4 | 40.7 | 897 | 2,104 | 0.94 (0.87 - 1.02) | 0.14 | 0.97 (0.89 - 1.06) | 0.58 |
| MDCR | 7,235 | 13,140 | 6.6 | 11.4 | 285 | 523 | 0.89 (0.76 - 1.03) | 0.13 | 0.90 (0.77 - 1.05) | 0.24 |
| MDCD | 5,785 | 13,053 | 3.9 | 8.1 | 273 | 591 | 0.88 (0.75 - 1.02) | 0.09 | 0.84 (0.72 - 0.98) | 0.06 |
| Open Claims | 374,458 | 719,384 | 321.8 | 670.5 | 8,141 | 16,352 | 0.98 (0.96 - 1.01) | 0.23 | 0.97 (0.88 - 1.06) | 0.50 |
| OEHR | 33,203 | 71,648 | 16.4 | 38.2 | 804 | 2,051 | 0.96 (0.88 - 1.05) | 0.38 | 0.97 (0.89 - 1.07) | 0.61 |
| VA | 1,662 | 10,429 | 1.2 | 7.9 | 47 | 296 | 1.11 (0.75 - 1.61) | 0.60 | 0.96 (0.66 - 1.42) | 0.82 |
| SIDIAP | 18,475 | 6,861 | 50.3 | 18.6 | 627 | 142 | 1.03 (0.84 - 1.27) | 0.80 | 1.00 (0.81 - 1.23) | 0.85 |
| Meta-analysis | 506,524 | 921,434 | 454.3 | 823.5 | 11,833 | 22,673 | 0.98 (0.95 - 1.00) | 0.04 | 0.96 (0.88 - 1.06) | 0.47 |
| Meta-analysis (w/o USOC) | 132,066 | 202,050 | 132.5 | 153.0 | 3,692 | 6,321 | 0.96 (0.91 - 1.00) | 0.05 | 0.96 (0.88 - 1.04) | 0.35 |
| **PS-matching / on-treatment TAR without escalation** | | | | | | | | | | |
| CCAE | 33,476 | 56,954 | 21.2 | 35.2 | 710 | 1,275 | 1.02 (0.90 - 1.16) | 0.77 | 1.00 (0.88 - 1.14) | 0.84 |
| DAG | 10,305 | 1,988 | 10.1 | 1.9 | 28 | 6 | 1.31 (0.40 - 5.93) | 0.69 | 1.41 (0.37 - 5.41) | 0.62 |
| ODOD | 22,398 | 49,215 | 14.5 | 36.6 | 700 | 1,906 | 0.91 (0.81 - 1.03) | 0.13 | 0.96 (0.85 - 1.08) | 0.55 |
| MDCR | 6,671 | 12,831 | 5.3 | 10.2 | 221 | 464 | 0.90 (0.72 - 1.12) | 0.35 | 0.90 (0.72 - 1.13) | 0.40 |
| MDCD | 5,573 | 12,971 | 3.0 | 6.8 | 211 | 500 | 0.90 (0.72 - 1.13) | 0.37 | 0.87 (0.69 - 1.09) | 0.26 |
| Open Claims | 366,241 | 719,300 | 250.6 | 568.4 | 6,265 | 13,841 | 0.98 (0.94 - 1.02) | 0.31 | 0.97 (0.89 - 1.06) | 0.57 |
| OEHR | 31,061 | 71,577 | 12.3 | 33.1 | 590 | 1,645 | 0.95 (0.84 - 1.08) | 0.43 | 0.96 (0.84 - 1.10) | 0.59 |
| VA | 1,386 | 7,321 | 0.8 | 4.7 | 31 | 160 | 1.00 (0.58 - 1.68) | 1.00 | 0.90 (0.53 - 1.54) | 0.71 |
| SIDIAP | 18,349 | 5,245 | 37.4 | 11.1 | 462 | 92 | 1.00 (0.74 - 1.37) | 0.99 | 1.07 (0.78 - 1.46) | 0.69 |
| Meta-analysis | 495,460 | 937,402 | 355.2 | 707.9 | 9,218 | 19,889 | 0.97 (0.94 - 1.00) | 0.09 | 0.97 (0.88 - 1.06) | 0.44 |
| Meta-analysis (w/o USOC) | 129,219 | 218,102 | 104.6 | 139.5 | 2,953 | 6,048 | 0.95 (0.89 - 1.01) | 0.12 | 0.96 (0.89 - 1.04) | 0.38 |
| **PS-stratification / on-treatment TAR without escalation** | | | | | | | | | | |
| CCAE | 38,768 | 58,736 | 24.8 | 36.2 | 865 | 1,306 | 0.96 (0.87 - 1.05) | 0.35 | 0.95 (0.86 - 1.05) | 0.35 |
| DAG | 10,336 | 2,309 | 10.1 | 2.3 | 28 | 7 | 0.93 (0.38 - 2.53) | 0.87 | 0.92 (0.36 - 2.38) | 0.86 |
| ODOD | 24,049 | 49,356 | 15.6 | 36.6 | 752 | 1,909 | 0.91 (0.84 - 1.00) | 0.05 | 0.96 (0.87 - 1.05) | 0.49 |
| MDCR | 7,235 | 13,140 | 5.8 | 10.5 | 252 | 472 | 0.90 (0.77 - 1.06) | 0.22 | 0.91 (0.77 - 1.08) | 0.34 |
| MDCD | 5,785 | 13,053 | 3.1 | 6.8 | 225 | 500 | 0.90 (0.76 - 1.06) | 0.21 | 0.86 (0.73 - 1.02) | 0.12 |
| Open Claims | 374,458 | 719,384 | 257.8 | 568.4 | 6,522 | 13,843 | 0.97 (0.95 - 1.00) | 0.10 | 0.96 (0.87 - 1.06) | 0.43 |
| OEHR | 33,203 | 71,648 | 13.3 | 33.1 | 641 | 1,645 | 1.00 (0.90 - 1.10) | 0.92 | 1.00 (0.90 - 1.11) | 0.82 |
| VA | 1,662 | 10,429 | 0.9 | 7.0 | 38 | 252 | 1.12 (0.73 - 1.70) | 0.59 | 0.99 (0.65 - 1.51) | 0.84 |
| SIDIAP | 18,475 | 6,861 | 37.5 | 15.1 | 466 | 112 | 1.02 (0.80 - 1.30) | 0.90 | 1.01 (0.79 - 1.28) | 0.85 |
| Meta-analysis | 506,524 | 921,434 | 365.0 | 702.1 | 9,526 | 19,294 | 0.97 (0.94 - 0.99) | 0.01 | 0.96 (0.89 - 1.05) | 0.39 |
| Meta-analysis (w/o USOC) | 132,066 | 202,050 | 107.1 | 133.7 | 3,004 | 5,451 | 0.95 (0.90 - 1.00) | 0.05 | 0.96 (0.89 - 1.04) | 0.39 |

##### 1.6.16 Genitourinary infection

**Supplementary Table 146:** Exposures, outcomes and risk estimates for Genitourinary infection in DPP4I (female) (T) vs SU (female) (C) new-user cohorts under four different propensity score (PS) adjustment / time-at-risk (TAR) designs across data sources. We report total patient exposure time in 1,000 patient-year (KYr) units, uncalibrated and calibrated hazard ratios (HRs) and their 95% confidence intervals (CIs). Greyed out entries did not pass study diagnostics and are included only for completeness.

| | Patients | | Exposure (KYr) | | Outcomes | | Uncalibrated | | Calibrated | |
|---|---|---|---|---|---|---|---|---|---|---|
| | T | C | T | C | T | C | HR (95% CI) | $p$ | HR (95% CI) | $p$ |
| **PS-matching / on-treatment TAR** | | | | | | | | | | |
| CCAE | 27,245 | 46,972 | 20.2 | 32.8 | 1,460 | 2,153 | 1.04 (0.95 - 1.15) | 0.39 | 1.01 (0.91 - 1.11) | 0.81 |
| DAG | 7,047 | 1,394 | 7.1 | 1.4 | 384 | 67 | 1.30 (0.93 - 1.85) | 0.14 | 1.36 (0.96 - 1.93) | 0.13 |
| ODOD | 18,104 | 40,283 | 13.3 | 31.9 | 1,331 | 3,262 | 0.99 (0.91 - 1.09) | 0.91 | 1.03 (0.93 - 1.13) | 0.64 |
| MDCR | 5,639 | 10,677 | 4.9 | 9.0 | 479 | 876 | 1.00 (0.84 - 1.17) | 0.97 | 1.01 (0.85 - 1.19) | 0.80 |
| MDCD | 4,784 | 10,812 | 3.2 | 6.6 | 328 | 700 | 1.01 (0.83 - 1.22) | 0.93 | 0.97 (0.80 - 1.19) | 0.78 |
| Open Claims | 289,230 | 578,011 | 243.1 | 534.8 | 12,925 | 27,146 | 1.00 (0.97 - 1.03) | 0.95 | 0.99 (0.91 - 1.09) | 0.85 |
| OEHR | 27,853 | 64,207 | 13.3 | 33.7 | 1,032 | 2,683 | 1.00 (0.91 - 1.10) | 0.95 | 1.02 (0.92 - 1.13) | 0.72 |
| VA | 1,178 | 6,123 | 0.8 | 4.4 | 45 | 202 | 1.17 (0.70 - 1.90) | 0.55 | 1.05 (0.64 - 1.74) | 0.82 |
| SIDIAP | 13,675 | 4,493 | 35.6 | 10.8 | 2,053 | 539 | 1.04 (0.91 - 1.18) | 0.61 | 1.08 (0.94 - 1.24) | 0.37 |
| Meta-analysis | 394,755 | 762,972 | 341.7 | 665.4 | 20,037 | 37,628 | 1.01 (0.98 - 1.03) | 0.70 | 0.99 (0.92 - 1.07) | 0.84 |
| Meta-analysis (w/o USOC) | 105,525 | 184,961 | 98.6 | 130.6 | 7,112 | 10,482 | 1.02 (0.97 - 1.07) | 0.41 | 1.02 (0.95 - 1.09) | 0.65 |
| **PS-stratification / on-treatment TAR** | | | | | | | | | | |
| CCAE | 31,471 | 48,414 | 23.4 | 33.7 | 1,751 | 2,226 | 1.10 (1.03 - 1.17) | <0.01 | 1.08 (1.01 - 1.16) | 0.05 |
| DAG | 7,079 | 1,619 | 7.2 | 1.6 | 386 | 82 | 1.19 (0.91 - 1.56) | 0.21 | 1.17 (0.89 - 1.54) | 0.33 |
| ODOD | 19,378 | 40,392 | 14.3 | 32.0 | 1,439 | 3,269 | 0.98 (0.92 - 1.05) | 0.64 | 1.01 (0.95 - 1.09) | 0.75 |
| MDCR | 6,012 | 11,044 | 5.3 | 9.3 | 505 | 900 | 0.99 (0.88 - 1.11) | 0.81 | 1.00 (0.89 - 1.13) | 0.83 |
| MDCD | 4,991 | 10,858 | 3.3 | 6.6 | 346 | 707 | 1.03 (0.90 - 1.18) | 0.66 | 0.99 (0.86 - 1.14) | 0.83 |
| Open Claims | 295,218 | 578,219 | 249.4 | 534.8 | 13,368 | 27,151 | 1.01 (0.99 - 1.03) | 0.42 | 0.99 (0.90 - 1.09) | 0.86 |
| OEHR | 29,709 | 64,264 | 14.4 | 33.7 | 1,118 | 2,686 | 1.02 (0.95 - 1.10) | 0.54 | 1.03 (0.95 - 1.12) | 0.49 |
| VA | 1,400 | 8,809 | 1.0 | 6.7 | 51 | 340 | 1.35 (0.92 - 1.96) | 0.12 | 1.18 (0.81 - 1.72) | 0.42 |
| SIDIAP | 13,757 | 5,991 | 35.7 | 15.0 | 2,058 | 737 | 1.04 (0.94 - 1.14) | 0.46 | 1.01 (0.91 - 1.11) | 0.74 |
| Meta-analysis | 402,624 | 749,943 | 349.6 | 660.1 | 20,625 | 37,051 | 1.02 (0.99 - 1.05) | 0.15 | 1.01 (0.92 - 1.11) | 0.84 |
| Meta-analysis (w/o USOC) | 107,406 | 171,724 | 100.2 | 125.3 | 7,257 | 9,900 | 1.03 (0.99 - 1.08) | 0.15 | 1.03 (0.95 - 1.12) | 0.45 |
| **PS-matching / on-treatment TAR without escalation** | | | | | | | | | | |
| CCAE | 27,245 | 46,972 | 16.7 | 28.6 | 1,222 | 1,852 | 1.07 (0.96 - 1.19) | 0.21 | 1.05 (0.94 - 1.17) | 0.44 |
| DAG | 7,047 | 1,394 | 6.5 | 1.3 | 352 | 60 | 1.26 (0.89 - 1.84) | 0.21 | 1.36 (0.94 - 1.97) | 0.16 |
| ODOD | 18,104 | 40,283 | 11.4 | 28.8 | 1,126 | 2,953 | 0.98 (0.89 - 1.07) | 0.62 | 1.02 (0.93 - 1.13) | 0.70 |
| MDCR | 5,639 | 10,677 | 4.3 | 8.3 | 407 | 787 | 0.96 (0.81 - 1.15) | 0.68 | 0.97 (0.81 - 1.16) | 0.73 |
| MDCD | 4,784 | 10,812 | 2.5 | 5.6 | 241 | 588 | 0.99 (0.80 - 1.22) | 0.91 | 0.95 (0.77 - 1.18) | 0.67 |
| Open Claims | 289,230 | 578,011 | 194.5 | 454.2 | 10,398 | 23,153 | 0.99 (0.96 - 1.03) | 0.66 | 0.99 (0.91 - 1.07) | 0.78 |
| OEHR | 27,853 | 64,207 | 10.9 | 29.2 | 804 | 2,197 | 1.02 (0.91 - 1.13) | 0.77 | 1.03 (0.92 - 1.15) | 0.67 |
| VA | 1,178 | 6,123 | 0.7 | 3.9 | 35 | 175 | 1.25 (0.72 - 2.11) | 0.41 | 1.13 (0.66 - 1.93) | 0.67 |
| SIDIAP | 13,675 | 4,493 | 27.0 | 8.9 | 1,502 | 441 | 0.99 (0.85 - 1.15) | 0.87 | 1.06 (0.90 - 1.24) | 0.55 |
| Meta-analysis | 394,755 | 762,972 | 274.6 | 568.8 | 16,087 | 32,206 | 1.00 (0.97 - 1.03) | 0.94 | 0.99 (0.91 - 1.09) | 0.86 |
| Meta-analysis (w/o USOC) | 105,525 | 184,961 | 80.0 | 114.6 | 5,689 | 9,053 | 1.01 (0.96 - 1.07) | 0.59 | 1.02 (0.96 - 1.09) | 0.57 |
| **PS-stratification / on-treatment TAR without escalation** | | | | | | | | | | |
| CCAE | 31,471 | 48,414 | 19.5 | 29.4 | 1,468 | 1,918 | 1.12 (1.04 - 1.20) | <0.01 | 1.11 (1.03 - 1.20) | 0.02 |
| DAG | 7,079 | 1,619 | 6.5 | 1.5 | 354 | 73 | 1.20 (0.91 - 1.61) | 0.20 | 1.20 (0.90 - 1.60) | 0.27 |
| ODOD | 19,378 | 40,392 | 12.2 | 28.9 | 1,220 | 2,959 | 0.97 (0.91 - 1.04) | 0.43 | 1.02 (0.94 - 1.10) | 0.74 |
| MDCR | 6,012 | 11,044 | 4.6 | 8.6 | 431 | 809 | 0.98 (0.87 - 1.11) | 0.79 | 0.99 (0.87 - 1.13) | 0.82 |
| MDCD | 4,991 | 10,858 | 2.6 | 5.6 | 255 | 595 | 0.95 (0.81 - 1.11) | 0.52 | 0.91 (0.78 - 1.07) | 0.31 |
| Open Claims | 295,218 | 578,219 | 199.6 | 454.3 | 10,757 | 23,157 | 1.00 (0.98 - 1.03) | 0.91 | 0.99 (0.89 - 1.09) | 0.78 |
| OEHR | 29,709 | 64,264 | 11.7 | 29.2 | 874 | 2,200 | 1.03 (0.94 - 1.12) | 0.52 | 1.03 (0.95 - 1.13) | 0.54 |
| VA | 1,400 | 8,809 | 0.8 | 5.9 | 40 | 297 | 1.42 (0.93 - 2.13) | 0.10 | 1.25 (0.82 - 1.89) | 0.32 |
| SIDIAP | 13,757 | 5,991 | 27.1 | 12.3 | 1,507 | 581 | 0.98 (0.88 - 1.10) | 0.77 | 0.98 (0.87 - 1.10) | 0.68 |
| Meta-analysis | 402,624 | 749,943 | 281.2 | 564.2 | 16,611 | 31,697 | 1.02 (0.98 - 1.06) | 0.37 | 1.02 (0.93 - 1.11) | 0.75 |
| Meta-analysis (w/o USOC) | 107,406 | 171,724 | 81.6 | 109.9 | 5,854 | 8,540 | 1.03 (0.97 - 1.09) | 0.36 | 1.04 (0.95 - 1.13) | 0.44 |

##### 1.6.17 Hyperkalemia

**Supplementary Table 147:** Exposures, outcomes and risk estimates for Hyperkalemia in DPP4I (female) (T) vs SU (female) (C) new-user cohorts under four different propensity score (PS) adjustment / time-at-risk (TAR) designs across data sources. We report total patient exposure time in 1,000 patient-year (KYr) units, uncalibrated and calibrated hazard ratios (HRs) and their 95% confidence intervals (CIs). Greyed out entries did not pass study diagnostics and are included only for completeness.

| | Patients | | Exposure (KYr) | | Outcomes | | Uncalibrated | | Calibrated | |
|---|---|---|---|---|---|---|---|---|---|---|
| | T | C | T | C | T | C | HR (95% CI) | $p$ | HR (95% CI) | $p$ |
| **PS-matching / on-treatment TAR** | | | | | | | | | | |
| CCAE | 38,521 | 65,087 | 30.0 | 47.2 | 172 | 368 | 0.85 (0.64 - 1.12) | 0.26 | 0.82 (0.62 - 1.09) | 0.18 |
| DAG | 9,775 | 1,951 | 10.4 | 2.0 | 176 | 22 | 1.74 (0.90 - 3.71) | 0.12 | 1.82 (0.90 - 3.70) | 0.11 |
| ODOD | 26,208 | 57,432 | 20.4 | 48.7 | 448 | 1,124 | 0.97 (0.83 - 1.14) | 0.72 | 1.01 (0.86 - 1.18) | 0.88 |
| MDCR | 7,925 | 14,952 | 7.4 | 13.2 | 111 | 203 | 0.89 (0.64 - 1.24) | 0.49 | 0.90 (0.64 - 1.26) | 0.54 |
| MDCD | 7,174 | 16,221 | 5.0 | 10.5 | 86 | 178 | 0.90 (0.61 - 1.31) | 0.57 | 0.87 (0.59 - 1.27) | 0.47 |
| Open Claims | 429,599 | 833,862 | 372.3 | 787.1 | 2,865 | 6,215 | 1.03 (0.96 - 1.10) | 0.42 | 1.02 (0.92 - 1.13) | 0.73 |
| OEHR | 35,700 | 81,633 | 18.1 | 45.8 | 242 | 642 | 1.18 (0.96 - 1.45) | 0.12 | 1.20 (0.97 - 1.47) | 0.10 |
| VA | 1,735 | 9,218 | 1.3 | 7.0 | 14 | 88 | 0.83 (0.35 - 1.78) | 0.66 | 0.75 (0.33 - 1.70) | 0.50 |
| SIDIAP | 18,570 | 5,270 | 50.4 | 13.6 | 453 | 128 | 0.83 (0.63 - 1.10) | 0.19 | 0.86 (0.65 - 1.15) | 0.34 |
| Meta-analysis | 575,207 | 1,085,626 | 515.3 | 975.3 | 4,567 | 8,968 | 1.01 (0.96 - 1.07) | 0.69 | 1.00 (0.91 - 1.09) | 0.90 |
| Meta-analysis (w/o USOC) | 145,608 | 251,764 | 143.0 | 188.1 | 1,702 | 2,753 | 0.97 (0.87 - 1.09) | 0.61 | 0.97 (0.86 - 1.10) | 0.68 |
| **PS-stratification / on-treatment TAR** | | | | | | | | | | |
| CCAE | 44,818 | 67,007 | 35.3 | 48.4 | 206 | 371 | 0.77 (0.64 - 0.92) | <0.01 | 0.76 (0.63 - 0.91) | <0.01 |
| DAG | 9,799 | 2,266 | 10.4 | 2.4 | 176 | 29 | 1.22 (0.79 - 1.93) | 0.39 | 1.20 (0.77 - 1.87) | 0.46 |
| ODOD | 28,155 | 57,590 | 22.0 | 48.8 | 480 | 1,125 | 1.01 (0.91 - 1.13) | 0.80 | 1.05 (0.93 - 1.17) | 0.51 |
| MDCR | 8,538 | 15,344 | 8.0 | 13.6 | 119 | 209 | 1.00 (0.79 - 1.27) | 0.98 | 1.02 (0.80 - 1.29) | 0.88 |
| MDCD | 7,515 | 16,325 | 5.3 | 10.5 | 94 | 179 | 1.12 (0.86 - 1.47) | 0.39 | 1.08 (0.82 - 1.42) | 0.59 |
| Open Claims | 439,411 | 833,960 | 383.1 | 787.1 | 2,989 | 6,215 | 1.00 (0.96 - 1.04) | 0.97 | 0.98 (0.89 - 1.09) | 0.73 |
| OEHR | 38,147 | 81,718 | 19.6 | 45.9 | 261 | 642 | 1.10 (0.95 - 1.28) | 0.21 | 1.12 (0.95 - 1.30) | 0.19 |
| VA | 2,107 | 12,693 | 1.5 | 10.0 | 15 | 139 | 0.98 (0.49 - 1.84) | 0.95 | 0.85 (0.44 - 1.64) | 0.64 |
| SIDIAP | 18,669 | 6,864 | 50.5 | 18.6 | 455 | 176 | 0.88 (0.72 - 1.08) | 0.23 | 0.85 (0.70 - 1.05) | 0.17 |
| Meta-analysis | 587,537 | 1,064,749 | 528.8 | 964.7 | 4,686 | 8,767 | 0.98 (0.90 - 1.07) | 0.60 | 0.97 (0.85 - 1.10) | 0.59 |
| Meta-analysis (w/o USOC) | 148,126 | 230,789 | 145.7 | 177.5 | 1,697 | 2,552 | 0.97 (0.86 - 1.09) | 0.59 | 0.97 (0.85 - 1.11) | 0.68 |
| **PS-matching / on-treatment TAR without escalation** | | | | | | | | | | |
| CCAE | 38,521 | 65,087 | 24.7 | 40.7 | 140 | 325 | 0.79 (0.58 - 1.06) | 0.12 | 0.77 (0.57 - 1.04) | 0.10 |
| DAG | 9,775 | 1,951 | 9.4 | 1.9 | 159 | 20 | 1.97 (0.99 - 4.38) | 0.07 | 2.12 (1.00 - 4.46) | 0.06 |
| ODOD | 26,208 | 57,432 | 17.1 | 43.5 | 367 | 981 | 0.94 (0.79 - 1.11) | 0.45 | 0.98 (0.83 - 1.17) | 0.84 |
| MDCR | 7,925 | 14,952 | 6.5 | 12.1 | 97 | 176 | 0.87 (0.61 - 1.22) | 0.43 | 0.88 (0.62 - 1.24) | 0.46 |
| MDCD | 7,174 | 16,221 | 3.9 | 8.7 | 65 | 155 | 0.82 (0.53 - 1.27) | 0.39 | 0.79 (0.51 - 1.23) | 0.31 |
| Open Claims | 429,599 | 833,862 | 295.7 | 662.8 | 2,271 | 5,177 | 1.01 (0.94 - 1.08) | 0.82 | 1.00 (0.91 - 1.11) | 0.90 |
| OEHR | 35,700 | 81,633 | 14.6 | 39.0 | 177 | 474 | 1.16 (0.91 - 1.47) | 0.22 | 1.18 (0.92 - 1.50) | 0.20 |
| VA | 1,735 | 9,218 | 1.0 | 6.1 | 12 | 73 | 1.05 (0.40 - 2.43) | 0.92 | 0.94 (0.38 - 2.33) | 0.88 |
| SIDIAP | 18,570 | 5,270 | 37.6 | 11.1 | 339 | 109 | 0.81 (0.59 - 1.13) | 0.21 | 0.87 (0.63 - 1.21) | 0.44 |
| Meta-analysis | 575,207 | 1,085,626 | 410.4 | 825.9 | 3,627 | 7,490 | 0.98 (0.92 - 1.05) | 0.60 | 0.98 (0.88 - 1.09) | 0.67 |
| Meta-analysis (w/o USOC) | 145,608 | 251,764 | 114.7 | 163.1 | 1,356 | 2,313 | 0.94 (0.82 - 1.08) | 0.39 | 0.95 (0.82 - 1.10) | 0.52 |
| **PS-stratification / on-treatment TAR without escalation** | | | | | | | | | | |
| CCAE | 44,818 | 67,007 | 29.0 | 41.7 | 167 | 328 | 0.75 (0.62 - 0.92) | <0.01 | 0.75 (0.61 - 0.91) | <0.01 |
| DAG | 9,799 | 2,266 | 9.4 | 2.2 | 159 | 26 | 1.32 (0.84 - 2.14) | 0.25 | 1.31 (0.82 - 2.10) | 0.28 |
| ODOD | 28,155 | 57,590 | 18.4 | 43.5 | 395 | 982 | 1.01 (0.89 - 1.14) | 0.91 | 1.05 (0.93 - 1.20) | 0.49 |
| MDCR | 8,538 | 15,344 | 7.0 | 12.5 | 104 | 181 | 1.04 (0.81 - 1.34) | 0.75 | 1.05 (0.81 - 1.36) | 0.73 |
| MDCD | 7,515 | 16,325 | 4.1 | 8.8 | 73 | 156 | 1.05 (0.78 - 1.41) | 0.75 | 1.01 (0.74 - 1.36) | 0.90 |
| Open Claims | 439,411 | 833,960 | 304.3 | 662.8 | 2,370 | 5,177 | 1.00 (0.95 - 1.05) | 0.95 | 0.99 (0.89 - 1.10) | 0.80 |
| OEHR | 38,147 | 81,718 | 15.8 | 39.1 | 193 | 474 | 1.12 (0.93 - 1.33) | 0.22 | 1.12 (0.94 - 1.35) | 0.23 |
| VA | 2,107 | 12,693 | 1.2 | 8.8 | 13 | 109 | 0.98 (0.45 - 1.98) | 0.96 | 0.86 (0.41 - 1.80) | 0.70 |
| SIDIAP | 18,669 | 6,864 | 37.7 | 15.0 | 341 | 147 | 0.80 (0.63 - 1.01) | 0.06 | 0.79 (0.63 - 1.00) | 0.08 |
| Meta-analysis | 587,537 | 1,064,749 | 421.7 | 816.9 | 3,729 | 7,315 | 0.97 (0.87 - 1.08) | 0.59 | 0.97 (0.84 - 1.11) | 0.62 |
| Meta-analysis (w/o USOC) | 148,126 | 230,789 | 117.3 | 154.0 | 1,359 | 2,138 | 0.96 (0.83 - 1.11) | 0.59 | 0.97 (0.83 - 1.14) | 0.73 |

##### 1.6.18 Hypoglycemia

**Supplementary Table 148:** Exposures, outcomes and risk estimates for Hypoglycemia in DPP4I (female) (T) vs SU (female) (C) new-user cohorts under four different propensity score (PS) adjustment / time-at-risk (TAR) designs across data sources. We report total patient exposure time in 1,000 patient-year (KYr) units, uncalibrated and calibrated hazard ratios (HRs) and their 95% confidence intervals (CIs). Greyed out entries did not pass study diagnostics and are included only for completeness.

| | Patients | | Exposure (KYr) | | Outcomes | | Uncalibrated | | Calibrated | |
|---|---|---|---|---|---|---|---|---|---|---|
| | T | C | T | C | T | C | HR (95% CI) | $p$ | HR (95% CI) | $p$ |
| **PS-matching / on-treatment TAR** | | | | | | | | | | |
| CCAE | 38,440 | 65,118 | 30.1 | 47.3 | 107 | 515 | 0.25 (0.18 - 0.34) | <0.01 | 0.24 (0.18 - 0.33) | <0.01 |
| DAG | 10,390 | 2,025 | 11.2 | 2.1 | 30 | 20 | 0.34 (0.14 - 0.81) | 0.01 | 0.35 (0.15 - 0.84) | 0.02 |
| ODOD | 27,017 | 59,247 | 21.4 | 50.9 | 165 | 1,320 | 0.26 (0.21 - 0.32) | <0.01 | 0.27 (0.22 - 0.34) | <0.01 |
| MDCR | 8,089 | 15,363 | 7.6 | 13.6 | 42 | 305 | 0.17 (0.10 - 0.27) | <0.01 | 0.17 (0.11 - 0.29) | <0.01 |
| MDCD | 7,197 | 16,312 | 5.1 | 10.6 | 60 | 340 | 0.35 (0.23 - 0.51) | <0.01 | 0.34 (0.23 - 0.51) | <0.01 |
| Open Claims | 431,606 | 837,656 | 376.5 | 794.4 | 1,208 | 8,598 | 0.23 (0.21 - 0.25) | <0.01 | 0.23 (0.20 - 0.26) | <0.01 |
| OEHR | 35,994 | 82,400 | 18.4 | 46.4 | 134 | 932 | 0.31 (0.24 - 0.39) | <0.01 | 0.31 (0.25 - 0.40) | <0.01 |
| VA | 1,785 | 9,513 | 1.3 | 7.3 | <10 | 92 | 0.26 (0.06 - 0.76) | 0.04 | 0.23 (0.07 - 0.83) | 0.02 |
| SIDIAP | 19,478 | 5,435 | 53.6 | 14.1 | 50 | 66 | 0.17 (0.10 - 0.29) | <0.01 | 0.18 (0.10 - 0.31) | <0.01 |
| Meta-analysis | 579,996 | 1,093,069 | 525.2 | 986.8 | <1,806 | 12,188 | 0.25 (0.22 - 0.29) | <0.01 | 0.25 (0.22 - 0.29) | <0.01 |
| Meta-analysis (w/o USOC) | 148,390 | 255,413 | 148.7 | 192.4 | <598 | 3,590 | 0.26 (0.22 - 0.31) | <0.01 | 0.26 (0.22 - 0.31) | <0.01 |
| **PS-stratification / on-treatment TAR** | | | | | | | | | | |
| CCAE | 44,599 | 67,013 | 35.3 | 48.5 | 120 | 533 | 0.31 (0.25 - 0.38) | <0.01 | 0.31 (0.25 - 0.38) | <0.01 |
| DAG | 10,421 | 2,349 | 11.3 | 2.5 | 30 | 24 | 0.31 (0.16 - 0.59) | <0.01 | 0.30 (0.16 - 0.57) | <0.01 |
| ODOD | 28,947 | 59,399 | 23.0 | 50.9 | 182 | 1,322 | 0.30 (0.26 - 0.35) | <0.01 | 0.31 (0.26 - 0.37) | <0.01 |
| MDCR | 8,701 | 15,683 | 8.2 | 13.9 | 45 | 312 | 0.24 (0.17 - 0.33) | <0.01 | 0.25 (0.18 - 0.34) | <0.01 |
| MDCD | 7,505 | 16,411 | 5.3 | 10.6 | 60 | 341 | 0.35 (0.26 - 0.46) | <0.01 | 0.34 (0.25 - 0.45) | <0.01 |
| Open Claims | 441,252 | 837,752 | 387.3 | 794.5 | 1,271 | 8,599 | 0.29 (0.27 - 0.31) | <0.01 | 0.28 (0.25 - 0.32) | <0.01 |
| OEHR | 38,401 | 82,487 | 19.8 | 46.4 | 139 | 934 | 0.36 (0.29 - 0.43) | <0.01 | 0.36 (0.30 - 0.43) | <0.01 |
| VA | 2,176 | 12,984 | 1.6 | 10.4 | <10 | 138 | 0.27 (0.09 - 0.65) | <0.01 | 0.24 (0.09 - 0.62) | <0.01 |
| SIDIAP | 19,590 | 7,064 | 53.7 | 19.3 | 50 | 84 | 0.19 (0.13 - 0.29) | <0.01 | 0.19 (0.13 - 0.28) | <0.01 |
| Meta-analysis | 591,911 | 1,071,747 | 538.5 | 976.0 | 1,837 | 11,808 | 0.30 (0.27 - 0.32) | <0.01 | 0.29 (0.26 - 0.33) | <0.01 |
| Meta-analysis (w/o USOC) | 150,659 | 233,995 | 151.2 | 181.6 | 566 | 3,209 | 0.29 (0.26 - 0.34) | <0.01 | 0.30 (0.25 - 0.35) | <0.01 |
| **PS-matching / on-treatment TAR without escalation** | | | | | | | | | | |
| CCAE | 38,440 | 65,118 | 24.7 | 40.8 | 60 | 430 | 0.21 (0.15 - 0.30) | <0.01 | 0.21 (0.15 - 0.30) | <0.01 |
| DAG | 10,390 | 2,025 | 10.1 | 2.0 | 26 | 20 | 0.34 (0.15 - 0.82) | 0.01 | 0.37 (0.15 - 0.87) | 0.03 |
| ODOD | 27,017 | 59,247 | 18.0 | 45.4 | 89 | 1,159 | 0.17 (0.13 - 0.22) | <0.01 | 0.18 (0.14 - 0.24) | <0.01 |
| MDCR | 8,089 | 15,363 | 6.6 | 12.5 | 26 | 276 | 0.13 (0.07 - 0.22) | <0.01 | 0.13 (0.07 - 0.24) | <0.01 |
| MDCD | 7,197 | 16,312 | 4.0 | 8.8 | 33 | 275 | 0.30 (0.18 - 0.47) | <0.01 | 0.29 (0.18 - 0.46) | <0.01 |
| Open Claims | 431,606 | 837,656 | 299.2 | 669.1 | 657 | 7,126 | 0.18 (0.16 - 0.20) | <0.01 | 0.18 (0.16 - 0.20) | <0.01 |
| OEHR | 35,994 | 82,400 | 14.8 | 39.5 | 63 | 686 | 0.20 (0.15 - 0.28) | <0.01 | 0.21 (0.15 - 0.29) | <0.01 |
| VA | 1,785 | 9,513 | 1.1 | 6.3 | <10 | 67 | 0.13 (0.01 - 0.64) | 0.07 | 0.12 (0.01 - 1.11) | 0.06 |
| SIDIAP | 19,478 | 5,435 | 39.9 | 11.5 | 14 | 51 | 0.06 (0.03 - 0.15) | <0.01 | 0.07 (0.03 - 0.17) | <0.01 |
| Meta-analysis | 579,996 | 1,093,069 | 418.4 | 835.9 | <978 | 10,090 | 0.18 (0.17 - 0.20) | <0.01 | 0.18 (0.16 - 0.21) | <0.01 |
| Meta-analysis (w/o USOC) | 148,390 | 255,413 | 119.2 | 166.8 | <321 | 2,964 | 0.19 (0.15 - 0.25) | <0.01 | 0.19 (0.15 - 0.25) | <0.01 |
| **PS-stratification / on-treatment TAR without escalation** | | | | | | | | | | |
| CCAE | 44,599 | 67,013 | 29.0 | 41.9 | 70 | 447 | 0.23 (0.17 - 0.29) | <0.01 | 0.22 (0.17 - 0.29) | <0.01 |
| DAG | 10,421 | 2,349 | 10.2 | 2.3 | 26 | 24 | 0.26 (0.14 - 0.52) | <0.01 | 0.26 (0.13 - 0.52) | <0.01 |
| ODOD | 28,947 | 59,399 | 19.3 | 45.4 | 101 | 1,161 | 0.20 (0.16 - 0.24) | <0.01 | 0.21 (0.17 - 0.25) | <0.01 |
| MDCR | 8,701 | 15,683 | 7.1 | 12.8 | 28 | 280 | 0.17 (0.11 - 0.25) | <0.01 | 0.17 (0.12 - 0.26) | <0.01 |
| MDCD | 7,505 | 16,411 | 4.2 | 8.8 | 33 | 276 | 0.25 (0.17 - 0.36) | <0.01 | 0.24 (0.17 - 0.35) | <0.01 |
| Open Claims | 441,252 | 837,752 | 307.9 | 669.2 | 691 | 7,127 | 0.19 (0.18 - 0.21) | <0.01 | 0.19 (0.17 - 0.22) | <0.01 |
| OEHR | 38,401 | 82,487 | 16.0 | 39.5 | 66 | 687 | 0.23 (0.18 - 0.29) | <0.01 | 0.23 (0.18 - 0.30) | <0.01 |
| VA | 2,176 | 12,984 | 1.3 | 9.1 | <10 | 104 | 0.28 (0.06 - 0.83) | 0.05 | 0.24 (0.07 - 0.88) | 0.03 |
| SIDIAP | 19,590 | 7,064 | 40.0 | 15.6 | 14 | 63 | 0.07 (0.04 - 0.14) | <0.01 | 0.07 (0.04 - 0.14) | <0.01 |
| Meta-analysis | 591,911 | 1,071,747 | 429.5 | 826.6 | 996 | 9,789 | 0.20 (0.18 - 0.21) | <0.01 | 0.20 (0.18 - 0.22) | <0.01 |
| Meta-analysis (w/o USOC) | 150,659 | 233,995 | 121.6 | 157.5 | 305 | 2,662 | 0.19 (0.16 - 0.24) | <0.01 | 0.20 (0.16 - 0.25) | <0.01 |

##### 1.6.19 Hypotension

**Supplementary Table 149:** Exposures, outcomes and risk estimates for Hypotension in DPP4I (female) (T) vs SU (female) (C) new-user cohorts under four different propensity score (PS) adjustment / time-at-risk (TAR) designs across data sources. We report total patient exposure time in 1,000 patient-year (KYr) units, uncalibrated and calibrated hazard ratios (HRs) and their 95% confidence intervals (CIs). Greyed out entries did not pass study diagnostics and are included only for completeness.

| | Patients | | Exposure (KYr) | | Outcomes | | Uncalibrated | | Calibrated | |
|---|---|---|---|---|---|---|---|---|---|---|
| | T | C | T | C | T | C | HR (95% CI) | $p$ | HR (95% CI) | $p$ |
| **PS-matching / on-treatment TAR** | | | | | | | | | | |
| CCAE | 38,381 | 64,846 | 30.0 | 47.2 | 211 | 343 | 1.10 (0.85 - 1.41) | 0.46 | 1.06 (0.82 - 1.37) | 0.66 |
| DAG | 10,116 | 1,972 | 10.9 | 2.0 | 69 | 6 | 1.89 (0.76 - 5.73) | 0.22 | 1.97 (0.72 - 5.43) | 0.20 |
| ODOD | 26,107 | 57,157 | 20.4 | 48.9 | 448 | 1,102 | 1.08 (0.92 - 1.25) | 0.34 | 1.12 (0.95 - 1.31) | 0.22 |
| MDCR | 7,752 | 14,741 | 7.3 | 13.0 | 159 | 297 | 0.83 (0.62 - 1.09) | 0.18 | 0.83 (0.63 - 1.11) | 0.22 |
| MDCD | 6,891 | 15,590 | 4.8 | 10.1 | 138 | 285 | 1.00 (0.72 - 1.37) | 0.99 | 0.96 (0.70 - 1.33) | 0.82 |
| Open Claims | 423,139 | 822,249 | 367.3 | 778.9 | 3,518 | 6,996 | 1.06 (1.00 - 1.12) | 0.05 | 1.05 (0.95 - 1.17) | 0.36 |
| OEHR | 35,607 | 81,377 | 18.0 | 45.6 | 262 | 715 | 0.93 (0.76 - 1.13) | 0.48 | 0.94 (0.77 - 1.15) | 0.59 |
| VA | 1,711 | 9,192 | 1.3 | 7.0 | 12 | 65 | 1.09 (0.41 - 2.63) | 0.85 | 0.99 (0.39 - 2.51) | 0.91 |
| SIDIAP | 19,382 | 5,423 | 53.3 | 14.2 | 95 | 18 | 0.97 (0.49 - 2.02) | 0.93 | 1.01 (0.50 - 2.05) | 0.93 |
| Meta-analysis | 569,086 | 1,072,547 | 513.3 | 967.1 | 4,912 | 9,827 | 1.05 (0.99 - 1.10) | 0.08 | 1.03 (0.94 - 1.13) | 0.48 |
| Meta-analysis (w/o USOC) | 145,947 | 250,298 | 146.0 | 188.2 | 1,394 | 2,831 | 1.01 (0.92 - 1.11) | 0.85 | 1.01 (0.91 - 1.12) | 0.85 |
| **PS-stratification / on-treatment TAR** | | | | | | | | | | |
| CCAE | 44,624 | 66,758 | 35.2 | 48.4 | 257 | 351 | 1.01 (0.85 - 1.20) | 0.87 | 1.00 (0.84 - 1.19) | 0.92 |
| DAG | 10,145 | 2,280 | 10.9 | 2.4 | 69 | 9 | 1.82 (0.89 - 4.12) | 0.13 | 1.79 (0.83 - 3.84) | 0.15 |
| ODOD | 28,041 | 57,316 | 22.0 | 49.0 | 486 | 1,103 | 1.05 (0.94 - 1.18) | 0.39 | 1.08 (0.96 - 1.22) | 0.23 |
| MDCR | 8,338 | 15,049 | 7.8 | 13.3 | 172 | 301 | 0.98 (0.80 - 1.19) | 0.82 | 0.99 (0.81 - 1.21) | 0.87 |
| MDCD | 7,199 | 15,691 | 5.0 | 10.1 | 153 | 287 | 1.09 (0.88 - 1.35) | 0.41 | 1.05 (0.85 - 1.30) | 0.67 |
| Open Claims | 432,724 | 822,422 | 377.9 | 778.9 | 3,664 | 6,996 | 1.06 (1.02 - 1.10) | <0.01 | 1.04 (0.94 - 1.15) | 0.42 |
| OEHR | 38,051 | 81,463 | 19.5 | 45.7 | 287 | 717 | 1.06 (0.91 - 1.22) | 0.45 | 1.07 (0.92 - 1.24) | 0.41 |
| VA | 2,102 | 12,553 | 1.5 | 10.0 | 15 | 100 | 1.22 (0.59 - 2.39) | 0.58 | 1.06 (0.53 - 2.13) | 0.87 |
| SIDIAP | 19,493 | 7,053 | 53.4 | 19.3 | 95 | 20 | 1.13 (0.68 - 1.96) | 0.65 | 1.09 (0.64 - 1.86) | 0.74 |
| Meta-analysis | 581,416 | 1,052,341 | 526.7 | 957.0 | 5,030 | 9,497 | 1.06 (1.02 - 1.09) | <0.01 | 1.04 (0.94 - 1.15) | 0.41 |
| Meta-analysis (w/o USOC) | 148,692 | 229,919 | 148.9 | 178.1 | 1,366 | 2,501 | 1.04 (0.97 - 1.12) | 0.26 | 1.04 (0.94 - 1.16) | 0.42 |
| **PS-matching / on-treatment TAR without escalation** | | | | | | | | | | |
| CCAE | 38,381 | 64,846 | 24.7 | 40.7 | 164 | 282 | 1.15 (0.88 - 1.51) | 0.30 | 1.13 (0.86 - 1.48) | 0.39 |
| DAG | 10,116 | 1,972 | 9.9 | 1.9 | 56 | 6 | 1.74 (0.68 - 5.35) | 0.29 | 1.87 (0.67 - 5.24) | 0.25 |
| ODOD | 26,107 | 57,157 | 17.2 | 43.6 | 383 | 971 | 1.10 (0.93 - 1.29) | 0.27 | 1.15 (0.97 - 1.36) | 0.13 |
| MDCR | 7,752 | 14,741 | 6.3 | 12.0 | 139 | 266 | 0.87 (0.64 - 1.16) | 0.33 | 0.87 (0.65 - 1.17) | 0.37 |
| MDCD | 6,891 | 15,590 | 3.8 | 8.4 | 91 | 224 | 0.87 (0.60 - 1.24) | 0.45 | 0.84 (0.58 - 1.21) | 0.35 |
| Open Claims | 423,139 | 822,249 | 291.9 | 655.8 | 2,798 | 5,815 | 1.06 (0.99 - 1.13) | 0.10 | 1.05 (0.95 - 1.16) | 0.37 |
| OEHR | 35,607 | 81,377 | 14.5 | 38.9 | 178 | 499 | 0.97 (0.77 - 1.23) | 0.81 | 0.98 (0.78 - 1.24) | 0.86 |
| VA | 1,711 | 9,192 | 1.0 | 6.1 | <10 | 57 | 0.99 (0.34 - 2.52) | 0.99 | 0.89 (0.33 - 2.43) | 0.83 |
| SIDIAP | 19,382 | 5,423 | 39.6 | 11.5 | 72 | 13 | 1.04 (0.50 - 2.34) | 0.92 | 1.12 (0.52 - 2.41) | 0.78 |
| Meta-analysis | 569,086 | 1,072,547 | 408.9 | 818.9 | <3,891 | 8,133 | 1.05 (0.99 - 1.11) | 0.09 | 1.04 (0.94 - 1.15) | 0.41 |
| Meta-analysis (w/o USOC) | 145,947 | 250,298 | 117.0 | 163.1 | <1,093 | 2,318 | 1.03 (0.93 - 1.14) | 0.58 | 1.04 (0.93 - 1.16) | 0.53 |
| **PS-stratification / on-treatment TAR without escalation** | | | | | | | | | | |
| CCAE | 44,624 | 66,758 | 29.0 | 41.7 | 202 | 289 | 1.02 (0.84 - 1.24) | 0.84 | 1.01 (0.83 - 1.23) | 0.90 |
| DAG | 10,145 | 2,280 | 9.9 | 2.2 | 56 | 9 | 1.56 (0.75 - 3.59) | 0.27 | 1.55 (0.71 - 3.41) | 0.28 |
| ODOD | 28,041 | 57,316 | 18.5 | 43.6 | 410 | 972 | 1.06 (0.94 - 1.20) | 0.34 | 1.11 (0.98 - 1.26) | 0.16 |
| MDCR | 8,338 | 15,049 | 6.8 | 12.2 | 151 | 269 | 0.99 (0.80 - 1.22) | 0.91 | 1.00 (0.80 - 1.23) | 0.88 |
| MDCD | 7,199 | 15,691 | 4.0 | 8.4 | 104 | 226 | 1.02 (0.79 - 1.30) | 0.90 | 0.97 (0.76 - 1.25) | 0.83 |
| Open Claims | 432,724 | 822,422 | 300.4 | 655.8 | 2,917 | 5,815 | 1.07 (1.02 - 1.12) | <0.01 | 1.05 (0.95 - 1.17) | 0.36 |
| OEHR | 38,051 | 81,463 | 15.8 | 38.9 | 200 | 501 | 1.09 (0.91 - 1.29) | 0.36 | 1.09 (0.91 - 1.30) | 0.36 |
| VA | 2,102 | 12,553 | 1.2 | 8.7 | 12 | 85 | 1.22 (0.55 - 2.56) | 0.61 | 1.08 (0.50 - 2.32) | 0.85 |
| SIDIAP | 19,493 | 7,053 | 39.7 | 15.7 | 72 | 13 | 1.32 (0.72 - 2.62) | 0.40 | 1.31 (0.68 - 2.51) | 0.43 |
| Meta-analysis | 581,416 | 1,052,341 | 420.0 | 810.2 | 4,008 | 7,868 | 1.07 (1.02 - 1.11) | <0.01 | 1.06 (0.97 - 1.16) | 0.20 |
| Meta-analysis (w/o USOC) | 148,692 | 229,919 | 119.7 | 154.4 | 1,091 | 2,053 | 1.06 (0.97 - 1.14) | 0.19 | 1.07 (0.96 - 1.18) | 0.25 |

##### 1.6.20 Joint pain

**Supplementary Table 150:** Exposures, outcomes and risk estimates for Joint pain in DPP4I (female) (T) vs SU (female) (C) new-user cohorts under four different propensity score (PS) adjustment / time-at-risk (TAR) designs across data sources. We report total patient exposure time in 1,000 patient-year (KYr) units, uncalibrated and calibrated hazard ratios (HRs) and their 95% confidence intervals (CIs). Greyed out entries did not pass study diagnostics and are included only for completeness.

| | Patients | | Exposure (KYr) | | Outcomes | | Uncalibrated | | Calibrated | |
|---|---|---|---|---|---|---|---|---|---|---|
| | T | C | T | C | T | C | HR (95% CI) | $p$ | HR (95% CI) | $p$ |
| **PS-matching / on-treatment TAR** | | | | | | | | | | |
| CCAE | 36,500 | 61,985 | 28.4 | 44.9 | 396 | 596 | 0.93 (0.77 - 1.12) | 0.45 | 0.90 (0.74 - 1.09) | 0.29 |
| DAG | 8,831 | 1,761 | 9.3 | 1.8 | 212 | 50 | 1.04 (0.69 - 1.60) | 0.87 | 1.08 (0.71 - 1.66) | 0.69 |
| ODOD | 25,431 | 56,095 | 19.9 | 48.3 | 385 | 814 | 1.06 (0.89 - 1.26) | 0.48 | 1.10 (0.92 - 1.31) | 0.33 |
| MDCR | 7,747 | 14,779 | 7.2 | 13.1 | 91 | 134 | 1.44 (0.97 - 2.13) | 0.07 | 1.45 (0.98 - 2.16) | 0.07 |
| MDCD | 6,596 | 15,176 | 4.5 | 9.8 | 143 | 230 | 1.16 (0.85 - 1.58) | 0.34 | 1.12 (0.82 - 1.53) | 0.49 |
| Open Claims | 406,341 | 794,621 | 352.7 | 754.5 | 3,374 | 6,218 | 1.08 (1.02 - 1.15) | 0.01 | 1.08 (0.97 - 1.19) | 0.19 |
| OEHR | 34,136 | 78,457 | 17.2 | 43.8 | 279 | 742 | 0.95 (0.79 - 1.16) | 0.64 | 0.97 (0.80 - 1.18) | 0.76 |
| VA | 1,424 | 7,508 | 1.1 | 5.7 | 10 | 79 | 0.89 (0.38 - 1.89) | 0.78 | 0.80 (0.36 - 1.79) | 0.60 |
| SIDIAP | 15,176 | 4,670 | 40.4 | 11.6 | 1,166 | 344 | 0.93 (0.79 - 1.10) | 0.39 | 0.97 (0.82 - 1.15) | 0.71 |
| Meta-analysis | 542,182 | 1,035,052 | 480.8 | 933.5 | 6,056 | 9,207 | 1.04 (0.97 - 1.11) | 0.29 | 1.02 (0.93 - 1.13) | 0.63 |
| Meta-analysis (w/o USOC) | 135,841 | 240,431 | 128.1 | 179.0 | 2,682 | 2,989 | 1.00 (0.92 - 1.09) | 1.00 | 1.00 (0.91 - 1.10) | 0.88 |
| **PS-stratification / on-treatment TAR** | | | | | | | | | | |
| CCAE | 42,281 | 63,796 | 33.2 | 46.0 | 487 | 601 | 1.00 (0.88 - 1.14) | 1.00 | 0.99 (0.87 - 1.13) | 0.85 |
| DAG | 8,855 | 2,055 | 9.3 | 2.1 | 213 | 61 | 0.89 (0.65 - 1.24) | 0.49 | 0.88 (0.63 - 1.22) | 0.48 |
| ODOD | 27,243 | 56,249 | 21.4 | 48.3 | 412 | 815 | 1.06 (0.94 - 1.20) | 0.35 | 1.09 (0.96 - 1.24) | 0.22 |
| MDCR | 8,335 | 15,094 | 7.8 | 13.4 | 100 | 135 | 1.27 (0.96 - 1.67) | 0.09 | 1.29 (0.98 - 1.70) | 0.09 |
| MDCD | 6,865 | 15,272 | 4.7 | 9.8 | 148 | 230 | 1.19 (0.96 - 1.49) | 0.11 | 1.15 (0.92 - 1.43) | 0.25 |
| Open Claims | 415,068 | 794,718 | 362.4 | 754.6 | 3,534 | 6,218 | 1.10 (1.05 - 1.14) | <0.01 | 1.08 (0.98 - 1.19) | 0.14 |
| OEHR | 36,357 | 78,542 | 18.5 | 43.8 | 310 | 742 | 0.95 (0.82 - 1.09) | 0.45 | 0.96 (0.83 - 1.11) | 0.58 |
| VA | 1,694 | 10,443 | 1.3 | 8.3 | 10 | 140 | 0.88 (0.40 - 1.79) | 0.74 | 0.77 (0.36 - 1.63) | 0.50 |
| SIDIAP | 15,301 | 6,185 | 40.5 | 16.0 | 1,167 | 465 | 1.01 (0.89 - 1.14) | 0.90 | 0.98 (0.86 - 1.11) | 0.72 |
| Meta-analysis | 553,440 | 1,016,639 | 493.2 | 924.3 | 6,223 | 9,037 | 1.05 (0.99 - 1.11) | 0.12 | 1.03 (0.93 - 1.15) | 0.56 |
| Meta-analysis (w/o USOC) | 138,372 | 221,921 | 130.8 | 169.7 | 2,689 | 2,819 | 1.01 (0.95 - 1.08) | 0.66 | 1.02 (0.93 - 1.12) | 0.75 |
| **PS-matching / on-treatment TAR without escalation** | | | | | | | | | | |
| CCAE | 36,500 | 61,985 | 23.4 | 38.7 | 328 | 512 | 0.92 (0.75 - 1.13) | 0.43 | 0.90 (0.73 - 1.11) | 0.35 |
| DAG | 8,831 | 1,761 | 8.4 | 1.7 | 198 | 47 | 0.99 (0.64 - 1.56) | 0.95 | 1.06 (0.67 - 1.67) | 0.72 |
| ODOD | 25,431 | 56,095 | 16.8 | 43.0 | 322 | 734 | 1.04 (0.86 - 1.25) | 0.69 | 1.09 (0.90 - 1.31) | 0.41 |
| MDCR | 7,747 | 14,779 | 6.3 | 12.0 | 79 | 122 | 1.51 (1.00 - 2.28) | 0.05 | 1.52 (1.00 - 2.29) | 0.05 |
| MDCD | 6,596 | 15,176 | 3.5 | 8.2 | 110 | 193 | 1.16 (0.81 - 1.64) | 0.41 | 1.11 (0.78 - 1.58) | 0.56 |
| Open Claims | 406,341 | 794,621 | 280.4 | 636.0 | 2,679 | 5,207 | 1.07 (1.00 - 1.14) | 0.05 | 1.06 (0.96 - 1.18) | 0.27 |
| OEHR | 34,136 | 78,457 | 13.8 | 37.3 | 220 | 602 | 1.03 (0.83 - 1.27) | 0.78 | 1.04 (0.84 - 1.29) | 0.72 |
| VA | 1,424 | 7,508 | 0.9 | 4.9 | <10 | 67 | 0.58 (0.19 - 1.50) | 0.31 | 0.52 (0.19 - 1.48) | 0.23 |
| SIDIAP | 15,176 | 4,670 | 30.1 | 9.6 | 859 | 278 | 1.01 (0.84 - 1.22) | 0.92 | 1.08 (0.90 - 1.31) | 0.47 |
| Meta-analysis | 542,182 | 1,035,052 | 383.7 | 791.5 | <4,805 | 7,762 | 1.05 (1.00 - 1.11) | 0.06 | 1.05 (0.95 - 1.16) | 0.36 |
| Meta-analysis (w/o USOC) | 135,841 | 240,431 | 103.3 | 155.5 | <2,126 | 2,555 | 1.03 (0.94 - 1.12) | 0.59 | 1.04 (0.94 - 1.14) | 0.54 |
| **PS-stratification / on-treatment TAR without escalation** | | | | | | | | | | |
| CCAE | 42,281 | 63,796 | 27.4 | 39.7 | 402 | 517 | 1.00 (0.87 - 1.15) | 0.98 | 0.99 (0.86 - 1.15) | 0.89 |
| DAG | 8,855 | 2,055 | 8.4 | 2.0 | 199 | 58 | 0.86 (0.62 - 1.22) | 0.39 | 0.86 (0.62 - 1.21) | 0.45 |
| ODOD | 27,243 | 56,249 | 18.0 | 43.1 | 345 | 734 | 1.05 (0.91 - 1.20) | 0.52 | 1.09 (0.95 - 1.26) | 0.27 |
| MDCR | 8,335 | 15,094 | 6.8 | 12.3 | 87 | 123 | 1.33 (0.99 - 1.77) | 0.06 | 1.34 (1.00 - 1.79) | 0.07 |
| MDCD | 6,865 | 15,272 | 3.7 | 8.2 | 115 | 193 | 1.16 (0.90 - 1.48) | 0.24 | 1.11 (0.87 - 1.42) | 0.45 |
| Open Claims | 415,068 | 794,718 | 288.2 | 636.0 | 2,807 | 5,207 | 1.09 (1.04 - 1.14) | <0.01 | 1.07 (0.96 - 1.19) | 0.20 |
| OEHR | 36,357 | 78,542 | 14.9 | 37.4 | 241 | 602 | 0.97 (0.83 - 1.14) | 0.73 | 0.98 (0.83 - 1.15) | 0.80 |
| VA | 1,694 | 10,443 | 1.0 | 7.3 | <10 | 121 | 0.55 (0.20 - 1.29) | 0.21 | 0.48 (0.19 - 1.23) | 0.13 |
| SIDIAP | 15,301 | 6,185 | 30.2 | 13.1 | 860 | 376 | 1.02 (0.89 - 1.18) | 0.78 | 1.01 (0.87 - 1.17) | 0.78 |
| Meta-analysis | 553,440 | 1,016,639 | 394.0 | 783.5 | 4,941 | 7,617 | 1.06 (1.01 - 1.11) | 0.03 | 1.05 (0.96 - 1.16) | 0.29 |
| Meta-analysis (w/o USOC) | 138,372 | 221,921 | 105.8 | 147.5 | 2,134 | 2,410 | 1.02 (0.95 - 1.09) | 0.56 | 1.03 (0.94 - 1.13) | 0.55 |

##### 1.6.21 Lower extremity amputation

**Supplementary Table 151:** Exposures, outcomes and risk estimates for Lower extremity amputation in DPP4I (female) (T) vs SU (female) (C) new-user cohorts under four different propensity score (PS) adjustment / time-at-risk (TAR) designs across data sources. We report total patient exposure time in 1,000 patient-year (KYr) units, uncalibrated and calibrated hazard ratios (HRs) and their 95% confidence intervals (CIs). Greyed out entries did not pass study diagnostics and are included only for completeness.

| | Patients | | Exposure (KYr) | | Outcomes | | Uncalibrated | | Calibrated | |
|---|---|---|---|---|---|---|---|---|---|---|
| | T | C | T | C | T | C | HR (95% CI) | $p$ | HR (95% CI) | $p$ |
| **PS-matching / on-treatment TAR** | | | | | | | | | | |
| CCAE | 39,120 | 66,027 | 30.7 | 48.2 | 9 | 15 | 2.09 (0.76 - 5.80) | 0.15 | 2.02 (0.73 - 5.56) | 0.18 |
| DAG | 10,485 | 2,040 | 11.4 | 2.1 | 0 | 0 | | | | |
| ODOD | 27,477 | 60,143 | 21.8 | 52.3 | 10 | 39 | 0.89 (0.31 - 2.30) | 0.82 | 0.92 (0.34 - 2.52) | 0.87 |
| MDCR | 8,154 | 15,485 | 7.7 | 13.8 | 6 | 9 | 0.98 (0.19 - 4.25) | 0.98 | 0.99 (0.21 - 4.65) | 0.98 |
| MDCD | 7,409 | 16,719 | 5.2 | 11.0 | <5 | 8 | 0.77 (0.03 - 9.19) | 0.85 | 0.74 (0.04 - 12.2) | 0.83 |
| Open Claims | 439,414 | 851,282 | 383.8 | 810.7 | 106 | 331 | 0.76 (0.55 - 1.05) | 0.10 | 0.76 (0.54 - 1.06) | 0.11 |
| OEHR | 36,406 | 83,227 | 18.7 | 47.4 | 7 | 28 | 1.28 (0.46 - 3.30) | 0.63 | 1.30 (0.48 - 3.48) | 0.61 |
| VA | 1,820 | 9,660 | 1.4 | 7.5 | 0 | <10 | | | | |
| SIDIAP | 19,504 | 5,435 | 53.7 | 14.2 | 8 | <5 | 1.34 (0.27 - 9.79) | 0.75 | 1.40 (0.23 - 8.41) | 0.72 |
| Meta-analysis | 542,417 | 1,060,679 | 455.0 | 958.5 | 132 | 413 | 1.00 (0.63 - 1.57) | 0.98 | 0.98 (0.62 - 1.56) | 0.94 |
| Meta-analysis (w/o USOC) | 103,003 | 209,397 | 71.1 | 147.9 | 26 | 82 | 1.33 (0.75 - 2.38) | 0.33 | 1.33 (0.75 - 2.38) | 0.33 |
| **PS-stratification / on-treatment TAR** | | | | | | | | | | |
| CCAE | 45,481 | 67,959 | 36.0 | 49.4 | 10 | 15 | 1.34 (0.54 - 3.23) | 0.52 | 1.33 (0.54 - 3.25) | 0.54 |
| DAG | 10,516 | 2,367 | 11.4 | 2.5 | 0 | 0 | | | | |
| ODOD | 29,479 | 60,301 | 23.5 | 52.3 | 10 | 39 | 0.73 (0.34 - 1.45) | 0.40 | 0.75 (0.36 - 1.57) | 0.45 |
| MDCR | 8,789 | 15,805 | 8.3 | 14.1 | 6 | 9 | 1.19 (0.38 - 3.51) | 0.76 | 1.21 (0.40 - 3.67) | 0.74 |
| MDCD | 7,756 | 16,825 | 5.5 | 11.0 | 5 | 8 | 1.46 (0.40 - 5.07) | 0.56 | 1.40 (0.39 - 5.00) | 0.60 |
| Open Claims | 449,397 | 851,380 | 395.0 | 810.7 | 110 | 331 | 0.78 (0.62 - 0.96) | 0.02 | 0.76 (0.60 - 0.97) | 0.03 |
| OEHR | 38,885 | 83,316 | 20.2 | 47.4 | 7 | 28 | 0.77 (0.30 - 1.76) | 0.57 | 0.78 (0.32 - 1.89) | 0.59 |
| VA | 2,218 | 13,208 | 1.6 | 10.6 | 0 | <10 | | | | |
| SIDIAP | 19,617 | 7,063 | 53.8 | 19.4 | 8 | <5 | 1.56 (0.38 - 7.89) | 0.57 | 1.51 (0.33 - 6.88) | 0.60 |
| Meta-analysis | 563,242 | 1,062,956 | 474.7 | 959.8 | 137 | 413 | 0.79 (0.65 - 0.97) | 0.02 | 0.78 (0.63 - 0.98) | 0.03 |
| Meta-analysis (w/o USOC) | 113,845 | 211,576 | 79.7 | 149.1 | 27 | 82 | 0.88 (0.55 - 1.42) | 0.61 | 0.88 (0.55 - 1.43) | 0.62 |
| **PS-matching / on-treatment TAR without escalation** | | | | | | | | | | |
| CCAE | 39,120 | 66,027 | 25.2 | 41.5 | 8 | 13 | 2.32 (0.79 - 6.87) | 0.13 | 2.28 (0.77 - 6.70) | 0.14 |
| DAG | 10,485 | 2,040 | 10.3 | 2.0 | 0 | 0 | | | | |
| ODOD | 27,477 | 60,143 | 18.3 | 46.5 | 8 | 32 | 0.83 (0.26 - 2.29) | 0.74 | 0.87 (0.29 - 2.60) | 0.81 |
| MDCR | 8,154 | 15,485 | 6.7 | 12.7 | 6 | 6 | 1.21 (0.23 - 5.94) | 0.82 | 1.22 (0.24 - 6.25) | 0.81 |
| MDCD | 7,409 | 16,719 | 4.1 | 9.1 | <5 | 5 | 0.77 (0.03 - 9.19) | 0.85 | 0.74 (0.04 - 12.2) | 0.83 |
| Open Claims | 439,414 | 851,282 | 304.7 | 681.8 | 84 | 252 | 0.73 (0.51 - 1.04) | 0.09 | 0.73 (0.50 - 1.05) | 0.09 |
| OEHR | 36,406 | 83,227 | 15.0 | 40.2 | <5 | 16 | 0.97 (0.25 - 3.31) | 0.96 | 0.98 (0.27 - 3.60) | 0.97 |
| VA | 1,820 | 9,660 | 1.1 | 6.5 | 0 | <10 | | | | |
| SIDIAP | 19,504 | 5,435 | 39.9 | 11.6 | 6 | <5 | 1.96 (0.27 - 40.2) | 0.60 | 2.11 (0.17 - 25.9) | 0.56 |
| Meta-analysis | 542,417 | 1,060,679 | 363.1 | 810.0 | <105 | 313 | 0.94 (0.57 - 1.56) | 0.81 | 0.94 (0.56 - 1.56) | 0.80 |
| Meta-analysis (w/o USOC) | 103,003 | 209,397 | 58.5 | 128.2 | <21 | 61 | 1.27 (0.66 - 2.46) | 0.47 | 1.28 (0.66 - 2.49) | 0.46 |
| **PS-stratification / on-treatment TAR without escalation** | | | | | | | | | | |
| CCAE | 45,481 | 67,959 | 29.6 | 42.6 | 9 | 13 | 1.45 (0.55 - 3.65) | 0.44 | 1.44 (0.56 - 3.69) | 0.45 |
| DAG | 10,516 | 2,367 | 10.3 | 2.3 | 0 | 0 | | | | |
| ODOD | 29,479 | 60,301 | 19.7 | 46.5 | 8 | 32 | 0.78 (0.32 - 1.67) | 0.55 | 0.81 (0.36 - 1.85) | 0.62 |
| MDCR | 8,789 | 15,805 | 7.2 | 12.9 | 6 | 6 | 1.90 (0.55 - 6.74) | 0.31 | 1.92 (0.55 - 6.72) | 0.31 |
| MDCD | 7,756 | 16,825 | 4.3 | 9.1 | <5 | 5 | 1.68 (0.30 - 8.68) | 0.55 | 1.60 (0.30 - 8.67) | 0.58 |
| Open Claims | 449,397 | 851,380 | 313.6 | 681.8 | 87 | 252 | 0.84 (0.65 - 1.07) | 0.17 | 0.83 (0.63 - 1.08) | 0.16 |
| OEHR | 38,885 | 83,316 | 16.3 | 40.2 | <5 | 16 | 0.68 (0.19 - 1.98) | 0.53 | 0.69 (0.21 - 2.23) | 0.53 |
| VA | 2,218 | 13,208 | 1.3 | 9.3 | 0 | <10 | | | | |
| SIDIAP | 19,617 | 7,063 | 40.0 | 15.7 | 6 | <5 | 1.52 (0.28 - 11.4) | 0.66 | 1.51 (0.24 - 9.55) | 0.66 |
| Meta-analysis | 563,242 | 1,062,956 | 379.1 | 811.1 | <109 | 313 | 0.85 (0.68 - 1.07) | 0.18 | 0.85 (0.67 - 1.08) | 0.19 |
| Meta-analysis (w/o USOC) | 113,845 | 211,576 | 65.5 | 129.3 | <22 | 61 | 0.93 (0.54 - 1.61) | 0.80 | 0.94 (0.54 - 1.64) | 0.83 |

##### 1.6.22 Nausea

**Supplementary Table 152:** Exposures, outcomes and risk estimates for Nausea in DPP4I (female) (T) vs SU (female) (C) new-user cohorts under four different propensity score (PS) adjustment / time-at-risk (TAR) designs across data sources. We report total patient exposure time in 1,000 patient-year (KYr) units, uncalibrated and calibrated hazard ratios (HRs) and their 95% confidence intervals (CIs). Greyed out entries did not pass study diagnostics and are included only for completeness.

| | Patients | | Exposure (KYr) | | Outcomes | | Uncalibrated | | Calibrated | |
|---|---|---|---|---|---|---|---|---|---|---|
| | T | C | T | C | T | C | HR (95% CI) | $p$ | HR (95% CI) | $p$ |
| **PS-matching / on-treatment TAR** | | | | | | | | | | |
| CCAE | 33,169 | 56,168 | 25.6 | 40.6 | 1,088 | 1,724 | 1.02 (0.91 - 1.14) | 0.72 | 0.98 (0.88 - 1.10) | 0.78 |
| DAG | 9,491 | 1,839 | 10.2 | 1.9 | 150 | 24 | 1.55 (0.89 - 2.83) | 0.14 | 1.62 (0.91 - 2.89) | 0.13 |
| ODOD | 22,223 | 48,605 | 17.1 | 41.0 | 924 | 2,379 | 0.91 (0.82 - 1.01) | 0.08 | 0.94 (0.84 - 1.05) | 0.41 |
| MDCR | 7,015 | 13,103 | 6.5 | 11.4 | 289 | 541 | 0.94 (0.77 - 1.14) | 0.54 | 0.95 (0.78 - 1.16) | 0.63 |
| MDCD | 4,961 | 11,206 | 3.3 | 7.1 | 387 | 724 | 1.02 (0.85 - 1.21) | 0.84 | 0.98 (0.82 - 1.18) | 0.79 |
| Open Claims | 358,720 | 698,954 | 310.6 | 663.8 | 9,048 | 19,011 | 1.01 (0.98 - 1.05) | 0.56 | 1.00 (0.92 - 1.10) | 0.89 |
| OEHR | 31,233 | 71,686 | 15.4 | 38.8 | 774 | 2,205 | 0.88 (0.79 - 0.98) | 0.03 | 0.89 (0.80 - 1.00) | 0.08 |
| VA | 1,492 | 7,880 | 1.1 | 6.0 | 29 | 141 | 1.17 (0.63 - 2.09) | 0.61 | 1.05 (0.58 - 1.92) | 0.84 |
| SIDIAP | 18,524 | 5,273 | 50.9 | 13.7 | 448 | 116 | 0.95 (0.71 - 1.27) | 0.71 | 0.99 (0.74 - 1.32) | 0.84 |
| Meta-analysis | 486,828 | 914,714 | 440.7 | 824.3 | 13,137 | 26,865 | 0.97 (0.92 - 1.03) | 0.33 | 0.96 (0.88 - 1.05) | 0.41 |
| Meta-analysis (w/o USOC) | 128,108 | 215,760 | 130.1 | 160.5 | 4,089 | 7,854 | 0.95 (0.90 - 1.01) | 0.11 | 0.95 (0.88 - 1.03) | 0.33 |
| **PS-stratification / on-treatment TAR** | | | | | | | | | | |
| CCAE | 38,599 | 57,863 | 30.0 | 41.6 | 1,285 | 1,755 | 1.01 (0.94 - 1.09) | 0.73 | 1.00 (0.92 - 1.09) | 0.84 |
| DAG | 9,527 | 2,130 | 10.2 | 2.3 | 150 | 30 | 1.44 (0.94 - 2.27) | 0.10 | 1.42 (0.91 - 2.21) | 0.14 |
| ODOD | 23,886 | 48,728 | 18.5 | 41.0 | 1,003 | 2,384 | 0.91 (0.85 - 0.99) | 0.03 | 0.94 (0.87 - 1.02) | 0.27 |
| MDCR | 7,552 | 13,481 | 7.0 | 11.8 | 317 | 552 | 0.91 (0.79 - 1.06) | 0.22 | 0.93 (0.80 - 1.07) | 0.38 |
| MDCD | 5,142 | 11,261 | 3.4 | 7.2 | 404 | 727 | 1.16 (1.02 - 1.32) | 0.03 | 1.11 (0.97 - 1.27) | 0.17 |
| Open Claims | 367,392 | 699,041 | 320.0 | 663.8 | 9,435 | 19,015 | 1.00 (0.98 - 1.03) | 0.79 | 0.99 (0.90 - 1.08) | 0.78 |
| OEHR | 33,348 | 71,751 | 16.7 | 38.8 | 842 | 2,206 | 0.92 (0.84 - 1.00) | 0.04 | 0.93 (0.85 - 1.01) | 0.15 |
| VA | 1,778 | 11,035 | 1.3 | 8.7 | 33 | 223 | 1.08 (0.67 - 1.68) | 0.75 | 0.94 (0.59 - 1.48) | 0.78 |
| SIDIAP | 18,621 | 6,878 | 51.0 | 18.7 | 449 | 152 | 0.85 (0.69 - 1.06) | 0.15 | 0.83 (0.67 - 1.03) | 0.11 |
| Meta-analysis | 498,925 | 899,872 | 453.3 | 818.1 | 13,481 | 26,094 | 0.96 (0.92 - 1.01) | 0.13 | 0.95 (0.86 - 1.06) | 0.35 |
| Meta-analysis (w/o USOC) | 131,533 | 200,831 | 133.3 | 154.2 | 4,046 | 7,079 | 0.94 (0.89 - 1.01) | 0.08 | 0.95 (0.86 - 1.04) | 0.28 |
| **PS-matching / on-treatment TAR without escalation** | | | | | | | | | | |
| CCAE | 33,169 | 56,168 | 21.2 | 35.3 | 892 | 1,457 | 1.05 (0.93 - 1.18) | 0.43 | 1.03 (0.91 - 1.16) | 0.68 |
| DAG | 9,491 | 1,839 | 9.2 | 1.8 | 135 | 21 | 1.80 (0.97 - 3.62) | 0.08 | 1.94 (1.00 - 3.75) | 0.06 |
| ODOD | 22,223 | 48,605 | 14.5 | 36.7 | 780 | 2,119 | 0.91 (0.82 - 1.02) | 0.12 | 0.96 (0.85 - 1.08) | 0.57 |
| MDCR | 7,015 | 13,103 | 5.7 | 10.5 | 244 | 486 | 0.92 (0.74 - 1.14) | 0.44 | 0.92 (0.75 - 1.15) | 0.50 |
| MDCD | 4,961 | 11,206 | 2.7 | 6.0 | 303 | 618 | 0.95 (0.78 - 1.16) | 0.62 | 0.92 (0.75 - 1.12) | 0.42 |
| Open Claims | 358,720 | 698,954 | 248.4 | 563.0 | 7,180 | 15,737 | 1.02 (0.98 - 1.06) | 0.45 | 1.01 (0.93 - 1.10) | 0.83 |
| OEHR | 31,233 | 71,686 | 12.5 | 33.6 | 584 | 1,757 | 0.87 (0.76 - 0.98) | 0.03 | 0.88 (0.77 - 1.00) | 0.07 |
| VA | 1,492 | 7,880 | 0.9 | 5.2 | 24 | 121 | 1.29 (0.67 - 2.39) | 0.44 | 1.16 (0.61 - 2.19) | 0.66 |
| SIDIAP | 18,524 | 5,273 | 37.9 | 11.2 | 331 | 93 | 0.96 (0.70 - 1.34) | 0.81 | 1.03 (0.74 - 1.43) | 0.82 |
| Meta-analysis | 486,828 | 914,714 | 353.0 | 703.2 | 10,473 | 22,409 | 0.97 (0.92 - 1.04) | 0.39 | 0.97 (0.87 - 1.08) | 0.55 |
| Meta-analysis (w/o USOC) | 128,108 | 215,760 | 104.6 | 140.2 | 3,293 | 6,672 | 0.95 (0.88 - 1.03) | 0.25 | 0.96 (0.88 - 1.05) | 0.49 |
| **PS-stratification / on-treatment TAR without escalation** | | | | | | | | | | |
| CCAE | 38,599 | 57,863 | 24.9 | 36.2 | 1,058 | 1,486 | 1.03 (0.95 - 1.12) | 0.46 | 1.03 (0.94 - 1.12) | 0.64 |
| DAG | 9,527 | 2,130 | 9.2 | 2.1 | 135 | 27 | 1.49 (0.95 - 2.41) | 0.09 | 1.49 (0.94 - 2.37) | 0.11 |
| ODOD | 23,886 | 48,728 | 15.7 | 36.8 | 846 | 2,124 | 0.91 (0.84 - 0.99) | 0.03 | 0.95 (0.87 - 1.05) | 0.45 |
| MDCR | 7,552 | 13,481 | 6.1 | 10.8 | 271 | 495 | 0.91 (0.78 - 1.07) | 0.26 | 0.92 (0.78 - 1.08) | 0.38 |
| MDCD | 5,142 | 11,261 | 2.8 | 6.1 | 316 | 621 | 1.11 (0.96 - 1.28) | 0.14 | 1.07 (0.92 - 1.23) | 0.48 |
| Open Claims | 367,392 | 699,041 | 256.1 | 563.0 | 7,482 | 15,741 | 1.01 (0.98 - 1.04) | 0.48 | 0.99 (0.90 - 1.10) | 0.88 |
| OEHR | 33,348 | 71,751 | 13.6 | 33.6 | 638 | 1,758 | 0.92 (0.83 - 1.01) | 0.09 | 0.92 (0.84 - 1.02) | 0.18 |
| VA | 1,778 | 11,035 | 1.0 | 7.6 | 26 | 189 | 1.01 (0.59 - 1.66) | 0.98 | 0.89 (0.53 - 1.48) | 0.65 |
| SIDIAP | 18,621 | 6,878 | 38.0 | 15.2 | 332 | 122 | 0.83 (0.65 - 1.07) | 0.15 | 0.83 (0.64 - 1.06) | 0.17 |
| Meta-analysis | 498,925 | 899,872 | 363.5 | 697.6 | 10,762 | 21,753 | 0.97 (0.92 - 1.02) | 0.24 | 0.96 (0.87 - 1.06) | 0.45 |
| Meta-analysis (w/o USOC) | 131,533 | 200,831 | 107.4 | 134.7 | 3,280 | 6,012 | 0.95 (0.88 - 1.02) | 0.18 | 0.96 (0.87 - 1.06) | 0.45 |

##### 1.6.23 Peripheral edema

**Supplementary Table 153:** Exposures, outcomes and risk estimates for Peripheral edema in DPP4I (female) (T) vs SU (female) (C) new-user cohorts under four different propensity score (PS) adjustment / time-at-risk (TAR) designs across data sources. We report total patient exposure time in 1,000 patient-year (KYr) units, uncalibrated and calibrated hazard ratios (HRs) and their 95% confidence intervals (CIs). Greyed out entries did not pass study diagnostics and are included only for completeness.

| | Patients | | Exposure (KYr) | | Outcomes | | Uncalibrated | | Calibrated | |
|---|---|---|---|---|---|---|---|---|---|---|
| | T | C | T | C | T | C | HR (95% CI) | $p$ | HR (95% CI) | $p$ |
| **PS-matching / on-treatment TAR** | | | | | | | | | | |
| CCAE | 30,818 | 52,614 | 23.2 | 36.9 | 1,179 | 2,139 | 0.95 (0.86 - 1.06) | 0.36 | 0.92 (0.83 - 1.02) | 0.17 |
| DAG | 8,516 | 1,689 | 9.0 | 1.7 | 312 | 71 | 1.04 (0.72 - 1.52) | 0.84 | 1.09 (0.75 - 1.59) | 0.64 |
| ODOD | 19,974 | 44,143 | 14.6 | 35.0 | 1,377 | 3,506 | 0.97 (0.89 - 1.06) | 0.47 | 1.00 (0.91 - 1.10) | 0.84 |
| MDCR | 5,797 | 11,111 | 5.1 | 9.3 | 468 | 998 | 0.85 (0.72 - 1.00) | 0.05 | 0.86 (0.73 - 1.02) | 0.10 |
| MDCD | 4,727 | 11,019 | 3.0 | 6.5 | 352 | 725 | 1.08 (0.89 - 1.30) | 0.43 | 1.04 (0.86 - 1.26) | 0.70 |
| Open Claims | 322,999 | 641,166 | 272.3 | 586.1 | 10,940 | 25,277 | 0.92 (0.89 - 0.95) | <0.01 | 0.91 (0.83 - 1.00) | 0.06 |
| OEHR | 29,650 | 68,002 | 14.3 | 35.5 | 806 | 2,574 | 0.90 (0.81 - 1.00) | 0.05 | 0.91 (0.82 - 1.02) | 0.13 |
| VA | 1,246 | 6,497 | 0.9 | 4.7 | 49 | 207 | 1.70 (1.09 - 2.62) | 0.02 | 1.54 (0.99 - 2.38) | 0.07 |
| SIDIAP | 17,390 | 5,086 | 47.0 | 12.9 | 926 | 235 | 0.90 (0.74 - 1.10) | 0.30 | 0.94 (0.77 - 1.15) | 0.59 |
| Meta-analysis | 441,117 | 841,327 | 389.5 | 728.6 | 16,409 | 35,732 | 0.93 (0.90 - 0.95) | <0.01 | 0.92 (0.85 - 0.99) | 0.03 |
| Meta-analysis (w/o USOC) | 118,118 | 200,161 | 117.1 | 142.5 | 5,469 | 10,455 | 0.96 (0.89 - 1.03) | 0.22 | 0.96 (0.88 - 1.04) | 0.40 |
| **PS-stratification / on-treatment TAR** | | | | | | | | | | |
| CCAE | 35,723 | 54,344 | 27.2 | 37.9 | 1,396 | 2,189 | 0.89 (0.83 - 0.95) | <0.01 | 0.88 (0.81 - 0.95) | <0.01 |
| DAG | 8,543 | 1,952 | 9.0 | 2.0 | 315 | 81 | 0.88 (0.67 - 1.17) | 0.37 | 0.87 (0.65 - 1.15) | 0.39 |
| ODOD | 21,408 | 44,278 | 15.7 | 35.1 | 1,486 | 3,509 | 0.97 (0.91 - 1.04) | 0.39 | 1.00 (0.93 - 1.07) | 0.84 |
| MDCR | 6,251 | 11,373 | 5.6 | 9.5 | 516 | 1,017 | 0.86 (0.77 - 0.96) | <0.01 | 0.87 (0.78 - 0.98) | 0.05 |
| MDCD | 4,930 | 11,097 | 3.1 | 6.6 | 377 | 726 | 1.12 (0.98 - 1.28) | 0.10 | 1.08 (0.94 - 1.23) | 0.36 |
| Open Claims | 329,855 | 641,293 | 279.7 | 586.1 | 11,328 | 25,279 | 0.92 (0.90 - 0.94) | <0.01 | 0.90 (0.82 - 0.99) | 0.04 |
| OEHR | 31,683 | 68,073 | 15.5 | 35.5 | 858 | 2,577 | 0.84 (0.78 - 0.92) | <0.01 | 0.85 (0.78 - 0.93) | <0.01 |
| VA | 1,478 | 9,426 | 1.0 | 7.1 | 55 | 346 | 1.44 (0.99 - 2.05) | 0.05 | 1.25 (0.87 - 1.80) | 0.25 |
| SIDIAP | 17,464 | 6,675 | 47.1 | 17.7 | 930 | 301 | 0.91 (0.78 - 1.06) | 0.21 | 0.88 (0.76 - 1.03) | 0.16 |
| Meta-analysis | 450,927 | 827,988 | 399.7 | 723.7 | 16,829 | 34,953 | 0.91 (0.87 - 0.94) | <0.01 | 0.89 (0.81 - 0.99) | 0.03 |
| Meta-analysis (w/o USOC) | 121,072 | 186,695 | 120.0 | 137.6 | 5,501 | 9,674 | 0.90 (0.85 - 0.95) | <0.01 | 0.90 (0.82 - 0.98) | 0.03 |
| **PS-matching / on-treatment TAR without escalation** | | | | | | | | | | |
| CCAE | 30,818 | 52,614 | 19.3 | 32.1 | 986 | 1,857 | 0.96 (0.86 - 1.07) | 0.43 | 0.94 (0.84 - 1.05) | 0.34 |
| DAG | 8,516 | 1,689 | 8.1 | 1.6 | 284 | 65 | 1.04 (0.71 - 1.55) | 0.86 | 1.11 (0.75 - 1.66) | 0.61 |
| ODOD | 19,974 | 44,143 | 12.3 | 31.5 | 1,144 | 3,125 | 0.96 (0.87 - 1.05) | 0.37 | 1.00 (0.91 - 1.11) | 0.82 |
| MDCR | 5,797 | 11,111 | 4.5 | 8.6 | 404 | 929 | 0.79 (0.67 - 0.94) | <0.01 | 0.80 (0.67 - 0.95) | 0.02 |
| MDCD | 4,727 | 11,019 | 2.4 | 5.6 | 275 | 620 | 1.10 (0.89 - 1.34) | 0.38 | 1.06 (0.86 - 1.30) | 0.63 |
| Open Claims | 322,999 | 641,166 | 218.1 | 497.3 | 8,695 | 21,267 | 0.91 (0.88 - 0.94) | <0.01 | 0.91 (0.84 - 0.98) | 0.04 |
| OEHR | 29,650 | 68,002 | 11.7 | 30.8 | 613 | 2,105 | 0.89 (0.79 - 1.01) | 0.07 | 0.90 (0.80 - 1.03) | 0.16 |
| VA | 1,246 | 6,497 | 0.7 | 4.1 | 41 | 176 | 1.82 (1.12 - 2.91) | 0.01 | 1.64 (1.02 - 2.65) | 0.05 |
| SIDIAP | 17,390 | 5,086 | 35.0 | 10.6 | 690 | 194 | 0.79 (0.63 - 0.99) | 0.04 | 0.84 (0.67 - 1.06) | 0.19 |
| Meta-analysis | 441,117 | 841,327 | 312.0 | 622.2 | 13,132 | 30,338 | 0.92 (0.89 - 0.95) | <0.01 | 0.91 (0.84 - 1.00) | 0.05 |
| Meta-analysis (w/o USOC) | 118,118 | 200,161 | 94.0 | 124.9 | 4,437 | 9,071 | 0.94 (0.86 - 1.03) | 0.21 | 0.95 (0.86 - 1.05) | 0.39 |
| **PS-stratification / on-treatment TAR without escalation** | | | | | | | | | | |
| CCAE | 35,723 | 54,344 | 22.6 | 33.1 | 1,160 | 1,900 | 0.90 (0.83 - 0.97) | <0.01 | 0.89 (0.82 - 0.97) | 0.01 |
| DAG | 8,543 | 1,952 | 8.2 | 1.8 | 287 | 73 | 0.89 (0.67 - 1.20) | 0.43 | 0.89 (0.66 - 1.19) | 0.48 |
| ODOD | 21,408 | 44,278 | 13.3 | 31.6 | 1,231 | 3,128 | 0.95 (0.89 - 1.02) | 0.20 | 1.00 (0.92 - 1.08) | 0.85 |
| MDCR | 6,251 | 11,373 | 4.9 | 8.8 | 449 | 944 | 0.84 (0.75 - 0.95) | <0.01 | 0.85 (0.75 - 0.96) | 0.02 |
| MDCD | 4,930 | 11,097 | 2.5 | 5.6 | 295 | 621 | 1.07 (0.92 - 1.24) | 0.35 | 1.03 (0.88 - 1.19) | 0.76 |
| Open Claims | 329,855 | 641,293 | 224.0 | 497.3 | 9,012 | 21,269 | 0.91 (0.89 - 0.94) | <0.01 | 0.90 (0.82 - 0.99) | 0.04 |
| OEHR | 31,683 | 68,073 | 12.6 | 30.8 | 658 | 2,106 | 0.83 (0.76 - 0.91) | <0.01 | 0.84 (0.76 - 0.92) | <0.01 |
| VA | 1,478 | 9,426 | 0.8 | 6.3 | 46 | 293 | 1.56 (1.04 - 2.31) | 0.03 | 1.37 (0.92 - 2.05) | 0.14 |
| SIDIAP | 17,464 | 6,675 | 35.0 | 14.4 | 694 | 243 | 0.90 (0.76 - 1.07) | 0.24 | 0.90 (0.75 - 1.07) | 0.29 |
| Meta-analysis | 450,927 | 827,988 | 320.5 | 617.8 | 13,491 | 29,663 | 0.90 (0.87 - 0.93) | <0.01 | 0.90 (0.82 - 0.98) | 0.02 |
| Meta-analysis (w/o USOC) | 121,072 | 186,695 | 96.6 | 120.5 | 4,479 | 8,394 | 0.89 (0.85 - 0.94) | <0.01 | 0.90 (0.83 - 0.98) | 0.03 |

##### 1.6.24 Photosensitivity

**Supplementary Table 154:** Exposures, outcomes and risk estimates for Photosensitivity in DPP4I (female) (T) vs SU (female) (C) new-user cohorts under four different propensity score (PS) adjustment / time-at-risk (TAR) designs across data sources. We report total patient exposure time in 1,000 patient-year (KYr) units, uncalibrated and calibrated hazard ratios (HRs) and their 95% confidence intervals (CIs). Greyed out entries did not pass study diagnostics and are included only for completeness.

| | Patients | | Exposure (KYr) | | Outcomes | | Uncalibrated | | Calibrated | |
|---|---|---|---|---|---|---|---|---|---|---|
| | T | C | T | C | T | C | HR (95% CI) | $p$ | HR (95% CI) | $p$ |
| **PS-matching / on-treatment TAR** | | | | | | | | | | |
| CCAE | 38,750 | 65,513 | 30.3 | 47.7 | 42 | 76 | 1.23 (0.70 - 2.17) | 0.47 | 1.18 (0.67 - 2.09) | 0.56 |
| DAG | 10,440 | 2,033 | 11.3 | 2.1 | <5 | <5 | | | | |
| ODOD | 27,224 | 59,640 | 21.5 | 51.7 | 42 | 81 | 1.66 (0.98 - 2.81) | 0.06 | 1.72 (1.02 - 2.91) | 0.05 |
| MDCR | 8,046 | 15,282 | 7.6 | 13.6 | 24 | 30 | 1.40 (0.70 - 2.81) | 0.34 | 1.41 (0.71 - 2.83) | 0.33 |
| MDCD | 7,363 | 16,649 | 5.2 | 10.9 | 10 | 10 | 0.85 (0.22 - 2.87) | 0.80 | 0.82 (0.23 - 2.97) | 0.76 |
| Open Claims | 435,042 | 844,230 | 379.6 | 802.8 | 199 | 387 | 1.01 (0.79 - 1.29) | 0.93 | 1.00 (0.77 - 1.30) | 0.96 |
| OEHR | 36,161 | 82,685 | 18.5 | 47.0 | 35 | 67 | 1.52 (0.89 - 2.55) | 0.12 | 1.54 (0.91 - 2.61) | 0.11 |
| VA | 1,802 | 9,559 | 1.4 | 7.4 | 0 | <10 | | | | |
| SIDIAP | 19,472 | 5,426 | 53.6 | 14.2 | 10 | <5 | 1.18 (0.26 - 8.44) | 0.86 | 1.23 (0.21 - 7.04) | 0.82 |
| Meta-analysis | 552,586 | 1,083,999 | 462.7 | 973.7 | 352 | 651 | 1.23 (0.97 - 1.55) | 0.08 | 1.21 (0.95 - 1.55) | 0.12 |
| Meta-analysis (w/o USOC) | 117,544 | 239,769 | 83.1 | 170.9 | 153 | 264 | 1.42 (1.08 - 1.88) | 0.01 | 1.42 (1.07 - 1.89) | 0.02 |
| **PS-stratification / on-treatment TAR** | | | | | | | | | | |
| CCAE | 45,015 | 67,427 | 35.5 | 48.8 | 50 | 77 | 0.93 (0.63 - 1.35) | 0.70 | 0.92 (0.63 - 1.34) | 0.66 |
| DAG | 10,471 | 2,360 | 11.3 | 2.5 | <5 | <5 | 0.22 (0.03 - 1.90) | 0.17 | 0.22 (0.03 - 1.87) | 0.17 |
| ODOD | 29,190 | 59,795 | 23.2 | 51.7 | 45 | 81 | 1.21 (0.82 - 1.78) | 0.33 | 1.25 (0.85 - 1.84) | 0.27 |
| MDCR | 8,668 | 15,603 | 8.2 | 13.9 | 25 | 30 | 1.44 (0.82 - 2.51) | 0.20 | 1.46 (0.83 - 2.56) | 0.19 |
| MDCD | 7,706 | 16,755 | 5.5 | 11.0 | 11 | 10 | 1.83 (0.74 - 4.56) | 0.19 | 1.76 (0.71 - 4.36) | 0.22 |
| Open Claims | 444,867 | 844,327 | 390.5 | 802.8 | 204 | 387 | 1.00 (0.84 - 1.19) | 0.97 | 0.99 (0.81 - 1.20) | 0.89 |
| OEHR | 38,618 | 82,774 | 20.0 | 47.0 | 37 | 67 | 1.69 (1.09 - 2.58) | 0.02 | 1.71 (1.11 - 2.63) | 0.02 |
| VA | 2,194 | 13,060 | 1.6 | 10.5 | 0 | <10 | | | | |
| SIDIAP | 19,584 | 7,060 | 53.7 | 19.4 | 10 | <5 | 0.79 (0.20 - 4.12) | 0.77 | 0.77 (0.17 - 3.51) | 0.74 |
| Meta-analysis | 566,358 | 1,069,926 | 477.4 | 964.3 | 361 | 642 | 1.16 (0.94 - 1.42) | 0.17 | 1.14 (0.91 - 1.43) | 0.26 |
| Meta-analysis (w/o USOC) | 121,491 | 225,599 | 86.8 | 161.5 | 157 | 255 | 1.25 (0.96 - 1.63) | 0.09 | 1.26 (0.96 - 1.65) | 0.10 |
| **PS-matching / on-treatment TAR without escalation** | | | | | | | | | | |
| CCAE | 38,750 | 65,513 | 24.9 | 41.1 | 39 | 68 | 1.21 (0.64 - 2.28) | 0.56 | 1.18 (0.63 - 2.23) | 0.60 |
| DAG | 10,440 | 2,033 | 10.2 | 2.0 | <5 | <5 | | | | |
| ODOD | 27,224 | 59,640 | 18.0 | 46.0 | 36 | 76 | 1.55 (0.90 - 2.67) | 0.11 | 1.63 (0.94 - 2.81) | 0.08 |
| MDCR | 8,046 | 15,282 | 6.6 | 12.5 | 22 | 27 | 1.43 (0.70 - 2.94) | 0.33 | 1.44 (0.70 - 2.94) | 0.32 |
| MDCD | 7,363 | 16,649 | 4.1 | 9.1 | 8 | 8 | 0.52 (0.08 - 2.32) | 0.46 | 0.50 (0.09 - 2.78) | 0.43 |
| Open Claims | 435,042 | 844,230 | 301.3 | 675.2 | 172 | 334 | 1.05 (0.80 - 1.37) | 0.71 | 1.05 (0.79 - 1.38) | 0.75 |
| OEHR | 36,161 | 82,685 | 14.9 | 39.8 | 30 | 57 | 1.38 (0.77 - 2.43) | 0.28 | 1.39 (0.78 - 2.48) | 0.26 |
| VA | 1,802 | 9,559 | 1.1 | 6.4 | 0 | <10 | | | | |
| SIDIAP | 19,472 | 5,426 | 39.9 | 11.5 | 9 | <5 | 2.62 (0.40 - 51.4) | 0.44 | 2.81 (0.25 - 31.7) | 0.40 |
| Meta-analysis | 545,223 | 1,067,350 | 365.7 | 814.5 | 299 | 562 | 1.19 (0.97 - 1.46) | 0.09 | 1.19 (0.95 - 1.48) | 0.13 |
| Meta-analysis (w/o USOC) | 110,181 | 223,120 | 64.4 | 139.3 | 127 | 228 | 1.40 (1.03 - 1.89) | 0.03 | 1.41 (1.04 - 1.92) | 0.03 |
| **PS-stratification / on-treatment TAR without escalation** | | | | | | | | | | |
| CCAE | 45,015 | 67,427 | 29.2 | 42.1 | 45 | 69 | 0.99 (0.66 - 1.47) | 0.95 | 0.98 (0.66 - 1.46) | 0.92 |
| DAG | 10,471 | 2,360 | 10.2 | 2.3 | <5 | <5 | 0.48 (0.04 - 10.6) | 0.60 | 0.48 (0.03 - 7.43) | 0.60 |
| ODOD | 29,190 | 59,795 | 19.4 | 46.0 | 37 | 76 | 1.14 (0.74 - 1.72) | 0.55 | 1.19 (0.78 - 1.81) | 0.42 |
| MDCR | 8,668 | 15,603 | 7.1 | 12.7 | 23 | 27 | 1.48 (0.82 - 2.65) | 0.19 | 1.49 (0.82 - 2.68) | 0.20 |
| MDCD | 7,706 | 16,755 | 4.3 | 9.1 | 9 | 8 | 1.77 (0.65 - 4.92) | 0.27 | 1.70 (0.62 - 4.67) | 0.31 |
| Open Claims | 444,867 | 844,327 | 310.0 | 675.2 | 176 | 334 | 1.05 (0.86 - 1.26) | 0.65 | 1.03 (0.83 - 1.27) | 0.80 |
| OEHR | 38,618 | 82,774 | 16.1 | 39.9 | 31 | 57 | 1.75 (1.08 - 2.78) | 0.02 | 1.76 (1.09 - 2.82) | 0.02 |
| VA | 2,194 | 13,060 | 1.3 | 9.2 | 0 | <10 | | | | |
| SIDIAP | 19,584 | 7,060 | 39.9 | 15.7 | 9 | <5 | 1.56 (0.32 - 11.9) | 0.63 | 1.54 (0.25 - 9.40) | 0.64 |
| Meta-analysis | 566,358 | 1,069,926 | 381.9 | 816.0 | 312 | 563 | 1.15 (0.96 - 1.38) | 0.12 | 1.15 (0.94 - 1.40) | 0.17 |
| Meta-analysis (w/o USOC) | 121,491 | 225,599 | 71.8 | 140.7 | 136 | 229 | 1.26 (0.97 - 1.63) | 0.08 | 1.27 (0.98 - 1.66) | 0.08 |

##### 1.6.25 Venous thromboembolism

**Supplementary Table 155:** Exposures, outcomes and risk estimates for Venous thromboembolism in DPP4I (female) (T) vs SU (female) (C) new-user cohorts under four different propensity score (PS) adjustment / time-at-risk (TAR) designs across data sources. We report total patient exposure time in 1,000 patient-year (KYr) units, uncalibrated and calibrated hazard ratios (HRs) and their 95% confidence intervals (CIs). Greyed out entries did not pass study diagnostics and are included only for completeness.

| | Patients | | Exposure (KYr) | | Outcomes | | Uncalibrated | | Calibrated | |
|---|---|---|---|---|---|---|---|---|---|---|
| | T | C | T | C | T | C | HR (95% CI) | $p$ | HR (95% CI) | $p$ |
| **PS-matching / on-treatment TAR** | | | | | | | | | | |
| CCAE | 38,199 | 64,583 | 29.8 | 47.0 | 160 | 302 | 0.72 (0.54 - 0.95) | 0.02 | 0.69 (0.52 - 0.92) | 0.01 |
| DAG | 9,609 | 1,856 | 10.3 | 1.9 | 113 | 22 | 0.86 (0.48 - 1.64) | 0.65 | 0.90 (0.49 - 1.69) | 0.74 |
| ODOD | 26,631 | 58,121 | 21.0 | 50.1 | 231 | 621 | 0.97 (0.78 - 1.19) | 0.76 | 1.00 (0.81 - 1.24) | 0.91 |
| MDCR | 7,810 | 14,717 | 7.4 | 13.0 | 101 | 227 | 0.82 (0.57 - 1.17) | 0.29 | 0.83 (0.58 - 1.19) | 0.33 |
| MDCD | 7,076 | 16,013 | 5.0 | 10.4 | 56 | 131 | 0.95 (0.59 - 1.51) | 0.84 | 0.92 (0.58 - 1.47) | 0.74 |
| Open Claims | 423,945 | 822,965 | 369.2 | 780.4 | 1,956 | 4,999 | 0.82 (0.76 - 0.88) | <0.01 | 0.81 (0.73 - 0.91) | <0.01 |
| OEHR | 35,455 | 81,178 | 18.0 | 45.6 | 131 | 472 | 0.75 (0.57 - 0.98) | 0.04 | 0.76 (0.58 - 1.01) | 0.06 |
| VA | 1,767 | 9,456 | 1.3 | 7.3 | <10 | 28 | 1.26 (0.17 - 5.77) | 0.79 | 1.14 (0.20 - 6.55) | 0.88 |
| SIDIAP | 18,840 | 5,317 | 51.8 | 13.8 | 264 | 63 | 1.08 (0.73 - 1.62) | 0.70 | 1.13 (0.76 - 1.68) | 0.57 |
| Meta-analysis | 569,332 | 1,074,206 | 513.7 | 969.6 | <3,022 | 6,865 | 0.83 (0.78 - 0.89) | <0.01 | 0.82 (0.75 - 0.91) | <0.01 |
| Meta-analysis (w/o USOC) | 145,387 | 251,241 | 144.5 | 189.2 | <1,066 | 1,866 | 0.86 (0.77 - 0.97) | 0.02 | 0.87 (0.76 - 0.98) | 0.04 |
| **PS-stratification / on-treatment TAR** | | | | | | | | | | |
| CCAE | 44,442 | 66,492 | 35.0 | 48.1 | 185 | 310 | 0.87 (0.72 - 1.06) | 0.16 | 0.86 (0.71 - 1.05) | 0.14 |
| DAG | 9,629 | 2,158 | 10.3 | 2.3 | 115 | 22 | 1.17 (0.72 - 1.97) | 0.55 | 1.15 (0.69 - 1.91) | 0.62 |
| ODOD | 28,540 | 58,276 | 22.6 | 50.2 | 240 | 622 | 0.99 (0.84 - 1.15) | 0.85 | 1.02 (0.87 - 1.19) | 0.85 |
| MDCR | 8,418 | 15,107 | 7.9 | 13.4 | 108 | 229 | 0.78 (0.61 - 0.99) | 0.05 | 0.79 (0.62 - 1.01) | 0.08 |
| MDCD | 7,411 | 16,115 | 5.2 | 10.5 | 61 | 131 | 1.03 (0.73 - 1.42) | 0.88 | 0.99 (0.71 - 1.37) | 0.91 |
| Open Claims | 433,673 | 823,062 | 380.1 | 780.4 | 2,031 | 4,999 | 0.83 (0.79 - 0.88) | <0.01 | 0.82 (0.74 - 0.91) | <0.01 |
| OEHR | 37,874 | 81,264 | 19.5 | 45.7 | 139 | 472 | 0.82 (0.67 - 0.99) | 0.05 | 0.82 (0.67 - 1.01) | 0.07 |
| VA | 2,154 | 12,911 | 1.6 | 10.4 | <10 | 42 | 0.89 (0.24 - 2.93) | 0.86 | 0.77 (0.22 - 2.73) | 0.69 |
| SIDIAP | 18,952 | 6,941 | 51.9 | 18.9 | 265 | 84 | 1.01 (0.77 - 1.35) | 0.92 | 0.98 (0.74 - 1.31) | 0.87 |
| Meta-analysis | 581,528 | 1,053,300 | 527.3 | 959.0 | 3,083 | 6,738 | 0.87 (0.81 - 0.94) | <0.01 | 0.86 (0.76 - 0.97) | 0.01 |
| Meta-analysis (w/o USOC) | 147,855 | 230,238 | 147.2 | 178.6 | 1,052 | 1,739 | 0.90 (0.82 - 0.99) | 0.03 | 0.90 (0.81 - 1.02) | 0.11 |
| **PS-matching / on-treatment TAR without escalation** | | | | | | | | | | |
| CCAE | 38,199 | 64,583 | 24.5 | 40.5 | 128 | 260 | 0.70 (0.52 - 0.94) | 0.02 | 0.69 (0.51 - 0.93) | 0.02 |
| DAG | 9,609 | 1,856 | 9.3 | 1.8 | 105 | 21 | 0.85 (0.46 - 1.62) | 0.60 | 0.91 (0.48 - 1.71) | 0.75 |
| ODOD | 26,631 | 58,121 | 17.6 | 44.6 | 204 | 563 | 1.02 (0.82 - 1.27) | 0.83 | 1.07 (0.86 - 1.34) | 0.55 |
| MDCR | 7,810 | 14,717 | 6.4 | 12.0 | 85 | 207 | 0.80 (0.54 - 1.16) | 0.24 | 0.80 (0.55 - 1.17) | 0.27 |
| MDCD | 7,076 | 16,013 | 3.9 | 8.7 | 39 | 108 | 0.77 (0.43 - 1.33) | 0.37 | 0.74 (0.43 - 1.30) | 0.31 |
| Open Claims | 423,945 | 822,965 | 293.2 | 656.7 | 1,576 | 4,270 | 0.80 (0.73 - 0.86) | <0.01 | 0.79 (0.71 - 0.88) | <0.01 |
| OEHR | 35,455 | 81,178 | 14.5 | 38.8 | 87 | 366 | 0.72 (0.52 - 0.98) | 0.04 | 0.73 (0.52 - 1.00) | 0.06 |
| VA | 1,767 | 9,456 | 1.1 | 6.3 | <10 | 21 | 1.13 (0.06 - 7.64) | 0.92 | 1.02 (0.09 - 11.8) | 0.96 |
| SIDIAP | 18,840 | 5,317 | 38.5 | 11.3 | 191 | 48 | 1.33 (0.80 - 2.28) | 0.29 | 1.42 (0.84 - 2.40) | 0.20 |
| Meta-analysis | 569,332 | 1,074,206 | 409.0 | 820.6 | <2,425 | 5,864 | 0.83 (0.74 - 0.94) | <0.01 | 0.83 (0.72 - 0.96) | 0.01 |
| Meta-analysis (w/o USOC) | 145,387 | 251,241 | 115.8 | 163.9 | <849 | 1,594 | 0.86 (0.74 - 1.00) | 0.05 | 0.87 (0.74 - 1.01) | 0.09 |
| **PS-stratification / on-treatment TAR without escalation** | | | | | | | | | | |
| CCAE | 44,442 | 66,492 | 28.8 | 41.5 | 147 | 266 | 0.84 (0.68 - 1.04) | 0.12 | 0.84 (0.67 - 1.04) | 0.12 |
| DAG | 9,629 | 2,158 | 9.3 | 2.1 | 107 | 21 | 1.16 (0.71 - 1.99) | 0.57 | 1.16 (0.69 - 1.95) | 0.59 |
| ODOD | 28,540 | 58,276 | 18.9 | 44.6 | 211 | 564 | 1.01 (0.85 - 1.19) | 0.92 | 1.06 (0.89 - 1.25) | 0.57 |
| MDCR | 8,418 | 15,107 | 6.9 | 12.3 | 89 | 209 | 0.75 (0.57 - 0.97) | 0.03 | 0.75 (0.58 - 0.98) | 0.05 |
| MDCD | 7,411 | 16,115 | 4.1 | 8.7 | 44 | 108 | 0.95 (0.64 - 1.38) | 0.79 | 0.91 (0.62 - 1.33) | 0.64 |
| Open Claims | 433,673 | 823,062 | 302.0 | 656.7 | 1,637 | 4,270 | 0.83 (0.78 - 0.88) | <0.01 | 0.81 (0.73 - 0.91) | <0.01 |
| OEHR | 37,874 | 81,264 | 15.7 | 38.9 | 95 | 366 | 0.74 (0.58 - 0.94) | 0.02 | 0.75 (0.59 - 0.95) | 0.02 |
| VA | 2,154 | 12,911 | 1.3 | 9.0 | <10 | 33 | 2.04 (0.51 - 6.87) | 0.28 | 1.79 (0.49 - 6.58) | 0.38 |
| SIDIAP | 18,952 | 6,941 | 38.6 | 15.4 | 192 | 61 | 1.05 (0.75 - 1.48) | 0.78 | 1.04 (0.74 - 1.46) | 0.82 |
| Meta-analysis | 581,528 | 1,053,300 | 420.3 | 811.5 | 2,478 | 5,757 | 0.86 (0.79 - 0.95) | <0.01 | 0.86 (0.76 - 0.97) | 0.02 |
| Meta-analysis (w/o USOC) | 147,855 | 230,238 | 118.3 | 154.7 | 841 | 1,487 | 0.89 (0.77 - 1.01) | 0.08 | 0.89 (0.77 - 1.04) | 0.16 |

##### 1.6.26 Vomiting

**Supplementary Table 156:** Exposures, outcomes and risk estimates for Vomiting in DPP4I (female) (T) vs SU (female) (C) new-user cohorts under four different propensity score (PS) adjustment / time-at-risk (TAR) designs across data sources. We report total patient exposure time in 1,000 patient-year (KYr) units, uncalibrated and calibrated hazard ratios (HRs) and their 95% confidence intervals (CIs). Greyed out entries did not pass study diagnostics and are included only for completeness.

| | Patients | | Exposure (KYr) | | Outcomes | | Uncalibrated | | Calibrated | |
|---|---|---|---|---|---|---|---|---|---|---|
| | T | C | T | C | T | C | HR (95% CI) | $p$ | HR (95% CI) | $p$ |
| **PS-matching / on-treatment TAR** | | | | | | | | | | |
| CCAE | 34,815 | 58,725 | 27.1 | 42.5 | 791 | 1,266 | 1.04 (0.92 - 1.18) | 0.53 | 1.00 (0.88 - 1.14) | 0.85 |
| DAG | 9,486 | 1,839 | 10.2 | 1.9 | 150 | 24 | 1.55 (0.90 - 2.84) | 0.14 | 1.62 (0.91 - 2.89) | 0.12 |
| ODOD | 23,788 | 51,899 | 18.5 | 44.2 | 672 | 1,854 | 0.87 (0.77 - 0.98) | 0.02 | 0.90 (0.79 - 1.02) | 0.18 |
| MDCR | 7,281 | 13,676 | 6.7 | 12.0 | 227 | 453 | 0.93 (0.74 - 1.16) | 0.51 | 0.94 (0.75 - 1.18) | 0.59 |
| MDCD | 5,459 | 12,359 | 3.7 | 7.9 | 288 | 562 | 1.15 (0.93 - 1.41) | 0.19 | 1.11 (0.90 - 1.36) | 0.37 |
| Open Claims | 377,793 | 733,701 | 328.5 | 699.2 | 6,955 | 14,507 | 1.04 (1.00 - 1.08) | 0.07 | 1.03 (0.94 - 1.13) | 0.56 |
| OEHR | 33,099 | 75,511 | 16.5 | 41.7 | 535 | 1,545 | 0.86 (0.75 - 0.98) | 0.03 | 0.87 (0.76 - 1.00) | 0.06 |
| VA | 1,579 | 8,275 | 1.2 | 6.3 | 18 | 106 | 0.86 (0.40 - 1.73) | 0.69 | 0.78 (0.38 - 1.62) | 0.51 |
| SIDIAP | 18,478 | 5,262 | 50.8 | 13.6 | 476 | 117 | 0.98 (0.74 - 1.31) | 0.88 | 1.02 (0.76 - 1.36) | 0.84 |
| Meta-analysis | 511,778 | 961,247 | 463.3 | 869.3 | 10,112 | 20,434 | 0.98 (0.91 - 1.06) | 0.65 | 0.97 (0.87 - 1.08) | 0.58 |
| Meta-analysis (w/o USOC) | 133,985 | 227,546 | 134.7 | 170.1 | 3,157 | 5,927 | 0.96 (0.88 - 1.06) | 0.46 | 0.97 (0.87 - 1.08) | 0.58 |
| **PS-stratification / on-treatment TAR** | | | | | | | | | | |
| CCAE | 40,551 | 60,512 | 31.7 | 43.6 | 925 | 1,294 | 1.02 (0.93 - 1.11) | 0.74 | 1.00 (0.91 - 1.10) | 0.86 |
| DAG | 9,522 | 2,129 | 10.2 | 2.3 | 150 | 31 | 1.41 (0.92 - 2.20) | 0.12 | 1.39 (0.89 - 2.14) | 0.17 |
| ODOD | 25,529 | 52,033 | 19.9 | 44.2 | 732 | 1,857 | 0.88 (0.80 - 0.96) | <0.01 | 0.91 (0.82 - 1.00) | 0.09 |
| MDCR | 7,844 | 14,058 | 7.3 | 12.3 | 245 | 461 | 0.87 (0.74 - 1.02) | 0.10 | 0.88 (0.75 - 1.04) | 0.19 |
| MDCD | 5,668 | 12,420 | 3.9 | 7.9 | 300 | 565 | 1.13 (0.97 - 1.31) | 0.10 | 1.09 (0.93 - 1.27) | 0.33 |
| Open Claims | 386,754 | 733,792 | 338.3 | 699.2 | 7,240 | 14,509 | 1.01 (0.98 - 1.04) | 0.42 | 1.00 (0.91 - 1.09) | 0.89 |
| OEHR | 35,368 | 75,582 | 17.9 | 41.7 | 580 | 1,545 | 0.91 (0.82 - 1.01) | 0.07 | 0.92 (0.83 - 1.02) | 0.17 |
| VA | 1,911 | 11,600 | 1.4 | 9.2 | 20 | 161 | 0.86 (0.48 - 1.49) | 0.61 | 0.75 (0.42 - 1.33) | 0.33 |
| SIDIAP | 18,575 | 6,868 | 50.9 | 18.7 | 477 | 154 | 0.87 (0.70 - 1.08) | 0.19 | 0.84 (0.68 - 1.04) | 0.15 |
| Meta-analysis | 524,143 | 944,974 | 476.2 | 862.0 | 10,349 | 19,851 | 0.95 (0.89 - 1.01) | 0.13 | 0.94 (0.84 - 1.05) | 0.29 |
| Meta-analysis (w/o USOC) | 137,389 | 211,182 | 137.9 | 162.8 | 3,109 | 5,342 | 0.93 (0.86 - 1.00) | 0.06 | 0.93 (0.84 - 1.03) | 0.21 |
| **PS-matching / on-treatment TAR without escalation** | | | | | | | | | | |
| CCAE | 34,815 | 58,725 | 22.4 | 36.9 | 648 | 1,064 | 1.07 (0.93 - 1.22) | 0.35 | 1.05 (0.91 - 1.20) | 0.56 |
| DAG | 9,486 | 1,839 | 9.2 | 1.8 | 135 | 21 | 1.78 (0.96 - 3.58) | 0.08 | 1.92 (0.99 - 3.71) | 0.07 |
| ODOD | 23,788 | 51,899 | 15.6 | 39.5 | 560 | 1,664 | 0.87 (0.76 - 0.99) | 0.03 | 0.91 (0.80 - 1.04) | 0.24 |
| MDCR | 7,281 | 13,676 | 5.9 | 11.0 | 187 | 398 | 0.98 (0.77 - 1.24) | 0.85 | 0.98 (0.77 - 1.25) | 0.83 |
| MDCD | 5,459 | 12,359 | 3.0 | 6.6 | 217 | 484 | 1.14 (0.91 - 1.43) | 0.25 | 1.10 (0.88 - 1.38) | 0.44 |
| Open Claims | 377,793 | 733,701 | 262.3 | 591.6 | 5,476 | 11,992 | 1.04 (0.99 - 1.09) | 0.09 | 1.03 (0.95 - 1.13) | 0.50 |
| OEHR | 33,099 | 75,511 | 13.4 | 35.8 | 406 | 1,228 | 0.85 (0.73 - 0.99) | 0.04 | 0.86 (0.74 - 1.00) | 0.07 |
| VA | 1,579 | 8,275 | 0.9 | 5.5 | 14 | 85 | 0.94 (0.41 - 1.99) | 0.87 | 0.84 (0.38 - 1.85) | 0.68 |
| SIDIAP | 18,478 | 5,262 | 37.8 | 11.1 | 345 | 92 | 0.99 (0.71 - 1.40) | 0.96 | 1.06 (0.76 - 1.49) | 0.73 |
| Meta-analysis | 511,778 | 961,247 | 370.6 | 739.7 | 7,988 | 17,028 | 0.99 (0.91 - 1.08) | 0.84 | 0.99 (0.87 - 1.11) | 0.82 |
| Meta-analysis (w/o USOC) | 133,985 | 227,546 | 108.3 | 148.2 | 2,512 | 5,036 | 0.98 (0.88 - 1.09) | 0.69 | 0.99 (0.88 - 1.11) | 0.85 |
| **PS-stratification / on-treatment TAR without escalation** | | | | | | | | | | |
| CCAE | 40,551 | 60,512 | 26.3 | 37.9 | 761 | 1,092 | 1.04 (0.94 - 1.14) | 0.47 | 1.03 (0.93 - 1.14) | 0.61 |
| DAG | 9,522 | 2,129 | 9.2 | 2.1 | 135 | 28 | 1.45 (0.93 - 2.33) | 0.11 | 1.45 (0.92 - 2.29) | 0.14 |
| ODOD | 25,529 | 52,033 | 16.9 | 39.5 | 609 | 1,667 | 0.86 (0.78 - 0.94) | <0.01 | 0.90 (0.81 - 1.00) | 0.10 |
| MDCR | 7,844 | 14,058 | 6.3 | 11.3 | 204 | 404 | 0.88 (0.73 - 1.05) | 0.15 | 0.88 (0.74 - 1.06) | 0.23 |
| MDCD | 5,668 | 12,420 | 3.1 | 6.7 | 227 | 487 | 1.04 (0.88 - 1.22) | 0.67 | 0.99 (0.84 - 1.18) | 0.84 |
| Open Claims | 386,754 | 733,792 | 270.1 | 591.6 | 5,707 | 11,994 | 1.01 (0.98 - 1.04) | 0.50 | 1.00 (0.90 - 1.10) | 0.89 |
| OEHR | 35,368 | 75,582 | 14.5 | 35.9 | 442 | 1,228 | 0.91 (0.81 - 1.02) | 0.12 | 0.92 (0.82 - 1.03) | 0.21 |
| VA | 1,911 | 11,600 | 1.1 | 8.1 | 15 | 131 | 0.87 (0.44 - 1.63) | 0.67 | 0.76 (0.39 - 1.47) | 0.43 |
| SIDIAP | 18,575 | 6,868 | 37.9 | 15.1 | 346 | 122 | 0.87 (0.68 - 1.11) | 0.27 | 0.86 (0.67 - 1.11) | 0.29 |
| Meta-analysis | 524,143 | 944,974 | 381.3 | 733.4 | 8,204 | 16,535 | 0.95 (0.88 - 1.03) | 0.19 | 0.95 (0.85 - 1.06) | 0.34 |
| Meta-analysis (w/o USOC) | 137,389 | 211,182 | 111.1 | 141.8 | 2,497 | 4,541 | 0.93 (0.85 - 1.03) | 0.15 | 0.94 (0.84 - 1.06) | 0.35 |

### 2 Baseline patient characteristics across data sources

#### 2.1 GLP1RA (female) vs SGLT2I (female)

##### 2.1.1 CCAE

**Supplementary Table 157:** Baseline patient characteristics for GLP1RA (female) (T) and SGLT2I (female) (C) new-users in the CCAE data source. We report proportion of initiators satisfying selected base-line characteristics and the standardized difference of population proportions (StdDiff) before and after propensity score adjustment. Less extreme StdDiffs through matching and stratification suggest improved balance between patient cohorts through adjustment.

| | Before adjustment | | | After matching | | | After stratification | | |
|---|---|---|---|---|---|---|---|---|---|
| Characteristic | T (%) | C (%) | StdDiff | T (%) | C (%) | StdDiff | T (%) | C (%) | StdDiff |
| **Age group** | | | | | | | | | |
| 15 - 19 | 0.2 | 0.1 | 0.02 | 0.2 | 0.2 | -0.01 | 0.2 | 0.2 | -0.01 |
| 20 - 24 | 0.9 | 0.6 | 0.04 | 0.8 | 0.8 | 0.00 | 0.8 | 0.8 | 0.00 |
| 25 - 29 | 1.4 | 0.8 | 0.05 | 1.2 | 1.0 | 0.02 | 1.1 | 1.0 | 0.02 |
| 30 - 34 | 3.7 | 2.1 | 0.10 | 2.9 | 2.7 | 0.02 | 2.9 | 2.6 | 0.01 |
| 35 - 39 | 7.1 | 4.8 | 0.10 | 5.8 | 5.6 | 0.00 | 5.7 | 5.5 | 0.01 |
| 45 - 49 | 16.2 | 14.9 | 0.03 | 15.8 | 15.5 | 0.01 | 15.8 | 15.5 | 0.01 |
| 50 - 54 | 20.8 | 20.3 | 0.01 | 21.2 | 20.8 | 0.01 | 21.0 | 20.6 | 0.01 |
| 55 - 59 | 21.0 | 24.3 | -0.08 | 22.2 | 23.0 | -0.02 | 22.2 | 23.3 | -0.03 |
| 60 - 64 | 16.4 | 21.5 | -0.13 | 18.8 | 18.6 | 0.00 | 19.4 | 19.4 | 0.00 |
| 65 - 69 | 1.0 | 1.6 | -0.05 | 1.2 | 1.5 | -0.03 | 1.2 | 1.5 | -0.02 |
| **Medical history: General** | | | | | | | | | |
| Acute respiratory disease | 35.7 | 33.9 | 0.04 | 34.6 | 34.5 | 0.00 | 34.5 | 34.7 | 0.00 |
| Attention deficit hyperactivity disorder | 2.0 | 1.3 | 0.05 | 2.0 | 1.8 | 0.02 | 1.9 | 1.7 | 0.02 |
| Chronic liver disease | 2.4 | 2.2 | 0.01 | 1.7 | 1.5 | 0.02 | 2.1 | 1.8 | 0.02 |
| Chronic obstructive lung disease | 2.5 | 2.6 | -0.01 | 2.4 | 2.7 | -0.02 | 2.5 | 2.7 | -0.01 |
| Crohn's disease | 0.3 | 0.3 | 0.00 | 0.3 | 0.4 | -0.01 | 0.3 | 0.4 | -0.01 |
| Dementia | 0.1 | 0.2 | -0.02 | 0.1 | 0.2 | -0.02 | 0.1 | 0.2 | -0.02 |
| Depressive disorder | 17.8 | 15.2 | 0.07 | 17.6 | 18.0 | -0.01 | 17.7 | 17.6 | 0.00 |
| Gastroesophageal reflux disease | 15.1 | 15.6 | -0.01 | 16.3 | 16.7 | -0.01 | 16.3 | 16.5 | 0.00 |
| Gastrointestinal hemorrhage | 2.0 | 1.6 | 0.03 | 1.6 | 1.7 | -0.01 | 1.7 | 1.8 | -0.01 |
| Human immunodeficiency virus infection | 0.1 | 0.1 | 0.00 | 0.2 | 0.1 | 0.01 | 0.2 | 0.2 | 0.01 |
| Hyperlipidemia | 61.7 | 69.6 | -0.17 | 67.8 | 67.3 | 0.01 | 67.9 | 67.8 | 0.00 |
| Hypertensive disorder | 61.8 | 69.0 | -0.15 | 67.3 | 67.2 | 0.00 | 68.0 | 67.7 | 0.01 |
| Lesion of liver | 0.6 | 0.5 | 0.01 | 0.7 | 0.6 | 0.02 | 0.6 | 0.6 | 0.01 |
| Obesity | 35.5 | 30.9 | 0.10 | 37.6 | 39.3 | -0.04 | 36.8 | 37.7 | -0.02 |
| Osteoarthritis | 22.3 | 20.3 | 0.05 | 22.0 | 21.9 | 0.00 | 21.8 | 21.7 | 0.00 |
| Pneumonia | 2.4 | 2.2 | 0.01 | 2.5 | 2.4 | 0.00 | 2.4 | 2.4 | 0.00 |
| Psoriasis | 2.0 | 1.7 | 0.02 | 2.0 | 1.8 | 0.02 | 2.1 | 1.8 | 0.02 |
| Renal impairment | 2.6 | 2.7 | -0.01 | 3.1 | 3.0 | 0.00 | 3.0 | 2.9 | 0.00 |
| Rheumatoid arthritis | 1.6 | 1.6 | 0.00 | 1.5 | 1.7 | -0.01 | 1.5 | 1.6 | -0.01 |
| Schizophrenia | 0.1 | 0.1 | -0.01 | 0.1 | 0.1 | -0.02 | 0.1 | 0.1 | -0.02 |
| Ulcerative colitis | 0.3 | 0.3 | 0.00 | 0.3 | 0.3 | 0.00 | 0.3 | 0.3 | 0.00 |
| Urinary tract infectious disease | 11.0 | 10.3 | 0.02 | 9.9 | 10.5 | -0.02 | 10.2 | 10.6 | -0.01 |
| Viral hepatitis C | 0.2 | 0.2 | 0.00 | 0.2 | 0.2 | 0.00 | 0.2 | 0.2 | 0.01 |
| Visual system disorder | 25.8 | 26.0 | 0.00 | 26.7 | 26.5 | 0.00 | 26.7 | 26.4 | 0.01 |
| **Medical history: Cardiovascular disease** | | | | | | | | | |
| Atrial fibrillation | 1.1 | 1.2 | -0.01 | 1.2 | 1.2 | 0.00 | 1.4 | 1.2 | 0.01 |
| Cerebrovascular disease | 1.9 | 2.2 | -0.02 | 1.8 | 2.1 | -0.02 | 1.9 | 2.1 | -0.02 |
| Coronary arteriosclerosis | 3.7 | 4.2 | -0.03 | 3.6 | 3.8 | -0.01 | 3.8 | 3.9 | -0.01 |
| Heart disease | 13.5 | 14.0 | -0.01 | 13.3 | 13.4 | 0.00 | 13.7 | 13.8 | 0.00 |
| Heart failure | 1.5 | 1.9 | -0.03 | 1.5 | 1.8 | -0.02 | 1.5 | 1.9 | -0.02 |
| Ischemic heart disease | 2.7 | 2.8 | -0.01 | 2.5 | 2.7 | -0.01 | 2.6 | 2.7 | 0.00 |
| Peripheral vascular disease | 3.2 | 3.0 | 0.01 | 3.4 | 3.2 | 0.01 | 3.3 | 3.1 | 0.01 |
| Pulmonary embolism | 0.5 | 0.3 | 0.02 | 0.5 | 0.4 | 0.02 | 0.5 | 0.3 | 0.02 |
| Venous thrombosis | 1.0 | 0.8 | 0.02 | 0.9 | 0.8 | 0.02 | 1.0 | 0.8 | 0.02 |
| **Medical history: Neoplasms** | | | | | | | | | |
| Hematologic neoplasm | 0.6 | 0.7 | -0.02 | 0.6 | 0.7 | -0.01 | 0.6 | 0.7 | -0.01 |
| Malignant lymphoma | 0.3 | 0.3 | -0.01 | 0.3 | 0.3 | 0.01 | 0.3 | 0.3 | 0.00 |
| Malignant neoplasm of anorectum | 0.1 | 0.1 | 0.00 | 0.1 | 0.1 | -0.01 | 0.1 | 0.1 | -0.01 |
| Malignant neoplastic disease | 6.0 | 6.3 | -0.02 | 6.2 | 6.3 | 0.00 | 6.2 | 6.3 | -0.01 |
| Malignant tumor of breast | 1.9 | 2.5 | -0.04 | 2.1 | 2.3 | -0.02 | 2.2 | 2.4 | -0.01 |

| (Continued) | Before ajustment | | | After matching | | | After stratification | | |
|---|---|---|---|---|---|---|---|---|---|
| Characteristic | T (%) | C (%) | StdDiff | T (%) | C (%) | StdDiff | T (%) | C (%) | StdDiff |
| Malignant tumor of colon | 0.2 | 0.2 | 0.00 | 0.2 | 0.2 | 0.01 | 0.2 | 0.2 | 0.01 |
| Malignant tumor of lung | 0.1 | 0.1 | -0.03 | 0.1 | 0.1 | -0.02 | 0.1 | 0.1 | -0.02 |
| Malignant tumor of urinary bladder | 0.1 | 0.1 | 0.01 | 0.1 | 0.1 | 0.00 | 0.1 | 0.1 | 0.01 |
| **Medication use** | | | | | | | | | |
| Agents acting on the renin-angiotensin system | 57.2 | 61.6 | -0.09 | 59.4 | 58.8 | 0.01 | 60.0 | 59.5 | 0.01 |
| Antibacterials for systemic use | 68.6 | 64.9 | 0.08 | 65.5 | 65.8 | -0.01 | 65.8 | 65.9 | 0.00 |
| Antidepressants | 45.1 | 37.5 | 0.15 | 41.1 | 41.9 | -0.01 | 40.7 | 41.1 | -0.01 |
| Antiepileptics | 18.7 | 16.1 | 0.07 | 17.4 | 17.9 | -0.01 | 17.6 | 17.6 | 0.00 |
| Antiinflammatory and antirheumatic products | 40.0 | 38.2 | 0.04 | 39.8 | 39.8 | 0.00 | 39.1 | 39.7 | -0.01 |
| Antineoplastic agents | 5.7 | 4.4 | 0.06 | 4.5 | 4.8 | -0.01 | 4.7 | 4.8 | 0.00 |
| Antipsoriatics | 0.8 | 0.6 | 0.02 | 0.7 | 0.6 | 0.02 | 0.8 | 0.6 | 0.02 |
| Antithrombotic agents | 7.2 | 7.8 | -0.02 | 7.5 | 7.6 | 0.00 | 7.5 | 7.7 | -0.01 |
| Beta blocking agents | 22.6 | 24.3 | -0.04 | 23.6 | 23.6 | 0.00 | 23.6 | 23.6 | 0.00 |
| Calcium channel blockers | 16.6 | 18.2 | -0.04 | 17.5 | 17.6 | 0.00 | 17.8 | 17.7 | 0.00 |
| Diuretics | 44.5 | 42.7 | 0.04 | 42.4 | 42.3 | 0.00 | 42.4 | 42.6 | 0.00 |
| Drugs for acid related disorders | 29.2 | 27.5 | 0.04 | 28.0 | 28.1 | 0.00 | 28.5 | 28.3 | 0.01 |
| Drugs for obstructive airway diseases | 46.5 | 44.0 | 0.05 | 45.7 | 46.4 | -0.01 | 45.6 | 46.0 | -0.01 |
| Immunosuppressants | 3.6 | 3.3 | 0.02 | 3.5 | 3.6 | 0.00 | 3.5 | 3.6 | 0.00 |
| Lipid modifying agents | 57.1 | 62.4 | -0.11 | 60.6 | 60.0 | 0.01 | 60.5 | 60.3 | 0.00 |
| Opioids | 30.0 | 27.6 | 0.05 | 27.7 | 28.3 | -0.01 | 28.1 | 28.3 | 0.00 |
| Psycholeptics | 31.1 | 26.6 | 0.10 | 28.4 | 28.8 | -0.01 | 28.4 | 28.8 | -0.01 |
| Psychostimulants | 6.0 | 4.2 | 0.08 | 5.2 | 5.0 | 0.01 | 5.1 | 4.9 | 0.01 |

##### 2.1.2 ODOD

**Supplementary Table 158:** Baseline patient characteristics for GLP1RA (female) (T) and SGLT2I (female) (C) new-users in the ODOD data source. We report proportion of initiators satisfying selected base-line characteristics and the standardized difference of population proportions (StdDiff) before and after propensity score adjustment. Less extreme StdDiffs through matching and stratification suggest improved balance between patient cohorts through adjustment.

| | Before adjustment | | | After matching | | | After stratification | | |
|---|---|---|---|---|---|---|---|---|---|
| Characteristic | T (%) | C (%) | StdDiff | T (%) | C (%) | StdDiff | T (%) | C (%) | StdDiff |
| **Age group** | | | | | | | | | |
| 20 - 24 | 0.6 | 0.2 | 0.06 | 0.4 | 0.3 | 0.01 | 0.4 | 0.3 | 0.01 |
| 25 - 29 | 1.4 | 0.4 | 0.10 | 0.9 | 0.6 | 0.03 | 0.9 | 0.6 | 0.03 |
| 30 - 34 | 2.7 | 1.3 | 0.10 | 1.9 | 1.6 | 0.02 | 2.0 | 1.6 | 0.03 |
| 35 - 39 | 5.1 | 3.1 | 0.10 | 3.7 | 3.6 | 0.01 | 3.7 | 3.6 | 0.01 |
| 40 - 44 | 8.1 | 5.4 | 0.11 | 6.2 | 6.2 | 0.00 | 6.4 | 6.2 | 0.01 |
| 45 - 49 | 11.3 | 8.9 | 0.08 | 10.0 | 9.8 | 0.01 | 9.8 | 9.7 | 0.00 |
| 50 - 54 | 14.4 | 11.7 | 0.08 | 12.6 | 12.6 | 0.00 | 12.8 | 12.6 | 0.00 |
| 55 - 59 | 15.1 | 14.5 | 0.02 | 15.1 | 14.8 | 0.01 | 14.3 | 14.7 | -0.01 |
| 60 - 64 | 13.4 | 13.6 | -0.01 | 14.4 | 13.9 | 0.02 | 13.9 | 13.8 | 0.00 |
| 65 - 69 | 13.1 | 16.5 | -0.10 | 15.5 | 16.6 | -0.03 | 15.1 | 15.9 | -0.02 |
| 70 - 74 | 9.4 | 14.2 | -0.15 | 12.1 | 12.1 | 0.00 | 12.8 | 12.3 | 0.01 |
| 75 - 79 | 3.9 | 6.6 | -0.12 | 5.3 | 5.4 | -0.01 | 5.6 | 5.6 | 0.00 |
| 80 - 84 | 0.9 | 2.5 | -0.12 | 1.4 | 1.6 | -0.01 | 1.6 | 1.9 | -0.03 |
| 85 - 89 | 0.4 | 1.0 | -0.07 | 0.6 | 0.8 | -0.03 | 0.6 | 0.8 | -0.03 |
| **Medical history: General** | | | | | | | | | |
| Acute respiratory disease | 33.5 | 30.4 | 0.07 | 31.3 | 31.8 | -0.01 | 31.3 | 31.8 | -0.01 |
| Attention deficit hyperactivity disorder | 1.8 | 1.3 | 0.04 | 1.3 | 1.6 | -0.03 | 1.5 | 1.6 | -0.01 |
| Chronic liver disease | 2.4 | 2.2 | 0.01 | 1.9 | 1.7 | 0.02 | 2.1 | 2.0 | 0.00 |
| Chronic obstructive lung disease | 7.5 | 8.2 | -0.03 | 8.2 | 8.1 | 0.00 | 8.2 | 8.1 | 0.00 |
| Crohn’s disease | 0.3 | 0.3 | 0.00 | 0.3 | 0.3 | 0.00 | 0.3 | 0.3 | 0.00 |
| Dementia | 0.9 | 1.0 | -0.01 | 1.0 | 0.9 | 0.01 | 1.1 | 0.9 | 0.02 |
| Depressive disorder | 26.0 | 21.8 | 0.10 | 23.6 | 25.6 | -0.05 | 23.8 | 25.0 | -0.03 |
| Gastroesophageal reflux disease | 23.2 | 22.9 | 0.00 | 23.0 | 23.7 | -0.02 | 23.1 | 23.9 | -0.02 |
| Gastrointestinal hemorrhage | 2.1 | 2.2 | -0.01 | 2.0 | 2.2 | -0.01 | 2.2 | 2.3 | -0.01 |
| Human immunodeficiency virus infection | 0.2 | 0.1 | 0.03 | 0.2 | 0.1 | 0.01 | 0.2 | 0.1 | 0.01 |
| Hyperlipidemia | 74.3 | 78.3 | -0.10 | 77.0 | 75.7 | 0.03 | 77.1 | 76.3 | 0.02 |
| Hypertensive disorder | 72.7 | 77.5 | -0.11 | 75.3 | 74.9 | 0.01 | 75.5 | 75.5 | 0.00 |
| Lesion of liver | 1.3 | 1.1 | 0.02 | 1.3 | 1.2 | 0.01 | 1.3 | 1.2 | 0.01 |
| Obesity | 46.8 | 34.9 | 0.24 | 40.9 | 43.2 | -0.05 | 40.5 | 42.2 | -0.04 |
| Osteoarthritis | 31.7 | 30.1 | 0.03 | 31.3 | 32.3 | -0.02 | 31.4 | 32.1 | -0.01 |
| Pneumonia | 3.5 | 3.5 | 0.00 | 3.4 | 3.6 | -0.01 | 3.6 | 3.7 | 0.00 |
| Psoriasis | 2.4 | 1.9 | 0.03 | 2.3 | 2.1 | 0.02 | 2.4 | 2.0 | 0.02 |
| Renal impairment | 9.0 | 10.3 | -0.04 | 10.4 | 10.2 | 0.01 | 10.7 | 10.2 | 0.02 |
| Rheumatoid arthritis | 2.8 | 2.6 | 0.01 | 2.8 | 2.9 | -0.01 | 2.8 | 2.9 | -0.01 |
| Schizophrenia | 0.2 | 0.3 | -0.01 | 0.2 | 0.4 | -0.03 | 0.2 | 0.3 | -0.02 |
| Urinary tract infectious disease | 12.8 | 12.1 | 0.02 | 12.2 | 12.1 | 0.00 | 12.3 | 12.1 | 0.00 |
| Viral hepatitis C | 0.4 | 0.3 | 0.02 | 0.5 | 0.4 | 0.01 | 0.4 | 0.4 | 0.01 |
| Visual system disorder | 36.9 | 40.7 | -0.08 | 39.1 | 38.3 | 0.01 | 39.5 | 39.1 | 0.01 |
| **Medical history: Cardiovascular disease** | | | | | | | | | |
| Atrial fibrillation | 3.3 | 4.4 | -0.06 | 3.9 | 3.5 | 0.02 | 4.3 | 3.8 | 0.02 |
| Cerebrovascular disease | 4.0 | 5.4 | -0.07 | 4.8 | 4.6 | 0.01 | 4.8 | 4.9 | -0.01 |
| Coronary arteriosclerosis | 7.7 | 11.1 | -0.12 | 9.3 | 8.7 | 0.02 | 10.1 | 9.7 | 0.01 |
| Heart disease | 21.5 | 25.7 | -0.10 | 23.9 | 22.7 | 0.03 | 25.0 | 23.9 | 0.03 |
| Heart failure | 4.7 | 6.6 | -0.08 | 5.5 | 5.2 | 0.01 | 6.1 | 5.8 | 0.01 |
| Ischemic heart disease | 4.7 | 6.7 | -0.09 | 5.4 | 5.2 | 0.01 | 5.9 | 5.8 | 0.00 |
| Peripheral vascular disease | 7.8 | 9.8 | -0.07 | 8.8 | 8.8 | 0.00 | 9.1 | 9.0 | 0.00 |
| Pulmonary embolism | 0.8 | 0.8 | 0.00 | 0.8 | 0.7 | 0.01 | 0.9 | 0.8 | 0.01 |
| Venous thrombosis | 1.2 | 1.3 | -0.01 | 1.3 | 1.3 | 0.00 | 1.4 | 1.3 | 0.01 |
| **Medical history: Neoplasms** | | | | | | | | | |
| Hematologic neoplasm | 0.8 | 0.8 | 0.00 | 0.8 | 0.8 | 0.01 | 0.8 | 0.8 | 0.01 |
| Malignant lymphoma | 0.4 | 0.4 | 0.00 | 0.5 | 0.4 | 0.02 | 0.5 | 0.4 | 0.02 |
| Malignant neoplasm of anorectum | 0.1 | 0.2 | -0.01 | 0.2 | 0.2 | 0.00 | 0.2 | 0.2 | 0.00 |
| Malignant neoplastic disease | 8.0 | 8.8 | -0.03 | 8.4 | 8.5 | 0.00 | 8.4 | 8.5 | 0.00 |
| Malignant tumor of breast | 2.9 | 3.2 | -0.02 | 3.3 | 3.1 | 0.01 | 3.2 | 3.1 | 0.01 |
| Malignant tumor of colon | 0.4 | 0.3 | 0.00 | 0.4 | 0.3 | 0.01 | 0.4 | 0.3 | 0.01 |
| Malignant tumor of lung | 0.2 | 0.3 | -0.04 | 0.2 | 0.3 | -0.02 | 0.2 | 0.4 | -0.04 |
| Malignant tumor of urinary bladder | 0.1 | 0.1 | -0.01 | 0.1 | 0.1 | 0.00 | 0.1 | 0.1 | -0.01 |

| (Continued) | Before ajustment | | | After matching | | | After stratification | | |
|---|---|---|---|---|---|---|---|---|---|
| Characteristic | T (%) | C (%) | StdDiff | T (%) | C (%) | StdDiff | T (%) | C (%) | StdDiff |
| **Medication use** | | | | | | | | | |
| Agents acting on the renin-angiotensin system | 60.3 | 67.5 | -0.15 | 64.9 | 64.4 | 0.01 | 64.9 | 64.4 | 0.01 |
| Antibacterials for systemic use | 66.0 | 63.5 | 0.05 | 63.5 | 64.4 | -0.02 | 63.8 | 64.8 | -0.02 |
| Antidepressants | 47.7 | 39.3 | 0.17 | 43.4 | 45.2 | -0.04 | 43.1 | 43.8 | -0.01 |
| Antiepileptics | 25.6 | 20.6 | 0.12 | 23.6 | 24.2 | -0.01 | 23.3 | 23.8 | -0.01 |
| Antiinflammatory and antirheumatic products | 39.7 | 37.0 | 0.06 | 38.1 | 39.2 | -0.02 | 38.1 | 39.2 | -0.02 |
| Antineoplastic agents | 6.2 | 5.4 | 0.03 | 5.9 | 5.7 | 0.01 | 6.0 | 5.7 | 0.02 |
| Antipsoriatics | 0.7 | 0.6 | 0.02 | 0.7 | 0.6 | 0.01 | 0.7 | 0.6 | 0.01 |
| Antithrombotic agents | 10.5 | 13.9 | -0.10 | 12.1 | 11.9 | 0.01 | 12.7 | 12.9 | -0.01 |
| Beta blocking agents | 28.7 | 33.0 | -0.10 | 30.8 | 30.3 | 0.01 | 31.2 | 31.2 | 0.00 |
| Calcium channel blockers | 20.5 | 24.4 | -0.09 | 23.1 | 22.7 | 0.01 | 23.3 | 23.1 | 0.00 |
| Diuretics | 46.0 | 45.9 | 0.00 | 46.3 | 45.6 | 0.01 | 46.0 | 45.8 | 0.00 |
| Drugs for acid related disorders | 31.9 | 31.0 | 0.02 | 32.0 | 32.0 | 0.00 | 31.6 | 31.9 | -0.01 |
| Drugs for obstructive airway diseases | 49.2 | 45.0 | 0.08 | 47.5 | 47.7 | -0.01 | 47.3 | 47.6 | 0.00 |
| Immunosuppressants | 4.0 | 3.8 | 0.01 | 3.9 | 4.2 | -0.01 | 4.0 | 4.2 | -0.01 |
| Lipid modifying agents | 64.0 | 72.2 | -0.18 | 69.8 | 69.1 | 0.01 | 69.8 | 69.5 | 0.01 |
| Opioids | 31.6 | 29.1 | 0.05 | 29.7 | 30.3 | -0.01 | 30.0 | 30.7 | -0.01 |
| Psycholeptics | 32.8 | 28.6 | 0.09 | 30.6 | 30.9 | -0.01 | 30.4 | 31.2 | -0.02 |
| Psychostimulants | 4.6 | 2.9 | 0.09 | 3.4 | 3.4 | 0.00 | 3.7 | 3.4 | 0.01 |

##### 2.1.3 MDCR

**Supplementary Table 159:** Baseline patient characteristics for GLP1RA (female) (T) and SGLT2I (female) (C) new-users in the MDCR data source. We report proportion of initiators satisfying selected base-line characteristics and the standardized difference of population proportions (StdDiff) before and after propensity score adjustment. Less extreme StdDiffs through matching and stratification suggest improved balance between patient cohorts through adjustment.

| | Before adjustment | | | After matching | | | After stratification | | |
|---|---|---|---|---|---|---|---|---|---|
| Characteristic | T (%) | C (%) | StdDiff | T (%) | C (%) | StdDiff | T (%) | C (%) | StdDiff |
| **Age group** | | | | | | | | | |
| 50 - 54 | 0.5 | <0.3 | 0.05 | <0.7 | <0.3 | -0.01 | <0.5 | <0.3 | -0.01 |
| 55 - 59 | 1.1 | 0.5 | 0.07 | <0.7 | 0.7 | -0.06 | <0.5 | 0.6 | -0.05 |
| 60 - 64 | 2.7 | 1.4 | 0.09 | 2.4 | 1.7 | 0.06 | 2.0 | 1.5 | 0.04 |
| 65 - 69 | 43.2 | 40.8 | 0.05 | 42.1 | 40.4 | 0.03 | 42.8 | 40.8 | 0.04 |
| 70 - 74 | 34.8 | 32.6 | 0.05 | 35.1 | 31.9 | 0.07 | 34.8 | 32.5 | 0.05 |
| 75 - 79 | 12.6 | 16.7 | -0.12 | 13.0 | 17.8 | -0.13 | 13.2 | 16.9 | -0.10 |
| 80 - 84 | 4.1 | 5.3 | -0.06 | 5.3 | 5.1 | 0.01 | 5.0 | 5.3 | -0.01 |
| 85 - 89 | 0.8 | 1.9 | -0.09 | 1.2 | 1.6 | -0.03 | 1.3 | 1.7 | -0.03 |
| 90 - 94 | <0.4 | 0.5 | -0.04 | <0.7 | 0.5 | -0.01 | 0.5 | 0.4 | 0.01 |
| **Medical history: General** | | | | | | | | | |
| Acute respiratory disease | 24.6 | 25.6 | -0.02 | 25.0 | 27.5 | -0.06 | 23.6 | 26.6 | -0.07 |
| Attention deficit hyperactivity disorder | 0.5 | <0.3 | 0.08 | <0.7 | <0.3 | 0.07 | <0.5 | <0.3 | 0.08 |
| Chronic liver disease | 2.2 | 2.6 | -0.03 | 1.6 | 2.6 | -0.07 | 1.4 | 2.6 | -0.08 |
| Chronic obstructive lung disease | 9.7 | 9.8 | 0.00 | 8.4 | 11.3 | -0.10 | 7.5 | 10.7 | -0.11 |
| Crohn’s disease | <0.4 | <0.3 | 0.02 | <0.7 | <0.3 | 0.04 | <0.5 | <0.3 | 0.01 |
| Dementia | 1.2 | 1.2 | 0.00 | 1.6 | 1.1 | 0.04 | 1.8 | 1.2 | 0.05 |
| Depressive disorder | 17.7 | 13.6 | 0.11 | 19.0 | 19.3 | -0.01 | 16.7 | 16.4 | 0.01 |
| Gastroesophageal reflux disease | 17.5 | 20.5 | -0.07 | 22.0 | 23.2 | -0.03 | 20.8 | 21.7 | -0.02 |
| Gastrointestinal hemorrhage | 2.6 | 3.0 | -0.02 | 2.6 | 3.3 | -0.04 | 2.4 | 3.2 | -0.04 |
| Hyperlipidemia | 64.1 | 79.0 | -0.33 | 79.6 | 79.5 | 0.00 | 79.0 | 79.3 | -0.01 |
| Hypertensive disorder | 72.9 | 83.2 | -0.25 | 80.3 | 82.9 | -0.07 | 81.3 | 83.4 | -0.05 |
| Lesion of liver | 0.6 | 1.2 | -0.06 | 0.8 | 1.2 | -0.04 | 0.7 | 1.2 | -0.05 |
| Obesity | 29.3 | 26.0 | 0.07 | 35.7 | 39.3 | -0.07 | 28.8 | 32.6 | -0.08 |
| Osteoarthritis | 40.9 | 35.9 | 0.10 | 42.5 | 42.7 | 0.00 | 38.7 | 38.6 | 0.00 |
| Pneumonia | 3.2 | 3.3 | -0.01 | 3.5 | 3.9 | -0.02 | 2.9 | 3.6 | -0.04 |
| Psoriasis | 2.3 | 2.5 | -0.01 | 2.6 | 2.9 | -0.02 | 2.1 | 2.6 | -0.04 |
| Renal impairment | 10.8 | 9.7 | 0.04 | 12.8 | 13.1 | -0.01 | 12.4 | 11.1 | 0.04 |
| Rheumatoid arthritis | 2.2 | 1.8 | 0.03 | 3.0 | 2.0 | 0.06 | 2.3 | 1.8 | 0.04 |
| Schizophrenia | <0.4 | <0.3 | 0.03 | <0.7 | <0.3 | 0.02 | <0.5 | <0.3 | 0.01 |
| Ulcerative colitis | <0.4 | 0.6 | -0.06 | <0.7 | 0.5 | -0.01 | <0.5 | 0.5 | -0.04 |
| Urinary tract infectious disease | 13.7 | 11.6 | 0.07 | 13.2 | 12.3 | 0.03 | 12.6 | 12.0 | 0.02 |
| Visual system disorder | 54.7 | 56.6 | -0.04 | 58.0 | 58.4 | -0.01 | 56.9 | 57.6 | -0.01 |
| **Medical history: Cardiovascular disease** | | | | | | | | | |
| Atrial fibrillation | 6.2 | 7.7 | -0.06 | 6.5 | 7.7 | -0.05 | 6.3 | 7.7 | -0.05 |
| Cerebrovascular disease | 8.9 | 10.6 | -0.06 | 9.8 | 10.5 | -0.03 | 9.5 | 10.7 | -0.04 |
| Coronary arteriosclerosis | 14.5 | 17.9 | -0.09 | 15.4 | 17.9 | -0.07 | 15.6 | 18.1 | -0.07 |
| Heart disease | 33.4 | 37.6 | -0.09 | 34.6 | 39.1 | -0.09 | 34.3 | 38.3 | -0.08 |
| Heart failure | 6.3 | 8.7 | -0.09 | 6.2 | 10.0 | -0.14 | 5.9 | 9.4 | -0.13 |
| Ischemic heart disease | 7.1 | 9.7 | -0.10 | 6.9 | 10.3 | -0.12 | 6.8 | 9.9 | -0.11 |
| Peripheral vascular disease | 10.3 | 12.2 | -0.06 | 11.3 | 13.1 | -0.06 | 10.8 | 12.5 | -0.06 |
| Pulmonary embolism | 0.4 | 0.8 | -0.05 | <0.7 | 1.0 | -0.07 | <0.5 | 0.9 | -0.07 |
| Venous thrombosis | 1.7 | 2.1 | -0.03 | 1.8 | 2.5 | -0.05 | 2.0 | 2.2 | -0.01 |
| **Medical history: Neoplasms** | | | | | | | | | |
| Hematologic neoplasm | 1.1 | 0.7 | 0.05 | 1.5 | 0.8 | 0.06 | 1.6 | 0.7 | 0.08 |
| Malignant lymphoma | 0.6 | 0.5 | 0.00 | 0.7 | 0.5 | 0.02 | 0.6 | 0.5 | 0.01 |
| Malignant neoplasm of anorectum | <0.4 | 0.3 | -0.04 | <0.7 | 0.4 | -0.02 | <0.5 | <0.3 | -0.02 |
| Malignant neoplastic disease | 13.2 | 14.0 | -0.02 | 13.7 | 14.3 | -0.02 | 13.3 | 14.0 | -0.02 |
| Malignant tumor of breast | 6.4 | 6.0 | 0.02 | 6.1 | 6.3 | -0.01 | 6.2 | 6.0 | 0.01 |
| Malignant tumor of colon | <0.4 | 0.7 | -0.06 | <0.7 | 0.8 | -0.08 | <0.5 | 0.7 | -0.07 |
| Malignant tumor of lung | <0.4 | 0.4 | -0.04 | <0.7 | <0.3 | -0.04 | <0.5 | 0.4 | -0.03 |
| Malignant tumor of urinary bladder | 0.9 | <0.3 | 0.08 | 1.2 | <0.3 | 0.13 | 1.1 | <0.3 | 0.11 |
| **Medication use** | | | | | | | | | |
| Agents acting on the renin-angiotensin system | 74.7 | 73.7 | 0.02 | 73.4 | 73.5 | 0.00 | 75.1 | 73.5 | 0.04 |
| Antibacterials for systemic use | 67.3 | 62.0 | 0.11 | 65.5 | 65.9 | -0.01 | 63.4 | 63.6 | 0.00 |
| Antidepressants | 44.7 | 34.1 | 0.22 | 41.8 | 44.0 | -0.04 | 37.0 | 38.7 | -0.04 |
| Antiepileptics | 21.9 | 19.5 | 0.06 | 21.6 | 23.3 | -0.04 | 20.0 | 21.2 | -0.03 |
| Antiinflammatory and antirheumatic products | 40.1 | 35.3 | 0.10 | 38.0 | 39.9 | -0.04 | 36.6 | 37.2 | -0.01 |

| (Continued) | Before ajustment | | | After matching | | | After stratification | | |
|---|---|---|---|---|---|---|---|---|---|
| Characteristic | T (%) | C (%) | StdDiff | T (%) | C (%) | StdDiff | T (%) | C (%) | StdDiff |
| Antineoplastic agents | 11.3 | 8.0 | 0.11 | 9.4 | 9.1 | 0.01 | 9.2 | 8.3 | 0.03 |
| Antipsoriatics | 1.1 | 1.1 | 0.01 | 1.1 | 1.0 | 0.01 | 0.9 | 1.1 | -0.02 |
| Antithrombotic agents | 17.8 | 18.3 | -0.01 | 17.1 | 19.5 | -0.06 | 16.7 | 18.8 | -0.06 |
| Beta blocking agents | 41.8 | 44.7 | -0.06 | 41.6 | 44.4 | -0.06 | 42.1 | 44.4 | -0.05 |
| Calcium channel blockers | 30.6 | 32.9 | -0.05 | 29.3 | 32.4 | -0.07 | 29.4 | 32.5 | -0.07 |
| Diuretics | 57.7 | 53.9 | 0.08 | 53.0 | 55.4 | -0.05 | 52.9 | 54.5 | -0.03 |
| Drugs for acid related disorders | 39.6 | 39.1 | 0.01 | 37.0 | 42.4 | -0.11 | 36.9 | 40.5 | -0.07 |
| Drugs for obstructive airway diseases | 53.5 | 49.7 | 0.07 | 51.6 | 53.5 | -0.04 | 49.0 | 51.5 | -0.05 |
| Immunosuppressants | 5.4 | 4.8 | 0.03 | 6.4 | 5.6 | 0.04 | 5.7 | 5.2 | 0.02 |
| Lipid modifying agents | 80.1 | 80.7 | -0.02 | 80.3 | 81.3 | -0.03 | 80.8 | 81.0 | -0.01 |
| Opioids | 30.5 | 27.2 | 0.07 | 26.8 | 32.3 | -0.12 | 24.9 | 29.5 | -0.10 |
| Psycholeptics | 31.4 | 27.2 | 0.09 | 27.3 | 31.8 | -0.10 | 26.4 | 29.4 | -0.07 |
| Psychostimulants | 2.4 | 2.0 | 0.03 | 1.9 | 3.0 | -0.07 | 2.1 | 2.5 | -0.03 |

##### 2.1.4 MDCD

**Supplementary Table 160:** Baseline patient characteristics for GLP1RA (female) (T) and SGLT2I (female) (C) new-users in the MDCD data source. We report proportion of initiators satisfying selected base-line characteristics and the standardized difference of population proportions (StdDiff) before and after propensity score adjustment. Less extreme StdDiffs through matching and stratification suggest improved balance between patient cohorts through adjustment.

| | Before adjustment | | | After matching | | | After stratification | | |
|---|---|---|---|---|---|---|---|---|---|
| Characteristic | T (%) | C (%) | StdDiff | T (%) | C (%) | StdDiff | T (%) | C (%) | StdDiff |
| **Age group** | | | | | | | | | |
| 15 - 19 | 1.7 | 0.6 | 0.11 | 1.6 | 0.8 | 0.07 | 1.4 | 0.7 | 0.06 |
| 20 - 24 | 1.8 | 1.0 | 0.06 | 1.5 | 1.4 | 0.01 | 1.3 | 1.3 | 0.01 |
| 25 - 29 | 4.4 | 3.9 | 0.02 | 3.7 | 4.8 | -0.05 | 3.9 | 4.4 | -0.03 |
| 30 - 34 | 8.8 | 7.2 | 0.06 | 8.2 | 8.5 | -0.01 | 7.6 | 8.0 | -0.01 |
| 35 - 39 | 13.3 | 10.7 | 0.08 | 13.3 | 11.5 | 0.06 | 12.5 | 11.4 | 0.04 |
| 40 - 44 | 15.2 | 14.0 | 0.04 | 15.8 | 14.4 | 0.04 | 15.4 | 14.5 | 0.03 |
| 45 - 49 | 15.6 | 14.4 | 0.03 | 15.8 | 15.0 | 0.02 | 15.7 | 14.5 | 0.03 |
| 50 - 54 | 14.4 | 15.9 | -0.04 | 13.9 | 15.2 | -0.04 | 14.3 | 15.4 | -0.03 |
| 55 - 59 | 12.4 | 16.1 | -0.10 | 13.4 | 14.4 | -0.03 | 14.2 | 15.2 | -0.03 |
| 60 - 64 | 8.7 | 11.8 | -0.10 | 9.4 | 10.5 | -0.04 | 9.6 | 10.9 | -0.04 |
| 65 - 69 | 2.4 | 2.4 | 0.00 | 2.4 | 2.0 | 0.03 | 2.7 | 2.1 | 0.04 |
| 70 - 74 | 0.6 | 1.2 | -0.06 | 0.6 | 1.0 | -0.04 | 0.8 | 1.1 | -0.03 |
| 75 - 79 | 0.3 | 0.2 | 0.02 | <0.3 | 0.2 | 0.01 | <0.3 | 0.2 | 0.00 |
| 80 - 84 | 0.3 | 0.3 | 0.00 | <0.3 | 0.2 | 0.00 | <0.3 | 0.2 | 0.00 |
| **Race** | | | | | | | | | |
| Black or African American | 27.2 | 26.2 | 0.02 | 27.0 | 27.0 | 0.00 | 27.7 | 26.3 | 0.03 |
| White | 61.2 | 60.6 | 0.01 | 60.8 | 59.9 | 0.02 | 60.2 | 60.4 | -0.01 |
| **Ethnicity** | | | | | | | | | |
| Hispanic or Latino | 3.6 | 2.6 | 0.06 | 4.0 | 3.0 | 0.06 | 3.9 | 2.8 | 0.06 |
| **Medical history: General** | | | | | | | | | |
| Acute respiratory disease | 44.3 | 41.1 | 0.06 | 42.7 | 42.7 | 0.00 | 42.7 | 42.7 | 0.00 |
| Attention deficit hyperactivity disorder | 4.2 | 2.3 | 0.11 | 3.9 | 3.0 | 0.05 | 3.8 | 2.8 | 0.05 |
| Chronic liver disease | 3.6 | 3.4 | 0.01 | 3.0 | 3.5 | -0.03 | 3.1 | 3.4 | -0.02 |
| Chronic obstructive lung disease | 17.8 | 18.3 | -0.01 | 17.2 | 17.9 | -0.02 | 17.9 | 17.9 | 0.00 |
| Crohn’s disease | 0.4 | 0.4 | 0.00 | 0.4 | 0.6 | -0.03 | 0.4 | 0.5 | -0.01 |
| Dementia | 1.0 | 0.9 | 0.01 | 1.0 | 0.9 | 0.01 | 1.3 | 0.9 | 0.03 |
| Depressive disorder | 43.6 | 40.1 | 0.07 | 42.2 | 43.3 | -0.02 | 41.3 | 42.1 | -0.02 |
| Gastroesophageal reflux disease | 29.8 | 30.7 | -0.02 | 29.7 | 31.2 | -0.03 | 29.7 | 31.4 | -0.04 |
| Gastrointestinal hemorrhage | 3.6 | 3.7 | 0.00 | 3.8 | 4.2 | -0.02 | 3.5 | 4.1 | -0.03 |
| Human immunodeficiency virus infection | 0.5 | 0.7 | -0.02 | <0.3 | 0.7 | -0.06 | 0.5 | 0.7 | -0.03 |
| Hyperlipidemia | 57.8 | 62.8 | -0.10 | 59.3 | 59.4 | 0.00 | 61.3 | 60.9 | 0.01 |
| Hypertensive disorder | 70.0 | 72.8 | -0.06 | 69.8 | 70.0 | 0.00 | 72.2 | 71.2 | 0.02 |
| Lesion of liver | 1.0 | 1.4 | -0.04 | 1.2 | 1.6 | -0.03 | 1.3 | 1.6 | -0.02 |
| Obesity | 58.2 | 47.3 | 0.22 | 54.4 | 60.4 | -0.12 | 49.3 | 54.9 | -0.11 |
| Osteoarthritis | 39.1 | 38.7 | 0.01 | 38.4 | 39.5 | -0.02 | 37.9 | 39.3 | -0.03 |
| Pneumonia | 5.8 | 5.5 | 0.01 | 5.3 | 5.9 | -0.03 | 5.0 | 5.8 | -0.03 |
| Psoriasis | 1.0 | 1.7 | -0.06 | 1.1 | 2.1 | -0.07 | 1.1 | 1.9 | -0.07 |
| Renal impairment | 5.7 | 5.9 | -0.01 | 5.7 | 6.0 | -0.01 | 6.0 | 6.0 | 0.00 |
| Rheumatoid arthritis | 2.3 | 2.3 | 0.00 | 2.0 | 2.4 | -0.03 | 2.0 | 2.3 | -0.02 |
| Schizophrenia | 2.1 | 2.7 | -0.04 | 1.8 | 3.1 | -0.09 | 1.8 | 3.1 | -0.08 |
| Ulcerative colitis | <0.2 | 0.2 | -0.03 | <0.3 | <0.2 | -0.03 | <0.3 | 0.2 | -0.05 |
| Urinary tract infectious disease | 15.8 | 13.3 | 0.07 | 14.3 | 14.8 | -0.02 | 14.0 | 14.5 | -0.01 |
| Viral hepatitis C | 1.7 | 1.6 | 0.01 | 1.8 | 1.6 | 0.01 | 1.7 | 1.6 | 0.01 |
| Visual system disorder | 43.5 | 46.0 | -0.05 | 43.1 | 45.3 | -0.04 | 44.7 | 46.0 | -0.03 |
| **Medical history: Cardiovascular disease** | | | | | | | | | |
| Atrial fibrillation | 2.0 | 2.1 | -0.01 | 1.9 | 1.9 | 0.00 | 2.0 | 2.0 | 0.00 |
| Cerebrovascular disease | 3.6 | 4.1 | -0.03 | 3.5 | 3.5 | 0.00 | 3.9 | 3.8 | 0.01 |
| Coronary arteriosclerosis | 6.9 | 8.5 | -0.06 | 7.1 | 7.4 | -0.01 | 7.1 | 7.4 | -0.01 |
| Heart disease | 23.3 | 23.8 | -0.01 | 22.2 | 23.4 | -0.03 | 23.1 | 23.7 | -0.01 |
| Heart failure | 6.8 | 6.5 | 0.01 | 6.2 | 6.7 | -0.02 | 6.1 | 6.7 | -0.03 |
| Ischemic heart disease | 4.7 | 5.9 | -0.05 | 4.7 | 5.4 | -0.03 | 5.3 | 5.6 | -0.01 |
| Peripheral vascular disease | 5.7 | 6.6 | -0.04 | 5.9 | 6.3 | -0.02 | 6.4 | 6.4 | 0.00 |
| Pulmonary embolism | 1.1 | 1.0 | 0.00 | 1.2 | 1.1 | 0.01 | 1.1 | 1.1 | 0.00 |
| Venous thrombosis | 1.7 | 1.4 | 0.03 | 1.5 | 1.4 | 0.01 | 1.5 | 1.4 | 0.01 |
| **Medical history: Neoplasms** | | | | | | | | | |
| Hematologic neoplasm | 0.6 | 0.8 | -0.02 | 0.6 | 0.6 | 0.00 | 0.6 | 0.7 | -0.01 |
| Malignant lymphoma | 0.2 | <0.2 | 0.02 | <0.3 | 0.2 | 0.00 | <0.3 | <0.2 | 0.02 |

| (Continued) | Before ajustment | | | After matching | | | After stratification | | |
|---|---|---|---|---|---|---|---|---|---|
| Characteristic | T (%) | C (%) | StdDiff | T (%) | C (%) | StdDiff | T (%) | C (%) | StdDiff |
| Malignant neoplastic disease | 4.6 | 6.0 | -0.06 | 4.7 | 5.8 | -0.05 | 4.9 | 6.0 | -0.04 |
| Malignant tumor of breast | 1.4 | 2.6 | -0.08 | 1.5 | 2.4 | -0.06 | 1.7 | 2.5 | -0.05 |
| Malignant tumor of lung | <0.2 | <0.2 | 0.02 | <0.3 | <0.2 | -0.02 | <0.3 | <0.2 | -0.02 |
| **Medication use** | | | | | | | | | |
| Agents acting on the renin-angiotensin system | 55.5 | 58.0 | -0.05 | 56.0 | 54.5 | 0.03 | 57.5 | 56.1 | 0.03 |
| Antibacterials for systemic use | 77.5 | 73.7 | 0.09 | 76.0 | 75.2 | 0.02 | 75.1 | 75.0 | 0.00 |
| Antidepressants | 64.1 | 59.3 | 0.10 | 62.4 | 60.6 | 0.04 | 61.3 | 60.5 | 0.02 |
| Antiepileptics | 47.0 | 44.4 | 0.05 | 46.9 | 46.1 | 0.02 | 45.6 | 45.6 | 0.00 |
| Antiinflammatory and antirheumatic products | 65.3 | 65.1 | 0.01 | 66.1 | 65.4 | 0.01 | 66.1 | 65.4 | 0.01 |
| Antineoplastic agents | 5.4 | 5.0 | 0.02 | 4.7 | 5.0 | -0.01 | 4.4 | 5.1 | -0.03 |
| Antipsoriatics | 0.9 | 0.7 | 0.03 | 0.9 | 0.7 | 0.01 | 0.9 | 0.7 | 0.02 |
| Antithrombotic agents | 18.6 | 21.3 | -0.07 | 19.4 | 20.2 | -0.02 | 20.5 | 20.8 | -0.01 |
| Beta blocking agents | 27.6 | 30.8 | -0.07 | 28.7 | 28.4 | 0.01 | 30.2 | 29.5 | 0.01 |
| Calcium channel blockers | 20.6 | 20.9 | -0.01 | 20.9 | 20.1 | 0.02 | 22.2 | 20.2 | 0.05 |
| Diuretics | 45.9 | 46.2 | -0.01 | 43.4 | 44.7 | -0.03 | 43.4 | 44.7 | -0.03 |
| Drugs for acid related disorders | 52.0 | 51.0 | 0.02 | 51.2 | 50.7 | 0.01 | 51.8 | 51.2 | 0.01 |
| Drugs for obstructive airway diseases | 66.4 | 64.1 | 0.05 | 64.2 | 65.7 | -0.03 | 63.7 | 65.2 | -0.03 |
| Immunosuppressants | 3.0 | 2.7 | 0.02 | 3.0 | 2.8 | 0.01 | 2.9 | 2.7 | 0.01 |
| Lipid modifying agents | 56.3 | 61.5 | -0.10 | 57.8 | 57.2 | 0.01 | 60.6 | 59.0 | 0.03 |
| Opioids | 49.7 | 47.9 | 0.04 | 46.9 | 49.6 | -0.05 | 46.3 | 48.8 | -0.05 |
| Psycholeptics | 54.4 | 51.3 | 0.06 | 52.3 | 52.9 | -0.01 | 51.0 | 52.8 | -0.04 |
| Psychostimulants | 9.4 | 6.2 | 0.12 | 8.6 | 7.4 | 0.04 | 8.1 | 7.0 | 0.04 |

##### 2.1.5 Open Claims

**Supplementary Table 161:** Baseline patient characteristics for GLP1RA (female) (T) and SGLT2I (female) (C) new-users in the Open Claims data source. We report proportion of initiators satisfying selected base-line characteristics and the standardized difference of population proportions (StdDiff) before and after propensity score adjustment. Less extreme StdDiffs through matching and stratification suggest improved balance between patient cohorts through adjustment.

| | Before adjustment | | | After matching | | | After stratification | | |
|---|---|---|---|---|---|---|---|---|---|
| Characteristic | T (%) | C (%) | StdDiff | T (%) | C (%) | StdDiff | T (%) | C (%) | StdDiff |
| **Age group** | | | | | | | | | |
| 15 - 19 | 0.2 | 0.1 | 0.02 | 0.2 | 0.2 | 0.01 | 0.2 | 0.2 | 0.01 |
| 20 - 24 | 0.9 | 0.5 | 0.05 | 0.7 | 0.7 | 0.00 | 0.7 | 0.7 | 0.00 |
| 25 - 29 | 1.7 | 1.0 | 0.06 | 1.3 | 1.2 | 0.00 | 1.3 | 1.3 | 0.00 |
| 30 - 34 | 3.1 | 1.9 | 0.08 | 2.4 | 2.4 | 0.00 | 2.5 | 2.4 | 0.00 |
| 35 - 39 | 5.4 | 3.6 | 0.09 | 4.3 | 4.3 | 0.00 | 4.5 | 4.4 | 0.00 |
| 40 - 44 | 8.5 | 6.3 | 0.08 | 7.2 | 7.4 | -0.01 | 7.2 | 7.3 | 0.00 |
| 45 - 49 | 12.0 | 9.8 | 0.07 | 10.9 | 11.0 | 0.00 | 10.7 | 10.8 | 0.00 |
| 50 - 54 | 15.5 | 13.9 | 0.04 | 14.8 | 15.0 | 0.00 | 14.5 | 14.7 | 0.00 |
| 55 - 59 | 16.4 | 16.6 | -0.01 | 17.0 | 16.7 | 0.01 | 16.7 | 16.4 | 0.01 |
| 60 - 64 | 14.5 | 16.3 | -0.05 | 15.8 | 15.7 | 0.00 | 15.4 | 15.5 | 0.00 |
| 65 - 69 | 10.8 | 12.8 | -0.06 | 12.1 | 12.2 | 0.00 | 11.9 | 12.0 | 0.00 |
| 70 - 74 | 6.5 | 8.7 | -0.08 | 7.8 | 7.6 | 0.01 | 7.9 | 7.7 | 0.01 |
| 75 - 79 | 3.0 | 5.2 | -0.11 | 3.7 | 3.7 | 0.00 | 4.2 | 4.2 | 0.00 |
| 80 - 84 | 1.3 | 2.9 | -0.11 | 1.8 | 1.8 | 0.00 | 2.1 | 2.2 | -0.01 |
| 85 - 89 | 0.1 | 0.2 | -0.03 | 0.1 | 0.2 | -0.01 | 0.1 | 0.2 | -0.01 |
| **Medical history: General** | | | | | | | | | |
| Acute respiratory disease | 16.8 | 16.3 | 0.01 | 15.9 | 16.1 | 0.00 | 16.5 | 16.7 | -0.01 |
| Attention deficit hyperactivity disorder | 1.2 | 0.6 | 0.06 | 0.8 | 0.8 | 0.00 | 0.9 | 0.9 | 0.00 |
| Chronic liver disease | 0.8 | 1.1 | -0.04 | 0.9 | 0.9 | 0.00 | 1.0 | 1.0 | 0.00 |
| Chronic obstructive lung disease | 4.1 | 4.5 | -0.02 | 4.3 | 4.4 | -0.01 | 4.4 | 4.5 | 0.00 |
| Crohn's disease | 0.2 | 0.2 | 0.00 | 0.2 | 0.2 | 0.00 | 0.2 | 0.2 | 0.00 |
| Dementia | 0.3 | 0.5 | -0.02 | 0.3 | 0.4 | 0.00 | 0.4 | 0.4 | 0.00 |
| Depressive disorder | 12.8 | 9.8 | 0.09 | 10.9 | 11.0 | 0.00 | 11.3 | 11.3 | 0.00 |
| Gastroesophageal reflux disease | 10.9 | 9.9 | 0.03 | 10.1 | 10.2 | 0.00 | 10.5 | 10.5 | 0.00 |
| Gastrointestinal hemorrhage | 1.1 | 1.2 | -0.01 | 1.2 | 1.2 | 0.00 | 1.2 | 1.2 | 0.00 |
| Human immunodeficiency virus infection | 0.2 | 0.2 | 0.01 | 0.2 | 0.2 | 0.00 | 0.2 | 0.2 | 0.00 |
| Hyperlipidemia | 37.9 | 42.7 | -0.10 | 39.1 | 39.4 | -0.01 | 40.3 | 40.8 | -0.01 |
| Hypertensive disorder | 44.9 | 48.2 | -0.07 | 45.4 | 45.6 | 0.00 | 46.6 | 47.0 | -0.01 |
| Lesion of liver | 0.5 | 0.6 | 0.00 | 0.5 | 0.6 | 0.00 | 0.5 | 0.6 | 0.00 |
| Obesity | 21.6 | 14.5 | 0.19 | 16.7 | 17.0 | -0.01 | 17.9 | 17.7 | 0.01 |
| Osteoarthritis | 17.2 | 15.6 | 0.04 | 15.8 | 16.2 | -0.01 | 16.4 | 16.6 | 0.00 |
| Pneumonia | 2.2 | 2.1 | 0.01 | 2.1 | 2.1 | 0.00 | 2.2 | 2.2 | 0.00 |
| Psoriasis | 1.4 | 1.1 | 0.03 | 1.2 | 1.2 | 0.00 | 1.2 | 1.2 | 0.00 |
| Renal impairment | 4.0 | 4.8 | -0.04 | 4.3 | 4.3 | 0.00 | 4.6 | 4.6 | 0.00 |
| Rheumatoid arthritis | 1.5 | 1.3 | 0.01 | 1.4 | 1.4 | 0.00 | 1.4 | 1.4 | 0.00 |
| Schizophrenia | 0.3 | 0.3 | 0.00 | 0.2 | 0.3 | 0.00 | 0.2 | 0.3 | 0.00 |
| Ulcerative colitis | 0.2 | 0.2 | 0.00 | 0.2 | 0.2 | 0.00 | 0.2 | 0.2 | 0.00 |
| Urinary tract infectious disease | 6.6 | 6.4 | 0.01 | 6.3 | 6.2 | 0.00 | 6.6 | 6.5 | 0.00 |
| Viral hepatitis C | 0.2 | 0.2 | -0.01 | 0.2 | 0.2 | 0.00 | 0.2 | 0.2 | 0.00 |
| Visual system disorder | 16.9 | 18.4 | -0.04 | 17.3 | 17.2 | 0.00 | 17.9 | 17.8 | 0.00 |
| **Medical history: Cardiovascular disease** | | | | | | | | | |
| Atrial fibrillation | 1.8 | 2.7 | -0.06 | 2.3 | 2.3 | 0.00 | 2.3 | 2.3 | 0.00 |
| Cerebrovascular disease | 1.8 | 2.4 | -0.04 | 2.0 | 1.9 | 0.00 | 2.2 | 2.1 | 0.00 |
| Coronary arteriosclerosis | 3.6 | 5.7 | -0.10 | 4.2 | 4.2 | 0.00 | 4.7 | 4.8 | 0.00 |
| Heart disease | 11.3 | 14.8 | -0.10 | 12.1 | 12.2 | 0.00 | 13.3 | 13.4 | 0.00 |
| Heart failure | 2.3 | 4.0 | -0.10 | 2.6 | 2.7 | -0.01 | 3.1 | 3.4 | -0.02 |
| Ischemic heart disease | 2.0 | 3.3 | -0.08 | 2.3 | 2.3 | 0.00 | 2.6 | 2.7 | -0.01 |
| Peripheral vascular disease | 3.2 | 3.8 | -0.04 | 3.3 | 3.4 | 0.00 | 3.5 | 3.6 | 0.00 |
| Pulmonary embolism | 0.5 | 0.5 | 0.01 | 0.5 | 0.5 | 0.01 | 0.5 | 0.5 | 0.01 |
| Venous thrombosis | 0.6 | 0.6 | 0.00 | 0.6 | 0.6 | 0.00 | 0.6 | 0.6 | 0.00 |
| **Medical history: Neoplasms** | | | | | | | | | |
| Hematologic neoplasm | 0.4 | 0.5 | -0.01 | 0.4 | 0.4 | 0.00 | 0.5 | 0.5 | 0.00 |
| Malignant lymphoma | 0.2 | 0.3 | -0.01 | 0.2 | 0.2 | 0.00 | 0.3 | 0.3 | 0.00 |
| Malignant neoplasm of anorectum | 0.1 | 0.1 | -0.01 | 0.1 | 0.1 | 0.00 | 0.1 | 0.1 | 0.00 |
| Malignant neoplastic disease | 4.4 | 5.0 | -0.03 | 4.5 | 4.5 | 0.00 | 4.8 | 4.7 | 0.00 |
| Malignant tumor of breast | 1.7 | 2.0 | -0.03 | 1.8 | 1.8 | 0.00 | 1.9 | 1.9 | 0.00 |
| Malignant tumor of colon | 0.1 | 0.2 | -0.01 | 0.2 | 0.2 | 0.00 | 0.2 | 0.2 | 0.00 |

| (Continued) | Before ajustment | | | After matching | | | After stratification | | |
|---|---|---|---|---|---|---|---|---|---|
| Characteristic | T (%) | C (%) | StdDiff | T (%) | C (%) | StdDiff | T (%) | C (%) | StdDiff |
| Malignant tumor of lung | 0.1 | 0.2 | -0.01 | 0.1 | 0.1 | 0.00 | 0.1 | 0.1 | 0.00 |
| Malignant tumor of urinary bladder | 0.1 | 0.1 | 0.00 | 0.1 | 0.1 | 0.00 | 0.1 | 0.1 | 0.00 |
| **Medication use** | | | | | | | | | |
| Agents acting on the renin-angiotensin system | 57.2 | 64.6 | -0.15 | 61.0 | 61.2 | 0.00 | 61.2 | 61.8 | -0.01 |
| Antibacterials for systemic use | 63.0 | 61.0 | 0.04 | 61.2 | 61.7 | -0.01 | 61.9 | 62.4 | -0.01 |
| Antidepressants | 47.3 | 38.3 | 0.18 | 42.4 | 42.9 | -0.01 | 42.7 | 42.9 | 0.00 |
| Antiepileptics | 26.4 | 21.9 | 0.11 | 23.8 | 24.1 | 0.00 | 24.2 | 24.3 | 0.00 |
| Antiinflammatory and antirheumatic products | 41.9 | 40.5 | 0.03 | 41.0 | 40.7 | 0.00 | 41.3 | 41.1 | 0.00 |
| Antineoplastic agents | 6.3 | 5.2 | 0.05 | 5.5 | 5.6 | 0.00 | 5.7 | 5.7 | 0.00 |
| Antipsoriatics | 2.1 | 1.5 | 0.04 | 1.7 | 1.7 | 0.00 | 1.7 | 1.7 | 0.00 |
| Beta blocking agents | 27.4 | 31.4 | -0.09 | 28.6 | 29.0 | -0.01 | 29.7 | 30.0 | -0.01 |
| Calcium channel blockers | 20.4 | 23.2 | -0.07 | 21.7 | 21.7 | 0.00 | 22.0 | 22.0 | 0.00 |
| Diuretics | 43.0 | 43.5 | -0.01 | 42.8 | 42.9 | 0.00 | 43.4 | 43.5 | 0.00 |
| Drugs for acid related disorders | 37.5 | 36.4 | 0.02 | 36.5 | 36.6 | 0.00 | 37.1 | 37.2 | 0.00 |
| Drugs for obstructive airway diseases | 46.8 | 43.2 | 0.07 | 44.6 | 44.9 | -0.01 | 45.0 | 45.3 | -0.01 |
| Immunosuppressants | 4.1 | 3.5 | 0.03 | 3.7 | 3.8 | 0.00 | 3.8 | 3.8 | 0.00 |
| Lipid modifying agents | 60.7 | 68.4 | -0.16 | 65.7 | 65.1 | 0.01 | 65.4 | 65.2 | 0.00 |
| Opioids | 26.9 | 25.7 | 0.03 | 25.8 | 25.9 | 0.00 | 26.6 | 26.4 | 0.00 |
| Psycholeptics | 30.3 | 26.4 | 0.09 | 27.6 | 28.0 | -0.01 | 28.4 | 28.4 | 0.00 |
| Psychostimulants | 5.6 | 3.5 | 0.10 | 4.2 | 4.2 | 0.00 | 4.5 | 4.4 | 0.00 |

##### 2.1.6 OEHR

**Supplementary Table 162:** Baseline patient characteristics for GLP1RA (female) (T) and SGLT2I (female) (C) new-users in the OEHR data source. We report proportion of initiators satisfying selected base-line characteristics and the standardized difference of population proportions (StdDiff) before and after propensity score adjustment. Less extreme StdDiffs through matching and stratification suggest improved balance between patient cohorts through adjustment.

| | Before adjustment | | | After matching | | | After stratification | | |
|---|---|---|---|---|---|---|---|---|---|
| Characteristic | T (%) | C (%) | StdDiff | T (%) | C (%) | StdDiff | T (%) | C (%) | StdDiff |
| **Age group** | | | | | | | | | |
| 15 - 19 | 0.2 | 0.1 | 0.02 | 0.2 | 0.2 | 0.02 | 0.2 | 0.1 | 0.02 |
| 20 - 24 | 1.0 | 0.6 | 0.04 | 0.9 | 0.7 | 0.02 | 0.9 | 0.7 | 0.02 |
| 25 - 29 | 2.1 | 1.2 | 0.07 | 1.5 | 1.4 | 0.01 | 1.6 | 1.4 | 0.02 |
| 30 - 34 | 3.8 | 2.3 | 0.08 | 3.1 | 2.9 | 0.01 | 3.2 | 3.0 | 0.01 |
| 35 - 39 | 7.1 | 4.7 | 0.10 | 5.8 | 5.6 | 0.01 | 5.6 | 5.6 | 0.00 |
| 40 - 44 | 10.0 | 7.8 | 0.08 | 8.5 | 8.6 | 0.00 | 8.6 | 8.6 | 0.00 |
| 45 - 49 | 13.7 | 11.8 | 0.06 | 13.2 | 12.8 | 0.01 | 12.7 | 12.7 | 0.00 |
| 50 - 54 | 16.3 | 16.4 | 0.00 | 16.4 | 16.3 | 0.00 | 16.6 | 16.2 | 0.01 |
| 55 - 59 | 16.6 | 18.1 | -0.04 | 17.4 | 17.6 | 0.00 | 17.4 | 17.6 | 0.00 |
| 60 - 64 | 14.6 | 16.5 | -0.05 | 15.8 | 15.8 | 0.00 | 15.5 | 15.8 | -0.01 |
| 65 - 69 | 8.1 | 10.1 | -0.07 | 9.1 | 9.2 | 0.00 | 9.3 | 9.3 | 0.00 |
| 70 - 74 | 4.3 | 5.9 | -0.07 | 5.2 | 5.3 | -0.01 | 5.2 | 5.3 | 0.00 |
| 75 - 79 | 1.6 | 2.9 | -0.09 | 2.0 | 2.2 | -0.01 | 2.3 | 2.4 | -0.01 |
| 80 - 84 | 0.5 | 1.2 | -0.07 | 0.6 | 0.8 | -0.03 | 0.6 | 0.9 | -0.03 |
| 85 - 89 | 0.1 | 0.4 | -0.05 | 0.2 | 0.4 | -0.05 | 0.2 | 0.4 | -0.04 |
| **Race** | | | | | | | | | |
| Asian | 1.5 | 2.6 | -0.08 | 1.7 | 1.9 | -0.01 | 1.9 | 2.2 | -0.02 |
| Black or African American | 14.5 | 13.8 | 0.02 | 14.5 | 14.4 | 0.00 | 14.5 | 14.2 | 0.01 |
| White | 76.9 | 75.5 | 0.03 | 76.0 | 75.9 | 0.00 | 76.0 | 75.9 | 0.00 |
| **Ethnicity** | | | | | | | | | |
| Hispanic or Latino | 5.9 | 6.8 | -0.04 | 6.0 | 6.1 | 0.00 | 6.1 | 6.3 | -0.01 |
| Not Hispanic or Latino | 85.9 | 85.1 | 0.02 | 85.5 | 85.4 | 0.00 | 85.7 | 85.5 | 0.01 |
| **Medical history: General** | | | | | | | | | |
| Acute respiratory disease | 22.7 | 23.8 | -0.03 | 22.5 | 22.7 | 0.00 | 23.3 | 23.4 | 0.00 |
| Attention deficit hyperactivity disorder | 1.4 | 1.1 | 0.03 | 1.1 | 1.4 | -0.02 | 1.1 | 1.4 | -0.02 |
| Chronic liver disease | 2.3 | 2.2 | 0.01 | 2.1 | 2.1 | 0.01 | 2.5 | 2.1 | 0.02 |
| Chronic obstructive lung disease | 4.1 | 4.6 | -0.03 | 4.3 | 4.5 | -0.01 | 4.4 | 4.6 | -0.01 |
| Crohn's disease | 0.2 | 0.2 | 0.00 | 0.2 | 0.3 | -0.01 | 0.2 | 0.3 | 0.00 |
| Dementia | 0.4 | 0.4 | 0.00 | 0.4 | 0.4 | 0.00 | 0.4 | 0.4 | 0.00 |
| Depressive disorder | 24.5 | 20.5 | 0.10 | 22.3 | 22.8 | -0.01 | 22.2 | 22.8 | -0.01 |
| Gastroesophageal reflux disease | 21.2 | 20.1 | 0.03 | 20.2 | 20.8 | -0.01 | 20.4 | 21.1 | -0.02 |
| Gastrointestinal hemorrhage | 1.3 | 1.1 | 0.01 | 1.2 | 1.1 | 0.01 | 1.2 | 1.1 | 0.01 |
| Human immunodeficiency virus infection | 0.1 | 0.1 | -0.01 | 0.1 | 0.1 | -0.01 | 0.1 | 0.1 | -0.02 |
| Hyperlipidemia | 58.4 | 63.0 | -0.10 | 60.3 | 59.2 | 0.02 | 61.4 | 60.6 | 0.02 |
| Hypertensive disorder | 59.0 | 63.6 | -0.10 | 60.8 | 60.5 | 0.01 | 62.0 | 61.7 | 0.01 |
| Lesion of liver | 0.6 | 0.8 | -0.02 | 0.6 | 0.9 | -0.03 | 0.6 | 0.8 | -0.03 |
| Obesity | 46.7 | 35.6 | 0.23 | 40.7 | 42.8 | -0.04 | 40.3 | 41.8 | -0.03 |
| Osteoarthritis | 18.5 | 17.3 | 0.03 | 17.7 | 18.0 | -0.01 | 18.1 | 18.1 | 0.00 |
| Pneumonia | 2.0 | 1.8 | 0.02 | 1.9 | 1.9 | 0.00 | 2.0 | 1.9 | 0.01 |
| Psoriasis | 2.0 | 1.7 | 0.02 | 1.9 | 1.8 | 0.01 | 1.9 | 1.8 | 0.01 |
| Renal impairment | 4.3 | 3.8 | 0.02 | 4.2 | 4.1 | 0.01 | 4.3 | 4.1 | 0.01 |
| Rheumatoid arthritis | 1.9 | 1.8 | 0.01 | 1.9 | 1.9 | 0.00 | 2.0 | 1.9 | 0.00 |
| Schizophrenia | 0.2 | 0.2 | 0.01 | 0.2 | 0.2 | 0.00 | 0.2 | 0.2 | 0.00 |
| Ulcerative colitis | 0.2 | 0.2 | 0.01 | 0.3 | 0.2 | 0.01 | 0.3 | 0.2 | 0.02 |
| Urinary tract infectious disease | 6.6 | 6.3 | 0.01 | 6.3 | 6.2 | 0.00 | 6.4 | 6.4 | 0.00 |
| Viral hepatitis C | 0.4 | 0.3 | 0.01 | 0.4 | 0.4 | 0.00 | 0.4 | 0.3 | 0.01 |
| Visual system disorder | 12.1 | 12.1 | 0.00 | 11.9 | 12.0 | 0.00 | 12.1 | 12.1 | 0.00 |
| **Medical history: Cardiovascular disease** | | | | | | | | | |
| Atrial fibrillation | 1.8 | 2.1 | -0.03 | 1.9 | 1.9 | 0.00 | 2.0 | 2.0 | 0.00 |
| Cerebrovascular disease | 1.7 | 2.0 | -0.03 | 2.0 | 1.9 | 0.00 | 2.0 | 1.9 | 0.01 |
| Coronary arteriosclerosis | 4.1 | 6.5 | -0.11 | 4.9 | 4.8 | 0.00 | 5.4 | 5.4 | 0.00 |
| Heart disease | 13.5 | 15.3 | -0.05 | 14.1 | 13.7 | 0.01 | 14.9 | 14.5 | 0.01 |
| Heart failure | 2.1 | 2.6 | -0.03 | 2.3 | 2.5 | -0.02 | 2.5 | 2.6 | -0.01 |
| Ischemic heart disease | 2.5 | 3.4 | -0.05 | 2.8 | 2.7 | 0.00 | 2.9 | 3.0 | -0.01 |
| Peripheral vascular disease | 3.5 | 3.9 | -0.02 | 3.7 | 3.9 | -0.01 | 3.7 | 3.9 | -0.01 |
| Pulmonary embolism | 0.6 | 0.4 | 0.02 | 0.5 | 0.4 | 0.02 | 0.5 | 0.5 | 0.00 |
| Venous thrombosis | 0.8 | 0.8 | 0.00 | 0.6 | 0.8 | -0.02 | 0.7 | 0.8 | -0.01 |

| (Continued) | Before ajustment | | | After matching | | | After stratification | | |
|---|---|---|---|---|---|---|---|---|---|
| Characteristic | T (%) | C (%) | StdDiff | T (%) | C (%) | StdDiff | T (%) | C (%) | StdDiff |
| **Medical history: Neoplasms** | | | | | | | | | |
| Hematologic neoplasm | 0.5 | 0.5 | -0.01 | 0.4 | 0.5 | -0.02 | 0.5 | 0.5 | -0.01 |
| Malignant lymphoma | 0.2 | 0.3 | -0.02 | 0.2 | 0.3 | -0.01 | 0.2 | 0.3 | -0.02 |
| Malignant neoplasm of anorectum | 0.1 | 0.1 | 0.01 | 0.1 | 0.1 | 0.01 | 0.1 | 0.1 | 0.01 |
| Malignant neoplastic disease | 5.3 | 5.5 | -0.01 | 5.2 | 5.5 | -0.01 | 5.4 | 5.6 | -0.01 |
| Malignant tumor of breast | 1.9 | 2.0 | 0.00 | 2.0 | 2.0 | 0.00 | 2.0 | 2.0 | 0.00 |
| Malignant tumor of colon | 0.2 | 0.2 | 0.01 | 0.3 | 0.2 | 0.02 | 0.3 | 0.2 | 0.02 |
| Malignant tumor of lung | 0.1 | 0.1 | -0.03 | 0.1 | 0.1 | -0.02 | 0.1 | 0.1 | -0.02 |
| **Medication use** | | | | | | | | | |
| Agents acting on the renin-angiotensin system | 55.9 | 61.3 | -0.11 | 59.0 | 58.2 | 0.02 | 59.3 | 59.0 | 0.01 |
| Antibacterials for systemic use | 50.8 | 49.2 | 0.03 | 49.3 | 49.3 | 0.00 | 49.5 | 49.7 | 0.00 |
| Antidepressants | 47.9 | 41.4 | 0.13 | 44.7 | 45.6 | -0.02 | 44.0 | 45.0 | -0.02 |
| Antiepileptics | 23.6 | 20.7 | 0.07 | 21.9 | 22.5 | -0.01 | 21.8 | 22.2 | -0.01 |
| Antiinflammatory and antirheumatic products | 56.4 | 56.3 | 0.00 | 56.3 | 56.2 | 0.00 | 56.5 | 56.7 | 0.00 |
| Antineoplastic agents | 5.4 | 4.8 | 0.03 | 4.9 | 5.0 | 0.00 | 5.1 | 5.0 | 0.01 |
| Antipsoriatics | 0.7 | 0.5 | 0.03 | 0.6 | 0.5 | 0.02 | 0.6 | 0.5 | 0.02 |
| Antithrombotic agents | 29.2 | 32.0 | -0.06 | 30.3 | 30.2 | 0.00 | 30.9 | 30.8 | 0.00 |
| Beta blocking agents | 26.5 | 29.7 | -0.07 | 28.0 | 27.8 | 0.00 | 28.6 | 28.5 | 0.00 |
| Calcium channel blockers | 17.4 | 18.9 | -0.04 | 18.0 | 18.1 | 0.00 | 18.0 | 18.1 | 0.00 |
| Diuretics | 41.5 | 41.0 | 0.01 | 40.8 | 40.8 | 0.00 | 41.1 | 41.1 | 0.00 |
| Drugs for acid related disorders | 43.0 | 41.6 | 0.03 | 41.8 | 42.6 | -0.02 | 41.9 | 42.6 | -0.01 |
| Drugs for obstructive airway diseases | 51.4 | 48.4 | 0.06 | 49.5 | 50.4 | -0.02 | 49.8 | 50.3 | -0.01 |
| Immunosuppressants | 4.0 | 3.3 | 0.03 | 3.9 | 3.6 | 0.01 | 3.8 | 3.6 | 0.01 |
| Lipid modifying agents | 60.5 | 65.9 | -0.11 | 63.8 | 63.2 | 0.01 | 64.0 | 63.4 | 0.01 |
| Opioids | 28.9 | 27.7 | 0.03 | 27.8 | 27.6 | 0.00 | 27.8 | 27.6 | 0.00 |
| Psycholeptics | 34.8 | 31.5 | 0.07 | 32.2 | 33.0 | -0.02 | 32.4 | 33.1 | -0.01 |
| Psychostimulants | 6.1 | 4.5 | 0.07 | 5.1 | 5.3 | -0.01 | 5.2 | 5.3 | -0.01 |

#### 2.2 SGLT2I (female) vs DPP4I (female)

##### 2.2.1 CCAE

**Supplementary Table 163:** Baseline patient characteristics for SGLT2I (female) (T) and DPP4I (female) (C) new-users in the CCAE data source. We report proportion of initiators satisfying selected base-line characteristics and the standardized difference of population proportions (StdDiff) before and after propensity score adjustment. Less extreme StdDiffs through matching and stratification suggest improved balance between patient cohorts through adjustment.

| | Before adjustment | | | After matching | | | After stratification | | |
|---|---|---|---|---|---|---|---|---|---|
| Characteristic | T (%) | C (%) | StdDiff | T (%) | C (%) | StdDiff | T (%) | C (%) | StdDiff |
| **Age group** | | | | | | | | | |
| 15 - 19 | 0.1 | 0.1 | -0.02 | 0.1 | 0.1 | -0.02 | 0.1 | 0.1 | -0.01 |
| 20 - 24 | 0.6 | 0.4 | -0.03 | 0.6 | 0.5 | -0.02 | 0.6 | 0.5 | -0.01 |
| 25 - 29 | 0.8 | 0.8 | 0.00 | 0.8 | 0.8 | 0.01 | 0.7 | 0.8 | 0.01 |
| 30 - 34 | 2.1 | 2.0 | -0.01 | 2.0 | 2.0 | 0.00 | 1.9 | 2.0 | 0.00 |
| 35 - 39 | 4.8 | 4.6 | -0.01 | 4.6 | 4.6 | 0.00 | 4.6 | 4.5 | -0.01 |
| 40 - 44 | 8.9 | 8.3 | -0.02 | 8.7 | 8.7 | 0.00 | 8.5 | 8.4 | 0.00 |
| 45 - 49 | 14.9 | 13.7 | -0.04 | 14.5 | 14.6 | 0.00 | 14.1 | 14.2 | 0.00 |
| 50 - 54 | 20.3 | 20.1 | 0.00 | 20.2 | 20.3 | 0.00 | 20.2 | 20.3 | 0.00 |
| 55 - 59 | 24.3 | 24.1 | 0.00 | 24.2 | 24.3 | 0.00 | 23.9 | 24.2 | 0.01 |
| 60 - 64 | 21.5 | 23.9 | 0.06 | 22.5 | 22.2 | -0.01 | 23.6 | 23.1 | -0.01 |
| 65 - 69 | 1.6 | 2.1 | 0.04 | 1.7 | 1.9 | 0.02 | 1.8 | 2.0 | 0.01 |
| **Medical history: General** | | | | | | | | | |
| Acute respiratory disease | 33.9 | 33.5 | -0.01 | 33.8 | 34.0 | 0.00 | 33.8 | 34.0 | 0.00 |
| Attention deficit hyperactivity disorder | 1.3 | 0.8 | -0.05 | 1.2 | 1.2 | 0.00 | 1.2 | 1.1 | -0.01 |
| Chronic liver disease | 2.2 | 3.9 | 0.10 | 2.9 | 2.9 | 0.00 | 2.9 | 2.9 | 0.00 |
| Chronic obstructive lung disease | 2.6 | 2.8 | 0.01 | 2.7 | 2.4 | -0.02 | 2.7 | 2.7 | 0.00 |
| Crohn's disease | 0.3 | 0.3 | 0.00 | 0.3 | 0.4 | 0.01 | 0.3 | 0.4 | 0.01 |
| Dementia | 0.2 | 0.2 | 0.00 | 0.2 | 0.2 | 0.00 | 0.3 | 0.2 | -0.01 |
| Depressive disorder | 15.2 | 12.7 | -0.07 | 14.9 | 15.1 | 0.01 | 14.7 | 15.0 | 0.01 |
| Gastroesophageal reflux disease | 15.6 | 13.1 | -0.07 | 15.7 | 15.8 | 0.00 | 15.7 | 15.8 | 0.00 |
| Gastrointestinal hemorrhage | 1.6 | 1.8 | 0.02 | 1.7 | 1.4 | -0.02 | 1.7 | 1.6 | -0.01 |
| Human immunodeficiency virus infection | 0.1 | 0.1 | 0.00 | 0.1 | 0.2 | 0.02 | 0.1 | 0.2 | 0.01 |
| Hyperlipidemia | 69.6 | 64.4 | -0.11 | 69.4 | 69.3 | 0.00 | 69.5 | 68.9 | -0.01 |
| Hypertensive disorder | 69.0 | 65.0 | -0.09 | 69.2 | 69.1 | 0.00 | 69.4 | 68.9 | -0.01 |
| Lesion of liver | 0.5 | 0.6 | 0.01 | 0.5 | 0.6 | 0.01 | 0.5 | 0.6 | 0.01 |
| Obesity | 30.9 | 19.4 | -0.27 | 29.3 | 29.7 | 0.01 | 27.9 | 28.2 | 0.01 |
| Osteoarthritis | 20.3 | 19.4 | -0.02 | 20.4 | 20.1 | -0.01 | 20.7 | 20.5 | 0.00 |
| Pneumonia | 2.2 | 2.1 | 0.00 | 2.2 | 2.1 | -0.01 | 2.3 | 2.2 | -0.01 |
| Psoriasis | 1.7 | 1.5 | -0.02 | 1.7 | 1.7 | 0.00 | 1.7 | 1.7 | 0.00 |
| Renal impairment | 2.7 | 3.1 | 0.02 | 2.9 | 2.9 | 0.00 | 3.4 | 3.3 | 0.00 |
| Rheumatoid arthritis | 1.6 | 1.6 | 0.00 | 1.6 | 1.4 | -0.01 | 1.6 | 1.5 | -0.01 |
| Schizophrenia | 0.1 | 0.1 | 0.01 | 0.1 | 0.1 | 0.00 | 0.1 | 0.1 | 0.01 |
| Ulcerative colitis | 0.3 | 0.3 | 0.00 | 0.4 | 0.3 | -0.01 | 0.4 | 0.3 | -0.01 |
| Urinary tract infectious disease | 10.3 | 11.8 | 0.05 | 10.6 | 10.2 | -0.01 | 11.2 | 10.7 | -0.02 |
| Viral hepatitis C | 0.2 | 0.3 | 0.02 | 0.2 | 0.3 | 0.02 | 0.2 | 0.3 | 0.01 |
| Visual system disorder | 26.0 | 27.0 | 0.02 | 26.7 | 26.4 | -0.01 | 27.6 | 27.2 | -0.01 |
| **Medical history: Cardiovascular disease** | | | | | | | | | |
| Atrial fibrillation | 1.2 | 1.3 | 0.00 | 1.2 | 1.2 | 0.00 | 1.3 | 1.3 | 0.00 |
| Cerebrovascular disease | 2.2 | 2.5 | 0.02 | 2.3 | 2.2 | 0.00 | 2.4 | 2.3 | -0.01 |
| Coronary arteriosclerosis | 4.2 | 4.3 | 0.01 | 4.1 | 3.8 | -0.01 | 4.2 | 3.9 | -0.01 |
| Heart disease | 14.0 | 14.3 | 0.01 | 13.8 | 13.4 | -0.01 | 14.3 | 13.8 | -0.01 |
| Heart failure | 1.9 | 1.7 | -0.01 | 1.8 | 1.8 | 0.00 | 1.8 | 1.9 | 0.01 |
| Ischemic heart disease | 2.8 | 2.8 | 0.00 | 2.7 | 2.5 | -0.01 | 2.9 | 2.6 | -0.01 |
| Peripheral vascular disease | 3.0 | 3.0 | 0.00 | 3.0 | 3.0 | 0.00 | 3.2 | 3.0 | -0.01 |
| Pulmonary embolism | 0.3 | 0.4 | 0.01 | 0.3 | 0.4 | 0.01 | 0.3 | 0.4 | 0.02 |
| Venous thrombosis | 0.8 | 1.0 | 0.02 | 0.8 | 0.9 | 0.01 | 0.9 | 0.9 | 0.01 |
| **Medical history: Neoplasms** | | | | | | | | | |
| Hematologic neoplasm | 0.7 | 0.8 | 0.01 | 0.7 | 0.8 | 0.01 | 0.7 | 0.8 | 0.01 |
| Malignant lymphoma | 0.3 | 0.3 | 0.01 | 0.3 | 0.3 | 0.01 | 0.3 | 0.4 | 0.01 |
| Malignant neoplasm of anorectum | 0.1 | 0.1 | 0.01 | 0.1 | 0.1 | 0.01 | 0.1 | 0.1 | 0.01 |
| Malignant neoplastic disease | 6.3 | 6.6 | 0.01 | 6.4 | 6.6 | 0.01 | 6.5 | 6.7 | 0.01 |
| Malignant tumor of breast | 2.5 | 2.7 | 0.01 | 2.6 | 2.7 | 0.01 | 2.7 | 2.7 | 0.00 |
| Malignant tumor of colon | 0.2 | 0.2 | 0.01 | 0.2 | 0.3 | 0.01 | 0.2 | 0.2 | 0.00 |
| Malignant tumor of lung | 0.1 | 0.2 | 0.01 | 0.1 | 0.2 | 0.01 | 0.2 | 0.2 | 0.01 |

| (Continued) | Before ajustment | | | After matching | | | After stratification | | |
|---|---|---|---|---|---|---|---|---|---|
| Characteristic | T (%) | C (%) | StdDiff | T (%) | C (%) | StdDiff | T (%) | C (%) | StdDiff |
| Malignant tumor of urinary bladder | 0.1 | 0.1 | 0.01 | 0.1 | 0.1 | 0.00 | 0.1 | 0.1 | 0.01 |
| **Medication use** | | | | | | | | | |
| Agents acting on the renin-angiotensin system | 61.6 | 63.5 | 0.04 | 62.1 | 62.1 | 0.00 | 62.4 | 62.6 | 0.00 |
| Antibacterials for systemic use | 64.9 | 66.8 | 0.04 | 65.3 | 64.9 | -0.01 | 65.7 | 65.5 | 0.00 |
| Antidepressants | 37.5 | 34.6 | -0.06 | 36.5 | 36.8 | 0.00 | 35.5 | 35.9 | 0.01 |
| Antiepileptics | 16.1 | 14.0 | -0.06 | 15.8 | 15.5 | -0.01 | 15.7 | 15.4 | -0.01 |
| Antiinflammatory and antirheumatic products | 38.2 | 35.0 | -0.06 | 38.0 | 38.0 | 0.00 | 38.1 | 37.6 | -0.01 |
| Antineoplastic agents | 4.4 | 4.8 | 0.02 | 4.4 | 4.3 | 0.00 | 4.4 | 4.4 | 0.00 |
| Antipsoriatics | 0.6 | 0.6 | 0.01 | 0.6 | 0.5 | -0.01 | 0.6 | 0.6 | 0.00 |
| Antithrombotic agents | 7.8 | 7.4 | -0.01 | 7.6 | 7.8 | 0.01 | 7.7 | 8.0 | 0.01 |
| Beta blocking agents | 24.3 | 24.7 | 0.01 | 24.1 | 23.8 | -0.01 | 24.4 | 24.1 | -0.01 |
| Calcium channel blockers | 18.2 | 18.6 | 0.01 | 18.3 | 18.4 | 0.00 | 18.6 | 18.7 | 0.00 |
| Diuretics | 42.7 | 43.4 | 0.01 | 42.5 | 42.6 | 0.00 | 42.6 | 42.4 | 0.00 |
| Drugs for acid related disorders | 27.5 | 27.8 | 0.01 | 27.7 | 27.6 | 0.00 | 28.3 | 27.8 | -0.01 |
| Drugs for obstructive airway diseases | 44.0 | 42.7 | -0.03 | 44.0 | 43.6 | -0.01 | 44.3 | 43.9 | -0.01 |
| Immunosuppressants | 3.3 | 3.0 | -0.02 | 3.4 | 3.1 | -0.02 | 3.4 | 3.2 | -0.01 |
| Lipid modifying agents | 62.4 | 63.4 | 0.02 | 62.4 | 62.1 | -0.01 | 62.8 | 62.6 | 0.00 |
| Opioids | 27.6 | 28.7 | 0.03 | 27.9 | 27.3 | -0.01 | 28.2 | 27.7 | -0.01 |
| Psychostimulants | 4.2 | 3.7 | -0.03 | 4.0 | 4.2 | 0.01 | 3.9 | 4.0 | 0.01 |

##### 2.2.2 DAG

**Supplementary Table 164:** Baseline patient characteristics for SGLT2I (female) (T) and DPP4I (female) (C) new-users in the DAG data source. We report proportion of initiators satisfying selected base-line characteristics and the standardized difference of population proportions (StdDiff) before and after propensity score adjustment. Less extreme StdDiffs through matching and stratification suggest improved balance between patient cohorts through adjustment.

| | Before adjustment | | | After matching | | | After stratification | | |
|---|---|---|---|---|---|---|---|---|---|
| Characteristic | T (%) | C (%) | StdDiff | T (%) | C (%) | StdDiff | T (%) | C (%) | StdDiff |
| **Age group** | | | | | | | | | |
| 20 - 24 | 0.2 | 0.1 | -0.04 | 0.2 | <0.2 | -0.05 | 0.5 | 0.1 | -0.08 |
| 25 - 29 | 0.3 | 0.2 | -0.02 | 0.3 | 0.2 | -0.02 | 0.2 | 0.2 | -0.02 |
| 30 - 34 | 1.1 | 0.6 | -0.06 | 1.1 | 0.6 | -0.06 | 1.0 | 0.5 | -0.05 |
| 35 - 39 | 1.8 | 1.1 | -0.05 | 1.8 | 1.4 | -0.04 | 2.0 | 1.2 | -0.07 |
| 40 - 44 | 2.7 | 2.5 | -0.01 | 2.7 | 3.0 | 0.02 | 3.2 | 2.5 | -0.04 |
| 45 - 49 | 6.1 | 4.7 | -0.06 | 5.9 | 6.3 | 0.02 | 4.7 | 4.9 | 0.01 |
| 50 - 54 | 9.8 | 7.9 | -0.07 | 9.6 | 9.6 | 0.00 | 8.8 | 8.1 | -0.03 |
| 55 - 59 | 13.4 | 11.2 | -0.07 | 13.4 | 14.1 | 0.02 | 11.5 | 11.7 | 0.01 |
| 60 - 64 | 15.5 | 13.4 | -0.06 | 15.4 | 14.9 | -0.01 | 14.7 | 13.7 | -0.03 |
| 65 - 69 | 15.8 | 14.2 | -0.04 | 15.5 | 16.4 | 0.02 | 15.0 | 15.0 | 0.00 |
| 70 - 74 | 12.2 | 14.0 | 0.05 | 12.3 | 12.7 | 0.01 | 12.0 | 13.0 | 0.03 |
| 75 - 79 | 10.0 | 13.6 | 0.11 | 10.3 | 9.4 | -0.03 | 12.3 | 12.8 | 0.01 |
| 80 - 84 | 7.7 | 10.3 | 0.09 | 7.8 | 7.7 | 0.00 | 9.7 | 10.1 | 0.01 |
| 85 - 89 | 2.8 | 4.7 | 0.10 | 3.0 | 2.7 | -0.02 | 3.6 | 4.6 | 0.05 |
| 90 - 94 | 0.6 | 1.4 | 0.09 | 0.6 | 0.8 | 0.03 | 0.6 | 1.5 | 0.09 |
| 95 - 99 | <0.2 | 0.1 | 0.02 | <0.1 | <0.2 | 0.01 | <0.1 | 0.1 | 0.03 |
| **Medical history: General** | | | | | | | | | |
| Acute respiratory disease | 18.0 | 18.5 | 0.01 | 17.7 | 17.1 | -0.01 | 19.4 | 18.1 | -0.03 |
| Chronic liver disease | 0.2 | 0.4 | 0.03 | 0.2 | 0.3 | 0.02 | 0.2 | 0.4 | 0.04 |
| Chronic obstructive lung disease | 4.8 | 5.9 | 0.05 | 4.7 | 4.6 | 0.00 | 5.9 | 5.6 | -0.01 |
| Crohn’s disease | <0.2 | 0.1 | -0.01 | <0.1 | <0.2 | 0.01 | <0.1 | 0.1 | -0.01 |
| Dementia | 1.3 | 2.3 | 0.08 | 1.3 | 1.2 | -0.01 | 1.5 | 2.2 | 0.05 |
| Depressive disorder | 11.3 | 11.8 | 0.02 | 11.0 | 10.6 | -0.01 | 11.2 | 11.9 | 0.02 |
| Gastroesophageal reflux disease | 2.3 | 3.0 | 0.04 | 2.3 | 2.3 | 0.00 | 2.2 | 2.8 | 0.03 |
| Gastrointestinal hemorrhage | 0.7 | 0.7 | 0.00 | 0.7 | 0.9 | 0.01 | 0.6 | 0.8 | 0.03 |
| Hyperlipidemia | 21.7 | 22.6 | 0.02 | 20.5 | 21.8 | 0.03 | 21.2 | 21.4 | 0.00 |
| Hypertensive disorder | 42.2 | 48.4 | 0.12 | 41.6 | 40.9 | -0.01 | 43.8 | 44.5 | 0.01 |
| Lesion of liver | 0.3 | 0.5 | 0.04 | 0.3 | 0.4 | 0.02 | 0.6 | 0.5 | -0.01 |
| Obesity | 11.2 | 9.5 | -0.06 | 10.3 | 10.4 | 0.00 | 10.2 | 9.6 | -0.02 |
| Osteoarthritis | 13.2 | 14.1 | 0.03 | 12.2 | 13.5 | 0.04 | 12.2 | 13.5 | 0.04 |
| Pneumonia | 1.2 | 2.0 | 0.06 | 1.2 | 1.6 | 0.03 | 1.5 | 1.9 | 0.03 |
| Psoriasis | 1.4 | 1.3 | -0.01 | 1.5 | 1.5 | 0.00 | 1.8 | 1.4 | -0.04 |
| Renal impairment | 5.0 | 7.3 | 0.10 | 5.1 | 4.5 | -0.03 | 6.3 | 6.9 | 0.02 |
| Rheumatoid arthritis | 2.0 | 1.7 | -0.02 | 1.9 | 1.6 | -0.02 | 2.1 | 1.7 | -0.03 |
| Schizophrenia | 0.3 | 0.2 | -0.02 | 0.3 | 0.2 | -0.03 | 0.2 | 0.2 | 0.00 |
| Ulcerative colitis | 0.2 | 0.3 | 0.02 | 0.2 | 0.4 | 0.04 | 0.3 | 0.4 | 0.02 |
| Urinary tract infectious disease | 7.2 | 8.6 | 0.06 | 7.3 | 7.0 | -0.01 | 8.1 | 8.3 | 0.01 |
| Viral hepatitis C | <0.2 | 0.1 | -0.01 | <0.1 | 0.1 | 0.00 | <0.1 | 0.1 | 0.00 |
| Visual system disorder | 7.7 | 9.5 | 0.06 | 7.6 | 7.7 | 0.00 | 7.3 | 8.8 | 0.05 |
| **Medical history: Cardiovascular disease** | | | | | | | | | |
| Atrial fibrillation | 2.3 | 3.0 | 0.04 | 2.2 | 2.2 | 0.00 | 2.8 | 3.0 | 0.01 |
| Cerebrovascular disease | 3.2 | 3.8 | 0.04 | 3.1 | 3.3 | 0.01 | 2.9 | 3.6 | 0.04 |
| Coronary arteriosclerosis | 4.0 | 2.9 | -0.06 | 3.4 | 3.6 | 0.01 | 3.6 | 3.3 | -0.02 |
| Heart disease | 21.2 | 23.7 | 0.06 | 19.9 | 18.7 | -0.03 | 20.5 | 22.5 | 0.05 |
| Heart failure | 7.1 | 7.3 | 0.01 | 6.4 | 5.1 | -0.06 | 7.5 | 6.9 | -0.03 |
| Ischemic heart disease | 8.1 | 8.7 | 0.02 | 7.5 | 6.7 | -0.03 | 7.3 | 8.1 | 0.03 |
| Peripheral vascular disease | 4.7 | 4.8 | 0.00 | 4.3 | 3.9 | -0.02 | 3.4 | 4.7 | 0.06 |
| Pulmonary embolism | 0.6 | 0.7 | 0.01 | 0.5 | 0.7 | 0.03 | 0.5 | 0.6 | 0.02 |
| Venous thrombosis | 1.4 | 1.8 | 0.03 | 1.4 | 1.4 | 0.01 | 1.6 | 1.7 | 0.01 |
| **Medical history: Neoplasms** | | | | | | | | | |
| Hematologic neoplasm | 0.2 | 0.4 | 0.03 | 0.2 | 0.3 | 0.01 | 0.6 | 0.3 | -0.04 |
| Malignant lymphoma | 0.2 | 0.3 | 0.02 | 0.2 | 0.3 | 0.02 | 0.2 | 0.3 | 0.01 |
| Malignant neoplasm of anorectum | <0.2 | 0.2 | 0.05 | <0.1 | <0.2 | 0.04 | <0.1 | 0.2 | 0.05 |
| Malignant neoplastic disease | 4.4 | 5.0 | 0.03 | 4.3 | 4.0 | -0.01 | 4.9 | 4.7 | -0.01 |
| Malignant tumor of breast | 1.4 | 1.2 | -0.02 | 1.3 | 1.0 | -0.03 | 1.8 | 1.1 | -0.06 |
| Malignant tumor of colon | 0.4 | 0.4 | 0.00 | 0.4 | 0.3 | -0.01 | 0.3 | 0.4 | 0.02 |
| Malignant tumor of lung | <0.2 | 0.1 | 0.01 | <0.1 | <0.2 | -0.01 | <0.1 | 0.1 | 0.00 |

| (Continued) | Before ajustment | | | After matching | | | After stratification | | |
|---|---|---|---|---|---|---|---|---|---|
| Characteristic | T (%) | C (%) | StdDiff | T (%) | C (%) | StdDiff | T (%) | C (%) | StdDiff |
| Malignant tumor of urinary bladder | <0.2 | 0.1 | 0.01 | <0.1 | <0.2 | 0.01 | <0.1 | 0.1 | 0.01 |
| **Medication use** | | | | | | | | | |
| Agents acting on the renin-angiotensin system | 67.7 | 67.1 | -0.01 | 66.7 | 64.7 | -0.04 | 65.4 | 66.8 | 0.03 |
| Antibacterials for systemic use | 24.2 | 30.2 | 0.13 | 25.0 | 24.5 | -0.01 | 29.8 | 28.3 | -0.03 |
| Antidepressants | 14.2 | 14.2 | 0.00 | 14.1 | 13.2 | -0.03 | 14.4 | 14.5 | 0.00 |
| Antiepileptics | 4.7 | 5.5 | 0.04 | 4.8 | 5.2 | 0.02 | 5.1 | 5.7 | 0.03 |
| Antiinflammatory and antirheumatic products | 39.6 | 41.7 | 0.04 | 38.9 | 39.4 | 0.01 | 39.7 | 40.6 | 0.02 |
| Antineoplastic agents | 1.5 | 1.7 | 0.01 | 1.6 | 1.5 | -0.01 | 1.5 | 1.8 | 0.03 |
| Antipsoriatics | 0.5 | 0.4 | 0.00 | 0.4 | 0.4 | 0.01 | 0.5 | 0.5 | 0.00 |
| Antithrombotic agents | 26.8 | 27.7 | 0.02 | 25.7 | 24.0 | -0.04 | 25.7 | 27.3 | 0.04 |
| Beta blocking agents | 47.1 | 50.2 | 0.06 | 46.5 | 43.7 | -0.06 | 48.6 | 48.9 | 0.01 |
| Calcium channel blockers | 33.9 | 33.6 | 0.00 | 33.5 | 33.3 | -0.01 | 32.6 | 34.3 | 0.04 |
| Diuretics | 48.8 | 52.9 | 0.08 | 48.0 | 46.1 | -0.04 | 50.8 | 50.8 | 0.00 |
| Drugs for acid related disorders | 36.6 | 38.8 | 0.04 | 36.7 | 35.5 | -0.03 | 39.4 | 38.9 | -0.01 |
| Drugs for obstructive airway diseases | 18.3 | 18.6 | 0.01 | 18.0 | 18.0 | 0.00 | 20.3 | 18.8 | -0.04 |
| Immunosuppressants | 0.8 | 0.8 | 0.00 | 0.9 | 0.8 | -0.01 | 0.8 | 0.9 | 0.01 |
| Lipid modifying agents | 45.9 | 42.0 | -0.08 | 43.5 | 44.0 | 0.01 | 41.6 | 42.8 | 0.03 |
| Opioids | 12.9 | 14.2 | 0.04 | 12.9 | 11.7 | -0.04 | 13.7 | 13.8 | 0.00 |
| Psycholeptics | 9.5 | 11.7 | 0.07 | 9.7 | 9.2 | -0.01 | 11.0 | 11.2 | 0.01 |
| Psychostimulants | 0.2 | 0.3 | 0.02 | 0.2 | 0.3 | 0.02 | 0.5 | 0.2 | -0.04 |

##### 2.2.3 ODOD

**Supplementary Table 165:** Baseline patient characteristics for SGLT2I (female) (T) and DPP4I (female) (C) new-users in the ODOD data source. We report proportion of initiators satisfying selected base-line characteristics and the standardized difference of population proportions (StdDiff) before and after propensity score adjustment. Less extreme StdDiffs through matching and stratification suggest improved balance between patient cohorts through adjustment.

| | Before adjustment | | | After matching | | | After stratification | | |
|---|---|---|---|---|---|---|---|---|---|
| Characteristic | T (%) | C (%) | StdDiff | T (%) | C (%) | StdDiff | T (%) | C (%) | StdDiff |
| **Age group** | | | | | | | | | |
| 20 - 24 | 0.2 | 0.2 | -0.01 | 0.2 | 0.2 | -0.01 | 0.2 | 0.2 | 0.00 |
| 25 - 29 | 0.4 | 0.4 | 0.00 | 0.4 | 0.3 | -0.02 | 0.4 | 0.3 | -0.02 |
| 30 - 34 | 1.3 | 1.3 | 0.00 | 1.3 | 1.2 | -0.01 | 1.1 | 1.1 | 0.00 |
| 35 - 39 | 3.1 | 2.5 | -0.03 | 2.9 | 2.7 | -0.01 | 2.6 | 2.3 | -0.02 |
| 40 - 44 | 5.4 | 4.4 | -0.05 | 5.0 | 4.8 | -0.01 | 4.5 | 4.2 | -0.01 |
| 45 - 49 | 8.9 | 7.3 | -0.06 | 8.3 | 8.5 | 0.01 | 7.5 | 7.2 | -0.01 |
| 50 - 54 | 11.7 | 10.1 | -0.05 | 11.1 | 11.4 | 0.01 | 10.0 | 9.9 | 0.00 |
| 55 - 59 | 14.5 | 12.0 | -0.07 | 13.6 | 14.1 | 0.02 | 13.0 | 12.2 | -0.02 |
| 60 - 64 | 13.6 | 12.5 | -0.03 | 13.3 | 13.4 | 0.00 | 12.3 | 12.1 | -0.01 |
| 65 - 69 | 16.5 | 15.5 | -0.03 | 16.8 | 17.6 | 0.02 | 15.7 | 17.1 | 0.04 |
| 70 - 74 | 14.2 | 15.1 | 0.03 | 15.4 | 14.9 | -0.01 | 16.0 | 16.3 | 0.01 |
| 75 - 79 | 6.6 | 9.5 | 0.11 | 7.4 | 6.8 | -0.02 | 8.8 | 9.0 | 0.00 |
| 80 - 84 | 2.5 | 5.8 | 0.17 | 2.9 | 2.6 | -0.02 | 5.2 | 5.0 | -0.01 |
| 85 - 89 | 1.0 | 3.1 | 0.15 | 1.1 | 1.0 | -0.02 | 2.7 | 2.8 | 0.00 |
| 90 - 94 | 0.1 | 0.2 | 0.03 | 0.1 | 0.4 | 0.05 | 0.1 | 0.3 | 0.05 |
| **Medical history: General** | | | | | | | | | |
| Acute respiratory disease | 30.4 | 31.5 | 0.02 | 30.5 | 30.2 | -0.01 | 30.9 | 30.5 | -0.01 |
| Attention deficit hyperactivity disorder | 1.3 | 0.6 | -0.07 | 1.2 | 0.9 | -0.03 | 1.1 | 0.8 | -0.03 |
| Chronic liver disease | 2.2 | 3.7 | 0.09 | 2.4 | 2.0 | -0.03 | 2.8 | 2.5 | -0.02 |
| Chronic obstructive lung disease | 8.2 | 9.2 | 0.04 | 8.5 | 8.8 | 0.01 | 9.8 | 9.7 | 0.00 |
| Crohn’s disease | 0.3 | 0.3 | 0.00 | 0.3 | 0.3 | 0.00 | 0.3 | 0.3 | 0.01 |
| Dementia | 1.0 | 2.8 | 0.13 | 1.2 | 1.2 | 0.00 | 2.6 | 2.6 | 0.00 |
| Depressive disorder | 21.8 | 18.9 | -0.07 | 21.2 | 21.4 | 0.00 | 21.2 | 21.4 | 0.00 |
| Gastroesophageal reflux disease | 22.9 | 21.6 | -0.03 | 23.0 | 22.9 | 0.00 | 24.0 | 23.5 | -0.01 |
| Gastrointestinal hemorrhage | 2.2 | 2.7 | 0.03 | 2.3 | 2.1 | -0.01 | 2.8 | 2.5 | -0.02 |
| Human immunodeficiency virus infection | 0.1 | 0.1 | 0.01 | 0.1 | 0.2 | 0.02 | 0.1 | 0.2 | 0.03 |
| Hyperlipidemia | 78.3 | 80.0 | 0.04 | 78.2 | 78.1 | 0.00 | 79.2 | 79.3 | 0.00 |
| Hypertensive disorder | 77.5 | 79.5 | 0.05 | 77.8 | 76.9 | -0.02 | 79.5 | 79.1 | -0.01 |
| Lesion of liver | 1.1 | 1.0 | 0.00 | 1.1 | 0.9 | -0.02 | 1.3 | 1.0 | -0.02 |
| Obesity | 34.9 | 24.3 | -0.23 | 32.0 | 32.6 | 0.01 | 30.0 | 30.1 | 0.00 |
| Osteoarthritis | 30.1 | 31.7 | 0.03 | 33.3 | 32.8 | -0.01 | 33.3 | 32.8 | -0.01 |
| Pneumonia | 3.5 | 4.0 | 0.03 | 3.5 | 3.3 | -0.01 | 4.2 | 4.0 | -0.01 |
| Psoriasis | 1.9 | 1.6 | -0.02 | 1.9 | 1.7 | -0.01 | 1.9 | 1.7 | -0.01 |
| Renal impairment | 10.3 | 14.8 | 0.14 | 11.1 | 9.9 | -0.04 | 14.4 | 14.5 | 0.00 |
| Rheumatoid arthritis | 2.6 | 2.6 | 0.00 | 2.9 | 2.7 | -0.01 | 2.9 | 2.7 | -0.01 |
| Schizophrenia | 0.3 | 0.4 | 0.02 | 0.3 | 0.5 | 0.04 | 0.3 | 0.5 | 0.04 |
| Ulcerative colitis | 0.4 | 0.4 | 0.01 | 0.4 | 0.3 | 0.00 | 0.4 | 0.4 | -0.01 |
| Urinary tract infectious disease | 12.1 | 17.0 | 0.14 | 13.0 | 12.4 | -0.02 | 15.1 | 15.3 | 0.00 |
| Viral hepatitis C | 0.3 | 0.5 | 0.02 | 0.3 | 0.4 | 0.01 | 0.3 | 0.5 | 0.02 |
| Visual system disorder | 40.7 | 43.3 | 0.05 | 41.9 | 40.9 | -0.02 | 44.1 | 43.6 | -0.01 |
| **Medical history: Cardiovascular disease** | | | | | | | | | |
| Atrial fibrillation | 4.4 | 5.4 | 0.05 | 4.5 | 4.0 | -0.02 | 5.3 | 5.4 | 0.01 |
| Cerebrovascular disease | 5.4 | 6.9 | 0.06 | 5.6 | 5.1 | -0.02 | 6.7 | 6.5 | -0.01 |
| Coronary arteriosclerosis | 11.1 | 11.4 | 0.01 | 10.6 | 10.1 | -0.01 | 11.8 | 11.5 | -0.01 |
| Heart disease | 25.7 | 27.8 | 0.05 | 25.3 | 24.8 | -0.01 | 27.9 | 27.7 | -0.01 |
| Heart failure | 6.6 | 7.0 | 0.02 | 6.2 | 5.8 | -0.01 | 7.2 | 7.3 | 0.00 |
| Ischemic heart disease | 6.7 | 6.4 | -0.01 | 6.3 | 5.6 | -0.03 | 6.5 | 6.4 | 0.00 |
| Peripheral vascular disease | 9.8 | 10.4 | 0.02 | 9.9 | 9.5 | -0.01 | 11.2 | 11.0 | 0.00 |
| Pulmonary embolism | 0.8 | 0.7 | -0.01 | 0.8 | 0.7 | -0.02 | 1.0 | 0.8 | -0.02 |
| Venous thrombosis | 1.3 | 1.7 | 0.03 | 1.3 | 1.3 | 0.00 | 1.7 | 1.6 | 0.00 |
| **Medical history: Neoplasms** | | | | | | | | | |
| Hematologic neoplasm | 0.8 | 1.2 | 0.04 | 0.8 | 1.1 | 0.02 | 0.9 | 1.2 | 0.03 |
| Malignant lymphoma | 0.4 | 0.5 | 0.01 | 0.5 | 0.5 | 0.01 | 0.5 | 0.5 | 0.00 |
| Malignant neoplasm of anorectum | 0.2 | 0.2 | 0.00 | 0.2 | 0.2 | 0.00 | 0.2 | 0.2 | 0.00 |
| Malignant neoplastic disease | 8.8 | 9.8 | 0.03 | 9.0 | 8.8 | -0.01 | 9.4 | 9.9 | 0.02 |
| Malignant tumor of breast | 3.2 | 3.7 | 0.03 | 3.3 | 3.2 | 0.00 | 3.4 | 3.7 | 0.02 |
| Malignant tumor of colon | 0.3 | 0.5 | 0.03 | 0.3 | 0.4 | 0.02 | 0.5 | 0.5 | 0.00 |

| (Continued) | Before ajustment | | | After matching | | | After stratification | | |
|---|---|---|---|---|---|---|---|---|---|
| Characteristic | T (%) | C (%) | StdDiff | T (%) | C (%) | StdDiff | T (%) | C (%) | StdDiff |
| Malignant tumor of lung | 0.3 | 0.3 | 0.00 | 0.4 | 0.3 | -0.01 | 0.3 | 0.4 | 0.01 |
| Malignant tumor of urinary bladder | 0.1 | 0.2 | 0.02 | 0.1 | 0.2 | 0.01 | 0.1 | 0.2 | 0.02 |
| **Medication use** | | | | | | | | | |
| Agents acting on the renin-angiotensin system | 67.5 | 68.8 | 0.03 | 67.8 | 66.9 | -0.02 | 68.6 | 68.6 | 0.00 |
| Antibacterials for systemic use | 63.5 | 65.0 | 0.03 | 63.5 | 63.4 | 0.00 | 64.7 | 64.0 | -0.01 |
| Antidepressants | 39.3 | 35.9 | -0.07 | 38.6 | 38.6 | 0.00 | 37.9 | 38.0 | 0.00 |
| Antiepileptics | 20.6 | 19.8 | -0.02 | 21.1 | 20.9 | 0.00 | 21.3 | 21.9 | 0.01 |
| Antiinflammatory and antirheumatic products | 37.0 | 34.5 | -0.05 | 36.5 | 37.0 | 0.01 | 36.6 | 36.3 | -0.01 |
| Antineoplastic agents | 5.4 | 6.4 | 0.04 | 5.6 | 5.6 | 0.00 | 5.8 | 6.0 | 0.01 |
| Antipsoriatics | 0.6 | 0.7 | 0.02 | 0.6 | 0.7 | 0.01 | 0.6 | 0.7 | 0.02 |
| Antithrombotic agents | 13.9 | 13.6 | -0.01 | 13.4 | 12.9 | -0.02 | 15.0 | 14.3 | -0.02 |
| Beta blocking agents | 33.0 | 34.3 | 0.03 | 32.8 | 31.8 | -0.02 | 34.8 | 34.4 | -0.01 |
| Calcium channel blockers | 24.4 | 27.0 | 0.06 | 25.0 | 24.7 | -0.01 | 27.5 | 27.1 | -0.01 |
| Diuretics | 45.9 | 48.0 | 0.04 | 45.8 | 45.5 | -0.01 | 45.8 | 45.5 | -0.01 |
| Drugs for acid related disorders | 31.0 | 32.3 | 0.03 | 31.8 | 31.9 | 0.00 | 33.6 | 33.5 | 0.00 |
| Drugs for obstructive airway diseases | 45.0 | 44.2 | -0.02 | 45.2 | 45.2 | 0.00 | 46.1 | 45.8 | -0.01 |
| Immunosuppressants | 3.8 | 3.7 | -0.01 | 4.0 | 3.8 | -0.01 | 4.2 | 3.9 | -0.01 |
| Lipid modifying agents | 72.2 | 72.5 | 0.00 | 72.4 | 72.3 | 0.00 | 72.8 | 73.5 | 0.01 |
| Opioids | 29.1 | 30.6 | 0.03 | 29.3 | 29.2 | 0.00 | 30.6 | 30.0 | -0.01 |
| Psycholeptics | 28.6 | 28.7 | 0.00 | 28.4 | 28.1 | -0.01 | 29.7 | 29.1 | -0.01 |
| Psychostimulants | 2.9 | 2.4 | -0.03 | 2.8 | 2.8 | 0.00 | 2.8 | 2.5 | -0.02 |

##### 2.2.4 MDCR

**Supplementary Table 166:** Baseline patient characteristics for SGLT2I (female) (T) and DPP4I (female) (C) new-users in the MDCR data source. We report proportion of initiators satisfying selected base-line characteristics and the standardized difference of population proportions (StdDiff) before and after propensity score adjustment. Less extreme StdDiffs through matching and stratification suggest improved balance between patient cohorts through adjustment.

| | Before adjustment | | | After matching | | | After stratification | | |
|---|---|---|---|---|---|---|---|---|---|
| Characteristic | T (%) | C (%) | StdDiff | T (%) | C (%) | StdDiff | T (%) | C (%) | StdDiff |
| **Age group** | | | | | | | | | |
| 50 - 54 | <0.3 | 0.1 | -0.01 | <0.3 | <0.4 | 0.03 | <0.3 | 0.1 | -0.01 |
| 55 - 59 | 0.5 | 0.5 | 0.00 | 0.6 | 0.8 | 0.02 | 0.6 | 0.5 | -0.01 |
| 60 - 64 | 1.4 | 1.2 | -0.02 | 1.4 | 1.0 | -0.03 | 1.1 | 1.0 | -0.01 |
| 65 - 69 | 40.8 | 29.3 | -0.24 | 40.6 | 33.8 | -0.14 | 36.4 | 30.9 | -0.12 |
| 70 - 74 | 32.6 | 29.6 | -0.06 | 31.8 | 35.1 | 0.07 | 29.1 | 31.1 | 0.04 |
| 75 - 79 | 16.7 | 18.5 | 0.05 | 16.9 | 18.5 | 0.04 | 17.3 | 18.2 | 0.02 |
| 80 - 84 | 5.3 | 12.3 | 0.25 | 5.7 | 5.4 | -0.01 | 9.4 | 10.3 | 0.03 |
| 85 - 89 | 1.9 | 6.0 | 0.21 | 2.1 | 2.4 | 0.01 | 4.5 | 5.4 | 0.04 |
| 90 - 94 | 0.5 | 1.9 | 0.13 | 0.5 | 2.3 | 0.15 | 1.3 | 2.0 | 0.06 |
| 95 - 99 | <0.3 | 0.4 | 0.07 | <0.3 | <0.4 | 0.05 | <0.3 | 0.5 | 0.08 |
| **Medical history: General** | | | | | | | | | |
| Acute respiratory disease | 25.6 | 25.7 | 0.00 | 26.0 | 26.6 | 0.01 | 27.2 | 26.7 | -0.01 |
| Attention deficit hyperactivity disorder | <0.3 | 0.2 | 0.03 | <0.3 | <0.4 | 0.02 | <0.3 | 0.3 | 0.06 |
| Chronic liver disease | 2.6 | 2.8 | 0.01 | 2.7 | 1.7 | -0.07 | 3.1 | 2.9 | -0.01 |
| Chronic obstructive lung disease | 9.8 | 10.1 | 0.01 | 9.6 | 8.7 | -0.03 | 9.5 | 10.2 | 0.02 |
| Crohn’s disease | <0.3 | 0.2 | 0.01 | <0.3 | <0.4 | 0.01 | <0.3 | 0.2 | 0.02 |
| Dementia | 1.2 | 3.3 | 0.14 | 1.4 | 2.4 | 0.07 | 2.6 | 3.7 | 0.06 |
| Depressive disorder | 13.6 | 10.1 | -0.11 | 12.7 | 11.8 | -0.03 | 11.9 | 13.0 | 0.04 |
| Gastroesophageal reflux disease | 20.5 | 15.1 | -0.14 | 20.0 | 21.0 | 0.03 | 19.7 | 19.5 | 0.00 |
| Gastrointestinal hemorrhage | 3.0 | 3.5 | 0.03 | 2.9 | 2.5 | -0.03 | 3.0 | 3.2 | 0.01 |
| Hyperlipidemia | 79.0 | 61.5 | -0.39 | 78.6 | 77.4 | -0.03 | 77.2 | 76.4 | -0.02 |
| Lesion of liver | 1.2 | 1.0 | -0.03 | 0.9 | 1.4 | 0.05 | 0.9 | 1.4 | 0.05 |
| Obesity | 26.0 | 10.7 | -0.40 | 22.6 | 24.5 | 0.04 | 16.6 | 18.0 | 0.04 |
| Osteoarthritis | 35.9 | 34.2 | -0.04 | 35.3 | 37.0 | 0.04 | 36.4 | 37.6 | 0.03 |
| Pneumonia | 3.3 | 4.7 | 0.07 | 3.0 | 3.4 | 0.02 | 3.5 | 4.4 | 0.04 |
| Psoriasis | 2.5 | 1.7 | -0.05 | 2.4 | 2.1 | -0.02 | 2.3 | 1.9 | -0.03 |
| Renal impairment | 9.7 | 13.1 | 0.11 | 10.3 | 10.4 | 0.01 | 10.4 | 15.0 | 0.14 |
| Rheumatoid arthritis | 1.8 | 2.3 | 0.04 | 1.8 | 3.1 | 0.09 | 1.6 | 2.5 | 0.06 |
| Schizophrenia | <0.3 | 0.2 | 0.03 | <0.3 | <0.4 | 0.04 | <0.3 | 0.2 | 0.03 |
| Ulcerative colitis | 0.6 | 0.6 | -0.01 | 0.7 | 0.6 | 0.00 | 0.6 | 0.6 | -0.01 |
| Urinary tract infectious disease | 11.6 | 15.4 | 0.11 | 11.7 | 11.7 | 0.00 | 13.7 | 14.8 | 0.03 |
| Viral hepatitis C | <0.3 | 0.2 | 0.01 | <0.3 | <0.4 | 0.02 | <0.3 | 0.2 | 0.04 |
| Visual system disorder | 56.6 | 54.7 | -0.04 | 56.2 | 58.1 | 0.04 | 56.8 | 58.1 | 0.03 |
| **Medical history: Cardiovascular disease** | | | | | | | | | |
| Atrial fibrillation | 7.7 | 10.5 | 0.10 | 8.0 | 8.2 | 0.01 | 10.8 | 10.4 | -0.01 |
| Cerebrovascular disease | 10.6 | 12.2 | 0.05 | 10.3 | 9.5 | -0.03 | 12.9 | 11.1 | -0.05 |
| Coronary arteriosclerosis | 17.9 | 17.4 | -0.01 | 17.0 | 15.4 | -0.04 | 19.4 | 17.2 | -0.06 |
| Heart disease | 37.6 | 39.6 | 0.04 | 36.8 | 35.2 | -0.03 | 42.8 | 38.9 | -0.08 |
| Heart failure | 8.7 | 9.1 | 0.01 | 8.8 | 6.1 | -0.10 | 10.5 | 9.3 | -0.04 |
| Ischemic heart disease | 9.7 | 8.5 | -0.04 | 9.2 | 7.7 | -0.06 | 10.3 | 8.4 | -0.07 |
| Peripheral vascular disease | 12.2 | 12.4 | 0.01 | 11.6 | 12.3 | 0.02 | 11.0 | 13.6 | 0.08 |
| Pulmonary embolism | 0.8 | 0.9 | 0.01 | 0.7 | 1.3 | 0.07 | 1.2 | 1.2 | 0.00 |
| Venous thrombosis | 2.1 | 2.2 | 0.01 | 2.1 | 1.7 | -0.03 | 2.9 | 2.3 | -0.04 |
| **Medical history: Neoplasms** | | | | | | | | | |
| Hematologic neoplasm | 0.7 | 1.5 | 0.08 | 0.7 | 1.5 | 0.08 | 0.8 | 1.5 | 0.06 |
| Malignant lymphoma | 0.5 | 0.8 | 0.04 | 0.6 | 0.7 | 0.01 | 0.7 | 0.8 | 0.02 |
| Malignant neoplasm of anorectum | 0.3 | 0.3 | 0.00 | 0.3 | <0.4 | -0.04 | 0.8 | 0.2 | -0.08 |
| Malignant neoplastic disease | 14.0 | 15.7 | 0.05 | 14.1 | 15.1 | 0.03 | 14.8 | 15.8 | 0.03 |
| Malignant tumor of breast | 6.0 | 5.8 | -0.01 | 6.0 | 6.0 | 0.00 | 5.9 | 5.9 | 0.00 |
| Malignant tumor of colon | 0.7 | 0.9 | 0.01 | 0.8 | 0.7 | -0.01 | 0.9 | 0.9 | -0.01 |
| Malignant tumor of lung | 0.4 | 0.6 | 0.03 | 0.4 | 0.6 | 0.03 | <0.3 | 0.7 | 0.06 |
| Malignant tumor of urinary bladder | <0.3 | 0.4 | 0.03 | <0.3 | <0.4 | -0.01 | <0.3 | 0.5 | 0.04 |
| **Medication use** | | | | | | | | | |
| Agents acting on the renin-angiotensin system | 73.7 | 75.3 | 0.04 | 73.8 | 72.9 | -0.02 | 73.7 | 74.3 | 0.01 |
| Antibacterials for systemic use | 62.0 | 66.8 | 0.10 | 62.7 | 63.1 | 0.01 | 64.1 | 65.1 | 0.02 |
| Antidepressants | 34.1 | 32.6 | -0.03 | 33.5 | 32.0 | -0.03 | 31.3 | 33.0 | 0.04 |
| Antiepileptics | 19.5 | 17.3 | -0.06 | 19.0 | 17.7 | -0.04 | 19.5 | 18.1 | -0.04 |

| (Continued) | Before ajustment | | | After matching | | | After stratification | | |
|---|---|---|---|---|---|---|---|---|---|
| Characteristic | T (%) | C (%) | StdDiff | T (%) | C (%) | StdDiff | T (%) | C (%) | StdDiff |
| Antiinflammatory and antirheumatic products | 35.3 | 31.2 | -0.09 | 34.4 | 33.4 | -0.02 | 30.1 | 31.5 | 0.03 |
| Antineoplastic agents | 8.0 | 9.4 | 0.05 | 7.7 | 8.4 | 0.03 | 7.4 | 8.0 | 0.02 |
| Antipsoriatics | 1.1 | 0.8 | -0.03 | 1.0 | 0.5 | -0.06 | 0.8 | 0.7 | -0.01 |
| Antithrombotic agents | 18.3 | 22.0 | 0.09 | 18.4 | 18.9 | 0.01 | 22.4 | 21.1 | -0.03 |
| Beta blocking agents | 44.7 | 48.8 | 0.08 | 45.0 | 44.1 | -0.02 | 49.9 | 47.3 | -0.05 |
| Calcium channel blockers | 32.9 | 37.1 | 0.09 | 33.4 | 34.1 | 0.01 | 35.6 | 36.1 | 0.01 |
| Diuretics | 53.9 | 57.4 | 0.07 | 54.4 | 53.0 | -0.03 | 56.0 | 54.6 | -0.03 |
| Drugs for acid related disorders | 39.1 | 37.3 | -0.04 | 38.8 | 39.0 | 0.00 | 35.8 | 37.4 | 0.03 |
| Drugs for obstructive airway diseases | 49.7 | 45.0 | -0.10 | 48.9 | 48.3 | -0.01 | 47.2 | 47.0 | 0.00 |
| Immunosuppressants | 4.8 | 4.5 | -0.01 | 4.8 | 4.9 | 0.00 | 4.1 | 4.6 | 0.03 |
| Lipid modifying agents | 80.7 | 79.4 | -0.03 | 80.3 | 79.2 | -0.03 | 80.3 | 79.8 | -0.01 |
| Opioids | 27.2 | 30.4 | 0.07 | 27.9 | 25.8 | -0.05 | 30.7 | 28.3 | -0.05 |
| Psycholeptics | 27.2 | 30.7 | 0.08 | 27.4 | 27.2 | 0.00 | 29.6 | 28.3 | -0.03 |
| Psychostimulants | 2.0 | 1.7 | -0.02 | 2.2 | 1.2 | -0.07 | 1.8 | 1.6 | -0.02 |

##### 2.2.5 MDCD

**Supplementary Table 167:** Baseline patient characteristics for SGLT2I (female) (T) and DPP4I (female) (C) new-users in the MDCD data source. We report proportion of initiators satisfying selected base-line characteristics and the standardized difference of population proportions (StdDiff) before and after propensity score adjustment. Less extreme StdDiffs through matching and stratification suggest improved balance between patient cohorts through adjustment.

| | Before adjustment | | | After matching | | | After stratification | | |
|---|---|---|---|---|---|---|---|---|---|
| Characteristic | T (%) | C (%) | StdDiff | T (%) | C (%) | StdDiff | T (%) | C (%) | StdDiff |
| **Age group** | | | | | | | | | |
| 15 - 19 | 0.6 | 0.6 | 0.01 | 0.6 | 0.6 | 0.00 | 0.5 | 0.7 | 0.03 |
| 20 - 24 | 1.0 | 1.1 | 0.00 | 1.0 | 1.5 | 0.04 | 1.0 | 1.2 | 0.01 |
| 25 - 29 | 3.9 | 2.7 | -0.07 | 3.9 | 3.2 | -0.04 | 3.5 | 2.8 | -0.04 |
| 30 - 34 | 7.2 | 5.6 | -0.06 | 7.2 | 6.7 | -0.02 | 6.9 | 5.7 | -0.05 |
| 35 - 39 | 10.7 | 9.2 | -0.05 | 10.6 | 10.2 | -0.01 | 9.5 | 9.9 | 0.01 |
| 40 - 44 | 14.0 | 11.7 | -0.07 | 13.8 | 13.1 | -0.02 | 12.9 | 12.2 | -0.02 |
| 45 - 49 | 14.4 | 13.3 | -0.03 | 14.5 | 14.5 | 0.00 | 14.5 | 13.7 | -0.02 |
| 50 - 54 | 15.9 | 15.9 | 0.00 | 15.9 | 15.5 | -0.01 | 16.5 | 16.0 | -0.01 |
| 55 - 59 | 16.1 | 16.1 | 0.00 | 16.3 | 15.5 | -0.02 | 16.4 | 16.2 | 0.00 |
| 60 - 64 | 11.8 | 13.0 | 0.04 | 11.9 | 12.0 | 0.00 | 13.0 | 12.6 | -0.01 |
| 65 - 69 | 2.4 | 4.6 | 0.12 | 2.4 | 3.4 | 0.06 | 2.7 | 4.0 | 0.07 |
| 70 - 74 | 1.2 | 2.4 | 0.09 | 1.2 | 1.3 | 0.00 | 1.6 | 2.0 | 0.03 |
| 75 - 79 | 0.2 | 1.9 | 0.16 | 0.3 | 1.2 | 0.11 | 0.2 | 1.6 | 0.15 |
| 80 - 84 | 0.3 | 1.1 | 0.09 | 0.3 | 0.9 | 0.07 | 0.4 | 0.8 | 0.06 |
| 85 - 89 | 0.2 | 0.7 | 0.08 | 0.2 | 0.5 | 0.04 | 0.5 | 0.7 | 0.03 |
| **Race** | | | | | | | | | |
| Black or African American | 26.2 | 26.5 | 0.01 | 26.7 | 24.9 | -0.04 | 29.8 | 26.6 | -0.07 |
| White | 60.6 | 57.0 | -0.07 | 59.9 | 59.3 | -0.01 | 56.8 | 57.1 | 0.01 |
| **Ethnicity** | | | | | | | | | |
| Hispanic or Latino | 2.6 | 3.0 | 0.02 | 2.7 | 3.6 | 0.06 | 2.4 | 3.2 | 0.05 |
| **Medical history: General** | | | | | | | | | |
| Acute respiratory disease | 41.1 | 40.8 | -0.01 | 41.0 | 41.2 | 0.00 | 41.3 | 40.7 | -0.01 |
| Attention deficit hyperactivity disorder | 2.3 | 1.9 | -0.03 | 2.2 | 2.4 | 0.01 | 2.0 | 2.1 | 0.01 |
| Chronic liver disease | 3.4 | 4.9 | 0.07 | 3.5 | 3.1 | -0.02 | 3.8 | 4.0 | 0.01 |
| Chronic obstructive lung disease | 18.3 | 20.2 | 0.05 | 18.4 | 17.7 | -0.02 | 18.6 | 19.2 | 0.01 |
| Crohn’s disease | 0.4 | 0.4 | 0.00 | 0.4 | 0.3 | -0.02 | 0.3 | 0.4 | 0.02 |
| Dementia | 0.9 | 1.9 | 0.09 | 0.9 | 1.6 | 0.06 | 0.8 | 1.9 | 0.09 |
| Depressive disorder | 40.1 | 38.1 | -0.04 | 39.9 | 40.9 | 0.02 | 38.0 | 39.1 | 0.02 |
| Gastroesophageal reflux disease | 30.7 | 29.7 | -0.02 | 30.7 | 31.3 | 0.01 | 30.8 | 30.8 | 0.00 |
| Gastrointestinal hemorrhage | 3.7 | 3.7 | 0.00 | 3.7 | 3.6 | 0.00 | 3.6 | 3.8 | 0.01 |
| Human immunodeficiency virus infection | 0.7 | 0.6 | 0.00 | 0.6 | 0.5 | -0.02 | 0.7 | 0.7 | 0.00 |
| Hyperlipidemia | 62.8 | 64.7 | 0.04 | 62.9 | 62.1 | -0.02 | 61.9 | 64.1 | 0.05 |
| Hypertensive disorder | 72.8 | 75.1 | 0.05 | 72.9 | 72.4 | -0.01 | 73.8 | 74.3 | 0.01 |
| Lesion of liver | 1.4 | 1.1 | -0.03 | 1.4 | 1.1 | -0.03 | 1.3 | 1.2 | -0.01 |
| Obesity | 47.3 | 39.6 | -0.16 | 46.5 | 48.8 | 0.05 | 42.5 | 43.5 | 0.02 |
| Osteoarthritis | 38.7 | 38.5 | 0.00 | 38.6 | 39.8 | 0.03 | 37.3 | 38.6 | 0.03 |
| Pneumonia | 5.5 | 6.0 | 0.02 | 5.2 | 5.7 | 0.03 | 5.2 | 5.7 | 0.03 |
| Psoriasis | 1.7 | 1.2 | -0.04 | 1.7 | 1.3 | -0.03 | 1.4 | 1.3 | 0.00 |
| Renal impairment | 5.9 | 8.1 | 0.09 | 6.1 | 5.8 | -0.01 | 6.8 | 7.8 | 0.04 |
| Rheumatoid arthritis | 2.3 | 2.8 | 0.03 | 2.4 | 2.7 | 0.02 | 2.5 | 2.6 | 0.01 |
| Schizophrenia | 2.7 | 3.8 | 0.06 | 2.7 | 2.8 | 0.01 | 2.9 | 3.4 | 0.03 |
| Urinary tract infectious disease | 13.3 | 16.5 | 0.09 | 13.4 | 14.0 | 0.02 | 13.4 | 14.0 | 0.02 |
| Viral hepatitis C | 1.6 | 1.7 | 0.01 | 1.6 | 1.4 | -0.02 | 1.5 | 1.7 | 0.02 |
| Visual system disorder | 46.0 | 44.9 | -0.02 | 46.0 | 43.9 | -0.04 | 47.5 | 45.6 | -0.04 |
| **Medical history: Cardiovascular disease** | | | | | | | | | |
| Atrial fibrillation | 2.1 | 3.2 | 0.07 | 2.1 | 2.1 | 0.00 | 2.2 | 2.9 | 0.05 |
| Cerebrovascular disease | 4.1 | 5.1 | 0.05 | 4.2 | 3.2 | -0.06 | 4.5 | 4.5 | 0.00 |
| Coronary arteriosclerosis | 8.5 | 10.5 | 0.07 | 8.6 | 8.1 | -0.02 | 9.1 | 9.4 | 0.01 |
| Heart disease | 23.8 | 27.5 | 0.09 | 24.0 | 22.0 | -0.05 | 25.2 | 24.9 | -0.01 |
| Heart failure | 6.5 | 8.1 | 0.06 | 6.6 | 6.4 | -0.01 | 7.1 | 7.4 | 0.01 |
| Ischemic heart disease | 5.9 | 7.2 | 0.05 | 6.0 | 5.5 | -0.02 | 6.0 | 6.2 | 0.01 |
| Peripheral vascular disease | 6.6 | 7.6 | 0.04 | 6.6 | 7.0 | 0.02 | 7.1 | 7.2 | 0.01 |
| Pulmonary embolism | 1.0 | 1.1 | 0.00 | 1.0 | 1.2 | 0.02 | 1.0 | 1.1 | 0.02 |
| Venous thrombosis | 1.4 | 1.7 | 0.03 | 1.4 | 1.2 | -0.01 | 1.2 | 1.5 | 0.03 |
| **Medical history: Neoplasms** | | | | | | | | | |
| Hematologic neoplasm | 0.8 | 0.8 | 0.00 | 0.8 | 0.4 | -0.05 | 0.8 | 0.7 | -0.01 |
| Malignant lymphoma | <0.2 | 0.3 | 0.03 | <0.2 | 0.3 | 0.03 | 0.3 | 0.3 | 0.00 |

| (Continued) | Before ajustment | | | After matching | | | After stratification | | |
|---|---|---|---|---|---|---|---|---|---|
| Characteristic | T (%) | C (%) | StdDiff | T (%) | C (%) | StdDiff | T (%) | C (%) | StdDiff |
| Malignant neoplasm of anorectum | <0.2 | 0.2 | 0.02 | <0.2 | 0.3 | 0.04 | <0.2 | 0.2 | 0.03 |
| Malignant neoplastic disease | 6.0 | 6.3 | 0.01 | 6.0 | 5.8 | -0.01 | 6.1 | 6.3 | 0.01 |
| Malignant tumor of breast | 2.6 | 2.6 | 0.00 | 2.6 | 2.1 | -0.03 | 3.0 | 2.6 | -0.02 |
| Malignant tumor of colon | <0.2 | 0.4 | 0.05 | <0.2 | 0.4 | 0.05 | <0.2 | 0.4 | 0.06 |
| Malignant tumor of lung | <0.2 | 0.3 | 0.03 | <0.2 | 0.2 | 0.03 | <0.2 | 0.3 | 0.04 |
| **Medication use** | | | | | | | | | |
| Agents acting on the renin-angiotensin system | 58.0 | 61.0 | 0.06 | 58.2 | 57.0 | -0.03 | 59.7 | 59.4 | -0.01 |
| Antibacterials for systemic use | 73.7 | 73.0 | -0.01 | 73.2 | 72.9 | -0.01 | 73.3 | 72.5 | -0.02 |
| Antidepressants | 59.3 | 57.7 | -0.03 | 59.0 | 60.2 | 0.02 | 56.8 | 57.9 | 0.02 |
| Antiepileptics | 44.4 | 42.8 | -0.03 | 44.1 | 44.5 | 0.01 | 42.7 | 43.8 | 0.02 |
| Antiinflammatory and antirheumatic products | 65.1 | 62.3 | -0.06 | 64.7 | 65.8 | 0.03 | 62.8 | 63.9 | 0.02 |
| Antineoplastic agents | 5.0 | 5.2 | 0.01 | 5.1 | 4.8 | -0.01 | 5.3 | 4.7 | -0.03 |
| Antipsoriatics | 0.7 | 0.5 | -0.02 | 0.7 | 0.6 | -0.01 | 0.5 | 0.5 | 0.00 |
| Antithrombotic agents | 21.3 | 23.7 | 0.06 | 21.5 | 20.2 | -0.03 | 21.9 | 22.7 | 0.02 |
| Beta blocking agents | 30.8 | 33.2 | 0.05 | 31.1 | 30.1 | -0.02 | 33.4 | 31.4 | -0.04 |
| Calcium channel blockers | 20.9 | 24.5 | 0.09 | 21.4 | 21.1 | -0.01 | 24.9 | 23.5 | -0.03 |
| Diuretics | 46.2 | 47.5 | 0.03 | 46.1 | 47.3 | 0.02 | 46.4 | 45.7 | -0.01 |
| Drugs for acid related disorders | 51.0 | 52.5 | 0.03 | 51.0 | 51.9 | 0.02 | 51.2 | 52.3 | 0.02 |
| Drugs for obstructive airway diseases | 64.1 | 62.3 | -0.04 | 64.0 | 63.3 | -0.01 | 63.9 | 63.1 | -0.02 |
| Immunosuppressants | 2.7 | 3.1 | 0.02 | 2.7 | 3.4 | 0.04 | 2.4 | 3.2 | 0.05 |
| Lipid modifying agents | 61.5 | 65.5 | 0.08 | 61.6 | 61.3 | -0.01 | 62.9 | 64.0 | 0.02 |
| Opioids | 47.9 | 49.0 | 0.02 | 47.7 | 45.3 | -0.05 | 47.9 | 47.2 | -0.01 |
| Psycholeptics | 51.3 | 50.2 | -0.02 | 51.0 | 51.4 | 0.01 | 49.3 | 49.4 | 0.00 |
| Psychostimulants | 6.2 | 6.0 | -0.01 | 6.1 | 6.2 | 0.00 | 6.1 | 6.2 | 0.01 |

##### 2.2.6 Open Claims

**Supplementary Table 168:** Baseline patient characteristics for SGLT2I (female) (T) and DPP4I (female) (C) new-users in the Open Claims data source. We report proportion of initiators satisfying selected base-line characteristics and the standardized difference of population proportions (StdDiff) before and after propensity score adjustment. Less extreme StdDiffs through matching and stratification suggest improved balance between patient cohorts through adjustment.

| | Before adjustment | | | After matching | | | After stratification | | |
|---|---|---|---|---|---|---|---|---|---|
| Characteristic | T (%) | C (%) | StdDiff | T (%) | C (%) | StdDiff | T (%) | C (%) | StdDiff |
| **Age group** | | | | | | | | | |
| 15 - 19 | 0.1 | 0.1 | 0.00 | 0.1 | 0.1 | 0.00 | 0.1 | 0.1 | 0.00 |
| 20 - 24 | 0.5 | 0.4 | -0.01 | 0.5 | 0.5 | 0.00 | 0.5 | 0.5 | 0.00 |
| 25 - 29 | 1.0 | 0.8 | -0.02 | 0.9 | 0.9 | 0.00 | 0.9 | 0.9 | 0.00 |
| 30 - 34 | 1.9 | 1.6 | -0.03 | 1.9 | 1.9 | 0.00 | 1.7 | 1.7 | 0.00 |
| 35 - 39 | 3.6 | 3.0 | -0.03 | 3.5 | 3.6 | 0.00 | 3.5 | 3.6 | 0.00 |
| 40 - 44 | 6.3 | 5.0 | -0.06 | 6.2 | 6.3 | 0.00 | 5.6 | 5.6 | 0.00 |
| 45 - 49 | 9.8 | 7.9 | -0.07 | 9.6 | 9.7 | 0.01 | 8.9 | 8.6 | -0.01 |
| 50 - 54 | 13.9 | 11.5 | -0.07 | 12.6 | 12.4 | -0.01 | 12.6 | 12.4 | -0.01 |
| 55 - 59 | 16.7 | 13.9 | -0.08 | 16.2 | 16.5 | 0.01 | 15.1 | 15.0 | 0.00 |
| 60 - 64 | 16.3 | 14.7 | -0.04 | 16.0 | 16.4 | 0.01 | 15.4 | 15.5 | 0.00 |
| 65 - 69 | 12.8 | 14.3 | 0.04 | 13.1 | 13.0 | 0.00 | 13.5 | 13.6 | 0.00 |
| 70 - 74 | 8.8 | 11.2 | 0.08 | 9.1 | 8.9 | -0.01 | 10.1 | 10.1 | 0.00 |
| 75 - 79 | 5.2 | 10.8 | 0.21 | 5.7 | 5.1 | -0.03 | 8.1 | 8.4 | 0.01 |
| 80 - 84 | 2.9 | 4.7 | 0.09 | 3.3 | 3.2 | 0.00 | 4.0 | 4.0 | 0.00 |
| 85 - 89 | 0.2 | 0.1 | -0.03 | 0.2 | 0.2 | 0.01 | 0.2 | 0.2 | 0.01 |
| **Medical history: General** | | | | | | | | | |
| Acute respiratory disease | 16.3 | 17.2 | 0.02 | 16.6 | 16.7 | 0.00 | 17.3 | 16.8 | -0.01 |
| Attention deficit hyperactivity disorder | 0.6 | 0.4 | -0.03 | 0.6 | 0.5 | 0.00 | 0.5 | 0.5 | -0.01 |
| Chronic liver disease | 1.1 | 1.6 | 0.04 | 1.2 | 1.2 | 0.00 | 1.5 | 1.4 | -0.01 |
| Chronic obstructive lung disease | 4.5 | 5.8 | 0.06 | 4.5 | 4.4 | -0.01 | 5.4 | 5.2 | -0.01 |
| Crohn’s disease | 0.2 | 0.2 | 0.00 | 0.2 | 0.2 | -0.01 | 0.2 | 0.2 | 0.00 |
| Dementia | 0.5 | 1.2 | 0.08 | 0.5 | 0.5 | 0.00 | 1.0 | 0.9 | 0.00 |
| Gastroesophageal reflux disease | 10.0 | 10.5 | 0.02 | 10.0 | 9.8 | -0.01 | 10.7 | 10.2 | -0.02 |
| Gastrointestinal hemorrhage | 1.2 | 1.5 | 0.03 | 1.2 | 1.2 | 0.00 | 1.4 | 1.4 | 0.00 |
| Human immunodeficiency virus infection | 0.2 | 0.2 | 0.01 | 0.2 | 0.2 | 0.01 | 0.2 | 0.2 | 0.01 |
| Hyperlipidemia | 42.8 | 44.4 | 0.03 | 42.8 | 42.5 | 0.00 | 44.2 | 43.7 | -0.01 |
| Hypertensive disorder | 48.2 | 51.7 | 0.07 | 48.5 | 47.9 | -0.01 | 50.6 | 50.1 | -0.01 |
| Lesion of liver | 0.6 | 0.7 | 0.01 | 0.6 | 0.6 | 0.00 | 0.6 | 0.6 | 0.00 |
| Obesity | 14.4 | 12.1 | -0.07 | 13.9 | 13.8 | 0.00 | 13.5 | 13.0 | -0.01 |
| Osteoarthritis | 15.5 | 18.0 | 0.07 | 15.9 | 15.6 | -0.01 | 17.5 | 16.9 | -0.02 |
| Pneumonia | 2.1 | 2.6 | 0.03 | 2.0 | 2.0 | 0.00 | 2.3 | 2.4 | 0.00 |
| Psoriasis | 1.1 | 0.9 | -0.01 | 1.0 | 1.0 | 0.00 | 1.0 | 1.0 | 0.00 |
| Renal impairment | 4.8 | 7.1 | 0.10 | 4.7 | 4.4 | -0.02 | 5.8 | 6.0 | 0.01 |
| Rheumatoid arthritis | 1.3 | 1.6 | 0.02 | 1.3 | 1.4 | 0.00 | 1.5 | 1.5 | 0.00 |
| Schizophrenia | 0.3 | 0.4 | 0.03 | 0.3 | 0.3 | 0.00 | 0.3 | 0.4 | 0.01 |
| Ulcerative colitis | 0.2 | 0.2 | 0.01 | 0.2 | 0.2 | 0.00 | 0.2 | 0.2 | 0.00 |
| Urinary tract infectious disease | 6.4 | 8.9 | 0.10 | 6.7 | 6.6 | -0.01 | 8.2 | 7.9 | -0.01 |
| Viral hepatitis C | 0.2 | 0.3 | 0.02 | 0.2 | 0.3 | 0.01 | 0.3 | 0.3 | 0.00 |
| Visual system disorder | 18.4 | 21.2 | 0.07 | 18.9 | 18.7 | 0.00 | 20.3 | 20.1 | -0.01 |
| **Medical history: Cardiovascular disease** | | | | | | | | | |
| Atrial fibrillation | 2.7 | 3.4 | 0.04 | 2.5 | 2.4 | -0.01 | 3.0 | 3.0 | 0.00 |
| Cerebrovascular disease | 2.4 | 3.6 | 0.07 | 2.5 | 2.4 | -0.01 | 3.2 | 3.1 | 0.00 |
| Coronary arteriosclerosis | 5.7 | 6.2 | 0.02 | 5.2 | 5.0 | -0.01 | 5.9 | 5.7 | -0.01 |
| Heart disease | 14.8 | 16.6 | 0.05 | 13.9 | 13.4 | -0.02 | 15.8 | 15.3 | -0.01 |
| Heart failure | 4.0 | 4.0 | 0.00 | 3.7 | 3.6 | -0.01 | 3.7 | 3.6 | -0.01 |
| Ischemic heart disease | 3.2 | 3.3 | 0.00 | 2.9 | 2.7 | -0.02 | 3.3 | 3.1 | -0.01 |
| Peripheral vascular disease | 3.9 | 4.5 | 0.03 | 4.2 | 4.3 | 0.00 | 4.2 | 4.3 | 0.00 |
| Pulmonary embolism | 0.5 | 0.5 | 0.01 | 0.4 | 0.5 | 0.00 | 0.5 | 0.5 | 0.00 |
| Venous thrombosis | 0.7 | 0.9 | 0.03 | 0.6 | 0.7 | 0.00 | 0.8 | 0.8 | 0.00 |
| **Medical history: Neoplasms** | | | | | | | | | |
| Hematologic neoplasm | 0.5 | 0.7 | 0.02 | 0.5 | 0.5 | 0.00 | 0.6 | 0.6 | 0.00 |
| Malignant lymphoma | 0.3 | 0.3 | 0.01 | 0.3 | 0.3 | 0.00 | 0.3 | 0.3 | 0.00 |
| Malignant neoplasm of anorectum | 0.1 | 0.1 | 0.01 | 0.1 | 0.1 | 0.01 | 0.1 | 0.1 | 0.01 |
| Malignant neoplastic disease | 5.0 | 6.1 | 0.05 | 5.1 | 5.1 | 0.00 | 5.7 | 5.8 | 0.00 |
| Malignant tumor of breast | 2.1 | 2.5 | 0.03 | 2.3 | 2.3 | 0.00 | 2.3 | 2.3 | 0.00 |
| Malignant tumor of colon | 0.2 | 0.3 | 0.03 | 0.2 | 0.2 | 0.01 | 0.2 | 0.3 | 0.01 |
| Malignant tumor of lung | 0.2 | 0.3 | 0.02 | 0.2 | 0.2 | 0.01 | 0.2 | 0.2 | 0.01 |

| (Continued) | Before ajustment | | | After matching | | | After stratification | | |
|---|---|---|---|---|---|---|---|---|---|
| Characteristic | T (%) | C (%) | StdDiff | T (%) | C (%) | StdDiff | T (%) | C (%) | StdDiff |
| Malignant tumor of urinary bladder | 0.1 | 0.1 | 0.02 | 0.1 | 0.1 | 0.01 | 0.1 | 0.1 | 0.00 |
| **Medication use** | | | | | | | | | |
| Agents acting on the renin-angiotensin system | 64.6 | 67.0 | 0.05 | 64.5 | 64.2 | -0.01 | 65.6 | 65.7 | 0.00 |
| Antibacterials for systemic use | 61.0 | 62.8 | 0.04 | 61.7 | 61.9 | 0.01 | 62.7 | 62.3 | -0.01 |
| Antidepressants | 38.3 | 36.9 | -0.03 | 37.7 | 37.8 | 0.00 | 37.7 | 37.4 | -0.01 |
| Antiepileptics | 21.9 | 23.1 | 0.03 | 21.8 | 21.9 | 0.00 | 22.9 | 22.6 | -0.01 |
| Antiinflammatory and antirheumatic products | 40.4 | 41.0 | 0.01 | 40.5 | 41.1 | 0.01 | 41.3 | 41.0 | -0.01 |
| Antineoplastic agents | 5.2 | 5.5 | 0.01 | 5.2 | 5.2 | 0.00 | 5.6 | 5.4 | -0.01 |
| Antipsoriatics | 1.5 | 1.3 | -0.02 | 1.4 | 1.4 | 0.00 | 1.4 | 1.4 | 0.00 |
| Antithrombotic agents | 15.3 | 18.1 | 0.07 | 15.0 | 14.5 | -0.01 | 16.8 | 16.6 | -0.01 |
| Beta blocking agents | 31.4 | 33.9 | 0.05 | 30.8 | 30.0 | -0.02 | 32.5 | 32.3 | -0.01 |
| Calcium channel blockers | 23.1 | 26.3 | 0.07 | 23.4 | 23.1 | -0.01 | 24.6 | 24.9 | 0.01 |
| Diuretics | 43.4 | 44.7 | 0.03 | 42.9 | 42.4 | -0.01 | 43.7 | 43.7 | 0.00 |
| Drugs for acid related disorders | 36.4 | 39.5 | 0.06 | 36.5 | 36.5 | 0.00 | 38.7 | 38.3 | -0.01 |
| Drugs for obstructive airway diseases | 43.2 | 43.5 | 0.00 | 43.1 | 43.2 | 0.00 | 44.0 | 43.4 | -0.01 |
| Immunosuppressants | 3.5 | 3.8 | 0.01 | 3.8 | 3.6 | -0.01 | 3.8 | 3.6 | -0.01 |
| Lipid modifying agents | 68.4 | 69.5 | 0.02 | 68.0 | 67.7 | -0.01 | 68.8 | 68.8 | 0.00 |
| Opioids | 25.7 | 28.4 | 0.06 | 26.4 | 26.5 | 0.00 | 27.9 | 27.3 | -0.01 |
| Psycholeptics | 26.4 | 27.9 | 0.03 | 26.4 | 26.7 | 0.01 | 27.5 | 27.4 | 0.00 |
| Psychostimulants | 3.5 | 3.0 | -0.03 | 3.4 | 3.2 | -0.01 | 3.4 | 3.2 | -0.01 |

##### 2.2.7 OEHR

**Supplementary Table 169:** Baseline patient characteristics for SGLT2I (female) (T) and DPP4I (female) (C) new-users in the OEHR data source. We report proportion of initiators satisfying selected base-line characteristics and the standardized difference of population proportions (StdDiff) before and after propensity score adjustment. Less extreme StdDiffs through matching and stratification suggest improved balance between patient cohorts through adjustment.

| | Before adjustment | | | After matching | | | After stratification | | |
|---|---|---|---|---|---|---|---|---|---|
| Characteristic | T (%) | C (%) | StdDiff | T (%) | C (%) | StdDiff | T (%) | C (%) | StdDiff |
| **Age group** | | | | | | | | | |
| 15 - 19 | 0.1 | 0.1 | -0.01 | 0.1 | 0.1 | -0.01 | 0.1 | 0.1 | -0.01 |
| 20 - 24 | 0.6 | 0.5 | -0.01 | 0.6 | 0.6 | 0.00 | 0.5 | 0.5 | 0.00 |
| 25 - 29 | 1.2 | 1.1 | -0.01 | 1.2 | 1.2 | 0.00 | 1.1 | 1.2 | 0.01 |
| 30 - 34 | 2.3 | 2.2 | -0.01 | 2.3 | 2.7 | 0.02 | 2.2 | 2.4 | 0.01 |
| 35 - 39 | 4.7 | 4.0 | -0.04 | 4.7 | 4.5 | -0.01 | 4.5 | 4.2 | -0.01 |
| 40 - 44 | 7.8 | 6.8 | -0.04 | 7.7 | 7.5 | -0.01 | 7.7 | 7.5 | -0.01 |
| 45 - 49 | 11.8 | 9.9 | -0.06 | 11.4 | 11.8 | 0.01 | 10.0 | 10.4 | 0.01 |
| 50 - 54 | 16.4 | 13.8 | -0.07 | 16.0 | 16.1 | 0.00 | 14.3 | 14.5 | 0.00 |
| 55 - 59 | 18.1 | 15.7 | -0.06 | 17.8 | 17.6 | -0.01 | 16.8 | 16.3 | -0.01 |
| 60 - 64 | 16.5 | 14.8 | -0.05 | 16.3 | 17.0 | 0.02 | 14.6 | 15.6 | 0.03 |
| 65 - 69 | 10.1 | 12.2 | 0.07 | 10.6 | 10.4 | -0.01 | 11.6 | 11.5 | 0.00 |
| 70 - 74 | 5.9 | 8.6 | 0.11 | 6.3 | 6.1 | -0.01 | 8.0 | 7.8 | -0.01 |
| 75 - 79 | 2.9 | 5.7 | 0.14 | 3.2 | 3.0 | -0.01 | 4.7 | 4.6 | -0.01 |
| 80 - 84 | 1.2 | 3.7 | 0.16 | 1.3 | 1.1 | -0.02 | 3.7 | 3.0 | -0.04 |
| 85 - 89 | 0.4 | 1.0 | 0.07 | 0.4 | 0.5 | 0.01 | 0.9 | 0.9 | 0.00 |
| **Race** | | | | | | | | | |
| Asian | 2.6 | 3.2 | 0.04 | 2.7 | 3.0 | 0.01 | 2.8 | 3.1 | 0.02 |
| Black or African American | 13.8 | 15.2 | 0.04 | 14.2 | 14.2 | 0.00 | 14.8 | 15.0 | 0.00 |
| White | 75.5 | 73.9 | -0.04 | 74.8 | 74.7 | 0.00 | 74.5 | 74.1 | -0.01 |
| **Ethnicity** | | | | | | | | | |
| Hispanic or Latino | 6.8 | 7.0 | 0.01 | 6.9 | 6.5 | -0.01 | 6.8 | 7.0 | 0.01 |
| Not Hispanic or Latino | 85.1 | 85.6 | 0.01 | 84.8 | 85.1 | 0.01 | 85.4 | 85.1 | -0.01 |
| **Medical history: General** | | | | | | | | | |
| Acute respiratory disease | 23.8 | 22.2 | -0.04 | 23.3 | 23.0 | -0.01 | 23.4 | 22.8 | -0.02 |
| Attention deficit hyperactivity disorder | 1.1 | 0.8 | -0.03 | 1.1 | 0.9 | -0.02 | 1.0 | 0.9 | -0.01 |
| Chronic liver disease | 2.2 | 3.1 | 0.06 | 2.3 | 2.4 | 0.00 | 2.4 | 2.8 | 0.03 |
| Chronic obstructive lung disease | 4.6 | 5.4 | 0.04 | 4.7 | 4.7 | 0.00 | 5.6 | 5.3 | -0.01 |
| Crohn's disease | 0.2 | 0.3 | 0.00 | 0.3 | 0.3 | 0.00 | 0.3 | 0.3 | 0.00 |
| Dementia | 0.4 | 1.0 | 0.07 | 0.4 | 0.5 | 0.01 | 0.7 | 0.9 | 0.03 |
| Depressive disorder | 20.5 | 18.7 | -0.04 | 19.9 | 19.5 | -0.01 | 19.8 | 19.5 | -0.01 |
| Gastroesophageal reflux disease | 20.1 | 19.7 | -0.01 | 19.8 | 19.3 | -0.01 | 20.9 | 20.2 | -0.02 |
| Gastrointestinal hemorrhage | 1.1 | 1.4 | 0.03 | 1.1 | 1.1 | 0.00 | 1.2 | 1.3 | 0.01 |
| Human immunodeficiency virus infection | 0.1 | 0.2 | 0.01 | 0.1 | 0.2 | 0.01 | 0.1 | 0.2 | 0.01 |
| Hyperlipidemia | 63.0 | 64.6 | 0.03 | 62.6 | 62.4 | 0.00 | 62.6 | 62.4 | 0.00 |
| Hypertensive disorder | 63.6 | 65.1 | 0.03 | 63.3 | 63.0 | -0.01 | 65.1 | 64.8 | -0.01 |
| Lesion of liver | 0.8 | 0.7 | -0.01 | 0.8 | 0.7 | -0.02 | 0.9 | 0.7 | -0.03 |
| Obesity | 35.6 | 27.9 | -0.17 | 34.0 | 34.7 | 0.01 | 31.7 | 31.4 | -0.01 |
| Osteoarthritis | 17.3 | 18.8 | 0.04 | 17.2 | 17.3 | 0.00 | 19.3 | 18.6 | -0.02 |
| Pneumonia | 1.8 | 2.1 | 0.02 | 1.9 | 2.0 | 0.01 | 1.9 | 2.0 | 0.01 |
| Psoriasis | 1.7 | 1.3 | -0.03 | 1.6 | 1.5 | -0.01 | 1.6 | 1.4 | -0.01 |
| Renal impairment | 3.8 | 6.2 | 0.11 | 4.0 | 4.0 | 0.00 | 6.0 | 5.6 | -0.02 |
| Rheumatoid arthritis | 1.8 | 1.6 | -0.02 | 1.7 | 1.5 | -0.02 | 1.7 | 1.6 | 0.00 |
| Schizophrenia | 0.2 | 0.3 | 0.03 | 0.2 | 0.2 | 0.02 | 0.2 | 0.3 | 0.03 |
| Ulcerative colitis | 0.2 | 0.2 | 0.01 | 0.2 | 0.3 | 0.01 | 0.2 | 0.3 | 0.01 |
| Urinary tract infectious disease | 6.3 | 8.7 | 0.09 | 6.5 | 6.3 | -0.01 | 8.9 | 7.9 | -0.04 |
| Viral hepatitis C | 0.3 | 0.4 | 0.01 | 0.3 | 0.3 | 0.00 | 0.3 | 0.4 | 0.01 |
| Visual system disorder | 12.1 | 13.6 | 0.04 | 12.3 | 12.0 | -0.01 | 13.3 | 13.2 | -0.01 |
| **Medical history: Cardiovascular disease** | | | | | | | | | |
| Atrial fibrillation | 2.1 | 3.3 | 0.07 | 2.2 | 2.2 | 0.00 | 3.1 | 3.0 | -0.01 |
| Cerebrovascular disease | 2.0 | 2.7 | 0.04 | 2.1 | 2.3 | 0.01 | 2.5 | 2.6 | 0.00 |
| Coronary arteriosclerosis | 6.5 | 6.4 | 0.00 | 6.3 | 6.0 | -0.01 | 6.5 | 6.3 | -0.01 |
| Heart disease | 15.3 | 17.0 | 0.05 | 15.1 | 14.7 | -0.01 | 16.4 | 16.5 | 0.00 |
| Heart failure | 2.6 | 2.8 | 0.01 | 2.5 | 2.2 | -0.02 | 2.8 | 2.6 | -0.01 |
| Ischemic heart disease | 3.4 | 3.3 | 0.00 | 3.2 | 3.2 | 0.00 | 3.4 | 3.3 | 0.00 |
| Peripheral vascular disease | 3.9 | 3.6 | -0.01 | 3.7 | 3.6 | -0.01 | 4.2 | 3.7 | -0.02 |
| Pulmonary embolism | 0.4 | 0.5 | 0.01 | 0.4 | 0.4 | -0.01 | 0.7 | 0.5 | -0.03 |
| Venous thrombosis | 0.8 | 1.0 | 0.02 | 0.8 | 0.8 | 0.00 | 0.9 | 0.9 | 0.01 |

| (Continued) | Before ajustment | | | After matching | | | After stratification | | |
|---|---|---|---|---|---|---|---|---|---|
| Characteristic | T (%) | C (%) | StdDiff | T (%) | C (%) | StdDiff | T (%) | C (%) | StdDiff |
| **Medical history: Neoplasms** | | | | | | | | | |
| Hematologic neoplasm | 0.5 | 0.7 | 0.02 | 0.6 | 0.6 | 0.01 | 0.6 | 0.7 | 0.02 |
| Malignant lymphoma | 0.3 | 0.4 | 0.01 | 0.3 | 0.3 | -0.01 | 0.4 | 0.4 | -0.01 |
| Malignant neoplasm of anorectum | 0.1 | 0.1 | 0.01 | 0.1 | 0.1 | 0.02 | 0.1 | 0.1 | -0.01 |
| Malignant neoplastic disease | 5.5 | 6.6 | 0.05 | 5.5 | 5.7 | 0.01 | 6.6 | 6.3 | -0.01 |
| Malignant tumor of breast | 2.0 | 2.7 | 0.04 | 2.0 | 2.2 | 0.01 | 2.4 | 2.5 | 0.00 |
| Malignant tumor of colon | 0.2 | 0.3 | 0.01 | 0.2 | 0.2 | 0.00 | 0.2 | 0.2 | 0.00 |
| Malignant tumor of lung | 0.1 | 0.2 | 0.01 | 0.1 | 0.1 | -0.01 | 0.2 | 0.2 | -0.02 |
| Malignant tumor of urinary bladder | 0.1 | 0.1 | 0.00 | 0.1 | 0.1 | -0.01 | 0.1 | 0.1 | 0.00 |
| **Medication use** | | | | | | | | | |
| Agents acting on the renin-angiotensin system | 61.3 | 62.0 | 0.02 | 61.4 | 61.8 | 0.01 | 61.4 | 61.8 | 0.01 |
| Antibacterials for systemic use | 49.2 | 49.4 | 0.00 | 48.8 | 49.3 | 0.01 | 50.3 | 49.3 | -0.02 |
| Antidepressants | 41.4 | 38.4 | -0.06 | 40.6 | 39.5 | -0.02 | 40.6 | 39.5 | -0.02 |
| Antiepileptics | 20.7 | 20.3 | -0.01 | 20.5 | 20.8 | 0.01 | 20.8 | 20.9 | 0.00 |
| Antiinflammatory and antirheumatic products | 56.3 | 56.5 | 0.00 | 56.1 | 56.3 | 0.00 | 56.3 | 56.6 | 0.01 |
| Antineoplastic agents | 4.8 | 5.0 | 0.01 | 4.7 | 4.9 | 0.01 | 5.1 | 5.0 | 0.00 |
| Antipsoriatics | 0.5 | 0.5 | 0.00 | 0.5 | 0.4 | -0.01 | 0.6 | 0.5 | -0.02 |
| Antithrombotic agents | 32.0 | 36.1 | 0.09 | 32.4 | 31.8 | -0.01 | 34.8 | 34.6 | 0.00 |
| Beta blocking agents | 29.7 | 31.2 | 0.03 | 29.6 | 29.3 | -0.01 | 31.8 | 30.8 | -0.02 |
| Calcium channel blockers | 18.9 | 21.2 | 0.06 | 19.1 | 19.3 | 0.00 | 20.8 | 20.6 | -0.01 |
| Diuretics | 41.0 | 42.4 | 0.03 | 40.7 | 40.5 | 0.00 | 41.7 | 41.6 | 0.00 |
| Drugs for acid related disorders | 41.6 | 43.7 | 0.04 | 41.7 | 41.5 | 0.00 | 44.4 | 43.1 | -0.03 |
| Drugs for obstructive airway diseases | 48.4 | 47.4 | -0.02 | 48.0 | 48.2 | 0.00 | 48.7 | 48.0 | -0.01 |
| Immunosuppressants | 3.3 | 3.2 | -0.01 | 3.3 | 3.2 | 0.00 | 3.2 | 3.3 | 0.00 |
| Lipid modifying agents | 65.9 | 67.0 | 0.03 | 65.7 | 66.0 | 0.01 | 66.6 | 66.6 | 0.00 |
| Opioids | 27.7 | 30.0 | 0.05 | 27.8 | 27.6 | 0.00 | 30.4 | 29.4 | -0.02 |
| Psycholeptics | 31.5 | 32.5 | 0.02 | 31.4 | 31.3 | 0.00 | 32.9 | 32.3 | -0.01 |
| Psychostimulants | 4.5 | 3.9 | -0.03 | 4.5 | 4.3 | 0.00 | 4.1 | 4.2 | 0.00 |

##### 2.2.8 SIDIAP

**Supplementary Table 170:** Baseline patient characteristics for SGLT2I (female) (T) and DPP4I (female) (C) new-users in the SIDIAP data source. We report proportion of initiators satisfying selected base-line characteristics and the standardized difference of population proportions (StdDiff) before and after propensity score adjustment. Less extreme StdDiffs through matching and stratification suggest improved balance between patient cohorts through adjustment.

| | Before adjustment | | | After matching | | | After stratification | | |
|---|---|---|---|---|---|---|---|---|---|
| Characteristic | T (%) | C (%) | StdDiff | T (%) | C (%) | StdDiff | T (%) | C (%) | StdDiff |
| **Age group** | | | | | | | | | |
| 25 - 29 | <0.2 | 0.1 | 0.01 | <0.2 | 0.2 | 0.02 | <0.1 | 0.1 | 0.02 |
| 30 - 34 | 0.5 | 0.3 | -0.04 | 0.6 | <0.2 | -0.07 | 1.0 | 0.3 | -0.10 |
| 35 - 39 | 0.9 | 0.7 | -0.02 | 0.9 | 0.5 | -0.05 | 1.0 | 0.6 | -0.05 |
| 40 - 44 | 3.6 | 2.0 | -0.09 | 3.6 | 3.5 | -0.01 | 2.5 | 2.2 | -0.02 |
| 45 - 49 | 6.2 | 3.7 | -0.12 | 6.2 | 5.9 | -0.01 | 4.2 | 4.0 | -0.01 |
| 50 - 54 | 9.7 | 7.1 | -0.10 | 9.9 | 10.0 | 0.00 | 7.3 | 7.3 | 0.00 |
| 55 - 59 | 15.0 | 10.2 | -0.15 | 14.7 | 15.9 | 0.03 | 10.4 | 10.5 | 0.00 |
| 60 - 64 | 15.7 | 12.4 | -0.10 | 15.6 | 16.7 | 0.03 | 14.0 | 12.5 | -0.04 |
| 65 - 69 | 17.1 | 14.4 | -0.07 | 17.0 | 17.2 | 0.00 | 14.6 | 14.3 | -0.01 |
| 70 - 74 | 13.3 | 15.4 | 0.06 | 13.5 | 13.2 | -0.01 | 15.1 | 15.4 | 0.01 |
| 75 - 79 | 10.0 | 13.3 | 0.10 | 10.0 | 9.4 | -0.02 | 11.6 | 12.8 | 0.04 |
| 80 - 84 | 5.0 | 10.9 | 0.22 | 5.2 | 4.7 | -0.02 | 10.8 | 10.4 | -0.01 |
| 85 - 89 | 2.0 | 6.5 | 0.23 | 2.0 | 1.8 | -0.01 | 5.7 | 6.6 | 0.04 |
| 90 - 94 | 0.5 | 2.4 | 0.15 | 1.4 | 2.5 | 0.08 | 1.4 | 2.5 | 0.08 |
| 95 - 99 | <0.2 | 0.4 | 0.07 | <0.2 | 0.4 | 0.05 | 0.2 | 0.5 | 0.06 |
| **Medical history: General** | | | | | | | | | |
| Acute respiratory disease | 17.4 | 14.4 | -0.08 | 17.3 | 17.3 | 0.00 | 16.0 | 16.2 | 0.00 |
| Chronic liver disease | <0.2 | 0.4 | 0.05 | <0.2 | 0.2 | 0.02 | 0.1 | 0.3 | 0.04 |
| Chronic obstructive lung disease | 0.8 | 0.8 | 0.00 | 0.9 | 0.6 | -0.03 | 0.8 | 0.7 | -0.01 |
| Dementia | 0.2 | 0.8 | 0.09 | 0.2 | 0.3 | 0.02 | 1.0 | 0.8 | -0.02 |
| Depressive disorder | 2.2 | 3.4 | 0.07 | 2.2 | 2.4 | 0.01 | 2.7 | 3.0 | 0.01 |
| Gastroesophageal reflux disease | 1.8 | 1.5 | -0.02 | 1.7 | 2.0 | 0.02 | 2.4 | 1.8 | -0.04 |
| Gastrointestinal hemorrhage | 1.2 | 1.4 | 0.02 | 1.1 | 1.2 | 0.01 | 1.4 | 1.4 | 0.00 |
| Hyperlipidemia | 2.8 | 3.2 | 0.02 | 2.8 | 2.7 | -0.01 | 2.5 | 2.6 | 0.01 |
| Hypertensive disorder | 4.2 | 3.8 | -0.02 | 4.1 | 4.0 | 0.00 | 3.7 | 3.4 | -0.02 |
| Lesion of liver | 0.2 | 0.2 | 0.02 | 0.2 | <0.2 | -0.04 | 0.1 | 0.2 | 0.02 |
| Obesity | 8.8 | 8.3 | -0.02 | 8.9 | 7.7 | -0.04 | 8.4 | 8.0 | -0.01 |
| Osteoarthritis | 4.8 | 6.6 | 0.07 | 4.9 | 3.9 | -0.05 | 4.9 | 5.8 | 0.04 |
| Pneumonia | 1.0 | 1.4 | 0.04 | 1.0 | 1.1 | 0.01 | 0.9 | 1.5 | 0.06 |
| Psoriasis | 0.3 | 0.3 | 0.00 | 0.4 | 0.5 | 0.02 | 0.5 | 0.3 | -0.03 |
| Renal impairment | 1.5 | 4.2 | 0.17 | 1.5 | 1.3 | -0.02 | 3.2 | 4.1 | 0.05 |
| Rheumatoid arthritis | 0.2 | 0.1 | -0.03 | 0.3 | <0.2 | -0.04 | 0.2 | 0.1 | -0.02 |
| Urinary tract infectious disease | 10.1 | 10.6 | 0.01 | 10.2 | 9.9 | -0.01 | 10.4 | 12.0 | 0.05 |
| Viral hepatitis C | <0.2 | 0.3 | 0.05 | <0.2 | 0.2 | 0.04 | <0.1 | 0.2 | 0.05 |
| Visual system disorder | 14.1 | 18.0 | 0.11 | 14.3 | 13.7 | -0.02 | 16.9 | 18.0 | 0.03 |
| **Medical history: Cardiovascular disease** | | | | | | | | | |
| Atrial fibrillation | 1.3 | 1.7 | 0.03 | 1.2 | 1.1 | -0.01 | 1.4 | 1.6 | 0.02 |
| Cerebrovascular disease | 0.6 | 1.2 | 0.06 | 0.6 | 0.5 | -0.02 | 1.0 | 1.0 | 0.00 |
| Heart disease | 8.8 | 7.8 | -0.04 | 6.9 | 6.9 | 0.00 | 7.3 | 7.5 | 0.01 |
| Heart failure | 2.4 | 2.0 | -0.02 | 1.8 | 1.5 | -0.03 | 2.1 | 2.0 | -0.01 |
| Ischemic heart disease | 3.3 | 1.5 | -0.12 | 2.1 | 2.2 | 0.01 | 1.7 | 1.5 | -0.02 |
| Peripheral vascular disease | 0.6 | 0.5 | -0.02 | 0.6 | 0.5 | -0.01 | 0.5 | 0.5 | 0.01 |
| Pulmonary embolism | 0.2 | 0.3 | 0.03 | <0.2 | 0.3 | 0.02 | 0.6 | 0.3 | -0.03 |
| Venous thrombosis | 1.4 | 1.2 | -0.01 | 1.3 | 0.6 | -0.08 | 2.0 | 1.2 | -0.07 |
| **Medical history: Neoplasms** | | | | | | | | | |
| Hematologic neoplasm | <0.2 | 0.1 | 0.03 | <0.2 | 0.2 | 0.05 | <0.1 | 0.1 | 0.04 |
| Malignant neoplasm of anorectum | <0.2 | 0.1 | 0.01 | <0.2 | <0.2 | 0.01 | <0.1 | 0.1 | 0.02 |
| Malignant neoplastic disease | 2.1 | 2.8 | 0.05 | 2.0 | 2.1 | 0.00 | 2.0 | 2.1 | 0.00 |
| Malignant tumor of breast | 0.5 | 0.6 | 0.02 | 0.6 | 0.5 | 0.00 | 0.6 | 0.5 | 0.00 |
| Malignant tumor of colon | 0.2 | 0.3 | 0.01 | 0.2 | <0.2 | -0.04 | 0.5 | 0.3 | -0.04 |
| **Medication use** | | | | | | | | | |
| Agents acting on the renin-angiotensin system | 61.9 | 61.7 | -0.01 | 60.4 | 59.4 | -0.02 | 62.1 | 61.8 | -0.01 |
| Antibacterials for systemic use | 44.7 | 43.4 | -0.03 | 44.6 | 44.5 | 0.00 | 44.1 | 45.5 | 0.03 |
| Antidepressants | 35.8 | 33.0 | -0.06 | 35.3 | 35.5 | 0.00 | 32.9 | 34.3 | 0.03 |
| Antiepileptics | 14.7 | 12.3 | -0.07 | 14.6 | 14.9 | 0.01 | 12.9 | 13.1 | 0.00 |
| Antiinflammatory and antirheumatic products | 52.3 | 53.6 | 0.03 | 51.4 | 51.0 | -0.01 | 52.0 | 50.5 | -0.03 |
| Antineoplastic agents | 2.5 | 2.7 | 0.01 | 2.5 | 2.7 | 0.01 | 2.7 | 2.5 | -0.01 |

| (Continued) | Before ajustment | | | After matching | | | After stratification | | |
|---|---|---|---|---|---|---|---|---|---|
| Characteristic | T (%) | C (%) | StdDiff | T (%) | C (%) | StdDiff | T (%) | C (%) | StdDiff |
| Antipsoriatics | 1.9 | 1.7 | -0.01 | 1.9 | 1.7 | -0.01 | 1.1 | 1.8 | 0.06 |
| Antithrombotic agents | 30.6 | 31.5 | 0.02 | 28.5 | 27.8 | -0.02 | 27.1 | 30.7 | 0.08 |
| Beta blocking agents | 31.7 | 26.5 | -0.11 | 29.4 | 29.2 | 0.00 | 27.0 | 28.0 | 0.02 |
| Calcium channel blockers | 22.7 | 23.8 | 0.03 | 22.1 | 21.8 | -0.01 | 23.6 | 23.8 | 0.00 |
| Diuretics | 50.6 | 52.1 | 0.03 | 49.1 | 48.2 | -0.02 | 52.7 | 52.1 | -0.01 |
| Drugs for acid related disorders | 56.7 | 61.8 | 0.10 | 56.2 | 55.9 | -0.01 | 59.4 | 60.4 | 0.02 |
| Drugs for obstructive airway diseases | 31.6 | 28.3 | -0.07 | 31.1 | 30.2 | -0.02 | 31.1 | 30.2 | -0.02 |
| Immunosuppressants | 2.1 | 1.5 | -0.04 | 2.1 | 2.3 | 0.02 | 1.6 | 1.8 | 0.02 |
| Lipid modifying agents | 61.2 | 61.4 | 0.00 | 60.1 | 61.2 | 0.02 | 62.5 | 60.5 | -0.04 |
| Opioids | 26.2 | 25.2 | -0.02 | 26.0 | 27.4 | 0.03 | 27.1 | 26.8 | -0.01 |
| Psycholeptics | 41.5 | 43.9 | 0.05 | 41.6 | 42.6 | 0.02 | 45.5 | 44.1 | -0.03 |
| Psychostimulants | 3.7 | 2.8 | -0.05 | 3.0 | 2.6 | -0.02 | 2.1 | 2.4 | 0.02 |

#### 2.3 SGLT2I (female) vs SU (female)

##### 2.3.1 CCAE

**Supplementary Table 171:** Baseline patient characteristics for SGLT2I (female) (T) and SU (female) (C) new-users in the CCAE data source. We report proportion of initiators satisfying selected base-line characteristics and the standardized difference of population proportions (StdDiff) before and after propensity score adjustment. Less extreme StdDiffs through matching and stratification suggest improved balance between patient cohorts through adjustment.

| | Before adjustment | | | After matching | | | After stratification | | |
|---|---|---|---|---|---|---|---|---|---|
| Characteristic | T (%) | C (%) | StdDiff | T (%) | C (%) | StdDiff | T (%) | C (%) | StdDiff |
| **Age group** | | | | | | | | | |
| 15 - 19 | 0.1 | 0.1 | 0.02 | 0.1 | 0.1 | 0.01 | 0.2 | 0.1 | 0.01 |
| 20 - 24 | 0.6 | 0.4 | 0.02 | 0.6 | 0.5 | 0.02 | 0.7 | 0.5 | 0.02 |
| 25 - 29 | 0.8 | 1.0 | -0.03 | 0.8 | 0.8 | 0.00 | 0.9 | 1.0 | -0.01 |
| 30 - 34 | 2.1 | 2.7 | -0.04 | 2.1 | 2.1 | 0.00 | 2.2 | 2.5 | -0.02 |
| 35 - 39 | 4.8 | 5.1 | -0.01 | 4.8 | 4.6 | 0.01 | 5.0 | 5.0 | 0.00 |
| 40 - 44 | 8.9 | 8.4 | 0.02 | 8.6 | 8.4 | 0.01 | 8.6 | 8.4 | 0.01 |
| 45 - 49 | 14.9 | 13.6 | 0.04 | 14.7 | 14.6 | 0.00 | 14.3 | 13.8 | 0.01 |
| 50 - 54 | 20.3 | 20.0 | 0.01 | 20.4 | 20.2 | 0.00 | 20.2 | 19.8 | 0.01 |
| 55 - 59 | 24.3 | 23.5 | 0.02 | 24.0 | 24.7 | -0.02 | 23.5 | 24.0 | -0.01 |
| 60 - 64 | 21.5 | 23.1 | -0.04 | 22.2 | 22.0 | 0.00 | 22.6 | 22.9 | 0.00 |
| 65 - 69 | 1.6 | 2.1 | -0.04 | 1.7 | 1.8 | -0.01 | 1.9 | 2.0 | -0.01 |
| **Medical history: General** | | | | | | | | | |
| Acute respiratory disease | 33.9 | 30.8 | 0.07 | 32.7 | 33.1 | -0.01 | 32.7 | 32.4 | 0.01 |
| Attention deficit hyperactivity disorder | 1.3 | 0.5 | 0.08 | 1.2 | 1.1 | 0.01 | 1.1 | 1.0 | 0.01 |
| Chronic liver disease | 2.2 | 3.6 | -0.08 | 2.3 | 2.3 | 0.00 | 2.3 | 2.3 | 0.00 |
| Chronic obstructive lung disease | 2.6 | 3.1 | -0.03 | 2.6 | 2.6 | 0.00 | 2.8 | 3.0 | -0.01 |
| Crohn’s disease | 0.3 | 0.3 | 0.01 | 0.3 | 0.2 | 0.01 | 0.3 | 0.2 | 0.00 |
| Dementia | 0.2 | 0.2 | 0.01 | 0.2 | 0.2 | 0.01 | 0.3 | 0.2 | 0.02 |
| Depressive disorder | 15.2 | 11.9 | 0.10 | 15.0 | 15.2 | -0.01 | 14.9 | 15.0 | 0.00 |
| Gastroesophageal reflux disease | 15.6 | 10.7 | 0.15 | 15.1 | 15.4 | -0.01 | 14.7 | 14.7 | 0.00 |
| Gastrointestinal hemorrhage | 1.6 | 1.8 | -0.02 | 1.6 | 1.5 | 0.01 | 1.6 | 1.6 | 0.00 |
| Human immunodeficiency virus infection | 0.1 | 0.1 | 0.01 | 0.1 | 0.1 | -0.01 | 0.1 | 0.1 | 0.00 |
| Hyperlipidemia | 69.6 | 56.3 | 0.28 | 67.9 | 68.9 | -0.02 | 66.3 | 66.3 | 0.00 |
| Hypertensive disorder | 69.0 | 60.7 | 0.17 | 68.7 | 69.0 | -0.01 | 68.0 | 67.6 | 0.01 |
| Lesion of liver | 0.5 | 0.7 | -0.02 | 0.5 | 0.6 | -0.01 | 0.5 | 0.7 | -0.03 |
| Obesity | 30.9 | 17.5 | 0.32 | 29.7 | 30.2 | -0.01 | 28.0 | 28.4 | -0.01 |
| Osteoarthritis | 20.3 | 17.0 | 0.09 | 19.7 | 19.9 | 0.00 | 19.8 | 19.4 | 0.01 |
| Pneumonia | 2.2 | 2.6 | -0.03 | 2.2 | 2.2 | 0.00 | 2.6 | 2.4 | 0.01 |
| Psoriasis | 1.7 | 1.4 | 0.03 | 1.6 | 1.8 | -0.01 | 1.6 | 1.7 | -0.01 |
| Renal impairment | 2.7 | 3.1 | -0.02 | 2.8 | 2.7 | 0.00 | 3.7 | 3.6 | 0.01 |
| Rheumatoid arthritis | 1.6 | 1.4 | 0.02 | 1.5 | 1.4 | 0.01 | 1.6 | 1.5 | 0.01 |
| Schizophrenia | 0.1 | 0.1 | 0.00 | 0.1 | 0.1 | 0.02 | 0.1 | 0.1 | 0.01 |
| Ulcerative colitis | 0.3 | 0.3 | 0.01 | 0.4 | 0.3 | 0.01 | 0.3 | 0.3 | 0.01 |
| Urinary tract infectious disease | 10.3 | 11.3 | -0.03 | 10.2 | 9.9 | 0.01 | 10.7 | 10.6 | 0.00 |
| Viral hepatitis C | 0.2 | 0.3 | -0.03 | 0.2 | 0.2 | -0.01 | 0.2 | 0.3 | 0.00 |
| Visual system disorder | 26.0 | 24.1 | 0.04 | 26.0 | 26.0 | 0.00 | 26.3 | 26.0 | 0.01 |
| **Medical history: Cardiovascular disease** | | | | | | | | | |
| Cerebrovascular disease | 2.2 | 2.4 | -0.01 | 2.1 | 2.1 | 0.00 | 2.1 | 2.1 | 0.00 |
| Coronary arteriosclerosis | 4.2 | 4.5 | -0.01 | 4.1 | 4.0 | 0.00 | 4.3 | 4.2 | 0.00 |
| Heart disease | 14.0 | 13.5 | 0.02 | 13.2 | 13.1 | 0.00 | 13.6 | 13.2 | 0.01 |
| Heart failure | 1.9 | 2.0 | -0.01 | 1.7 | 1.8 | 0.00 | 1.7 | 1.9 | -0.02 |
| Ischemic heart disease | 2.8 | 3.2 | -0.02 | 2.7 | 2.5 | 0.01 | 2.9 | 2.7 | 0.01 |
| Peripheral vascular disease | 3.0 | 2.4 | 0.04 | 2.9 | 3.0 | 0.00 | 3.0 | 2.8 | 0.01 |
| Pulmonary embolism | 0.3 | 0.3 | 0.00 | 0.3 | 0.4 | -0.01 | 0.3 | 0.4 | -0.01 |
| Venous thrombosis | 0.8 | 0.8 | -0.01 | 0.8 | 0.8 | 0.00 | 0.9 | 0.9 | 0.01 |
| **Medical history: Neoplasms** | | | | | | | | | |
| Hematologic neoplasm | 0.7 | 0.8 | -0.01 | 0.7 | 0.8 | 0.00 | 0.8 | 0.9 | -0.01 |
| Malignant lymphoma | 0.3 | 0.3 | 0.00 | 0.3 | 0.3 | -0.01 | 0.3 | 0.3 | -0.01 |
| Malignant neoplasm of anorectum | 0.1 | 0.2 | -0.02 | 0.1 | 0.1 | -0.01 | 0.1 | 0.1 | -0.01 |
| Malignant neoplastic disease | 6.3 | 6.1 | 0.01 | 6.1 | 6.2 | 0.00 | 6.3 | 6.3 | 0.00 |
| Malignant tumor of breast | 2.5 | 2.5 | 0.00 | 2.5 | 2.5 | 0.00 | 2.7 | 2.5 | 0.01 |
| Malignant tumor of colon | 0.2 | 0.3 | -0.02 | 0.2 | 0.2 | -0.01 | 0.2 | 0.3 | -0.01 |
| Malignant tumor of lung | 0.1 | 0.2 | -0.01 | 0.2 | 0.1 | 0.02 | 0.1 | 0.2 | 0.00 |
| **Medication use** | | | | | | | | | |

| (Continued) | Before ajustment | | | After matching | | | After stratification | | |
|---|---|---|---|---|---|---|---|---|---|
| Characteristic | T (%) | C (%) | StdDiff | T (%) | C (%) | StdDiff | T (%) | C (%) | StdDiff |
| Agents acting on the renin-angiotensin system | 61.6 | 61.9 | -0.01 | 61.4 | 61.6 | 0.00 | 61.2 | 61.5 | -0.01 |
| Antibacterials for systemic use | 64.9 | 64.7 | 0.00 | 64.0 | 64.2 | 0.00 | 64.2 | 64.2 | 0.00 |
| Antidepressants | 37.5 | 33.8 | 0.08 | 36.1 | 36.4 | 0.00 | 35.2 | 35.3 | 0.00 |
| Antiepileptics | 16.1 | 13.4 | 0.08 | 16.2 | 15.6 | 0.01 | 16.2 | 15.6 | 0.01 |
| Antiinflammatory and antirheumatic products | 38.2 | 34.1 | 0.09 | 37.4 | 37.7 | -0.01 | 37.6 | 37.1 | 0.01 |
| Antineoplastic agents | 4.4 | 4.7 | -0.01 | 4.2 | 4.1 | 0.00 | 4.2 | 4.1 | 0.01 |
| Antipsoriatics | 0.6 | 0.6 | -0.01 | 0.5 | 0.6 | -0.01 | 0.5 | 0.6 | -0.01 |
| Antithrombotic agents | 7.8 | 7.4 | 0.01 | 7.7 | 7.6 | 0.00 | 7.9 | 8.1 | -0.01 |
| Beta blocking agents | 24.3 | 25.2 | -0.02 | 23.7 | 23.9 | 0.00 | 24.8 | 24.4 | 0.01 |
| Calcium channel blockers | 18.2 | 18.1 | 0.00 | 18.1 | 18.2 | 0.00 | 18.4 | 18.2 | 0.00 |
| Diuretics | 42.7 | 43.3 | -0.01 | 42.0 | 42.1 | 0.00 | 41.8 | 41.6 | 0.00 |
| Drugs for acid related disorders | 27.5 | 26.1 | 0.03 | 26.8 | 26.9 | 0.00 | 27.0 | 26.5 | 0.01 |
| Drugs for obstructive airway diseases | 44.0 | 39.3 | 0.10 | 42.9 | 43.1 | 0.00 | 43.3 | 42.4 | 0.02 |
| Immunosuppressants | 3.3 | 2.6 | 0.04 | 3.2 | 3.0 | 0.01 | 3.2 | 3.0 | 0.01 |
| Lipid modifying agents | 62.4 | 60.0 | 0.05 | 61.9 | 61.9 | 0.00 | 61.7 | 61.4 | 0.01 |
| Opioids | 27.6 | 29.3 | -0.04 | 27.2 | 26.9 | 0.01 | 28.6 | 28.1 | 0.01 |
| Psycholeptics | 26.6 | 25.7 | 0.02 | 26.0 | 25.9 | 0.00 | 26.6 | 26.0 | 0.01 |
| Psychostimulants | 4.2 | 3.1 | 0.06 | 3.8 | 3.8 | 0.00 | 3.5 | 3.6 | -0.01 |

##### 2.3.2 DAG

**Supplementary Table 172:** Baseline patient characteristics for SGLT2I (female) (T) and SU (female) (C) new-users in the DAG data source. We report proportion of initiators satisfying selected base-line characteristics and the standardized difference of population proportions (StdDiff) before and after propensity score adjustment. Less extreme StdDiffs through matching and stratification suggest improved balance between patient cohorts through adjustment.

| | Before adjustment | | | After matching | | | After stratification | | |
|---|---|---|---|---|---|---|---|---|---|
| Characteristic | T (%) | C (%) | StdDiff | T (%) | C (%) | StdDiff | T (%) | C (%) | StdDiff |
| **Age group** | | | | | | | | | |
| 20 - 24 | 0.2 | <0.2 | 0.04 | <0.7 | <0.2 | 0.04 | 0.2 | <0.2 | 0.06 |
| 25 - 29 | 0.3 | 0.2 | 0.02 | <0.7 | <0.2 | 0.07 | 0.3 | <0.2 | 0.05 |
| 30 - 34 | 1.1 | 0.4 | 0.08 | 1.3 | 0.4 | 0.10 | 1.5 | <0.2 | 0.14 |
| 35 - 39 | 1.8 | 1.0 | 0.06 | 2.3 | 1.3 | 0.08 | 1.9 | 1.5 | 0.03 |
| 40 - 44 | 2.7 | 2.2 | 0.03 | 2.9 | 2.5 | 0.03 | 2.8 | 3.8 | -0.06 |
| 45 - 49 | 6.1 | 4.1 | 0.09 | 5.6 | 4.5 | 0.05 | 5.6 | 7.2 | -0.07 |
| 50 - 54 | 9.8 | 6.8 | 0.11 | 9.4 | 9.3 | 0.00 | 9.6 | 10.9 | -0.04 |
| 55 - 59 | 13.4 | 9.5 | 0.12 | 12.2 | 10.8 | 0.04 | 12.2 | 10.8 | 0.05 |
| 60 - 64 | 15.5 | 12.6 | 0.08 | 13.8 | 12.8 | 0.03 | 14.9 | 12.3 | 0.07 |
| 65 - 69 | 15.8 | 16.3 | -0.01 | 15.4 | 16.2 | -0.02 | 15.9 | 18.7 | -0.07 |
| 70 - 74 | 12.2 | 17.0 | -0.14 | 11.6 | 12.7 | -0.03 | 11.8 | 11.0 | 0.03 |
| 75 - 79 | 10.0 | 15.6 | -0.17 | 11.9 | 12.9 | -0.03 | 11.1 | 10.3 | 0.03 |
| 80 - 84 | 7.7 | 9.7 | -0.07 | 8.5 | 10.2 | -0.06 | 8.2 | 7.6 | 0.03 |
| 85 - 89 | 2.8 | 3.8 | -0.05 | 4.0 | 4.7 | -0.04 | 3.3 | 4.2 | -0.04 |
| 90 - 94 | 0.6 | 0.8 | -0.03 | <0.7 | 1.6 | -0.11 | 0.5 | 1.5 | -0.10 |
| **Medical history: General** | | | | | | | | | |
| Acute respiratory disease | 18.0 | 19.5 | -0.04 | 20.7 | 18.2 | 0.06 | 19.1 | 16.3 | 0.07 |
| Chronic liver disease | 0.2 | 0.2 | 0.01 | <0.7 | 0.3 | -0.04 | 0.2 | <0.2 | 0.00 |
| Chronic obstructive lung disease | 4.8 | 6.6 | -0.08 | 7.0 | 5.8 | 0.05 | 5.3 | 6.9 | -0.07 |
| Crohn’s disease | <0.2 | <0.2 | 0.01 | <0.7 | 0.3 | -0.03 | <0.1 | 0.3 | -0.05 |
| Dementia | 1.3 | 2.7 | -0.10 | 1.6 | 2.2 | -0.04 | 1.4 | 1.9 | -0.04 |
| Depressive disorder | 11.3 | 12.6 | -0.04 | 13.2 | 13.0 | 0.00 | 12.5 | 13.9 | -0.04 |
| Gastroesophageal reflux disease | 2.3 | 2.9 | -0.03 | 2.9 | 2.3 | 0.04 | 2.6 | 1.6 | 0.07 |
| Gastrointestinal hemorrhage | 0.7 | 0.8 | -0.01 | <0.7 | 0.7 | -0.01 | 0.7 | 0.4 | 0.03 |
| Hyperlipidemia | 21.7 | 24.6 | -0.07 | 21.1 | 19.3 | 0.04 | 21.1 | 22.8 | -0.04 |
| Hypertensive disorder | 42.2 | 59.5 | -0.35 | 46.3 | 45.4 | 0.02 | 44.1 | 42.8 | 0.03 |
| Lesion of liver | 0.3 | 0.5 | -0.03 | <0.7 | 0.7 | -0.06 | 0.3 | 2.0 | -0.16 |
| Obesity | 11.2 | 9.1 | 0.07 | 11.0 | 9.6 | 0.05 | 11.4 | 10.0 | 0.05 |
| Osteoarthritis | 13.2 | 16.6 | -0.10 | 13.0 | 12.9 | 0.00 | 13.0 | 12.9 | 0.00 |
| Pneumonia | 1.2 | 2.0 | -0.06 | 1.0 | 1.3 | -0.03 | 1.3 | 1.5 | -0.02 |
| Psoriasis | 1.4 | 1.2 | 0.03 | 1.8 | 1.1 | 0.06 | 1.6 | 0.6 | 0.10 |
| Renal impairment | 5.0 | 4.7 | 0.01 | 4.7 | 5.3 | -0.03 | 5.0 | 4.6 | 0.02 |
| Rheumatoid arthritis | 2.0 | 1.8 | 0.01 | 2.2 | 1.6 | 0.04 | 2.1 | 0.9 | 0.10 |
| Schizophrenia | 0.3 | 0.3 | -0.01 | <0.7 | <0.2 | 0.04 | 0.2 | <0.2 | 0.04 |
| Urinary tract infectious disease | 7.2 | 9.5 | -0.09 | 6.9 | 7.8 | -0.03 | 7.6 | 6.9 | 0.02 |
| Viral hepatitis C | <0.2 | <0.2 | 0.02 | <0.1 | <0.2 | -0.03 | <0.1 | <0.2 | -0.03 |
| Visual system disorder | 7.7 | 10.6 | -0.10 | 8.2 | 9.3 | -0.04 | 7.9 | 7.3 | 0.02 |
| **Medical history: Cardiovascular disease** | | | | | | | | | |
| Atrial fibrillation | 2.3 | 2.3 | 0.00 | 1.8 | 1.6 | 0.01 | 2.0 | 1.5 | 0.04 |
| Cerebrovascular disease | 3.2 | 4.3 | -0.06 | 3.8 | 3.7 | 0.00 | 3.3 | 4.6 | -0.07 |
| Coronary arteriosclerosis | 4.0 | 2.8 | 0.07 | 3.2 | 2.8 | 0.03 | 3.9 | 3.7 | 0.01 |
| Heart disease | 21.2 | 27.7 | -0.15 | 20.1 | 18.0 | 0.05 | 20.8 | 20.3 | 0.01 |
| Heart failure | 7.1 | 9.6 | -0.09 | 6.5 | 6.4 | 0.00 | 7.4 | 5.8 | 0.06 |
| Ischemic heart disease | 8.1 | 12.9 | -0.16 | 8.2 | 9.5 | -0.04 | 8.2 | 9.5 | -0.04 |
| Peripheral vascular disease | 4.7 | 4.9 | -0.01 | 4.3 | 5.5 | -0.06 | 4.4 | 6.2 | -0.08 |
| Pulmonary embolism | 0.6 | 0.6 | -0.01 | <0.7 | 0.5 | 0.00 | 0.5 | 0.3 | 0.03 |
| Venous thrombosis | 1.4 | 1.8 | -0.03 | 1.8 | 1.9 | -0.01 | 1.3 | 3.4 | -0.14 |
| **Medical history: Neoplasms** | | | | | | | | | |
| Hematologic neoplasm | 0.2 | 0.3 | -0.01 | <0.7 | 0.3 | 0.00 | 0.3 | <0.2 | 0.02 |
| Malignant neoplasm of anorectum | <0.2 | 0.2 | -0.05 | <0.1 | <0.2 | -0.03 | <0.1 | <0.2 | -0.03 |
| Malignant neoplastic disease | 4.4 | 5.6 | -0.05 | 4.8 | 4.0 | 0.04 | 4.0 | 3.2 | 0.04 |
| Malignant tumor of breast | 1.4 | 2.1 | -0.05 | 1.2 | 1.6 | -0.03 | 1.2 | 1.1 | 0.01 |
| Malignant tumor of colon | 0.4 | 0.5 | -0.01 | 0.9 | 0.2 | 0.09 | 0.3 | <0.2 | 0.04 |
| Malignant tumor of lung | <0.2 | 0.2 | -0.04 | <0.7 | 0.2 | 0.01 | <0.1 | <0.2 | 0.00 |
| **Medication use** | | | | | | | | | |
| Agents acting on the renin-angiotensin system | 67.7 | 65.3 | 0.05 | 63.5 | 63.0 | 0.01 | 65.4 | 62.4 | 0.06 |
| Antibacterials for systemic use | 24.2 | 33.8 | -0.21 | 30.2 | 28.6 | 0.04 | 27.6 | 27.7 | 0.00 |

| (Continued) | Before ajustment | | | After matching | | | After stratification | | |
|---|---|---|---|---|---|---|---|---|---|
| Characteristic | T (%) | C (%) | StdDiff | T (%) | C (%) | StdDiff | T (%) | C (%) | StdDiff |
| Antidepressants | 14.2 | 13.1 | 0.03 | 14.7 | 13.1 | 0.05 | 14.3 | 15.0 | -0.02 |
| Antiepileptics | 4.7 | 3.7 | 0.05 | 6.3 | 4.1 | 0.10 | 5.1 | 3.9 | 0.06 |
| Antiinflammatory and antirheumatic products | 39.6 | 47.6 | -0.16 | 41.6 | 40.0 | 0.03 | 40.8 | 37.7 | 0.06 |
| Antineoplastic agents | 1.5 | 1.3 | 0.02 | 1.5 | 1.3 | 0.02 | 1.4 | 1.1 | 0.02 |
| Antipsoriatics | 0.5 | 0.2 | 0.05 | <0.7 | 0.2 | 0.05 | 0.7 | <0.2 | 0.09 |
| Antithrombotic agents | 26.8 | 27.0 | 0.00 | 23.0 | 23.0 | 0.00 | 26.0 | 21.9 | 0.10 |
| Beta blocking agents | 47.1 | 49.8 | -0.05 | 48.8 | 48.9 | 0.00 | 47.7 | 44.6 | 0.06 |
| Calcium channel blockers | 33.9 | 32.1 | 0.04 | 34.0 | 32.9 | 0.02 | 34.3 | 30.7 | 0.07 |
| Diuretics | 48.8 | 57.6 | -0.18 | 53.1 | 49.5 | 0.07 | 49.7 | 46.0 | 0.07 |
| Drugs for acid related disorders | 36.6 | 36.6 | 0.00 | 40.6 | 40.0 | 0.01 | 38.3 | 36.7 | 0.03 |
| Drugs for obstructive airway diseases | 18.3 | 17.9 | 0.01 | 18.6 | 18.1 | 0.01 | 18.6 | 18.1 | 0.01 |
| Immunosuppressants | 0.8 | 0.9 | 0.00 | 1.0 | 1.0 | 0.00 | 0.7 | 0.5 | 0.03 |
| Lipid modifying agents | 45.9 | 37.0 | 0.18 | 37.0 | 36.8 | 0.00 | 41.8 | 45.5 | -0.07 |
| Opioids | 12.9 | 14.7 | -0.05 | 13.9 | 13.1 | 0.02 | 13.6 | 11.8 | 0.05 |
| Psycholeptics | 9.5 | 14.4 | -0.15 | 10.7 | 11.4 | -0.02 | 10.0 | 9.8 | 0.01 |
| Psychostimulants | 0.2 | 0.4 | -0.05 | <0.7 | <0.2 | 0.02 | 0.4 | <0.2 | 0.07 |

##### 2.3.3 ODOD

**Supplementary Table 173:** Baseline patient characteristics for SGLT2I (female) (T) and SU (female) (C) new-users in the ODOD data source. We report proportion of initiators satisfying selected base-line characteristics and the standardized difference of population proportions (StdDiff) before and after propensity score adjustment. Less extreme StdDiffs through matching and stratification suggest improved balance between patient cohorts through adjustment.

| | Before adjustment | | | After matching | | | After stratification | | |
|---|---|---|---|---|---|---|---|---|---|
| Characteristic | T (%) | C (%) | StdDiff | T (%) | C (%) | StdDiff | T (%) | C (%) | StdDiff |
| **Age group** | | | | | | | | | |
| 25 - 29 | 0.4 | 0.5 | -0.01 | 0.4 | 0.4 | 0.00 | 0.5 | 0.4 | 0.01 |
| 30 - 34 | 1.3 | 1.4 | -0.01 | 1.4 | 1.1 | 0.02 | 1.2 | 1.2 | 0.00 |
| 35 - 39 | 3.1 | 2.5 | 0.04 | 3.0 | 3.0 | 0.00 | 2.4 | 2.3 | 0.00 |
| 40 - 44 | 5.4 | 3.9 | 0.07 | 5.1 | 5.3 | -0.01 | 3.8 | 3.8 | 0.00 |
| 45 - 49 | 8.9 | 6.1 | 0.10 | 8.5 | 8.9 | -0.01 | 6.2 | 6.2 | 0.00 |
| 50 - 54 | 11.7 | 8.8 | 0.09 | 11.4 | 11.7 | -0.01 | 8.6 | 8.7 | 0.00 |
| 55 - 59 | 14.5 | 10.8 | 0.11 | 14.0 | 14.7 | -0.02 | 11.7 | 11.1 | 0.02 |
| 60 - 64 | 13.6 | 11.2 | 0.07 | 13.4 | 13.5 | 0.00 | 11.1 | 11.3 | -0.01 |
| 65 - 69 | 16.5 | 17.5 | -0.03 | 17.1 | 17.2 | 0.00 | 17.9 | 18.6 | -0.02 |
| 70 - 74 | 14.2 | 16.9 | -0.07 | 14.9 | 14.2 | 0.02 | 18.1 | 17.7 | 0.01 |
| 75 - 79 | 6.6 | 10.9 | -0.15 | 6.9 | 6.4 | 0.02 | 10.3 | 10.0 | 0.01 |
| 80 - 84 | 2.5 | 6.2 | -0.18 | 2.6 | 2.3 | 0.02 | 5.3 | 5.3 | 0.00 |
| 85 - 89 | 1.0 | 2.9 | -0.14 | 1.0 | 1.0 | 0.01 | 2.7 | 2.8 | 0.00 |
| 90 - 94 | 0.1 | 0.2 | -0.01 | 0.1 | 0.1 | 0.00 | 0.1 | 0.2 | -0.02 |
| **Medical history: General** | | | | | | | | | |
| Acute respiratory disease | 30.4 | 29.5 | 0.02 | 29.9 | 30.3 | -0.01 | 30.0 | 29.2 | 0.02 |
| Attention deficit hyperactivity disorder | 1.3 | 0.5 | 0.08 | 1.2 | 1.0 | 0.02 | 0.8 | 0.7 | 0.01 |
| Chronic liver disease | 2.2 | 3.5 | -0.07 | 2.2 | 2.1 | 0.01 | 2.9 | 2.7 | 0.02 |
| Chronic obstructive lung disease | 8.2 | 10.2 | -0.07 | 8.2 | 8.2 | 0.00 | 10.5 | 10.2 | 0.01 |
| Crohn’s disease | 0.3 | 0.3 | 0.00 | 0.3 | 0.2 | 0.01 | 0.3 | 0.3 | 0.00 |
| Dementia | 1.0 | 2.8 | -0.13 | 1.1 | 1.1 | 0.00 | 2.9 | 2.5 | 0.02 |
| Depressive disorder | 21.8 | 19.1 | 0.07 | 21.4 | 21.8 | -0.01 | 21.8 | 21.4 | 0.01 |
| Gastroesophageal reflux disease | 22.9 | 20.6 | 0.06 | 22.6 | 22.3 | 0.01 | 23.5 | 22.5 | 0.02 |
| Gastrointestinal hemorrhage | 2.2 | 2.8 | -0.04 | 2.2 | 2.1 | 0.01 | 2.9 | 2.5 | 0.02 |
| Human immunodeficiency virus infection | 0.1 | 0.1 | -0.01 | 0.1 | 0.2 | -0.03 | 0.1 | 0.2 | -0.03 |
| Hyperlipidemia | 78.3 | 77.0 | 0.03 | 77.6 | 77.6 | 0.00 | 77.2 | 77.5 | -0.01 |
| Hypertensive disorder | 77.5 | 79.5 | -0.05 | 77.5 | 76.9 | 0.01 | 79.4 | 79.1 | 0.01 |
| Lesion of liver | 1.1 | 1.3 | -0.02 | 1.0 | 1.1 | 0.00 | 1.5 | 1.3 | 0.01 |
| Obesity | 34.9 | 24.3 | 0.23 | 33.4 | 34.5 | -0.02 | 28.7 | 29.3 | -0.01 |
| Osteoarthritis | 30.1 | 30.4 | 0.00 | 29.9 | 29.8 | 0.00 | 33.4 | 31.7 | 0.04 |
| Pneumonia | 3.5 | 4.6 | -0.05 | 3.6 | 3.4 | 0.01 | 4.6 | 4.5 | 0.00 |
| Psoriasis | 1.9 | 1.5 | 0.03 | 1.9 | 1.9 | 0.00 | 1.7 | 1.7 | 0.00 |
| Renal impairment | 10.3 | 15.9 | -0.17 | 10.6 | 9.7 | 0.03 | 16.2 | 15.8 | 0.01 |
| Schizophrenia | 0.3 | 0.5 | -0.03 | 0.3 | 0.4 | -0.02 | 0.3 | 0.5 | -0.04 |
| Ulcerative colitis | 0.4 | 0.4 | 0.00 | 0.4 | 0.3 | 0.01 | 0.5 | 0.3 | 0.03 |
| Urinary tract infectious disease | 12.1 | 16.6 | -0.13 | 12.3 | 11.8 | 0.01 | 15.5 | 15.1 | 0.01 |
| Viral hepatitis C | 0.3 | 0.5 | -0.02 | 0.3 | 0.4 | -0.01 | 0.4 | 0.5 | -0.01 |
| Visual system disorder | 40.7 | 42.9 | -0.04 | 40.9 | 40.1 | 0.01 | 43.6 | 43.5 | 0.00 |
| **Medical history: Cardiovascular disease** | | | | | | | | | |
| Atrial fibrillation | 4.4 | 5.8 | -0.06 | 4.3 | 4.1 | 0.01 | 5.7 | 5.6 | 0.01 |
| Cerebrovascular disease | 5.4 | 6.8 | -0.06 | 5.2 | 4.9 | 0.01 | 6.4 | 6.2 | 0.01 |
| Coronary arteriosclerosis | 11.1 | 11.7 | -0.02 | 10.8 | 10.5 | 0.01 | 11.6 | 11.8 | 0.00 |
| Heart disease | 25.7 | 28.0 | -0.05 | 25.1 | 24.3 | 0.02 | 27.4 | 27.4 | 0.00 |
| Heart failure | 6.6 | 7.5 | -0.04 | 6.2 | 5.7 | 0.02 | 7.3 | 7.4 | 0.00 |
| Ischemic heart disease | 6.7 | 7.0 | -0.01 | 6.4 | 6.4 | 0.00 | 6.7 | 6.9 | -0.01 |
| Peripheral vascular disease | 9.8 | 10.4 | -0.02 | 9.7 | 9.5 | 0.01 | 10.8 | 11.1 | -0.01 |
| Pulmonary embolism | 0.8 | 0.9 | 0.00 | 0.8 | 0.7 | 0.01 | 1.2 | 0.9 | 0.03 |
| Venous thrombosis | 1.3 | 1.8 | -0.04 | 1.3 | 1.3 | 0.00 | 1.6 | 1.7 | -0.01 |
| **Medical history: Neoplasms** | | | | | | | | | |
| Hematologic neoplasm | 0.8 | 1.4 | -0.06 | 0.8 | 1.0 | -0.01 | 1.0 | 1.3 | -0.02 |
| Malignant neoplasm of anorectum | 0.2 | 0.2 | -0.01 | 0.2 | 0.1 | 0.02 | 0.2 | 0.2 | 0.01 |
| Malignant neoplastic disease | 8.8 | 10.1 | -0.04 | 8.8 | 8.7 | 0.00 | 9.8 | 9.9 | 0.00 |
| Malignant tumor of breast | 3.2 | 3.8 | -0.04 | 3.2 | 3.2 | 0.00 | 3.4 | 3.7 | -0.02 |
| Malignant tumor of colon | 0.3 | 0.5 | -0.03 | 0.3 | 0.3 | 0.00 | 0.6 | 0.5 | 0.02 |
| Malignant tumor of lung | 0.3 | 0.5 | -0.03 | 0.3 | 0.3 | 0.01 | 0.5 | 0.5 | 0.01 |
| Malignant tumor of urinary bladder | 0.1 | 0.2 | -0.02 | 0.1 | 0.1 | 0.00 | 0.1 | 0.2 | -0.01 |
| **Medication use** | | | | | | | | | |

| (Continued) | Before ajustment | | | After matching | | | After stratification | | |
|---|---|---|---|---|---|---|---|---|---|
| Characteristic | T (%) | C (%) | StdDiff | T (%) | C (%) | StdDiff | T (%) | C (%) | StdDiff |
| Agents acting on the renin-angiotensin system | 67.5 | 69.1 | -0.04 | 67.8 | 67.0 | 0.02 | 69.1 | 69.1 | 0.00 |
| Antibacterials for systemic use | 63.5 | 63.1 | 0.01 | 62.9 | 62.7 | 0.00 | 63.7 | 62.5 | 0.02 |
| Antidepressants | 39.3 | 34.8 | 0.09 | 38.6 | 38.6 | 0.00 | 36.7 | 36.8 | 0.00 |
| Antiepileptics | 20.6 | 19.7 | 0.02 | 20.6 | 20.7 | 0.00 | 22.9 | 22.0 | 0.02 |
| Antiinflammatory and antirheumatic products | 37.0 | 32.5 | 0.09 | 36.3 | 36.9 | -0.01 | 35.4 | 34.6 | 0.02 |
| Antineoplastic agents | 5.4 | 5.6 | -0.01 | 5.6 | 5.6 | 0.00 | 5.6 | 5.6 | 0.00 |
| Antipsoriatics | 0.6 | 0.6 | 0.00 | 0.6 | 0.6 | -0.01 | 0.5 | 0.6 | -0.02 |
| Antithrombotic agents | 13.9 | 14.5 | -0.02 | 13.6 | 13.4 | 0.01 | 15.2 | 14.9 | 0.01 |
| Beta blocking agents | 33.0 | 35.9 | -0.06 | 32.8 | 32.2 | 0.01 | 35.7 | 35.4 | 0.01 |
| Calcium channel blockers | 24.4 | 27.7 | -0.08 | 24.6 | 24.3 | 0.01 | 28.4 | 27.6 | 0.02 |
| Diuretics | 45.9 | 48.7 | -0.06 | 45.6 | 45.2 | 0.01 | 47.0 | 47.3 | -0.01 |
| Drugs for acid related disorders | 31.0 | 31.8 | -0.02 | 30.7 | 30.6 | 0.00 | 33.7 | 33.0 | 0.01 |
| Drugs for obstructive airway diseases | 45.0 | 40.9 | 0.08 | 44.3 | 44.6 | -0.01 | 44.2 | 43.1 | 0.02 |
| Immunosuppressants | 3.8 | 3.2 | 0.04 | 3.7 | 3.6 | 0.00 | 3.7 | 3.5 | 0.01 |
| Lipid modifying agents | 72.2 | 70.5 | 0.04 | 72.2 | 72.1 | 0.00 | 72.2 | 72.1 | 0.00 |
| Opioids | 29.1 | 30.3 | -0.03 | 28.7 | 28.3 | 0.01 | 31.4 | 29.6 | 0.04 |
| Psycholeptics | 28.6 | 27.8 | 0.02 | 28.2 | 27.9 | 0.01 | 30.2 | 28.5 | 0.04 |
| Psychostimulants | 2.9 | 1.9 | 0.06 | 2.6 | 2.7 | 0.00 | 2.3 | 2.1 | 0.02 |

##### 2.3.4 MDCR

**Supplementary Table 174:** Baseline patient characteristics for SGLT2I (female) (T) and SU (female) (C) new-users in the MDCR data source. We report proportion of initiators satisfying selected base-line characteristics and the standardized difference of population proportions (StdDiff) before and after propensity score adjustment. Less extreme StdDiffs through matching and stratification suggest improved balance between patient cohorts through adjustment.

| | Before adjustment | | | After matching | | | After stratification | | |
|---|---|---|---|---|---|---|---|---|---|
| Characteristic | T (%) | C (%) | StdDiff | T (%) | C (%) | StdDiff | T (%) | C (%) | StdDiff |
| **Age group** | | | | | | | | | |
| 50 - 54 | <0.3 | 0.1 | 0.01 | <0.4 | 0.1 | 0.03 | <0.2 | 0.1 | 0.02 |
| 55 - 59 | 0.5 | 0.6 | -0.01 | 0.5 | 0.8 | -0.03 | 0.5 | 0.6 | -0.02 |
| 60 - 64 | 1.4 | 1.1 | 0.03 | 1.4 | 1.4 | 0.00 | 1.1 | 1.5 | -0.03 |
| 65 - 69 | 40.8 | 26.5 | 0.31 | 39.5 | 36.2 | 0.07 | 34.4 | 31.0 | 0.07 |
| 70 - 74 | 32.6 | 28.2 | 0.10 | 32.3 | 34.6 | -0.05 | 28.9 | 29.2 | -0.01 |
| 75 - 79 | 16.7 | 21.2 | -0.12 | 17.3 | 17.6 | -0.01 | 16.2 | 18.7 | -0.06 |
| 80 - 84 | 5.3 | 13.3 | -0.28 | 5.9 | 5.5 | 0.02 | 7.7 | 10.8 | -0.11 |
| 85 - 89 | 1.9 | 6.5 | -0.23 | 2.1 | 2.4 | -0.02 | 7.2 | 5.8 | 0.06 |
| 90 - 94 | 0.5 | 2.1 | -0.14 | 0.5 | 1.2 | -0.07 | 3.8 | 2.0 | 0.10 |
| 95 - 99 | <0.3 | 0.3 | -0.05 | <0.4 | 0.2 | -0.03 | <0.2 | 0.3 | -0.06 |
| **Medical history: General** | | | | | | | | | |
| Acute respiratory disease | 25.6 | 22.8 | 0.06 | 25.4 | 26.0 | -0.01 | 26.1 | 25.6 | 0.01 |
| Attention deficit hyperactivity disorder | <0.3 | 0.1 | -0.01 | <0.4 | 0.5 | -0.08 | <0.2 | 0.2 | -0.06 |
| Chronic liver disease | 2.6 | 2.4 | 0.02 | 2.4 | 1.8 | 0.04 | 3.1 | 2.5 | 0.04 |
| Chronic obstructive lung disease | 9.8 | 10.3 | -0.01 | 9.8 | 10.1 | -0.01 | 8.5 | 11.0 | -0.08 |
| Crohn’s disease | <0.3 | 0.3 | -0.02 | <0.4 | 0.4 | -0.03 | <0.2 | 0.4 | -0.06 |
| Dementia | 1.2 | 3.4 | -0.15 | 1.4 | 1.7 | -0.03 | 2.6 | 4.1 | -0.08 |
| Depressive disorder | 13.6 | 9.0 | 0.14 | 13.7 | 14.5 | -0.03 | 14.5 | 14.3 | 0.00 |
| Gastroesophageal reflux disease | 20.5 | 11.6 | 0.24 | 19.1 | 19.6 | -0.01 | 16.3 | 18.4 | -0.06 |
| Gastrointestinal hemorrhage | 3.0 | 3.6 | -0.04 | 3.1 | 2.8 | 0.02 | 2.2 | 3.2 | -0.06 |
| Hyperlipidemia | 79.0 | 48.0 | 0.68 | 78.1 | 80.1 | -0.05 | 77.6 | 74.3 | 0.08 |
| Hypertensive disorder | 83.2 | 68.1 | 0.36 | 82.9 | 83.3 | -0.01 | 83.9 | 82.4 | 0.04 |
| Lesion of liver | 1.2 | 1.2 | 0.00 | 0.8 | 1.0 | -0.01 | 0.6 | 1.2 | -0.06 |
| Obesity | 26.0 | 8.4 | 0.48 | 24.5 | 25.3 | -0.02 | 17.5 | 19.0 | -0.04 |
| Osteoarthritis | 35.9 | 29.8 | 0.13 | 35.0 | 36.3 | -0.03 | 41.1 | 36.5 | 0.09 |
| Pneumonia | 3.3 | 5.8 | -0.12 | 2.8 | 2.5 | 0.02 | 6.1 | 4.7 | 0.06 |
| Psoriasis | 2.5 | 1.3 | 0.09 | 2.6 | 1.9 | 0.04 | 1.9 | 1.9 | 0.00 |
| Renal impairment | 9.7 | 11.6 | -0.06 | 10.3 | 9.7 | 0.02 | 9.6 | 16.2 | -0.20 |
| Rheumatoid arthritis | 1.8 | 2.1 | -0.02 | 1.7 | 2.2 | -0.04 | 1.6 | 2.5 | -0.07 |
| Schizophrenia | <0.3 | 0.2 | -0.03 | <0.2 | 0.1 | -0.02 | <0.2 | 0.1 | -0.02 |
| Ulcerative colitis | 0.6 | 0.4 | 0.03 | 0.6 | 0.3 | 0.05 | 0.4 | 0.3 | 0.01 |
| Urinary tract infectious disease | 11.6 | 15.0 | -0.10 | 11.5 | 11.6 | 0.00 | 12.7 | 15.9 | -0.09 |
| Viral hepatitis C | <0.3 | 0.3 | -0.02 | <0.4 | 0.2 | -0.03 | <0.2 | 0.3 | -0.05 |
| Visual system disorder | 56.6 | 50.0 | 0.13 | 56.4 | 56.5 | 0.00 | 51.4 | 55.2 | -0.07 |
| **Medical history: Cardiovascular disease** | | | | | | | | | |
| Atrial fibrillation | 7.7 | 9.5 | -0.06 | 7.9 | 7.9 | 0.00 | 11.3 | 9.8 | 0.05 |
| Cerebrovascular disease | 10.6 | 11.4 | -0.03 | 10.1 | 8.8 | 0.04 | 12.2 | 10.4 | 0.06 |
| Coronary arteriosclerosis | 17.9 | 17.0 | 0.03 | 16.7 | 17.2 | -0.01 | 15.9 | 17.0 | -0.03 |
| Heart disease | 37.6 | 37.9 | -0.01 | 35.7 | 36.0 | -0.01 | 40.5 | 37.4 | 0.06 |
| Heart failure | 8.7 | 9.9 | -0.04 | 8.3 | 6.6 | 0.06 | 13.4 | 9.4 | 0.13 |
| Ischemic heart disease | 9.7 | 8.9 | 0.03 | 8.9 | 8.2 | 0.03 | 7.4 | 8.0 | -0.02 |
| Peripheral vascular disease | 12.2 | 10.1 | 0.06 | 11.5 | 11.7 | 0.00 | 8.2 | 11.9 | -0.12 |
| Pulmonary embolism | 0.8 | 0.9 | -0.01 | 0.8 | 0.9 | -0.02 | 0.8 | 0.9 | -0.02 |
| Venous thrombosis | 2.1 | 2.3 | -0.01 | 2.2 | 1.8 | 0.03 | 2.0 | 2.6 | -0.04 |
| **Medical history: Neoplasms** | | | | | | | | | |
| Hematologic neoplasm | 0.7 | 1.6 | -0.09 | 0.8 | 1.2 | -0.05 | 0.7 | 1.8 | -0.10 |
| Malignant lymphoma | 0.5 | 0.7 | -0.02 | 0.5 | 0.6 | -0.02 | 0.5 | 0.8 | -0.04 |
| Malignant neoplasm of anorectum | 0.3 | 0.2 | 0.02 | 0.4 | 0.1 | 0.05 | 0.6 | 0.2 | 0.06 |
| Malignant neoplastic disease | 14.0 | 14.1 | 0.00 | 14.3 | 14.3 | 0.00 | 12.5 | 15.1 | -0.08 |
| Malignant tumor of breast | 6.0 | 5.7 | 0.01 | 6.1 | 6.0 | 0.00 | 5.1 | 6.1 | -0.04 |
| Malignant tumor of colon | 0.7 | 0.7 | 0.00 | 0.8 | 0.4 | 0.05 | 0.8 | 0.6 | 0.03 |
| Malignant tumor of lung | 0.4 | 0.7 | -0.04 | 0.5 | 0.6 | -0.03 | <0.2 | 0.7 | -0.08 |
| Malignant tumor of urinary bladder | <0.3 | 0.5 | -0.03 | <0.4 | 0.4 | -0.01 | <0.2 | 0.4 | -0.04 |
| **Medication use** | | | | | | | | | |
| Agents acting on the renin-angiotensin system | 73.7 | 72.0 | 0.04 | 74.2 | 73.8 | 0.01 | 74.6 | 74.1 | 0.01 |
| Antibacterials for systemic use | 62.0 | 64.3 | -0.05 | 62.2 | 62.2 | 0.00 | 63.0 | 64.5 | -0.03 |
| Antidepressants | 34.1 | 31.5 | 0.06 | 34.1 | 33.5 | 0.01 | 28.4 | 32.8 | -0.10 |

| (Continued) | Before ajustment | | | After matching | | | After stratification | | |
|---|---|---|---|---|---|---|---|---|---|
| Characteristic | T (%) | C (%) | StdDiff | T (%) | C (%) | StdDiff | T (%) | C (%) | StdDiff |
| Antiepileptics | 19.5 | 16.4 | 0.08 | 19.5 | 18.3 | 0.03 | 18.8 | 18.7 | 0.00 |
| Antiinflammatory and antirheumatic products | 35.3 | 29.2 | 0.13 | 34.1 | 34.9 | -0.02 | 34.9 | 30.9 | 0.09 |
| Antineoplastic agents | 8.0 | 9.4 | -0.05 | 8.1 | 7.8 | 0.01 | 7.6 | 8.0 | -0.01 |
| Antipsoriatics | 1.1 | 0.7 | 0.04 | 1.0 | 1.1 | -0.01 | 0.6 | 0.9 | -0.04 |
| Antithrombotic agents | 18.3 | 21.8 | -0.09 | 18.2 | 18.1 | 0.00 | 21.9 | 20.8 | 0.03 |
| Beta blocking agents | 44.7 | 47.4 | -0.06 | 44.8 | 44.4 | 0.01 | 52.1 | 47.0 | 0.10 |
| Calcium channel blockers | 32.9 | 35.2 | -0.05 | 33.5 | 32.2 | 0.03 | 40.2 | 34.5 | 0.12 |
| Diuretics | 53.9 | 59.2 | -0.11 | 54.3 | 52.3 | 0.04 | 54.9 | 55.3 | -0.01 |
| Drugs for acid related disorders | 39.1 | 35.2 | 0.08 | 37.8 | 37.8 | 0.00 | 32.0 | 35.8 | -0.08 |
| Drugs for obstructive airway diseases | 49.7 | 40.0 | 0.20 | 48.5 | 49.4 | -0.02 | 41.7 | 45.2 | -0.07 |
| Immunosuppressants | 4.8 | 3.5 | 0.06 | 4.9 | 4.2 | 0.03 | 4.3 | 4.4 | -0.01 |
| Lipid modifying agents | 80.7 | 73.6 | 0.17 | 80.6 | 81.3 | -0.02 | 82.2 | 77.8 | 0.11 |
| Opioids | 27.2 | 30.8 | -0.08 | 27.6 | 27.4 | 0.00 | 33.7 | 29.8 | 0.09 |
| Psycholeptics | 27.2 | 29.4 | -0.05 | 27.5 | 27.0 | 0.01 | 29.3 | 28.3 | 0.02 |
| Psychostimulants | 2.0 | 1.4 | 0.04 | 2.3 | 1.9 | 0.03 | 1.5 | 1.5 | 0.00 |

##### 2.3.5 MDCD

**Supplementary Table 175:** Baseline patient characteristics for SGLT2I (female) (T) and SU (female) (C) new-users in the MDCD data source. We report proportion of initiators satisfying selected base-line characteristics and the standardized difference of population proportions (StdDiff) before and after propensity score adjustment. Less extreme StdDiffs through matching and stratification suggest improved balance between patient cohorts through adjustment.

| | Before adjustment | | | After matching | | | After stratification | | |
|---|---|---|---|---|---|---|---|---|---|
| Characteristic | T (%) | C (%) | StdDiff | T (%) | C (%) | StdDiff | T (%) | C (%) | StdDiff |
| **Age group** | | | | | | | | | |
| 15 - 19 | 0.6 | 0.6 | 0.00 | 0.6 | 0.5 | 0.01 | 0.5 | 0.6 | -0.01 |
| 20 - 24 | 1.0 | 1.4 | -0.04 | 1.0 | 1.1 | -0.01 | 1.8 | 1.4 | 0.03 |
| 25 - 29 | 3.9 | 3.6 | 0.02 | 3.9 | 3.1 | 0.04 | 3.1 | 3.3 | -0.01 |
| 30 - 34 | 7.2 | 6.4 | 0.03 | 7.2 | 6.6 | 0.03 | 6.2 | 6.4 | -0.01 |
| 35 - 39 | 10.7 | 9.3 | 0.05 | 10.8 | 10.4 | 0.01 | 10.2 | 10.0 | 0.00 |
| 40 - 44 | 14.0 | 11.5 | 0.07 | 14.0 | 13.8 | 0.00 | 12.6 | 12.2 | 0.01 |
| 45 - 49 | 14.4 | 13.0 | 0.04 | 14.3 | 14.2 | 0.00 | 15.1 | 13.4 | 0.05 |
| 50 - 54 | 15.9 | 15.3 | 0.02 | 16.0 | 15.4 | 0.02 | 15.9 | 15.1 | 0.02 |
| 55 - 59 | 16.1 | 16.1 | 0.00 | 16.0 | 16.4 | -0.01 | 16.3 | 16.8 | -0.01 |
| 60 - 64 | 11.8 | 13.2 | -0.04 | 11.8 | 12.2 | -0.01 | 12.9 | 13.2 | -0.01 |
| 65 - 69 | 2.4 | 4.3 | -0.11 | 2.4 | 2.9 | -0.03 | 2.9 | 3.6 | -0.04 |
| 70 - 74 | 1.2 | 2.2 | -0.08 | 1.2 | 1.7 | -0.04 | 1.6 | 1.8 | -0.02 |
| 75 - 79 | 0.2 | 1.5 | -0.14 | 0.3 | 0.8 | -0.07 | 0.2 | 1.1 | -0.12 |
| 80 - 84 | 0.3 | 1.0 | -0.08 | 0.3 | 0.5 | -0.03 | 0.3 | 0.6 | -0.05 |
| 85 - 89 | 0.2 | 0.6 | -0.06 | 0.2 | 0.2 | -0.01 | 0.5 | 0.4 | 0.02 |
| **Race** | | | | | | | | | |
| Black or African American | 26.2 | 30.9 | -0.10 | 26.6 | 23.8 | 0.06 | 32.5 | 29.8 | 0.06 |
| White | 60.6 | 51.1 | 0.19 | 60.1 | 62.4 | -0.05 | 53.2 | 52.9 | 0.01 |
| **Ethnicity** | | | | | | | | | |
| Hispanic or Latino | 2.6 | 2.7 | 0.00 | 2.7 | 2.7 | 0.00 | 2.3 | 3.0 | -0.04 |
| **Medical history: General** | | | | | | | | | |
| Acute respiratory disease | 41.1 | 41.0 | 0.00 | 41.2 | 42.4 | -0.02 | 40.3 | 41.1 | -0.02 |
| Attention deficit hyperactivity disorder | 2.3 | 1.5 | 0.06 | 2.3 | 2.5 | -0.02 | 1.6 | 1.9 | -0.03 |
| Chronic liver disease | 3.4 | 5.7 | -0.11 | 3.4 | 3.4 | 0.00 | 4.4 | 4.8 | -0.02 |
| Chronic obstructive lung disease | 18.3 | 18.5 | -0.01 | 18.2 | 18.1 | 0.00 | 17.7 | 18.5 | -0.02 |
| Crohn's disease | 0.4 | 0.3 | 0.02 | 0.4 | 0.3 | 0.01 | 0.4 | 0.3 | 0.01 |
| Dementia | 0.9 | 1.5 | -0.06 | 0.9 | 1.1 | -0.02 | 0.8 | 1.4 | -0.06 |
| Depressive disorder | 40.1 | 36.5 | 0.07 | 39.8 | 41.0 | -0.02 | 40.2 | 39.0 | 0.02 |
| Gastroesophageal reflux disease | 30.7 | 27.1 | 0.08 | 30.4 | 31.4 | -0.02 | 31.2 | 29.4 | 0.04 |
| Gastrointestinal hemorrhage | 3.7 | 3.8 | 0.00 | 3.8 | 3.3 | 0.02 | 3.4 | 3.6 | -0.01 |
| Human immunodeficiency virus infection | 0.7 | 0.6 | 0.00 | 0.7 | 0.5 | 0.03 | 0.7 | 0.6 | 0.01 |
| Hyperlipidemia | 62.8 | 59.7 | 0.06 | 62.4 | 63.4 | -0.02 | 54.8 | 60.6 | -0.12 |
| Hypertensive disorder | 72.8 | 74.1 | -0.03 | 72.8 | 72.9 | 0.00 | 72.1 | 74.0 | -0.04 |
| Lesion of liver | 1.4 | 1.4 | 0.00 | 1.4 | 1.4 | 0.00 | 1.2 | 1.4 | -0.02 |
| Obesity | 47.3 | 39.5 | 0.16 | 47.7 | 47.6 | 0.00 | 43.7 | 44.3 | -0.01 |
| Osteoarthritis | 38.7 | 34.5 | 0.09 | 38.5 | 39.8 | -0.03 | 35.7 | 36.2 | -0.01 |
| Pneumonia | 5.5 | 6.7 | -0.05 | 5.5 | 5.4 | 0.00 | 5.8 | 6.4 | -0.02 |
| Psoriasis | 1.7 | 1.2 | 0.04 | 1.6 | 1.8 | -0.01 | 1.2 | 1.5 | -0.02 |
| Renal impairment | 5.9 | 8.1 | -0.09 | 5.8 | 5.8 | 0.00 | 8.0 | 8.2 | -0.01 |
| Rheumatoid arthritis | 2.3 | 2.3 | 0.00 | 2.3 | 2.4 | -0.01 | 1.8 | 2.3 | -0.04 |
| Schizophrenia | 2.7 | 4.2 | -0.08 | 2.7 | 2.6 | 0.01 | 3.0 | 3.6 | -0.03 |
| Ulcerative colitis | 0.2 | 0.2 | 0.00 | 0.2 | 0.2 | 0.01 | 0.4 | 0.2 | 0.04 |
| Urinary tract infectious disease | 13.3 | 18.6 | -0.15 | 13.5 | 12.5 | 0.03 | 17.8 | 16.7 | 0.03 |
| Viral hepatitis C | 1.6 | 2.1 | -0.04 | 1.6 | 1.8 | -0.02 | 2.2 | 2.0 | 0.01 |
| Visual system disorder | 46.0 | 41.5 | 0.09 | 45.9 | 45.8 | 0.00 | 44.9 | 43.4 | 0.03 |
| **Medical history: Cardiovascular disease** | | | | | | | | | |
| Atrial fibrillation | 2.1 | 2.8 | -0.04 | 2.1 | 2.1 | 0.00 | 1.7 | 2.4 | -0.05 |
| Cerebrovascular disease | 4.1 | 5.1 | -0.05 | 3.9 | 3.4 | 0.03 | 4.5 | 4.4 | 0.00 |
| Coronary arteriosclerosis | 8.5 | 9.3 | -0.03 | 8.3 | 7.9 | 0.01 | 8.4 | 8.2 | 0.01 |
| Heart disease | 23.8 | 26.8 | -0.07 | 23.7 | 22.7 | 0.02 | 24.2 | 24.8 | -0.01 |
| Heart failure | 6.5 | 8.4 | -0.07 | 6.4 | 6.2 | 0.01 | 6.6 | 7.5 | -0.04 |
| Ischemic heart disease | 5.9 | 6.9 | -0.04 | 5.8 | 5.7 | 0.00 | 5.5 | 6.2 | -0.03 |
| Peripheral vascular disease | 6.6 | 6.9 | -0.01 | 6.5 | 6.7 | -0.01 | 6.2 | 6.7 | -0.02 |
| Pulmonary embolism | 1.0 | 1.1 | -0.01 | 1.0 | 1.0 | 0.00 | 0.7 | 1.2 | -0.05 |
| Venous thrombosis | 1.4 | 1.7 | -0.03 | 1.4 | 1.4 | 0.00 | 1.3 | 1.6 | -0.03 |
| **Medical history: Neoplasms** | | | | | | | | | |
| Hematologic neoplasm | 0.8 | 1.0 | -0.03 | 0.8 | 1.0 | -0.02 | 0.9 | 1.0 | -0.01 |

| (Continued) | Before ajustment | | | After matching | | | After stratification | | |
|---|---|---|---|---|---|---|---|---|---|
| Characteristic | T (%) | C (%) | StdDiff | T (%) | C (%) | StdDiff | T (%) | C (%) | StdDiff |
| Malignant lymphoma | <0.2 | 0.3 | -0.04 | <0.2 | 0.2 | -0.01 | 0.3 | 0.3 | 0.01 |
| Malignant neoplasm of anorectum | <0.2 | 0.2 | -0.01 | <0.2 | 0.1 | 0.01 | <0.1 | 0.1 | -0.01 |
| Malignant neoplastic disease | 6.0 | 6.5 | -0.02 | 6.0 | 6.3 | -0.01 | 7.2 | 6.5 | 0.03 |
| Malignant tumor of breast | 2.6 | 2.4 | 0.01 | 2.6 | 2.5 | 0.01 | 3.9 | 2.4 | 0.09 |
| Malignant tumor of colon | <0.2 | 0.4 | -0.06 | <0.2 | 0.3 | -0.03 | 0.2 | 0.4 | -0.03 |
| Malignant tumor of lung | <0.2 | 0.3 | -0.05 | <0.2 | 0.3 | -0.03 | <0.1 | 0.3 | -0.06 |
| **Medication use** | | | | | | | | | |
| Agents acting on the renin-angiotensin system | 58.0 | 60.7 | -0.06 | 57.8 | 57.2 | 0.01 | 58.3 | 58.9 | -0.01 |
| Antibacterials for systemic use | 73.7 | 72.6 | 0.03 | 73.4 | 74.0 | -0.01 | 73.2 | 72.4 | 0.02 |
| Antidepressants | 59.3 | 53.9 | 0.11 | 58.9 | 61.1 | -0.04 | 54.7 | 55.6 | -0.02 |
| Antiepileptics | 44.4 | 39.1 | 0.11 | 44.2 | 44.9 | -0.01 | 39.1 | 41.9 | -0.06 |
| Antiinflammatory and antirheumatic products | 65.1 | 59.9 | 0.11 | 65.0 | 64.6 | 0.01 | 61.5 | 63.0 | -0.03 |
| Antineoplastic agents | 5.0 | 4.3 | 0.04 | 5.0 | 4.5 | 0.02 | 4.5 | 4.2 | 0.01 |
| Antipsoriatics | 0.7 | 0.4 | 0.03 | 0.6 | 0.5 | 0.01 | 0.4 | 0.5 | 0.00 |
| Antithrombotic agents | 21.3 | 22.4 | -0.03 | 21.3 | 20.8 | 0.01 | 21.5 | 22.9 | -0.03 |
| Beta blocking agents | 30.8 | 31.8 | -0.02 | 30.8 | 30.1 | 0.01 | 32.5 | 30.9 | 0.04 |
| Calcium channel blockers | 20.9 | 24.8 | -0.09 | 21.1 | 20.6 | 0.01 | 27.1 | 23.7 | 0.08 |
| Diuretics | 46.2 | 48.4 | -0.04 | 46.1 | 46.0 | 0.00 | 48.2 | 46.5 | 0.04 |
| Drugs for acid related disorders | 51.0 | 48.6 | 0.05 | 50.8 | 51.8 | -0.02 | 52.9 | 49.6 | 0.07 |
| Drugs for obstructive airway diseases | 64.1 | 58.1 | 0.12 | 63.6 | 64.0 | -0.01 | 58.0 | 60.0 | -0.04 |
| Immunosuppressants | 2.7 | 2.4 | 0.02 | 2.7 | 2.9 | -0.01 | 1.9 | 2.6 | -0.05 |
| Lipid modifying agents | 61.5 | 60.1 | 0.03 | 61.4 | 61.6 | 0.00 | 60.5 | 60.5 | 0.00 |
| Opioids | 47.9 | 49.4 | -0.03 | 47.9 | 46.8 | 0.02 | 49.0 | 47.8 | 0.02 |
| Psycholeptics | 51.3 | 49.0 | 0.05 | 51.2 | 51.9 | -0.01 | 48.3 | 49.0 | -0.01 |
| Psychostimulants | 6.2 | 5.2 | 0.04 | 6.2 | 6.5 | -0.01 | 5.2 | 5.6 | -0.02 |

##### 2.3.6 Open Claims

**Supplementary Table 176:** Baseline patient characteristics for SGLT2I (female) (T) and SU (female) (C) new-users in the Open Claims data source. We report proportion of initiators satisfying selected base-line characteristics and the standardized difference of population proportions (StdDiff) before and after propensity score adjustment. Less extreme StdDiffs through matching and stratification suggest improved balance between patient cohorts through adjustment.

| | Before adjustment | | | After matching | | | After stratification | | |
|---|---|---|---|---|---|---|---|---|---|
| Characteristic | T (%) | C (%) | StdDiff | T (%) | C (%) | StdDiff | T (%) | C (%) | StdDiff |
| **Age group** | | | | | | | | | |
| 15 - 19 | 0.1 | 0.1 | 0.01 | 0.1 | 0.1 | 0.00 | 0.1 | 0.1 | 0.00 |
| 25 - 29 | 1.0 | 1.1 | -0.01 | 1.0 | 0.9 | 0.00 | 1.0 | 1.0 | 0.00 |
| 30 - 34 | 1.9 | 2.0 | -0.01 | 2.0 | 2.1 | -0.01 | 2.0 | 2.1 | -0.01 |
| 35 - 39 | 3.6 | 3.3 | 0.01 | 3.5 | 3.4 | 0.01 | 3.5 | 3.4 | 0.01 |
| 40 - 44 | 6.3 | 5.1 | 0.05 | 6.4 | 6.3 | 0.00 | 5.7 | 5.5 | 0.01 |
| 45 - 49 | 9.8 | 7.8 | 0.07 | 9.8 | 10.1 | -0.01 | 8.5 | 8.4 | 0.00 |
| 50 - 54 | 13.9 | 11.3 | 0.08 | 13.8 | 14.0 | -0.01 | 13.8 | 14.0 | -0.01 |
| 55 - 59 | 16.6 | 13.5 | 0.09 | 16.5 | 16.2 | 0.01 | 14.7 | 14.2 | 0.01 |
| 60 - 64 | 16.3 | 14.0 | 0.06 | 16.1 | 16.4 | -0.01 | 14.5 | 14.5 | 0.00 |
| 65 - 69 | 12.8 | 14.8 | -0.06 | 12.9 | 13.0 | 0.00 | 14.2 | 14.3 | 0.00 |
| 70 - 74 | 8.8 | 11.5 | -0.09 | 8.9 | 8.7 | 0.01 | 10.6 | 10.8 | 0.00 |
| 75 - 79 | 5.2 | 10.8 | -0.21 | 5.3 | 4.8 | 0.02 | 8.6 | 9.3 | -0.02 |
| 80 - 84 | 2.9 | 4.1 | -0.06 | 3.0 | 2.9 | 0.00 | 3.7 | 3.8 | 0.00 |
| 85 - 89 | 0.2 | 0.1 | 0.04 | 0.2 | 0.2 | 0.00 | 0.1 | 0.1 | 0.00 |
| **Medical history: General** | | | | | | | | | |
| Acute respiratory disease | 16.3 | 15.8 | 0.01 | 16.0 | 16.2 | -0.01 | 16.6 | 16.1 | 0.01 |
| Attention deficit hyperactivity disorder | 0.6 | 0.4 | 0.04 | 0.6 | 0.6 | 0.00 | 0.5 | 0.4 | 0.00 |
| Chronic liver disease | 1.1 | 1.6 | -0.04 | 1.2 | 1.1 | 0.00 | 1.6 | 1.5 | 0.01 |
| Chronic obstructive lung disease | 4.5 | 5.9 | -0.06 | 4.4 | 4.5 | 0.00 | 5.9 | 5.6 | 0.01 |
| Crohn's disease | 0.2 | 0.2 | 0.00 | 0.2 | 0.2 | 0.00 | 0.2 | 0.2 | 0.00 |
| Dementia | 0.5 | 1.0 | -0.07 | 0.5 | 0.5 | -0.01 | 0.8 | 0.9 | -0.01 |
| Depressive disorder | 9.8 | 9.4 | 0.01 | 9.6 | 9.7 | 0.00 | 10.3 | 9.4 | 0.03 |
| Gastroesophageal reflux disease | 9.9 | 9.2 | 0.03 | 9.7 | 9.8 | 0.00 | 9.9 | 9.5 | 0.01 |
| Gastrointestinal hemorrhage | 1.2 | 1.4 | -0.02 | 1.2 | 1.2 | 0.00 | 1.4 | 1.4 | 0.00 |
| Human immunodeficiency virus infection | 0.2 | 0.2 | 0.00 | 0.2 | 0.2 | 0.00 | 0.2 | 0.2 | 0.00 |
| Hyperlipidemia | 42.7 | 39.1 | 0.07 | 41.8 | 41.7 | 0.00 | 40.8 | 40.0 | 0.02 |
| Hypertensive disorder | 48.2 | 48.3 | 0.00 | 47.5 | 47.4 | 0.00 | 48.9 | 48.4 | 0.01 |
| Lesion of liver | 0.6 | 0.7 | -0.02 | 0.7 | 0.7 | 0.00 | 0.7 | 0.7 | 0.00 |
| Obesity | 14.4 | 11.7 | 0.08 | 14.0 | 14.0 | 0.00 | 13.0 | 12.3 | 0.02 |
| Osteoarthritis | 15.5 | 16.2 | -0.02 | 15.3 | 15.2 | 0.00 | 16.9 | 16.0 | 0.03 |
| Pneumonia | 2.1 | 2.8 | -0.04 | 2.0 | 2.1 | -0.01 | 2.6 | 2.6 | 0.00 |
| Psoriasis | 1.0 | 0.8 | 0.02 | 1.0 | 1.0 | 0.00 | 0.9 | 0.9 | 0.00 |
| Renal impairment | 4.8 | 6.9 | -0.09 | 4.7 | 4.3 | 0.02 | 4.7 | 4.3 | 0.02 |
| Rheumatoid arthritis | 1.3 | 1.3 | 0.00 | 1.3 | 1.4 | -0.01 | 1.5 | 1.4 | 0.00 |
| Schizophrenia | 0.3 | 0.4 | -0.02 | 0.3 | 0.3 | 0.00 | 0.4 | 0.4 | 0.00 |
| Ulcerative colitis | 0.2 | 0.2 | 0.00 | 0.2 | 0.2 | 0.00 | 0.2 | 0.2 | 0.00 |
| Urinary tract infectious disease | 6.4 | 8.2 | -0.07 | 6.3 | 6.5 | 0.00 | 8.2 | 7.8 | 0.02 |
| Viral hepatitis C | 0.2 | 0.4 | -0.03 | 0.2 | 0.3 | -0.01 | 0.3 | 0.3 | 0.00 |
| Visual system disorder | 18.4 | 19.4 | -0.03 | 18.3 | 17.8 | 0.01 | 19.4 | 19.1 | 0.01 |
| **Medical history: Cardiovascular disease** | | | | | | | | | |
| Atrial fibrillation | 2.7 | 3.2 | -0.03 | 2.5 | 2.5 | 0.00 | 3.1 | 3.0 | 0.00 |
| Cerebrovascular disease | 2.4 | 3.2 | -0.05 | 2.3 | 2.4 | 0.00 | 3.0 | 3.0 | 0.00 |
| Coronary arteriosclerosis | 5.7 | 5.8 | 0.00 | 5.3 | 4.9 | 0.02 | 5.7 | 5.6 | 0.01 |
| Heart disease | 14.8 | 15.4 | -0.02 | 13.9 | 13.4 | 0.01 | 15.4 | 15.0 | 0.01 |
| Heart failure | 4.0 | 4.0 | 0.00 | 3.4 | 3.2 | 0.01 | 3.8 | 3.9 | 0.00 |
| Ischemic heart disease | 3.2 | 3.3 | 0.00 | 3.0 | 2.8 | 0.01 | 3.2 | 3.1 | 0.00 |
| Peripheral vascular disease | 3.9 | 4.2 | -0.02 | 3.7 | 3.7 | 0.00 | 4.3 | 4.0 | 0.01 |
| Pulmonary embolism | 0.5 | 0.5 | -0.01 | 0.4 | 0.4 | 0.00 | 0.5 | 0.5 | 0.00 |
| Venous thrombosis | 0.6 | 0.8 | -0.02 | 0.6 | 0.7 | -0.01 | 0.8 | 0.8 | 0.00 |
| **Medical history: Neoplasms** | | | | | | | | | |
| Hematologic neoplasm | 0.5 | 0.7 | -0.03 | 0.5 | 0.5 | 0.00 | 0.6 | 0.7 | 0.00 |
| Malignant neoplasm of anorectum | 0.1 | 0.1 | -0.01 | 0.1 | 0.1 | 0.00 | 0.1 | 0.1 | 0.00 |
| Malignant neoplastic disease | 5.0 | 5.8 | -0.03 | 4.9 | 4.9 | 0.00 | 5.7 | 5.6 | 0.00 |
| Malignant tumor of breast | 2.0 | 2.2 | -0.01 | 2.0 | 2.0 | 0.00 | 2.2 | 2.2 | 0.00 |
| Malignant tumor of colon | 0.2 | 0.3 | -0.02 | 0.2 | 0.2 | 0.00 | 0.3 | 0.3 | 0.00 |
| Malignant tumor of lung | 0.2 | 0.3 | -0.02 | 0.2 | 0.2 | 0.00 | 0.2 | 0.2 | 0.00 |
| Malignant tumor of urinary bladder | 0.1 | 0.1 | -0.02 | 0.1 | 0.1 | 0.01 | 0.1 | 0.1 | 0.01 |

| (Continued) | Before ajustment | | | After matching | | | After stratification | | |
|---|---|---|---|---|---|---|---|---|---|
| Characteristic | T (%) | C (%) | StdDiff | T (%) | C (%) | StdDiff | T (%) | C (%) | StdDiff |
| **Medication use** | | | | | | | | | |
| Agents acting on the renin-angiotensin system | 64.6 | 65.8 | -0.03 | 64.1 | 64.1 | 0.00 | 64.7 | 65.5 | -0.02 |
| Antibacterials for systemic use | 61.0 | 60.6 | 0.01 | 60.7 | 61.2 | -0.01 | 61.8 | 61.1 | 0.01 |
| Antidepressants | 38.3 | 35.9 | 0.05 | 37.7 | 37.8 | 0.00 | 37.6 | 36.6 | 0.02 |
| Antiepileptics | 21.9 | 22.8 | -0.02 | 21.6 | 22.0 | -0.01 | 23.4 | 22.8 | 0.01 |
| Antiinflammatory and antirheumatic products | 40.4 | 39.3 | 0.02 | 40.2 | 40.7 | -0.01 | 40.4 | 40.0 | 0.01 |
| Antineoplastic agents | 5.2 | 4.6 | 0.03 | 5.0 | 5.2 | -0.01 | 5.0 | 4.9 | 0.01 |
| Antipsoriatics | 1.5 | 1.1 | 0.04 | 1.4 | 1.4 | 0.00 | 1.2 | 1.2 | 0.00 |
| Antithrombotic agents | 15.3 | 17.3 | -0.05 | 14.8 | 14.5 | 0.01 | 16.5 | 16.7 | -0.01 |
| Beta blocking agents | 31.4 | 33.3 | -0.04 | 30.6 | 30.2 | 0.01 | 32.2 | 32.5 | -0.01 |
| Calcium channel blockers | 23.2 | 25.6 | -0.06 | 23.1 | 22.8 | 0.01 | 24.5 | 24.9 | -0.01 |
| Diuretics | 43.5 | 44.1 | -0.01 | 42.8 | 42.2 | 0.01 | 43.6 | 43.6 | 0.00 |
| Drugs for acid related disorders | 36.3 | 36.6 | -0.01 | 35.7 | 36.0 | -0.01 | 37.4 | 36.6 | 0.01 |
| Drugs for obstructive airway diseases | 43.2 | 39.9 | 0.07 | 42.4 | 43.1 | -0.01 | 41.3 | 40.9 | 0.01 |
| Immunosuppressants | 3.5 | 3.0 | 0.03 | 3.3 | 3.3 | 0.00 | 3.3 | 3.1 | 0.01 |
| Lipid modifying agents | 68.4 | 66.1 | 0.05 | 67.7 | 67.8 | 0.00 | 66.4 | 66.7 | 0.00 |
| Opioids | 25.7 | 28.2 | -0.06 | 25.5 | 26.1 | -0.01 | 28.5 | 27.7 | 0.02 |
| Psycholeptics | 26.3 | 26.8 | -0.01 | 26.0 | 26.1 | 0.00 | 27.9 | 26.8 | 0.03 |
| Psychostimulants | 3.5 | 2.8 | 0.04 | 3.4 | 3.5 | -0.01 | 3.0 | 3.0 | 0.00 |

##### 2.3.7 OEHR

**Supplementary Table 177:** Baseline patient characteristics for SGLT2I (female) (T) and SU (female) (C) new-users in the OEHR data source. We report proportion of initiators satisfying selected base-line characteristics and the standardized difference of population proportions (StdDiff) before and after propensity score adjustment. Less extreme StdDiffs through matching and stratification suggest improved balance between patient cohorts through adjustment.

| | Before adjustment | | | After matching | | | After stratification | | |
|---|---|---|---|---|---|---|---|---|---|
| Characteristic | T (%) | C (%) | StdDiff | T (%) | C (%) | StdDiff | T (%) | C (%) | StdDiff |
| **Age group** | | | | | | | | | |
| 15 - 19 | 0.1 | 0.1 | 0.01 | 0.1 | 0.1 | 0.01 | 0.2 | 0.1 | 0.03 |
| 20 - 24 | 0.6 | 0.6 | 0.01 | 0.5 | 0.6 | -0.01 | 0.5 | 0.6 | -0.01 |
| 25 - 29 | 1.2 | 1.3 | -0.02 | 1.2 | 1.3 | -0.01 | 1.5 | 1.4 | 0.01 |
| 30 - 34 | 2.3 | 2.4 | 0.00 | 2.4 | 2.3 | 0.00 | 2.7 | 2.4 | 0.01 |
| 35 - 39 | 4.7 | 4.2 | 0.03 | 4.8 | 4.8 | 0.00 | 4.1 | 4.4 | -0.01 |
| 40 - 44 | 7.8 | 6.2 | 0.06 | 7.7 | 7.7 | 0.00 | 6.9 | 6.5 | 0.01 |
| 45 - 49 | 11.8 | 9.6 | 0.07 | 11.6 | 11.6 | 0.00 | 9.3 | 9.9 | -0.02 |
| 50 - 54 | 16.4 | 12.9 | 0.10 | 16.1 | 16.4 | -0.01 | 13.8 | 13.5 | 0.01 |
| 55 - 59 | 18.1 | 15.1 | 0.08 | 17.9 | 17.8 | 0.00 | 16.0 | 15.7 | 0.01 |
| 60 - 64 | 16.5 | 14.3 | 0.06 | 16.3 | 16.6 | -0.01 | 13.0 | 14.9 | -0.05 |
| 65 - 69 | 10.1 | 12.2 | -0.07 | 10.3 | 10.2 | 0.00 | 12.4 | 11.8 | 0.02 |
| 70 - 74 | 5.9 | 9.3 | -0.13 | 6.1 | 5.9 | 0.01 | 9.1 | 8.6 | 0.02 |
| 75 - 79 | 2.9 | 6.7 | -0.18 | 3.1 | 2.9 | 0.01 | 3.1 | 2.9 | 0.01 |
| 80 - 84 | 1.2 | 4.1 | -0.19 | 1.2 | 1.1 | 0.01 | 4.4 | 3.9 | 0.03 |
| 85 - 89 | 0.4 | 0.9 | -0.07 | 0.4 | 0.5 | -0.01 | 0.8 | 1.0 | -0.02 |
| **Race** | | | | | | | | | |
| Asian | 2.6 | 2.8 | -0.01 | 2.6 | 2.7 | 0.00 | 2.2 | 2.8 | -0.03 |
| Black or African American | 13.8 | 13.9 | 0.00 | 13.9 | 13.5 | 0.01 | 13.2 | 14.1 | -0.03 |
| White | 75.5 | 74.7 | 0.02 | 75.1 | 75.4 | -0.01 | 76.2 | 74.4 | 0.04 |
| **Ethnicity** | | | | | | | | | |
| Hispanic or Latino | 6.8 | 8.3 | -0.06 | 7.0 | 6.9 | 0.00 | 7.5 | 7.9 | -0.02 |
| Not Hispanic or Latino | 85.1 | 84.5 | 0.02 | 85.0 | 85.2 | 0.00 | 85.6 | 84.6 | 0.03 |
| **Medical history: General** | | | | | | | | | |
| Acute respiratory disease | 23.8 | 20.3 | 0.08 | 23.3 | 23.4 | 0.00 | 21.8 | 21.4 | 0.01 |
| Attention deficit hyperactivity disorder | 1.1 | 0.7 | 0.04 | 1.1 | 1.1 | 0.00 | 0.8 | 0.8 | -0.01 |
| Chronic liver disease | 2.2 | 3.2 | -0.06 | 2.2 | 2.2 | 0.00 | 2.4 | 3.0 | -0.04 |
| Chronic obstructive lung disease | 4.6 | 5.7 | -0.05 | 4.7 | 4.7 | 0.00 | 6.7 | 5.7 | 0.04 |
| Crohn’s disease | 0.2 | 0.3 | 0.00 | 0.3 | 0.3 | 0.00 | 0.2 | 0.3 | 0.00 |
| Dementia | 0.4 | 1.0 | -0.07 | 0.4 | 0.4 | 0.00 | 0.8 | 0.9 | -0.01 |
| Depressive disorder | 20.5 | 18.6 | 0.05 | 20.1 | 20.2 | 0.00 | 19.4 | 19.5 | 0.00 |
| Gastroesophageal reflux disease | 20.1 | 17.9 | 0.06 | 19.1 | 19.0 | 0.00 | 19.1 | 19.0 | 0.00 |
| Gastrointestinal hemorrhage | 1.1 | 1.4 | -0.03 | 1.1 | 1.1 | 0.00 | 1.2 | 1.4 | -0.01 |
| Human immunodeficiency virus infection | 0.1 | 0.1 | 0.00 | 0.1 | 0.1 | 0.01 | 0.1 | 0.1 | 0.00 |
| Hyperlipidemia | 63.0 | 61.0 | 0.04 | 61.9 | 62.1 | 0.00 | 62.3 | 60.8 | 0.03 |
| Hypertensive disorder | 63.6 | 63.7 | 0.00 | 63.1 | 63.2 | 0.00 | 64.6 | 63.7 | 0.02 |
| Lesion of liver | 0.8 | 0.8 | 0.00 | 0.8 | 0.7 | 0.01 | 1.4 | 0.8 | 0.06 |
| Obesity | 35.6 | 26.3 | 0.20 | 34.7 | 35.5 | -0.02 | 34.7 | 35.5 | -0.02 |
| Osteoarthritis | 17.3 | 18.1 | -0.02 | 17.1 | 17.2 | 0.00 | 19.4 | 18.4 | 0.03 |
| Pneumonia | 1.8 | 2.4 | -0.04 | 1.8 | 1.9 | -0.01 | 2.8 | 2.3 | 0.03 |
| Psoriasis | 1.7 | 1.3 | 0.03 | 1.6 | 1.6 | 0.00 | 1.5 | 1.5 | 0.00 |
| Renal impairment | 3.8 | 6.8 | -0.13 | 4.0 | 3.7 | 0.01 | 7.1 | 6.5 | 0.03 |
| Rheumatoid arthritis | 1.8 | 1.5 | 0.02 | 1.7 | 1.7 | 0.00 | 1.6 | 1.6 | 0.00 |
| Schizophrenia | 0.2 | 0.4 | -0.04 | 0.1 | 0.2 | -0.01 | 0.1 | 0.3 | -0.04 |
| Ulcerative colitis | 0.2 | 0.3 | -0.01 | 0.2 | 0.2 | -0.01 | 0.2 | 0.3 | 0.00 |
| Urinary tract infectious disease | 6.3 | 8.6 | -0.09 | 6.4 | 6.3 | 0.00 | 9.5 | 8.1 | 0.05 |
| Viral hepatitis C | 0.3 | 0.5 | -0.02 | 0.3 | 0.3 | 0.00 | 0.4 | 0.4 | -0.01 |
| Visual system disorder | 12.1 | 15.5 | -0.10 | 12.3 | 12.2 | 0.00 | 15.1 | 14.9 | 0.01 |
| **Medical history: Cardiovascular disease** | | | | | | | | | |
| Atrial fibrillation | 2.1 | 3.2 | -0.07 | 2.2 | 2.0 | 0.01 | 3.3 | 3.0 | 0.02 |
| Cerebrovascular disease | 2.0 | 2.8 | -0.05 | 2.0 | 2.1 | -0.01 | 2.3 | 2.7 | -0.03 |
| Coronary arteriosclerosis | 6.5 | 6.5 | 0.00 | 6.2 | 6.1 | 0.00 | 6.9 | 6.4 | 0.02 |
| Heart disease | 15.3 | 16.5 | -0.03 | 14.9 | 14.7 | 0.00 | 17.1 | 16.3 | 0.02 |
| Heart failure | 2.6 | 3.3 | -0.04 | 2.5 | 2.4 | 0.01 | 2.7 | 3.1 | -0.02 |
| Peripheral vascular disease | 3.9 | 3.8 | 0.01 | 3.7 | 3.6 | 0.01 | 4.4 | 3.9 | 0.03 |
| Pulmonary embolism | 0.4 | 0.5 | -0.01 | 0.4 | 0.4 | 0.00 | 1.5 | 0.5 | 0.10 |
| Venous thrombosis | 0.8 | 1.0 | -0.02 | 0.8 | 0.8 | 0.00 | 0.8 | 1.0 | -0.01 |
| **Medical history: Neoplasms** | | | | | | | | | |

| (Continued) | Before ajustment | | | After matching | | | After stratification | | |
|---|---|---|---|---|---|---|---|---|---|
| Characteristic | T (%) | C (%) | StdDiff | T (%) | C (%) | StdDiff | T (%) | C (%) | StdDiff |
| Hematologic neoplasm | 0.5 | 0.7 | -0.02 | 0.6 | 0.6 | -0.01 | 0.6 | 0.7 | -0.01 |
| Malignant lymphoma | 0.3 | 0.3 | 0.00 | 0.3 | 0.3 | 0.01 | 0.3 | 0.3 | 0.01 |
| Malignant neoplasm of anorectum | 0.1 | 0.1 | -0.02 | 0.1 | 0.1 | 0.00 | 0.5 | 0.1 | 0.07 |
| Malignant neoplastic disease | 5.5 | 6.6 | -0.05 | 5.5 | 5.5 | 0.00 | 6.3 | 6.4 | 0.00 |
| Malignant tumor of breast | 2.0 | 2.5 | -0.04 | 2.0 | 2.1 | 0.00 | 2.1 | 2.4 | -0.02 |
| Malignant tumor of colon | 0.2 | 0.3 | -0.02 | 0.2 | 0.2 | 0.00 | 0.2 | 0.3 | -0.01 |
| Malignant tumor of lung | 0.1 | 0.2 | -0.02 | 0.2 | 0.1 | 0.00 | 0.6 | 0.2 | 0.06 |
| Malignant tumor of urinary bladder | 0.1 | 0.1 | -0.01 | 0.1 | 0.1 | 0.00 | 0.1 | 0.1 | 0.00 |
| **Medication use** | | | | | | | | | |
| Agents acting on the renin-angiotensin system | 61.3 | 61.7 | -0.01 | 61.1 | 60.8 | 0.00 | 61.3 | 61.4 | 0.00 |
| Antibacterials for systemic use | 49.2 | 47.4 | 0.04 | 48.6 | 49.0 | -0.01 | 49.6 | 47.7 | 0.04 |
| Antidepressants | 41.4 | 37.2 | 0.09 | 40.7 | 41.0 | -0.01 | 39.8 | 38.1 | 0.03 |
| Antiepileptics | 20.7 | 19.7 | 0.02 | 20.5 | 20.6 | 0.00 | 21.6 | 20.6 | 0.02 |
| Antiinflammatory and antirheumatic products | 56.3 | 57.6 | -0.03 | 55.9 | 55.7 | 0.00 | 55.2 | 57.4 | -0.04 |
| Antineoplastic agents | 4.8 | 4.4 | 0.02 | 4.6 | 4.8 | -0.01 | 4.2 | 4.5 | -0.01 |
| Antipsoriatics | 0.5 | 0.5 | 0.00 | 0.5 | 0.5 | 0.00 | 0.8 | 0.5 | 0.05 |
| Antithrombotic agents | 32.0 | 37.8 | -0.12 | 32.1 | 31.5 | 0.01 | 35.6 | 36.2 | -0.01 |
| Beta blocking agents | 29.7 | 32.1 | -0.05 | 29.4 | 29.2 | 0.00 | 32.5 | 31.4 | 0.02 |
| Calcium channel blockers | 18.9 | 20.4 | -0.04 | 18.9 | 18.8 | 0.00 | 21.0 | 20.2 | 0.02 |
| Diuretics | 41.0 | 42.8 | -0.04 | 40.6 | 40.4 | 0.00 | 42.0 | 41.9 | 0.00 |
| Drugs for acid related disorders | 41.6 | 42.7 | -0.02 | 41.0 | 41.1 | 0.00 | 43.7 | 42.6 | 0.02 |
| Drugs for obstructive airway diseases | 48.4 | 45.2 | 0.06 | 47.7 | 47.8 | 0.00 | 46.9 | 46.3 | 0.01 |
| Immunosuppressants | 3.3 | 2.8 | 0.03 | 2.9 | 3.0 | -0.01 | 2.9 | 3.0 | -0.01 |
| Lipid modifying agents | 65.9 | 65.2 | 0.01 | 64.9 | 65.2 | -0.01 | 64.9 | 65.2 | -0.01 |
| Opioids | 27.7 | 30.6 | -0.06 | 27.6 | 27.4 | 0.00 | 31.6 | 30.0 | 0.04 |
| Psycholeptics | 31.5 | 31.3 | 0.01 | 31.0 | 31.3 | -0.01 | 32.6 | 31.4 | 0.02 |
| Psychostimulants | 4.5 | 3.3 | 0.06 | 4.4 | 4.4 | 0.00 | 3.6 | 3.7 | 0.00 |

##### 2.3.8 SIDIAP

**Supplementary Table 178:** Baseline patient characteristics for SGLT2I (female) (T) and SU (female) (C) new-users in the SIDIAP data source. We report proportion of initiators satisfying selected base-line characteristics and the standardized difference of population proportions (StdDiff) before and after propensity score adjustment. Less extreme StdDiffs through matching and stratification suggest improved balance between patient cohorts through adjustment.

| | Before adjustment | | | After matching | | | After stratification | | |
|---|---|---|---|---|---|---|---|---|---|
| Characteristic | T (%) | C (%) | StdDiff | T (%) | C (%) | StdDiff | T (%) | C (%) | StdDiff |
| **Age group** | | | | | | | | | |
| 25 - 29 | <0.2 | 0.1 | -0.01 | <0.1 | <0.2 | -0.03 | <0.1 | <0.2 | -0.03 |
| 30 - 34 | 0.5 | 0.3 | 0.04 | 0.9 | 0.3 | 0.08 | 1.2 | 0.3 | 0.11 |
| 35 - 39 | 0.9 | 0.8 | 0.01 | 1.5 | 0.7 | 0.08 | 1.0 | 0.6 | 0.05 |
| 40 - 44 | 3.6 | 2.5 | 0.06 | 3.9 | 3.3 | 0.03 | 3.9 | 2.9 | 0.06 |
| 45 - 49 | 6.2 | 5.1 | 0.05 | 7.8 | 6.6 | 0.05 | 7.0 | 6.0 | 0.04 |
| 50 - 54 | 9.7 | 8.4 | 0.05 | 9.8 | 10.2 | -0.01 | 9.7 | 11.1 | -0.04 |
| 55 - 59 | 15.0 | 11.2 | 0.11 | 13.9 | 12.7 | 0.03 | 13.5 | 11.4 | 0.06 |
| 60 - 64 | 15.7 | 14.7 | 0.03 | 16.2 | 15.5 | 0.02 | 16.6 | 13.9 | 0.07 |
| 65 - 69 | 17.1 | 15.8 | 0.04 | 16.6 | 16.7 | 0.00 | 15.9 | 18.1 | -0.06 |
| 70 - 74 | 13.3 | 14.8 | -0.04 | 15.1 | 14.8 | 0.01 | 15.8 | 16.4 | -0.02 |
| 75 - 79 | 10.0 | 12.6 | -0.08 | 7.8 | 8.9 | -0.04 | 8.5 | 7.3 | 0.04 |
| 80 - 84 | 5.0 | 8.3 | -0.13 | 4.2 | 5.3 | -0.05 | 4.4 | 6.7 | -0.10 |
| 85 - 89 | 2.0 | 4.2 | -0.13 | 1.7 | 3.0 | -0.09 | 1.9 | 2.9 | -0.06 |
| 90 - 94 | 0.5 | 1.1 | -0.06 | <0.5 | 1.4 | -0.12 | 0.4 | 2.1 | -0.15 |
| 95 - 99 | <0.2 | 0.2 | -0.02 | <0.5 | 0.3 | -0.02 | <0.1 | <0.2 | -0.02 |
| **Medical history: General** | | | | | | | | | |
| Acute respiratory disease | 17.4 | 11.7 | 0.16 | 18.3 | 17.7 | 0.02 | 17.0 | 14.2 | 0.08 |
| Chronic liver disease | <0.2 | 0.4 | -0.04 | <0.5 | 0.2 | -0.03 | <0.1 | <0.2 | -0.01 |
| Chronic obstructive lung disease | 0.8 | 0.7 | 0.02 | 0.8 | 0.3 | 0.07 | 0.8 | 0.3 | 0.06 |
| Dementia | 0.2 | 0.9 | -0.10 | <0.5 | 0.7 | -0.06 | 0.4 | 0.6 | -0.02 |
| Depressive disorder | 2.2 | 4.6 | -0.13 | 2.0 | 3.2 | -0.08 | 2.6 | 3.9 | -0.07 |
| Gastroesophageal reflux disease | 1.8 | 1.0 | 0.07 | 2.1 | 2.5 | -0.03 | 2.3 | 1.9 | 0.03 |
| Gastrointestinal hemorrhage | 1.2 | 1.0 | 0.02 | 1.5 | 1.1 | 0.03 | 1.6 | 0.9 | 0.07 |
| Hyperlipidemia | 2.8 | 4.2 | -0.07 | 2.2 | 3.3 | -0.07 | 2.3 | 3.3 | -0.06 |
| Hypertensive disorder | 4.2 | 5.7 | -0.07 | 3.4 | 3.1 | 0.02 | 3.7 | 3.9 | -0.01 |
| Lesion of liver | 0.2 | 0.2 | -0.01 | <0.1 | <0.2 | 0.00 | <0.1 | <0.2 | 0.00 |
| Obesity | 8.8 | 9.4 | -0.02 | 10.3 | 9.4 | 0.03 | 9.3 | 8.5 | 0.03 |
| Osteoarthritis | 4.8 | 7.5 | -0.11 | 5.1 | 4.6 | 0.02 | 4.6 | 4.0 | 0.03 |
| Pneumonia | 1.0 | 1.2 | -0.02 | <0.5 | 1.4 | -0.10 | 0.8 | 0.9 | -0.01 |
| Psoriasis | 0.3 | 0.4 | 0.00 | <0.5 | 0.3 | 0.03 | 0.4 | 0.2 | 0.03 |
| Renal impairment | 1.5 | 1.9 | -0.03 | 1.6 | 1.3 | 0.02 | 1.9 | 1.2 | 0.06 |
| Rheumatoid arthritis | 0.2 | 0.2 | 0.01 | <0.5 | <0.1 | 0.06 | 0.2 | <0.2 | 0.04 |
| Urinary tract infectious disease | 10.1 | 5.7 | 0.16 | 10.8 | 10.6 | 0.01 | 10.3 | 9.4 | 0.03 |
| Viral hepatitis C | <0.2 | 0.1 | -0.02 | <0.5 | <0.1 | 0.00 | <0.1 | <0.2 | 0.00 |
| Visual system disorder | 14.1 | 17.3 | -0.09 | 17.5 | 17.5 | 0.00 | 15.9 | 16.8 | -0.02 |
| **Medical history: Cardiovascular disease** | | | | | | | | | |
| Atrial fibrillation | 1.3 | 1.5 | -0.02 | <0.5 | 0.8 | -0.06 | 0.9 | 0.6 | 0.03 |
| Cerebrovascular disease | 0.6 | 1.3 | -0.06 | 1.0 | 1.0 | 0.00 | 0.8 | 1.3 | -0.05 |
| Heart disease | 8.8 | 7.4 | 0.05 | 5.2 | 5.5 | -0.01 | 6.9 | 6.0 | 0.04 |
| Heart failure | 2.4 | 2.0 | 0.03 | 0.9 | 0.5 | 0.06 | 1.6 | 0.5 | 0.10 |
| Ischemic heart disease | 3.3 | 1.7 | 0.10 | 1.6 | 1.5 | 0.00 | 2.8 | 1.8 | 0.07 |
| Peripheral vascular disease | 0.6 | 0.7 | -0.01 | 0.5 | 0.6 | -0.01 | 0.5 | 0.6 | -0.01 |
| Pulmonary embolism | 0.2 | 0.2 | -0.01 | <0.5 | 0.2 | 0.01 | <0.1 | 0.2 | -0.01 |
| Venous thrombosis | 1.4 | 1.2 | 0.01 | 2.1 | 1.0 | 0.09 | 1.4 | 0.9 | 0.05 |
| **Medical history: Neoplasms** | | | | | | | | | |
| Malignant neoplastic disease | 2.1 | 2.4 | -0.03 | 1.7 | 2.3 | -0.04 | 1.8 | 1.8 | 0.00 |
| Malignant tumor of breast | 0.5 | 0.8 | -0.04 | <0.5 | 0.8 | -0.05 | 0.5 | 0.6 | -0.02 |
| Malignant tumor of colon | 0.2 | 0.2 | 0.01 | <0.5 | 0.2 | 0.01 | 0.2 | <0.2 | 0.01 |
| Malignant tumor of lung | <0.2 | <0.1 | 0.00 | <0.1 | <0.2 | -0.01 | <0.1 | <0.2 | -0.01 |
| Malignant tumor of urinary bladder | <0.2 | <0.1 | 0.01 | <0.1 | <0.2 | 0.01 | <0.1 | <0.2 | 0.01 |
| **Medication use** | | | | | | | | | |
| Agents acting on the renin-angiotensin system | 61.9 | 55.9 | 0.12 | 58.0 | 56.7 | 0.03 | 59.9 | 56.8 | 0.06 |
| Antibacterials for systemic use | 44.7 | 35.6 | 0.19 | 46.9 | 44.1 | 0.06 | 41.8 | 47.5 | -0.11 |
| Antidepressants | 35.8 | 28.4 | 0.16 | 32.7 | 32.3 | 0.01 | 34.4 | 36.5 | -0.04 |
| Antiepileptics | 14.7 | 8.2 | 0.21 | 12.5 | 13.1 | -0.01 | 13.1 | 14.2 | -0.03 |
| Antiinflammatory and antirheumatic products | 52.3 | 58.3 | -0.12 | 50.9 | 50.5 | 0.01 | 51.2 | 51.3 | 0.00 |
| Antineoplastic agents | 2.5 | 2.0 | 0.03 | 2.4 | 2.8 | -0.02 | 2.2 | 2.8 | -0.04 |

| (Continued) | Before ajustment | | | After matching | | | After stratification | | |
|---|---|---|---|---|---|---|---|---|---|
| Characteristic | T (%) | C (%) | StdDiff | T (%) | C (%) | StdDiff | T (%) | C (%) | StdDiff |
| Antipsoriatics | 1.9 | 0.9 | 0.09 | 1.0 | 1.2 | -0.02 | 1.5 | 1.1 | 0.03 |
| Antithrombotic agents | 30.6 | 29.3 | 0.03 | 22.6 | 23.6 | -0.02 | 26.6 | 22.9 | 0.09 |
| Beta blocking agents | 31.7 | 22.4 | 0.21 | 23.6 | 23.1 | 0.01 | 27.8 | 27.5 | 0.01 |
| Calcium channel blockers | 22.7 | 19.0 | 0.09 | 19.5 | 19.9 | -0.01 | 21.2 | 19.6 | 0.04 |
| Diuretics | 50.6 | 48.2 | 0.05 | 49.4 | 47.5 | 0.04 | 51.0 | 49.6 | 0.03 |
| Drugs for acid related disorders | 56.7 | 57.6 | -0.02 | 55.2 | 54.8 | 0.01 | 56.3 | 55.6 | 0.01 |
| Drugs for obstructive airway diseases | 31.6 | 23.3 | 0.19 | 28.1 | 28.5 | -0.01 | 29.3 | 30.2 | -0.02 |
| Immunosuppressants | 2.1 | 0.9 | 0.09 | 1.7 | 0.9 | 0.07 | 1.7 | 0.9 | 0.07 |
| Lipid modifying agents | 61.2 | 54.1 | 0.14 | 55.6 | 55.0 | 0.01 | 60.1 | 56.3 | 0.08 |
| Opioids | 26.2 | 19.7 | 0.16 | 25.7 | 25.4 | 0.01 | 25.3 | 23.6 | 0.04 |
| Psycholeptics | 41.5 | 40.1 | 0.03 | 43.3 | 42.5 | 0.02 | 42.1 | 45.2 | -0.06 |
| Psychostimulants | 3.7 | 3.2 | 0.02 | 1.6 | 1.2 | 0.03 | 2.7 | 1.4 | 0.09 |

#### 2.4 GLP1RA (female) vs DPP4I (female)

##### 2.4.1 CCAE

**Supplementary Table 179:** Baseline patient characteristics for GLP1RA (female) (T) and DPP4I (female) (C) new-users in the CCAE data source. We report proportion of initiators satisfying selected base-line characteristics and the standardized difference of population proportions (StdDiff) before and after propensity score adjustment. Less extreme StdDiffs through matching and stratification suggest improved balance between patient cohorts through adjustment.

| Characteristic | Before adjustment | | | After matching | | | After stratification | | |
|---|---|---|---|---|---|---|---|---|---|
| | T (%) | C (%) | StdDiff | T (%) | C (%) | StdDiff | T (%) | C (%) | StdDiff |
| **Age group** | | | | | | | | | |
| 15 - 19 | 0.2 | 0.1 | -0.04 | 0.2 | 0.1 | -0.02 | 0.2 | 0.1 | -0.03 |
| 20 - 24 | 0.9 | 0.4 | -0.07 | 0.7 | 0.7 | -0.01 | 0.7 | 0.5 | -0.02 |
| 25 - 29 | 1.4 | 0.8 | -0.05 | 1.2 | 1.1 | -0.01 | 1.1 | 0.9 | -0.02 |
| 30 - 34 | 3.7 | 2.0 | -0.10 | 2.9 | 2.8 | -0.01 | 2.7 | 2.4 | -0.02 |
| 35 - 39 | 7.1 | 4.6 | -0.11 | 6.1 | 6.1 | 0.00 | 5.3 | 5.3 | 0.00 |
| 40 - 44 | 11.3 | 8.3 | -0.10 | 10.3 | 10.1 | -0.01 | 9.5 | 9.2 | -0.01 |
| 45 - 49 | 16.2 | 13.7 | -0.07 | 15.7 | 15.4 | -0.01 | 14.5 | 14.5 | 0.00 |
| 50 - 54 | 20.8 | 20.1 | -0.02 | 21.1 | 21.0 | 0.00 | 20.6 | 20.4 | -0.01 |
| 55 - 59 | 21.0 | 24.1 | 0.07 | 22.0 | 22.8 | 0.02 | 22.4 | 23.3 | 0.02 |
| 60 - 64 | 16.4 | 23.9 | 0.19 | 18.6 | 18.5 | 0.00 | 21.3 | 21.6 | 0.00 |
| 65 - 69 | 1.0 | 2.1 | 0.08 | 1.3 | 1.4 | 0.01 | 1.7 | 1.8 | 0.01 |
| **Medical history: General** | | | | | | | | | |
| Acute respiratory disease | 35.7 | 33.5 | -0.05 | 35.1 | 35.7 | 0.01 | 34.1 | 34.4 | 0.00 |
| Attention deficit hyperactivity disorder | 2.0 | 0.8 | -0.10 | 1.5 | 1.3 | -0.02 | 1.2 | 1.1 | -0.01 |
| Chronic liver disease | 2.4 | 3.9 | 0.09 | 2.7 | 2.7 | 0.00 | 3.5 | 3.4 | 0.00 |
| Chronic obstructive lung disease | 2.5 | 2.8 | 0.02 | 2.6 | 2.6 | 0.00 | 2.9 | 2.7 | -0.01 |
| Crohn’s disease | 0.3 | 0.3 | 0.00 | 0.3 | 0.4 | 0.01 | 0.3 | 0.3 | 0.01 |
| Dementia | 0.1 | 0.2 | 0.01 | 0.1 | 0.2 | 0.01 | 0.1 | 0.2 | 0.02 |
| Depressive disorder | 17.8 | 12.7 | -0.14 | 15.8 | 16.3 | 0.01 | 14.4 | 14.5 | 0.00 |
| Gastroesophageal reflux disease | 15.1 | 13.1 | -0.06 | 14.3 | 14.5 | 0.00 | 13.6 | 13.9 | 0.01 |
| Gastrointestinal hemorrhage | 2.0 | 1.8 | -0.01 | 1.9 | 1.8 | -0.01 | 2.0 | 1.8 | -0.01 |
| Human immunodeficiency virus infection | 0.1 | 0.1 | -0.01 | 0.1 | 0.2 | 0.01 | 0.1 | 0.1 | 0.01 |
| Hyperlipidemia | 61.7 | 64.4 | 0.06 | 63.2 | 63.7 | 0.01 | 63.9 | 64.0 | 0.00 |
| Hypertensive disorder | 61.8 | 65.0 | 0.07 | 63.5 | 63.6 | 0.00 | 63.5 | 63.6 | 0.00 |
| Lesion of liver | 0.6 | 0.6 | 0.00 | 0.6 | 0.6 | 0.00 | 0.6 | 0.6 | 0.00 |
| Obesity | 35.5 | 19.4 | -0.37 | 29.2 | 30.3 | 0.02 | 24.7 | 24.8 | 0.00 |
| Osteoarthritis | 22.3 | 19.4 | -0.07 | 21.2 | 21.3 | 0.00 | 20.6 | 20.2 | -0.01 |
| Pneumonia | 2.4 | 2.1 | -0.02 | 2.3 | 2.3 | 0.00 | 2.3 | 2.2 | -0.01 |
| Psoriasis | 2.0 | 1.5 | -0.04 | 1.8 | 1.8 | 0.00 | 1.8 | 1.6 | -0.02 |
| Renal impairment | 2.6 | 3.1 | 0.03 | 2.7 | 2.9 | 0.01 | 2.9 | 3.0 | 0.00 |
| Rheumatoid arthritis | 1.6 | 1.6 | -0.01 | 1.6 | 1.6 | 0.01 | 1.5 | 1.6 | 0.00 |
| Schizophrenia | 0.1 | 0.1 | 0.02 | 0.1 | 0.1 | 0.02 | 0.1 | 0.1 | 0.02 |
| Ulcerative colitis | 0.3 | 0.3 | 0.00 | 0.4 | 0.3 | -0.01 | 0.4 | 0.3 | -0.01 |
| Urinary tract infectious disease | 11.0 | 11.8 | 0.02 | 11.2 | 11.3 | 0.00 | 11.7 | 11.6 | 0.00 |
| Viral hepatitis C | 0.2 | 0.3 | 0.02 | 0.2 | 0.2 | 0.00 | 0.2 | 0.2 | 0.00 |
| Visual system disorder | 25.8 | 27.0 | 0.03 | 26.2 | 26.4 | 0.00 | 26.6 | 26.7 | 0.00 |
| **Medical history: Cardiovascular disease** | | | | | | | | | |
| Atrial fibrillation | 1.1 | 1.3 | 0.01 | 1.2 | 1.2 | 0.00 | 1.4 | 1.2 | -0.01 |
| Cerebrovascular disease | 1.9 | 2.5 | 0.04 | 2.0 | 2.0 | 0.00 | 2.0 | 2.3 | 0.02 |
| Coronary arteriosclerosis | 3.7 | 4.3 | 0.03 | 3.7 | 3.5 | -0.01 | 3.9 | 4.0 | 0.01 |
| Heart disease | 13.5 | 14.3 | 0.02 | 13.5 | 13.4 | 0.00 | 14.0 | 14.0 | 0.00 |
| Heart failure | 1.5 | 1.7 | 0.02 | 1.5 | 1.7 | 0.01 | 1.6 | 1.8 | 0.01 |
| Ischemic heart disease | 2.7 | 2.8 | 0.01 | 2.6 | 2.6 | 0.00 | 2.9 | 2.7 | -0.01 |
| Peripheral vascular disease | 3.2 | 3.0 | -0.01 | 3.1 | 3.1 | 0.00 | 2.8 | 3.0 | 0.01 |
| Pulmonary embolism | 0.5 | 0.4 | -0.01 | 0.4 | 0.5 | 0.01 | 0.3 | 0.4 | 0.02 |
| **Medical history: Neoplasms** | | | | | | | | | |
| Hematologic neoplasm | 0.6 | 0.8 | 0.03 | 0.6 | 0.7 | 0.01 | 0.6 | 0.8 | 0.02 |
| Malignant lymphoma | 0.3 | 0.3 | 0.01 | 0.3 | 0.4 | 0.01 | 0.3 | 0.3 | 0.01 |
| Malignant neoplasm of anorectum | 0.1 | 0.1 | 0.01 | 0.1 | 0.1 | 0.02 | 0.1 | 0.1 | 0.01 |
| Malignant neoplastic disease | 6.0 | 6.6 | 0.03 | 6.1 | 6.2 | 0.00 | 6.5 | 6.4 | 0.00 |
| Malignant tumor of breast | 1.9 | 2.7 | 0.05 | 2.1 | 2.3 | 0.01 | 2.3 | 2.5 | 0.01 |
| Malignant tumor of colon | 0.2 | 0.2 | 0.01 | 0.2 | 0.2 | 0.00 | 0.2 | 0.2 | 0.00 |
| Malignant tumor of lung | 0.1 | 0.2 | 0.04 | 0.1 | 0.1 | 0.01 | 0.1 | 0.1 | 0.01 |
| Malignant tumor of urinary bladder | 0.1 | 0.1 | 0.01 | 0.1 | 0.1 | 0.01 | 0.1 | 0.1 | 0.01 |

| (Continued) | Before ajustment | | | After matching | | | After stratification | | |
|---|---|---|---|---|---|---|---|---|---|
| Characteristic | T (%) | C (%) | StdDiff | T (%) | C (%) | StdDiff | T (%) | C (%) | StdDiff |
| **Medication use** | | | | | | | | | |
| Agents acting on the renin-angiotensin system | 57.2 | 63.5 | 0.13 | 59.8 | 59.9 | 0.00 | 61.7 | 61.8 | 0.00 |
| Antibacterials for systemic use | 68.6 | 66.8 | -0.04 | 67.8 | 68.6 | 0.02 | 67.2 | 67.5 | 0.01 |
| Antidepressants | 45.1 | 34.6 | -0.22 | 41.4 | 42.8 | 0.03 | 38.2 | 38.0 | 0.00 |
| Antiepileptics | 18.7 | 14.0 | -0.13 | 16.9 | 17.4 | 0.01 | 16.9 | 17.4 | 0.01 |
| Antiinflammatory and antirheumatic products | 40.0 | 35.0 | -0.10 | 38.4 | 38.8 | 0.01 | 36.0 | 36.7 | 0.01 |
| Antineoplastic agents | 5.7 | 4.8 | -0.04 | 5.2 | 5.5 | 0.01 | 5.2 | 5.0 | -0.01 |
| Antipsoriatics | 0.8 | 0.6 | -0.02 | 0.7 | 0.6 | -0.02 | 0.8 | 0.6 | -0.02 |
| Antithrombotic agents | 7.2 | 7.4 | 0.01 | 7.2 | 7.4 | 0.01 | 7.0 | 7.4 | 0.02 |
| Beta blocking agents | 22.6 | 24.7 | 0.05 | 23.0 | 22.7 | -0.01 | 24.4 | 24.1 | 0.00 |
| Calcium channel blockers | 16.6 | 18.6 | 0.05 | 17.2 | 17.5 | 0.01 | 18.0 | 18.1 | 0.00 |
| Diuretics | 44.5 | 43.4 | -0.02 | 44.0 | 44.4 | 0.01 | 43.5 | 43.8 | 0.00 |
| Drugs for acid related disorders | 29.2 | 27.8 | -0.03 | 28.5 | 28.9 | 0.01 | 28.2 | 28.4 | 0.00 |
| Drugs for obstructive airway diseases | 46.5 | 42.7 | -0.08 | 45.5 | 45.8 | 0.01 | 44.5 | 44.2 | 0.00 |
| Immunosuppressants | 3.6 | 3.0 | -0.03 | 3.4 | 3.4 | 0.00 | 3.3 | 3.2 | -0.01 |
| Lipid modifying agents | 57.1 | 63.4 | 0.13 | 59.7 | 60.1 | 0.01 | 61.1 | 61.8 | 0.01 |
| Opioids | 30.0 | 28.7 | -0.03 | 29.5 | 29.8 | 0.00 | 29.2 | 29.1 | 0.00 |
| Psycholeptics | 31.1 | 26.6 | -0.10 | 29.2 | 30.0 | 0.02 | 28.1 | 28.0 | 0.00 |
| Psychostimulants | 6.0 | 3.7 | -0.11 | 5.0 | 5.1 | 0.00 | 4.4 | 4.3 | 0.00 |

##### 2.4.2 ODOD

**Supplementary Table 180:** Baseline patient characteristics for GLP1RA (female) (T) and DPP4I (female) (C) new-users in the ODOD data source. We report proportion of initiators satisfying selected base-line characteristics and the standardized difference of population proportions (StdDiff) before and after propensity score adjustment. Less extreme StdDiffs through matching and stratification suggest improved balance between patient cohorts through adjustment.

| | Before adjustment | | | After matching | | | After stratification | | |
|---|---|---|---|---|---|---|---|---|---|
| Characteristic | T (%) | C (%) | StdDiff | T (%) | C (%) | StdDiff | T (%) | C (%) | StdDiff |
| **Age group** | | | | | | | | | |
| 20 - 24 | 0.6 | 0.2 | -0.06 | 0.4 | 0.2 | -0.03 | 0.3 | 0.2 | -0.02 |
| 25 - 29 | 1.4 | 0.4 | -0.11 | 1.0 | 0.7 | -0.03 | 0.8 | 0.6 | -0.03 |
| 30 - 34 | 2.7 | 1.3 | -0.10 | 2.2 | 1.9 | -0.02 | 1.9 | 1.6 | -0.02 |
| 35 - 39 | 5.1 | 2.5 | -0.13 | 4.2 | 4.0 | -0.01 | 3.4 | 3.3 | -0.01 |
| 40 - 44 | 8.1 | 4.4 | -0.15 | 6.7 | 6.8 | 0.00 | 6.0 | 5.4 | -0.03 |
| 45 - 49 | 11.3 | 7.3 | -0.14 | 10.0 | 9.8 | -0.01 | 8.1 | 8.3 | 0.01 |
| 50 - 54 | 14.4 | 10.1 | -0.13 | 13.1 | 13.5 | 0.01 | 11.4 | 11.3 | 0.00 |
| 55 - 59 | 15.1 | 12.0 | -0.09 | 14.5 | 14.2 | -0.01 | 12.9 | 12.9 | 0.00 |
| 60 - 64 | 13.4 | 12.5 | -0.03 | 13.9 | 14.1 | 0.00 | 13.5 | 12.8 | -0.02 |
| 65 - 69 | 13.1 | 15.5 | 0.07 | 14.6 | 15.9 | 0.04 | 13.9 | 15.1 | 0.04 |
| 70 - 74 | 9.4 | 15.1 | 0.17 | 14.1 | 13.5 | -0.02 | 14.1 | 13.5 | -0.02 |
| 75 - 79 | 3.9 | 9.5 | 0.22 | 5.4 | 4.6 | -0.04 | 8.8 | 7.9 | -0.03 |
| 80 - 84 | 0.9 | 5.8 | 0.27 | 1.4 | 1.3 | -0.01 | 2.9 | 4.4 | 0.08 |
| 85 - 89 | 0.4 | 3.1 | 0.21 | 0.6 | 0.6 | 0.01 | 2.0 | 2.4 | 0.03 |
| **Medical history: General** | | | | | | | | | |
| Acute respiratory disease | 33.5 | 31.5 | -0.04 | 32.7 | 33.6 | 0.02 | 32.5 | 32.4 | 0.00 |
| Attention deficit hyperactivity disorder | 1.8 | 0.6 | -0.11 | 1.1 | 1.2 | 0.01 | 0.9 | 0.9 | 0.00 |
| Chronic liver disease | 2.4 | 3.7 | 0.08 | 2.8 | 2.6 | -0.01 | 3.6 | 3.3 | -0.01 |
| Chronic obstructive lung disease | 7.5 | 9.2 | 0.06 | 8.0 | 8.4 | 0.01 | 8.6 | 9.0 | 0.01 |
| Crohn’s disease | 0.3 | 0.3 | 0.01 | 0.2 | 0.3 | 0.00 | 0.2 | 0.3 | 0.02 |
| Dementia | 0.9 | 2.8 | 0.14 | 1.1 | 1.1 | 0.00 | 1.8 | 2.3 | 0.03 |
| Depressive disorder | 26.0 | 18.9 | -0.17 | 22.9 | 24.3 | 0.03 | 20.2 | 21.2 | 0.03 |
| Gastroesophageal reflux disease | 23.2 | 21.6 | -0.04 | 22.2 | 22.4 | 0.01 | 21.8 | 22.4 | 0.01 |
| Gastrointestinal hemorrhage | 2.1 | 2.7 | 0.04 | 2.1 | 2.4 | 0.01 | 2.4 | 2.6 | 0.01 |
| Human immunodeficiency virus infection | 0.2 | 0.1 | -0.02 | 0.2 | 0.2 | 0.00 | 0.2 | 0.2 | 0.00 |
| Hyperlipidemia | 74.3 | 80.0 | 0.14 | 76.3 | 76.9 | 0.01 | 79.3 | 78.9 | -0.01 |
| Hypertensive disorder | 72.7 | 79.5 | 0.16 | 74.5 | 74.7 | 0.00 | 77.4 | 78.0 | 0.01 |
| Lesion of liver | 1.3 | 1.0 | -0.02 | 1.3 | 1.1 | -0.01 | 1.0 | 1.0 | 0.00 |
| Obesity | 46.8 | 24.3 | -0.48 | 36.8 | 39.8 | 0.06 | 30.2 | 31.1 | 0.02 |
| Osteoarthritis | 31.7 | 31.7 | 0.00 | 31.4 | 31.7 | 0.01 | 31.7 | 31.8 | 0.00 |
| Pneumonia | 3.5 | 4.0 | 0.03 | 3.3 | 3.4 | 0.00 | 5.0 | 3.9 | -0.06 |
| Psoriasis | 2.4 | 1.6 | -0.05 | 2.2 | 1.9 | -0.02 | 1.8 | 1.8 | 0.00 |
| Renal impairment | 9.0 | 14.8 | 0.18 | 10.4 | 10.5 | 0.00 | 13.8 | 13.3 | -0.02 |
| Rheumatoid arthritis | 2.8 | 2.6 | -0.01 | 2.7 | 2.7 | 0.00 | 2.8 | 2.6 | -0.01 |
| Schizophrenia | 0.2 | 0.4 | 0.04 | 0.2 | 0.4 | 0.03 | 0.2 | 0.4 | 0.04 |
| Ulcerative colitis | 0.4 | 0.4 | 0.01 | 0.3 | 0.3 | 0.00 | 0.3 | 0.4 | 0.01 |
| Urinary tract infectious disease | 12.8 | 17.0 | 0.12 | 13.5 | 13.1 | -0.01 | 15.2 | 15.7 | 0.02 |
| Viral hepatitis C | 0.4 | 0.5 | 0.00 | 0.5 | 0.4 | 0.00 | 0.4 | 0.5 | 0.02 |
| Visual system disorder | 36.9 | 43.3 | 0.13 | 39.1 | 38.7 | -0.01 | 42.9 | 41.5 | -0.03 |
| **Medical history: Cardiovascular disease** | | | | | | | | | |
| Atrial fibrillation | 3.3 | 5.4 | 0.10 | 3.7 | 3.4 | -0.02 | 5.4 | 4.8 | -0.03 |
| Cerebrovascular disease | 4.0 | 6.9 | 0.13 | 4.7 | 4.3 | -0.02 | 5.5 | 6.0 | 0.02 |
| Coronary arteriosclerosis | 7.7 | 11.4 | 0.12 | 8.8 | 8.1 | -0.02 | 10.6 | 10.3 | -0.01 |
| Heart disease | 21.5 | 27.8 | 0.15 | 22.8 | 22.6 | 0.00 | 27.3 | 26.2 | -0.03 |
| Heart failure | 4.7 | 7.0 | 0.10 | 5.0 | 5.1 | 0.01 | 6.8 | 6.5 | -0.01 |
| Ischemic heart disease | 4.7 | 6.4 | 0.07 | 5.0 | 4.3 | -0.03 | 6.1 | 5.8 | -0.01 |
| Peripheral vascular disease | 7.8 | 10.4 | 0.09 | 8.5 | 8.2 | -0.01 | 9.8 | 9.7 | 0.00 |
| Pulmonary embolism | 0.8 | 0.7 | -0.01 | 0.8 | 0.7 | -0.01 | 0.6 | 0.7 | 0.01 |
| Venous thrombosis | 1.2 | 1.7 | 0.04 | 1.3 | 1.3 | 0.00 | 1.9 | 1.6 | -0.03 |
| **Medical history: Neoplasms** | | | | | | | | | |
| Hematologic neoplasm | 0.8 | 1.2 | 0.04 | 0.8 | 1.0 | 0.02 | 0.9 | 1.1 | 0.02 |
| Malignant lymphoma | 0.4 | 0.5 | 0.01 | 0.4 | 0.4 | 0.00 | 0.5 | 0.4 | -0.01 |
| Malignant neoplasm of anorectum | 0.1 | 0.2 | 0.01 | 0.1 | 0.1 | 0.00 | 0.1 | 0.1 | 0.01 |
| Malignant neoplastic disease | 8.0 | 9.8 | 0.06 | 8.4 | 8.4 | 0.00 | 8.8 | 9.3 | 0.02 |
| Malignant tumor of breast | 2.9 | 3.7 | 0.04 | 3.3 | 3.1 | -0.01 | 3.5 | 3.5 | 0.00 |
| Malignant tumor of colon | 0.4 | 0.5 | 0.02 | 0.4 | 0.4 | -0.01 | 0.6 | 0.5 | -0.02 |
| Malignant tumor of lung | 0.2 | 0.3 | 0.04 | 0.2 | 0.2 | 0.02 | 0.1 | 0.3 | 0.04 |

| (Continued) | Before ajustment | | | After matching | | | After stratification | | |
|---|---|---|---|---|---|---|---|---|---|
| Characteristic | T (%) | C (%) | StdDiff | T (%) | C (%) | StdDiff | T (%) | C (%) | StdDiff |
| Malignant tumor of urinary bladder | 0.1 | 0.2 | 0.03 | 0.1 | 0.2 | 0.01 | 0.1 | 0.2 | 0.02 |
| **Medication use** | | | | | | | | | |
| Agents acting on the renin-angiotensin system | 60.3 | 68.8 | 0.18 | 66.4 | 66.8 | 0.01 | 66.4 | 66.8 | 0.01 |
| Antibacterials for systemic use | 66.0 | 65.0 | -0.02 | 65.2 | 65.7 | 0.01 | 64.2 | 65.6 | 0.03 |
| Antidepressants | 47.7 | 35.9 | -0.24 | 43.5 | 45.2 | 0.03 | 39.2 | 39.6 | 0.01 |
| Antiepileptics | 25.6 | 19.8 | -0.14 | 23.5 | 24.9 | 0.03 | 21.6 | 21.5 | 0.00 |
| Antiinflammatory and antirheumatic products | 39.7 | 34.5 | -0.11 | 37.7 | 39.1 | 0.03 | 34.9 | 36.3 | 0.03 |
| Antineoplastic agents | 6.2 | 6.4 | 0.01 | 6.0 | 5.5 | -0.02 | 6.4 | 6.3 | 0.00 |
| Antipsoriatics | 0.7 | 0.7 | 0.00 | 0.7 | 0.7 | -0.01 | 0.6 | 0.8 | 0.02 |
| Antithrombotic agents | 10.5 | 13.6 | 0.10 | 10.9 | 11.3 | 0.01 | 12.3 | 12.9 | 0.02 |
| Beta blocking agents | 28.7 | 34.3 | 0.12 | 33.0 | 32.8 | 0.00 | 33.0 | 32.8 | 0.00 |
| Calcium channel blockers | 20.5 | 27.0 | 0.15 | 21.8 | 22.1 | 0.01 | 24.8 | 25.3 | 0.01 |
| Diuretics | 46.0 | 48.0 | 0.04 | 46.1 | 46.8 | 0.01 | 48.5 | 47.8 | -0.01 |
| Drugs for acid related disorders | 31.9 | 32.3 | 0.01 | 31.7 | 32.0 | 0.01 | 32.8 | 32.6 | 0.00 |
| Drugs for obstructive airway diseases | 49.2 | 44.2 | -0.10 | 47.3 | 48.3 | 0.02 | 46.1 | 46.2 | 0.00 |
| Immunosuppressants | 4.0 | 3.7 | -0.02 | 3.8 | 4.0 | 0.01 | 3.9 | 3.8 | 0.00 |
| Lipid modifying agents | 64.0 | 72.5 | 0.18 | 67.4 | 67.8 | 0.01 | 69.3 | 70.5 | 0.03 |
| Opioids | 31.6 | 30.6 | -0.02 | 31.1 | 32.3 | 0.03 | 30.5 | 31.1 | 0.01 |
| Psycholeptics | 32.8 | 28.7 | -0.09 | 30.9 | 31.8 | 0.02 | 27.8 | 29.9 | 0.05 |
| Psychostimulants | 4.6 | 2.4 | -0.12 | 3.5 | 3.6 | 0.01 | 3.0 | 2.9 | 0.00 |

##### 2.4.3 MDCR

**Supplementary Table 181:** Baseline patient characteristics for GLP1RA (female) (T) and DPP4I (female) (C) new-users in the MDCR data source. We report proportion of initiators satisfying selected base-line characteristics and the standardized difference of population proportions (StdDiff) before and after propensity score adjustment. Less extreme StdDiffs through matching and stratification suggest improved balance between patient cohorts through adjustment.

| | Before adjustment | | | After matching | | | After stratification | | |
|---|---|---|---|---|---|---|---|---|---|
| Characteristic | T (%) | C (%) | StdDiff | T (%) | C (%) | StdDiff | T (%) | C (%) | StdDiff |
| **Age group** | | | | | | | | | |
| 50 - 54 | 0.5 | 0.1 | -0.06 | 0.6 | <0.5 | -0.04 | 0.5 | 0.2 | -0.05 |
| 55 - 59 | 1.1 | 0.5 | -0.07 | 1.1 | 0.9 | -0.02 | 0.6 | 0.6 | 0.00 |
| 60 - 64 | 2.7 | 1.2 | -0.11 | 2.5 | 1.5 | -0.07 | 2.0 | 1.2 | -0.06 |
| 65 - 69 | 43.2 | 29.3 | -0.29 | 43.1 | 37.8 | -0.11 | 36.1 | 30.3 | -0.12 |
| 70 - 74 | 34.8 | 29.6 | -0.11 | 33.6 | 36.9 | 0.07 | 28.9 | 30.4 | 0.03 |
| 75 - 79 | 12.6 | 18.5 | 0.16 | 13.2 | 14.2 | 0.03 | 17.1 | 18.0 | 0.02 |
| 80 - 84 | 4.1 | 12.3 | 0.30 | 4.6 | 4.1 | -0.03 | 9.6 | 11.4 | 0.06 |
| 85 - 89 | 0.8 | 6.0 | 0.29 | 1.0 | 2.1 | 0.09 | 5.1 | 5.5 | 0.02 |
| 90 - 94 | <0.4 | 1.9 | 0.16 | <0.4 | 1.7 | 0.14 | <0.2 | 1.9 | 0.18 |
| **Medical history: General** | | | | | | | | | |
| Acute respiratory disease | 24.6 | 25.7 | 0.02 | 25.5 | 24.4 | -0.02 | 23.7 | 25.6 | 0.04 |
| Attention deficit hyperactivity disorder | 0.5 | 0.2 | -0.06 | <0.4 | 0.6 | 0.04 | <0.2 | 0.2 | 0.02 |
| Chronic liver disease | 2.2 | 2.8 | 0.04 | 2.6 | 2.8 | 0.01 | 4.7 | 2.8 | -0.10 |
| Chronic obstructive lung disease | 9.7 | 10.1 | 0.01 | 9.5 | 10.1 | 0.02 | 10.2 | 10.1 | 0.00 |
| Crohn’s disease | <0.4 | 0.2 | -0.02 | <0.4 | <0.5 | 0.05 | <0.2 | 0.2 | 0.05 |
| Dementia | 1.2 | 3.3 | 0.14 | 1.5 | 2.4 | 0.07 | 2.4 | 3.2 | 0.05 |
| Depressive disorder | 17.7 | 10.1 | -0.22 | 16.6 | 17.8 | 0.03 | 10.1 | 11.0 | 0.03 |
| Gastroesophageal reflux disease | 17.5 | 15.1 | -0.07 | 17.0 | 17.6 | 0.02 | 10.8 | 15.6 | 0.14 |
| Gastrointestinal hemorrhage | 2.6 | 3.5 | 0.05 | 2.5 | 2.0 | -0.03 | 3.7 | 3.4 | -0.02 |
| Hyperlipidemia | 64.1 | 61.5 | -0.05 | 65.6 | 67.4 | 0.04 | 65.8 | 62.3 | -0.07 |
| Hypertensive disorder | 72.9 | 75.9 | 0.07 | 74.2 | 74.9 | 0.02 | 81.6 | 76.0 | -0.14 |
| Lesion of liver | 0.6 | 1.0 | 0.03 | 0.7 | 1.2 | 0.05 | 0.3 | 1.0 | 0.09 |
| Obesity | 29.3 | 10.7 | -0.48 | 25.4 | 30.5 | 0.12 | 12.3 | 12.9 | 0.02 |
| Osteoarthritis | 40.9 | 34.2 | -0.14 | 38.9 | 39.8 | 0.02 | 34.9 | 34.8 | 0.00 |
| Pneumonia | 3.2 | 4.7 | 0.08 | 3.1 | 3.4 | 0.02 | 2.8 | 4.5 | 0.09 |
| Psoriasis | 2.3 | 1.7 | -0.04 | 2.0 | 2.2 | 0.01 | 1.4 | 1.8 | 0.03 |
| Renal impairment | 10.8 | 13.1 | 0.07 | 11.2 | 13.2 | 0.06 | 16.3 | 13.1 | -0.09 |
| Rheumatoid arthritis | 2.2 | 2.3 | 0.01 | 2.0 | 1.3 | -0.05 | 1.2 | 2.3 | 0.08 |
| Schizophrenia | <0.4 | 0.2 | 0.00 | 0.2 | 0.2 | -0.01 | 0.2 | 0.2 | -0.01 |
| Ulcerative colitis | <0.4 | 0.6 | 0.05 | <0.4 | <0.5 | 0.03 | <0.2 | 0.5 | 0.09 |
| Urinary tract infectious disease | 13.7 | 15.4 | 0.05 | 14.0 | 13.5 | -0.02 | 16.9 | 15.2 | -0.05 |
| **Medical history: Cardiovascular disease** | | | | | | | | | |
| Atrial fibrillation | 6.2 | 10.5 | 0.16 | 6.2 | 6.3 | 0.00 | 11.1 | 10.1 | -0.03 |
| Cerebrovascular disease | 8.9 | 12.2 | 0.11 | 9.0 | 8.1 | -0.03 | 14.5 | 11.8 | -0.08 |
| Coronary arteriosclerosis | 14.5 | 17.4 | 0.08 | 14.4 | 14.6 | 0.00 | 19.9 | 17.0 | -0.07 |
| Heart disease | 33.4 | 39.6 | 0.13 | 33.3 | 33.0 | -0.01 | 41.0 | 39.0 | -0.04 |
| Heart failure | 6.3 | 9.1 | 0.11 | 6.4 | 6.3 | 0.00 | 12.4 | 8.8 | -0.11 |
| Ischemic heart disease | 7.1 | 8.5 | 0.05 | 7.2 | 6.9 | -0.01 | 9.4 | 8.2 | -0.04 |
| Peripheral vascular disease | 10.3 | 12.4 | 0.06 | 10.7 | 12.6 | 0.06 | 9.9 | 12.4 | 0.08 |
| Pulmonary embolism | 0.4 | 0.9 | 0.06 | <0.4 | 0.9 | 0.07 | <0.2 | 0.9 | 0.11 |
| Venous thrombosis | 1.7 | 2.2 | 0.04 | 1.8 | 2.0 | 0.02 | 3.4 | 2.2 | -0.07 |
| **Medical history: Neoplasms** | | | | | | | | | |
| Hematologic neoplasm | 1.1 | 1.5 | 0.03 | 1.3 | 1.2 | 0.00 | 1.0 | 1.5 | 0.04 |
| Malignant lymphoma | 0.6 | 0.8 | 0.03 | 0.5 | <0.5 | -0.04 | <0.2 | 0.8 | 0.09 |
| Malignant neoplasm of anorectum | <0.4 | 0.3 | 0.03 | <0.4 | <0.5 | 0.02 | <0.2 | 0.3 | 0.03 |
| Malignant neoplastic disease | 13.2 | 15.7 | 0.07 | 13.4 | 15.2 | 0.05 | 13.8 | 15.6 | 0.05 |
| Malignant tumor of breast | 6.4 | 5.8 | -0.02 | 6.5 | 6.8 | 0.01 | 6.8 | 5.8 | -0.04 |
| Malignant tumor of colon | <0.4 | 0.9 | 0.07 | <0.4 | 0.7 | 0.05 | <0.2 | 0.9 | 0.11 |
| Malignant tumor of lung | <0.4 | 0.6 | 0.07 | <0.4 | <0.5 | 0.04 | 0.8 | 0.6 | -0.03 |
| Malignant tumor of urinary bladder | 0.9 | 0.4 | -0.06 | 1.0 | 0.6 | -0.05 | 1.6 | 0.4 | -0.12 |
| **Medication use** | | | | | | | | | |
| Agents acting on the renin-angiotensin system | 74.7 | 75.3 | 0.01 | 75.6 | 75.5 | 0.00 | 79.7 | 75.0 | -0.11 |
| Antibacterials for systemic use | 67.3 | 66.8 | -0.01 | 68.8 | 67.5 | -0.03 | 65.1 | 66.8 | 0.04 |
| Antidepressants | 44.7 | 32.6 | -0.25 | 43.4 | 43.8 | 0.01 | 30.9 | 33.9 | 0.06 |
| Antiepileptics | 21.9 | 17.3 | -0.12 | 20.7 | 21.8 | 0.03 | 18.2 | 17.9 | -0.01 |
| Antiinflammatory and antirheumatic products | 40.1 | 31.2 | -0.19 | 38.5 | 38.0 | -0.01 | 34.3 | 32.0 | -0.05 |
| Antineoplastic agents | 11.3 | 9.4 | -0.06 | 10.5 | 9.3 | -0.04 | 10.5 | 9.3 | -0.04 |

| (Continued) | Before ajustment | | | After matching | | | After stratification | | |
|---|---|---|---|---|---|---|---|---|---|
| Characteristic | T (%) | C (%) | StdDiff | T (%) | C (%) | StdDiff | T (%) | C (%) | StdDiff |
| Antipsoriatics | 1.1 | 0.8 | -0.04 | 1.2 | 0.9 | -0.03 | 1.0 | 0.8 | -0.03 |
| Antithrombotic agents | 17.8 | 22.0 | 0.10 | 17.5 | 16.9 | -0.02 | 23.7 | 21.6 | -0.05 |
| Beta blocking agents | 41.8 | 48.8 | 0.14 | 42.4 | 43.4 | 0.02 | 47.5 | 48.2 | 0.01 |
| Calcium channel blockers | 30.6 | 37.1 | 0.14 | 30.8 | 31.1 | 0.01 | 38.8 | 36.4 | -0.05 |
| Diuretics | 57.7 | 57.4 | 0.00 | 57.3 | 55.3 | -0.04 | 63.5 | 57.2 | -0.13 |
| Drugs for acid related disorders | 39.6 | 37.3 | -0.05 | 38.0 | 40.2 | 0.04 | 32.7 | 37.7 | 0.10 |
| Drugs for obstructive airway diseases | 53.5 | 45.0 | -0.17 | 52.9 | 52.8 | 0.00 | 48.5 | 45.9 | -0.05 |
| Immunosuppressants | 5.4 | 4.5 | -0.04 | 5.1 | 4.6 | -0.03 | 3.9 | 4.6 | 0.03 |
| Lipid modifying agents | 80.1 | 79.4 | -0.02 | 79.7 | 81.2 | 0.04 | 78.8 | 79.6 | 0.02 |
| Opioids | 30.5 | 30.4 | 0.00 | 30.8 | 30.5 | 0.00 | 30.9 | 30.4 | -0.01 |
| Psycholeptics | 31.4 | 30.7 | -0.01 | 32.7 | 31.6 | -0.02 | 33.0 | 30.8 | -0.05 |
| Psychostimulants | 2.4 | 1.7 | -0.05 | 2.3 | 2.4 | 0.01 | 0.9 | 1.8 | 0.08 |

##### 2.4.4 MDCD

**Supplementary Table 182:** Baseline patient characteristics for GLP1RA (female) (T) and DPP4I (female) (C) new-users in the MDCD data source. We report proportion of initiators satisfying selected base-line characteristics and the standardized difference of population proportions (StdDiff) before and after propensity score adjustment. Less extreme StdDiffs through matching and stratification suggest improved balance between patient cohorts through adjustment.

| | Before adjustment | | | After matching | | | After stratification | | |
|---|---|---|---|---|---|---|---|---|---|
| Characteristic | T (%) | C (%) | StdDiff | T (%) | C (%) | StdDiff | T (%) | C (%) | StdDiff |
| **Age group** | | | | | | | | | |
| 15 - 19 | 1.7 | 0.6 | -0.10 | 1.6 | 0.8 | -0.08 | 1.0 | 0.7 | -0.03 |
| 20 - 24 | 1.8 | 1.1 | -0.06 | 1.6 | 1.8 | 0.01 | 1.0 | 1.2 | 0.02 |
| 25 - 29 | 4.4 | 2.7 | -0.09 | 4.3 | 4.0 | -0.02 | 3.6 | 3.0 | -0.03 |
| 30 - 34 | 8.8 | 5.6 | -0.12 | 8.4 | 8.3 | 0.00 | 7.2 | 6.1 | -0.04 |
| 35 - 39 | 13.3 | 9.2 | -0.13 | 12.5 | 12.2 | -0.01 | 10.2 | 10.0 | -0.01 |
| 40 - 44 | 15.2 | 11.7 | -0.10 | 14.8 | 14.7 | 0.00 | 12.6 | 12.3 | -0.01 |
| 45 - 49 | 15.6 | 13.3 | -0.06 | 15.5 | 15.2 | -0.01 | 14.1 | 13.6 | -0.01 |
| 50 - 54 | 14.4 | 15.9 | 0.04 | 15.0 | 14.0 | -0.03 | 16.9 | 15.6 | -0.04 |
| 55 - 59 | 12.4 | 16.1 | 0.11 | 12.9 | 11.8 | -0.03 | 14.4 | 15.5 | 0.03 |
| 60 - 64 | 8.7 | 13.0 | 0.14 | 9.2 | 9.6 | 0.02 | 12.2 | 12.1 | 0.00 |
| 65 - 69 | 2.4 | 4.6 | 0.12 | 2.6 | 3.2 | 0.04 | 3.7 | 4.3 | 0.03 |
| 70 - 74 | 0.6 | 2.4 | 0.14 | 0.7 | 1.6 | 0.09 | 1.1 | 2.1 | 0.09 |
| 75 - 79 | 0.3 | 1.9 | 0.15 | 0.4 | 1.7 | 0.13 | 0.9 | 1.7 | 0.07 |
| 80 - 84 | 0.3 | 1.1 | 0.09 | 0.3 | 0.6 | 0.04 | 1.1 | 1.0 | -0.01 |
| **Race** | | | | | | | | | |
| Black or African American | 27.2 | 26.5 | -0.02 | 27.4 | 25.9 | -0.03 | 28.4 | 26.3 | -0.05 |
| White | 61.2 | 57.0 | -0.09 | 60.6 | 60.9 | 0.01 | 57.3 | 57.8 | 0.01 |
| **Ethnicity** | | | | | | | | | |
| Hispanic or Latino | 3.6 | 3.0 | -0.04 | 3.6 | 2.8 | -0.05 | 3.9 | 3.0 | -0.05 |
| **Medical history: General** | | | | | | | | | |
| Acute respiratory disease | 44.3 | 40.8 | -0.07 | 44.0 | 43.9 | 0.00 | 40.9 | 41.5 | 0.01 |
| Attention deficit hyperactivity disorder | 4.2 | 1.9 | -0.14 | 3.8 | 3.0 | -0.04 | 2.3 | 2.1 | -0.01 |
| Chronic liver disease | 3.6 | 4.9 | 0.06 | 3.9 | 3.6 | -0.01 | 4.2 | 4.6 | 0.02 |
| Chronic obstructive lung disease | 17.8 | 20.2 | 0.06 | 18.2 | 20.1 | 0.05 | 20.1 | 20.0 | 0.00 |
| Crohn’s disease | 0.4 | 0.4 | 0.00 | 0.3 | 0.3 | 0.00 | 0.3 | 0.3 | 0.00 |
| Dementia | 1.0 | 1.9 | 0.08 | 1.0 | 1.4 | 0.04 | 1.7 | 1.8 | 0.01 |
| Depressive disorder | 43.6 | 38.1 | -0.11 | 42.1 | 45.8 | 0.08 | 40.9 | 39.7 | -0.02 |
| Gastroesophageal reflux disease | 29.8 | 29.7 | 0.00 | 29.5 | 30.7 | 0.03 | 29.9 | 30.0 | 0.00 |
| Gastrointestinal hemorrhage | 3.6 | 3.7 | 0.00 | 3.7 | 4.3 | 0.03 | 4.4 | 3.7 | -0.04 |
| Human immunodeficiency virus infection | 0.5 | 0.6 | 0.02 | 0.5 | 1.0 | 0.06 | 0.3 | 0.7 | 0.05 |
| Hyperlipidemia | 57.8 | 64.7 | 0.14 | 58.9 | 59.7 | 0.02 | 62.8 | 63.4 | 0.01 |
| Hypertensive disorder | 70.0 | 75.1 | 0.12 | 71.2 | 68.8 | -0.05 | 75.1 | 74.0 | -0.03 |
| Lesion of liver | 1.0 | 1.1 | 0.01 | 1.1 | 1.4 | 0.03 | 1.1 | 1.2 | 0.01 |
| Obesity | 58.2 | 39.6 | -0.38 | 55.1 | 59.6 | 0.09 | 40.6 | 44.3 | 0.07 |
| Osteoarthritis | 39.1 | 38.5 | -0.01 | 38.6 | 38.5 | 0.00 | 38.0 | 38.7 | 0.01 |
| Pneumonia | 5.8 | 6.0 | 0.01 | 5.9 | 5.6 | -0.01 | 5.1 | 6.0 | 0.04 |
| Psoriasis | 1.0 | 1.2 | 0.02 | 1.0 | 1.6 | 0.05 | 1.0 | 1.3 | 0.03 |
| Renal impairment | 5.7 | 8.1 | 0.09 | 5.9 | 6.6 | 0.03 | 6.1 | 7.7 | 0.06 |
| Rheumatoid arthritis | 2.3 | 2.8 | 0.03 | 2.2 | 2.4 | 0.01 | 2.1 | 2.7 | 0.04 |
| Schizophrenia | 2.1 | 3.8 | 0.10 | 2.1 | 2.0 | 0.00 | 2.9 | 3.6 | 0.04 |
| Ulcerative colitis | <0.2 | 0.2 | 0.03 | <0.2 | <0.3 | 0.01 | <0.2 | 0.2 | 0.04 |
| Urinary tract infectious disease | 15.8 | 16.5 | 0.02 | 15.8 | 15.8 | 0.00 | 15.3 | 16.6 | 0.04 |
| Viral hepatitis C | 1.7 | 1.7 | 0.00 | 1.7 | 1.4 | -0.02 | 1.9 | 1.7 | -0.02 |
| Visual system disorder | 43.5 | 44.9 | 0.03 | 43.4 | 42.6 | -0.02 | 44.2 | 44.8 | 0.01 |
| **Medical history: Cardiovascular disease** | | | | | | | | | |
| Atrial fibrillation | 2.0 | 3.2 | 0.08 | 2.0 | 2.7 | 0.04 | 1.8 | 3.1 | 0.08 |
| Cerebrovascular disease | 3.6 | 5.1 | 0.08 | 3.6 | 3.3 | -0.01 | 5.0 | 4.7 | -0.01 |
| Coronary arteriosclerosis | 6.9 | 10.5 | 0.13 | 7.1 | 6.6 | -0.02 | 10.5 | 9.8 | -0.03 |
| Heart disease | 23.3 | 27.5 | 0.10 | 23.7 | 23.1 | -0.01 | 28.2 | 26.6 | -0.04 |
| Heart failure | 6.8 | 8.1 | 0.05 | 7.0 | 7.3 | 0.01 | 7.4 | 7.9 | 0.02 |
| Ischemic heart disease | 4.7 | 7.2 | 0.11 | 4.8 | 4.9 | 0.00 | 6.4 | 6.7 | 0.01 |
| Peripheral vascular disease | 5.7 | 7.6 | 0.07 | 5.7 | 6.3 | 0.03 | 6.5 | 7.3 | 0.03 |
| Venous thrombosis | 1.7 | 1.7 | 0.00 | 1.7 | 1.5 | -0.01 | 1.6 | 1.7 | 0.00 |
| **Medical history: Neoplasms** | | | | | | | | | |
| Hematologic neoplasm | 0.6 | 0.8 | 0.02 | 0.7 | 0.9 | 0.02 | 0.7 | 0.8 | 0.01 |
| Malignant lymphoma | 0.2 | 0.3 | 0.01 | 0.3 | <0.3 | -0.01 | 0.3 | 0.3 | 0.00 |
| Malignant neoplastic disease | 4.6 | 6.3 | 0.08 | 4.8 | 5.7 | 0.04 | 5.3 | 6.1 | 0.04 |

| (Continued) | Before ajustment | | | After matching | | | After stratification | | |
|---|---|---|---|---|---|---|---|---|---|
| Characteristic | T (%) | C (%) | StdDiff | T (%) | C (%) | StdDiff | T (%) | C (%) | StdDiff |
| Malignant tumor of breast | 1.4 | 2.6 | 0.08 | 1.6 | 1.7 | 0.01 | 1.7 | 2.4 | 0.05 |
| Malignant tumor of lung | <0.2 | 0.3 | 0.01 | 0.2 | 0.2 | 0.00 | 0.2 | 0.2 | 0.00 |
| Malignant tumor of urinary bladder | <0.2 | 0.1 | -0.01 | <0.2 | <0.3 | -0.02 | <0.2 | 0.1 | -0.02 |
| **Medication use** | | | | | | | | | |
| Agents acting on the renin-angiotensin system | 55.5 | 61.0 | 0.11 | 56.8 | 53.0 | -0.08 | 63.1 | 59.5 | -0.07 |
| Antibacterials for systemic use | 77.5 | 73.0 | -0.10 | 77.3 | 76.2 | -0.03 | 74.6 | 73.9 | -0.02 |
| Antidepressants | 64.1 | 57.7 | -0.13 | 62.9 | 63.7 | 0.01 | 59.9 | 58.9 | -0.02 |
| Antiepileptics | 47.0 | 42.8 | -0.08 | 46.1 | 46.1 | 0.00 | 42.3 | 43.7 | 0.03 |
| Antiinflammatory and antirheumatic products | 65.3 | 62.3 | -0.06 | 64.5 | 65.1 | 0.01 | 63.2 | 63.2 | 0.00 |
| Antineoplastic agents | 5.4 | 5.2 | -0.01 | 5.3 | 5.2 | -0.01 | 5.3 | 5.2 | -0.01 |
| Antipsoriatics | 0.9 | 0.5 | -0.05 | 0.9 | 0.6 | -0.03 | 0.8 | 0.5 | -0.04 |
| Antithrombotic agents | 18.6 | 23.7 | 0.12 | 19.0 | 18.5 | -0.01 | 23.4 | 22.7 | -0.01 |
| Beta blocking agents | 27.6 | 33.2 | 0.12 | 28.2 | 27.0 | -0.03 | 33.0 | 32.0 | -0.02 |
| Calcium channel blockers | 20.6 | 24.5 | 0.10 | 21.2 | 20.6 | -0.02 | 25.4 | 23.6 | -0.04 |
| Diuretics | 45.9 | 47.5 | 0.03 | 46.6 | 44.8 | -0.04 | 48.8 | 47.1 | -0.03 |
| Drugs for acid related disorders | 52.0 | 52.5 | 0.01 | 52.0 | 52.5 | 0.01 | 53.2 | 52.5 | -0.01 |
| Drugs for obstructive airway diseases | 66.4 | 62.3 | -0.09 | 66.2 | 65.9 | -0.01 | 64.1 | 63.2 | -0.02 |
| Immunosuppressants | 3.0 | 3.1 | 0.01 | 3.0 | 2.8 | -0.01 | 3.1 | 3.2 | 0.00 |
| Lipid modifying agents | 56.3 | 65.5 | 0.19 | 58.0 | 57.4 | -0.01 | 65.6 | 63.4 | -0.05 |
| Opioids | 49.7 | 49.0 | -0.01 | 49.6 | 49.3 | -0.01 | 47.8 | 49.0 | 0.02 |
| Psycholeptics | 54.4 | 50.2 | -0.08 | 53.5 | 52.6 | -0.02 | 50.9 | 51.0 | 0.00 |
| Psychostimulants | 9.4 | 6.0 | -0.13 | 8.5 | 7.9 | -0.02 | 6.1 | 6.5 | 0.02 |

##### 2.4.5 Open Claims

**Supplementary Table 183:** Baseline patient characteristics for GLP1RA (female) (T) and DPP4I (female) (C) new-users in the Open Claims data source. We report proportion of initiators satisfying selected base-line characteristics and the standardized difference of population proportions (StdDiff) before and after propensity score adjustment. Less extreme StdDiffs through matching and stratification suggest improved balance between patient cohorts through adjustment.

| | Before adjustment | | | After matching | | | After stratification | | |
|---|---|---|---|---|---|---|---|---|---|
| Characteristic | T (%) | C (%) | StdDiff | T (%) | C (%) | StdDiff | T (%) | C (%) | StdDiff |
| **Age group** | | | | | | | | | |
| 15 - 19 | 0.2 | 0.1 | -0.03 | 0.2 | 0.2 | -0.01 | 0.2 | 0.2 | -0.01 |
| 20 - 24 | 0.9 | 0.4 | -0.06 | 0.8 | 0.6 | -0.01 | 0.6 | 0.6 | -0.01 |
| 25 - 29 | 1.7 | 0.8 | -0.08 | 1.3 | 1.3 | 0.00 | 1.3 | 1.3 | 0.00 |
| 30 - 34 | 3.1 | 1.6 | -0.10 | 2.4 | 2.4 | 0.00 | 2.2 | 2.1 | 0.00 |
| 35 - 39 | 5.4 | 3.0 | -0.12 | 4.3 | 4.5 | 0.01 | 3.9 | 3.8 | 0.00 |
| 40 - 44 | 8.5 | 5.0 | -0.14 | 7.2 | 7.1 | 0.00 | 6.4 | 6.2 | -0.01 |
| 45 - 49 | 12.0 | 7.9 | -0.14 | 10.7 | 10.7 | 0.00 | 9.4 | 9.3 | 0.00 |
| 50 - 54 | 15.5 | 11.5 | -0.12 | 14.3 | 14.1 | -0.01 | 12.9 | 12.7 | -0.01 |
| 55 - 59 | 16.4 | 13.9 | -0.07 | 16.1 | 16.3 | 0.00 | 14.8 | 14.7 | 0.00 |
| 60 - 64 | 14.5 | 14.7 | 0.00 | 15.4 | 15.6 | 0.01 | 14.8 | 14.7 | 0.00 |
| 65 - 69 | 10.8 | 14.3 | 0.10 | 12.5 | 12.5 | 0.00 | 13.2 | 13.2 | 0.00 |
| 70 - 74 | 6.5 | 11.2 | 0.16 | 8.2 | 8.4 | 0.00 | 9.5 | 9.6 | 0.00 |
| 75 - 79 | 3.0 | 10.8 | 0.31 | 4.2 | 4.1 | -0.01 | 7.2 | 8.1 | 0.03 |
| 80 - 84 | 1.3 | 4.6 | 0.20 | 2.1 | 2.2 | 0.01 | 3.6 | 3.5 | 0.00 |
| 85 - 89 | 0.1 | 0.1 | 0.01 | 0.2 | 0.2 | 0.01 | 0.1 | 0.1 | 0.01 |
| **Medical history: General** | | | | | | | | | |
| Acute respiratory disease | 16.8 | 17.3 | 0.01 | 16.5 | 16.9 | 0.01 | 16.5 | 16.9 | 0.01 |
| Attention deficit hyperactivity disorder | 1.2 | 0.4 | -0.09 | 0.8 | 0.7 | -0.01 | 0.7 | 0.7 | -0.01 |
| Chronic liver disease | 0.8 | 1.6 | 0.08 | 1.0 | 1.0 | 0.00 | 1.4 | 1.4 | 0.00 |
| Chronic obstructive lung disease | 4.1 | 5.8 | 0.07 | 4.6 | 4.9 | 0.01 | 5.2 | 5.4 | 0.01 |
| Crohn’s disease | 0.2 | 0.2 | 0.00 | 0.2 | 0.2 | 0.00 | 0.2 | 0.2 | 0.00 |
| Dementia | 0.3 | 1.3 | 0.11 | 0.4 | 0.4 | 0.00 | 0.9 | 0.9 | 0.00 |
| Depressive disorder | 12.8 | 9.7 | -0.10 | 11.2 | 11.3 | 0.00 | 11.2 | 10.9 | -0.01 |
| Gastroesophageal reflux disease | 10.9 | 10.4 | -0.02 | 10.4 | 10.5 | 0.00 | 10.9 | 10.6 | -0.01 |
| Gastrointestinal hemorrhage | 1.1 | 1.5 | 0.04 | 1.2 | 1.2 | 0.00 | 1.4 | 1.4 | 0.00 |
| Human immunodeficiency virus infection | 0.2 | 0.2 | 0.01 | 0.2 | 0.3 | 0.01 | 0.2 | 0.2 | 0.01 |
| Hyperlipidemia | 37.9 | 44.4 | 0.13 | 39.2 | 39.4 | 0.00 | 42.4 | 42.5 | 0.00 |
| Hypertensive disorder | 44.9 | 51.6 | 0.14 | 46.1 | 46.1 | 0.00 | 49.2 | 49.5 | 0.01 |
| Lesion of liver | 0.5 | 0.6 | 0.01 | 0.5 | 0.6 | 0.00 | 0.6 | 0.6 | 0.01 |
| Obesity | 21.6 | 12.1 | -0.26 | 16.7 | 16.7 | 0.00 | 16.0 | 15.2 | -0.02 |
| Osteoarthritis | 17.2 | 18.0 | 0.02 | 16.7 | 16.9 | 0.01 | 17.9 | 17.7 | -0.01 |
| Pneumonia | 2.2 | 2.6 | 0.02 | 2.1 | 2.2 | 0.01 | 2.4 | 2.6 | 0.01 |
| Psoriasis | 1.4 | 0.9 | -0.04 | 1.1 | 1.2 | 0.00 | 1.1 | 1.1 | 0.00 |
| Renal impairment | 4.0 | 7.1 | 0.14 | 4.6 | 4.7 | 0.01 | 5.8 | 6.1 | 0.01 |
| Rheumatoid arthritis | 1.5 | 1.6 | 0.00 | 1.4 | 1.5 | 0.01 | 1.5 | 1.6 | 0.01 |
| Schizophrenia | 0.3 | 0.4 | 0.03 | 0.3 | 0.3 | 0.01 | 0.3 | 0.4 | 0.01 |
| Ulcerative colitis | 0.2 | 0.2 | 0.00 | 0.2 | 0.2 | 0.00 | 0.2 | 0.2 | 0.00 |
| Urinary tract infectious disease | 6.6 | 8.9 | 0.09 | 8.3 | 8.2 | 0.00 | 8.3 | 8.2 | 0.00 |
| Viral hepatitis C | 0.2 | 0.3 | 0.03 | 0.2 | 0.2 | 0.00 | 0.3 | 0.3 | -0.01 |
| Visual system disorder | 16.9 | 21.2 | 0.11 | 17.8 | 18.1 | 0.01 | 19.7 | 19.9 | 0.01 |
| **Medical history: Cardiovascular disease** | | | | | | | | | |
| Atrial fibrillation | 1.8 | 3.4 | 0.10 | 2.1 | 2.1 | 0.00 | 2.8 | 2.9 | 0.01 |
| Cerebrovascular disease | 1.8 | 3.6 | 0.11 | 2.1 | 2.2 | 0.00 | 2.9 | 3.0 | 0.01 |
| Coronary arteriosclerosis | 3.6 | 6.2 | 0.12 | 4.1 | 4.1 | 0.00 | 5.3 | 5.3 | 0.00 |
| Heart disease | 11.3 | 16.7 | 0.16 | 12.2 | 12.3 | 0.00 | 14.7 | 15.1 | 0.01 |
| Heart failure | 2.3 | 4.0 | 0.10 | 2.5 | 2.6 | 0.01 | 3.1 | 3.5 | 0.02 |
| Ischemic heart disease | 2.0 | 3.3 | 0.08 | 2.3 | 2.2 | 0.00 | 2.8 | 2.9 | 0.01 |
| Peripheral vascular disease | 3.2 | 4.5 | 0.07 | 3.4 | 3.4 | 0.00 | 4.0 | 4.1 | 0.00 |
| Venous thrombosis | 0.6 | 0.9 | 0.03 | 0.6 | 0.7 | 0.01 | 0.8 | 0.8 | 0.00 |
| **Medical history: Neoplasms** | | | | | | | | | |
| Hematologic neoplasm | 0.4 | 0.7 | 0.03 | 0.5 | 0.5 | 0.00 | 0.6 | 0.6 | 0.01 |
| Malignant lymphoma | 0.2 | 0.3 | 0.02 | 0.3 | 0.3 | 0.00 | 0.3 | 0.3 | 0.00 |
| Malignant neoplasm of anorectum | 0.1 | 0.1 | 0.03 | 0.1 | 0.1 | 0.00 | 0.1 | 0.1 | 0.01 |
| Malignant neoplastic disease | 4.4 | 6.2 | 0.08 | 4.8 | 4.8 | 0.00 | 5.5 | 5.6 | 0.00 |
| Malignant tumor of breast | 1.7 | 2.5 | 0.06 | 1.8 | 2.0 | 0.01 | 2.1 | 2.3 | 0.01 |
| Malignant tumor of colon | 0.1 | 0.3 | 0.04 | 0.2 | 0.2 | -0.01 | 0.2 | 0.3 | 0.01 |
| Malignant tumor of lung | 0.1 | 0.3 | 0.04 | 0.1 | 0.2 | 0.01 | 0.1 | 0.2 | 0.02 |

| (Continued) | Before ajustment | | | After matching | | | After stratification | | |
|---|---|---|---|---|---|---|---|---|---|
| Characteristic | T (%) | C (%) | StdDiff | T (%) | C (%) | StdDiff | T (%) | C (%) | StdDiff |
| Malignant tumor of urinary bladder | 0.1 | 0.1 | 0.02 | 0.1 | 0.1 | 0.01 | 0.1 | 0.1 | 0.01 |
| **Medication use** | | | | | | | | | |
| Agents acting on the renin-angiotensin system | 57.2 | 67.1 | 0.20 | 61.1 | 61.1 | 0.00 | 63.6 | 64.0 | 0.01 |
| Antibacterials for systemic use | 63.0 | 62.8 | 0.00 | 62.3 | 62.7 | 0.01 | 63.0 | 63.1 | 0.00 |
| Antidepressants | 47.3 | 36.9 | -0.21 | 42.6 | 42.8 | 0.00 | 41.5 | 40.5 | -0.02 |
| Antiepileptics | 26.4 | 23.0 | -0.08 | 24.6 | 24.9 | 0.01 | 24.8 | 24.5 | -0.01 |
| Antiinflammatory and antirheumatic products | 41.9 | 41.1 | -0.02 | 41.5 | 41.5 | 0.00 | 41.7 | 41.4 | -0.01 |
| Antineoplastic agents | 6.3 | 5.5 | -0.03 | 5.7 | 5.6 | 0.00 | 5.9 | 5.7 | -0.01 |
| Antipsoriatics | 2.1 | 1.3 | -0.06 | 1.7 | 1.6 | 0.00 | 1.6 | 1.5 | -0.01 |
| Antithrombotic agents | 11.7 | 18.1 | 0.18 | 15.5 | 16.0 | 0.01 | 15.5 | 16.0 | 0.01 |
| Beta blocking agents | 27.4 | 33.9 | 0.14 | 28.7 | 29.1 | 0.01 | 31.3 | 31.9 | 0.01 |
| Calcium channel blockers | 20.4 | 26.4 | 0.14 | 21.9 | 22.1 | 0.00 | 23.7 | 24.3 | 0.01 |
| Diuretics | 43.0 | 44.6 | 0.03 | 42.8 | 42.7 | 0.00 | 43.8 | 44.0 | 0.00 |
| Drugs for acid related disorders | 37.5 | 39.5 | 0.04 | 37.3 | 37.9 | 0.01 | 38.8 | 39.1 | 0.01 |
| Drugs for obstructive airway diseases | 46.8 | 43.6 | -0.06 | 45.2 | 45.8 | 0.01 | 44.7 | 44.9 | 0.00 |
| Immunosuppressants | 4.1 | 3.7 | -0.02 | 3.8 | 3.9 | 0.00 | 3.8 | 3.9 | 0.00 |
| Lipid modifying agents | 60.7 | 69.5 | 0.18 | 64.9 | 64.8 | 0.00 | 66.8 | 67.0 | 0.00 |
| Opioids | 26.9 | 28.3 | 0.03 | 28.5 | 28.0 | -0.01 | 28.5 | 28.0 | -0.01 |
| Psycholeptics | 30.3 | 27.9 | -0.05 | 28.6 | 28.5 | 0.00 | 29.4 | 28.8 | -0.01 |
| Psychostimulants | 5.6 | 3.1 | -0.12 | 4.1 | 4.3 | 0.01 | 4.0 | 4.0 | 0.00 |

##### 2.4.6 OEHR

**Supplementary Table 184:** Baseline patient characteristics for GLP1RA (female) (T) and DPP4I (female) (C) new-users in the OEHR data source. We report proportion of initiators satisfying selected base-line characteristics and the standardized difference of population proportions (StdDiff) before and after propensity score adjustment. Less extreme StdDiffs through matching and stratification suggest improved balance between patient cohorts through adjustment.

| | Before adjustment | | | After matching | | | After stratification | | |
|---|---|---|---|---|---|---|---|---|---|
| Characteristic | T (%) | C (%) | StdDiff | T (%) | C (%) | StdDiff | T (%) | C (%) | StdDiff |
| **Age group** | | | | | | | | | |
| 15 - 19 | 0.2 | 0.1 | -0.03 | 0.2 | 0.1 | -0.02 | 0.2 | 0.1 | -0.01 |
| 20 - 24 | 1.0 | 0.5 | -0.06 | 0.9 | 0.7 | -0.02 | 0.7 | 0.6 | -0.01 |
| 25 - 29 | 2.1 | 1.1 | -0.08 | 1.9 | 1.8 | -0.01 | 1.5 | 1.3 | -0.01 |
| 30 - 34 | 3.8 | 2.2 | -0.09 | 3.4 | 3.2 | -0.01 | 2.9 | 2.6 | -0.02 |
| 35 - 39 | 7.1 | 4.0 | -0.14 | 6.1 | 6.3 | 0.01 | 4.8 | 4.8 | 0.00 |
| 40 - 44 | 10.0 | 6.8 | -0.12 | 8.9 | 9.1 | 0.00 | 7.5 | 7.5 | 0.00 |
| 45 - 49 | 13.7 | 9.9 | -0.12 | 12.8 | 12.9 | 0.00 | 10.6 | 10.9 | 0.01 |
| 50 - 54 | 16.3 | 13.8 | -0.07 | 16.2 | 16.1 | 0.00 | 14.8 | 14.5 | -0.01 |
| 55 - 59 | 16.6 | 15.7 | -0.02 | 17.0 | 17.3 | 0.01 | 16.1 | 16.0 | 0.00 |
| 60 - 64 | 14.6 | 14.8 | 0.01 | 15.3 | 15.6 | 0.01 | 14.9 | 14.8 | 0.00 |
| 65 - 69 | 8.1 | 12.2 | 0.14 | 9.2 | 9.0 | -0.01 | 11.9 | 11.1 | -0.03 |
| 70 - 74 | 4.3 | 8.6 | 0.18 | 5.2 | 4.9 | -0.01 | 7.5 | 7.4 | 0.00 |
| 75 - 79 | 1.6 | 5.7 | 0.22 | 2.0 | 1.9 | -0.01 | 4.2 | 4.6 | 0.02 |
| 80 - 84 | 0.5 | 3.7 | 0.23 | 0.6 | 0.7 | 0.01 | 1.9 | 2.9 | 0.06 |
| 85 - 89 | 0.1 | 1.0 | 0.12 | 0.2 | 0.3 | 0.03 | 0.6 | 0.8 | 0.03 |
| **Race** | | | | | | | | | |
| Asian | 1.5 | 3.2 | 0.11 | 1.8 | 1.8 | 0.00 | 2.4 | 2.7 | 0.02 |
| Black or African American | 14.5 | 15.2 | 0.02 | 15.0 | 15.1 | 0.00 | 15.5 | 15.1 | -0.01 |
| White | 76.9 | 73.9 | -0.07 | 75.8 | 75.8 | 0.00 | 75.0 | 74.6 | -0.01 |
| **Ethnicity** | | | | | | | | | |
| Hispanic or Latino | 5.9 | 7.0 | 0.04 | 6.1 | 6.5 | 0.02 | 6.0 | 6.7 | 0.03 |
| Not Hispanic or Latino | 85.9 | 85.6 | -0.01 | 85.4 | 85.0 | -0.01 | 86.2 | 85.7 | -0.01 |
| **Medical history: General** | | | | | | | | | |
| Acute respiratory disease | 22.7 | 22.2 | -0.01 | 22.1 | 22.0 | 0.00 | 22.4 | 22.3 | 0.00 |
| Attention deficit hyperactivity disorder | 1.4 | 0.8 | -0.06 | 1.2 | 1.2 | 0.00 | 0.9 | 1.0 | 0.01 |
| Chronic liver disease | 2.3 | 3.1 | 0.05 | 2.6 | 2.4 | -0.01 | 3.3 | 2.9 | -0.03 |
| Chronic obstructive lung disease | 4.1 | 5.4 | 0.06 | 4.4 | 4.3 | 0.00 | 5.3 | 5.1 | -0.01 |
| Crohn's disease | 0.2 | 0.3 | 0.00 | 0.2 | 0.3 | 0.01 | 0.2 | 0.3 | 0.01 |
| Dementia | 0.4 | 1.0 | 0.07 | 0.4 | 0.5 | 0.01 | 0.7 | 0.9 | 0.02 |
| Depressive disorder | 24.5 | 18.7 | -0.14 | 22.1 | 22.5 | 0.01 | 20.3 | 20.3 | 0.00 |
| Gastroesophageal reflux disease | 21.2 | 19.7 | -0.04 | 19.9 | 19.9 | 0.00 | 19.9 | 20.1 | 0.00 |
| Gastrointestinal hemorrhage | 1.3 | 1.4 | 0.01 | 1.3 | 1.4 | 0.01 | 1.5 | 1.4 | 0.00 |
| Human immunodeficiency virus infection | 0.1 | 0.2 | 0.02 | 0.1 | 0.2 | 0.03 | 0.1 | 0.2 | 0.03 |
| Hyperlipidemia | 58.4 | 64.6 | 0.13 | 59.1 | 57.7 | -0.03 | 63.1 | 62.7 | -0.01 |
| Hypertensive disorder | 59.0 | 65.1 | 0.13 | 59.6 | 59.4 | 0.00 | 63.6 | 63.5 | 0.00 |
| Lesion of liver | 0.6 | 0.7 | 0.01 | 0.6 | 0.7 | 0.01 | 0.5 | 0.7 | 0.02 |
| Obesity | 46.7 | 27.9 | -0.40 | 39.5 | 41.1 | 0.03 | 32.8 | 32.9 | 0.00 |
| Osteoarthritis | 18.5 | 18.8 | 0.01 | 17.7 | 17.5 | 0.00 | 18.7 | 18.7 | 0.00 |
| Pneumonia | 2.0 | 2.1 | 0.00 | 1.9 | 2.1 | 0.01 | 2.0 | 2.0 | 0.00 |
| Psoriasis | 2.0 | 1.3 | -0.05 | 1.8 | 1.6 | -0.01 | 1.5 | 1.4 | -0.01 |
| Renal impairment | 4.3 | 6.2 | 0.09 | 4.4 | 4.5 | 0.01 | 5.3 | 5.7 | 0.02 |
| Rheumatoid arthritis | 1.9 | 1.6 | -0.02 | 1.8 | 1.5 | -0.02 | 1.8 | 1.6 | -0.02 |
| Schizophrenia | 0.2 | 0.3 | 0.02 | 0.2 | 0.3 | 0.02 | 0.2 | 0.3 | 0.01 |
| Urinary tract infectious disease | 6.6 | 8.7 | 0.08 | 6.8 | 6.9 | 0.00 | 8.0 | 8.2 | 0.01 |
| Viral hepatitis C | 0.4 | 0.4 | 0.00 | 0.5 | 0.4 | -0.03 | 0.5 | 0.4 | -0.03 |
| Visual system disorder | 12.1 | 13.6 | 0.05 | 11.8 | 11.6 | -0.01 | 12.5 | 13.1 | 0.02 |
| **Medical history: Cardiovascular disease** | | | | | | | | | |
| Atrial fibrillation | 1.8 | 3.3 | 0.10 | 2.3 | 2.9 | 0.04 | 2.3 | 2.9 | 0.04 |
| Cerebrovascular disease | 1.7 | 2.7 | 0.07 | 1.8 | 2.0 | 0.01 | 2.6 | 2.5 | 0.00 |
| Coronary arteriosclerosis | 4.1 | 6.4 | 0.10 | 4.3 | 4.5 | 0.01 | 5.7 | 5.8 | 0.00 |
| Heart disease | 13.5 | 17.0 | 0.10 | 13.6 | 13.5 | 0.00 | 15.9 | 16.0 | 0.00 |
| Heart failure | 2.1 | 2.8 | 0.04 | 2.1 | 2.0 | -0.01 | 2.4 | 2.6 | 0.01 |
| Ischemic heart disease | 2.5 | 3.3 | 0.05 | 2.6 | 2.7 | 0.01 | 3.0 | 3.1 | 0.00 |
| Peripheral vascular disease | 3.5 | 3.6 | 0.01 | 3.2 | 3.2 | 0.00 | 3.1 | 3.5 | 0.03 |
| Pulmonary embolism | 0.6 | 0.5 | -0.01 | 0.5 | 0.4 | -0.02 | 0.5 | 0.5 | 0.00 |
| Venous thrombosis | 0.8 | 1.0 | 0.02 | 0.7 | 0.8 | 0.01 | 0.7 | 1.0 | 0.03 |
| **Medical history: Neoplasms** | | | | | | | | | |

| (Continued) | Before ajustment | | | After matching | | | After stratification | | |
|---|---|---|---|---|---|---|---|---|---|
| Characteristic | T (%) | C (%) | StdDiff | T (%) | C (%) | StdDiff | T (%) | C (%) | StdDiff |
| Hematologic neoplasm | 0.5 | 0.7 | 0.04 | 0.4 | 0.7 | 0.03 | 0.4 | 0.7 | 0.04 |
| Malignant lymphoma | 0.2 | 0.4 | 0.03 | 0.2 | 0.3 | 0.01 | 0.2 | 0.4 | 0.03 |
| Malignant neoplasm of anorectum | 0.1 | 0.1 | 0.01 | 0.1 | 0.1 | 0.01 | 0.1 | 0.1 | -0.01 |
| Malignant neoplastic disease | 5.3 | 6.6 | 0.05 | 5.4 | 5.7 | 0.01 | 5.9 | 6.3 | 0.01 |
| Malignant tumor of breast | 1.9 | 2.7 | 0.05 | 2.0 | 2.1 | 0.01 | 2.3 | 2.5 | 0.01 |
| Malignant tumor of colon | 0.2 | 0.3 | 0.01 | 0.3 | 0.2 | -0.01 | 0.4 | 0.2 | -0.02 |
| Malignant tumor of lung | 0.1 | 0.2 | 0.04 | 0.1 | 0.1 | 0.02 | 0.1 | 0.1 | 0.02 |
| **Medication use** | | | | | | | | | |
| Agents acting on the renin-angiotensin system | 55.9 | 62.0 | 0.12 | 58.2 | 57.4 | -0.02 | 61.9 | 60.6 | -0.03 |
| Antibacterials for systemic use | 50.8 | 49.4 | -0.03 | 49.5 | 49.1 | -0.01 | 49.3 | 49.7 | 0.01 |
| Antidepressants | 47.9 | 38.4 | -0.19 | 44.7 | 45.6 | 0.02 | 41.5 | 40.9 | -0.01 |
| Antiepileptics | 23.6 | 20.3 | -0.08 | 22.2 | 23.0 | 0.02 | 21.4 | 21.2 | 0.00 |
| Antiinflammatory and antirheumatic products | 56.4 | 56.5 | 0.00 | 56.1 | 56.4 | 0.01 | 57.5 | 56.6 | -0.02 |
| Antineoplastic agents | 5.4 | 5.0 | -0.01 | 5.0 | 5.2 | 0.01 | 5.2 | 5.1 | 0.00 |
| Antipsoriatics | 0.7 | 0.5 | -0.03 | 0.6 | 0.4 | -0.02 | 0.6 | 0.5 | -0.01 |
| Antithrombotic agents | 29.2 | 36.1 | 0.15 | 30.8 | 31.1 | 0.01 | 34.9 | 34.4 | -0.01 |
| Beta blocking agents | 26.5 | 31.2 | 0.10 | 26.9 | 27.3 | 0.01 | 26.9 | 27.3 | 0.01 |
| Calcium channel blockers | 17.4 | 21.2 | 0.10 | 17.9 | 18.1 | 0.00 | 20.4 | 20.2 | 0.00 |
| Diuretics | 41.5 | 42.4 | 0.02 | 42.5 | 42.0 | -0.01 | 42.5 | 42.0 | -0.01 |
| Drugs for acid related disorders | 43.0 | 43.7 | 0.01 | 42.2 | 42.5 | 0.01 | 43.7 | 43.5 | 0.00 |
| Drugs for obstructive airway diseases | 51.4 | 47.4 | -0.08 | 49.6 | 50.2 | 0.01 | 48.7 | 48.5 | 0.00 |
| Immunosuppressants | 4.0 | 3.2 | -0.04 | 3.8 | 3.6 | -0.01 | 3.6 | 3.4 | -0.01 |
| Lipid modifying agents | 60.5 | 67.0 | 0.14 | 62.5 | 62.0 | -0.01 | 66.3 | 65.5 | -0.02 |
| Opioids | 28.9 | 30.0 | 0.02 | 29.3 | 29.8 | 0.01 | 29.3 | 29.8 | 0.01 |
| Psycholeptics | 34.8 | 32.5 | -0.05 | 33.4 | 33.9 | 0.01 | 32.5 | 33.1 | 0.01 |
| Psychostimulants | 6.1 | 3.9 | -0.10 | 5.4 | 5.7 | 0.01 | 4.4 | 4.6 | 0.01 |

#### 2.5 GLP1RA (female) vs SU (female)

##### 2.5.1 CCAE

**Supplementary Table 185:** Baseline patient characteristics for GLP1RA (female) (T) and SU (female) (C) new-users in the CCAE data source. We report proportion of initiators satisfying selected base-line characteristics and the standardized difference of population proportions (StdDiff) before and after propensity score adjustment. Less extreme StdDiffs through matching and stratification suggest improved balance between patient cohorts through adjustment.

| | Before adjustment | | | After matching | | | After stratification | | |
|---|---|---|---|---|---|---|---|---|---|
| Characteristic | T (%) | C (%) | StdDiff | T (%) | C (%) | StdDiff | T (%) | C (%) | StdDiff |
| **Age group** | | | | | | | | | |
| 15 - 19 | 0.2 | 0.1 | 0.04 | 0.2 | 0.1 | 0.02 | 0.3 | 0.1 | 0.05 |
| 20 - 24 | 0.9 | 0.4 | 0.06 | 0.7 | 0.6 | 0.01 | 0.6 | 0.5 | 0.02 |
| 25 - 29 | 1.4 | 1.0 | 0.03 | 1.3 | 1.1 | 0.01 | 1.0 | 1.1 | -0.01 |
| 30 - 34 | 3.7 | 2.7 | 0.06 | 3.1 | 3.1 | 0.00 | 2.9 | 2.9 | 0.00 |
| 35 - 39 | 7.1 | 5.1 | 0.08 | 6.4 | 6.3 | 0.00 | 5.6 | 5.6 | 0.00 |
| 40 - 44 | 11.3 | 8.4 | 0.10 | 10.4 | 10.6 | -0.01 | 9.3 | 9.0 | 0.01 |
| 45 - 49 | 16.2 | 13.6 | 0.07 | 15.9 | 15.8 | 0.00 | 14.0 | 14.1 | 0.00 |
| 50 - 54 | 20.8 | 20.0 | 0.02 | 20.8 | 21.1 | -0.01 | 20.1 | 20.1 | 0.00 |
| 55 - 59 | 21.0 | 23.5 | -0.06 | 21.9 | 22.3 | -0.01 | 22.4 | 23.1 | -0.02 |
| 60 - 64 | 16.4 | 23.1 | -0.17 | 18.1 | 17.6 | 0.01 | 21.9 | 21.6 | 0.01 |
| 65 - 69 | 1.0 | 2.1 | -0.09 | 1.2 | 1.3 | -0.01 | 1.9 | 1.9 | 0.00 |
| **Medical history: General** | | | | | | | | | |
| Acute respiratory disease | 35.7 | 30.8 | 0.10 | 34.3 | 34.7 | -0.01 | 34.3 | 34.7 | -0.01 |
| Attention deficit hyperactivity disorder | 2.0 | 0.5 | 0.13 | 1.3 | 1.2 | 0.00 | 0.9 | 0.8 | 0.00 |
| Chronic liver disease | 2.4 | 3.6 | -0.07 | 2.7 | 2.6 | 0.01 | 3.6 | 3.4 | 0.01 |
| Chronic obstructive lung disease | 2.5 | 3.1 | -0.04 | 2.6 | 2.7 | 0.00 | 3.3 | 3.0 | 0.01 |
| Crohn’s disease | 0.3 | 0.3 | 0.00 | 0.3 | 0.3 | 0.00 | 0.2 | 0.3 | -0.01 |
| Dementia | 0.1 | 0.2 | -0.01 | 0.1 | 0.2 | -0.01 | 0.1 | 0.2 | -0.02 |
| Depressive disorder | 17.8 | 11.9 | 0.17 | 15.7 | 16.2 | -0.01 | 14.7 | 13.7 | 0.03 |
| Gastroesophageal reflux disease | 15.1 | 10.7 | 0.13 | 13.3 | 13.7 | -0.01 | 12.3 | 12.1 | 0.01 |
| Gastrointestinal hemorrhage | 2.0 | 1.8 | 0.01 | 2.0 | 1.8 | 0.01 | 2.0 | 1.8 | 0.01 |
| Human immunodeficiency virus infection | 0.1 | 0.1 | 0.01 | 0.1 | 0.1 | 0.01 | 0.1 | 0.1 | 0.00 |
| Hyperlipidemia | 61.7 | 56.3 | 0.11 | 61.1 | 61.5 | -0.01 | 60.3 | 59.1 | 0.02 |
| Hypertensive disorder | 61.8 | 60.7 | 0.02 | 62.4 | 63.0 | -0.01 | 63.4 | 62.3 | 0.03 |
| Lesion of liver | 0.6 | 0.7 | -0.01 | 0.6 | 0.6 | 0.00 | 0.8 | 0.7 | 0.01 |
| Obesity | 35.5 | 17.5 | 0.42 | 28.9 | 30.2 | -0.03 | 22.1 | 22.4 | -0.01 |
| Osteoarthritis | 22.3 | 17.0 | 0.13 | 20.4 | 21.0 | -0.02 | 19.5 | 18.5 | 0.03 |
| Pneumonia | 2.4 | 2.6 | -0.01 | 2.3 | 2.3 | 0.00 | 2.8 | 2.6 | 0.02 |
| Psoriasis | 2.0 | 1.4 | 0.04 | 1.8 | 1.8 | 0.00 | 1.7 | 1.5 | 0.01 |
| Renal impairment | 2.6 | 3.1 | -0.03 | 2.5 | 2.6 | -0.01 | 3.1 | 3.1 | 0.00 |
| Rheumatoid arthritis | 1.6 | 1.4 | 0.02 | 1.5 | 1.6 | 0.00 | 1.5 | 1.5 | 0.00 |
| Schizophrenia | 0.1 | 0.1 | -0.01 | 0.1 | 0.1 | -0.01 | 0.1 | 0.1 | -0.01 |
| Ulcerative colitis | 0.3 | 0.3 | 0.00 | 0.3 | 0.3 | 0.01 | 0.4 | 0.3 | 0.02 |
| Urinary tract infectious disease | 11.0 | 11.3 | -0.01 | 10.8 | 10.9 | 0.00 | 11.8 | 11.2 | 0.02 |
| Viral hepatitis C | 0.2 | 0.3 | -0.03 | 0.2 | 0.2 | 0.00 | 0.2 | 0.3 | -0.01 |
| Visual system disorder | 25.8 | 24.1 | 0.04 | 25.3 | 25.3 | 0.00 | 25.3 | 25.3 | 0.00 |
| **Medical history: Cardiovascular disease** | | | | | | | | | |
| Atrial fibrillation | 1.1 | 1.2 | -0.01 | 1.2 | 1.1 | 0.00 | 1.6 | 1.2 | 0.04 |
| Cerebrovascular disease | 1.9 | 2.4 | -0.03 | 1.9 | 1.9 | 0.00 | 1.9 | 2.3 | -0.02 |
| Coronary arteriosclerosis | 3.7 | 4.5 | -0.04 | 3.8 | 3.8 | 0.00 | 4.3 | 4.3 | 0.00 |
| Heart disease | 13.5 | 13.5 | 0.00 | 13.1 | 13.2 | 0.00 | 14.4 | 13.3 | 0.03 |
| Heart failure | 1.5 | 2.0 | -0.04 | 1.5 | 1.6 | -0.01 | 2.1 | 1.9 | 0.02 |
| Ischemic heart disease | 2.7 | 3.2 | -0.03 | 2.8 | 2.7 | 0.01 | 3.2 | 2.9 | 0.01 |
| Peripheral vascular disease | 3.2 | 2.4 | 0.05 | 3.0 | 2.9 | 0.00 | 2.3 | 2.6 | -0.02 |
| Pulmonary embolism | 0.5 | 0.3 | 0.02 | 0.4 | 0.4 | 0.01 | 0.3 | 0.4 | -0.02 |
| Venous thrombosis | 1.0 | 0.8 | 0.01 | 0.9 | 0.9 | 0.00 | 0.9 | 0.9 | 0.00 |
| **Medical history: Neoplasms** | | | | | | | | | |
| Hematologic neoplasm | 0.6 | 0.8 | -0.03 | 0.5 | 0.6 | -0.01 | 0.7 | 0.8 | -0.01 |
| Malignant lymphoma | 0.3 | 0.3 | -0.01 | 0.3 | 0.2 | 0.00 | 0.3 | 0.3 | 0.00 |
| Malignant neoplasm of anorectum | 0.1 | 0.2 | -0.02 | 0.1 | 0.1 | -0.01 | 0.2 | 0.1 | 0.01 |
| Malignant neoplastic disease | 6.0 | 6.1 | -0.01 | 5.7 | 5.7 | 0.00 | 5.8 | 6.1 | -0.01 |
| Malignant tumor of breast | 1.9 | 2.5 | -0.04 | 2.0 | 2.0 | 0.00 | 2.3 | 2.4 | -0.01 |
| Malignant tumor of colon | 0.2 | 0.3 | -0.02 | 0.2 | 0.2 | 0.00 | 0.3 | 0.3 | 0.00 |
| Malignant tumor of lung | 0.1 | 0.2 | -0.04 | 0.1 | 0.1 | -0.01 | 0.1 | 0.1 | -0.01 |

| (Continued) | Before ajustment | | | After matching | | | After stratification | | |
|---|---|---|---|---|---|---|---|---|---|
| Characteristic | T (%) | C (%) | StdDiff | T (%) | C (%) | StdDiff | T (%) | C (%) | StdDiff |
| Malignant tumor of urinary bladder | 0.1 | 0.1 | -0.01 | 0.1 | 0.1 | 0.00 | 0.1 | 0.1 | -0.01 |
| **Medication use** | | | | | | | | | |
| Agents acting on the renin-angiotensin system | 57.2 | 61.9 | -0.10 | 61.6 | 61.7 | 0.00 | 61.6 | 61.7 | 0.00 |
| Antibacterials for systemic use | 68.6 | 64.7 | 0.08 | 67.3 | 67.8 | -0.01 | 65.8 | 65.8 | 0.00 |
| Antidepressants | 45.1 | 33.8 | 0.23 | 41.7 | 42.6 | -0.02 | 38.2 | 36.5 | 0.04 |
| Antiepileptics | 18.7 | 13.4 | 0.15 | 16.7 | 17.2 | -0.01 | 16.0 | 14.8 | 0.03 |
| Antiinflammatory and antirheumatic products | 40.0 | 34.1 | 0.12 | 38.2 | 38.8 | -0.01 | 36.5 | 35.8 | 0.01 |
| Antineoplastic agents | 5.7 | 4.7 | 0.04 | 5.3 | 5.2 | 0.00 | 5.0 | 4.6 | 0.02 |
| Antipsoriatics | 0.8 | 0.6 | 0.02 | 0.7 | 0.7 | 0.00 | 0.7 | 0.6 | 0.01 |
| Antithrombotic agents | 7.2 | 7.4 | -0.01 | 7.0 | 7.5 | -0.02 | 7.0 | 7.5 | -0.02 |
| Beta blocking agents | 22.6 | 25.2 | -0.06 | 22.9 | 23.1 | 0.00 | 26.1 | 24.8 | 0.03 |
| Calcium channel blockers | 16.6 | 18.1 | -0.04 | 17.1 | 17.4 | -0.01 | 18.8 | 18.0 | 0.02 |
| Diuretics | 44.5 | 43.3 | 0.03 | 44.2 | 44.5 | -0.01 | 42.8 | 43.5 | -0.01 |
| Drugs for acid related disorders | 29.2 | 26.1 | 0.07 | 28.0 | 28.6 | -0.01 | 27.3 | 26.9 | 0.01 |
| Drugs for obstructive airway diseases | 46.5 | 39.3 | 0.15 | 44.5 | 45.1 | -0.01 | 42.5 | 41.5 | 0.02 |
| Immunosuppressants | 3.6 | 2.6 | 0.06 | 3.3 | 3.2 | 0.00 | 3.0 | 2.9 | 0.01 |
| Lipid modifying agents | 57.1 | 60.0 | -0.06 | 59.0 | 59.1 | 0.00 | 61.2 | 60.6 | 0.01 |
| Opioids | 30.0 | 29.3 | 0.01 | 29.3 | 29.5 | 0.00 | 30.8 | 29.7 | 0.02 |
| Psycholeptics | 31.1 | 25.7 | 0.12 | 28.8 | 29.3 | -0.01 | 28.1 | 27.1 | 0.02 |
| Psychostimulants | 6.0 | 3.1 | 0.14 | 4.7 | 4.8 | 0.00 | 3.7 | 3.8 | -0.01 |

##### 2.5.2 ODOD

**Supplementary Table 186:** Baseline patient characteristics for GLP1RA (female) (T) and SU (female) (C) new-users in the ODOD data source. We report proportion of initiators satisfying selected base-line characteristics and the standardized difference of population proportions (StdDiff) before and after propensity score adjustment. Less extreme StdDiffs through matching and stratification suggest improved balance between patient cohorts through adjustment.

| | Before adjustment | | | After matching | | | After stratification | | |
|---|---|---|---|---|---|---|---|---|---|
| Characteristic | T (%) | C (%) | StdDiff | T (%) | C (%) | StdDiff | T (%) | C (%) | StdDiff |
| **Age group** | | | | | | | | | |
| 20 - 24 | 0.6 | 0.2 | 0.06 | 0.5 | 0.3 | 0.03 | 0.4 | 0.2 | 0.04 |
| 25 - 29 | 1.4 | 0.5 | 0.10 | 1.2 | 1.0 | 0.01 | 0.7 | 0.6 | 0.01 |
| 30 - 34 | 2.7 | 1.4 | 0.09 | 2.3 | 2.1 | 0.01 | 1.6 | 1.5 | 0.00 |
| 35 - 39 | 5.1 | 2.5 | 0.14 | 4.2 | 4.3 | -0.01 | 2.8 | 2.8 | 0.00 |
| 40 - 44 | 8.1 | 3.9 | 0.17 | 6.7 | 7.1 | -0.01 | 4.7 | 4.6 | 0.01 |
| 45 - 49 | 11.3 | 6.1 | 0.18 | 10.4 | 10.4 | 0.00 | 10.4 | 10.4 | 0.00 |
| 50 - 54 | 14.4 | 8.8 | 0.17 | 13.4 | 13.6 | 0.00 | 9.2 | 9.7 | -0.02 |
| 55 - 59 | 15.1 | 10.8 | 0.13 | 14.8 | 15.0 | 0.00 | 11.0 | 11.5 | -0.01 |
| 60 - 64 | 13.4 | 11.2 | 0.07 | 13.9 | 13.8 | 0.00 | 11.7 | 11.7 | 0.00 |
| 65 - 69 | 13.1 | 17.5 | -0.12 | 14.8 | 15.2 | -0.01 | 16.1 | 17.0 | -0.02 |
| 70 - 74 | 9.4 | 16.9 | -0.22 | 11.1 | 10.7 | 0.01 | 17.9 | 15.6 | 0.06 |
| 75 - 79 | 3.9 | 10.9 | -0.27 | 4.9 | 4.6 | 0.01 | 11.2 | 9.8 | 0.05 |
| 80 - 84 | 0.9 | 6.2 | -0.28 | 1.2 | 1.1 | 0.01 | 3.4 | 5.3 | -0.09 |
| 85 - 89 | 0.4 | 2.9 | -0.20 | 0.5 | 0.6 | -0.01 | 2.2 | 2.5 | -0.02 |
| **Medical history: General** | | | | | | | | | |
| Acute respiratory disease | 33.5 | 29.5 | 0.09 | 32.2 | 32.6 | -0.01 | 32.7 | 30.1 | 0.06 |
| Attention deficit hyperactivity disorder | 1.8 | 0.5 | 0.12 | 1.1 | 1.2 | -0.01 | 0.7 | 0.7 | 0.00 |
| Chronic liver disease | 2.4 | 3.5 | -0.06 | 2.6 | 2.4 | 0.02 | 2.9 | 3.2 | -0.02 |
| Chronic obstructive lung disease | 7.5 | 10.2 | -0.10 | 8.0 | 8.2 | -0.01 | 9.5 | 9.9 | -0.01 |
| Crohn's disease | 0.3 | 0.3 | 0.00 | 0.2 | 0.2 | 0.00 | 0.2 | 0.3 | -0.02 |
| Dementia | 0.9 | 2.8 | -0.14 | 2.1 | 2.5 | -0.03 | 2.1 | 2.5 | -0.03 |
| Depressive disorder | 26.0 | 19.1 | 0.16 | 23.9 | 24.7 | -0.02 | 19.7 | 20.3 | -0.02 |
| Gastroesophageal reflux disease | 23.2 | 20.6 | 0.06 | 22.0 | 22.7 | -0.02 | 20.7 | 21.1 | -0.01 |
| Gastrointestinal hemorrhage | 2.1 | 2.8 | -0.05 | 2.1 | 2.2 | -0.01 | 2.9 | 2.7 | 0.01 |
| Human immunodeficiency virus infection | 0.2 | 0.1 | 0.02 | 0.2 | 0.2 | 0.00 | 0.1 | 0.2 | -0.01 |
| Hyperlipidemia | 74.3 | 77.0 | -0.06 | 75.1 | 75.2 | 0.00 | 79.5 | 76.7 | 0.07 |
| Hypertensive disorder | 72.7 | 79.5 | -0.16 | 74.6 | 74.1 | 0.01 | 79.2 | 78.5 | 0.01 |
| Lesion of liver | 1.3 | 1.3 | 0.00 | 1.3 | 1.2 | 0.01 | 1.1 | 1.3 | -0.01 |
| Obesity | 46.8 | 24.3 | 0.48 | 40.4 | 42.4 | -0.04 | 27.3 | 27.9 | -0.01 |
| Osteoarthritis | 31.7 | 30.4 | 0.03 | 30.9 | 31.2 | 0.00 | 30.7 | 30.6 | 0.00 |
| Pneumonia | 3.5 | 4.6 | -0.06 | 3.4 | 3.5 | 0.00 | 6.5 | 4.4 | 0.09 |
| Psoriasis | 2.4 | 1.5 | 0.06 | 2.1 | 2.1 | 0.00 | 1.6 | 1.6 | 0.00 |
| Renal impairment | 9.0 | 15.9 | -0.21 | 10.0 | 9.8 | 0.01 | 17.8 | 14.8 | 0.08 |
| Rheumatoid arthritis | 2.8 | 2.6 | 0.01 | 2.6 | 2.8 | -0.01 | 3.1 | 2.6 | 0.03 |
| Schizophrenia | 0.2 | 0.5 | -0.04 | 0.2 | 0.4 | -0.03 | 0.2 | 0.5 | -0.06 |
| Ulcerative colitis | 0.4 | 0.4 | 0.00 | 0.4 | 0.3 | 0.01 | 0.4 | 0.4 | 0.01 |
| Urinary tract infectious disease | 12.8 | 16.6 | -0.11 | 12.8 | 12.9 | 0.00 | 16.8 | 16.0 | 0.02 |
| Viral hepatitis C | 0.4 | 0.5 | 0.00 | 0.4 | 0.4 | 0.01 | 0.4 | 0.5 | 0.00 |
| Visual system disorder | 36.9 | 42.9 | -0.12 | 38.1 | 37.7 | 0.01 | 44.0 | 41.9 | 0.04 |
| **Medical history: Cardiovascular disease** | | | | | | | | | |
| Atrial fibrillation | 3.3 | 5.8 | -0.12 | 3.5 | 3.2 | 0.02 | 6.3 | 5.3 | 0.04 |
| Cerebrovascular disease | 4.0 | 6.8 | -0.12 | 4.3 | 4.1 | 0.01 | 5.5 | 6.4 | -0.04 |
| Coronary arteriosclerosis | 7.7 | 11.7 | -0.14 | 8.4 | 8.1 | 0.01 | 11.2 | 11.1 | 0.00 |
| Heart disease | 21.5 | 28.0 | -0.15 | 21.9 | 21.4 | 0.01 | 28.8 | 26.9 | 0.04 |
| Heart failure | 4.7 | 7.5 | -0.12 | 4.9 | 4.7 | 0.01 | 8.3 | 7.1 | 0.05 |
| Ischemic heart disease | 4.7 | 7.0 | -0.10 | 4.8 | 4.9 | 0.00 | 6.8 | 6.6 | 0.00 |
| Peripheral vascular disease | 7.8 | 10.4 | -0.09 | 8.2 | 8.1 | 0.00 | 12.2 | 10.0 | 0.07 |
| Pulmonary embolism | 0.8 | 0.9 | 0.00 | 0.8 | 0.7 | 0.00 | 0.7 | 0.9 | -0.02 |
| Venous thrombosis | 1.2 | 1.8 | -0.05 | 2.5 | 1.7 | 0.05 | 2.5 | 1.7 | 0.05 |
| **Medical history: Neoplasms** | | | | | | | | | |
| Hematologic neoplasm | 0.8 | 1.4 | -0.06 | 0.8 | 0.8 | 0.00 | 0.8 | 1.3 | -0.04 |
| Malignant lymphoma | 0.4 | 0.5 | -0.02 | 0.4 | 0.4 | -0.01 | 0.5 | 0.5 | -0.01 |
| Malignant neoplasm of anorectum | 0.1 | 0.2 | -0.02 | 0.1 | 0.1 | 0.01 | 0.1 | 0.2 | -0.03 |
| Malignant neoplastic disease | 8.0 | 10.1 | -0.07 | 8.2 | 8.1 | 0.00 | 9.2 | 9.7 | -0.02 |
| Malignant tumor of breast | 2.9 | 3.8 | -0.05 | 3.2 | 3.0 | 0.01 | 3.5 | 3.7 | -0.01 |
| Malignant tumor of colon | 0.4 | 0.5 | -0.03 | 0.4 | 0.3 | 0.02 | 0.6 | 0.5 | 0.01 |
| Malignant tumor of lung | 0.2 | 0.5 | -0.06 | 0.1 | 0.2 | -0.01 | 0.1 | 0.5 | -0.07 |

| (Continued) | Before ajustment | | | After matching | | | After stratification | | |
|---|---|---|---|---|---|---|---|---|---|
| Characteristic | T (%) | C (%) | StdDiff | T (%) | C (%) | StdDiff | T (%) | C (%) | StdDiff |
| Malignant tumor of urinary bladder | 0.1 | 0.2 | -0.03 | 0.1 | 0.1 | 0.00 | 0.1 | 0.2 | -0.04 |
| **Medication use** | | | | | | | | | |
| Agents acting on the renin-angiotensin system | 60.3 | 69.1 | -0.18 | 64.0 | 63.0 | 0.02 | 69.2 | 67.9 | 0.03 |
| Antibacterials for systemic use | 66.0 | 63.1 | 0.06 | 63.8 | 63.4 | 0.01 | 63.8 | 63.4 | 0.01 |
| Antidepressants | 47.7 | 34.8 | 0.26 | 44.5 | 45.5 | -0.02 | 37.2 | 36.9 | 0.01 |
| Antiepileptics | 25.6 | 19.7 | 0.14 | 24.1 | 24.6 | -0.01 | 21.4 | 20.7 | 0.02 |
| Antiinflammatory and antirheumatic products | 39.7 | 32.5 | 0.15 | 37.9 | 38.9 | -0.02 | 33.4 | 33.8 | -0.01 |
| Antineoplastic agents | 6.2 | 5.6 | 0.03 | 5.9 | 5.7 | 0.01 | 5.8 | 5.7 | 0.01 |
| Antipsoriatics | 0.7 | 0.6 | 0.02 | 0.6 | 0.7 | -0.01 | 0.6 | 0.6 | 0.00 |
| Antithrombotic agents | 10.5 | 14.5 | -0.12 | 10.7 | 11.0 | -0.01 | 15.4 | 13.9 | 0.04 |
| Beta blocking agents | 28.7 | 35.9 | -0.16 | 29.5 | 28.9 | 0.01 | 35.9 | 34.7 | 0.03 |
| Calcium channel blockers | 20.5 | 27.7 | -0.17 | 21.7 | 21.4 | 0.01 | 28.1 | 26.6 | 0.03 |
| Diuretics | 46.0 | 48.7 | -0.06 | 46.2 | 45.8 | 0.01 | 49.9 | 48.3 | 0.03 |
| Drugs for acid related disorders | 31.9 | 31.8 | 0.00 | 31.4 | 32.0 | -0.01 | 33.7 | 31.9 | 0.04 |
| Drugs for obstructive airway diseases | 49.2 | 40.9 | 0.17 | 46.7 | 47.7 | -0.02 | 44.9 | 42.3 | 0.05 |
| Immunosuppressants | 4.0 | 3.2 | 0.05 | 3.6 | 3.6 | 0.00 | 4.0 | 3.3 | 0.04 |
| Lipid modifying agents | 64.0 | 70.5 | -0.14 | 66.5 | 66.4 | 0.00 | 70.4 | 69.7 | 0.02 |
| Opioids | 31.6 | 30.3 | 0.03 | 30.4 | 30.8 | -0.01 | 31.1 | 30.5 | 0.01 |
| Psycholeptics | 32.8 | 27.8 | 0.11 | 30.7 | 31.0 | -0.01 | 27.2 | 28.5 | -0.03 |
| Psychostimulants | 4.6 | 1.9 | 0.15 | 3.5 | 3.5 | 0.00 | 2.4 | 2.3 | 0.01 |

##### 2.5.3 MDCR

**Supplementary Table 187:** Baseline patient characteristics for GLP1RA (female) (T) and SU (female) (C) new-users in the MDCR data source. We report proportion of initiators satisfying selected base-line characteristics and the standardized difference of population proportions (StdDiff) before and after propensity score adjustment. Less extreme StdDiffs through matching and stratification suggest improved balance between patient cohorts through adjustment.

| | Before adjustment | | | After matching | | | After stratification | | |
|---|---|---|---|---|---|---|---|---|---|
| Characteristic | T (%) | C (%) | StdDiff | T (%) | C (%) | StdDiff | T (%) | C (%) | StdDiff |
| **Age group** | | | | | | | | | |
| 50 - 54 | 0.5 | 0.1 | 0.06 | 0.5 | 0.2 | 0.06 | 0.2 | 0.1 | 0.01 |
| 55 - 59 | 1.1 | 0.6 | 0.06 | 1.1 | 1.2 | -0.01 | 1.0 | 0.7 | 0.04 |
| 60 - 64 | 2.7 | 1.1 | 0.12 | 2.2 | 2.0 | 0.02 | 1.2 | 1.3 | 0.00 |
| 65 - 69 | 43.2 | 26.5 | 0.36 | 43.3 | 41.6 | 0.03 | 35.3 | 28.5 | 0.15 |
| 70 - 74 | 34.8 | 28.2 | 0.14 | 34.0 | 36.0 | -0.04 | 29.2 | 28.4 | 0.02 |
| 75 - 79 | 12.6 | 21.2 | -0.23 | 13.3 | 12.7 | 0.02 | 17.3 | 19.8 | -0.06 |
| 80 - 84 | 4.1 | 13.3 | -0.33 | 4.5 | 4.0 | 0.02 | 10.0 | 12.6 | -0.08 |
| 85 - 89 | 0.8 | 6.5 | -0.31 | 0.9 | 1.2 | -0.03 | 5.7 | 6.2 | -0.02 |
| 90 - 94 | <0.4 | 2.1 | -0.17 | <0.5 | 0.9 | -0.09 | <0.2 | 2.0 | -0.20 |
| **Medical history: General** | | | | | | | | | |
| Acute respiratory disease | 24.6 | 22.8 | 0.04 | 24.0 | 24.4 | -0.01 | 22.9 | 23.6 | -0.02 |
| Attention deficit hyperactivity disorder | 0.5 | 0.1 | 0.07 | <0.5 | 0.5 | -0.08 | <0.2 | 0.1 | -0.01 |
| Chronic liver disease | 2.2 | 2.4 | -0.01 | 2.5 | 2.1 | 0.02 | 3.7 | 2.6 | 0.07 |
| Chronic obstructive lung disease | 9.7 | 10.3 | -0.02 | 9.3 | 10.5 | -0.04 | 11.3 | 10.6 | 0.02 |
| Crohn's disease | <0.4 | 0.3 | 0.00 | <0.5 | 0.4 | 0.00 | 0.2 | 0.4 | -0.02 |
| Dementia | 1.2 | 3.4 | -0.15 | 1.4 | 1.6 | -0.01 | 2.9 | 3.5 | -0.03 |
| Depressive disorder | 17.7 | 9.0 | 0.26 | 15.8 | 17.5 | -0.05 | 11.8 | 10.6 | 0.04 |
| Gastroesophageal reflux disease | 17.5 | 11.6 | 0.17 | 16.1 | 17.0 | -0.02 | 8.0 | 13.0 | -0.16 |
| Gastrointestinal hemorrhage | 2.6 | 3.6 | -0.06 | 2.6 | 2.5 | 0.00 | 4.6 | 3.5 | 0.06 |
| Hyperlipidemia | 64.1 | 48.0 | 0.33 | 61.7 | 64.9 | -0.06 | 63.4 | 53.4 | 0.20 |
| Hypertensive disorder | 72.9 | 68.1 | 0.10 | 72.0 | 74.7 | -0.06 | 78.2 | 71.1 | 0.16 |
| Lesion of liver | 0.6 | 1.2 | -0.06 | 0.6 | 1.0 | -0.04 | 0.4 | 1.2 | -0.10 |
| Obesity | 29.3 | 8.4 | 0.56 | 25.0 | 28.0 | -0.07 | 12.6 | 10.7 | 0.06 |
| Osteoarthritis | 40.9 | 29.8 | 0.23 | 38.5 | 39.7 | -0.03 | 34.7 | 31.7 | 0.06 |
| Pneumonia | 3.2 | 5.8 | -0.13 | 2.9 | 3.3 | -0.02 | 2.9 | 5.6 | -0.13 |
| Psoriasis | 2.3 | 1.3 | 0.07 | 2.1 | 1.8 | 0.02 | 1.4 | 1.5 | -0.01 |
| Renal impairment | 10.8 | 11.6 | -0.03 | 10.6 | 11.8 | -0.04 | 17.4 | 13.0 | 0.12 |
| Rheumatoid arthritis | 2.2 | 2.1 | 0.00 | 1.8 | 2.6 | -0.05 | 1.0 | 2.3 | -0.10 |
| Schizophrenia | <0.4 | 0.2 | 0.00 | <0.5 | 0.1 | 0.03 | 0.2 | 0.1 | 0.01 |
| Ulcerative colitis | <0.4 | 0.4 | -0.03 | <0.5 | 0.3 | -0.02 | <0.2 | 0.4 | -0.06 |
| Urinary tract infectious disease | 13.7 | 15.0 | -0.04 | 13.5 | 12.6 | 0.03 | 16.9 | 15.3 | 0.04 |
| Visual system disorder | 54.7 | 50.0 | 0.09 | 53.3 | 55.7 | -0.05 | 53.0 | 51.3 | 0.03 |
| **Medical history: Cardiovascular disease** | | | | | | | | | |
| Atrial fibrillation | 6.2 | 9.5 | -0.12 | 6.1 | 5.9 | 0.01 | 9.0 | 9.5 | -0.02 |
| Cerebrovascular disease | 8.9 | 11.4 | -0.08 | 8.2 | 8.5 | -0.01 | 15.6 | 11.1 | 0.13 |
| Coronary arteriosclerosis | 14.5 | 17.0 | -0.07 | 14.3 | 14.7 | -0.01 | 22.5 | 16.9 | 0.14 |
| Heart disease | 33.4 | 37.9 | -0.09 | 32.9 | 33.6 | -0.02 | 42.2 | 37.6 | 0.10 |
| Heart failure | 6.3 | 9.9 | -0.14 | 6.1 | 6.2 | 0.00 | 14.4 | 9.6 | 0.15 |
| Ischemic heart disease | 7.1 | 8.9 | -0.07 | 10.5 | 8.4 | 0.07 | 10.5 | 8.4 | 0.07 |
| Peripheral vascular disease | 10.3 | 10.1 | 0.01 | 9.7 | 9.8 | 0.00 | 9.3 | 10.5 | -0.04 |
| Pulmonary embolism | 0.4 | 0.9 | -0.06 | <0.5 | 1.1 | -0.09 | 0.2 | 1.0 | -0.11 |
| Venous thrombosis | 1.7 | 2.3 | -0.04 | 1.6 | 1.9 | -0.02 | 3.7 | 2.4 | 0.07 |
| **Medical history: Neoplasms** | | | | | | | | | |
| Hematologic neoplasm | 1.1 | 1.6 | -0.04 | 1.3 | 1.3 | 0.00 | 0.9 | 1.7 | -0.07 |
| Malignant lymphoma | 0.6 | 0.7 | -0.01 | 0.6 | 0.6 | 0.00 | <0.2 | 0.7 | -0.10 |
| Malignant neoplasm of anorectum | <0.4 | 0.2 | -0.02 | <0.5 | 0.1 | 0.02 | 0.2 | 0.2 | 0.00 |
| Malignant neoplastic disease | 13.2 | 14.1 | -0.03 | 12.7 | 14.0 | -0.04 | 13.6 | 14.3 | -0.02 |
| Malignant tumor of breast | 6.4 | 5.7 | 0.03 | 6.1 | 6.5 | -0.02 | 6.2 | 5.7 | 0.02 |
| Malignant tumor of colon | <0.4 | 0.7 | -0.05 | <0.5 | 0.3 | -0.01 | <0.2 | 0.6 | -0.09 |
| Malignant tumor of lung | <0.4 | 0.7 | -0.08 | <0.5 | 0.5 | -0.06 | 1.1 | 0.7 | 0.04 |
| Malignant tumor of urinary bladder | 0.9 | 0.5 | 0.05 | 0.9 | 0.4 | 0.06 | 1.2 | 0.4 | 0.08 |
| **Medication use** | | | | | | | | | |
| Agents acting on the renin-angiotensin system | 74.7 | 72.0 | 0.06 | 75.6 | 75.3 | 0.01 | 81.2 | 74.3 | 0.17 |
| Antibacterials for systemic use | 67.3 | 64.3 | 0.06 | 66.7 | 66.9 | 0.00 | 64.1 | 65.2 | -0.02 |
| Antidepressants | 44.7 | 31.5 | 0.27 | 42.9 | 44.2 | -0.03 | 31.9 | 33.1 | -0.03 |
| Antiepileptics | 21.9 | 16.4 | 0.14 | 19.9 | 22.6 | -0.07 | 20.7 | 17.9 | 0.07 |
| Antiinflammatory and antirheumatic products | 40.1 | 29.2 | 0.23 | 38.1 | 39.5 | -0.03 | 34.1 | 30.0 | 0.09 |

| (Continued) | Before ajustment | | | After matching | | | After stratification | | |
|---|---|---|---|---|---|---|---|---|---|
| Characteristic | T (%) | C (%) | StdDiff | T (%) | C (%) | StdDiff | T (%) | C (%) | StdDiff |
| Antineoplastic agents | 11.3 | 9.4 | 0.06 | 11.1 | 10.8 | 0.01 | 8.3 | 9.0 | -0.02 |
| Antipsoriatics | 1.1 | 0.7 | 0.04 | 0.4 | 0.8 | -0.04 | 0.4 | 0.8 | -0.04 |
| Antithrombotic agents | 17.8 | 21.8 | -0.10 | 17.0 | 17.8 | -0.02 | 21.9 | 21.8 | 0.00 |
| Beta blocking agents | 41.8 | 47.4 | -0.11 | 42.3 | 42.2 | 0.00 | 50.1 | 48.4 | 0.04 |
| Calcium channel blockers | 30.6 | 35.2 | -0.10 | 30.4 | 30.3 | 0.00 | 36.1 | 35.4 | 0.01 |
| Diuretics | 57.7 | 59.2 | -0.03 | 57.6 | 57.6 | 0.00 | 64.5 | 59.4 | 0.10 |
| Drugs for acid related disorders | 39.6 | 35.2 | 0.09 | 38.6 | 39.5 | -0.02 | 32.9 | 35.9 | -0.06 |
| Drugs for obstructive airway diseases | 53.5 | 40.0 | 0.27 | 51.2 | 52.9 | -0.03 | 45.0 | 42.2 | 0.06 |
| Immunosuppressants | 5.4 | 3.5 | 0.09 | 5.0 | 4.8 | 0.01 | 3.4 | 3.9 | -0.03 |
| Lipid modifying agents | 80.1 | 73.6 | 0.15 | 79.9 | 80.6 | -0.02 | 79.7 | 76.8 | 0.07 |
| Opioids | 30.5 | 30.8 | 0.00 | 30.5 | 32.2 | -0.04 | 34.1 | 31.5 | 0.06 |
| Psycholeptics | 31.4 | 29.4 | 0.04 | 31.3 | 32.3 | -0.02 | 34.1 | 30.0 | 0.09 |
| Psychostimulants | 2.4 | 1.4 | 0.07 | 2.0 | 2.6 | -0.04 | 0.9 | 1.6 | -0.06 |

##### 2.5.4 MDCD

**Supplementary Table 188:** Baseline patient characteristics for GLP1RA (female) (T) and SU (female) (C) new-users in the MDCD data source. We report proportion of initiators satisfying selected base-line characteristics and the standardized difference of population proportions (StdDiff) before and after propensity score adjustment. Less extreme StdDiffs through matching and stratification suggest improved balance between patient cohorts through adjustment.

| | Before adjustment | | | After matching | | | After stratification | | |
|---|---|---|---|---|---|---|---|---|---|
| Characteristic | T (%) | C (%) | StdDiff | T (%) | C (%) | StdDiff | T (%) | C (%) | StdDiff |
| **Age group** | | | | | | | | | |
| 15 - 19 | 1.7 | 0.6 | 0.10 | 1.5 | 0.9 | 0.06 | 0.8 | 0.6 | 0.02 |
| 20 - 24 | 1.8 | 1.4 | 0.03 | 1.8 | 1.4 | 0.03 | 0.7 | 1.4 | -0.07 |
| 25 - 29 | 4.4 | 3.6 | 0.04 | 4.5 | 3.7 | 0.04 | 3.7 | 3.6 | 0.01 |
| 30 - 34 | 8.8 | 6.4 | 0.09 | 8.3 | 8.1 | 0.01 | 6.3 | 6.6 | -0.01 |
| 35 - 39 | 13.3 | 9.3 | 0.13 | 12.8 | 13.2 | -0.01 | 11.5 | 9.7 | 0.06 |
| 40 - 44 | 15.2 | 11.5 | 0.11 | 14.9 | 15.1 | 0.00 | 11.6 | 11.9 | -0.01 |
| 45 - 49 | 15.6 | 13.0 | 0.07 | 15.5 | 15.3 | 0.00 | 13.1 | 13.3 | -0.01 |
| 50 - 54 | 14.4 | 15.3 | -0.02 | 14.7 | 14.1 | 0.02 | 14.6 | 15.1 | -0.01 |
| 55 - 59 | 12.4 | 16.1 | -0.11 | 12.8 | 13.9 | -0.03 | 13.5 | 15.8 | -0.06 |
| 60 - 64 | 8.7 | 13.2 | -0.15 | 9.2 | 9.2 | 0.00 | 18.3 | 12.8 | 0.15 |
| 65 - 69 | 2.4 | 4.3 | -0.10 | 2.6 | 2.8 | -0.01 | 3.0 | 4.1 | -0.06 |
| 70 - 74 | 0.6 | 2.2 | -0.13 | 0.7 | 1.1 | -0.04 | 1.2 | 2.1 | -0.07 |
| 75 - 79 | 0.3 | 1.5 | -0.12 | 0.4 | 0.7 | -0.04 | 0.6 | 1.4 | -0.08 |
| 80 - 84 | 0.3 | 1.0 | -0.08 | 0.4 | 0.5 | -0.01 | 0.8 | 0.9 | -0.01 |
| **Race** | | | | | | | | | |
| Black or African American | 27.2 | 30.9 | -0.08 | 27.8 | 25.6 | 0.05 | 27.4 | 30.3 | -0.06 |
| White | 61.2 | 51.1 | 0.20 | 60.3 | 62.6 | -0.05 | 53.5 | 52.3 | 0.02 |
| **Ethnicity** | | | | | | | | | |
| Hispanic or Latino | 3.6 | 2.7 | 0.05 | 3.6 | 3.0 | 0.03 | 3.3 | 2.8 | 0.03 |
| **Medical history: General** | | | | | | | | | |
| Acute respiratory disease | 44.3 | 41.0 | 0.07 | 44.4 | 45.2 | -0.02 | 37.4 | 41.4 | -0.08 |
| Attention deficit hyperactivity disorder | 4.2 | 1.5 | 0.16 | 3.6 | 3.3 | 0.01 | 1.7 | 1.7 | 0.00 |
| Chronic liver disease | 3.6 | 5.7 | -0.10 | 3.9 | 4.1 | -0.01 | 4.0 | 5.5 | -0.07 |
| Chronic obstructive lung disease | 17.8 | 18.5 | -0.02 | 18.2 | 18.8 | -0.02 | 18.7 | 18.6 | 0.00 |
| Crohn’s disease | 0.4 | 0.3 | 0.02 | 0.4 | 0.3 | 0.01 | 0.5 | 0.3 | 0.03 |
| Dementia | 1.0 | 1.5 | -0.05 | 1.0 | 0.9 | 0.01 | 4.3 | 1.5 | 0.17 |
| Depressive disorder | 43.6 | 36.5 | 0.14 | 42.6 | 44.6 | -0.04 | 41.4 | 37.4 | 0.08 |
| Gastroesophageal reflux disease | 29.8 | 27.1 | 0.06 | 29.2 | 30.7 | -0.03 | 28.6 | 27.5 | 0.02 |
| Gastrointestinal hemorrhage | 3.6 | 3.8 | -0.01 | 3.4 | 3.6 | -0.01 | 4.3 | 3.8 | 0.03 |
| Human immunodeficiency virus infection | 0.5 | 0.6 | -0.02 | 0.5 | 0.4 | 0.01 | 0.2 | 0.6 | -0.07 |
| Hyperlipidemia | 57.8 | 59.7 | -0.04 | 58.2 | 59.1 | -0.02 | 61.0 | 59.6 | 0.03 |
| Hypertensive disorder | 70.0 | 74.1 | -0.09 | 70.9 | 69.9 | 0.02 | 77.2 | 73.7 | 0.08 |
| Lesion of liver | 1.0 | 1.4 | -0.03 | 1.1 | 1.3 | -0.02 | 0.8 | 1.4 | -0.06 |
| Obesity | 58.2 | 39.5 | 0.38 | 56.0 | 60.3 | -0.09 | 40.9 | 41.7 | -0.01 |
| Osteoarthritis | 39.1 | 34.5 | 0.10 | 38.2 | 40.1 | -0.04 | 35.8 | 35.1 | 0.01 |
| Pneumonia | 5.8 | 6.7 | -0.04 | 6.0 | 6.0 | 0.00 | 4.9 | 6.6 | -0.07 |
| Psoriasis | 1.0 | 1.2 | -0.02 | 1.0 | 1.7 | -0.06 | 3.0 | 1.3 | 0.12 |
| Renal impairment | 5.7 | 8.1 | -0.10 | 5.8 | 5.8 | 0.00 | 5.8 | 7.9 | -0.08 |
| Rheumatoid arthritis | 2.3 | 2.3 | 0.00 | 2.2 | 2.6 | -0.03 | 1.5 | 2.4 | -0.07 |
| Schizophrenia | 2.1 | 4.2 | -0.12 | 2.2 | 2.3 | 0.00 | 2.7 | 4.0 | -0.07 |
| Ulcerative colitis | <0.2 | 0.2 | -0.03 | <0.3 | 0.1 | 0.00 | <0.1 | 0.2 | -0.04 |
| Urinary tract infectious disease | 15.8 | 18.6 | -0.07 | 15.6 | 15.3 | 0.01 | 16.5 | 18.3 | -0.05 |
| Viral hepatitis C | 1.7 | 2.1 | -0.04 | 1.7 | 1.9 | -0.01 | 2.4 | 2.1 | 0.02 |
| Visual system disorder | 43.5 | 41.5 | 0.04 | 43.3 | 44.9 | -0.03 | 46.1 | 41.8 | 0.09 |
| **Medical history: Cardiovascular disease** | | | | | | | | | |
| Atrial fibrillation | 2.0 | 2.8 | -0.05 | 2.1 | 1.9 | 0.01 | 2.0 | 2.7 | -0.05 |
| Cerebrovascular disease | 3.6 | 5.1 | -0.07 | 3.6 | 3.5 | 0.00 | 4.1 | 4.9 | -0.04 |
| Coronary arteriosclerosis | 6.9 | 9.3 | -0.09 | 7.2 | 6.3 | 0.03 | 10.7 | 9.0 | 0.06 |
| Heart disease | 23.3 | 26.8 | -0.08 | 23.6 | 22.4 | 0.03 | 26.5 | 26.3 | 0.00 |
| Heart failure | 6.8 | 8.4 | -0.06 | 7.2 | 6.6 | 0.03 | 6.8 | 8.2 | -0.06 |
| Ischemic heart disease | 4.7 | 6.9 | -0.10 | 4.8 | 4.9 | 0.00 | 7.4 | 6.7 | 0.03 |
| Peripheral vascular disease | 5.7 | 6.9 | -0.05 | 5.7 | 6.2 | -0.02 | 4.8 | 6.8 | -0.09 |
| Pulmonary embolism | 1.1 | 1.1 | 0.00 | 1.1 | 1.1 | 0.00 | 1.6 | 1.1 | 0.04 |
| Venous thrombosis | 1.7 | 1.7 | 0.00 | 1.7 | 1.6 | 0.01 | 2.3 | 1.7 | 0.04 |
| **Medical history: Neoplasms** | | | | | | | | | |
| Hematologic neoplasm | 0.6 | 1.0 | -0.04 | 0.6 | 0.8 | -0.02 | 0.8 | 1.0 | -0.02 |
| Malignant lymphoma | 0.2 | 0.3 | -0.02 | <0.3 | 0.4 | -0.03 | 0.2 | 0.3 | -0.03 |

(Continued)

| Characteristic | Before ajustment | | | After matching | | | After stratification | | |
|---|---|---|---|---|---|---|---|---|---|
| | T (%) | C (%) | StdDiff | T (%) | C (%) | StdDiff | T (%) | C (%) | StdDiff |
| Malignant neoplastic disease | 4.6 | 6.5 | -0.08 | 4.8 | 5.3 | -0.02 | 8.1 | 6.4 | 0.07 |
| Malignant tumor of breast | 1.4 | 2.4 | -0.07 | 1.5 | 1.6 | 0.00 | 1.9 | 2.3 | -0.03 |
| Malignant tumor of lung | <0.2 | 0.3 | -0.03 | <0.3 | 0.3 | -0.01 | 0.2 | 0.3 | -0.02 |
| Malignant tumor of urinary bladder | <0.2 | 0.1 | 0.00 | <0.3 | 0.1 | 0.01 | <0.1 | 0.1 | -0.01 |
| **Medication use** | | | | | | | | | |
| Agents acting on the renin-angiotensin system | 55.5 | 60.7 | -0.11 | 56.2 | 55.1 | 0.02 | 68.3 | 60.1 | 0.17 |
| Antibacterials for systemic use | 77.5 | 72.6 | 0.12 | 76.8 | 77.1 | -0.01 | 73.7 | 73.0 | 0.01 |
| Antidepressants | 64.1 | 53.9 | 0.21 | 63.0 | 65.1 | -0.04 | 57.1 | 55.1 | 0.04 |
| Antiepileptics | 47.0 | 39.1 | 0.16 | 45.8 | 47.4 | -0.03 | 31.9 | 40.0 | -0.17 |
| Antiinflammatory and antirheumatic products | 65.3 | 59.9 | 0.11 | 64.5 | 65.7 | -0.03 | 61.4 | 60.5 | 0.02 |
| Antineoplastic agents | 5.4 | 4.3 | 0.05 | 5.1 | 4.7 | 0.02 | 4.0 | 4.3 | -0.02 |
| Antipsoriatics | 0.9 | 0.4 | 0.06 | 0.8 | 0.6 | 0.03 | 0.6 | 0.5 | 0.01 |
| Antithrombotic agents | 18.6 | 22.4 | -0.09 | 19.3 | 19.1 | 0.00 | 27.4 | 22.0 | 0.12 |
| Beta blocking agents | 27.6 | 31.8 | -0.09 | 28.0 | 27.7 | 0.01 | 38.4 | 31.3 | 0.15 |
| Calcium channel blockers | 20.6 | 24.8 | -0.10 | 21.4 | 20.1 | 0.03 | 23.8 | 24.3 | -0.01 |
| Diuretics | 45.9 | 48.4 | -0.05 | 46.6 | 46.0 | 0.01 | 52.7 | 48.1 | 0.09 |
| Drugs for acid related disorders | 52.0 | 48.6 | 0.07 | 51.1 | 51.6 | -0.01 | 47.2 | 48.9 | -0.03 |
| Drugs for obstructive airway diseases | 66.4 | 58.1 | 0.17 | 66.1 | 67.7 | -0.03 | 62.5 | 59.1 | 0.07 |
| Immunosuppressants | 3.0 | 2.4 | 0.04 | 2.9 | 2.8 | 0.00 | 2.8 | 2.4 | 0.02 |
| Lipid modifying agents | 56.3 | 60.1 | -0.08 | 57.5 | 57.0 | 0.01 | 61.4 | 59.8 | 0.03 |
| Opioids | 49.7 | 49.4 | 0.00 | 49.0 | 48.9 | 0.00 | 46.7 | 49.4 | -0.05 |
| Psycholeptics | 54.4 | 49.0 | 0.11 | 53.5 | 54.2 | -0.01 | 48.8 | 49.6 | -0.02 |
| Psychostimulants | 9.4 | 5.2 | 0.16 | 8.2 | 8.5 | -0.01 | 4.6 | 5.5 | -0.04 |

##### 2.5.5 Open Claims

**Supplementary Table 189:** Baseline patient characteristics for GLP1RA (female) (T) and SU (female) (C) new-users in the Open Claims data source. We report proportion of initiators satisfying selected base-line characteristics and the standardized difference of population proportions (StdDiff) before and after propensity score adjustment. Less extreme StdDiffs through matching and stratification suggest improved balance between patient cohorts through adjustment.

| | Before adjustment | | | After matching | | | After stratification | | |
|---|---|---|---|---|---|---|---|---|---|
| Characteristic | T (%) | C (%) | StdDiff | T (%) | C (%) | StdDiff | T (%) | C (%) | StdDiff |
| **Age group** | | | | | | | | | |
| 15 - 19 | 0.2 | 0.1 | 0.03 | 0.2 | 0.2 | 0.00 | 0.2 | 0.1 | 0.01 |
| 20 - 24 | 0.9 | 0.5 | 0.04 | 0.8 | 0.7 | 0.01 | 0.6 | 0.6 | 0.00 |
| 25 - 29 | 1.7 | 1.1 | 0.06 | 1.6 | 1.4 | 0.01 | 1.2 | 1.2 | 0.00 |
| 30 - 34 | 3.1 | 2.0 | 0.07 | 2.2 | 2.3 | 0.00 | 2.2 | 2.3 | 0.00 |
| 35 - 39 | 5.4 | 3.4 | 0.10 | 4.8 | 4.7 | 0.00 | 3.8 | 3.8 | 0.00 |
| 40 - 44 | 8.5 | 5.2 | 0.13 | 7.7 | 7.6 | 0.00 | 7.7 | 7.6 | 0.00 |
| 45 - 49 | 12.0 | 7.8 | 0.14 | 11.2 | 11.3 | 0.00 | 8.7 | 8.8 | 0.00 |
| 50 - 54 | 15.5 | 11.2 | 0.13 | 14.6 | 14.8 | 0.00 | 11.9 | 12.1 | -0.01 |
| 55 - 59 | 16.4 | 13.6 | 0.08 | 16.2 | 16.2 | 0.00 | 14.1 | 14.1 | 0.00 |
| 60 - 64 | 14.5 | 14.0 | 0.02 | 15.2 | 14.8 | 0.01 | 14.3 | 14.2 | 0.00 |
| 65 - 69 | 10.8 | 14.7 | -0.12 | 12.0 | 12.2 | -0.01 | 14.2 | 14.0 | 0.01 |
| 70 - 74 | 6.5 | 11.4 | -0.17 | 10.9 | 10.4 | 0.01 | 10.9 | 10.4 | 0.01 |
| 75 - 79 | 3.0 | 10.7 | -0.31 | 3.6 | 3.6 | 0.00 | 8.2 | 9.0 | -0.03 |
| 80 - 84 | 1.3 | 4.1 | -0.17 | 1.7 | 1.8 | -0.01 | 3.7 | 3.5 | 0.01 |
| 85 - 89 | 0.1 | 0.1 | 0.01 | 0.1 | 0.2 | -0.02 | 0.1 | 0.1 | -0.01 |
| **Medical history: General** | | | | | | | | | |
| Acute respiratory disease | 16.8 | 16.0 | 0.02 | 15.7 | 16.5 | -0.02 | 16.5 | 16.4 | 0.00 |
| Attention deficit hyperactivity disorder | 1.2 | 0.4 | 0.10 | 0.8 | 0.9 | 0.00 | 0.6 | 0.5 | 0.01 |
| Chronic liver disease | 0.8 | 1.6 | -0.08 | 0.9 | 1.0 | -0.01 | 1.4 | 1.4 | -0.01 |
| Chronic obstructive lung disease | 4.1 | 5.9 | -0.08 | 4.3 | 4.7 | -0.02 | 5.9 | 5.7 | 0.01 |
| Crohn’s disease | 0.2 | 0.2 | 0.00 | 0.2 | 0.2 | -0.01 | 0.2 | 0.2 | -0.01 |
| Dementia | 0.3 | 1.0 | -0.09 | 0.3 | 0.4 | -0.01 | 0.9 | 0.9 | 0.00 |
| Depressive disorder | 12.8 | 9.5 | 0.10 | 11.3 | 11.8 | -0.02 | 10.8 | 10.3 | 0.02 |
| Gastroesophageal reflux disease | 10.9 | 9.2 | 0.06 | 9.8 | 10.3 | -0.02 | 10.1 | 9.7 | 0.01 |
| Gastrointestinal hemorrhage | 1.1 | 1.4 | -0.03 | 1.1 | 1.1 | 0.00 | 1.4 | 1.4 | 0.00 |
| Human immunodeficiency virus infection | 0.2 | 0.2 | 0.00 | 0.2 | 0.2 | 0.00 | 0.2 | 0.2 | 0.01 |
| Hyperlipidemia | 37.9 | 39.2 | -0.03 | 36.9 | 37.6 | -0.01 | 40.0 | 39.1 | 0.02 |
| Hypertensive disorder | 44.9 | 48.3 | -0.07 | 44.4 | 44.7 | -0.01 | 48.3 | 47.7 | 0.01 |
| Lesion of liver | 0.5 | 0.7 | -0.02 | 0.5 | 0.6 | -0.01 | 0.6 | 0.7 | -0.02 |
| Obesity | 21.6 | 11.8 | 0.27 | 17.1 | 17.5 | -0.01 | 14.3 | 13.7 | 0.02 |
| Osteoarthritis | 17.2 | 16.1 | 0.03 | 16.0 | 16.6 | -0.02 | 17.4 | 16.5 | 0.02 |
| Pneumonia | 2.2 | 2.8 | -0.04 | 2.2 | 2.2 | -0.01 | 2.7 | 2.6 | 0.00 |
| Psoriasis | 1.4 | 0.8 | 0.05 | 1.1 | 1.2 | 0.00 | 1.0 | 0.9 | 0.01 |
| Renal impairment | 4.0 | 7.0 | -0.13 | 4.2 | 4.3 | -0.01 | 4.2 | 4.3 | -0.01 |
| Rheumatoid arthritis | 1.5 | 1.4 | 0.01 | 1.4 | 1.4 | 0.00 | 1.6 | 1.5 | 0.01 |
| Schizophrenia | 0.3 | 0.4 | -0.03 | 0.3 | 0.3 | -0.01 | 0.3 | 0.4 | -0.01 |
| Ulcerative colitis | 0.2 | 0.2 | 0.00 | 0.2 | 0.2 | -0.01 | 0.2 | 0.2 | -0.01 |
| Urinary tract infectious disease | 6.6 | 8.2 | -0.06 | 6.5 | 6.8 | -0.01 | 8.3 | 8.0 | 0.01 |
| Viral hepatitis C | 0.2 | 0.4 | -0.03 | 0.2 | 0.2 | -0.01 | 0.3 | 0.3 | -0.01 |
| Visual system disorder | 16.9 | 19.3 | -0.06 | 16.8 | 17.3 | -0.01 | 19.0 | 18.9 | 0.00 |
| **Medical history: Cardiovascular disease** | | | | | | | | | |
| Atrial fibrillation | 1.8 | 3.2 | -0.09 | 1.9 | 2.0 | 0.00 | 3.1 | 2.9 | 0.01 |
| Cerebrovascular disease | 1.8 | 3.1 | -0.09 | 1.9 | 1.9 | 0.00 | 3.1 | 2.9 | 0.01 |
| Coronary arteriosclerosis | 3.6 | 5.7 | -0.10 | 3.7 | 4.1 | -0.02 | 5.3 | 5.4 | 0.00 |
| Heart disease | 11.3 | 15.3 | -0.12 | 11.2 | 11.8 | -0.02 | 15.0 | 14.7 | 0.01 |
| Heart failure | 2.3 | 4.0 | -0.10 | 2.3 | 2.6 | -0.02 | 3.6 | 3.7 | -0.01 |
| Ischemic heart disease | 2.0 | 3.2 | -0.07 | 2.1 | 2.2 | -0.01 | 2.8 | 3.0 | -0.01 |
| Peripheral vascular disease | 3.2 | 4.2 | -0.05 | 3.3 | 3.3 | 0.00 | 4.2 | 3.9 | 0.01 |
| Pulmonary embolism | 0.5 | 0.5 | 0.01 | 0.5 | 0.5 | 0.00 | 0.6 | 0.5 | 0.01 |
| Venous thrombosis | 0.6 | 0.8 | -0.03 | 0.6 | 0.7 | -0.01 | 0.8 | 0.8 | -0.01 |
| **Medical history: Neoplasms** | | | | | | | | | |
| Hematologic neoplasm | 0.4 | 0.7 | -0.04 | 0.4 | 0.4 | 0.00 | 0.6 | 0.7 | -0.01 |
| Malignant lymphoma | 0.2 | 0.4 | -0.02 | 0.2 | 0.3 | 0.00 | 0.3 | 0.3 | 0.00 |
| Malignant neoplasm of anorectum | 0.1 | 0.1 | -0.02 | 0.1 | 0.1 | 0.00 | 0.1 | 0.1 | -0.01 |
| Malignant neoplastic disease | 4.4 | 5.7 | -0.06 | 4.4 | 4.6 | -0.01 | 5.4 | 5.5 | 0.00 |
| Malignant tumor of breast | 1.7 | 2.2 | -0.04 | 1.7 | 1.8 | 0.00 | 1.7 | 1.8 | 0.00 |
| Malignant tumor of colon | 0.1 | 0.3 | -0.03 | 0.2 | 0.1 | 0.01 | 0.2 | 0.3 | -0.01 |

| (Continued) | Before ajustment | | | After matching | | | After stratification | | |
|---|---|---|---|---|---|---|---|---|---|
| Characteristic | T (%) | C (%) | StdDiff | T (%) | C (%) | StdDiff | T (%) | C (%) | StdDiff |
| Malignant tumor of lung | 0.1 | 0.3 | -0.04 | 0.1 | 0.2 | -0.01 | 0.2 | 0.3 | -0.01 |
| Malignant tumor of urinary bladder | 0.1 | 0.1 | -0.02 | 0.1 | 0.1 | 0.00 | 0.1 | 0.1 | -0.01 |
| **Medication use** | | | | | | | | | |
| Agents acting on the renin-angiotensin system | 57.2 | 65.7 | -0.17 | 59.4 | 59.9 | -0.01 | 59.4 | 59.9 | -0.01 |
| Antibacterials for systemic use | 63.0 | 60.5 | 0.05 | 61.3 | 62.4 | -0.02 | 61.7 | 61.3 | 0.01 |
| Antidepressants | 47.3 | 35.9 | 0.23 | 43.2 | 44.0 | -0.02 | 40.1 | 38.4 | 0.03 |
| Antiepileptics | 26.4 | 22.8 | 0.08 | 24.5 | 25.3 | -0.02 | 25.0 | 23.8 | 0.03 |
| Antiinflammatory and antirheumatic products | 41.9 | 39.3 | 0.05 | 40.5 | 41.5 | -0.02 | 40.5 | 40.1 | 0.01 |
| Antineoplastic agents | 6.3 | 4.6 | 0.08 | 5.3 | 5.6 | -0.01 | 5.3 | 5.0 | 0.01 |
| Antipsoriatics | 2.1 | 1.1 | 0.08 | 1.6 | 1.6 | 0.01 | 1.3 | 1.2 | 0.01 |
| Antithrombotic agents | 11.7 | 17.2 | -0.16 | 12.2 | 12.7 | -0.02 | 16.1 | 16.2 | 0.00 |
| Beta blocking agents | 27.4 | 33.3 | -0.13 | 27.7 | 28.5 | -0.02 | 32.3 | 32.3 | 0.00 |
| Calcium channel blockers | 20.4 | 25.5 | -0.12 | 21.1 | 21.3 | 0.00 | 24.3 | 24.4 | 0.00 |
| Diuretics | 43.0 | 44.0 | -0.02 | 42.3 | 42.5 | 0.00 | 44.1 | 43.8 | 0.01 |
| Drugs for acid related disorders | 37.5 | 36.6 | 0.02 | 35.8 | 37.0 | -0.03 | 37.3 | 37.1 | 0.00 |
| Drugs for obstructive airway diseases | 46.8 | 39.9 | 0.14 | 44.2 | 45.0 | -0.01 | 42.6 | 41.3 | 0.03 |
| Immunosuppressants | 4.1 | 2.9 | 0.06 | 3.5 | 3.8 | -0.01 | 3.3 | 3.2 | 0.00 |
| Lipid modifying agents | 60.7 | 66.2 | -0.11 | 62.3 | 63.5 | -0.03 | 65.9 | 65.4 | 0.01 |
| Opioids | 26.9 | 28.2 | -0.03 | 26.2 | 27.0 | -0.02 | 29.2 | 28.2 | 0.02 |
| Psycholeptics | 30.3 | 26.7 | 0.08 | 28.3 | 28.9 | -0.01 | 29.0 | 27.6 | 0.03 |
| Psychostimulants | 5.6 | 2.7 | 0.14 | 4.2 | 4.4 | -0.01 | 3.6 | 3.3 | 0.01 |

##### 2.5.6 OEHR

**Supplementary Table 190:** Baseline patient characteristics for GLP1RA (female) (T) and SU (female) (C) new-users in the OEHR data source. We report proportion of initiators satisfying selected base-line characteristics and the standardized difference of population proportions (StdDiff) before and after propensity score adjustment. Less extreme StdDiffs through matching and stratification suggest improved balance between patient cohorts through adjustment.

| | Before adjustment | | | After matching | | | After stratification | | |
|---|---|---|---|---|---|---|---|---|---|
| Characteristic | T (%) | C (%) | StdDiff | T (%) | C (%) | StdDiff | T (%) | C (%) | StdDiff |
| **Age group** | | | | | | | | | |
| 15 - 19 | 0.2 | 0.1 | 0.03 | 0.2 | 0.1 | 0.01 | 0.1 | 0.1 | 0.00 |
| 20 - 24 | 1.0 | 0.6 | 0.05 | 0.9 | 0.8 | 0.01 | 0.7 | 0.6 | 0.01 |
| 25 - 29 | 2.1 | 1.3 | 0.06 | 2.0 | 1.9 | 0.01 | 1.6 | 1.4 | 0.01 |
| 30 - 34 | 3.8 | 2.4 | 0.08 | 3.5 | 3.4 | 0.00 | 2.8 | 2.6 | 0.01 |
| 35 - 39 | 7.1 | 4.2 | 0.12 | 6.4 | 6.6 | 0.00 | 4.7 | 4.6 | 0.00 |
| 40 - 44 | 10.0 | 6.2 | 0.14 | 9.3 | 9.4 | 0.00 | 6.6 | 6.7 | -0.01 |
| 45 - 49 | 13.7 | 9.6 | 0.13 | 13.1 | 13.1 | 0.00 | 9.5 | 10.1 | -0.02 |
| 50 - 54 | 16.3 | 12.9 | 0.10 | 16.3 | 16.3 | 0.00 | 14.2 | 13.5 | 0.02 |
| 55 - 59 | 16.6 | 15.1 | 0.04 | 16.8 | 17.1 | -0.01 | 14.6 | 15.4 | -0.02 |
| 60 - 64 | 14.6 | 14.3 | 0.01 | 15.2 | 15.2 | 0.00 | 14.5 | 14.4 | 0.00 |
| 65 - 69 | 8.1 | 12.2 | -0.14 | 8.7 | 8.5 | 0.01 | 13.3 | 11.6 | 0.05 |
| 70 - 74 | 4.3 | 9.3 | -0.20 | 4.9 | 4.8 | 0.00 | 4.9 | 4.8 | 0.00 |
| 75 - 79 | 1.6 | 6.7 | -0.26 | 1.9 | 1.8 | 0.01 | 4.9 | 6.0 | -0.04 |
| 80 - 84 | 0.5 | 4.1 | -0.25 | 0.6 | 0.6 | 0.00 | 3.1 | 3.6 | -0.03 |
| 85 - 89 | 0.1 | 0.9 | -0.11 | 0.2 | 0.3 | -0.02 | 1.0 | 0.8 | 0.02 |
| **Race** | | | | | | | | | |
| Asian | 1.5 | 2.8 | -0.09 | 1.6 | 1.6 | 0.00 | 2.3 | 2.6 | -0.02 |
| Black or African American | 14.5 | 13.9 | 0.02 | 14.5 | 14.2 | 0.01 | 14.6 | 13.9 | 0.02 |
| White | 76.9 | 74.7 | 0.05 | 76.6 | 76.9 | -0.01 | 75.7 | 75.1 | 0.01 |
| **Ethnicity** | | | | | | | | | |
| Hispanic or Latino | 5.9 | 8.3 | -0.09 | 6.2 | 6.2 | 0.00 | 7.7 | 8.0 | -0.01 |
| Not Hispanic or Latino | 85.9 | 84.5 | 0.04 | 85.9 | 85.8 | 0.00 | 85.6 | 84.7 | 0.02 |
| **Medical history: General** | | | | | | | | | |
| Acute respiratory disease | 22.7 | 20.3 | 0.06 | 21.9 | 22.4 | -0.01 | 21.8 | 20.7 | 0.03 |
| Attention deficit hyperactivity disorder | 1.4 | 0.7 | 0.07 | 1.2 | 1.3 | -0.01 | 0.9 | 0.8 | 0.01 |
| Chronic liver disease | 2.3 | 3.2 | -0.05 | 2.5 | 2.3 | 0.01 | 3.5 | 3.0 | 0.03 |
| Chronic obstructive lung disease | 4.1 | 5.7 | -0.07 | 4.3 | 4.5 | -0.01 | 6.7 | 5.6 | 0.05 |
| Crohn’s disease | 0.2 | 0.3 | -0.01 | 0.2 | 0.3 | -0.01 | 0.2 | 0.3 | -0.01 |
| Dementia | 0.4 | 1.0 | -0.07 | 0.4 | 0.4 | 0.01 | 0.8 | 0.9 | -0.02 |
| Depressive disorder | 24.5 | 18.6 | 0.14 | 23.0 | 23.4 | -0.01 | 20.7 | 19.4 | 0.03 |
| Gastroesophageal reflux disease | 21.2 | 17.9 | 0.08 | 19.9 | 20.1 | -0.01 | 18.7 | 18.3 | 0.01 |
| Gastrointestinal hemorrhage | 1.3 | 1.4 | -0.02 | 1.2 | 1.2 | 0.00 | 1.8 | 1.4 | 0.03 |
| Human immunodeficiency virus infection | 0.1 | 0.1 | 0.00 | 0.1 | 0.1 | 0.00 | 0.1 | 0.1 | 0.00 |
| Hyperlipidemia | 58.4 | 61.0 | -0.05 | 57.4 | 57.3 | 0.00 | 61.3 | 60.5 | 0.02 |
| Hypertensive disorder | 59.0 | 63.7 | -0.10 | 58.9 | 58.9 | 0.00 | 64.5 | 63.0 | 0.03 |
| Lesion of liver | 0.6 | 0.8 | -0.02 | 0.6 | 0.7 | -0.02 | 0.8 | 0.8 | 0.00 |
| Obesity | 46.7 | 26.3 | 0.43 | 41.2 | 42.6 | -0.03 | 30.8 | 29.0 | 0.04 |
| Osteoarthritis | 18.5 | 18.1 | 0.01 | 17.6 | 17.8 | -0.01 | 18.6 | 18.1 | 0.01 |
| Pneumonia | 2.0 | 2.4 | -0.03 | 2.0 | 2.1 | 0.00 | 1.9 | 2.4 | -0.03 |
| Psoriasis | 2.0 | 1.3 | 0.05 | 1.8 | 1.7 | 0.01 | 1.2 | 1.4 | -0.02 |
| Renal impairment | 4.3 | 6.8 | -0.11 | 4.3 | 4.4 | 0.00 | 6.2 | 6.4 | -0.01 |
| Rheumatoid arthritis | 1.9 | 1.5 | 0.03 | 1.8 | 1.7 | 0.01 | 1.8 | 1.5 | 0.02 |
| Schizophrenia | 0.2 | 0.4 | -0.03 | 0.2 | 0.3 | -0.02 | 0.2 | 0.4 | -0.03 |
| Ulcerative colitis | 0.2 | 0.3 | 0.00 | 0.3 | 0.3 | 0.00 | 0.2 | 0.3 | -0.01 |
| Urinary tract infectious disease | 6.6 | 8.6 | -0.08 | 6.6 | 6.6 | 0.00 | 8.1 | 8.3 | -0.01 |
| Viral hepatitis C | 0.4 | 0.5 | -0.01 | 0.4 | 0.3 | 0.02 | 1.0 | 0.4 | 0.07 |
| Visual system disorder | 12.1 | 15.5 | -0.10 | 12.0 | 12.0 | 0.00 | 13.3 | 15.0 | -0.05 |
| **Medical history: Cardiovascular disease** | | | | | | | | | |
| Atrial fibrillation | 1.8 | 3.2 | -0.09 | 1.8 | 1.8 | 0.01 | 3.1 | 3.0 | 0.00 |
| Cerebrovascular disease | 1.7 | 2.8 | -0.08 | 1.7 | 1.8 | 0.00 | 2.6 | 2.7 | 0.00 |
| Coronary arteriosclerosis | 4.1 | 6.5 | -0.11 | 4.2 | 4.1 | 0.00 | 6.3 | 6.1 | 0.01 |
| Heart disease | 13.5 | 16.5 | -0.08 | 13.5 | 13.2 | 0.01 | 17.1 | 16.0 | 0.03 |
| Heart failure | 2.1 | 3.3 | -0.07 | 2.2 | 2.3 | 0.00 | 3.6 | 3.1 | 0.02 |
| Ischemic heart disease | 2.5 | 3.4 | -0.05 | 2.6 | 2.3 | 0.01 | 3.7 | 3.2 | 0.03 |
| Peripheral vascular disease | 3.5 | 3.8 | -0.01 | 3.4 | 3.7 | -0.01 | 3.4 | 3.7 | -0.01 |
| Pulmonary embolism | 0.6 | 0.5 | 0.01 | 0.5 | 0.4 | 0.01 | 0.6 | 0.5 | 0.02 |
| Venous thrombosis | 0.8 | 1.0 | -0.02 | 0.8 | 0.8 | 0.00 | 0.7 | 1.0 | -0.04 |

| (Continued) | Before ajustment | | | After matching | | | After stratification | | |
|---|---|---|---|---|---|---|---|---|---|
| Characteristic | T (%) | C (%) | StdDiff | T (%) | C (%) | StdDiff | T (%) | C (%) | StdDiff |
| **Medical history: Neoplasms** | | | | | | | | | |
| Hematologic neoplasm | 0.5 | 0.7 | -0.03 | 0.4 | 0.5 | -0.01 | 0.4 | 0.7 | -0.04 |
| Malignant lymphoma | 0.2 | 0.3 | -0.02 | 0.2 | 0.2 | 0.00 | 0.3 | 0.3 | -0.01 |
| Malignant neoplasm of anorectum | 0.1 | 0.1 | -0.01 | 0.1 | 0.1 | 0.00 | 0.1 | 0.1 | -0.02 |
| Malignant neoplastic disease | 5.3 | 6.6 | -0.05 | 5.3 | 5.2 | 0.00 | 5.8 | 6.4 | -0.02 |
| Malignant tumor of breast | 1.9 | 2.5 | -0.04 | 2.0 | 1.9 | 0.01 | 2.1 | 2.4 | -0.02 |
| Malignant tumor of colon | 0.2 | 0.3 | -0.01 | 0.2 | 0.2 | 0.01 | 0.4 | 0.3 | 0.02 |
| Malignant tumor of lung | 0.1 | 0.2 | -0.05 | 0.1 | 0.1 | -0.02 | 0.1 | 0.1 | -0.02 |
| **Medication use** | | | | | | | | | |
| Agents acting on the renin-angiotensin system | 55.9 | 61.7 | -0.12 | 57.7 | 57.3 | 0.01 | 63.2 | 61.0 | 0.04 |
| Antibacterials for systemic use | 50.8 | 47.4 | 0.07 | 49.3 | 49.6 | -0.01 | 48.2 | 47.8 | 0.01 |
| Antidepressants | 47.9 | 37.2 | 0.22 | 45.5 | 46.3 | -0.02 | 41.5 | 38.6 | 0.06 |
| Antiepileptics | 23.6 | 19.7 | 0.10 | 22.5 | 22.9 | -0.01 | 22.5 | 20.3 | 0.05 |
| Antiinflammatory and antirheumatic products | 56.4 | 57.6 | -0.02 | 56.4 | 56.2 | 0.00 | 58.7 | 57.4 | 0.03 |
| Antineoplastic agents | 5.4 | 4.4 | 0.04 | 4.8 | 5.1 | -0.02 | 4.8 | 4.5 | 0.01 |
| Antipsoriatics | 0.7 | 0.5 | 0.03 | 0.6 | 0.5 | 0.01 | 0.4 | 0.5 | -0.01 |
| Antithrombotic agents | 29.2 | 37.8 | -0.18 | 30.3 | 29.9 | 0.01 | 37.1 | 36.6 | 0.01 |
| Beta blocking agents | 26.5 | 32.1 | -0.12 | 26.9 | 26.6 | 0.00 | 31.1 | 31.3 | 0.00 |
| Calcium channel blockers | 17.4 | 20.4 | -0.07 | 22.9 | 19.9 | 0.07 | 22.9 | 19.9 | 0.07 |
| Diuretics | 41.5 | 42.8 | -0.03 | 40.8 | 40.6 | 0.00 | 44.9 | 42.5 | 0.05 |
| Drugs for acid related disorders | 43.0 | 42.7 | 0.01 | 42.1 | 42.3 | 0.00 | 44.0 | 42.7 | 0.03 |
| Drugs for obstructive airway diseases | 51.4 | 45.2 | 0.12 | 49.4 | 50.3 | -0.02 | 46.8 | 46.1 | 0.01 |
| Immunosuppressants | 4.0 | 2.8 | 0.06 | 3.7 | 3.6 | 0.00 | 3.4 | 3.0 | 0.02 |
| Lipid modifying agents | 60.5 | 65.2 | -0.10 | 61.7 | 61.6 | 0.00 | 65.8 | 64.7 | 0.02 |
| Opioids | 28.9 | 30.6 | -0.04 | 28.7 | 28.3 | 0.01 | 31.0 | 30.2 | 0.02 |
| Psycholeptics | 34.8 | 31.3 | 0.07 | 33.4 | 33.6 | 0.00 | 31.8 | 31.7 | 0.00 |
| Psychostimulants | 6.1 | 3.3 | 0.13 | 5.3 | 5.6 | -0.01 | 3.6 | 3.7 | 0.00 |

#### 2.6 DPP4I (female) vs SU (female)

##### 2.6.1 CCAE

**Supplementary Table 191:** Baseline patient characteristics for DPP4I (female) (T) and SU (female) (C) new-users in the CCAE data source. We report proportion of initiators satisfying selected base-line characteristics and the standardized difference of population proportions (StdDiff) before and after propensity score adjustment. Less extreme StdDiffs through matching and stratification suggest improved balance between patient cohorts through adjustment.

| | Before adjustment | | | After matching | | | After stratification | | |
|---|---|---|---|---|---|---|---|---|---|
| Characteristic | T (%) | C (%) | StdDiff | T (%) | C (%) | StdDiff | T (%) | C (%) | StdDiff |
| **Age group** | | | | | | | | | |
| 20 - 24 | 0.4 | 0.4 | -0.01 | 0.4 | 0.4 | 0.00 | 0.4 | 0.4 | 0.00 |
| 25 - 29 | 0.8 | 1.0 | -0.03 | 0.9 | 0.8 | 0.01 | 0.9 | 1.0 | -0.01 |
| 30 - 34 | 2.0 | 2.7 | -0.04 | 2.1 | 2.0 | 0.00 | 2.1 | 2.4 | -0.02 |
| 35 - 39 | 4.6 | 5.1 | -0.03 | 4.7 | 4.6 | 0.01 | 4.8 | 5.0 | -0.01 |
| 40 - 44 | 8.3 | 8.4 | 0.00 | 8.4 | 8.3 | 0.00 | 8.3 | 8.4 | 0.00 |
| 45 - 49 | 13.7 | 13.6 | 0.00 | 13.8 | 13.7 | 0.00 | 13.7 | 13.5 | 0.01 |
| 50 - 54 | 20.1 | 20.0 | 0.00 | 19.9 | 20.0 | 0.00 | 20.0 | 19.8 | 0.01 |
| 55 - 59 | 24.1 | 23.5 | 0.01 | 23.8 | 24.0 | 0.00 | 23.8 | 24.0 | 0.00 |
| 60 - 64 | 23.9 | 23.1 | 0.02 | 23.8 | 23.9 | 0.00 | 23.8 | 23.7 | 0.00 |
| 65 - 69 | 2.1 | 2.1 | 0.00 | 2.1 | 2.2 | -0.01 | 2.1 | 2.2 | 0.00 |
| **Medical history: General** | | | | | | | | | |
| Acute respiratory disease | 33.5 | 30.8 | 0.06 | 32.4 | 32.6 | 0.00 | 32.4 | 32.2 | 0.00 |
| Attention deficit hyperactivity disorder | 0.8 | 0.5 | 0.03 | 0.7 | 0.7 | 0.00 | 0.7 | 0.7 | 0.00 |
| Chronic liver disease | 3.9 | 3.6 | 0.02 | 3.8 | 3.8 | 0.00 | 3.9 | 3.8 | 0.00 |
| Chronic obstructive lung disease | 2.8 | 3.1 | -0.02 | 2.8 | 2.8 | 0.00 | 3.2 | 3.1 | 0.01 |
| Crohn’s disease | 0.3 | 0.3 | 0.00 | 0.3 | 0.3 | 0.00 | 0.3 | 0.3 | 0.00 |
| Dementia | 0.2 | 0.2 | 0.00 | 0.2 | 0.2 | 0.00 | 0.2 | 0.2 | 0.01 |
| Depressive disorder | 12.7 | 11.9 | 0.03 | 12.6 | 12.8 | -0.01 | 13.0 | 12.8 | 0.00 |
| Gastroesophageal reflux disease | 13.1 | 10.7 | 0.07 | 12.4 | 12.6 | 0.00 | 12.4 | 12.6 | 0.00 |
| Gastrointestinal hemorrhage | 1.8 | 1.8 | 0.00 | 1.8 | 1.9 | -0.01 | 1.8 | 1.8 | 0.00 |
| Human immunodeficiency virus infection | 0.1 | 0.1 | 0.01 | 0.1 | 0.1 | 0.00 | 0.1 | 0.1 | 0.00 |
| Hyperlipidemia | 64.4 | 56.3 | 0.17 | 62.9 | 63.5 | -0.01 | 61.6 | 61.5 | 0.00 |
| Hypertensive disorder | 65.0 | 60.7 | 0.09 | 64.4 | 64.7 | -0.01 | 64.5 | 63.9 | 0.01 |
| Lesion of liver | 0.6 | 0.7 | -0.01 | 0.6 | 0.6 | 0.00 | 0.7 | 0.7 | 0.00 |
| Obesity | 19.4 | 17.5 | 0.05 | 19.4 | 19.4 | 0.00 | 19.4 | 19.6 | -0.01 |
| Osteoarthritis | 19.4 | 17.0 | 0.06 | 18.6 | 18.9 | -0.01 | 18.8 | 18.4 | 0.01 |
| Pneumonia | 2.1 | 2.6 | -0.03 | 2.1 | 2.2 | 0.00 | 2.5 | 2.4 | 0.01 |
| Psoriasis | 1.5 | 1.4 | 0.01 | 1.5 | 1.5 | 0.00 | 1.4 | 1.5 | 0.00 |
| Renal impairment | 3.1 | 3.1 | 0.00 | 3.1 | 3.1 | 0.00 | 3.5 | 3.3 | 0.01 |
| Rheumatoid arthritis | 1.6 | 1.4 | 0.01 | 1.5 | 1.5 | 0.00 | 1.6 | 1.5 | 0.00 |
| Schizophrenia | 0.1 | 0.1 | 0.00 | 0.1 | 0.1 | 0.01 | 0.1 | 0.1 | 0.01 |
| Ulcerative colitis | 0.3 | 0.3 | 0.01 | 0.3 | 0.3 | 0.00 | 0.3 | 0.3 | 0.00 |
| Urinary tract infectious disease | 11.8 | 11.3 | 0.01 | 11.3 | 11.4 | 0.00 | 11.7 | 11.5 | 0.01 |
| Viral hepatitis C | 0.3 | 0.3 | -0.01 | 0.3 | 0.3 | 0.00 | 0.3 | 0.3 | 0.00 |
| Visual system disorder | 27.0 | 24.1 | 0.06 | 25.9 | 26.1 | 0.00 | 25.3 | 25.5 | 0.00 |
| **Medical history: Cardiovascular disease** | | | | | | | | | |
| Atrial fibrillation | 1.3 | 1.2 | 0.00 | 1.3 | 1.3 | 0.00 | 1.4 | 1.3 | 0.01 |
| Cerebrovascular disease | 2.5 | 2.4 | 0.01 | 2.3 | 2.4 | -0.01 | 2.5 | 2.5 | 0.00 |
| Coronary arteriosclerosis | 4.3 | 4.5 | -0.01 | 4.2 | 4.3 | -0.01 | 4.6 | 4.4 | 0.01 |
| Heart disease | 14.3 | 13.5 | 0.03 | 14.3 | 13.6 | 0.02 | 14.3 | 13.6 | 0.02 |
| Heart failure | 1.7 | 2.0 | -0.02 | 1.7 | 1.8 | 0.00 | 2.0 | 1.9 | 0.01 |
| Ischemic heart disease | 2.8 | 3.2 | -0.02 | 2.7 | 2.8 | 0.00 | 3.1 | 2.9 | 0.01 |
| Peripheral vascular disease | 3.0 | 2.4 | 0.03 | 2.8 | 2.8 | 0.00 | 2.8 | 2.7 | 0.01 |
| Pulmonary embolism | 0.4 | 0.3 | 0.01 | 0.4 | 0.4 | 0.01 | 0.4 | 0.4 | 0.01 |
| Venous thrombosis | 1.0 | 0.8 | 0.01 | 0.9 | 0.9 | 0.00 | 0.9 | 0.9 | 0.00 |
| **Medical history: Neoplasms** | | | | | | | | | |
| Hematologic neoplasm | 0.8 | 0.8 | 0.00 | 0.8 | 0.8 | 0.00 | 0.9 | 0.8 | 0.01 |
| Malignant lymphoma | 0.3 | 0.3 | 0.00 | 0.3 | 0.3 | 0.00 | 0.3 | 0.3 | 0.00 |
| Malignant neoplasm of anorectum | 0.1 | 0.2 | -0.01 | 0.1 | 0.1 | 0.00 | 0.1 | 0.1 | 0.00 |
| Malignant neoplastic disease | 6.6 | 6.1 | 0.02 | 6.3 | 6.5 | 0.00 | 6.5 | 6.4 | 0.00 |
| Malignant tumor of breast | 2.7 | 2.5 | 0.01 | 2.6 | 2.7 | 0.00 | 2.6 | 2.6 | 0.00 |
| Malignant tumor of colon | 0.2 | 0.3 | -0.01 | 0.2 | 0.3 | -0.01 | 0.3 | 0.3 | 0.00 |
| Malignant tumor of lung | 0.2 | 0.2 | 0.00 | 0.2 | 0.1 | 0.01 | 0.2 | 0.2 | 0.01 |
| **Medication use** | | | | | | | | | |
| Agents acting on the renin-angiotensin system | 63.5 | 61.9 | 0.03 | 63.1 | 63.0 | 0.00 | 63.4 | 63.0 | 0.01 |
| Antibacterials for systemic use | 66.8 | 64.7 | 0.04 | 65.7 | 66.0 | -0.01 | 66.0 | 65.7 | 0.01 |

| (Continued) | Before ajustment | | | After matching | | | After stratification | | |
|---|---|---|---|---|---|---|---|---|---|
| Characteristic | T (%) | C (%) | StdDiff | T (%) | C (%) | StdDiff | T (%) | C (%) | StdDiff |
| Antidepressants | 34.6 | 33.8 | 0.02 | 34.0 | 34.2 | 0.00 | 35.0 | 34.2 | 0.01 |
| Antiepileptics | 14.0 | 13.4 | 0.02 | 13.9 | 13.7 | 0.00 | 14.5 | 13.9 | 0.02 |
| Antiinflammatory and antirheumatic products | 35.0 | 34.1 | 0.02 | 34.4 | 34.6 | 0.00 | 34.9 | 34.7 | 0.00 |
| Antineoplastic agents | 4.8 | 4.7 | 0.00 | 4.5 | 4.5 | 0.00 | 4.5 | 4.4 | 0.00 |
| Antithrombotic agents | 7.4 | 7.4 | 0.00 | 7.4 | 7.4 | 0.00 | 7.9 | 7.6 | 0.01 |
| Beta blocking agents | 24.7 | 25.2 | -0.01 | 24.6 | 24.6 | 0.00 | 25.8 | 25.2 | 0.01 |
| Calcium channel blockers | 18.6 | 18.1 | 0.01 | 18.5 | 18.6 | 0.00 | 18.6 | 18.4 | 0.00 |
| Diuretics | 43.4 | 43.3 | 0.00 | 43.0 | 43.0 | 0.00 | 43.9 | 43.1 | 0.02 |
| Drugs for acid related disorders | 27.8 | 26.1 | 0.04 | 26.8 | 26.8 | 0.00 | 27.0 | 26.8 | 0.00 |
| Drugs for obstructive airway diseases | 42.7 | 39.3 | 0.07 | 41.5 | 41.6 | 0.00 | 41.8 | 41.4 | 0.01 |
| Immunosuppressants | 3.0 | 2.6 | 0.03 | 2.8 | 2.9 | 0.00 | 2.9 | 2.8 | 0.00 |
| Lipid modifying agents | 63.4 | 60.0 | 0.07 | 62.3 | 62.5 | 0.00 | 62.4 | 62.3 | 0.00 |
| Opioids | 28.7 | 29.3 | -0.01 | 28.6 | 28.5 | 0.00 | 30.1 | 29.3 | 0.02 |
| Psycholeptics | 26.6 | 25.7 | 0.02 | 26.2 | 26.4 | 0.00 | 27.2 | 26.4 | 0.02 |
| Psychostimulants | 3.7 | 3.1 | 0.03 | 3.5 | 3.4 | 0.00 | 3.6 | 3.4 | 0.01 |

##### 2.6.2 DAG

**Supplementary Table 192:** Baseline patient characteristics for DPP4I (female) (T) and SU (female) (C) new-users in the DAG data source. We report proportion of initiators satisfying selected base-line characteristics and the standardized difference of population proportions (StdDiff) before and after propensity score adjustment. Less extreme StdDiffs through matching and stratification suggest improved balance between patient cohorts through adjustment.

| | Before adjustment | | | After matching | | | After stratification | | |
|---|---|---|---|---|---|---|---|---|---|
| Characteristic | T (%) | C (%) | StdDiff | T (%) | C (%) | StdDiff | T (%) | C (%) | StdDiff |
| **Age group** | | | | | | | | | |
| 20 - 24 | 0.1 | <0.2 | 0.00 | <0.2 | <0.2 | -0.01 | 0.1 | <0.1 | 0.01 |
| 25 - 29 | 0.2 | 0.2 | 0.01 | <0.2 | 0.2 | -0.01 | 0.2 | <0.1 | 0.01 |
| 30 - 34 | 0.6 | 0.4 | 0.02 | 0.5 | 0.4 | 0.01 | 0.6 | 0.3 | 0.05 |
| 35 - 39 | 1.1 | 1.0 | 0.01 | 1.1 | 1.2 | 0.00 | 1.2 | 1.1 | 0.01 |
| 40 - 44 | 2.5 | 2.2 | 0.02 | 2.9 | 2.4 | 0.03 | 2.5 | 2.2 | 0.02 |
| 45 - 49 | 4.7 | 4.1 | 0.03 | 4.4 | 3.7 | 0.04 | 4.6 | 4.1 | 0.02 |
| 50 - 54 | 7.9 | 6.8 | 0.04 | 7.5 | 7.0 | 0.02 | 7.8 | 8.0 | -0.01 |
| 55 - 59 | 11.2 | 9.5 | 0.06 | 9.1 | 9.7 | -0.02 | 10.9 | 10.6 | 0.01 |
| 60 - 64 | 13.4 | 12.6 | 0.03 | 12.5 | 12.1 | 0.01 | 13.2 | 12.8 | 0.01 |
| 65 - 69 | 14.2 | 16.3 | -0.06 | 14.7 | 13.9 | 0.02 | 14.2 | 14.4 | -0.01 |
| 70 - 74 | 14.0 | 17.0 | -0.08 | 16.5 | 16.1 | 0.01 | 14.3 | 15.2 | -0.02 |
| 75 - 79 | 13.6 | 15.6 | -0.06 | 15.1 | 16.1 | -0.03 | 14.1 | 12.9 | 0.04 |
| 80 - 84 | 10.3 | 9.7 | 0.02 | 10.1 | 10.8 | -0.02 | 10.3 | 11.5 | -0.04 |
| 85 - 89 | 4.7 | 3.8 | 0.05 | 4.4 | 5.2 | -0.04 | 4.6 | 5.4 | -0.03 |
| 90 - 94 | 1.4 | 0.8 | 0.06 | 0.7 | 1.2 | -0.05 | 1.3 | 1.4 | -0.01 |
| **Medical history: General** | | | | | | | | | |
| Acute respiratory disease | 18.5 | 19.5 | -0.03 | 20.5 | 19.6 | 0.02 | 18.9 | 17.8 | 0.03 |
| Attention deficit hyperactivity disorder | 0.2 | 0.4 | -0.04 | 0.3 | 0.6 | -0.04 | 0.2 | 0.6 | -0.07 |
| Chronic liver disease | 0.4 | 0.2 | 0.04 | 0.3 | <0.2 | 0.04 | 0.4 | 0.3 | 0.03 |
| Chronic obstructive lung disease | 5.9 | 6.6 | -0.03 | 6.5 | 6.6 | 0.00 | 5.9 | 5.8 | 0.00 |
| Crohn's disease | 0.1 | <0.2 | 0.00 | <0.2 | <0.2 | -0.03 | 0.1 | <0.1 | -0.01 |
| Dementia | 2.3 | 2.7 | -0.03 | 2.3 | 3.0 | -0.04 | 2.3 | 2.0 | 0.02 |
| Depressive disorder | 11.8 | 12.6 | -0.03 | 12.6 | 13.1 | -0.01 | 12.0 | 11.7 | 0.01 |
| Gastroesophageal reflux disease | 3.0 | 2.9 | 0.01 | 3.4 | 2.9 | 0.03 | 3.1 | 2.4 | 0.04 |
| Gastrointestinal hemorrhage | 0.7 | 0.8 | -0.01 | 0.4 | 0.8 | -0.04 | 0.7 | 0.5 | 0.02 |
| Hyperlipidemia | 22.6 | 24.6 | -0.05 | 24.8 | 24.2 | 0.01 | 23.0 | 21.9 | 0.03 |
| Hypertensive disorder | 48.4 | 59.5 | -0.22 | 57.5 | 56.8 | 0.02 | 50.3 | 48.0 | 0.05 |
| Lesion of liver | 0.5 | 0.5 | 0.01 | 0.5 | 0.5 | 0.00 | 0.5 | 0.6 | -0.01 |
| Obesity | 9.5 | 9.1 | 0.02 | 8.8 | 8.5 | 0.01 | 9.4 | 8.8 | 0.02 |
| Osteoarthritis | 14.1 | 16.6 | -0.07 | 15.9 | 15.4 | 0.01 | 14.4 | 14.3 | 0.00 |
| Psoriasis | 1.3 | 1.2 | 0.01 | 0.9 | 1.0 | -0.02 | 1.2 | 1.1 | 0.01 |
| Renal impairment | 7.3 | 4.7 | 0.11 | 5.6 | 5.5 | 0.01 | 5.6 | 5.5 | 0.01 |
| Rheumatoid arthritis | 1.7 | 1.8 | -0.01 | 1.5 | 1.8 | -0.02 | 1.6 | 1.7 | 0.00 |
| Schizophrenia | 0.2 | 0.3 | -0.03 | <0.2 | 0.2 | -0.02 | <0.2 | 0.2 | -0.02 |
| Ulcerative colitis | 0.3 | 0.2 | 0.03 | 0.3 | <0.2 | 0.05 | 0.3 | <0.1 | 0.06 |
| Urinary tract infectious disease | 8.6 | 9.5 | -0.03 | 9.7 | 9.8 | 0.00 | 8.8 | 8.8 | 0.00 |
| Viral hepatitis C | 0.1 | <0.2 | 0.02 | <0.2 | <0.2 | 0.02 | 0.1 | <0.1 | 0.01 |
| Visual system disorder | 9.5 | 10.6 | -0.04 | 11.0 | 10.3 | 0.03 | 9.8 | 8.9 | 0.03 |
| **Medical history: Cardiovascular disease** | | | | | | | | | |
| Atrial fibrillation | 3.0 | 2.3 | 0.04 | 2.4 | 2.6 | -0.01 | 2.9 | 2.5 | 0.02 |
| Cerebrovascular disease | 3.8 | 4.3 | -0.03 | 4.2 | 3.8 | 0.02 | 4.0 | 3.8 | 0.01 |
| Coronary arteriosclerosis | 2.9 | 2.8 | 0.01 | 2.4 | 3.1 | -0.05 | 2.8 | 3.6 | -0.05 |
| Heart disease | 23.7 | 27.7 | -0.09 | 27.3 | 25.3 | 0.04 | 24.3 | 23.3 | 0.02 |
| Heart failure | 7.3 | 9.6 | -0.08 | 8.2 | 8.2 | 0.00 | 7.5 | 8.0 | -0.02 |
| Ischemic heart disease | 8.7 | 12.9 | -0.14 | 11.1 | 10.5 | 0.02 | 9.2 | 9.2 | 0.00 |
| Peripheral vascular disease | 4.8 | 4.9 | -0.01 | 5.1 | 5.5 | -0.02 | 4.8 | 5.3 | -0.02 |
| Pulmonary embolism | 0.7 | 0.6 | 0.00 | 0.5 | 0.7 | -0.02 | 0.7 | 0.8 | -0.01 |
| Venous thrombosis | 1.8 | 1.8 | 0.00 | 1.9 | 2.0 | -0.01 | 1.9 | 2.2 | -0.02 |
| **Medical history: Neoplasms** | | | | | | | | | |
| Hematologic neoplasm | 0.4 | 0.3 | 0.01 | 0.5 | 0.4 | 0.02 | 0.4 | 0.3 | 0.01 |
| Malignant lymphoma | 0.3 | 0.2 | 0.02 | 0.4 | 0.2 | 0.04 | 0.3 | <0.1 | 0.04 |
| Malignant neoplasm of anorectum | 0.2 | 0.2 | 0.00 | 0.2 | <0.2 | 0.03 | 0.2 | <0.1 | 0.01 |
| Malignant neoplastic disease | 5.0 | 5.6 | -0.03 | 5.0 | 5.6 | -0.02 | 5.0 | 4.6 | 0.02 |
| Malignant tumor of breast | 1.2 | 2.1 | -0.07 | 1.3 | 2.0 | -0.05 | 1.2 | 1.5 | -0.02 |
| Malignant tumor of colon | 0.4 | 0.5 | -0.01 | 0.3 | 0.3 | -0.01 | 0.4 | 0.3 | 0.01 |
| Malignant tumor of lung | 0.1 | 0.2 | -0.02 | <0.2 | 0.2 | -0.03 | 0.1 | 0.2 | -0.01 |
| Malignant tumor of urinary bladder | 0.1 | <0.2 | 0.02 | <0.2 | <0.2 | 0.02 | 0.1 | <0.1 | 0.03 |
| **Medication use** | | | | | | | | | |

| (Continued) | Before ajustment | | | After matching | | | After stratification | | |
|---|---|---|---|---|---|---|---|---|---|
| Characteristic | T (%) | C (%) | StdDiff | T (%) | C (%) | StdDiff | T (%) | C (%) | StdDiff |
| Agents acting on the renin-angiotensin system | 67.1 | 65.3 | 0.04 | 68.4 | 67.1 | 0.03 | 67.2 | 65.6 | 0.03 |
| Antibacterials for systemic use | 30.2 | 33.8 | -0.08 | 34.9 | 33.6 | 0.03 | 31.2 | 31.3 | 0.00 |
| Antidepressants | 14.2 | 13.1 | 0.03 | 13.2 | 13.4 | -0.01 | 14.1 | 11.9 | 0.07 |
| Antiepileptics | 5.5 | 3.7 | 0.09 | 4.3 | 4.5 | -0.01 | 5.3 | 4.6 | 0.03 |
| Antiinflammatory and antirheumatic products | 41.7 | 47.6 | -0.12 | 46.2 | 45.7 | 0.01 | 42.7 | 41.1 | 0.03 |
| Antineoplastic agents | 1.7 | 1.3 | 0.04 | 1.3 | 1.4 | -0.01 | 1.6 | 1.3 | 0.02 |
| Antipsoriatics | 0.4 | 0.2 | 0.04 | 0.2 | <0.2 | 0.02 | 0.4 | 0.2 | 0.04 |
| Antithrombotic agents | 27.7 | 27.0 | 0.02 | 26.5 | 26.4 | 0.00 | 27.5 | 26.9 | 0.01 |
| Beta blocking agents | 50.2 | 49.8 | 0.01 | 53.0 | 51.1 | 0.04 | 50.6 | 50.6 | 0.00 |
| Calcium channel blockers | 33.6 | 32.1 | 0.03 | 33.2 | 33.4 | 0.00 | 33.5 | 32.4 | 0.02 |
| Diuretics | 52.9 | 57.6 | -0.10 | 58.1 | 57.4 | 0.01 | 54.1 | 52.8 | 0.03 |
| Drugs for acid related disorders | 38.8 | 36.6 | 0.05 | 40.0 | 39.5 | 0.01 | 39.0 | 38.7 | 0.01 |
| Drugs for obstructive airway diseases | 18.6 | 17.9 | 0.02 | 18.6 | 18.7 | 0.00 | 18.6 | 17.9 | 0.02 |
| Immunosuppressants | 0.8 | 0.9 | 0.00 | 0.7 | 1.0 | -0.03 | 0.8 | 0.8 | 0.00 |
| Lipid modifying agents | 42.0 | 37.0 | 0.10 | 39.3 | 39.0 | 0.01 | 41.3 | 40.3 | 0.02 |
| Opioids | 14.2 | 14.7 | -0.01 | 15.2 | 14.5 | 0.02 | 14.3 | 13.8 | 0.01 |
| Psycholeptics | 11.7 | 14.4 | -0.08 | 14.0 | 13.6 | 0.01 | 12.1 | 11.6 | 0.01 |
| Psychostimulants | 0.3 | 0.4 | -0.02 | 0.3 | <0.2 | 0.05 | 0.3 | <0.1 | 0.05 |

##### 2.6.3 ODOD

**Supplementary Table 193:** Baseline patient characteristics for DPP4I (female) (T) and SU (female) (C) new-users in the ODOD data source. We report proportion of initiators satisfying selected base-line characteristics and the standardized difference of population proportions (StdDiff) before and after propensity score adjustment. Less extreme StdDiffs through matching and stratification suggest improved balance between patient cohorts through adjustment.

| | Before adjustment | | | After matching | | | After stratification | | |
|---|---|---|---|---|---|---|---|---|---|
| Characteristic | T (%) | C (%) | StdDiff | T (%) | C (%) | StdDiff | T (%) | C (%) | StdDiff |
| **Age group** | | | | | | | | | |
| 20 - 24 | 0.2 | 0.2 | 0.00 | 0.2 | 0.1 | 0.01 | 0.2 | 0.2 | 0.01 |
| 25 - 29 | 0.4 | 0.5 | -0.01 | 0.4 | 0.4 | 0.01 | 0.4 | 0.5 | 0.00 |
| 30 - 34 | 1.3 | 1.4 | -0.01 | 1.3 | 1.1 | 0.02 | 1.3 | 1.3 | 0.00 |
| 35 - 39 | 2.5 | 2.5 | 0.00 | 2.5 | 2.3 | 0.01 | 2.4 | 2.4 | 0.00 |
| 40 - 44 | 4.4 | 3.9 | 0.02 | 4.3 | 4.2 | 0.00 | 4.3 | 4.2 | 0.00 |
| 45 - 49 | 7.3 | 6.1 | 0.05 | 7.1 | 7.2 | 0.00 | 6.4 | 6.5 | 0.00 |
| 50 - 54 | 10.1 | 8.8 | 0.04 | 9.9 | 10.0 | 0.00 | 9.2 | 9.2 | 0.00 |
| 55 - 59 | 12.0 | 10.8 | 0.04 | 11.7 | 11.8 | 0.00 | 11.1 | 11.2 | 0.00 |
| 60 - 64 | 12.5 | 11.2 | 0.04 | 11.6 | 11.6 | 0.00 | 11.6 | 11.6 | 0.00 |
| 65 - 69 | 15.5 | 17.5 | -0.05 | 15.7 | 16.7 | -0.03 | 16.5 | 17.1 | -0.02 |
| 70 - 74 | 15.1 | 16.9 | -0.05 | 15.5 | 15.4 | 0.00 | 16.6 | 16.3 | 0.01 |
| 75 - 79 | 9.5 | 10.9 | -0.04 | 9.8 | 9.7 | 0.00 | 10.6 | 10.4 | 0.00 |
| 80 - 84 | 5.8 | 6.2 | -0.02 | 5.9 | 5.7 | 0.01 | 6.2 | 6.0 | 0.01 |
| 85 - 89 | 3.1 | 2.9 | 0.01 | 3.1 | 3.0 | 0.01 | 3.1 | 3.0 | 0.01 |
| 90 - 94 | 0.2 | 0.2 | 0.01 | 0.3 | 0.2 | 0.02 | 0.3 | 0.2 | 0.02 |
| **Medical history: General** | | | | | | | | | |
| Acute respiratory disease | 31.5 | 29.5 | 0.04 | 30.8 | 31.1 | 0.00 | 30.0 | 30.2 | 0.00 |
| Attention deficit hyperactivity disorder | 0.6 | 0.5 | 0.01 | 0.6 | 0.6 | 0.00 | 0.5 | 0.5 | 0.00 |
| Chronic liver disease | 3.7 | 3.5 | 0.01 | 3.5 | 3.7 | -0.01 | 3.4 | 3.6 | -0.01 |
| Chronic obstructive lung disease | 9.2 | 10.2 | -0.03 | 10.0 | 10.0 | 0.00 | 10.0 | 10.0 | 0.00 |
| Crohn’s disease | 0.3 | 0.3 | 0.01 | 0.3 | 0.3 | 0.01 | 0.4 | 0.3 | 0.01 |
| Dementia | 2.8 | 2.8 | 0.00 | 2.8 | 2.7 | 0.00 | 2.9 | 2.8 | 0.01 |
| Depressive disorder | 18.9 | 19.1 | 0.00 | 18.8 | 19.2 | -0.01 | 19.1 | 19.2 | 0.00 |
| Gastroesophageal reflux disease | 21.6 | 20.6 | 0.03 | 21.2 | 21.7 | -0.01 | 21.0 | 21.1 | 0.00 |
| Gastrointestinal hemorrhage | 2.7 | 2.8 | -0.01 | 2.7 | 2.9 | -0.01 | 2.7 | 2.9 | -0.01 |
| Human immunodeficiency virus infection | 0.1 | 0.1 | 0.00 | 0.2 | 0.2 | 0.00 | 0.1 | 0.1 | 0.00 |
| Hyperlipidemia | 80.0 | 77.0 | 0.07 | 79.2 | 79.9 | -0.02 | 77.9 | 78.1 | 0.00 |
| Lesion of liver | 1.0 | 1.3 | -0.02 | 1.0 | 1.2 | -0.01 | 1.1 | 1.2 | -0.01 |
| Osteoarthritis | 31.7 | 30.4 | 0.03 | 31.2 | 31.8 | -0.01 | 30.9 | 31.0 | 0.00 |
| Pneumonia | 4.0 | 4.6 | -0.03 | 4.1 | 4.1 | 0.00 | 4.5 | 4.4 | 0.00 |
| Psoriasis | 1.6 | 1.5 | 0.01 | 1.6 | 1.6 | 0.00 | 1.6 | 1.6 | 0.00 |
| Renal impairment | 14.8 | 15.9 | -0.03 | 15.0 | 14.9 | 0.00 | 15.9 | 15.6 | 0.01 |
| Rheumatoid arthritis | 2.6 | 2.6 | 0.00 | 2.6 | 2.7 | -0.01 | 2.6 | 2.6 | 0.00 |
| Schizophrenia | 0.4 | 0.5 | -0.01 | 0.5 | 0.5 | -0.01 | 0.5 | 0.5 | 0.00 |
| Ulcerative colitis | 0.4 | 0.4 | 0.01 | 0.4 | 0.4 | 0.01 | 0.4 | 0.4 | 0.01 |
| Urinary tract infectious disease | 17.0 | 16.6 | 0.01 | 16.9 | 17.1 | -0.01 | 16.8 | 16.9 | 0.00 |
| Viral hepatitis C | 0.5 | 0.5 | 0.00 | 0.5 | 0.5 | 0.00 | 0.5 | 0.5 | 0.00 |
| Visual system disorder | 43.3 | 42.9 | 0.01 | 43.2 | 43.5 | -0.01 | 43.2 | 43.2 | 0.00 |
| **Medical history: Cardiovascular disease** | | | | | | | | | |
| Atrial fibrillation | 5.4 | 5.8 | -0.02 | 5.5 | 5.4 | 0.01 | 5.9 | 5.7 | 0.01 |
| Cerebrovascular disease | 6.9 | 6.8 | 0.00 | 6.7 | 6.9 | -0.01 | 6.7 | 6.9 | -0.01 |
| Coronary arteriosclerosis | 11.4 | 11.7 | -0.01 | 11.3 | 11.4 | 0.00 | 11.8 | 11.7 | 0.01 |
| Heart disease | 27.8 | 28.0 | 0.00 | 27.6 | 27.7 | 0.00 | 28.2 | 28.0 | 0.00 |
| Heart failure | 7.0 | 7.5 | -0.02 | 7.2 | 6.9 | 0.01 | 7.6 | 7.4 | 0.01 |
| Ischemic heart disease | 6.4 | 7.0 | -0.02 | 6.5 | 6.6 | 0.00 | 6.9 | 6.9 | 0.00 |
| Pulmonary embolism | 0.7 | 0.9 | -0.01 | 0.7 | 0.8 | -0.01 | 0.8 | 0.9 | 0.00 |
| Venous thrombosis | 1.7 | 1.8 | -0.01 | 1.7 | 1.7 | 0.00 | 1.7 | 1.8 | 0.00 |
| **Medical history: Neoplasms** | | | | | | | | | |
| Hematologic neoplasm | 1.2 | 1.4 | -0.01 | 1.2 | 1.2 | 0.00 | 1.3 | 1.3 | 0.00 |
| Malignant lymphoma | 0.5 | 0.5 | -0.01 | 0.5 | 0.5 | 0.00 | 0.5 | 0.5 | 0.00 |
| Malignant neoplasm of anorectum | 0.2 | 0.2 | -0.01 | 0.1 | 0.2 | -0.01 | 0.2 | 0.2 | -0.01 |
| Malignant neoplastic disease | 9.8 | 10.1 | -0.01 | 9.7 | 9.9 | 0.00 | 9.7 | 9.9 | 0.00 |
| Malignant tumor of breast | 3.7 | 3.8 | 0.00 | 3.8 | 3.7 | 0.00 | 3.9 | 3.8 | 0.01 |
| Malignant tumor of colon | 0.5 | 0.5 | 0.00 | 0.5 | 0.5 | 0.00 | 0.5 | 0.5 | 0.00 |
| Malignant tumor of lung | 0.3 | 0.5 | -0.03 | 0.4 | 0.4 | 0.00 | 0.4 | 0.5 | 0.00 |
| Malignant tumor of urinary bladder | 0.2 | 0.2 | 0.00 | 0.2 | 0.2 | 0.00 | 0.3 | 0.2 | 0.01 |
| **Medication use** | | | | | | | | | |
| Agents acting on the renin-angiotensin system | 68.8 | 69.1 | 0.00 | 68.8 | 68.6 | 0.00 | 69.6 | 68.9 | 0.01 |

| (Continued) | Before ajustment | | | After matching | | | After stratification | | |
|---|---|---|---|---|---|---|---|---|---|
| Characteristic | T (%) | C (%) | StdDiff | T (%) | C (%) | StdDiff | T (%) | C (%) | StdDiff |
| Antibacterials for systemic use | 65.0 | 63.1 | 0.04 | 64.3 | 64.6 | -0.01 | 63.5 | 63.8 | -0.01 |
| Antidepressants | 35.9 | 34.8 | 0.02 | 35.4 | 35.7 | -0.01 | 35.2 | 35.2 | 0.00 |
| Antiepileptics | 19.8 | 19.7 | 0.00 | 19.7 | 20.0 | -0.01 | 19.9 | 19.8 | 0.00 |
| Antiinflammatory and antirheumatic products | 34.5 | 32.5 | 0.04 | 33.9 | 34.3 | -0.01 | 33.0 | 33.3 | -0.01 |
| Antineoplastic agents | 6.4 | 5.6 | 0.03 | 6.1 | 6.2 | 0.00 | 5.9 | 5.9 | 0.00 |
| Antipsoriatics | 0.7 | 0.6 | 0.02 | 0.7 | 0.7 | 0.00 | 0.6 | 0.6 | 0.00 |
| Antithrombotic agents | 13.6 | 14.5 | -0.03 | 13.7 | 13.8 | 0.00 | 14.5 | 14.3 | 0.01 |
| Beta blocking agents | 34.3 | 35.9 | -0.03 | 34.5 | 34.3 | 0.00 | 35.9 | 35.4 | 0.01 |
| Calcium channel blockers | 27.0 | 27.7 | -0.02 | 27.9 | 27.6 | 0.01 | 27.9 | 27.6 | 0.01 |
| Diuretics | 48.0 | 48.7 | -0.01 | 47.8 | 47.7 | 0.00 | 48.9 | 48.4 | 0.01 |
| Drugs for acid related disorders | 32.3 | 31.8 | 0.01 | 31.9 | 32.2 | 0.00 | 32.4 | 32.1 | 0.01 |
| Drugs for obstructive airway diseases | 44.2 | 40.9 | 0.07 | 43.3 | 44.0 | -0.01 | 41.8 | 42.2 | -0.01 |
| Immunosuppressants | 3.7 | 3.2 | 0.03 | 3.5 | 3.6 | 0.00 | 3.4 | 3.4 | 0.00 |
| Lipid modifying agents | 72.5 | 70.5 | 0.04 | 72.0 | 72.3 | -0.01 | 71.4 | 71.2 | 0.01 |
| Opioids | 30.6 | 30.3 | 0.01 | 30.4 | 30.4 | 0.00 | 30.6 | 30.4 | 0.00 |
| Psycholeptics | 28.7 | 27.8 | 0.02 | 28.3 | 28.5 | 0.00 | 28.3 | 28.2 | 0.00 |
| Psychostimulants | 2.4 | 1.9 | 0.03 | 2.2 | 2.3 | -0.01 | 2.1 | 2.1 | 0.00 |

##### 2.6.4 MDCR

**Supplementary Table 194:** Baseline patient characteristics for DPP4I (female) (T) and SU (female) (C) new-users in the MDCR data source. We report proportion of initiators satisfying selected base-line characteristics and the standardized difference of population proportions (StdDiff) before and after propensity score adjustment. Less extreme StdDiffs through matching and stratification suggest improved balance between patient cohorts through adjustment.

| | Before adjustment | | | After matching | | | After stratification | | |
|---|---|---|---|---|---|---|---|---|---|
| Characteristic | T (%) | C (%) | StdDiff | T (%) | C (%) | StdDiff | T (%) | C (%) | StdDiff |
| **Age group** | | | | | | | | | |
| 50 - 54 | 0.1 | 0.1 | 0.00 | 0.1 | 0.2 | -0.01 | 0.2 | 0.1 | 0.00 |
| 55 - 59 | 0.5 | 0.6 | -0.01 | 0.5 | 0.6 | -0.01 | 0.5 | 0.6 | -0.02 |
| 60 - 64 | 1.2 | 1.1 | 0.01 | 1.2 | 1.3 | -0.01 | 1.2 | 1.3 | 0.00 |
| 65 - 69 | 29.3 | 26.5 | 0.06 | 29.3 | 28.8 | 0.01 | 28.9 | 28.5 | 0.01 |
| 70 - 74 | 29.6 | 28.2 | 0.03 | 29.1 | 29.2 | 0.00 | 28.5 | 28.5 | 0.00 |
| 75 - 79 | 18.5 | 21.2 | -0.07 | 18.7 | 18.8 | 0.00 | 19.3 | 19.4 | 0.00 |
| 80 - 84 | 12.3 | 13.3 | -0.03 | 12.4 | 12.6 | -0.01 | 12.4 | 12.8 | -0.01 |
| 85 - 89 | 6.0 | 6.5 | -0.02 | 6.1 | 6.5 | -0.01 | 6.5 | 6.5 | 0.00 |
| 90 - 94 | 1.9 | 2.1 | -0.02 | 2.0 | 1.8 | 0.01 | 2.0 | 2.0 | 0.00 |
| 95 - 99 | 0.4 | 0.3 | 0.02 | 0.4 | 0.3 | 0.03 | 0.5 | 0.3 | 0.03 |
| **Medical history: General** | | | | | | | | | |
| Acute respiratory disease | 25.7 | 22.8 | 0.07 | 24.8 | 24.8 | 0.00 | 24.5 | 24.3 | 0.00 |
| Attention deficit hyperactivity disorder | 0.2 | 0.1 | 0.01 | 0.2 | 0.2 | 0.00 | 0.1 | 0.1 | 0.00 |
| Chronic liver disease | 2.8 | 2.4 | 0.03 | 2.7 | 2.7 | 0.00 | 2.7 | 2.7 | 0.00 |
| Chronic obstructive lung disease | 10.1 | 10.3 | -0.01 | 9.8 | 10.0 | -0.01 | 11.1 | 10.5 | 0.02 |
| Crohn’s disease | 0.2 | 0.3 | -0.01 | 0.2 | 0.4 | -0.03 | 0.2 | 0.4 | -0.04 |
| Dementia | 3.3 | 3.4 | -0.01 | 3.4 | 3.5 | 0.00 | 3.7 | 3.5 | 0.01 |
| Depressive disorder | 10.1 | 9.0 | 0.04 | 10.1 | 10.3 | 0.00 | 10.5 | 10.5 | 0.00 |
| Gastroesophageal reflux disease | 15.1 | 11.6 | 0.10 | 14.3 | 15.0 | -0.02 | 13.6 | 14.2 | -0.02 |
| Gastrointestinal hemorrhage | 3.5 | 3.6 | 0.00 | 3.4 | 3.7 | -0.02 | 3.6 | 3.6 | 0.00 |
| Hyperlipidemia | 61.5 | 48.0 | 0.27 | 59.9 | 61.2 | -0.03 | 56.8 | 58.1 | -0.03 |
| Hypertensive disorder | 75.9 | 68.1 | 0.17 | 75.0 | 75.9 | -0.02 | 73.6 | 74.0 | -0.01 |
| Lesion of liver | 1.0 | 1.2 | -0.02 | 1.0 | 1.1 | -0.02 | 1.0 | 1.2 | -0.02 |
| Obesity | 10.7 | 8.4 | 0.08 | 10.5 | 11.0 | -0.02 | 10.3 | 10.8 | -0.02 |
| Osteoarthritis | 34.2 | 29.8 | 0.09 | 33.4 | 34.3 | -0.02 | 32.6 | 33.0 | -0.01 |
| Pneumonia | 4.7 | 5.8 | -0.05 | 4.8 | 4.4 | 0.02 | 5.6 | 5.2 | 0.02 |
| Psoriasis | 1.7 | 1.3 | 0.04 | 1.7 | 1.7 | 0.01 | 1.6 | 1.6 | 0.00 |
| Renal impairment | 13.1 | 11.6 | 0.05 | 13.1 | 13.3 | -0.01 | 13.9 | 13.7 | 0.01 |
| Rheumatoid arthritis | 2.3 | 2.1 | 0.01 | 2.2 | 2.3 | -0.01 | 2.2 | 2.3 | -0.01 |
| Schizophrenia | 0.2 | 0.2 | 0.00 | 0.2 | 0.1 | 0.02 | 0.2 | 0.1 | 0.01 |
| Ulcerative colitis | 0.6 | 0.4 | 0.03 | 0.5 | 0.4 | 0.02 | 0.5 | 0.4 | 0.02 |
| Urinary tract infectious disease | 15.4 | 15.0 | 0.01 | 15.3 | 15.6 | -0.01 | 15.7 | 15.6 | 0.00 |
| Viral hepatitis C | 0.2 | 0.3 | -0.01 | 0.2 | 0.3 | -0.01 | 0.2 | 0.3 | -0.01 |
| Visual system disorder | 54.7 | 50.0 | 0.09 | 53.8 | 54.6 | -0.01 | 51.8 | 52.7 | -0.02 |
| **Medical history: Cardiovascular disease** | | | | | | | | | |
| Atrial fibrillation | 10.5 | 9.5 | 0.03 | 10.8 | 10.1 | 0.03 | 10.8 | 10.1 | 0.03 |
| Cerebrovascular disease | 12.2 | 11.4 | 0.03 | 11.6 | 11.9 | -0.01 | 12.0 | 11.5 | 0.01 |
| Coronary arteriosclerosis | 17.4 | 17.0 | 0.01 | 17.0 | 17.6 | -0.02 | 17.6 | 17.4 | 0.00 |
| Heart disease | 39.6 | 37.9 | 0.04 | 38.8 | 39.4 | -0.01 | 39.2 | 38.5 | 0.01 |
| Heart failure | 9.1 | 9.9 | -0.03 | 9.2 | 9.0 | 0.01 | 10.0 | 9.5 | 0.02 |
| Ischemic heart disease | 8.5 | 8.9 | -0.02 | 8.8 | 8.4 | 0.02 | 8.8 | 8.4 | 0.02 |
| Peripheral vascular disease | 12.4 | 10.1 | 0.07 | 11.9 | 12.1 | 0.00 | 11.7 | 11.3 | 0.01 |
| Venous thrombosis | 2.2 | 2.3 | 0.00 | 2.2 | 2.6 | -0.02 | 2.2 | 2.5 | -0.02 |
| **Medical history: Neoplasms** | | | | | | | | | |
| Hematologic neoplasm | 1.5 | 1.6 | -0.01 | 1.4 | 1.8 | -0.03 | 1.4 | 1.8 | -0.03 |
| Malignant lymphoma | 0.8 | 0.7 | 0.02 | 0.8 | 0.8 | -0.01 | 0.8 | 0.8 | 0.00 |
| Malignant neoplasm of anorectum | 0.3 | 0.2 | 0.01 | 0.3 | 0.2 | 0.01 | 0.4 | 0.2 | 0.03 |
| Malignant neoplastic disease | 15.7 | 14.1 | 0.04 | 15.6 | 15.4 | 0.01 | 15.3 | 15.0 | 0.01 |
| Malignant tumor of breast | 5.8 | 5.7 | 0.01 | 5.9 | 6.0 | 0.00 | 5.8 | 5.9 | 0.00 |
| Malignant tumor of colon | 0.9 | 0.7 | 0.02 | 0.9 | 0.7 | 0.03 | 0.9 | 0.7 | 0.03 |
| Malignant tumor of lung | 0.6 | 0.7 | -0.02 | 0.6 | 0.7 | -0.02 | 0.7 | 0.8 | -0.01 |
| Malignant tumor of urinary bladder | 0.4 | 0.5 | 0.00 | 0.4 | 0.5 | 0.00 | 0.4 | 0.5 | -0.01 |
| **Medication use** | | | | | | | | | |
| Agents acting on the renin-angiotensin system | 75.3 | 72.0 | 0.07 | 75.3 | 74.9 | 0.01 | 75.1 | 74.8 | 0.01 |
| Antibacterials for systemic use | 66.8 | 64.3 | 0.05 | 65.8 | 66.6 | -0.02 | 65.7 | 65.9 | 0.00 |
| Antidepressants | 32.6 | 31.5 | 0.02 | 32.8 | 32.6 | 0.01 | 32.8 | 32.6 | 0.01 |
| Antiepileptics | 17.3 | 16.4 | 0.02 | 17.2 | 17.5 | -0.01 | 17.8 | 17.8 | 0.00 |
| Antiinflammatory and antirheumatic products | 31.2 | 29.2 | 0.04 | 30.5 | 30.6 | 0.00 | 29.5 | 29.9 | -0.01 |

| (Continued) | Before ajustment | | | After matching | | | After stratification | | |
|---|---|---|---|---|---|---|---|---|---|
| Characteristic | T (%) | C (%) | StdDiff | T (%) | C (%) | StdDiff | T (%) | C (%) | StdDiff |
| Antipsoriatics | 0.8 | 0.7 | 0.01 | 0.7 | 0.9 | -0.02 | 0.7 | 0.8 | -0.02 |
| Antithrombotic agents | 22.0 | 21.8 | 0.00 | 21.8 | 21.9 | 0.00 | 22.6 | 22.1 | 0.01 |
| Beta blocking agents | 48.8 | 47.4 | 0.03 | 48.6 | 48.8 | 0.00 | 49.7 | 49.1 | 0.01 |
| Calcium channel blockers | 37.1 | 35.2 | 0.04 | 36.4 | 36.4 | 0.00 | 36.4 | 36.4 | 0.00 |
| Diuretics | 57.4 | 59.2 | -0.04 | 57.7 | 57.2 | 0.01 | 58.9 | 58.4 | 0.01 |
| Drugs for acid related disorders | 37.3 | 35.2 | 0.04 | 36.3 | 36.8 | -0.01 | 36.2 | 36.5 | -0.01 |
| Drugs for obstructive airway diseases | 45.0 | 40.0 | 0.10 | 44.0 | 44.2 | 0.00 | 43.0 | 43.1 | 0.00 |
| Immunosuppressants | 4.5 | 3.5 | 0.05 | 4.4 | 4.1 | 0.01 | 4.3 | 4.0 | 0.01 |
| Lipid modifying agents | 79.4 | 73.6 | 0.14 | 78.7 | 78.9 | 0.00 | 77.7 | 78.0 | -0.01 |
| Opioids | 30.4 | 30.8 | -0.01 | 30.1 | 30.2 | 0.00 | 31.2 | 31.0 | 0.00 |
| Psycholeptics | 30.7 | 29.4 | 0.03 | 30.1 | 30.1 | 0.00 | 30.5 | 30.1 | 0.01 |
| Psychostimulants | 1.7 | 1.4 | 0.02 | 1.7 | 1.6 | 0.01 | 1.7 | 1.6 | 0.01 |

##### 2.6.5 MDCD

**Supplementary Table 195:** Baseline patient characteristics for DPP4I (female) (T) and SU (female) (C) new-users in the MDCD data source. We report proportion of initiators satisfying selected base-line characteristics and the standardized difference of population proportions (StdDiff) before and after propensity score adjustment. Less extreme StdDiffs through matching and stratification suggest improved balance between patient cohorts through adjustment.

| | Before adjustment | | | After matching | | | After stratification | | |
|---|---|---|---|---|---|---|---|---|---|
| Characteristic | T (%) | C (%) | StdDiff | T (%) | C (%) | StdDiff | T (%) | C (%) | StdDiff |
| **Age group** | | | | | | | | | |
| 15 - 19 | 0.6 | 0.6 | 0.00 | 0.6 | 0.5 | 0.02 | 0.5 | 0.6 | 0.00 |
| 20 - 24 | 1.1 | 1.4 | -0.03 | 1.0 | 1.0 | 0.00 | 1.0 | 1.3 | -0.03 |
| 25 - 29 | 2.7 | 3.6 | -0.05 | 2.8 | 2.6 | 0.02 | 3.3 | 3.3 | 0.00 |
| 30 - 34 | 5.6 | 6.4 | -0.03 | 5.7 | 5.2 | 0.02 | 6.1 | 6.0 | 0.00 |
| 35 - 39 | 9.2 | 9.3 | 0.00 | 9.3 | 8.9 | 0.02 | 9.4 | 9.1 | 0.01 |
| 40 - 44 | 11.7 | 11.5 | 0.01 | 11.8 | 11.8 | 0.00 | 11.8 | 11.6 | 0.00 |
| 45 - 49 | 13.3 | 13.0 | 0.01 | 13.3 | 13.5 | -0.01 | 13.0 | 13.2 | -0.01 |
| 50 - 54 | 15.9 | 15.3 | 0.02 | 15.8 | 15.6 | 0.00 | 15.8 | 15.4 | 0.01 |
| 55 - 59 | 16.1 | 16.1 | 0.00 | 16.0 | 17.1 | -0.03 | 15.6 | 16.4 | -0.02 |
| 60 - 64 | 13.0 | 13.2 | -0.01 | 12.8 | 13.5 | -0.02 | 12.8 | 13.5 | -0.02 |
| 65 - 69 | 4.6 | 4.3 | 0.02 | 4.6 | 4.7 | 0.00 | 4.6 | 4.4 | 0.01 |
| 70 - 74 | 2.4 | 2.2 | 0.01 | 2.4 | 2.5 | -0.01 | 2.2 | 2.3 | -0.01 |
| 75 - 79 | 1.9 | 1.5 | 0.03 | 1.9 | 1.5 | 0.03 | 1.7 | 1.5 | 0.02 |
| 80 - 84 | 1.1 | 1.0 | 0.01 | 1.1 | 1.0 | 0.01 | 1.1 | 1.0 | 0.01 |
| 85 - 89 | 0.7 | 0.6 | 0.02 | 0.7 | 0.6 | 0.02 | 0.7 | 0.6 | 0.01 |
| **Race** | | | | | | | | | |
| Black or African American | 26.5 | 30.9 | -0.10 | 27.0 | 26.1 | 0.02 | 29.7 | 29.2 | 0.01 |
| White | 57.0 | 51.1 | 0.12 | 56.2 | 57.5 | -0.03 | 52.5 | 53.3 | -0.02 |
| **Ethnicity** | | | | | | | | | |
| Hispanic or Latino | 3.0 | 2.7 | 0.02 | 3.0 | 2.9 | 0.01 | 2.9 | 2.8 | 0.01 |
| **Medical history: General** | | | | | | | | | |
| Acute respiratory disease | 40.8 | 41.0 | 0.00 | 40.4 | 41.0 | -0.01 | 40.6 | 41.1 | -0.01 |
| Attention deficit hyperactivity disorder | 1.9 | 1.5 | 0.03 | 1.8 | 1.9 | 0.00 | 1.7 | 1.6 | 0.00 |
| Chronic liver disease | 4.9 | 5.7 | -0.04 | 4.9 | 5.2 | -0.01 | 5.4 | 5.5 | 0.00 |
| Chronic obstructive lung disease | 20.2 | 18.5 | 0.04 | 19.5 | 20.2 | -0.02 | 19.2 | 19.3 | 0.00 |
| Crohn’s disease | 0.4 | 0.3 | 0.02 | 0.4 | 0.3 | 0.02 | 0.4 | 0.3 | 0.01 |
| Dementia | 1.9 | 1.5 | 0.03 | 1.9 | 1.7 | 0.02 | 1.8 | 1.6 | 0.02 |
| Depressive disorder | 38.1 | 36.5 | 0.03 | 37.6 | 38.6 | -0.02 | 36.9 | 37.3 | -0.01 |
| Gastroesophageal reflux disease | 29.7 | 27.1 | 0.06 | 29.1 | 29.5 | -0.01 | 27.5 | 28.1 | -0.01 |
| Gastrointestinal hemorrhage | 3.7 | 3.8 | -0.01 | 3.6 | 3.8 | -0.01 | 3.6 | 3.8 | -0.01 |
| Hyperlipidemia | 64.7 | 59.7 | 0.10 | 63.8 | 64.9 | -0.02 | 61.5 | 61.6 | 0.00 |
| Hypertensive disorder | 75.1 | 74.1 | 0.02 | 74.9 | 74.7 | 0.00 | 75.4 | 74.4 | 0.02 |
| Lesion of liver | 1.1 | 1.4 | -0.02 | 1.1 | 1.3 | -0.01 | 1.3 | 1.4 | 0.00 |
| Obesity | 39.6 | 39.5 | 0.00 | 39.3 | 40.0 | -0.01 | 39.1 | 39.7 | -0.01 |
| Osteoarthritis | 38.5 | 34.5 | 0.08 | 37.6 | 38.5 | -0.02 | 35.8 | 36.0 | 0.00 |
| Pneumonia | 6.0 | 6.7 | -0.03 | 5.9 | 6.0 | 0.00 | 6.4 | 6.5 | 0.00 |
| Renal impairment | 8.1 | 8.1 | 0.00 | 7.9 | 8.4 | -0.02 | 8.0 | 8.2 | -0.01 |
| Rheumatoid arthritis | 2.8 | 2.3 | 0.03 | 2.6 | 2.5 | 0.00 | 2.7 | 2.4 | 0.02 |
| Schizophrenia | 3.8 | 4.2 | -0.02 | 3.9 | 3.5 | 0.02 | 4.4 | 3.9 | 0.02 |
| Ulcerative colitis | 0.2 | 0.2 | 0.00 | 0.2 | 0.2 | -0.01 | 0.2 | 0.2 | -0.01 |
| Urinary tract infectious disease | 16.5 | 18.6 | -0.06 | 16.7 | 16.5 | 0.01 | 17.6 | 18.0 | -0.01 |
| Viral hepatitis C | 1.7 | 2.1 | -0.03 | 1.7 | 1.9 | -0.01 | 2.0 | 2.1 | -0.01 |
| Visual system disorder | 44.9 | 41.5 | 0.07 | 44.3 | 45.9 | -0.03 | 41.5 | 43.0 | -0.03 |
| **Medical history: Cardiovascular disease** | | | | | | | | | |
| Atrial fibrillation | 3.2 | 2.8 | 0.03 | 3.2 | 2.9 | 0.01 | 3.3 | 2.8 | 0.03 |
| Cerebrovascular disease | 5.1 | 5.1 | 0.00 | 5.0 | 5.3 | -0.01 | 5.1 | 5.2 | 0.00 |
| Coronary arteriosclerosis | 10.5 | 9.3 | 0.04 | 10.2 | 10.1 | 0.00 | 9.8 | 9.7 | 0.00 |
| Heart disease | 27.5 | 26.8 | 0.01 | 27.1 | 26.9 | 0.00 | 27.8 | 27.0 | 0.02 |
| Heart failure | 8.1 | 8.4 | -0.01 | 8.1 | 7.9 | 0.01 | 8.7 | 8.3 | 0.01 |
| Ischemic heart disease | 7.2 | 6.9 | 0.01 | 7.1 | 7.0 | 0.00 | 7.2 | 7.0 | 0.01 |
| Peripheral vascular disease | 7.6 | 6.9 | 0.03 | 7.3 | 7.6 | -0.01 | 7.0 | 7.1 | 0.00 |
| Pulmonary embolism | 1.1 | 1.1 | 0.00 | 1.1 | 1.1 | -0.01 | 1.2 | 1.1 | 0.00 |
| Venous thrombosis | 1.7 | 1.7 | 0.00 | 1.8 | 1.8 | 0.00 | 1.8 | 1.8 | 0.00 |
| **Medical history: Neoplasms** | | | | | | | | | |
| Hematologic neoplasm | 0.8 | 1.0 | -0.02 | 0.8 | 1.0 | -0.02 | 0.9 | 1.0 | -0.02 |
| Malignant lymphoma | 0.3 | 0.3 | 0.00 | 0.4 | 0.3 | 0.01 | 0.4 | 0.3 | 0.01 |
| Malignant neoplasm of anorectum | 0.2 | 0.2 | 0.01 | 0.2 | 0.1 | 0.02 | 0.2 | 0.2 | 0.01 |
| Malignant neoplastic disease | 6.3 | 6.5 | -0.01 | 6.2 | 6.6 | -0.01 | 6.2 | 6.5 | -0.01 |

| (Continued) | Before ajustment | | | After matching | | | After stratification | | |
|---|---|---|---|---|---|---|---|---|---|
| Characteristic | T (%) | C (%) | StdDiff | T (%) | C (%) | StdDiff | T (%) | C (%) | StdDiff |
| Malignant tumor of breast | 2.6 | 2.4 | 0.01 | 2.5 | 2.6 | 0.00 | 2.4 | 2.5 | 0.00 |
| Malignant tumor of colon | 0.4 | 0.4 | 0.00 | 0.4 | 0.4 | 0.00 | 0.4 | 0.4 | 0.00 |
| Malignant tumor of lung | 0.3 | 0.3 | -0.02 | 0.3 | 0.3 | -0.01 | 0.2 | 0.3 | -0.02 |
| Malignant tumor of urinary bladder | 0.1 | 0.1 | -0.01 | <0.1 | 0.1 | 0.00 | 0.1 | 0.1 | -0.01 |
| **Medication use** | | | | | | | | | |
| Agents acting on the renin-angiotensin system | 61.0 | 60.7 | 0.01 | 60.9 | 60.9 | 0.00 | 61.2 | 60.8 | 0.01 |
| Antibacterials for systemic use | 73.0 | 72.6 | 0.01 | 72.6 | 72.9 | -0.01 | 72.6 | 72.8 | 0.00 |
| Antidepressants | 57.7 | 53.9 | 0.08 | 56.6 | 57.9 | -0.03 | 55.3 | 55.4 | 0.00 |
| Antiepileptics | 42.8 | 39.1 | 0.07 | 41.9 | 42.9 | -0.02 | 40.3 | 40.5 | 0.00 |
| Antiinflammatory and antirheumatic products | 62.3 | 59.9 | 0.05 | 61.9 | 62.5 | -0.01 | 60.3 | 60.8 | -0.01 |
| Antineoplastic agents | 5.2 | 4.3 | 0.04 | 4.9 | 4.9 | 0.00 | 4.6 | 4.5 | 0.00 |
| Antipsoriatics | 0.5 | 0.4 | 0.01 | 0.5 | 0.5 | -0.01 | 0.5 | 0.5 | 0.00 |
| Antithrombotic agents | 23.7 | 22.4 | 0.03 | 23.3 | 24.1 | -0.02 | 22.7 | 23.0 | -0.01 |
| Beta blocking agents | 33.2 | 31.8 | 0.03 | 32.9 | 33.3 | -0.01 | 32.6 | 32.4 | 0.00 |
| Calcium channel blockers | 24.5 | 24.8 | 0.00 | 24.5 | 24.5 | 0.00 | 25.1 | 24.7 | 0.01 |
| Diuretics | 47.5 | 48.4 | -0.02 | 47.4 | 47.3 | 0.00 | 48.9 | 48.1 | 0.02 |
| Drugs for acid related disorders | 52.5 | 48.6 | 0.08 | 51.4 | 52.6 | -0.02 | 49.4 | 50.1 | -0.01 |
| Drugs for obstructive airway diseases | 62.3 | 58.1 | 0.09 | 61.3 | 62.1 | -0.02 | 59.4 | 59.6 | 0.00 |
| Immunosuppressants | 3.1 | 2.4 | 0.05 | 2.9 | 2.7 | 0.02 | 2.8 | 2.5 | 0.02 |
| Lipid modifying agents | 65.5 | 60.1 | 0.11 | 64.6 | 66.2 | -0.03 | 61.9 | 62.3 | -0.01 |
| Opioids | 49.0 | 49.4 | -0.01 | 48.6 | 48.6 | 0.00 | 49.2 | 49.2 | 0.00 |
| Psycholeptics | 50.2 | 49.0 | 0.02 | 49.6 | 50.2 | -0.01 | 49.4 | 49.6 | 0.00 |
| Psychostimulants | 6.0 | 5.2 | 0.04 | 6.0 | 5.7 | 0.01 | 5.6 | 5.4 | 0.01 |

##### 2.6.6 Open Claims

**Supplementary Table 196:** Baseline patient characteristics for DPP4I (female) (T) and SU (female) (C) new-users in the Open Claims data source. We report proportion of initiators satisfying selected base-line characteristics and the standardized difference of population proportions (StdDiff) before and after propensity score adjustment. Less extreme StdDiffs through matching and stratification suggest improved balance between patient cohorts through adjustment.

| | Before adjustment | | | After matching | | | After stratification | | |
|---|---|---|---|---|---|---|---|---|---|
| Characteristic | T (%) | C (%) | StdDiff | T (%) | C (%) | StdDiff | T (%) | C (%) | StdDiff |
| **Age group** | | | | | | | | | |
| 15 - 19 | 0.1 | 0.1 | 0.00 | 0.1 | 0.1 | 0.00 | 0.1 | 0.1 | 0.00 |
| 20 - 24 | 0.4 | 0.6 | -0.02 | 0.4 | 0.4 | 0.00 | 0.5 | 0.5 | -0.01 |
| 25 - 29 | 0.8 | 1.0 | -0.02 | 0.8 | 0.9 | 0.00 | 0.9 | 1.0 | -0.01 |
| 30 - 34 | 1.6 | 2.0 | -0.03 | 1.6 | 1.6 | 0.00 | 1.9 | 1.9 | 0.00 |
| 35 - 39 | 3.0 | 3.3 | -0.02 | 3.0 | 2.9 | 0.01 | 3.2 | 3.2 | 0.00 |
| 40 - 44 | 5.0 | 5.2 | -0.01 | 5.1 | 4.9 | 0.01 | 5.2 | 5.1 | 0.00 |
| 45 - 49 | 8.0 | 7.9 | 0.00 | 8.0 | 7.6 | 0.01 | 8.0 | 7.8 | 0.01 |
| 50 - 54 | 11.4 | 11.1 | 0.01 | 11.5 | 11.3 | 0.01 | 11.3 | 11.2 | 0.00 |
| 55 - 59 | 13.9 | 13.5 | 0.01 | 13.9 | 13.8 | 0.00 | 13.7 | 13.6 | 0.00 |
| 60 - 64 | 14.6 | 14.1 | 0.01 | 14.5 | 14.8 | -0.01 | 14.1 | 14.2 | 0.00 |
| 65 - 69 | 14.3 | 14.8 | -0.01 | 14.3 | 14.7 | -0.01 | 14.4 | 14.7 | -0.01 |
| 70 - 74 | 11.2 | 11.5 | -0.01 | 11.2 | 10.9 | 0.01 | 11.7 | 11.4 | 0.01 |
| 75 - 79 | 10.9 | 10.7 | 0.00 | 10.7 | 11.1 | -0.01 | 10.7 | 10.9 | -0.01 |
| 80 - 84 | 4.6 | 4.1 | 0.03 | 4.6 | 4.7 | -0.01 | 4.2 | 4.3 | 0.00 |
| 85 - 89 | 0.1 | 0.1 | 0.01 | 0.1 | 0.1 | 0.01 | 0.1 | 0.1 | 0.01 |
| **Medical history: General** | | | | | | | | | |
| Acute respiratory disease | 17.2 | 15.9 | 0.04 | 17.0 | 17.5 | -0.01 | 16.3 | 16.5 | 0.00 |
| Attention deficit hyperactivity disorder | 0.4 | 0.4 | 0.01 | 0.4 | 0.4 | 0.00 | 0.4 | 0.4 | 0.00 |
| Chronic liver disease | 1.6 | 1.6 | 0.00 | 1.6 | 1.6 | 0.00 | 1.6 | 1.6 | 0.00 |
| Chronic obstructive lung disease | 5.8 | 6.0 | -0.01 | 5.8 | 5.9 | -0.01 | 5.9 | 5.9 | 0.00 |
| Crohn's disease | 0.2 | 0.2 | 0.00 | 0.2 | 0.2 | 0.00 | 0.2 | 0.2 | 0.00 |
| Dementia | 1.3 | 1.0 | 0.02 | 1.2 | 1.3 | -0.01 | 1.1 | 1.1 | 0.00 |
| Depressive disorder | 9.8 | 9.6 | 0.01 | 9.7 | 10.0 | -0.01 | 9.7 | 10.0 | -0.01 |
| Gastroesophageal reflux disease | 10.5 | 9.2 | 0.04 | 10.3 | 10.5 | -0.01 | 9.7 | 9.6 | 0.00 |
| Gastrointestinal hemorrhage | 1.5 | 1.4 | 0.01 | 1.5 | 1.6 | 0.00 | 1.5 | 1.5 | 0.00 |
| Human immunodeficiency virus infection | 0.2 | 0.2 | 0.01 | 0.2 | 0.2 | 0.01 | 0.2 | 0.2 | 0.00 |
| Hyperlipidemia | 44.4 | 39.3 | 0.10 | 43.7 | 44.1 | -0.01 | 41.1 | 41.0 | 0.00 |
| Hypertensive disorder | 51.6 | 48.5 | 0.06 | 51.2 | 51.5 | -0.01 | 49.8 | 49.5 | 0.01 |
| Lesion of liver | 0.6 | 0.7 | -0.01 | 0.7 | 0.7 | 0.00 | 0.7 | 0.7 | 0.00 |
| Obesity | 12.1 | 11.7 | 0.01 | 12.2 | 12.1 | 0.00 | 11.9 | 11.8 | 0.00 |
| Osteoarthritis | 18.0 | 16.3 | 0.05 | 17.6 | 17.8 | 0.00 | 17.0 | 16.9 | 0.00 |
| Pneumonia | 2.6 | 2.7 | -0.01 | 2.6 | 2.7 | -0.01 | 2.7 | 2.7 | 0.00 |
| Psoriasis | 0.9 | 0.8 | 0.01 | 0.9 | 0.9 | 0.00 | 0.9 | 0.9 | 0.00 |
| Renal impairment | 7.1 | 7.0 | 0.01 | 7.1 | 7.3 | -0.01 | 7.0 | 7.1 | 0.00 |
| Rheumatoid arthritis | 1.6 | 1.4 | 0.02 | 1.6 | 1.5 | 0.00 | 1.5 | 1.4 | 0.01 |
| Schizophrenia | 0.4 | 0.4 | 0.00 | 0.4 | 0.4 | 0.00 | 0.4 | 0.4 | 0.00 |
| Ulcerative colitis | 0.2 | 0.2 | 0.01 | 0.2 | 0.2 | 0.00 | 0.2 | 0.2 | 0.00 |
| Urinary tract infectious disease | 8.9 | 8.3 | 0.02 | 8.6 | 8.5 | 0.00 | 8.6 | 8.5 | 0.00 |
| Viral hepatitis C | 0.3 | 0.4 | -0.01 | 0.3 | 0.4 | 0.00 | 0.3 | 0.4 | 0.00 |
| Visual system disorder | 21.3 | 19.5 | 0.04 | 20.9 | 21.3 | -0.01 | 20.2 | 20.1 | 0.00 |
| **Medical history: Cardiovascular disease** | | | | | | | | | |
| Atrial fibrillation | 3.4 | 3.2 | 0.01 | 3.4 | 3.4 | 0.00 | 3.4 | 3.3 | 0.00 |
| Cerebrovascular disease | 3.7 | 3.2 | 0.03 | 3.5 | 3.6 | 0.00 | 3.3 | 3.3 | 0.00 |
| Coronary arteriosclerosis | 6.3 | 5.8 | 0.02 | 6.0 | 5.9 | 0.01 | 6.0 | 5.9 | 0.01 |
| Heart disease | 16.7 | 15.4 | 0.04 | 16.4 | 16.3 | 0.00 | 16.2 | 15.8 | 0.01 |
| Heart failure | 4.0 | 4.0 | 0.00 | 4.0 | 4.0 | 0.00 | 4.1 | 4.0 | 0.00 |
| Ischemic heart disease | 3.4 | 3.2 | 0.01 | 3.3 | 3.3 | 0.00 | 3.3 | 3.3 | 0.00 |
| Peripheral vascular disease | 4.6 | 4.2 | 0.02 | 4.4 | 4.5 | 0.00 | 4.3 | 4.4 | 0.00 |
| Pulmonary embolism | 0.5 | 0.5 | 0.00 | 0.5 | 0.5 | 0.00 | 0.5 | 0.5 | 0.00 |
| Venous thrombosis | 0.9 | 0.9 | 0.01 | 0.9 | 0.9 | 0.00 | 0.9 | 0.9 | 0.00 |
| **Medical history: Neoplasms** | | | | | | | | | |
| Hematologic neoplasm | 0.7 | 0.7 | 0.00 | 0.7 | 0.7 | 0.00 | 0.7 | 0.7 | 0.00 |
| Malignant lymphoma | 0.3 | 0.3 | 0.00 | 0.3 | 0.3 | 0.00 | 0.3 | 0.3 | 0.00 |
| Malignant neoplasm of anorectum | 0.1 | 0.1 | 0.00 | 0.1 | 0.1 | 0.01 | 0.1 | 0.1 | 0.01 |
| Malignant neoplastic disease | 6.1 | 5.8 | 0.01 | 6.1 | 6.3 | -0.01 | 5.9 | 5.9 | 0.00 |
| Malignant tumor of breast | 2.5 | 2.2 | 0.01 | 2.4 | 2.5 | -0.01 | 2.3 | 2.4 | 0.00 |
| Malignant tumor of colon | 0.3 | 0.3 | 0.00 | 0.3 | 0.3 | 0.00 | 0.3 | 0.3 | 0.00 |

| (Continued) | Before ajustment | | | After matching | | | After stratification | | |
|---|---|---|---|---|---|---|---|---|---|
| Characteristic | T (%) | C (%) | StdDiff | T (%) | C (%) | StdDiff | T (%) | C (%) | StdDiff |
| Malignant tumor of lung | 0.3 | 0.3 | -0.01 | 0.3 | 0.3 | 0.00 | 0.3 | 0.3 | 0.00 |
| Malignant tumor of urinary bladder | 0.1 | 0.1 | 0.00 | 0.1 | 0.1 | 0.00 | 0.1 | 0.1 | 0.01 |
| **Medication use** | | | | | | | | | |
| Agents acting on the renin-angiotensin system | 66.9 | 65.9 | 0.02 | 66.7 | 66.8 | 0.00 | 66.5 | 66.1 | 0.01 |
| Antibacterials for systemic use | 62.7 | 60.7 | 0.04 | 62.5 | 62.8 | -0.01 | 61.2 | 61.6 | -0.01 |
| Antidepressants | 36.9 | 35.9 | 0.02 | 36.7 | 37.0 | -0.01 | 36.7 | 37.0 | -0.01 |
| Antiepileptics | 23.0 | 22.8 | 0.00 | 22.8 | 23.1 | -0.01 | 23.2 | 23.1 | 0.00 |
| Antiinflammatory and antirheumatic products | 41.1 | 39.4 | 0.04 | 40.7 | 40.9 | 0.00 | 40.3 | 39.9 | 0.01 |
| Antineoplastic agents | 5.5 | 4.6 | 0.04 | 5.4 | 5.3 | 0.00 | 5.1 | 4.9 | 0.01 |
| Antipsoriatics | 1.3 | 1.0 | 0.02 | 1.2 | 1.2 | 0.00 | 1.2 | 1.1 | 0.01 |
| Antithrombotic agents | 18.0 | 17.4 | 0.02 | 17.8 | 18.3 | -0.01 | 17.6 | 17.7 | 0.00 |
| Beta blocking agents | 33.8 | 33.3 | 0.01 | 33.6 | 34.2 | -0.01 | 33.5 | 33.7 | 0.00 |
| Calcium channel blockers | 26.3 | 25.6 | 0.02 | 26.1 | 26.2 | 0.00 | 26.1 | 26.2 | 0.00 |
| Diuretics | 44.6 | 44.0 | 0.01 | 44.4 | 44.3 | 0.00 | 44.6 | 44.1 | 0.01 |
| Drugs for acid related disorders | 39.5 | 36.8 | 0.06 | 39.1 | 39.3 | -0.01 | 37.9 | 37.8 | 0.00 |
| Drugs for obstructive airway diseases | 43.5 | 39.9 | 0.07 | 43.0 | 43.3 | 0.00 | 41.4 | 41.2 | 0.00 |
| Immunosuppressants | 3.7 | 3.0 | 0.04 | 3.6 | 3.5 | 0.00 | 3.4 | 3.2 | 0.01 |
| Lipid modifying agents | 69.4 | 66.2 | 0.07 | 69.2 | 69.5 | -0.01 | 67.3 | 67.4 | 0.00 |
| Opioids | 28.4 | 28.2 | 0.01 | 28.3 | 28.6 | -0.01 | 28.3 | 28.4 | 0.00 |
| Psycholeptics | 27.9 | 26.8 | 0.03 | 27.6 | 27.9 | -0.01 | 27.6 | 27.9 | -0.01 |
| Psychostimulants | 3.1 | 2.8 | 0.02 | 3.1 | 3.0 | 0.00 | 2.9 | 2.8 | 0.00 |

##### 2.6.7 OEHR

**Supplementary Table 197:** Baseline patient characteristics for DPP4I (female) (T) and SU (female) (C) new-users in the OEHR data source. We report proportion of initiators satisfying selected base-line characteristics and the standardized difference of population proportions (StdDiff) before and after propensity score adjustment. Less extreme StdDiffs through matching and stratification suggest improved balance between patient cohorts through adjustment.

| | Before adjustment | | | After matching | | | After stratification | | |
|---|---|---|---|---|---|---|---|---|---|
| Characteristic | T (%) | C (%) | StdDiff | T (%) | C (%) | StdDiff | T (%) | C (%) | StdDiff |
| **Age group** | | | | | | | | | |
| 15 - 19 | 0.1 | 0.1 | 0.00 | 0.1 | 0.1 | 0.00 | 0.1 | 0.1 | 0.00 |
| 20 - 24 | 0.5 | 0.6 | -0.01 | 0.5 | 0.5 | 0.00 | 0.5 | 0.5 | 0.00 |
| 25 - 29 | 1.1 | 1.3 | -0.02 | 1.1 | 1.1 | 0.00 | 1.2 | 1.3 | -0.01 |
| 30 - 34 | 2.2 | 2.4 | -0.01 | 2.3 | 2.1 | 0.01 | 2.3 | 2.1 | 0.01 |
| 35 - 39 | 4.0 | 4.2 | -0.01 | 4.1 | 4.0 | 0.01 | 4.1 | 4.0 | 0.01 |
| 40 - 44 | 6.8 | 6.2 | 0.02 | 6.7 | 6.8 | 0.00 | 6.4 | 6.4 | 0.00 |
| 45 - 49 | 9.9 | 9.6 | 0.01 | 9.8 | 9.8 | 0.00 | 9.7 | 9.7 | 0.00 |
| 50 - 54 | 13.8 | 12.9 | 0.02 | 13.6 | 13.7 | 0.00 | 13.6 | 13.7 | 0.00 |
| 60 - 64 | 14.8 | 14.3 | 0.01 | 14.8 | 14.9 | 0.00 | 14.4 | 14.5 | 0.00 |
| 65 - 69 | 12.2 | 12.2 | 0.00 | 12.2 | 12.3 | 0.00 | 12.2 | 12.2 | 0.00 |
| 75 - 79 | 5.7 | 6.7 | -0.04 | 5.8 | 5.8 | 0.00 | 6.5 | 6.4 | 0.00 |
| 80 - 84 | 3.7 | 4.1 | -0.02 | 4.0 | 4.0 | 0.00 | 4.0 | 4.0 | 0.00 |
| 85 - 89 | 1.0 | 0.9 | 0.01 | 1.0 | 1.0 | 0.00 | 1.0 | 0.9 | 0.00 |
| **Race** | | | | | | | | | |
| Asian | 3.2 | 2.8 | 0.02 | 3.1 | 3.0 | 0.00 | 2.8 | 2.9 | 0.00 |
| Black or African American | 15.2 | 13.9 | 0.04 | 14.2 | 14.4 | 0.00 | 14.2 | 14.4 | 0.00 |
| White | 73.9 | 74.7 | -0.02 | 74.0 | 74.1 | 0.00 | 74.9 | 74.5 | 0.01 |
| **Ethnicity** | | | | | | | | | |
| Hispanic or Latino | 7.0 | 8.3 | -0.05 | 7.1 | 6.9 | 0.01 | 7.7 | 7.8 | 0.00 |
| Not Hispanic or Latino | 85.6 | 84.5 | 0.03 | 85.5 | 85.8 | -0.01 | 85.2 | 84.9 | 0.01 |
| **Medical history: General** | | | | | | | | | |
| Acute respiratory disease | 22.2 | 20.3 | 0.05 | 21.7 | 21.8 | 0.00 | 21.0 | 21.0 | 0.00 |
| Attention deficit hyperactivity disorder | 0.8 | 0.7 | 0.01 | 0.7 | 0.8 | -0.01 | 0.7 | 0.8 | -0.01 |
| Chronic liver disease | 3.1 | 3.2 | 0.00 | 3.1 | 3.1 | 0.00 | 3.2 | 3.2 | 0.00 |
| Chronic obstructive lung disease | 5.4 | 5.7 | -0.01 | 5.4 | 5.5 | -0.01 | 5.8 | 5.7 | 0.00 |
| Crohn's disease | 0.3 | 0.3 | 0.00 | 0.3 | 0.3 | 0.00 | 0.3 | 0.3 | 0.00 |
| Dementia | 1.0 | 1.0 | 0.00 | 1.0 | 1.0 | 0.01 | 1.1 | 1.0 | 0.01 |
| Depressive disorder | 18.7 | 18.6 | 0.00 | 18.8 | 18.7 | 0.00 | 18.8 | 18.7 | 0.00 |
| Gastroesophageal reflux disease | 19.7 | 17.9 | 0.05 | 19.0 | 19.3 | -0.01 | 18.7 | 18.5 | 0.00 |
| Gastrointestinal hemorrhage | 1.4 | 1.4 | 0.00 | 1.4 | 1.4 | -0.01 | 1.4 | 1.5 | 0.00 |
| Human immunodeficiency virus infection | 0.2 | 0.1 | 0.01 | 0.1 | 0.1 | 0.02 | 0.2 | 0.1 | 0.02 |
| Hyperlipidemia | 64.6 | 61.0 | 0.07 | 63.4 | 64.0 | -0.01 | 62.2 | 62.3 | 0.00 |
| Hypertensive disorder | 65.1 | 63.7 | 0.03 | 64.5 | 64.9 | -0.01 | 64.6 | 64.2 | 0.01 |
| Lesion of liver | 0.7 | 0.8 | -0.01 | 0.7 | 0.7 | -0.01 | 0.7 | 0.8 | 0.00 |
| Obesity | 27.9 | 26.3 | 0.04 | 27.5 | 27.7 | 0.00 | 26.8 | 26.9 | 0.00 |
| Osteoarthritis | 18.8 | 18.1 | 0.02 | 18.4 | 18.6 | 0.00 | 18.6 | 18.4 | 0.01 |
| Pneumonia | 2.1 | 2.4 | -0.02 | 2.1 | 2.1 | 0.00 | 2.4 | 2.3 | 0.00 |
| Psoriasis | 1.3 | 1.3 | 0.00 | 1.3 | 1.3 | 0.00 | 1.3 | 1.4 | -0.01 |
| Renal impairment | 6.2 | 6.8 | -0.03 | 6.2 | 6.2 | 0.00 | 6.8 | 6.6 | 0.01 |
| Rheumatoid arthritis | 1.6 | 1.5 | 0.01 | 1.5 | 1.6 | -0.01 | 1.5 | 1.5 | 0.00 |
| Schizophrenia | 0.3 | 0.4 | -0.01 | 0.3 | 0.3 | 0.00 | 0.4 | 0.4 | 0.00 |
| Ulcerative colitis | 0.2 | 0.3 | 0.00 | 0.2 | 0.3 | 0.00 | 0.2 | 0.3 | 0.00 |
| Urinary tract infectious disease | 8.7 | 8.6 | 0.00 | 8.6 | 8.5 | 0.00 | 8.8 | 8.7 | 0.00 |
| Viral hepatitis C | 0.4 | 0.5 | -0.01 | 0.4 | 0.4 | 0.00 | 0.5 | 0.4 | 0.00 |
| Visual system disorder | 13.6 | 15.5 | -0.05 | 13.7 | 13.5 | 0.01 | 15.1 | 14.9 | 0.01 |
| **Medical history: Cardiovascular disease** | | | | | | | | | |
| Atrial fibrillation | 3.3 | 3.2 | 0.00 | 3.2 | 3.2 | 0.00 | 3.5 | 3.3 | 0.01 |
| Cerebrovascular disease | 2.7 | 2.8 | -0.01 | 2.6 | 2.7 | -0.01 | 2.8 | 2.8 | 0.00 |
| Coronary arteriosclerosis | 6.4 | 6.5 | 0.00 | 6.3 | 6.4 | 0.00 | 6.6 | 6.5 | 0.00 |
| Heart disease | 17.0 | 16.5 | 0.02 | 16.7 | 16.8 | 0.00 | 17.2 | 16.7 | 0.01 |
| Heart failure | 2.8 | 3.3 | -0.03 | 2.8 | 2.8 | 0.00 | 3.3 | 3.2 | 0.01 |
| Ischemic heart disease | 3.3 | 3.4 | 0.00 | 3.2 | 3.2 | 0.00 | 3.5 | 3.3 | 0.01 |
| Peripheral vascular disease | 3.6 | 3.8 | -0.01 | 3.5 | 3.6 | 0.00 | 3.8 | 3.7 | 0.00 |
| Pulmonary embolism | 0.5 | 0.5 | 0.00 | 0.5 | 0.5 | 0.00 | 0.6 | 0.5 | 0.01 |
| Venous thrombosis | 1.0 | 1.0 | 0.00 | 1.0 | 1.0 | 0.00 | 1.1 | 1.0 | 0.01 |
| **Medical history: Neoplasms** | | | | | | | | | |
| Hematologic neoplasm | 0.7 | 0.7 | 0.00 | 0.7 | 0.8 | 0.00 | 0.7 | 0.7 | 0.00 |
| Malignant lymphoma | 0.4 | 0.3 | 0.01 | 0.4 | 0.4 | 0.00 | 0.4 | 0.4 | 0.00 |

| (Continued) | Before ajustment | | | After matching | | | After stratification | | |
|---|---|---|---|---|---|---|---|---|---|
| Characteristic | T (%) | C (%) | StdDiff | T (%) | C (%) | StdDiff | T (%) | C (%) | StdDiff |
| Malignant neoplasm of anorectum | 0.1 | 0.1 | -0.01 | 0.1 | 0.1 | 0.00 | 0.1 | 0.1 | 0.00 |
| Malignant neoplastic disease | 6.6 | 6.6 | 0.00 | 6.5 | 6.7 | 0.00 | 6.7 | 6.6 | 0.00 |
| Malignant tumor of breast | 2.7 | 2.5 | 0.01 | 2.6 | 2.7 | 0.00 | 2.6 | 2.6 | 0.00 |
| Malignant tumor of colon | 0.3 | 0.3 | -0.01 | 0.3 | 0.3 | 0.00 | 0.3 | 0.3 | 0.00 |
| Malignant tumor of lung | 0.2 | 0.2 | -0.01 | 0.2 | 0.2 | -0.01 | 0.2 | 0.2 | -0.01 |
| Malignant tumor of urinary bladder | 0.1 | 0.1 | -0.01 | 0.1 | 0.1 | 0.00 | 0.1 | 0.1 | 0.00 |
| **Medication use** | | | | | | | | | |
| Agents acting on the renin-angiotensin system | 62.0 | 61.7 | 0.01 | 61.8 | 61.9 | 0.00 | 62.5 | 61.8 | 0.01 |
| Antibacterials for systemic use | 49.4 | 47.4 | 0.04 | 48.5 | 48.6 | 0.00 | 48.2 | 48.0 | 0.00 |
| Antidepressants | 38.4 | 37.2 | 0.03 | 37.9 | 38.1 | 0.00 | 37.6 | 37.6 | 0.00 |
| Antiepileptics | 20.3 | 19.7 | 0.01 | 20.2 | 19.9 | 0.01 | 20.2 | 19.9 | 0.01 |
| Antiinflammatory and antirheumatic products | 56.5 | 57.6 | -0.02 | 56.2 | 56.1 | 0.00 | 57.6 | 57.2 | 0.01 |
| Antineoplastic agents | 5.0 | 4.4 | 0.03 | 4.8 | 5.0 | -0.01 | 4.6 | 4.6 | 0.00 |
| Antipsoriatics | 0.5 | 0.5 | 0.00 | 0.5 | 0.5 | 0.00 | 0.5 | 0.5 | 0.00 |
| Antithrombotic agents | 36.1 | 37.8 | -0.03 | 36.0 | 35.9 | 0.00 | 37.9 | 37.2 | 0.01 |
| Beta blocking agents | 31.2 | 32.1 | -0.02 | 31.0 | 30.8 | 0.00 | 32.1 | 31.7 | 0.01 |
| Calcium channel blockers | 21.2 | 20.4 | 0.02 | 20.8 | 21.1 | 0.00 | 20.9 | 20.7 | 0.01 |
| Diuretics | 42.4 | 42.8 | -0.01 | 42.2 | 42.1 | 0.00 | 43.2 | 42.7 | 0.01 |
| Drugs for acid related disorders | 43.7 | 42.7 | 0.02 | 43.0 | 43.2 | 0.00 | 43.1 | 43.0 | 0.00 |
| Drugs for obstructive airway diseases | 47.4 | 45.2 | 0.04 | 46.6 | 46.9 | -0.01 | 46.1 | 46.0 | 0.00 |
| Immunosuppressants | 3.2 | 2.8 | 0.02 | 3.1 | 3.1 | 0.00 | 2.9 | 2.9 | 0.00 |
| Lipid modifying agents | 67.0 | 65.2 | 0.04 | 66.3 | 66.5 | 0.00 | 66.1 | 65.8 | 0.01 |
| Opioids | 30.0 | 30.6 | -0.01 | 29.7 | 29.7 | 0.00 | 30.8 | 30.4 | 0.01 |
| Psycholeptics | 32.5 | 31.3 | 0.03 | 32.0 | 32.2 | 0.00 | 32.0 | 31.7 | 0.01 |
| Psychostimulants | 3.9 | 3.3 | 0.03 | 3.7 | 3.8 | 0.00 | 3.5 | 3.5 | 0.00 |

### 2.6.8 VA

**Supplementary Table 198:** Baseline patient characteristics for DPP4I (female) (T) and SU (female) (C) new-users in the VA data source. We report proportion of initiators satisfying selected base-line characteristics and the standardized difference of population proportions (StdDiff) before and after propensity score adjustment. Less extreme StdDiffs through matching and stratification suggest improved balance between patient cohorts through adjustment.

| | Before adjustment | | | After matching | | | After stratification | | |
|---|---|---|---|---|---|---|---|---|---|
| Characteristic | T (%) | C (%) | StdDiff | T (%) | C (%) | StdDiff | T (%) | C (%) | StdDiff |
| **Age group** | | | | | | | | | |
| 20 - 24 | <0.5 | 0.1 | -0.01 | <0.2 | <0.1 | -0.02 | <0.2 | <0.1 | -0.02 |
| 25 - 29 | <0.5 | 0.5 | -0.04 | <0.5 | 0.5 | -0.04 | <0.2 | 0.5 | -0.06 |
| 30 - 34 | 1.4 | 1.7 | -0.03 | 1.4 | 1.7 | -0.03 | 3.9 | 1.8 | 0.13 |
| 35 - 39 | 4.1 | 4.1 | 0.00 | 4.0 | 4.8 | -0.04 | 2.7 | 4.4 | -0.09 |
| 40 - 44 | 6.9 | 7.5 | -0.02 | 7.0 | 6.6 | 0.01 | 5.3 | 7.2 | -0.08 |
| 45 - 49 | 10.7 | 12.4 | -0.05 | 9.8 | 10.3 | -0.02 | 5.9 | 11.5 | -0.20 |
| 50 - 54 | 16.5 | 19.6 | -0.08 | 15.9 | 16.5 | -0.02 | 14.9 | 18.9 | -0.11 |
| 55 - 59 | 21.4 | 21.3 | 0.00 | 21.4 | 20.9 | 0.01 | 26.4 | 21.7 | 0.11 |
| 60 - 64 | 19.9 | 17.7 | 0.06 | 21.0 | 19.2 | 0.04 | 25.9 | 18.8 | 0.17 |
| 65 - 69 | 11.1 | 8.1 | 0.10 | 11.8 | 12.3 | -0.01 | 8.7 | 9.2 | -0.02 |
| 70 - 74 | 4.6 | 3.3 | 0.07 | 4.3 | 4.5 | -0.01 | 3.5 | 3.4 | 0.00 |
| 75 - 79 | 1.5 | 1.6 | -0.01 | 1.4 | 1.6 | -0.02 | 1.3 | 1.4 | 0.00 |
| 80 - 84 | 1.1 | 1.3 | -0.02 | 1.1 | 0.6 | 0.06 | 0.5 | 0.6 | -0.01 |
| 85 - 89 | <0.5 | 0.7 | -0.05 | <0.5 | 0.3 | 0.01 | 0.2 | 0.5 | -0.05 |
| 90 - 94 | <0.5 | 0.2 | 0.00 | 0.3 | 0.2 | 0.03 | 0.3 | 0.2 | 0.03 |
| 95 - 99 | <0.5 | <0.1 | 0.01 | <0.2 | <0.1 | 0.02 | <0.2 | <0.1 | 0.02 |
| **Race** | | | | | | | | | |
| Asian | 1.9 | 1.1 | 0.06 | 1.8 | 1.4 | 0.04 | 1.8 | 1.4 | 0.04 |
| Black or African American | 37.2 | 31.3 | 0.12 | 36.9 | 38.0 | -0.02 | 26.2 | 33.8 | -0.17 |
| White | 52.0 | 57.1 | -0.10 | 52.5 | 51.3 | 0.02 | 65.3 | 55.5 | 0.20 |
| Unknown | 5.7 | 8.1 | -0.09 | 5.5 | 6.2 | -0.03 | 3.6 | 6.9 | -0.15 |
| Native Hawaiian or Other Pacific Islander | 2.1 | 1.4 | 0.06 | 2.1 | 1.6 | 0.04 | 2.3 | 1.4 | 0.06 |
| American Indian or Alaska Native | 1.1 | 0.9 | 0.01 | 1.1 | 1.1 | 0.00 | 0.7 | 1.0 | -0.02 |
| **Ethnicity** | | | | | | | | | |
| Hispanic or Latino | 5.7 | 6.0 | -0.01 | 5.9 | 5.2 | 0.03 | 6.7 | 5.9 | 0.03 |
| Not Hispanic or Latino | 92.7 | 90.5 | 0.08 | 92.7 | 92.5 | 0.01 | 92.6 | 91.5 | 0.04 |
| **Medical history: General** | | | | | | | | | |
| Acute respiratory disease | 16.3 | 18.2 | -0.05 | 16.6 | 16.3 | 0.01 | 24.0 | 17.8 | 0.15 |
| Attention deficit hyperactivity disorder | 1.3 | 0.8 | 0.05 | 1.3 | 1.1 | 0.02 | 4.0 | 0.9 | 0.20 |
| Chronic liver disease | 1.5 | 2.4 | -0.07 | 1.7 | 1.5 | 0.01 | 2.0 | 2.2 | -0.02 |
| Chronic obstructive lung disease | 7.0 | 7.1 | 0.00 | 6.7 | 7.4 | -0.03 | 9.5 | 7.1 | 0.09 |
| Crohn's disease | <0.5 | 0.3 | -0.01 | <0.5 | 0.2 | -0.01 | <0.2 | 0.3 | -0.05 |
| Dementia | 0.6 | 0.5 | 0.01 | 0.7 | 0.5 | 0.02 | 0.3 | 0.4 | -0.02 |
| Depressive disorder | 41.3 | 39.0 | 0.05 | 39.5 | 39.9 | -0.01 | 40.9 | 40.0 | 0.02 |
| Gastroesophageal reflux disease | 25.8 | 23.9 | 0.04 | 24.0 | 25.0 | -0.02 | 22.9 | 24.6 | -0.04 |
| Gastrointestinal hemorrhage | 1.3 | 1.5 | -0.01 | 1.2 | 1.3 | -0.01 | 1.2 | 1.5 | -0.02 |
| Human immunodeficiency virus infection | <0.5 | 0.2 | 0.01 | <0.5 | 0.2 | -0.01 | <0.2 | 0.2 | -0.03 |
| Hyperlipidemia | 65.0 | 68.9 | -0.08 | 63.7 | 62.9 | 0.02 | 62.2 | 68.6 | -0.13 |
| Hypertensive disorder | 63.6 | 69.7 | -0.13 | 63.8 | 63.5 | 0.01 | 62.2 | 68.3 | -0.13 |
| Lesion of liver | 1.0 | 0.8 | 0.02 | 1.2 | 1.1 | 0.01 | 0.8 | 0.9 | -0.01 |
| Obesity | 41.1 | 42.4 | -0.03 | 40.2 | 40.8 | -0.01 | 32.1 | 43.0 | -0.23 |
| Osteoarthritis | 22.2 | 25.3 | -0.07 | 21.6 | 21.7 | 0.00 | 18.7 | 24.9 | -0.15 |
| Pneumonia | 1.4 | 1.6 | -0.02 | 1.3 | 1.5 | -0.02 | 3.5 | 1.6 | 0.12 |
| Psoriasis | 1.5 | 1.6 | 0.00 | 1.5 | 1.6 | -0.01 | 3.7 | 1.6 | 0.13 |
| Renal impairment | 4.5 | 3.5 | 0.05 | 4.4 | 4.6 | -0.01 | 3.7 | 4.0 | -0.01 |
| Rheumatoid arthritis | 1.5 | 1.5 | 0.00 | 1.4 | 1.7 | -0.02 | 0.7 | 1.5 | -0.08 |
| Schizophrenia | 1.3 | 1.8 | -0.04 | 1.1 | 1.3 | -0.02 | 0.5 | 1.5 | -0.10 |
| Ulcerative colitis | <0.5 | 0.3 | 0.02 | <0.5 | 0.3 | 0.01 | 3.4 | 0.3 | 0.23 |
| Urinary tract infectious disease | 6.6 | 8.2 | -0.06 | 10.4 | 7.8 | 0.09 | 10.4 | 7.8 | 0.09 |
| Viral hepatitis C | 0.6 | 1.2 | -0.07 | 0.7 | 0.8 | -0.01 | 0.6 | 1.0 | -0.05 |
| Visual system disorder | 44.4 | 42.1 | 0.05 | 43.7 | 43.4 | 0.01 | 40.0 | 42.8 | -0.06 |
| **Medical history: Cardiovascular disease** | | | | | | | | | |
| Atrial fibrillation | 1.8 | 1.5 | 0.02 | 1.6 | 1.9 | -0.02 | 1.5 | 1.6 | -0.01 |
| Cerebrovascular disease | 2.1 | 2.5 | -0.03 | 2.0 | 1.9 | 0.01 | 1.4 | 2.3 | -0.06 |
| Coronary arteriosclerosis | 3.4 | 4.6 | -0.06 | 3.6 | 3.9 | -0.02 | 2.6 | 4.1 | -0.08 |
| Heart disease | 13.2 | 14.6 | -0.04 | 13.1 | 14.1 | -0.03 | 14.4 | 13.9 | 0.01 |
| Heart failure | 1.9 | 2.2 | -0.02 | 2.0 | 2.4 | -0.02 | 6.5 | 2.2 | 0.21 |
| Ischemic heart disease | 2.7 | 4.3 | -0.09 | 2.8 | 2.4 | 0.03 | 8.2 | 3.5 | 0.20 |

| (Continued) | Before ajustment | | | After matching | | | After stratification | | |
|---|---|---|---|---|---|---|---|---|---|
| Characteristic | T (%) | C (%) | StdDiff | T (%) | C (%) | StdDiff | T (%) | C (%) | StdDiff |
| Peripheral vascular disease | 1.2 | 1.8 | -0.05 | 1.3 | 1.4 | -0.01 | 1.0 | 1.7 | -0.06 |
| Pulmonary embolism | <0.5 | 0.5 | -0.03 | <0.5 | 0.7 | -0.09 | <0.2 | 0.5 | -0.09 |
| Venous thrombosis | 0.6 | 0.7 | -0.01 | 0.6 | 0.7 | -0.02 | 0.5 | 0.7 | -0.04 |
| **Medical history: Neoplasms** | | | | | | | | | |
| Hematologic neoplasm | 0.6 | 0.5 | 0.02 | 0.6 | 0.6 | 0.00 | 0.5 | 0.5 | 0.00 |
| Malignant lymphoma | 0.6 | 0.2 | 0.05 | <0.5 | 0.3 | 0.04 | 0.2 | 0.3 | -0.01 |
| Malignant neoplastic disease | 5.5 | 5.8 | -0.01 | 5.1 | 5.4 | -0.01 | 6.8 | 5.7 | 0.05 |
| Malignant tumor of breast | 2.0 | 2.6 | -0.04 | 1.9 | 1.9 | 0.00 | 4.0 | 2.4 | 0.09 |
| Malignant tumor of colon | <0.5 | 0.2 | -0.04 | <0.5 | 0.2 | -0.02 | <0.2 | 0.2 | -0.05 |
| Malignant tumor of lung | <0.5 | 0.2 | -0.02 | <0.5 | 0.3 | -0.04 | <0.2 | 0.2 | -0.01 |
| Malignant tumor of urinary bladder | <0.5 | 0.1 | 0.00 | <0.5 | 0.1 | 0.01 | <0.2 | 0.1 | -0.01 |
| **Medication use** | | | | | | | | | |
| Agents acting on the renin-angiotensin system | 51.1 | 59.6 | -0.17 | 51.5 | 50.6 | 0.02 | 53.2 | 57.2 | -0.08 |
| Antibacterials for systemic use | 39.6 | 45.5 | -0.12 | 39.7 | 39.7 | 0.00 | 55.6 | 44.3 | 0.23 |
| Antidepressants | 53.6 | 52.3 | 0.03 | 52.6 | 53.0 | -0.01 | 59.3 | 52.8 | 0.13 |
| Antiepileptics | 34.0 | 28.6 | 0.12 | 32.6 | 33.8 | -0.02 | 33.3 | 30.9 | 0.05 |
| Antiinflammatory and antirheumatic products | 57.9 | 56.2 | 0.04 | 56.9 | 58.8 | -0.04 | 59.7 | 56.7 | 0.06 |
| Antineoplastic agents | 5.8 | 4.0 | 0.08 | 5.3 | 5.8 | -0.02 | 3.0 | 4.4 | -0.07 |
| Antipsoriatics | 1.3 | 0.7 | 0.06 | 1.0 | 0.5 | 0.06 | 0.5 | 0.7 | -0.02 |
| Antithrombotic agents | 25.0 | 26.2 | -0.03 | 24.7 | 24.8 | 0.00 | 25.9 | 25.8 | 0.00 |
| Beta blocking agents | 26.8 | 29.8 | -0.07 | 26.8 | 26.1 | 0.02 | 32.3 | 28.8 | 0.07 |
| Calcium channel blockers | 23.1 | 22.3 | 0.02 | 22.4 | 22.9 | -0.01 | 21.9 | 22.3 | -0.01 |
| Diuretics | 36.7 | 43.6 | -0.14 | 36.9 | 36.2 | 0.01 | 48.4 | 42.1 | 0.13 |
| Drugs for acid related disorders | 49.3 | 52.3 | -0.06 | 48.0 | 48.9 | -0.02 | 60.4 | 51.3 | 0.18 |
| Drugs for obstructive airway diseases | 51.0 | 44.5 | 0.13 | 48.6 | 50.1 | -0.03 | 57.7 | 46.2 | 0.23 |
| Immunosuppressants | 4.7 | 2.8 | 0.10 | 4.1 | 4.2 | -0.01 | 2.1 | 3.2 | -0.07 |
| Lipid modifying agents | 71.3 | 72.6 | -0.03 | 70.3 | 70.1 | 0.00 | 74.6 | 73.1 | 0.04 |
| Opioids | 19.9 | 27.7 | -0.18 | 19.2 | 18.5 | 0.02 | 41.5 | 26.3 | 0.32 |
| Psycholeptics | 36.5 | 34.1 | 0.05 | 35.2 | 36.0 | -0.01 | 38.0 | 35.2 | 0.06 |
| Psychostimulants | 4.3 | 4.1 | 0.01 | 4.3 | 4.4 | 0.00 | 6.3 | 4.1 | 0.10 |

##### 2.6.9 SIDIAP

**Supplementary Table 199:** Baseline patient characteristics for DPP4I (female) (T) and SU (female) (C) new-users in the SIDIAP data source. We report proportion of initiators satisfying selected base-line characteristics and the standardized difference of population proportions (StdDiff) before and after propensity score adjustment. Less extreme StdDiffs through matching and stratification suggest improved balance between patient cohorts through adjustment.

| | Before adjustment | | | After matching | | | After stratification | | |
|---|---|---|---|---|---|---|---|---|---|
| Characteristic | T (%) | C (%) | StdDiff | T (%) | C (%) | StdDiff | T (%) | C (%) | StdDiff |
| **Age group** | | | | | | | | | |
| 25 - 29 | 0.1 | 0.1 | 0.00 | 0.2 | 0.2 | 0.01 | 0.2 | 0.2 | 0.00 |
| 30 - 34 | 0.3 | 0.3 | 0.01 | 0.5 | 0.3 | 0.03 | 0.4 | 0.2 | 0.02 |
| 35 - 39 | 0.7 | 0.8 | -0.01 | 0.9 | 0.8 | 0.02 | 0.7 | 0.6 | 0.02 |
| 40 - 44 | 2.0 | 2.5 | -0.03 | 2.5 | 2.8 | -0.02 | 2.2 | 2.9 | -0.05 |
| 45 - 49 | 3.7 | 5.1 | -0.07 | 4.1 | 4.8 | -0.03 | 4.1 | 4.8 | -0.03 |
| 50 - 54 | 7.1 | 8.4 | -0.05 | 8.6 | 8.4 | 0.01 | 7.5 | 8.7 | -0.05 |
| 55 - 59 | 10.2 | 11.2 | -0.04 | 12.0 | 11.6 | 0.01 | 10.6 | 10.4 | 0.01 |
| 60 - 64 | 12.4 | 14.7 | -0.07 | 14.2 | 14.1 | 0.00 | 13.0 | 13.5 | -0.02 |
| 65 - 69 | 14.4 | 15.8 | -0.04 | 15.7 | 15.8 | 0.00 | 14.9 | 15.9 | -0.03 |
| 70 - 74 | 15.4 | 14.8 | 0.02 | 14.4 | 14.7 | -0.01 | 15.0 | 17.0 | -0.05 |
| 75 - 79 | 13.3 | 12.6 | 0.02 | 12.4 | 12.3 | 0.00 | 13.1 | 11.1 | 0.06 |
| 80 - 84 | 10.9 | 8.3 | 0.09 | 7.7 | 8.4 | -0.03 | 9.9 | 8.7 | 0.04 |
| 85 - 89 | 6.5 | 4.2 | 0.10 | 4.4 | 4.3 | 0.01 | 5.8 | 4.1 | 0.08 |
| 90 - 94 | 2.4 | 1.1 | 0.10 | 1.2 | 1.2 | 0.00 | 2.1 | 1.6 | 0.04 |
| 95 - 99 | 0.4 | 0.2 | 0.05 | 0.3 | 0.2 | 0.03 | 0.4 | 0.3 | 0.02 |
| **Medical history: General** | | | | | | | | | |
| Acute respiratory disease | 14.4 | 11.7 | 0.08 | 12.6 | 12.7 | 0.00 | 13.9 | 13.2 | 0.02 |
| Chronic liver disease | 0.4 | 0.4 | 0.00 | 0.5 | 0.3 | 0.02 | 0.4 | 0.3 | 0.03 |
| Chronic obstructive lung disease | 0.8 | 0.7 | 0.01 | 1.0 | 0.7 | 0.03 | 0.8 | 0.5 | 0.04 |
| Dementia | 0.8 | 0.9 | 0.00 | 0.8 | 0.8 | 0.00 | 0.8 | 0.8 | 0.00 |
| Depressive disorder | 3.4 | 4.6 | -0.06 | 4.6 | 4.7 | 0.00 | 3.8 | 3.7 | 0.00 |
| Gastroesophageal reflux disease | 1.5 | 1.0 | 0.05 | 0.9 | 1.2 | -0.03 | 1.3 | 1.6 | -0.03 |
| Gastrointestinal hemorrhage | 1.4 | 1.0 | 0.04 | 1.3 | 1.2 | 0.01 | 1.3 | 1.0 | 0.03 |
| Hyperlipidemia | 3.2 | 4.2 | -0.05 | 4.1 | 4.3 | -0.01 | 3.5 | 3.8 | -0.02 |
| Hypertensive disorder | 3.8 | 5.7 | -0.09 | 5.2 | 5.1 | 0.01 | 4.2 | 3.9 | 0.01 |
| Lesion of liver | 0.2 | 0.2 | 0.00 | 0.3 | 0.2 | 0.02 | 0.2 | 0.2 | 0.02 |
| Obesity | 8.3 | 9.4 | -0.04 | 9.8 | 9.2 | 0.02 | 8.7 | 8.3 | 0.01 |
| Osteoarthritis | 6.6 | 7.5 | -0.04 | 7.4 | 7.5 | -0.01 | 6.8 | 5.8 | 0.04 |
| Pneumonia | 1.4 | 1.2 | 0.02 | 1.1 | 1.2 | -0.01 | 1.3 | 1.2 | 0.01 |
| Psoriasis | 0.3 | 0.4 | 0.00 | 0.4 | 0.3 | 0.01 | 0.3 | 0.3 | 0.01 |
| Renal impairment | 4.2 | 1.9 | 0.14 | 2.3 | 2.4 | 0.00 | 3.6 | 2.5 | 0.07 |
| Rheumatoid arthritis | 0.1 | 0.2 | -0.02 | 0.1 | 0.2 | -0.02 | 0.1 | 0.1 | -0.01 |
| Schizophrenia | 0.1 | 0.1 | -0.01 | <0.1 | <0.1 | 0.00 | 0.1 | <0.1 | 0.01 |
| Urinary tract infectious disease | 10.6 | 5.7 | 0.18 | 7.2 | 7.2 | 0.00 | 9.4 | 9.0 | 0.01 |
| Viral hepatitis C | 0.3 | 0.1 | 0.03 | 0.3 | 0.1 | 0.04 | 0.3 | 0.1 | 0.04 |
| Visual system disorder | 18.0 | 17.3 | 0.02 | 17.8 | 18.4 | -0.02 | 17.9 | 17.7 | 0.00 |
| **Medical history: Cardiovascular disease** | | | | | | | | | |
| Atrial fibrillation | 1.7 | 1.5 | 0.01 | 1.4 | 1.4 | 0.00 | 1.6 | 1.2 | 0.04 |
| Cerebrovascular disease | 1.2 | 1.3 | -0.01 | 1.2 | 1.3 | 0.00 | 1.2 | 1.2 | -0.01 |
| Heart disease | 7.8 | 7.4 | 0.01 | 7.0 | 7.4 | -0.01 | 7.5 | 6.7 | 0.03 |
| Heart failure | 2.0 | 2.0 | 0.00 | 1.6 | 2.0 | -0.03 | 1.9 | 1.4 | 0.04 |
| Ischemic heart disease | 1.5 | 1.7 | -0.01 | 1.5 | 1.8 | -0.02 | 1.5 | 1.5 | 0.00 |
| Peripheral vascular disease | 0.5 | 0.7 | -0.03 | 0.3 | 0.7 | -0.06 | 0.4 | 0.8 | -0.05 |
| Pulmonary embolism | 0.3 | 0.2 | 0.02 | 0.2 | 0.2 | -0.01 | 0.3 | 0.2 | 0.01 |
| Venous thrombosis | 1.2 | 1.2 | 0.00 | 1.2 | 1.3 | -0.01 | 1.2 | 1.1 | 0.01 |
| **Medical history: Neoplasms** | | | | | | | | | |
| Hematologic neoplasm | 0.1 | 0.1 | 0.01 | <0.1 | <0.1 | 0.02 | 0.1 | <0.1 | 0.03 |
| Malignant lymphoma | 0.1 | <0.1 | 0.00 | <0.1 | <0.1 | 0.01 | 0.1 | <0.1 | 0.02 |
| Malignant neoplasm of anorectum | 0.1 | 0.1 | -0.01 | 0.1 | 0.1 | 0.00 | 0.1 | <0.1 | 0.01 |
| Malignant neoplastic disease | 2.8 | 2.4 | 0.02 | 2.7 | 2.5 | 0.01 | 2.8 | 2.3 | 0.03 |
| Malignant tumor of breast | 0.6 | 0.8 | -0.02 | 0.8 | 0.8 | 0.00 | 0.6 | 0.7 | 0.00 |
| Malignant tumor of colon | 0.3 | 0.2 | 0.02 | 0.3 | 0.2 | 0.02 | 0.3 | 0.2 | 0.01 |
| **Medication use** | | | | | | | | | |
| Agents acting on the renin-angiotensin system | 61.7 | 55.9 | 0.12 | 57.8 | 58.0 | 0.00 | 60.1 | 58.6 | 0.03 |
| Antibacterials for systemic use | 43.4 | 35.6 | 0.16 | 38.6 | 39.5 | -0.02 | 41.8 | 41.6 | 0.00 |
| Antidepressants | 33.0 | 28.4 | 0.10 | 29.3 | 30.2 | -0.02 | 32.0 | 31.1 | 0.02 |
| Antiepileptics | 12.3 | 8.2 | 0.14 | 9.9 | 9.5 | 0.01 | 11.5 | 12.7 | -0.04 |
| Antiinflammatory and antirheumatic products | 53.6 | 58.3 | -0.10 | 59.3 | 59.1 | 0.00 | 55.3 | 54.0 | 0.03 |

| (Continued) | Before ajustment | | | After matching | | | After stratification | | |
|---|---|---|---|---|---|---|---|---|---|
| Characteristic | T (%) | C (%) | StdDiff | T (%) | C (%) | StdDiff | T (%) | C (%) | StdDiff |
| Antineoplastic agents | 2.7 | 2.0 | 0.04 | 2.6 | 2.3 | 0.01 | 2.6 | 2.7 | -0.01 |
| Antipsoriatics | 1.7 | 0.9 | 0.07 | 1.4 | 1.0 | 0.03 | 1.6 | 1.4 | 0.02 |
| Antithrombotic agents | 31.5 | 29.3 | 0.05 | 29.0 | 30.0 | -0.02 | 30.8 | 28.4 | 0.05 |
| Beta blocking agents | 26.5 | 22.4 | 0.10 | 22.3 | 23.0 | -0.02 | 25.4 | 23.7 | 0.04 |
| Calcium channel blockers | 23.8 | 19.0 | 0.12 | 20.6 | 20.6 | 0.00 | 22.5 | 22.2 | 0.01 |
| Diuretics | 52.1 | 48.2 | 0.08 | 48.7 | 49.6 | -0.02 | 51.0 | 49.6 | 0.03 |
| Drugs for acid related disorders | 61.8 | 57.6 | 0.09 | 60.3 | 60.6 | -0.01 | 60.9 | 59.2 | 0.04 |
| Drugs for obstructive airway diseases | 28.3 | 23.3 | 0.12 | 24.5 | 25.2 | -0.01 | 27.3 | 26.7 | 0.01 |
| Immunosuppressants | 1.5 | 0.9 | 0.05 | 0.8 | 1.1 | -0.03 | 1.3 | 1.2 | 0.01 |
| Lipid modifying agents | 61.4 | 54.1 | 0.15 | 58.2 | 58.7 | -0.01 | 59.7 | 59.5 | 0.00 |
| Opioids | 25.2 | 19.7 | 0.13 | 24.0 | 23.6 | 0.01 | 24.0 | 23.6 | 0.01 |
| Psycholeptics | 43.9 | 40.1 | 0.08 | 40.9 | 42.0 | -0.02 | 40.9 | 42.0 | -0.02 |
| Psychostimulants | 2.8 | 3.2 | -0.02 | 3.3 | 3.2 | 0.00 | 3.0 | 2.4 | 0.04 |

### 3 Cohort balance and systematic error diagnostics across data sources

#### 3.1 GLP1RA (female) vs SGLT2I (female)

##### 3.1.1 CCAE

**Supplementary Figure 1:** Preference score distributions, propensity score stratified (middle column) and matched (right column) covariate balance and negative control error distributions (bottom row) for GLP1RA (female) vs SGLT2I (female) in the CCAE data source.

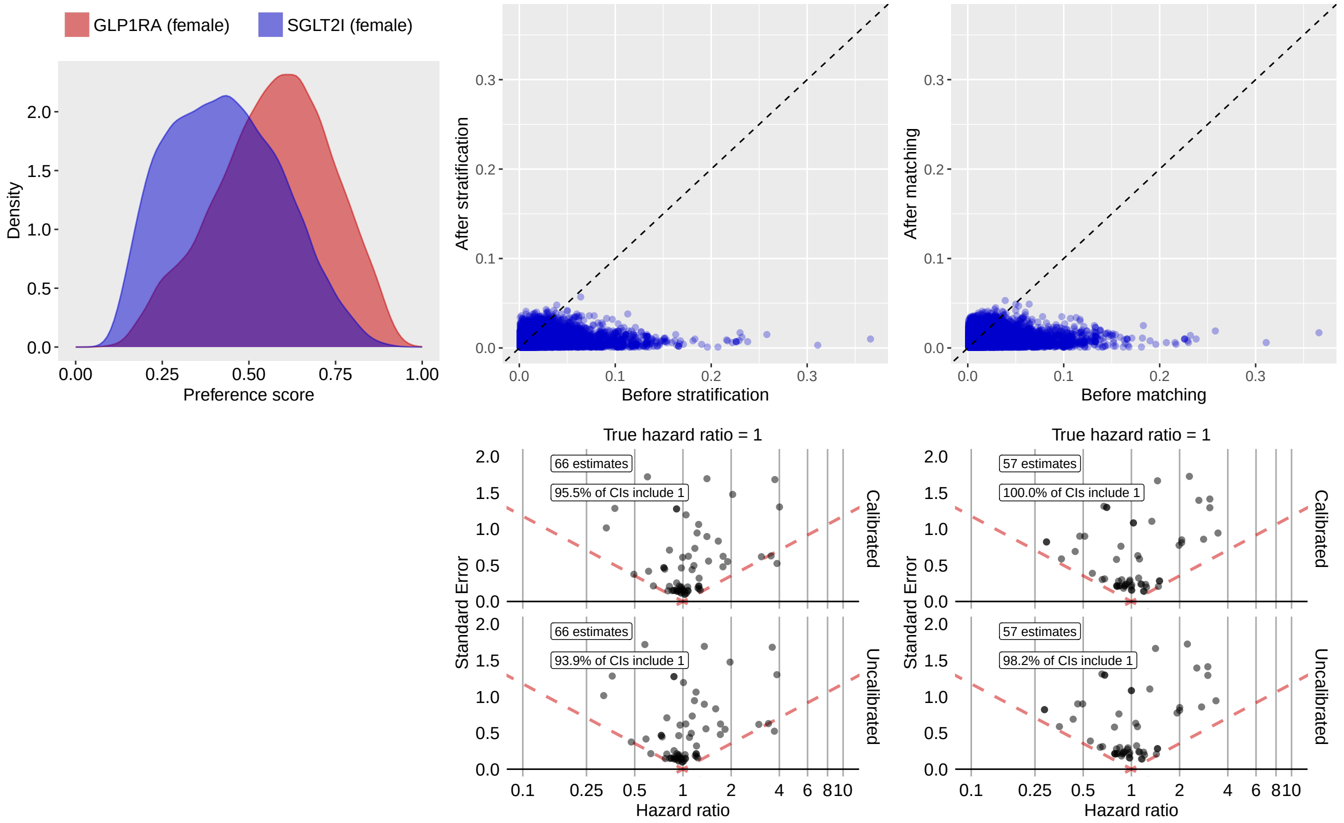

**Supplementary Table 200:** Most extreme patient characteristics balance diagnostics after stratification for GLP1RA (female) vs SGLT2I (female) in the CCAE data source.

| | Before stratification | | | After stratification | | |
|---|---|---|---|---|---|---|
| Characteristic (total count = 11726) | T (%) | C (%) | SDf | T (%) | C (%) | SDf |
| drug_era group during day -365 through 0 days relative to index: sacubitril | 0.1 | 0.3 | -0.06 | 0.1 | 0.3 | -0.06 |
| condition_era group during day -365 through 0 days relative to index: Pancreatitis | 0.2 | 0.5 | -0.04 | 0.2 | 0.5 | -0.05 |
| condition_era group during day -30 through 0 days relative to index: Thrombophilia | 0.1 | 0.2 | -0.03 | 0.1 | 0.2 | -0.04 |
| measurement within normal range during day -365 through 0 days relative to index: Glucose [Mass/volume] in Serum or Plasma | 0.6 | 0.3 | 0.05 | 0.5 | 0.2 | 0.04 |
| observation during day -365 through 0 days relative to index: Inadequate immune status | 0.3 | 0.6 | -0.04 | 0.4 | 0.7 | -0.04 |
| condition_era group during day -30 through 0 days relative to index: Hirsutism | 0.4 | 0.1 | 0.05 | 0.4 | 0.2 | 0.04 |

| (Continued) | T (%) | C (%) | SDf | T (%) | C (%) | SDf |
|---|---|---|---|---|---|---|
| procedure_occurrence during day -365 through 0 days relative to index: Duplex scan of extracranial arteries; unilateral or limited study | 0.2 | 0.1 | 0.03 | 0.2 | 0.1 | 0.04 |
| measurement below normal range during day -365 through 0 days relative to index: Protein [Mass/volume] in Serum or Plasma | 0.2 | 0.1 | 0.01 | 0.3 | 0.1 | 0.04 |
| procedure_occurrence during day -365 through 0 days relative to index: Endovenous ablation therapy of incompetent vein, extremity, inclusive of all imaging guidance and monitoring, percutaneous, radiofrequency; first vein treated | 0.2 | 0.1 | 0.03 | 0.3 | 0.1 | 0.04 |
| procedure_occurrence during day -365 through 0 days relative to index: Destruction (eg, laser surgery, electrosurgery, cryosurgery, chemosurgery, surgical curettement), of benign lesions other than skin tags or cutaneous vascular proliferative lesions; 15 or more lesions | 0.1 | 0.2 | -0.02 | 0.1 | 0.2 | -0.04 |

**Supplementary Table 201:** Most extreme patient characteristics balance diagnostics after matching for GLP1RA (female) vs SGLT2I (female) in the CCAE data source.

| | Before matching | | | After matching | | |
|---|---|---|---|---|---|---|
| Characteristic (total count = 11622) | T (%) | C (%) | SDf | T (%) | C (%) | SDf |
| condition_era group during day -365 through 0 days relative to index: Pancreatitis | 0.2 | 0.5 | -0.04 | 0.2 | 0.5 | -0.05 |
| drug_era group during day -365 through 0 days relative to index: sacubitril | 0.1 | 0.3 | -0.06 | 0.1 | 0.3 | -0.05 |
| condition_era group during day -30 through 0 days relative to index: Hirsutism | 0.4 | 0.1 | 0.05 | 0.4 | 0.1 | 0.05 |
| observation during day -365 through 0 days relative to index: History of cardiovascular surgery | 0.1 | 0.4 | -0.05 | 0.2 | 0.4 | -0.05 |
| condition_era group during day -365 through 0 days relative to index: Acute pancreatitis | 0.2 | 0.4 | -0.03 | 0.2 | 0.4 | -0.04 |
| condition_era group during day -30 through 0 days relative to index: Thrombophilia | 0.1 | 0.2 | -0.03 | 0.1 | 0.2 | -0.04 |
| measurement within normal range during day -365 through 0 days relative to index: Glucose [Mass/volume] in Serum or Plasma | 0.6 | 0.3 | 0.05 | 0.5 | 0.2 | 0.04 |
| observation during day -365 through 0 days relative to index: Inadequate immune status | 0.3 | 0.6 | -0.04 | 0.4 | 0.7 | -0.04 |
| procedure_occurrence during day -365 through 0 days relative to index: Endovenous ablation therapy of incompetent vein, extremity, inclusive of all imaging guidance and monitoring, percutaneous, radiofrequency; first vein treated | 0.2 | 0.1 | 0.03 | 0.2 | 0.1 | 0.04 |
| condition_era group during day -365 through 0 days relative to index: Exacerbation of intermittent asthma | 0.3 | 0.5 | -0.02 | 0.4 | 0.7 | -0.04 |

##### 3.1.2 ODOD

**Supplementary Figure 2:** Preference score distributions, propensity score stratified (middle column) and matched (right column) covariate balance and negative control error distributions (bottom row) for GLP1RA (female) vs SGLT2I (female) in the ODOD data source.

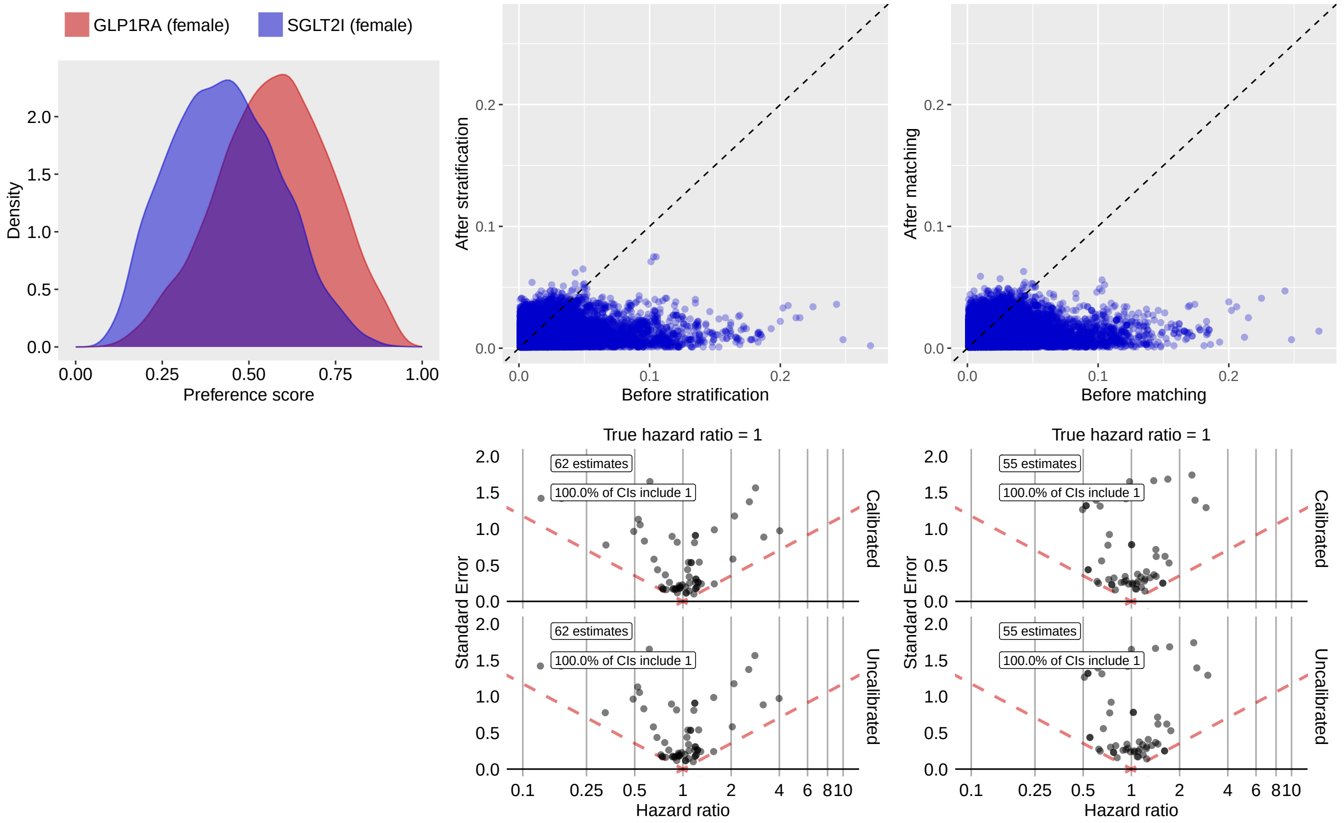

**Supplementary Table 202:** Most extreme patient characteristics balance diagnostics after stratification for GLP1RA (female) vs SGLT2I (female) in the ODOD data source.

| | Before stratification | | | After stratification | | |
|---|---|---|---|---|---|---|
| Characteristic (total count = 16088) | T (%) | C (%) | SDf | T (%) | C (%) | SDf |
| drug_era group during day -365 through 0 days relative to index: sacubitril | 0.2 | 0.9 | -0.10 | 0.2 | 0.7 | -0.07 |
| drug_era group during day 0 through 0 days relative to index: sacubitril | 0.1 | 0.8 | -0.10 | 0.2 | 0.6 | -0.07 |
| drug_era group during day -30 through 0 days relative to index: sacubitril | 0.1 | 0.9 | -0.10 | 0.2 | 0.7 | -0.07 |
| observation during day -365 through 0 days relative to index: Transportation of portable x-ray equipment and personnel to home or nursing home, per trip to facility or location, more than one patient seen | 0.3 | 0.1 | 0.05 | 0.3 | 0.1 | 0.07 |
| observation during day -365 through 0 days relative to index: Set-up portable x-ray equipment | 0.4 | 0.2 | 0.04 | 0.5 | 0.1 | 0.06 |
| drug_era group during day -365 through 0 days relative to index: lansoprazole | 0.4 | 0.5 | -0.01 | 0.2 | 0.5 | -0.05 |
| condition_era group during day -30 through 0 days relative to index: Hypokalemia | 0.6 | 1.1 | -0.05 | 0.6 | 1.1 | -0.05 |

| (Continued) | T (%) | C (%) | SDf | T (%) | C (%) | SDf |
|---|---|---|---|---|---|---|
| condition_era group during day -30 through 0 days relative to index: Arthritis of shoulder region joint | 0.1 | 0.2 | -0.04 | 0.1 | 0.3 | -0.05 |
| condition_era group during day -365 through 0 days relative to index: Pancreatitis | 0.3 | 0.6 | -0.05 | 0.3 | 0.7 | -0.05 |
| condition_era group during day -365 through 0 days relative to index: Lump of lower inner quadrant of breast | 0.3 | 0.5 | -0.03 | 0.3 | 0.6 | -0.05 |

**Supplementary Table 203:** Most extreme patient characteristics balance diagnostics after matching for GLP1RA (female) vs SGLT2I (female) in the ODOD data source.

| | Before matching | | | After matching | | |
|---|---|---|---|---|---|---|
| Characteristic (total count = 16422) | T (%) | C (%) | SDf | T (%) | C (%) | SDf |
| observation during day -365 through 0 days relative to index: Set-up portable x-ray equipment | 0.4 | 0.2 | 0.04 | 0.5 | 0.1 | 0.06 |
| drug_era group during day -365 through 0 days relative to index: lansoprazole | 0.4 | 0.5 | -0.01 | 0.2 | 0.5 | -0.06 |
| condition_era group during day -365 through 0 days relative to index: Lump of lower inner quadrant of breast | 0.3 | 0.5 | -0.03 | 0.3 | 0.7 | -0.06 |
| drug_era group during day 0 through 0 days relative to index: sacubitril | 0.1 | 0.8 | -0.10 | 0.2 | 0.5 | -0.06 |
| drug_era group during day -365 through 0 days relative to index: Progestogens | 2.4 | 1.8 | 0.04 | 1.4 | 2.1 | -0.05 |
| measurement within normal range during day -365 through 0 days relative to index: Fasting glucose [Mass/volume] in Serum or Plasma | 0.3 | 0.1 | 0.05 | 0.3 | 0.1 | 0.05 |
| condition_era group during day -365 through 0 days relative to index: Pancreatitis | 0.3 | 0.6 | -0.05 | 0.3 | 0.7 | -0.05 |
| drug_era group during day -365 through 0 days relative to index: sacubitril | 0.2 | 0.9 | -0.10 | 0.2 | 0.5 | -0.05 |
| condition_era group during day -30 through 0 days relative to index: Arthritis of shoulder region joint | 0.1 | 0.2 | -0.04 | -0.1 | 0.3 | -0.05 |
| observation during day -365 through 0 days relative to index: Influenza immunization was not administered, reason not given | 0.4 | 0.8 | -0.05 | 0.3 | 0.7 | -0.05 |

##### 3.1.3 MDCR

**Supplementary Figure 3:** Preference score distributions, propensity score stratified (middle column) and matched (right column) covariate balance and negative control error distributions (bottom row) for GLP1RA (female) vs SGLT2I (female) in the MDCR data source.

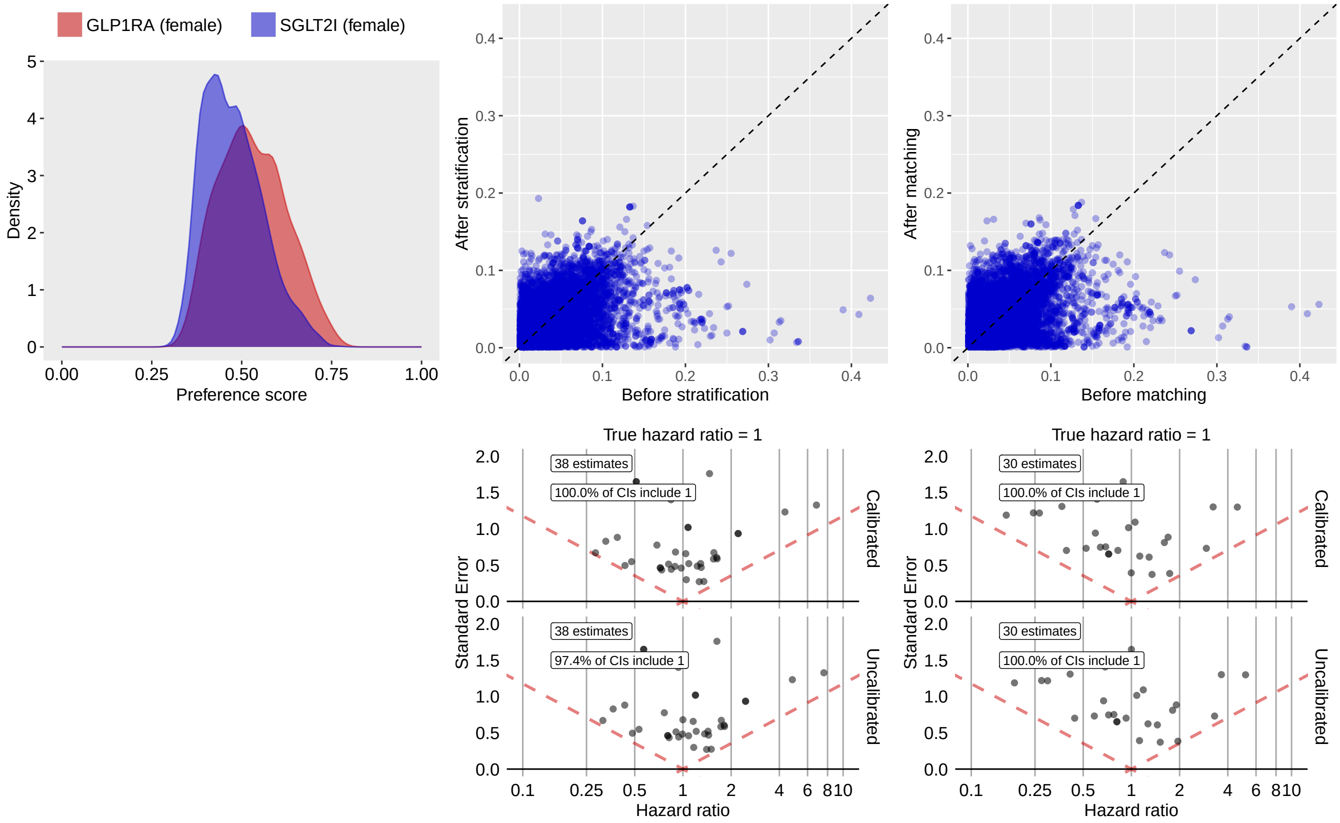

**Supplementary Table 204:** Most extreme patient characteristics balance diagnostics after stratification for GLP1RA (female) vs SGLT2I (female) in the MDCR data source.

| Characteristic (total count = 11909) | Before stratification | | | After stratification | | |
|---|---|---|---|---|---|---|
| | T (%) | C (%) | SDf | T (%) | C (%) | SDf |
| index year: 2021 | 20.1 | 19.1 | 0.02 | 28.7 | 20.4 | 0.19 |
| condition_era group during day -365 through 0 days relative to index: Pain characterized by provoking factor | 1.8 | 4.1 | -0.14 | 1.3 | 4.4 | -0.18 |
| condition_era group during day -365 through 0 days relative to index: Pain characterized by relieving factor | 1.7 | 3.9 | -0.13 | 1.2 | 4.1 | -0.18 |
| condition_era group during day -365 through 0 days relative to index: Pain provoked by exertion | 1.7 | 3.9 | -0.13 | 1.2 | 4.1 | -0.18 |
| condition_era group during day -365 through 0 days relative to index: Pain relief by rest | 1.7 | 3.9 | -0.13 | 1.2 | 4.1 | -0.18 |
| condition_era group during day -365 through 0 days relative to index: Intermittent pain | 1.7 | 3.9 | -0.13 | 1.2 | 4.1 | -0.18 |
| measurement during day -30 through 0 days relative to index: Blood count; complete (CBC), automated (Hgb, Hct, RBC, WBC and platelet count) | 1.2 | 3.1 | -0.13 | 1.0 | 3.4 | -0.17 |
| drug_era group during day -30 through 0 days relative to index: albuterol | 4.1 | 5.7 | -0.08 | 2.7 | 6.1 | -0.16 |

| (Continued) | T (%) | C (%) | SDf | T (%) | C (%) | SDf |
|---|---|---|---|---|---|---|
| drug_era group during day -30 through 0 days relative to index: ADRENERGICS FOR SYSTEMIC USE | 4.1 | 5.7 | -0.08 | 2.7 | 6.1 | -0.16 |
| drug_era group during day -30 through 0 days relative to index: Selective beta-2-adrenoreceptor agonists | 4.1 | 5.7 | -0.08 | 2.7 | 6.1 | -0.16 |

**Supplementary Table 205:** Most extreme patient characteristics balance diagnostics after matching for GLP1RA (female) vs SGLT2I (female) in the MDCR data source.

| | Before matching | | | After matching | | |
|---|---|---|---|---|---|---|
| Characteristic (total count = 10989) | T (%) | C (%) | SDf | T (%) | C (%) | SDf |
| condition_era group during day -365 through 0 days relative to index: Pain characterized by provoking factor | 1.8 | 4.1 | -0.14 | 1.5 | 4.7 | -0.19 |
| condition_era group during day -365 through 0 days relative to index: Pain characterized by relieving factor | 1.7 | 3.9 | -0.13 | 1.4 | 4.4 | -0.18 |
| condition_era group during day -365 through 0 days relative to index: Pain provoked by exertion | 1.7 | 3.9 | -0.13 | 1.4 | 4.4 | -0.18 |
| condition_era group during day -365 through 0 days relative to index: Pain relief by rest | 1.7 | 3.9 | -0.13 | 1.4 | 4.4 | -0.18 |
| condition_era group during day -365 through 0 days relative to index: Intermittent pain | 1.7 | 3.9 | -0.13 | 1.4 | 4.4 | -0.18 |
| condition_era group during day -365 through 0 days relative to index: Prediabetes | 2.9 | 1.1 | 0.13 | 4.2 | 1.3 | 0.18 |
| measurement during day -30 through 0 days relative to index: Blood count; complete (CBC), automated (Hgb, Hct, RBC, WBC and platelet count) | 1.2 | 3.1 | -0.13 | 1.0 | 3.4 | -0.17 |
| condition_era group during day -30 through 0 days relative to index: Vascular disorder of extremity | 0.6 | 1.5 | -0.09 | -0.7 | 1.8 | -0.17 |
| condition_era group during day -365 through 0 days relative to index: Squamous neoplasm of surface epithelium | 0.8 | 2.9 | -0.15 | 0.7 | 2.9 | -0.17 |
| Diabetes Comorbidity Severity Index (DCSI) | 342.3 | 350.8 | -0.03 | 315.4 | 359.7 | -0.17 |

##### 3.1.4 MDCD

**Supplementary Figure 4:** Preference score distributions, propensity score stratified (middle column) and matched (right column) covariate balance and negative control error distributions (bottom row) for GLP1RA (female) vs SGLT2I (female) in the MDCD data source.

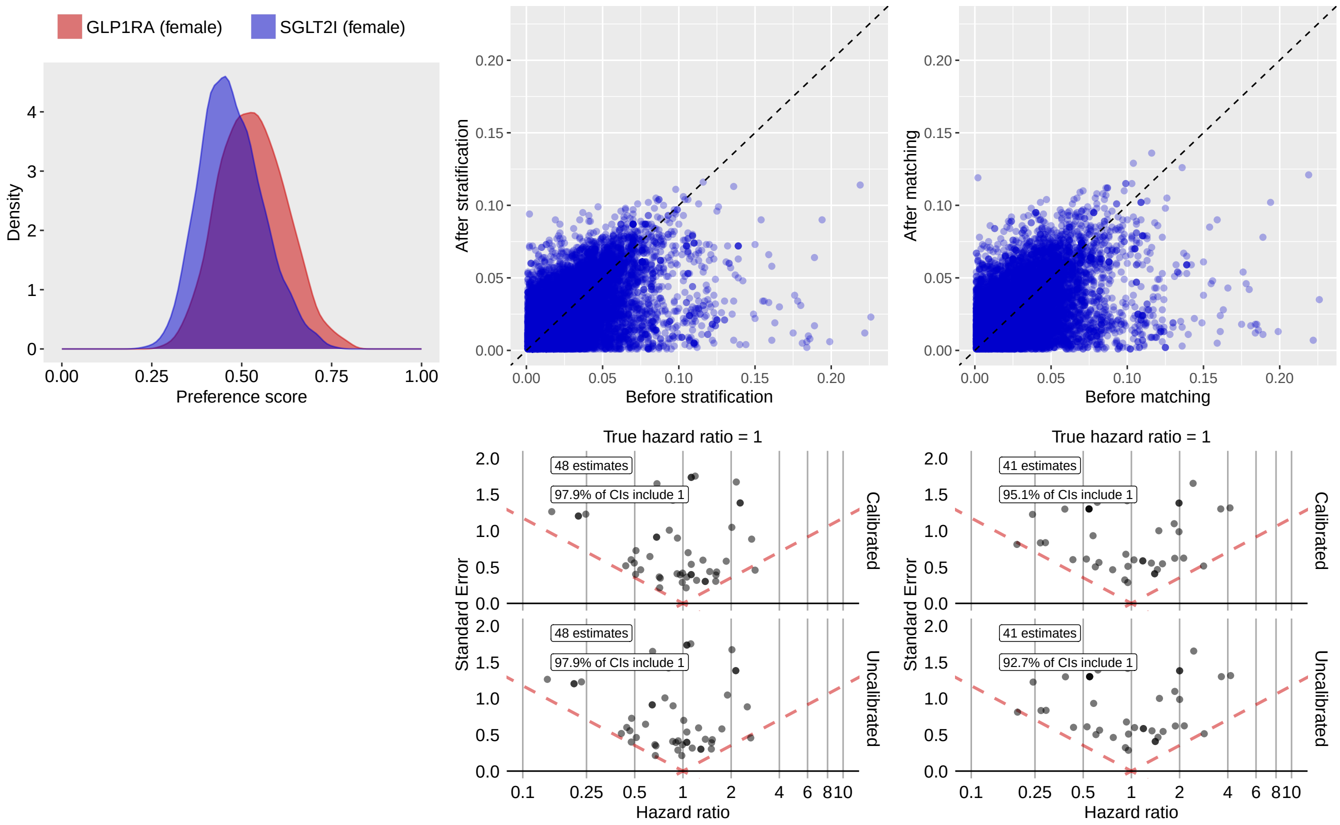

**Supplementary Table 206:** Most extreme patient characteristics balance diagnostics after stratification for GLP1RA (female) vs SGLT2I (female) in the MDCD data source.

| Characteristic (total count = 15721) | Before stratification | | | After stratification | | |
|---|---|---|---|---|---|---|
| | T (%) | C (%) | SDf | T (%) | C (%) | SDf |
| drug_era group during day -365 through 0 days relative to index: liothyronine | 1.0 | -0.2 | 0.12 | 0.9 | -0.2 | 0.12 |
| condition_era group during day -365 through 0 days relative to index: Obesity | 58.2 | 47.3 | 0.22 | 49.3 | 54.9 | -0.11 |
| condition_era group during day -30 through 0 days relative to index: Metabolic syndrome X | 2.4 | 0.7 | 0.14 | 2.0 | 0.7 | 0.11 |
| condition_era group during day -365 through 0 days relative to index: Somatic dysfunction of sacral region | 0.3 | 1.1 | -0.10 | -0.3 | 1.1 | -0.11 |
| condition_era group during day -30 through 0 days relative to index: Intervertebral disc disorder with myelopathy | 1.1 | 0.2 | 0.10 | 1.0 | -0.2 | 0.11 |
| observation during day -30 through 0 days relative to index: Handling and/or conveyance of specimen for transfer from the office to a laboratory | 1.2 | 0.5 | 0.09 | 1.4 | 0.4 | 0.10 |
| drug_era group during day -30 through 0 days relative to index: Sodium | 2.1 | 3.9 | -0.11 | 2.0 | 3.8 | -0.10 |

| (Continued) | T (%) | C (%) | SDf | T (%) | C (%) | SDf |
|---|---|---|---|---|---|---|
| condition_era group during day -365 through 0 days relative to index: Achilles tendinitis | 0.3 | 1.3 | -0.10 | 0.4 | 1.4 | -0.10 |
| drug_era group during day 0 through 0 days relative to index: sulfamethoxazole | 1.8 | 0.8 | 0.09 | 2.1 | 0.8 | 0.10 |
| drug_era group during day 0 through 0 days relative to index: Intermediate-acting sulfonamides | 1.8 | 0.8 | 0.09 | 2.1 | 0.8 | 0.10 |

**Supplementary Table 207:** Most extreme patient characteristics balance diagnostics after matching for GLP1RA (female) vs SGLT2I (female) in the MDCD data source.

| | Before matching | | | After matching | | |
|---|---|---|---|---|---|---|
| Characteristic (total count = 14556) | T (%) | C (%) | SDf | T (%) | C (%) | SDf |
| drug_era group during day -365 through 0 days relative to index: liothyronine | 1.0 | -0.2 | 0.12 | 1.1 | -0.2 | 0.14 |
| condition_era group during day -365 through 0 days relative to index: Achilles tendinitis | 0.3 | 1.3 | -0.10 | -0.3 | 1.5 | -0.13 |
| condition_era group during day -30 through 0 days relative to index: Metabolic syndrome X | 2.4 | 0.7 | 0.14 | 2.3 | 0.7 | 0.13 |
| condition_era group during day -365 through 0 days relative to index: Obesity | 58.2 | 47.3 | 0.22 | 54.4 | 60.4 | -0.12 |
| device_exposure during day -365 through 0 days relative to index: Spring-powered device for lancet, each | 0.8 | 0.8 | 0.00 | -0.3 | 0.9 | -0.12 |
| condition_era group during day -30 through 0 days relative to index: Intervertebral disc disorder with myelopathy | 1.1 | 0.2 | 0.10 | 1.1 | -0.2 | 0.12 |
| drug_era group during day -30 through 0 days relative to index: liothyronine | 0.8 | -0.2 | 0.10 | 0.9 | -0.2 | 0.12 |
| drug_era group during day 0 through 0 days relative to index: liothyronine | 0.8 | -0.2 | 0.10 | 0.9 | -0.2 | 0.12 |
| drug_era group during day 0 through 0 days relative to index: sulfamethoxazole | 1.8 | 0.8 | 0.09 | 2.0 | 0.7 | 0.11 |
| drug_era group during day 0 through 0 days relative to index: Intermediate-acting sulfonamides | 1.8 | 0.8 | 0.09 | 2.0 | 0.7 | 0.11 |

##### 3.1.5 Open Claims

**Supplementary Figure 5:** Preference score distributions, propensity score stratified (middle column) and matched (right column) covariate balance and negative control error distributions (bottom row) for GLP1RA (female) vs SGLT2I (female) in the Open Claims data source.

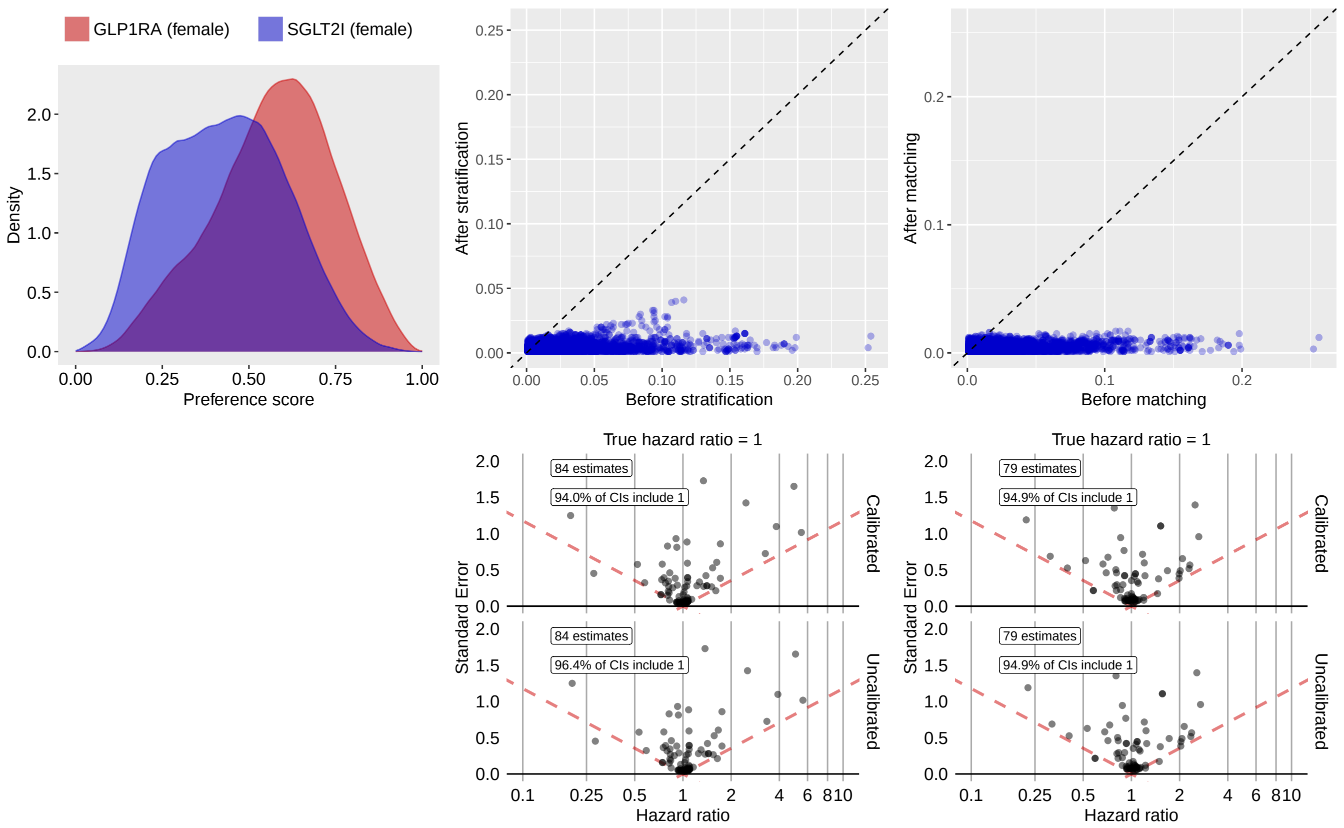

**Supplementary Table 208:** Most extreme patient characteristics balance diagnostics after stratification for GLP1RA (female) vs SGLT2I (female) in the Open Claims data source.

| | Before stratification | | | After stratification | | |
|---|---|---|---|---|---|---|
| Characteristic (total count = 10118) | T (%) | C (%) | SDf | T (%) | C (%) | SDf |
| drug_era group during day -365 through 0 days relative to index: sacubitril | 0.2 | 1.2 | -0.12 | 0.5 | 0.8 | -0.04 |
| drug_era group during day -30 through 0 days relative to index: sacubitril | 0.2 | 1.0 | -0.11 | 0.4 | 0.7 | -0.04 |
| drug_era group during day 0 through 0 days relative to index: sacubitril | 0.2 | 1.0 | -0.11 | 0.4 | 0.6 | -0.04 |
| condition_era group during day -30 through 0 days relative to index: Systolic dysfunction | 0.2 | 0.8 | -0.09 | 0.4 | 0.6 | -0.03 |
| condition_era group during day -30 through 0 days relative to index: Systolic heart failure | 0.2 | 0.8 | -0.09 | 0.3 | 0.6 | -0.03 |
| condition_era group during day -30 through 0 days relative to index: Abnormal cardiovascular function | 0.4 | 1.3 | -0.09 | 0.7 | 1.0 | -0.03 |
| condition_era group during day -30 through 0 days relative to index: Chronic systolic heart failure | 0.1 | 0.6 | -0.08 | 0.3 | 0.4 | -0.03 |
| condition_era group during day -365 through 0 days relative to index: Systolic heart failure | 0.5 | 1.6 | -0.10 | 0.9 | 1.2 | -0.03 |

| (Continued) | T (%) | C (%) | SDf | T (%) | C (%) | SDf |
|---|---|---|---|---|---|---|
| condition_era group during day -30 through 0 days relative to index: Heart failure | 0.7 | 1.9 | -0.10 | 1.2 | 1.5 | -0.03 |
| condition_era group during day -30 through 0 days relative to index: Congestive heart failure | 0.5 | 1.3 | -0.09 | 0.8 | 1.0 | -0.03 |

**Supplementary Table 209:** Most extreme patient characteristics balance diagnostics after matching for GLP1RA (female) vs SGLT2I (female) in the Open Claims data source.

| | Before matching | | | After matching | | |
|---|---|---|---|---|---|---|
| Characteristic (total count = 9995) | T (%) | C (%) | SDf | T (%) | C (%) | SDf |
| drug_era group during day -365 through 0 days relative to index: sacubitril | 0.2 | 1.2 | -0.12 | 0.3 | 0.4 | -0.02 |
| drug_era group during day -30 through 0 days relative to index: sacubitril | 0.2 | 1.0 | -0.11 | 0.2 | 0.3 | -0.02 |
| drug_era group during day 0 through 0 days relative to index: sacubitril | 0.2 | 1.0 | -0.11 | 0.2 | 0.3 | -0.02 |
| condition_era group during day -365 through 0 days relative to index: Lower urinary tract obstructive syndrome | 0.1 | 0.1 | -0.02 | 0.1 | 0.1 | -0.02 |
| condition_era group during day -365 through 0 days relative to index: Urinary outflow obstruction | 0.1 | 0.1 | -0.02 | 0.1 | 0.1 | -0.02 |
| index year: 2014 | 1.8 | 5.5 | -0.20 | 2.4 | 2.6 | -0.01 |
| condition_era group during day -30 through 0 days relative to index: Heart failure | 0.7 | 1.9 | -0.10 | 0.9 | 1.0 | -0.01 |
| condition_era group during day -30 through 0 days relative to index: Abnormal cardiovascular function | 0.4 | 1.3 | -0.09 | 0.5 | 0.6 | -0.01 |
| condition_era group during day -365 through 0 days relative to index: Cyst of abdomen | 4.5 | 2.9 | 0.09 | 3.0 | 3.3 | -0.01 |
| drug_era group during day -365 through 0 days relative to index: Other antipsychotics | 3.6 | 2.5 | 0.07 | 2.8 | 3.0 | -0.01 |

##### 3.1.6 OEHR

**Supplementary Figure 6:** Preference score distributions, propensity score stratified (middle column) and matched (right column) covariate balance and negative control error distributions (bottom row) for GLP1RA (female) vs SGLT2I (female) in the OEHR data source.

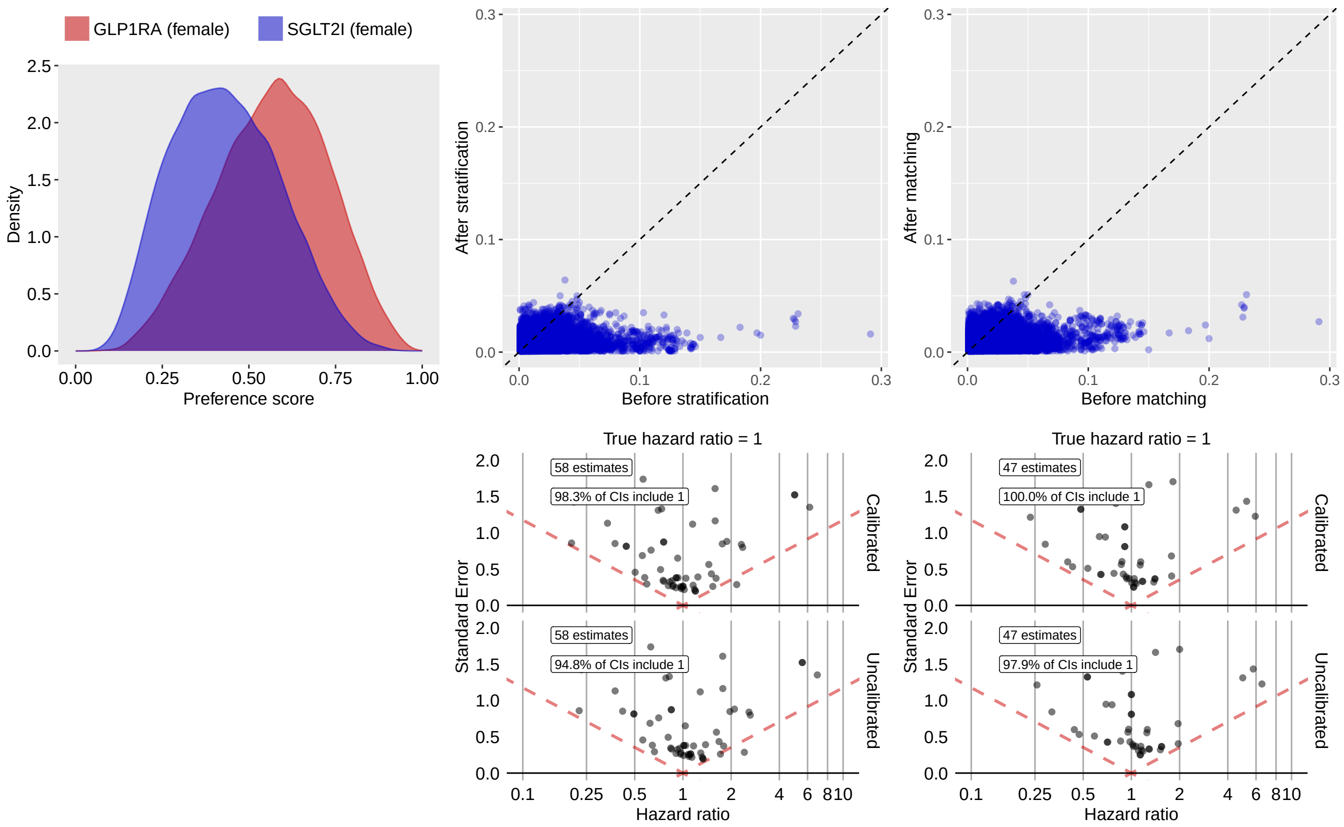

**Supplementary Table 210:** Most extreme patient characteristics balance diagnostics after stratification for GLP1RA (female) vs SGLT2I (female) in the OEHR data source.

| | Before stratification | | | After stratification | | |
|---|---|---|---|---|---|---|
| Characteristic (total count = 12308) | T (%) | C (%) | SDf | T (%) | C (%) | SDf |
| observation during day -365 through 0 days relative to index: Intensive Care - Psychiatric | 0.2 | 0.1 | 0.04 | 0.3 | 0.1 | 0.06 |
| drug_era group during day -365 through 0 days relative to index: sacubitril | 0.1 | 0.3 | -0.05 | 0.1 | 0.4 | -0.05 |
| drug_era group during day -365 through 0 days relative to index: water | 0.2 | 0.3 | -0.03 | 0.1 | 0.4 | -0.05 |
| observation during day -365 through 0 days relative to index: Other Therapeutic Services - Cardiac Rehabilitation | 0.1 | 0.3 | -0.05 | 0.1 | 0.2 | -0.05 |
| drug_era group during day -30 through 0 days relative to index: sacubitril | 0.1 | 0.3 | -0.04 | 0.1 | 0.3 | -0.04 |
| age group: 85 - 89 | 0.1 | 0.4 | -0.05 | 0.2 | 0.4 | -0.04 |
| observation during day -30 through 0 days relative to index: Colorectal cancer screening results documented and reviewed (PV) | 0.3 | 0.2 | 0.01 | 0.4 | 0.2 | 0.04 |
| observation during day -365 through 0 days relative to index: Pharmacy - Generic Drugs | 0.3 | 0.6 | -0.05 | 0.3 | 0.6 | -0.04 |

| (Continued) | T (%) | C (%) | SDf | T (%) | C (%) | SDf |
|---|---|---|---|---|---|---|
| procedure_occurrence during day -365 through 0 days relative to index: Physician or other qualified health care professional services for outpatient cardiac rehabilitation; with continuous ECG monitoring (per session) | 0.1 | 0.3 | -0.04 | 0.1 | 0.2 | -0.04 |
| drug_era group during day 0 through 0 days relative to index: sacubitril | 0.1 | 0.3 | -0.04 | 0.1 | 0.3 | -0.04 |

**Supplementary Table 211:** Most extreme patient characteristics balance diagnostics after matching for GLP1RA (female) vs SGLT2I (female) in the OEHR data source.

| | Before matching | | | After matching | | |
|---|---|---|---|---|---|---|
| Characteristic (total count = 12482) | T (%) | C (%) | SDf | T (%) | C (%) | SDf |
| observation during day -365 through 0 days relative to index: Intensive Care - Psychiatric | 0.2 | 0.1 | 0.04 | 0.3 | 0.1 | 0.06 |
| condition_era group during day -365 through 0 days relative to index: Morbid obesity | 28.3 | 18.6 | 0.23 | 22.5 | 24.7 | -0.05 |
| age group: 85 - 89 | 0.1 | 0.4 | -0.05 | 0.2 | 0.4 | -0.05 |
| condition_era group during day -365 through 0 days relative to index: Congenital abnormality of lower limb and pelvic girdle | 0.7 | 0.3 | 0.05 | 0.7 | 0.3 | 0.05 |
| observation during day -365 through 0 days relative to index: Other Therapeutic Services - Cardiac Rehabilitation | 0.1 | 0.3 | -0.05 | 0.1 | 0.2 | -0.05 |
| drug_era group during day -365 through 0 days relative to index: water | 0.2 | 0.3 | -0.03 | 0.2 | 0.4 | -0.05 |
| condition_era group during day -30 through 0 days relative to index: Bacterial upper respiratory infection | 0.2 | 0.1 | 0.04 | 0.2 | 0.1 | 0.05 |
| procedure_occurrence during day -365 through 0 days relative to index: Physician or other qualified health care professional services for outpatient cardiac rehabilitation; with continuous ECG monitoring (per session) | 0.1 | 0.3 | -0.04 | 0.1 | 0.2 | -0.04 |
| condition_era group during day -365 through 0 days relative to index: Congenital anomaly of abdomen | 0.4 | 0.2 | 0.04 | 0.4 | 0.2 | 0.04 |
| drug_era group during day -365 through 0 days relative to index: sacubitril | 0.1 | 0.3 | -0.05 | 0.2 | 0.4 | -0.04 |

#### 3.2 SGLT2I (female) vs DPP4I (female)

##### 3.2.1 CCAE

**Supplementary Figure 7:** Preference score distributions, propensity score stratified (middle column) and matched (right column) covariate balance and negative control error distributions (bottom row) for SGLT2I (female) vs DPP4I (female) in the CCAE data source.

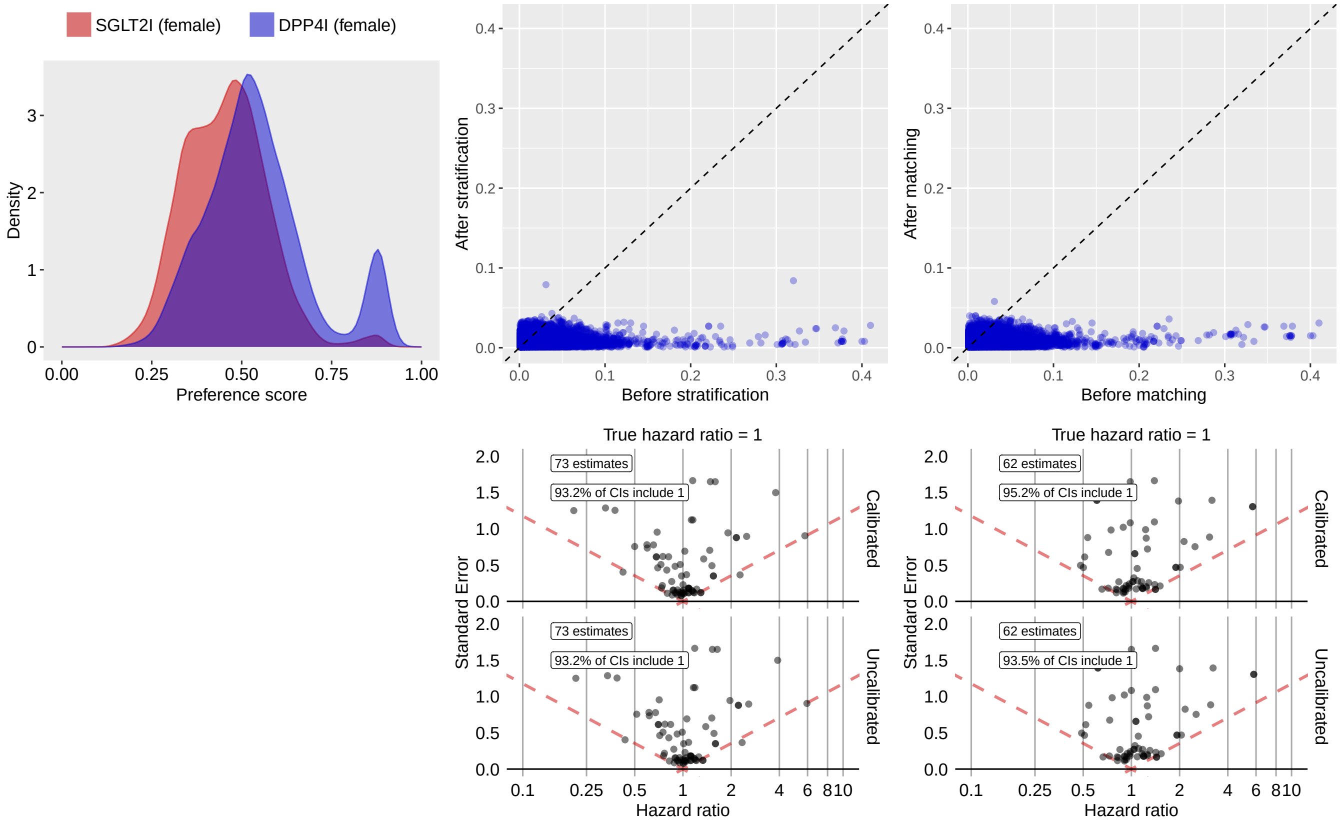

**Supplementary Table 212:** Most extreme patient characteristics balance diagnostics after stratification for SGLT2I (female) vs DPP4I (female) in the CCAE data source.

| | Before stratification | | | After stratification | | |
|---|---|---|---|---|---|---|
| Characteristic (total count = 11683) | T (%) | C (%) | SDf | T (%) | C (%) | SDf |
| index year: 2013 | 1.3 | 8.0 | 0.32 | 4.6 | 6.5 | 0.08 |
| index year: 2014 | 9.7 | 8.8 | -0.03 | 15.1 | 12.4 | -0.08 |
| condition_era group during day -30 through 0 days relative to index: Atelectasis | 0.1 | 0.2 | 0.04 | 0.1 | 0.2 | 0.04 |
| measurement within normal range during day -365 through 0 days relative to index: Platelet mean volume [Entitic volume] in Blood by Automated count | 0.1 | 0.1 | 0.03 | 0.1 | 0.2 | 0.04 |
| condition_era group during day -365 through 0 days relative to index: Periocular open wound | 0.3 | 0.1 | -0.05 | 0.4 | 0.2 | -0.04 |
| condition_era group during day -30 through 0 days relative to index: Renal failure syndrome | 0.2 | 0.5 | 0.06 | 0.3 | 0.5 | 0.04 |
| procedure_occurrence during day -365 through 0 days relative to index: Bone and/or joint imaging; whole body | 0.1 | 0.4 | 0.04 | 0.2 | 0.3 | 0.04 |

| (Continued) | T (%) | C (%) | SDf | T (%) | C (%) | SDf |
|---|---|---|---|---|---|---|
| condition_era group during day -30 through 0 days relative to index: Hypoglycemic disorder | 0.3 | 0.1 | -0.04 | 0.3 | 0.1 | -0.04 |
| condition_era group during day -30 through 0 days relative to index: Acute nephropathy | 0.1 | 0.4 | 0.05 | 0.2 | 0.4 | 0.04 |
| measurement above normal range during day -365 through 0 days relative to index: Cholesterol in LDL/Cholesterol in HDL [Mass Ratio] in Serum or Plasma | 0.5 | 0.3 | -0.03 | 0.6 | 0.3 | -0.04 |

**Supplementary Table 213:** Most extreme patient characteristics balance diagnostics after matching for SGLT2I (female) vs DPP4I (female) in the CCAE data source.

| | Before matching | | | After matching | | |
|---|---|---|---|---|---|---|
| Characteristic (total count = 11527) | T (%) | C (%) | SDf | T (%) | C (%) | SDf |
| index year: 2014 | 9.7 | 8.8 | -0.03 | 11.1 | 9.4 | -0.06 |
| condition_era group during day -30 through 0 days relative to index: Thyroid atrophy | 0.2 | 0.2 | -0.01 | 0.2 | 0.4 | 0.04 |
| condition_era group during day -30 through 0 days relative to index: Atrophy of thyroid - acquired | 0.2 | 0.2 | -0.01 | 0.2 | 0.4 | 0.04 |
| measurement during day -365 through 0 days relative to index: Thyroid disorder screening | 0.4 | 0.4 | 0.00 | 0.4 | 0.2 | -0.04 |
| measurement during day -365 through 0 days relative to index: Antistreptolysin 0; titer | 0.3 | 0.3 | -0.01 | 0.3 | 0.1 | -0.04 |
| condition_era group during day -30 through 0 days relative to index: Acute nephropathy | 0.1 | 0.4 | 0.05 | 0.1 | 0.3 | 0.04 |
| observation during day -365 through 0 days relative to index: Influenza immunization was not administered, reason not given | 0.5 | 0.4 | -0.02 | 0.5 | 0.8 | 0.04 |
| condition_era group during day -30 through 0 days relative to index: Primary insomnia | 0.2 | 0.1 | -0.01 | 0.2 | 0.4 | 0.04 |
| procedure_occurrence during day -365 through 0 days relative to index: Computer-aided detection (computer algorithm analysis of digital image data for lesion detection) with further review for interpretation, with or without digitization of film radiographic images; screening mammography (List separately in addition to code | 19.5 | 29.6 | 0.23 | 21.8 | 20.4 | -0.04 |
| condition_era group during day -30 through 0 days relative to index: Renal failure syndrome | 0.2 | 0.5 | 0.06 | 0.2 | 0.4 | 0.04 |

##### 3.2.2 DAG

**Supplementary Figure 8:** Preference score distributions, propensity score stratified (middle column) and matched (right column) covariate balance and negative control error distributions (bottom row) for SGLT2I (female) vs DPP4I (female) in the DAG data source.

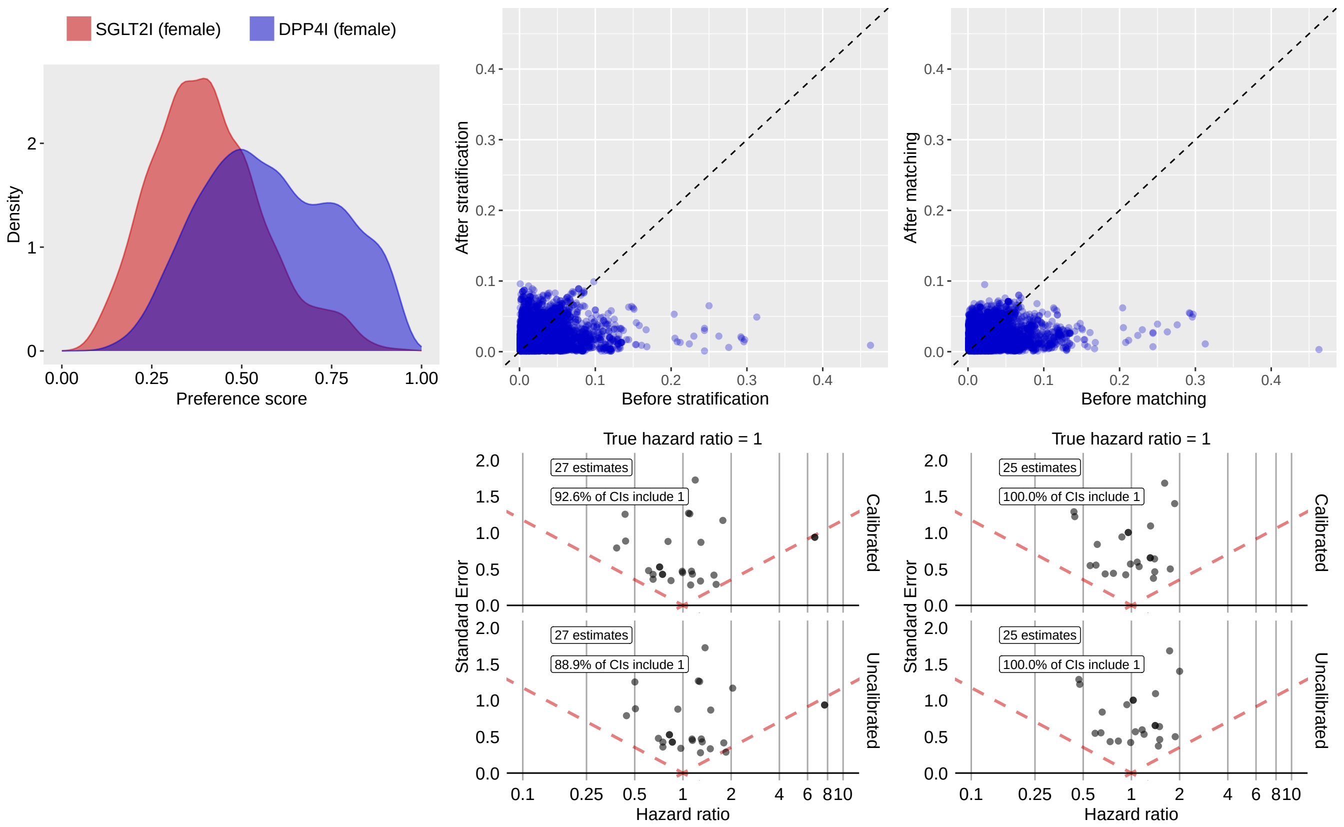

**Supplementary Table 214:** Most extreme patient characteristics balance diagnostics after stratification for SGLT2I (female) vs DPP4I (female) in the DAG data source.

| Characteristic (total count = 6753) | Before stratification | | | After stratification | | |
|---|---|---|---|---|---|---|
| | T (%) | C (%) | SDf | T (%) | C (%) | SDf |
| condition_era group during day -365 through 0 days relative to index: Extrapyramidal disease | -0.2 | 0.7 | 0.10 | -0.1 | 0.7 | 0.10 |
| condition_era group during day -365 through 0 days relative to index: Toxic liver disease | 0.2 | 0.2 | -0.00 | 0.8 | 0.2 | -0.10 |
| procedure_occurrence during day -365 through 0 days relative to index: Developing a treatment plan | 0.9 | 0.8 | -0.01 | 1.6 | 0.7 | -0.09 |
| condition_era group during day -365 through 0 days relative to index: Senility | 0.9 | 1.7 | 0.08 | 0.8 | 1.8 | 0.09 |
| condition_era group during day -365 through 0 days relative to index: Aging | 0.9 | 1.7 | 0.08 | 0.8 | 1.8 | 0.09 |
| condition_era group during day -365 through 0 days relative to index: Old-age | 0.9 | 1.7 | 0.08 | 0.8 | 1.8 | 0.09 |
| drug_era group during day -365 through 0 days relative to index: ferric cation | 0.6 | 0.5 | -0.02 | 1.3 | 0.5 | -0.09 |
| condition_era group during day -365 through 0 days relative to index: Acute exacerbation of chronic obstructive airways disease | 1.0 | 0.8 | -0.02 | 1.9 | 0.9 | -0.09 |

| (Continued) | T (%) | C (%) | SDf | T (%) | C (%) | SDf |
|---|---|---|---|---|---|---|
| drug_era group during day 0 through 0 days relative to index: aliskiren | 0.3 | 0.2 | -0.01 | 0.7 | 0.1 | -0.09 |
| drug_era group during day 0 through 0 days relative to index: OTHER AGENTS ACTING ON THE RENIN-ANGIOTENSIN SYSTEM | 0.3 | 0.2 | -0.01 | 0.7 | 0.1 | -0.09 |

**Supplementary Table 215:** Most extreme patient characteristics balance diagnostics after matching for SGLT2I (female) vs DPP4I (female) in the DAG data source.

| | Before matching | | | After matching | | |
|---|---|---|---|---|---|---|
| Characteristic (total count = 6232) | T (%) | C (%) | SDf | T (%) | C (%) | SDf |
| Diabetes Comorbidity Severity Index (DCSI) | 150.8 | 147.1 | -0.02 | 146.6 | 131.3 | -0.10 |
| drug_era group during day -30 through 0 days relative to index: prasugrel | 0.5 | 0.1 | -0.07 | 0.4 | -0.2 | -0.08 |
| drug_era group during day 0 through 0 days relative to index: prasugrel | 0.5 | 0.1 | -0.07 | 0.4 | -0.2 | -0.08 |
| condition_era group during day -30 through 0 days relative to index: Thrombosis | 0.2 | 0.6 | 0.07 | 0.1 | 0.6 | 0.08 |
| condition_era group during day -365 through 0 days relative to index: Developmental mental disorder | 0.2 | 0.4 | 0.05 | -0.1 | 0.6 | 0.08 |
| condition_era group during day -30 through 0 days relative to index: Thrombosis of blood vessel | 0.2 | 0.6 | 0.07 | 0.1 | 0.6 | 0.07 |
| condition_era group during day -365 through 0 days relative to index: Cranial nerve disorder | 1.0 | 0.6 | -0.04 | 1.0 | 0.4 | -0.07 |
| condition_era group during day -365 through 0 days relative to index: Lesion of eyelid | -0.2 | 0.1 | 0.03 | -0.1 | 0.3 | 0.07 |
| condition_era group during day -30 through 0 days relative to index: Phlebitis of deep veins of lower extremity | -0.2 | 0.2 | 0.06 | -0.1 | 0.3 | 0.07 |
| condition_era group during day -30 through 0 days relative to index: Deep vein phlebitis and thrombophlebitis of the leg | -0.2 | 0.2 | 0.05 | -0.1 | 0.3 | 0.07 |

##### 3.2.3 ODOD

**Supplementary Figure 9:** Preference score distributions, propensity score stratified (middle column) and matched (right column) covariate balance and negative control error distributions (bottom row) for SGLT2I (female) vs DPP4I (female) in the ODOD data source.

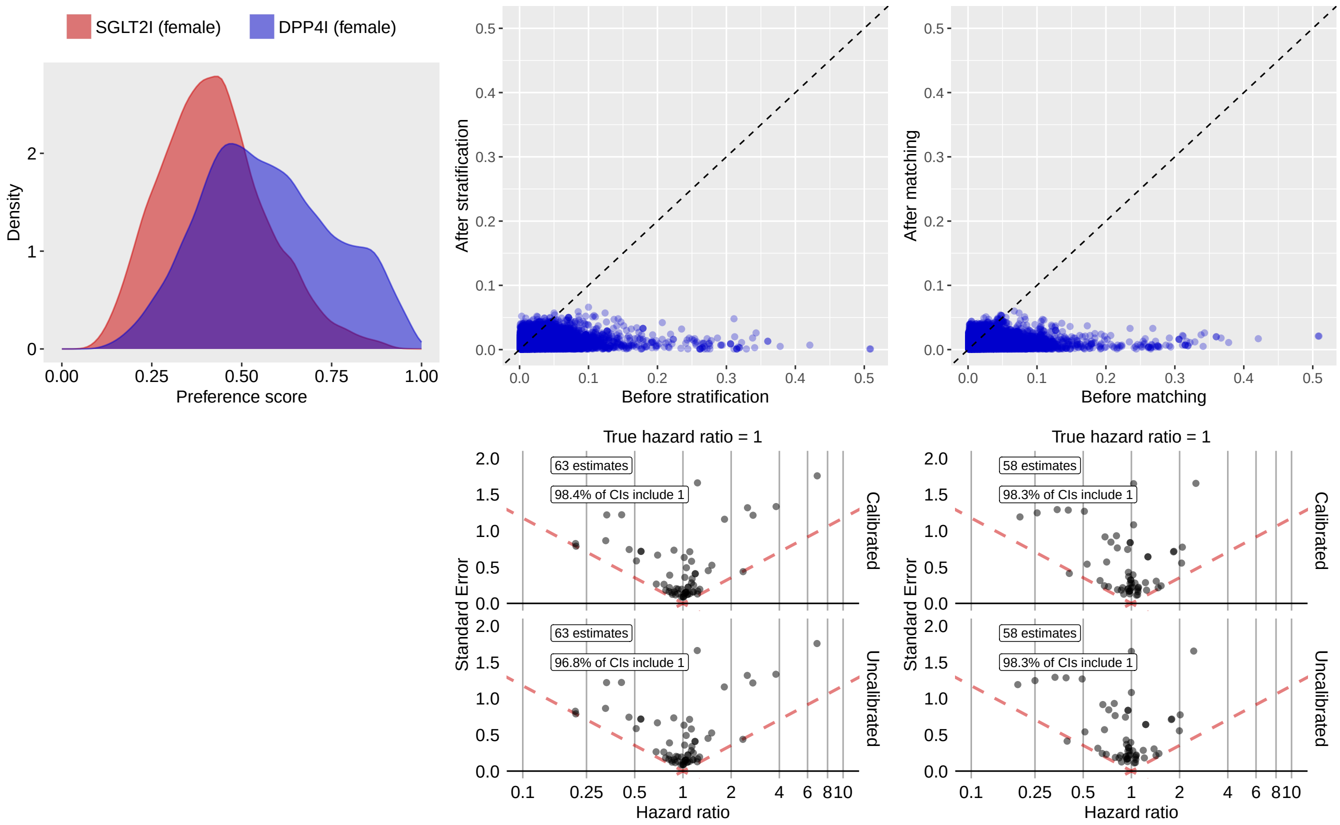

**Supplementary Table 216:** Most extreme patient characteristics balance diagnostics after stratification for SGLT2I (female) vs DPP4I (female) in the ODOD data source.

| | Before stratification | | | After stratification | | |
|---|---|---|---|---|---|---|
| Characteristic (total count = 16017) | T (%) | C (%) | SDf | T (%) | C (%) | SDf |
| condition_era group during day -365 through 0 days relative to index: Chronic kidney disease stage 4 | 0.3 | 1.1 | 0.10 | 0.4 | 1.0 | 0.07 |
| condition_era group during day -30 through 0 days relative to index: Chronic kidney disease stage 4 | 0.1 | 0.7 | 0.09 | 0.2 | 0.6 | 0.06 |
| device_exposure during day -365 through 0 days relative to index: CONTOUR TEST STRIPS | 0.1 | 1.0 | 0.12 | 0.1 | 0.4 | 0.06 |
| measurement during day -365 through 0 days relative to index: Drug screen, qualitative; single drug class method (eg, immunoassay, enzyme assay), each drug class | 0.2 | 0.4 | 0.05 | 0.5 | 0.2 | -0.06 |
| drug_era group during day -365 through 0 days relative to index: calcitriol | 0.1 | 0.4 | 0.05 | 0.1 | 0.4 | 0.05 |
| drug_era group during day -30 through 0 days relative to index: sacubitril | 0.9 | 0.1 | -0.11 | 0.5 | 0.2 | -0.05 |
| condition_era group during day -30 through 0 days relative to index: Hypertensive renal failure | 0.1 | 0.6 | 0.10 | 0.1 | 0.3 | 0.05 |

| (Continued) | T (%) | C (%) | SDf | T (%) | C (%) | SDf |
|---|---|---|---|---|---|---|
| condition_era group during day -30 through 0 days relative to index: Abnormal breathing | 0.2 | 0.8 | 0.09 | 0.2 | 0.5 | 0.05 |
| condition_era group during day -30 through 0 days relative to index: Chronic kidney disease stage 2 | 1.2 | 0.9 | -0.03 | 1.6 | 1.0 | -0.05 |
| condition_era group during day -365 through 0 days relative to index: Closed fracture of intracapsular section of femur | 0.1 | 0.1 | -0.00 | 0.3 | 0.1 | -0.05 |

**Supplementary Table 217:** Most extreme patient characteristics balance diagnostics after matching for SGLT2I (female) vs DPP4I (female) in the ODOD data source.

| | Before matching | | | After matching | | |
|---|---|---|---|---|---|---|
| Characteristic (total count = 15761) | T (%) | C (%) | SDf | T (%) | C (%) | SDf |
| condition_era group during day -30 through 0 days relative to index: Chronic kidney disease stage 3A | 0.5 | 0.1 | -0.07 | 0.5 | 0.2 | -0.06 |
| condition_era group during day -365 through 0 days relative to index: Chronic kidney disease stage 3A | 0.7 | 0.2 | -0.07 | 0.7 | 0.3 | -0.06 |
| measurement within normal range during day -365 through 0 days relative to index: Codfish IgE Ab [Units/volume] in Serum | 0.3 | 0.1 | -0.05 | 0.3 | 0.1 | -0.05 |
| measurement during day -365 through 0 days relative to index: Egg white IgE Ab [Units/volume] in Serum | 0.4 | 0.1 | -0.05 | 0.4 | 0.1 | -0.05 |
| measurement within normal range during day -365 through 0 days relative to index: Egg white IgE Ab [Units/volume] in Serum | 0.3 | 0.1 | -0.05 | 0.3 | 0.1 | -0.05 |
| measurement during day -365 through 0 days relative to index: Codfish IgE Ab [Units/volume] in Serum | 0.3 | 0.1 | -0.05 | 0.3 | 0.1 | -0.05 |
| condition_era group during day -365 through 0 days relative to index: Chronic congestive heart failure | 1.5 | 0.7 | -0.08 | 1.3 | 0.8 | -0.05 |
| age group: 90 - 94 | 0.1 | 0.2 | 0.03 | 0.1 | 0.4 | 0.05 |
| condition_era group during day -365 through 0 days relative to index: Chronic systolic heart failure | 2.2 | 1.4 | -0.06 | 1.8 | 1.2 | -0.05 |
| measurement during day -365 through 0 days relative to index: Peanut IgE Ab [Units/volume] in Serum | 0.4 | 0.2 | -0.04 | 0.4 | 0.2 | -0.05 |

##### 3.2.4 MDCR

**Supplementary Figure 10:** Preference score distributions, propensity score stratified (middle column) and matched (right column) covariate balance and negative control error distributions (bottom row) for SGLT2I (female) vs DPP4I (female) in the MDCR data source.

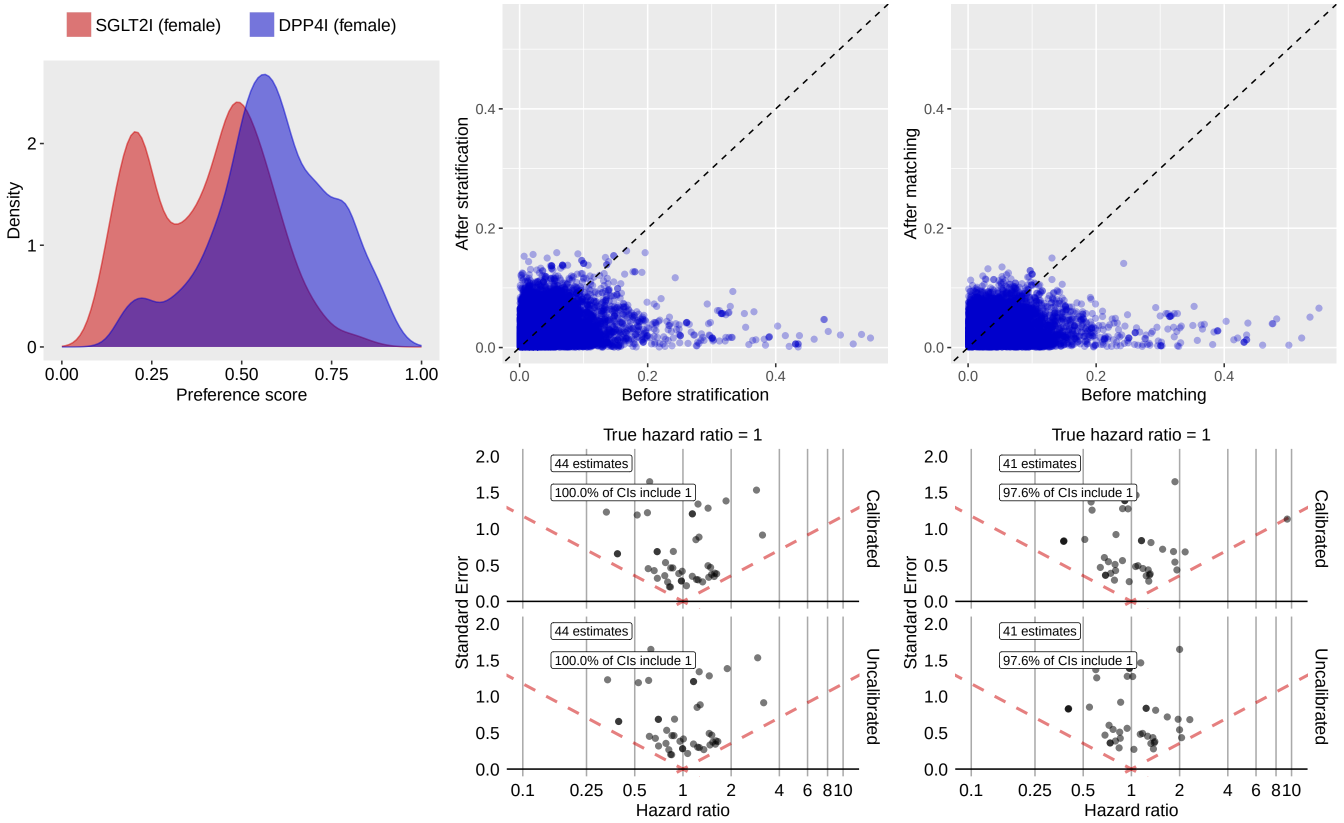

**Supplementary Table 218:** Most extreme patient characteristics balance diagnostics after stratification for SGLT2I (female) vs DPP4I (female) in the MDCR data source.

| Characteristic (total count = 15404) | Before stratification | | | After stratification | | |
|---|---|---|---|---|---|---|
| | T (%) | C (%) | SDf | T (%) | C (%) | SDf |
| condition_era group during day -30 through 0 days relative to index: Renal failure syndrome | 0.5 | 2.6 | 0.17 | 0.5 | 2.5 | 0.16 |
| condition_era group during day -365 through 0 days relative to index: Renal failure syndrome | 2.3 | 6.2 | 0.20 | 2.8 | 6.1 | 0.16 |
| procedure_occurrence during day -30 through 0 days relative to index: Scanning computerized ophthalmic diagnostic imaging, posterior segment, with interpretation and report, unilateral or bilateral; retina | 1.8 | 1.1 | -0.06 | 3.9 | 1.3 | -0.16 |
| condition_era group during day -30 through 0 days relative to index: Renal impairment | 5.0 | 7.1 | 0.09 | 4.6 | 8.5 | 0.16 |
| procedure_occurrence during day -365 through 0 days relative to index: Spinal puncture, lumbar, diagnostic | 0.3 | 0.2 | -0.02 | 1.7 | 0.2 | -0.16 |
| condition_era group during day -365 through 0 days relative to index: Acute renal failure syndrome | 1.8 | 4.3 | 0.15 | 1.8 | 4.5 | 0.15 |
| condition_era group during day -365 through 0 days relative to index: Acute renal impairment | 1.8 | 4.3 | 0.15 | 1.8 | 4.5 | 0.15 |

| (Continued) | T (%) | C (%) | SDf | T (%) | C (%) | SDf |
|---|---|---|---|---|---|---|
| measurement during day -30 through 0 days relative to index: Myoglobin | -0.3 | 0.2 | -0.01 | 1.5 | 0.1 | -0.15 |
| condition_era group during day -365 through 0 days relative to index: Acute nephropathy | 2.1 | 4.5 | 0.13 | 2.0 | 4.6 | 0.15 |
| drug_era group during day -365 through 0 days relative to index: tolterodine | 2.1 | 2.7 | 0.04 | 4.0 | 1.5 | -0.15 |

**Supplementary Table 219:** Most extreme patient characteristics balance diagnostics after matching for SGLT2I (female) vs DPP4I (female) in the MDCR data source.

| | Before matching | | | After matching | | |
|---|---|---|---|---|---|---|
| Characteristic (total count = 11333) | T (%) | C (%) | SDf | T (%) | C (%) | SDf |
| age group: 90 - 94 | 0.5 | 1.9 | 0.13 | 0.5 | 2.3 | 0.15 |
| age group: 65 - 69 | 40.8 | 29.3 | -0.24 | 40.6 | 33.8 | -0.14 |
| drug_era group during day -365 through 0 days relative to index: Insulins and analogues for injection, intermediate- or long-acting combined with fast-acting | 1.0 | 0.5 | -0.05 | 1.1 | -0.4 | -0.14 |
| drug_era group during day -30 through 0 days relative to index: nitroglycerin | 1.3 | 1.0 | -0.03 | 1.4 | -0.4 | -0.13 |
| condition_era group during day -30 through 0 days relative to index: Finding related to ability to perform gross motor function | 0.3 | 1.2 | 0.10 | -0.3 | 1.5 | 0.13 |
| condition_era group during day -30 through 0 days relative to index: Finding related to ability to move | 0.3 | 1.2 | 0.10 | -0.3 | 1.5 | 0.13 |
| observation during day -30 through 0 days relative to index: Advance Care Planning discussed and documented advance care plan or surrogate decision maker documented in the medical record (DEM) (GER, Pall Cr) | -0.3 | 0.7 | 0.06 | -0.3 | 1.4 | 0.12 |
| condition_era group during day -30 through 0 days relative to index: Difficulty moving | -0.3 | 1.1 | 0.10 | -0.3 | 1.4 | 0.12 |
| condition_era group during day -30 through 0 days relative to index: Finding related to ability to walk | -0.3 | 1.1 | 0.10 | -0.3 | 1.4 | 0.12 |
| condition_era group during day -30 through 0 days relative to index: Difficulty walking | -0.3 | 1.1 | 0.10 | -0.3 | 1.4 | 0.12 |

##### 3.2.5 MDCD

**Supplementary Figure 11:** Preference score distributions, propensity score stratified (middle column) and matched (right column) covariate balance and negative control error distributions (bottom row) for SGLT2I (female) vs DPP4I (female) in the MDCD data source.

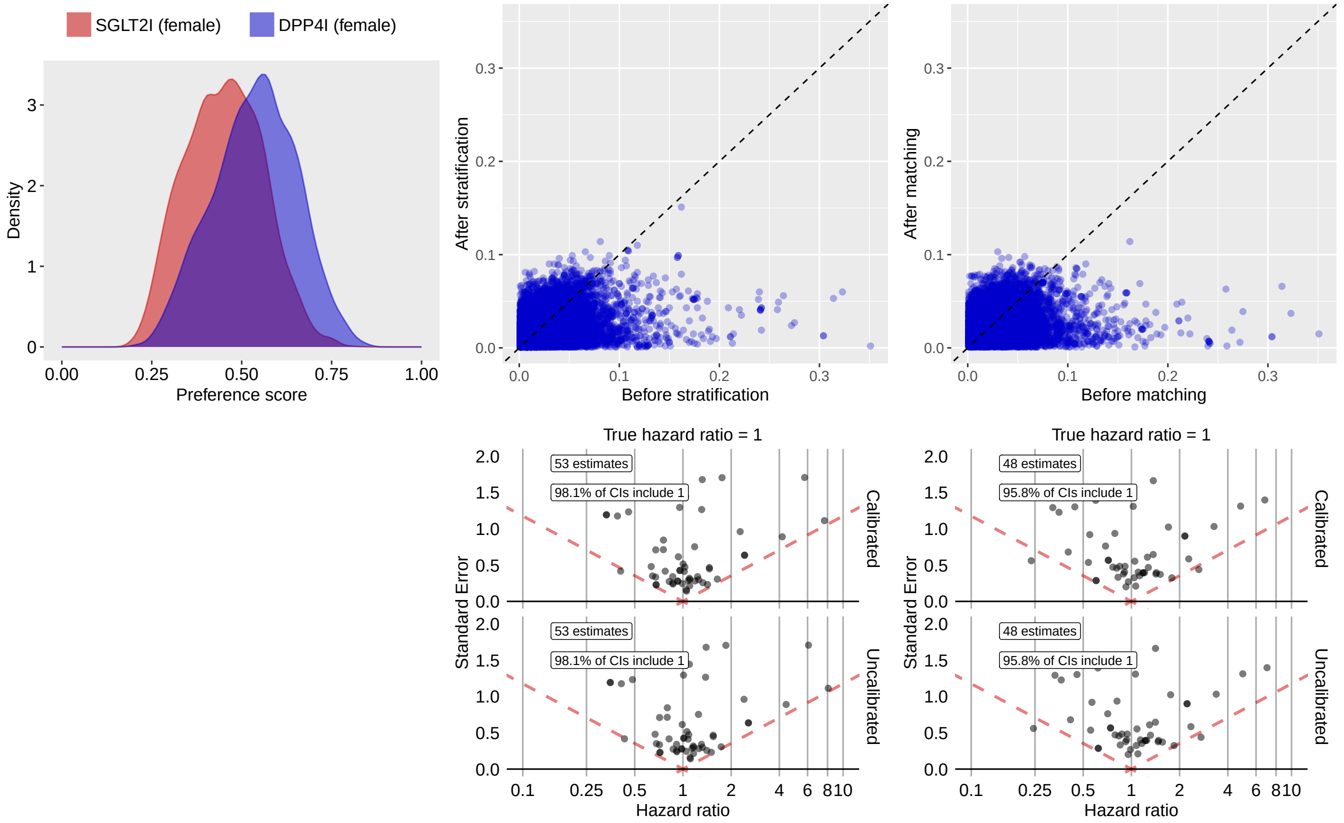

**Supplementary Table 220:** Most extreme patient characteristics balance diagnostics after stratification for SGLT2I (female) vs DPP4I (female) in the MDCD data source.

| Characteristic (total count = 19075) | Before stratification | | | After stratification | | |
|---|---|---|---|---|---|---|
| | T (%) | C (%) | SDf | T (%) | C (%) | SDf |
| age group: 75 - 79 | 0.2 | 1.9 | 0.16 | 0.2 | 1.6 | 0.15 |
| condition_era group during day -30 through 0 days relative to index: Organic mental disorder | 0.8 | 1.7 | 0.08 | 0.6 | 1.8 | 0.11 |
| condition_era group during day -30 through 0 days relative to index: Acute genitourinary disorder | 0.6 | 1.9 | 0.12 | 0.6 | 1.9 | 0.11 |
| condition_era group during day -30 through 0 days relative to index: Acute nephropathy | 0.3 | 1.2 | 0.11 | 0.3 | 1.2 | 0.10 |
| condition_era group during day -30 through 0 days relative to index: Acute renal failure syndrome | 0.2 | 1.2 | 0.11 | 0.3 | 1.1 | 0.10 |
| condition_era group during day -30 through 0 days relative to index: Acute renal impairment | 0.2 | 1.2 | 0.11 | 0.3 | 1.1 | 0.10 |
| procedure_occurrence during day -365 through 0 days relative to index: Electroretinography with interpretation and report | 0.4 | 0.1 | -0.07 | 0.7 | 0.1 | -0.10 |
| condition_era group during day -365 through 0 days relative to index: Allergic disorder of respiratory system | -0.2 | 1.7 | 0.16 | 0.3 | 1.2 | 0.10 |

| (Continued) | T (%) | C (%) | SDf | T (%) | C (%) | SDf |
|---|---|---|---|---|---|---|
| condition_era group during day -365 through 0 days relative to index: Allergic disorder of respiratory tract | -0.2 | 1.7 | 0.16 | 0.3 | 1.2 | 0.10 |
| condition_era group during day -30 through 0 days relative to index: Dementia | -0.2 | 0.8 | 0.09 | -0.2 | 0.7 | 0.10 |

**Supplementary Table 221:** Most extreme patient characteristics balance diagnostics after matching for SGLT2I (female) vs DPP4I (female) in the MDCD data source.

| | Before matching | | | After matching | | |
|---|---|---|---|---|---|---|
| Characteristic (total count = 14544) | T (%) | C (%) | SDf | T (%) | C (%) | SDf |
| age group: 75 - 79 | 0.2 | 1.9 | 0.16 | 0.3 | 1.2 | 0.11 |
| observation during day -365 through 0 days relative to index: Most recent diastolic blood pressure 80-89 mm Hg (HTN, CKD, CAD) (DM) | 2.8 | 2.4 | -0.03 | 2.6 | 4.6 | 0.10 |
| procedure_occurrence during day -365 through 0 days relative to index: Radiologic examination, ribs, unilateral; including posteroanterior chest, minimum of 3 views | 0.6 | 1.1 | 0.06 | 0.6 | 1.6 | 0.10 |
| device_exposure during day -365 through 0 days relative to index: ACCU-CHEK SMARTVIEW STRIP | 1.1 | 0.7 | -0.04 | 1.1 | 0.3 | -0.10 |
| index month: 1 | 9.1 | 10.1 | 0.03 | 8.7 | 11.6 | 0.10 |
| condition_era group during day -365 through 0 days relative to index: Urinary incontinence of non-organic origin | -0.2 | 0.5 | 0.05 | -0.2 | 0.8 | 0.09 |
| condition_era group during day -365 through 0 days relative to index: Functional urinary incontinence | -0.2 | 0.5 | 0.05 | -0.2 | 0.8 | 0.09 |
| condition_era group during day -30 through 0 days relative to index: Acute genitourinary disorder | 0.6 | 1.9 | 0.12 | 0.6 | 1.5 | 0.09 |
| observation during day -30 through 0 days relative to index: Blood pressure measured (CKD)(DM) | -0.2 | 0.3 | 0.06 | -0.2 | 0.5 | 0.09 |
| condition_era group during day -30 through 0 days relative to index: Schizoaffective disorder, bipolar type | 0.4 | 0.8 | 0.05 | 0.4 | 1.2 | 0.09 |

##### 3.2.6 Open Claims

**Supplementary Figure 12:** Preference score distributions, propensity score stratified (middle column) and matched (right column) covariate balance and negative control error distributions (bottom row) for SGLT2I (female) vs DPP4I (female) in the Open Claims data source.

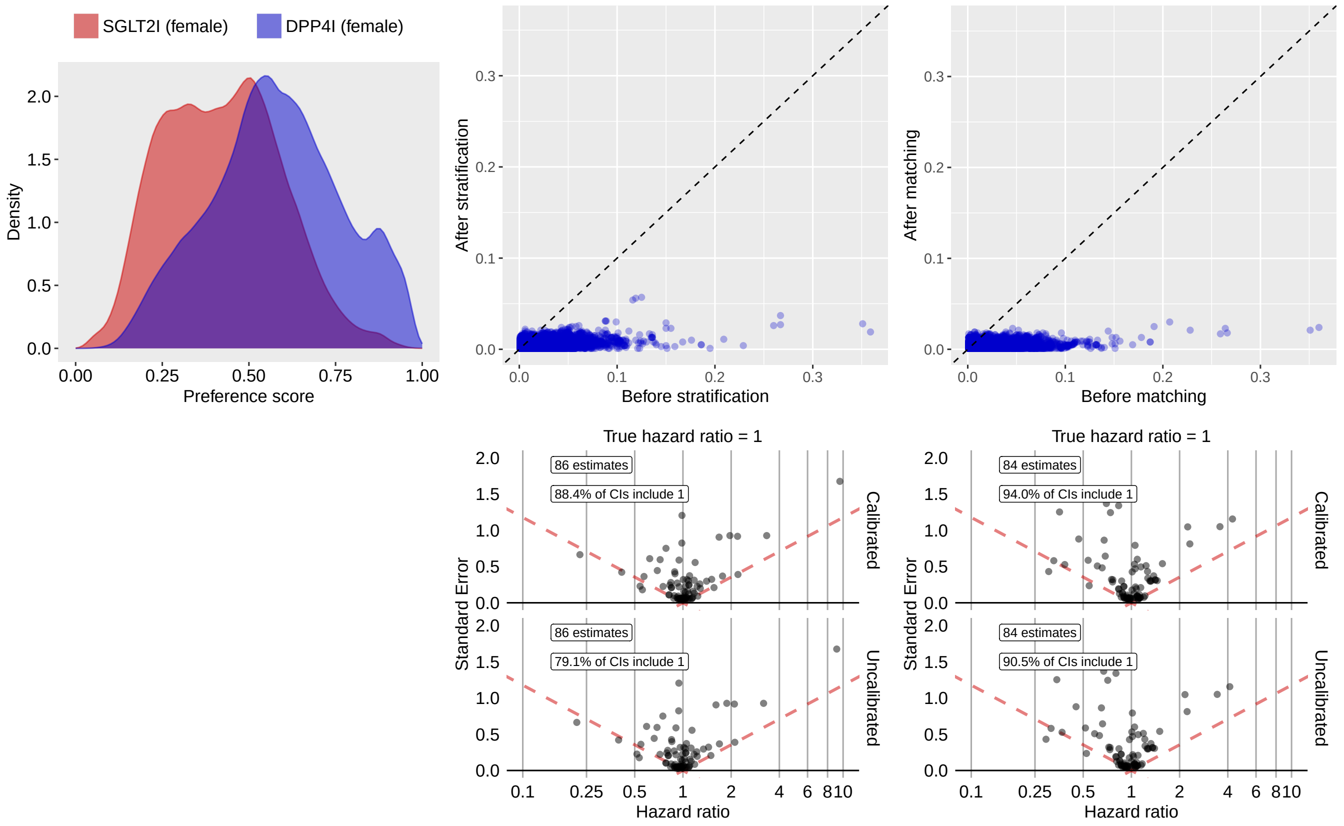

**Supplementary Table 222:** Most extreme patient characteristics balance diagnostics after stratification for SGLT2I (female) vs DPP4I (female) in the Open Claims data source.

| | Before stratification | | | After stratification | | |
|---|---|---|---|---|---|---|
| Characteristic (total count = 10642) | T (%) | C (%) | SDf | T (%) | C (%) | SDf |
| drug_era group during day -365 through 0 days relative to index: sacubitril | 1.2 | 0.2 | -0.12 | 0.6 | 0.3 | -0.06 |
| drug_era group during day -30 through 0 days relative to index: sacubitril | 1.0 | 0.1 | -0.12 | 0.6 | 0.2 | -0.06 |
| drug_era group during day 0 through 0 days relative to index: sacubitril | 1.0 | 0.1 | -0.12 | 0.5 | 0.2 | -0.05 |
| index year: 2022 | 8.2 | 2.3 | -0.27 | 4.9 | 4.1 | -0.04 |
| condition_era group during day -30 through 0 days relative to index: Acute nephropathy | 0.3 | 1.0 | 0.09 | 0.5 | 0.7 | 0.03 |
| condition_era group during day -30 through 0 days relative to index: Acute renal impairment | 0.2 | 0.9 | 0.09 | 0.4 | 0.7 | 0.03 |
| condition_era group during day -30 through 0 days relative to index: Acute renal failure syndrome | 0.2 | 0.9 | 0.09 | 0.4 | 0.7 | 0.03 |
| condition_era group during day -30 through 0 days relative to index: Renal failure syndrome | 0.3 | 1.1 | 0.10 | 0.6 | 0.8 | 0.03 |

| (Continued) | T (%) | C (%) | SDf | T (%) | C (%) | SDf |
|---|---|---|---|---|---|---|
| observation during day -365 through 0 days relative to index: Prescription(s) generated and transmitted via a qualified erx system | 0.2 | 1.7 | 0.15 | 0.9 | 1.1 | 0.03 |
| index year: 2013 | 0.8 | 7.7 | 0.35 | 4.1 | 4.7 | 0.03 |

**Supplementary Table 223:** Most extreme patient characteristics balance diagnostics after matching for SGLT2I (female) vs DPP4I (female) in the Open Claims data source.

| | Before matching | | | After matching | | |
|---|---|---|---|---|---|---|
| Characteristic (total count = 10281) | T (%) | C (%) | SDf | T (%) | C (%) | SDf |
| age group: 75 - 79 | 5.2 | 10.8 | 0.21 | 5.7 | 5.1 | -0.03 |
| CHADS2VASc | 343.0 | 366.7 | 0.19 | 344.3 | 341.3 | -0.03 |
| index year: 2021 | 21.5 | 8.8 | -0.36 | 16.6 | 15.7 | -0.02 |
| index year: 2022 | 8.2 | 2.4 | -0.26 | 5.0 | 4.5 | -0.02 |
| drug_era group during day -365 through 0 days relative to index: evolocumab | 0.2 | 0.1 | -0.05 | 0.2 | 0.1 | -0.02 |
| index year: 2013 | 0.8 | 7.7 | 0.35 | 0.9 | 0.7 | -0.02 |
| index year: 2014 | 5.5 | 11.8 | 0.23 | 6.5 | 6.0 | -0.02 |
| condition_era group during day -365 through 0 days relative to index: Chronic kidney disease stage 3B | 0.4 | 0.1 | -0.05 | 0.3 | 0.2 | -0.02 |
| CHADS2 | 212.9 | 227.9 | 0.14 | 213.3 | 211.3 | -0.02 |
| condition_era group during day -30 through 0 days relative to index: Chronic kidney disease stage 3B | 0.2 | 0.1 | -0.04 | 0.2 | 0.1 | -0.02 |

##### 3.2.7 OEHR

**Supplementary Figure 13:** Preference score distributions, propensity score stratified (middle column) and matched (right column) covariate balance and negative control error distributions (bottom row) for SGLT2I (female) vs DPP4I (female) in the OEHR data source.

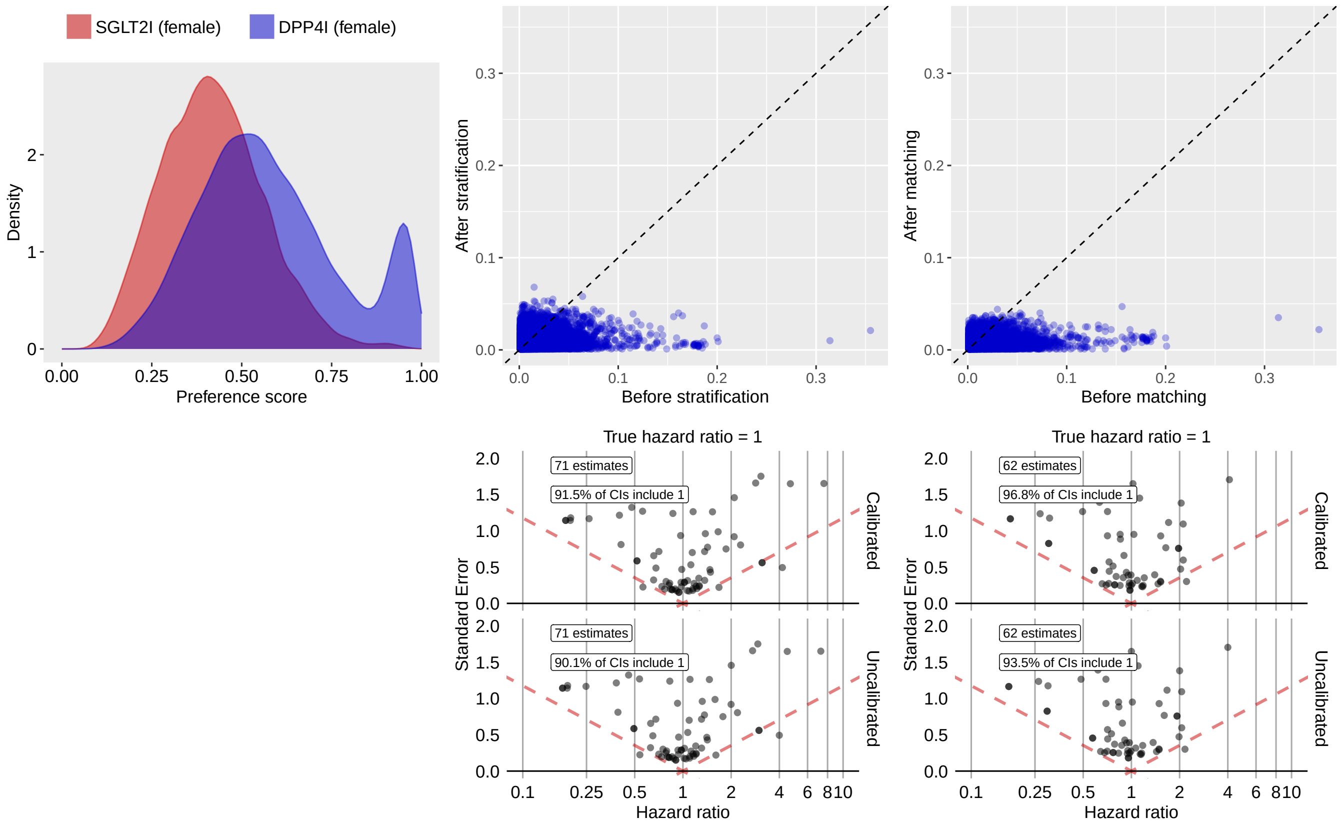

**Supplementary Table 224:** Most extreme patient characteristics balance diagnostics after stratification for SGLT2I (female) vs DPP4I (female) in the OEHR data source.

| | Before stratification | | | After stratification | | |
|---|---|---|---|---|---|---|
| Characteristic (total count = 12320) | T (%) | C (%) | SDf | T (%) | C (%) | SDf |
| measurement during day -30 through 0 days relative to index: Oral temperature | 0.9 | 0.8 | -0.01 | 1.4 | 0.7 | -0.07 |
| condition_era group during day -365 through 0 days relative to index: Chronic kidney disease stage 4 | 0.1 | 0.3 | 0.06 | 0.1 | 0.3 | 0.06 |
| condition_era group during day -365 through 0 days relative to index: Hypertrophy of tonsils | 0.2 | 0.1 | -0.03 | 0.3 | 0.1 | -0.06 |
| measurement during day -365 through 0 days relative to index: Microorganisms seen [Type] in Urine sediment by Light microscopy | 17.5 | 18.6 | 0.03 | 20.6 | 18.5 | -0.05 |
| measurement within normal range during day -365 through 0 days relative to index: Bile.microscopic observation [Identifier] in Tissue by Fouchet stain | 2.0 | 2.3 | 0.03 | 3.1 | 2.2 | -0.05 |
| drug_era group during day 0 through 0 days relative to index: doxazosin | 0.2 | 0.2 | 0.01 | 0.5 | 0.2 | -0.05 |
| drug_era group during day -30 through 0 days relative to index: doxazosin | 0.2 | 0.2 | 0.02 | 0.5 | 0.2 | -0.05 |

| (Continued) | T (%) | C (%) | SDf | T (%) | C (%) | SDf |
|---|---|---|---|---|---|---|
| condition_era group during day -365 through 0 days relative to index: Enlargement of tonsil or adenoid | 0.2 | 0.1 | -0.03 | 0.4 | 0.1 | -0.05 |
| condition_era group during day -365 through 0 days relative to index: Tonsil and/or adenoid hypertrophy | 0.2 | 0.1 | -0.03 | 0.4 | 0.1 | -0.05 |
| drug_era group during day 0 through 0 days relative to index: ANTI-ADRENERGIC AGENTS, PERIPHERALLY ACTING | 0.4 | 0.4 | -0.01 | 0.7 | 0.4 | -0.05 |

**Supplementary Table 225:** Most extreme patient characteristics balance diagnostics after matching for SGLT2I (female) vs DPP4I (female) in the OEHR data source.

| | Before matching | | | After matching | | |
|---|---|---|---|---|---|---|
| Characteristic (total count = 12133) | T (%) | C (%) | SDf | T (%) | C (%) | SDf |
| index year: 2014 | 5.1 | 9.1 | 0.16 | 5.7 | 4.6 | -0.05 |
| condition_era group during day -365 through 0 days relative to index: Elevated C-reactive protein | 0.3 | 0.2 | -0.03 | 0.3 | 0.1 | -0.04 |
| observation during day -30 through 0 days relative to index: Discharge medications reconciled with the current medication list in outpatient medical record (COA) (GER) | 0.5 | 0.1 | -0.07 | 0.4 | 0.2 | -0.04 |
| condition_era group during day -30 through 0 days relative to index: Infective pneumonia | 0.1 | 0.3 | 0.05 | 0.1 | 0.2 | 0.04 |
| condition_era group during day -30 through 0 days relative to index: Hypoxemia | 0.2 | 0.5 | 0.04 | 0.2 | 0.4 | 0.04 |
| drug_era group during day -30 through 0 days relative to index: sacubitril | 0.3 | 0.1 | -0.06 | 0.3 | 0.1 | -0.04 |
| measurement during day -365 through 0 days relative to index: Cortisol; free | 0.2 | 0.1 | -0.02 | 0.2 | 0.1 | -0.04 |
| condition_era group during day -30 through 0 days relative to index: Infectious disease of lung | 0.1 | 0.3 | 0.05 | 0.1 | 0.2 | 0.04 |
| observation during day -30 through 0 days relative to index: Most recent systolic blood pressure less than 130 mm Hg (DM), (HTN, CKD, CAD) | 2.7 | 2.0 | -0.05 | 2.6 | 3.2 | 0.04 |
| measurement during day -30 through 0 days relative to index: Lactate dehydrogenase 1/Lactate dehydrogenase.total in Urine by Electrophoresis | 0.2 | 0.4 | 0.04 | 0.2 | 0.4 | 0.04 |

##### 3.2.8 SIDIAP

**Supplementary Figure 14:** Preference score distributions, propensity score stratified (middle column) and matched (right column) covariate balance and negative control error distributions (bottom row) for SGLT2I (female) vs DPP4I (female) in the SIDIAP data source.

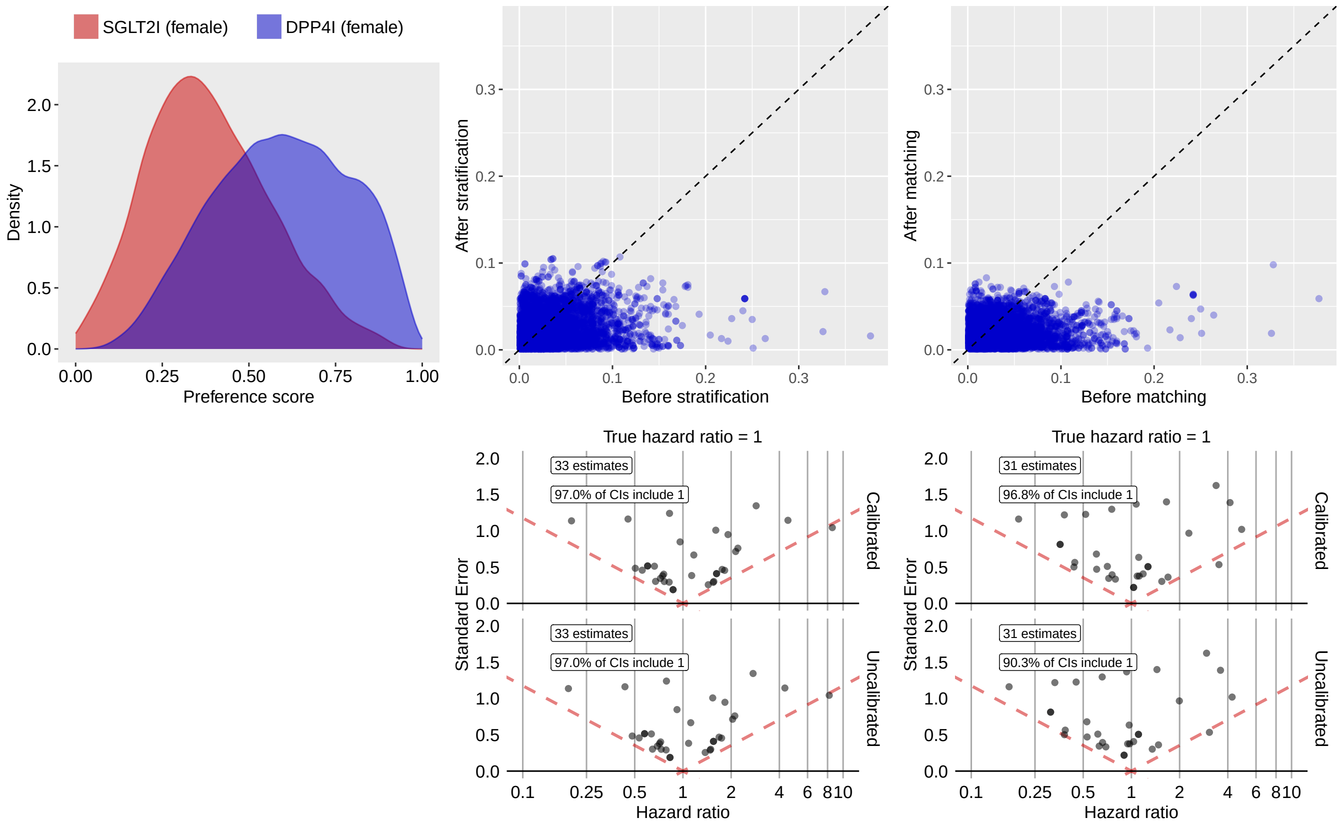

**Supplementary Table 226:** Most extreme patient characteristics balance diagnostics after stratification for SGLT2I (female) vs DPP4I (female) in the SIDIAP data source.

| | Before stratification | | | After stratification | | |
|---|---|---|---|---|---|---|
| Characteristic (total count = 6803) | T (%) | C (%) | SDf | T (%) | C (%) | SDf |
| procedure_occurrence during day -365 through 0 days relative to index: Computerized axial tomography of head | -0.2 | 0.7 | 0.11 | -0.1 | 0.6 | 0.11 |
| condition_era group during day -365 through 0 days relative to index: Meralgia paresthetica | 0.2 | 0.1 | -0.04 | 0.7 | 0.1 | -0.10 |
| condition_era group during day -365 through 0 days relative to index: Lateral cutaneous nerve of thigh lesion | 0.2 | 0.1 | -0.04 | 0.7 | 0.1 | -0.10 |
| condition_era group during day -365 through 0 days relative to index: Compression neuropathy of lower limb | 0.2 | 0.1 | -0.03 | 0.7 | 0.1 | -0.10 |
| condition_era group during day -365 through 0 days relative to index: Mononeuropathy of lower limb | 0.2 | 0.1 | -0.03 | 0.7 | 0.1 | -0.10 |
| drug_era group during day -365 through 0 days relative to index: levodopa | -0.2 | 0.7 | 0.09 | -0.1 | 0.7 | 0.10 |
| drug_era group during day 0 through 0 days relative to index: Dopa and dopa derivatives | -0.2 | 0.7 | 0.09 | -0.1 | 0.7 | 0.10 |
| drug_era group during day 0 through 0 days relative to index: levodopa | -0.2 | 0.7 | 0.09 | -0.1 | 0.7 | 0.10 |

| (Continued) | T (%) | C (%) | SDf | T (%) | C (%) | SDf |
|---|---|---|---|---|---|---|
| drug_era group during day -30 through 0 days relative to index: Dopa and dopa derivatives | -0.2 | 0.7 | 0.09 | -0.1 | 0.7 | 0.10 |
| drug_era group during day -30 through 0 days relative to index: levodopa | -0.2 | 0.7 | 0.09 | -0.1 | 0.7 | 0.10 |

**Supplementary Table 227:** Most extreme patient characteristics balance diagnostics after matching for SGLT2I (female) vs DPP4I (female) in the SIDIAP data source.

| | Before matching | | | After matching | | |
|---|---|---|---|---|---|---|
| Characteristic (total count = 6320) | T (%) | C (%) | SDf | T (%) | C (%) | SDf |
| index year: 2013 | -0.2 | 5.2 | 0.33 | -0.2 | 0.6 | 0.10 |
| condition_era group during day -365 through 0 days relative to index: Diaphragmatic hernia | 0.8 | 1.0 | 0.02 | 0.8 | 0.2 | -0.08 |
| procedure_occurrence during day -365 through 0 days relative to index: Computerized axial tomography of head | -0.2 | 0.7 | 0.11 | -0.2 | 0.3 | 0.08 |
| condition_era group during day -365 through 0 days relative to index: Venous thrombosis | 1.4 | 1.2 | -0.01 | 1.3 | 0.6 | -0.08 |
| condition_era group during day -365 through 0 days relative to index: Thrombosis of blood vessel | 1.4 | 1.3 | -0.01 | 1.4 | 0.6 | -0.08 |
| condition_era group during day -365 through 0 days relative to index: Thrombosis | 1.4 | 1.3 | -0.01 | 1.4 | 0.6 | -0.08 |
| index year: 2019 | 20.7 | 12.4 | -0.22 | 20.6 | 17.7 | -0.07 |
| procedure_occurrence during day -365 through 0 days relative to index: Transfusion of packed cells | -0.2 | 0.5 | 0.09 | -0.2 | 0.4 | 0.07 |
| condition_era group during day -365 through 0 days relative to index: Phlebitis | 1.2 | 0.9 | -0.03 | 1.1 | 0.5 | -0.07 |
| age group: 30 - 34 | 0.5 | 0.3 | -0.04 | 0.6 | -0.2 | -0.07 |

#### 3.3 SGLT2I (female) vs SU (female)

##### 3.3.1 CCAE

**Supplementary Figure 15:** Preference score distributions, propensity score stratified (middle column) and matched (right column) covariate balance and negative control error distributions (bottom row) for SGLT2I (female) vs SU (female) in the CCAE data source.

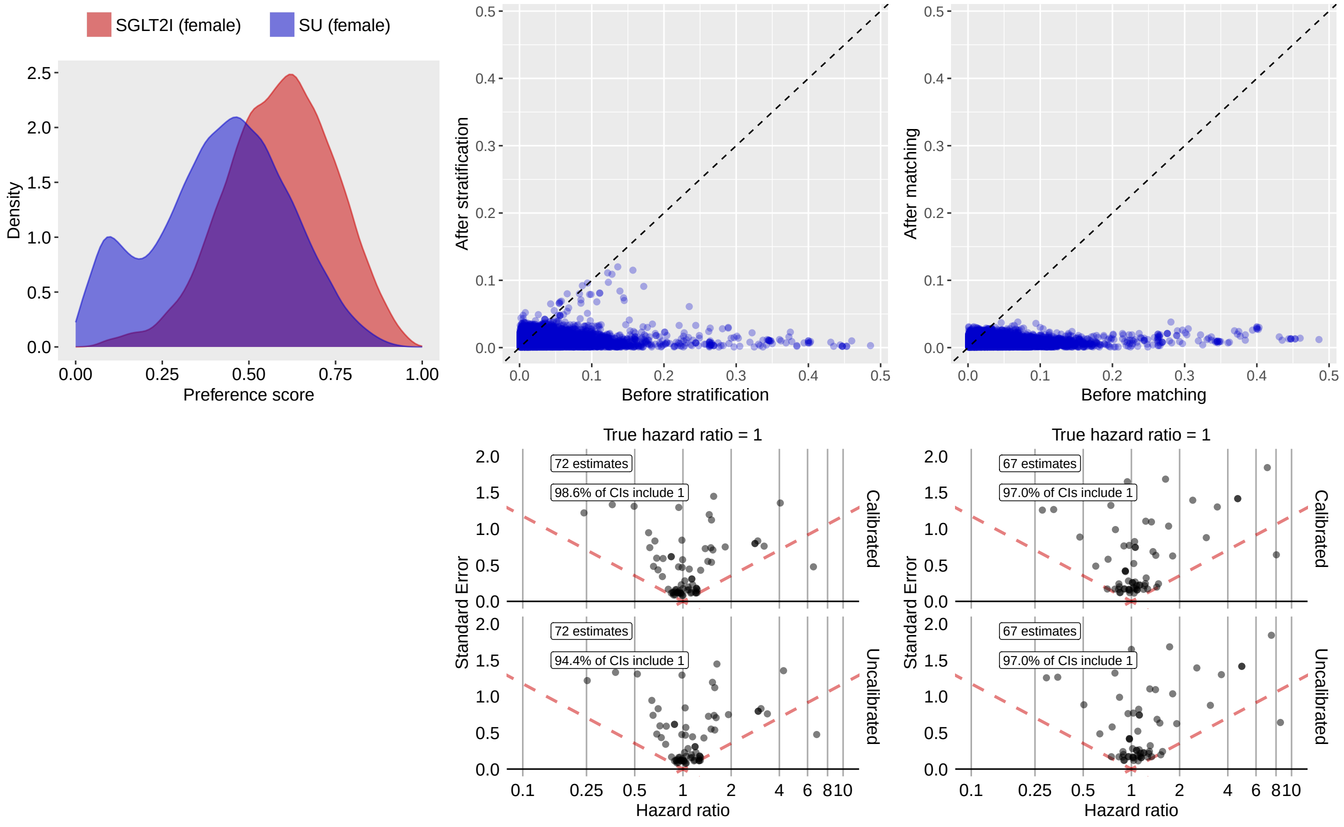

**Supplementary Table 228:** Most extreme patient characteristics balance diagnostics after stratification for SGLT2I (female) vs SU (female) in the CCAE data source.

| Characteristic (total count = 11213) | Before stratification | | | After stratification | | |
|---|---|---|---|---|---|---|
| | T (%) | C (%) | SDf | T (%) | C (%) | SDf |
| observation during day -365 through 0 days relative to index: Routine antenatal care | 0.1 | 1.2 | -0.14 | 0.1 | 1.0 | -0.12 |
| condition_era group during day -365 through 0 days relative to index: Mother delivered | 0.1 | 1.4 | -0.16 | 0.2 | 1.1 | -0.12 |
| condition_era group during day -365 through 0 days relative to index: Pregnant | 0.1 | 1.0 | -0.12 | 0.1 | 0.9 | -0.11 |
| procedure_occurrence during day -365 through 0 days relative to index: Ultrasound, pregnant uterus, real time with image documentation, transvaginal | 0.1 | 1.1 | -0.13 | 0.1 | 0.9 | -0.11 |
| condition_era group during day -365 through 0 days relative to index: Complication occurring during pregnancy | 0.1 | 1.1 | -0.12 | 0.2 | 0.9 | -0.10 |
| observation during day -365 through 0 days relative to index: Supervision of high risk pregnancy | 0.1 | 0.6 | -0.09 | 0.1 | 0.6 | -0.10 |

| (Continued) | T (%) | C (%) | SDf | T (%) | C (%) | SDf |
|---|---|---|---|---|---|---|
| condition_era group during day -365 through 0 days relative to index: High risk pregnancy | 0.1 | 0.6 | -0.09 | 0.1 | 0.6 | -0.09 |
| condition_era group during day -30 through 0 days relative to index: Finding related to pregnancy | 0.1 | 1.7 | -0.17 | 0.5 | 1.3 | -0.09 |
| condition_era group during day -365 through 0 days relative to index: Complication of pregnancy, childbirth and/or the puerperium | 0.3 | 1.6 | -0.14 | 0.5 | 1.3 | -0.08 |
| condition_era group during day -365 through 0 days relative to index: Hyperglycemic disorder in pregnancy | 0.1 | 1.0 | -0.11 | 0.3 | 0.9 | -0.08 |

**Supplementary Table 229:** Most extreme patient characteristics balance diagnostics after matching for SGLT2I (female) vs SU (female) in the CCAE data source.

| | Before matching | | | After matching | | |
|---|---|---|---|---|---|---|
| Characteristic (total count = 10976) | T (%) | C (%) | SDf | T (%) | C (%) | SDf |
| index year: 2013 | 1.3 | 6.9 | -0.28 | 1.6 | 1.1 | 0.04 |
| drug_era group during day -30 through 0 days relative to index: pitavastatin | 0.4 | 0.1 | 0.07 | 0.3 | 0.1 | 0.04 |
| condition_era group during day -30 through 0 days relative to index: Hypoglycemic disorder | 0.3 | 0.1 | 0.05 | 0.3 | 0.1 | 0.04 |
| index year: 2014 | 9.7 | 7.1 | 0.09 | 10.8 | 9.7 | 0.04 |
| procedure_occurrence during day -365 through 0 days relative to index: Ambulatory continuous glucose monitoring of interstitial tissue fluid via a subcutaneous sensor for a minimum of 72 hours; analysis, interpretation and report | 0.3 | 0.1 | 0.06 | 0.3 | 0.1 | 0.03 |
| device_exposure during day -365 through 0 days relative to index: Frames, purchases | 0.1 | 0.5 | -0.06 | 0.1 | 0.3 | -0.03 |
| condition_era group during day -365 through 0 days relative to index: Undernutrition | 25.8 | 10.6 | 0.40 | 22.6 | 23.9 | -0.03 |
| condition_era group during day -365 through 0 days relative to index: Tumor of eyelid | 0.2 | 0.3 | -0.01 | 0.2 | 0.3 | -0.03 |
| condition_era group during day -365 through 0 days relative to index: Abscess of groin | 0.1 | 0.1 | 0.00 | 0.1 | 0.2 | -0.03 |
| condition_era group during day -365 through 0 days relative to index: Deficiency of micronutrients | 25.7 | 10.5 | 0.40 | 22.5 | 23.8 | -0.03 |

##### 3.3.2 DAG

**Supplementary Figure 16:** Preference score distributions, propensity score stratified (middle column) and matched (right column) covariate balance and negative control error distributions (bottom row) for SGLT2I (female) vs SU (female) in the DAG data source.

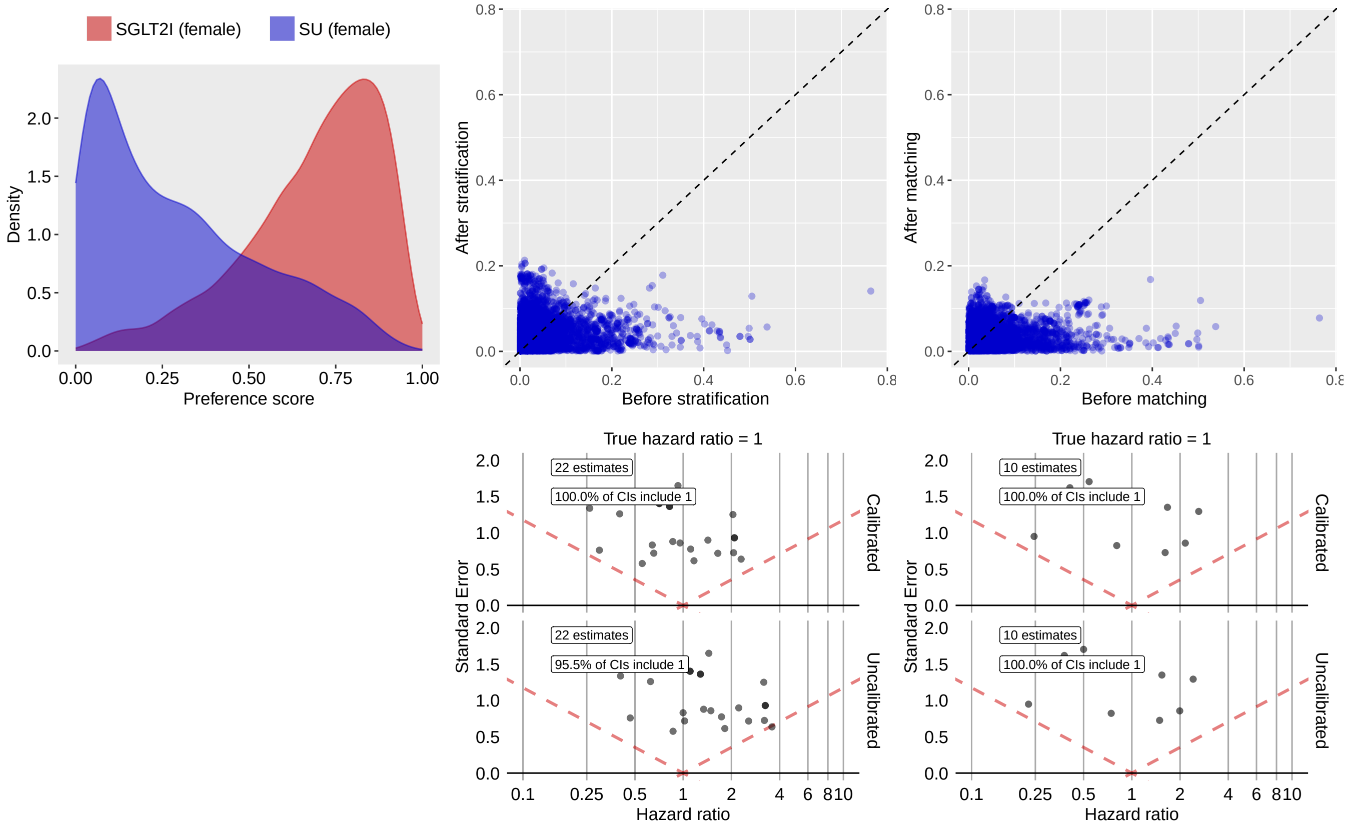

**Supplementary Table 230:** Most extreme patient characteristics balance diagnostics after stratification for SGLT2I (female) vs SU (female) in the DAG data source.

| Characteristic (total count = 6584) | Before stratification | | | After stratification | | |
|---|---|---|---|---|---|---|
| | T (%) | C (%) | SDf | T (%) | C (%) | SDf |
| condition_era group during day -365 through 0 days relative to index: Kidney stone | 0.6 | 0.7 | -0.01 | 0.5 | 3.5 | -0.21 |
| condition_era group during day -365 through 0 days relative to index: Bacterial gastrointestinal infectious disease | 0.2 | 0.2 | -0.01 | -0.1 | 2.4 | -0.21 |
| condition_era group during day -365 through 0 days relative to index: Bacterial intestinal infectious disease | 0.2 | 0.2 | -0.01 | -0.1 | 2.4 | -0.21 |
| condition_era group during day -365 through 0 days relative to index: Dysthymia | 0.2 | 0.3 | -0.00 | 0.2 | 2.6 | -0.20 |
| condition_era group during day -365 through 0 days relative to index: Bacterial infection of the digestive tract | 0.2 | 0.2 | 0.00 | 0.1 | 2.4 | -0.20 |
| condition_era group during day -365 through 0 days relative to index: Infectious enteritis | 0.2 | 0.2 | 0.00 | 0.2 | 2.3 | -0.20 |
| condition_era group during day -365 through 0 days relative to index: Generalized anxiety disorder | 0.4 | 0.8 | -0.05 | 0.4 | 2.8 | -0.20 |
| condition_era group during day -365 through 0 days relative to index: Indigestion | 0.3 | 0.6 | -0.05 | 0.3 | 2.5 | -0.19 |

| (Continued) | T (%) | C (%) | SDf | T (%) | C (%) | SDf |
|---|---|---|---|---|---|---|
| condition_era group during day -365 through 0 days relative to index: Nonulcer dyspepsia | 0.3 | 0.6 | -0.05 | 0.3 | 2.5 | -0.19 |
| device_exposure during day -365 through 0 days relative to index: INKONTINENT.BEDARF EIN-/VORLAGE | -0.2 | 0.4 | -0.07 | -0.1 | 1.8 | -0.18 |

**Supplementary Table 231:** Most extreme patient characteristics balance diagnostics after matching for SGLT2I (female) vs SU (female) in the DAG data source.

| | Before matching | | | After matching | | |
|---|---|---|---|---|---|---|
| Characteristic (total count = 5109) | T (%) | C (%) | SDf | T (%) | C (%) | SDf |
| index year: 2019 | 14.1 | 3.2 | 0.40 | 7.8 | 12.9 | -0.17 |
| drug_era group during day 0 through 0 days relative to index: Other nasal preparations | 0.7 | 0.5 | 0.04 | 1.5 | -0.2 | 0.17 |
| drug_era group during day -30 through 0 days relative to index: Other antiallergics | 0.6 | 0.5 | 0.02 | 1.2 | -0.2 | 0.15 |
| drug_era group during day -30 through 0 days relative to index: Other nasal preparations | 0.9 | 0.6 | 0.03 | 1.9 | 0.3 | 0.15 |
| drug_era group during day -365 through 0 days relative to index: ALLERGENS | 0.6 | 0.8 | -0.02 | -0.7 | 1.7 | -0.14 |
| drug_era group during day -365 through 0 days relative to index: ALLERGENS | 0.6 | 0.8 | -0.02 | -0.7 | 1.7 | -0.14 |
| drug_era group during day -365 through 0 days relative to index: Allergen extracts | 0.6 | 0.8 | -0.02 | -0.7 | 1.7 | -0.14 |
| procedure_occurrence during day -365 through 0 days relative to index: Developing a treatment plan | 0.9 | 0.7 | 0.03 | 1.5 | -0.2 | 0.14 |
| drug_era group during day -365 through 0 days relative to index: gabapentin | 1.6 | 1.3 | 0.03 | 3.2 | 1.1 | 0.14 |
| drug_era group during day 0 through 0 days relative to index: ipratropium | 0.4 | 0.3 | 0.01 | 1.0 | -0.2 | 0.14 |

##### 3.3.3 ODOD

**Supplementary Figure 17:** Preference score distributions, propensity score stratified (middle column) and matched (right column) covariate balance and negative control error distributions (bottom row) for SGLT2I (female) vs SU (female) in the ODOD data source.

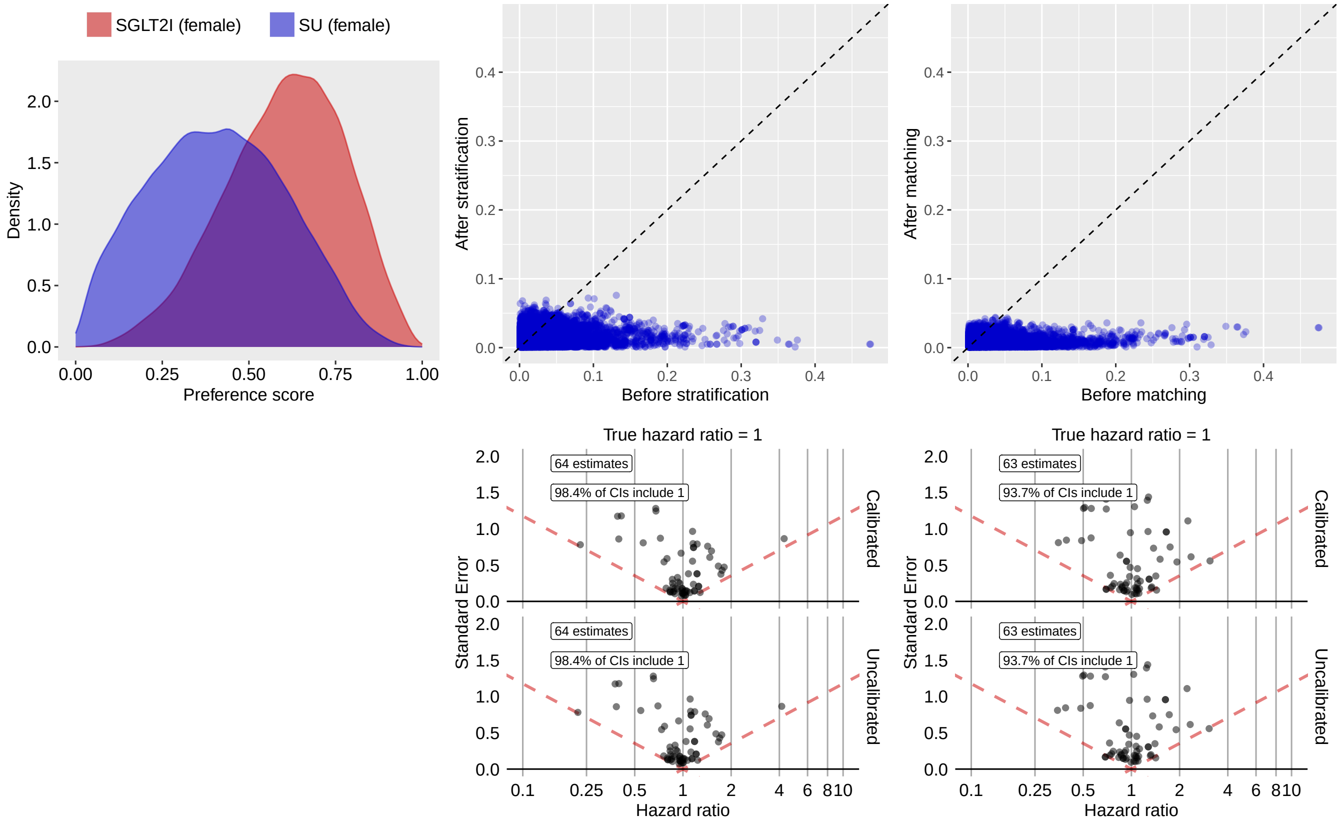

**Supplementary Table 232:** Most extreme patient characteristics balance diagnostics after stratification for SGLT2I (female) vs SU (female) in the ODOD data source.

| Characteristic (total count = 15689) | Before stratification | | | After stratification | | |
|---|---|---|---|---|---|---|
| | T (%) | C (%) | SDf | T (%) | C (%) | SDf |
| condition_era group during day -30 through 0 days relative to index: Hypertensive renal failure | 0.1 | 1.0 | -0.13 | 0.1 | 0.5 | -0.08 |
| procedure_occurrence during day -365 through 0 days relative to index: Ultrasound, pregnant uterus, real time with image documentation, transvaginal | 0.1 | 0.6 | -0.09 | 0.1 | 0.4 | -0.07 |
| condition_era group during day -365 through 0 days relative to index: Complication occurring during pregnancy | 0.1 | 0.7 | -0.10 | 0.1 | 0.5 | -0.07 |
| condition_era group during day -365 through 0 days relative to index: Idiopathic scoliosis of lumbar spine | 0.2 | 0.1 | 0.04 | 0.5 | 0.1 | 0.07 |
| condition_era group during day -365 through 0 days relative to index: Closed fracture of intracapsular section of femur | 0.1 | 0.1 | -0.00 | 0.4 | 0.1 | 0.07 |
| measurement during day -365 through 0 days relative to index: Blood group antibody screen [Presence] in Serum or Plasma | 0.1 | 0.5 | -0.07 | 0.1 | 0.4 | -0.06 |
| measurement within normal range during day -365 through 0 days relative to index: Blood group antibody screen [Presence] in Serum or Plasma | 0.1 | 0.5 | -0.07 | 0.1 | 0.4 | -0.06 |

| (Continued) | T (%) | C (%) | SDf | T (%) | C (%) | SDf |
|---|---|---|---|---|---|---|
| procedure_occurrence during day -30 through 0 days relative to index: Intravenous infusion, hydration; each additional hour (List separately in addition to code for primary procedure) | 0.8 | 1.1 | -0.04 | 2.0 | 1.2 | 0.06 |
| device_exposure during day -30 through 0 days relative to index: Low osmolar contrast material, 300-399 mg/ml iodine concentration, per ml | 1.5 | 1.5 | -0.00 | 2.5 | 1.6 | 0.06 |
| condition_era group during day -365 through 0 days relative to index: Idiopathic scoliosis | 0.3 | 0.1 | 0.04 | 0.5 | 0.2 | 0.06 |

**Supplementary Table 233:** Most extreme patient characteristics balance diagnostics after matching for SGLT2I (female) vs SU (female) in the ODOD data source.

| | Before matching | | | After matching | | |
|---|---|---|---|---|---|---|
| Characteristic (total count = 15535) | T (%) | C (%) | SDf | T (%) | C (%) | SDf |
| measurement above normal range during day -365 through 0 days relative to index: Microalbumin/Creatinine [Mass Ratio] in 24 hour Urine | 0.2 | 0.1 | 0.04 | 0.2 | 0.1 | 0.04 |
| measurement within normal range during day -365 through 0 days relative to index: Egg white IgE Ab [Units/volume] in Serum | 0.3 | 0.1 | 0.05 | 0.3 | 0.1 | 0.04 |
| measurement during day -365 through 0 days relative to index: Microalbumin/Creatinine [Mass Ratio] in 24 hour Urine | 0.3 | 0.1 | 0.04 | 0.3 | 0.1 | 0.04 |
| condition_era group during day -30 through 0 days relative to index: Ventricular tachycardia | 0.3 | 0.2 | 0.02 | 0.3 | 0.1 | 0.04 |
| measurement during day -365 through 0 days relative to index: Egg white IgE Ab [Units/volume] in Serum | 0.4 | 0.1 | 0.05 | 0.4 | 0.1 | 0.04 |
| drug_era group during day 0 through 0 days relative to index: pitavastatin | 0.3 | 0.1 | 0.05 | 0.3 | 0.1 | 0.04 |
| measurement within normal range during day -365 through 0 days relative to index: Shrimp IgE Ab [Units/volume] in Serum | 0.3 | 0.1 | 0.05 | 0.3 | 0.1 | 0.04 |
| procedure_occurrence during day -365 through 0 days relative to index: Anesthesia for all procedures on nerves, muscles, tendons, fascia, and bursae of shoulder and axilla | 0.1 | 0.3 | -0.03 | 0.1 | 0.3 | -0.04 |
| measurement during day -365 through 0 days relative to index: Hemoglobin [Mass/volume] in Urine by Automated test strip | 0.1 | 0.2 | -0.03 | 0.1 | 0.3 | -0.04 |
| measurement within normal range during day -365 through 0 days relative to index: Hemoglobin [Mass/volume] in Urine by Automated test strip | 0.1 | 0.2 | -0.03 | 0.1 | 0.3 | -0.04 |

##### 3.3.4 MDCR

**Supplementary Figure 18:** Preference score distributions, propensity score stratified (middle column) and matched (right column) covariate balance and negative control error distributions (bottom row) for SGLT2I (female) vs SU (female) in the MDCR data source.

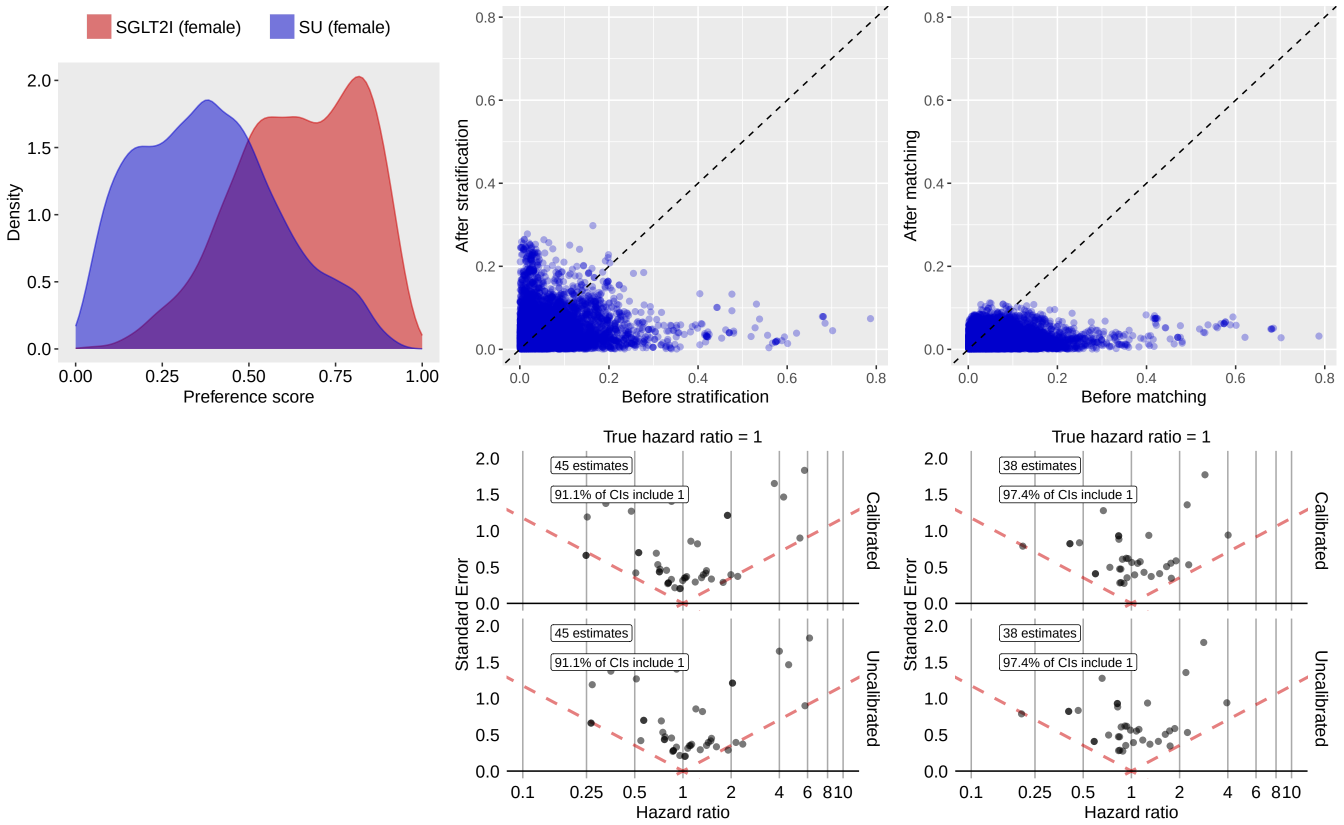

**Supplementary Table 234:** Most extreme patient characteristics balance diagnostics after stratification for SGLT2I (female) vs SU (female) in the MDCR data source.

| Characteristic (total count = 11902) | Before stratification | | | After stratification | | |
|---|---|---|---|---|---|---|
| | T (%) | C (%) | SDf | T (%) | C (%) | SDf |
| condition_era group during day -30 through 0 days relative to index: Chronic pain | 4.0 | 1.4 | 0.16 | 10.4 | 3.0 | 0.30 |
| measurement during day -30 through 0 days relative to index: Myoglobin | -0.3 | 0.1 | 0.02 | 4.1 | 0.1 | 0.28 |
| procedure_occurrence during day -365 through 0 days relative to index: Lumbar puncture | -0.3 | 0.1 | 0.01 | 3.6 | -0.1 | 0.27 |
| drug_era group during day 0 through 0 days relative to index: tolterodine | 1.2 | 1.9 | -0.05 | 5.3 | 0.8 | 0.26 |
| procedure_occurrence during day -365 through 0 days relative to index: Spinal puncture, lumbar, diagnostic | 0.3 | 0.3 | 0.01 | 3.9 | 0.2 | 0.26 |
| measurement during day -365 through 0 days relative to index: Creatine kinase (CK), (CPK); isoenzymes | -0.3 | 0.1 | 0.00 | 3.3 | -0.1 | 0.26 |
| drug_era group during day -30 through 0 days relative to index: tolterodine | 1.3 | 2.1 | -0.07 | 5.4 | 0.9 | 0.26 |

| (Continued) | T (%) | C (%) | SDf | T (%) | C (%) | SDf |
|---|---|---|---|---|---|---|
| procedure_occurrence during day -30 through 0 days relative to index: Radiologic examination, ribs, unilateral; including posteroanterior chest, minimum of 3 views | -0.3 | 0.1 | 0.02 | 3.4 | 0.1 | 0.26 |
| measurement during day -365 through 0 days relative to index: Myoglobin | 0.7 | 0.7 | 0.01 | 4.5 | 0.5 | 0.26 |
| procedure_occurrence during day -365 through 0 days relative to index: Closed treatment of distal radial fracture (eg, Colles or Smith type) or epiphyseal separation, includes closed treatment of fracture of ulnar styloid, when performed; without manipulation | 0.3 | 0.2 | 0.04 | 3.5 | 0.2 | 0.25 |

**Supplementary Table 235:** Most extreme patient characteristics balance diagnostics after matching for SGLT2I (female) vs SU (female) in the MDCR data source.

| | Before matching | | | After matching | | |
|---|---|---|---|---|---|---|
| Characteristic (total count = 10930) | T (%) | C (%) | SDf | T (%) | C (%) | SDf |
| drug_era group during day -365 through 0 days relative to index: Insulins and analogues for injection, intermediate- or long-acting combined with fast-acting | 1.0 | 0.6 | 0.05 | 1.1 | 0.2 | 0.11 |
| drug_era group during day -365 through 0 days relative to index: INSULINS AND ANALOGUES | 1.1 | 0.6 | 0.05 | 1.1 | 0.2 | 0.11 |
| condition_era group during day -30 through 0 days relative to index: Movement disorder | -0.3 | 0.5 | -0.08 | -0.4 | 0.8 | -0.11 |
| condition_era group during day -30 through 0 days relative to index: System disorder of the nervous system | -0.3 | 0.5 | -0.08 | -0.4 | 0.8 | -0.11 |
| drug_era group during day -365 through 0 days relative to index: Phenothiazines with piperazine structure | -0.3 | 1.6 | -0.15 | -0.4 | 1.0 | -0.11 |
| procedure_occurrence during day -365 through 0 days relative to index: Fluoroscopic guidance for needle placement (eg, biopsy, aspiration, injection, localization device) (List separately in addition to code for primary procedure) | 0.4 | 0.6 | -0.03 | 0.4 | 1.4 | -0.11 |
| drug_era group during day -365 through 0 days relative to index: prochlorperazine | -0.3 | 1.3 | -0.13 | -0.4 | 0.9 | -0.10 |
| condition_era group during day -30 through 0 days relative to index: Cellulitis | -0.3 | 0.9 | -0.08 | -0.4 | 1.1 | -0.10 |
| drug_era group during day -365 through 0 days relative to index: Insulins and analogues for injection, long-acting | 0.9 | 0.2 | 0.09 | 0.9 | 0.2 | 0.10 |
| drug_era group during day -365 through 0 days relative to index: Insulins and analogues for injection, fast-acting | 0.7 | 0.4 | 0.04 | 0.8 | 0.1 | 0.10 |

##### 3.3.5 MDCD

**Supplementary Figure 19:** Preference score distributions, propensity score stratified (middle column) and matched (right column) covariate balance and negative control error distributions (bottom row) for SGLT2I (female) vs SU (female) in the MDCD data source.

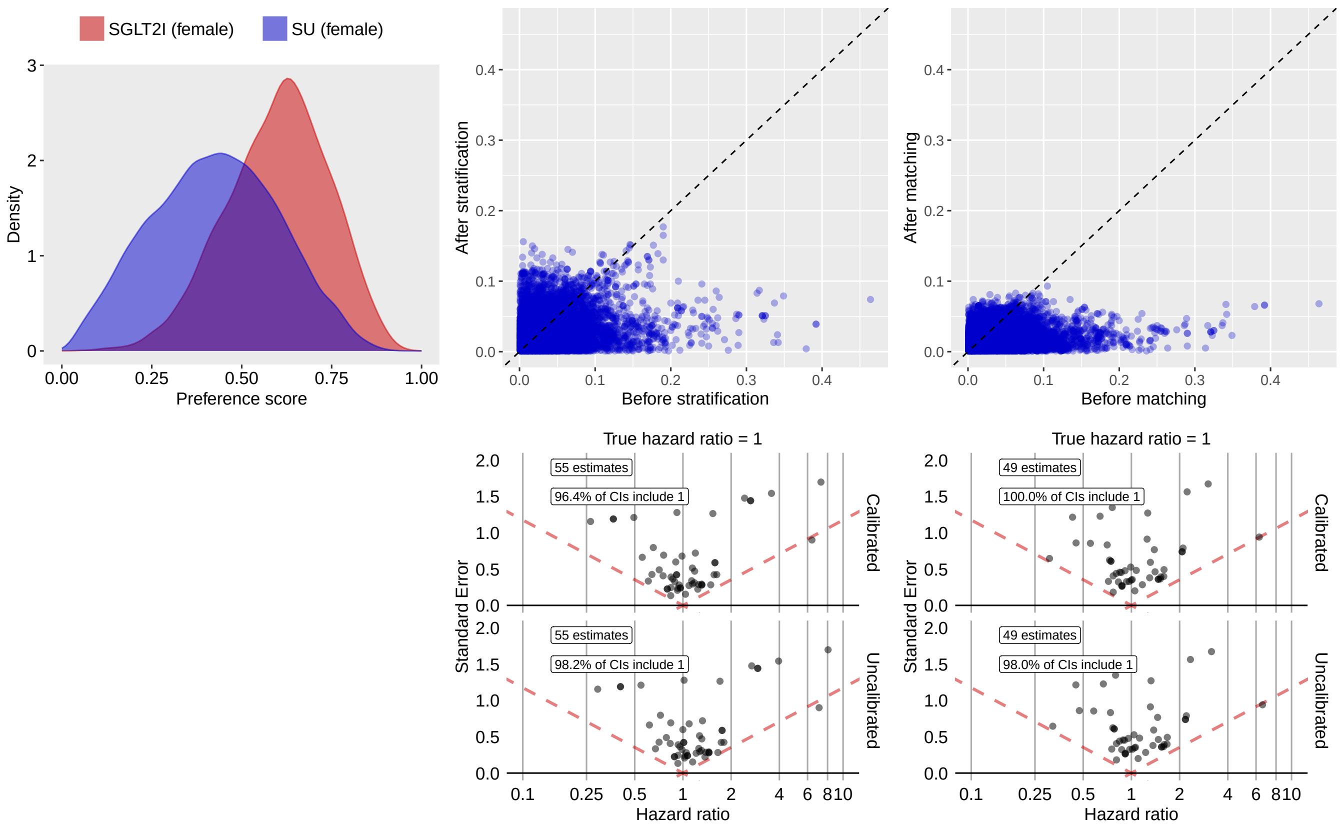

**Supplementary Table 236:** Most extreme patient characteristics balance diagnostics after stratification for SGLT2I (female) vs SU (female) in the MDCD data source.

| Characteristic (total count = 14542) | Before stratification | | | After stratification | | |
|---|---|---|---|---|---|---|
| | T (%) | C (%) | SDf | T (%) | C (%) | SDf |
| condition_era group during day -30 through 0 days relative to index: Mother delivered | -0.2 | 2.2 | -0.19 | 0.1 | 1.9 | -0.18 |
| observation during day -365 through 0 days relative to index: Routine antenatal care | 0.2 | 2.3 | -0.19 | 0.3 | 2.0 | -0.17 |
| device_exposure during day -365 through 0 days relative to index: WALGREENS ULTRA THIN LANCETS | -0.2 | 0.1 | -0.01 | 1.6 | 0.2 | 0.16 |
| observation during day -365 through 0 days relative to index: Supervision of high risk pregnancy | 0.2 | 1.6 | -0.15 | 0.2 | 1.6 | -0.15 |
| condition_era group during day -365 through 0 days relative to index: Mother delivered | 0.4 | 2.5 | -0.18 | 0.5 | 2.2 | -0.15 |
| condition_era group during day -365 through 0 days relative to index: Pregnant | 0.4 | 2.0 | -0.15 | 0.3 | 1.9 | -0.15 |
| drug_era group during day -30 through 0 days relative to index: letrozole | 0.3 | 0.2 | 0.02 | 1.7 | 0.2 | 0.15 |
| procedure_occurrence during day -365 through 0 days relative to index: Mental health service plan development by non-physician | 0.5 | 2.1 | -0.14 | 0.4 | 1.9 | -0.15 |

| (Continued) | T (%) | C (%) | SDf | T (%) | C (%) | SDf |
|---|---|---|---|---|---|---|
| index month: 9 | 6.9 | 7.4 | -0.02 | 11.5 | 7.2 | 0.15 |
| drug_era group during day -365 through 0 days relative to index: Insulins and analogues for injection, long-acting | 1.4 | 0.8 | 0.06 | 2.7 | 0.8 | 0.14 |

**Supplementary Table 237:** Most extreme patient characteristics balance diagnostics after matching for SGLT2I (female) vs SU (female) in the MDCD data source.

| | Before matching | | | After matching | | |
|---|---|---|---|---|---|---|
| Characteristic (total count = 13813) | T (%) | C (%) | SDf | T (%) | C (%) | SDf |
| device_exposure during day -365 through 0 days relative to index: OPTICHAMBER DIAMOND VHC | 1.0 | 0.2 | 0.10 | 1.0 | 0.3 | 0.09 |
| condition_era group during day -365 through 0 days relative to index: Chronic alcoholism in remission | 0.9 | 0.2 | 0.09 | 0.9 | 0.3 | 0.08 |
| procedure_occurrence during day -30 through 0 days relative to index: Radiologic examination, spine, cervical; 2 or 3 views | -0.2 | 0.4 | -0.06 | -0.2 | 0.6 | -0.08 |
| condition_era group during day -30 through 0 days relative to index: Primary malignant neoplasm of intra-abdominal organs | -0.2 | 0.6 | -0.09 | -0.2 | 0.5 | -0.08 |
| measurement during day -365 through 0 days relative to index: Zinc | 0.2 | 0.4 | -0.03 | -0.2 | 0.7 | -0.08 |
| procedure_occurrence during day -365 through 0 days relative to index: Positron emission tomography (PET) with concurrently acquired computed tomography (CT) for attenuation correction and anatomical localization imaging; skull base to mid-thigh | -0.2 | 0.6 | -0.08 | -0.2 | 0.6 | -0.08 |
| condition_era group during day -30 through 0 days relative to index: Malignant tumor of pelvis | -0.2 | 0.4 | -0.08 | -0.2 | 0.4 | -0.08 |
| condition_era group during day -30 through 0 days relative to index: Malignant neoplasm of intraabdominal organ | -0.2 | 0.8 | -0.10 | -0.2 | 0.6 | -0.08 |
| condition_era group during day -30 through 0 days relative to index: Borderline glaucoma | 0.8 | 0.4 | 0.06 | 0.8 | 0.2 | 0.08 |
| condition_era group during day -365 through 0 days relative to index: Renal function tests abnormal | -0.2 | 0.5 | -0.08 | -0.2 | 0.4 | -0.08 |

##### 3.3.6 Open Claims

**Supplementary Figure 20:** Preference score distributions, propensity score stratified (middle column) and matched (right column) covariate balance and negative control error distributions (bottom row) for SGLT2I (female) vs SU (female) in the Open Claims data source.

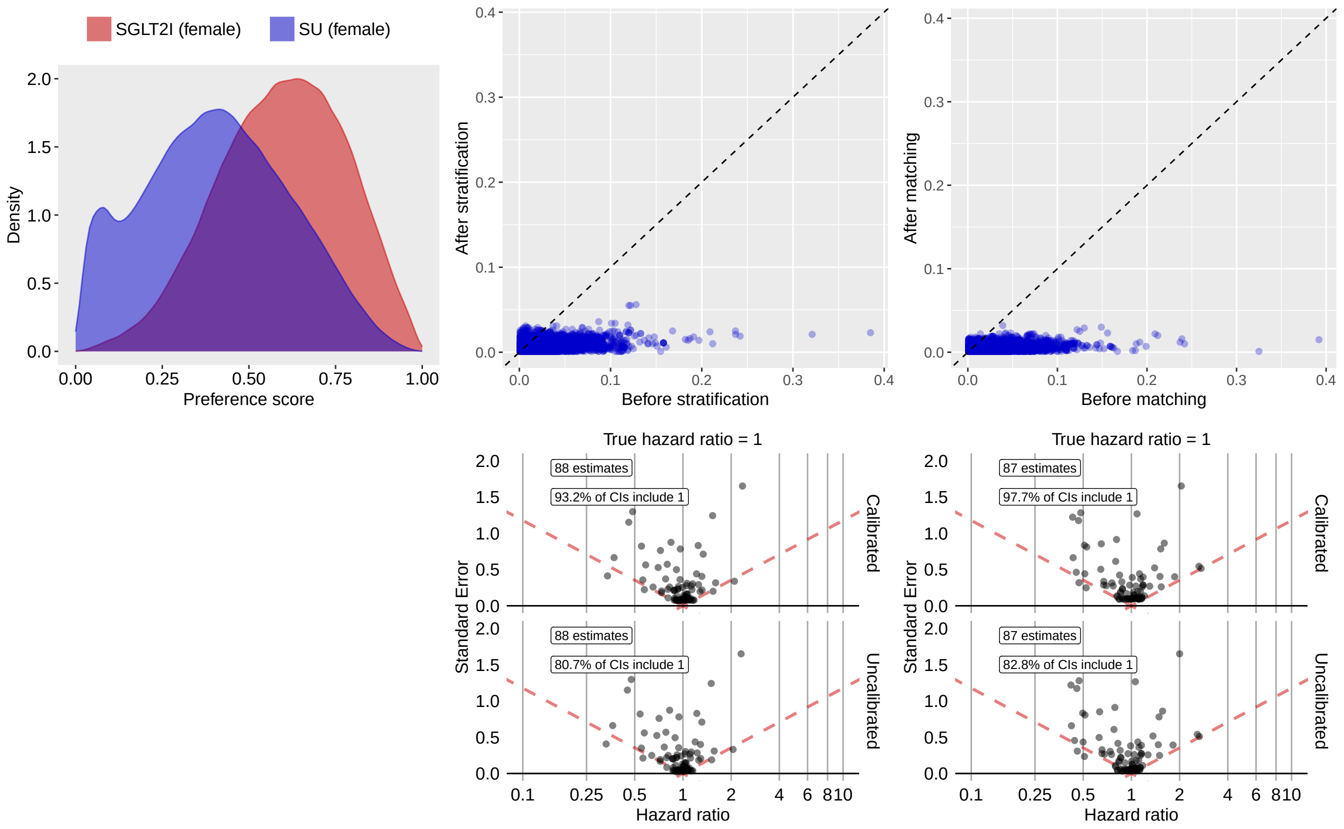

**Supplementary Table 238:** Most extreme patient characteristics balance diagnostics after stratification for SGLT2I (female) vs SU (female) in the Open Claims data source.

| | Before stratification | | | After stratification | | |
|---|---|---|---|---|---|---|
| Characteristic (total count = 10577) | T (%) | C (%) | SDf | T (%) | C (%) | SDf |
| drug_era group during day -365 through 0 days relative to index: sacubitril | 1.2 | 0.1 | 0.13 | 0.5 | 0.2 | 0.06 |
| drug_era group during day -30 through 0 days relative to index: sacubitril | 1.0 | 0.1 | 0.12 | 0.5 | 0.2 | 0.06 |
| drug_era group during day 0 through 0 days relative to index: sacubitril | 1.0 | 0.1 | 0.12 | 0.4 | 0.1 | 0.06 |
| condition_era group during day -30 through 0 days relative to index: Finding related to pregnancy | 0.1 | 0.5 | -0.09 | 0.2 | 0.4 | -0.04 |
| drug_era group during day -30 through 0 days relative to index: icosapent ethyl | 0.9 | 0.1 | 0.10 | 0.4 | 0.2 | 0.03 |
| drug_era group during day 0 through 0 days relative to index: icosapent ethyl | 0.8 | 0.1 | 0.10 | 0.4 | 0.2 | 0.03 |
| drug_era group during day -365 through 0 days relative to index: icosapent ethyl | 1.2 | 0.2 | 0.12 | 0.6 | 0.4 | 0.03 |
| condition_era group during day -365 through 0 days relative to index: Major depressive disorder | 7.9 | 6.9 | 0.04 | 7.8 | 7.0 | 0.03 |

| (Continued) | T (%) | C (%) | SDf | T (%) | C (%) | SDf |
|---|---|---|---|---|---|---|
| drug_era group during day -365 through 0 days relative to index: azilsartan | 0.2 | 0.1 | 0.04 | 0.2 | 0.1 | 0.03 |
| condition_era group during day -365 through 0 days relative to index: Mood disorder | 10.8 | 10.6 | 0.01 | 11.5 | 10.6 | 0.03 |

**Supplementary Table 239:** Most extreme patient characteristics balance diagnostics after matching for SGLT2I (female) vs SU (female) in the Open Claims data source.

| | Before matching | | | After matching | | |
|---|---|---|---|---|---|---|
| Characteristic (total count = 10167) | T (%) | C (%) | SDf | T (%) | C (%) | SDf |
| drug_era group during day -365 through 0 days relative to index: azilsartan | 0.2 | 0.1 | 0.04 | 0.2 | 0.1 | 0.03 |
| index year: 2020 | 15.2 | 10.3 | 0.15 | 15.2 | 16.3 | -0.03 |
| drug_era group during day -365 through 0 days relative to index: sacubitril | 1.2 | 0.1 | 0.13 | 0.6 | 0.4 | 0.03 |
| drug_era group during day 0 through 0 days relative to index: sacubitril | 1.0 | 0.1 | 0.12 | 0.4 | 0.3 | 0.03 |
| drug_era group during day -30 through 0 days relative to index: sacubitril | 1.0 | 0.1 | 0.12 | 0.5 | 0.3 | 0.03 |
| CHADS2VASc | 343.0 | 362.3 | -0.16 | 341.8 | 339.2 | 0.02 |
| drug_era group during day -365 through 0 days relative to index: ticagrelor | 0.5 | 0.2 | 0.05 | 0.5 | 0.3 | 0.02 |
| drug_era group during day 0 through 0 days relative to index: ticagrelor | 0.3 | 0.1 | 0.04 | 0.3 | 0.2 | 0.02 |
| age group: 75 - 79 | 5.2 | 10.8 | -0.21 | 5.3 | 4.8 | 0.02 |
| drug_era group during day -365 through 0 days relative to index: Aldosterone antagonists | 5.0 | 3.5 | 0.07 | 4.4 | 4.0 | 0.02 |

##### 3.3.7 OEHR

**Supplementary Figure 21:** Preference score distributions, propensity score stratified (middle column) and matched (right column) covariate balance and negative control error distributions (bottom row) for SGLT2I (female) vs SU (female) in the OEHR data source.

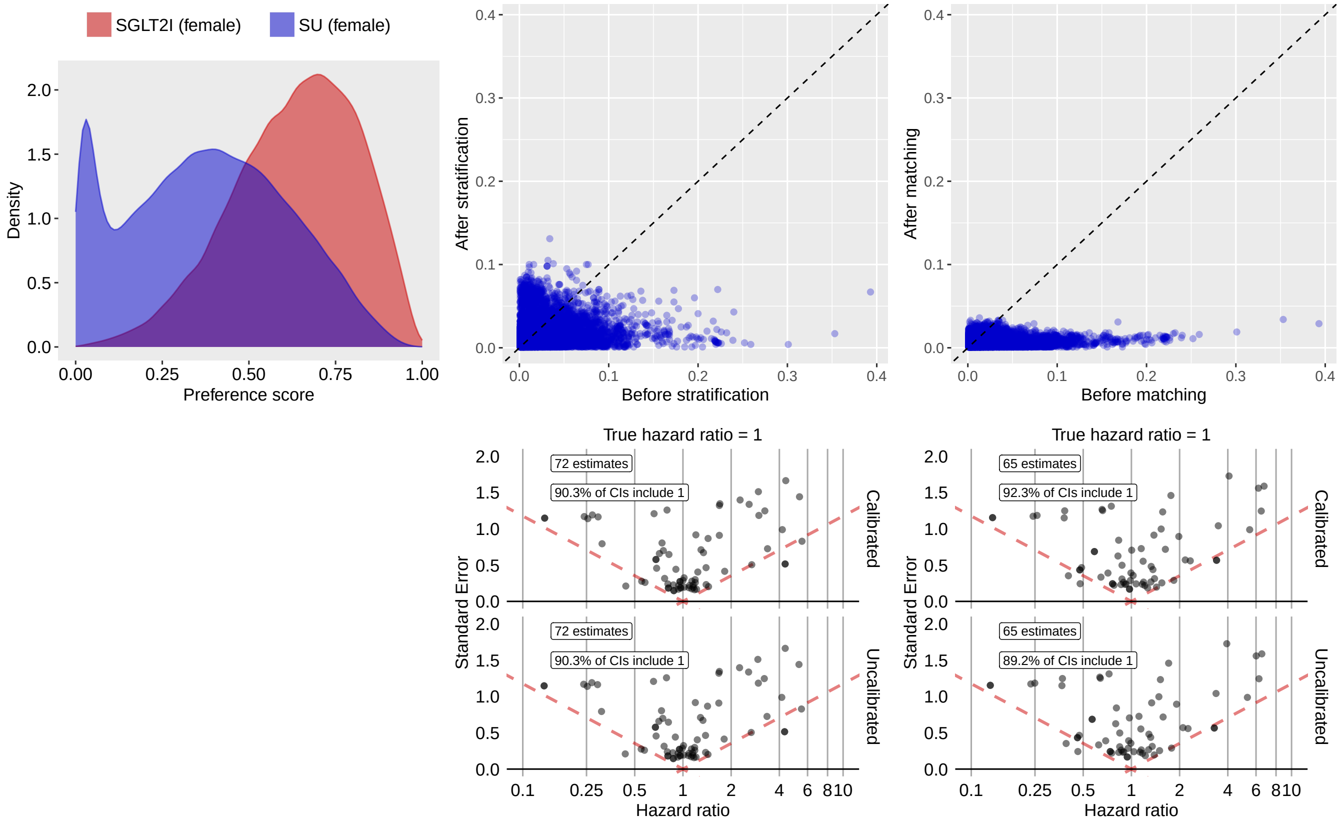

**Supplementary Table 240:** Most extreme patient characteristics balance diagnostics after stratification for SGLT2I (female) vs SU (female) in the OEHR data source.

| | Before stratification | | | After stratification | | |
|---|---|---|---|---|---|---|
| Characteristic (total count = 12087) | T (%) | C (%) | SDf | T (%) | C (%) | SDf |
| measurement within normal range during day -365 through 0 days relative to index: Bile.microscopic observation [Identifier] in Tissue by Fouchet stain | 2.0 | 2.5 | -0.03 | 4.9 | 2.4 | 0.13 |
| procedure_occurrence during day -365 through 0 days relative to index: Application of a modality to 1 or more areas; hot or cold packs | 0.6 | 0.4 | 0.03 | 1.3 | 0.4 | 0.10 |
| procedure_occurrence during day -365 through 0 days relative to index: Counseling | 0.8 | 1.1 | -0.04 | 2.4 | 1.1 | 0.10 |
| measurement during day -365 through 0 days relative to index: Bile.microscopic observation [Identifier] in Tissue by Fouchet stain | 8.3 | 10.5 | -0.08 | 12.9 | 9.8 | 0.10 |
| observation during day -365 through 0 days relative to index: Physical rehabilitation therapy procedure | 0.4 | 1.0 | -0.07 | 2.0 | 0.8 | 0.10 |
| procedure_occurrence during day -30 through 0 days relative to index: Counseling | 0.3 | 0.3 | -0.02 | 1.2 | 0.3 | 0.10 |
| condition_era group during day -365 through 0 days relative to index: Pulmonary embolism | 0.4 | 0.5 | -0.01 | 1.5 | 0.5 | 0.10 |

| (Continued) | T (%) | C (%) | SDf | T (%) | C (%) | SDf |
|---|---|---|---|---|---|---|
| condition_era group during day -365 through 0 days relative to index: Pulmonary necrosis | 0.2 | 0.3 | -0.03 | 1.1 | 0.3 | 0.10 |
| condition_era group during day -365 through 0 days relative to index: Pulmonary embolism with pulmonary infarction | 0.2 | 0.3 | -0.03 | 1.1 | 0.3 | 0.10 |
| condition_era group during day -365 through 0 days relative to index: Pulmonary infarction | 0.2 | 0.3 | -0.03 | 1.1 | 0.3 | 0.10 |

**Supplementary Table 241:** Most extreme patient characteristics balance diagnostics after matching for SGLT2I (female) vs SU (female) in the OEHR data source.

| | Before matching | | | After matching | | |
|---|---|---|---|---|---|---|
| Characteristic (total count = 11938) | T (%) | C (%) | SDf | T (%) | C (%) | SDf |
| procedure_occurrence during day -365 through 0 days relative to index: Electroretinography with interpretation and report | 0.1 | 0.1 | -0.01 | 0.1 | 0.2 | -0.04 |
| index year: 2020 | 18.7 | 7.1 | 0.35 | 17.3 | 18.5 | -0.03 |
| measurement during day -365 through 0 days relative to index: Cortisol; free | 0.2 | 0.1 | 0.04 | 0.2 | 0.1 | 0.03 |
| drug_era group during day -30 through 0 days relative to index: ANDROGENS | 0.2 | 0.1 | 0.03 | 0.2 | 0.1 | 0.03 |
| drug_era group during day -30 through 0 days relative to index: 3-oxoandrosten (4) derivatives | 0.2 | 0.1 | 0.03 | 0.2 | 0.1 | 0.03 |
| observation during day -30 through 0 days relative to index: Functional status assessed (COA) (RA) | 0.1 | 0.1 | -0.00 | 0.1 | 0.2 | -0.03 |
| drug_era group during day 0 through 0 days relative to index: ANDROGENS | 0.2 | 0.1 | 0.04 | 0.2 | 0.1 | 0.03 |
| drug_era group during day 0 through 0 days relative to index: 3-oxoandrosten (4) derivatives | 0.2 | 0.1 | 0.04 | 0.2 | 0.1 | 0.03 |
| drug_era group during day -365 through 0 days relative to index: febuxostat | 0.2 | 0.1 | 0.02 | 0.2 | 0.1 | 0.03 |
| index year: 2014 | 5.1 | 9.5 | -0.17 | 5.5 | 4.8 | 0.03 |

##### 3.3.8 SIDIAP

**Supplementary Figure 22:** Preference score distributions, propensity score stratified (middle column) and matched (right column) covariate balance and negative control error distributions (bottom row) for SGLT2I (female) vs SU (female) in the SIDIAP data source.

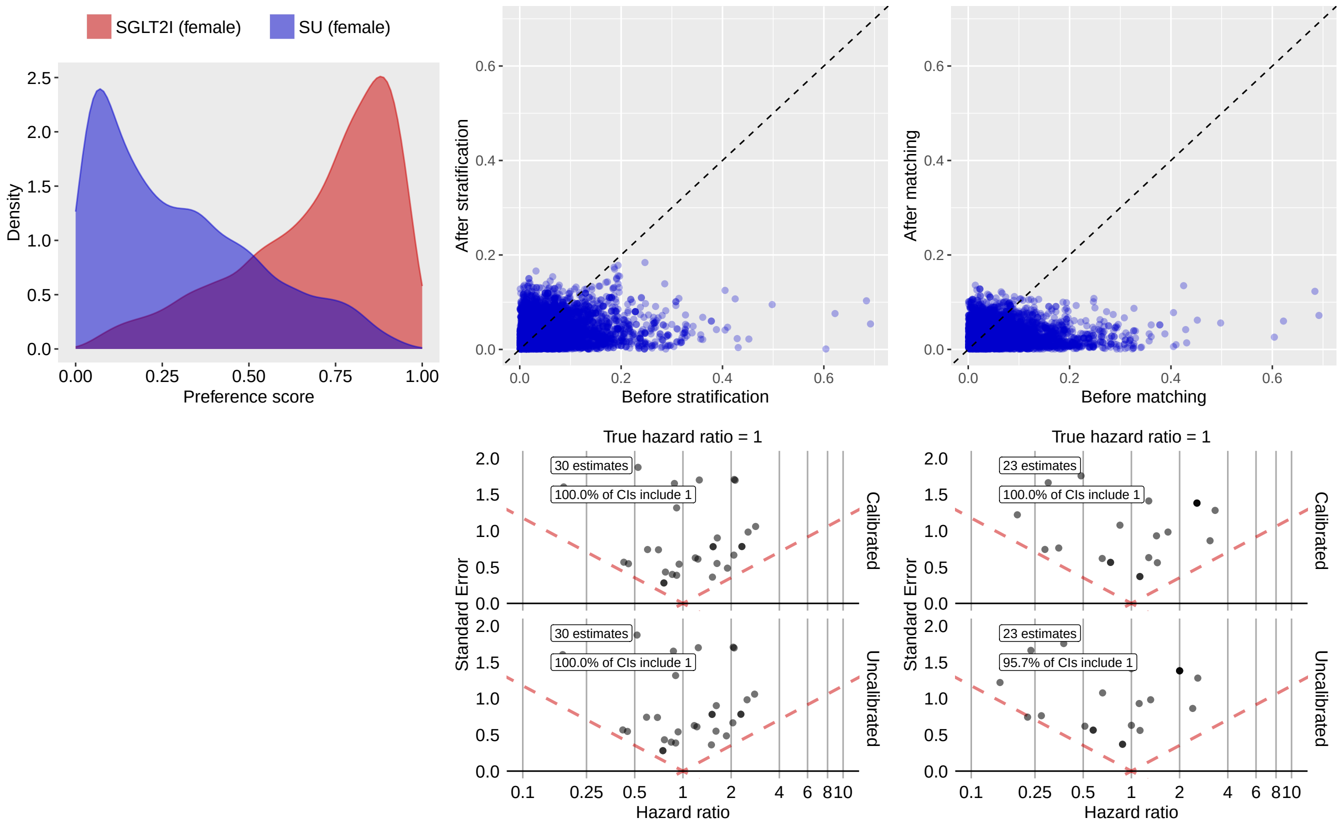

**Supplementary Table 242:** Most extreme patient characteristics balance diagnostics after stratification for SGLT2I (female) vs SU (female) in the SIDIAP data source.

| Characteristic (total count = 7187) | Before stratification | | | After stratification | | |
|---|---|---|---|---|---|---|
| | T (%) | C (%) | SDf | T (%) | C (%) | SDf |
| procedure_occurrence during day -365 through 0 days relative to index: Ultrasonography of Right and Left Heart | 3.0 | -0.1 | 0.25 | 2.0 | -0.2 | 0.18 |
| drug_era group during day -30 through 0 days relative to index: Aldosterone antagonists | 5.2 | 1.7 | 0.19 | 4.0 | 1.2 | 0.18 |
| drug_era group during day -365 through 0 days relative to index: Aldosterone antagonists | 5.5 | 2.0 | 0.19 | 4.3 | 1.4 | 0.17 |
| drug_era group during day 0 through 0 days relative to index: Aldosterone antagonists | 4.9 | 1.6 | 0.19 | 3.8 | 1.2 | 0.17 |
| drug_era group during day -365 through 0 days relative to index: solifenacin | 1.3 | 1.0 | 0.03 | 2.4 | 0.4 | 0.17 |
| drug_era group during day 0 through 0 days relative to index: rosuvastatin | 2.9 | 0.4 | 0.19 | 2.1 | 0.4 | 0.16 |
| drug_era group during day -365 through 0 days relative to index: rosuvastatin | 3.5 | 0.7 | 0.20 | 2.4 | 0.6 | 0.15 |
| age group: 90 - 94 | 0.5 | 1.1 | -0.07 | 0.4 | 2.1 | -0.15 |

| (Continued) | T (%) | C (%) | SDf | T (%) | C (%) | SDf |
|---|---|---|---|---|---|---|
| drug_era group during day -30 through 0 days relative to index: rosuvastatin | 3.0 | 0.5 | 0.19 | 2.2 | 0.5 | 0.15 |
| condition_era group during day -30 through 0 days relative to index: Thyrotoxicosis | 0.4 | 0.3 | 0.02 | 0.4 | 2.0 | -0.15 |

**Supplementary Table 243:** Most extreme patient characteristics balance diagnostics after matching for SGLT2I (female) vs SU (female) in the SIDIAP data source.

| | Before matching | | | After matching | | |
|---|---|---|---|---|---|---|
| Characteristic (total count = 6078) | T (%) | C (%) | SDf | T (%) | C (%) | SDf |
| drug_era group during day -365 through 0 days relative to index: Oxicams | 0.9 | 1.0 | -0.01 | 1.5 | 0.2 | 0.14 |
| index year: 2013 | -0.2 | 8.4 | -0.42 | -0.5 | 1.2 | -0.14 |
| condition_era group during day -365 through 0 days relative to index: Fever | 0.8 | 0.6 | 0.02 | -0.5 | 1.1 | -0.13 |
| condition_era group during day -365 through 0 days relative to index: Abnormal body temperature | 0.8 | 0.6 | 0.02 | -0.5 | 1.1 | -0.13 |
| condition_era group during day -365 through 0 days relative to index: Body temperature above reference range | 0.8 | 0.6 | 0.02 | -0.5 | 1.1 | -0.13 |
| drug_era group during day -365 through 0 days relative to index: meloxicam | 0.5 | 0.4 | 0.02 | 0.9 | -0.1 | 0.13 |
| index year: 2021 | 21.6 | 1.1 | 0.68 | 5.4 | 8.5 | -0.12 |
| age group: 90 - 94 | 0.5 | 1.1 | -0.07 | -0.5 | 1.4 | -0.12 |
| condition_era group during day -30 through 0 days relative to index: Musculoskeletal disorder of the neck | 0.2 | 0.6 | -0.07 | -0.5 | 1.0 | -0.12 |
| drug_era group during day -365 through 0 days relative to index: telmisartan | 1.3 | 1.8 | -0.04 | 1.6 | 0.4 | 0.12 |

#### 3.4 GLP1RA (female) vs DPP4I (female)

##### 3.4.1 CCAE

**Supplementary Figure 23:** Preference score distributions, propensity score stratified (middle column) and matched (right column) covariate balance and negative control error distributions (bottom row) for GLP1RA (female) vs DPP4I (female) in the CCAE data source.

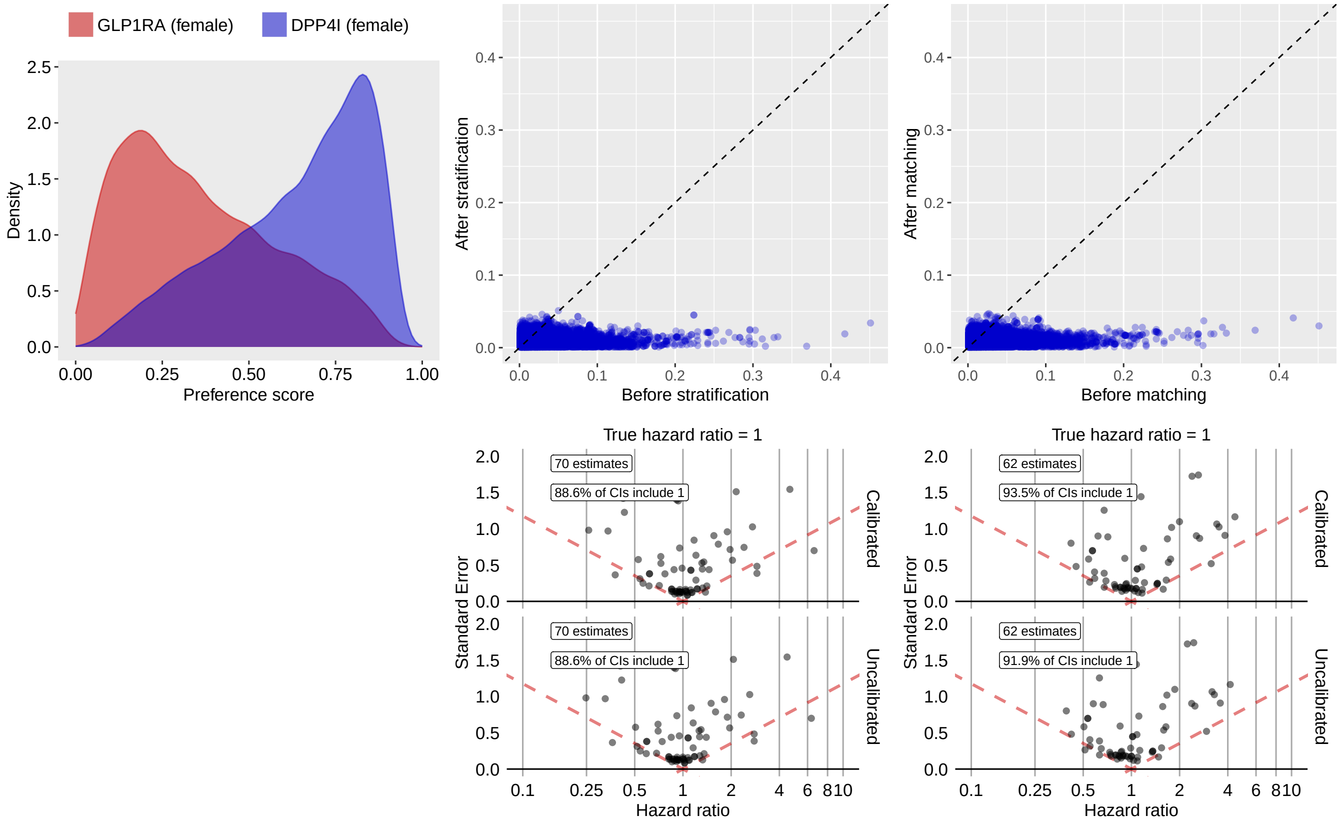

**Supplementary Table 244:** Most extreme patient characteristics balance diagnostics after stratification for GLP1RA (female) vs DPP4I (female) in the CCAE data source.

| | Before stratification | | | After stratification | | |
|---|---|---|---|---|---|---|
| Characteristic (total count = 11788) | T (%) | C (%) | SDf | T (%) | C (%) | SDf |
| procedure_occurrence during day -365 through 0 days relative to index: Fluoroscopic guidance for central venous access device placement, replacement (catheter only or complete), or removal (includes fluoroscopic guidance for vascular access and catheter manipulation, any necessary contrast injections through access site or c | 0.1 | 0.3 | 0.05 | 0.1 | 0.3 | 0.05 |
| procedure_occurrence during day -365 through 0 days relative to index: Ultrasound guidance for vascular access requiring ultrasound evaluation of potential access sites, documentation of selected vessel patency, concurrent realtime ultrasound visualization of vascular needle entry, with permanent recording and reporting (Lis | 0.2 | 0.4 | 0.03 | 0.2 | 0.4 | 0.05 |
| condition_era group during day -30 through 0 days relative to index: Benign hypertension | 4.6 | 10.4 | 0.22 | 9.8 | 8.6 | -0.04 |
| condition_era group during day -30 through 0 days relative to index: Benign essential hypertension | 4.6 | 10.4 | 0.22 | 9.8 | 8.5 | -0.04 |

| (Continued) | T (%) | C (%) | SDf | T (%) | C (%) | SDf |
|---|---|---|---|---|---|---|
| measurement during day -365 through 0 days relative to index: Cell Fractions/Differential [Interpretation] in Blood | 0.2 | 0.6 | 0.07 | 0.2 | 0.5 | 0.04 |
| measurement within normal range during day -365 through 0 days relative to index: Cell Fractions/Differential [Interpretation] in Blood | 0.2 | 0.6 | 0.07 | 0.2 | 0.5 | 0.04 |
| condition_era group during day -30 through 0 days relative to index: Respiratory failure | 0.1 | 0.2 | 0.04 | 0.1 | 0.2 | 0.04 |
| condition_era group during day -30 through 0 days relative to index: Respiratory insufficiency | 0.1 | 0.2 | 0.04 | 0.1 | 0.2 | 0.04 |
| device_exposure during day -30 through 0 days relative to index: ACCU-CHEK MULTICLIX LANCETS | 0.3 | 0.4 | 0.01 | 0.7 | 0.4 | -0.04 |
| procedure_occurrence during day -365 through 0 days relative to index: Insertion of tunneled centrally inserted central venous access device, with subcutaneous port; age 5 years or older | 0.1 | 0.3 | 0.04 | 0.1 | 0.2 | 0.04 |

**Supplementary Table 245:** Most extreme patient characteristics balance diagnostics after matching for GLP1RA (female) vs DPP4I (female) in the CCAE data source.

| | Before matching | | | After matching | | |
|---|---|---|---|---|---|---|
| Characteristic (total count = 11812) | T (%) | C (%) | SDf | T (%) | C (%) | SDf |
| procedure_occurrence during day -30 through 0 days relative to index: Blood typing, serologic; ABO | 0.2 | 0.3 | 0.03 | 0.1 | 0.4 | 0.05 |
| drug_era group during day -365 through 0 days relative to index: ALIMENTARY TRACT AND METABOLISM | 98.9 | 99.1 | 0.03 | 98.8 | 99.3 | 0.05 |
| procedure_occurrence during day -30 through 0 days relative to index: Antibody screen, RBC, each serum technique | 0.1 | 0.2 | 0.03 | 0.1 | 0.3 | 0.04 |
| condition_era group during day -30 through 0 days relative to index: Hypertensive crisis | 0.2 | 0.5 | 0.06 | 0.2 | 0.4 | 0.04 |
| procedure_occurrence during day -30 through 0 days relative to index: Blood typing, serologic; Rh (D) | 0.2 | 0.3 | 0.02 | 0.2 | 0.4 | 0.04 |
| condition_era group during day -30 through 0 days relative to index: Female genital organ symptoms | 0.1 | 0.3 | 0.04 | 0.1 | 0.3 | 0.04 |
| observation during day -365 through 0 days relative to index: Body mass index 20-24 - normal | 0.2 | 0.3 | 0.01 | 0.2 | 0.5 | 0.04 |
| condition_era group during day -30 through 0 days relative to index: Respiratory failure | 0.1 | 0.2 | 0.04 | 0.1 | 0.3 | 0.04 |
| procedure_occurrence during day -365 through 0 days relative to index: Computer-aided detection (computer algorithm analysis of digital image data for lesion detection) with further review for interpretation, with or without digitization of film radiographic images; screening mammography (List separately in addition to code | 12.8 | 29.6 | 0.42 | 17.6 | 16.1 | -0.04 |
| drug_era group during day -365 through 0 days relative to index: DRUGS USED IN DIABETES | 95.3 | 97.1 | 0.09 | 95.9 | 96.7 | 0.04 |

##### 3.4.2 ODOD

**Supplementary Figure 24:** Preference score distributions, propensity score stratified (middle column) and matched (right column) covariate balance and negative control error distributions (bottom row) for GLP1RA (female) vs DPP4I (female) in the ODOD data source.

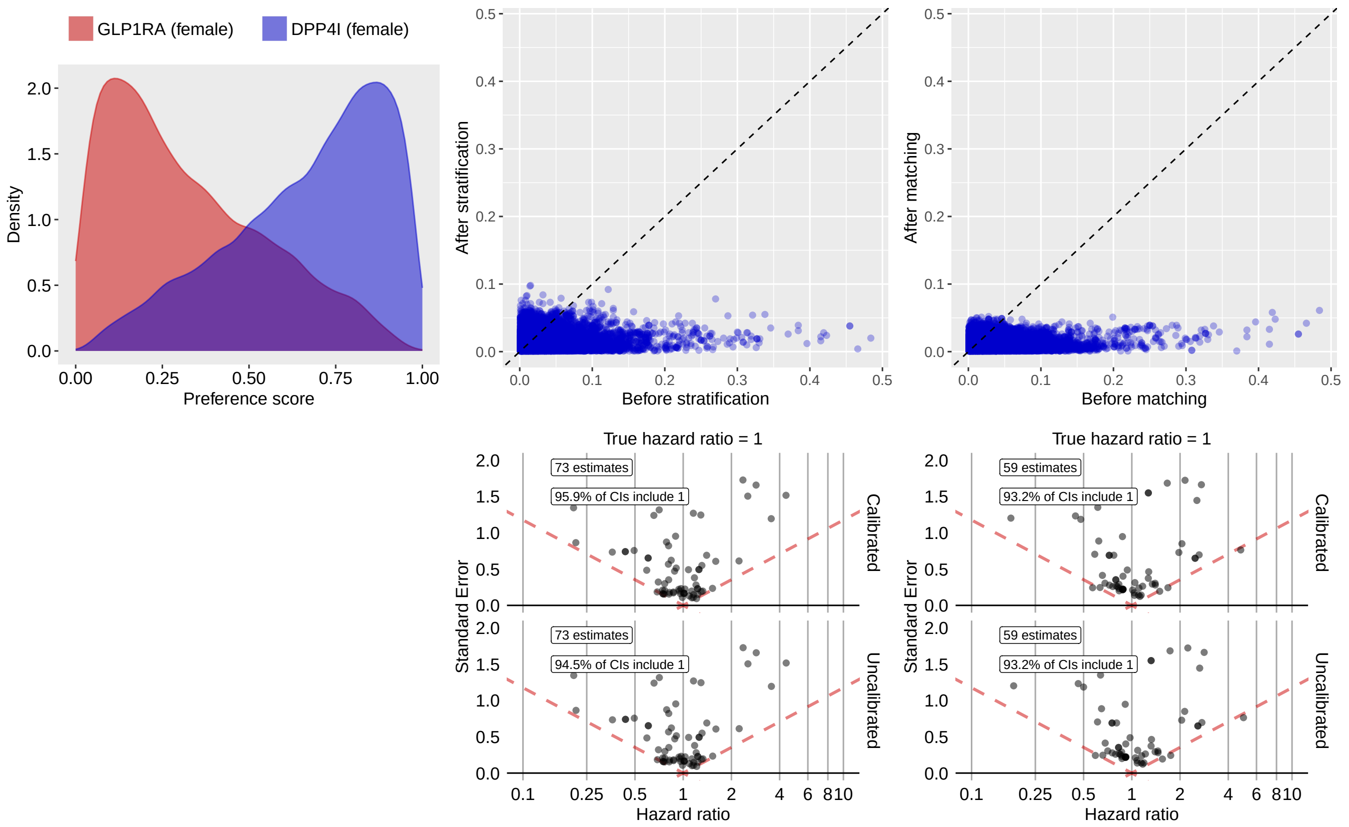

**Supplementary Table 246:** Most extreme patient characteristics balance diagnostics after stratification for GLP1RA (female) vs DPP4I (female) in the ODOD data source.

| Characteristic (total count = 15839) | Before stratification | | | After stratification | | |
|---|---|---|---|---|---|---|
| | T (%) | C (%) | SDf | T (%) | C (%) | SDf |
| measurement within normal range during day -365 through 0 days relative to index: Erythrocyte distribution width [Ratio] | 0.1 | 0.1 | 0.01 | 0.7 | 0.1 | -0.10 |
| measurement during day -365 through 0 days relative to index: Erythrocyte distribution width [Ratio] | 0.1 | 0.1 | 0.01 | 0.7 | 0.1 | -0.10 |
| condition_era group during day -365 through 0 days relative to index: Hypertensive renal failure | 0.3 | 1.5 | 0.12 | 2.3 | 1.1 | -0.09 |
| measurement during day -365 through 0 days relative to index: Eye disorder screening | 0.1 | 0.2 | 0.03 | 0.6 | 0.1 | -0.08 |
| observation during day -30 through 0 days relative to index: H/O: pneumonia | 0.1 | 0.1 | -0.01 | 0.5 | 0.1 | -0.08 |
| condition_era group during day -30 through 0 days relative to index: Multiple valve disease | 0.2 | 0.2 | 0.01 | 0.7 | 0.2 | -0.08 |
| condition_era group during day -365 through 0 days relative to index: Hypertensive heart AND renal disease | 0.8 | 1.4 | 0.06 | 2.2 | 1.2 | -0.08 |
| age group: 80 - 84 | 0.9 | 5.8 | 0.27 | 2.9 | 4.4 | 0.08 |

| (Continued) | T (%) | C (%) | SDf | T (%) | C (%) | SDf |
|---|---|---|---|---|---|---|
| procedure_occurrence during day -365 through 0 days relative to index: Closed treatment of distal radial fracture (eg, Colles or Smith type) or epiphyseal separation, includes closed treatment of fracture of ulnar styloid, when performed; without manipulation | 0.1 | 0.1 | 0.01 | 0.5 | 0.1 | -0.08 |
| condition_era group during day -365 through 0 days relative to index: Ulcerative stomatitis | 0.1 | 0.1 | -0.00 | 0.4 | 0.1 | -0.08 |

**Supplementary Table 247:** Most extreme patient characteristics balance diagnostics after matching for GLP1RA (female) vs DPP4I (female) in the ODOD data source.

| | Before matching | | | After matching | | |
|---|---|---|---|---|---|---|
| Characteristic (total count = 16329) | T (%) | C (%) | SDf | T (%) | C (%) | SDf |
| condition_era group during day -365 through 0 days relative to index: Obesity | 46.8 | 24.3 | -0.48 | 36.8 | 39.8 | 0.06 |
| index year: 2020 | 25.0 | 9.5 | -0.42 | 21.4 | 19.1 | -0.06 |
| condition_era group during day -30 through 0 days relative to index: Old myocardial infarction | 0.3 | 0.5 | 0.03 | 0.4 | 0.1 | -0.05 |
| condition_era group during day -365 through 0 days relative to index: Disorder due to type 2 diabetes mellitus | 43.2 | 33.5 | -0.20 | 41.8 | 44.3 | 0.05 |
| measurement during day -365 through 0 days relative to index: Immunoassay for tumor antigen, quantitative; CA 125 | 1.2 | 1.0 | -0.02 | 1.2 | 0.7 | -0.05 |
| condition_era group during day -365 through 0 days relative to index: Centriacinar emphysema | 0.2 | 0.3 | 0.01 | 0.2 | 0.5 | 0.05 |
| procedure_occurrence during day -365 through 0 days relative to index: Acoustic immittance testing, includes tympanometry (impedance testing), acoustic reflex threshold testing, and acoustic reflex decay testing | 0.1 | 0.1 | -0.01 | 0.2 | -0.1 | -0.05 |
| drug_era group during day -365 through 0 days relative to index: aliskiren | 0.1 | 0.3 | 0.05 | 0.1 | 0.4 | 0.05 |
| drug_era group during day -365 through 0 days relative to index: OTHER AGENTS ACTING ON THE RENIN-ANGIOTENSIN SYSTEM | 0.1 | 0.3 | 0.05 | 0.1 | 0.4 | 0.05 |
| drug_era group during day -365 through 0 days relative to index: Renin-inhibitors | 0.1 | 0.3 | 0.05 | 0.1 | 0.4 | 0.05 |

##### 3.4.3 MDCR

**Supplementary Figure 25:** Preference score distributions, propensity score stratified (middle column) and matched (right column) covariate balance and negative control error distributions (bottom row) for GLP1RA (female) vs DPP4I (female) in the MDCR data source.

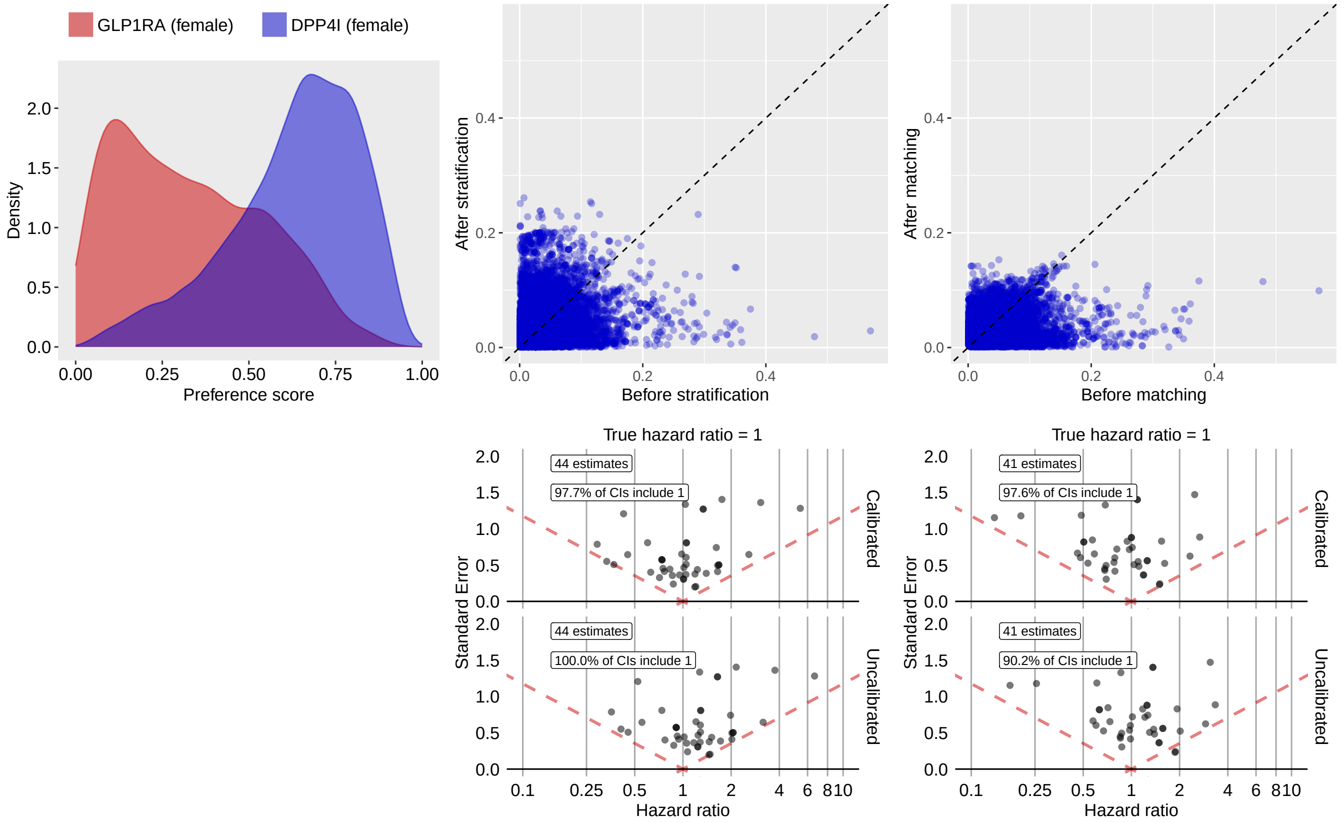

**Supplementary Table 248:** Most extreme patient characteristics balance diagnostics after stratification for GLP1RA (female) vs DPP4I (female) in the MDCR data source.

| Characteristic (total count = 15082) | Before stratification | | | After stratification | | |
|---|---|---|---|---|---|---|
| | T (%) | C (%) | SDf | T (%) | C (%) | SDf |
| observation during day -365 through 0 days relative to index: Aspirin or clopidogrel prescribed or currently being taken (CAD) | -0.4 | 0.4 | 0.01 | 4.3 | 0.4 | -0.26 |
| condition_era group during day -365 through 0 days relative to index: Cerebral infarction due to thrombosis of cerebral arteries | 0.7 | 2.1 | 0.12 | 7.2 | 1.9 | -0.25 |
| condition_era group during day -365 through 0 days relative to index: Thrombotic infarction | 0.7 | 2.1 | 0.12 | 7.2 | 1.9 | -0.25 |
| condition_era group during day -365 through 0 days relative to index: Chronic diastolic heart failure | 2.2 | 2.2 | 0.00 | 7.6 | 2.2 | -0.25 |
| condition_era group during day -365 through 0 days relative to index: Acute diastolic heart failure | 0.8 | 1.5 | 0.06 | 5.9 | 1.5 | -0.24 |
| condition_era group during day -365 through 0 days relative to index: Diastolic heart failure | 2.6 | 3.2 | 0.03 | 8.7 | 3.1 | -0.24 |
| observation during day -30 through 0 days relative to index: Patient encounter was documented using an ehr system that has been certified by an authorized testing and certification body (atcb) | 0.4 | 0.6 | 0.03 | 4.2 | 0.6 | -0.24 |
| index year: 2014 | 3.2 | 10.5 | 0.29 | 4.1 | 10.0 | 0.23 |

| (Continued) | T (%) | C (%) | SDf | T (%) | C (%) | SDf |
|---|---|---|---|---|---|---|
| device_exposure during day -365 through 0 days relative to index: Anchor/screw for opposing bone-to-bone or soft tissue-to-bone (implantable) | 1.5 | 0.3 | -0.13 | 3.4 | 0.3 | -0.23 |
| condition_era group during day -365 through 0 days relative to index: Diastolic dysfunction | 2.7 | 3.3 | 0.04 | 8.7 | 3.3 | -0.23 |

**Supplementary Table 249:** Most extreme patient characteristics balance diagnostics after matching for GLP1RA (female) vs DPP4I (female) in the MDCR data source.

| | Before matching | | | After matching | | |
|---|---|---|---|---|---|---|
| Characteristic (total count = 10615) | T (%) | C (%) | SDf | T (%) | C (%) | SDf |
| measurement during day -365 through 0 days relative to index: Globulin [Mass/volume] in Serum by calculation | 0.5 | 2.2 | 0.15 | 0.6 | 2.6 | 0.16 |
| observation during day -365 through 0 days relative to index: Most recent systolic blood pressure 130-139 mm Hg (DM),(HTN, CKD, CAD) | 2.0 | 1.4 | -0.05 | 1.7 | 4.4 | 0.15 |
| measurement during day -365 through 0 days relative to index: Albumin/Globulin [Mass Ratio] in Serum or Plasma | 0.6 | 2.3 | 0.14 | 0.8 | 2.7 | 0.15 |
| observation during day -365 through 0 days relative to index: Hospital outpatient clinic visit for assessment and management of a patient | 13.8 | 7.5 | -0.21 | 12.7 | 18.0 | 0.14 |
| condition_era group during day -30 through 0 days relative to index: Kidney lesion | 2.1 | 1.6 | -0.04 | 2.3 | 0.6 | -0.14 |
| age group: 90 - 94 | -0.4 | 1.9 | 0.16 | -0.4 | 1.7 | 0.14 |
| measurement above normal range during day -365 through 0 days relative to index: Creatinine [Mass/volume] in Serum or Plasma | -0.4 | 2.0 | 0.15 | 0.4 | 1.9 | 0.14 |
| measurement above normal range during day -365 through 0 days relative to index: Aspartate aminotransferase [Enzymatic activity/volume] in Serum or Plasma | -0.4 | 1.3 | 0.12 | -0.4 | 1.7 | 0.14 |
| condition_era group during day -365 through 0 days relative to index: Intermittent claudication of right lower limb co-occurrent and due to atherosclerosis | -0.4 | 0.4 | 0.02 | -0.4 | 1.8 | 0.14 |
| condition_era group during day -365 through 0 days relative to index: Intermittent claudication of left lower limb co-occurrent and due to atherosclerosis | -0.4 | 0.4 | 0.01 | -0.4 | 1.8 | 0.14 |

##### 3.4.4 MDCD

**Supplementary Figure 26:** Preference score distributions, propensity score stratified (middle column) and matched (right column) covariate balance and negative control error distributions (bottom row) for GLP1RA (female) vs DPP4I (female) in the MDCD data source.

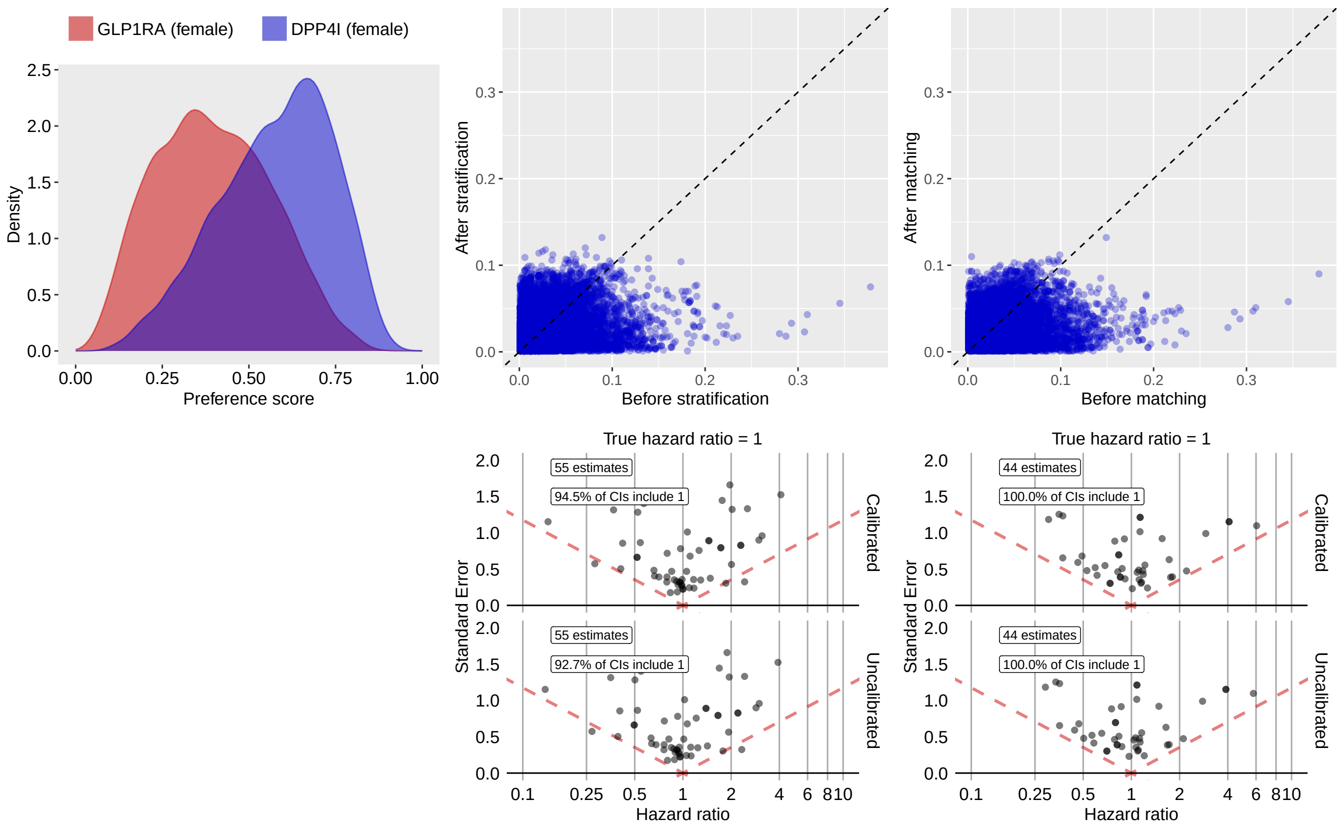

**Supplementary Table 250:** Most extreme patient characteristics balance diagnostics after stratification for GLP1RA (female) vs DPP4I (female) in the MDCD data source.

| | Before stratification | | | After stratification | | |
|---|---|---|---|---|---|---|
| Characteristic (total count = 18999) | T (%) | C (%) | SDf | T (%) | C (%) | SDf |
| condition_era group during day -365 through 0 days relative to index: Pleural effusion | 1.1 | 2.3 | 0.09 | 0.7 | 2.3 | 0.13 |
| device_exposure during day -365 through 0 days relative to index: Gradient compression stocking, below knee, 18-30 mmhg, each | 0.9 | 0.3 | -0.07 | 1.5 | 0.4 | -0.12 |
| condition_era group during day -365 through 0 days relative to index: Mucous polyp of cervix | -0.2 | 0.1 | -0.03 | 0.9 | 0.1 | -0.12 |
| measurement during day -30 through 0 days relative to index: Sedimentation rate, erythrocyte; non-automated | 0.7 | 0.5 | -0.03 | 1.7 | 0.5 | -0.12 |
| device_exposure during day -30 through 0 days relative to index: Blood glucose test or reagent strips for home blood glucose monitor, per 50 strips | 2.4 | 2.1 | -0.02 | 3.9 | 2.0 | -0.11 |
| procedure_occurrence during day -365 through 0 days relative to index: Alcohol and/or drug services; methadone administration and/or service (provision of the drug by a licensed program) | 0.4 | 0.1 | -0.06 | 0.9 | 0.1 | -0.11 |
| condition_era group during day -365 through 0 days relative to index: Lump of cervix | 1.8 | 1.0 | -0.07 | 2.5 | 1.0 | -0.11 |

| (Continued) | T (%) | C (%) | SDf | T (%) | C (%) | SDf |
|---|---|---|---|---|---|---|
| procedure_occurrence during day -365 through 0 days relative to index: Individual psychotherapy, insight oriented, behavior modifying and/or supportive, in an office or outpatient facility, approximately 45 to 50 minutes face-to-face with the patient | 2.0 | 1.5 | -0.04 | 3.2 | 1.5 | -0.11 |
| condition_era group during day -365 through 0 days relative to index: Intestinal obstruction | -0.2 | 0.8 | 0.08 | -0.2 | 0.8 | 0.11 |
| condition_era group during day -365 through 0 days relative to index: Degenerative joint disease involving multiple joints | 1.9 | 2.0 | 0.01 | 3.6 | 1.8 | -0.11 |

**Supplementary Table 251:** Most extreme patient characteristics balance diagnostics after matching for GLP1RA (female) vs DPP4I (female) in the MDCD data source.

| | Before matching | | | After matching | | |
|---|---|---|---|---|---|---|
| Characteristic (total count = 14189) | T (%) | C (%) | SDf | T (%) | C (%) | SDf |
| age group: 75 - 79 | 0.3 | 1.9 | 0.15 | 0.4 | 1.7 | 0.13 |
| drug_era group during day -365 through 0 days relative to index: Insulins and analogues for injection, long-acting | 2.1 | 0.9 | -0.10 | 2.1 | 0.8 | -0.11 |
| index year: 2008 | 1.9 | 1.9 | 0.00 | 2.0 | 0.8 | -0.11 |
| drug_era group during day -365 through 0 days relative to index: INSULINS AND ANALOGUES | 2.9 | 1.6 | -0.09 | 2.9 | 1.3 | -0.11 |
| observation during day -30 through 0 days relative to index: Long-term current use of insulin | 1.7 | 0.7 | -0.10 | 1.7 | 0.6 | -0.11 |
| drug_era group during day -365 through 0 days relative to index: Insulins and analogues for injection, intermediate- or long-acting combined with fast-acting | 2.8 | 1.4 | -0.09 | 2.8 | 1.3 | -0.10 |
| Charlson index - Romano adaptation | 371.5 | 394.9 | 0.10 | 372.5 | 397.0 | 0.10 |
| condition_era group during day -30 through 0 days relative to index: Non-rheumatic heart valve disorder | -0.2 | 0.7 | 0.09 | -0.2 | 0.8 | 0.10 |
| condition_era group during day -365 through 0 days relative to index: Tendon sheath disorder | 2.6 | 1.6 | -0.07 | 2.5 | 1.1 | -0.10 |
| observation during day -365 through 0 days relative to index: Other specified case management service not elsewhere classified | 0.2 | 0.8 | 0.08 | 0.3 | 1.1 | 0.10 |

##### 3.4.5 Open Claims

**Supplementary Figure 27:** Preference score distributions, propensity score stratified (middle column) and matched (right column) covariate balance and negative control error distributions (bottom row) for GLP1RA (female) vs DPP4I (female) in the Open Claims data source.

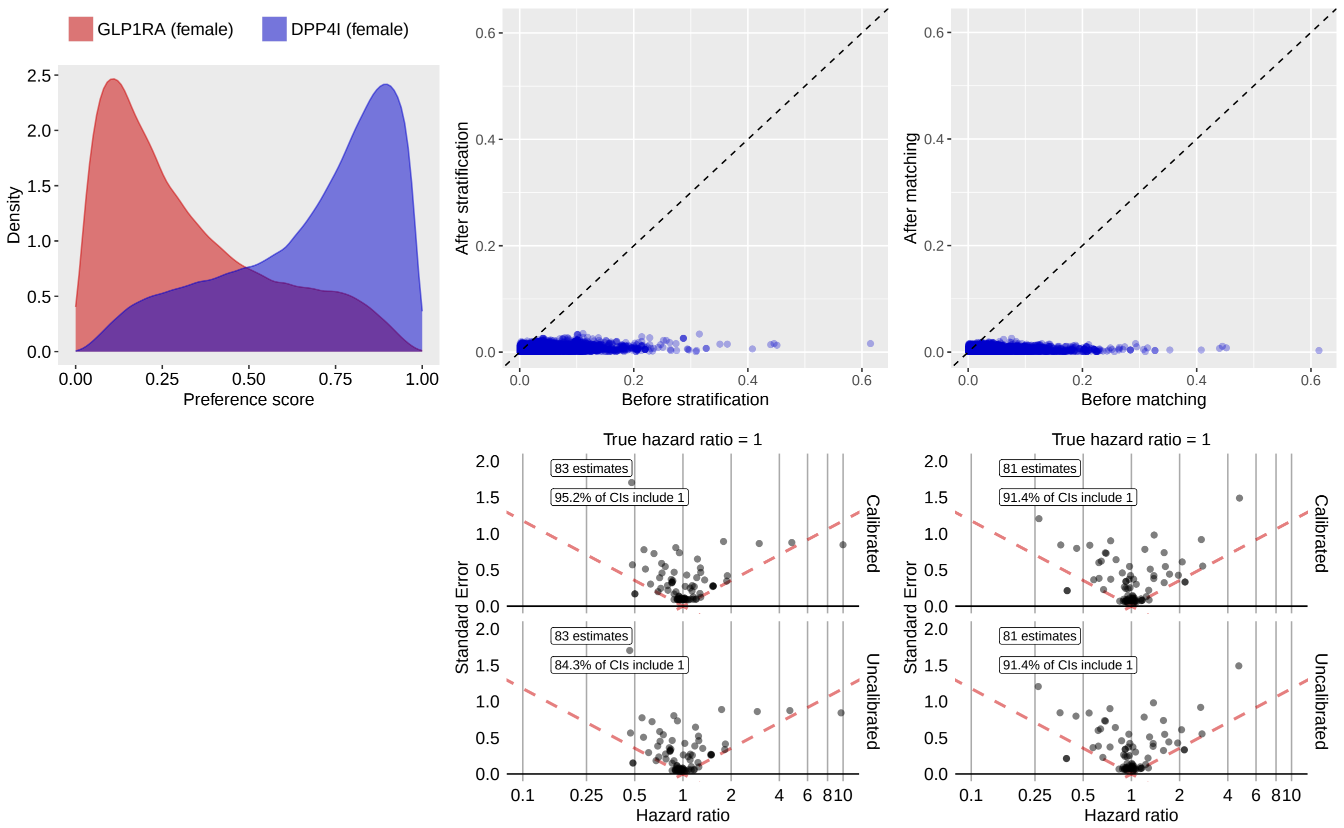

**Supplementary Table 252:** Most extreme patient characteristics balance diagnostics after stratification for GLP1RA (female) vs DPP4I (female) in the Open Claims data source.

| Characteristic (total count = 10554) | Before stratification | | | After stratification | | |
|---|---|---|---|---|---|---|
| | T (%) | C (%) | SDf | T (%) | C (%) | SDf |
| procedure_occurrence during day -30 through 0 days relative to index: Radiologic examination, chest; single view, frontal | 0.1 | 0.8 | 0.11 | 0.4 | 0.6 | 0.04 |
| age group: 75 - 79 | 3.0 | 10.8 | 0.32 | 7.2 | 8.1 | 0.03 |
| condition_era group during day -30 through 0 days relative to index: Acute nephropathy | 0.2 | 1.0 | 0.10 | 0.5 | 0.7 | 0.03 |
| condition_era group during day -30 through 0 days relative to index: Acute renal impairment | 0.2 | 0.9 | 0.10 | 0.4 | 0.7 | 0.03 |
| condition_era group during day -30 through 0 days relative to index: Acute renal failure syndrome | 0.2 | 0.9 | 0.10 | 0.4 | 0.7 | 0.03 |
| condition_era group during day -365 through 0 days relative to index: Prediabetes | 4.4 | 1.0 | -0.21 | 2.2 | 1.8 | -0.03 |
| condition_era group during day -365 through 0 days relative to index: Metabolic syndrome X | 2.0 | 0.7 | -0.12 | 1.2 | 0.9 | -0.03 |
| condition_era group during day -30 through 0 days relative to index: Renal failure syndrome | 0.2 | 1.1 | 0.11 | 0.6 | 0.8 | 0.03 |

| (Continued) | T (%) | C (%) | SDf | T (%) | C (%) | SDf |
|---|---|---|---|---|---|---|
| drug_era group during day -365 through 0 days relative to index: phentermine | 5.7 | 1.5 | -0.23 | 3.0 | 2.5 | -0.03 |
| condition_era group during day -30 through 0 days relative to index: Prediabetes | 1.5 | 0.2 | -0.14 | 0.7 | 0.5 | -0.03 |

**Supplementary Table 253:** Most extreme patient characteristics balance diagnostics after matching for GLP1RA (female) vs DPP4I (female) in the Open Claims data source.

| | Before matching | | | After matching | | |
|---|---|---|---|---|---|---|
| Characteristic (total count = 10331) | T (%) | C (%) | SDf | T (%) | C (%) | SDf |
| Charlson index - Romano adaptation | 323.2 | 340.4 | 0.08 | 328.4 | 334.2 | 0.03 |
| index month: 3 | 11.2 | 9.3 | -0.06 | 9.6 | 10.3 | 0.02 |
| drug_era group during day -365 through 0 days relative to index: liothyronine | 1.1 | 0.4 | -0.08 | 0.7 | 0.6 | -0.02 |
| device_exposure during day -365 through 0 days relative to index: TRUEPLUS SUPER THIN 28G LANCET | 0.4 | 0.8 | 0.06 | 0.4 | 0.6 | 0.02 |
| drug_era group during day 0 through 0 days relative to index: liothyronine | 0.8 | 0.2 | -0.07 | 0.5 | 0.4 | -0.02 |
| device_exposure during day -30 through 0 days relative to index: Walker, folding, wheeled, adjustable or fixed height | 0.1 | 0.2 | 0.04 | 0.1 | 0.1 | 0.02 |
| drug_era group during day 0 through 0 days relative to index: varicella zoster virus glycoprotein E | 0.2 | 0.1 | -0.04 | 0.2 | 0.1 | -0.02 |
| drug_era group during day -30 through 0 days relative to index: liothyronine | 0.8 | 0.3 | -0.07 | 0.6 | 0.4 | -0.02 |
| drug_era group during day -30 through 0 days relative to index: nifedipine | 0.7 | 1.1 | 0.04 | 0.8 | 0.9 | 0.02 |
| drug_era group during day -365 through 0 days relative to index: evolocumab | 0.2 | 0.1 | -0.04 | 0.2 | 0.1 | -0.02 |

##### 3.4.6 OEHR

**Supplementary Figure 28:** Preference score distributions, propensity score stratified (middle column) and matched (right column) covariate balance and negative control error distributions (bottom row) for GLP1RA (female) vs DPP4I (female) in the OEHR data source.

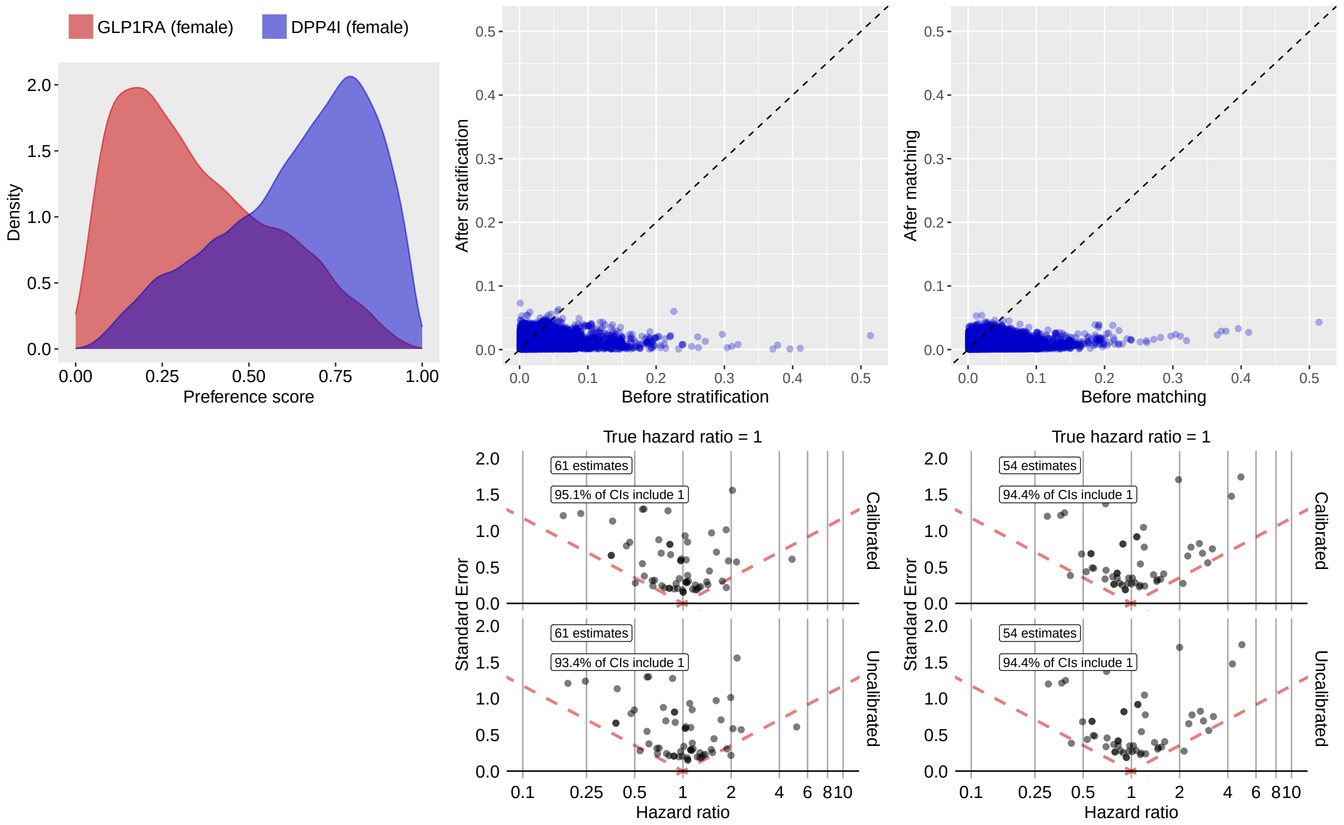

**Supplementary Table 254:** Most extreme patient characteristics balance diagnostics after stratification for GLP1RA (female) vs DPP4I (female) in the OEHR data source.

| Characteristic (total count = 12239) | Before stratification | | | After stratification | | |
|---|---|---|---|---|---|---|
| | T (%) | C (%) | SDf | T (%) | C (%) | SDf |
| measurement during day -365 through 0 days relative to index: Complex medical conditions action plan | 0.3 | 0.3 | -0.00 | 0.9 | 0.3 | -0.07 |
| condition_era group during day -365 through 0 days relative to index: Primary open angle glaucoma | 0.1 | 0.4 | 0.06 | 0.1 | 0.4 | 0.06 |
| age group: 80 - 84 | 0.5 | 3.7 | 0.23 | 1.9 | 2.9 | 0.06 |
| condition_era group during day -365 through 0 days relative to index: Open-angle glaucoma | 0.2 | 0.5 | 0.06 | 0.1 | 0.4 | 0.06 |
| measurement during day -30 through 0 days relative to index: Oral temperature | 0.6 | 0.8 | 0.01 | 1.3 | 0.7 | -0.06 |
| observation during day -30 through 0 days relative to index: Screening mammography results documented and reviewed (PV) | 0.2 | 0.5 | 0.05 | 0.8 | 0.4 | -0.06 |
| condition_era group during day -365 through 0 days relative to index: Loss of appetite | 0.1 | 0.3 | 0.04 | 0.1 | 0.3 | 0.05 |
| measurement during day -365 through 0 days relative to index: Education note | 0.2 | 0.2 | -0.01 | 0.5 | 0.2 | -0.05 |

| (Continued) | T (%) | C (%) | SDf | T (%) | C (%) | SDf |
|---|---|---|---|---|---|---|
| measurement during day -30 through 0 days relative to index: Education note | 0.1 | 0.1 | -0.01 | 0.3 | 0.1 | -0.05 |
| measurement during day -30 through 0 days relative to index: Complex medical conditions action plan | 0.2 | 0.2 | -0.00 | 0.4 | 0.1 | -0.05 |

**Supplementary Table 255:** Most extreme patient characteristics balance diagnostics after matching for GLP1RA (female) vs DPP4I (female) in the OEHR data source.

| | Before matching | | | After matching | | |
|---|---|---|---|---|---|---|
| Characteristic (total count = 12255) | T (%) | C (%) | SDf | T (%) | C (%) | SDf |
| observation during day -30 through 0 days relative to index: Surveillance of contraception | 0.1 | 0.1 | 0.02 | 0.1 | 0.3 | 0.05 |
| measurement during day -365 through 0 days relative to index: Education note | 0.2 | 0.2 | -0.01 | 0.3 | 0.1 | -0.05 |
| observation during day -30 through 0 days relative to index: Discharge medications reconciled with the current medication list in outpatient medical record (COA) (GER) | 0.4 | 0.1 | -0.05 | 0.3 | 0.1 | -0.05 |
| observation during day -365 through 0 days relative to index: Bmi documented outside normal parameters, no follow-up plan documented, no reason given | 0.4 | 0.7 | 0.04 | 0.5 | 0.2 | -0.05 |
| measurement during day -30 through 0 days relative to index: Estradiol | 0.4 | 0.1 | -0.06 | 0.3 | 0.1 | -0.04 |
| condition_era group during day -365 through 0 days relative to index: Breathing painful | 0.4 | 0.5 | 0.03 | 0.4 | 0.2 | -0.04 |
| index year: 2020 | 27.1 | 8.1 | -0.51 | 19.9 | 21.6 | 0.04 |
| drug_era group during day -365 through 0 days relative to index: Other parasympathomimetics | 0.3 | 0.2 | -0.03 | 0.3 | 0.1 | -0.04 |
| condition_era group during day -30 through 0 days relative to index: Inflammation of large intestine | 0.2 | 0.4 | 0.04 | 0.2 | 0.5 | 0.04 |
| observation during day -30 through 0 days relative to index: Coordinated care fee, maintenance rate | 0.2 | 0.1 | -0.03 | 0.2 | 0.1 | -0.04 |

#### 3.5 GLP1RA (female) vs SU (female)

##### 3.5.1 CCAE

**Supplementary Figure 29:** Preference score distributions, propensity score stratified (middle column) and matched (right column) covariate balance and negative control error distributions (bottom row) for GLP1RA (female) vs SU (female) in the CCAE data source.

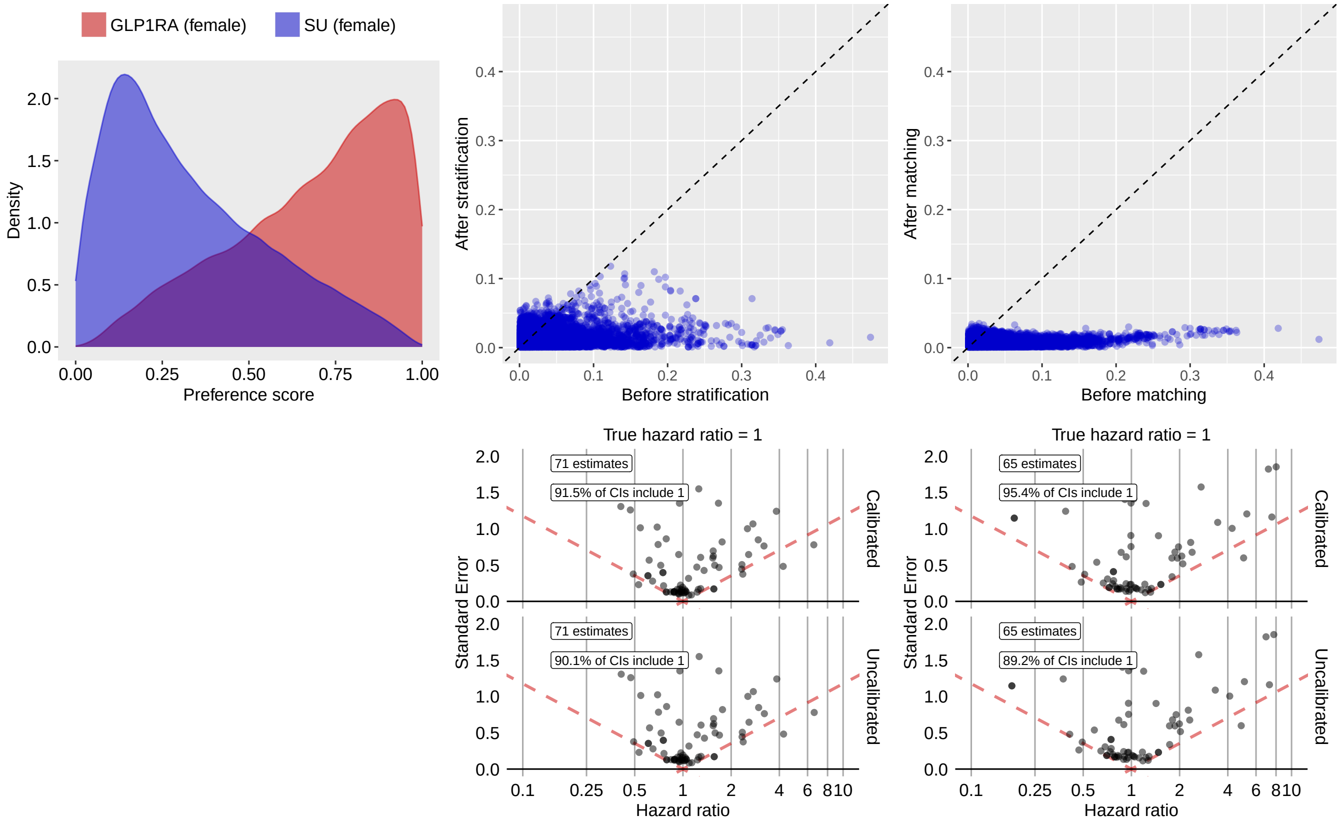

**Supplementary Table 256:** Most extreme patient characteristics balance diagnostics after stratification for GLP1RA (female) vs SU (female) in the CCAE data source.

| Characteristic (total count = 11392) | Before stratification | | | After stratification | | |
|---|---|---|---|---|---|---|
| | T (%) | C (%) | SDf | T (%) | C (%) | SDf |
| observation during day -365 through 0 days relative to index: Routine antenatal care | 0.2 | 1.2 | -0.12 | 0.2 | 1.1 | -0.12 |
| measurement during day -365 through 0 days relative to index: Potassium; serum, plasma or whole blood | 2.7 | 6.5 | -0.18 | 3.2 | 5.5 | -0.11 |
| condition_era group during day -365 through 0 days relative to index: Mother delivered | 0.2 | 1.4 | -0.14 | 0.3 | 1.2 | -0.11 |
| procedure_occurrence during day -365 through 0 days relative to index: Ultrasound, pregnant uterus, real time with image documentation, fetal and maternal evaluation, first trimester ($<$ 14 weeks 0 days), transabdominal approach; single or first gestation | 0.1 | 0.8 | -0.11 | 0.1 | 0.7 | -0.10 |
| measurement during day -365 through 0 days relative to index: Cholesterol, serum or whole blood, total | 1.1 | 4.2 | -0.20 | 1.6 | 3.2 | -0.10 |
| measurement during day -365 through 0 days relative to index: Hemoglobin A1c/Hemoglobin.total in Blood by calculation | 0.1 | 1.2 | -0.14 | 0.2 | 1.0 | -0.10 |

| (Continued) | T (%) | C (%) | SDf | T (%) | C (%) | SDf |
|---|---|---|---|---|---|---|
| measurement above normal range during day -365 through 0 days relative to index: Hemoglobin A1c/Hemoglobin.total in Blood by calculation | 0.1 | 1.2 | -0.14 | 0.2 | 1.0 | -0.10 |
| measurement during day -30 through 0 days relative to index: Creatinine; blood | 1.5 | 4.8 | -0.19 | 2.3 | 4.0 | -0.10 |
| measurement during day -365 through 0 days relative to index: Hematocrit [Volume Fraction] of Blood | 0.1 | 0.8 | -0.10 | 0.1 | 0.7 | -0.09 |
| condition_era group during day -30 through 0 days relative to index: Complication of pregnancy, childbirth and/or the puerperium | 0.1 | 1.2 | -0.14 | 0.3 | 1.0 | -0.09 |

**Supplementary Table 257:** Most extreme patient characteristics balance diagnostics after matching for GLP1RA (female) vs SU (female) in the CCAE data source.

| | Before matching | | | After matching | | |
|---|---|---|---|---|---|---|
| Characteristic (total count = 11418) | T (%) | C (%) | SDf | T (%) | C (%) | SDf |
| condition_era group during day -365 through 0 days relative to index: Lichen sclerosus et atrophicus | 0.1 | 0.1 | 0.01 | 0.1 | 0.2 | -0.04 |
| drug_era group during day -365 through 0 days relative to index: hepatitis B surface antigen vaccine | 0.1 | 0.1 | -0.01 | 0.1 | 0.2 | -0.03 |
| procedure_occurrence during day -365 through 0 days relative to index: Evaluation procedure | 0.1 | 0.3 | -0.06 | 0.1 | 0.2 | -0.03 |
| condition_era group during day -30 through 0 days relative to index: Polyuria | 0.1 | 0.1 | -0.02 | 0.1 | 0.2 | -0.03 |
| condition_era group during day -365 through 0 days relative to index: Victim status | 0.2 | 0.2 | 0.01 | 0.1 | 0.3 | -0.03 |
| observation during day -365 through 0 days relative to index: Motor vehicle on road in collision with another motor vehicle | 0.2 | 0.2 | -0.01 | 0.1 | 0.3 | -0.03 |
| condition_era group during day -365 through 0 days relative to index: Pain in left lower limb | 7.2 | 2.6 | 0.22 | 5.5 | 6.2 | -0.03 |
| condition_era group during day -365 through 0 days relative to index: Victim of vehicular AND/OR traffic accident | 0.2 | 0.2 | 0.01 | 0.1 | 0.3 | -0.03 |
| condition_era group during day -365 through 0 days relative to index: Driver in vehicular AND/OR traffic accident | 0.1 | 0.1 | 0.01 | 0.1 | 0.2 | -0.03 |
| condition_era group during day -365 through 0 days relative to index: Motor vehicle accident, driver | 0.1 | 0.1 | 0.01 | 0.1 | 0.2 | -0.03 |

##### 3.5.2 ODOD

**Supplementary Figure 30:** Preference score distributions, propensity score stratified (middle column) and matched (right column) covariate balance and negative control error distributions (bottom row) for GLP1RA (female) vs SU (female) in the ODOD data source.

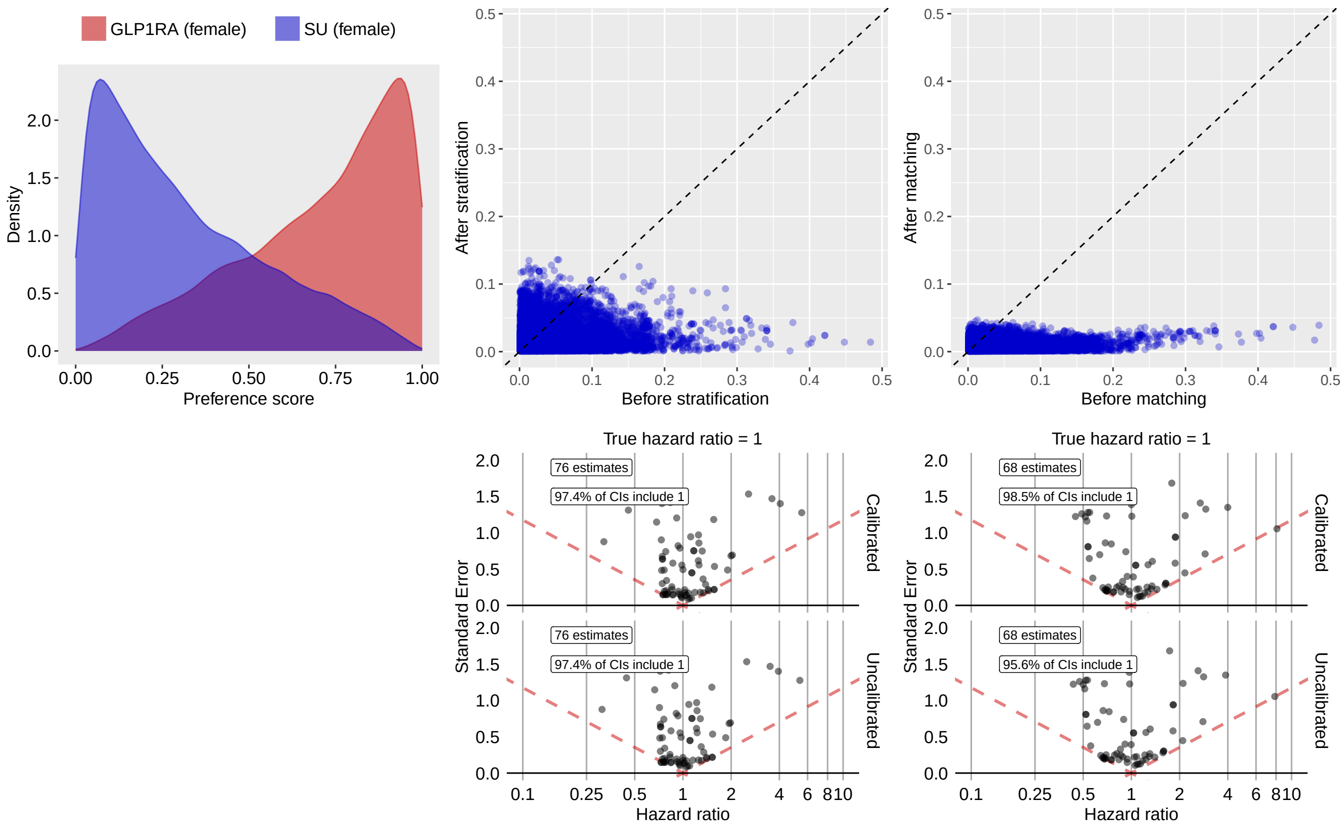

**Supplementary Table 258:** Most extreme patient characteristics balance diagnostics after stratification for GLP1RA (female) vs SU (female) in the ODOD data source.

| Characteristic (total count = 15280) | Before stratification | | | After stratification | | |
|---|---|---|---|---|---|---|
| | T (%) | C (%) | SDf | T (%) | C (%) | SDf |
| condition_era group during day -365 through 0 days relative to index: Hypertensive heart AND renal disease | 0.8 | 1.4 | -0.05 | 3.3 | 1.3 | 0.14 |
| measurement during day -365 through 0 days relative to index: Eye disorder screening | 0.1 | 0.3 | -0.05 | 1.5 | 0.2 | 0.14 |
| observation during day -30 through 0 days relative to index: H/O: pneumonia | 0.1 | 0.1 | 0.01 | 1.1 | 0.1 | 0.14 |
| condition_era group during day -30 through 0 days relative to index: Hypertensive heart AND renal disease | 0.3 | 0.6 | -0.04 | 2.0 | 0.6 | 0.13 |
| condition_era group during day -365 through 0 days relative to index: Hypertensive renal failure | 0.3 | 2.2 | -0.17 | 4.0 | 1.8 | 0.13 |
| condition_era group during day -30 through 0 days relative to index: Diseases of mitral and aortic valves | 0.1 | 0.1 | -0.01 | 1.1 | 0.1 | 0.13 |
| measurement within normal range during day -365 through 0 days relative to index: Erythrocyte distribution width [Ratio] | 0.1 | 0.1 | -0.02 | 1.1 | 0.1 | 0.12 |
| condition_era group during day -30 through 0 days relative to index: Multiple valve disease | 0.2 | 0.2 | -0.01 | 1.2 | 0.2 | 0.12 |

| (Continued) | T (%) | C (%) | SDf | T (%) | C (%) | SDf |
|---|---|---|---|---|---|---|
| measurement during day -365 through 0 days relative to index: Erythrocyte distribution width [Ratio] | 0.1 | 0.1 | -0.02 | 1.1 | 0.1 | 0.12 |
| condition_era group during day -365 through 0 days relative to index: Chronic combined systolic and diastolic heart failure | 0.3 | 0.6 | -0.05 | 1.8 | 0.5 | 0.12 |

**Supplementary Table 259:** Most extreme patient characteristics balance diagnostics after matching for GLP1RA (female) vs SU (female) in the ODOD data source.

| | Before matching | | | After matching | | |
|---|---|---|---|---|---|---|
| Characteristic (total count = 15908) | T (%) | C (%) | SDf | T (%) | C (%) | SDf |
| procedure_occurrence during day -365 through 0 days relative to index: Collection of venous blood by venipuncture or urine sample by catheterization from an individual in a skilled nursing facility (snf) or by a laboratory on behalf of a home health agency (hha) | 0.2 | 0.1 | 0.01 | 0.2 | 0.1 | 0.05 |
| drug_era group during day -365 through 0 days relative to index: acetic acid | 0.1 | 0.2 | -0.01 | 0.1 | 0.2 | -0.04 |
| condition_era group during day -365 through 0 days relative to index: High risk pregnancy | 0.1 | 0.4 | -0.06 | -0.1 | 0.2 | -0.04 |
| condition_era group during day -30 through 0 days relative to index: Gingival and periodontal disease | 0.1 | 0.2 | -0.03 | -0.1 | 0.2 | -0.04 |
| condition_era group during day -365 through 0 days relative to index: First trimester pregnancy | 0.1 | 0.2 | -0.04 | -0.1 | 0.2 | -0.04 |
| procedure_occurrence during day -365 through 0 days relative to index: Ultrasound, pregnant uterus, real time with image documentation, transvaginal | 0.1 | 0.6 | -0.09 | 0.1 | 0.2 | -0.04 |
| procedure_occurrence during day -365 through 0 days relative to index: Injection(s); single or multiple trigger point(s), 3 or more muscles | 0.6 | 0.4 | 0.03 | 0.4 | 0.7 | -0.04 |
| measurement during day -365 through 0 days relative to index: Elastase, pancreatic (EL-1), fecal, qualitative or semi-quantitative | 0.1 | 0.1 | -0.00 | -0.1 | 0.2 | -0.04 |
| procedure_occurrence during day -365 through 0 days relative to index: Computed tomography, head or brain; without contrast material, followed by contrast material(s) and further sections | 0.1 | 0.3 | -0.05 | 0.1 | 0.2 | -0.04 |
| procedure_occurrence during day -365 through 0 days relative to index: Radiologic examination, spine, entire thoracic and lumbar, including skull, cervical and sacral spine if performed (eg, scoliosis evaluation); 2 or 3 views | 0.3 | 0.1 | 0.05 | 0.2 | 0.1 | 0.04 |

##### 3.5.3 MDCR

**Supplementary Figure 31:** Preference score distributions, propensity score stratified (middle column) and matched (right column) covariate balance and negative control error distributions (bottom row) for GLP1RA (female) vs SU (female) in the MDCR data source.

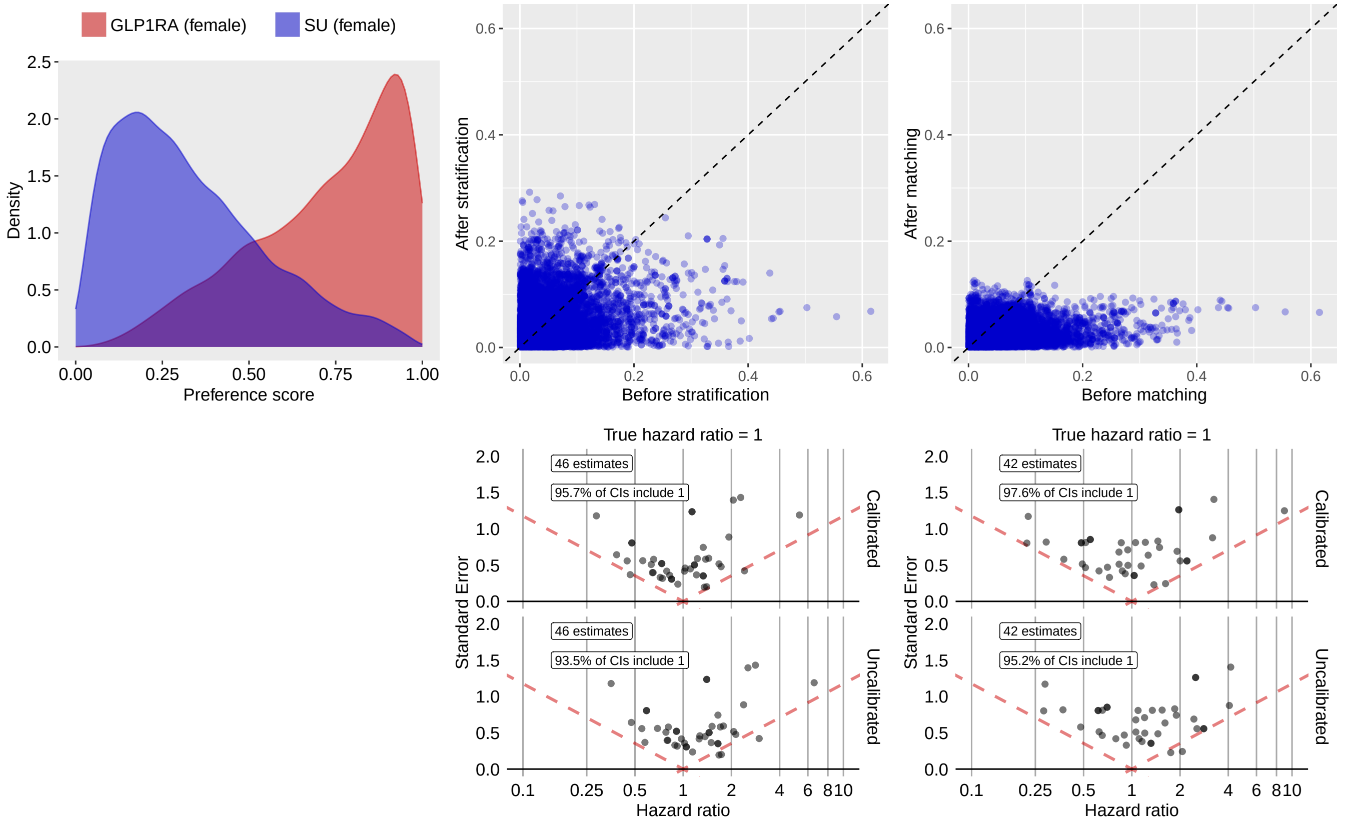

**Supplementary Table 260:** Most extreme patient characteristics balance diagnostics after stratification for GLP1RA (female) vs SU (female) in the MDCR data source.

| Characteristic (total count = 11936) | Before stratification | | | After stratification | | |
|---|---|---|---|---|---|---|
| | T (%) | C (%) | SDf | T (%) | C (%) | SDf |
| observation during day -365 through 0 days relative to index: Aspirin or clopidogrel prescribed or currently being taken (CAD) | -0.4 | 0.2 | 0.02 | 4.8 | 0.3 | 0.29 |
| condition_era group during day -365 through 0 days relative to index: Scar conditions and fibrosis of skin | 1.5 | 0.8 | 0.07 | 6.0 | 0.9 | 0.28 |
| condition_era group during day -365 through 0 days relative to index: Chronic diastolic heart failure | 2.2 | 1.8 | 0.03 | 8.1 | 2.0 | 0.28 |
| condition_era group during day -365 through 0 days relative to index: Diastolic heart failure | 2.6 | 2.5 | 0.00 | 9.5 | 2.9 | 0.28 |
| condition_era group during day -365 through 0 days relative to index: Diastolic dysfunction | 2.7 | 2.6 | 0.01 | 9.5 | 2.9 | 0.27 |
| drug_era group during day -365 through 0 days relative to index: formoterol | 3.9 | 1.7 | 0.13 | 7.9 | 2.1 | 0.27 |
| condition_era group during day -365 through 0 days relative to index: Cerebral infarction due to thrombosis of cerebral arteries | 0.7 | 2.2 | -0.12 | 8.0 | 2.2 | 0.27 |
| drug_era group during day -365 through 0 days relative to index: budesonide | 4.2 | 2.4 | 0.10 | 8.8 | 2.7 | 0.27 |

| (Continued) | T (%) | C (%) | SDf | T (%) | C (%) | SDf |
|---|---|---|---|---|---|---|
| procedure_occurrence during day -365 through 0 days relative to index: Therapeutic, prophylactic, or diagnostic injection (specify substance or drug); intravenous push, single or initial substance/drug | 5.3 | 3.6 | 0.08 | 11.4 | 4.3 | 0.27 |
| condition_era group during day -365 through 0 days relative to index: Thrombotic infarction | 0.7 | 2.2 | -0.12 | 8.0 | 2.2 | 0.26 |

**Supplementary Table 261:** Most extreme patient characteristics balance diagnostics after matching for GLP1RA (female) vs SU (female) in the MDCR data source.

| | Before matching | | | After matching | | |
|---|---|---|---|---|---|---|
| Characteristic (total count = 11138) | T (%) | C (%) | SDf | T (%) | C (%) | SDf |
| condition_era group during day -30 through 0 days relative to index: Cellulitis | -0.4 | 0.9 | -0.10 | -0.5 | 1.0 | -0.13 |
| condition_era group during day -365 through 0 days relative to index: Severe depression | 0.6 | 0.7 | -0.01 | 0.5 | 1.8 | -0.13 |
| condition_era group during day -365 through 0 days relative to index: Severe major depression | 0.6 | 0.7 | -0.01 | 0.5 | 1.7 | -0.12 |
| condition_era group during day -365 through 0 days relative to index: Severe major depression without psychotic features | 0.6 | 0.7 | -0.01 | 0.5 | 1.7 | -0.12 |
| condition_era group during day -365 through 0 days relative to index: Severe recurrent major depression without psychotic features | 0.5 | 0.6 | -0.01 | -0.5 | 1.5 | -0.12 |
| condition_era group during day -365 through 0 days relative to index: Severe recurrent major depression | 0.5 | 0.6 | -0.01 | -0.5 | 1.5 | -0.12 |
| condition_era group during day -365 through 0 days relative to index: Greater trochanteric pain syndrome | 1.4 | 0.4 | 0.10 | 0.5 | 1.8 | -0.12 |
| condition_era group during day -365 through 0 days relative to index: Bursitis of hip | 1.5 | 0.5 | 0.11 | 0.6 | 2.0 | -0.12 |
| condition_era group during day -365 through 0 days relative to index: Bursitis of pelvic region | 1.5 | 0.5 | 0.11 | 0.6 | 2.0 | -0.12 |
| observation during day -365 through 0 days relative to index: Abnormal weight loss | 0.4 | 2.1 | -0.15 | -0.5 | 1.3 | -0.12 |

##### 3.5.4 MDCD

**Supplementary Figure 32:** Preference score distributions, propensity score stratified (middle column) and matched (right column) covariate balance and negative control error distributions (bottom row) for GLP1RA (female) vs SU (female) in the MDCD data source.

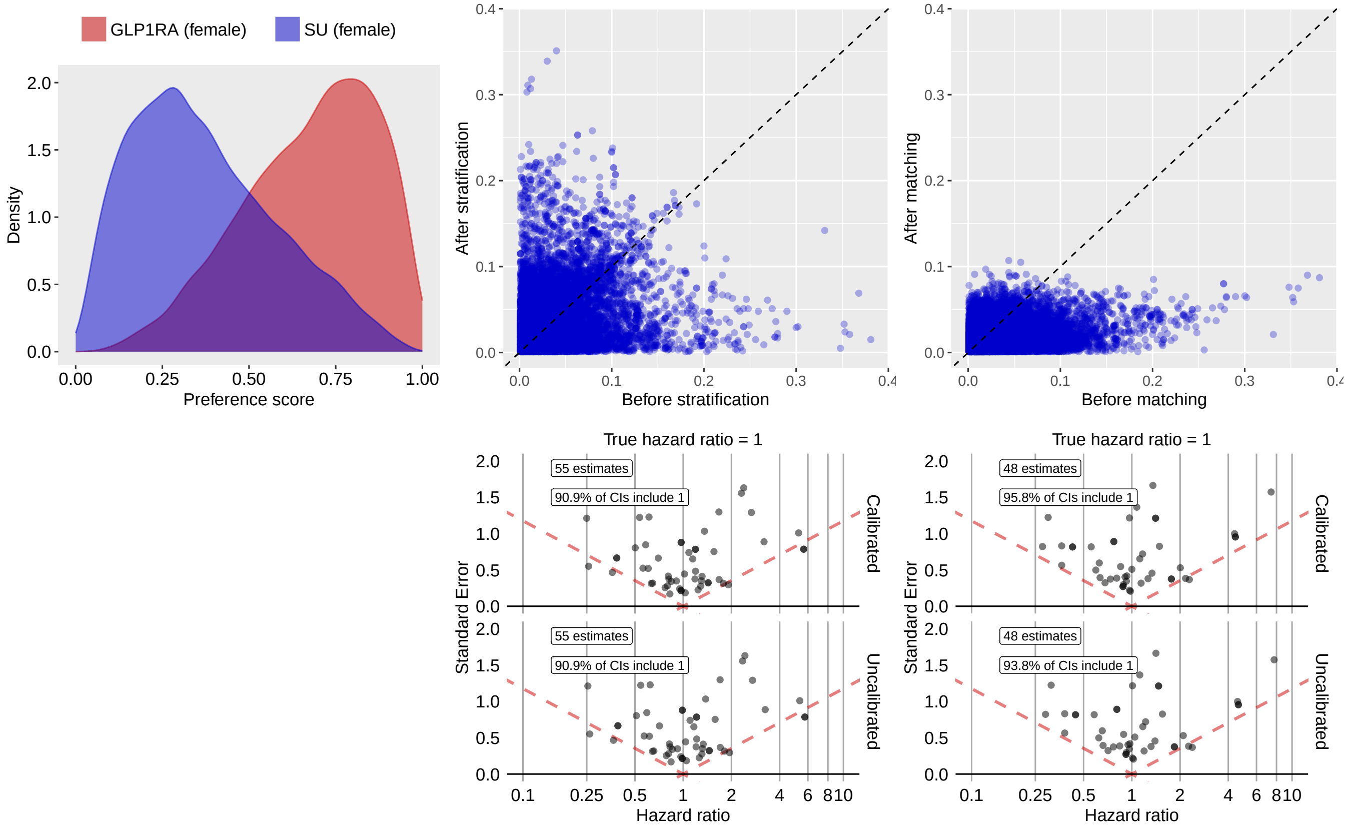

**Supplementary Table 262:** Most extreme patient characteristics balance diagnostics after stratification for GLP1RA (female) vs SU (female) in the MDCD data source.

| Characteristic (total count = 15060) | Before stratification | | | After stratification | | |
|---|---|---|---|---|---|---|
| | T (%) | C (%) | SDf | T (%) | C (%) | SDf |
| condition_era group during day -365 through 0 days relative to index: Lump of cervix | 1.8 | 1.3 | 0.04 | 8.9 | 1.3 | 0.35 |
| condition_era group during day -365 through 0 days relative to index: Lesion of cervix | 1.9 | 1.5 | 0.03 | 9.0 | 1.5 | 0.34 |
| condition_era group during day -365 through 0 days relative to index: Mucous polyp of cervix | -0.2 | 0.1 | 0.01 | 5.2 | 0.1 | 0.32 |
| condition_era group during day -365 through 0 days relative to index: Mass of uterus | 5.6 | 5.4 | 0.01 | 14.6 | 5.4 | 0.31 |
| condition_era group during day -365 through 0 days relative to index: Polyp of cervix | 0.3 | 0.3 | 0.01 | 5.2 | 0.3 | 0.31 |
| condition_era group during day -365 through 0 days relative to index: Lesion of uterus | 5.9 | 5.7 | 0.01 | 14.7 | 5.6 | 0.30 |
| drug_era group during day -30 through 0 days relative to index: Antibiotics | 6.2 | 4.4 | 0.08 | 11.5 | 4.6 | 0.26 |
| drug_era group during day -30 through 0 days relative to index: clindamycin | 2.2 | 1.4 | 0.06 | 6.3 | 1.5 | 0.25 |

| (Continued) | T (%) | C (%) | SDf | T (%) | C (%) | SDf |
|---|---|---|---|---|---|---|
| drug_era group during day -30 through 0 days relative to index: Lincosamides | 2.2 | 1.4 | 0.06 | 6.3 | 1.5 | 0.25 |
| drug_era group during day -30 through 0 days relative to index: ceftriaxone | 1.0 | 1.1 | -0.01 | 5.2 | 1.0 | 0.24 |

**Supplementary Table 263:** Most extreme patient characteristics balance diagnostics after matching for GLP1RA (female) vs SU (female) in the MDCD data source.

| | Before matching | | | After matching | | |
|---|---|---|---|---|---|---|
| Characteristic (total count = 13791) | T (%) | C (%) | SDf | T (%) | C (%) | SDf |
| condition_era group during day -365 through 0 days relative to index: Somatic dysfunction of sacral region | 0.3 | 0.6 | -0.04 | -0.3 | 1.1 | -0.11 |
| condition_era group during day -365 through 0 days relative to index: Intracranial tumor | -0.2 | 0.5 | -0.06 | -0.3 | 0.8 | -0.10 |
| condition_era group during day -365 through 0 days relative to index: Achilles tendinitis | 0.3 | 0.5 | -0.02 | -0.3 | 1.1 | -0.10 |
| condition_era group during day -365 through 0 days relative to index: Neoplasm of nervous system | 0.4 | 0.8 | -0.04 | 0.3 | 1.1 | -0.09 |
| observation during day -30 through 0 days relative to index: Non-compliance with medication regimen | -0.2 | 0.4 | -0.06 | -0.3 | 0.6 | -0.09 |
| condition_era group during day -365 through 0 days relative to index: Neoplasm of uncertain behavior of nervous system | -0.2 | 0.3 | -0.02 | -0.3 | 0.6 | -0.09 |
| observation during day -365 through 0 days relative to index: Exposure to potentially harmful entity | 0.8 | 0.9 | -0.01 | 0.7 | 1.7 | -0.09 |
| observation during day -365 through 0 days relative to index: Long-term current use of drug therapy | 39.7 | 22.9 | 0.37 | 37.7 | 42.1 | -0.09 |
| condition_era group during day -365 through 0 days relative to index: Limbic disorder | 0.3 | 0.7 | -0.05 | -0.3 | 1.0 | -0.09 |
| condition_era group during day -365 through 0 days relative to index: Laceration of head and neck | -0.2 | 0.3 | -0.05 | -0.3 | 0.5 | -0.09 |

##### 3.5.5 Open Claims

**Supplementary Figure 33:** Preference score distributions, propensity score stratified (middle column) and matched (right column) covariate balance and negative control error distributions (bottom row) for GLP1RA (female) vs SU (female) in the Open Claims data source.

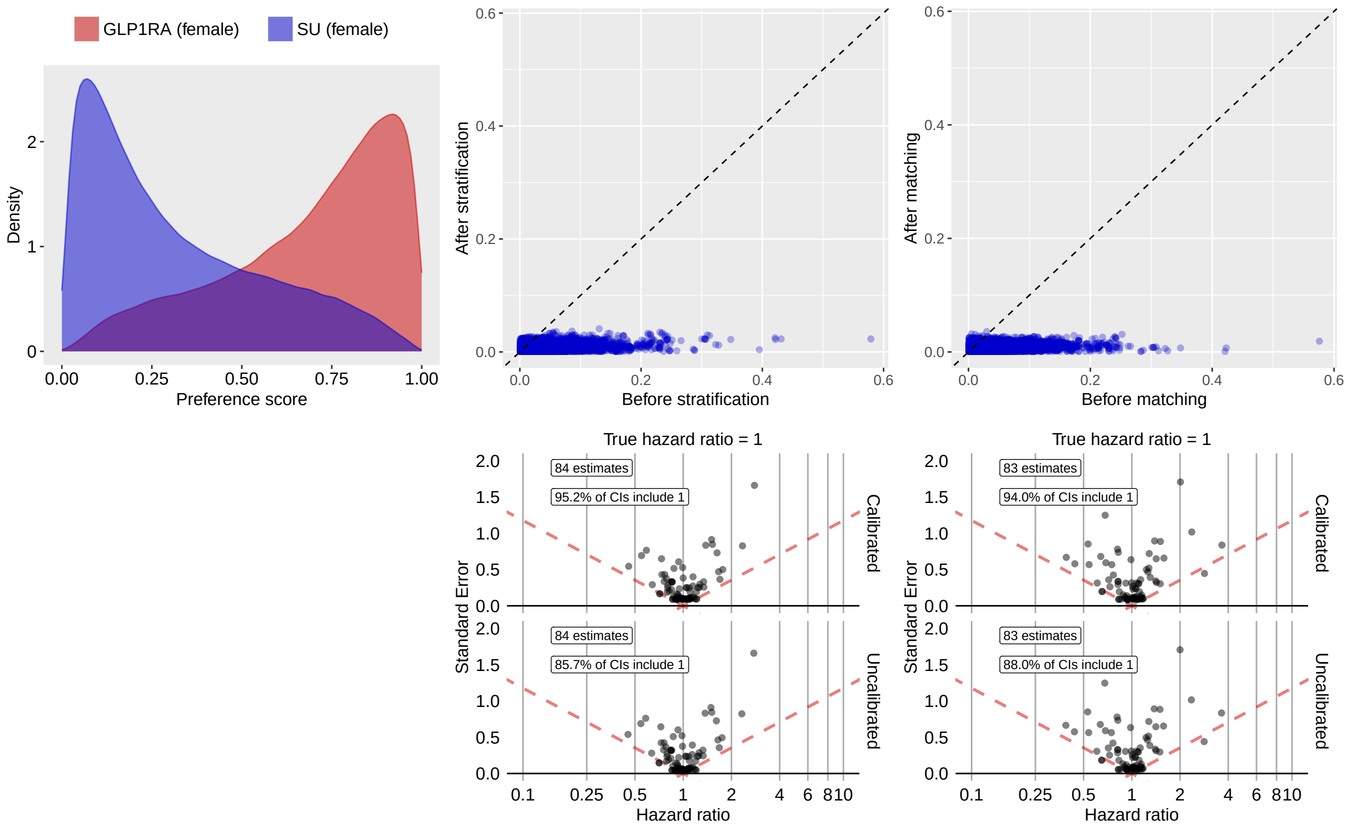

**Supplementary Table 264:** Most extreme patient characteristics balance diagnostics after stratification for GLP1RA (female) vs SU (female) in the Open Claims data source.

| | Before stratification | | | After stratification | | |
|---|---|---|---|---|---|---|
| Characteristic (total count = 10405) | T (%) | C (%) | SDf | T (%) | C (%) | SDf |
| procedure_occurrence during day -30 through 0 days relative to index: Radiologic examination, chest; single view, frontal | 0.1 | 1.1 | -0.13 | 0.5 | 0.9 | -0.04 |
| condition_era group during day -30 through 0 days relative to index: Finding related to pregnancy | 0.1 | 0.5 | -0.09 | 0.2 | 0.4 | -0.04 |
| drug_era group during day -30 through 0 days relative to index: Other antiepileptics | 16.4 | 14.2 | 0.06 | 16.1 | 14.8 | 0.04 |
| condition_era group during day -365 through 0 days relative to index: Prediabetes | 4.4 | 0.8 | 0.23 | 1.7 | 1.2 | 0.04 |
| drug_era group during day 0 through 0 days relative to index: Other antiepileptics | 14.8 | 12.9 | 0.06 | 14.6 | 13.4 | 0.04 |
| drug_era group during day -365 through 0 days relative to index: ANTIDEPRESSANTS | 47.3 | 35.8 | 0.23 | 40.1 | 38.4 | 0.03 |
| drug_era group during day 0 through 0 days relative to index: ANTIDEPRESSANTS | 37.1 | 27.3 | 0.21 | 31.1 | 29.6 | 0.03 |
| condition_era group during day -365 through 0 days relative to index: Metabolic syndrome X | 2.0 | 0.4 | 0.15 | 0.8 | 0.6 | 0.03 |

| (Continued) | T (%) | C (%) | SDf | T (%) | C (%) | SDf |
|---|---|---|---|---|---|---|
| condition_era group during day -30 through 0 days relative to index: Prediabetes | 1.5 | 0.2 | 0.14 | 0.5 | 0.3 | 0.03 |
| drug_era group during day -30 through 0 days relative to index: ANTIEPILEPTICS | 18.6 | 16.5 | 0.06 | 18.4 | 17.2 | 0.03 |

**Supplementary Table 265:** Most extreme patient characteristics balance diagnostics after matching for GLP1RA (female) vs SU (female) in the Open Claims data source.

| | Before matching | | | After matching | | |
|---|---|---|---|---|---|---|
| Characteristic (total count = 10292) | T (%) | C (%) | SDf | T (%) | C (%) | SDf |
| Charlson index - Romano adaptation | 323.2 | 330.0 | -0.03 | 322.2 | 330.0 | -0.04 |
| condition_era group during day -365 through 0 days relative to index: Inflammatory disorder of digestive system | 10.4 | 10.3 | 0.01 | 9.5 | 10.5 | -0.03 |
| Diabetes Comorbidity Severity Index (DCSI) | 213.1 | 274.8 | -0.25 | 220.6 | 227.9 | -0.03 |
| condition_era group during day -365 through 0 days relative to index: Inflammatory disorder of digestive tract | 9.1 | 8.8 | 0.01 | 8.3 | 9.2 | -0.03 |
| device_exposure during day -365 through 0 days relative to index: Water chamber for humidifier, used with positive airway pressure device, replacement, each | 4.3 | 1.4 | 0.17 | 3.1 | 3.6 | -0.03 |
| procedure_occurrence during day -365 through 0 days relative to index: Collection of venous blood by venipuncture | 35.2 | 36.3 | -0.02 | 33.4 | 34.8 | -0.03 |
| drug_era group during day -365 through 0 days relative to index: baclofen | 2.7 | 2.4 | 0.02 | 2.5 | 3.0 | -0.03 |
| condition_era group during day -365 through 0 days relative to index: Breathing-related sleep disorder | 13.6 | 6.4 | 0.24 | 10.7 | 11.5 | -0.03 |
| drug_era group during day -365 through 0 days relative to index: HMG CoA reductase inhibitors in combination with other lipid modifying agents | 59.6 | 64.8 | -0.11 | 61.2 | 62.5 | -0.03 |
| drug_era group during day 0 through 0 days relative to index: OTHER ANALGESICS AND ANTIPYRETICS | 28.8 | 33.7 | -0.11 | 28.4 | 29.6 | -0.03 |

##### 3.5.6 OEHR

**Supplementary Figure 34:** Preference score distributions, propensity score stratified (middle column) and matched (right column) covariate balance and negative control error distributions (bottom row) for GLP1RA (female) vs SU (female) in the OEHR data source.

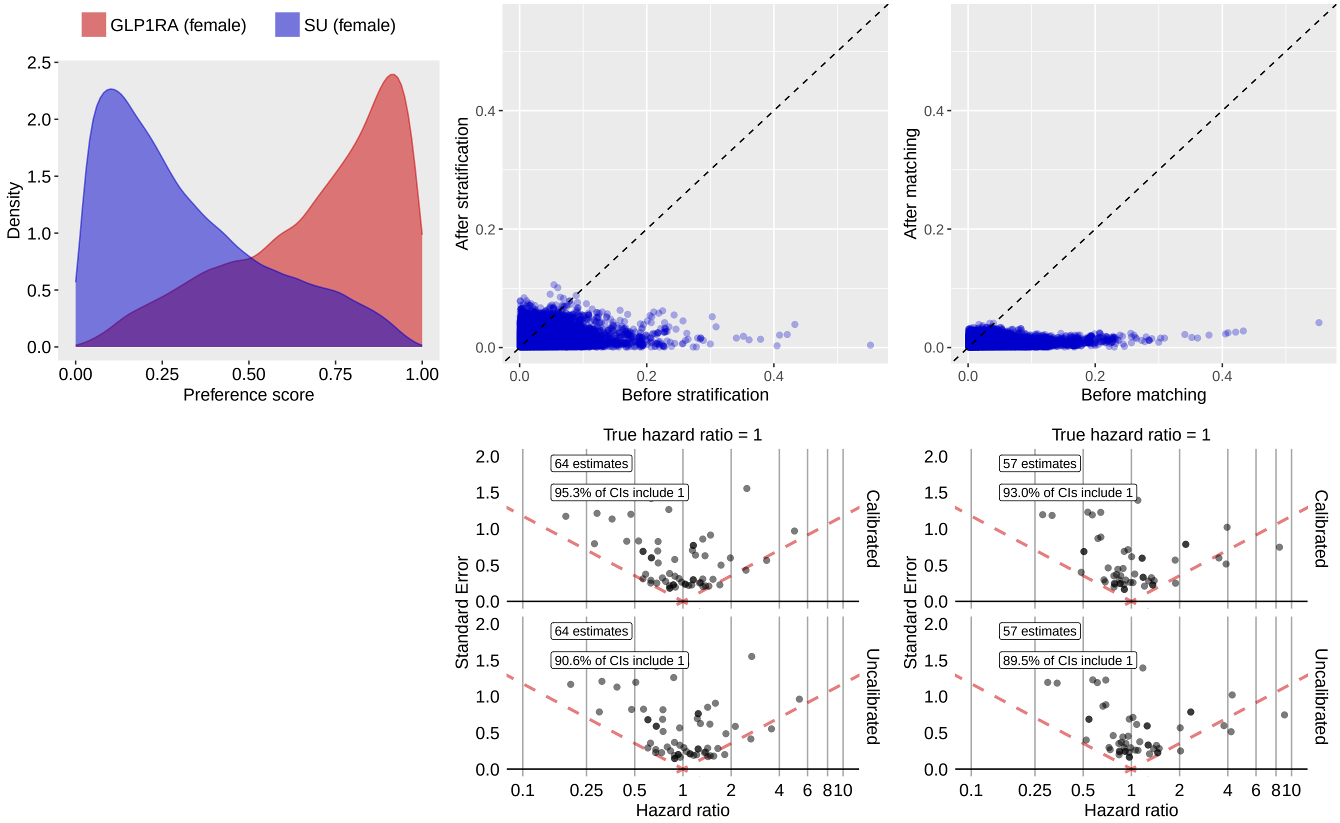

**Supplementary Table 266:** Most extreme patient characteristics balance diagnostics after stratification for GLP1RA (female) vs SU (female) in the OEHR data source.

| Characteristic (total count = 12014) | Before stratification | | | After stratification | | |
|---|---|---|---|---|---|---|
| | T (%) | C (%) | SDf | T (%) | C (%) | SDf |
| drug_era group during day 0 through 0 days relative to index: varicella-zoster virus vaccine live (Oka-Merck) strain | 0.2 | 0.6 | -0.05 | 1.6 | 0.5 | 0.11 |
| drug_era group during day -30 through 0 days relative to index: varicella-zoster virus vaccine live (Oka-Merck) strain | 0.3 | 0.7 | -0.06 | 1.7 | 0.6 | 0.10 |
| drug_era group during day 0 through 0 days relative to index: Varicella zoster vaccines | 1.5 | 1.0 | 0.05 | 2.2 | 1.1 | 0.09 |
| observation during day -365 through 0 days relative to index: Screening mammography results documented and reviewed (PV) | 0.3 | 1.1 | -0.09 | 2.0 | 0.9 | 0.09 |
| drug_era group during day -30 through 0 days relative to index: Varicella zoster vaccines | 2.0 | 1.2 | 0.06 | 2.5 | 1.3 | 0.09 |
| measurement during day -365 through 0 days relative to index: Carbamazepine; total | 0.1 | 0.1 | -0.01 | 0.5 | 0.1 | 0.08 |
| observation during day -30 through 0 days relative to index: Screening mammography results documented and reviewed (PV) | 0.2 | 0.7 | -0.07 | 1.4 | 0.6 | 0.08 |
| condition_era group during day -365 through 0 days relative to index: Nuclear cataract | 1.6 | 2.7 | -0.08 | 1.4 | 2.5 | -0.08 |

| (Continued) | T (%) | C (%) | SDf | T (%) | C (%) | SDf |
|---|---|---|---|---|---|---|
| measurement during day -365 through 0 days relative to index: carBAMazepine [Mass] of Dose | 0.1 | 0.1 | -0.01 | 0.5 | 0.1 | 0.08 |
| drug_era group during day 0 through 0 days relative to index: High-ceiling diuretics and potassium-sparing agents | 11.0 | 11.0 | 0.00 | 13.5 | 10.9 | 0.08 |

**Supplementary Table 267:** Most extreme patient characteristics balance diagnostics after matching for GLP1RA (female) vs SU (female) in the OEHR data source.

| | Before matching | | | After matching | | |
|---|---|---|---|---|---|---|
| Characteristic (total count = 12132) | T (%) | C (%) | SDf | T (%) | C (%) | SDf |
| index year: 2020 | 27.1 | 7.1 | 0.55 | 22.5 | 24.3 | -0.04 |
| condition_era group during day -365 through 0 days relative to index: Inflammation of upper respiratory tract caused by fumes | 0.2 | 0.1 | 0.04 | 0.2 | 0.1 | 0.04 |
| measurement during day -30 through 0 days relative to index: Measurement of Severe acute respiratory syndrome coronavirus 2 (SARS-CoV-2) | 0.2 | 0.1 | 0.03 | 0.1 | 0.2 | -0.04 |
| observation during day -30 through 0 days relative to index: Surveillance of contraception | 0.1 | 0.1 | -0.02 | 0.1 | 0.2 | -0.04 |
| condition_era group during day -365 through 0 days relative to index: Perioperative injury | 0.1 | 0.2 | -0.02 | 0.1 | 0.2 | -0.04 |
| observation during day -365 through 0 days relative to index: Free-Standing Clinic - Other Freestanding Clinic | 0.1 | 0.2 | -0.04 | 0.1 | 0.2 | -0.04 |
| condition_era group during day -30 through 0 days relative to index: Lesion of vulva | 0.1 | 0.2 | -0.02 | 0.1 | 0.2 | -0.04 |
| observation during day -365 through 0 days relative to index: Foot examination performed (includes examination through visual inspection, sensory exam with 10-g monofilament plus testing any one of the following: vibration using 128-hz tuning fork, pinprick sensation, ankle reflexes, or vibration perception thres... | 0.3 | 0.1 | 0.05 | 0.3 | 0.1 | 0.03 |
| condition_era group during day -30 through 0 days relative to index: Chronic kidney disease stage 4 | 0.1 | 0.3 | -0.05 | 0.1 | 0.2 | -0.03 |
| condition_era group during day -365 through 0 days relative to index: Excess intake of micronutrients | 0.2 | 0.1 | 0.04 | 0.2 | 0.1 | 0.03 |

#### 3.6 DPP4I (female) vs SU (female)

##### 3.6.1 CCAE

**Supplementary Figure 35:** Preference score distributions, propensity score stratified (middle column) and matched (right column) covariate balance and negative control error distributions (bottom row) for DPP4I (female) vs SU (female) in the CCAE data source.

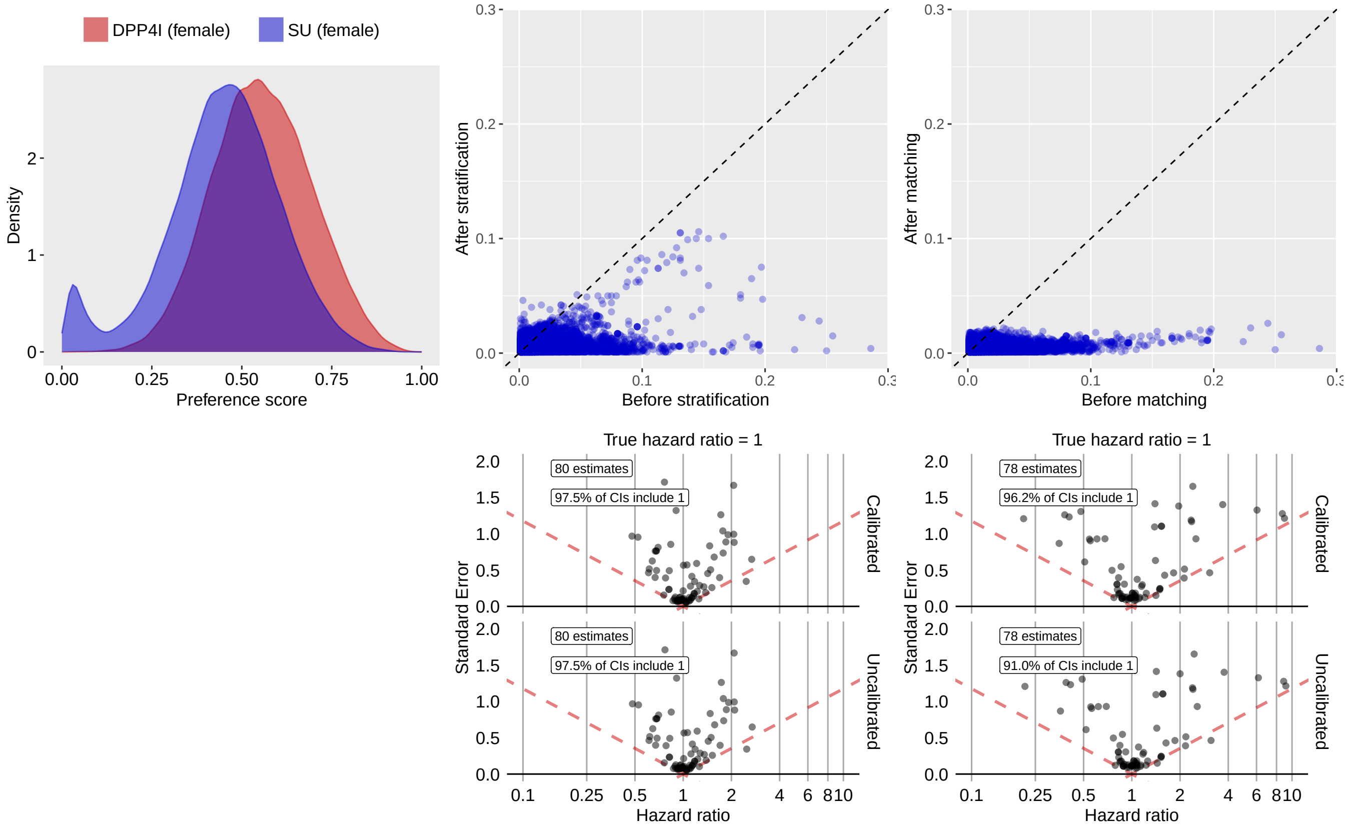

**Supplementary Table 268:** Most extreme patient characteristics balance diagnostics after stratification for DPP4I (female) vs SU (female) in the CCAE data source.

| | Before stratification | | | After stratification | | |
|---|---|---|---|---|---|---|
| Characteristic (total count = 11920) | T (%) | C (%) | SDf | T (%) | C (%) | SDf |
| condition_era group during day -30 through 0 days relative to index: Complication of pregnancy, childbirth and/or the puerperium | 0.1 | 1.2 | -0.15 | 0.1 | 0.9 | -0.11 |
| measurement during day -365 through 0 days relative to index: Hemoglobin A1c/Hemoglobin.total in Blood by calculation | 0.1 | 1.2 | -0.13 | 0.2 | 0.9 | -0.10 |
| measurement above normal range during day -365 through 0 days relative to index: Hemoglobin A1c/Hemoglobin.total in Blood by calculation | 0.1 | 1.2 | -0.13 | 0.2 | 0.9 | -0.10 |
| condition_era group during day -30 through 0 days relative to index: Finding related to pregnancy | 0.1 | 1.7 | -0.17 | 0.3 | 1.2 | -0.10 |
| condition_era group during day -365 through 0 days relative to index: Mother delivered | 0.1 | 1.4 | -0.15 | 0.3 | 1.1 | -0.10 |
| observation during day -365 through 0 days relative to index: Routine antenatal care | 0.1 | 1.2 | -0.14 | 0.2 | 0.9 | -0.10 |

| (Continued) | T (%) | C (%) | SDf | T (%) | C (%) | SDf |
|---|---|---|---|---|---|---|
| measurement during day -365 through 0 days relative to index: Immunoassay for infectious agent antibody, qualitative or semiquantitative, single step method (eg, reagent strip) | 0.2 | 1.5 | -0.14 | 0.3 | 1.1 | -0.10 |
| procedure_occurrence during day -365 through 0 days relative to index: Ultrasound, pregnant uterus, real time with image documentation, transvaginal | 0.1 | 1.1 | -0.13 | 0.2 | 0.8 | -0.09 |
| procedure_occurrence during day -365 through 0 days relative to index: Ultrasound, pregnant uterus, real time with image documentation, fetal and maternal evaluation, first trimester ($<$ 14 weeks 0 days), transabdominal approach; single or first gestation | 0.1 | 0.8 | -0.12 | 0.1 | 0.6 | -0.09 |
| condition_era group during day -365 through 0 days relative to index: Pregnant | 0.1 | 1.0 | -0.12 | 0.2 | 0.8 | -0.08 |

**Supplementary Table 269:** Most extreme patient characteristics balance diagnostics after matching for DPP4I (female) vs SU (female) in the CCAE data source.

| | Before matching | | | After matching | | |
|---|---|---|---|---|---|---|
| Characteristic (total count = 11727) | T (%) | C (%) | SDf | T (%) | C (%) | SDf |
| measurement during day -365 through 0 days relative to index: Lipid panel This panel must include the following: Cholesterol, serum, total (82465) Lipoprotein, direct measurement, high density cholesterol (HDL cholesterol) (83718) Triglycerides (84478) | 84.2 | 74.4 | 0.24 | 82.9 | 83.8 | -0.03 |
| procedure_occurrence during day -365 through 0 days relative to index: Office or other outpatient visit for the evaluation and management of an established patient, which requires at least 2 of these 3 key components: A detailed history; A detailed examination; Medical decision making of moderate complexity. Counseling and/o | 85.5 | 76.5 | 0.23 | 84.3 | 85.1 | -0.02 |
| drug_era group during day -365 through 0 days relative to index: febuxostat | 0.1 | 0.1 | 0.03 | 0.1 | 0.1 | 0.02 |
| measurement during day -365 through 0 days relative to index: Creatinine; blood | 7.0 | 12.8 | -0.20 | 7.3 | 6.7 | 0.02 |
| observation during day -365 through 0 days relative to index: Disease management program, follow-up/reassessment | 0.1 | 0.1 | 0.03 | 0.1 | 0.1 | 0.02 |
| condition_era group during day -30 through 0 days relative to index: Lesion of female perineum | 0.1 | 0.1 | -0.02 | 0.1 | 0.1 | -0.02 |
| condition_era group during day -365 through 0 days relative to index: Mechanical complication of internal orthopedic device, implant AND/OR graft | 0.1 | 0.1 | -0.01 | 0.1 | 0.1 | -0.02 |
| measurement during day -365 through 0 days relative to index: Potassium; serum, plasma or whole blood | 2.6 | 6.5 | -0.19 | 2.7 | 2.4 | 0.02 |
| drug_era group during day -30 through 0 days relative to index: pitavastatin | 0.2 | 0.1 | 0.04 | 0.2 | 0.1 | 0.02 |
| observation during day -365 through 0 days relative to index: Diabetic management program, dietitian visit | 0.2 | 0.1 | 0.04 | 0.2 | 0.1 | 0.02 |

##### 3.6.2 DAG

**Supplementary Figure 36:** Preference score distributions, propensity score stratified (middle column) and matched (right column) covariate balance and negative control error distributions (bottom row) for DPP4I (female) vs SU (female) in the DAG data source.

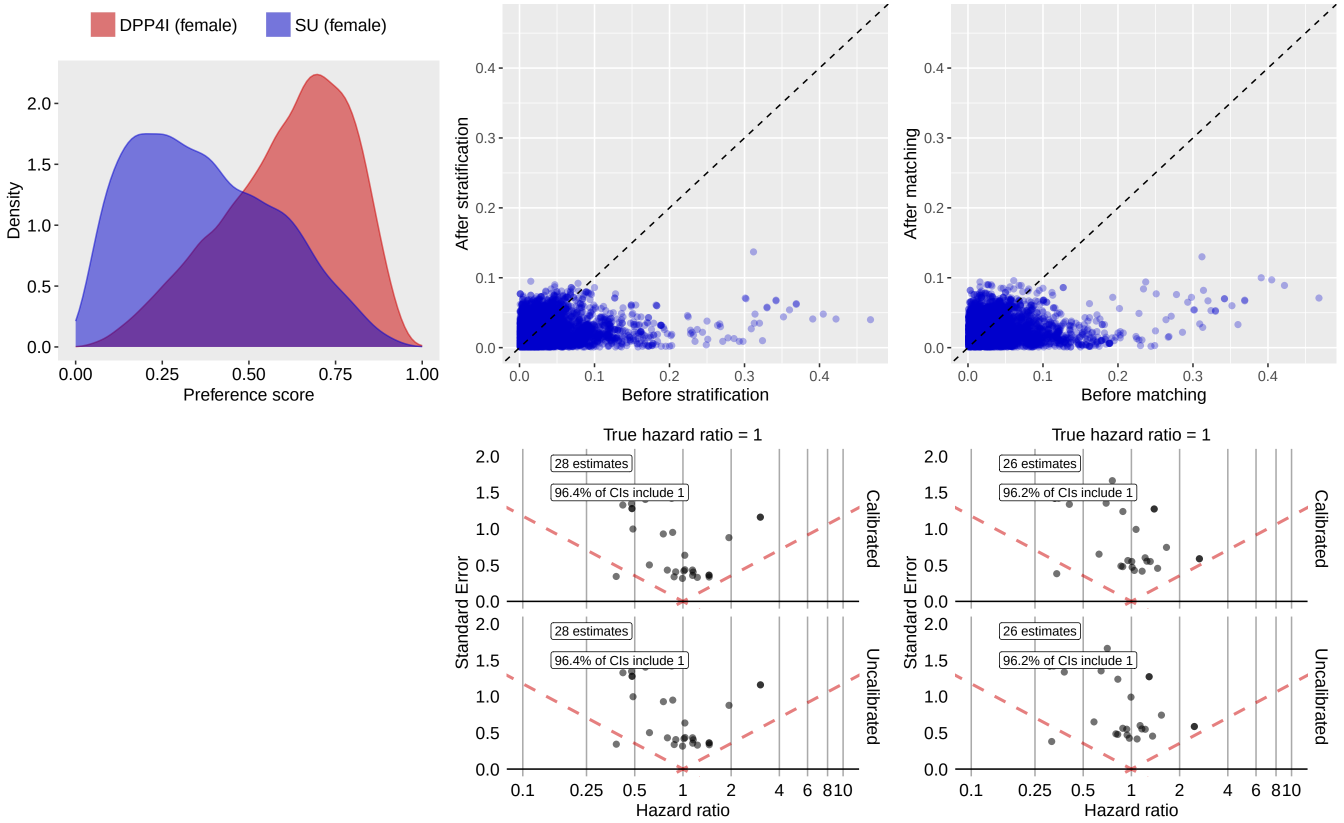

**Supplementary Table 270:** Most extreme patient characteristics balance diagnostics after stratification for DPP4I (female) vs SU (female) in the DAG data source.

| | Before stratification | | | After stratification | | |
|---|---|---|---|---|---|---|
| Characteristic (total count = 6495) | T (%) | C (%) | SDf | T (%) | C (%) | SDf |
| index year: 2019 | 11.2 | 3.2 | 0.31 | 10.3 | 6.5 | 0.14 |
| condition_era group during day -365 through 0 days relative to index: Degenerative disorder of cartilage | 0.4 | 0.5 | -0.01 | 0.4 | 1.2 | -0.10 |
| condition_era group during day -365 through 0 days relative to index: Aortic valve disorder | 1.5 | 0.7 | 0.08 | 1.4 | 0.5 | 0.09 |
| condition_era group during day -365 through 0 days relative to index: Chondromalacia of patella | 0.2 | 0.4 | -0.05 | 0.2 | 0.8 | -0.09 |
| condition_era group during day -365 through 0 days relative to index: Diffuse cervicobrachial syndrome | 1.7 | 2.4 | -0.05 | 1.7 | 3.0 | -0.09 |
| drug_era group during day 0 through 0 days relative to index: febuxostat | 0.6 | -0.2 | 0.10 | 0.6 | -0.1 | 0.09 |
| condition_era group during day -365 through 0 days relative to index: Tendon sheath disorder | 0.9 | 0.4 | 0.07 | 0.9 | 0.3 | 0.09 |
| condition_era group during day -365 through 0 days relative to index: Tenosynovitis | 0.9 | 0.4 | 0.07 | 0.9 | 0.3 | 0.09 |

| (Continued) | T (%) | C (%) | SDf | T (%) | C (%) | SDf |
|---|---|---|---|---|---|---|
| drug_era group during day -30 through 0 days relative to index: perindopril | 0.7 | 0.2 | 0.07 | 0.7 | 0.2 | 0.08 |
| condition_era group during day -365 through 0 days relative to index: Articular cartilage disorder of knee | 0.2 | 0.4 | -0.04 | 0.2 | 0.8 | -0.08 |

**Supplementary Table 271:** Most extreme patient characteristics balance diagnostics after matching for DPP4I (female) vs SU (female) in the DAG data source.

| | Before matching | | | After matching | | |
|---|---|---|---|---|---|---|
| Characteristic (total count = 5964) | T (%) | C (%) | SDf | T (%) | C (%) | SDf |
| index year: 2019 | 11.2 | 3.2 | 0.31 | 7.8 | 4.6 | 0.13 |
| measurement during day -365 through 0 days relative to index: Platelets [#/volume] in Body fluid by Automated count | 40.5 | 22.6 | 0.39 | 27.7 | 32.3 | -0.10 |
| measurement during day -365 through 0 days relative to index: Leukocytes [#/volume] in Body fluid | 40.3 | 22.0 | 0.41 | 27.0 | 31.4 | -0.10 |
| condition_era group during day -30 through 0 days relative to index: Neoplasm of abdomen | 0.7 | 0.3 | 0.06 | 0.9 | 0.2 | 0.10 |
| measurement during day -30 through 0 days relative to index: Leukocytes [#/volume] in Body fluid | 19.7 | 11.2 | 0.24 | 12.5 | 15.8 | -0.09 |
| condition_era group during day -30 through 0 days relative to index: Major depressive disorder | 1.5 | 1.3 | 0.02 | 0.6 | 1.6 | -0.09 |
| condition_era group during day -30 through 0 days relative to index: Recurrent major depression | 0.5 | 0.7 | -0.03 | 0.2 | 0.9 | -0.09 |
| condition_era group during day -30 through 0 days relative to index: Chronic mood disorder | 0.5 | 0.7 | -0.03 | 0.2 | 0.9 | -0.09 |
| condition_era group during day -30 through 0 days relative to index: Recurrent depression | 0.5 | 0.7 | -0.03 | 0.2 | 0.9 | -0.09 |
| condition_era group during day -30 through 0 days relative to index: Chronic mental disorder | 0.5 | 0.7 | -0.03 | 0.2 | 0.9 | -0.09 |

##### 3.6.3 ODOD

**Supplementary Figure 37:** Preference score distributions, propensity score stratified (middle column) and matched (right column) covariate balance and negative control error distributions (bottom row) for DPP4I (female) vs SU (female) in the ODOD data source.

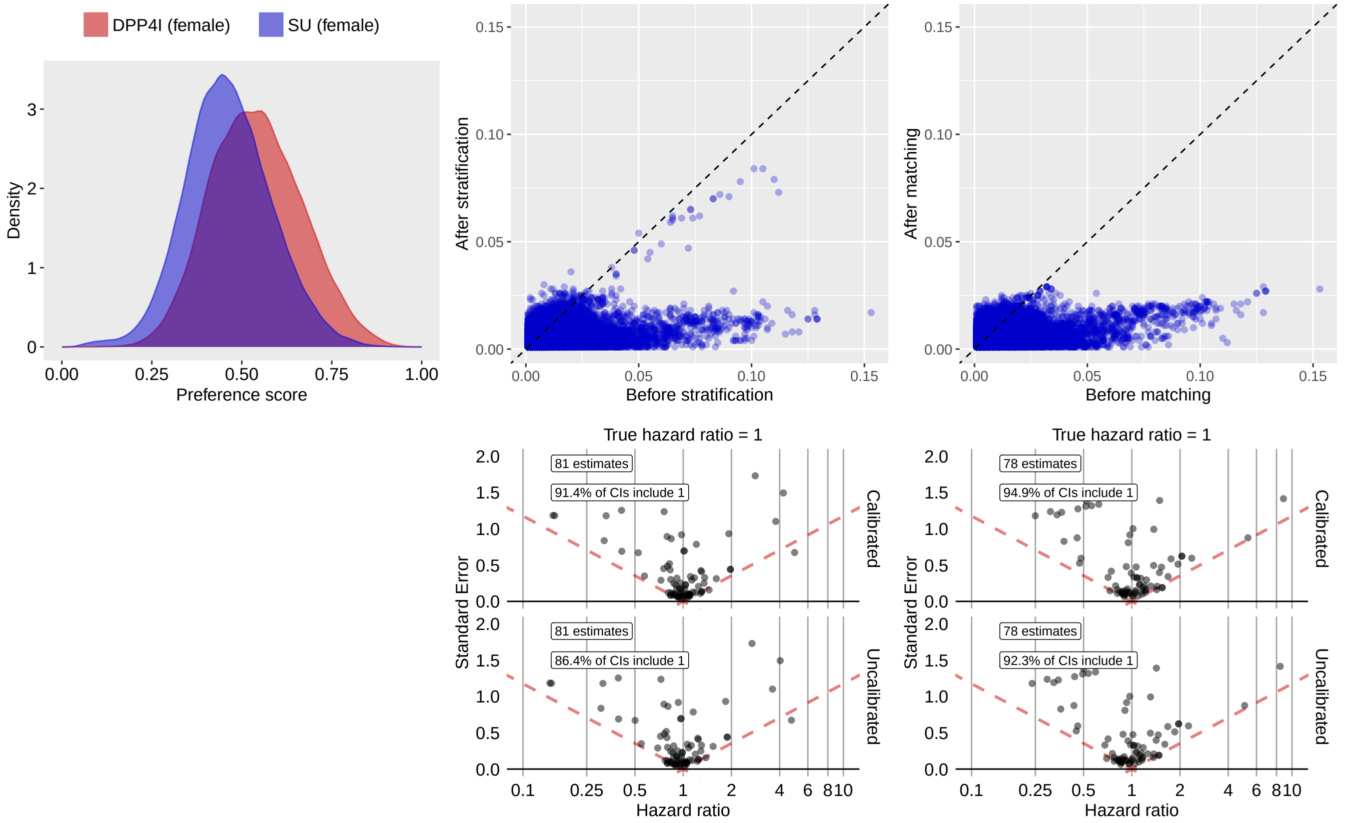

**Supplementary Table 272:** Most extreme patient characteristics balance diagnostics after stratification for DPP4I (female) vs SU (female) in the ODOD data source.

| Characteristic (total count = 16503) | Before stratification | | | After stratification | | |
|---|---|---|---|---|---|---|
| | T (%) | C (%) | SDf | T (%) | C (%) | SDf |
| condition_era group during day -30 through 0 days relative to index: Complication of pregnancy, childbirth and/or the puerperium | 0.1 | 0.7 | -0.10 | 0.1 | 0.5 | -0.08 |
| observation during day -365 through 0 days relative to index: Routine antenatal care | 0.1 | 0.7 | -0.10 | 0.1 | 0.5 | -0.08 |
| condition_era group during day -365 through 0 days relative to index: Mother delivered | 0.1 | 0.8 | -0.11 | 0.2 | 0.7 | -0.08 |
| condition_era group during day -365 through 0 days relative to index: Pregnant | 0.1 | 0.6 | -0.10 | 0.1 | 0.5 | -0.08 |
| condition_era group during day -30 through 0 days relative to index: Finding related to pregnancy | 0.1 | 1.0 | -0.11 | 0.2 | 0.8 | -0.07 |
| condition_era group during day -365 through 0 days relative to index: Complication occurring during pregnancy | 0.1 | 0.7 | -0.09 | 0.1 | 0.6 | -0.07 |
| procedure_occurrence during day -365 through 0 days relative to index: Ultrasound, pregnant uterus, real time with image documentation, transvaginal | 0.1 | 0.6 | -0.09 | 0.1 | 0.5 | -0.07 |

| (Continued) | T (%) | C (%) | SDf | T (%) | C (%) | SDf |
|---|---|---|---|---|---|---|
| condition_era group during day -365 through 0 days relative to index: Hyperglycemic disorder in pregnancy | 0.1 | 0.6 | -0.08 | 0.1 | 0.5 | -0.07 |
| condition_era group during day -365 through 0 days relative to index: Diabetes mellitus during pregnancy, childbirth and the puerperium | 0.1 | 0.6 | -0.08 | 0.1 | 0.5 | -0.07 |
| measurement during day -365 through 0 days relative to index: Blood group antibody screen [Presence] in Serum or Plasma | 0.1 | 0.5 | -0.07 | 0.1 | 0.4 | -0.07 |

**Supplementary Table 273:** Most extreme patient characteristics balance diagnostics after matching for DPP4I (female) vs SU (female) in the ODOD data source.

| | Before matching | | | After matching | | |
|---|---|---|---|---|---|---|
| Characteristic (total count = 16360) | T (%) | C (%) | SDf | T (%) | C (%) | SDf |
| condition_era group during day -365 through 0 days relative to index: Vitamin D deficiency | 22.1 | 17.1 | 0.13 | 20.7 | 21.9 | -0.03 |
| condition_era group during day -30 through 0 days relative to index: Primary malignant neoplasm of upper outer quadrant of female breast | 0.2 | 0.3 | -0.03 | 0.2 | 0.3 | -0.03 |
| condition_era group during day -30 through 0 days relative to index: Malignant neoplasm of upper-outer quadrant of female breast | 0.2 | 0.3 | -0.03 | 0.2 | 0.3 | -0.03 |
| condition_era group during day -30 through 0 days relative to index: Neoplasm of upper outer quadrant of female breast | 0.2 | 0.3 | -0.03 | 0.2 | 0.3 | -0.03 |
| condition_era group during day -30 through 0 days relative to index: Malignant neoplasm of breast upper outer quadrant | 0.2 | 0.3 | -0.03 | 0.2 | 0.3 | -0.03 |
| condition_era group during day -30 through 0 days relative to index: Primary malignant neoplasm of breast upper outer quadrant | 0.2 | 0.3 | -0.03 | 0.2 | 0.3 | -0.03 |
| measurement during day -365 through 0 days relative to index: Vitamin D; 25 hydroxy, includes fraction(s), if performed | 32.9 | 25.9 | 0.15 | 31.1 | 32.4 | -0.03 |
| drug_era group during day -365 through 0 days relative to index: aliskiren | 0.3 | 0.1 | 0.03 | 0.3 | 0.1 | 0.03 |
| drug_era group during day -365 through 0 days relative to index: OTHER AGENTS ACTING ON THE RENIN-ANGIOTENSIN SYSTEM | 0.3 | 0.1 | 0.03 | 0.3 | 0.1 | 0.03 |
| drug_era group during day -365 through 0 days relative to index: Renin-inhibitors | 0.3 | 0.1 | 0.03 | 0.3 | 0.1 | 0.03 |

##### 3.6.4 MDCR

**Supplementary Figure 38:** Preference score distributions, propensity score stratified (middle column) and matched (right column) covariate balance and negative control error distributions (bottom row) for DPP4I (female) vs SU (female) in the MDCR data source.

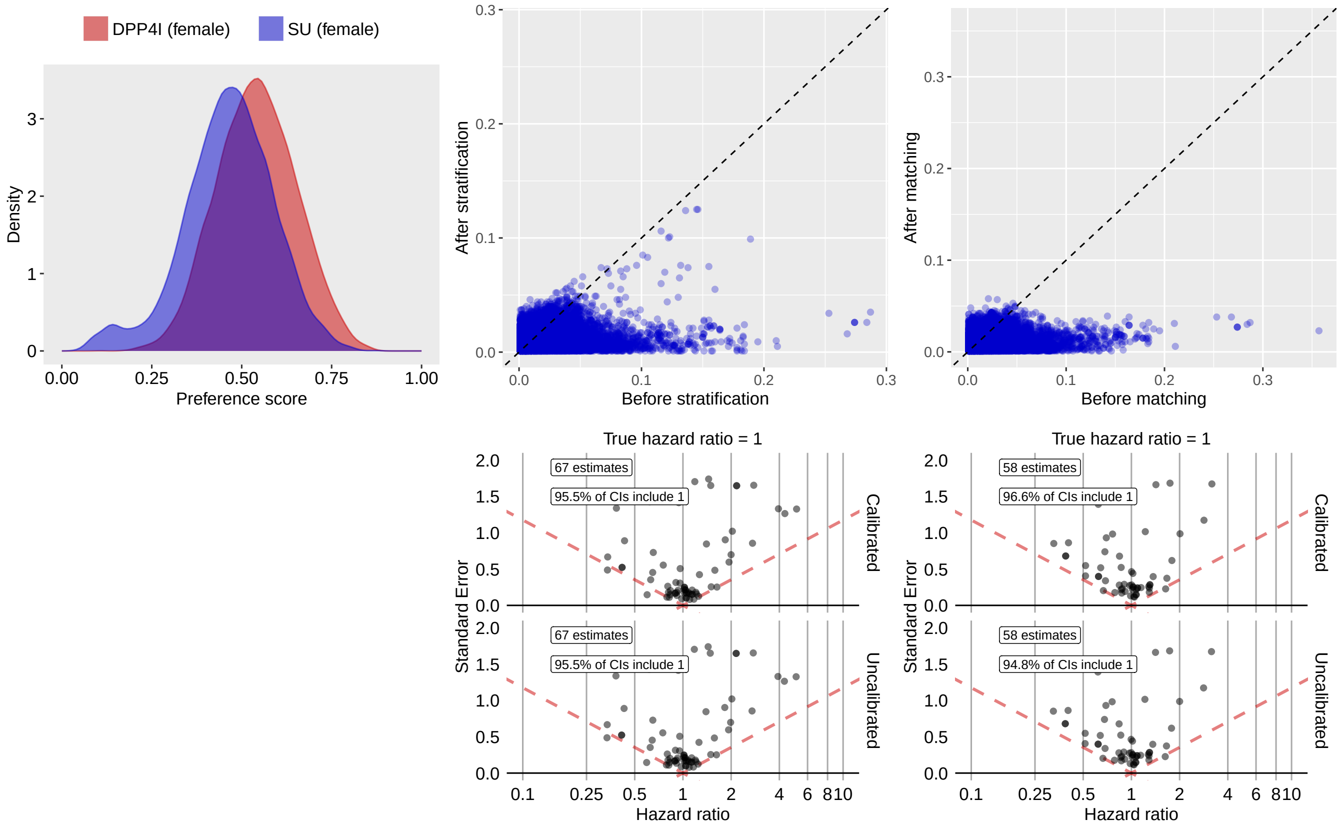

**Supplementary Table 274:** Most extreme patient characteristics balance diagnostics after stratification for DPP4I (female) vs SU (female) in the MDCR data source.

| Characteristic (total count = 13708) | Before stratification | | | After stratification | | |
|---|---|---|---|---|---|---|
| | T (%) | C (%) | SDf | T (%) | C (%) | SDf |
| measurement during day -365 through 0 days relative to index: Hemoglobin A1c/Hemoglobin.total in Blood by calculation | -0.1 | 1.2 | -0.15 | 0.1 | 1.1 | -0.12 |
| measurement above normal range during day -365 through 0 days relative to index: Hemoglobin A1c/Hemoglobin.total in Blood by calculation | -0.1 | 1.1 | -0.14 | 0.1 | 1.1 | -0.12 |
| measurement during day -365 through 0 days relative to index: Immunoassay for infectious agent antibody, qualitative or semiquantitative, single step method (eg, reagent strip) | 0.1 | 1.1 | -0.14 | 0.1 | 0.9 | -0.12 |
| measurement during day -30 through 0 days relative to index: Hemoglobin A1c/Hemoglobin.total in Blood by calculation | -0.1 | 0.7 | -0.12 | -0.1 | 0.7 | -0.11 |
| measurement during day -365 through 0 days relative to index: Glucose [Mass/volume] in Serum or Plasma –8 hours fasting | -0.1 | 0.8 | -0.12 | 0.1 | 0.8 | -0.10 |
| measurement above normal range during day -365 through 0 days relative to index: Glucose [Mass/volume] in Serum or Plasma –8 hours fasting | -0.1 | 0.8 | -0.12 | 0.1 | 0.8 | -0.10 |

| (Continued) | T (%) | C (%) | SDf | T (%) | C (%) | SDf |
|---|---|---|---|---|---|---|
| measurement during day -365 through 0 days relative to index: Cholesterol, serum or whole blood, total | 0.4 | 2.8 | -0.19 | 0.9 | 2.1 | -0.10 |
| device_exposure during day -365 through 0 days relative to index: ONE TOUCH ULTRA CONTROL SOLN | -0.1 | 0.6 | -0.10 | 0.1 | 0.6 | -0.09 |
| measurement within normal range during day -365 through 0 days relative to index: Hematocrit [Volume Fraction] of Blood | 0.1 | 0.7 | -0.10 | 0.2 | 0.7 | -0.08 |
| measurement during day -30 through 0 days relative to index: Cholesterol, serum or whole blood, total | 0.1 | 1.2 | -0.13 | 0.3 | 0.9 | -0.08 |

**Supplementary Table 275:** Most extreme patient characteristics balance diagnostics after matching for DPP4I (female) vs SU (female) in the MDCR data source.

| | Before matching | | | After matching | | |
|---|---|---|---|---|---|---|
| Characteristic (total count = 13505) | T (%) | C (%) | SDf | T (%) | C (%) | SDf |
| procedure_occurrence during day -365 through 0 days relative to index: Trimming of dystrophic nails, any number | 0.8 | 1.0 | -0.02 | 0.6 | 1.2 | -0.06 |
| procedure_occurrence during day -365 through 0 days relative to index: Injection procedure for sacroiliac joint; provision of anesthetic, steroid and/or other therapeutic agent, with or without arthrography | 0.1 | 0.3 | -0.03 | 0.1 | 0.4 | -0.06 |
| condition_era group during day -365 through 0 days relative to index: Neoplasm of pancreas | 0.2 | 0.4 | -0.05 | 0.2 | 0.4 | -0.05 |
| condition_era group during day -30 through 0 days relative to index: Neoplasm of pancreas | -0.1 | 0.2 | -0.05 | -0.1 | 0.2 | -0.05 |
| drug_era group during day -365 through 0 days relative to index: bupivacaine | -0.1 | 0.2 | -0.04 | -0.1 | 0.2 | -0.05 |
| condition_era group during day -30 through 0 days relative to index: Primary malignant neoplasm of pancreas | -0.1 | 0.2 | -0.05 | -0.1 | 0.2 | -0.05 |
| condition_era group during day -30 through 0 days relative to index: Malignant tumor of pancreas | -0.1 | 0.2 | -0.05 | -0.1 | 0.2 | -0.05 |
| condition_era group during day -30 through 0 days relative to index: Mass of pancreas | 0.1 | 0.3 | -0.04 | 0.1 | 0.3 | -0.05 |
| condition_era group during day -365 through 0 days relative to index: Unstable angina co-occurrent and due to coronary arteriosclerosis | 0.1 | 0.2 | -0.02 | 0.1 | 0.3 | -0.05 |
| measurement during day -365 through 0 days relative to index: Cytopathology, evaluation of fine needle aspirate; interpretation and report | 1.1 | 1.1 | 0.00 | 1.0 | 1.5 | -0.05 |

##### 3.6.5 MDCD

**Supplementary Figure 39:** Preference score distributions, propensity score stratified (middle column) and matched (right column) covariate balance and negative control error distributions (bottom row) for DPP4I (female) vs SU (female) in the MDCD data source.

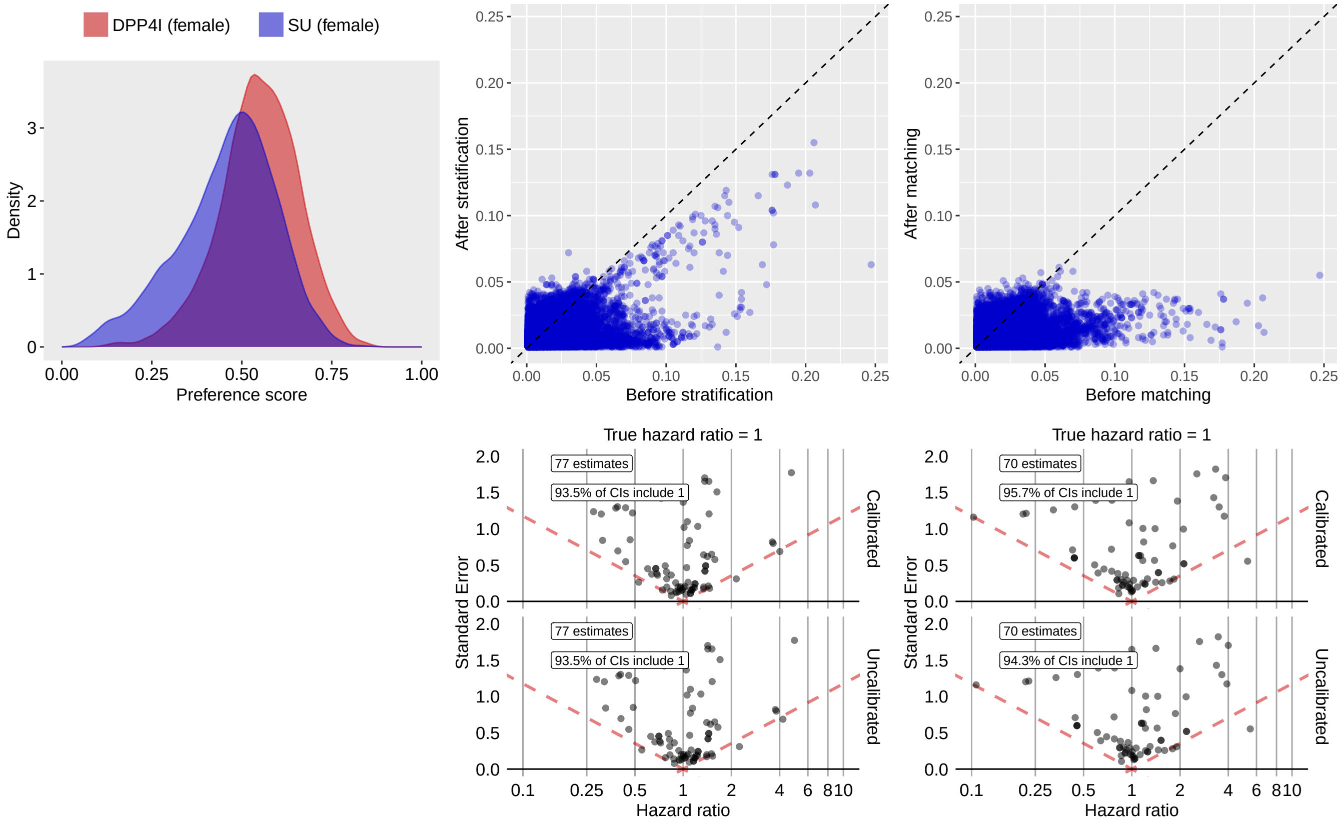

**Supplementary Table 276:** Most extreme patient characteristics balance diagnostics after stratification for DPP4I (female) vs SU (female) in the MDCD data source.

| Characteristic (total count = 16048) | Before stratification | | | After stratification | | |
|---|---|---|---|---|---|---|
| | T (%) | C (%) | SDf | T (%) | C (%) | SDf |
| condition_era group during day -30 through 0 days relative to index: Mother delivered | -0.1 | 2.2 | -0.21 | 0.2 | 1.6 | -0.15 |
| condition_era group during day -365 through 0 days relative to index: Mother delivered | 0.2 | 2.5 | -0.20 | 0.5 | 1.9 | -0.13 |
| condition_era group during day -30 through 0 days relative to index: Complication of pregnancy, childbirth and/or the puerperium | 0.1 | 2.1 | -0.20 | 0.3 | 1.6 | -0.13 |
| condition_era group during day -30 through 0 days relative to index: Hyperglycemic disorder in pregnancy | -0.1 | 1.7 | -0.18 | 0.1 | 1.2 | -0.13 |
| condition_era group during day -30 through 0 days relative to index: Diabetes mellitus during pregnancy, childbirth and the puerperium | -0.1 | 1.7 | -0.18 | 0.1 | 1.2 | -0.13 |
| condition_era group during day -30 through 0 days relative to index: Complication occurring during pregnancy | -0.1 | 1.6 | -0.18 | 0.1 | 1.2 | -0.13 |
| observation during day -365 through 0 days relative to index: Routine antenatal care | 0.2 | 2.3 | -0.19 | 0.5 | 1.7 | -0.12 |
| observation during day -30 through 0 days relative to index: Supervision of high risk pregnancy | -0.1 | 1.0 | -0.14 | -0.1 | 0.8 | -0.12 |

| (Continued) | T (%) | C (%) | SDf | T (%) | C (%) | SDf |
|---|---|---|---|---|---|---|
| condition_era group during day -365 through 0 days relative to index: Pregnant | 0.3 | 2.0 | -0.17 | 0.4 | 1.5 | -0.12 |
| condition_era group during day -30 through 0 days relative to index: Pregnant | -0.1 | 1.1 | -0.14 | -0.1 | 0.8 | -0.12 |

**Supplementary Table 277:** Most extreme patient characteristics balance diagnostics after matching for DPP4I (female) vs SU (female) in the MDCD data source.

| | Before matching | | | After matching | | |
|---|---|---|---|---|---|---|
| Characteristic (total count = 15381) | T (%) | C (%) | SDf | T (%) | C (%) | SDf |
| procedure_occurrence during day -365 through 0 days relative to index: Chronic care management services, at least 20 minutes of clinical staff time directed by a physician or other qualified health care professional, per calendar month, with the following required elements: multiple (two or more) chronic conditions expected | 0.4 | 0.1 | 0.06 | 0.4 | 0.1 | 0.06 |
| drug_era group during day -365 through 0 days relative to index: trospium | 0.5 | 0.1 | 0.07 | 0.5 | 0.1 | 0.06 |
| drug_era group during day 0 through 0 days relative to index: nebivolol | 0.4 | 0.1 | 0.06 | 0.4 | 0.1 | 0.06 |
| procedure_occurrence during day -365 through 0 days relative to index: Application of a modality to 1 or more areas; iontophoresis, each 15 minutes | -0.1 | 0.2 | -0.05 | -0.1 | 0.2 | -0.06 |
| measurement during day -365 through 0 days relative to index: Lipoprotein, direct measurement; high density cholesterol (HDL cholesterol) | 3.8 | 10.0 | -0.25 | 4.0 | 3.0 | 0.06 |
| drug_era group during day -30 through 0 days relative to index: nebivolol | 0.5 | 0.1 | 0.06 | 0.5 | 0.2 | 0.05 |
| condition_era group during day -365 through 0 days relative to index: Burns classified according to percentage of body surface involved | 0.3 | 0.1 | 0.03 | 0.3 | 0.1 | 0.05 |
| device_exposure during day -365 through 0 days relative to index: Port, indwelling (implantable) | -0.1 | 0.2 | -0.06 | -0.1 | 0.2 | -0.05 |
| condition_era group during day -365 through 0 days relative to index: Burn any degree involving less than 10 percent of body surface | 0.3 | 0.1 | 0.03 | 0.3 | 0.1 | 0.05 |
| observation during day -365 through 0 days relative to index: Accident while engaged in household activity | 1.1 | 0.9 | 0.02 | 1.1 | 0.6 | 0.05 |

##### 3.6.6 Open Claims

**Supplementary Figure 40:** Preference score distributions, propensity score stratified (middle column) and matched (right column) covariate balance and negative control error distributions (bottom row) for DPP4I (female) vs SU (female) in the Open Claims data source.

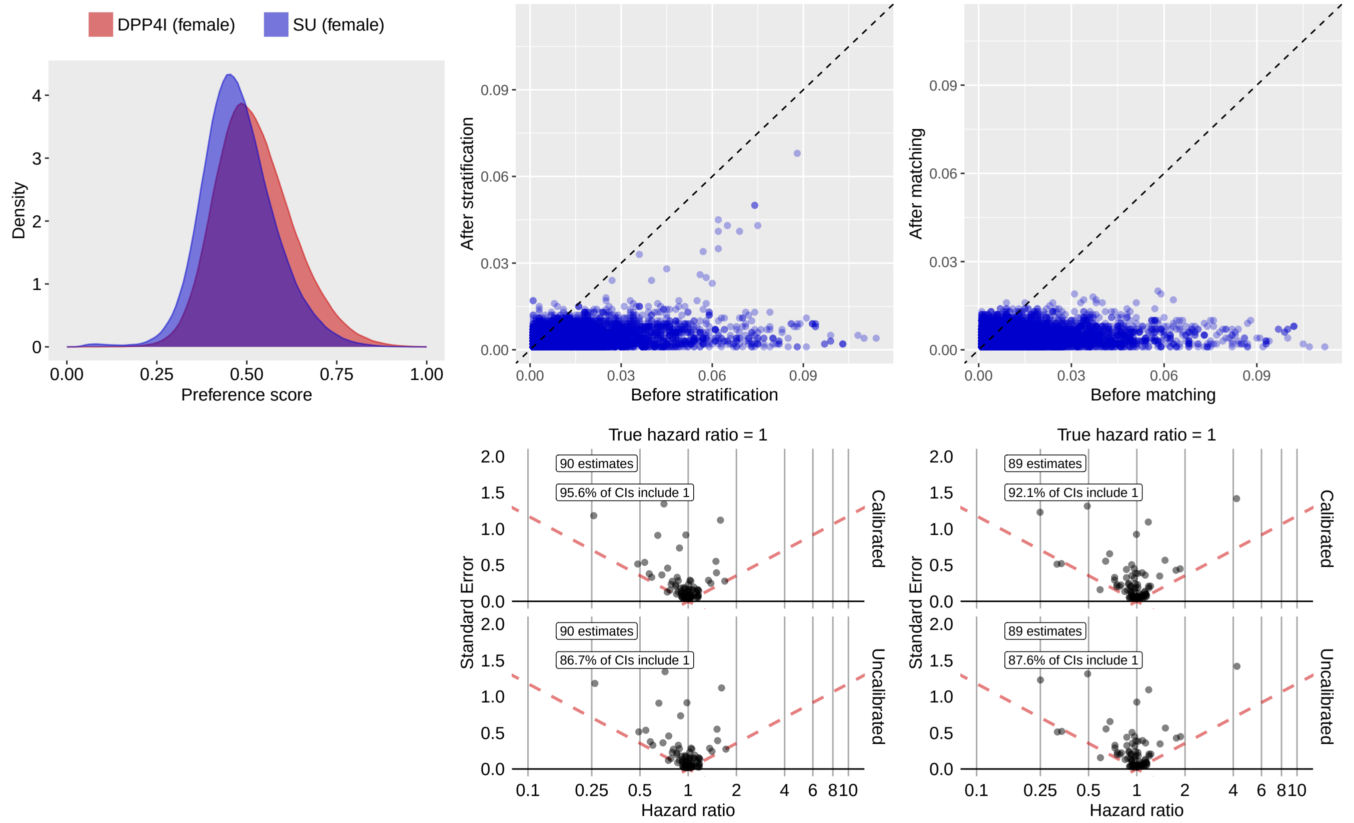

**Supplementary Table 278:** Most extreme patient characteristics balance diagnostics after stratification for DPP4I (female) vs SU (female) in the Open Claims data source.

| Characteristic (total count = 10484) | Before stratification | | | After stratification | | |
|---|---|---|---|---|---|---|
| | T (%) | C (%) | SDf | T (%) | C (%) | SDf |
| condition_era group during day -30 through 0 days relative to index: Finding related to pregnancy | 0.1 | 0.5 | -0.09 | 0.1 | 0.4 | -0.07 |
| condition_era group during day -365 through 0 days relative to index: Hyperglycemic disorder in pregnancy | 0.1 | 0.5 | -0.07 | 0.1 | 0.4 | -0.05 |
| condition_era group during day -365 through 0 days relative to index: Diabetes mellitus during pregnancy, childbirth and the puerperium | 0.1 | 0.5 | -0.07 | 0.1 | 0.4 | -0.05 |
| condition_era group during day -365 through 0 days relative to index: Pregnant | 0.1 | 0.4 | -0.06 | 0.1 | 0.3 | -0.04 |
| condition_era group during day -365 through 0 days relative to index: Finding related to pregnancy | 0.3 | 0.9 | -0.07 | 0.5 | 0.8 | -0.04 |
| observation during day -365 through 0 days relative to index: Routine antenatal care | 0.1 | 0.4 | -0.07 | 0.1 | 0.3 | -0.04 |
| condition_era group during day -365 through 0 days relative to index: Complication of pregnancy, childbirth and/or the puerperium | 0.2 | 0.7 | -0.07 | 0.3 | 0.6 | -0.04 |
| condition_era group during day -365 through 0 days relative to index: First trimester pregnancy | 0.1 | 0.3 | -0.06 | 0.1 | 0.3 | -0.04 |

| (Continued) | T (%) | C (%) | SDf | T (%) | C (%) | SDf |
|---|---|---|---|---|---|---|
| condition_era group during day -365 through 0 days relative to index: Complication occurring during pregnancy | 0.1 | 0.5 | -0.06 | 0.2 | 0.4 | -0.04 |
| procedure_occurrence during day -365 through 0 days relative to index: Ultrasound, pregnant uterus, real time with image documentation, transvaginal | 0.1 | 0.3 | -0.06 | 0.1 | 0.3 | -0.03 |

**Supplementary Table 279:** Most extreme patient characteristics balance diagnostics after matching for DPP4I (female) vs SU (female) in the Open Claims data source.

| | Before matching | | | After matching | | |
|---|---|---|---|---|---|---|
| Characteristic (total count = 10555) | T (%) | C (%) | SDf | T (%) | C (%) | SDf |
| drug_era group during day -30 through 0 days relative to index: icosapent ethyl | 0.5 | 0.1 | 0.06 | 0.3 | 0.2 | 0.02 |
| drug_era group during day 0 through 0 days relative to index: icosapent ethyl | 0.4 | 0.1 | 0.06 | 0.3 | 0.2 | 0.02 |
| drug_era group during day -365 through 0 days relative to index: azilsartan | 0.2 | 0.1 | 0.03 | 0.1 | 0.1 | 0.02 |
| drug_era group during day -30 through 0 days relative to index: pitavastatin | 0.2 | 0.1 | 0.04 | 0.2 | 0.1 | 0.02 |
| drug_era group during day -365 through 0 days relative to index: icosapent ethyl | 0.6 | 0.2 | 0.06 | 0.5 | 0.4 | 0.02 |
| drug_era group during day -365 through 0 days relative to index: pitavastatin | 0.3 | 0.1 | 0.04 | 0.3 | 0.2 | 0.02 |
| drug_era group during day 0 through 0 days relative to index: pitavastatin | 0.2 | 0.1 | 0.03 | 0.2 | 0.1 | 0.02 |
| drug_era group during day -30 through 0 days relative to index: omega-3 acid ethyl esters (USP) | 1.0 | 0.5 | 0.05 | 0.9 | 0.8 | 0.02 |
| drug_era group during day -30 through 0 days relative to index: colesevelam | 0.4 | 0.2 | 0.04 | 0.4 | 0.3 | 0.02 |
| drug_era group during day 0 through 0 days relative to index: colesevelam | 0.4 | 0.2 | 0.04 | 0.3 | 0.2 | 0.02 |

##### 3.6.7 OEHR

**Supplementary Figure 41:** Preference score distributions, propensity score stratified (middle column) and matched (right column) covariate balance and negative control error distributions (bottom row) for DPP4I (female) vs SU (female) in the OEHR data source.

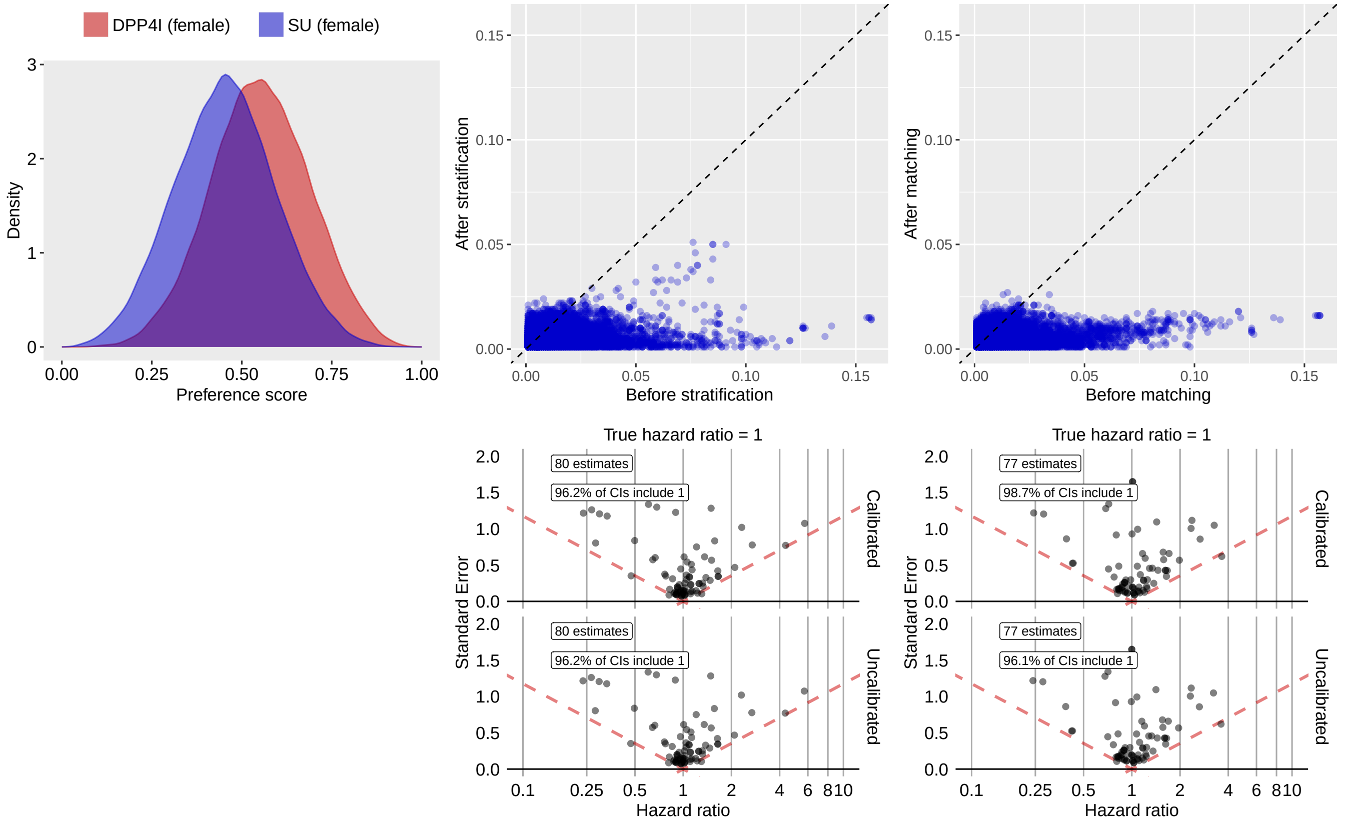

**Supplementary Table 280:** Most extreme patient characteristics balance diagnostics after stratification for DPP4I (female) vs SU (female) in the OEHR data source.

| Characteristic (total count = 12515) | Before stratification | | | After stratification | | |
|---|---|---|---|---|---|---|
| | T (%) | C (%) | SDf | T (%) | C (%) | SDf |
| condition_era group during day -30 through 0 days relative to index: Complication occurring during pregnancy | 0.1 | 0.5 | -0.08 | 0.1 | 0.3 | -0.05 |
| condition_era group during day -30 through 0 days relative to index: Mother delivered | 0.1 | 0.6 | -0.09 | 0.2 | 0.4 | -0.05 |
| condition_era group during day -30 through 0 days relative to index: Hyperglycemic disorder in pregnancy | 0.1 | 0.5 | -0.09 | 0.1 | 0.4 | -0.05 |
| condition_era group during day -30 through 0 days relative to index: Diabetes mellitus during pregnancy, childbirth and the puerperium | 0.1 | 0.5 | -0.09 | 0.1 | 0.4 | -0.05 |
| observation during day -365 through 0 days relative to index: Supervision of high risk pregnancy | 0.1 | 0.5 | -0.08 | 0.1 | 0.4 | -0.05 |
| condition_era group during day -365 through 0 days relative to index: Mother delivered | 0.1 | 0.6 | -0.09 | 0.2 | 0.5 | -0.04 |
| condition_era group during day -365 through 0 days relative to index: Hyperglycemic disorder in pregnancy | 0.2 | 0.8 | -0.08 | 0.3 | 0.6 | -0.04 |
| condition_era group during day -365 through 0 days relative to index: Diabetes mellitus during pregnancy, childbirth and the puerperium | 0.2 | 0.8 | -0.08 | 0.3 | 0.6 | -0.04 |

| (Continued) | T (%) | C (%) | SDf | T (%) | C (%) | SDf |
|---|---|---|---|---|---|---|
| condition_era group during day -365 through 0 days relative to index: Complication occurring during pregnancy | 0.3 | 0.7 | -0.07 | 0.3 | 0.6 | -0.04 |
| procedure_occurrence during day -365 through 0 days relative to index: Ultrasound, pregnant uterus, real time with image documentation, transvaginal | 0.1 | 0.3 | -0.06 | 0.1 | 0.3 | -0.04 |

**Supplementary Table 281:** Most extreme patient characteristics balance diagnostics after matching for DPP4I (female) vs SU (female) in the OEHR data source.

| | Before matching | | | After matching | | |
|---|---|---|---|---|---|---|
| Characteristic (total count = 12351) | T (%) | C (%) | SDf | T (%) | C (%) | SDf |
| measurement during day -30 through 0 days relative to index: Blood test | 0.1 | 0.1 | -0.01 | 0.1 | 0.2 | -0.03 |
| measurement during day -365 through 0 days relative to index: Oxygen/Inspired gas Respiratory system –on ventilator | 0.2 | 0.1 | 0.03 | 0.2 | 0.1 | 0.03 |
| observation during day -365 through 0 days relative to index: Exposure to viral hepatitis | 0.1 | 0.1 | -0.01 | 0.1 | 0.1 | -0.03 |
| drug_era group during day -365 through 0 days relative to index: Typhoid vaccines | 0.1 | 0.2 | -0.02 | 0.1 | 0.2 | -0.02 |
| drug_era group during day -365 through 0 days relative to index: Salmonella typhi Ty21a live antigen | 0.1 | 0.1 | -0.02 | 0.1 | 0.1 | -0.02 |
| observation during day -365 through 0 days relative to index: Patient not identified as an unhealthy alcohol user when screened for unhealthy alcohol use using a systematic screening method | 0.1 | 0.1 | -0.01 | 0.1 | 0.2 | -0.02 |
| drug_era group during day -365 through 0 days relative to index: febuxostat | 0.2 | 0.1 | 0.03 | 0.2 | 0.1 | 0.02 |
| drug_era group during day -365 through 0 days relative to index: aliskiren | 0.2 | 0.1 | 0.03 | 0.2 | 0.1 | 0.02 |
| drug_era group during day -365 through 0 days relative to index: OTHER AGENTS ACTING ON THE RENIN-ANGIOTENSIN SYSTEM | 0.2 | 0.1 | 0.03 | 0.2 | 0.1 | 0.02 |
| drug_era group during day -365 through 0 days relative to index: Renin-inhibitors | 0.2 | 0.1 | 0.03 | 0.2 | 0.1 | 0.02 |

### 3.6.8 VA

**Supplementary Figure 42:** Preference score distributions, propensity score stratified (middle column) and matched (right column) covariate balance and negative control error distributions (bottom row) for DPP4I (female) vs SU (female) in the VA data source.

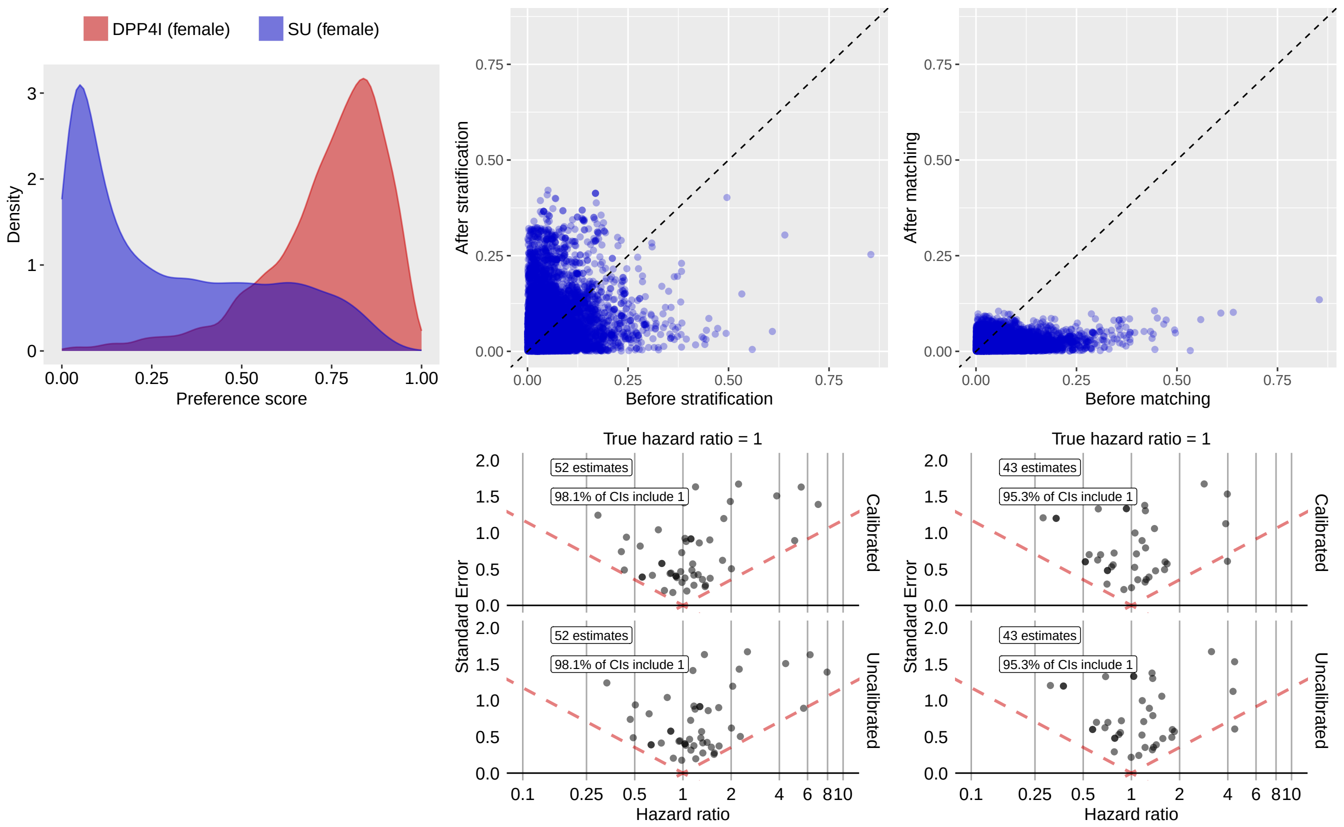

**Supplementary Table 282:** Most extreme patient characteristics balance diagnostics after stratification for DPP4I (female) vs SU (female) in the VA data source.

| Characteristic (total count = 20843) | Before stratification | | | After stratification | | |
|---|---|---|---|---|---|---|
| | T (%) | C (%) | SDf | T (%) | C (%) | SDf |
| condition_era group during day -365 through 0 days relative to index: Bacterial infection of the digestive tract | 9.5 | 8.0 | 0.05 | 23.8 | 8.6 | 0.42 |
| drug_era group during day -365 through 0 days relative to index: QUINOLONE ANTIBACTERIALS | 7.5 | 12.6 | -0.17 | 27.5 | 11.4 | 0.41 |
| drug_era group during day -365 through 0 days relative to index: Fluoroquinolones | 7.5 | 12.6 | -0.17 | 27.5 | 11.4 | 0.41 |
| drug_era group during day -365 through 0 days relative to index: Fluoroquinolones | 7.5 | 12.6 | -0.17 | 27.5 | 11.4 | 0.41 |
| condition_era group during day -365 through 0 days relative to index: Infectious disease of digestive tract | 10.2 | 8.7 | 0.05 | 24.3 | 9.3 | 0.41 |
| procedure_occurrence during day -30 through 0 days relative to index: Counseling | 1.3 | 14.2 | -0.50 | 27.2 | 11.5 | 0.40 |
| drug_era group during day 0 through 0 days relative to index: Tramadol | 3.0 | 4.3 | -0.07 | 16.2 | 4.3 | 0.40 |
| drug_era group during day 0 through 0 days relative to index: Other opioids | 3.0 | 4.3 | -0.07 | 16.2 | 4.3 | 0.40 |

| (Continued) | T (%) | C (%) | SDf | T (%) | C (%) | SDf |
|---|---|---|---|---|---|---|
| condition_era group during day -365 through 0 days relative to index: Infection of digestive system | 10.9 | 10.0 | 0.03 | 25.2 | 10.4 | 0.39 |
| drug_era group during day -365 through 0 days relative to index: Codeine | 4.0 | 8.1 | -0.17 | 20.2 | 7.1 | 0.39 |

**Supplementary Table 283:** Most extreme patient characteristics balance diagnostics after matching for DPP4I (female) vs SU (female) in the VA data source.

| | Before matching | | | After matching | | |
|---|---|---|---|---|---|---|
| Characteristic (total count = 15047) | T (%) | C (%) | SDf | T (%) | C (%) | SDf |
| procedure_occurrence during day -365 through 0 days relative to index: Counseling | 6.8 | 40.2 | -0.85 | 8.2 | 4.8 | 0.14 |
| index year: 2021 | 16.4 | 3.4 | 0.44 | 13.5 | 17.3 | -0.11 |
| procedure_occurrence during day -365 through 0 days relative to index: Arthrocentesis, aspiration and/or injection, intermediate joint or bursa (eg, temporomandibular, acromioclavicular, wrist, elbow or ankle, olecranon bursa); without ultrasound guidance | -0.5 | 0.7 | -0.06 | -0.5 | 0.9 | -0.10 |
| observation during day -365 through 0 days relative to index: Requires influenza virus vaccination | 3.5 | 24.6 | -0.64 | 4.2 | 2.4 | 0.10 |
| index year: 2020 | 28.3 | 6.3 | 0.61 | 27.6 | 23.2 | 0.10 |
| measurement during day -365 through 0 days relative to index: INR in Blood by Coagulation assay | -0.5 | 0.8 | -0.07 | -0.5 | 1.0 | -0.10 |
| measurement during day -30 through 0 days relative to index: Respiratory rate | 63.6 | 76.7 | -0.29 | 66.4 | 61.7 | 0.10 |
| measurement during day -365 through 0 days relative to index: Methadone cutoff [Mass/volume] in Urine for Screen method | 0.9 | 0.9 | 0.01 | 1.1 | 0.3 | 0.10 |
| measurement within normal range during day -365 through 0 days relative to index: Lipoprotein.alpha [Mass/volume] in Serum or Plasma | -0.5 | 0.5 | -0.09 | -0.5 | 0.6 | -0.10 |
| procedure_occurrence during day -365 through 0 days relative to index: Duplex scan of arterial inflow and venous outflow of abdominal, pelvic, scrotal contents and/or retroperitoneal organs; complete study | 1.1 | 0.4 | 0.08 | 1.3 | 0.4 | 0.10 |

##### 3.6.9 SIDIAP

**Supplementary Figure 43:** Preference score distributions, propensity score stratified (middle column) and matched (right column) covariate balance and negative control error distributions (bottom row) for DPP4I (female) vs SU (female) in the SIDIAP data source.

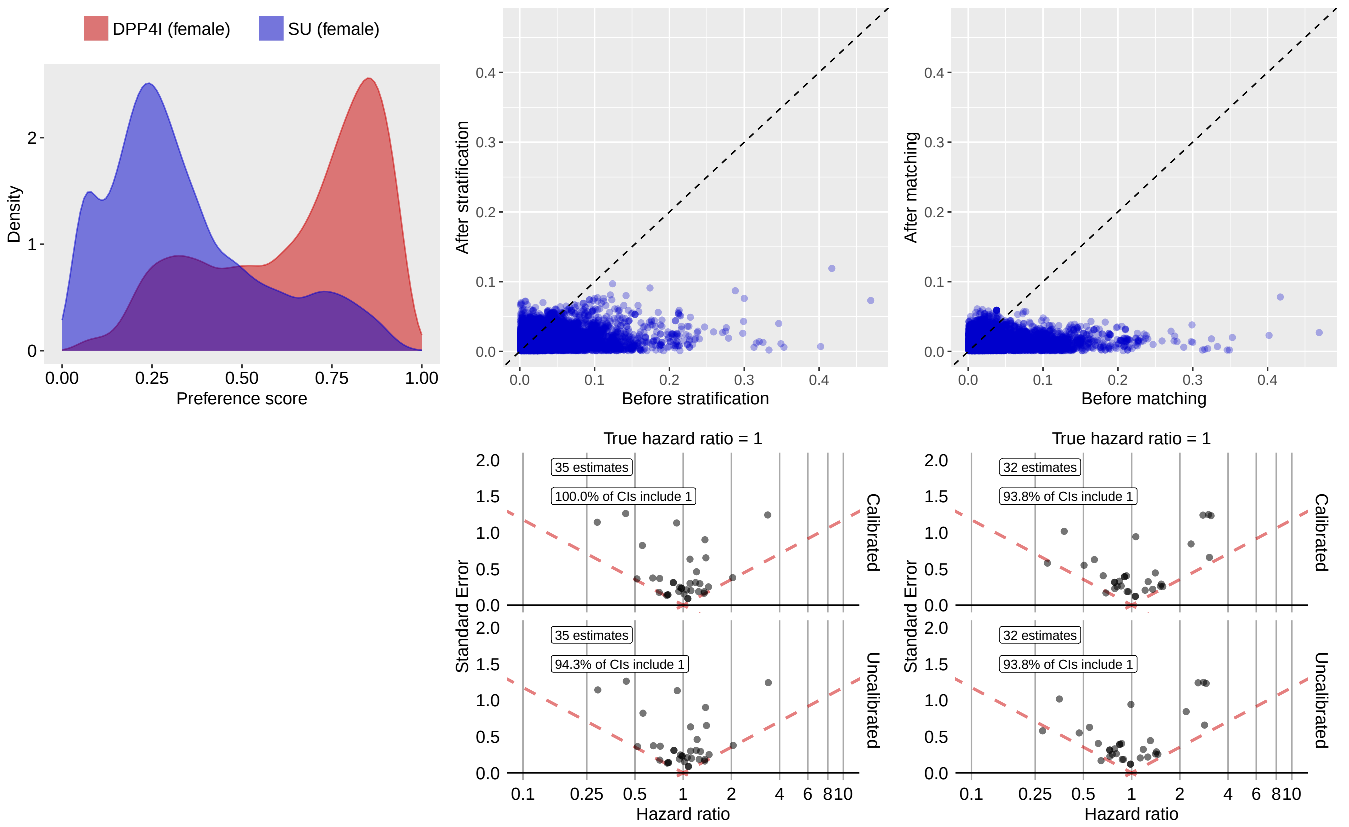

**Supplementary Table 284:** Most extreme patient characteristics balance diagnostics after stratification for DPP4I (female) vs SU (female) in the SIDIAP data source.

| Characteristic (total count = 7197) | Before stratification | | | After stratification | | |
|---|---|---|---|---|---|---|
| | T (%) | C (%) | SDf | T (%) | C (%) | SDf |
| index year: 2019 | 12.4 | 1.9 | 0.42 | 9.9 | 6.6 | 0.12 |
| procedure_occurrence during day -365 through 0 days relative to index: Measurement of Cardiac Electrical Activity, External Approach | 0.8 | -0.1 | 0.12 | 0.6 | 0.1 | 0.10 |
| CHADS2VASc | 391.5 | 369.8 | 0.17 | 384.0 | 372.7 | 0.09 |
| Charlson index - Romano adaptation | 205.8 | 166.8 | 0.29 | 195.5 | 183.6 | 0.09 |
| drug_era group during day -365 through 0 days relative to index: rivaroxaban | 0.8 | -0.1 | 0.12 | 0.7 | 0.1 | 0.08 |
| CHADS2 | 217.7 | 205.2 | 0.14 | 213.3 | 206.3 | 0.08 |
| age group: 85 - 89 | 6.5 | 4.2 | 0.10 | 5.8 | 4.1 | 0.08 |
| Diabetes Comorbidity Severity Index (DCSI) | 96.0 | 58.8 | 0.30 | 85.8 | 76.2 | 0.08 |
| condition_era group during day -365 through 0 days relative to index: Impaired cognition | 1.9 | 0.6 | 0.11 | 1.6 | 0.8 | 0.08 |
| drug_era group during day -30 through 0 days relative to index: denosumab | 0.6 | -0.1 | 0.10 | 0.5 | 0.1 | 0.08 |

**Supplementary Table 285:** Most extreme patient characteristics balance diagnostics after matching for DPP4I (female) vs SU (female) in the SIDIAP data source.

| Characteristic (total count = 7152) | Before matching | | | After matching | | |
|---|---|---|---|---|---|---|
| | T (%) | C (%) | SDf | T (%) | C (%) | SDf |
| index year: 2019 | 12.4 | 1.9 | 0.42 | 4.0 | 2.6 | 0.08 |
| drug_era group during day -30 through 0 days relative to index: Other hypnotics and sedatives | 0.9 | 1.0 | -0.01 | 0.5 | 1.1 | -0.06 |
| drug_era group during day -30 through 0 days relative to index: chlormethiazole | 0.4 | 0.6 | -0.02 | 0.2 | 0.6 | -0.06 |
| condition_era group during day -30 through 0 days relative to index: Acquired trigger finger | 0.3 | 0.1 | 0.04 | 0.5 | 0.1 | 0.06 |
| condition_era group during day -30 through 0 days relative to index: Acquired deformity of finger | 0.3 | 0.1 | 0.04 | 0.5 | 0.1 | 0.06 |
| condition_era group during day -30 through 0 days relative to index: Acquired deformity of upper limb | 0.3 | 0.1 | 0.04 | 0.5 | 0.1 | 0.06 |
| condition_era group during day -30 through 0 days relative to index: Deformity of hand | 0.3 | 0.1 | 0.04 | 0.5 | 0.1 | 0.06 |
| condition_era group during day -30 through 0 days relative to index: Deformity of upper limb | 0.3 | 0.1 | 0.04 | 0.5 | 0.1 | 0.06 |
| condition_era group during day -30 through 0 days relative to index: Acquired deformity of hand | 0.3 | 0.1 | 0.04 | 0.5 | 0.1 | 0.06 |
| condition_era group during day -30 through 0 days relative to index: Triggering of digit | 0.3 | 0.1 | 0.04 | 0.5 | 0.1 | 0.06 |

### 4 Unadjusted incidence rates across data sources and outcomes

#### 4.1 GLP1RA (female) vs SGLT2I (female)

##### 4.1.1 3-point MACE

**Supplementary Table 286:** Exposures, outcomes and risk estimates for 3-point MACE in GLP1RA (female) (T) vs SGLT2I (female) (C) new-user cohorts under four different propensity score (PS) adjustment / time-at-risk (TAR) designs across data sources. We report total patient exposure time in 1,000 patient-year (KYr) units, uncalibrated and calibrated hazard ratios (HRs) and their 95% confidence intervals (CIs). Greyed out entries did not pass study diagnostics and are included only for completeness.

| | Patients | | Exposure (KYr) | | Outcomes | | Incidence (per KYr) | |
|---|---|---|---|---|---|---|---|---|
| | T | C | T | C | T | C | T | C |
| **PS-matching / on-treatment TAR** | | | | | | | | |
| CCAE | 10,652 | 18,456 | 6.9 | 13.4 | 31 | 88 | 4.47 (3.04 - 6.34) | 6.59 (5.28 - 8.12) |
| ODOD | 7,259 | 12,275 | 4.1 | 7.6 | 46 | 85 | 11.2 (8.21 - 15.0) | 11.2 (8.98 - 13.9) |
| MDCR | 698 | 1,406 | 0.4 | 0.9 | 5 | 12 | 11.7 (3.80 - 27.3) | 13.3 (6.89 - 23.3) |
| MDCD | 1,407 | 2,280 | 0.8 | 1.2 | 7 | 20 | 8.82 (3.54 - 18.2) | 16.4 (10.0 - 25.4) |
| Open Claims | 164,256 | 263,893 | 111.1 | 196.2 | 558 | 1,135 | 5.02 (4.62 - 5.46) | 5.78 (5.45 - 6.13) |
| OEHR | 9,548 | 16,375 | 2.8 | 5.1 | 20 | 31 | 7.21 (4.41 - 11.1) | 6.02 (4.09 - 8.55) |
| Meta-analysis | 193,122 | 313,279 | 125.7 | 223.5 | 662 | 1,359 | 5.27 (4.87 - 5.68) | 6.08 (5.76 - 6.41) |
| Meta-analysis (w/o USOC) | 28,866 | 49,386 | 14.6 | 27.3 | 104 | 224 | 7.12 (5.82 - 8.63) | 8.21 (7.17 - 9.36) |
| **PS-stratification / on-treatment TAR** | | | | | | | | |
| CCAE | 14,071 | 18,456 | 8.6 | 13.4 | 39 | 88 | 4.52 (3.21 - 6.18) | 6.59 (5.28 - 8.12) |
| ODOD | 9,783 | 12,278 | 5.2 | 7.6 | 59 | 85 | 11.4 (8.64 - 14.6) | 11.2 (8.98 - 13.9) |
| MDCR | 775 | 1,406 | 0.5 | 0.9 | 5 | 12 | 10.9 (3.54 - 25.5) | 13.3 (6.89 - 23.3) |
| MDCD | 1,662 | 2,284 | 0.9 | 1.2 | 8 | 20 | 8.60 (3.71 - 16.9) | 16.4 (10.0 - 25.3) |
| Open Claims | 223,161 | 263,915 | 139.1 | 196.3 | 676 | 1,135 | 4.86 (4.50 - 5.24) | 5.78 (5.45 - 6.13) |
| OEHR | 12,715 | 16,377 | 3.7 | 5.1 | 25 | 31 | 6.85 (4.43 - 10.1) | 6.02 (4.09 - 8.55) |
| Meta-analysis | 261,392 | 313,310 | 157.5 | 223.5 | 807 | 1,359 | 5.12 (4.78 - 5.49) | 6.08 (5.76 - 6.41) |
| Meta-analysis (w/o USOC) | 38,231 | 49,395 | 18.4 | 27.3 | 131 | 224 | 7.12 (5.95 - 8.45) | 8.21 (7.17 - 9.36) |
| **PS-matching / on-treatment TAR without escalation** | | | | | | | | |
| CCAE | 10,652 | 18,456 | 6.3 | 11.5 | 25 | 71 | 4.00 (2.59 - 5.90) | 6.16 (4.81 - 7.76) |
| ODOD | 7,259 | 12,275 | 3.7 | 6.7 | 41 | 77 | 10.9 (7.85 - 14.8) | 11.5 (9.07 - 14.4) |
| MDCR | 698 | 1,406 | 0.4 | 0.8 | 5 | 10 | 12.4 (4.03 - 29.0) | 12.5 (6.00 - 23.0) |
| MDCD | 1,407 | 2,280 | 0.7 | 1.0 | 6 | 12 | 8.74 (3.21 - 19.0) | 11.8 (6.12 - 20.7) |
| Open Claims | 164,256 | 263,893 | 99.3 | 165.0 | 483 | 947 | 4.87 (4.44 - 5.32) | 5.74 (5.38 - 6.12) |
| OEHR | 9,548 | 16,375 | 2.4 | 4.5 | 15 | 26 | 6.27 (3.51 - 10.3) | 5.81 (3.79 - 8.51) |
| Meta-analysis | 193,122 | 313,279 | 112.3 | 188.7 | 570 | 1,133 | 5.07 (4.67 - 5.51) | 6.00 (5.66 - 6.36) |
| Meta-analysis (w/o USOC) | 28,866 | 49,386 | 13.1 | 23.7 | 87 | 186 | 6.65 (5.33 - 8.20) | 7.84 (6.75 - 9.05) |
| **PS-stratification / on-treatment TAR without escalation** | | | | | | | | |
| CCAE | 14,071 | 18,456 | 7.8 | 11.5 | 31 | 71 | 3.96 (2.69 - 5.62) | 6.16 (4.81 - 7.76) |
| ODOD | 9,783 | 12,278 | 4.8 | 6.7 | 53 | 77 | 11.1 (8.31 - 14.5) | 11.5 (9.06 - 14.3) |
| MDCR | 775 | 1,406 | 0.4 | 0.8 | 5 | 10 | 11.7 (3.78 - 27.2) | 12.5 (6.00 - 23.0) |
| MDCD | 1,662 | 2,284 | 0.8 | 1.0 | 7 | 12 | 8.73 (3.51 - 18.0) | 11.8 (6.10 - 20.6) |
| Open Claims | 223,161 | 263,915 | 125.6 | 165.0 | 596 | 947 | 4.75 (4.37 - 5.14) | 5.74 (5.38 - 6.12) |
| OEHR | 12,715 | 16,377 | 3.2 | 4.5 | 20 | 26 | 6.29 (3.84 - 9.72) | 5.81 (3.79 - 8.51) |
| Meta-analysis | 261,392 | 313,310 | 142.2 | 188.8 | 707 | 1,133 | 4.97 (4.61 - 5.35) | 6.00 (5.66 - 6.36) |
| Meta-analysis (w/o USOC) | 38,231 | 49,395 | 16.6 | 23.7 | 111 | 186 | 6.69 (5.51 - 8.06) | 7.84 (6.75 - 9.05) |

##### 4.1.2 4-point MACE

**Supplementary Table 287:** Exposures, outcomes and risk estimates for 4-point MACE in GLP1RA (female) (T) vs SGLT2I (female) (C) new-user cohorts under four different propensity score (PS) adjustment / time-at-risk (TAR) designs across data sources. We report total patient exposure time in 1,000 patient-year (KYr) units, uncalibrated and calibrated hazard ratios (HRs) and their 95% confidence intervals (CIs). Greyed out entries did not pass study diagnostics and are included only for completeness.

| | Patients | | Exposure (KYr) | | Outcomes | | Incidence (per KYr) | |
|---|---|---|---|---|---|---|---|---|
| | T | C | T | C | T | C | T | C |
| **PS-matching / on-treatment TAR** | | | | | | | | |
| CCAE | 10,491 | 18,202 | 6.8 | 13.2 | 38 | 121 | 5.57 (3.94 - 7.64) | 9.18 (7.62 - 11.0) |
| ODOD | 7,024 | 11,795 | 4.0 | 7.3 | 83 | 129 | 20.9 (16.6 - 25.9) | 17.7 (14.8 - 21.0) |
| MDCR | 652 | 1,334 | 0.4 | 0.9 | 8 | 20 | 19.5 (8.42 - 38.4) | 23.3 (14.2 - 35.9) |
| MDCD | 1,331 | 2,154 | 0.8 | 1.2 | 11 | 35 | 14.5 (7.25 - 26.0) | 30.3 (21.1 - 42.2) |
| Open Claims | 160,005 | 255,636 | 108.2 | 191.0 | 852 | 1,703 | 7.87 (7.35 - 8.42) | 8.91 (8.50 - 9.35) |
| OEHR | 9,389 | 16,143 | 2.7 | 5.1 | 32 | 59 | 11.8 (8.04 - 16.6) | 11.6 (8.87 - 15.0) |
| Meta-analysis | 188,240 | 303,930 | 122.5 | 217.7 | 1,016 | 2,047 | 8.29 (7.79 - 8.82) | 9.40 (9.00 - 9.82) |
| Meta-analysis (w/o USOC) | 28,235 | 48,294 | 14.3 | 26.7 | 164 | 344 | 11.5 (9.80 - 13.4) | 12.9 (11.6 - 14.3) |
| **PS-stratification / on-treatment TAR** | | | | | | | | |
| CCAE | 13,882 | 18,202 | 8.5 | 13.2 | 46 | 121 | 5.41 (3.96 - 7.22) | 9.18 (7.62 - 11.0) |
| ODOD | 9,492 | 11,798 | 5.0 | 7.3 | 99 | 130 | 19.7 (16.0 - 23.9) | 17.8 (14.9 - 21.1) |
| MDCR | 736 | 1,334 | 0.4 | 0.9 | 9 | 20 | 20.3 (9.26 - 38.5) | 23.3 (14.2 - 35.9) |
| MDCD | 1,563 | 2,158 | 0.9 | 1.2 | 17 | 35 | 19.5 (11.4 - 31.3) | 30.2 (21.1 - 42.1) |
| Open Claims | 218,158 | 255,645 | 135.9 | 191.0 | 1,041 | 1,703 | 7.66 (7.20 - 8.14) | 8.91 (8.50 - 9.35) |
| OEHR | 12,521 | 16,145 | 3.6 | 5.1 | 41 | 59 | 11.4 (8.20 - 15.5) | 11.6 (8.86 - 15.0) |
| Meta-analysis | 255,616 | 303,948 | 153.9 | 217.7 | 1,244 | 2,048 | 8.08 (7.64 - 8.54) | 9.41 (9.00 - 9.82) |
| Meta-analysis (w/o USOC) | 37,458 | 48,303 | 18.0 | 26.7 | 203 | 345 | 11.3 (9.78 - 12.9) | 12.9 (11.6 - 14.4) |
| **PS-matching / on-treatment TAR without escalation** | | | | | | | | |
| CCAE | 10,491 | 18,202 | 6.1 | 11.4 | 31 | 102 | 5.04 (3.43 - 7.16) | 8.95 (7.30 - 10.9) |
| ODOD | 7,024 | 11,795 | 3.6 | 6.5 | 74 | 112 | 20.4 (16.0 - 25.6) | 17.3 (14.2 - 20.8) |
| MDCR | 652 | 1,334 | 0.4 | 0.8 | 8 | 17 | 20.6 (8.91 - 40.6) | 22.3 (13.0 - 35.8) |
| MDCD | 1,331 | 2,154 | 0.7 | 1.0 | 10 | 24 | 15.3 (7.33 - 28.1) | 25.0 (16.0 - 37.2) |
| Open Claims | 160,005 | 255,636 | 96.8 | 160.6 | 732 | 1,411 | 7.56 (7.02 - 8.13) | 8.78 (8.33 - 9.25) |
| OEHR | 9,389 | 16,143 | 2.4 | 4.4 | 24 | 49 | 10.2 (6.53 - 15.2) | 11.1 (8.22 - 14.7) |
| Meta-analysis | 188,240 | 303,930 | 109.6 | 183.9 | 871 | 1,698 | 7.95 (7.43 - 8.49) | 9.23 (8.80 - 9.68) |
| Meta-analysis (w/o USOC) | 28,235 | 48,294 | 12.8 | 23.2 | 139 | 287 | 10.9 (9.14 - 12.8) | 12.3 (11.0 - 13.9) |
| **PS-stratification / on-treatment TAR without escalation** | | | | | | | | |
| CCAE | 13,882 | 18,202 | 7.7 | 11.4 | 37 | 102 | 4.79 (3.37 - 6.60) | 8.95 (7.30 - 10.9) |
| ODOD | 9,492 | 11,798 | 4.6 | 6.5 | 89 | 112 | 19.2 (15.5 - 23.7) | 17.3 (14.2 - 20.8) |
| MDCR | 736 | 1,334 | 0.4 | 0.8 | 9 | 17 | 21.6 (9.88 - 41.0) | 22.3 (13.0 - 35.8) |
| MDCD | 1,563 | 2,158 | 0.8 | 1.0 | 14 | 24 | 18.7 (10.2 - 31.3) | 24.9 (16.0 - 37.1) |
| Open Claims | 218,158 | 255,645 | 122.8 | 160.7 | 905 | 1,411 | 7.37 (6.90 - 7.87) | 8.78 (8.33 - 9.25) |
| OEHR | 12,521 | 16,145 | 3.1 | 4.4 | 33 | 49 | 10.5 (7.26 - 14.8) | 11.1 (8.22 - 14.7) |
| Meta-analysis | 255,616 | 303,948 | 139.0 | 183.9 | 1,078 | 1,698 | 7.75 (7.30 - 8.23) | 9.23 (8.80 - 9.68) |
| Meta-analysis (w/o USOC) | 37,458 | 48,303 | 16.2 | 23.2 | 173 | 287 | 10.7 (9.13 - 12.4) | 12.3 (11.0 - 13.9) |

##### 4.1.3 Acute myocardial infarction

**Supplementary Table 288:** Exposures, outcomes and risk estimates for Acute myocardial infarction in GLP1RA (female) (T) vs SGLT2I (female) (C) new-user cohorts under four different propensity score (PS) adjustment / time-at-risk (TAR) designs across data sources. We report total patient exposure time in 1,000 patient-year (KYr) units, uncalibrated and calibrated hazard ratios (HRs) and their 95% confidence intervals (CIs). Greyed out entries did not pass study diagnostics and are included only for completeness.

| | Patients | | Exposure (KYr) | | Outcomes | | Incidence (per KYr) | |
|---|---|---|---|---|---|---|---|---|
| | T | C | T | C | T | C | T | C |
| **PS-matching / on-treatment TAR** | | | | | | | | |
| CCAE | 10,787 | 18,670 | 7.0 | 13.5 | 14 | 35 | 1.99 (1.09 - 3.34) | 2.59 (1.80 - 3.60) |
| ODOD | 7,385 | 12,541 | 4.2 | 7.7 | 20 | 34 | 4.77 (2.91 - 7.37) | 4.40 (3.05 - 6.15) |
| MDCR | 723 | 1,439 | 0.4 | 0.9 | <5 | 7 | < 11.5 (3.73 - 26.8) | 7.63 (3.07 - 15.7) |
| MDCD | 1,449 | 2,357 | 0.8 | 1.3 | <5 | 8 | < 6.14 (1.99 - 14.3) | 6.32 (2.73 - 12.4) |
| Open Claims | 167,713 | 269,863 | 113.4 | 200.4 | 237 | 547 | 2.09 (1.83 - 2.37) | 2.73 (2.51 - 2.97) |
| OEHR | 9,618 | 16,489 | 2.8 | 5.2 | 12 | 17 | 4.28 (2.21 - 7.48) | 3.28 (1.91 - 5.25) |
| Meta-analysis | 195,503 | 317,563 | 127.4 | 226.8 | 283 | 633 | 2.22 (1.97 - 2.50) | 2.79 (2.58 - 3.02) |
| Meta-analysis (w/o USOC) | 27,790 | 47,700 | 14.0 | 26.4 | 46 | 86 | 3.28 (2.40 - 4.37) | 3.25 (2.60 - 4.02) |
| **PS-stratification / on-treatment TAR** | | | | | | | | |
| CCAE | 14,254 | 18,670 | 8.7 | 13.5 | 18 | 35 | 2.06 (1.22 - 3.25) | 2.59 (1.80 - 3.60) |
| ODOD | 9,933 | 12,545 | 5.3 | 7.7 | 25 | 34 | 4.71 (3.05 - 6.96) | 4.40 (3.04 - 6.14) |
| MDCR | 805 | 1,439 | 0.5 | 0.9 | <5 | 7 | < 10.6 (3.46 - 24.8) | 7.63 (3.07 - 15.7) |
| MDCD | 1,709 | 2,362 | 0.9 | 1.3 | <5 | 8 | < 5.27 (1.71 - 12.3) | 6.30 (2.72 - 12.4) |
| Open Claims | 227,503 | 269,891 | 141.8 | 200.4 | 281 | 548 | 1.98 (1.76 - 2.23) | 2.73 (2.51 - 2.97) |
| OEHR | 12,808 | 16,491 | 3.7 | 5.2 | 15 | 17 | 4.06 (2.27 - 6.70) | 3.28 (1.91 - 5.25) |
| Meta-analysis | 264,498 | 317,597 | 159.5 | 226.8 | 339 | 634 | 2.13 (1.90 - 2.36) | 2.80 (2.58 - 3.02) |
| Meta-analysis (w/o USOC) | 36,995 | 47,706 | 17.7 | 26.5 | 58 | 86 | 3.27 (2.48 - 4.23) | 3.25 (2.60 - 4.02) |
| **PS-matching / on-treatment TAR without escalation** | | | | | | | | |
| CCAE | 10,787 | 18,670 | 6.3 | 11.7 | 12 | 28 | 1.89 (0.98 - 3.31) | 2.40 (1.59 - 3.47) |
| ODOD | 7,385 | 12,541 | 3.8 | 6.9 | 18 | 29 | 4.70 (2.79 - 7.43) | 4.23 (2.83 - 6.08) |
| MDCR | 723 | 1,439 | 0.4 | 0.8 | <5 | 6 | < 12.2 (3.97 - 28.5) | 7.39 (2.71 - 16.1) |
| MDCD | 1,449 | 2,357 | 0.7 | 1.0 | <5 | 6 | < 7.11 (2.31 - 16.6) | 5.74 (2.11 - 12.5) |
| Open Claims | 167,713 | 269,863 | 101.3 | 168.5 | 210 | 457 | 2.07 (1.80 - 2.37) | 2.71 (2.47 - 2.97) |
| OEHR | 9,618 | 16,489 | 2.4 | 4.5 | 9 | 16 | 3.72 (1.70 - 7.06) | 3.55 (2.03 - 5.76) |
| Meta-analysis | 195,503 | 317,563 | 113.9 | 191.5 | 249 | 530 | 2.19 (1.92 - 2.48) | 2.77 (2.54 - 3.01) |
| Meta-analysis (w/o USOC) | 27,790 | 47,700 | 12.6 | 23.0 | 39 | 73 | 3.10 (2.20 - 4.23) | 3.17 (2.48 - 3.99) |
| **PS-stratification / on-treatment TAR without escalation** | | | | | | | | |
| CCAE | 14,254 | 18,670 | 7.9 | 11.7 | 16 | 28 | 2.01 (1.15 - 3.27) | 2.40 (1.59 - 3.47) |
| ODOD | 9,933 | 12,545 | 4.9 | 6.9 | 23 | 29 | 4.72 (2.99 - 7.08) | 4.23 (2.83 - 6.07) |
| MDCR | 805 | 1,439 | 0.4 | 0.8 | <5 | 6 | < 11.4 (3.70 - 26.6) | 7.39 (2.71 - 16.1) |
| MDCD | 1,709 | 2,362 | 0.8 | 1.0 | <5 | 6 | < 6.11 (1.98 - 14.2) | 5.72 (2.10 - 12.4) |
| Open Claims | 227,503 | 269,891 | 128.0 | 168.5 | 251 | 458 | 1.96 (1.73 - 2.22) | 2.72 (2.48 - 2.98) |
| OEHR | 12,808 | 16,491 | 3.2 | 4.5 | 12 | 16 | 3.74 (1.93 - 6.53) | 3.55 (2.03 - 5.76) |
| Meta-analysis | 264,498 | 317,597 | 144.0 | 191.5 | 302 | 531 | 2.10 (1.87 - 2.35) | 2.77 (2.54 - 3.02) |
| Meta-analysis (w/o USOC) | 36,995 | 47,706 | 16.0 | 23.0 | 51 | 73 | 3.18 (2.37 - 4.18) | 3.17 (2.48 - 3.98) |

##### 4.1.4 Acute renal failure

**Supplementary Table 289:** Exposures, outcomes and risk estimates for Acute renal failure in GLP1RA (female) (T) vs SGLT2I (female) (C) new-user cohorts under four different propensity score (PS) adjustment / time-at-risk (TAR) designs across data sources. We report total patient exposure time in 1,000 patient-year (KYr) units, uncalibrated and calibrated hazard ratios (HRs) and their 95% confidence intervals (CIs). Greyed out entries did not pass study diagnostics and are included only for completeness.

| | Patients | | Exposure (KYr) | | Outcomes | | Incidence (per KYr) | |
|---|---|---|---|---|---|---|---|---|
| | T | C | T | C | T | C | T | C |
| **PS-matching / on-treatment TAR** | | | | | | | | |
| CCAE | 10,761 | 18,692 | 7.0 | 13.5 | 39 | 62 | 5.56 (3.95 - 7.60) | 4.58 (3.51 - 5.87) |
| ODOD | 7,251 | 12,443 | 4.1 | 7.6 | 58 | 120 | 14.2 (10.8 - 18.3) | 15.7 (13.0 - 18.8) |
| MDCR | 702 | 1,448 | 0.4 | 0.9 | 8 | 13 | 18.9 (8.14 - 37.2) | 14.0 (7.48 - 24.0) |
| MDCD | 1,404 | 2,308 | 0.8 | 1.2 | 13 | 21 | 16.8 (8.94 - 28.7) | 17.0 (10.5 - 25.9) |
| Open Claims | 166,240 | 269,436 | 112.0 | 200.1 | 766 | 1,385 | 6.84 (6.36 - 7.34) | 6.92 (6.56 - 7.30) |
| OEHR | 9,599 | 16,499 | 2.8 | 5.2 | 35 | 39 | 12.5 (8.74 - 17.4) | 7.53 (5.36 - 10.3) |
| Meta-analysis | 195,255 | 319,378 | 126.7 | 227.7 | 911 | 1,627 | 7.19 (6.73 - 7.67) | 7.15 (6.80 - 7.50) |
| Meta-analysis (w/o USOC) | 29,015 | 49,942 | 14.7 | 27.6 | 145 | 242 | 9.88 (8.34 - 11.6) | 8.77 (7.70 - 9.95) |
| **PS-stratification / on-treatment TAR** | | | | | | | | |
| CCAE | 14,196 | 18,692 | 8.7 | 13.5 | 56 | 62 | 6.43 (4.86 - 8.36) | 4.58 (3.51 - 5.87) |
| ODOD | 9,772 | 12,447 | 5.2 | 7.6 | 71 | 120 | 13.7 (10.7 - 17.2) | 15.7 (13.0 - 18.8) |
| MDCR | 783 | 1,449 | 0.5 | 0.9 | 8 | 13 | 17.6 (7.59 - 34.6) | 14.0 (7.47 - 24.0) |
| MDCD | 1,659 | 2,313 | 0.9 | 1.2 | 16 | 21 | 17.7 (10.1 - 28.7) | 16.9 (10.5 - 25.8) |
| Open Claims | 225,203 | 269,448 | 140.0 | 200.1 | 936 | 1,386 | 6.69 (6.26 - 7.13) | 6.93 (6.57 - 7.30) |
| OEHR | 12,747 | 16,501 | 3.7 | 5.2 | 43 | 39 | 11.7 (8.49 - 15.8) | 7.53 (5.36 - 10.3) |
| Meta-analysis | 263,577 | 319,401 | 158.5 | 227.7 | 1,122 | 1,628 | 7.08 (6.67 - 7.51) | 7.15 (6.81 - 7.51) |
| Meta-analysis (w/o USOC) | 38,374 | 49,953 | 18.5 | 27.6 | 186 | 242 | 10.1 (8.67 - 11.6) | 8.77 (7.70 - 9.95) |
| **PS-matching / on-treatment TAR without escalation** | | | | | | | | |
| CCAE | 10,761 | 18,692 | 6.3 | 11.7 | 34 | 53 | 5.38 (3.72 - 7.51) | 4.54 (3.40 - 5.94) |
| ODOD | 7,251 | 12,443 | 3.7 | 6.8 | 51 | 103 | 13.6 (10.1 - 17.9) | 15.2 (12.4 - 18.4) |
| MDCR | 702 | 1,448 | 0.4 | 0.8 | 7 | 12 | 17.5 (7.05 - 36.1) | 14.6 (7.54 - 25.5) |
| MDCD | 1,404 | 2,308 | 0.7 | 1.0 | 12 | 14 | 17.9 (9.27 - 31.3) | 13.7 (7.47 - 22.9) |
| Open Claims | 166,240 | 269,436 | 100.1 | 168.4 | 650 | 1,116 | 6.49 (6.00 - 7.01) | 6.63 (6.25 - 7.03) |
| OEHR | 9,599 | 16,499 | 2.4 | 4.5 | 23 | 29 | 9.52 (6.04 - 14.3) | 6.44 (4.31 - 9.25) |
| Meta-analysis | 195,255 | 319,378 | 113.2 | 192.3 | 770 | 1,315 | 6.80 (6.33 - 7.30) | 6.84 (6.47 - 7.22) |
| Meta-analysis (w/o USOC) | 29,015 | 49,942 | 13.2 | 24.0 | 120 | 199 | 9.12 (7.57 - 10.9) | 8.30 (7.19 - 9.54) |
| **PS-stratification / on-treatment TAR without escalation** | | | | | | | | |
| CCAE | 14,196 | 18,692 | 7.9 | 11.7 | 50 | 53 | 6.33 (4.70 - 8.34) | 4.54 (3.40 - 5.94) |
| ODOD | 9,772 | 12,447 | 4.8 | 6.8 | 64 | 103 | 13.4 (10.3 - 17.1) | 15.2 (12.4 - 18.4) |
| MDCR | 783 | 1,449 | 0.4 | 0.8 | 7 | 12 | 16.4 (6.58 - 33.7) | 14.6 (7.53 - 25.5) |
| MDCD | 1,659 | 2,313 | 0.8 | 1.0 | 14 | 14 | 17.9 (9.78 - 30.0) | 13.6 (7.44 - 22.8) |
| Open Claims | 225,203 | 269,448 | 126.4 | 168.4 | 809 | 1,117 | 6.40 (5.97 - 6.86) | 6.63 (6.25 - 7.04) |
| OEHR | 12,747 | 16,501 | 3.2 | 4.5 | 29 | 29 | 9.07 (6.08 - 13.0) | 6.44 (4.31 - 9.24) |
| Meta-analysis | 263,577 | 319,401 | 143.0 | 192.4 | 966 | 1,316 | 6.75 (6.33 - 7.19) | 6.84 (6.48 - 7.22) |
| Meta-analysis (w/o USOC) | 38,374 | 49,953 | 16.7 | 24.0 | 157 | 199 | 9.43 (8.01 - 11.0) | 8.30 (7.19 - 9.53) |

##### 4.1.5 Glycemic control

**Supplementary Table 290:** Exposures, outcomes and risk estimates for Glycemic control in GLP1RA (female) (T) vs SGLT2I (female) (C) new-user cohorts under four different propensity score (PS) adjustment / time-at-risk (TAR) designs across data sources. We report total patient exposure time in 1,000 patient-year (KYr) units, uncalibrated and calibrated hazard ratios (HRs) and their 95% confidence intervals (CIs). Greyed out entries did not pass study diagnostics and are included only for completeness.

| | Patients | | Exposure (KYr) | | Outcomes | | Incidence (per KYr) | |
|---|---|---|---|---|---|---|---|---|
| | T | C | T | C | T | C | T | C |
| **PS-matching / on-treatment TAR** | | | | | | | | |
| CCAE | 10,790 | 18,683 | 7.0 | 13.5 | 10 | 22 | 1.42 (0.68 - 2.61) | 1.62 (1.02 - 2.46) |
| ODOD | 4,647 | 7,822 | 2.5 | 4.4 | 290 | 456 | 117 (104 - 131) | 103 (93.6 - 113) |
| MDCR | 732 | 1,481 | 0.4 | 0.9 | 0 | 0 | | |
| MDCD | 1,485 | 2,430 | 0.8 | 1.3 | 0 | 0 | | |
| Open Claims | 170,783 | 276,514 | 115.3 | 205.0 | 0 | 0 | | |
| OEHR | 9,714 | 16,664 | 2.8 | 5.2 | 0 | 0 | | |
| Meta-analysis | 15,437 | 26,505 | 9.5 | 18.0 | 300 | 478 | 31.5 (28.1 - 35.3) | 26.6 (24.3 - 29.1) |
| Meta-analysis (w/o USOC) | 15,437 | 26,505 | 9.5 | 18.0 | 300 | 478 | 31.5 (28.1 - 35.3) | 26.6 (24.3 - 29.1) |
| **PS-stratification / on-treatment TAR** | | | | | | | | |
| CCAE | 14,261 | 18,683 | 8.7 | 13.5 | 11 | 22 | 1.26 (0.63 - 2.25) | 1.62 (1.02 - 2.46) |
| ODOD | 6,078 | 7,839 | 3.1 | 4.4 | 346 | 456 | 113 (101 - 126) | 103 (93.4 - 113) |
| MDCR | 815 | 1,482 | 0.5 | 0.9 | 0 | 0 | | |
| MDCD | 1,747 | 2,435 | 1.0 | 1.3 | 0 | 0 | | |
| Open Claims | 230,932 | 276,521 | 143.9 | 205.0 | 0 | 0 | | |
| OEHR | 12,899 | 16,666 | 3.7 | 5.2 | 0 | 0 | | |
| Meta-analysis | 20,339 | 26,522 | 11.8 | 18.0 | 357 | 478 | 30.3 (27.2 - 33.6) | 26.6 (24.2 - 29.1) |
| Meta-analysis (w/o USOC) | 20,339 | 26,522 | 11.8 | 18.0 | 357 | 478 | 30.3 (27.2 - 33.6) | 26.6 (24.2 - 29.1) |
| **PS-matching / on-treatment TAR without escalation** | | | | | | | | |
| CCAE | 10,790 | 18,683 | 6.3 | 11.7 | 10 | 18 | 1.58 (0.76 - 2.90) | 1.54 (0.91 - 2.44) |
| ODOD | 4,647 | 7,822 | 2.3 | 3.9 | 261 | 397 | 116 (102 - 131) | 101 (91.4 - 112) |
| MDCR | 732 | 1,481 | 0.4 | 0.8 | 0 | 0 | | |
| MDCD | 1,485 | 2,430 | 0.7 | 1.1 | 0 | 0 | | |
| Open Claims | 170,783 | 276,514 | 103.0 | 172.3 | 0 | 0 | | |
| OEHR | 9,714 | 16,664 | 2.4 | 4.6 | 0 | 0 | | |
| Meta-analysis | 15,437 | 26,505 | 8.6 | 15.6 | 271 | 415 | 31.5 (27.9 - 35.5) | 26.6 (24.1 - 29.3) |
| Meta-analysis (w/o USOC) | 15,437 | 26,505 | 8.6 | 15.6 | 271 | 415 | 31.5 (27.9 - 35.5) | 26.6 (24.1 - 29.3) |
| **PS-stratification / on-treatment TAR without escalation** | | | | | | | | |
| CCAE | 14,261 | 18,683 | 7.9 | 11.7 | 11 | 18 | 1.39 (0.69 - 2.48) | 1.54 (0.91 - 2.44) |
| ODOD | 6,078 | 7,839 | 2.8 | 3.9 | 313 | 397 | 112 (100 - 125) | 101 (91.3 - 111) |
| MDCR | 815 | 1,482 | 0.4 | 0.8 | 0 | 0 | | |
| MDCD | 1,747 | 2,435 | 0.8 | 1.1 | 0 | 0 | | |
| Open Claims | 230,932 | 276,521 | 129.8 | 172.3 | 0 | 0 | | |
| OEHR | 12,899 | 16,666 | 3.2 | 4.6 | 0 | 0 | | |
| Meta-analysis | 20,339 | 26,522 | 10.7 | 15.6 | 324 | 415 | 30.2 (27.0 - 33.7) | 26.6 (24.1 - 29.3) |
| Meta-analysis (w/o USOC) | 20,339 | 26,522 | 10.7 | 15.6 | 324 | 415 | 30.2 (27.0 - 33.7) | 26.6 (24.1 - 29.3) |

##### 4.1.6 Hospitalization with heart failure

**Supplementary Table 291:** Exposures, outcomes and risk estimates for Hospitalization with heart failure in GLP1RA (female) (T) vs SGLT2I (female) (C) new-user cohorts under four different propensity score (PS) adjustment / time-at-risk (TAR) designs across data sources. We report total patient exposure time in 1,000 patient-year (KYr) units, uncalibrated and calibrated hazard ratios (HRs) and their 95% confidence intervals (CIs). Greyed out entries did not pass study diagnostics and are included only for completeness.

| | Patients | | Exposure (KYr) | | Outcomes | | Incidence (per KYr) | |
|---|---|---|---|---|---|---|---|---|
| | T | C | T | C | T | C | T | C |
| **PS-matching / on-treatment TAR** | | | | | | | | |
| CCAE | 10,691 | 18,562 | 7.0 | 13.5 | 12 | 47 | 1.72 (0.89 - 3.00) | 3.49 (2.56 - 4.64) |
| ODOD | 7,194 | 12,161 | 4.1 | 7.5 | 55 | 82 | 13.5 (10.2 - 17.6) | 10.9 (8.68 - 13.5) |
| MDCR | 684 | 1,394 | 0.4 | 0.9 | 7 | 14 | 16.5 (6.62 - 33.9) | 15.7 (8.60 - 26.4) |
| MDCD | 1,382 | 2,261 | 0.8 | 1.2 | 11 | 20 | 14.0 (7.01 - 25.1) | 16.4 (10.0 - 25.3) |
| Open Claims | 165,236 | 265,014 | 111.7 | 197.8 | 493 | 960 | 4.42 (4.03 - 4.82) | 4.85 (4.55 - 5.17) |
| OEHR | 9,525 | 16,385 | 2.8 | 5.1 | 20 | 36 | 7.22 (4.41 - 11.1) | 7.00 (4.90 - 9.69) |
| Meta-analysis | 194,028 | 314,383 | 126.3 | 225.1 | 591 | 1,145 | 4.68 (4.31 - 5.07) | 5.09 (4.80 - 5.39) |
| Meta-analysis (w/o USOC) | 28,792 | 49,369 | 14.6 | 27.3 | 98 | 185 | 6.71 (5.45 - 8.17) | 6.76 (5.82 - 7.81) |
| **PS-stratification / on-treatment TAR** | | | | | | | | |
| CCAE | 14,134 | 18,562 | 8.7 | 13.5 | 12 | 47 | 1.38 (0.71 - 2.42) | 3.49 (2.56 - 4.64) |
| ODOD | 9,699 | 12,165 | 5.2 | 7.5 | 60 | 82 | 11.6 (8.85 - 14.9) | 10.9 (8.67 - 13.5) |
| MDCR | 771 | 1,395 | 0.5 | 0.9 | 8 | 14 | 17.5 (7.54 - 34.4) | 15.7 (8.58 - 26.3) |
| MDCD | 1,619 | 2,266 | 0.9 | 1.2 | 16 | 20 | 17.8 (10.2 - 29.0) | 16.3 (9.98 - 25.2) |
| Open Claims | 224,509 | 265,019 | 139.9 | 197.8 | 587 | 960 | 4.20 (3.86 - 4.55) | 4.85 (4.55 - 5.17) |
| OEHR | 12,672 | 16,387 | 3.6 | 5.1 | 25 | 36 | 6.86 (4.44 - 10.1) | 7.00 (4.90 - 9.69) |
| Meta-analysis | 262,633 | 314,399 | 158.3 | 225.1 | 700 | 1,145 | 4.42 (4.10 - 4.76) | 5.09 (4.80 - 5.39) |
| Meta-analysis (w/o USOC) | 38,124 | 49,380 | 18.4 | 27.4 | 113 | 185 | 6.14 (5.06 - 7.39) | 6.76 (5.82 - 7.81) |
| **PS-matching / on-treatment TAR without escalation** | | | | | | | | |
| CCAE | 10,691 | 18,562 | 6.3 | 11.6 | 9 | 42 | 1.43 (0.65 - 2.72) | 3.61 (2.60 - 4.89) |
| ODOD | 7,194 | 12,161 | 3.7 | 6.7 | 49 | 70 | 13.1 (9.72 - 17.4) | 10.5 (8.17 - 13.2) |
| MDCR | 684 | 1,394 | 0.4 | 0.8 | 6 | 13 | 14.9 (5.47 - 32.5) | 16.5 (8.79 - 28.2) |
| MDCD | 1,382 | 2,261 | 0.7 | 1.0 | 10 | 14 | 14.8 (7.08 - 27.2) | 13.9 (7.59 - 23.3) |
| Open Claims | 165,236 | 265,014 | 99.8 | 166.2 | 425 | 794 | 4.26 (3.86 - 4.68) | 4.78 (4.45 - 5.12) |
| OEHR | 9,525 | 16,385 | 2.4 | 4.5 | 15 | 29 | 6.25 (3.50 - 10.3) | 6.48 (4.34 - 9.31) |
| Meta-analysis | 194,028 | 314,383 | 112.9 | 190.0 | 508 | 949 | 4.50 (4.12 - 4.91) | 5.00 (4.68 - 5.32) |
| Meta-analysis (w/o USOC) | 28,792 | 49,369 | 13.1 | 23.8 | 83 | 155 | 6.34 (5.05 - 7.86) | 6.52 (5.53 - 7.63) |
| **PS-stratification / on-treatment TAR without escalation** | | | | | | | | |
| CCAE | 14,134 | 18,562 | 7.9 | 11.6 | 9 | 42 | 1.14 (0.52 - 2.17) | 3.61 (2.60 - 4.89) |
| ODOD | 9,699 | 12,165 | 4.7 | 6.7 | 54 | 70 | 11.4 (8.54 - 14.8) | 10.5 (8.17 - 13.2) |
| MDCR | 771 | 1,395 | 0.4 | 0.8 | 7 | 13 | 16.3 (6.55 - 33.6) | 16.5 (8.78 - 28.2) |
| MDCD | 1,619 | 2,266 | 0.8 | 1.0 | 13 | 14 | 16.8 (8.94 - 28.7) | 13.8 (7.57 - 23.2) |
| Open Claims | 224,509 | 265,019 | 126.3 | 166.2 | 507 | 794 | 4.01 (3.67 - 4.38) | 4.78 (4.45 - 5.12) |
| OEHR | 12,672 | 16,387 | 3.2 | 4.5 | 20 | 29 | 6.29 (3.84 - 9.71) | 6.48 (4.34 - 9.31) |
| Meta-analysis | 262,633 | 314,399 | 142.9 | 190.0 | 603 | 949 | 4.22 (3.89 - 4.57) | 5.00 (4.68 - 5.32) |
| Meta-analysis (w/o USOC) | 38,124 | 49,380 | 16.6 | 23.8 | 96 | 155 | 5.79 (4.69 - 7.07) | 6.52 (5.53 - 7.63) |

##### 4.1.7 Measured renal dysfunction

**Supplementary Table 292:** Exposures, outcomes and risk estimates for Measured renal dysfunction in GLP1RA (female) (T) vs SGLT2I (female) (C) new-user cohorts under four different propensity score (PS) adjustment / time-at-risk (TAR) designs across data sources. We report total patient exposure time in 1,000 patient-year (KYr) units, uncalibrated and calibrated hazard ratios (HRs) and their 95% confidence intervals (CIs). Greyed out entries did not pass study diagnostics and are included only for completeness.

| | Patients | | Exposure (KYr) | | Outcomes | | Incidence (per KYr) | |
|---|---|---|---|---|---|---|---|---|
| | T | C | T | C | T | C | T | C |
| **PS-matching / on-treatment TAR** | | | | | | | | |
| CCAE | 10,862 | 18,849 | 7.1 | 13.7 | <5 | <5 | < 0.70 (0.23 - 1.64) | < 0.37 (0.12 - 0.85) |
| ODOD | 7,487 | 12,830 | 4.2 | 7.9 | <5 | 10 | < 1.18 (0.38 - 2.75) | 1.27 (0.61 - 2.34) |
| MDCR | 736 | 1,488 | 0.4 | 0.9 | 0 | 0 | | |
| MDCD | 1,485 | 2,430 | 0.8 | 1.3 | 0 | 0 | | |
| Open Claims | 170,783 | 276,514 | 115.3 | 205.0 | 0 | 0 | | |
| OEHR | 9,714 | 16,664 | 2.8 | 5.2 | 0 | 0 | | |
| **PS-stratification / on-treatment TAR** | | | | | | | | |
| CCAE | 14,338 | 18,849 | 8.8 | 13.7 | <5 | <5 | < 0.57 (0.18 - 1.32) | < 0.37 (0.12 - 0.85) |
| ODOD | 10,060 | 12,834 | 5.4 | 7.9 | <5 | 10 | < 0.93 (0.30 - 2.17) | 1.27 (0.61 - 2.34) |
| MDCR | 819 | 1,489 | 0.5 | 0.9 | 0 | 0 | | |
| MDCD | 1,747 | 2,435 | 1.0 | 1.3 | 0 | 0 | | |
| Open Claims | 230,932 | 276,521 | 143.9 | 205.0 | 0 | 0 | | |
| OEHR | 12,899 | 16,666 | 3.7 | 5.2 | 0 | 0 | | |
| **PS-matching / on-treatment TAR without escalation** | | | | | | | | |
| CCAE | 10,862 | 18,849 | 6.4 | 11.8 | <5 | <5 | < 0.78 (0.25 - 1.82) | < 0.42 (0.14 - 0.99) |
| ODOD | 7,487 | 12,830 | 3.9 | 7.0 | <5 | 8 | < 1.29 (0.42 - 3.01) | 1.15 (0.50 - 2.26) |
| MDCR | 736 | 1,488 | 0.4 | 0.8 | 0 | 0 | | |
| MDCD | 1,485 | 2,430 | 0.7 | 1.1 | 0 | 0 | | |
| Open Claims | 170,783 | 276,514 | 103.0 | 172.3 | 0 | 0 | | |
| OEHR | 9,714 | 16,664 | 2.4 | 4.6 | 0 | 0 | | |
| **PS-stratification / on-treatment TAR without escalation** | | | | | | | | |
| CCAE | 14,338 | 18,849 | 8.0 | 11.8 | <5 | <5 | < 0.63 (0.20 - 1.46) | < 0.42 (0.14 - 0.99) |
| ODOD | 10,060 | 12,834 | 4.9 | 7.0 | <5 | 8 | < 1.01 (0.33 - 2.37) | 1.15 (0.50 - 2.26) |
| MDCR | 819 | 1,489 | 0.4 | 0.8 | 0 | 0 | | |
| MDCD | 1,747 | 2,435 | 0.8 | 1.1 | 0 | 0 | | |
| Open Claims | 230,932 | 276,521 | 129.8 | 172.3 | 0 | 0 | | |
| OEHR | 12,899 | 16,666 | 3.2 | 4.6 | 0 | 0 | | |

##### 4.1.8 Stroke

**Supplementary Table 293:** Exposures, outcomes and risk estimates for Stroke in GLP1RA (female) (T) vs SGLT2I (female) (C) new-user cohorts under four different propensity score (PS) adjustment / time-at-risk (TAR) designs across data sources. We report total patient exposure time in 1,000 patient-year (KYr) units, uncalibrated and calibrated hazard ratios (HRs) and their 95% confidence intervals (CIs). Greyed out entries did not pass study diagnostics and are included only for completeness.

| | Patients | | Exposure (KYr) | | Outcomes | | Incidence (per KYr) | |
|---|---|---|---|---|---|---|---|---|
| | T | C | T | C | T | C | T | C |
| **PS-matching / on-treatment TAR** | | | | | | | | |
| CCAE | 10,750 | 18,673 | 7.0 | 13.5 | 17 | 51 | 2.42 (1.41 - 3.88) | 3.77 (2.81 - 4.96) |
| ODOD | 7,366 | 12,560 | 4.2 | 7.7 | 26 | 53 | 6.26 (4.09 - 9.17) | 6.89 (5.16 - 9.02) |
| MDCR | 711 | 1,452 | 0.4 | 0.9 | <5 | 5 | < 11.6 (3.75 - 27.0) | 5.40 (1.75 - 12.6) |
| MDCD | 1,435 | 2,340 | 0.8 | 1.3 | 6 | 12 | 7.40 (2.72 - 16.1) | 9.58 (4.95 - 16.7) |
| Open Claims | 167,178 | 270,181 | 113.0 | 200.6 | 324 | 602 | 2.87 (2.56 - 3.20) | 3.00 (2.77 - 3.25) |
| OEHR | 9,640 | 16,550 | 2.8 | 5.2 | 7 | 14 | 2.50 (1.00 - 5.15) | 2.69 (1.47 - 4.52) |
| Meta-analysis | 196,369 | 320,304 | 127.7 | 228.3 | 380 | 732 | 2.97 (2.68 - 3.29) | 3.21 (2.98 - 3.45) |
| Meta-analysis (w/o USOC) | 29,191 | 50,123 | 14.8 | 27.7 | 56 | 130 | 3.79 (2.86 - 4.92) | 4.70 (3.93 - 5.58) |
| **PS-stratification / on-treatment TAR** | | | | | | | | |
| CCAE | 14,188 | 18,673 | 8.7 | 13.5 | 22 | 51 | 2.52 (1.58 - 3.82) | 3.77 (2.81 - 4.96) |
| ODOD | 9,918 | 12,563 | 5.3 | 7.7 | 33 | 53 | 6.27 (4.32 - 8.81) | 6.89 (5.16 - 9.01) |
| MDCR | 791 | 1,453 | 0.5 | 0.9 | <5 | 5 | < 10.7 (3.49 - 25.1) | 5.40 (1.75 - 12.6) |
| MDCD | 1,694 | 2,344 | 0.9 | 1.3 | 7 | 12 | 7.41 (2.98 - 15.3) | 9.56 (4.94 - 16.7) |
| Open Claims | 226,441 | 270,188 | 141.2 | 200.6 | 396 | 602 | 2.81 (2.54 - 3.10) | 3.00 (2.77 - 3.25) |
| OEHR | 12,803 | 16,552 | 3.7 | 5.2 | 9 | 14 | 2.45 (1.12 - 4.64) | 2.69 (1.47 - 4.52) |
| Meta-analysis | 265,044 | 320,320 | 159.8 | 228.3 | 467 | 732 | 2.92 (2.66 - 3.20) | 3.21 (2.98 - 3.45) |
| Meta-analysis (w/o USOC) | 38,603 | 50,132 | 18.6 | 27.7 | 71 | 130 | 3.82 (2.98 - 4.81) | 4.70 (3.93 - 5.58) |
| **PS-matching / on-treatment TAR without escalation** | | | | | | | | |
| CCAE | 10,750 | 18,673 | 6.3 | 11.7 | 13 | 41 | 2.05 (1.09 - 3.51) | 3.51 (2.52 - 4.77) |
| ODOD | 7,366 | 12,560 | 3.8 | 6.8 | 23 | 50 | 6.05 (3.84 - 9.08) | 7.34 (5.44 - 9.67) |
| MDCR | 711 | 1,452 | 0.4 | 0.8 | <5 | <5 | < 12.3 (3.99 - 28.6) | < 6.08 (1.97 - 14.2) |
| MDCD | 1,435 | 2,340 | 0.7 | 1.0 | 5 | 6 | 7.13 (2.31 - 16.6) | 5.77 (2.12 - 12.6) |
| Open Claims | 167,178 | 270,181 | 100.9 | 168.7 | 278 | 505 | 2.75 (2.44 - 3.10) | 2.99 (2.74 - 3.27) |
| OEHR | 9,640 | 16,550 | 2.4 | 4.5 | 5 | 9 | 2.07 (0.67 - 4.83) | 1.99 (0.91 - 3.78) |
| Meta-analysis | 185,294 | 301,414 | 111.0 | 187.2 | 314 | 596 | 2.83 (2.52 - 3.16) | 3.18 (2.93 - 3.45) |
| Meta-analysis (w/o USOC) | 18,116 | 31,233 | 10.1 | 18.5 | 36 | 91 | 3.56 (2.49 - 4.92) | 4.92 (3.96 - 6.05) |
| **PS-stratification / on-treatment TAR without escalation** | | | | | | | | |
| CCAE | 14,188 | 18,673 | 7.9 | 11.7 | 16 | 41 | 2.02 (1.16 - 3.29) | 3.51 (2.52 - 4.77) |
| ODOD | 9,918 | 12,563 | 4.8 | 6.8 | 29 | 50 | 6.00 (4.02 - 8.61) | 7.33 (5.44 - 9.66) |
| MDCR | 791 | 1,453 | 0.4 | 0.8 | <5 | <5 | < 11.5 (3.72 - 26.7) | < 6.07 (1.97 - 14.2) |
| MDCD | 1,694 | 2,344 | 0.8 | 1.0 | 6 | 6 | 7.36 (2.70 - 16.0) | 5.75 (2.11 - 12.5) |
| Open Claims | 226,441 | 270,188 | 127.4 | 168.7 | 348 | 505 | 2.73 (2.45 - 3.03) | 2.99 (2.74 - 3.27) |
| OEHR | 12,803 | 16,552 | 3.2 | 4.5 | 7 | 9 | 2.19 (0.88 - 4.50) | 1.99 (0.91 - 3.78) |
| Meta-analysis | 250,547 | 301,424 | 140.1 | 187.2 | 393 | 596 | 2.81 (2.53 - 3.10) | 3.18 (2.93 - 3.45) |
| Meta-analysis (w/o USOC) | 24,106 | 31,236 | 12.7 | 18.5 | 45 | 91 | 3.53 (2.58 - 4.73) | 4.92 (3.96 - 6.04) |

###### 4.1.9 Sudden cardiac death

**Supplementary Table 294:** Exposures, outcomes and risk estimates for Sudden cardiac death in GLP1RA (female) (T) vs SGLT2I (female) (C) new-user cohorts under four different propensity score (PS) adjustment / time-at-risk (TAR) designs across data sources. We report total patient exposure time in 1,000 patient-year (KYr) units, uncalibrated and calibrated hazard ratios (HRs) and their 95% confidence intervals (CIs). Greyed out entries did not pass study diagnostics and are included only for completeness.

| | Patients | | Exposure (KYr) | | Outcomes | | Incidence (per KYr) | |
|---|---|---|---|---|---|---|---|---|
| | T | C | T | C | T | C | T | C |
| **PS-matching / on-treatment TAR** | | | | | | | | |
| CCAE | 10,874 | 18,867 | 7.1 | 13.7 | 5 | 8 | 0.70 (0.23 - 1.64) | 0.58 (0.25 - 1.15) |
| ODOD | 7,487 | 12,824 | 4.2 | 7.9 | <5 | 6 | < 1.18 (0.38 - 2.75) | 0.76 (0.28 - 1.66) |
| MDCR | 734 | 1,486 | 0.4 | 0.9 | 0 | 0 | | |
| MDCD | 1,485 | 2,428 | 0.8 | 1.3 | 0 | <5 | | < 3.84 (1.25 - 8.95) |
| Open Claims | 170,550 | 276,019 | 115.2 | 204.6 | 56 | 121 | 0.49 (0.37 - 0.63) | 0.59 (0.49 - 0.71) |
| OEHR | 9,707 | 16,655 | 2.8 | 5.2 | <5 | <5 | < 1.77 (0.57 - 4.12) | < 0.96 (0.31 - 2.23) |
| Meta-analysis | 170,550 | 276,019 | 115.2 | 204.6 | 56 | 121 | 0.49 (0.37 - 0.63) | 0.59 (0.49 - 0.71) |
| **PS-stratification / on-treatment TAR** | | | | | | | | |
| CCAE | 14,348 | 18,867 | 8.8 | 13.7 | 5 | 8 | 0.57 (0.18 - 1.32) | 0.58 (0.25 - 1.15) |
| ODOD | 10,066 | 12,828 | 5.4 | 7.9 | 6 | 6 | 1.12 (0.41 - 2.43) | 0.76 (0.28 - 1.66) |
| MDCR | 815 | 1,487 | 0.5 | 0.9 | 0 | 0 | | |
| MDCD | 1,746 | 2,433 | 1.0 | 1.3 | 0 | <5 | | < 3.83 (1.24 - 8.93) |
| Open Claims | 230,619 | 276,028 | 143.7 | 204.6 | 66 | 121 | 0.46 (0.36 - 0.58) | 0.59 (0.49 - 0.71) |
| OEHR | 12,892 | 16,657 | 3.7 | 5.2 | <5 | <5 | < 1.34 (0.44 - 3.14) | < 0.96 (0.31 - 2.23) |
| Meta-analysis | 230,619 | 276,028 | 143.7 | 204.6 | 66 | 121 | 0.46 (0.36 - 0.58) | 0.59 (0.49 - 0.71) |
| **PS-matching / on-treatment TAR without escalation** | | | | | | | | |
| CCAE | 10,874 | 18,867 | 6.4 | 11.8 | <5 | 7 | < 0.78 (0.25 - 1.82) | 0.59 (0.24 - 1.22) |
| ODOD | 7,487 | 12,824 | 3.9 | 7.0 | <5 | 6 | < 1.29 (0.42 - 3.01) | 0.86 (0.32 - 1.87) |
| MDCR | 734 | 1,486 | 0.4 | 0.8 | 0 | 0 | | |
| MDCD | 1,485 | 2,428 | 0.7 | 1.1 | 0 | <5 | | < 4.65 (1.51 - 10.8) |
| Open Claims | 170,550 | 276,019 | 102.9 | 172.0 | 48 | 105 | 0.47 (0.34 - 0.62) | 0.61 (0.50 - 0.74) |
| OEHR | 9,707 | 16,655 | 2.4 | 4.5 | <5 | <5 | < 2.05 (0.66 - 4.77) | < 1.10 (0.36 - 2.57) |
| Meta-analysis | 170,550 | 276,019 | 102.9 | 172.0 | 48 | 105 | 0.47 (0.34 - 0.62) | 0.61 (0.50 - 0.74) |
| **PS-stratification / on-treatment TAR without escalation** | | | | | | | | |
| CCAE | 14,348 | 18,867 | 8.0 | 11.8 | <5 | 7 | < 0.63 (0.20 - 1.46) | 0.59 (0.24 - 1.22) |
| ODOD | 10,066 | 12,828 | 4.9 | 7.0 | 6 | 6 | 1.22 (0.45 - 2.65) | 0.86 (0.32 - 1.87) |
| MDCR | 815 | 1,487 | 0.4 | 0.8 | 0 | 0 | | |
| MDCD | 1,746 | 2,433 | 0.8 | 1.1 | 0 | <5 | | < 4.63 (1.50 - 10.8) |
| Open Claims | 230,619 | 276,028 | 129.7 | 172.0 | 57 | 105 | 0.44 (0.33 - 0.57) | 0.61 (0.50 - 0.74) |
| OEHR | 12,892 | 16,657 | 3.2 | 4.5 | <5 | <5 | < 1.54 (0.50 - 3.61) | < 1.10 (0.36 - 2.56) |
| Meta-analysis | 230,619 | 276,028 | 129.7 | 172.0 | 57 | 105 | 0.44 (0.33 - 0.57) | 0.61 (0.50 - 0.74) |

##### 4.1.10 Abnormal weight gain

**Supplementary Table 295:** Exposures, outcomes and risk estimates for Abnormal weight gain in GLP1RA (female) (T) vs SGLT2I (female) (C) new-user cohorts under four different propensity score (PS) adjustment / time-at-risk (TAR) designs across data sources. We report total patient exposure time in 1,000 patient-year (KYr) units, uncalibrated and calibrated hazard ratios (HRs) and their 95% confidence intervals (CIs). Greyed out entries did not pass study diagnostics and are included only for completeness.

| | Patients | | Exposure (KYr) | | Outcomes | | Incidence (per KYr) | |
|---|---|---|---|---|---|---|---|---|
| | T | C | T | C | T | C | T | C |
| **PS-matching / on-treatment TAR** | | | | | | | | |
| CCAE | 9,579 | 17,049 | 6.2 | 12.4 | 64 | 80 | 10.3 (7.91 - 13.1) | 6.46 (5.12 - 8.04) |
| ODOD | 6,779 | 11,831 | 3.8 | 7.2 | 38 | 54 | 9.98 (7.06 - 13.7) | 7.54 (5.67 - 9.84) |
| MDCR | 701 | 1,454 | 0.4 | 0.9 | <5 | <5 | < 11.9 (3.86 - 27.7) | < 5.44 (1.77 - 12.7) |
| MDCD | 1,271 | 2,172 | 0.7 | 1.2 | 8 | 5 | 11.1 (4.78 - 21.8) | 4.27 (1.39 - 9.96) |
| Open Claims | 154,383 | 255,338 | 103.9 | 188.9 | 379 | 486 | 3.65 (3.29 - 4.03) | 2.57 (2.35 - 2.81) |
| OEHR | 8,813 | 15,453 | 2.5 | 4.8 | 30 | 29 | 11.8 (7.97 - 16.9) | 6.02 (4.03 - 8.64) |
| Meta-analysis | 179,554 | 299,671 | 116.5 | 213.3 | 511 | 649 | 4.39 (4.01 - 4.78) | 3.04 (2.81 - 3.29) |
| Meta-analysis (w/o USOC) | 25,171 | 44,333 | 12.6 | 24.4 | 132 | 163 | 10.5 (8.78 - 12.4) | 6.69 (5.70 - 7.80) |
| **PS-stratification / on-treatment TAR** | | | | | | | | |
| CCAE | 12,191 | 17,049 | 7.5 | 12.4 | 85 | 80 | 11.3 (9.05 - 14.0) | 6.46 (5.12 - 8.04) |
| ODOD | 8,795 | 11,835 | 4.7 | 7.2 | 47 | 54 | 10.1 (7.39 - 13.4) | 7.54 (5.66 - 9.84) |
| MDCR | 769 | 1,455 | 0.4 | 0.9 | <5 | <5 | < 11.2 (3.63 - 26.1) | < 5.43 (1.76 - 12.7) |
| MDCD | 1,464 | 2,177 | 0.8 | 1.2 | 10 | 5 | 12.3 (5.91 - 22.7) | 4.26 (1.38 - 9.93) |
| Open Claims | 201,930 | 255,345 | 126.3 | 188.9 | 548 | 486 | 4.34 (3.98 - 4.72) | 2.57 (2.35 - 2.81) |
| OEHR | 11,328 | 15,455 | 3.2 | 4.8 | 45 | 29 | 13.9 (10.2 - 18.6) | 6.02 (4.03 - 8.64) |
| Meta-analysis | 234,244 | 299,684 | 141.7 | 213.3 | 725 | 649 | 5.12 (4.75 - 5.50) | 3.04 (2.81 - 3.29) |
| Meta-analysis (w/o USOC) | 32,314 | 44,339 | 15.4 | 24.4 | 177 | 163 | 11.5 (9.86 - 13.3) | 6.69 (5.70 - 7.80) |
| **PS-matching / on-treatment TAR without escalation** | | | | | | | | |
| CCAE | 9,579 | 17,049 | 5.6 | 10.7 | 60 | 68 | 10.7 (8.17 - 13.8) | 6.38 (4.96 - 8.09) |
| ODOD | 6,779 | 11,831 | 3.5 | 6.4 | 35 | 48 | 10.1 (7.01 - 14.0) | 7.54 (5.56 - 10.0) |
| MDCR | 701 | 1,454 | 0.4 | 0.8 | <5 | <5 | < 12.7 (4.12 - 29.6) | < 6.13 (1.99 - 14.3) |
| MDCD | 1,271 | 2,172 | 0.6 | 1.0 | 6 | <5 | 9.61 (3.53 - 20.9) | < 5.19 (1.68 - 12.1) |
| Open Claims | 154,383 | 255,338 | 92.8 | 159.0 | 331 | 388 | 3.57 (3.19 - 3.97) | 2.44 (2.20 - 2.70) |
| OEHR | 8,813 | 15,453 | 2.2 | 4.2 | 24 | 24 | 10.9 (6.98 - 16.2) | 5.71 (3.66 - 8.49) |
| Meta-analysis | 179,554 | 299,671 | 104.1 | 180.2 | 450 | 528 | 4.32 (3.93 - 4.74) | 2.93 (2.69 - 3.19) |
| Meta-analysis (w/o USOC) | 25,171 | 44,333 | 11.3 | 21.2 | 119 | 140 | 10.5 (8.74 - 12.6) | 6.60 (5.55 - 7.78) |
| **PS-stratification / on-treatment TAR without escalation** | | | | | | | | |
| CCAE | 12,191 | 17,049 | 6.8 | 10.7 | 80 | 68 | 11.8 (9.34 - 14.7) | 6.38 (4.96 - 8.09) |
| ODOD | 8,795 | 11,835 | 4.3 | 6.4 | 44 | 48 | 10.3 (7.47 - 13.8) | 7.54 (5.56 - 10.00) |
| MDCR | 769 | 1,455 | 0.4 | 0.8 | <5 | <5 | < 12.0 (3.89 - 28.0) | < 6.12 (1.99 - 14.3) |
| MDCD | 1,464 | 2,177 | 0.7 | 1.0 | 8 | <5 | 11.4 (4.92 - 22.5) | < 5.17 (1.68 - 12.1) |
| Open Claims | 201,930 | 255,345 | 113.7 | 159.0 | 498 | 388 | 4.38 (4.00 - 4.78) | 2.44 (2.20 - 2.70) |
| OEHR | 11,328 | 15,455 | 2.8 | 4.2 | 38 | 24 | 13.5 (9.54 - 18.5) | 5.71 (3.66 - 8.49) |
| Meta-analysis | 234,244 | 299,684 | 127.6 | 180.2 | 660 | 528 | 5.17 (4.78 - 5.58) | 2.93 (2.69 - 3.19) |
| Meta-analysis (w/o USOC) | 32,314 | 44,339 | 13.9 | 21.2 | 162 | 140 | 11.7 (9.93 - 13.6) | 6.59 (5.55 - 7.78) |

###### 4.1.11 Abnormal weight loss

**Supplementary Table 296:** Exposures, outcomes and risk estimates for Abnormal weight loss in GLP1RA (female) (T) vs SGLT2I (female) (C) new-user cohorts under four different propensity score (PS) adjustment / time-at-risk (TAR) designs across data sources. We report total patient exposure time in 1,000 patient-year (KYr) units, uncalibrated and calibrated hazard ratios (HRs) and their 95% confidence intervals (CIs). Greyed out entries did not pass study diagnostics and are included only for completeness.

| | Patients | | Exposure (KYr) | | Outcomes | | Incidence (per KYr) | |
|---|---|---|---|---|---|---|---|---|
| | T | C | T | C | T | C | T | C |
| **PS-matching / on-treatment TAR** | | | | | | | | |
| CCAE | 10,615 | 18,444 | 6.9 | 13.3 | 67 | 112 | 9.71 (7.52 - 12.3) | 8.41 (6.93 - 10.1) |
| ODOD | 7,288 | 12,376 | 4.1 | 7.5 | 68 | 114 | 16.6 (12.9 - 21.0) | 15.1 (12.5 - 18.2) |
| MDCR | 719 | 1,437 | 0.4 | 0.9 | 9 | 11 | 20.9 (9.56 - 39.7) | 12.0 (6.00 - 21.5) |
| MDCD | 1,418 | 2,325 | 0.8 | 1.2 | 12 | 24 | 15.3 (7.90 - 26.7) | 19.4 (12.4 - 28.8) |
| Open Claims | 165,133 | 266,944 | 111.3 | 197.5 | 581 | 1,078 | 5.22 (4.80 - 5.66) | 5.46 (5.14 - 5.79) |
| OEHR | 9,470 | 16,230 | 2.7 | 5.1 | 23 | 46 | 8.37 (5.30 - 12.6) | 9.07 (6.64 - 12.1) |
| Meta-analysis | 193,924 | 316,319 | 125.9 | 224.7 | 751 | 1,374 | 5.97 (5.55 - 6.41) | 6.12 (5.80 - 6.45) |
| Meta-analysis (w/o USOC) | 28,791 | 49,375 | 14.5 | 27.2 | 170 | 296 | 11.7 (10.0 - 13.6) | 10.9 (9.69 - 12.2) |
| **PS-stratification / on-treatment TAR** | | | | | | | | |
| CCAE | 13,986 | 18,444 | 8.5 | 13.3 | 89 | 112 | 10.4 (8.36 - 12.8) | 8.41 (6.93 - 10.1) |
| ODOD | 9,740 | 12,379 | 5.2 | 7.5 | 81 | 115 | 15.7 (12.4 - 19.5) | 15.2 (12.6 - 18.3) |
| MDCR | 800 | 1,438 | 0.5 | 0.9 | 9 | 11 | 19.4 (8.87 - 36.8) | 12.0 (5.99 - 21.5) |
| MDCD | 1,674 | 2,330 | 0.9 | 1.2 | 15 | 24 | 16.4 (9.15 - 27.0) | 19.3 (12.4 - 28.7) |
| Open Claims | 223,055 | 266,948 | 138.7 | 197.5 | 738 | 1,078 | 5.32 (4.94 - 5.72) | 5.46 (5.14 - 5.79) |
| OEHR | 12,562 | 16,231 | 3.6 | 5.1 | 32 | 46 | 8.88 (6.07 - 12.5) | 9.07 (6.64 - 12.1) |
| Meta-analysis | 261,017 | 316,332 | 157.0 | 224.7 | 955 | 1,375 | 6.08 (5.70 - 6.48) | 6.12 (5.80 - 6.45) |
| Meta-analysis (w/o USOC) | 37,962 | 49,384 | 18.2 | 27.2 | 217 | 297 | 11.9 (10.4 - 13.6) | 10.9 (9.72 - 12.2) |
| **PS-matching / on-treatment TAR without escalation** | | | | | | | | |
| CCAE | 10,615 | 18,444 | 6.2 | 11.5 | 56 | 93 | 8.99 (6.79 - 11.7) | 8.10 (6.54 - 9.92) |
| ODOD | 7,288 | 12,376 | 3.7 | 6.7 | 60 | 99 | 16.0 (12.2 - 20.6) | 14.8 (12.0 - 18.0) |
| MDCR | 719 | 1,437 | 0.4 | 0.8 | 9 | 10 | 22.3 (10.2 - 42.3) | 12.3 (5.90 - 22.6) |
| MDCD | 1,418 | 2,325 | 0.7 | 1.0 | 9 | 21 | 13.3 (6.07 - 25.2) | 20.5 (12.7 - 31.3) |
| Open Claims | 165,133 | 266,944 | 99.5 | 166.1 | 517 | 907 | 5.20 (4.76 - 5.66) | 5.46 (5.11 - 5.83) |
| OEHR | 9,470 | 16,230 | 2.4 | 4.4 | 19 | 39 | 8.00 (4.82 - 12.5) | 8.82 (6.27 - 12.1) |
| Meta-analysis | 193,924 | 316,319 | 112.5 | 189.8 | 661 | 1,159 | 5.87 (5.43 - 6.34) | 6.11 (5.76 - 6.47) |
| Meta-analysis (w/o USOC) | 28,791 | 49,375 | 13.0 | 23.6 | 144 | 252 | 11.1 (9.32 - 13.0) | 10.7 (9.40 - 12.1) |
| **PS-stratification / on-treatment TAR without escalation** | | | | | | | | |
| CCAE | 13,986 | 18,444 | 7.8 | 11.5 | 74 | 93 | 9.52 (7.48 - 12.0) | 8.10 (6.54 - 9.92) |
| ODOD | 9,740 | 12,379 | 4.7 | 6.7 | 72 | 100 | 15.2 (11.9 - 19.1) | 15.0 (12.2 - 18.2) |
| MDCR | 800 | 1,438 | 0.4 | 0.8 | 9 | 10 | 20.8 (9.51 - 39.5) | 12.3 (5.89 - 22.6) |
| MDCD | 1,674 | 2,330 | 0.8 | 1.0 | 10 | 21 | 12.6 (6.06 - 23.2) | 20.4 (12.6 - 31.2) |
| Open Claims | 223,055 | 266,948 | 125.2 | 166.1 | 666 | 907 | 5.32 (4.92 - 5.74) | 5.46 (5.11 - 5.83) |
| OEHR | 12,562 | 16,231 | 3.1 | 4.4 | 27 | 39 | 8.60 (5.67 - 12.5) | 8.82 (6.27 - 12.1) |
| Meta-analysis | 261,017 | 316,332 | 141.7 | 189.8 | 849 | 1,160 | 5.99 (5.60 - 6.41) | 6.11 (5.77 - 6.48) |
| Meta-analysis (w/o USOC) | 37,962 | 49,384 | 16.4 | 23.6 | 183 | 253 | 11.1 (9.57 - 12.9) | 10.7 (9.43 - 12.1) |

###### 4.1.12 Acute pancreatitis

**Supplementary Table 297:** Exposures, outcomes and risk estimates for Acute pancreatitis in GLP1RA (female) (T) vs SGLT2I (female) (C) new-user cohorts under four different propensity score (PS) adjustment / time-at-risk (TAR) designs across data sources. We report total patient exposure time in 1,000 patient-year (KYr) units, uncalibrated and calibrated hazard ratios (HRs) and their 95% confidence intervals (CIs). Greyed out entries did not pass study diagnostics and are included only for completeness.

| | Patients | | Exposure (KYr) | | Outcomes | | Incidence (per KYr) | |
|---|---|---|---|---|---|---|---|---|
| | T | C | T | C | T | C | T | C |
| **PS-matching / on-treatment TAR** | | | | | | | | |
| CCAE | 10,791 | 18,710 | 7.1 | 13.6 | 10 | 22 | 1.42 (0.68 - 2.61) | 1.62 (1.02 - 2.45) |
| ODOD | 7,436 | 12,737 | 4.2 | 7.8 | 13 | 17 | 3.08 (1.64 - 5.27) | 2.18 (1.27 - 3.49) |
| MDCR | 730 | 1,472 | 0.4 | 0.9 | <5 | <5 | < 11.4 (3.72 - 26.7) | < 5.33 (1.73 - 12.4) |
| MDCD | 1,464 | 2,385 | 0.8 | 1.3 | <5 | 9 | < 6.09 (1.98 - 14.2) | 7.07 (3.23 - 13.4) |
| Open Claims | 168,780 | 272,682 | 114.1 | 202.2 | 177 | 275 | 1.55 (1.33 - 1.80) | 1.36 (1.20 - 1.53) |
| OEHR | 9,670 | 16,580 | 2.8 | 5.2 | 9 | 9 | 3.20 (1.46 - 6.07) | 1.73 (0.79 - 3.28) |
| Meta-analysis | 196,677 | 320,709 | 128.2 | 228.8 | 209 | 323 | 1.63 (1.42 - 1.87) | 1.41 (1.26 - 1.57) |
| Meta-analysis (w/o USOC) | 27,897 | 48,027 | 14.1 | 26.6 | 32 | 48 | 2.27 (1.55 - 3.21) | 1.81 (1.33 - 2.39) |
| **PS-stratification / on-treatment TAR** | | | | | | | | |
| CCAE | 14,283 | 18,712 | 8.8 | 13.6 | 11 | 22 | 1.25 (0.63 - 2.24) | 1.62 (1.02 - 2.45) |
| ODOD | 10,013 | 12,741 | 5.3 | 7.8 | 16 | 17 | 3.00 (1.71 - 4.86) | 2.18 (1.27 - 3.49) |
| MDCR | 816 | 1,473 | 0.5 | 0.9 | <5 | <5 | < 10.6 (3.43 - 24.6) | < 5.33 (1.73 - 12.4) |
| MDCD | 1,724 | 2,390 | 1.0 | 1.3 | <5 | 9 | < 5.23 (1.70 - 12.2) | 7.05 (3.22 - 13.4) |
| Open Claims | 228,642 | 272,688 | 142.6 | 202.2 | 216 | 275 | 1.51 (1.32 - 1.73) | 1.36 (1.20 - 1.53) |
| OEHR | 12,842 | 16,582 | 3.7 | 5.2 | 10 | 9 | 2.70 (1.30 - 4.97) | 1.73 (0.79 - 3.28) |
| Meta-analysis | 265,780 | 320,723 | 160.4 | 228.8 | 253 | 323 | 1.58 (1.39 - 1.78) | 1.41 (1.26 - 1.57) |
| Meta-analysis (w/o USOC) | 37,138 | 48,035 | 17.8 | 26.6 | 37 | 48 | 2.08 (1.46 - 2.86) | 1.81 (1.33 - 2.39) |
| **PS-matching / on-treatment TAR without escalation** | | | | | | | | |
| CCAE | 10,791 | 18,710 | 6.4 | 11.7 | 8 | 17 | 1.26 (0.54 - 2.48) | 1.45 (0.85 - 2.33) |
| ODOD | 7,436 | 12,737 | 3.9 | 6.9 | 12 | 13 | 3.12 (1.61 - 5.44) | 1.88 (1.00 - 3.21) |
| MDCR | 730 | 1,472 | 0.4 | 0.8 | <5 | <5 | < 12.2 (3.95 - 28.4) | < 6.02 (1.96 - 14.1) |
| MDCD | 1,464 | 2,385 | 0.7 | 1.1 | <5 | 8 | < 7.00 (2.27 - 16.3) | 7.61 (3.28 - 15.0) |
| Open Claims | 168,780 | 272,682 | 101.9 | 170.1 | 156 | 211 | 1.53 (1.30 - 1.79) | 1.24 (1.08 - 1.42) |
| OEHR | 9,670 | 16,580 | 2.4 | 4.5 | 8 | 7 | 3.29 (1.42 - 6.48) | 1.54 (0.62 - 3.18) |
| Meta-analysis | 187,007 | 304,129 | 112.1 | 188.7 | 176 | 241 | 1.57 (1.35 - 1.82) | 1.28 (1.12 - 1.45) |
| Meta-analysis (w/o USOC) | 18,227 | 31,447 | 10.2 | 18.6 | 20 | 30 | 1.96 (1.20 - 3.03) | 1.61 (1.09 - 2.30) |
| **PS-stratification / on-treatment TAR without escalation** | | | | | | | | |
| CCAE | 14,283 | 18,712 | 8.0 | 11.7 | 9 | 17 | 1.13 (0.52 - 2.14) | 1.45 (0.85 - 2.33) |
| ODOD | 10,013 | 12,741 | 4.9 | 6.9 | 14 | 13 | 2.85 (1.56 - 4.79) | 1.88 (1.00 - 3.21) |
| MDCR | 816 | 1,473 | 0.4 | 0.8 | <5 | <5 | < 11.3 (3.67 - 26.4) | < 6.01 (1.95 - 14.0) |
| MDCD | 1,724 | 2,390 | 0.8 | 1.1 | <5 | 8 | < 6.03 (1.96 - 14.1) | 7.58 (3.27 - 14.9) |
| Open Claims | 228,642 | 272,688 | 128.7 | 170.1 | 192 | 211 | 1.49 (1.29 - 1.72) | 1.24 (1.08 - 1.42) |
| OEHR | 12,842 | 16,582 | 3.2 | 4.5 | 9 | 7 | 2.79 (1.28 - 5.30) | 1.54 (0.62 - 3.18) |
| Meta-analysis | 252,938 | 304,141 | 141.5 | 188.7 | 215 | 241 | 1.52 (1.32 - 1.74) | 1.28 (1.12 - 1.45) |
| Meta-analysis (w/o USOC) | 24,296 | 31,453 | 12.9 | 18.6 | 23 | 30 | 1.79 (1.13 - 2.68) | 1.61 (1.09 - 2.30) |

##### 4.1.13 Bone fracture

**Supplementary Table 298:** Exposures, outcomes and risk estimates for Bone fracture in GLP1RA (female) (T) vs SGLT2I (female) (C) new-user cohorts under four different propensity score (PS) adjustment / time-at-risk (TAR) designs across data sources. We report total patient exposure time in 1,000 patient-year (KYr) units, uncalibrated and calibrated hazard ratios (HRs) and their 95% confidence intervals (CIs). Greyed out entries did not pass study diagnostics and are included only for completeness.

| | Patients | | Exposure (KYr) | | Outcomes | | Incidence (per KYr) | |
|---|---|---|---|---|---|---|---|---|
| | T | C | T | C | T | C | T | C |
| **PS-matching / on-treatment TAR** | | | | | | | | |
| CCAE | 9,687 | 16,973 | 6.2 | 12.0 | 117 | 230 | 18.8 (15.6 - 22.6) | 19.1 (16.7 - 21.7) |
| ODOD | 6,688 | 11,475 | 3.7 | 6.9 | 114 | 222 | 30.8 (25.4 - 37.0) | 32.1 (28.0 - 36.7) |
| MDCR | 619 | 1,279 | 0.4 | 0.8 | 9 | 35 | 23.9 (10.9 - 45.4) | 45.0 (31.3 - 62.5) |
| MDCD | 1,275 | 2,097 | 0.7 | 1.1 | 26 | 33 | 37.0 (24.1 - 54.1) | 30.1 (20.7 - 42.2) |
| Open Claims | 144,944 | 237,925 | 96.8 | 174.9 | 1,400 | 2,816 | 14.5 (13.7 - 15.2) | 16.1 (15.5 - 16.7) |
| OEHR | 8,944 | 15,398 | 2.6 | 4.8 | 34 | 69 | 13.2 (9.12 - 18.4) | 14.3 (11.2 - 18.1) |
| Meta-analysis | 171,538 | 283,868 | 110.0 | 199.8 | 1,691 | 3,370 | 15.4 (14.6 - 16.1) | 16.9 (16.3 - 17.4) |
| Meta-analysis (w/o USOC) | 26,594 | 45,943 | 13.2 | 24.9 | 291 | 554 | 22.0 (19.6 - 24.7) | 22.3 (20.5 - 24.2) |
| **PS-stratification / on-treatment TAR** | | | | | | | | |
| CCAE | 12,748 | 16,973 | 7.7 | 12.0 | 150 | 230 | 19.5 (16.5 - 22.9) | 19.1 (16.7 - 21.7) |
| ODOD | 8,968 | 11,478 | 4.7 | 6.9 | 139 | 222 | 29.7 (24.9 - 35.0) | 32.1 (28.0 - 36.6) |
| MDCR | 684 | 1,280 | 0.4 | 0.8 | 10 | 35 | 24.8 (11.9 - 45.6) | 44.9 (31.3 - 62.4) |
| MDCD | 1,490 | 2,102 | 0.8 | 1.1 | 30 | 33 | 37.0 (25.0 - 52.8) | 30.0 (20.6 - 42.1) |
| Open Claims | 194,529 | 237,944 | 120.1 | 175.0 | 1,753 | 2,817 | 14.6 (13.9 - 15.3) | 16.1 (15.5 - 16.7) |
| OEHR | 11,791 | 15,400 | 3.4 | 4.8 | 44 | 69 | 13.1 (9.51 - 17.6) | 14.3 (11.2 - 18.1) |
| Meta-analysis | 229,526 | 283,897 | 136.7 | 199.8 | 2,116 | 3,371 | 15.5 (14.8 - 16.2) | 16.9 (16.3 - 17.4) |
| Meta-analysis (w/o USOC) | 34,997 | 45,953 | 16.6 | 24.9 | 363 | 554 | 21.9 (19.7 - 24.3) | 22.3 (20.5 - 24.2) |
| **PS-matching / on-treatment TAR without escalation** | | | | | | | | |
| CCAE | 9,687 | 16,973 | 5.6 | 10.4 | 104 | 196 | 18.6 (15.2 - 22.5) | 18.9 (16.3 - 21.7) |
| ODOD | 6,688 | 11,475 | 3.4 | 6.1 | 98 | 192 | 28.9 (23.5 - 35.2) | 31.2 (27.0 - 36.0) |
| MDCR | 619 | 1,279 | 0.4 | 0.7 | 9 | 30 | 25.3 (11.6 - 48.0) | 43.5 (29.3 - 62.1) |
| MDCD | 1,275 | 2,097 | 0.6 | 0.9 | 24 | 28 | 39.3 (25.2 - 58.5) | 30.8 (20.5 - 44.6) |
| Open Claims | 144,944 | 237,925 | 86.6 | 147.3 | 1,217 | 2,345 | 14.0 (13.3 - 14.9) | 15.9 (15.3 - 16.6) |
| OEHR | 8,944 | 15,398 | 2.2 | 4.2 | 30 | 59 | 13.5 (9.09 - 19.2) | 14.1 (10.7 - 18.2) |
| Meta-analysis | 171,538 | 283,868 | 98.5 | 168.9 | 1,473 | 2,820 | 15.0 (14.2 - 15.7) | 16.7 (16.1 - 17.3) |
| Meta-analysis (w/o USOC) | 26,594 | 45,943 | 11.8 | 21.6 | 256 | 475 | 21.6 (19.1 - 24.5) | 22.0 (20.0 - 24.0) |
| **PS-stratification / on-treatment TAR without escalation** | | | | | | | | |
| CCAE | 12,748 | 16,973 | 7.0 | 10.4 | 135 | 196 | 19.3 (16.2 - 22.9) | 18.9 (16.3 - 21.7) |
| ODOD | 8,968 | 11,478 | 4.3 | 6.2 | 122 | 192 | 28.3 (23.5 - 33.8) | 31.2 (27.0 - 36.0) |
| MDCR | 684 | 1,280 | 0.4 | 0.7 | 10 | 30 | 26.4 (12.7 - 48.6) | 43.4 (29.3 - 61.9) |
| MDCD | 1,490 | 2,102 | 0.7 | 0.9 | 27 | 28 | 38.4 (25.3 - 55.9) | 30.7 (20.4 - 44.4) |
| Open Claims | 194,529 | 237,944 | 108.5 | 147.3 | 1,546 | 2,346 | 14.3 (13.6 - 15.0) | 15.9 (15.3 - 16.6) |
| OEHR | 11,791 | 15,400 | 2.9 | 4.2 | 40 | 59 | 13.7 (9.78 - 18.6) | 14.1 (10.7 - 18.2) |
| Meta-analysis | 229,526 | 283,897 | 123.4 | 168.9 | 1,870 | 2,821 | 15.2 (14.5 - 15.9) | 16.7 (16.1 - 17.3) |
| Meta-analysis (w/o USOC) | 34,997 | 45,953 | 14.9 | 21.6 | 324 | 475 | 21.7 (19.4 - 24.2) | 22.0 (20.0 - 24.0) |

###### 4.1.14 Diabetic ketoacidosis

**Supplementary Table 299:** Exposures, outcomes and risk estimates for Diabetic ketoacidosis in GLP1RA (female) (T) vs SGLT2I (female) (C) new-user cohorts under four different propensity score (PS) adjustment / time-at-risk (TAR) designs across data sources. We report total patient exposure time in 1,000 patient-year (KYr) units, uncalibrated and calibrated hazard ratios (HRs) and their 95% confidence intervals (CIs). Greyed out entries did not pass study diagnostics and are included only for completeness.

| | Patients | | Exposure (KYr) | | Outcomes | | Incidence (per KYr) | |
|---|---|---|---|---|---|---|---|---|
| | T | C | T | C | T | C | T | C |
| **PS-matching / on-treatment TAR** | | | | | | | | |
| CCAE | 10,874 | 18,865 | 7.1 | 13.7 | 5 | 31 | 0.70 (0.23 - 1.64) | 2.27 (1.54 - 3.22) |
| ODOD | 7,491 | 12,829 | 4.2 | 7.9 | <5 | 24 | < 1.18 (0.38 - 2.75) | 3.05 (1.96 - 4.54) |
| MDCR | 735 | 1,485 | 0.4 | 0.9 | 0 | <5 | | < 5.30 (1.72 - 12.4) |
| MDCD | 1,478 | 2,422 | 0.8 | 1.3 | <5 | 5 | < 6.06 (1.97 - 14.1) | 3.84 (1.25 - 8.96) |
| Open Claims | 170,551 | 276,150 | 115.2 | 204.7 | 61 | 254 | 0.53 (0.41 - 0.68) | 1.24 (1.09 - 1.40) |
| OEHR | 9,710 | 16,655 | 2.8 | 5.2 | 5 | 11 | 1.77 (0.57 - 4.12) | 2.10 (1.05 - 3.76) |
| Meta-analysis | 188,916 | 307,844 | 126.5 | 226.2 | <71 | 309 | < 0.56 (0.44 - 0.71) | 1.37 (1.22 - 1.53) |
| Meta-analysis (w/o USOC) | 18,365 | 31,694 | 11.3 | 21.5 | <10 | 55 | < 0.88 (0.42 - 1.62) | 2.55 (1.92 - 3.33) |
| **PS-stratification / on-treatment TAR** | | | | | | | | |
| CCAE | 14,354 | 18,865 | 8.8 | 13.7 | 5 | 31 | 0.57 (0.18 - 1.32) | 2.27 (1.54 - 3.22) |
| ODOD | 10,062 | 12,833 | 5.4 | 7.9 | 6 | 24 | 1.12 (0.41 - 2.43) | 3.05 (1.96 - 4.54) |
| MDCR | 819 | 1,486 | 0.5 | 0.9 | 0 | <5 | | < 5.29 (1.72 - 12.3) |
| MDCD | 1,739 | 2,427 | 1.0 | 1.3 | <5 | 5 | < 5.21 (1.69 - 12.2) | 3.83 (1.24 - 8.94) |
| Open Claims | 230,627 | 276,157 | 143.7 | 204.7 | 78 | 254 | 0.54 (0.43 - 0.68) | 1.24 (1.09 - 1.40) |
| OEHR | 12,891 | 16,657 | 3.7 | 5.2 | 8 | 11 | 2.15 (0.93 - 4.24) | 2.10 (1.05 - 3.76) |
| Meta-analysis | 267,934 | 324,512 | 161.7 | 231.4 | 97 | 320 | 0.60 (0.49 - 0.73) | 1.38 (1.24 - 1.54) |
| Meta-analysis (w/o USOC) | 37,307 | 48,355 | 17.9 | 26.8 | 19 | 66 | 1.06 (0.64 - 1.66) | 2.47 (1.91 - 3.14) |
| **PS-matching / on-treatment TAR without escalation** | | | | | | | | |
| CCAE | 10,874 | 18,865 | 6.4 | 11.8 | <5 | 24 | < 0.78 (0.25 - 1.82) | 2.04 (1.31 - 3.03) |
| ODOD | 7,491 | 12,829 | 3.9 | 7.0 | <5 | 20 | < 1.29 (0.42 - 3.01) | 2.87 (1.75 - 4.43) |
| MDCR | 735 | 1,485 | 0.4 | 0.8 | 0 | <5 | | < 5.98 (1.94 - 13.9) |
| MDCD | 1,478 | 2,422 | 0.7 | 1.1 | <5 | <5 | < 7.00 (2.27 - 16.3) | < 4.65 (1.51 - 10.9) |
| Open Claims | 170,551 | 276,150 | 102.9 | 172.1 | 52 | 200 | 0.51 (0.38 - 0.66) | 1.16 (1.01 - 1.34) |
| OEHR | 9,710 | 16,655 | 2.4 | 4.6 | <5 | 7 | < 2.05 (0.66 - 4.77) | 1.54 (0.62 - 3.17) |
| Meta-analysis | 188,916 | 307,844 | 113.2 | 190.8 | <62 | 244 | < 0.55 (0.42 - 0.70) | 1.28 (1.12 - 1.45) |
| Meta-analysis (w/o USOC) | 18,365 | 31,694 | 10.3 | 18.8 | <10 | 44 | < 0.97 (0.47 - 1.79) | 2.35 (1.70 - 3.15) |
| **PS-stratification / on-treatment TAR without escalation** | | | | | | | | |
| CCAE | 14,354 | 18,865 | 8.0 | 11.8 | <5 | 24 | < 0.62 (0.20 - 1.46) | 2.04 (1.31 - 3.03) |
| ODOD | 10,062 | 12,833 | 4.9 | 7.0 | 5 | 20 | 1.01 (0.33 - 2.37) | 2.87 (1.75 - 4.43) |
| MDCR | 819 | 1,486 | 0.4 | 0.8 | 0 | <5 | | < 5.97 (1.94 - 13.9) |
| MDCD | 1,739 | 2,427 | 0.8 | 1.1 | <5 | <5 | < 6.03 (1.96 - 14.1) | < 4.64 (1.51 - 10.8) |
| Open Claims | 230,627 | 276,157 | 129.7 | 172.1 | 67 | 200 | 0.52 (0.40 - 0.66) | 1.16 (1.01 - 1.33) |
| OEHR | 12,891 | 16,657 | 3.2 | 4.6 | <5 | 7 | < 1.54 (0.50 - 3.61) | 1.54 (0.62 - 3.17) |
| Meta-analysis | 255,043 | 307,855 | 142.6 | 190.8 | <77 | 244 | < 0.54 (0.43 - 0.67) | 1.28 (1.12 - 1.45) |
| Meta-analysis (w/o USOC) | 24,416 | 31,698 | 12.9 | 18.8 | <10 | 44 | < 0.77 (0.37 - 1.42) | 2.35 (1.70 - 3.15) |

##### 4.1.15 Diarrhea

**Supplementary Table 300:** Exposures, outcomes and risk estimates for Diarrhea in GLP1RA (female) (T) vs SGLT2I (female) (C) new-user cohorts under four different propensity score (PS) adjustment / time-at-risk (TAR) designs across data sources. We report total patient exposure time in 1,000 patient-year (KYr) units, uncalibrated and calibrated hazard ratios (HRs) and their 95% confidence intervals (CIs). Greyed out entries did not pass study diagnostics and are included only for completeness.

| | Patients | | Exposure (KYr) | | Outcomes | | Incidence (per KYr) | |
|---|---|---|---|---|---|---|---|---|
| | T | C | T | C | T | C | T | C |
| **PS-matching / on-treatment TAR** | | | | | | | | |
| CCAE | 9,039 | 15,863 | 5.7 | 11.4 | 285 | 371 | 50.2 (44.6 - 56.4) | 32.6 (29.4 - 36.1) |
| ODOD | 6,071 | 10,571 | 3.4 | 6.4 | 181 | 261 | 53.9 (46.3 - 62.4) | 41.0 (36.2 - 46.3) |
| MDCR | 608 | 1,227 | 0.4 | 0.8 | 10 | 29 | 28.2 (13.5 - 51.9) | 37.1 (24.9 - 53.3) |
| MDCD | 1,098 | 1,789 | 0.6 | 0.9 | 49 | 57 | 81.0 (59.9 - 107) | 60.6 (45.9 - 78.5) |
| Open Claims | 139,745 | 230,065 | 93.2 | 169.4 | 2,342 | 3,165 | 25.1 (24.1 - 26.2) | 18.7 (18.0 - 19.3) |
| OEHR | 8,173 | 14,216 | 2.3 | 4.4 | 140 | 177 | 60.6 (51.0 - 71.5) | 40.7 (34.9 - 47.1) |
| Meta-analysis | 164,126 | 272,504 | 105.2 | 192.4 | 2,997 | 4,031 | 28.5 (27.5 - 29.5) | 20.9 (20.3 - 21.6) |
| Meta-analysis (w/o USOC) | 24,381 | 42,439 | 11.9 | 23.0 | 655 | 866 | 54.8 (50.7 - 59.2) | 37.6 (35.1 - 40.2) |
| **PS-stratification / on-treatment TAR** | | | | | | | | |
| CCAE | 11,815 | 15,864 | 7.0 | 11.4 | 360 | 371 | 51.5 (46.4 - 57.2) | 32.6 (29.4 - 36.1) |
| ODOD | 8,096 | 10,574 | 4.2 | 6.4 | 244 | 262 | 58.2 (51.1 - 66.0) | 41.1 (36.3 - 46.4) |
| MDCR | 680 | 1,228 | 0.4 | 0.8 | 12 | 29 | 31.2 (16.1 - 54.5) | 37.1 (24.8 - 53.2) |
| MDCD | 1,299 | 1,793 | 0.7 | 0.9 | 64 | 57 | 91.1 (70.2 - 116) | 60.4 (45.7 - 78.2) |
| Open Claims | 186,585 | 230,071 | 115.0 | 169.4 | 3,055 | 3,165 | 26.6 (25.6 - 27.5) | 18.7 (18.0 - 19.3) |
| OEHR | 10,772 | 14,218 | 3.0 | 4.4 | 190 | 177 | 63.5 (54.7 - 73.1) | 40.7 (34.9 - 47.1) |
| Meta-analysis | 218,567 | 272,520 | 129.9 | 192.5 | 3,913 | 4,032 | 30.1 (29.2 - 31.1) | 20.9 (20.3 - 21.6) |
| Meta-analysis (w/o USOC) | 31,982 | 42,449 | 14.9 | 23.0 | 858 | 867 | 57.7 (53.9 - 61.7) | 37.6 (35.2 - 40.2) |
| **PS-matching / on-treatment TAR without escalation** | | | | | | | | |
| CCAE | 9,039 | 15,863 | 5.2 | 9.9 | 255 | 322 | 49.5 (43.6 - 55.9) | 32.5 (29.1 - 36.3) |
| ODOD | 6,071 | 10,571 | 3.1 | 5.7 | 171 | 233 | 55.7 (47.6 - 64.7) | 41.0 (35.9 - 46.6) |
| MDCR | 608 | 1,227 | 0.3 | 0.7 | 9 | 26 | 27.1 (12.4 - 51.5) | 37.7 (24.6 - 55.2) |
| MDCD | 1,098 | 1,789 | 0.5 | 0.8 | 43 | 47 | 79.8 (57.8 - 108) | 60.0 (44.1 - 79.8) |
| Open Claims | 139,745 | 230,065 | 83.6 | 143.4 | 2,096 | 2,586 | 25.1 (24.0 - 26.2) | 18.0 (17.3 - 18.7) |
| OEHR | 8,173 | 14,216 | 2.0 | 3.8 | 126 | 147 | 62.7 (52.3 - 74.7) | 38.5 (32.5 - 45.2) |
| Meta-analysis | 164,126 | 272,504 | 94.4 | 163.6 | 2,691 | 3,335 | 28.5 (27.5 - 29.6) | 20.4 (19.7 - 21.1) |
| Meta-analysis (w/o USOC) | 24,381 | 42,439 | 10.8 | 20.2 | 595 | 749 | 55.2 (50.9 - 59.8) | 37.1 (34.5 - 39.9) |
| **PS-stratification / on-treatment TAR without escalation** | | | | | | | | |
| CCAE | 11,815 | 15,864 | 6.4 | 9.9 | 325 | 322 | 50.9 (45.5 - 56.8) | 32.5 (29.1 - 36.3) |
| ODOD | 8,096 | 10,574 | 3.9 | 5.7 | 232 | 234 | 60.1 (52.6 - 68.3) | 41.2 (36.1 - 46.8) |
| MDCR | 680 | 1,228 | 0.4 | 0.7 | 10 | 26 | 27.9 (13.4 - 51.3) | 37.6 (24.6 - 55.1) |
| MDCD | 1,299 | 1,793 | 0.6 | 0.8 | 56 | 47 | 89.5 (67.6 - 116) | 59.8 (43.9 - 79.5) |
| Open Claims | 186,585 | 230,071 | 104.0 | 143.4 | 2,764 | 2,586 | 26.6 (25.6 - 27.6) | 18.0 (17.3 - 18.7) |
| OEHR | 10,772 | 14,218 | 2.6 | 3.8 | 171 | 147 | 65.0 (55.6 - 75.5) | 38.4 (32.5 - 45.2) |
| Meta-analysis | 218,567 | 272,520 | 117.5 | 163.6 | 3,548 | 3,336 | 30.2 (29.2 - 31.2) | 20.4 (19.7 - 21.1) |
| Meta-analysis (w/o USOC) | 31,982 | 42,449 | 13.5 | 20.2 | 784 | 750 | 58.1 (54.1 - 62.3) | 37.1 (34.5 - 39.9) |

##### 4.1.16 Genitourinary infection

**Supplementary Table 301:** Exposures, outcomes and risk estimates for Genitourinary infection in GLP1RA (female) (T) vs SGLT2I (female) (C) new-user cohorts under four different propensity score (PS) adjustment / time-at-risk (TAR) designs across data sources. We report total patient exposure time in 1,000 patient-year (KYr) units, uncalibrated and calibrated hazard ratios (HRs) and their 95% confidence intervals (CIs). Greyed out entries did not pass study diagnostics and are included only for completeness.

| | Patients | | Exposure (KYr) | | Outcomes | | Incidence (per KYr) | |
|---|---|---|---|---|---|---|---|---|
| | T | C | T | C | T | C | T | C |
| **PS-matching / on-treatment TAR** | | | | | | | | |
| CCAE | 7,431 | 12,966 | 4.6 | 9.0 | 340 | 553 | 74.1 (66.4 - 82.4) | 61.4 (56.4 - 66.7) |
| ODOD | 5,164 | 8,864 | 2.8 | 5.2 | 246 | 402 | 89.4 (78.6 - 101) | 77.0 (69.7 - 84.9) |
| MDCR | 503 | 1,114 | 0.3 | 0.7 | 24 | 54 | 84.9 (54.4 - 126) | 77.9 (58.5 - 102) |
| MDCD | 920 | 1,600 | 0.5 | 0.8 | 52 | 79 | 102 (76.4 - 134) | 95.5 (75.6 - 119) |
| Open Claims | 109,958 | 183,236 | 73.2 | 136.5 | 2,971 | 5,340 | 40.6 (39.2 - 42.1) | 39.1 (38.1 - 40.2) |
| OEHR | 7,379 | 12,670 | 2.1 | 3.8 | 159 | 254 | 77.5 (66.0 - 90.6) | 66.5 (58.6 - 75.2) |
| Meta-analysis | 130,852 | 219,336 | 83.1 | 155.4 | 3,768 | 6,628 | 45.4 (43.9 - 46.8) | 42.7 (41.6 - 43.7) |
| Meta-analysis (w/o USOC) | 20,894 | 36,100 | 9.9 | 18.9 | 797 | 1,288 | 80.5 (75.0 - 86.3) | 68.2 (64.5 - 72.0) |
| **PS-stratification / on-treatment TAR** | | | | | | | | |
| CCAE | 9,602 | 12,966 | 5.6 | 9.0 | 411 | 553 | 73.9 (66.9 - 81.4) | 61.4 (56.4 - 66.7) |
| ODOD | 6,814 | 8,866 | 3.5 | 5.2 | 298 | 402 | 86.3 (76.8 - 96.7) | 76.9 (69.6 - 84.8) |
| MDCR | 545 | 1,115 | 0.3 | 0.7 | 25 | 54 | 83.9 (54.3 - 124) | 77.8 (58.4 - 102) |
| MDCD | 1,123 | 1,602 | 0.6 | 0.8 | 64 | 79 | 105 (80.7 - 134) | 95.5 (75.6 - 119) |
| Open Claims | 144,211 | 183,262 | 89.1 | 136.5 | 3,714 | 5,341 | 41.7 (40.4 - 43.1) | 39.1 (38.1 - 40.2) |
| OEHR | 9,671 | 12,672 | 2.6 | 3.8 | 218 | 254 | 82.4 (71.8 - 94.1) | 66.5 (58.5 - 75.2) |
| Meta-analysis | 171,421 | 219,368 | 101.3 | 155.4 | 4,705 | 6,629 | 46.4 (45.1 - 47.8) | 42.7 (41.6 - 43.7) |
| Meta-analysis (w/o USOC) | 27,210 | 36,106 | 12.3 | 18.9 | 991 | 1,288 | 80.8 (75.8 - 86.0) | 68.2 (64.5 - 72.0) |
| **PS-matching / on-treatment TAR without escalation** | | | | | | | | |
| CCAE | 7,431 | 12,966 | 4.2 | 7.8 | 309 | 485 | 73.9 (65.9 - 82.6) | 61.8 (56.4 - 67.5) |
| ODOD | 5,164 | 8,864 | 2.5 | 4.7 | 223 | 355 | 88.0 (76.8 - 100) | 76.0 (68.3 - 84.3) |
| MDCR | 503 | 1,114 | 0.3 | 0.6 | 19 | 49 | 69.6 (41.9 - 109) | 79.6 (58.9 - 105) |
| MDCD | 920 | 1,600 | 0.4 | 0.7 | 40 | 59 | 90.5 (64.7 - 123) | 84.7 (64.5 - 109) |
| Open Claims | 109,958 | 183,236 | 65.5 | 115.3 | 2,616 | 4,528 | 40.0 (38.4 - 41.5) | 39.3 (38.1 - 40.4) |
| OEHR | 7,379 | 12,670 | 1.8 | 3.3 | 135 | 218 | 75.1 (63.0 - 88.9) | 65.4 (57.0 - 74.7) |
| Meta-analysis | 130,852 | 219,336 | 74.4 | 131.8 | 3,323 | 5,645 | 44.6 (43.1 - 46.2) | 42.8 (41.7 - 43.9) |
| Meta-analysis (w/o USOC) | 20,894 | 36,100 | 9.0 | 16.6 | 707 | 1,117 | 79.0 (73.2 - 85.0) | 67.5 (63.6 - 71.6) |
| **PS-stratification / on-treatment TAR without escalation** | | | | | | | | |
| CCAE | 9,602 | 12,966 | 5.1 | 7.8 | 378 | 485 | 74.1 (66.9 - 82.0) | 61.8 (56.4 - 67.5) |
| ODOD | 6,814 | 8,866 | 3.2 | 4.7 | 272 | 355 | 85.1 (75.3 - 95.9) | 75.9 (68.2 - 84.2) |
| MDCR | 545 | 1,115 | 0.3 | 0.6 | 19 | 49 | 66.4 (40.0 - 104) | 79.4 (58.8 - 105) |
| MDCD | 1,123 | 1,602 | 0.5 | 0.7 | 50 | 59 | 93.1 (69.1 - 123) | 84.7 (64.5 - 109) |
| Open Claims | 144,211 | 183,262 | 80.4 | 115.3 | 3,305 | 4,529 | 41.1 (39.7 - 42.5) | 39.3 (38.1 - 40.4) |
| OEHR | 9,671 | 12,672 | 2.3 | 3.3 | 186 | 218 | 79.5 (68.5 - 91.8) | 65.4 (57.0 - 74.6) |
| Meta-analysis | 171,421 | 219,368 | 91.6 | 131.9 | 4,191 | 5,646 | 45.8 (44.4 - 47.2) | 42.8 (41.7 - 44.0) |
| Meta-analysis (w/o USOC) | 27,210 | 36,106 | 11.2 | 16.6 | 886 | 1,117 | 79.3 (74.2 - 84.7) | 67.5 (63.6 - 71.5) |

##### 4.1.17 Hyperkalemia

**Supplementary Table 302:** Exposures, outcomes and risk estimates for Hyperkalemia in GLP1RA (female) (T) vs SGLT2I (female) (C) new-user cohorts under four different propensity score (PS) adjustment / time-at-risk (TAR) designs across data sources. We report total patient exposure time in 1,000 patient-year (KYr) units, uncalibrated and calibrated hazard ratios (HRs) and their 95% confidence intervals (CIs). Greyed out entries did not pass study diagnostics and are included only for completeness.

| | Patients | | Exposure (KYr) | | Outcomes | | Incidence (per KYr) | |
|---|---|---|---|---|---|---|---|---|
| | T | C | T | C | T | C | T | C |
| **PS-matching / on-treatment TAR** | | | | | | | | |
| CCAE | 10,686 | 18,560 | 6.9 | 13.4 | 32 | 76 | 4.61 (3.15 - 6.50) | 5.69 (4.48 - 7.12) |
| ODOD | 7,258 | 12,377 | 4.1 | 7.6 | 56 | 109 | 13.7 (10.4 - 17.8) | 14.4 (11.8 - 17.4) |
| MDCR | 717 | 1,450 | 0.4 | 0.9 | <5 | 5 | < 11.8 (3.84 - 27.6) | 5.38 (1.75 - 12.6) |
| MDCD | 1,450 | 2,358 | 0.8 | 1.3 | 5 | 7 | 6.19 (2.01 - 14.4) | 5.53 (2.22 - 11.4) |
| Open Claims | 167,890 | 271,508 | 113.2 | 201.2 | 361 | 716 | 3.19 (2.87 - 3.54) | 3.56 (3.30 - 3.83) |
| OEHR | 9,580 | 16,393 | 2.8 | 5.1 | 24 | 32 | 8.60 (5.51 - 12.8) | 6.23 (4.26 - 8.80) |
| Meta-analysis | 195,414 | 318,838 | 127.0 | 227.2 | 473 | 933 | 3.72 (3.40 - 4.08) | 4.11 (3.85 - 4.38) |
| Meta-analysis (w/o USOC) | 27,524 | 47,330 | 13.8 | 26.1 | 112 | 217 | 8.11 (6.67 - 9.75) | 8.33 (7.26 - 9.51) |
| **PS-stratification / on-treatment TAR** | | | | | | | | |
| CCAE | 14,113 | 18,560 | 8.6 | 13.4 | 36 | 76 | 4.17 (2.92 - 5.77) | 5.69 (4.48 - 7.12) |
| ODOD | 9,779 | 12,381 | 5.2 | 7.6 | 65 | 109 | 12.5 (9.66 - 16.0) | 14.4 (11.8 - 17.4) |
| MDCR | 798 | 1,451 | 0.5 | 0.9 | 5 | 5 | 11.0 (3.57 - 25.7) | 5.37 (1.75 - 12.5) |
| MDCD | 1,710 | 2,363 | 0.9 | 1.3 | 6 | 7 | 6.38 (2.34 - 13.9) | 5.52 (2.22 - 11.4) |
| Open Claims | 227,232 | 271,518 | 141.3 | 201.2 | 425 | 717 | 3.01 (2.73 - 3.31) | 3.56 (3.31 - 3.83) |
| OEHR | 12,701 | 16,395 | 3.7 | 5.1 | 29 | 32 | 7.93 (5.31 - 11.4) | 6.23 (4.26 - 8.80) |
| Meta-analysis | 263,825 | 318,854 | 158.8 | 227.2 | 555 | 934 | 3.49 (3.21 - 3.80) | 4.11 (3.85 - 4.38) |
| Meta-analysis (w/o USOC) | 36,593 | 47,336 | 17.5 | 26.1 | 130 | 217 | 7.43 (6.21 - 8.83) | 8.32 (7.25 - 9.51) |
| **PS-matching / on-treatment TAR without escalation** | | | | | | | | |
| CCAE | 10,686 | 18,560 | 6.3 | 11.5 | 30 | 67 | 4.79 (3.23 - 6.84) | 5.81 (4.50 - 7.38) |
| ODOD | 7,258 | 12,377 | 3.7 | 6.7 | 55 | 97 | 14.7 (11.1 - 19.2) | 14.5 (11.7 - 17.6) |
| MDCR | 717 | 1,450 | 0.4 | 0.8 | <5 | 5 | < 12.6 (4.09 - 29.4) | 6.08 (1.97 - 14.2) |
| MDCD | 1,450 | 2,358 | 0.7 | 1.0 | 5 | 7 | 7.14 (2.32 - 16.7) | 6.69 (2.69 - 13.8) |
| Open Claims | 167,890 | 271,508 | 101.1 | 169.2 | 316 | 579 | 3.12 (2.79 - 3.49) | 3.42 (3.15 - 3.71) |
| OEHR | 9,580 | 16,393 | 2.4 | 4.5 | 22 | 26 | 9.13 (5.72 - 13.8) | 5.83 (3.81 - 8.54) |
| Meta-analysis | 195,414 | 318,838 | 113.5 | 191.9 | 423 | 769 | 3.73 (3.38 - 4.10) | 4.01 (3.73 - 4.30) |
| Meta-analysis (w/o USOC) | 27,524 | 47,330 | 12.4 | 22.7 | 107 | 190 | 8.63 (7.07 - 10.4) | 8.37 (7.22 - 9.65) |
| **PS-stratification / on-treatment TAR without escalation** | | | | | | | | |
| CCAE | 14,113 | 18,560 | 7.8 | 11.5 | 34 | 67 | 4.34 (3.00 - 6.06) | 5.81 (4.50 - 7.38) |
| ODOD | 9,779 | 12,381 | 4.8 | 6.7 | 63 | 97 | 13.2 (10.2 - 16.9) | 14.5 (11.7 - 17.6) |
| MDCR | 798 | 1,451 | 0.4 | 0.8 | 5 | 5 | 11.8 (3.83 - 27.5) | 6.07 (1.97 - 14.2) |
| MDCD | 1,710 | 2,363 | 0.8 | 1.0 | 6 | 7 | 7.38 (2.71 - 16.1) | 6.67 (2.68 - 13.7) |
| Open Claims | 227,232 | 271,518 | 127.6 | 169.2 | 372 | 580 | 2.92 (2.63 - 3.23) | 3.43 (3.15 - 3.72) |
| OEHR | 12,701 | 16,395 | 3.2 | 4.5 | 27 | 26 | 8.48 (5.59 - 12.3) | 5.82 (3.81 - 8.53) |
| Meta-analysis | 263,825 | 318,854 | 143.3 | 191.9 | 496 | 770 | 3.46 (3.16 - 3.78) | 4.01 (3.73 - 4.31) |
| Meta-analysis (w/o USOC) | 36,593 | 47,336 | 15.8 | 22.7 | 124 | 190 | 7.85 (6.53 - 9.36) | 8.37 (7.22 - 9.64) |

##### 4.1.18 Hypoglycemia

**Supplementary Table 303:** Exposures, outcomes and risk estimates for Hypoglycemia in GLP1RA (female) (T) vs SGLT2I (female) (C) new-user cohorts under four different propensity score (PS) adjustment / time-at-risk (TAR) designs across data sources. We report total patient exposure time in 1,000 patient-year (KYr) units, uncalibrated and calibrated hazard ratios (HRs) and their 95% confidence intervals (CIs). Greyed out entries did not pass study diagnostics and are included only for completeness.

| | Patients | | Exposure (KYr) | | Outcomes | | Incidence (per KYr) | |
|---|---|---|---|---|---|---|---|---|
| | T | C | T | C | T | C | T | C |
| **PS-matching / on-treatment TAR** | | | | | | | | |
| CCAE | 10,584 | 18,425 | 6.9 | 13.3 | 26 | 50 | 3.76 (2.46 - 5.51) | 3.76 (2.79 - 4.96) |
| ODOD | 7,341 | 12,576 | 4.1 | 7.7 | 32 | 50 | 7.71 (5.28 - 10.9) | 6.51 (4.83 - 8.58) |
| MDCR | 721 | 1,472 | 0.4 | 0.9 | <5 | 0 | < 11.6 (3.76 - 27.0) | |
| MDCD | 1,427 | 2,344 | 0.8 | 1.3 | <5 | 12 | < 6.22 (2.02 - 14.5) | 9.56 (4.94 - 16.7) |
| Open Claims | 167,043 | 270,929 | 112.8 | 200.8 | 311 | 416 | 2.76 (2.46 - 3.08) | 2.07 (1.88 - 2.28) |
| OEHR | 9,568 | 16,442 | 2.8 | 5.2 | 17 | 18 | 6.08 (3.54 - 9.74) | 3.49 (2.07 - 5.52) |
| Meta-analysis | 194,536 | 318,372 | 126.6 | 226.9 | 386 | 534 | 3.05 (2.75 - 3.37) | 2.35 (2.16 - 2.56) |
| Meta-analysis (w/o USOC) | 27,493 | 47,443 | 13.9 | 26.1 | 75 | 118 | 5.41 (4.26 - 6.78) | 4.51 (3.74 - 5.41) |
| **PS-stratification / on-treatment TAR** | | | | | | | | |
| CCAE | 13,925 | 18,425 | 8.6 | 13.3 | 43 | 50 | 5.03 (3.64 - 6.77) | 3.76 (2.79 - 4.96) |
| ODOD | 9,812 | 12,580 | 5.2 | 7.7 | 45 | 50 | 8.60 (6.28 - 11.5) | 6.50 (4.83 - 8.57) |
| MDCR | 804 | 1,473 | 0.5 | 0.9 | <5 | 0 | < 10.7 (3.47 - 24.9) | |
| MDCD | 1,676 | 2,349 | 0.9 | 1.3 | 8 | 12 | 8.59 (3.71 - 16.9) | 9.54 (4.93 - 16.7) |
| Open Claims | 224,703 | 270,936 | 140.1 | 200.8 | 400 | 416 | 2.85 (2.58 - 3.15) | 2.07 (1.88 - 2.28) |
| OEHR | 12,655 | 16,444 | 3.6 | 5.2 | 30 | 18 | 8.24 (5.56 - 11.8) | 3.49 (2.07 - 5.52) |
| Meta-analysis | 262,771 | 320,734 | 158.5 | 228.2 | 526 | 546 | 3.32 (3.04 - 3.62) | 2.39 (2.20 - 2.60) |
| Meta-analysis (w/o USOC) | 38,068 | 49,798 | 18.4 | 27.4 | 126 | 130 | 6.86 (5.72 - 8.17) | 4.74 (3.96 - 5.63) |
| **PS-matching / on-treatment TAR without escalation** | | | | | | | | |
| CCAE | 10,584 | 18,425 | 6.2 | 11.5 | 21 | 37 | 3.37 (2.09 - 5.15) | 3.22 (2.27 - 4.44) |
| ODOD | 7,341 | 12,576 | 3.8 | 6.8 | 26 | 37 | 6.86 (4.48 - 10.1) | 5.42 (3.82 - 7.48) |
| MDCR | 721 | 1,472 | 0.4 | 0.8 | <5 | 0 | < 12.3 (4.00 - 28.8) | |
| MDCD | 1,427 | 2,344 | 0.7 | 1.0 | <5 | 9 | < 7.14 (2.32 - 16.7) | 8.71 (3.98 - 16.5) |
| Open Claims | 167,043 | 270,929 | 100.8 | 168.9 | 260 | 277 | 2.58 (2.28 - 2.91) | 1.64 (1.45 - 1.85) |
| OEHR | 9,568 | 16,442 | 2.4 | 4.5 | 12 | 9 | 4.97 (2.57 - 8.68) | 2.01 (0.92 - 3.81) |
| Meta-analysis | 194,536 | 318,372 | 113.2 | 191.6 | 319 | 360 | 2.82 (2.52 - 3.14) | 1.88 (1.69 - 2.08) |
| Meta-analysis (w/o USOC) | 27,493 | 47,443 | 12.4 | 22.8 | 59 | 83 | 4.75 (3.61 - 6.12) | 3.64 (2.90 - 4.52) |
| **PS-stratification / on-treatment TAR without escalation** | | | | | | | | |
| CCAE | 13,925 | 18,425 | 7.8 | 11.5 | 37 | 37 | 4.76 (3.35 - 6.57) | 3.22 (2.27 - 4.44) |
| ODOD | 9,812 | 12,580 | 4.8 | 6.8 | 39 | 37 | 8.12 (5.77 - 11.1) | 5.42 (3.82 - 7.47) |
| MDCR | 804 | 1,473 | 0.4 | 0.8 | <5 | 0 | < 11.4 (3.72 - 26.7) | |
| MDCD | 1,676 | 2,349 | 0.8 | 1.0 | 6 | 9 | 7.43 (2.73 - 16.2) | 8.68 (3.97 - 16.5) |
| Open Claims | 224,703 | 270,936 | 126.5 | 168.9 | 337 | 277 | 2.66 (2.39 - 2.97) | 1.64 (1.45 - 1.85) |
| OEHR | 12,655 | 16,444 | 3.2 | 4.5 | 23 | 9 | 7.25 (4.59 - 10.9) | 2.01 (0.92 - 3.81) |
| Meta-analysis | 261,095 | 318,385 | 142.2 | 191.7 | 436 | 360 | 3.07 (2.78 - 3.37) | 1.88 (1.69 - 2.08) |
| Meta-analysis (w/o USOC) | 36,392 | 47,449 | 15.7 | 22.8 | 99 | 83 | 6.29 (5.11 - 7.66) | 3.64 (2.90 - 4.51) |

##### 4.1.19 Hypotension

**Supplementary Table 304:** Exposures, outcomes and risk estimates for Hypotension in GLP1RA (female) (T) vs SGLT2I (female) (C) new-user cohorts under four different propensity score (PS) adjustment / time-at-risk (TAR) designs across data sources. We report total patient exposure time in 1,000 patient-year (KYr) units, uncalibrated and calibrated hazard ratios (HRs) and their 95% confidence intervals (CIs). Greyed out entries did not pass study diagnostics and are included only for completeness.

| | Patients | | Exposure (KYr) | | Outcomes | | Incidence (per KYr) | |
|---|---|---|---|---|---|---|---|---|
| | T | C | T | C | T | C | T | C |
| **PS-matching / on-treatment TAR** | | | | | | | | |
| CCAE | 10,655 | 18,471 | 6.9 | 13.3 | 60 | 110 | 8.68 (6.63 - 11.2) | 8.25 (6.78 - 9.94) |
| ODOD | 7,194 | 12,288 | 4.0 | 7.5 | 70 | 148 | 17.3 (13.5 - 21.9) | 19.8 (16.7 - 23.2) |
| MDCR | 692 | 1,424 | 0.4 | 0.9 | 7 | 25 | 16.9 (6.79 - 34.8) | 27.6 (17.9 - 40.8) |
| MDCD | 1,396 | 2,259 | 0.8 | 1.2 | 22 | 42 | 28.8 (18.1 - 43.7) | 35.5 (25.6 - 48.0) |
| Open Claims | 165,007 | 267,166 | 111.1 | 198.1 | 734 | 1,306 | 6.61 (6.14 - 7.10) | 6.59 (6.24 - 6.96) |
| OEHR | 9,495 | 16,278 | 2.7 | 5.1 | 44 | 60 | 16.0 (11.6 - 21.5) | 11.8 (8.97 - 15.1) |
| Meta-analysis | 193,747 | 316,462 | 125.5 | 225.2 | 930 | 1,666 | 7.41 (6.94 - 7.90) | 7.40 (7.05 - 7.76) |
| Meta-analysis (w/o USOC) | 28,740 | 49,296 | 14.5 | 27.1 | 196 | 360 | 13.5 (11.7 - 15.6) | 13.3 (11.9 - 14.7) |
| **PS-stratification / on-treatment TAR** | | | | | | | | |
| CCAE | 14,028 | 18,471 | 8.6 | 13.3 | 81 | 110 | 9.45 (7.51 - 11.8) | 8.25 (6.78 - 9.94) |
| ODOD | 9,654 | 12,292 | 5.1 | 7.5 | 90 | 148 | 17.6 (14.1 - 21.6) | 19.8 (16.7 - 23.2) |
| MDCR | 776 | 1,425 | 0.5 | 0.9 | 7 | 25 | 15.5 (6.25 - 32.0) | 27.6 (17.9 - 40.8) |
| MDCD | 1,644 | 2,263 | 0.9 | 1.2 | 25 | 42 | 28.1 (18.2 - 41.5) | 35.4 (25.5 - 47.9) |
| Open Claims | 222,878 | 267,171 | 138.4 | 198.1 | 937 | 1,308 | 6.77 (6.34 - 7.22) | 6.60 (6.25 - 6.97) |
| OEHR | 12,645 | 16,280 | 3.6 | 5.1 | 55 | 60 | 15.2 (11.5 - 19.8) | 11.8 (8.97 - 15.1) |
| Meta-analysis | 260,849 | 316,477 | 156.6 | 225.2 | 1,188 | 1,668 | 7.59 (7.16 - 8.03) | 7.41 (7.06 - 7.77) |
| Meta-analysis (w/o USOC) | 37,971 | 49,306 | 18.2 | 27.1 | 251 | 360 | 13.8 (12.1 - 15.6) | 13.3 (11.9 - 14.7) |
| **PS-matching / on-treatment TAR without escalation** | | | | | | | | |
| CCAE | 10,655 | 18,471 | 6.2 | 11.5 | 55 | 84 | 8.83 (6.65 - 11.5) | 7.30 (5.82 - 9.04) |
| ODOD | 7,194 | 12,288 | 3.7 | 6.6 | 64 | 126 | 17.3 (13.3 - 22.1) | 19.0 (15.8 - 22.6) |
| MDCR | 692 | 1,424 | 0.4 | 0.8 | 7 | 24 | 17.9 (7.19 - 36.9) | 29.9 (19.1 - 44.5) |
| MDCD | 1,396 | 2,259 | 0.7 | 1.0 | 19 | 32 | 28.7 (17.3 - 44.8) | 32.6 (22.3 - 46.0) |
| Open Claims | 165,007 | 267,166 | 99.3 | 166.7 | 621 | 1,065 | 6.25 (5.77 - 6.76) | 6.39 (6.01 - 6.78) |
| OEHR | 9,495 | 16,278 | 2.4 | 4.4 | 34 | 42 | 14.3 (9.91 - 20.0) | 9.46 (6.81 - 12.8) |
| Meta-analysis | 193,747 | 316,462 | 112.3 | 190.3 | 793 | 1,349 | 7.06 (6.58 - 7.57) | 7.09 (6.72 - 7.48) |
| Meta-analysis (w/o USOC) | 28,740 | 49,296 | 13.0 | 23.6 | 172 | 284 | 13.3 (11.3 - 15.4) | 12.0 (10.7 - 13.5) |
| **PS-stratification / on-treatment TAR without escalation** | | | | | | | | |
| CCAE | 14,028 | 18,471 | 7.8 | 11.5 | 73 | 84 | 9.39 (7.36 - 11.8) | 7.30 (5.82 - 9.04) |
| ODOD | 9,654 | 12,292 | 4.7 | 6.6 | 82 | 126 | 17.4 (13.9 - 21.6) | 19.0 (15.8 - 22.6) |
| MDCR | 776 | 1,425 | 0.4 | 0.8 | 7 | 24 | 16.6 (6.67 - 34.2) | 29.8 (19.1 - 44.4) |
| MDCD | 1,644 | 2,263 | 0.8 | 1.0 | 20 | 32 | 25.9 (15.8 - 40.0) | 32.5 (22.2 - 45.8) |
| Open Claims | 222,878 | 267,171 | 125.0 | 166.7 | 807 | 1,066 | 6.46 (6.02 - 6.92) | 6.40 (6.02 - 6.79) |
| OEHR | 12,645 | 16,280 | 3.2 | 4.4 | 43 | 42 | 13.6 (9.86 - 18.4) | 9.45 (6.81 - 12.8) |
| Meta-analysis | 260,849 | 316,477 | 141.4 | 190.3 | 1,025 | 1,350 | 7.25 (6.81 - 7.71) | 7.10 (6.72 - 7.48) |
| Meta-analysis (w/o USOC) | 37,971 | 49,306 | 16.4 | 23.6 | 218 | 284 | 13.3 (11.6 - 15.2) | 12.0 (10.7 - 13.5) |

##### 4.1.20 Joint pain

**Supplementary Table 305:** Exposures, outcomes and risk estimates for Joint pain in GLP1RA (female) (T) vs SGLT2I (female) (C) new-user cohorts under four different propensity score (PS) adjustment / time-at-risk (TAR) designs across data sources. We report total patient exposure time in 1,000 patient-year (KYr) units, uncalibrated and calibrated hazard ratios (HRs) and their 95% confidence intervals (CIs). Greyed out entries did not pass study diagnostics and are included only for completeness.

| | Patients | | Exposure (KYr) | | Outcomes | | Incidence (per KYr) | |
|---|---|---|---|---|---|---|---|---|
| | T | C | T | C | T | C | T | C |
| **PS-matching / on-treatment TAR** | | | | | | | | |
| CCAE | 9,829 | 17,271 | 6.4 | 12.5 | 115 | 175 | 18.1 (14.9 - 21.7) | 14.0 (12.0 - 16.2) |
| ODOD | 6,780 | 11,746 | 3.8 | 7.1 | 78 | 129 | 20.5 (16.2 - 25.6) | 18.0 (15.1 - 21.4) |
| MDCR | 677 | 1,411 | 0.4 | 0.9 | 5 | 14 | 12.0 (3.90 - 28.1) | 15.7 (8.58 - 26.3) |
| MDCD | 1,299 | 2,153 | 0.7 | 1.2 | 22 | 31 | 30.6 (19.2 - 46.3) | 26.8 (18.2 - 38.1) |
| Open Claims | 155,108 | 253,902 | 104.5 | 188.4 | 828 | 1,464 | 7.92 (7.39 - 8.48) | 7.77 (7.38 - 8.18) |
| OEHR | 8,894 | 15,385 | 2.6 | 4.8 | 47 | 80 | 18.4 (13.5 - 24.5) | 16.8 (13.3 - 20.9) |
| Meta-analysis | 181,910 | 300,457 | 118.0 | 214.0 | 1,090 | 1,879 | 9.24 (8.70 - 9.81) | 8.78 (8.39 - 9.19) |
| Meta-analysis (w/o USOC) | 26,802 | 46,555 | 13.4 | 25.6 | 262 | 415 | 19.5 (17.2 - 22.0) | 16.2 (14.7 - 17.9) |
| **PS-stratification / on-treatment TAR** | | | | | | | | |
| CCAE | 12,834 | 17,271 | 7.8 | 12.5 | 140 | 175 | 17.9 (15.1 - 21.2) | 14.0 (12.0 - 16.2) |
| ODOD | 8,959 | 11,750 | 4.7 | 7.2 | 97 | 129 | 20.4 (16.6 - 24.9) | 18.0 (15.1 - 21.4) |
| MDCR | 743 | 1,412 | 0.4 | 0.9 | 5 | 14 | 11.4 (3.69 - 26.5) | 15.7 (8.57 - 26.3) |
| MDCD | 1,513 | 2,157 | 0.8 | 1.2 | 26 | 31 | 31.7 (20.7 - 46.5) | 26.8 (18.2 - 38.0) |
| Open Claims | 207,412 | 253,928 | 129.2 | 188.5 | 1,119 | 1,465 | 8.66 (8.16 - 9.19) | 7.77 (7.38 - 8.18) |
| OEHR | 11,723 | 15,387 | 3.3 | 4.8 | 61 | 80 | 18.4 (14.1 - 23.6) | 16.8 (13.3 - 20.9) |
| Meta-analysis | 242,441 | 300,493 | 145.9 | 214.0 | 1,443 | 1,880 | 9.89 (9.39 - 10.4) | 8.78 (8.39 - 9.19) |
| Meta-analysis (w/o USOC) | 35,029 | 46,565 | 16.7 | 25.6 | 324 | 415 | 19.4 (17.4 - 21.6) | 16.2 (14.7 - 17.9) |
| **PS-matching / on-treatment TAR without escalation** | | | | | | | | |
| CCAE | 9,829 | 17,271 | 5.7 | 10.8 | 101 | 150 | 17.6 (14.3 - 21.4) | 13.9 (11.8 - 16.3) |
| ODOD | 6,780 | 11,746 | 3.5 | 6.3 | 66 | 113 | 19.0 (14.7 - 24.1) | 17.8 (14.7 - 21.4) |
| MDCR | 677 | 1,411 | 0.4 | 0.8 | 5 | 13 | 12.8 (4.16 - 29.9) | 16.3 (8.68 - 27.9) |
| MDCD | 1,299 | 2,153 | 0.6 | 1.0 | 21 | 28 | 33.9 (21.0 - 51.8) | 29.3 (19.5 - 42.4) |
| Open Claims | 155,108 | 253,902 | 93.3 | 158.6 | 735 | 1,232 | 7.87 (7.32 - 8.46) | 7.77 (7.34 - 8.21) |
| OEHR | 8,894 | 15,385 | 2.2 | 4.2 | 40 | 72 | 18.0 (12.9 - 24.5) | 17.3 (13.6 - 21.8) |
| Meta-analysis | 181,910 | 300,457 | 105.4 | 180.8 | 963 | 1,595 | 9.14 (8.57 - 9.73) | 8.82 (8.39 - 9.26) |
| Meta-analysis (w/o USOC) | 26,802 | 46,555 | 12.1 | 22.2 | 228 | 363 | 18.9 (16.5 - 21.5) | 16.3 (14.7 - 18.1) |
| **PS-stratification / on-treatment TAR without escalation** | | | | | | | | |
| CCAE | 12,834 | 17,271 | 7.1 | 10.8 | 124 | 150 | 17.5 (14.5 - 20.8) | 13.9 (11.8 - 16.3) |
| ODOD | 8,959 | 11,750 | 4.4 | 6.3 | 84 | 113 | 19.3 (15.4 - 23.8) | 17.8 (14.7 - 21.4) |
| MDCR | 743 | 1,412 | 0.4 | 0.8 | 5 | 13 | 12.2 (3.95 - 28.4) | 16.3 (8.66 - 27.8) |
| MDCD | 1,513 | 2,157 | 0.7 | 1.0 | 25 | 28 | 35.2 (22.8 - 52.0) | 29.2 (19.4 - 42.3) |
| Open Claims | 207,412 | 253,928 | 116.4 | 158.6 | 1,006 | 1,233 | 8.64 (8.12 - 9.19) | 7.77 (7.35 - 8.22) |
| OEHR | 11,723 | 15,387 | 2.9 | 4.2 | 53 | 72 | 18.2 (13.7 - 23.9) | 17.3 (13.6 - 21.8) |
| Meta-analysis | 242,441 | 300,493 | 131.5 | 180.9 | 1,292 | 1,596 | 9.83 (9.30 - 10.4) | 8.82 (8.40 - 9.27) |
| Meta-analysis (w/o USOC) | 35,029 | 46,565 | 15.1 | 22.2 | 286 | 363 | 19.0 (16.8 - 21.3) | 16.3 (14.7 - 18.1) |

##### 4.1.21 Lower extremity amputation

**Supplementary Table 306:** Exposures, outcomes and risk estimates for Lower extremity amputation in GLP1RA (female) (T) vs SGLT2I (female) (C) new-user cohorts under four different propensity score (PS) adjustment / time-at-risk (TAR) designs across data sources. We report total patient exposure time in 1,000 patient-year (KYr) units, uncalibrated and calibrated hazard ratios (HRs) and their 95% confidence intervals (CIs). Greyed out entries did not pass study diagnostics and are included only for completeness.

| | Patients | | Exposure (KYr) | | Outcomes | | Incidence (per KYr) | |
|---|---|---|---|---|---|---|---|---|
| | T | C | T | C | T | C | T | C |
| **PS-matching / on-treatment TAR** | | | | | | | | |
| CCAE | 10,881 | 18,874 | 7.1 | 13.7 | <5 | 5 | < 0.70 (0.23 - 1.64) | 0.37 (0.12 - 0.85) |
| ODOD | 7,484 | 12,827 | 4.2 | 7.9 | 0 | 5 | | 0.64 (0.21 - 1.49) |
| MDCR | 734 | 1,485 | 0.4 | 0.9 | 0 | <5 | | < 5.32 (1.73 - 12.4) |
| MDCD | 1,483 | 2,426 | 0.8 | 1.3 | 0 | 0 | | |
| Open Claims | 170,652 | 276,297 | 115.2 | 204.7 | 21 | 39 | 0.18 (0.11 - 0.28) | 0.19 (0.14 - 0.26) |
| OEHR | 9,708 | 16,652 | 2.8 | 5.2 | 0 | <5 | | < 0.96 (0.31 - 2.23) |
| Meta-analysis | 170,652 | 276,297 | 115.2 | 204.7 | 21 | 39 | 0.18 (0.11 - 0.28) | 0.19 (0.14 - 0.26) |
| **PS-stratification / on-treatment TAR** | | | | | | | | |
| CCAE | 14,357 | 18,874 | 8.8 | 13.7 | <5 | 5 | < 0.57 (0.18 - 1.32) | 0.37 (0.12 - 0.85) |
| ODOD | 10,063 | 12,831 | 5.4 | 7.9 | 0 | 5 | | 0.64 (0.21 - 1.48) |
| MDCR | 818 | 1,486 | 0.5 | 0.9 | 0 | <5 | | < 5.31 (1.73 - 12.4) |
| MDCD | 1,745 | 2,431 | 1.0 | 1.3 | <5 | 0 | < 5.18 (1.68 - 12.1) | |
| Open Claims | 230,754 | 276,304 | 143.8 | 204.7 | 22 | 39 | 0.15 (0.10 - 0.23) | 0.19 (0.14 - 0.26) |
| OEHR | 12,890 | 16,654 | 3.7 | 5.2 | 0 | <5 | | < 0.96 (0.31 - 2.23) |
| Meta-analysis | 230,754 | 276,304 | 143.8 | 204.7 | 22 | 39 | 0.15 (0.10 - 0.23) | 0.19 (0.14 - 0.26) |
| **PS-matching / on-treatment TAR without escalation** | | | | | | | | |
| CCAE | 10,881 | 18,874 | 6.4 | 11.8 | <5 | <5 | < 0.78 (0.25 - 1.82) | < 0.42 (0.14 - 0.99) |
| ODOD | 7,484 | 12,827 | 3.9 | 7.0 | 0 | <5 | | < 0.72 (0.23 - 1.68) |
| MDCR | 734 | 1,485 | 0.4 | 0.8 | 0 | <5 | | < 6.01 (1.95 - 14.0) |
| MDCD | 1,483 | 2,426 | 0.7 | 1.1 | 0 | 0 | | |
| Open Claims | 170,652 | 276,297 | 103.0 | 172.1 | 15 | 27 | 0.15 (0.08 - 0.24) | 0.16 (0.10 - 0.23) |
| OEHR | 9,708 | 16,652 | 2.4 | 4.5 | 0 | <5 | | < 1.10 (0.36 - 2.56) |
| Meta-analysis | 170,652 | 276,297 | 103.0 | 172.1 | 15 | 27 | 0.15 (0.08 - 0.24) | 0.16 (0.10 - 0.23) |
| **PS-stratification / on-treatment TAR without escalation** | | | | | | | | |
| CCAE | 14,357 | 18,874 | 8.0 | 11.8 | <5 | <5 | < 0.62 (0.20 - 1.46) | < 0.42 (0.14 - 0.99) |
| ODOD | 10,063 | 12,831 | 4.9 | 7.0 | 0 | <5 | | < 0.72 (0.23 - 1.67) |
| MDCR | 818 | 1,486 | 0.4 | 0.8 | 0 | <5 | | < 6.00 (1.95 - 14.0) |
| MDCD | 1,745 | 2,431 | 0.8 | 1.1 | <5 | 0 | < 6.00 (1.95 - 14.0) | |
| Open Claims | 230,754 | 276,304 | 129.7 | 172.1 | 16 | 27 | 0.12 (0.07 - 0.20) | 0.16 (0.10 - 0.23) |
| OEHR | 12,890 | 16,654 | 3.2 | 4.6 | 0 | <5 | | < 1.10 (0.36 - 2.56) |
| Meta-analysis | 230,754 | 276,304 | 129.7 | 172.1 | 16 | 27 | 0.12 (0.07 - 0.20) | 0.16 (0.10 - 0.23) |

##### 4.1.22 Nausea

**Supplementary Table 307:** Exposures, outcomes and risk estimates for Nausea in GLP1RA (female) (T) vs SGLT2I (female) (C) new-user cohorts under four different propensity score (PS) adjustment / time-at-risk (TAR) designs across data sources. We report total patient exposure time in 1,000 patient-year (KYr) units, uncalibrated and calibrated hazard ratios (HRs) and their 95% confidence intervals (CIs). Greyed out entries did not pass study diagnostics and are included only for completeness.

| | Patients | | Exposure (KYr) | | Outcomes | | Incidence (per KYr) | |
|---|---|---|---|---|---|---|---|---|
| | T | C | T | C | T | C | T | C |
| **PS-matching / on-treatment TAR** | | | | | | | | |
| CCAE | 8,590 | 15,256 | 5.4 | 10.9 | 385 | 515 | 71.1 (64.2 - 78.6) | 47.2 (43.2 - 51.4) |
| ODOD | 5,818 | 10,199 | 3.2 | 6.2 | 292 | 321 | 91.4 (81.2 - 103) | 51.9 (46.4 - 57.9) |
| MDCR | 643 | 1,258 | 0.4 | 0.8 | 23 | 32 | 60.9 (38.6 - 91.3) | 40.6 (27.8 - 57.4) |
| MDCD | 909 | 1,538 | 0.5 | 0.8 | 87 | 79 | 175 (140 - 215) | 96.3 (76.2 - 120) |
| Open Claims | 132,962 | 221,448 | 89.5 | 165.5 | 3,475 | 3,960 | 38.8 (37.5 - 40.1) | 23.9 (23.2 - 24.7) |
| OEHR | 7,954 | 14,002 | 2.2 | 4.3 | 203 | 230 | 90.6 (78.6 - 104) | 53.8 (47.1 - 61.2) |
| Meta-analysis | 156,233 | 262,443 | 100.9 | 187.7 | 4,442 | 5,105 | 44.0 (42.7 - 45.3) | 27.2 (26.5 - 28.0) |
| Meta-analysis (w/o USOC) | 23,271 | 40,995 | 11.3 | 22.2 | 967 | 1,145 | 85.2 (79.9 - 90.8) | 51.6 (48.6 - 54.7) |
| **PS-stratification / on-treatment TAR** | | | | | | | | |
| CCAE | 11,176 | 15,258 | 6.6 | 10.9 | 504 | 515 | 76.0 (69.5 - 82.9) | 47.1 (43.2 - 51.4) |
| ODOD | 7,686 | 10,200 | 4.0 | 6.2 | 406 | 321 | 103 (92.9 - 113) | 51.9 (46.4 - 57.9) |
| MDCR | 721 | 1,258 | 0.4 | 0.8 | 25 | 32 | 61.6 (39.9 - 90.9) | 40.6 (27.8 - 57.4) |
| MDCD | 1,056 | 1,543 | 0.6 | 0.8 | 93 | 80 | 165 (133 - 202) | 97.2 (77.1 - 121) |
| Open Claims | 175,582 | 221,452 | 109.4 | 165.5 | 4,646 | 3,960 | 42.5 (41.3 - 43.7) | 23.9 (23.2 - 24.7) |
| OEHR | 10,399 | 14,004 | 2.9 | 4.3 | 273 | 230 | 94.3 (83.4 - 106) | 53.8 (47.1 - 61.2) |
| Meta-analysis | 205,899 | 262,457 | 123.4 | 187.7 | 5,922 | 5,106 | 48.0 (46.8 - 49.2) | 27.2 (26.5 - 28.0) |
| Meta-analysis (w/o USOC) | 30,317 | 41,005 | 14.1 | 22.2 | 1,276 | 1,146 | 90.8 (85.9 - 95.9) | 51.6 (48.7 - 54.7) |
| **PS-matching / on-treatment TAR without escalation** | | | | | | | | |
| CCAE | 8,590 | 15,256 | 4.9 | 9.5 | 353 | 450 | 71.8 (64.5 - 79.7) | 47.3 (43.1 - 51.9) |
| ODOD | 5,818 | 10,199 | 2.9 | 5.5 | 272 | 277 | 92.7 (82.0 - 104) | 50.1 (44.3 - 56.3) |
| MDCR | 643 | 1,258 | 0.4 | 0.7 | 20 | 29 | 56.2 (34.3 - 86.8) | 41.6 (27.9 - 59.8) |
| MDCD | 909 | 1,538 | 0.4 | 0.7 | 80 | 64 | 186 (147 - 231) | 91.8 (70.7 - 117) |
| Open Claims | 132,962 | 221,448 | 80.3 | 140.3 | 3,121 | 3,228 | 38.9 (37.5 - 40.3) | 23.0 (22.2 - 23.8) |
| OEHR | 7,954 | 14,002 | 1.9 | 3.8 | 178 | 190 | 91.9 (78.9 - 106) | 50.6 (43.7 - 58.3) |
| Meta-analysis | 156,233 | 262,443 | 90.5 | 159.8 | 4,004 | 4,209 | 44.3 (42.9 - 45.6) | 26.3 (25.6 - 27.2) |
| Meta-analysis (w/o USOC) | 23,271 | 40,995 | 10.2 | 19.5 | 883 | 981 | 86.4 (80.8 - 92.3) | 50.3 (47.2 - 53.6) |
| **PS-stratification / on-treatment TAR without escalation** | | | | | | | | |
| CCAE | 11,176 | 15,258 | 6.1 | 9.5 | 465 | 450 | 76.8 (70.0 - 84.1) | 47.3 (43.1 - 51.9) |
| ODOD | 7,686 | 10,200 | 3.7 | 5.5 | 382 | 277 | 105 (94.4 - 116) | 50.1 (44.3 - 56.3) |
| MDCR | 721 | 1,258 | 0.4 | 0.7 | 22 | 29 | 57.3 (35.9 - 86.8) | 41.6 (27.9 - 59.8) |
| MDCD | 1,056 | 1,543 | 0.5 | 0.7 | 86 | 65 | 175 (140 - 216) | 93.0 (71.8 - 118) |
| Open Claims | 175,582 | 221,452 | 98.9 | 140.3 | 4,231 | 3,228 | 42.8 (41.5 - 44.1) | 23.0 (22.2 - 23.8) |
| OEHR | 10,399 | 14,004 | 2.5 | 3.8 | 239 | 190 | 94.4 (82.9 - 107) | 50.6 (43.7 - 58.3) |
| Meta-analysis | 205,899 | 262,457 | 111.6 | 159.8 | 5,403 | 4,210 | 48.4 (47.1 - 49.7) | 26.4 (25.6 - 27.2) |
| Meta-analysis (w/o USOC) | 30,317 | 41,005 | 12.7 | 19.5 | 1,172 | 982 | 92.1 (86.9 - 97.5) | 50.4 (47.3 - 53.6) |

##### 4.1.23 Peripheral edema

**Supplementary Table 308:** Exposures, outcomes and risk estimates for Peripheral edema in GLP1RA (female) (T) vs SGLT2I (female) (C) new-user cohorts under four different propensity score (PS) adjustment / time-at-risk (TAR) designs across data sources. We report total patient exposure time in 1,000 patient-year (KYr) units, uncalibrated and calibrated hazard ratios (HRs) and their 95% confidence intervals (CIs). Greyed out entries did not pass study diagnostics and are included only for completeness.

| | Patients | | Exposure (KYr) | | Outcomes | | Incidence (per KYr) | |
|---|---|---|---|---|---|---|---|---|
| | T | C | T | C | T | C | T | C |
| **PS-matching / on-treatment TAR** | | | | | | | | |
| CCAE | 8,076 | 14,268 | 5.1 | 10.1 | 258 | 434 | 50.2 (44.3 - 56.7) | 43.2 (39.2 - 47.4) |
| ODOD | 5,356 | 9,262 | 3.0 | 5.5 | 169 | 324 | 57.0 (48.8 - 66.3) | 58.7 (52.5 - 65.4) |
| MDCR | 511 | 1,086 | 0.3 | 0.7 | 23 | 30 | 77.8 (49.3 - 117) | 44.7 (30.2 - 63.9) |
| MDCD | 937 | 1,533 | 0.5 | 0.8 | 44 | 69 | 84.5 (61.4 - 113) | 89.3 (69.5 - 113) |
| Open Claims | 121,526 | 201,851 | 81.0 | 149.5 | 2,047 | 3,611 | 25.3 (24.2 - 26.4) | 24.1 (23.4 - 24.9) |
| OEHR | 7,661 | 13,267 | 2.2 | 4.1 | 97 | 157 | 44.8 (36.3 - 54.7) | 38.5 (32.7 - 45.0) |
| Meta-analysis | 143,556 | 240,181 | 91.8 | 170.0 | 2,615 | 4,595 | 28.5 (27.4 - 29.6) | 27.0 (26.3 - 27.8) |
| Meta-analysis (w/o USOC) | 22,030 | 38,330 | 10.8 | 20.4 | 568 | 984 | 52.7 (48.4 - 57.2) | 48.2 (45.2 - 51.3) |
| **PS-stratification / on-treatment TAR** | | | | | | | | |
| CCAE | 10,423 | 14,270 | 6.2 | 10.1 | 318 | 434 | 51.1 (45.6 - 57.0) | 43.2 (39.2 - 47.4) |
| ODOD | 7,141 | 9,264 | 3.7 | 5.5 | 219 | 325 | 59.5 (51.9 - 67.9) | 58.9 (52.6 - 65.6) |
| MDCR | 569 | 1,087 | 0.3 | 0.7 | 25 | 30 | 78.4 (50.8 - 116) | 44.7 (30.1 - 63.7) |
| MDCD | 1,097 | 1,536 | 0.6 | 0.8 | 48 | 69 | 80.8 (59.6 - 107) | 89.1 (69.3 - 113) |
| Open Claims | 160,550 | 201,879 | 99.1 | 149.5 | 2,527 | 3,614 | 25.5 (24.5 - 26.5) | 24.2 (23.4 - 25.0) |
| OEHR | 9,971 | 13,269 | 2.8 | 4.1 | 129 | 157 | 46.6 (38.9 - 55.3) | 38.5 (32.7 - 45.0) |
| Meta-analysis | 189,182 | 240,218 | 112.4 | 170.0 | 3,241 | 4,599 | 28.8 (27.9 - 29.9) | 27.1 (26.3 - 27.8) |
| Meta-analysis (w/o USOC) | 28,632 | 38,339 | 13.3 | 20.4 | 714 | 985 | 53.8 (49.9 - 57.9) | 48.2 (45.2 - 51.3) |
| **PS-matching / on-treatment TAR without escalation** | | | | | | | | |
| CCAE | 8,076 | 14,268 | 4.6 | 8.7 | 232 | 372 | 50.0 (43.7 - 56.8) | 42.7 (38.5 - 47.3) |
| ODOD | 5,356 | 9,262 | 2.7 | 4.9 | 158 | 287 | 58.1 (49.4 - 67.9) | 58.1 (51.6 - 65.3) |
| MDCR | 511 | 1,086 | 0.3 | 0.6 | 22 | 28 | 79.8 (50.0 - 121) | 46.2 (30.7 - 66.8) |
| MDCD | 937 | 1,533 | 0.5 | 0.6 | 37 | 60 | 81.5 (57.4 - 112) | 94.2 (71.9 - 121) |
| Open Claims | 121,526 | 201,851 | 72.4 | 126.4 | 1,762 | 2,984 | 24.3 (23.2 - 25.5) | 23.6 (22.8 - 24.5) |
| OEHR | 7,661 | 13,267 | 1.9 | 3.6 | 89 | 131 | 47.3 (37.9 - 58.2) | 36.8 (30.8 - 43.7) |
| Meta-analysis | 143,556 | 240,181 | 82.1 | 144.3 | 2,278 | 3,834 | 27.7 (26.6 - 28.9) | 26.6 (25.7 - 27.4) |
| Meta-analysis (w/o USOC) | 22,030 | 38,330 | 9.7 | 17.8 | 516 | 850 | 53.2 (48.7 - 58.0) | 47.6 (44.5 - 51.0) |
| **PS-stratification / on-treatment TAR without escalation** | | | | | | | | |
| CCAE | 10,423 | 14,270 | 5.6 | 8.7 | 283 | 372 | 50.1 (44.4 - 56.3) | 42.7 (38.5 - 47.3) |
| ODOD | 7,141 | 9,264 | 3.4 | 4.9 | 204 | 288 | 60.1 (52.1 - 68.9) | 58.3 (51.8 - 65.5) |
| MDCR | 569 | 1,087 | 0.3 | 0.6 | 24 | 28 | 81.5 (52.2 - 121) | 46.1 (30.6 - 66.7) |
| MDCD | 1,097 | 1,536 | 0.5 | 0.6 | 40 | 60 | 77.2 (55.1 - 105) | 93.9 (71.6 - 121) |
| Open Claims | 160,550 | 201,879 | 89.4 | 126.4 | 2,212 | 2,985 | 24.7 (23.7 - 25.8) | 23.6 (22.8 - 24.5) |
| OEHR | 9,971 | 13,269 | 2.4 | 3.6 | 119 | 131 | 49.1 (40.7 - 58.8) | 36.8 (30.8 - 43.7) |
| Meta-analysis | 189,182 | 240,218 | 101.4 | 144.3 | 2,858 | 3,836 | 28.2 (27.2 - 29.2) | 26.6 (25.8 - 27.4) |
| Meta-analysis (w/o USOC) | 28,632 | 38,339 | 12.0 | 17.8 | 646 | 851 | 53.9 (49.8 - 58.2) | 47.7 (44.5 - 51.0) |

##### 4.1.24 Photosensitivity

**Supplementary Table 309:** Exposures, outcomes and risk estimates for Photosensitivity in GLP1RA (female) (T) vs SGLT2I (female) (C) new-user cohorts under four different propensity score (PS) adjustment / time-at-risk (TAR) designs across data sources. We report total patient exposure time in 1,000 patient-year (KYr) units, uncalibrated and calibrated hazard ratios (HRs) and their 95% confidence intervals (CIs). Greyed out entries did not pass study diagnostics and are included only for completeness.

| | Patients | | Exposure (KYr) | | Outcomes | | Incidence (per KYr) | |
|---|---|---|---|---|---|---|---|---|
| | T | C | T | C | T | C | T | C |
| **PS-matching / on-treatment TAR** | | | | | | | | |
| CCAE | 10,789 | 18,681 | 7.0 | 13.5 | <5 | 11 | < 0.71 (0.23 - 1.66) | 0.81 (0.41 - 1.46) |
| ODOD | 7,449 | 12,733 | 4.2 | 7.8 | 7 | <5 | 1.67 (0.67 - 3.43) | < 0.64 (0.21 - 1.50) |
| MDCR | 722 | 1,472 | 0.4 | 0.9 | <5 | <5 | < 11.7 (3.80 - 27.3) | < 5.36 (1.74 - 12.5) |
| MDCD | 1,473 | 2,411 | 0.8 | 1.3 | <5 | <5 | < 6.05 (1.96 - 14.1) | < 3.85 (1.25 - 8.99) |
| Open Claims | 168,804 | 273,492 | 113.8 | 202.5 | 42 | 72 | 0.37 (0.27 - 0.50) | 0.36 (0.28 - 0.45) |
| OEHR | 9,639 | 16,533 | 2.8 | 5.2 | 6 | <5 | 2.14 (0.78 - 4.65) | < 0.96 (0.31 - 2.25) |
| Meta-analysis | 168,804 | 273,492 | 113.8 | 202.5 | 42 | 72 | 0.37 (0.27 - 0.50) | 0.36 (0.28 - 0.45) |
| **PS-stratification / on-treatment TAR** | | | | | | | | |
| CCAE | 14,215 | 18,681 | 8.7 | 13.5 | 7 | 11 | 0.80 (0.32 - 1.65) | 0.81 (0.41 - 1.46) |
| ODOD | 10,003 | 12,737 | 5.3 | 7.8 | 7 | <5 | 1.31 (0.53 - 2.71) | < 0.64 (0.21 - 1.50) |
| MDCR | 797 | 1,473 | 0.5 | 0.9 | <5 | <5 | < 10.9 (3.54 - 25.4) | < 5.35 (1.74 - 12.5) |
| MDCD | 1,727 | 2,416 | 1.0 | 1.3 | <5 | <5 | < 5.22 (1.70 - 12.2) | < 3.84 (1.25 - 8.97) |
| Open Claims | 228,019 | 273,499 | 141.9 | 202.5 | 50 | 72 | 0.35 (0.26 - 0.46) | 0.36 (0.28 - 0.45) |
| OEHR | 12,790 | 16,535 | 3.7 | 5.2 | 8 | <5 | 2.17 (0.94 - 4.28) | < 0.96 (0.31 - 2.25) |
| Meta-analysis | 242,234 | 292,180 | 150.6 | 216.0 | 57 | 83 | 0.38 (0.29 - 0.49) | 0.38 (0.31 - 0.48) |
| Meta-analysis (w/o USOC) | 14,215 | 18,681 | 8.7 | 13.5 | 7 | 11 | 0.80 (0.32 - 1.65) | 0.81 (0.41 - 1.46) |
| **PS-matching / on-treatment TAR without escalation** | | | | | | | | |
| CCAE | 10,789 | 18,681 | 6.3 | 11.7 | <5 | 10 | < 0.79 (0.26 - 1.84) | 0.86 (0.41 - 1.58) |
| ODOD | 7,449 | 12,733 | 3.8 | 6.9 | 7 | <5 | 1.82 (0.73 - 3.76) | < 0.72 (0.24 - 1.69) |
| MDCR | 722 | 1,472 | 0.4 | 0.8 | <5 | <5 | < 12.5 (4.06 - 29.1) | < 6.05 (1.96 - 14.1) |
| MDCD | 1,473 | 2,411 | 0.7 | 1.1 | <5 | <5 | < 6.98 (2.27 - 16.3) | < 4.67 (1.52 - 10.9) |
| Open Claims | 168,804 | 273,492 | 101.7 | 170.2 | 31 | 59 | 0.30 (0.21 - 0.43) | 0.35 (0.26 - 0.45) |
| OEHR | 9,639 | 16,533 | 2.4 | 4.5 | <5 | <5 | < 2.06 (0.67 - 4.82) | < 1.11 (0.36 - 2.58) |
| Meta-analysis | 168,804 | 273,492 | 101.7 | 170.2 | 31 | 59 | 0.30 (0.21 - 0.43) | 0.35 (0.26 - 0.45) |
| **PS-stratification / on-treatment TAR without escalation** | | | | | | | | |
| CCAE | 14,215 | 18,681 | 7.9 | 11.7 | 7 | 10 | 0.88 (0.36 - 1.82) | 0.86 (0.41 - 1.58) |
| ODOD | 10,003 | 12,737 | 4.9 | 6.9 | 7 | <5 | 1.43 (0.58 - 2.95) | < 0.72 (0.23 - 1.69) |
| MDCR | 797 | 1,473 | 0.4 | 0.8 | <5 | <5 | < 11.6 (3.78 - 27.2) | < 6.04 (1.96 - 14.1) |
| MDCD | 1,727 | 2,416 | 0.8 | 1.1 | <5 | <5 | < 6.04 (1.96 - 14.1) | < 4.65 (1.51 - 10.9) |
| Open Claims | 228,019 | 273,499 | 128.0 | 170.2 | 38 | 59 | 0.30 (0.21 - 0.41) | 0.35 (0.26 - 0.45) |
| OEHR | 12,790 | 16,535 | 3.2 | 4.5 | 5 | <5 | 1.56 (0.51 - 3.64) | < 1.11 (0.36 - 2.58) |
| Meta-analysis | 242,234 | 292,180 | 136.0 | 181.9 | 45 | 69 | 0.33 (0.24 - 0.44) | 0.38 (0.30 - 0.48) |
| Meta-analysis (w/o USOC) | 14,215 | 18,681 | 7.9 | 11.7 | 7 | 10 | 0.88 (0.36 - 1.82) | 0.86 (0.41 - 1.58) |

##### 4.1.25 Venous thromboembolism

**Supplementary Table 310:** Exposures, outcomes and risk estimates for Venous thromboembolism in GLP1RA (female) (T) vs SGLT2I (female) (C) new-user cohorts under four different propensity score (PS) adjustment / time-at-risk (TAR) designs across data sources. We report total patient exposure time in 1,000 patient-year (KYr) units, uncalibrated and calibrated hazard ratios (HRs) and their 95% confidence intervals (CIs). Greyed out entries did not pass study diagnostics and are included only for completeness.

| | Patients | | Exposure (KYr) | | Outcomes | | Incidence (per KYr) | |
|---|---|---|---|---|---|---|---|---|
| | T | C | T | C | T | C | T | C |
| **PS-matching / on-treatment TAR** | | | | | | | | |
| CCAE | 10,610 | 18,464 | 6.9 | 13.4 | 31 | 59 | 4.49 (3.05 - 6.37) | 4.42 (3.36 - 5.70) |
| ODOD | 7,217 | 12,413 | 4.1 | 7.6 | 32 | 60 | 7.83 (5.35 - 11.1) | 7.91 (6.04 - 10.2) |
| MDCR | 704 | 1,425 | 0.4 | 0.9 | 5 | 5 | 12.4 (4.04 - 29.0) | 5.48 (1.78 - 12.8) |
| MDCD | 1,412 | 2,319 | 0.8 | 1.2 | 5 | 10 | 6.29 (2.04 - 14.7) | 8.02 (3.85 - 14.8) |
| Open Claims | 164,239 | 267,171 | 110.9 | 198.1 | 406 | 730 | 3.66 (3.31 - 4.04) | 3.69 (3.42 - 3.96) |
| OEHR | 9,470 | 16,263 | 2.8 | 5.1 | 17 | 38 | 6.15 (3.58 - 9.84) | 7.47 (5.28 - 10.2) |
| Meta-analysis | 191,536 | 314,311 | 124.6 | 224.1 | 486 | 887 | 3.90 (3.56 - 4.26) | 3.96 (3.70 - 4.23) |
| Meta-analysis (w/o USOC) | 27,297 | 47,140 | 13.8 | 26.0 | 80 | 157 | 5.81 (4.61 - 7.24) | 6.03 (5.13 - 7.05) |
| **PS-stratification / on-treatment TAR** | | | | | | | | |
| CCAE | 13,935 | 18,464 | 8.5 | 13.4 | 37 | 59 | 4.35 (3.06 - 5.99) | 4.42 (3.36 - 5.70) |
| ODOD | 9,685 | 12,418 | 5.2 | 7.6 | 43 | 60 | 8.33 (6.03 - 11.2) | 7.91 (6.03 - 10.2) |
| MDCR | 778 | 1,426 | 0.4 | 0.9 | 5 | 5 | 11.3 (3.67 - 26.4) | 5.48 (1.78 - 12.8) |
| MDCD | 1,661 | 2,324 | 0.9 | 1.2 | 6 | 10 | 6.54 (2.40 - 14.2) | 8.00 (3.84 - 14.7) |
| Open Claims | 221,626 | 267,177 | 138.1 | 198.1 | 506 | 730 | 3.66 (3.35 - 4.00) | 3.69 (3.42 - 3.96) |
| OEHR | 12,519 | 16,265 | 3.6 | 5.1 | 25 | 38 | 6.93 (4.48 - 10.2) | 7.47 (5.28 - 10.2) |
| Meta-analysis | 257,765 | 314,324 | 155.4 | 224.1 | 611 | 887 | 3.93 (3.63 - 4.26) | 3.96 (3.70 - 4.23) |
| Meta-analysis (w/o USOC) | 36,139 | 47,147 | 17.3 | 26.0 | 105 | 157 | 6.08 (4.97 - 7.36) | 6.03 (5.12 - 7.05) |
| **PS-matching / on-treatment TAR without escalation** | | | | | | | | |
| CCAE | 10,610 | 18,464 | 6.2 | 11.5 | 30 | 49 | 4.82 (3.25 - 6.88) | 4.25 (3.15 - 5.62) |
| ODOD | 7,217 | 12,413 | 3.7 | 6.7 | 28 | 53 | 7.49 (4.98 - 10.8) | 7.88 (5.90 - 10.3) |
| MDCR | 704 | 1,425 | 0.4 | 0.8 | <5 | <5 | < 13.2 (4.27 - 30.7) | < 6.20 (2.01 - 14.5) |
| MDCD | 1,412 | 2,319 | 0.7 | 1.0 | <5 | 7 | < 7.24 (2.35 - 16.9) | 6.83 (2.74 - 14.1) |
| Open Claims | 164,239 | 267,171 | 99.1 | 166.7 | 352 | 597 | 3.55 (3.19 - 3.94) | 3.58 (3.30 - 3.88) |
| OEHR | 9,470 | 16,263 | 2.4 | 4.4 | 15 | 32 | 6.28 (3.52 - 10.4) | 7.23 (4.94 - 10.2) |
| Meta-analysis | 191,536 | 314,311 | 111.5 | 189.3 | 425 | 731 | 3.81 (3.46 - 4.19) | 3.86 (3.59 - 4.15) |
| Meta-analysis (w/o USOC) | 27,297 | 47,140 | 12.4 | 22.7 | 73 | 134 | 5.91 (4.63 - 7.43) | 5.91 (4.95 - 7.00) |
| **PS-stratification / on-treatment TAR without escalation** | | | | | | | | |
| CCAE | 13,935 | 18,464 | 7.7 | 11.5 | 35 | 49 | 4.53 (3.16 - 6.30) | 4.25 (3.15 - 5.62) |
| ODOD | 9,685 | 12,418 | 4.7 | 6.7 | 39 | 53 | 8.23 (5.85 - 11.2) | 7.88 (5.90 - 10.3) |
| MDCR | 778 | 1,426 | 0.4 | 0.8 | <5 | <5 | < 12.0 (3.89 - 28.0) | < 6.19 (2.01 - 14.4) |
| MDCD | 1,661 | 2,324 | 0.8 | 1.0 | <5 | 7 | < 6.27 (2.04 - 14.6) | 6.80 (2.73 - 14.0) |
| Open Claims | 221,626 | 267,177 | 124.6 | 166.7 | 444 | 597 | 3.56 (3.24 - 3.91) | 3.58 (3.30 - 3.88) |
| OEHR | 12,519 | 16,265 | 3.1 | 4.4 | 22 | 32 | 7.01 (4.39 - 10.6) | 7.23 (4.94 - 10.2) |
| Meta-analysis | 257,765 | 314,324 | 140.2 | 189.3 | 540 | 731 | 3.85 (3.53 - 4.19) | 3.86 (3.59 - 4.15) |
| Meta-analysis (w/o USOC) | 36,139 | 47,147 | 15.6 | 22.7 | 96 | 134 | 6.15 (4.98 - 7.51) | 5.91 (4.95 - 7.00) |

##### 4.1.26 Vomiting

**Supplementary Table 311:** Exposures, outcomes and risk estimates for Vomiting in GLP1RA (female) (T) vs SGLT2I (female) (C) new-user cohorts under four different propensity score (PS) adjustment / time-at-risk (TAR) designs across data sources. We report total patient exposure time in 1,000 patient-year (KYr) units, uncalibrated and calibrated hazard ratios (HRs) and their 95% confidence intervals (CIs). Greyed out entries did not pass study diagnostics and are included only for completeness.

| | Patients | | Exposure (KYr) | | Outcomes | | Incidence (per KYr) | |
|---|---|---|---|---|---|---|---|---|
| | T | C | T | C | T | C | T | C |
| **PS-matching / on-treatment TAR** | | | | | | | | |
| CCAE | 9,342 | 16,473 | 6.0 | 11.9 | 303 | 353 | 50.6 (45.1 - 56.6) | 29.7 (26.7 - 33.0) |
| ODOD | 6,403 | 11,061 | 3.6 | 6.7 | 215 | 209 | 60.2 (52.4 - 68.8) | 31.0 (27.0 - 35.6) |
| MDCR | 663 | 1,345 | 0.4 | 0.8 | 17 | 21 | 43.3 (25.2 - 69.4) | 24.8 (15.3 - 37.9) |
| MDCD | 1,035 | 1,722 | 0.6 | 0.9 | 63 | 70 | 110 (84.5 - 141) | 74.9 (58.4 - 94.7) |
| Open Claims | 143,217 | 235,916 | 96.6 | 176.1 | 2,492 | 2,889 | 25.8 (24.8 - 26.8) | 16.4 (15.8 - 17.0) |
| OEHR | 8,617 | 15,026 | 2.5 | 4.6 | 126 | 137 | 51.0 (42.5 - 60.7) | 29.5 (24.8 - 34.9) |
| Meta-analysis | 168,614 | 280,198 | 109.2 | 200.2 | 3,199 | 3,658 | 29.3 (28.3 - 30.3) | 18.3 (17.7 - 18.9) |
| Meta-analysis (w/o USOC) | 25,397 | 44,282 | 12.6 | 24.2 | 707 | 769 | 56.1 (52.0 - 60.4) | 31.8 (29.6 - 34.1) |
| **PS-stratification / on-treatment TAR** | | | | | | | | |
| CCAE | 12,234 | 16,475 | 7.4 | 11.9 | 393 | 353 | 53.2 (48.1 - 58.7) | 29.7 (26.7 - 33.0) |
| ODOD | 8,551 | 11,062 | 4.5 | 6.7 | 299 | 210 | 66.8 (59.4 - 74.8) | 31.2 (27.1 - 35.7) |
| MDCR | 743 | 1,346 | 0.4 | 0.8 | 19 | 21 | 45.2 (27.2 - 70.6) | 24.7 (15.3 - 37.8) |
| MDCD | 1,209 | 1,727 | 0.6 | 0.9 | 72 | 71 | 111 (86.7 - 140) | 75.8 (59.2 - 95.6) |
| Open Claims | 191,085 | 235,920 | 119.1 | 176.1 | 3,332 | 2,889 | 28.0 (27.0 - 28.9) | 16.4 (15.8 - 17.0) |
| OEHR | 11,301 | 15,028 | 3.2 | 4.6 | 169 | 137 | 52.8 (45.1 - 61.4) | 29.5 (24.8 - 34.9) |
| Meta-analysis | 224,380 | 280,212 | 134.8 | 200.2 | 4,265 | 3,660 | 31.6 (30.7 - 32.6) | 18.3 (17.7 - 18.9) |
| Meta-analysis (w/o USOC) | 33,295 | 44,292 | 15.7 | 24.2 | 933 | 771 | 59.4 (55.6 - 63.3) | 31.9 (29.7 - 34.2) |
| **PS-matching / on-treatment TAR without escalation** | | | | | | | | |
| CCAE | 9,342 | 16,473 | 5.4 | 10.3 | 279 | 301 | 51.5 (45.7 - 57.9) | 29.2 (26.0 - 32.7) |
| ODOD | 6,403 | 11,061 | 3.3 | 6.0 | 198 | 178 | 60.7 (52.6 - 69.8) | 29.7 (25.5 - 34.4) |
| MDCR | 663 | 1,345 | 0.4 | 0.7 | 16 | 20 | 43.3 (24.7 - 70.3) | 26.7 (16.3 - 41.3) |
| MDCD | 1,035 | 1,722 | 0.5 | 0.8 | 58 | 55 | 117 (88.8 - 151) | 70.1 (52.8 - 91.2) |
| Open Claims | 143,217 | 235,916 | 86.6 | 148.9 | 2,232 | 2,319 | 25.8 (24.7 - 26.9) | 15.6 (15.0 - 16.2) |
| OEHR | 8,617 | 15,026 | 2.1 | 4.1 | 109 | 112 | 51.0 (41.9 - 61.5) | 27.6 (22.7 - 33.2) |
| Meta-analysis | 168,614 | 280,198 | 97.9 | 170.0 | 2,876 | 2,965 | 29.4 (28.3 - 30.5) | 17.4 (16.8 - 18.1) |
| Meta-analysis (w/o USOC) | 25,397 | 44,282 | 11.3 | 21.1 | 644 | 646 | 56.9 (52.6 - 61.5) | 30.6 (28.3 - 33.0) |
| **PS-stratification / on-treatment TAR without escalation** | | | | | | | | |
| CCAE | 12,234 | 16,475 | 6.7 | 10.3 | 363 | 301 | 54.0 (48.5 - 59.8) | 29.2 (26.0 - 32.7) |
| ODOD | 8,551 | 11,062 | 4.1 | 6.0 | 276 | 179 | 67.1 (59.4 - 75.5) | 29.9 (25.7 - 34.6) |
| MDCR | 743 | 1,346 | 0.4 | 0.7 | 17 | 20 | 43.0 (25.1 - 68.9) | 26.7 (16.3 - 41.2) |
| MDCD | 1,209 | 1,727 | 0.6 | 0.8 | 66 | 56 | 117 (90.4 - 149) | 71.2 (53.8 - 92.4) |
| Open Claims | 191,085 | 235,920 | 107.6 | 148.9 | 3,019 | 2,319 | 28.0 (27.1 - 29.1) | 15.6 (15.0 - 16.2) |
| OEHR | 11,301 | 15,028 | 2.8 | 4.1 | 146 | 112 | 52.2 (44.1 - 61.4) | 27.6 (22.7 - 33.2) |
| Meta-analysis | 224,380 | 280,212 | 121.9 | 170.0 | 3,870 | 2,967 | 31.8 (30.8 - 32.8) | 17.5 (16.8 - 18.1) |
| Meta-analysis (w/o USOC) | 33,295 | 44,292 | 14.2 | 21.1 | 851 | 648 | 59.9 (56.0 - 64.1) | 30.7 (28.4 - 33.1) |

#### 4.2 SGLT2I (female) vs DPP4I (female)

##### 4.2.1 3-point MACE

**Supplementary Table 312:** Exposures, outcomes and risk estimates for 3-point MACE in SGLT2I (female) (T) vs DPP4I (female) (C) new-user cohorts under four different propensity score (PS) adjustment / time-at-risk (TAR) designs across data sources. We report total patient exposure time in 1,000 patient-year (KYr) units, uncalibrated and calibrated hazard ratios (HRs) and their 95% confidence intervals (CIs). Greyed out entries did not pass study diagnostics and are included only for completeness.

| | Patients | | Exposure (KYr) | | Outcomes | | Incidence (per KYr) | |
|---|---|---|---|---|---|---|---|---|
| | T | C | T | C | T | C | T | C |
| **PS-matching / on-treatment TAR** | | | | | | | | |
| CCAE | 16,056 | 25,285 | 12.1 | 19.0 | 79 | 105 | 6.54 (5.18 - 8.16) | 5.54 (4.53 - 6.70) |
| DAG | 2,939 | 8,792 | 2.4 | 9.3 | 0 | 0 | | |
| ODOD | 10,337 | 19,536 | 6.7 | 14.8 | 78 | 306 | 11.6 (9.14 - 14.4) | 20.6 (18.4 - 23.1) |
| MDCR | 1,234 | 4,463 | 0.8 | 3.8 | 10 | 99 | 11.8 (5.68 - 21.8) | 26.2 (21.3 - 31.9) |
| MDCD | 2,197 | 5,957 | 1.2 | 4.2 | 20 | 86 | 16.8 (10.2 - 25.9) | 20.7 (16.6 - 25.6) |
| Open Claims | 222,562 | 424,612 | 175.7 | 370.6 | 1,044 | 3,895 | 5.94 (5.59 - 6.31) | 10.5 (10.2 - 10.8) |
| OEHR | 14,837 | 33,015 | 4.7 | 17.4 | 29 | 142 | 6.18 (4.14 - 8.88) | 8.17 (6.88 - 9.63) |
| SIDIAP | 2,719 | 14,071 | 3.8 | 30.4 | 13 | 94 | 3.46 (1.84 - 5.92) | 3.09 (2.50 - 3.78) |
| Meta-analysis | 268,708 | 522,476 | 204.2 | 456.4 | 1,263 | 4,628 | 6.19 (5.85 - 6.54) | 10.1 (9.85 - 10.4) |
| Meta-analysis (w/o USOC) | 46,146 | 97,864 | 28.5 | 85.7 | 219 | 733 | 7.70 (6.71 - 8.79) | 8.55 (7.94 - 9.19) |
| **PS-stratification / on-treatment TAR** | | | | | | | | |
| CCAE | 18,456 | 25,317 | 13.4 | 19.0 | 88 | 105 | 6.59 (5.28 - 8.12) | 5.53 (4.52 - 6.69) |
| DAG | 3,243 | 8,833 | 2.5 | 9.3 | 0 | 0 | | |
| ODOD | 12,278 | 19,723 | 7.6 | 15.0 | 85 | 317 | 11.2 (8.98 - 13.9) | 21.2 (18.9 - 23.6) |
| MDCR | 1,395 | 4,506 | 0.9 | 3.8 | 12 | 102 | 13.3 (6.89 - 23.3) | 26.8 (21.8 - 32.5) |
| MDCD | 2,284 | 5,975 | 1.2 | 4.2 | 20 | 89 | 16.4 (10.0 - 25.3) | 21.4 (17.2 - 26.3) |
| Open Claims | 263,915 | 424,925 | 196.3 | 370.9 | 1,135 | 3,909 | 5.78 (5.45 - 6.13) | 10.5 (10.2 - 10.9) |
| OEHR | 16,377 | 34,132 | 5.1 | 17.9 | 31 | 151 | 6.02 (4.09 - 8.55) | 8.42 (7.13 - 9.87) |
| SIDIAP | 2,858 | 14,547 | 3.9 | 31.9 | 13 | 99 | 3.35 (1.78 - 5.72) | 3.10 (2.52 - 3.78) |
| Meta-analysis | 313,884 | 518,644 | 226.2 | 454.7 | 1,352 | 4,581 | 5.98 (5.66 - 6.30) | 10.1 (9.78 - 10.4) |
| Meta-analysis (w/o USOC) | 49,969 | 93,719 | 30.0 | 83.8 | 217 | 672 | 7.24 (6.31 - 8.28) | 8.02 (7.42 - 8.65) |
| **PS-matching / on-treatment TAR without escalation** | | | | | | | | |
| CCAE | 16,056 | 25,285 | 10.4 | 15.5 | 63 | 85 | 6.07 (4.67 - 7.77) | 5.50 (4.39 - 6.80) |
| DAG | 2,939 | 8,792 | 2.2 | 8.3 | 0 | 0 | | |
| ODOD | 10,337 | 19,536 | 6.0 | 12.5 | 70 | 270 | 11.7 (9.15 - 14.8) | 21.7 (19.2 - 24.4) |
| MDCR | 1,234 | 4,463 | 0.7 | 3.3 | 9 | 78 | 12.0 (5.51 - 22.9) | 23.6 (18.7 - 29.5) |
| MDCD | 2,197 | 5,957 | 1.0 | 3.2 | 12 | 66 | 12.1 (6.26 - 21.2) | 20.4 (15.8 - 26.0) |
| Open Claims | 222,562 | 424,612 | 146.8 | 294.7 | 870 | 3,161 | 5.92 (5.54 - 6.33) | 10.7 (10.4 - 11.1) |
| OEHR | 14,837 | 33,015 | 4.1 | 14.0 | 24 | 94 | 5.88 (3.77 - 8.75) | 6.72 (5.43 - 8.23) |
| SIDIAP | 2,719 | 14,071 | 3.3 | 24.1 | 12 | 77 | 3.64 (1.88 - 6.37) | 3.20 (2.53 - 4.00) |
| Meta-analysis | 268,708 | 522,476 | 171.5 | 363.9 | 1,051 | 3,753 | 6.13 (5.76 - 6.51) | 10.3 (9.99 - 10.6) |
| Meta-analysis (w/o USOC) | 46,146 | 97,864 | 24.7 | 69.2 | 181 | 592 | 7.33 (6.30 - 8.48) | 8.56 (7.88 - 9.27) |
| **PS-stratification / on-treatment TAR without escalation** | | | | | | | | |
| CCAE | 18,456 | 25,317 | 11.5 | 15.5 | 71 | 85 | 6.16 (4.81 - 7.76) | 5.49 (4.39 - 6.79) |
| DAG | 3,243 | 8,833 | 2.3 | 8.4 | 0 | 0 | | |
| ODOD | 12,278 | 19,723 | 6.7 | 12.6 | 77 | 281 | 11.5 (9.06 - 14.3) | 22.3 (19.8 - 25.0) |
| MDCR | 1,395 | 4,506 | 0.8 | 3.3 | 10 | 80 | 12.5 (6.00 - 23.0) | 24.1 (19.1 - 29.9) |
| MDCD | 2,284 | 5,975 | 1.0 | 3.2 | 12 | 68 | 11.8 (6.10 - 20.6) | 21.0 (16.3 - 26.6) |
| Open Claims | 263,915 | 424,925 | 165.0 | 294.9 | 947 | 3,175 | 5.74 (5.38 - 6.12) | 10.8 (10.4 - 11.1) |
| OEHR | 16,377 | 34,132 | 4.5 | 14.4 | 26 | 99 | 5.81 (3.79 - 8.51) | 6.85 (5.57 - 8.35) |
| SIDIAP | 2,858 | 14,547 | 3.4 | 25.3 | 12 | 79 | 3.53 (1.82 - 6.16) | 3.13 (2.48 - 3.90) |
| Meta-analysis | 313,884 | 518,644 | 191.2 | 362.7 | 1,133 | 3,719 | 5.93 (5.59 - 6.28) | 10.3 (9.93 - 10.6) |
| Meta-analysis (w/o USOC) | 49,969 | 93,719 | 26.1 | 67.8 | 186 | 544 | 7.12 (6.13 - 8.22) | 8.03 (7.36 - 8.73) |

##### 4.2.2 4-point MACE

**Supplementary Table 313:** Exposures, outcomes and risk estimates for 4-point MACE in SGLT2I (female) (T) vs DPP4I (female) (C) new-user cohorts under four different propensity score (PS) adjustment / time-at-risk (TAR) designs across data sources. We report total patient exposure time in 1,000 patient-year (KYr) units, uncalibrated and calibrated hazard ratios (HRs) and their 95% confidence intervals (CIs). Greyed out entries did not pass study diagnostics and are included only for completeness.

| | Patients | | Exposure (KYr) | | Outcomes | | Incidence (per KYr) | |
|---|---|---|---|---|---|---|---|---|
| | T | C | T | C | T | C | T | C |
| **PS-matching / on-treatment TAR** | | | | | | | | |
| CCAE | 15,870 | 24,934 | 11.9 | 18.7 | 110 | 170 | 9.22 (7.58 - 11.1) | 9.09 (7.78 - 10.6) |
| DAG | 2,939 | 8,792 | 2.4 | 9.3 | 0 | 0 | | |
| ODOD | 9,977 | 18,473 | 6.5 | 13.9 | 117 | 500 | 17.9 (14.8 - 21.4) | 36.0 (32.9 - 39.3) |
| MDCR | 1,170 | 4,153 | 0.8 | 3.5 | 18 | 147 | 22.3 (13.2 - 35.2) | 41.8 (35.3 - 49.2) |
| MDCD | 2,075 | 5,549 | 1.1 | 3.8 | 35 | 147 | 31.0 (21.6 - 43.1) | 38.6 (32.6 - 45.4) |
| Open Claims | 217,181 | 410,711 | 171.9 | 357.2 | 1,536 | 6,409 | 8.94 (8.49 - 9.39) | 17.9 (17.5 - 18.4) |
| OEHR | 14,644 | 32,499 | 4.6 | 17.0 | 57 | 284 | 12.3 (9.35 - 16.0) | 16.7 (14.8 - 18.8) |
| SIDIAP | 2,701 | 14,001 | 3.7 | 30.3 | 13 | 118 | 3.47 (1.85 - 5.93) | 3.90 (3.22 - 4.67) |
| Meta-analysis | 262,448 | 506,167 | 199.9 | 440.9 | 1,868 | 7,628 | 9.35 (8.93 - 9.78) | 17.3 (16.9 - 17.7) |
| Meta-analysis (w/o USOC) | 45,267 | 95,456 | 28.0 | 83.7 | 332 | 1,219 | 11.9 (10.6 - 13.2) | 14.6 (13.8 - 15.4) |
| **PS-stratification / on-treatment TAR** | | | | | | | | |
| CCAE | 18,202 | 24,965 | 13.2 | 18.7 | 121 | 170 | 9.18 (7.62 - 11.0) | 9.08 (7.77 - 10.6) |
| DAG | 3,243 | 8,833 | 2.5 | 9.3 | 0 | 0 | | |
| ODOD | 11,798 | 18,772 | 7.3 | 14.1 | 130 | 528 | 17.8 (14.9 - 21.1) | 37.4 (34.2 - 40.7) |
| MDCR | 1,323 | 4,178 | 0.9 | 3.5 | 20 | 148 | 23.3 (14.2 - 35.9) | 41.9 (35.4 - 49.2) |
| MDCD | 2,158 | 5,565 | 1.2 | 3.8 | 35 | 149 | 30.2 (21.1 - 42.1) | 39.0 (33.0 - 45.8) |
| Open Claims | 255,645 | 410,993 | 191.0 | 357.5 | 1,703 | 6,427 | 8.91 (8.50 - 9.35) | 18.0 (17.5 - 18.4) |
| OEHR | 16,145 | 33,583 | 5.1 | 17.5 | 59 | 306 | 11.6 (8.86 - 15.0) | 17.5 (15.6 - 19.5) |
| SIDIAP | 2,834 | 14,477 | 3.9 | 31.8 | 14 | 127 | 3.62 (1.98 - 6.08) | 4.00 (3.33 - 4.76) |
| Meta-analysis | 304,624 | 502,790 | 220.5 | 439.6 | 2,027 | 7,558 | 9.19 (8.80 - 9.60) | 17.2 (16.8 - 17.6) |
| Meta-analysis (w/o USOC) | 48,979 | 91,797 | 29.4 | 82.1 | 324 | 1,131 | 11.0 (9.85 - 12.3) | 13.8 (13.0 - 14.6) |
| **PS-matching / on-treatment TAR without escalation** | | | | | | | | |
| CCAE | 15,870 | 24,934 | 10.3 | 15.2 | 92 | 143 | 8.96 (7.22 - 11.0) | 9.38 (7.91 - 11.1) |
| DAG | 2,939 | 8,792 | 2.2 | 8.3 | 0 | 0 | | |
| ODOD | 9,977 | 18,473 | 5.8 | 11.7 | 100 | 427 | 17.3 (14.1 - 21.0) | 36.6 (33.2 - 40.2) |
| MDCR | 1,170 | 4,153 | 0.7 | 3.1 | 16 | 122 | 22.4 (12.8 - 36.4) | 39.6 (32.9 - 47.3) |
| MDCD | 2,075 | 5,549 | 0.9 | 3.0 | 24 | 112 | 25.7 (16.4 - 38.2) | 37.7 (31.0 - 45.3) |
| Open Claims | 217,181 | 410,711 | 143.7 | 284.2 | 1,264 | 5,166 | 8.80 (8.32 - 9.29) | 18.2 (17.7 - 18.7) |
| OEHR | 14,644 | 32,499 | 4.0 | 13.7 | 47 | 194 | 11.7 (8.59 - 15.6) | 14.2 (12.2 - 16.3) |
| SIDIAP | 2,701 | 14,001 | 3.3 | 24.0 | 12 | 95 | 3.65 (1.89 - 6.38) | 3.96 (3.21 - 4.85) |
| Meta-analysis | 262,448 | 506,167 | 168.0 | 351.8 | 1,539 | 6,137 | 9.16 (8.71 - 9.63) | 17.4 (17.0 - 17.9) |
| Meta-analysis (w/o USOC) | 45,267 | 95,456 | 24.3 | 67.6 | 275 | 971 | 11.3 (10.0 - 12.7) | 14.4 (13.5 - 15.3) |
| **PS-stratification / on-treatment TAR without escalation** | | | | | | | | |
| CCAE | 18,202 | 24,965 | 11.4 | 15.3 | 102 | 143 | 8.95 (7.30 - 10.9) | 9.37 (7.90 - 11.0) |
| DAG | 3,243 | 8,833 | 2.3 | 8.4 | 0 | 0 | | |
| ODOD | 11,798 | 18,772 | 6.5 | 11.9 | 112 | 453 | 17.3 (14.2 - 20.8) | 38.1 (34.6 - 41.7) |
| MDCR | 1,323 | 4,178 | 0.8 | 3.1 | 17 | 122 | 22.3 (13.0 - 35.8) | 39.5 (32.8 - 47.1) |
| MDCD | 2,158 | 5,565 | 1.0 | 3.0 | 24 | 113 | 24.9 (16.0 - 37.1) | 37.9 (31.2 - 45.6) |
| Open Claims | 255,645 | 410,993 | 160.7 | 284.4 | 1,411 | 5,184 | 8.78 (8.33 - 9.25) | 18.2 (17.7 - 18.7) |
| OEHR | 16,145 | 33,583 | 4.4 | 14.1 | 49 | 209 | 11.1 (8.22 - 14.7) | 14.8 (12.8 - 16.9) |
| SIDIAP | 2,834 | 14,477 | 3.4 | 25.2 | 13 | 99 | 3.84 (2.04 - 6.56) | 3.94 (3.20 - 4.79) |
| Meta-analysis | 304,624 | 502,790 | 186.3 | 350.9 | 1,687 | 6,088 | 9.05 (8.63 - 9.50) | 17.4 (16.9 - 17.8) |
| Meta-analysis (w/o USOC) | 48,979 | 91,797 | 25.7 | 66.5 | 276 | 904 | 10.7 (9.52 - 12.1) | 13.6 (12.7 - 14.5) |

##### 4.2.3 Acute myocardial infarction

**Supplementary Table 314:** Exposures, outcomes and risk estimates for Acute myocardial infarction in SGLT2I (female) (T) vs DPP4I (female) (C) new-user cohorts under four different propensity score (PS) adjustment / time-at-risk (TAR) designs across data sources. We report total patient exposure time in 1,000 patient-year (KYr) units, uncalibrated and calibrated hazard ratios (HRs) and their 95% confidence intervals (CIs). Greyed out entries did not pass study diagnostics and are included only for completeness.

| | Patients | | Exposure (KYr) | | Outcomes | | Incidence (per KYr) | |
|---|---|---|---|---|---|---|---|---|
| | T | C | T | C | T | C | T | C |
| **PS-matching / on-treatment TAR** | | | | | | | | |
| CCAE | 16,247 | 25,644 | 12.2 | 19.2 | 34 | 42 | 2.78 (1.93 - 3.89) | 2.18 (1.57 - 2.95) |
| DAG | 2,939 | 8,792 | 2.4 | 9.3 | 0 | 0 | | |
| ODOD | 10,559 | 20,205 | 6.9 | 15.4 | 31 | 106 | 4.49 (3.05 - 6.37) | 6.88 (5.63 - 8.32) |
| MDCR | 1,262 | 4,678 | 0.9 | 4.0 | 7 | 45 | 8.14 (3.27 - 16.8) | 11.3 (8.25 - 15.1) |
| MDCD | 2,273 | 6,190 | 1.2 | 4.3 | 8 | 28 | 6.45 (2.78 - 12.7) | 6.45 (4.29 - 9.33) |
| Open Claims | 227,528 | 436,789 | 179.4 | 382.1 | 488 | 1,651 | 2.72 (2.48 - 2.97) | 4.32 (4.12 - 4.53) |
| OEHR | 14,941 | 33,414 | 4.7 | 17.6 | 15 | 65 | 3.18 (1.78 - 5.24) | 3.69 (2.84 - 4.70) |
| SIDIAP | 2,735 | 14,188 | 3.8 | 30.7 | <5 | 28 | < 1.32 (0.43 - 3.08) | 0.91 (0.61 - 1.32) |
| Meta-analysis | 274,283 | 536,430 | 208.3 | 469.4 | <581 | 1,920 | < 2.79 (2.57 - 3.03) | 4.09 (3.91 - 4.28) |
| Meta-analysis (w/o USOC) | 46,755 | 99,641 | 28.9 | 87.3 | <93 | 269 | < 3.22 (2.60 - 3.95) | 3.08 (2.72 - 3.47) |
| **PS-stratification / on-treatment TAR** | | | | | | | | |
| CCAE | 18,670 | 25,678 | 13.5 | 19.3 | 35 | 42 | 2.59 (1.80 - 3.60) | 2.18 (1.57 - 2.95) |
| DAG | 3,243 | 8,833 | 2.5 | 9.3 | 0 | 0 | | |
| ODOD | 12,545 | 20,404 | 7.7 | 15.6 | 34 | 108 | 4.40 (3.04 - 6.14) | 6.94 (5.69 - 8.38) |
| MDCR | 1,428 | 4,740 | 0.9 | 4.0 | 7 | 47 | 7.63 (3.07 - 15.7) | 11.7 (8.58 - 15.5) |
| MDCD | 2,362 | 6,208 | 1.3 | 4.4 | 8 | 28 | 6.30 (2.72 - 12.4) | 6.42 (4.27 - 9.28) |
| Open Claims | 269,891 | 437,223 | 200.4 | 382.5 | 548 | 1,660 | 2.73 (2.51 - 2.97) | 4.34 (4.13 - 4.55) |
| OEHR | 16,491 | 34,445 | 5.2 | 18.2 | 17 | 67 | 3.28 (1.91 - 5.25) | 3.69 (2.86 - 4.68) |
| SIDIAP | 2,880 | 14,673 | 3.9 | 32.2 | <5 | 28 | < 1.28 (0.41 - 2.98) | 0.87 (0.58 - 1.26) |
| Meta-analysis | 320,477 | 532,423 | 230.7 | 467.7 | <639 | 1,905 | < 2.77 (2.56 - 2.99) | 4.07 (3.89 - 4.26) |
| Meta-analysis (w/o USOC) | 50,586 | 95,200 | 30.4 | 85.2 | <91 | 245 | < 3.00 (2.41 - 3.68) | 2.88 (2.53 - 3.26) |
| **PS-matching / on-treatment TAR without escalation** | | | | | | | | |
| CCAE | 16,247 | 25,644 | 10.5 | 15.7 | 27 | 29 | 2.57 (1.70 - 3.74) | 1.85 (1.24 - 2.66) |
| DAG | 2,939 | 8,792 | 2.2 | 8.3 | 0 | 0 | | |
| ODOD | 10,559 | 20,205 | 6.1 | 12.9 | 26 | 96 | 4.26 (2.78 - 6.24) | 7.41 (6.01 - 9.05) |
| MDCR | 1,262 | 4,678 | 0.8 | 3.5 | 6 | 39 | 7.91 (2.90 - 17.2) | 11.3 (8.00 - 15.4) |
| MDCD | 2,273 | 6,190 | 1.0 | 3.4 | 6 | 19 | 5.86 (2.15 - 12.8) | 5.64 (3.39 - 8.80) |
| Open Claims | 227,528 | 436,789 | 149.9 | 303.7 | 406 | 1,333 | 2.71 (2.45 - 2.99) | 4.39 (4.16 - 4.63) |
| OEHR | 14,941 | 33,414 | 4.1 | 14.2 | 14 | 40 | 3.41 (1.86 - 5.72) | 2.82 (2.02 - 3.84) |
| SIDIAP | 2,735 | 14,188 | 3.3 | 24.3 | <5 | 21 | < 1.50 (0.49 - 3.51) | 0.86 (0.54 - 1.32) |
| Meta-analysis | 271,548 | 522,242 | 171.6 | 349.9 | 479 | 1,517 | 2.79 (2.55 - 3.05) | 4.34 (4.12 - 4.56) |
| Meta-analysis (w/o USOC) | 44,020 | 85,453 | 21.7 | 46.2 | 73 | 184 | 3.36 (2.63 - 4.23) | 3.99 (3.43 - 4.61) |
| **PS-stratification / on-treatment TAR without escalation** | | | | | | | | |
| CCAE | 18,670 | 25,678 | 11.7 | 15.7 | 28 | 29 | 2.40 (1.59 - 3.47) | 1.85 (1.24 - 2.65) |
| DAG | 3,243 | 8,833 | 2.3 | 8.4 | 0 | 0 | | |
| ODOD | 12,545 | 20,404 | 6.9 | 13.1 | 29 | 98 | 4.23 (2.83 - 6.07) | 7.48 (6.07 - 9.12) |
| MDCR | 1,428 | 4,740 | 0.8 | 3.5 | 6 | 41 | 7.40 (2.71 - 16.1) | 11.7 (8.41 - 15.9) |
| MDCD | 2,362 | 6,208 | 1.0 | 3.4 | 6 | 19 | 5.72 (2.10 - 12.4) | 5.62 (3.38 - 8.78) |
| Open Claims | 269,891 | 437,223 | 168.5 | 304.1 | 458 | 1,342 | 2.72 (2.48 - 2.98) | 4.41 (4.18 - 4.66) |
| OEHR | 16,491 | 34,445 | 4.5 | 14.6 | 16 | 42 | 3.55 (2.03 - 5.76) | 2.88 (2.07 - 3.89) |
| SIDIAP | 2,880 | 14,673 | 3.4 | 25.5 | <5 | 21 | < 1.46 (0.47 - 3.40) | 0.82 (0.51 - 1.26) |
| Meta-analysis | 317,597 | 517,750 | 191.5 | 347.5 | 531 | 1,511 | 2.77 (2.54 - 3.02) | 4.35 (4.13 - 4.57) |
| Meta-analysis (w/o USOC) | 47,706 | 80,527 | 23.0 | 43.4 | 73 | 169 | 3.17 (2.48 - 3.98) | 3.89 (3.33 - 4.53) |

##### 4.2.4 Acute renal failure

**Supplementary Table 315:** Exposures, outcomes and risk estimates for Acute renal failure in SGLT2I (female) (T) vs DPP4I (female) (C) new-user cohorts under four different propensity score (PS) adjustment / time-at-risk (TAR) designs across data sources. We report total patient exposure time in 1,000 patient-year (KYr) units, uncalibrated and calibrated hazard ratios (HRs) and their 95% confidence intervals (CIs). Greyed out entries did not pass study diagnostics and are included only for completeness.

| | Patients | | Exposure (KYr) | | Outcomes | | Incidence (per KYr) | |
|---|---|---|---|---|---|---|---|---|
| | T | C | T | C | T | C | T | C |
| **PS-matching / on-treatment TAR** | | | | | | | | |
| CCAE | 16,249 | 25,527 | 12.2 | 19.2 | 57 | 138 | 4.67 (3.54 - 6.05) | 7.20 (6.05 - 8.51) |
| DAG | 2,939 | 8,792 | 2.4 | 9.3 | 0 | 0 | | |
| ODOD | 10,409 | 19,326 | 6.8 | 14.5 | 109 | 481 | 16.0 (13.2 - 19.3) | 33.1 (30.2 - 36.2) |
| MDCR | 1,266 | 4,529 | 0.9 | 3.8 | 12 | 135 | 13.8 (7.14 - 24.1) | 35.4 (29.7 - 41.9) |
| MDCD | 2,226 | 5,994 | 1.2 | 4.1 | 19 | 141 | 15.7 (9.43 - 24.5) | 34.4 (29.0 - 40.6) |
| Open Claims | 226,364 | 430,258 | 178.7 | 373.9 | 1,226 | 5,473 | 6.86 (6.48 - 7.25) | 14.6 (14.3 - 15.0) |
| OEHR | 14,927 | 33,215 | 4.7 | 17.4 | 36 | 241 | 7.65 (5.36 - 10.6) | 13.8 (12.1 - 15.7) |
| SIDIAP | 2,778 | 14,248 | 3.9 | 30.8 | <5 | 12 | < 1.29 (0.42 - 3.02) | 0.39 (0.20 - 0.68) |
| Meta-analysis | 270,175 | 514,320 | 203.7 | 429.2 | 1,447 | 6,474 | 7.11 (6.74 - 7.48) | 15.1 (14.7 - 15.5) |
| Meta-analysis (w/o USOC) | 43,811 | 84,062 | 24.9 | 55.2 | 221 | 1,001 | 8.87 (7.74 - 10.1) | 18.1 (17.0 - 19.3) |
| **PS-stratification / on-treatment TAR** | | | | | | | | |
| CCAE | 18,692 | 25,555 | 13.5 | 19.2 | 62 | 139 | 4.58 (3.51 - 5.87) | 7.25 (6.09 - 8.56) |
| DAG | 3,243 | 8,833 | 2.5 | 9.3 | 0 | 0 | | |
| ODOD | 12,447 | 19,568 | 7.6 | 14.7 | 120 | 495 | 15.7 (13.0 - 18.8) | 33.6 (30.7 - 36.7) |
| MDCR | 1,438 | 4,575 | 0.9 | 3.8 | 13 | 144 | 14.0 (7.47 - 24.0) | 37.6 (31.7 - 44.3) |
| MDCD | 2,313 | 6,013 | 1.2 | 4.1 | 21 | 141 | 16.9 (10.5 - 25.8) | 34.2 (28.8 - 40.4) |
| Open Claims | 269,448 | 430,532 | 200.1 | 374.2 | 1,386 | 5,492 | 6.93 (6.57 - 7.30) | 14.7 (14.3 - 15.1) |
| OEHR | 16,501 | 34,223 | 5.2 | 17.9 | 39 | 253 | 7.53 (5.36 - 10.3) | 14.1 (12.4 - 16.0) |
| SIDIAP | 2,956 | 14,730 | 4.0 | 32.3 | <5 | 13 | < 1.24 (0.40 - 2.90) | 0.40 (0.21 - 0.69) |
| Meta-analysis | 317,088 | 509,878 | 226.5 | 426.0 | 1,607 | 6,379 | 7.10 (6.75 - 7.45) | 15.0 (14.6 - 15.3) |
| Meta-analysis (w/o USOC) | 47,640 | 79,346 | 26.4 | 51.8 | 221 | 887 | 8.38 (7.32 - 9.57) | 17.1 (16.0 - 18.3) |
| **PS-matching / on-treatment TAR without escalation** | | | | | | | | |
| CCAE | 16,249 | 25,527 | 10.5 | 15.6 | 48 | 113 | 4.58 (3.38 - 6.08) | 7.24 (5.97 - 8.70) |
| DAG | 2,939 | 8,792 | 2.2 | 8.3 | 0 | 0 | | |
| ODOD | 10,409 | 19,326 | 6.0 | 12.3 | 95 | 401 | 15.8 (12.8 - 19.3) | 32.7 (29.6 - 36.1) |
| MDCR | 1,266 | 4,529 | 0.8 | 3.3 | 11 | 114 | 14.3 (7.14 - 25.6) | 34.2 (28.2 - 41.1) |
| MDCD | 2,226 | 5,994 | 1.0 | 3.2 | 13 | 109 | 13.0 (6.91 - 22.2) | 34.0 (27.9 - 41.0) |
| Open Claims | 226,364 | 430,258 | 149.4 | 297.6 | 976 | 4,352 | 6.53 (6.13 - 6.96) | 14.6 (14.2 - 15.1) |
| OEHR | 14,927 | 33,215 | 4.1 | 14.0 | 26 | 165 | 6.34 (4.14 - 9.30) | 11.8 (10.0 - 13.7) |
| SIDIAP | 2,778 | 14,248 | 3.4 | 24.4 | <5 | 12 | < 1.48 (0.48 - 3.44) | 0.49 (0.25 - 0.86) |
| Meta-analysis | 270,175 | 514,320 | 171.0 | 342.7 | 1,158 | 5,140 | 6.77 (6.39 - 7.17) | 15.0 (14.6 - 15.4) |
| Meta-analysis (w/o USOC) | 43,811 | 84,062 | 21.6 | 45.1 | 182 | 788 | 8.43 (7.25 - 9.75) | 17.5 (16.3 - 18.7) |
| **PS-stratification / on-treatment TAR without escalation** | | | | | | | | |
| CCAE | 18,692 | 25,555 | 11.7 | 15.6 | 53 | 113 | 4.54 (3.40 - 5.94) | 7.23 (5.96 - 8.69) |
| DAG | 3,243 | 8,833 | 2.3 | 8.4 | 0 | 0 | | |
| ODOD | 12,447 | 19,568 | 6.8 | 12.4 | 103 | 413 | 15.2 (12.4 - 18.4) | 33.2 (30.1 - 36.6) |
| MDCR | 1,438 | 4,575 | 0.8 | 3.4 | 12 | 120 | 14.6 (7.53 - 25.5) | 35.8 (29.7 - 42.8) |
| MDCD | 2,313 | 6,013 | 1.0 | 3.2 | 14 | 109 | 13.6 (7.44 - 22.8) | 33.9 (27.8 - 40.9) |
| Open Claims | 269,448 | 430,532 | 168.4 | 297.8 | 1,117 | 4,368 | 6.63 (6.25 - 7.04) | 14.7 (14.2 - 15.1) |
| OEHR | 16,501 | 34,223 | 4.5 | 14.5 | 29 | 173 | 6.44 (4.31 - 9.24) | 12.0 (10.2 - 13.9) |
| SIDIAP | 2,956 | 14,730 | 3.5 | 25.6 | <5 | 13 | < 1.42 (0.46 - 3.31) | 0.51 (0.27 - 0.87) |
| Meta-analysis | 317,088 | 509,878 | 191.3 | 340.3 | 1,302 | 5,067 | 6.81 (6.44 - 7.19) | 14.9 (14.5 - 15.3) |
| Meta-analysis (w/o USOC) | 47,640 | 79,346 | 23.0 | 42.5 | 185 | 699 | 8.06 (6.94 - 9.31) | 16.4 (15.2 - 17.7) |

##### 4.2.5 Glycemic control

**Supplementary Table 316:** Exposures, outcomes and risk estimates for Glycemic control in SGLT2I (female) (T) vs DPP4I (female) (C) new-user cohorts under four different propensity score (PS) adjustment / time-at-risk (TAR) designs across data sources. We report total patient exposure time in 1,000 patient-year (KYr) units, uncalibrated and calibrated hazard ratios (HRs) and their 95% confidence intervals (CIs). Greyed out entries did not pass study diagnostics and are included only for completeness.

| | Patients | | Exposure (KYr) | | Outcomes | | Incidence (per KYr) | |
|---|---|---|---|---|---|---|---|---|
| | T | C | T | C | T | C | T | C |
| **PS-matching / on-treatment TAR** | | | | | | | | |
| CCAE | 16,210 | 25,462 | 12.2 | 19.2 | 22 | 41 | 1.80 (1.13 - 2.73) | 2.14 (1.53 - 2.90) |
| DAG | 1,012 | 3,111 | 0.7 | 2.7 | 95 | 380 | 135 (110 - 166) | 140 (126 - 154) |
| ODOD | 6,625 | 12,653 | 3.9 | 8.5 | 403 | 977 | 102 (92.5 - 113) | 114 (107 - 122) |
| MDCR | 1,294 | 4,793 | 0.9 | 4.1 | 0 | 6 | | 1.47 (0.54 - 3.20) |
| MDCD | 2,343 | 6,366 | 1.3 | 4.5 | 0 | 0 | | |
| Open Claims | 231,830 | 446,127 | 182.9 | 391.2 | 0 | 0 | | |
| OEHR | 15,066 | 33,703 | 4.8 | 17.8 | 0 | 0 | | |
| SIDIAP | 186 | 964 | 0.2 | 1.2 | 52 | 367 | 257 (192 - 337) | 294 (264 - 325) |
| Meta-analysis | 24,033 | 42,190 | 17.0 | 31.7 | 572 | 1,765 | 33.6 (30.9 - 36.4) | 55.7 (53.1 - 58.4) |
| Meta-analysis (w/o USOC) | 24,033 | 42,190 | 17.0 | 31.7 | 572 | 1,765 | 33.6 (30.9 - 36.4) | 55.7 (53.1 - 58.4) |
| **PS-stratification / on-treatment TAR** | | | | | | | | |
| CCAE | 18,683 | 25,497 | 13.5 | 19.2 | 22 | 42 | 1.62 (1.02 - 2.46) | 2.19 (1.58 - 2.96) |
| DAG | 1,073 | 3,152 | 0.7 | 2.8 | 100 | 385 | 136 (111 - 166) | 140 (126 - 155) |
| ODOD | 7,839 | 12,767 | 4.4 | 8.6 | 456 | 984 | 103 (93.4 - 113) | 114 (107 - 121) |
| MDCR | 1,471 | 4,858 | 0.9 | 4.1 | 0 | 6 | | 1.45 (0.53 - 3.16) |
| MDCD | 2,435 | 6,385 | 1.3 | 4.5 | 0 | 0 | | |
| Open Claims | 276,521 | 446,563 | 205.0 | 391.6 | 0 | 0 | | |
| OEHR | 16,666 | 34,746 | 5.2 | 18.4 | 0 | 0 | | |
| SIDIAP | 193 | 1,065 | 0.2 | 1.4 | 55 | 418 | 266 (200 - 346) | 297 (270 - 327) |
| Meta-analysis | 27,788 | 42,481 | 18.9 | 32.0 | 633 | 1,829 | 33.4 (30.9 - 36.2) | 57.2 (54.6 - 59.9) |
| Meta-analysis (w/o USOC) | 27,788 | 42,481 | 18.9 | 32.0 | 633 | 1,829 | 33.4 (30.9 - 36.2) | 57.2 (54.6 - 59.9) |
| **PS-matching / on-treatment TAR without escalation** | | | | | | | | |
| CCAE | 16,210 | 25,462 | 10.5 | 15.6 | 18 | 36 | 1.72 (1.02 - 2.72) | 2.31 (1.62 - 3.20) |
| DAG | 1,012 | 3,111 | 0.6 | 2.5 | 78 | 334 | 121 (96.0 - 152) | 136 (122 - 151) |
| ODOD | 6,625 | 12,653 | 3.5 | 7.2 | 349 | 820 | 100 (90.2 - 112) | 114 (107 - 122) |
| MDCR | 1,294 | 4,793 | 0.8 | 3.6 | 0 | <5 | | < 1.41 (0.46 - 3.28) |
| MDCD | 2,343 | 6,366 | 1.1 | 3.5 | 0 | 0 | | |
| Open Claims | 231,830 | 446,127 | 152.7 | 310.8 | 0 | 0 | | |
| OEHR | 15,066 | 33,703 | 4.1 | 14.3 | 0 | 0 | | |
| SIDIAP | 186 | 964 | 0.2 | 1.0 | 45 | 294 | 256 (187 - 343) | 299 (266 - 335) |
| Meta-analysis | 24,033 | 42,190 | 14.8 | 26.2 | 490 | 1,484 | 33.2 (30.3 - 36.3) | 56.6 (53.8 - 59.6) |
| Meta-analysis (w/o USOC) | 24,033 | 42,190 | 14.8 | 26.2 | 490 | 1,484 | 33.2 (30.3 - 36.3) | 56.6 (53.8 - 59.6) |
| **PS-stratification / on-treatment TAR without escalation** | | | | | | | | |
| CCAE | 18,683 | 25,497 | 11.7 | 15.6 | 18 | 37 | 1.54 (0.91 - 2.44) | 2.37 (1.67 - 3.27) |
| DAG | 1,073 | 3,152 | 0.7 | 2.5 | 82 | 339 | 122 (97.4 - 152) | 136 (122 - 152) |
| ODOD | 7,839 | 12,767 | 3.9 | 7.3 | 397 | 827 | 101 (91.3 - 111) | 114 (106 - 122) |
| MDCR | 1,471 | 4,858 | 0.8 | 3.6 | 0 | <5 | | < 1.39 (0.45 - 3.25) |
| MDCD | 2,435 | 6,385 | 1.1 | 3.5 | 0 | 0 | | |
| Open Claims | 276,521 | 446,563 | 172.3 | 311.2 | 0 | 0 | | |
| OEHR | 16,666 | 34,746 | 4.6 | 14.8 | 0 | 0 | | |
| SIDIAP | 193 | 1,065 | 0.2 | 1.1 | 48 | 338 | 267 (197 - 354) | 311 (279 - 346) |
| Meta-analysis | 27,788 | 42,481 | 16.4 | 26.5 | 545 | 1,541 | 33.1 (30.4 - 36.0) | 58.3 (55.4 - 61.2) |
| Meta-analysis (w/o USOC) | 27,788 | 42,481 | 16.4 | 26.5 | 545 | 1,541 | 33.1 (30.4 - 36.0) | 58.3 (55.4 - 61.2) |

##### 4.2.6 Hospitalization with heart failure

**Supplementary Table 317:** Exposures, outcomes and risk estimates for Hospitalization with heart failure in SGLT2I (female) (T) vs DPP4I (female) (C) new-user cohorts under four different propensity score (PS) adjustment / time-at-risk (TAR) designs across data sources. We report total patient exposure time in 1,000 patient-year (KYr) units, uncalibrated and calibrated hazard ratios (HRs) and their 95% confidence intervals (CIs). Greyed out entries did not pass study diagnostics and are included only for completeness.

| | Patients | | Exposure (KYr) | | Outcomes | | Incidence (per KYr) | |
|---|---|---|---|---|---|---|---|---|
| | T | C | T | C | T | C | T | C |
| **PS-matching / on-treatment TAR** | | | | | | | | |
| CCAE | 16,166 | 25,458 | 12.2 | 19.1 | 45 | 96 | 3.70 (2.70 - 4.95) | 5.02 (4.07 - 6.13) |
| DAG | 2,939 | 8,792 | 2.4 | 9.3 | 0 | 0 | | |
| ODOD | 10,265 | 19,245 | 6.7 | 14.5 | 68 | 360 | 10.1 (7.86 - 12.8) | 24.8 (22.3 - 27.5) |
| MDCR | 1,218 | 4,401 | 0.8 | 3.7 | 13 | 102 | 15.5 (8.26 - 26.5) | 27.2 (22.2 - 33.1) |
| MDCD | 2,184 | 5,838 | 1.2 | 4.0 | 20 | 96 | 16.8 (10.2 - 25.9) | 23.8 (19.3 - 29.1) |
| Open Claims | 224,538 | 427,200 | 177.7 | 372.7 | 839 | 4,237 | 4.72 (4.41 - 5.05) | 11.4 (11.0 - 11.7) |
| OEHR | 14,834 | 32,983 | 4.7 | 17.3 | 36 | 211 | 7.70 (5.39 - 10.7) | 12.2 (10.6 - 14.0) |
| SIDIAP | 2,760 | 14,201 | 3.9 | 30.8 | <5 | 29 | < 1.30 (0.42 - 3.03) | 0.94 (0.63 - 1.35) |
| Meta-analysis | 270,747 | 524,925 | 206.3 | 458.5 | <1,013 | 5,029 | < 4.91 (4.61 - 5.22) | 11.0 (10.7 - 11.3) |
| Meta-analysis (w/o USOC) | 46,209 | 97,725 | 28.6 | 85.8 | <174 | 792 | < 6.08 (5.21 - 7.05) | 9.24 (8.60 - 9.90) |
| **PS-stratification / on-treatment TAR** | | | | | | | | |
| CCAE | 18,562 | 25,492 | 13.5 | 19.2 | 47 | 96 | 3.49 (2.56 - 4.64) | 5.01 (4.06 - 6.12) |
| DAG | 3,243 | 8,833 | 2.5 | 9.3 | 0 | 0 | | |
| ODOD | 12,165 | 19,513 | 7.5 | 14.7 | 82 | 378 | 10.9 (8.67 - 13.5) | 25.6 (23.1 - 28.4) |
| MDCR | 1,384 | 4,436 | 0.9 | 3.8 | 14 | 103 | 15.7 (8.58 - 26.3) | 27.3 (22.3 - 33.1) |
| MDCD | 2,266 | 5,856 | 1.2 | 4.1 | 20 | 99 | 16.3 (9.98 - 25.2) | 24.4 (19.9 - 29.8) |
| Open Claims | 265,019 | 427,498 | 197.8 | 373.0 | 960 | 4,257 | 4.85 (4.55 - 5.17) | 11.4 (11.1 - 11.8) |
| OEHR | 16,387 | 34,086 | 5.1 | 17.9 | 36 | 225 | 7.00 (4.90 - 9.69) | 12.6 (11.0 - 14.4) |
| SIDIAP | 2,929 | 14,683 | 4.0 | 32.3 | <5 | 33 | < 1.25 (0.41 - 2.92) | 1.02 (0.70 - 1.44) |
| Meta-analysis | 315,062 | 521,272 | 227.9 | 457.0 | <1,130 | 4,989 | < 4.96 (4.67 - 5.26) | 10.9 (10.6 - 11.2) |
| Meta-analysis (w/o USOC) | 50,043 | 93,774 | 30.1 | 84.0 | <170 | 732 | < 5.64 (4.83 - 6.56) | 8.71 (8.09 - 9.37) |
| **PS-matching / on-treatment TAR without escalation** | | | | | | | | |
| CCAE | 16,166 | 25,458 | 10.5 | 15.6 | 40 | 84 | 3.83 (2.73 - 5.21) | 5.39 (4.30 - 6.67) |
| DAG | 2,939 | 8,792 | 2.2 | 8.3 | 0 | 0 | | |
| ODOD | 10,265 | 19,245 | 5.9 | 12.2 | 57 | 300 | 9.58 (7.26 - 12.4) | 24.6 (21.9 - 27.5) |
| MDCR | 1,218 | 4,401 | 0.7 | 3.3 | 12 | 84 | 16.2 (8.39 - 28.4) | 25.7 (20.5 - 31.8) |
| MDCD | 2,184 | 5,838 | 1.0 | 3.1 | 14 | 73 | 14.2 (7.77 - 23.9) | 23.2 (18.2 - 29.2) |
| Open Claims | 224,538 | 427,200 | 148.4 | 296.4 | 683 | 3,393 | 4.60 (4.26 - 4.96) | 11.4 (11.1 - 11.8) |
| OEHR | 14,834 | 32,983 | 4.1 | 13.9 | 29 | 148 | 7.13 (4.77 - 10.2) | 10.6 (8.97 - 12.5) |
| SIDIAP | 2,760 | 14,201 | 3.4 | 24.3 | <5 | 23 | < 1.48 (0.48 - 3.45) | 0.95 (0.60 - 1.42) |
| Meta-analysis | 267,987 | 510,724 | 169.9 | 341.3 | 823 | 3,998 | 4.84 (4.52 - 5.19) | 11.7 (11.4 - 12.1) |
| Meta-analysis (w/o USOC) | 43,449 | 83,524 | 21.5 | 44.9 | 140 | 605 | 6.52 (5.49 - 7.70) | 13.5 (12.4 - 14.6) |
| **PS-stratification / on-treatment TAR without escalation** | | | | | | | | |
| CCAE | 18,562 | 25,492 | 11.6 | 15.6 | 42 | 84 | 3.61 (2.60 - 4.89) | 5.38 (4.29 - 6.66) |
| DAG | 3,243 | 8,833 | 2.3 | 8.4 | 0 | 0 | | |
| ODOD | 12,165 | 19,513 | 6.7 | 12.4 | 70 | 317 | 10.5 (8.17 - 13.2) | 25.6 (22.8 - 28.5) |
| MDCR | 1,384 | 4,436 | 0.8 | 3.3 | 13 | 85 | 16.5 (8.78 - 28.2) | 25.9 (20.7 - 32.0) |
| MDCD | 2,266 | 5,856 | 1.0 | 3.2 | 14 | 73 | 13.8 (7.57 - 23.2) | 23.1 (18.1 - 29.0) |
| Open Claims | 265,019 | 427,498 | 166.2 | 296.7 | 794 | 3,410 | 4.78 (4.45 - 5.12) | 11.5 (11.1 - 11.9) |
| OEHR | 16,387 | 34,086 | 4.5 | 14.4 | 29 | 159 | 6.48 (4.34 - 9.31) | 11.0 (9.39 - 12.9) |
| SIDIAP | 2,929 | 14,683 | 3.5 | 25.5 | <5 | 25 | < 1.42 (0.46 - 3.32) | 0.98 (0.63 - 1.45) |
| Meta-analysis | 315,062 | 521,272 | 192.5 | 364.6 | <940 | 3,995 | < 4.88 (4.58 - 5.21) | 11.0 (10.6 - 11.3) |
| Meta-analysis (w/o USOC) | 50,043 | 93,774 | 26.3 | 68.0 | <146 | 585 | < 5.55 (4.69 - 6.53) | 8.61 (7.93 - 9.34) |

##### 4.2.7 Measured renal dysfunction

**Supplementary Table 318:** Exposures, outcomes and risk estimates for Measured renal dysfunction in SGLT2I (female) (T) vs DPP4I (female) (C) new-user cohorts under four different propensity score (PS) adjustment / time-at-risk (TAR) designs across data sources. We report total patient exposure time in 1,000 patient-year (KYr) units, uncalibrated and calibrated hazard ratios (HRs) and their 95% confidence intervals (CIs). Greyed out entries did not pass study diagnostics and are included only for completeness.

| | Patients | | Exposure (KYr) | | Outcomes | | Incidence (per KYr) | |
|---|---|---|---|---|---|---|---|---|
| | T | C | T | C | T | C | T | C |
| **PS-matching / on-treatment TAR** | | | | | | | | |
| CCAE | 16,380 | 25,894 | 12.3 | 19.5 | <5 | 7 | < 0.41 (0.13 - 0.95) | 0.36 (0.14 - 0.74) |
| DAG | 2,939 | 8,792 | 2.4 | 9.3 | 0 | 0 | | |
| ODOD | 10,719 | 20,598 | 7.0 | 15.7 | 10 | 41 | 1.43 (0.69 - 2.63) | 2.61 (1.87 - 3.54) |
| MDCR | 1,303 | 4,842 | 0.9 | 4.1 | 0 | 0 | | |
| MDCD | 2,343 | 6,366 | 1.3 | 4.5 | 0 | 0 | | |
| Open Claims | 231,830 | 446,127 | 182.9 | 391.2 | 0 | 0 | | |
| OEHR | 15,066 | 33,703 | 4.8 | 17.8 | 0 | 0 | | |
| SIDIAP | 2,772 | 14,209 | 3.9 | 30.7 | 0 | 95 | | 3.09 (2.50 - 3.78) |
| Meta-analysis | 10,719 | 20,598 | 7.0 | 15.7 | 10 | 41 | 1.43 (0.69 - 2.63) | 2.61 (1.87 - 3.54) |
| Meta-analysis (w/o USOC) | 10,719 | 20,598 | 7.0 | 15.7 | 10 | 41 | 1.43 (0.69 - 2.63) | 2.61 (1.87 - 3.54) |
| **PS-stratification / on-treatment TAR** | | | | | | | | |
| CCAE | 18,849 | 25,930 | 13.7 | 19.5 | <5 | 7 | < 0.37 (0.12 - 0.85) | 0.36 (0.14 - 0.74) |
| DAG | 3,243 | 8,833 | 2.5 | 9.3 | 0 | 0 | | |
| ODOD | 12,834 | 20,806 | 7.9 | 15.9 | 10 | 46 | 1.27 (0.61 - 2.34) | 2.90 (2.12 - 3.87) |
| MDCR | 1,478 | 4,909 | 0.9 | 4.2 | 0 | 0 | | |
| MDCD | 2,435 | 6,385 | 1.3 | 4.5 | 0 | 0 | | |
| Open Claims | 276,521 | 446,563 | 205.0 | 391.6 | 0 | 0 | | |
| OEHR | 16,666 | 34,746 | 5.2 | 18.4 | 0 | 0 | | |
| SIDIAP | 2,949 | 14,687 | 4.0 | 32.2 | 0 | 107 | | 3.32 (2.72 - 4.02) |
| Meta-analysis | 12,834 | 20,806 | 7.9 | 15.9 | 10 | 46 | 1.27 (0.61 - 2.34) | 2.90 (2.12 - 3.87) |
| Meta-analysis (w/o USOC) | 12,834 | 20,806 | 7.9 | 15.9 | 10 | 46 | 1.27 (0.61 - 2.34) | 2.90 (2.12 - 3.87) |
| **PS-matching / on-treatment TAR without escalation** | | | | | | | | |
| CCAE | 16,380 | 25,894 | 10.6 | 15.9 | <5 | 6 | < 0.47 (0.15 - 1.10) | 0.38 (0.14 - 0.82) |
| DAG | 2,939 | 8,792 | 2.2 | 8.3 | 0 | 0 | | |
| ODOD | 10,719 | 20,598 | 6.2 | 13.2 | 8 | 33 | 1.30 (0.56 - 2.56) | 2.50 (1.72 - 3.51) |
| MDCR | 1,303 | 4,842 | 0.8 | 3.6 | 0 | 0 | | |
| MDCD | 2,343 | 6,366 | 1.1 | 3.5 | 0 | 0 | | |
| Open Claims | 231,830 | 446,127 | 152.7 | 310.8 | 0 | 0 | | |
| OEHR | 15,066 | 33,703 | 4.1 | 14.3 | 0 | 0 | | |
| SIDIAP | 2,772 | 14,209 | 3.4 | 24.3 | 0 | 80 | | 3.29 (2.61 - 4.10) |
| Meta-analysis | 10,719 | 20,598 | 6.2 | 13.2 | 8 | 33 | 1.30 (0.56 - 2.56) | 2.50 (1.72 - 3.51) |
| Meta-analysis (w/o USOC) | 10,719 | 20,598 | 6.2 | 13.2 | 8 | 33 | 1.30 (0.56 - 2.56) | 2.50 (1.72 - 3.51) |
| **PS-stratification / on-treatment TAR without escalation** | | | | | | | | |
| CCAE | 18,849 | 25,930 | 11.8 | 15.9 | <5 | 6 | < 0.42 (0.14 - 0.99) | 0.38 (0.14 - 0.82) |
| DAG | 3,243 | 8,833 | 2.3 | 8.4 | 0 | 0 | | |
| ODOD | 12,834 | 20,806 | 7.0 | 13.3 | 8 | 38 | 1.15 (0.50 - 2.26) | 2.85 (2.01 - 3.91) |
| MDCR | 1,478 | 4,909 | 0.8 | 3.6 | 0 | 0 | | |
| MDCD | 2,435 | 6,385 | 1.1 | 3.5 | 0 | 0 | | |
| Open Claims | 276,521 | 446,563 | 172.3 | 311.2 | 0 | 0 | | |
| OEHR | 16,666 | 34,746 | 4.6 | 14.8 | 0 | 0 | | |
| SIDIAP | 2,949 | 14,687 | 3.5 | 25.5 | 0 | 89 | | 3.49 (2.81 - 4.30) |
| Meta-analysis | 12,834 | 20,806 | 7.0 | 13.3 | 8 | 38 | 1.15 (0.50 - 2.26) | 2.85 (2.01 - 3.91) |
| Meta-analysis (w/o USOC) | 12,834 | 20,806 | 7.0 | 13.3 | 8 | 38 | 1.15 (0.50 - 2.26) | 2.85 (2.01 - 3.91) |

##### 4.2.8 Stroke

**Supplementary Table 319:** Exposures, outcomes and risk estimates for Stroke in SGLT2I (female) (T) vs DPP4I (female) (C) new-user cohorts under four different propensity score (PS) adjustment / time-at-risk (TAR) designs across data sources. We report total patient exposure time in 1,000 patient-year (KYr) units, uncalibrated and calibrated hazard ratios (HRs) and their 95% confidence intervals (CIs). Greyed out entries did not pass study diagnostics and are included only for completeness.

| | Patients | | Exposure (KYr) | | Outcomes | | Incidence (per KYr) | |
|---|---|---|---|---|---|---|---|---|
| | T | C | T | C | T | C | T | C |
| **PS-matching / on-treatment TAR** | | | | | | | | |
| CCAE | 16,223 | 25,555 | 12.2 | 19.2 | 44 | 69 | 3.61 (2.62 - 4.84) | 3.59 (2.79 - 4.54) |
| DAG | 2,939 | 8,792 | 2.4 | 9.3 | 0 | 0 | | |
| ODOD | 10,508 | 19,921 | 6.8 | 15.2 | 50 | 198 | 7.31 (5.43 - 9.64) | 13.1 (11.3 - 15.0) |
| MDCR | 1,273 | 4,612 | 0.9 | 3.9 | <5 | 63 | < 5.75 (1.87 - 13.4) | 16.2 (12.4 - 20.7) |
| MDCD | 2,255 | 6,115 | 1.2 | 4.3 | 12 | 61 | 9.77 (5.05 - 17.1) | 14.3 (10.9 - 18.4) |
| Open Claims | 226,598 | 433,211 | 179.1 | 379.0 | 562 | 2,320 | 3.14 (2.88 - 3.41) | 6.12 (5.87 - 6.38) |
| OEHR | 14,961 | 33,293 | 4.7 | 17.6 | 13 | 74 | 2.75 (1.46 - 4.70) | 4.21 (3.31 - 5.29) |
| SIDIAP | 2,762 | 14,143 | 3.8 | 30.6 | 10 | 72 | 2.61 (1.25 - 4.81) | 2.35 (1.84 - 2.96) |
| Meta-analysis | 273,307 | 532,238 | 207.9 | 465.8 | 691 | 2,794 | 3.32 (3.08 - 3.58) | 6.00 (5.78 - 6.22) |
| Meta-analysis (w/o USOC) | 46,709 | 99,027 | 28.8 | 86.8 | 129 | 474 | 4.47 (3.74 - 5.32) | 5.46 (4.98 - 5.98) |
| **PS-stratification / on-treatment TAR** | | | | | | | | |
| CCAE | 18,673 | 25,589 | 13.5 | 19.3 | 51 | 69 | 3.77 (2.81 - 4.96) | 3.58 (2.79 - 4.54) |
| DAG | 3,243 | 8,833 | 2.5 | 9.3 | 0 | 0 | | |
| ODOD | 12,563 | 20,120 | 7.7 | 15.3 | 53 | 206 | 6.89 (5.16 - 9.01) | 13.4 (11.7 - 15.4) |
| MDCR | 1,442 | 4,659 | 0.9 | 3.9 | 5 | 64 | 5.40 (1.75 - 12.6) | 16.3 (12.5 - 20.8) |
| MDCD | 2,344 | 6,133 | 1.3 | 4.3 | 12 | 62 | 9.56 (4.94 - 16.7) | 14.5 (11.1 - 18.6) |
| Open Claims | 270,188 | 433,555 | 200.6 | 379.4 | 602 | 2,328 | 3.00 (2.77 - 3.25) | 6.14 (5.89 - 6.39) |
| OEHR | 16,552 | 34,421 | 5.2 | 18.1 | 14 | 79 | 2.69 (1.47 - 4.52) | 4.35 (3.45 - 5.43) |
| SIDIAP | 2,932 | 14,618 | 4.0 | 32.1 | 10 | 77 | 2.51 (1.20 - 4.61) | 2.40 (1.89 - 3.00) |
| Meta-analysis | 320,908 | 528,303 | 231.0 | 464.1 | 730 | 2,759 | 3.16 (2.93 - 3.40) | 5.94 (5.72 - 6.17) |
| Meta-analysis (w/o USOC) | 50,720 | 94,748 | 30.4 | 84.8 | 128 | 431 | 4.21 (3.51 - 5.01) | 5.08 (4.61 - 5.59) |
| **PS-matching / on-treatment TAR without escalation** | | | | | | | | |
| CCAE | 16,223 | 25,555 | 10.5 | 15.7 | 35 | 60 | 3.34 (2.33 - 4.64) | 3.83 (2.92 - 4.93) |
| DAG | 2,939 | 8,792 | 2.2 | 8.3 | 0 | 0 | | |
| ODOD | 10,508 | 19,921 | 6.0 | 12.7 | 47 | 177 | 7.78 (5.72 - 10.3) | 13.9 (11.9 - 16.1) |
| MDCR | 1,273 | 4,612 | 0.8 | 3.4 | <5 | 48 | < 6.48 (2.10 - 15.1) | 14.1 (10.4 - 18.7) |
| MDCD | 2,255 | 6,115 | 1.0 | 3.3 | 6 | 52 | 5.89 (2.16 - 12.8) | 15.7 (11.7 - 20.5) |
| Open Claims | 226,598 | 433,211 | 149.5 | 301.2 | 470 | 1,875 | 3.14 (2.87 - 3.44) | 6.22 (5.95 - 6.51) |
| OEHR | 14,961 | 33,293 | 4.1 | 14.1 | 9 | 53 | 2.19 (1.00 - 4.15) | 3.75 (2.81 - 4.91) |
| SIDIAP | 2,762 | 14,143 | 3.4 | 24.2 | 10 | 61 | 2.98 (1.43 - 5.49) | 2.52 (1.93 - 3.24) |
| Meta-analysis | 273,307 | 532,238 | 174.5 | 371.3 | 577 | 2,278 | 3.31 (3.04 - 3.59) | 6.14 (5.89 - 6.39) |
| Meta-analysis (w/o USOC) | 46,709 | 99,027 | 25.0 | 70.0 | 107 | 403 | 4.28 (3.51 - 5.17) | 5.75 (5.21 - 6.34) |
| **PS-stratification / on-treatment TAR without escalation** | | | | | | | | |
| CCAE | 18,673 | 25,589 | 11.7 | 15.7 | 41 | 60 | 3.51 (2.52 - 4.77) | 3.82 (2.92 - 4.92) |
| DAG | 3,243 | 8,833 | 2.3 | 8.4 | 0 | 0 | | |
| ODOD | 12,563 | 20,120 | 6.8 | 12.9 | 50 | 185 | 7.33 (5.44 - 9.66) | 14.3 (12.3 - 16.6) |
| MDCR | 1,442 | 4,659 | 0.8 | 3.4 | <5 | 48 | < 6.07 (1.97 - 14.2) | 14.0 (10.3 - 18.5) |
| MDCD | 2,344 | 6,133 | 1.0 | 3.3 | 6 | 52 | 5.75 (2.11 - 12.5) | 15.6 (11.7 - 20.5) |
| Open Claims | 270,188 | 433,555 | 168.7 | 301.5 | 505 | 1,883 | 2.99 (2.74 - 3.27) | 6.24 (5.97 - 6.53) |
| OEHR | 16,552 | 34,421 | 4.5 | 14.6 | 9 | 56 | 1.99 (0.91 - 3.78) | 3.84 (2.90 - 4.99) |
| SIDIAP | 2,932 | 14,618 | 3.5 | 25.4 | 10 | 63 | 2.86 (1.37 - 5.27) | 2.48 (1.91 - 3.18) |
| Meta-analysis | 320,908 | 528,303 | 195.2 | 370.1 | 615 | 2,247 | 3.15 (2.91 - 3.41) | 6.07 (5.82 - 6.33) |
| Meta-analysis (w/o USOC) | 50,720 | 94,748 | 26.5 | 68.6 | 110 | 364 | 4.15 (3.41 - 5.00) | 5.31 (4.78 - 5.88) |

##### 4.2.9 Sudden cardiac death

**Supplementary Table 320:** Exposures, outcomes and risk estimates for Sudden cardiac death in SGLT2I (female) (T) vs DPP4I (female) (C) new-user cohorts under four different propensity score (PS) adjustment / time-at-risk (TAR) designs across data sources. We report total patient exposure time in 1,000 patient-year (KYr) units, uncalibrated and calibrated hazard ratios (HRs) and their 95% confidence intervals (CIs). Greyed out entries did not pass study diagnostics and are included only for completeness.

| | Patients | | Exposure (KYr) | | Outcomes | | Incidence (per KYr) | |
|---|---|---|---|---|---|---|---|---|
| | T | C | T | C | T | C | T | C |
| **PS-matching / on-treatment TAR** | | | | | | | | |
| CCAE | 16,402 | 25,916 | 12.3 | 19.5 | 7 | 11 | 0.57 (0.23 - 1.17) | 0.56 (0.28 - 1.01) |
| DAG | 2,939 | 8,792 | 2.4 | 9.3 | 0 | 0 | | |
| ODOD | 10,719 | 20,603 | 7.0 | 15.7 | 5 | 54 | 0.72 (0.23 - 1.67) | 3.43 (2.58 - 4.47) |
| MDCR | 1,303 | 4,830 | 0.9 | 4.1 | 0 | 5 | | 1.22 (0.40 - 2.84) |
| MDCD | 2,341 | 6,354 | 1.3 | 4.4 | <5 | 14 | < 3.91 (1.27 - 9.13) | 3.15 (1.72 - 5.28) |
| Open Claims | 231,530 | 445,456 | 182.6 | 390.5 | 102 | 475 | 0.56 (0.46 - 0.68) | 1.22 (1.11 - 1.33) |
| OEHR | 15,059 | 33,690 | 4.8 | 17.8 | <5 | 17 | < 1.05 (0.34 - 2.45) | 0.95 (0.56 - 1.53) |
| SIDIAP | 2,778 | 14,273 | 3.9 | 30.9 | 0 | 0 | | |
| Meta-analysis | 273,710 | 525,665 | 206.7 | 443.5 | <119 | 557 | < 0.58 (0.48 - 0.69) | 1.26 (1.15 - 1.36) |
| Meta-analysis (w/o USOC) | 42,180 | 80,209 | 24.1 | 53.1 | <17 | 82 | < 0.71 (0.41 - 1.13) | 1.55 (1.23 - 1.92) |
| **PS-stratification / on-treatment TAR** | | | | | | | | |
| CCAE | 18,867 | 25,952 | 13.7 | 19.5 | 8 | 11 | 0.58 (0.25 - 1.15) | 0.56 (0.28 - 1.01) |
| DAG | 3,243 | 8,833 | 2.5 | 9.3 | 0 | 0 | | |
| ODOD | 12,828 | 20,812 | 7.9 | 15.9 | 6 | 56 | 0.76 (0.28 - 1.66) | 3.52 (2.66 - 4.57) |
| MDCR | 1,476 | 4,897 | 0.9 | 4.2 | 0 | 5 | | 1.20 (0.39 - 2.81) |
| MDCD | 2,433 | 6,373 | 1.3 | 4.5 | <5 | 17 | < 3.83 (1.24 - 8.93) | 3.80 (2.21 - 6.08) |
| Open Claims | 276,028 | 445,883 | 204.6 | 390.9 | 121 | 476 | 0.59 (0.49 - 0.71) | 1.22 (1.11 - 1.33) |
| OEHR | 16,657 | 34,732 | 5.2 | 18.4 | <5 | 19 | < 0.96 (0.31 - 2.23) | 1.03 (0.62 - 1.62) |
| SIDIAP | 2,955 | 14,757 | 4.0 | 32.4 | 0 | 0 | | |
| Meta-analysis | 324,380 | 527,379 | 231.4 | 444.7 | <140 | 562 | < 0.61 (0.51 - 0.71) | 1.26 (1.16 - 1.37) |
| Meta-analysis (w/o USOC) | 48,352 | 81,496 | 26.8 | 53.8 | <19 | 86 | < 0.71 (0.43 - 1.11) | 1.60 (1.28 - 1.97) |
| **PS-matching / on-treatment TAR without escalation** | | | | | | | | |
| CCAE | 16,402 | 25,916 | 10.6 | 15.9 | 6 | 7 | 0.57 (0.21 - 1.23) | 0.44 (0.18 - 0.91) |
| DAG | 2,939 | 8,792 | 2.2 | 8.3 | 0 | 0 | | |
| ODOD | 10,719 | 20,603 | 6.2 | 13.2 | 5 | 42 | 0.81 (0.26 - 1.89) | 3.17 (2.29 - 4.29) |
| MDCR | 1,303 | 4,830 | 0.8 | 3.6 | 0 | 5 | | 1.40 (0.45 - 3.26) |
| MDCD | 2,341 | 6,354 | 1.1 | 3.5 | <5 | 8 | < 4.75 (1.54 - 11.1) | 2.31 (1.00 - 4.56) |
| Open Claims | 231,530 | 445,456 | 152.4 | 310.3 | 88 | 387 | 0.58 (0.46 - 0.71) | 1.25 (1.13 - 1.38) |
| OEHR | 15,059 | 33,690 | 4.1 | 14.3 | <5 | 10 | < 1.21 (0.39 - 2.82) | 0.70 (0.34 - 1.29) |
| SIDIAP | 2,778 | 14,273 | 3.4 | 24.4 | 0 | 0 | | |
| Meta-analysis | 242,249 | 466,059 | 158.6 | 323.5 | 93 | 429 | 0.59 (0.47 - 0.72) | 1.33 (1.20 - 1.46) |
| Meta-analysis (w/o USOC) | 10,719 | 20,603 | 6.2 | 13.2 | 5 | 42 | 0.81 (0.26 - 1.89) | 3.17 (2.29 - 4.29) |
| **PS-stratification / on-treatment TAR without escalation** | | | | | | | | |
| CCAE | 18,867 | 25,952 | 11.8 | 15.9 | 7 | 7 | 0.59 (0.24 - 1.22) | 0.44 (0.18 - 0.91) |
| DAG | 3,243 | 8,833 | 2.3 | 8.4 | 0 | 0 | | |
| ODOD | 12,828 | 20,812 | 7.0 | 13.4 | 6 | 44 | 0.86 (0.32 - 1.87) | 3.28 (2.39 - 4.41) |
| MDCR | 1,476 | 4,897 | 0.8 | 3.6 | 0 | 5 | | 1.38 (0.45 - 3.23) |
| MDCD | 2,433 | 6,373 | 1.1 | 3.5 | <5 | 10 | < 4.63 (1.50 - 10.8) | 2.88 (1.38 - 5.30) |
| Open Claims | 276,028 | 445,883 | 172.0 | 310.7 | 105 | 388 | 0.61 (0.50 - 0.74) | 1.25 (1.13 - 1.38) |
| OEHR | 16,657 | 34,732 | 4.5 | 14.7 | <5 | 10 | < 1.10 (0.36 - 2.56) | 0.68 (0.33 - 1.25) |
| SIDIAP | 2,955 | 14,757 | 3.5 | 25.6 | 0 | 0 | | |
| Meta-analysis | 288,856 | 466,695 | 179.0 | 324.0 | 111 | 432 | 0.62 (0.51 - 0.75) | 1.33 (1.21 - 1.46) |
| Meta-analysis (w/o USOC) | 12,828 | 20,812 | 7.0 | 13.4 | 6 | 44 | 0.86 (0.32 - 1.87) | 3.28 (2.39 - 4.41) |

##### 4.2.10 Abnormal weight gain

**Supplementary Table 321:** Exposures, outcomes and risk estimates for Abnormal weight gain in SGLT2I (female) (T) vs DPP4I (female) (C) new-user cohorts under four different propensity score (PS) adjustment / time-at-risk (TAR) designs across data sources. We report total patient exposure time in 1,000 patient-year (KYr) units, uncalibrated and calibrated hazard ratios (HRs) and their 95% confidence intervals (CIs). Greyed out entries did not pass study diagnostics and are included only for completeness.

| | Patients | | Exposure (KYr) | | Outcomes | | Incidence (per KYr) | |
|---|---|---|---|---|---|---|---|---|
| | T | C | T | C | T | C | T | C |
| **PS-matching / on-treatment TAR** | | | | | | | | |
| CCAE | 14,880 | 23,938 | 11.2 | 18.0 | 74 | 144 | 6.58 (5.17 - 8.26) | 8.01 (6.75 - 9.43) |
| DAG | 2,894 | 8,701 | 2.3 | 9.2 | <5 | 13 | < 2.14 (0.70 - 5.00) | 1.42 (0.75 - 2.42) |
| ODOD | 9,959 | 19,506 | 6.4 | 14.8 | 45 | 114 | 7.04 (5.13 - 9.42) | 7.68 (6.33 - 9.22) |
| MDCR | 1,274 | 4,750 | 0.9 | 4.0 | <5 | 14 | < 5.79 (1.88 - 13.5) | 3.47 (1.90 - 5.82) |
| MDCD | 2,097 | 5,787 | 1.1 | 4.0 | 5 | 41 | 4.35 (1.41 - 10.2) | 10.2 (7.31 - 13.8) |
| Open Claims | 215,276 | 422,234 | 169.3 | 370.3 | 427 | 1,112 | 2.52 (2.29 - 2.77) | 3.00 (2.83 - 3.18) |
| OEHR | 14,026 | 31,802 | 4.4 | 16.7 | 25 | 145 | 5.67 (3.67 - 8.38) | 8.70 (7.34 - 10.2) |
| SIDIAP | 2,750 | 14,120 | 3.8 | 30.5 | <5 | 28 | < 1.31 (0.43 - 3.05) | 0.92 (0.61 - 1.32) |
| Meta-analysis | 258,988 | 517,387 | 196.3 | 454.4 | <581 | 1,584 | < 2.96 (2.72 - 3.21) | 3.49 (3.32 - 3.66) |
| Meta-analysis (w/o USOC) | 43,712 | 95,153 | 27.0 | 84.1 | <154 | 472 | < 5.70 (4.84 - 6.68) | 5.61 (5.12 - 6.14) |
| **PS-stratification / on-treatment TAR** | | | | | | | | |
| CCAE | 17,049 | 23,974 | 12.4 | 18.0 | 80 | 145 | 6.46 (5.12 - 8.04) | 8.05 (6.79 - 9.47) |
| DAG | 3,193 | 8,742 | 2.5 | 9.2 | <5 | 13 | < 2.01 (0.65 - 4.70) | 1.41 (0.75 - 2.41) |
| ODOD | 11,835 | 19,719 | 7.2 | 15.0 | 54 | 114 | 7.54 (5.66 - 9.84) | 7.59 (6.26 - 9.11) |
| MDCR | 1,444 | 4,816 | 0.9 | 4.1 | <5 | 14 | < 5.43 (1.76 - 12.7) | 3.43 (1.87 - 5.75) |
| MDCD | 2,177 | 5,803 | 1.2 | 4.0 | 5 | 41 | 4.26 (1.38 - 9.93) | 10.1 (7.28 - 13.8) |
| Open Claims | 255,345 | 422,689 | 188.9 | 370.8 | 486 | 1,116 | 2.57 (2.35 - 2.81) | 3.01 (2.84 - 3.19) |
| OEHR | 15,455 | 32,773 | 4.8 | 17.2 | 29 | 150 | 6.02 (4.03 - 8.64) | 8.74 (7.39 - 10.3) |
| SIDIAP | 2,925 | 14,600 | 4.0 | 32.0 | <5 | 29 | < 1.26 (0.41 - 2.94) | 0.91 (0.61 - 1.30) |
| Meta-analysis | 302,609 | 513,755 | 217.3 | 453.0 | <654 | 1,554 | < 3.01 (2.78 - 3.25) | 3.43 (3.26 - 3.61) |
| Meta-analysis (w/o USOC) | 47,264 | 91,066 | 28.3 | 82.2 | <168 | 438 | < 5.93 (5.06 - 6.89) | 5.33 (4.84 - 5.85) |
| **PS-matching / on-treatment TAR without escalation** | | | | | | | | |
| CCAE | 14,880 | 23,938 | 9.6 | 14.7 | 63 | 115 | 6.54 (5.02 - 8.37) | 7.84 (6.47 - 9.41) |
| DAG | 2,894 | 8,701 | 2.1 | 8.3 | <5 | 10 | < 2.34 (0.76 - 5.47) | 1.21 (0.58 - 2.23) |
| ODOD | 9,959 | 19,506 | 5.7 | 12.5 | 39 | 92 | 6.89 (4.90 - 9.41) | 7.35 (5.93 - 9.02) |
| MDCR | 1,274 | 4,750 | 0.8 | 3.5 | <5 | 13 | < 6.55 (2.13 - 15.3) | 3.70 (1.97 - 6.32) |
| MDCD | 2,097 | 5,787 | 0.9 | 3.1 | <5 | 33 | < 5.29 (1.72 - 12.4) | 10.6 (7.31 - 14.9) |
| Open Claims | 215,276 | 422,234 | 141.5 | 294.8 | 339 | 846 | 2.40 (2.15 - 2.66) | 2.87 (2.68 - 3.07) |
| OEHR | 14,026 | 31,802 | 3.8 | 13.4 | 20 | 98 | 5.20 (3.18 - 8.03) | 7.31 (5.93 - 8.90) |
| SIDIAP | 2,750 | 14,120 | 3.3 | 24.2 | <5 | 23 | < 1.49 (0.48 - 3.48) | 0.95 (0.60 - 1.43) |
| Meta-analysis | 256,238 | 503,267 | 161.6 | 338.5 | <466 | 1,184 | < 2.88 (2.63 - 3.16) | 3.50 (3.30 - 3.70) |
| Meta-analysis (w/o USOC) | 40,962 | 81,033 | 20.1 | 43.7 | <127 | 338 | < 6.32 (5.27 - 7.52) | 7.73 (6.93 - 8.60) |
| **PS-stratification / on-treatment TAR without escalation** | | | | | | | | |
| CCAE | 17,049 | 23,974 | 10.7 | 14.7 | 68 | 115 | 6.38 (4.96 - 8.09) | 7.82 (6.46 - 9.39) |
| DAG | 3,193 | 8,742 | 2.3 | 8.3 | <5 | 10 | < 2.20 (0.72 - 5.14) | 1.20 (0.58 - 2.22) |
| ODOD | 11,835 | 19,719 | 6.4 | 12.7 | 48 | 92 | 7.54 (5.56 - 10.00) | 7.26 (5.85 - 8.91) |
| MDCR | 1,444 | 4,816 | 0.8 | 3.6 | <5 | 13 | < 6.12 (1.99 - 14.3) | 3.66 (1.95 - 6.26) |
| MDCD | 2,177 | 5,803 | 1.0 | 3.1 | <5 | 33 | < 5.17 (1.68 - 12.1) | 10.6 (7.29 - 14.9) |
| Open Claims | 255,345 | 422,689 | 159.0 | 295.2 | 388 | 850 | 2.44 (2.20 - 2.70) | 2.88 (2.69 - 3.08) |
| OEHR | 15,455 | 32,773 | 4.2 | 13.8 | 24 | 101 | 5.71 (3.66 - 8.49) | 7.31 (5.95 - 8.88) |
| SIDIAP | 2,925 | 14,600 | 3.5 | 25.4 | <5 | 24 | < 1.43 (0.47 - 3.35) | 0.95 (0.61 - 1.41) |
| Meta-analysis | 299,684 | 499,155 | 180.2 | 336.4 | 528 | 1,158 | 2.93 (2.69 - 3.19) | 3.44 (3.25 - 3.65) |
| Meta-analysis (w/o USOC) | 44,339 | 76,466 | 21.2 | 41.2 | 140 | 308 | 6.59 (5.55 - 7.78) | 7.48 (6.67 - 8.36) |

##### 4.2.11 Abnormal weight loss

**Supplementary Table 322:** Exposures, outcomes and risk estimates for Abnormal weight loss in SGLT2I (female) (T) vs DPP4I (female) (C) new-user cohorts under four different propensity score (PS) adjustment / time-at-risk (TAR) designs across data sources. We report total patient exposure time in 1,000 patient-year (KYr) units, uncalibrated and calibrated hazard ratios (HRs) and their 95% confidence intervals (CIs). Greyed out entries did not pass study diagnostics and are included only for completeness.

| | Patients | | Exposure (KYr) | | Outcomes | | Incidence (per KYr) | |
|---|---|---|---|---|---|---|---|---|
| | T | C | T | C | T | C | T | C |
| **PS-matching / on-treatment TAR** | | | | | | | | |
| CCAE | 16,030 | 25,270 | 12.0 | 18.9 | 104 | 154 | 8.67 (7.09 - 10.5) | 8.15 (6.91 - 9.54) |
| DAG | 2,860 | 8,535 | 2.3 | 8.9 | 21 | 70 | 9.09 (5.63 - 13.9) | 7.84 (6.11 - 9.90) |
| ODOD | 10,339 | 19,502 | 6.7 | 14.8 | 99 | 223 | 14.8 (12.0 - 18.0) | 15.1 (13.1 - 17.2) |
| MDCR | 1,247 | 4,569 | 0.9 | 3.9 | 9 | 66 | 10.5 (4.81 - 19.9) | 17.1 (13.3 - 21.8) |
| MDCD | 2,245 | 6,043 | 1.2 | 4.2 | 24 | 76 | 19.8 (12.7 - 29.4) | 18.2 (14.3 - 22.7) |
| Open Claims | 223,805 | 427,915 | 176.2 | 373.0 | 956 | 2,614 | 5.43 (5.09 - 5.78) | 7.01 (6.74 - 7.28) |
| OEHR | 14,682 | 32,695 | 4.6 | 17.1 | 42 | 195 | 9.10 (6.56 - 12.3) | 11.4 (9.84 - 13.1) |
| SIDIAP | 2,749 | 14,055 | 3.8 | 30.3 | 17 | 131 | 4.45 (2.59 - 7.12) | 4.32 (3.61 - 5.13) |
| Meta-analysis | 272,710 | 534,015 | 206.8 | 467.3 | 1,263 | 3,463 | 6.11 (5.77 - 6.45) | 7.41 (7.17 - 7.66) |
| Meta-analysis (w/o USOC) | 48,905 | 106,100 | 30.6 | 94.3 | 307 | 849 | 10.0 (8.93 - 11.2) | 9.01 (8.41 - 9.63) |
| **PS-stratification / on-treatment TAR** | | | | | | | | |
| CCAE | 18,444 | 25,301 | 13.3 | 18.9 | 112 | 154 | 8.41 (6.93 - 10.1) | 8.14 (6.91 - 9.53) |
| DAG | 3,163 | 8,568 | 2.5 | 9.0 | 21 | 70 | 8.54 (5.29 - 13.1) | 7.80 (6.08 - 9.86) |
| ODOD | 12,379 | 19,749 | 7.5 | 15.0 | 115 | 229 | 15.2 (12.6 - 18.3) | 15.3 (13.4 - 17.4) |
| MDCR | 1,427 | 4,653 | 0.9 | 3.9 | 11 | 68 | 12.0 (5.99 - 21.5) | 17.4 (13.5 - 22.0) |
| MDCD | 2,330 | 6,060 | 1.2 | 4.2 | 24 | 76 | 19.3 (12.4 - 28.7) | 18.1 (14.2 - 22.6) |
| Open Claims | 266,948 | 428,313 | 197.5 | 373.4 | 1,078 | 2,622 | 5.46 (5.14 - 5.79) | 7.02 (6.76 - 7.30) |
| OEHR | 16,231 | 33,687 | 5.1 | 17.6 | 46 | 206 | 9.07 (6.64 - 12.1) | 11.7 (10.1 - 13.4) |
| SIDIAP | 2,925 | 14,525 | 4.0 | 31.8 | 17 | 140 | 4.27 (2.49 - 6.84) | 4.40 (3.70 - 5.20) |
| Meta-analysis | 320,090 | 530,143 | 229.9 | 465.7 | 1,389 | 3,421 | 6.04 (5.73 - 6.37) | 7.35 (7.10 - 7.60) |
| Meta-analysis (w/o USOC) | 53,142 | 101,830 | 32.4 | 92.3 | 311 | 799 | 9.61 (8.57 - 10.7) | 8.66 (8.07 - 9.28) |
| **PS-matching / on-treatment TAR without escalation** | | | | | | | | |
| CCAE | 16,030 | 25,270 | 10.3 | 15.4 | 86 | 128 | 8.36 (6.69 - 10.3) | 8.31 (6.93 - 9.87) |
| DAG | 2,860 | 8,535 | 2.1 | 8.0 | 19 | 61 | 9.05 (5.45 - 14.1) | 7.59 (5.80 - 9.75) |
| ODOD | 10,339 | 19,502 | 5.9 | 12.4 | 89 | 184 | 15.1 (12.1 - 18.5) | 14.8 (12.7 - 17.1) |
| MDCR | 1,247 | 4,569 | 0.8 | 3.3 | 8 | 60 | 10.6 (4.56 - 20.8) | 17.9 (13.7 - 23.1) |
| MDCD | 2,245 | 6,043 | 1.0 | 3.3 | 21 | 53 | 20.9 (13.0 - 32.0) | 16.2 (12.2 - 21.2) |
| Open Claims | 223,805 | 427,915 | 147.2 | 296.4 | 810 | 2,130 | 5.50 (5.13 - 5.90) | 7.19 (6.88 - 7.50) |
| OEHR | 14,682 | 32,695 | 4.0 | 13.8 | 36 | 155 | 8.95 (6.27 - 12.4) | 11.2 (9.55 - 13.2) |
| SIDIAP | 2,749 | 14,055 | 3.4 | 24.0 | 15 | 106 | 4.47 (2.50 - 7.38) | 4.42 (3.62 - 5.34) |
| Meta-analysis | 272,710 | 534,015 | 173.9 | 373.3 | 1,076 | 2,817 | 6.19 (5.82 - 6.57) | 7.55 (7.27 - 7.83) |
| Meta-analysis (w/o USOC) | 48,905 | 106,100 | 26.7 | 76.9 | 266 | 687 | 9.97 (8.81 - 11.2) | 8.93 (8.27 - 9.62) |
| **PS-stratification / on-treatment TAR without escalation** | | | | | | | | |
| CCAE | 18,444 | 25,301 | 11.5 | 15.4 | 93 | 128 | 8.10 (6.54 - 9.92) | 8.29 (6.92 - 9.86) |
| DAG | 3,163 | 8,568 | 2.2 | 8.1 | 19 | 61 | 8.49 (5.11 - 13.3) | 7.55 (5.78 - 9.70) |
| ODOD | 12,379 | 19,749 | 6.7 | 12.6 | 100 | 190 | 15.0 (12.2 - 18.2) | 15.1 (13.0 - 17.4) |
| MDCR | 1,427 | 4,653 | 0.8 | 3.4 | 10 | 62 | 12.3 (5.89 - 22.6) | 18.2 (14.0 - 23.4) |
| MDCD | 2,330 | 6,060 | 1.0 | 3.3 | 21 | 53 | 20.4 (12.6 - 31.2) | 16.2 (12.1 - 21.1) |
| Open Claims | 266,948 | 428,313 | 166.1 | 296.7 | 907 | 2,138 | 5.46 (5.11 - 5.83) | 7.21 (6.90 - 7.52) |
| OEHR | 16,231 | 33,687 | 4.4 | 14.2 | 39 | 161 | 8.82 (6.27 - 12.1) | 11.3 (9.65 - 13.2) |
| SIDIAP | 2,925 | 14,525 | 3.5 | 25.2 | 15 | 113 | 4.30 (2.40 - 7.09) | 4.49 (3.70 - 5.40) |
| Meta-analysis | 320,090 | 530,143 | 194.5 | 372.2 | 1,173 | 2,791 | 6.03 (5.69 - 6.39) | 7.50 (7.22 - 7.78) |
| Meta-analysis (w/o USOC) | 53,142 | 101,830 | 28.3 | 75.5 | 266 | 653 | 9.39 (8.30 - 10.6) | 8.65 (8.00 - 9.34) |

##### 4.2.12 Acute pancreatitis

**Supplementary Table 323:** Exposures, outcomes and risk estimates for Acute pancreatitis in SGLT2I (female) (T) vs DPP4I (female) (C) new-user cohorts under four different propensity score (PS) adjustment / time-at-risk (TAR) designs across data sources. We report total patient exposure time in 1,000 patient-year (KYr) units, uncalibrated and calibrated hazard ratios (HRs) and their 95% confidence intervals (CIs). Greyed out entries did not pass study diagnostics and are included only for completeness.

| | Patients | | Exposure (KYr) | | Outcomes | | Incidence (per KYr) | |
|---|---|---|---|---|---|---|---|---|
| | T | C | T | C | T | C | T | C |
| **PS-matching / on-treatment TAR** | | | | | | | | |
| CCAE | 16,286 | 25,742 | 12.3 | 19.3 | 21 | 45 | 1.71 (1.06 - 2.62) | 2.33 (1.70 - 3.11) |
| DAG | 2,939 | 8,792 | 2.4 | 9.3 | 0 | 0 | | |
| ODOD | 10,645 | 20,480 | 6.9 | 15.6 | 16 | 49 | 2.31 (1.32 - 3.75) | 3.13 (2.32 - 4.14) |
| MDCR | 1,287 | 4,814 | 0.9 | 4.1 | <5 | 6 | < 5.68 (1.84 - 13.3) | 1.47 (0.54 - 3.19) |
| MDCD | 2,301 | 6,249 | 1.2 | 4.4 | 9 | 21 | 7.21 (3.30 - 13.7) | 4.80 (2.97 - 7.33) |
| Open Claims | 228,911 | 441,231 | 180.6 | 386.7 | 242 | 831 | 1.34 (1.18 - 1.52) | 2.15 (2.01 - 2.30) |
| OEHR | 14,999 | 33,550 | 4.7 | 17.8 | 9 | 34 | 1.90 (0.87 - 3.60) | 1.91 (1.33 - 2.68) |
| SIDIAP | 2,770 | 14,242 | 3.9 | 30.8 | <5 | 12 | < 1.30 (0.42 - 3.03) | 0.39 (0.20 - 0.68) |
| Meta-analysis | 273,142 | 527,252 | 205.7 | 443.8 | 297 | 980 | 1.44 (1.28 - 1.62) | 2.21 (2.07 - 2.35) |
| Meta-analysis (w/o USOC) | 44,231 | 86,021 | 25.2 | 57.1 | 55 | 149 | 2.18 (1.65 - 2.84) | 2.61 (2.21 - 3.06) |
| **PS-stratification / on-treatment TAR** | | | | | | | | |
| CCAE | 18,712 | 25,778 | 13.6 | 19.4 | 22 | 45 | 1.62 (1.02 - 2.45) | 2.32 (1.70 - 3.11) |
| DAG | 3,243 | 8,833 | 2.5 | 9.3 | 0 | 0 | | |
| ODOD | 12,741 | 20,685 | 7.8 | 15.8 | 17 | 52 | 2.18 (1.27 - 3.49) | 3.29 (2.46 - 4.31) |
| MDCR | 1,462 | 4,881 | 0.9 | 4.1 | <5 | 6 | < 5.33 (1.73 - 12.4) | 1.45 (0.53 - 3.15) |
| MDCD | 2,390 | 6,268 | 1.3 | 4.4 | 9 | 21 | 7.05 (3.22 - 13.4) | 4.77 (2.95 - 7.29) |
| Open Claims | 272,688 | 441,635 | 202.2 | 387.1 | 275 | 835 | 1.36 (1.20 - 1.53) | 2.16 (2.01 - 2.31) |
| OEHR | 16,582 | 34,586 | 5.2 | 18.3 | 9 | 36 | 1.73 (0.79 - 3.28) | 1.97 (1.38 - 2.73) |
| SIDIAP | 2,947 | 14,724 | 4.0 | 32.3 | <5 | 13 | < 1.25 (0.40 - 2.91) | 0.40 (0.21 - 0.69) |
| Meta-analysis | 320,723 | 522,684 | 228.8 | 440.5 | 323 | 968 | 1.41 (1.26 - 1.57) | 2.20 (2.06 - 2.34) |
| Meta-analysis (w/o USOC) | 48,035 | 81,049 | 26.6 | 53.5 | 48 | 133 | 1.81 (1.33 - 2.39) | 2.49 (2.08 - 2.95) |
| **PS-matching / on-treatment TAR without escalation** | | | | | | | | |
| CCAE | 16,286 | 25,742 | 10.5 | 15.8 | 16 | 37 | 1.52 (0.87 - 2.47) | 2.35 (1.65 - 3.24) |
| DAG | 2,939 | 8,792 | 2.2 | 8.3 | 0 | 0 | | |
| ODOD | 10,645 | 20,480 | 6.1 | 13.1 | 12 | 39 | 1.96 (1.01 - 3.42) | 2.97 (2.11 - 4.05) |
| MDCR | 1,287 | 4,814 | 0.8 | 3.6 | <5 | 5 | < 6.43 (2.09 - 15.0) | 1.40 (0.46 - 3.27) |
| MDCD | 2,301 | 6,249 | 1.0 | 3.4 | 8 | 14 | 7.78 (3.36 - 15.3) | 4.12 (2.25 - 6.90) |
| Open Claims | 228,911 | 441,231 | 150.9 | 307.3 | 189 | 643 | 1.25 (1.08 - 1.44) | 2.09 (1.93 - 2.26) |
| OEHR | 14,999 | 33,550 | 4.1 | 14.2 | 7 | 26 | 1.70 (0.68 - 3.50) | 1.82 (1.19 - 2.67) |
| SIDIAP | 2,770 | 14,242 | 3.4 | 24.4 | <5 | 8 | < 1.48 (0.48 - 3.45) | 0.33 (0.14 - 0.65) |
| Meta-analysis | 273,142 | 527,252 | 172.7 | 353.9 | 232 | 759 | 1.34 (1.18 - 1.53) | 2.14 (1.99 - 2.30) |
| Meta-analysis (w/o USOC) | 44,231 | 86,021 | 21.8 | 46.6 | 43 | 116 | 1.97 (1.43 - 2.66) | 2.49 (2.06 - 2.99) |
| **PS-stratification / on-treatment TAR without escalation** | | | | | | | | |
| CCAE | 18,712 | 25,778 | 11.7 | 15.8 | 17 | 37 | 1.45 (0.85 - 2.33) | 2.34 (1.65 - 3.23) |
| DAG | 3,243 | 8,833 | 2.3 | 8.4 | 0 | 0 | | |
| ODOD | 12,741 | 20,685 | 6.9 | 13.3 | 13 | 42 | 1.88 (1.00 - 3.21) | 3.16 (2.27 - 4.27) |
| MDCR | 1,462 | 4,881 | 0.8 | 3.6 | <5 | 5 | < 6.01 (1.95 - 14.0) | 1.39 (0.45 - 3.24) |
| MDCD | 2,390 | 6,268 | 1.1 | 3.4 | 8 | 14 | 7.58 (3.27 - 14.9) | 4.10 (2.24 - 6.88) |
| Open Claims | 272,688 | 441,635 | 170.1 | 307.7 | 211 | 646 | 1.24 (1.08 - 1.42) | 2.10 (1.94 - 2.27) |
| OEHR | 16,582 | 34,586 | 4.5 | 14.7 | 7 | 27 | 1.54 (0.62 - 3.18) | 1.84 (1.21 - 2.67) |
| SIDIAP | 2,947 | 14,724 | 3.5 | 25.6 | <5 | 9 | < 1.42 (0.46 - 3.31) | 0.35 (0.16 - 0.67) |
| Meta-analysis | 320,723 | 522,684 | 193.2 | 351.4 | 248 | 752 | 1.28 (1.13 - 1.45) | 2.14 (1.99 - 2.30) |
| Meta-analysis (w/o USOC) | 48,035 | 81,049 | 23.2 | 43.8 | 37 | 106 | 1.60 (1.12 - 2.20) | 2.42 (1.98 - 2.93) |

##### 4.2.13 Bone fracture

**Supplementary Table 324:** Exposures, outcomes and risk estimates for Bone fracture in SGLT2I (female) (T) vs DPP4I (female) (C) new-user cohorts under four different propensity score (PS) adjustment / time-at-risk (TAR) designs across data sources. We report total patient exposure time in 1,000 patient-year (KYr) units, uncalibrated and calibrated hazard ratios (HRs) and their 95% confidence intervals (CIs). Greyed out entries did not pass study diagnostics and are included only for completeness.

| | Patients | | Exposure (KYr) | | Outcomes | | Incidence (per KYr) | |
|---|---|---|---|---|---|---|---|---|
| | T | C | T | C | T | C | T | C |
| **PS-matching / on-treatment TAR** | | | | | | | | |
| CCAE | 14,745 | 23,331 | 10.8 | 17.1 | 203 | 386 | 18.7 (16.3 - 21.5) | 22.6 (20.4 - 24.9) |
| DAG | 2,574 | 7,485 | 2.1 | 7.8 | 30 | 182 | 14.6 (9.86 - 20.9) | 23.5 (20.2 - 27.1) |
| ODOD | 9,580 | 18,098 | 6.1 | 13.5 | 206 | 484 | 33.7 (29.2 - 38.6) | 35.8 (32.7 - 39.2) |
| MDCR | 1,130 | 3,987 | 0.7 | 3.3 | 34 | 148 | 46.2 (32.0 - 64.6) | 45.3 (38.3 - 53.3) |
| MDCD | 2,026 | 5,487 | 1.1 | 3.7 | 33 | 151 | 30.7 (21.1 - 43.1) | 40.4 (34.2 - 47.4) |
| Open Claims | 200,013 | 381,565 | 156.2 | 327.8 | 2,577 | 7,138 | 16.5 (15.9 - 17.2) | 21.8 (21.3 - 22.3) |
| OEHR | 13,943 | 30,967 | 4.4 | 16.1 | 66 | 348 | 15.1 (11.7 - 19.2) | 21.6 (19.4 - 23.9) |
| SIDIAP | 2,357 | 11,917 | 3.3 | 25.2 | 50 | 559 | 15.2 (11.3 - 20.0) | 22.2 (20.4 - 24.1) |
| Meta-analysis | 246,566 | 483,639 | 185.2 | 416.9 | 3,177 | 9,325 | 17.2 (16.6 - 17.8) | 22.4 (21.9 - 22.8) |
| Meta-analysis (w/o USOC) | 46,553 | 102,074 | 29.0 | 89.1 | 600 | 2,187 | 20.7 (19.0 - 22.4) | 24.5 (23.5 - 25.6) |
| **PS-stratification / on-treatment TAR** | | | | | | | | |
| CCAE | 16,973 | 23,365 | 12.0 | 17.1 | 230 | 388 | 19.1 (16.7 - 21.7) | 22.7 (20.5 - 25.0) |
| DAG | 2,825 | 7,508 | 2.2 | 7.8 | 31 | 186 | 14.2 (9.66 - 20.2) | 23.9 (20.6 - 27.6) |
| ODOD | 11,478 | 18,317 | 6.9 | 13.7 | 222 | 500 | 32.1 (28.0 - 36.6) | 36.6 (33.5 - 40.0) |
| MDCR | 1,272 | 4,110 | 0.8 | 3.4 | 35 | 159 | 44.9 (31.3 - 62.4) | 47.4 (40.3 - 55.3) |
| MDCD | 2,102 | 5,506 | 1.1 | 3.8 | 33 | 154 | 30.0 (20.6 - 42.1) | 41.0 (34.8 - 48.0) |
| Open Claims | 237,944 | 382,107 | 175.0 | 328.3 | 2,817 | 7,167 | 16.1 (15.5 - 16.7) | 21.8 (21.3 - 22.3) |
| OEHR | 15,400 | 31,977 | 4.8 | 16.6 | 69 | 368 | 14.3 (11.2 - 18.1) | 22.1 (19.9 - 24.5) |
| SIDIAP | 2,515 | 12,279 | 3.4 | 26.3 | 55 | 605 | 16.0 (12.1 - 20.8) | 23.0 (21.2 - 24.9) |
| Meta-analysis | 287,135 | 475,553 | 204.3 | 409.8 | 3,424 | 9,214 | 16.8 (16.2 - 17.3) | 22.5 (22.0 - 22.9) |
| Meta-analysis (w/o USOC) | 49,191 | 93,446 | 29.4 | 81.5 | 607 | 2,047 | 20.7 (19.1 - 22.4) | 25.1 (24.0 - 26.2) |
| **PS-matching / on-treatment TAR without escalation** | | | | | | | | |
| CCAE | 14,745 | 23,331 | 9.3 | 14.1 | 172 | 302 | 18.5 (15.9 - 21.5) | 21.5 (19.1 - 24.0) |
| DAG | 2,574 | 7,485 | 1.9 | 7.0 | 27 | 167 | 14.4 (9.50 - 21.0) | 24.0 (20.5 - 27.9) |
| ODOD | 9,580 | 18,098 | 5.4 | 11.4 | 177 | 399 | 32.6 (28.0 - 37.8) | 35.1 (31.7 - 38.7) |
| MDCR | 1,130 | 3,987 | 0.7 | 2.9 | 29 | 123 | 44.6 (29.8 - 64.0) | 42.7 (35.5 - 50.9) |
| MDCD | 2,026 | 5,487 | 0.9 | 2.9 | 28 | 117 | 31.6 (21.0 - 45.6) | 40.4 (33.4 - 48.4) |
| Open Claims | 200,013 | 381,565 | 130.6 | 261.2 | 2,137 | 5,665 | 16.4 (15.7 - 17.1) | 21.7 (21.1 - 22.3) |
| OEHR | 13,943 | 30,967 | 3.8 | 13.0 | 56 | 256 | 14.7 (11.1 - 19.1) | 19.7 (17.3 - 22.2) |
| SIDIAP | 2,357 | 11,917 | 2.9 | 20.0 | 41 | 457 | 14.2 (10.2 - 19.2) | 22.9 (20.8 - 25.1) |
| Meta-analysis | 246,566 | 483,639 | 156.0 | 334.1 | 2,649 | 7,425 | 17.0 (16.3 - 17.6) | 22.2 (21.7 - 22.7) |
| Meta-analysis (w/o USOC) | 46,553 | 102,074 | 25.3 | 72.9 | 512 | 1,760 | 20.2 (18.5 - 22.0) | 24.1 (23.0 - 25.3) |
| **PS-stratification / on-treatment TAR without escalation** | | | | | | | | |
| CCAE | 16,973 | 23,365 | 10.4 | 14.1 | 196 | 304 | 18.9 (16.3 - 21.7) | 21.6 (19.2 - 24.2) |
| DAG | 2,825 | 7,508 | 2.0 | 7.0 | 28 | 171 | 14.1 (9.36 - 20.4) | 24.5 (21.0 - 28.4) |
| ODOD | 11,478 | 18,317 | 6.2 | 11.5 | 192 | 413 | 31.2 (27.0 - 36.0) | 35.8 (32.5 - 39.5) |
| MDCR | 1,272 | 4,110 | 0.7 | 3.0 | 30 | 134 | 43.4 (29.3 - 61.9) | 45.3 (37.9 - 53.6) |
| MDCD | 2,102 | 5,506 | 0.9 | 2.9 | 28 | 118 | 30.7 (20.4 - 44.4) | 40.5 (33.5 - 48.5) |
| Open Claims | 237,944 | 382,107 | 147.3 | 261.6 | 2,346 | 5,689 | 15.9 (15.3 - 16.6) | 21.7 (21.2 - 22.3) |
| OEHR | 15,400 | 31,977 | 4.2 | 13.4 | 59 | 272 | 14.1 (10.7 - 18.2) | 20.3 (17.9 - 22.8) |
| SIDIAP | 2,515 | 12,279 | 3.0 | 20.8 | 46 | 497 | 15.2 (11.2 - 20.3) | 23.8 (21.8 - 26.0) |
| Meta-analysis | 287,135 | 475,553 | 173.0 | 328.4 | 2,867 | 7,346 | 16.6 (16.0 - 17.2) | 22.4 (21.9 - 22.9) |
| Meta-analysis (w/o USOC) | 49,191 | 93,446 | 25.7 | 66.9 | 521 | 1,657 | 20.3 (18.6 - 22.1) | 24.8 (23.6 - 26.0) |

##### 4.2.14 Diabetic ketoacidosis

**Supplementary Table 325:** Exposures, outcomes and risk estimates for Diabetic ketoacidosis in SGLT2I (female) (T) vs DPP4I (female) (C) new-user cohorts under four different propensity score (PS) adjustment / time-at-risk (TAR) designs across data sources. We report total patient exposure time in 1,000 patient-year (KYr) units, uncalibrated and calibrated hazard ratios (HRs) and their 95% confidence intervals (CIs). Greyed out entries did not pass study diagnostics and are included only for completeness.

| | Patients | | Exposure (KYr) | | Outcomes | | Incidence (per KYr) | |
|---|---|---|---|---|---|---|---|---|
| | T | C | T | C | T | C | T | C |
| **PS-matching / on-treatment TAR** | | | | | | | | |
| CCAE | 16,394 | 25,898 | 12.3 | 19.5 | 25 | 19 | 2.03 (1.31 - 2.99) | 0.98 (0.59 - 1.52) |
| DAG | 2,939 | 8,792 | 2.4 | 9.3 | 0 | 0 | | |
| ODOD | 10,722 | 20,613 | 7.0 | 15.7 | 21 | 28 | 3.01 (1.86 - 4.60) | 1.78 (1.18 - 2.57) |
| MDCR | 1,300 | 4,828 | 0.9 | 4.1 | <5 | 5 | < 5.64 (1.83 - 13.2) | 1.22 (0.40 - 2.84) |
| MDCD | 2,335 | 6,347 | 1.3 | 4.4 | <5 | 16 | < 3.92 (1.27 - 9.15) | 3.60 (2.06 - 5.85) |
| Open Claims | 231,509 | 445,431 | 182.6 | 390.4 | 212 | 359 | 1.16 (1.01 - 1.33) | 0.92 (0.83 - 1.02) |
| OEHR | 15,056 | 33,689 | 4.8 | 17.8 | 8 | 22 | 1.68 (0.73 - 3.31) | 1.23 (0.77 - 1.87) |
| SIDIAP | 2,778 | 14,274 | 3.9 | 30.9 | 0 | 0 | | |
| Meta-analysis | 276,016 | 531,978 | 207.9 | 447.9 | <271 | 444 | < 1.30 (1.15 - 1.47) | 0.99 (0.90 - 1.09) |
| Meta-analysis (w/o USOC) | 44,507 | 86,547 | 25.3 | 57.5 | <59 | 85 | < 2.33 (1.77 - 3.00) | 1.48 (1.18 - 1.83) |
| **PS-stratification / on-treatment TAR** | | | | | | | | |
| CCAE | 18,865 | 25,934 | 13.7 | 19.5 | 31 | 19 | 2.27 (1.54 - 3.22) | 0.97 (0.59 - 1.52) |
| DAG | 3,243 | 8,833 | 2.5 | 9.3 | 0 | 0 | | |
| ODOD | 12,833 | 20,819 | 7.9 | 15.9 | 24 | 28 | 3.05 (1.96 - 4.54) | 1.76 (1.17 - 2.54) |
| MDCR | 1,475 | 4,895 | 0.9 | 4.2 | <5 | 5 | < 5.29 (1.72 - 12.3) | 1.20 (0.39 - 2.81) |
| MDCD | 2,427 | 6,366 | 1.3 | 4.5 | 5 | 16 | 3.83 (1.24 - 8.94) | 3.58 (2.05 - 5.82) |
| Open Claims | 276,157 | 445,863 | 204.7 | 390.8 | 254 | 359 | 1.24 (1.09 - 1.40) | 0.92 (0.83 - 1.02) |
| OEHR | 16,657 | 34,731 | 5.2 | 18.4 | 11 | 24 | 2.10 (1.05 - 3.76) | 1.31 (0.84 - 1.95) |
| SIDIAP | 2,956 | 14,758 | 4.0 | 32.4 | 0 | 0 | | |
| Meta-analysis | 324,512 | 527,347 | 231.4 | 444.6 | 320 | 430 | 1.38 (1.24 - 1.54) | 0.97 (0.88 - 1.06) |
| Meta-analysis (w/o USOC) | 48,355 | 81,484 | 26.8 | 53.8 | 66 | 71 | 2.47 (1.91 - 3.14) | 1.32 (1.03 - 1.66) |
| **PS-matching / on-treatment TAR without escalation** | | | | | | | | |
| CCAE | 16,394 | 25,898 | 10.6 | 15.9 | 18 | 13 | 1.70 (1.01 - 2.69) | 0.82 (0.44 - 1.40) |
| DAG | 2,939 | 8,792 | 2.2 | 8.3 | 0 | 0 | | |
| ODOD | 10,722 | 20,613 | 6.2 | 13.2 | 19 | 22 | 3.08 (1.85 - 4.81) | 1.66 (1.04 - 2.52) |
| MDCR | 1,300 | 4,828 | 0.8 | 3.6 | <5 | <5 | < 6.38 (2.07 - 14.9) | < 1.40 (0.45 - 3.26) |
| MDCD | 2,335 | 6,347 | 1.1 | 3.5 | <5 | 11 | < 4.76 (1.54 - 11.1) | 3.18 (1.59 - 5.70) |
| Open Claims | 231,509 | 445,431 | 152.5 | 310.4 | 164 | 250 | 1.08 (0.92 - 1.25) | 0.81 (0.71 - 0.91) |
| OEHR | 15,056 | 33,689 | 4.1 | 14.3 | <5 | 13 | < 1.21 (0.39 - 2.82) | 0.91 (0.48 - 1.55) |
| SIDIAP | 2,778 | 14,274 | 3.4 | 24.4 | 0 | 0 | | |
| Meta-analysis | 258,625 | 491,942 | 169.2 | 339.5 | 201 | 285 | 1.19 (1.03 - 1.36) | 0.84 (0.74 - 0.94) |
| Meta-analysis (w/o USOC) | 27,116 | 46,511 | 16.7 | 29.1 | 37 | 35 | 2.21 (1.56 - 3.05) | 1.20 (0.84 - 1.67) |
| **PS-stratification / on-treatment TAR without escalation** | | | | | | | | |
| CCAE | 18,865 | 25,934 | 11.8 | 15.9 | 24 | 13 | 2.04 (1.31 - 3.03) | 0.82 (0.44 - 1.40) |
| DAG | 3,243 | 8,833 | 2.3 | 8.4 | 0 | 0 | | |
| ODOD | 12,833 | 20,819 | 7.0 | 13.4 | 20 | 22 | 2.87 (1.75 - 4.43) | 1.64 (1.03 - 2.49) |
| MDCR | 1,475 | 4,895 | 0.8 | 3.6 | <5 | <5 | < 5.97 (1.94 - 13.9) | < 1.38 (0.45 - 3.23) |
| MDCD | 2,427 | 6,366 | 1.1 | 3.5 | <5 | 11 | < 4.64 (1.51 - 10.8) | 3.17 (1.58 - 5.68) |
| Open Claims | 276,157 | 445,863 | 172.1 | 310.7 | 200 | 250 | 1.16 (1.01 - 1.33) | 0.80 (0.71 - 0.91) |
| OEHR | 16,657 | 34,731 | 4.6 | 14.8 | 7 | 14 | 1.54 (0.62 - 3.17) | 0.95 (0.52 - 1.59) |
| SIDIAP | 2,956 | 14,758 | 3.5 | 25.6 | 0 | 0 | | |
| Meta-analysis | 324,512 | 527,347 | 195.4 | 354.8 | 251 | 299 | 1.28 (1.13 - 1.45) | 0.84 (0.75 - 0.94) |
| Meta-analysis (w/o USOC) | 48,355 | 81,484 | 23.3 | 44.0 | 51 | 49 | 2.19 (1.63 - 2.88) | 1.11 (0.82 - 1.47) |

##### 4.2.15 Diarrhea

**Supplementary Table 326:** Exposures, outcomes and risk estimates for Diarrhea in SGLT2I (female) (T) vs DPP4I (female) (C) new-user cohorts under four different propensity score (PS) adjustment / time-at-risk (TAR) designs across data sources. We report total patient exposure time in 1,000 patient-year (KYr) units, uncalibrated and calibrated hazard ratios (HRs) and their 95% confidence intervals (CIs). Greyed out entries did not pass study diagnostics and are included only for completeness.

| | Patients | | Exposure (KYr) | | Outcomes | | Incidence (per KYr) | |
|---|---|---|---|---|---|---|---|---|
| | T | C | T | C | T | C | T | C |
| **PS-matching / on-treatment TAR** | | | | | | | | |
| CCAE | 13,703 | 21,595 | 10.2 | 15.7 | 329 | 589 | 32.1 (28.7 - 35.8) | 37.5 (34.5 - 40.7) |
| DAG | 2,883 | 8,641 | 2.3 | 9.1 | 7 | 22 | 3.01 (1.21 - 6.20) | 2.42 (1.52 - 3.67) |
| ODOD | 8,810 | 16,500 | 5.6 | 12.1 | 238 | 625 | 42.2 (37.0 - 47.9) | 51.6 (47.7 - 55.8) |
| MDCR | 1,060 | 3,818 | 0.7 | 3.1 | 27 | 131 | 37.2 (24.5 - 54.1) | 42.0 (35.1 - 49.8) |
| MDCD | 1,724 | 4,705 | 0.9 | 3.1 | 55 | 216 | 59.7 (45.0 - 77.8) | 69.2 (60.3 - 79.1) |
| Open Claims | 193,021 | 370,847 | 151.0 | 317.9 | 2,852 | 8,032 | 18.9 (18.2 - 19.6) | 25.3 (24.7 - 25.8) |
| OEHR | 12,832 | 28,602 | 4.0 | 14.4 | 159 | 695 | 40.2 (34.2 - 46.9) | 48.4 (44.8 - 52.1) |
| SIDIAP | 2,601 | 13,232 | 3.6 | 28.3 | 53 | 432 | 14.8 (11.1 - 19.3) | 15.2 (13.8 - 16.7) |
| Meta-analysis | 236,667 | 468,127 | 178.8 | 405.3 | 3,713 | 10,717 | 20.8 (20.1 - 21.4) | 26.4 (25.9 - 26.9) |
| Meta-analysis (w/o USOC) | 43,646 | 97,280 | 27.7 | 87.4 | 861 | 2,685 | 31.0 (29.0 - 33.2) | 30.7 (29.6 - 31.9) |
| **PS-stratification / on-treatment TAR** | | | | | | | | |
| CCAE | 15,864 | 21,619 | 11.4 | 15.7 | 371 | 589 | 32.6 (29.4 - 36.1) | 37.5 (34.5 - 40.6) |
| DAG | 3,170 | 8,679 | 2.5 | 9.1 | 7 | 23 | 2.84 (1.14 - 5.85) | 2.52 (1.60 - 3.78) |
| ODOD | 10,574 | 16,684 | 6.4 | 12.2 | 262 | 634 | 41.1 (36.3 - 46.4) | 51.9 (47.9 - 56.1) |
| MDCR | 1,217 | 3,941 | 0.8 | 3.2 | 29 | 139 | 37.1 (24.8 - 53.2) | 43.3 (36.4 - 51.1) |
| MDCD | 1,793 | 4,717 | 0.9 | 3.1 | 57 | 217 | 60.4 (45.7 - 78.2) | 69.3 (60.4 - 79.2) |
| Open Claims | 230,071 | 371,526 | 169.4 | 318.5 | 3,165 | 8,067 | 18.7 (18.0 - 19.3) | 25.3 (24.8 - 25.9) |
| OEHR | 14,218 | 29,436 | 4.4 | 14.8 | 177 | 720 | 40.7 (34.9 - 47.1) | 48.7 (45.2 - 52.4) |
| SIDIAP | 2,780 | 13,682 | 3.8 | 29.7 | 58 | 451 | 15.5 (11.7 - 20.0) | 15.2 (13.8 - 16.7) |
| Meta-analysis | 276,677 | 461,626 | 197.7 | 400.1 | 4,040 | 10,484 | 20.4 (19.8 - 21.1) | 26.2 (25.7 - 26.7) |
| Meta-analysis (w/o USOC) | 46,606 | 90,100 | 28.3 | 81.5 | 875 | 2,417 | 30.9 (28.9 - 33.0) | 29.6 (28.5 - 30.8) |
| **PS-matching / on-treatment TAR without escalation** | | | | | | | | |
| CCAE | 13,703 | 21,595 | 8.9 | 13.0 | 282 | 479 | 31.8 (28.2 - 35.8) | 36.8 (33.6 - 40.2) |
| DAG | 2,883 | 8,641 | 2.1 | 8.2 | 7 | 21 | 3.31 (1.33 - 6.81) | 2.57 (1.59 - 3.92) |
| ODOD | 8,810 | 16,500 | 5.0 | 10.3 | 213 | 522 | 42.5 (37.0 - 48.6) | 50.7 (46.5 - 55.3) |
| MDCR | 1,060 | 3,818 | 0.6 | 2.7 | 24 | 114 | 37.6 (24.1 - 55.9) | 41.7 (34.4 - 50.1) |
| MDCD | 1,724 | 4,705 | 0.8 | 2.5 | 45 | 177 | 58.9 (43.0 - 78.8) | 71.1 (61.0 - 82.4) |
| Open Claims | 193,021 | 370,847 | 127.1 | 254.9 | 2,324 | 6,448 | 18.3 (17.6 - 19.0) | 25.3 (24.7 - 25.9) |
| OEHR | 12,832 | 28,602 | 3.5 | 11.7 | 131 | 549 | 37.7 (31.5 - 44.7) | 47.0 (43.2 - 51.1) |
| SIDIAP | 2,601 | 13,232 | 3.2 | 22.5 | 46 | 340 | 14.6 (10.7 - 19.4) | 15.1 (13.5 - 16.8) |
| Meta-analysis | 236,667 | 468,127 | 151.4 | 326.9 | 3,067 | 8,624 | 20.3 (19.5 - 21.0) | 26.4 (25.8 - 26.9) |
| Meta-analysis (w/o USOC) | 43,646 | 97,280 | 24.3 | 72.0 | 743 | 2,176 | 30.5 (28.4 - 32.8) | 30.2 (29.0 - 31.5) |
| **PS-stratification / on-treatment TAR without escalation** | | | | | | | | |
| CCAE | 15,864 | 21,619 | 9.9 | 13.0 | 322 | 479 | 32.5 (29.1 - 36.3) | 36.8 (33.5 - 40.2) |
| DAG | 3,170 | 8,679 | 2.2 | 8.2 | 7 | 22 | 3.12 (1.25 - 6.43) | 2.67 (1.68 - 4.05) |
| ODOD | 10,574 | 16,684 | 5.7 | 10.4 | 234 | 530 | 41.2 (36.1 - 46.8) | 50.9 (46.7 - 55.5) |
| MDCR | 1,217 | 3,941 | 0.7 | 2.8 | 26 | 121 | 37.6 (24.6 - 55.1) | 43.1 (35.8 - 51.5) |
| MDCD | 1,793 | 4,717 | 0.8 | 2.5 | 47 | 178 | 59.8 (43.9 - 79.5) | 71.4 (61.3 - 82.7) |
| Open Claims | 230,071 | 371,526 | 143.4 | 255.4 | 2,586 | 6,473 | 18.0 (17.3 - 18.7) | 25.3 (24.7 - 26.0) |
| OEHR | 14,218 | 29,436 | 3.8 | 12.0 | 147 | 568 | 38.4 (32.5 - 45.2) | 47.2 (43.4 - 51.3) |
| SIDIAP | 2,780 | 13,682 | 3.3 | 23.6 | 50 | 357 | 15.2 (11.2 - 20.0) | 15.1 (13.6 - 16.8) |
| Meta-analysis | 276,677 | 461,626 | 168.3 | 322.7 | 3,346 | 8,429 | 19.9 (19.2 - 20.6) | 26.1 (25.6 - 26.7) |
| Meta-analysis (w/o USOC) | 46,606 | 90,100 | 24.9 | 67.3 | 760 | 1,956 | 30.5 (28.3 - 32.7) | 29.1 (27.8 - 30.4) |

##### 4.2.16 Genitourinary infection

**Supplementary Table 327:** Exposures, outcomes and risk estimates for Genitourinary infection in SGLT2I (female) (T) vs DPP4I (female) (C) new-user cohorts under four different propensity score (PS) adjustment / time-at-risk (TAR) designs across data sources. We report total patient exposure time in 1,000 patient-year (KYr) units, uncalibrated and calibrated hazard ratios (HRs) and their 95% confidence intervals (CIs). Greyed out entries did not pass study diagnostics and are included only for completeness.

| | Patients | | Exposure (KYr) | | Outcomes | | Incidence (per KYr) | |
|---|---|---|---|---|---|---|---|---|
| | T | C | T | C | T | C | T | C |
| **PS-matching / on-treatment TAR** | | | | | | | | |
| CCAE | 11,146 | 17,561 | 8.0 | 12.5 | 498 | 857 | 61.9 (56.6 - 67.6) | 68.6 (64.0 - 73.3) |
| DAG | 1,996 | 5,825 | 1.6 | 5.8 | 83 | 302 | 52.8 (42.0 - 65.4) | 52.3 (46.6 - 58.6) |
| ODOD | 7,390 | 13,219 | 4.6 | 9.5 | 365 | 945 | 79.3 (71.4 - 87.9) | 99.9 (93.6 - 106) |
| MDCR | 963 | 3,293 | 0.6 | 2.6 | 51 | 246 | 79.3 (59.1 - 104) | 94.9 (83.4 - 108) |
| MDCD | 1,545 | 4,091 | 0.8 | 2.7 | 78 | 265 | 96.2 (76.1 - 120) | 98.8 (87.3 - 111) |
| Open Claims | 153,187 | 292,252 | 121.4 | 246.4 | 4,837 | 13,138 | 39.9 (38.7 - 41.0) | 53.3 (52.4 - 54.2) |
| OEHR | 11,514 | 25,196 | 3.5 | 12.5 | 227 | 934 | 65.3 (57.1 - 74.4) | 74.9 (70.2 - 79.9) |
| SIDIAP | 1,802 | 9,133 | 2.6 | 18.9 | 146 | 1,140 | 57.2 (48.3 - 67.3) | 60.3 (56.8 - 63.9) |
| Meta-analysis | 189,714 | 371,177 | 143.5 | 312.7 | 6,258 | 17,692 | 43.6 (42.5 - 44.7) | 56.6 (55.7 - 57.4) |
| Meta-analysis (w/o USOC) | 36,527 | 78,925 | 22.2 | 66.4 | 1,421 | 4,554 | 64.1 (60.8 - 67.5) | 68.6 (66.7 - 70.7) |
| **PS-stratification / on-treatment TAR** | | | | | | | | |
| CCAE | 12,966 | 17,577 | 9.0 | 12.5 | 553 | 857 | 61.4 (56.4 - 66.7) | 68.5 (64.0 - 73.3) |
| DAG | 2,201 | 5,902 | 1.7 | 5.9 | 89 | 311 | 53.2 (42.7 - 65.5) | 53.1 (47.4 - 59.4) |
| ODOD | 8,866 | 13,327 | 5.2 | 9.5 | 402 | 959 | 76.9 (69.6 - 84.8) | 100 (94.2 - 107) |
| MDCR | 1,106 | 3,317 | 0.7 | 2.6 | 54 | 249 | 77.8 (58.4 - 102) | 95.5 (84.0 - 108) |
| MDCD | 1,602 | 4,111 | 0.8 | 2.7 | 79 | 269 | 95.5 (75.6 - 119) | 99.3 (87.8 - 112) |
| Open Claims | 183,262 | 292,863 | 136.5 | 246.8 | 5,341 | 13,216 | 39.1 (38.1 - 40.2) | 53.5 (52.6 - 54.5) |
| OEHR | 12,672 | 26,236 | 3.8 | 12.9 | 254 | 987 | 66.5 (58.5 - 75.2) | 76.3 (71.6 - 81.2) |
| SIDIAP | 1,919 | 9,437 | 2.7 | 19.8 | 154 | 1,212 | 57.8 (49.0 - 67.7) | 61.3 (57.9 - 64.9) |
| Meta-analysis | 221,886 | 365,342 | 158.9 | 307.5 | 6,793 | 17,542 | 42.7 (41.7 - 43.8) | 57.1 (56.2 - 57.9) |
| Meta-analysis (w/o USOC) | 38,624 | 72,479 | 22.4 | 60.6 | 1,452 | 4,326 | 64.8 (61.5 - 68.3) | 71.4 (69.3 - 73.5) |
| **PS-matching / on-treatment TAR without escalation** | | | | | | | | |
| CCAE | 11,146 | 17,561 | 7.0 | 10.3 | 435 | 706 | 62.3 (56.6 - 68.5) | 68.5 (63.6 - 73.8) |
| DAG | 1,996 | 5,825 | 1.4 | 5.2 | 75 | 279 | 52.4 (41.2 - 65.7) | 53.3 (47.2 - 59.9) |
| ODOD | 7,390 | 13,219 | 4.1 | 8.1 | 321 | 805 | 78.2 (69.9 - 87.3) | 99.8 (93.0 - 107) |
| MDCR | 963 | 3,293 | 0.6 | 2.3 | 46 | 202 | 80.8 (59.1 - 108) | 89.0 (77.1 - 102) |
| MDCD | 1,545 | 4,091 | 0.7 | 2.1 | 58 | 192 | 85.0 (64.5 - 110) | 90.2 (77.9 - 104) |
| Open Claims | 153,187 | 292,252 | 101.8 | 197.4 | 4,088 | 10,580 | 40.1 (38.9 - 41.4) | 53.6 (52.6 - 54.6) |
| OEHR | 11,514 | 25,196 | 3.0 | 10.1 | 196 | 727 | 64.5 (55.8 - 74.2) | 71.7 (66.6 - 77.2) |
| SIDIAP | 1,802 | 9,133 | 2.2 | 15.2 | 134 | 905 | 59.7 (50.0 - 70.7) | 59.6 (55.7 - 63.6) |
| Meta-analysis | 189,714 | 371,177 | 121.3 | 252.2 | 5,329 | 14,286 | 43.9 (42.8 - 45.1) | 56.7 (55.7 - 57.6) |
| Meta-analysis (w/o USOC) | 36,527 | 78,925 | 19.5 | 54.8 | 1,241 | 3,706 | 63.7 (60.2 - 67.4) | 67.6 (65.5 - 69.8) |
| **PS-stratification / on-treatment TAR without escalation** | | | | | | | | |
| CCAE | 12,966 | 17,577 | 7.8 | 10.3 | 485 | 706 | 61.8 (56.4 - 67.5) | 68.5 (63.5 - 73.7) |
| DAG | 2,201 | 5,902 | 1.5 | 5.3 | 81 | 288 | 53.1 (42.2 - 66.1) | 54.3 (48.2 - 60.9) |
| ODOD | 8,866 | 13,327 | 4.7 | 8.2 | 355 | 819 | 75.9 (68.2 - 84.2) | 100 (93.7 - 108) |
| MDCR | 1,106 | 3,317 | 0.6 | 2.3 | 49 | 205 | 79.4 (58.8 - 105) | 89.9 (78.0 - 103) |
| MDCD | 1,602 | 4,111 | 0.7 | 2.2 | 59 | 195 | 84.7 (64.5 - 109) | 90.6 (78.4 - 104) |
| Open Claims | 183,262 | 292,863 | 115.3 | 197.8 | 4,529 | 10,651 | 39.3 (38.1 - 40.4) | 53.9 (52.8 - 54.9) |
| OEHR | 12,672 | 26,236 | 3.3 | 10.5 | 218 | 770 | 65.4 (57.0 - 74.6) | 73.2 (68.2 - 78.6) |
| SIDIAP | 1,919 | 9,437 | 2.3 | 15.9 | 141 | 968 | 60.1 (50.6 - 70.9) | 60.9 (57.1 - 64.9) |
| Meta-analysis | 221,886 | 365,342 | 135.0 | 247.9 | 5,809 | 14,202 | 43.0 (41.9 - 44.1) | 57.3 (56.3 - 58.2) |
| Meta-analysis (w/o USOC) | 38,624 | 72,479 | 19.7 | 50.2 | 1,280 | 3,551 | 64.9 (61.4 - 68.5) | 70.8 (68.5 - 73.1) |

##### 4.2.17 Hyperkalemia

**Supplementary Table 328:** Exposures, outcomes and risk estimates for Hyperkalemia in SGLT2I (female) (T) vs DPP4I (female) (C) new-user cohorts under four different propensity score (PS) adjustment / time-at-risk (TAR) designs across data sources. We report total patient exposure time in 1,000 patient-year (KYr) units, uncalibrated and calibrated hazard ratios (HRs) and their 95% confidence intervals (CIs). Greyed out entries did not pass study diagnostics and are included only for completeness.

| | Patients | | Exposure (KYr) | | Outcomes | | Incidence (per KYr) | |
|---|---|---|---|---|---|---|---|---|
| | T | C | T | C | T | C | T | C |
| **PS-matching / on-treatment TAR** | | | | | | | | |
| CCAE | 16,149 | 25,490 | 12.1 | 19.0 | 74 | 127 | 6.13 (4.82 - 7.70) | 6.67 (5.56 - 7.93) |
| DAG | 2,759 | 8,171 | 2.2 | 8.5 | 25 | 145 | 11.2 (7.25 - 16.5) | 17.1 (14.5 - 20.2) |
| ODOD | 10,330 | 19,477 | 6.7 | 14.6 | 98 | 361 | 14.6 (11.9 - 17.8) | 24.8 (22.3 - 27.5) |
| MDCR | 1,274 | 4,650 | 0.9 | 3.9 | 5 | 66 | 5.72 (1.86 - 13.3) | 16.8 (13.0 - 21.4) |
| MDCD | 2,274 | 6,148 | 1.2 | 4.2 | 7 | 69 | 5.65 (2.27 - 11.6) | 16.3 (12.7 - 20.7) |
| Open Claims | 227,815 | 435,705 | 179.5 | 378.9 | 641 | 2,940 | 3.57 (3.30 - 3.86) | 7.76 (7.48 - 8.05) |
| OEHR | 14,821 | 33,041 | 4.7 | 17.3 | 30 | 225 | 6.43 (4.33 - 9.17) | 13.0 (11.4 - 14.9) |
| SIDIAP | 2,677 | 13,494 | 3.7 | 28.9 | 35 | 233 | 9.40 (6.55 - 13.1) | 8.05 (7.05 - 9.16) |
| Meta-analysis | 276,825 | 541,526 | 210.1 | 471.4 | 910 | 4,100 | 4.33 (4.05 - 4.62) | 8.70 (8.43 - 8.97) |
| Meta-analysis (w/o USOC) | 49,010 | 105,821 | 30.6 | 92.5 | 269 | 1,160 | 8.78 (7.76 - 9.89) | 12.5 (11.8 - 13.3) |
| **PS-stratification / on-treatment TAR** | | | | | | | | |
| CCAE | 18,560 | 25,525 | 13.4 | 19.1 | 76 | 127 | 5.69 (4.48 - 7.12) | 6.66 (5.55 - 7.92) |
| DAG | 3,033 | 8,195 | 2.4 | 8.5 | 26 | 145 | 11.0 (7.17 - 16.1) | 17.1 (14.4 - 20.1) |
| ODOD | 12,381 | 19,703 | 7.6 | 14.7 | 109 | 371 | 14.4 (11.8 - 17.4) | 25.2 (22.7 - 27.9) |
| MDCR | 1,440 | 4,709 | 0.9 | 4.0 | 5 | 71 | 5.37 (1.75 - 12.5) | 17.9 (14.0 - 22.6) |
| MDCD | 2,363 | 6,166 | 1.3 | 4.2 | 7 | 70 | 5.52 (2.22 - 11.4) | 16.5 (12.9 - 20.9) |
| Open Claims | 271,518 | 436,158 | 201.2 | 379.3 | 717 | 2,958 | 3.56 (3.31 - 3.83) | 7.80 (7.52 - 8.08) |
| OEHR | 16,395 | 34,043 | 5.1 | 17.8 | 32 | 237 | 6.23 (4.26 - 8.80) | 13.3 (11.7 - 15.1) |
| SIDIAP | 2,857 | 13,932 | 3.9 | 30.2 | 40 | 254 | 10.3 (7.35 - 14.0) | 8.40 (7.40 - 9.50) |
| Meta-analysis | 324,744 | 537,556 | 233.5 | 469.7 | 1,000 | 4,092 | 4.28 (4.02 - 4.56) | 8.71 (8.45 - 8.98) |
| Meta-analysis (w/o USOC) | 53,226 | 101,398 | 32.3 | 90.3 | 283 | 1,134 | 8.76 (7.77 - 9.84) | 12.6 (11.8 - 13.3) |
| **PS-matching / on-treatment TAR without escalation** | | | | | | | | |
| CCAE | 16,149 | 25,490 | 10.4 | 15.5 | 65 | 102 | 6.27 (4.84 - 7.99) | 6.57 (5.36 - 7.97) |
| DAG | 2,759 | 8,171 | 2.0 | 7.6 | 22 | 128 | 10.8 (6.77 - 16.4) | 16.7 (14.0 - 19.9) |
| ODOD | 10,330 | 19,477 | 5.9 | 12.3 | 86 | 299 | 14.5 (11.6 - 17.9) | 24.4 (21.7 - 27.3) |
| MDCR | 1,274 | 4,650 | 0.8 | 3.4 | 5 | 57 | 6.48 (2.10 - 15.1) | 16.7 (12.6 - 21.6) |
| MDCD | 2,274 | 6,148 | 1.0 | 3.3 | 7 | 55 | 6.84 (2.75 - 14.1) | 16.8 (12.6 - 21.8) |
| Open Claims | 227,815 | 435,705 | 150.0 | 301.3 | 511 | 2,333 | 3.41 (3.12 - 3.72) | 7.74 (7.43 - 8.06) |
| OEHR | 14,821 | 33,041 | 4.1 | 13.9 | 25 | 167 | 6.16 (3.99 - 9.09) | 12.0 (10.3 - 14.0) |
| SIDIAP | 2,677 | 13,494 | 3.3 | 22.9 | 31 | 195 | 9.45 (6.42 - 13.4) | 8.50 (7.35 - 9.79) |
| Meta-analysis | 276,825 | 541,526 | 176.7 | 376.8 | 747 | 3,279 | 4.23 (3.93 - 4.54) | 8.70 (8.41 - 9.00) |
| Meta-analysis (w/o USOC) | 49,010 | 105,821 | 26.7 | 75.5 | 236 | 946 | 8.84 (7.75 - 10.0) | 12.5 (11.7 - 13.3) |
| **PS-stratification / on-treatment TAR without escalation** | | | | | | | | |
| CCAE | 18,560 | 25,525 | 11.5 | 15.6 | 67 | 102 | 5.81 (4.50 - 7.38) | 6.56 (5.35 - 7.96) |
| DAG | 3,033 | 8,195 | 2.2 | 7.7 | 23 | 128 | 10.6 (6.74 - 16.0) | 16.7 (13.9 - 19.8) |
| ODOD | 12,381 | 19,703 | 6.7 | 12.4 | 97 | 308 | 14.5 (11.7 - 17.6) | 24.8 (22.1 - 27.7) |
| MDCR | 1,440 | 4,709 | 0.8 | 3.5 | 5 | 61 | 6.07 (1.97 - 14.2) | 17.7 (13.5 - 22.7) |
| MDCD | 2,363 | 6,166 | 1.0 | 3.3 | 7 | 56 | 6.67 (2.68 - 13.7) | 17.0 (12.8 - 22.1) |
| Open Claims | 271,518 | 436,158 | 169.2 | 301.7 | 580 | 2,348 | 3.43 (3.15 - 3.72) | 7.78 (7.47 - 8.11) |
| OEHR | 16,395 | 34,043 | 4.5 | 14.3 | 26 | 174 | 5.82 (3.81 - 8.53) | 12.2 (10.4 - 14.1) |
| SIDIAP | 2,857 | 13,932 | 3.4 | 24.0 | 35 | 212 | 10.2 (7.12 - 14.2) | 8.84 (7.69 - 10.1) |
| Meta-analysis | 324,744 | 537,556 | 197.5 | 375.6 | 828 | 3,272 | 4.19 (3.91 - 4.49) | 8.71 (8.42 - 9.02) |
| Meta-analysis (w/o USOC) | 53,226 | 101,398 | 28.3 | 73.9 | 248 | 924 | 8.76 (7.71 - 9.93) | 12.5 (11.7 - 13.3) |

##### 4.2.18 Hypoglycemia

**Supplementary Table 329:** Exposures, outcomes and risk estimates for Hypoglycemia in SGLT2I (female) (T) vs DPP4I (female) (C) new-user cohorts under four different propensity score (PS) adjustment / time-at-risk (TAR) designs across data sources. We report total patient exposure time in 1,000 patient-year (KYr) units, uncalibrated and calibrated hazard ratios (HRs) and their 95% confidence intervals (CIs). Greyed out entries did not pass study diagnostics and are included only for completeness.

| | Patients | | Exposure (KYr) | | Outcomes | | Incidence (per KYr) | |
|---|---|---|---|---|---|---|---|---|
| | T | C | T | C | T | C | T | C |
| **PS-matching / on-treatment TAR** | | | | | | | | |
| CCAE | 16,052 | 25,420 | 12.0 | 19.1 | 46 | 73 | 3.82 (2.80 - 5.10) | 3.82 (3.00 - 4.81) |
| DAG | 2,924 | 8,714 | 2.4 | 9.2 | <5 | 26 | < 2.10 (0.68 - 4.91) | 2.84 (1.85 - 4.16) |
| ODOD | 10,519 | 20,248 | 6.8 | 15.5 | 46 | 135 | 6.73 (4.93 - 8.97) | 8.74 (7.33 - 10.3) |
| MDCR | 1,288 | 4,765 | 0.9 | 4.0 | 0 | 25 | | 6.19 (4.00 - 9.13) |
| MDCD | 2,263 | 6,160 | 1.2 | 4.3 | 12 | 47 | 9.74 (5.03 - 17.0) | 11.0 (8.05 - 14.6) |
| Open Claims | 227,219 | 437,467 | 179.1 | 383.0 | 367 | 1,254 | 2.05 (1.84 - 2.27) | 3.27 (3.10 - 3.46) |
| OEHR | 14,853 | 33,249 | 4.7 | 17.5 | 17 | 131 | 3.63 (2.12 - 5.81) | 7.49 (6.26 - 8.89) |
| SIDIAP | 2,773 | 14,240 | 3.9 | 30.8 | 0 | 28 | | 0.91 (0.60 - 1.31) |
| Meta-analysis | 273,830 | 531,258 | 206.3 | 448.5 | <493 | 1,666 | < 2.39 (2.18 - 2.61) | 3.71 (3.54 - 3.90) |
| Meta-analysis (w/o USOC) | 46,611 | 93,791 | 27.2 | 65.5 | <126 | 412 | < 4.64 (3.86 - 5.52) | 6.29 (5.70 - 6.93) |
| **PS-stratification / on-treatment TAR** | | | | | | | | |
| CCAE | 18,425 | 25,454 | 13.3 | 19.1 | 50 | 73 | 3.76 (2.79 - 4.96) | 3.82 (2.99 - 4.80) |
| DAG | 3,226 | 8,750 | 2.5 | 9.2 | <5 | 26 | < 1.98 (0.64 - 4.62) | 2.82 (1.84 - 4.13) |
| ODOD | 12,580 | 20,448 | 7.7 | 15.6 | 50 | 137 | 6.50 (4.83 - 8.57) | 8.77 (7.36 - 10.4) |
| MDCR | 1,462 | 4,831 | 0.9 | 4.1 | 0 | 25 | | 6.12 (3.96 - 9.03) |
| MDCD | 2,349 | 6,175 | 1.3 | 4.3 | 12 | 47 | 9.54 (4.93 - 16.7) | 10.9 (8.02 - 14.5) |
| Open Claims | 270,936 | 438,007 | 200.8 | 383.5 | 416 | 1,258 | 2.07 (1.88 - 2.28) | 3.28 (3.10 - 3.47) |
| OEHR | 16,444 | 34,279 | 5.2 | 18.0 | 18 | 133 | 3.49 (2.07 - 5.52) | 7.38 (6.18 - 8.75) |
| SIDIAP | 2,950 | 14,724 | 4.0 | 32.3 | <5 | 29 | < 1.25 (0.40 - 2.91) | 0.90 (0.60 - 1.29) |
| Meta-analysis | 324,561 | 541,662 | 233.4 | 477.8 | <544 | 1,656 | < 2.33 (2.14 - 2.53) | 3.47 (3.30 - 3.64) |
| Meta-analysis (w/o USOC) | 53,625 | 103,655 | 32.7 | 94.3 | <128 | 398 | < 3.92 (3.27 - 4.66) | 4.22 (3.82 - 4.66) |
| **PS-matching / on-treatment TAR without escalation** | | | | | | | | |
| CCAE | 16,052 | 25,420 | 10.3 | 15.6 | 34 | 41 | 3.29 (2.28 - 4.60) | 2.63 (1.89 - 3.57) |
| DAG | 2,924 | 8,714 | 2.2 | 8.3 | <5 | 23 | < 2.31 (0.75 - 5.39) | 2.79 (1.77 - 4.18) |
| ODOD | 10,519 | 20,248 | 6.0 | 13.0 | 33 | 76 | 5.46 (3.76 - 7.66) | 5.84 (4.60 - 7.31) |
| MDCR | 1,288 | 4,765 | 0.8 | 3.5 | 0 | 16 | | 4.55 (2.60 - 7.38) |
| MDCD | 2,263 | 6,160 | 1.0 | 3.3 | 9 | 25 | 8.88 (4.06 - 16.9) | 7.47 (4.84 - 11.0) |
| Open Claims | 227,219 | 437,467 | 149.6 | 304.8 | 238 | 682 | 1.59 (1.39 - 1.81) | 2.24 (2.07 - 2.41) |
| OEHR | 14,853 | 33,249 | 4.1 | 14.1 | 9 | 63 | 2.21 (1.01 - 4.19) | 4.48 (3.44 - 5.73) |
| SIDIAP | 2,773 | 14,240 | 3.4 | 24.4 | 0 | 12 | | 0.49 (0.25 - 0.86) |
| Meta-analysis | 273,830 | 531,258 | 173.3 | 359.0 | <328 | 910 | < 1.89 (1.69 - 2.11) | 2.53 (2.37 - 2.70) |
| Meta-analysis (w/o USOC) | 46,611 | 93,791 | 23.6 | 54.3 | <90 | 228 | < 3.81 (3.06 - 4.68) | 4.20 (3.67 - 4.78) |
| **PS-stratification / on-treatment TAR without escalation** | | | | | | | | |
| CCAE | 18,425 | 25,454 | 11.5 | 15.6 | 37 | 41 | 3.22 (2.27 - 4.44) | 2.63 (1.89 - 3.57) |
| DAG | 3,226 | 8,750 | 2.3 | 8.3 | <5 | 23 | < 2.17 (0.71 - 5.07) | 2.77 (1.76 - 4.16) |
| ODOD | 12,580 | 20,448 | 6.8 | 13.2 | 37 | 77 | 5.42 (3.82 - 7.47) | 5.85 (4.62 - 7.31) |
| MDCR | 1,462 | 4,831 | 0.8 | 3.6 | 0 | 16 | | 4.50 (2.57 - 7.31) |
| MDCD | 2,349 | 6,175 | 1.0 | 3.4 | 9 | 25 | 8.68 (3.97 - 16.5) | 7.45 (4.82 - 11.0) |
| Open Claims | 270,936 | 438,007 | 168.9 | 305.2 | 277 | 684 | 1.64 (1.45 - 1.85) | 2.24 (2.08 - 2.42) |
| OEHR | 16,444 | 34,279 | 4.5 | 14.5 | 9 | 63 | 2.01 (0.92 - 3.81) | 4.34 (3.34 - 5.55) |
| SIDIAP | 2,950 | 14,724 | 3.5 | 25.6 | <5 | 12 | < 1.42 (0.46 - 3.31) | 0.47 (0.24 - 0.82) |
| Meta-analysis | 321,611 | 526,938 | 194.0 | 356.8 | <365 | 888 | < 1.88 (1.69 - 2.09) | 2.49 (2.33 - 2.66) |
| Meta-analysis (w/o USOC) | 50,675 | 88,931 | 25.1 | 51.6 | <88 | 204 | < 3.51 (2.81 - 4.32) | 3.96 (3.43 - 4.54) |

###### 4.2.19 Hypotension

**Supplementary Table 330:** Exposures, outcomes and risk estimates for Hypotension in SGLT2I (female) (T) vs DPP4I (female) (C) new-user cohorts under four different propensity score (PS) adjustment / time-at-risk (TAR) designs across data sources. We report total patient exposure time in 1,000 patient-year (KYr) units, uncalibrated and calibrated hazard ratios (HRs) and their 95% confidence intervals (CIs). Greyed out entries did not pass study diagnostics and are included only for completeness.

| | Patients | | Exposure (KYr) | | Outcomes | | Incidence (per KYr) | |
|---|---|---|---|---|---|---|---|---|
| | T | C | T | C | T | C | T | C |
| **PS-matching / on-treatment TAR** | | | | | | | | |
| CCAE | 16,059 | 25,318 | 12.0 | 18.9 | 99 | 150 | 8.23 (6.69 - 10.0) | 7.92 (6.70 - 9.29) |
| DAG | 2,827 | 8,489 | 2.3 | 8.9 | 6 | 54 | 2.62 (0.96 - 5.70) | 6.05 (4.55 - 7.90) |
| ODOD | 10,269 | 19,416 | 6.7 | 14.6 | 130 | 365 | 19.5 (16.3 - 23.2) | 24.9 (22.4 - 27.6) |
| MDCR | 1,244 | 4,557 | 0.8 | 3.9 | 25 | 93 | 29.5 (19.1 - 43.5) | 24.1 (19.4 - 29.5) |
| MDCD | 2,178 | 5,890 | 1.2 | 4.0 | 39 | 120 | 33.7 (24.0 - 46.1) | 29.8 (24.7 - 35.6) |
| Open Claims | 224,435 | 428,978 | 176.9 | 373.7 | 1,160 | 3,602 | 6.56 (6.18 - 6.95) | 9.64 (9.33 - 9.96) |
| OEHR | 14,728 | 32,933 | 4.6 | 17.2 | 57 | 256 | 12.3 (9.31 - 15.9) | 14.9 (13.1 - 16.8) |
| SIDIAP | 2,758 | 14,150 | 3.8 | 30.6 | 8 | 59 | 2.09 (0.90 - 4.12) | 1.93 (1.47 - 2.49) |
| Meta-analysis | 274,695 | 540,426 | 208.9 | 474.2 | <1,504 | 4,630 | < 7.20 (6.84 - 7.57) | 9.76 (9.48 - 10.0) |
| Meta-analysis (w/o USOC) | 50,260 | 111,448 | 32.0 | 100.5 | <344 | 1,028 | < 10.7 (9.64 - 11.9) | 10.2 (9.61 - 10.9) |
| **PS-stratification / on-treatment TAR** | | | | | | | | |
| CCAE | 18,471 | 25,349 | 13.3 | 19.0 | 110 | 150 | 8.25 (6.78 - 9.94) | 7.91 (6.70 - 9.28) |
| DAG | 3,116 | 8,520 | 2.4 | 9.0 | 7 | 55 | 2.88 (1.16 - 5.94) | 6.14 (4.62 - 7.99) |
| ODOD | 12,292 | 19,645 | 7.5 | 14.8 | 148 | 379 | 19.8 (16.7 - 23.2) | 25.6 (23.1 - 28.3) |
| MDCR | 1,415 | 4,606 | 0.9 | 3.9 | 25 | 98 | 27.6 (17.9 - 40.8) | 25.2 (20.4 - 30.7) |
| MDCD | 2,263 | 5,907 | 1.2 | 4.0 | 42 | 120 | 35.4 (25.5 - 47.9) | 29.6 (24.6 - 35.4) |
| Open Claims | 267,171 | 429,492 | 198.1 | 374.2 | 1,308 | 3,619 | 6.60 (6.25 - 6.97) | 9.67 (9.36 - 9.99) |
| OEHR | 16,280 | 33,949 | 5.1 | 17.7 | 60 | 269 | 11.8 (8.97 - 15.1) | 15.2 (13.4 - 17.1) |
| SIDIAP | 2,935 | 14,629 | 4.0 | 32.1 | 8 | 64 | 2.01 (0.87 - 3.95) | 2.00 (1.54 - 2.55) |
| Meta-analysis | 320,265 | 531,584 | 230.4 | 466.7 | 1,641 | 4,536 | 7.12 (6.78 - 7.47) | 9.72 (9.44 - 10.0) |
| Meta-analysis (w/o USOC) | 53,094 | 102,092 | 32.3 | 92.5 | 333 | 917 | 10.3 (9.22 - 11.5) | 9.91 (9.28 - 10.6) |
| **PS-matching / on-treatment TAR without escalation** | | | | | | | | |
| CCAE | 16,059 | 25,318 | 10.3 | 15.5 | 76 | 115 | 7.35 (5.79 - 9.20) | 7.44 (6.14 - 8.93) |
| DAG | 2,827 | 8,489 | 2.1 | 8.0 | 6 | 43 | 2.88 (1.06 - 6.27) | 5.35 (3.87 - 7.20) |
| ODOD | 10,269 | 19,416 | 5.9 | 12.4 | 110 | 308 | 18.7 (15.4 - 22.5) | 24.9 (22.2 - 27.9) |
| MDCR | 1,244 | 4,557 | 0.8 | 3.4 | 24 | 87 | 32.0 (20.5 - 47.6) | 25.8 (20.7 - 31.9) |
| MDCD | 2,178 | 5,890 | 1.0 | 3.2 | 29 | 80 | 30.2 (20.2 - 43.4) | 25.3 (20.1 - 31.5) |
| Open Claims | 224,435 | 428,978 | 147.9 | 297.3 | 936 | 2,869 | 6.33 (5.93 - 6.75) | 9.65 (9.30 - 10.0) |
| OEHR | 14,728 | 32,933 | 4.0 | 13.8 | 41 | 176 | 10.2 (7.29 - 13.8) | 12.7 (10.9 - 14.7) |
| SIDIAP | 2,758 | 14,150 | 3.4 | 24.2 | 7 | 47 | 2.08 (0.84 - 4.29) | 1.94 (1.43 - 2.58) |
| Meta-analysis | 274,695 | 540,426 | 175.9 | 379.4 | <1,210 | 3,659 | < 6.88 (6.50 - 7.28) | 9.64 (9.33 - 9.96) |
| Meta-analysis (w/o USOC) | 50,260 | 111,448 | 27.9 | 82.1 | <274 | 790 | < 9.81 (8.68 - 11.0) | 9.62 (8.96 - 10.3) |
| **PS-stratification / on-treatment TAR without escalation** | | | | | | | | |
| CCAE | 18,471 | 25,349 | 11.5 | 15.5 | 84 | 115 | 7.30 (5.82 - 9.04) | 7.43 (6.13 - 8.92) |
| DAG | 3,116 | 8,520 | 2.2 | 8.1 | 7 | 43 | 3.17 (1.27 - 6.53) | 5.32 (3.85 - 7.16) |
| ODOD | 12,292 | 19,645 | 6.6 | 12.5 | 126 | 320 | 19.0 (15.8 - 22.6) | 25.6 (22.8 - 28.5) |
| MDCR | 1,415 | 4,606 | 0.8 | 3.4 | 24 | 91 | 29.8 (19.1 - 44.4) | 26.8 (21.6 - 33.0) |
| MDCD | 2,263 | 5,907 | 1.0 | 3.2 | 32 | 80 | 32.5 (22.2 - 45.8) | 25.3 (20.0 - 31.4) |
| Open Claims | 267,171 | 429,492 | 166.7 | 297.7 | 1,066 | 2,886 | 6.40 (6.02 - 6.79) | 9.69 (9.34 - 10.1) |
| OEHR | 16,280 | 33,949 | 4.4 | 14.3 | 42 | 186 | 9.45 (6.81 - 12.8) | 13.0 (11.2 - 15.0) |
| SIDIAP | 2,935 | 14,629 | 3.5 | 25.4 | 7 | 51 | 2.00 (0.80 - 4.12) | 2.01 (1.50 - 2.64) |
| Meta-analysis | 320,265 | 531,584 | 195.0 | 373.5 | 1,332 | 3,601 | 6.83 (6.47 - 7.21) | 9.64 (9.33 - 9.96) |
| Meta-analysis (w/o USOC) | 53,094 | 102,092 | 28.3 | 75.8 | 266 | 715 | 9.39 (8.30 - 10.6) | 9.44 (8.76 - 10.2) |

##### 4.2.20 Joint pain

**Supplementary Table 331:** Exposures, outcomes and risk estimates for Joint pain in SGLT2I (female) (T) vs DPP4I (female) (C) new-user cohorts under four different propensity score (PS) adjustment / time-at-risk (TAR) designs across data sources. We report total patient exposure time in 1,000 patient-year (KYr) units, uncalibrated and calibrated hazard ratios (HRs) and their 95% confidence intervals (CIs). Greyed out entries did not pass study diagnostics and are included only for completeness.

| | Patients | | Exposure (KYr) | | Outcomes | | Incidence (per KYr) | |
|---|---|---|---|---|---|---|---|---|
| | T | C | T | C | T | C | T | C |
| **PS-matching / on-treatment TAR** | | | | | | | | |
| CCAE | 15,034 | 23,820 | 11.3 | 17.7 | 152 | 297 | 13.4 (11.4 - 15.8) | 16.8 (15.0 - 18.8) |
| DAG | 2,422 | 7,333 | 1.9 | 7.5 | 41 | 178 | 21.5 (15.4 - 29.1) | 23.6 (20.3 - 27.4) |
| ODOD | 9,854 | 18,883 | 6.4 | 14.2 | 112 | 286 | 17.6 (14.5 - 21.2) | 20.1 (17.9 - 22.6) |
| MDCR | 1,242 | 4,550 | 0.8 | 3.9 | 11 | 61 | 13.1 (6.55 - 23.5) | 15.8 (12.1 - 20.4) |
| MDCD | 2,068 | 5,622 | 1.1 | 3.8 | 30 | 122 | 26.6 (17.9 - 38.0) | 31.9 (26.5 - 38.1) |
| Open Claims | 213,048 | 411,581 | 168.2 | 358.5 | 1,309 | 3,498 | 7.78 (7.36 - 8.21) | 9.76 (9.44 - 10.1) |
| OEHR | 13,960 | 31,427 | 4.3 | 16.3 | 75 | 277 | 17.3 (13.6 - 21.6) | 17.0 (15.1 - 19.1) |
| SIDIAP | 1,982 | 10,479 | 2.8 | 22.0 | 58 | 606 | 21.0 (16.0 - 27.2) | 27.5 (25.3 - 29.8) |
| Meta-analysis | 259,656 | 513,802 | 197.3 | 445.5 | 1,784 | 5,325 | 9.04 (8.63 - 9.47) | 12.0 (11.6 - 12.3) |
| Meta-analysis (w/o USOC) | 46,608 | 102,221 | 29.1 | 87.0 | 475 | 1,827 | 16.3 (14.9 - 17.9) | 21.0 (20.0 - 22.0) |
| **PS-stratification / on-treatment TAR** | | | | | | | | |
| CCAE | 17,271 | 23,853 | 12.5 | 17.7 | 175 | 297 | 14.0 (12.0 - 16.2) | 16.8 (14.9 - 18.8) |
| DAG | 2,670 | 7,357 | 2.0 | 7.6 | 46 | 179 | 22.6 (16.6 - 30.2) | 23.7 (20.3 - 27.4) |
| ODOD | 11,750 | 19,065 | 7.2 | 14.4 | 129 | 288 | 18.0 (15.1 - 21.4) | 20.1 (17.8 - 22.5) |
| MDCR | 1,402 | 4,613 | 0.9 | 3.9 | 14 | 61 | 15.7 (8.57 - 26.3) | 15.7 (12.0 - 20.1) |
| MDCD | 2,157 | 5,641 | 1.2 | 3.8 | 31 | 123 | 26.8 (18.2 - 38.0) | 32.0 (26.6 - 38.2) |
| Open Claims | 253,928 | 411,935 | 188.5 | 358.8 | 1,465 | 3,504 | 7.77 (7.38 - 8.18) | 9.77 (9.45 - 10.1) |
| OEHR | 15,387 | 32,362 | 4.8 | 16.8 | 80 | 288 | 16.8 (13.3 - 20.9) | 17.2 (15.3 - 19.3) |
| SIDIAP | 2,113 | 10,863 | 2.9 | 23.1 | 59 | 654 | 20.5 (15.6 - 26.5) | 28.3 (26.1 - 30.5) |
| Meta-analysis | 303,119 | 505,435 | 217.8 | 438.3 | 1,954 | 5,210 | 8.97 (8.58 - 9.38) | 11.9 (11.6 - 12.2) |
| Meta-analysis (w/o USOC) | 49,191 | 93,500 | 29.3 | 79.5 | 489 | 1,706 | 16.7 (15.2 - 18.2) | 21.5 (20.5 - 22.5) |
| **PS-matching / on-treatment TAR without escalation** | | | | | | | | |
| CCAE | 15,034 | 23,820 | 9.7 | 14.4 | 129 | 241 | 13.3 (11.1 - 15.8) | 16.7 (14.6 - 18.9) |
| DAG | 2,422 | 7,333 | 1.7 | 6.8 | 39 | 167 | 22.6 (16.1 - 30.9) | 24.6 (21.0 - 28.7) |
| ODOD | 9,854 | 18,883 | 5.6 | 12.0 | 97 | 238 | 17.2 (14.0 - 21.0) | 19.9 (17.5 - 22.6) |
| MDCR | 1,242 | 4,550 | 0.7 | 3.4 | 10 | 52 | 13.4 (6.41 - 24.6) | 15.5 (11.6 - 20.3) |
| MDCD | 2,068 | 5,622 | 0.9 | 3.0 | 27 | 97 | 29.0 (19.1 - 42.2) | 32.5 (26.4 - 39.7) |
| Open Claims | 213,048 | 411,581 | 140.6 | 285.4 | 1,098 | 2,775 | 7.81 (7.35 - 8.28) | 9.72 (9.37 - 10.1) |
| OEHR | 13,960 | 31,427 | 3.8 | 13.1 | 68 | 215 | 17.9 (13.9 - 22.7) | 16.4 (14.3 - 18.7) |
| SIDIAP | 1,982 | 10,479 | 2.4 | 17.5 | 44 | 465 | 18.1 (13.2 - 24.3) | 26.6 (24.2 - 29.1) |
| Meta-analysis | 259,656 | 513,802 | 166.0 | 356.6 | 1,509 | 4,247 | 9.09 (8.64 - 9.56) | 11.9 (11.6 - 12.3) |
| Meta-analysis (w/o USOC) | 46,608 | 102,221 | 25.4 | 71.2 | 411 | 1,472 | 16.2 (14.7 - 17.9) | 20.7 (19.6 - 21.8) |
| **PS-stratification / on-treatment TAR without escalation** | | | | | | | | |
| CCAE | 17,271 | 23,853 | 10.8 | 14.5 | 150 | 241 | 13.9 (11.8 - 16.3) | 16.7 (14.6 - 18.9) |
| DAG | 2,670 | 7,357 | 1.8 | 6.8 | 44 | 167 | 23.9 (17.4 - 32.1) | 24.5 (20.9 - 28.5) |
| ODOD | 11,750 | 19,065 | 6.3 | 12.1 | 113 | 240 | 17.8 (14.7 - 21.4) | 19.8 (17.4 - 22.5) |
| MDCR | 1,402 | 4,613 | 0.8 | 3.4 | 13 | 52 | 16.3 (8.66 - 27.8) | 15.3 (11.5 - 20.1) |
| MDCD | 2,157 | 5,641 | 1.0 | 3.0 | 28 | 97 | 29.2 (19.4 - 42.3) | 32.4 (26.3 - 39.5) |
| Open Claims | 253,928 | 411,935 | 158.6 | 285.6 | 1,233 | 2,781 | 7.77 (7.35 - 8.22) | 9.74 (9.38 - 10.1) |
| OEHR | 15,387 | 32,362 | 4.2 | 13.5 | 72 | 225 | 17.3 (13.6 - 21.8) | 16.6 (14.5 - 19.0) |
| SIDIAP | 2,113 | 10,863 | 2.5 | 18.4 | 45 | 505 | 17.7 (12.9 - 23.7) | 27.4 (25.1 - 29.9) |
| Meta-analysis | 303,119 | 505,435 | 184.3 | 350.9 | 1,657 | 4,159 | 8.99 (8.56 - 9.44) | 11.9 (11.5 - 12.2) |
| Meta-analysis (w/o USOC) | 49,191 | 93,500 | 25.7 | 65.3 | 424 | 1,378 | 16.5 (15.0 - 18.2) | 21.1 (20.0 - 22.3) |

##### 4.2.21 Lower extremity amputation

**Supplementary Table 332:** Exposures, outcomes and risk estimates for Lower extremity amputation in SGLT2I (female) (T) vs DPP4I (female) (C) new-user cohorts under four different propensity score (PS) adjustment / time-at-risk (TAR) designs across data sources. We report total patient exposure time in 1,000 patient-year (KYr) units, uncalibrated and calibrated hazard ratios (HRs) and their 95% confidence intervals (CIs). Greyed out entries did not pass study diagnostics and are included only for completeness.

| | Patients | | Exposure (KYr) | | Outcomes | | Incidence (per KYr) | |
|---|---|---|---|---|---|---|---|---|
| | T | C | T | C | T | C | T | C |
| **PS-matching / on-treatment TAR** | | | | | | | | |
| CCAE | 16,405 | 25,915 | 12.3 | 19.5 | 5 | <5 | 0.41 (0.13 - 0.95) | < 0.26 (0.08 - 0.60) |
| DAG | 2,939 | 8,792 | 2.4 | 9.3 | 0 | 0 | | |
| ODOD | 10,719 | 20,618 | 7.0 | 15.8 | 5 | 6 | 0.72 (0.23 - 1.67) | 0.38 (0.14 - 0.83) |
| MDCR | 1,302 | 4,829 | 0.9 | 4.1 | <5 | <5 | < 5.67 (1.84 - 13.2) | < 1.22 (0.39 - 2.84) |
| MDCD | 2,339 | 6,352 | 1.3 | 4.4 | 0 | <5 | | < 1.13 (0.37 - 2.63) |
| Open Claims | 231,643 | 445,651 | 182.7 | 390.7 | 35 | 108 | 0.19 (0.13 - 0.27) | 0.28 (0.23 - 0.33) |
| OEHR | 15,057 | 33,693 | 4.8 | 17.8 | <5 | 7 | < 1.05 (0.34 - 2.45) | 0.39 (0.16 - 0.81) |
| SIDIAP | 2,778 | 14,267 | 3.9 | 30.9 | 0 | <5 | | < 0.16 (0.05 - 0.38) |
| Meta-analysis | 231,643 | 445,651 | 182.7 | 390.7 | 35 | 108 | 0.19 (0.13 - 0.27) | 0.28 (0.23 - 0.33) |
| **PS-stratification / on-treatment TAR** | | | | | | | | |
| CCAE | 18,874 | 25,951 | 13.7 | 19.5 | 5 | <5 | 0.37 (0.12 - 0.85) | < 0.26 (0.08 - 0.60) |
| DAG | 3,243 | 8,833 | 2.5 | 9.3 | 0 | 0 | | |
| ODOD | 12,831 | 20,824 | 7.9 | 15.9 | 5 | 6 | 0.64 (0.21 - 1.48) | 0.38 (0.14 - 0.82) |
| MDCR | 1,475 | 4,896 | 0.9 | 4.2 | <5 | <5 | < 5.31 (1.73 - 12.4) | < 1.20 (0.39 - 2.81) |
| MDCD | 2,431 | 6,371 | 1.3 | 4.5 | 0 | 5 | | 1.12 (0.36 - 2.61) |
| Open Claims | 276,304 | 446,087 | 204.7 | 391.1 | 39 | 108 | 0.19 (0.14 - 0.26) | 0.28 (0.23 - 0.33) |
| OEHR | 16,654 | 34,735 | 5.2 | 18.4 | <5 | 7 | < 0.96 (0.31 - 2.23) | 0.38 (0.15 - 0.79) |
| SIDIAP | 2,956 | 14,749 | 4.0 | 32.4 | 0 | <5 | | < 0.15 (0.05 - 0.36) |
| Meta-analysis | 276,304 | 446,087 | 204.7 | 391.1 | 39 | 108 | 0.19 (0.14 - 0.26) | 0.28 (0.23 - 0.33) |
| **PS-matching / on-treatment TAR without escalation** | | | | | | | | |
| CCAE | 16,405 | 25,915 | 10.6 | 15.9 | <5 | <5 | < 0.47 (0.15 - 1.10) | < 0.31 (0.10 - 0.73) |
| DAG | 2,939 | 8,792 | 2.2 | 8.3 | 0 | 0 | | |
| ODOD | 10,719 | 20,618 | 6.2 | 13.2 | <5 | 6 | < 0.81 (0.26 - 1.89) | 0.45 (0.17 - 0.99) |
| MDCR | 1,302 | 4,829 | 0.8 | 3.6 | <5 | <5 | < 6.41 (2.08 - 15.0) | < 1.40 (0.45 - 3.26) |
| MDCD | 2,339 | 6,352 | 1.1 | 3.5 | 0 | <5 | | < 1.45 (0.47 - 3.38) |
| Open Claims | 231,643 | 445,651 | 152.5 | 310.5 | 25 | 86 | 0.16 (0.11 - 0.24) | 0.28 (0.22 - 0.34) |
| OEHR | 15,057 | 33,693 | 4.1 | 14.3 | <5 | <5 | < 1.21 (0.39 - 2.82) | < 0.35 (0.11 - 0.82) |
| SIDIAP | 2,778 | 14,267 | 3.4 | 24.4 | 0 | <5 | | < 0.20 (0.07 - 0.48) |
| Meta-analysis | 231,643 | 445,651 | 152.5 | 310.5 | 25 | 86 | 0.16 (0.11 - 0.24) | 0.28 (0.22 - 0.34) |
| **PS-stratification / on-treatment TAR without escalation** | | | | | | | | |
| CCAE | 18,874 | 25,951 | 11.8 | 15.9 | <5 | <5 | < 0.42 (0.14 - 0.99) | < 0.31 (0.10 - 0.73) |
| DAG | 3,243 | 8,833 | 2.3 | 8.4 | 0 | 0 | | |
| ODOD | 12,831 | 20,824 | 7.0 | 13.4 | <5 | 6 | < 0.72 (0.23 - 1.67) | 0.45 (0.16 - 0.97) |
| MDCR | 1,475 | 4,896 | 0.8 | 3.6 | <5 | <5 | < 6.00 (1.95 - 14.0) | < 1.38 (0.45 - 3.23) |
| MDCD | 2,431 | 6,371 | 1.1 | 3.5 | 0 | <5 | | < 1.44 (0.47 - 3.37) |
| Open Claims | 276,304 | 446,087 | 172.1 | 310.9 | 27 | 86 | 0.16 (0.10 - 0.23) | 0.28 (0.22 - 0.34) |
| OEHR | 16,654 | 34,735 | 4.6 | 14.8 | <5 | <5 | < 1.10 (0.36 - 2.56) | < 0.34 (0.11 - 0.79) |
| SIDIAP | 2,956 | 14,749 | 3.5 | 25.6 | 0 | <5 | | < 0.20 (0.06 - 0.46) |
| Meta-analysis | 276,304 | 446,087 | 172.1 | 310.9 | 27 | 86 | 0.16 (0.10 - 0.23) | 0.28 (0.22 - 0.34) |

##### 4.2.22 Nausea

**Supplementary Table 333:** Exposures, outcomes and risk estimates for Nausea in SGLT2I (female) (T) vs DPP4I (female) (C) new-user cohorts under four different propensity score (PS) adjustment / time-at-risk (TAR) designs across data sources. We report total patient exposure time in 1,000 patient-year (KYr) units, uncalibrated and calibrated hazard ratios (HRs) and their 95% confidence intervals (CIs). Greyed out entries did not pass study diagnostics and are included only for completeness.

| | Patients | | Exposure (KYr) | | Outcomes | | Incidence (per KYr) | |
|---|---|---|---|---|---|---|---|---|
| | T | C | T | C | T | C | T | C |
| **PS-matching / on-treatment TAR** | | | | | | | | |
| CCAE | 13,325 | 21,231 | 9.9 | 15.6 | 461 | 771 | 46.6 (42.5 - 51.1) | 49.3 (45.9 - 52.9) |
| DAG | 2,604 | 7,941 | 2.1 | 8.3 | 27 | 114 | 12.9 (8.48 - 18.7) | 13.8 (11.4 - 16.6) |
| ODOD | 8,575 | 16,310 | 5.5 | 12.1 | 276 | 718 | 50.2 (44.4 - 56.5) | 59.4 (55.1 - 63.9) |
| MDCR | 1,099 | 4,068 | 0.7 | 3.4 | 30 | 178 | 40.4 (27.3 - 57.7) | 53.0 (45.5 - 61.4) |
| MDCD | 1,504 | 4,121 | 0.8 | 2.7 | 79 | 324 | 97.5 (77.2 - 122) | 119 (106 - 133) |
| Open Claims | 186,937 | 363,943 | 148.2 | 316.2 | 3,541 | 9,321 | 23.9 (23.1 - 24.7) | 29.5 (28.9 - 30.1) |
| OEHR | 12,664 | 28,430 | 3.9 | 14.5 | 211 | 742 | 54.4 (47.3 - 62.2) | 51.2 (47.6 - 55.0) |
| SIDIAP | 2,595 | 13,378 | 3.6 | 28.9 | 18 | 287 | 5.01 (2.97 - 7.92) | 9.93 (8.81 - 11.1) |
| Meta-analysis | 229,573 | 460,318 | 175.3 | 404.2 | 4,623 | 12,303 | 26.4 (25.6 - 27.1) | 30.4 (29.9 - 31.0) |
| Meta-analysis (w/o USOC) | 42,636 | 96,375 | 27.1 | 88.1 | 1,082 | 2,982 | 39.9 (37.6 - 42.4) | 33.9 (32.7 - 35.1) |
| **PS-stratification / on-treatment TAR** | | | | | | | | |
| CCAE | 15,258 | 21,256 | 10.9 | 15.7 | 515 | 774 | 47.1 (43.2 - 51.4) | 49.4 (46.0 - 53.1) |
| DAG | 2,873 | 7,967 | 2.2 | 8.3 | 27 | 114 | 12.1 (8.00 - 17.7) | 13.7 (11.3 - 16.5) |
| ODOD | 10,200 | 16,497 | 6.2 | 12.2 | 321 | 730 | 51.9 (46.4 - 57.9) | 59.7 (55.4 - 64.2) |
| MDCR | 1,249 | 4,115 | 0.8 | 3.4 | 32 | 179 | 40.6 (27.8 - 57.4) | 52.8 (45.3 - 61.1) |
| MDCD | 1,543 | 4,133 | 0.8 | 2.7 | 80 | 325 | 97.2 (77.1 - 121) | 119 (106 - 132) |
| Open Claims | 221,452 | 364,477 | 165.5 | 316.7 | 3,960 | 9,346 | 23.9 (23.2 - 24.7) | 29.5 (28.9 - 30.1) |
| OEHR | 14,004 | 29,486 | 4.3 | 15.0 | 230 | 773 | 53.8 (47.1 - 61.2) | 51.5 (48.0 - 55.3) |
| SIDIAP | 2,767 | 13,824 | 3.7 | 30.3 | 21 | 300 | 5.60 (3.47 - 8.56) | 9.90 (8.81 - 11.1) |
| Meta-analysis | 266,554 | 453,507 | 192.8 | 398.2 | 5,074 | 12,037 | 26.3 (25.6 - 27.1) | 30.2 (29.7 - 30.8) |
| Meta-analysis (w/o USOC) | 45,102 | 89,030 | 27.4 | 81.5 | 1,114 | 2,691 | 40.7 (38.4 - 43.2) | 33.0 (31.8 - 34.3) |
| **PS-matching / on-treatment TAR without escalation** | | | | | | | | |
| CCAE | 13,325 | 21,231 | 8.6 | 12.9 | 400 | 637 | 46.6 (42.2 - 51.4) | 49.5 (45.7 - 53.5) |
| DAG | 2,604 | 7,941 | 1.9 | 7.4 | 23 | 100 | 12.1 (7.65 - 18.1) | 13.4 (10.9 - 16.4) |
| ODOD | 8,575 | 16,310 | 4.9 | 10.3 | 244 | 607 | 49.7 (43.7 - 56.4) | 59.1 (54.5 - 64.0) |
| MDCR | 1,099 | 4,068 | 0.7 | 2.9 | 27 | 149 | 41.3 (27.2 - 60.1) | 50.7 (42.9 - 59.5) |
| MDCD | 1,504 | 4,121 | 0.7 | 2.2 | 64 | 251 | 93.3 (71.8 - 119) | 115 (101 - 130) |
| Open Claims | 186,937 | 363,943 | 124.9 | 253.3 | 2,881 | 7,391 | 23.1 (22.2 - 23.9) | 29.2 (28.5 - 29.9) |
| OEHR | 12,664 | 28,430 | 3.4 | 11.7 | 174 | 565 | 51.0 (43.7 - 59.2) | 48.1 (44.2 - 52.2) |
| SIDIAP | 2,595 | 13,378 | 3.2 | 22.9 | 14 | 220 | 4.43 (2.42 - 7.43) | 9.59 (8.37 - 10.9) |
| Meta-analysis | 229,573 | 460,318 | 148.7 | 325.6 | 3,809 | 9,796 | 25.6 (24.8 - 26.4) | 30.1 (29.5 - 30.7) |
| Meta-analysis (w/o USOC) | 42,636 | 96,375 | 23.8 | 72.3 | 928 | 2,405 | 38.9 (36.5 - 41.5) | 33.3 (32.0 - 34.6) |
| **PS-stratification / on-treatment TAR without escalation** | | | | | | | | |
| CCAE | 15,258 | 21,256 | 9.5 | 12.9 | 450 | 640 | 47.3 (43.1 - 51.9) | 49.7 (45.9 - 53.7) |
| DAG | 2,873 | 7,967 | 2.0 | 7.5 | 23 | 100 | 11.4 (7.22 - 17.1) | 13.4 (10.9 - 16.3) |
| ODOD | 10,200 | 16,497 | 5.5 | 10.4 | 277 | 618 | 50.1 (44.3 - 56.3) | 59.4 (54.8 - 64.3) |
| MDCR | 1,249 | 4,115 | 0.7 | 3.0 | 29 | 150 | 41.6 (27.9 - 59.8) | 50.6 (42.8 - 59.4) |
| MDCD | 1,543 | 4,133 | 0.7 | 2.2 | 65 | 252 | 93.0 (71.8 - 118) | 115 (102 - 130) |
| Open Claims | 221,452 | 364,477 | 140.3 | 253.7 | 3,228 | 7,416 | 23.0 (22.2 - 23.8) | 29.2 (28.6 - 29.9) |
| OEHR | 14,004 | 29,486 | 3.8 | 12.2 | 190 | 589 | 50.6 (43.7 - 58.3) | 48.4 (44.6 - 52.5) |
| SIDIAP | 2,767 | 13,824 | 3.3 | 24.1 | 16 | 229 | 4.85 (2.77 - 7.87) | 9.51 (8.32 - 10.8) |
| Meta-analysis | 266,554 | 453,507 | 164.4 | 320.7 | 4,184 | 9,592 | 25.5 (24.7 - 26.2) | 29.9 (29.3 - 30.5) |
| Meta-analysis (w/o USOC) | 45,102 | 89,030 | 24.1 | 67.0 | 956 | 2,176 | 39.6 (37.2 - 42.2) | 32.5 (31.1 - 33.9) |

##### 4.2.23 Peripheral edema

**Supplementary Table 334:** Exposures, outcomes and risk estimates for Peripheral edema in SGLT2I (female) (T) vs DPP4I (female) (C) new-user cohorts under four different propensity score (PS) adjustment / time-at-risk (TAR) designs across data sources. We report total patient exposure time in 1,000 patient-year (KYr) units, uncalibrated and calibrated hazard ratios (HRs) and their 95% confidence intervals (CIs). Greyed out entries did not pass study diagnostics and are included only for completeness.

| | Patients | | Exposure (KYr) | | Outcomes | | Incidence (per KYr) | |
|---|---|---|---|---|---|---|---|---|
| | T | C | T | C | T | C | T | C |
| **PS-matching / on-treatment TAR** | | | | | | | | |
| CCAE | 12,430 | 19,749 | 9.1 | 14.4 | 392 | 705 | 43.0 (38.8 - 47.5) | 49.1 (45.5 - 52.8) |
| DAG | 2,349 | 7,030 | 1.9 | 7.2 | 31 | 264 | 16.3 (11.0 - 23.1) | 36.6 (32.3 - 41.3) |
| ODOD | 7,836 | 14,517 | 4.9 | 10.3 | 290 | 992 | 58.6 (52.1 - 65.8) | 96.5 (90.6 - 103) |
| MDCR | 947 | 3,271 | 0.6 | 2.6 | 28 | 253 | 44.5 (29.6 - 64.3) | 97.1 (85.5 - 110) |
| MDCD | 1,489 | 4,025 | 0.8 | 2.5 | 68 | 292 | 89.6 (69.5 - 114) | 115 (102 - 129) |
| Open Claims | 170,651 | 326,752 | 134.0 | 276.5 | 3,312 | 11,135 | 24.7 (23.9 - 25.6) | 40.3 (39.5 - 41.0) |
| OEHR | 12,122 | 27,295 | 3.8 | 13.6 | 150 | 721 | 39.9 (33.7 - 46.8) | 52.9 (49.1 - 56.9) |
| SIDIAP | 2,316 | 12,341 | 3.3 | 26.1 | 45 | 587 | 13.7 (10.0 - 18.4) | 22.5 (20.7 - 24.3) |
| Meta-analysis | 210,322 | 415,605 | 158.9 | 355.3 | 4,299 | 14,785 | 27.1 (26.3 - 27.9) | 41.6 (40.9 - 42.3) |
| Meta-analysis (w/o USOC) | 39,671 | 88,853 | 24.9 | 78.8 | 987 | 3,650 | 39.7 (37.3 - 42.3) | 46.3 (44.8 - 47.9) |
| **PS-stratification / on-treatment TAR** | | | | | | | | |
| CCAE | 14,270 | 19,774 | 10.1 | 14.4 | 434 | 706 | 43.2 (39.2 - 47.4) | 49.1 (45.5 - 52.8) |
| DAG | 2,574 | 7,105 | 2.0 | 7.3 | 33 | 272 | 16.4 (11.3 - 23.1) | 37.2 (32.9 - 41.9) |
| ODOD | 9,264 | 14,641 | 5.5 | 10.4 | 325 | 1,009 | 58.9 (52.6 - 65.6) | 97.4 (91.5 - 104) |
| MDCR | 1,079 | 3,319 | 0.7 | 2.6 | 30 | 262 | 44.7 (30.1 - 63.7) | 99.5 (87.8 - 112) |
| MDCD | 1,536 | 4,052 | 0.8 | 2.6 | 69 | 299 | 89.1 (69.3 - 113) | 117 (104 - 131) |
| Open Claims | 201,879 | 327,260 | 149.5 | 276.9 | 3,614 | 11,193 | 24.2 (23.4 - 25.0) | 40.4 (39.7 - 41.2) |
| OEHR | 13,269 | 28,054 | 4.1 | 14.0 | 157 | 756 | 38.5 (32.7 - 45.0) | 54.0 (50.2 - 58.0) |
| SIDIAP | 2,465 | 12,766 | 3.4 | 27.4 | 48 | 623 | 14.1 (10.4 - 18.7) | 22.7 (21.0 - 24.6) |
| Meta-analysis | 243,721 | 409,600 | 174.6 | 350.4 | 4,611 | 14,559 | 26.4 (25.6 - 27.2) | 41.6 (40.9 - 42.2) |
| Meta-analysis (w/o USOC) | 41,842 | 82,340 | 25.1 | 73.4 | 997 | 3,366 | 39.8 (37.3 - 42.3) | 45.8 (44.3 - 47.4) |
| **PS-matching / on-treatment TAR without escalation** | | | | | | | | |
| CCAE | 12,430 | 19,749 | 7.9 | 11.8 | 334 | 587 | 42.5 (38.1 - 47.3) | 49.7 (45.7 - 53.9) |
| DAG | 2,349 | 7,030 | 1.7 | 6.5 | 29 | 241 | 16.8 (11.2 - 24.1) | 37.1 (32.5 - 42.0) |
| ODOD | 7,836 | 14,517 | 4.4 | 8.7 | 254 | 823 | 57.7 (50.8 - 65.2) | 94.4 (88.0 - 101) |
| MDCR | 947 | 3,271 | 0.6 | 2.3 | 26 | 223 | 45.8 (29.9 - 67.1) | 97.8 (85.4 - 111) |
| MDCD | 1,489 | 4,025 | 0.6 | 2.0 | 60 | 221 | 95.8 (73.1 - 123) | 109 (94.9 - 124) |
| Open Claims | 170,651 | 326,752 | 112.6 | 221.6 | 2,729 | 8,869 | 24.2 (23.3 - 25.2) | 40.0 (39.2 - 40.9) |
| OEHR | 12,122 | 27,295 | 3.3 | 11.1 | 126 | 555 | 38.4 (32.0 - 45.7) | 50.0 (46.0 - 54.4) |
| SIDIAP | 2,316 | 12,341 | 2.9 | 20.8 | 39 | 463 | 13.5 (9.57 - 18.4) | 22.3 (20.3 - 24.4) |
| Meta-analysis | 210,322 | 415,605 | 134.4 | 286.2 | 3,576 | 11,832 | 26.6 (25.7 - 27.5) | 41.3 (40.6 - 42.1) |
| Meta-analysis (w/o USOC) | 39,671 | 88,853 | 21.8 | 64.7 | 847 | 2,963 | 38.9 (36.3 - 41.6) | 45.8 (44.2 - 47.5) |
| **PS-stratification / on-treatment TAR without escalation** | | | | | | | | |
| CCAE | 14,270 | 19,774 | 8.7 | 11.8 | 372 | 588 | 42.7 (38.5 - 47.3) | 49.7 (45.8 - 53.9) |
| DAG | 2,574 | 7,105 | 1.8 | 6.6 | 31 | 248 | 17.0 (11.6 - 24.1) | 37.7 (33.1 - 42.7) |
| ODOD | 9,264 | 14,641 | 4.9 | 8.8 | 288 | 839 | 58.3 (51.8 - 65.5) | 95.5 (89.1 - 102) |
| MDCR | 1,079 | 3,319 | 0.6 | 2.3 | 28 | 229 | 46.1 (30.6 - 66.7) | 99.3 (86.9 - 113) |
| MDCD | 1,536 | 4,052 | 0.6 | 2.0 | 60 | 228 | 93.9 (71.6 - 121) | 111 (97.4 - 127) |
| Open Claims | 201,879 | 327,260 | 126.4 | 221.9 | 2,985 | 8,914 | 23.6 (22.8 - 24.5) | 40.2 (39.3 - 41.0) |
| OEHR | 13,269 | 28,054 | 3.6 | 11.4 | 131 | 580 | 36.8 (30.8 - 43.7) | 50.9 (46.9 - 55.2) |
| SIDIAP | 2,465 | 12,766 | 3.0 | 21.8 | 41 | 493 | 13.6 (9.76 - 18.5) | 22.6 (20.7 - 24.7) |
| Meta-analysis | 243,721 | 409,600 | 148.5 | 282.3 | 3,848 | 11,662 | 25.9 (25.1 - 26.7) | 41.3 (40.6 - 42.1) |
| Meta-analysis (w/o USOC) | 41,842 | 82,340 | 22.0 | 60.4 | 863 | 2,748 | 39.1 (36.6 - 41.8) | 45.5 (43.8 - 47.2) |

##### 4.2.24 Photosensitivity

**Supplementary Table 335:** Exposures, outcomes and risk estimates for Photosensitivity in SGLT2I (female) (T) vs DPP4I (female) (C) new-user cohorts under four different propensity score (PS) adjustment / time-at-risk (TAR) designs across data sources. We report total patient exposure time in 1,000 patient-year (KYr) units, uncalibrated and calibrated hazard ratios (HRs) and their 95% confidence intervals (CIs). Greyed out entries did not pass study diagnostics and are included only for completeness.

| | Patients | | Exposure (KYr) | | Outcomes | | Incidence (per KYr) | |
|---|---|---|---|---|---|---|---|---|
| | T | C | T | C | T | C | T | C |
| **PS-matching / on-treatment TAR** | | | | | | | | |
| CCAE | 16,226 | 25,629 | 12.2 | 19.2 | 9 | 14 | 0.74 (0.34 - 1.40) | 0.73 (0.40 - 1.22) |
| DAG | 2,924 | 8,758 | 2.4 | 9.2 | <5 | <5 | < 2.11 (0.68 - 4.92) | < 0.54 (0.18 - 1.26) |
| ODOD | 10,639 | 20,438 | 6.9 | 15.6 | <5 | 17 | < 0.72 (0.23 - 1.69) | 1.09 (0.64 - 1.75) |
| MDCR | 1,291 | 4,759 | 0.9 | 4.0 | <5 | 7 | < 5.70 (1.85 - 13.3) | 1.73 (0.70 - 3.57) |
| MDCD | 2,325 | 6,314 | 1.3 | 4.4 | <5 | 6 | < 3.93 (1.28 - 9.17) | 1.36 (0.50 - 2.97) |
| Open Claims | 229,224 | 441,027 | 180.6 | 386.2 | 67 | 195 | 0.37 (0.29 - 0.47) | 0.50 (0.44 - 0.58) |
| OEHR | 14,958 | 33,449 | 4.7 | 17.7 | <5 | 30 | < 1.06 (0.34 - 2.47) | 1.70 (1.15 - 2.42) |
| SIDIAP | 2,767 | 14,239 | 3.9 | 30.8 | 0 | 7 | | 0.23 (0.09 - 0.47) |
| Meta-analysis | 271,047 | 520,543 | 204.4 | 438.7 | <86 | 256 | < 0.42 (0.34 - 0.52) | 0.58 (0.51 - 0.66) |
| Meta-analysis (w/o USOC) | 41,823 | 79,516 | 23.8 | 52.5 | <19 | 61 | < 0.80 (0.48 - 1.25) | 1.16 (0.89 - 1.49) |
| **PS-stratification / on-treatment TAR** | | | | | | | | |
| CCAE | 18,681 | 25,665 | 13.5 | 19.3 | 11 | 14 | 0.81 (0.41 - 1.46) | 0.73 (0.40 - 1.22) |
| DAG | 3,228 | 8,796 | 2.5 | 9.3 | <5 | <5 | < 1.98 (0.64 - 4.63) | < 0.54 (0.17 - 1.26) |
| ODOD | 12,737 | 20,640 | 7.8 | 15.8 | <5 | 17 | < 0.64 (0.21 - 1.50) | 1.08 (0.63 - 1.73) |
| MDCR | 1,462 | 4,825 | 0.9 | 4.1 | <5 | 7 | < 5.35 (1.74 - 12.5) | 1.71 (0.69 - 3.52) |
| MDCD | 2,416 | 6,333 | 1.3 | 4.4 | <5 | 6 | < 3.84 (1.25 - 8.97) | 1.35 (0.50 - 2.95) |
| Open Claims | 273,499 | 441,578 | 202.5 | 386.7 | 72 | 195 | 0.36 (0.28 - 0.45) | 0.50 (0.44 - 0.58) |
| OEHR | 16,535 | 34,484 | 5.2 | 18.2 | <5 | 30 | < 0.96 (0.31 - 2.25) | 1.65 (1.11 - 2.35) |
| SIDIAP | 2,944 | 14,723 | 4.0 | 32.3 | 0 | 8 | | 0.25 (0.11 - 0.49) |
| Meta-analysis | 321,452 | 522,367 | 229.0 | 439.9 | <93 | 256 | < 0.41 (0.33 - 0.50) | 0.58 (0.51 - 0.66) |
| Meta-analysis (w/o USOC) | 47,953 | 80,789 | 26.5 | 53.2 | <21 | 61 | < 0.79 (0.49 - 1.21) | 1.15 (0.88 - 1.47) |
| **PS-matching / on-treatment TAR without escalation** | | | | | | | | |
| CCAE | 16,226 | 25,629 | 10.4 | 15.7 | 9 | 13 | 0.86 (0.39 - 1.64) | 0.83 (0.44 - 1.42) |
| DAG | 2,924 | 8,758 | 2.2 | 8.3 | <5 | <5 | < 2.32 (0.75 - 5.41) | < 0.60 (0.20 - 1.41) |
| ODOD | 10,639 | 20,438 | 6.1 | 13.1 | <5 | 16 | < 0.82 (0.27 - 1.91) | 1.22 (0.70 - 1.98) |
| MDCR | 1,291 | 4,759 | 0.8 | 3.5 | <5 | 6 | < 6.46 (2.10 - 15.1) | 1.70 (0.63 - 3.71) |
| MDCD | 2,325 | 6,314 | 1.0 | 3.4 | <5 | <5 | < 4.77 (1.55 - 11.1) | < 1.46 (0.47 - 3.40) |
| Open Claims | 229,224 | 441,027 | 150.8 | 306.9 | 55 | 168 | 0.36 (0.27 - 0.47) | 0.55 (0.47 - 0.64) |
| OEHR | 14,958 | 33,449 | 4.1 | 14.2 | <5 | 26 | < 1.22 (0.39 - 2.84) | 1.83 (1.20 - 2.68) |
| SIDIAP | 2,767 | 14,239 | 3.4 | 24.4 | 0 | 6 | | 0.25 (0.09 - 0.54) |
| Meta-analysis | 271,047 | 520,543 | 171.4 | 349.8 | <74 | 223 | < 0.43 (0.34 - 0.54) | 0.64 (0.56 - 0.73) |
| Meta-analysis (w/o USOC) | 41,823 | 79,516 | 20.7 | 43.0 | <19 | 55 | < 0.92 (0.55 - 1.43) | 1.28 (0.96 - 1.67) |
| **PS-stratification / on-treatment TAR without escalation** | | | | | | | | |
| CCAE | 18,681 | 25,665 | 11.7 | 15.7 | 10 | 13 | 0.86 (0.41 - 1.58) | 0.83 (0.44 - 1.42) |
| DAG | 3,228 | 8,796 | 2.3 | 8.3 | <5 | <5 | < 2.18 (0.71 - 5.08) | < 0.60 (0.19 - 1.40) |
| ODOD | 12,737 | 20,640 | 6.9 | 13.2 | <5 | 16 | < 0.72 (0.23 - 1.69) | 1.21 (0.69 - 1.96) |
| MDCR | 1,462 | 4,825 | 0.8 | 3.6 | <5 | 6 | < 6.04 (1.96 - 14.1) | 1.69 (0.62 - 3.67) |
| MDCD | 2,416 | 6,333 | 1.1 | 3.4 | <5 | <5 | < 4.65 (1.51 - 10.9) | < 1.45 (0.47 - 3.39) |
| Open Claims | 273,499 | 441,578 | 170.2 | 307.3 | 59 | 168 | 0.35 (0.26 - 0.45) | 0.55 (0.47 - 0.64) |
| OEHR | 16,535 | 34,484 | 4.5 | 14.6 | <5 | 26 | < 1.11 (0.36 - 2.58) | 1.78 (1.16 - 2.60) |
| SIDIAP | 2,944 | 14,723 | 3.5 | 25.6 | 0 | 7 | | 0.27 (0.11 - 0.56) |
| Meta-analysis | 321,452 | 522,367 | 193.3 | 350.9 | <79 | 223 | < 0.41 (0.32 - 0.51) | 0.64 (0.55 - 0.72) |
| Meta-analysis (w/o USOC) | 47,953 | 80,789 | 23.1 | 43.6 | <20 | 55 | < 0.87 (0.53 - 1.34) | 1.26 (0.95 - 1.64) |

##### 4.2.25 Venous thromboembolism

**Supplementary Table 336:** Exposures, outcomes and risk estimates for Venous thromboembolism in SGLT2I (female) (T) vs DPP4I (female) (C) new-user cohorts under four different propensity score (PS) adjustment / time-at-risk (TAR) designs across data sources. We report total patient exposure time in 1,000 patient-year (KYr) units, uncalibrated and calibrated hazard ratios (HRs) and their 95% confidence intervals (CIs). Greyed out entries did not pass study diagnostics and are included only for completeness.

| | Patients | | Exposure (KYr) | | Outcomes | | Incidence (per KYr) | |
|---|---|---|---|---|---|---|---|---|
| | T | C | T | C | T | C | T | C |
| **PS-matching / on-treatment TAR** | | | | | | | | |
| CCAE | 16,050 | 25,231 | 12.1 | 18.9 | 55 | 101 | 4.56 (3.44 - 5.94) | 5.34 (4.35 - 6.49) |
| DAG | 2,690 | 8,055 | 2.1 | 8.4 | 13 | 94 | 6.06 (3.23 - 10.4) | 11.2 (9.04 - 13.7) |
| ODOD | 10,381 | 19,876 | 6.7 | 15.1 | 53 | 165 | 7.86 (5.89 - 10.3) | 10.9 (9.32 - 12.7) |
| MDCR | 1,249 | 4,593 | 0.9 | 3.9 | 5 | 49 | 5.83 (1.89 - 13.6) | 12.6 (9.29 - 16.6) |
| MDCD | 2,233 | 6,071 | 1.2 | 4.2 | 10 | 42 | 8.22 (3.94 - 15.1) | 9.99 (7.20 - 13.5) |
| Open Claims | 224,268 | 429,979 | 176.8 | 375.9 | 655 | 1,999 | 3.70 (3.43 - 4.00) | 5.32 (5.09 - 5.56) |
| OEHR | 14,678 | 32,822 | 4.6 | 17.2 | 36 | 118 | 7.79 (5.46 - 10.8) | 6.86 (5.67 - 8.21) |
| SIDIAP | 2,655 | 13,693 | 3.7 | 29.6 | 13 | 150 | 3.52 (1.88 - 6.03) | 5.07 (4.29 - 5.95) |
| Meta-analysis | 274,352 | 540,752 | 208.7 | 475.2 | <840 | 2,695 | < 4.03 (3.76 - 4.31) | 5.67 (5.46 - 5.89) |
| Meta-analysis (w/o USOC) | 50,084 | 110,773 | 31.8 | 99.3 | <185 | 696 | < 5.81 (5.00 - 6.71) | 7.01 (6.49 - 7.55) |
| **PS-stratification / on-treatment TAR** | | | | | | | | |
| CCAE | 18,464 | 25,266 | 13.4 | 18.9 | 59 | 101 | 4.42 (3.36 - 5.70) | 5.34 (4.35 - 6.48) |
| DAG | 2,972 | 8,077 | 2.3 | 8.4 | 15 | 94 | 6.56 (3.67 - 10.8) | 11.1 (9.01 - 13.6) |
| ODOD | 12,418 | 20,057 | 7.6 | 15.3 | 60 | 170 | 7.91 (6.03 - 10.2) | 11.1 (9.53 - 13.0) |
| MDCR | 1,415 | 4,651 | 0.9 | 3.9 | 5 | 49 | 5.48 (1.78 - 12.8) | 12.4 (9.21 - 16.5) |
| MDCD | 2,324 | 6,091 | 1.2 | 4.2 | 10 | 42 | 8.00 (3.84 - 14.7) | 9.93 (7.15 - 13.4) |
| Open Claims | 267,177 | 430,441 | 198.1 | 376.3 | 730 | 2,014 | 3.69 (3.42 - 3.96) | 5.35 (5.12 - 5.59) |
| OEHR | 16,265 | 33,807 | 5.1 | 17.7 | 38 | 124 | 7.47 (5.28 - 10.2) | 7.00 (5.82 - 8.34) |
| SIDIAP | 2,822 | 14,164 | 3.8 | 31.0 | 15 | 161 | 3.91 (2.19 - 6.44) | 5.19 (4.42 - 6.05) |
| Meta-analysis | 320,118 | 531,812 | 230.2 | 467.7 | 917 | 2,664 | 3.98 (3.73 - 4.25) | 5.70 (5.48 - 5.92) |
| Meta-analysis (w/o USOC) | 52,941 | 101,371 | 32.2 | 91.4 | 187 | 650 | 5.81 (5.01 - 6.71) | 7.11 (6.58 - 7.68) |
| **PS-matching / on-treatment TAR without escalation** | | | | | | | | |
| CCAE | 16,050 | 25,231 | 10.4 | 15.4 | 46 | 80 | 4.44 (3.25 - 5.92) | 5.19 (4.12 - 6.46) |
| DAG | 2,690 | 8,055 | 2.0 | 7.6 | 11 | 86 | 5.64 (2.82 - 10.1) | 11.3 (9.07 - 14.0) |
| ODOD | 10,381 | 19,876 | 6.0 | 12.7 | 46 | 147 | 7.72 (5.65 - 10.3) | 11.6 (9.78 - 13.6) |
| MDCR | 1,249 | 4,593 | 0.8 | 3.4 | <5 | 43 | < 6.61 (2.15 - 15.4) | 12.7 (9.18 - 17.1) |
| MDCD | 2,233 | 6,071 | 1.0 | 3.3 | 7 | 30 | 7.00 (2.82 - 14.4) | 9.15 (6.17 - 13.1) |
| Open Claims | 224,268 | 429,979 | 147.8 | 298.9 | 528 | 1,611 | 3.57 (3.27 - 3.89) | 5.39 (5.13 - 5.66) |
| OEHR | 14,678 | 32,822 | 4.0 | 13.9 | 31 | 82 | 7.71 (5.24 - 10.9) | 5.92 (4.71 - 7.34) |
| SIDIAP | 2,655 | 13,693 | 3.2 | 23.4 | 11 | 116 | 3.39 (1.69 - 6.07) | 4.96 (4.10 - 5.95) |
| Meta-analysis | 274,352 | 540,752 | 175.5 | 380.0 | <685 | 2,172 | < 3.90 (3.62 - 4.21) | 5.72 (5.48 - 5.96) |
| Meta-analysis (w/o USOC) | 50,084 | 110,773 | 27.8 | 81.0 | <157 | 561 | < 5.66 (4.81 - 6.61) | 6.92 (6.36 - 7.52) |
| **PS-stratification / on-treatment TAR without escalation** | | | | | | | | |
| CCAE | 18,464 | 25,266 | 11.5 | 15.4 | 49 | 80 | 4.25 (3.15 - 5.62) | 5.19 (4.11 - 6.45) |
| DAG | 2,972 | 8,077 | 2.1 | 7.6 | 13 | 86 | 6.25 (3.33 - 10.7) | 11.3 (9.03 - 13.9) |
| ODOD | 12,418 | 20,057 | 6.7 | 12.8 | 53 | 152 | 7.88 (5.90 - 10.3) | 11.9 (10.0 - 13.9) |
| MDCR | 1,415 | 4,651 | 0.8 | 3.4 | <5 | 43 | < 6.19 (2.01 - 14.4) | 12.6 (9.11 - 16.9) |
| MDCD | 2,324 | 6,091 | 1.0 | 3.3 | 7 | 30 | 6.80 (2.73 - 14.0) | 9.11 (6.15 - 13.0) |
| Open Claims | 267,177 | 430,441 | 166.7 | 299.3 | 597 | 1,625 | 3.58 (3.30 - 3.88) | 5.43 (5.17 - 5.70) |
| OEHR | 16,265 | 33,807 | 4.4 | 14.3 | 32 | 86 | 7.23 (4.94 - 10.2) | 6.02 (4.82 - 7.44) |
| SIDIAP | 2,822 | 14,164 | 3.4 | 24.6 | 13 | 124 | 3.85 (2.05 - 6.58) | 5.05 (4.20 - 6.02) |
| Meta-analysis | 320,118 | 531,812 | 194.8 | 374.0 | 757 | 2,153 | 3.89 (3.61 - 4.17) | 5.76 (5.52 - 6.00) |
| Meta-analysis (w/o USOC) | 52,941 | 101,371 | 28.1 | 74.7 | 160 | 528 | 5.69 (4.84 - 6.64) | 7.07 (6.48 - 7.69) |

##### 4.2.26 Vomiting

**Supplementary Table 337:** Exposures, outcomes and risk estimates for Vomiting in SGLT2I (female) (T) vs DPP4I (female) (C) new-user cohorts under four different propensity score (PS) adjustment / time-at-risk (TAR) designs across data sources. We report total patient exposure time in 1,000 patient-year (KYr) units, uncalibrated and calibrated hazard ratios (HRs) and their 95% confidence intervals (CIs). Greyed out entries did not pass study diagnostics and are included only for completeness.

| | Patients | | Exposure (KYr) | | Outcomes | | Incidence (per KYr) | |
|---|---|---|---|---|---|---|---|---|
| | T | C | T | C | T | C | T | C |
| **PS-matching / on-treatment TAR** | | | | | | | | |
| CCAE | 14,340 | 22,669 | 10.7 | 16.9 | 318 | 531 | 29.6 (26.4 - 33.0) | 31.5 (28.9 - 34.3) |
| DAG | 2,601 | 7,938 | 2.1 | 8.3 | 27 | 115 | 12.9 (8.49 - 18.7) | 13.9 (11.5 - 16.7) |
| ODOD | 9,254 | 17,588 | 6.0 | 13.2 | 179 | 512 | 30.1 (25.8 - 34.8) | 38.9 (35.6 - 42.4) |
| MDCR | 1,171 | 4,258 | 0.8 | 3.5 | 20 | 130 | 25.1 (15.3 - 38.8) | 36.8 (30.8 - 43.7) |
| MDCD | 1,673 | 4,598 | 0.9 | 3.1 | 70 | 243 | 76.4 (59.5 - 96.5) | 78.5 (68.9 - 89.0) |
| Open Claims | 198,473 | 383,325 | 157.4 | 334.5 | 2,551 | 7,135 | 16.2 (15.6 - 16.9) | 21.3 (20.8 - 21.8) |
| OEHR | 13,565 | 30,253 | 4.2 | 15.6 | 125 | 502 | 29.6 (24.7 - 35.3) | 32.1 (29.4 - 35.1) |
| SIDIAP | 2,588 | 13,345 | 3.6 | 28.8 | 23 | 309 | 6.43 (4.08 - 9.65) | 10.7 (9.55 - 12.0) |
| Meta-analysis | 242,494 | 479,716 | 184.9 | 420.4 | 3,293 | 9,347 | 17.8 (17.2 - 18.4) | 22.2 (21.8 - 22.7) |
| Meta-analysis (w/o USOC) | 44,021 | 96,391 | 27.5 | 85.9 | 742 | 2,212 | 27.0 (25.1 - 29.0) | 25.8 (24.7 - 26.9) |
| **PS-stratification / on-treatment TAR** | | | | | | | | |
| CCAE | 16,475 | 22,697 | 11.9 | 16.9 | 353 | 533 | 29.7 (26.7 - 33.0) | 31.6 (29.0 - 34.4) |
| DAG | 2,869 | 7,964 | 2.2 | 8.3 | 27 | 115 | 12.2 (8.01 - 17.7) | 13.8 (11.4 - 16.6) |
| ODOD | 11,062 | 17,785 | 6.7 | 13.3 | 210 | 522 | 31.2 (27.1 - 35.7) | 39.2 (35.9 - 42.7) |
| MDCR | 1,336 | 4,310 | 0.8 | 3.6 | 21 | 132 | 24.7 (15.3 - 37.8) | 37.0 (31.0 - 43.9) |
| MDCD | 1,727 | 4,612 | 0.9 | 3.1 | 71 | 245 | 75.8 (59.2 - 95.6) | 78.8 (69.3 - 89.3) |
| Open Claims | 235,920 | 383,765 | 176.1 | 334.9 | 2,889 | 7,163 | 16.4 (15.8 - 17.0) | 21.4 (20.9 - 21.9) |
| OEHR | 15,028 | 31,401 | 4.6 | 16.2 | 137 | 529 | 29.5 (24.8 - 34.9) | 32.7 (29.9 - 35.6) |
| SIDIAP | 2,759 | 13,789 | 3.7 | 30.2 | 26 | 323 | 6.97 (4.55 - 10.2) | 10.7 (9.55 - 11.9) |
| Meta-analysis | 284,113 | 477,401 | 205.3 | 419.8 | 3,642 | 9,185 | 17.7 (17.2 - 18.3) | 21.9 (21.4 - 22.3) |
| Meta-analysis (w/o USOC) | 48,193 | 93,636 | 29.2 | 84.9 | 753 | 2,022 | 25.8 (24.0 - 27.7) | 23.8 (22.8 - 24.9) |
| **PS-matching / on-treatment TAR without escalation** | | | | | | | | |
| CCAE | 14,340 | 22,669 | 9.3 | 13.8 | 269 | 431 | 29.0 (25.6 - 32.7) | 31.1 (28.3 - 34.2) |
| DAG | 2,601 | 7,938 | 1.9 | 7.4 | 23 | 101 | 12.1 (7.65 - 18.1) | 13.6 (11.1 - 16.5) |
| ODOD | 9,254 | 17,588 | 5.3 | 11.2 | 158 | 427 | 30.0 (25.5 - 35.0) | 38.2 (34.7 - 42.0) |
| MDCR | 1,171 | 4,258 | 0.7 | 3.1 | 19 | 107 | 27.1 (16.3 - 42.3) | 34.7 (28.4 - 41.9) |
| MDCD | 1,673 | 4,598 | 0.8 | 2.5 | 56 | 181 | 72.9 (55.1 - 94.7) | 73.4 (63.1 - 84.9) |
| Open Claims | 198,473 | 383,325 | 132.2 | 267.3 | 2,040 | 5,628 | 15.4 (14.8 - 16.1) | 21.1 (20.5 - 21.6) |
| OEHR | 13,565 | 30,253 | 3.7 | 12.6 | 102 | 385 | 27.7 (22.6 - 33.6) | 30.5 (27.5 - 33.7) |
| SIDIAP | 2,588 | 13,345 | 3.2 | 22.9 | 18 | 231 | 5.71 (3.38 - 9.02) | 10.1 (8.83 - 11.5) |
| Meta-analysis | 242,494 | 479,716 | 156.2 | 337.8 | 2,666 | 7,384 | 17.1 (16.4 - 17.7) | 21.9 (21.4 - 22.4) |
| Meta-analysis (w/o USOC) | 44,021 | 96,391 | 24.1 | 70.4 | 626 | 1,756 | 26.0 (24.0 - 28.1) | 24.9 (23.8 - 26.1) |
| **PS-stratification / on-treatment TAR without escalation** | | | | | | | | |
| CCAE | 16,475 | 22,697 | 10.3 | 13.9 | 301 | 433 | 29.2 (26.0 - 32.7) | 31.2 (28.4 - 34.3) |
| DAG | 2,869 | 7,964 | 2.0 | 7.5 | 23 | 101 | 11.4 (7.22 - 17.1) | 13.5 (11.0 - 16.4) |
| ODOD | 11,062 | 17,785 | 6.0 | 11.3 | 179 | 434 | 29.9 (25.7 - 34.6) | 38.4 (34.9 - 42.2) |
| MDCR | 1,336 | 4,310 | 0.7 | 3.1 | 20 | 109 | 26.7 (16.3 - 41.2) | 35.0 (28.7 - 42.2) |
| MDCD | 1,727 | 4,612 | 0.8 | 2.5 | 56 | 183 | 71.2 (53.8 - 92.4) | 74.0 (63.6 - 85.5) |
| Open Claims | 235,920 | 383,765 | 148.9 | 267.7 | 2,319 | 5,654 | 15.6 (15.0 - 16.2) | 21.1 (20.6 - 21.7) |
| OEHR | 15,028 | 31,401 | 4.1 | 13.1 | 112 | 407 | 27.6 (22.7 - 33.2) | 31.1 (28.1 - 34.2) |
| SIDIAP | 2,759 | 13,789 | 3.3 | 24.0 | 20 | 241 | 6.07 (3.71 - 9.38) | 10.0 (8.80 - 11.4) |
| Meta-analysis | 284,113 | 477,401 | 174.5 | 337.4 | 2,954 | 7,270 | 16.9 (16.3 - 17.5) | 21.5 (21.1 - 22.0) |
| Meta-analysis (w/o USOC) | 48,193 | 93,636 | 25.7 | 69.8 | 635 | 1,616 | 24.8 (22.9 - 26.8) | 23.2 (22.0 - 24.3) |

#### 4.3 SGLT2I (female) vs SU (female)

##### 4.3.1 3-point MACE

**Supplementary Table 338:** Exposures, outcomes and risk estimates for 3-point MACE in SGLT2I (female) (T) vs SU (female) (C) new-user cohorts under four different propensity score (PS) adjustment / time-at-risk (TAR) designs across data sources. We report total patient exposure time in 1,000 patient-year (KYr) units, uncalibrated and calibrated hazard ratios (HRs) and their 95% confidence intervals (CIs). Greyed out entries did not pass study diagnostics and are included only for completeness.

| | Patients | | Exposure (KYr) | | Outcomes | | Incidence (per KYr) | |
|---|---|---|---|---|---|---|---|---|
| | T | C | T | C | T | C | T | C |
| **PS-matching / on-treatment TAR** | | | | | | | | |
| CCAE | 15,697 | 34,271 | 11.6 | 24.4 | 75 | 178 | 6.48 (5.10 - 8.12) | 7.29 (6.26 - 8.44) |
| DAG | 3,180 | 682 | 2.5 | 0.7 | 0 | 0 | | |
| ODOD | 11,353 | 38,676 | 7.1 | 31.9 | 78 | 748 | 10.9 (8.65 - 13.7) | 23.4 (21.8 - 25.2) |
| MDCR | 1,260 | 6,287 | 0.8 | 5.0 | 10 | 147 | 12.0 (5.74 - 22.0) | 29.1 (24.6 - 34.3) |
| MDCD | 2,240 | 10,840 | 1.2 | 6.7 | 20 | 127 | 16.6 (10.2 - 25.7) | 18.8 (15.7 - 22.4) |
| Open Claims | 250,635 | 792,791 | 188.5 | 747.0 | 1,095 | 8,730 | 5.81 (5.47 - 6.16) | 11.7 (11.4 - 11.9) |
| OEHR | 15,228 | 62,852 | 4.8 | 35.7 | 31 | 432 | 6.49 (4.41 - 9.21) | 12.1 (11.0 - 13.3) |
| SIDIAP | 2,768 | 954 | 3.8 | 1.7 | 13 | <5 | 3.39 (1.80 - 5.79) | < 3.00 (0.97 - 6.99) |
| Meta-analysis | 296,413 | 945,717 | 214.1 | 850.8 | 1,309 | 10,362 | 6.12 (5.79 - 6.46) | 12.2 (11.9 - 12.4) |
| Meta-analysis (w/o USOC) | 45,778 | 152,926 | 25.5 | 103.9 | 214 | 1,632 | 8.39 (7.30 - 9.59) | 15.7 (15.0 - 16.5) |
| **PS-stratification / on-treatment TAR** | | | | | | | | |
| CCAE | 18,456 | 34,871 | 13.4 | 24.6 | 88 | 182 | 6.59 (5.28 - 8.12) | 7.39 (6.35 - 8.54) |
| DAG | 3,231 | 1,228 | 2.5 | 1.2 | 0 | 0 | | |
| ODOD | 12,278 | 39,450 | 7.6 | 32.7 | 85 | 781 | 11.2 (8.98 - 13.9) | 23.9 (22.2 - 25.6) |
| MDCR | 1,406 | 6,710 | 0.9 | 5.4 | 12 | 159 | 13.3 (6.89 - 23.3) | 29.5 (25.1 - 34.5) |
| MDCD | 2,284 | 11,111 | 1.2 | 6.9 | 20 | 131 | 16.4 (10.0 - 25.3) | 18.9 (15.8 - 22.4) |
| Open Claims | 263,915 | 801,770 | 196.3 | 755.2 | 1,135 | 9,013 | 5.78 (5.45 - 6.13) | 11.9 (11.7 - 12.2) |
| OEHR | 16,377 | 69,021 | 5.1 | 39.4 | 31 | 512 | 6.02 (4.09 - 8.55) | 13.0 (11.9 - 14.2) |
| SIDIAP | 2,852 | 1,992 | 3.9 | 4.3 | 13 | 11 | 3.35 (1.78 - 5.72) | 2.57 (1.28 - 4.59) |
| Meta-analysis | 311,026 | 945,112 | 222.3 | 851.9 | 1,339 | 10,488 | 6.02 (5.70 - 6.35) | 12.3 (12.1 - 12.5) |
| Meta-analysis (w/o USOC) | 47,111 | 143,342 | 26.1 | 96.7 | 204 | 1,475 | 7.83 (6.79 - 8.98) | 15.2 (14.5 - 16.0) |
| **PS-matching / on-treatment TAR without escalation** | | | | | | | | |
| CCAE | 15,697 | 34,271 | 10.0 | 21.0 | 59 | 154 | 5.92 (4.50 - 7.63) | 7.33 (6.22 - 8.59) |
| DAG | 3,180 | 682 | 2.3 | 0.6 | 0 | 0 | | |
| ODOD | 11,353 | 38,676 | 6.3 | 28.5 | 70 | 660 | 11.1 (8.65 - 14.0) | 23.1 (21.4 - 25.0) |
| MDCR | 1,260 | 6,287 | 0.7 | 4.6 | 10 | 135 | 13.5 (6.49 - 24.9) | 29.3 (24.6 - 34.7) |
| MDCD | 2,240 | 10,840 | 1.0 | 5.6 | 12 | 107 | 12.0 (6.20 - 21.0) | 19.2 (15.7 - 23.2) |
| Open Claims | 250,635 | 792,791 | 158.3 | 629.5 | 911 | 7,430 | 5.75 (5.39 - 6.14) | 11.8 (11.5 - 12.1) |
| OEHR | 15,228 | 62,852 | 4.2 | 30.4 | 26 | 327 | 6.26 (4.09 - 9.18) | 10.8 (9.63 - 12.0) |
| SIDIAP | 2,768 | 954 | 3.4 | 1.4 | 12 | <5 | 3.57 (1.84 - 6.24) | < 3.55 (1.15 - 8.30) |
| Meta-analysis | 296,413 | 945,717 | 180.5 | 719.6 | 1,088 | 8,813 | 6.03 (5.68 - 6.40) | 12.2 (12.0 - 12.5) |
| Meta-analysis (w/o USOC) | 45,778 | 152,926 | 22.2 | 90.1 | 177 | 1,383 | 7.98 (6.85 - 9.25) | 15.3 (14.5 - 16.2) |
| **PS-stratification / on-treatment TAR without escalation** | | | | | | | | |
| CCAE | 18,456 | 34,871 | 11.5 | 21.2 | 71 | 157 | 6.16 (4.81 - 7.76) | 7.40 (6.29 - 8.66) |
| DAG | 3,231 | 1,228 | 2.3 | 1.2 | 0 | 0 | | |
| ODOD | 12,278 | 39,450 | 6.7 | 29.2 | 77 | 691 | 11.5 (9.06 - 14.3) | 23.6 (21.9 - 25.5) |
| MDCR | 1,406 | 6,710 | 0.8 | 4.9 | 10 | 146 | 12.5 (6.00 - 23.0) | 29.7 (25.1 - 34.9) |
| MDCD | 2,284 | 11,111 | 1.0 | 5.7 | 12 | 110 | 11.8 (6.10 - 20.6) | 19.2 (15.7 - 23.1) |
| Open Claims | 263,915 | 801,770 | 165.0 | 636.6 | 947 | 7,674 | 5.74 (5.38 - 6.12) | 12.1 (11.8 - 12.3) |
| OEHR | 16,377 | 69,021 | 4.5 | 33.5 | 26 | 381 | 5.81 (3.79 - 8.51) | 11.4 (10.3 - 12.6) |
| SIDIAP | 2,852 | 1,992 | 3.4 | 3.5 | 12 | 11 | 3.53 (1.82 - 6.16) | 3.12 (1.56 - 5.59) |
| Meta-analysis | 311,026 | 945,112 | 187.8 | 720.6 | 1,121 | 8,903 | 5.97 (5.63 - 6.33) | 12.4 (12.1 - 12.6) |
| Meta-analysis (w/o USOC) | 47,111 | 143,342 | 22.7 | 83.9 | 174 | 1,229 | 7.66 (6.56 - 8.89) | 14.6 (13.8 - 15.5) |

##### 4.3.2 4-point MACE

**Supplementary Table 339:** Exposures, outcomes and risk estimates for 4-point MACE in SGLT2I (female) (T) vs SU (female) (C) new-user cohorts under four different propensity score (PS) adjustment / time-at-risk (TAR) designs across data sources. We report total patient exposure time in 1,000 patient-year (KYr) units, uncalibrated and calibrated hazard ratios (HRs) and their 95% confidence intervals (CIs). Greyed out entries did not pass study diagnostics and are included only for completeness.

| | Patients | | Exposure (KYr) | | Outcomes | | Incidence (per KYr) | |
|---|---|---|---|---|---|---|---|---|
| | T | C | T | C | T | C | T | C |
| **PS-matching / on-treatment TAR** | | | | | | | | |
| CCAE | 15,500 | 33,636 | 11.4 | 23.9 | 105 | 289 | 9.19 (7.52 - 11.1) | 12.1 (10.7 - 13.5) |
| DAG | 3,180 | 682 | 2.5 | 0.7 | 0 | 0 | | |
| ODOD | 10,944 | 36,544 | 6.9 | 29.9 | 119 | 1,184 | 17.3 (14.3 - 20.7) | 39.6 (37.3 - 41.9) |
| MDCR | 1,196 | 5,866 | 0.8 | 4.7 | 18 | 225 | 22.6 (13.4 - 35.7) | 48.3 (42.2 - 55.1) |
| MDCD | 2,122 | 10,183 | 1.1 | 6.3 | 35 | 206 | 30.6 (21.3 - 42.6) | 32.7 (28.4 - 37.5) |
| Open Claims | 243,852 | 766,558 | 184.1 | 718.2 | 1,628 | 14,529 | 8.84 (8.42 - 9.28) | 20.2 (19.9 - 20.6) |
| OEHR | 15,021 | 61,694 | 4.7 | 34.7 | 59 | 805 | 12.5 (9.55 - 16.2) | 23.2 (21.6 - 24.9) |
| SIDIAP | 2,745 | 952 | 3.8 | 1.7 | 14 | <5 | 3.66 (2.00 - 6.15) | < 3.00 (0.97 - 7.00) |
| Meta-analysis | 288,635 | 914,481 | 209.1 | 817.7 | 1,964 | 17,238 | 9.39 (8.98 - 9.82) | 21.1 (20.8 - 21.4) |
| Meta-analysis (w/o USOC) | 44,783 | 147,923 | 25.0 | 99.5 | 336 | 2,709 | 13.5 (12.1 - 15.0) | 27.2 (26.2 - 28.3) |
| **PS-stratification / on-treatment TAR** | | | | | | | | |
| CCAE | 18,202 | 34,311 | 13.2 | 24.2 | 121 | 294 | 9.18 (7.62 - 11.0) | 12.1 (10.8 - 13.6) |
| DAG | 3,231 | 1,228 | 2.5 | 1.2 | 0 | 0 | | |
| ODOD | 11,798 | 37,406 | 7.3 | 30.8 | 130 | 1,261 | 17.8 (14.9 - 21.1) | 41.0 (38.8 - 43.3) |
| MDCR | 1,334 | 6,198 | 0.9 | 4.9 | 20 | 245 | 23.3 (14.2 - 35.9) | 50.0 (43.9 - 56.7) |
| MDCD | 2,158 | 10,389 | 1.2 | 6.4 | 35 | 208 | 30.2 (21.1 - 42.1) | 32.3 (28.0 - 37.0) |
| Open Claims | 255,645 | 774,956 | 191.0 | 725.9 | 1,703 | 15,047 | 8.91 (8.50 - 9.35) | 20.7 (20.4 - 21.1) |
| OEHR | 16,145 | 67,681 | 5.1 | 38.2 | 59 | 943 | 11.6 (8.86 - 15.0) | 24.7 (23.1 - 26.3) |
| SIDIAP | 2,828 | 1,990 | 3.9 | 4.3 | 14 | 12 | 3.62 (1.98 - 6.08) | 2.80 (1.45 - 4.89) |
| Meta-analysis | 301,790 | 914,354 | 216.6 | 819.0 | 2,013 | 17,545 | 9.29 (8.89 - 9.71) | 21.4 (21.1 - 21.7) |
| Meta-analysis (w/o USOC) | 46,145 | 139,398 | 25.5 | 93.1 | 310 | 2,498 | 12.1 (10.8 - 13.6) | 26.8 (25.8 - 27.9) |
| **PS-matching / on-treatment TAR without escalation** | | | | | | | | |
| CCAE | 15,500 | 33,636 | 9.9 | 20.6 | 86 | 250 | 8.72 (6.98 - 10.8) | 12.1 (10.7 - 13.7) |
| DAG | 3,180 | 682 | 2.3 | 0.6 | 0 | 0 | | |
| ODOD | 10,944 | 36,544 | 6.1 | 26.8 | 101 | 1,043 | 16.5 (13.5 - 20.1) | 38.9 (36.6 - 41.4) |
| MDCR | 1,196 | 5,866 | 0.7 | 4.3 | 17 | 209 | 24.2 (14.1 - 38.7) | 49.0 (42.5 - 56.1) |
| MDCD | 2,122 | 10,183 | 0.9 | 5.2 | 24 | 167 | 25.3 (16.2 - 37.6) | 31.9 (27.3 - 37.2) |
| Open Claims | 243,852 | 766,558 | 154.6 | 605.6 | 1,344 | 12,307 | 8.69 (8.23 - 9.17) | 20.3 (20.0 - 20.7) |
| OEHR | 15,021 | 61,694 | 4.1 | 29.6 | 49 | 626 | 12.0 (8.87 - 15.8) | 21.1 (19.5 - 22.8) |
| SIDIAP | 2,745 | 952 | 3.3 | 1.4 | 13 | <5 | 3.89 (2.07 - 6.64) | < 3.56 (1.16 - 8.31) |
| Meta-analysis | 288,635 | 914,481 | 176.3 | 692.2 | 1,621 | 14,602 | 9.19 (8.75 - 9.65) | 21.1 (20.8 - 21.4) |
| Meta-analysis (w/o USOC) | 44,783 | 147,923 | 21.7 | 86.5 | 277 | 2,295 | 12.8 (11.3 - 14.4) | 26.5 (25.4 - 27.6) |
| **PS-stratification / on-treatment TAR without escalation** | | | | | | | | |
| CCAE | 18,202 | 34,311 | 11.4 | 20.9 | 102 | 253 | 8.95 (7.30 - 10.9) | 12.1 (10.7 - 13.7) |
| DAG | 3,231 | 1,228 | 2.3 | 1.2 | 0 | 0 | | |
| ODOD | 11,798 | 37,406 | 6.5 | 27.5 | 112 | 1,109 | 17.3 (14.2 - 20.8) | 40.3 (37.9 - 42.7) |
| MDCR | 1,334 | 6,198 | 0.8 | 4.5 | 17 | 228 | 22.3 (13.0 - 35.8) | 50.8 (44.4 - 57.8) |
| MDCD | 2,158 | 10,389 | 1.0 | 5.3 | 24 | 169 | 24.9 (16.0 - 37.1) | 31.6 (27.0 - 36.8) |
| Open Claims | 255,645 | 774,956 | 160.7 | 612.4 | 1,411 | 12,760 | 8.78 (8.33 - 9.25) | 20.8 (20.5 - 21.2) |
| OEHR | 16,145 | 67,681 | 4.4 | 32.6 | 49 | 722 | 11.1 (8.22 - 14.7) | 22.1 (20.5 - 23.8) |
| SIDIAP | 2,828 | 1,990 | 3.4 | 3.5 | 13 | 12 | 3.84 (2.04 - 6.56) | 3.41 (1.76 - 5.95) |
| Meta-analysis | 301,790 | 914,354 | 182.9 | 693.4 | 1,674 | 14,844 | 9.15 (8.72 - 9.60) | 21.4 (21.1 - 21.8) |
| Meta-analysis (w/o USOC) | 46,145 | 139,398 | 22.3 | 81.0 | 263 | 2,084 | 11.8 (10.4 - 13.3) | 25.7 (24.6 - 26.8) |

##### 4.3.3 Acute myocardial infarction

**Supplementary Table 340:** Exposures, outcomes and risk estimates for Acute myocardial infarction in SGLT2I (female) (T) vs SU (female) (C) new-user cohorts under four different propensity score (PS) adjustment / time-at-risk (TAR) designs across data sources. We report total patient exposure time in 1,000 patient-year (KYr) units, uncalibrated and calibrated hazard ratios (HRs) and their 95% confidence intervals (CIs). Greyed out entries did not pass study diagnostics and are included only for completeness.

| | Patients | | Exposure (KYr) | | Outcomes | | Incidence (per KYr) | |
|---|---|---|---|---|---|---|---|---|
| | T | C | T | C | T | C | T | C |
| **PS-matching / on-treatment TAR** | | | | | | | | |
| CCAE | 15,900 | 34,793 | 11.7 | 24.9 | 31 | 63 | 2.65 (1.80 - 3.76) | 2.53 (1.95 - 3.24) |
| DAG | 3,180 | 682 | 2.5 | 0.7 | 0 | 0 | | |
| ODOD | 11,601 | 40,002 | 7.3 | 33.3 | 32 | 330 | 4.39 (3.00 - 6.20) | 9.90 (8.86 - 11.0) |
| MDCR | 1,290 | 6,575 | 0.9 | 5.3 | 6 | 58 | 7.03 (2.58 - 15.3) | 10.9 (8.29 - 14.1) |
| MDCD | 2,318 | 11,314 | 1.3 | 7.1 | 8 | 48 | 6.39 (2.76 - 12.6) | 6.78 (5.00 - 8.99) |
| Open Claims | 256,294 | 815,039 | 192.5 | 770.0 | 527 | 3,768 | 2.74 (2.51 - 2.98) | 4.89 (4.74 - 5.05) |
| OEHR | 15,344 | 63,602 | 4.8 | 36.3 | 17 | 213 | 3.53 (2.05 - 5.64) | 5.87 (5.11 - 6.71) |
| SIDIAP | 2,787 | 963 | 3.9 | 1.7 | <5 | <5 | < 1.29 (0.42 - 3.01) | < 2.97 (0.97 - 6.94) |
| Meta-analysis | 302,747 | 971,325 | 218.4 | 876.9 | 621 | 4,480 | 2.84 (2.62 - 3.08) | 5.11 (4.96 - 5.26) |
| Meta-analysis (w/o USOC) | 46,453 | 156,286 | 25.9 | 106.9 | 94 | 712 | 3.62 (2.93 - 4.44) | 6.66 (6.18 - 7.17) |
| **PS-stratification / on-treatment TAR** | | | | | | | | |
| CCAE | 18,670 | 35,410 | 13.5 | 25.1 | 35 | 66 | 2.59 (1.80 - 3.60) | 2.63 (2.03 - 3.35) |
| DAG | 3,231 | 1,228 | 2.5 | 1.2 | 0 | 0 | | |
| ODOD | 12,545 | 40,829 | 7.7 | 34.1 | 34 | 346 | 4.40 (3.04 - 6.14) | 10.1 (9.09 - 11.3) |
| MDCR | 1,439 | 7,041 | 0.9 | 5.7 | 7 | 65 | 7.63 (3.07 - 15.7) | 11.4 (8.81 - 14.5) |
| MDCD | 2,362 | 11,609 | 1.3 | 7.3 | 8 | 53 | 6.30 (2.72 - 12.4) | 7.28 (5.45 - 9.52) |
| Open Claims | 269,891 | 824,655 | 200.4 | 779.0 | 548 | 3,897 | 2.73 (2.51 - 2.97) | 5.00 (4.85 - 5.16) |
| OEHR | 16,491 | 69,757 | 5.2 | 40.0 | 17 | 248 | 3.28 (1.91 - 5.25) | 6.20 (5.46 - 7.03) |
| SIDIAP | 2,874 | 2,005 | 3.9 | 4.3 | <5 | 5 | < 1.28 (0.41 - 2.98) | 1.16 (0.38 - 2.70) |
| Meta-analysis | 317,597 | 970,651 | 226.8 | 878.2 | 634 | 4,557 | 2.80 (2.58 - 3.02) | 5.19 (5.04 - 5.34) |
| Meta-analysis (w/o USOC) | 47,706 | 145,996 | 26.5 | 99.2 | 86 | 660 | 3.25 (2.60 - 4.02) | 6.65 (6.16 - 7.18) |
| **PS-matching / on-treatment TAR without escalation** | | | | | | | | |
| CCAE | 15,900 | 34,793 | 10.1 | 21.4 | 24 | 56 | 2.38 (1.52 - 3.54) | 2.62 (1.98 - 3.40) |
| DAG | 3,180 | 682 | 2.3 | 0.6 | 0 | 0 | | |
| ODOD | 11,601 | 40,002 | 6.5 | 29.7 | 27 | 288 | 4.18 (2.76 - 6.09) | 9.68 (8.60 - 10.9) |
| MDCR | 1,290 | 6,575 | 0.8 | 4.8 | 6 | 54 | 7.97 (2.93 - 17.3) | 11.2 (8.38 - 14.6) |
| MDCD | 2,318 | 11,314 | 1.0 | 5.9 | 6 | 39 | 5.81 (2.13 - 12.6) | 6.66 (4.74 - 9.11) |
| Open Claims | 256,294 | 815,039 | 161.6 | 648.6 | 439 | 3,217 | 2.72 (2.47 - 2.98) | 4.96 (4.79 - 5.13) |
| OEHR | 15,344 | 63,602 | 4.2 | 30.8 | 16 | 157 | 3.82 (2.18 - 6.20) | 5.09 (4.33 - 5.95) |
| SIDIAP | 2,787 | 963 | 3.4 | 1.4 | <5 | <5 | < 1.47 (0.48 - 3.44) | < 3.53 (1.14 - 8.23) |
| Meta-analysis | 302,747 | 971,325 | 184.1 | 741.2 | 518 | 3,811 | 2.81 (2.58 - 3.07) | 5.14 (4.98 - 5.31) |
| Meta-analysis (w/o USOC) | 46,453 | 156,286 | 22.5 | 92.6 | 79 | 594 | 3.51 (2.78 - 4.37) | 6.41 (5.91 - 6.95) |
| **PS-stratification / on-treatment TAR without escalation** | | | | | | | | |
| CCAE | 18,670 | 35,410 | 11.7 | 21.6 | 28 | 58 | 2.40 (1.59 - 3.47) | 2.69 (2.04 - 3.47) |
| DAG | 3,231 | 1,228 | 2.3 | 1.2 | 0 | 0 | | |
| ODOD | 12,545 | 40,829 | 6.9 | 30.5 | 29 | 303 | 4.23 (2.83 - 6.07) | 9.94 (8.85 - 11.1) |
| MDCR | 1,439 | 7,041 | 0.8 | 5.2 | 6 | 61 | 7.39 (2.71 - 16.1) | 11.8 (8.99 - 15.1) |
| MDCD | 2,362 | 11,609 | 1.0 | 6.0 | 6 | 42 | 5.72 (2.10 - 12.4) | 6.98 (5.03 - 9.43) |
| Open Claims | 269,891 | 824,655 | 168.5 | 656.4 | 458 | 3,333 | 2.72 (2.48 - 2.98) | 5.08 (4.91 - 5.25) |
| OEHR | 16,491 | 69,757 | 4.5 | 33.9 | 16 | 184 | 3.55 (2.03 - 5.76) | 5.42 (4.67 - 6.26) |
| SIDIAP | 2,874 | 2,005 | 3.4 | 3.6 | <5 | 5 | < 1.46 (0.47 - 3.40) | 1.41 (0.46 - 3.28) |
| Meta-analysis | 317,597 | 970,651 | 191.5 | 742.4 | 531 | 3,878 | 2.77 (2.54 - 3.02) | 5.22 (5.06 - 5.39) |
| Meta-analysis (w/o USOC) | 47,706 | 145,996 | 23.0 | 86.0 | 73 | 545 | 3.17 (2.48 - 3.98) | 6.34 (5.82 - 6.89) |

##### 4.3.4 Acute renal failure

**Supplementary Table 341:** Exposures, outcomes and risk estimates for Acute renal failure in SGLT2I (female) (T) vs SU (female) (C) new-user cohorts under four different propensity score (PS) adjustment / time-at-risk (TAR) designs across data sources. We report total patient exposure time in 1,000 patient-year (KYr) units, uncalibrated and calibrated hazard ratios (HRs) and their 95% confidence intervals (CIs). Greyed out entries did not pass study diagnostics and are included only for completeness.

| | Patients | | Exposure (KYr) | | Outcomes | | Incidence (per KYr) | |
|---|---|---|---|---|---|---|---|---|
| | T | C | T | C | T | C | T | C |
| **PS-matching / on-treatment TAR** | | | | | | | | |
| CCAE | 15,896 | 34,558 | 11.7 | 24.6 | 52 | 224 | 4.44 (3.32 - 5.82) | 9.09 (7.94 - 10.4) |
| DAG | 3,180 | 682 | 2.5 | 0.7 | 0 | 0 | | |
| ODOD | 11,479 | 38,470 | 7.2 | 31.6 | 112 | 1,096 | 15.6 (12.8 - 18.8) | 34.6 (32.6 - 36.7) |
| MDCR | 1,294 | 6,359 | 0.9 | 5.1 | 12 | 190 | 13.9 (7.20 - 24.3) | 37.4 (32.3 - 43.2) |
| MDCD | 2,271 | 10,872 | 1.2 | 6.7 | 20 | 206 | 16.3 (9.97 - 25.2) | 30.7 (26.7 - 35.2) |
| Open Claims | 255,466 | 803,475 | 192.0 | 754.8 | 1,320 | 11,815 | 6.87 (6.51 - 7.26) | 15.7 (15.4 - 15.9) |
| OEHR | 15,348 | 63,238 | 4.8 | 35.8 | 38 | 574 | 7.90 (5.59 - 10.8) | 16.0 (14.7 - 17.4) |
| SIDIAP | 2,849 | 965 | 4.0 | 1.7 | <5 | <5 | < 1.26 (0.41 - 2.94) | < 2.96 (0.96 - 6.92) |
| Meta-analysis | 301,754 | 956,972 | 217.8 | 858.7 | 1,554 | 14,105 | 7.14 (6.78 - 7.50) | 16.4 (16.2 - 16.7) |
| Meta-analysis (w/o USOC) | 46,288 | 153,497 | 25.8 | 103.9 | 234 | 2,290 | 9.07 (7.95 - 10.3) | 22.0 (21.1 - 23.0) |
| **PS-stratification / on-treatment TAR** | | | | | | | | |
| CCAE | 18,692 | 35,143 | 13.5 | 24.9 | 62 | 226 | 4.58 (3.51 - 5.87) | 9.09 (7.95 - 10.4) |
| DAG | 3,231 | 1,228 | 2.5 | 1.2 | 0 | 0 | | |
| ODOD | 12,447 | 39,271 | 7.6 | 32.4 | 120 | 1,156 | 15.7 (13.0 - 18.8) | 35.7 (33.6 - 37.8) |
| MDCR | 1,449 | 6,718 | 0.9 | 5.4 | 13 | 214 | 14.0 (7.47 - 24.0) | 39.9 (34.8 - 45.6) |
| MDCD | 2,313 | 11,156 | 1.2 | 6.9 | 21 | 214 | 16.9 (10.5 - 25.8) | 31.0 (27.0 - 35.4) |
| Open Claims | 269,448 | 812,179 | 200.1 | 762.8 | 1,386 | 12,217 | 6.93 (6.57 - 7.30) | 16.0 (15.7 - 16.3) |
| OEHR | 16,501 | 69,263 | 5.2 | 39.4 | 39 | 669 | 7.53 (5.36 - 10.3) | 17.0 (15.7 - 18.3) |
| SIDIAP | 2,950 | 2,012 | 4.0 | 4.3 | <5 | <5 | < 1.24 (0.40 - 2.90) | < 1.15 (0.37 - 2.69) |
| Meta-analysis | 317,088 | 955,856 | 226.5 | 859.5 | 1,607 | 14,268 | 7.10 (6.75 - 7.45) | 16.6 (16.3 - 16.9) |
| Meta-analysis (w/o USOC) | 47,640 | 143,677 | 26.4 | 96.7 | 221 | 2,051 | 8.38 (7.32 - 9.57) | 21.2 (20.3 - 22.2) |
| **PS-matching / on-treatment TAR without escalation** | | | | | | | | |
| CCAE | 15,896 | 34,558 | 10.1 | 21.2 | 44 | 185 | 4.37 (3.17 - 5.86) | 8.72 (7.51 - 10.1) |
| DAG | 3,180 | 682 | 2.3 | 0.6 | 0 | 0 | | |
| ODOD | 11,479 | 38,470 | 6.4 | 28.3 | 98 | 961 | 15.4 (12.5 - 18.8) | 33.9 (31.8 - 36.2) |
| MDCR | 1,294 | 6,359 | 0.8 | 4.6 | 11 | 170 | 14.4 (7.20 - 25.8) | 36.7 (31.4 - 42.7) |
| MDCD | 2,271 | 10,872 | 1.0 | 5.6 | 13 | 160 | 12.8 (6.83 - 21.9) | 28.7 (24.5 - 33.5) |
| Open Claims | 255,466 | 803,475 | 161.3 | 636.5 | 1,059 | 9,892 | 6.56 (6.18 - 6.97) | 15.5 (15.2 - 15.8) |
| OEHR | 15,348 | 63,238 | 4.2 | 30.6 | 28 | 405 | 6.69 (4.45 - 9.67) | 13.3 (12.0 - 14.6) |
| SIDIAP | 2,849 | 965 | 3.5 | 1.4 | <5 | <5 | < 1.44 (0.47 - 3.36) | < 3.52 (1.14 - 8.20) |
| Meta-analysis | 301,754 | 956,972 | 183.7 | 726.8 | 1,253 | 11,773 | 6.82 (6.45 - 7.21) | 16.2 (15.9 - 16.5) |
| Meta-analysis (w/o USOC) | 46,288 | 153,497 | 22.4 | 90.3 | 194 | 1,881 | 8.66 (7.49 - 9.97) | 20.8 (19.9 - 21.8) |
| **PS-stratification / on-treatment TAR without escalation** | | | | | | | | |
| CCAE | 18,692 | 35,143 | 11.7 | 21.4 | 53 | 187 | 4.54 (3.40 - 5.94) | 8.73 (7.53 - 10.1) |
| DAG | 3,231 | 1,228 | 2.3 | 1.2 | 0 | 0 | | |
| ODOD | 12,447 | 39,271 | 6.8 | 29.0 | 103 | 1,018 | 15.2 (12.4 - 18.4) | 35.1 (33.0 - 37.3) |
| MDCR | 1,449 | 6,718 | 0.8 | 4.9 | 12 | 192 | 14.6 (7.53 - 25.5) | 39.3 (33.9 - 45.2) |
| MDCD | 2,313 | 11,156 | 1.0 | 5.7 | 14 | 166 | 13.6 (7.44 - 22.8) | 29.0 (24.7 - 33.7) |
| Open Claims | 269,448 | 812,179 | 168.4 | 643.5 | 1,117 | 10,227 | 6.63 (6.25 - 7.04) | 15.9 (15.6 - 16.2) |
| OEHR | 16,501 | 69,263 | 4.5 | 33.6 | 29 | 473 | 6.44 (4.31 - 9.24) | 14.1 (12.8 - 15.4) |
| SIDIAP | 2,950 | 2,012 | 3.5 | 3.6 | <5 | <5 | < 1.42 (0.46 - 3.31) | < 1.40 (0.45 - 3.26) |
| Meta-analysis | 317,088 | 955,856 | 191.3 | 727.5 | 1,302 | 11,905 | 6.81 (6.44 - 7.19) | 16.4 (16.1 - 16.7) |
| Meta-analysis (w/o USOC) | 47,640 | 143,677 | 23.0 | 84.0 | 185 | 1,678 | 8.06 (6.94 - 9.31) | 20.0 (19.0 - 20.9) |

##### 4.3.5 Glycemic control

**Supplementary Table 342:** Exposures, outcomes and risk estimates for Glycemic control in SGLT2I (female) (T) vs SU (female) (C) new-user cohorts under four different propensity score (PS) adjustment / time-at-risk (TAR) designs across data sources. We report total patient exposure time in 1,000 patient-year (KYr) units, uncalibrated and calibrated hazard ratios (HRs) and their 95% confidence intervals (CIs). Greyed out entries did not pass study diagnostics and are included only for completeness.

| | Patients | | Exposure (KYr) | | Outcomes | | Incidence (per KYr) | |
|---|---|---|---|---|---|---|---|---|
| | T | C | T | C | T | C | T | C |
| **PS-matching / on-treatment TAR** | | | | | | | | |
| CCAE | 15,937 | 34,825 | 11.8 | 24.9 | 18 | 39 | 1.53 (0.91 - 2.42) | 1.57 (1.11 - 2.14) |
| DAG | 1,045 | 274 | 0.7 | 0.2 | 97 | 29 | 135 (109 - 165) | 124 (82.8 - 178) |
| ODOD | 7,412 | 26,878 | 4.3 | 19.9 | 430 | 2,143 | 100 (91.2 - 110) | 108 (103 - 112) |
| MDCR | 1,327 | 6,736 | 0.9 | 5.5 | 0 | 9 | | 1.65 (0.75 - 3.12) |
| MDCD | 2,388 | 11,679 | 1.3 | 7.3 | 0 | 0 | | |
| Open Claims | 261,894 | 832,892 | 196.5 | 789.1 | 0 | 0 | | |
| OEHR | 15,494 | 64,240 | 4.9 | 36.8 | 0 | 0 | | |
| SIDIAP | 95 | 222 | 0.1 | 0.3 | 32 | 75 | 257 (176 - 363) | 264 (208 - 331) |
| Meta-analysis | 23,444 | 61,925 | 16.2 | 45.1 | 480 | 2,257 | 29.7 (27.1 - 32.5) | 50.1 (48.0 - 52.2) |
| Meta-analysis (w/o USOC) | 23,444 | 61,925 | 16.2 | 45.1 | 480 | 2,257 | 29.7 (27.1 - 32.5) | 50.1 (48.0 - 52.2) |
| **PS-stratification / on-treatment TAR** | | | | | | | | |
| CCAE | 18,683 | 35,323 | 13.5 | 25.1 | 22 | 47 | 1.62 (1.02 - 2.46) | 1.87 (1.38 - 2.49) |
| DAG | 1,071 | 535 | 0.7 | 0.5 | 100 | 62 | 136 (111 - 166) | 129 (98.6 - 165) |
| ODOD | 7,839 | 27,419 | 4.4 | 20.4 | 456 | 2,208 | 103 (93.4 - 113) | 108 (104 - 113) |
| MDCR | 1,482 | 7,185 | 0.9 | 5.9 | 0 | 10 | | 1.70 (0.82 - 3.13) |
| MDCD | 2,435 | 11,946 | 1.3 | 7.5 | 0 | 0 | | |
| Open Claims | 276,521 | 842,971 | 205.0 | 798.5 | 0 | 0 | | |
| OEHR | 16,666 | 70,488 | 5.2 | 40.6 | 0 | 0 | | |
| SIDIAP | 193 | 251 | 0.2 | 0.3 | 55 | 87 | 266 (200 - 346) | 266 (213 - 328) |
| Meta-analysis | 26,522 | 62,742 | 18.0 | 45.5 | 478 | 2,255 | 26.6 (24.2 - 29.1) | 49.6 (47.6 - 51.7) |
| Meta-analysis (w/o USOC) | 26,522 | 62,742 | 18.0 | 45.5 | 478 | 2,255 | 26.6 (24.2 - 29.1) | 49.6 (47.6 - 51.7) |
| **PS-matching / on-treatment TAR without escalation** | | | | | | | | |
| CCAE | 15,937 | 34,825 | 10.1 | 21.4 | 16 | 36 | 1.58 (0.91 - 2.57) | 1.68 (1.18 - 2.33) |
| DAG | 1,045 | 274 | 0.7 | 0.2 | 79 | 29 | 121 (95.5 - 150) | 132 (88.6 - 190) |
| ODOD | 7,412 | 26,878 | 3.8 | 17.8 | 373 | 1,953 | 98.6 (88.8 - 109) | 110 (105 - 115) |
| MDCR | 1,327 | 6,736 | 0.8 | 5.0 | 0 | 9 | | 1.81 (0.83 - 3.44) |
| MDCD | 2,388 | 11,679 | 1.1 | 6.1 | 0 | 0 | | |
| Open Claims | 261,894 | 832,892 | 164.9 | 664.4 | 0 | 0 | | |
| OEHR | 15,494 | 64,240 | 4.2 | 31.2 | 0 | 0 | | |
| SIDIAP | 95 | 222 | 0.1 | 0.2 | 26 | 63 | 245 (160 - 359) | 258 (198 - 330) |
| Meta-analysis | 23,444 | 61,925 | 14.0 | 39.4 | 415 | 2,052 | 29.7 (26.9 - 32.7) | 52.1 (49.9 - 54.4) |
| Meta-analysis (w/o USOC) | 23,444 | 61,925 | 14.0 | 39.4 | 415 | 2,052 | 29.7 (26.9 - 32.7) | 52.1 (49.9 - 54.4) |
| **PS-stratification / on-treatment TAR without escalation** | | | | | | | | |
| CCAE | 18,683 | 35,323 | 11.7 | 21.6 | 18 | 43 | 1.54 (0.91 - 2.44) | 2.00 (1.44 - 2.69) |
| DAG | 1,071 | 535 | 0.7 | 0.5 | 82 | 62 | 123 (97.4 - 152) | 138 (106 - 176) |
| ODOD | 7,839 | 27,419 | 3.9 | 18.2 | 397 | 2,016 | 101 (91.3 - 111) | 111 (106 - 116) |
| MDCR | 1,482 | 7,185 | 0.8 | 5.3 | 0 | 10 | | 1.87 (0.90 - 3.45) |
| MDCD | 2,435 | 11,946 | 1.1 | 6.2 | 0 | 0 | | |
| Open Claims | 276,521 | 842,971 | 172.3 | 672.6 | 0 | 0 | | |
| OEHR | 16,666 | 70,488 | 4.6 | 34.4 | 0 | 0 | | |
| SIDIAP | 193 | 251 | 0.2 | 0.3 | 48 | 75 | 267 (197 - 354) | 265 (208 - 332) |
| Meta-analysis | 26,522 | 62,742 | 15.6 | 39.7 | 415 | 2,059 | 26.6 (24.1 - 29.3) | 51.8 (49.6 - 54.1) |
| Meta-analysis (w/o USOC) | 26,522 | 62,742 | 15.6 | 39.7 | 415 | 2,059 | 26.6 (24.1 - 29.3) | 51.8 (49.6 - 54.1) |

##### 4.3.6 Hospitalization with heart failure

**Supplementary Table 343:** Exposures, outcomes and risk estimates for Hospitalization with heart failure in SGLT2I (female) (T) vs SU (female) (C) new-user cohorts under four different propensity score (PS) adjustment / time-at-risk (TAR) designs across data sources. We report total patient exposure time in 1,000 patient-year (KYr) units, uncalibrated and calibrated hazard ratios (HRs) and their 95% confidence intervals (CIs). Greyed out entries did not pass study diagnostics and are included only for completeness.

| | Patients | | Exposure (KYr) | | Outcomes | | Incidence (per KYr) | |
|---|---|---|---|---|---|---|---|---|
| | T | C | T | C | T | C | T | C |
| **PS-matching / on-treatment TAR** | | | | | | | | |
| CCAE | 15,826 | 34,412 | 11.7 | 24.6 | 42 | 148 | 3.60 (2.59 - 4.86) | 6.02 (5.09 - 7.07) |
| DAG | 3,180 | 682 | 2.5 | 0.7 | 0 | 0 | | |
| ODOD | 11,278 | 38,129 | 7.1 | 31.5 | 75 | 836 | 10.6 (8.31 - 13.2) | 26.5 (24.7 - 28.4) |
| MDCR | 1,252 | 6,184 | 0.8 | 4.9 | 14 | 152 | 16.9 (9.24 - 28.4) | 30.7 (26.1 - 36.0) |
| MDCD | 2,226 | 10,744 | 1.2 | 6.7 | 20 | 146 | 16.6 (10.1 - 25.6) | 21.9 (18.5 - 25.8) |
| Open Claims | 252,612 | 797,448 | 190.5 | 750.2 | 903 | 9,400 | 4.74 (4.44 - 5.06) | 12.5 (12.3 - 12.8) |
| OEHR | 15,240 | 62,764 | 4.8 | 35.5 | 35 | 552 | 7.32 (5.10 - 10.2) | 15.6 (14.3 - 16.9) |
| SIDIAP | 2,824 | 963 | 3.9 | 1.7 | <5 | <5 | < 1.27 (0.41 - 2.96) | < 2.97 (0.96 - 6.93) |
| Meta-analysis | 298,434 | 949,681 | 216.1 | 853.4 | 1,089 | 11,234 | 5.04 (4.74 - 5.35) | 13.2 (12.9 - 13.4) |
| Meta-analysis (w/o USOC) | 45,822 | 152,233 | 25.6 | 103.2 | 186 | 1,834 | 7.27 (6.26 - 8.39) | 17.8 (17.0 - 18.6) |
| **PS-stratification / on-treatment TAR** | | | | | | | | |
| CCAE | 18,562 | 35,113 | 13.5 | 24.9 | 47 | 153 | 3.49 (2.56 - 4.64) | 6.15 (5.22 - 7.21) |
| DAG | 3,231 | 1,228 | 2.5 | 1.2 | 0 | 0 | | |
| ODOD | 12,165 | 39,032 | 7.5 | 32.4 | 82 | 895 | 10.9 (8.67 - 13.5) | 27.6 (25.8 - 29.5) |
| MDCR | 1,395 | 6,557 | 0.9 | 5.2 | 14 | 169 | 15.7 (8.58 - 26.3) | 32.3 (27.6 - 37.5) |
| MDCD | 2,266 | 10,936 | 1.2 | 6.8 | 20 | 147 | 16.3 (9.98 - 25.2) | 21.7 (18.3 - 25.5) |
| Open Claims | 265,019 | 806,578 | 197.8 | 758.7 | 960 | 9,828 | 4.85 (4.55 - 5.17) | 13.0 (12.7 - 13.2) |
| OEHR | 16,387 | 68,880 | 5.1 | 39.1 | 36 | 646 | 7.00 (4.90 - 9.69) | 16.5 (15.3 - 17.8) |
| SIDIAP | 2,923 | 2,011 | 4.0 | 4.3 | <5 | <5 | < 1.25 (0.41 - 2.92) | < 1.15 (0.37 - 2.69) |
| Meta-analysis | 312,133 | 949,603 | 223.9 | 855.1 | 1,125 | 11,522 | 5.02 (4.74 - 5.33) | 13.5 (13.2 - 13.7) |
| Meta-analysis (w/o USOC) | 47,114 | 143,025 | 26.1 | 96.3 | 165 | 1,694 | 6.31 (5.39 - 7.35) | 17.6 (16.8 - 18.4) |
| **PS-matching / on-treatment TAR without escalation** | | | | | | | | |
| CCAE | 15,826 | 34,412 | 10.1 | 21.1 | 37 | 130 | 3.68 (2.59 - 5.07) | 6.15 (5.14 - 7.30) |
| DAG | 3,180 | 682 | 2.3 | 0.6 | 0 | 0 | | |
| ODOD | 11,278 | 38,129 | 6.3 | 28.2 | 63 | 735 | 10.0 (7.69 - 12.8) | 26.1 (24.2 - 28.0) |
| MDCR | 1,252 | 6,184 | 0.7 | 4.5 | 13 | 141 | 17.8 (9.47 - 30.4) | 31.2 (26.3 - 36.8) |
| MDCD | 2,226 | 10,744 | 1.0 | 5.5 | 14 | 111 | 14.0 (7.68 - 23.6) | 20.1 (16.6 - 24.2) |
| Open Claims | 252,612 | 797,448 | 159.9 | 632.3 | 742 | 7,921 | 4.64 (4.31 - 4.99) | 12.5 (12.3 - 12.8) |
| OEHR | 15,240 | 62,764 | 4.2 | 30.3 | 28 | 421 | 6.74 (4.48 - 9.74) | 13.9 (12.6 - 15.3) |
| SIDIAP | 2,824 | 963 | 3.5 | 1.4 | <5 | <5 | < 1.45 (0.47 - 3.37) | < 3.52 (1.14 - 8.22) |
| Meta-analysis | 298,434 | 949,681 | 182.1 | 721.9 | 897 | 9,459 | 4.93 (4.61 - 5.26) | 13.1 (12.8 - 13.4) |
| Meta-analysis (w/o USOC) | 45,822 | 152,233 | 22.2 | 89.6 | 155 | 1,538 | 6.97 (5.92 - 8.16) | 17.2 (16.3 - 18.0) |
| **PS-stratification / on-treatment TAR without escalation** | | | | | | | | |
| CCAE | 18,562 | 35,113 | 11.6 | 21.4 | 42 | 134 | 3.61 (2.60 - 4.89) | 6.26 (5.25 - 7.42) |
| DAG | 3,231 | 1,228 | 2.3 | 1.2 | 0 | 0 | | |
| ODOD | 12,165 | 39,032 | 6.7 | 29.0 | 70 | 785 | 10.5 (8.17 - 13.2) | 27.1 (25.2 - 29.1) |
| MDCR | 1,395 | 6,557 | 0.8 | 4.8 | 13 | 158 | 16.5 (8.78 - 28.2) | 33.0 (28.0 - 38.5) |
| MDCD | 2,266 | 10,936 | 1.0 | 5.6 | 14 | 112 | 13.8 (7.57 - 23.2) | 19.9 (16.4 - 24.0) |
| Open Claims | 265,019 | 806,578 | 166.2 | 639.7 | 794 | 8,291 | 4.78 (4.45 - 5.12) | 13.0 (12.7 - 13.2) |
| OEHR | 16,387 | 68,880 | 4.5 | 33.3 | 29 | 487 | 6.48 (4.34 - 9.31) | 14.6 (13.3 - 16.0) |
| SIDIAP | 2,923 | 2,011 | 3.5 | 3.6 | <5 | <5 | < 1.42 (0.46 - 3.32) | < 1.40 (0.45 - 3.26) |
| Meta-analysis | 312,133 | 949,603 | 189.0 | 723.4 | 935 | 9,697 | 4.95 (4.64 - 5.28) | 13.4 (13.1 - 13.7) |
| Meta-analysis (w/o USOC) | 47,114 | 143,025 | 22.8 | 83.7 | 141 | 1,406 | 6.19 (5.21 - 7.30) | 16.8 (15.9 - 17.7) |

##### 4.3.7 Measured renal dysfunction

**Supplementary Table 344:** Exposures, outcomes and risk estimates for Measured renal dysfunction in SGLT2I (female) (T) vs SU (female) (C) new-user cohorts under four different propensity score (PS) adjustment / time-at-risk (TAR) designs across data sources. We report total patient exposure time in 1,000 patient-year (KYr) units, uncalibrated and calibrated hazard ratios (HRs) and their 95% confidence intervals (CIs). Greyed out entries did not pass study diagnostics and are included only for completeness.

| | Patients | | Exposure (KYr) | | Outcomes | | Incidence (per KYr) | |
|---|---|---|---|---|---|---|---|---|
| | T | C | T | C | T | C | T | C |
| **PS-matching / on-treatment TAR** | | | | | | | | |
| CCAE | 16,054 | 35,135 | 11.8 | 25.1 | <5 | 13 | < 0.42 (0.14 - 0.99) | 0.52 (0.28 - 0.89) |
| DAG | 3,180 | 682 | 2.5 | 0.7 | 0 | 0 | | |
| ODOD | 11,834 | 40,860 | 7.4 | 34.2 | 10 | 56 | 1.35 (0.65 - 2.49) | 1.64 (1.24 - 2.13) |
| MDCR | 1,333 | 6,800 | 0.9 | 5.5 | 0 | <5 | | < 0.91 (0.29 - 2.12) |
| MDCD | 2,388 | 11,679 | 1.3 | 7.3 | 0 | 0 | | |
| Open Claims | 261,894 | 832,892 | 196.5 | 789.1 | 0 | 0 | | |
| OEHR | 15,494 | 64,240 | 4.9 | 36.8 | 0 | 0 | | |
| SIDIAP | 2,843 | 962 | 4.0 | 1.7 | 0 | <5 | | < 2.97 (0.96 - 6.92) |
| Meta-analysis | 11,834 | 40,860 | 7.4 | 34.2 | 10 | 56 | 1.35 (0.65 - 2.49) | 1.64 (1.24 - 2.13) |
| Meta-analysis (w/o USOC) | 11,834 | 40,860 | 7.4 | 34.2 | 10 | 56 | 1.35 (0.65 - 2.49) | 1.64 (1.24 - 2.13) |
| **PS-stratification / on-treatment TAR** | | | | | | | | |
| CCAE | 18,849 | 35,766 | 13.7 | 25.3 | <5 | 13 | < 0.37 (0.12 - 0.85) | 0.51 (0.27 - 0.88) |
| DAG | 3,231 | 1,228 | 2.5 | 1.2 | 0 | 0 | | |
| ODOD | 12,834 | 41,754 | 7.9 | 35.0 | 10 | 64 | 1.27 (0.61 - 2.34) | 1.83 (1.41 - 2.33) |
| MDCR | 1,489 | 7,293 | 0.9 | 5.9 | 0 | <5 | | < 0.84 (0.27 - 1.97) |
| MDCD | 2,435 | 11,946 | 1.3 | 7.5 | 0 | 0 | | |
| Open Claims | 276,521 | 842,971 | 205.0 | 798.5 | 0 | 0 | | |
| OEHR | 16,666 | 70,488 | 5.2 | 40.6 | 0 | 0 | | |
| SIDIAP | 2,943 | 2,006 | 4.0 | 4.3 | 0 | 5 | | 1.15 (0.37 - 2.69) |
| Meta-analysis | 12,834 | 41,754 | 7.9 | 35.0 | 10 | 64 | 1.27 (0.61 - 2.34) | 1.83 (1.41 - 2.33) |
| Meta-analysis (w/o USOC) | 12,834 | 41,754 | 7.9 | 35.0 | 10 | 64 | 1.27 (0.61 - 2.34) | 1.83 (1.41 - 2.33) |
| **PS-matching / on-treatment TAR without escalation** | | | | | | | | |
| CCAE | 16,054 | 35,135 | 10.2 | 21.6 | <5 | 12 | < 0.49 (0.16 - 1.15) | 0.56 (0.29 - 0.97) |
| DAG | 3,180 | 682 | 2.3 | 0.6 | 0 | 0 | | |
| ODOD | 11,834 | 40,860 | 6.5 | 30.5 | 8 | 47 | 1.22 (0.53 - 2.41) | 1.54 (1.13 - 2.05) |
| MDCR | 1,333 | 6,800 | 0.8 | 5.0 | 0 | <5 | | < 1.00 (0.32 - 2.33) |
| MDCD | 2,388 | 11,679 | 1.1 | 6.1 | 0 | 0 | | |
| Open Claims | 261,894 | 832,892 | 164.9 | 664.4 | 0 | 0 | | |
| OEHR | 15,494 | 64,240 | 4.2 | 31.2 | 0 | 0 | | |
| SIDIAP | 2,843 | 962 | 3.5 | 1.4 | 0 | <5 | | < 3.52 (1.14 - 8.22) |
| Meta-analysis | 11,834 | 40,860 | 6.5 | 30.5 | 8 | 47 | 1.22 (0.53 - 2.41) | 1.54 (1.13 - 2.05) |
| Meta-analysis (w/o USOC) | 11,834 | 40,860 | 6.5 | 30.5 | 8 | 47 | 1.22 (0.53 - 2.41) | 1.54 (1.13 - 2.05) |
| **PS-stratification / on-treatment TAR without escalation** | | | | | | | | |
| CCAE | 18,849 | 35,766 | 11.8 | 21.8 | <5 | 12 | < 0.42 (0.14 - 0.99) | 0.55 (0.28 - 0.96) |
| DAG | 3,231 | 1,228 | 2.3 | 1.2 | 0 | 0 | | |
| ODOD | 12,834 | 41,754 | 7.0 | 31.3 | 8 | 55 | 1.15 (0.50 - 2.26) | 1.76 (1.32 - 2.29) |
| MDCR | 1,489 | 7,293 | 0.8 | 5.4 | 0 | <5 | | < 0.93 (0.30 - 2.17) |
| MDCD | 2,435 | 11,946 | 1.1 | 6.2 | 0 | 0 | | |
| Open Claims | 276,521 | 842,971 | 172.3 | 672.6 | 0 | 0 | | |
| OEHR | 16,666 | 70,488 | 4.6 | 34.4 | 0 | 0 | | |
| SIDIAP | 2,943 | 2,006 | 3.5 | 3.6 | 0 | 5 | | 1.40 (0.46 - 3.27) |
| Meta-analysis | 12,834 | 41,754 | 7.0 | 31.3 | 8 | 55 | 1.15 (0.50 - 2.26) | 1.76 (1.32 - 2.29) |
| Meta-analysis (w/o USOC) | 12,834 | 41,754 | 7.0 | 31.3 | 8 | 55 | 1.15 (0.50 - 2.26) | 1.76 (1.32 - 2.29) |

##### 4.3.8 Stroke

**Supplementary Table 345:** Exposures, outcomes and risk estimates for Stroke in SGLT2I (female) (T) vs SU (female) (C) new-user cohorts under four different propensity score (PS) adjustment / time-at-risk (TAR) designs across data sources. We report total patient exposure time in 1,000 patient-year (KYr) units, uncalibrated and calibrated hazard ratios (HRs) and their 95% confidence intervals (CIs). Greyed out entries did not pass study diagnostics and are included only for completeness.

| | Patients | | Exposure (KYr) | | Outcomes | | Incidence (per KYr) | |
|---|---|---|---|---|---|---|---|---|
| | T | C | T | C | T | C | T | C |
| **PS-matching / on-treatment TAR** | | | | | | | | |
| CCAE | 15,885 | 34,645 | 11.7 | 24.7 | 43 | 108 | 3.67 (2.65 - 4.94) | 4.37 (3.59 - 5.28) |
| DAG | 3,180 | 682 | 2.5 | 0.7 | 0 | 0 | | |
| ODOD | 11,589 | 39,553 | 7.2 | 32.9 | 48 | 409 | 6.64 (4.90 - 8.80) | 12.4 (11.3 - 13.7) |
| MDCR | 1,300 | 6,486 | 0.9 | 5.2 | <5 | 93 | < 5.82 (1.89 - 13.6) | 17.8 (14.4 - 21.8) |
| MDCD | 2,298 | 11,202 | 1.2 | 7.0 | 12 | 87 | 9.70 (5.01 - 17.0) | 12.5 (9.98 - 15.4) |
| Open Claims | 255,962 | 809,471 | 192.4 | 764.9 | 577 | 5,068 | 3.00 (2.76 - 3.25) | 6.63 (6.44 - 6.81) |
| OEHR | 15,375 | 63,445 | 4.8 | 36.2 | 14 | 209 | 2.90 (1.59 - 4.87) | 5.77 (5.01 - 6.60) |
| SIDIAP | 2,830 | 956 | 3.9 | 1.7 | 10 | <5 | 2.55 (1.22 - 4.68) | < 2.99 (0.97 - 6.97) |
| Meta-analysis | 302,409 | 964,802 | 218.3 | 870.9 | <699 | 5,974 | < 3.20 (2.97 - 3.45) | 6.86 (6.69 - 7.04) |
| Meta-analysis (w/o USOC) | 46,447 | 155,331 | 25.9 | 106.0 | <122 | 906 | < 4.72 (3.92 - 5.63) | 8.54 (8.00 - 9.12) |
| **PS-stratification / on-treatment TAR** | | | | | | | | |
| CCAE | 18,673 | 35,257 | 13.5 | 24.9 | 51 | 109 | 3.77 (2.81 - 4.96) | 4.37 (3.59 - 5.27) |
| DAG | 3,231 | 1,228 | 2.5 | 1.2 | 0 | 0 | | |
| ODOD | 12,563 | 40,405 | 7.7 | 33.7 | 53 | 429 | 6.89 (5.16 - 9.01) | 12.7 (11.6 - 14.0) |
| MDCR | 1,453 | 6,939 | 0.9 | 5.6 | 5 | 102 | 5.40 (1.75 - 12.6) | 18.2 (14.9 - 22.1) |
| MDCD | 2,344 | 11,432 | 1.3 | 7.2 | 12 | 87 | 9.56 (4.94 - 16.7) | 12.2 (9.74 - 15.0) |
| Open Claims | 270,188 | 818,848 | 200.6 | 773.4 | 602 | 5,250 | 3.00 (2.77 - 3.25) | 6.79 (6.61 - 6.97) |
| OEHR | 16,552 | 69,698 | 5.2 | 40.0 | 14 | 251 | 2.69 (1.47 - 4.52) | 6.28 (5.53 - 7.10) |
| SIDIAP | 2,926 | 1,998 | 4.0 | 4.3 | 10 | 6 | 2.51 (1.20 - 4.61) | 1.39 (0.51 - 3.03) |
| Meta-analysis | 317,976 | 964,208 | 227.1 | 872.1 | 720 | 6,039 | 3.17 (2.94 - 3.41) | 6.92 (6.75 - 7.10) |
| Meta-analysis (w/o USOC) | 47,788 | 145,360 | 26.4 | 98.6 | 118 | 789 | 4.47 (3.70 - 5.35) | 8.00 (7.45 - 8.58) |
| **PS-matching / on-treatment TAR without escalation** | | | | | | | | |
| CCAE | 15,885 | 34,645 | 10.1 | 21.2 | 34 | 92 | 3.37 (2.33 - 4.71) | 4.33 (3.49 - 5.31) |
| DAG | 3,180 | 682 | 2.3 | 0.6 | 0 | 0 | | |
| ODOD | 11,589 | 39,553 | 6.4 | 29.4 | 45 | 361 | 7.03 (5.13 - 9.41) | 12.3 (11.1 - 13.6) |
| MDCR | 1,300 | 6,486 | 0.8 | 4.8 | <5 | 85 | < 6.56 (2.13 - 15.3) | 17.8 (14.2 - 22.1) |
| MDCD | 2,298 | 11,202 | 1.0 | 5.8 | 6 | 75 | 5.85 (2.15 - 12.7) | 13.0 (10.2 - 16.3) |
| Open Claims | 255,962 | 809,471 | 161.5 | 644.2 | 482 | 4,307 | 2.98 (2.72 - 3.26) | 6.69 (6.49 - 6.89) |
| OEHR | 15,375 | 63,445 | 4.2 | 30.7 | 9 | 167 | 2.15 (0.98 - 4.08) | 5.43 (4.64 - 6.32) |
| SIDIAP | 2,830 | 956 | 3.4 | 1.4 | 10 | <5 | 2.91 (1.39 - 5.35) | < 3.54 (1.15 - 8.27) |
| Meta-analysis | 302,409 | 964,802 | 184.0 | 736.1 | <581 | 5,087 | < 3.16 (2.91 - 3.43) | 6.91 (6.72 - 7.10) |
| Meta-analysis (w/o USOC) | 46,447 | 155,331 | 22.5 | 91.9 | <99 | 780 | < 4.41 (3.58 - 5.36) | 8.49 (7.90 - 9.11) |
| **PS-stratification / on-treatment TAR without escalation** | | | | | | | | |
| CCAE | 18,673 | 35,257 | 11.7 | 21.4 | 41 | 93 | 3.51 (2.52 - 4.77) | 4.34 (3.50 - 5.31) |
| DAG | 3,231 | 1,228 | 2.3 | 1.2 | 0 | 0 | | |
| ODOD | 12,563 | 40,405 | 6.8 | 30.1 | 50 | 380 | 7.33 (5.44 - 9.66) | 12.6 (11.4 - 14.0) |
| MDCR | 1,453 | 6,939 | 0.8 | 5.1 | <5 | 93 | < 6.07 (1.97 - 14.2) | 18.2 (14.7 - 22.3) |
| MDCD | 2,344 | 11,432 | 1.0 | 5.9 | 6 | 75 | 5.75 (2.11 - 12.5) | 12.7 (9.96 - 15.9) |
| Open Claims | 270,188 | 818,848 | 168.7 | 651.7 | 505 | 4,463 | 2.99 (2.74 - 3.27) | 6.85 (6.65 - 7.05) |
| OEHR | 16,552 | 69,698 | 4.5 | 33.9 | 9 | 194 | 1.99 (0.91 - 3.78) | 5.72 (4.95 - 6.59) |
| SIDIAP | 2,926 | 1,998 | 3.5 | 3.5 | 10 | 6 | 2.86 (1.37 - 5.27) | 1.69 (0.62 - 3.69) |
| Meta-analysis | 317,976 | 964,208 | 191.7 | 737.2 | 605 | 5,130 | 3.16 (2.91 - 3.42) | 6.96 (6.77 - 7.15) |
| Meta-analysis (w/o USOC) | 47,788 | 145,360 | 23.0 | 85.4 | 100 | 667 | 4.35 (3.54 - 5.29) | 7.81 (7.23 - 8.42) |

##### 4.3.9 Sudden cardiac death

**Supplementary Table 346:** Exposures, outcomes and risk estimates for Sudden cardiac death in SGLT2I (female) (T) vs SU (female) (C) new-user cohorts under four different propensity score (PS) adjustment / time-at-risk (TAR) designs across data sources. We report total patient exposure time in 1,000 patient-year (KYr) units, uncalibrated and calibrated hazard ratios (HRs) and their 95% confidence intervals (CIs). Greyed out entries did not pass study diagnostics and are included only for completeness.

| | Patients | | Exposure (KYr) | | Outcomes | | Incidence (per KYr) | |
|---|---|---|---|---|---|---|---|---|
| | T | C | T | C | T | C | T | C |
| **PS-matching / on-treatment TAR** | | | | | | | | |
| CCAE | 16,073 | 35,171 | 11.9 | 25.1 | 7 | 20 | 0.59 (0.24 - 1.22) | 0.80 (0.49 - 1.23) |
| DAG | 3,180 | 682 | 2.5 | 0.7 | 0 | 0 | | |
| ODOD | 11,832 | 40,927 | 7.4 | 34.3 | 6 | 118 | 0.81 (0.30 - 1.77) | 3.44 (2.85 - 4.12) |
| MDCR | 1,332 | 6,788 | 0.9 | 5.5 | 0 | 17 | | 3.10 (1.80 - 4.96) |
| MDCD | 2,386 | 11,636 | 1.3 | 7.3 | <5 | 20 | < 3.88 (1.26 - 9.06) | 2.74 (1.68 - 4.24) |
| Open Claims | 261,446 | 831,503 | 196.2 | 787.7 | 115 | 1,013 | 0.59 (0.48 - 0.70) | 1.29 (1.21 - 1.37) |
| OEHR | 15,487 | 64,209 | 4.9 | 36.8 | <5 | 52 | < 1.03 (0.33 - 2.40) | 1.41 (1.05 - 1.85) |
| SIDIAP | 2,849 | 965 | 4.0 | 1.7 | 0 | 0 | | |
| Meta-analysis | 304,838 | 971,810 | 220.3 | 883.9 | <133 | 1,203 | < 0.60 (0.51 - 0.72) | 1.36 (1.29 - 1.44) |
| Meta-analysis (w/o USOC) | 43,392 | 140,307 | 24.1 | 96.2 | <18 | 190 | < 0.75 (0.44 - 1.18) | 1.97 (1.70 - 2.28) |
| **PS-stratification / on-treatment TAR** | | | | | | | | |
| CCAE | 18,867 | 35,802 | 13.7 | 25.4 | 8 | 20 | 0.58 (0.25 - 1.15) | 0.79 (0.48 - 1.22) |
| DAG | 3,231 | 1,228 | 2.5 | 1.2 | 0 | 0 | | |
| ODOD | 12,828 | 41,824 | 7.9 | 35.1 | 6 | 127 | 0.76 (0.28 - 1.66) | 3.61 (3.01 - 4.30) |
| MDCR | 1,487 | 7,275 | 0.9 | 5.9 | 0 | 17 | | 2.88 (1.68 - 4.61) |
| MDCD | 2,433 | 11,899 | 1.3 | 7.5 | <5 | 20 | < 3.83 (1.24 - 8.93) | 2.67 (1.63 - 4.13) |
| Open Claims | 276,028 | 841,576 | 204.6 | 797.0 | 121 | 1,054 | 0.59 (0.49 - 0.71) | 1.32 (1.24 - 1.40) |
| OEHR | 16,657 | 70,454 | 5.2 | 40.6 | <5 | 69 | < 0.96 (0.31 - 2.23) | 1.70 (1.32 - 2.15) |
| SIDIAP | 2,949 | 2,013 | 4.0 | 4.3 | 0 | 0 | | |
| Meta-analysis | 324,380 | 989,656 | 231.4 | 898.1 | <140 | 1,270 | < 0.61 (0.51 - 0.71) | 1.41 (1.34 - 1.49) |
| Meta-analysis (w/o USOC) | 48,352 | 148,080 | 26.8 | 101.1 | <19 | 216 | < 0.71 (0.43 - 1.11) | 2.14 (1.86 - 2.44) |
| **PS-matching / on-treatment TAR without escalation** | | | | | | | | |
| CCAE | 16,073 | 35,171 | 10.2 | 21.6 | 6 | 17 | 0.59 (0.22 - 1.28) | 0.79 (0.46 - 1.26) |
| DAG | 3,180 | 682 | 2.3 | 0.6 | 0 | 0 | | |
| ODOD | 11,832 | 40,927 | 6.5 | 30.6 | 6 | 99 | 0.92 (0.34 - 2.00) | 3.24 (2.63 - 3.94) |
| MDCR | 1,332 | 6,788 | 0.8 | 5.0 | 0 | 17 | | 3.40 (1.98 - 5.45) |
| MDCD | 2,386 | 11,636 | 1.1 | 6.0 | <5 | 18 | < 4.71 (1.53 - 11.0) | 2.99 (1.77 - 4.72) |
| Open Claims | 261,446 | 831,503 | 164.7 | 663.2 | 99 | 849 | 0.60 (0.49 - 0.73) | 1.28 (1.20 - 1.37) |
| OEHR | 15,487 | 64,209 | 4.2 | 31.2 | <5 | 34 | < 1.18 (0.38 - 2.76) | 1.09 (0.75 - 1.52) |
| SIDIAP | 2,849 | 965 | 3.5 | 1.4 | 0 | 0 | | |
| Meta-analysis | 304,838 | 971,810 | 185.6 | 746.6 | <116 | 999 | < 0.62 (0.52 - 0.75) | 1.34 (1.26 - 1.42) |
| Meta-analysis (w/o USOC) | 43,392 | 140,307 | 21.0 | 83.4 | <17 | 150 | < 0.81 (0.47 - 1.30) | 1.80 (1.52 - 2.11) |
| **PS-stratification / on-treatment TAR without escalation** | | | | | | | | |
| CCAE | 18,867 | 35,802 | 11.8 | 21.8 | 7 | 17 | 0.59 (0.24 - 1.22) | 0.78 (0.45 - 1.25) |
| DAG | 3,231 | 1,228 | 2.3 | 1.2 | 0 | 0 | | |
| ODOD | 12,828 | 41,824 | 7.0 | 31.4 | 6 | 108 | 0.86 (0.32 - 1.87) | 3.44 (2.83 - 4.16) |
| MDCR | 1,487 | 7,275 | 0.8 | 5.4 | 0 | 17 | | 3.16 (1.84 - 5.06) |
| MDCD | 2,433 | 11,899 | 1.1 | 6.2 | <5 | 18 | < 4.63 (1.50 - 10.8) | 2.91 (1.72 - 4.60) |
| Open Claims | 276,028 | 841,576 | 172.0 | 671.4 | 105 | 887 | 0.61 (0.50 - 0.74) | 1.32 (1.24 - 1.41) |
| OEHR | 16,657 | 70,454 | 4.5 | 34.4 | <5 | 45 | < 1.10 (0.36 - 2.56) | 1.31 (0.96 - 1.75) |
| SIDIAP | 2,949 | 2,013 | 3.5 | 3.6 | 0 | 0 | | |
| Meta-analysis | 324,380 | 989,656 | 195.3 | 758.9 | <123 | 1,057 | < 0.63 (0.52 - 0.75) | 1.39 (1.31 - 1.48) |
| Meta-analysis (w/o USOC) | 48,352 | 148,080 | 23.3 | 87.5 | <18 | 170 | < 0.77 (0.46 - 1.22) | 1.94 (1.66 - 2.26) |

##### 4.3.10 Abnormal weight gain

**Supplementary Table 347:** Exposures, outcomes and risk estimates for Abnormal weight gain in SGLT2I (female) (T) vs SU (female) (C) new-user cohorts under four different propensity score (PS) adjustment / time-at-risk (TAR) designs across data sources. We report total patient exposure time in 1,000 patient-year (KYr) units, uncalibrated and calibrated hazard ratios (HRs) and their 95% confidence intervals (CIs). Greyed out entries did not pass study diagnostics and are included only for completeness.

| | Patients | | Exposure (KYr) | | Outcomes | | Incidence (per KYr) | |
|---|---|---|---|---|---|---|---|---|
| | T | C | T | C | T | C | T | C |
| **PS-matching / on-treatment TAR** | | | | | | | | |
| CCAE | 14,730 | 33,074 | 10.9 | 23.6 | 68 | 216 | 6.27 (4.87 - 7.94) | 9.16 (7.98 - 10.5) |
| DAG | 3,132 | 671 | 2.5 | 0.6 | <5 | <5 | < 2.04 (0.66 - 4.75) | < 7.73 (2.51 - 18.0) |
| ODOD | 11,025 | 39,200 | 6.8 | 32.7 | 49 | 283 | 7.22 (5.34 - 9.54) | 8.66 (7.68 - 9.73) |
| MDCR | 1,302 | 6,684 | 0.9 | 5.4 | <5 | 27 | < 5.86 (1.90 - 13.7) | 5.01 (3.30 - 7.28) |
| MDCD | 2,135 | 10,801 | 1.2 | 6.8 | 5 | 84 | 4.32 (1.40 - 10.1) | 12.4 (9.85 - 15.3) |
| Open Claims | 242,662 | 794,057 | 181.8 | 752.7 | 448 | 2,374 | 2.46 (2.24 - 2.70) | 3.15 (3.03 - 3.28) |
| OEHR | 14,423 | 61,417 | 4.5 | 34.9 | 27 | 328 | 6.01 (3.96 - 8.74) | 9.39 (8.40 - 10.5) |
| SIDIAP | 2,818 | 956 | 3.9 | 1.7 | <5 | <5 | < 1.28 (0.41 - 2.98) | < 3.00 (0.97 - 7.01) |
| Meta-analysis | 286,277 | 945,233 | 205.9 | 856.0 | <602 | 3,312 | < 2.92 (2.69 - 3.17) | 3.87 (3.74 - 4.00) |
| Meta-analysis (w/o USOC) | 43,615 | 151,176 | 24.1 | 103.4 | <154 | 938 | < 6.38 (5.41 - 7.47) | 9.07 (8.50 - 9.67) |
| **PS-stratification / on-treatment TAR** | | | | | | | | |
| CCAE | 17,049 | 33,731 | 12.4 | 23.8 | 80 | 218 | 6.46 (5.12 - 8.04) | 9.14 (7.97 - 10.4) |
| DAG | 3,181 | 1,213 | 2.5 | 1.2 | <5 | <5 | < 2.02 (0.65 - 4.70) | < 4.09 (1.33 - 9.54) |
| ODOD | 11,835 | 40,059 | 7.2 | 33.5 | 54 | 299 | 7.54 (5.66 - 9.84) | 8.93 (7.94 - 10.00) |
| MDCR | 1,455 | 7,166 | 0.9 | 5.8 | <5 | 29 | < 5.43 (1.76 - 12.7) | 5.00 (3.35 - 7.18) |
| MDCD | 2,177 | 11,027 | 1.2 | 7.0 | 5 | 86 | 4.26 (1.38 - 9.93) | 12.4 (9.88 - 15.3) |
| Open Claims | 255,345 | 804,381 | 188.9 | 762.3 | 486 | 2,422 | 2.57 (2.35 - 2.81) | 3.18 (3.05 - 3.31) |
| OEHR | 15,455 | 67,433 | 4.8 | 38.5 | 29 | 369 | 6.02 (4.03 - 8.64) | 9.58 (8.63 - 10.6) |
| SIDIAP | 2,919 | 1,994 | 4.0 | 4.3 | <5 | 6 | < 1.26 (0.41 - 2.94) | 1.40 (0.51 - 3.04) |
| Meta-analysis | 299,684 | 945,604 | 213.3 | 858.1 | 649 | 3,308 | 3.04 (2.81 - 3.29) | 3.85 (3.72 - 3.99) |
| Meta-analysis (w/o USOC) | 44,339 | 141,223 | 24.4 | 95.9 | 163 | 886 | 6.69 (5.70 - 7.80) | 9.24 (8.64 - 9.87) |
| **PS-matching / on-treatment TAR without escalation** | | | | | | | | |
| CCAE | 14,730 | 33,074 | 9.3 | 20.3 | 56 | 176 | 6.00 (4.53 - 7.79) | 8.66 (7.43 - 10.0) |
| DAG | 3,132 | 671 | 2.2 | 0.6 | <5 | <5 | < 2.23 (0.72 - 5.19) | < 8.38 (2.72 - 19.5) |
| ODOD | 11,025 | 39,200 | 6.0 | 29.2 | 44 | 242 | 7.30 (5.31 - 9.81) | 8.28 (7.27 - 9.39) |
| MDCR | 1,302 | 6,684 | 0.8 | 4.9 | <5 | 21 | < 6.63 (2.15 - 15.5) | 4.27 (2.65 - 6.53) |
| MDCD | 2,135 | 10,801 | 1.0 | 5.6 | <5 | 70 | < 5.25 (1.70 - 12.3) | 12.4 (9.69 - 15.7) |
| Open Claims | 242,662 | 794,057 | 152.7 | 635.2 | 357 | 1,960 | 2.34 (2.10 - 2.59) | 3.09 (2.95 - 3.23) |
| OEHR | 14,423 | 61,417 | 3.9 | 29.7 | 22 | 259 | 5.62 (3.52 - 8.51) | 8.73 (7.70 - 9.86) |
| SIDIAP | 2,818 | 956 | 3.4 | 1.4 | <5 | <5 | < 1.46 (0.47 - 3.40) | < 3.56 (1.16 - 8.32) |
| Meta-analysis | 284,975 | 938,549 | 172.9 | 720.0 | <484 | 2,707 | < 2.80 (2.56 - 3.06) | 3.76 (3.62 - 3.90) |
| Meta-analysis (w/o USOC) | 42,313 | 144,492 | 20.2 | 84.9 | <127 | 747 | < 6.28 (5.24 - 7.47) | 8.80 (8.18 - 9.46) |
| **PS-stratification / on-treatment TAR without escalation** | | | | | | | | |
| CCAE | 17,049 | 33,731 | 10.7 | 20.6 | 68 | 178 | 6.38 (4.96 - 8.09) | 8.66 (7.43 - 10.0) |
| DAG | 3,181 | 1,213 | 2.3 | 1.1 | <5 | <5 | < 2.21 (0.72 - 5.15) | < 4.43 (1.44 - 10.3) |
| ODOD | 11,835 | 40,059 | 6.4 | 30.0 | 48 | 257 | 7.54 (5.56 - 10.00) | 8.57 (7.56 - 9.69) |
| MDCR | 1,455 | 7,166 | 0.8 | 5.3 | <5 | 23 | < 6.12 (1.99 - 14.3) | 4.36 (2.76 - 6.54) |
| MDCD | 2,177 | 11,027 | 1.0 | 5.8 | <5 | 72 | < 5.17 (1.68 - 12.1) | 12.5 (9.76 - 15.7) |
| Open Claims | 255,345 | 804,381 | 159.0 | 643.5 | 388 | 2,004 | 2.44 (2.20 - 2.70) | 3.11 (2.98 - 3.25) |
| OEHR | 15,455 | 67,433 | 4.2 | 32.7 | 24 | 290 | 5.71 (3.66 - 8.49) | 8.86 (7.87 - 9.94) |
| SIDIAP | 2,919 | 1,994 | 3.5 | 3.5 | <5 | 6 | < 1.43 (0.47 - 3.35) | 1.69 (0.62 - 3.68) |
| Meta-analysis | 299,684 | 945,604 | 180.2 | 726.8 | 528 | 2,729 | 2.93 (2.69 - 3.19) | 3.75 (3.62 - 3.90) |
| Meta-analysis (w/o USOC) | 44,339 | 141,223 | 21.2 | 83.3 | 140 | 725 | 6.59 (5.55 - 7.78) | 8.71 (8.09 - 9.37) |

##### 4.3.11 Abnormal weight loss

**Supplementary Table 348:** Exposures, outcomes and risk estimates for Abnormal weight loss in SGLT2I (female) (T) vs SU (female) (C) new-user cohorts under four different propensity score (PS) adjustment / time-at-risk (TAR) designs across data sources. We report total patient exposure time in 1,000 patient-year (KYr) units, uncalibrated and calibrated hazard ratios (HRs) and their 95% confidence intervals (CIs). Greyed out entries did not pass study diagnostics and are included only for completeness.

| | Patients | | Exposure (KYr) | | Outcomes | | Incidence (per KYr) | |
|---|---|---|---|---|---|---|---|---|
| | T | C | T | C | T | C | T | C |
| **PS-matching / on-treatment TAR** | | | | | | | | |
| CCAE | 15,708 | 34,218 | 11.5 | 24.4 | 93 | 147 | 8.07 (6.51 - 9.88) | 6.02 (5.08 - 7.07) |
| DAG | 3,100 | 659 | 2.4 | 0.6 | 21 | <5 | 8.63 (5.35 - 13.2) | < 7.94 (2.58 - 18.5) |
| ODOD | 11,398 | 39,009 | 7.1 | 32.4 | 107 | 469 | 15.1 (12.4 - 18.3) | 14.5 (13.2 - 15.9) |
| MDCR | 1,287 | 6,450 | 0.9 | 5.2 | 10 | 70 | 11.7 (5.62 - 21.6) | 13.5 (10.5 - 17.0) |
| MDCD | 2,284 | 11,065 | 1.2 | 6.8 | 24 | 111 | 19.6 (12.5 - 29.1) | 16.3 (13.4 - 19.6) |
| Open Claims | 252,941 | 802,749 | 189.5 | 758.2 | 1,026 | 3,898 | 5.42 (5.09 - 5.76) | 5.14 (4.98 - 5.31) |
| OEHR | 15,065 | 62,231 | 4.7 | 35.3 | 43 | 380 | 9.13 (6.60 - 12.3) | 10.8 (9.70 - 11.9) |
| SIDIAP | 2,818 | 953 | 3.9 | 1.7 | 17 | 6 | 4.33 (2.52 - 6.94) | 3.60 (1.32 - 7.83) |
| Meta-analysis | 301,501 | 956,675 | 218.8 | 864.0 | 1,320 | 5,081 | 6.03 (5.71 - 6.37) | 5.88 (5.72 - 6.04) |
| Meta-analysis (w/o USOC) | 48,560 | 153,926 | 29.3 | 105.8 | 294 | 1,183 | 10.0 (8.91 - 11.2) | 11.2 (10.6 - 11.8) |
| **PS-stratification / on-treatment TAR** | | | | | | | | |
| CCAE | 18,444 | 34,899 | 13.3 | 24.7 | 112 | 149 | 8.41 (6.93 - 10.1) | 6.03 (5.10 - 7.09) |
| DAG | 3,151 | 1,186 | 2.5 | 1.2 | 21 | 8 | 8.55 (5.29 - 13.1) | 6.78 (2.92 - 13.3) |
| ODOD | 12,379 | 39,757 | 7.5 | 33.1 | 115 | 489 | 15.2 (12.6 - 18.3) | 14.8 (13.5 - 16.1) |
| MDCR | 1,438 | 6,921 | 0.9 | 5.6 | 11 | 79 | 12.0 (5.99 - 21.5) | 14.2 (11.2 - 17.7) |
| MDCD | 2,330 | 11,349 | 1.2 | 7.0 | 24 | 114 | 19.3 (12.4 - 28.7) | 16.2 (13.4 - 19.5) |
| Open Claims | 266,948 | 812,384 | 197.5 | 767.2 | 1,078 | 4,035 | 5.46 (5.14 - 5.79) | 5.26 (5.10 - 5.42) |
| OEHR | 16,231 | 68,316 | 5.1 | 39.0 | 46 | 440 | 9.07 (6.64 - 12.1) | 11.3 (10.3 - 12.4) |
| SIDIAP | 2,919 | 1,995 | 4.0 | 4.3 | 17 | 12 | 4.27 (2.49 - 6.84) | 2.79 (1.44 - 4.88) |
| Meta-analysis | 314,002 | 955,356 | 223.5 | 863.9 | 1,351 | 5,113 | 6.05 (5.73 - 6.38) | 5.92 (5.76 - 6.08) |
| Meta-analysis (w/o USOC) | 47,054 | 142,972 | 25.9 | 96.8 | 273 | 1,078 | 10.5 (9.32 - 11.9) | 11.1 (10.5 - 11.8) |
| **PS-matching / on-treatment TAR without escalation** | | | | | | | | |
| CCAE | 15,708 | 34,218 | 9.9 | 21.0 | 76 | 119 | 7.65 (6.03 - 9.58) | 5.67 (4.70 - 6.78) |
| DAG | 3,100 | 659 | 2.2 | 0.6 | 19 | <5 | 8.58 (5.17 - 13.4) | < 8.60 (2.79 - 20.1) |
| ODOD | 11,398 | 39,009 | 6.3 | 28.9 | 94 | 415 | 15.0 (12.1 - 18.4) | 14.3 (13.0 - 15.8) |
| MDCR | 1,287 | 6,450 | 0.8 | 4.7 | 9 | 60 | 11.9 (5.45 - 22.6) | 12.7 (9.67 - 16.3) |
| MDCD | 2,284 | 11,065 | 1.0 | 5.7 | 21 | 97 | 20.7 (12.8 - 31.7) | 17.1 (13.9 - 20.9) |
| Open Claims | 252,941 | 802,749 | 159.1 | 638.7 | 864 | 3,279 | 5.43 (5.07 - 5.80) | 5.13 (4.96 - 5.31) |
| OEHR | 15,065 | 62,231 | 4.1 | 30.0 | 37 | 298 | 9.02 (6.35 - 12.4) | 9.94 (8.84 - 11.1) |
| SIDIAP | 2,818 | 953 | 3.4 | 1.4 | 15 | <5 | 4.36 (2.44 - 7.20) | < 3.57 (1.16 - 8.33) |
| Meta-analysis | 298,683 | 955,722 | 181.2 | 729.0 | 1,101 | 4,268 | 6.08 (5.72 - 6.45) | 5.85 (5.68 - 6.03) |
| Meta-analysis (w/o USOC) | 45,742 | 152,973 | 22.1 | 90.3 | 237 | 989 | 10.7 (9.42 - 12.2) | 11.0 (10.3 - 11.7) |
| **PS-stratification / on-treatment TAR without escalation** | | | | | | | | |
| CCAE | 18,444 | 34,899 | 11.5 | 21.2 | 93 | 121 | 8.10 (6.54 - 9.92) | 5.70 (4.73 - 6.81) |
| DAG | 3,151 | 1,186 | 2.2 | 1.1 | 19 | 7 | 8.50 (5.12 - 13.3) | 6.42 (2.58 - 13.2) |
| ODOD | 12,379 | 39,757 | 6.7 | 29.6 | 100 | 435 | 15.0 (12.2 - 18.2) | 14.7 (13.4 - 16.2) |
| MDCR | 1,438 | 6,921 | 0.8 | 5.1 | 10 | 68 | 12.3 (5.89 - 22.6) | 13.4 (10.4 - 17.0) |
| MDCD | 2,330 | 11,349 | 1.0 | 5.8 | 21 | 100 | 20.4 (12.6 - 31.2) | 17.2 (14.0 - 20.9) |
| Open Claims | 266,948 | 812,384 | 166.1 | 646.5 | 907 | 3,398 | 5.46 (5.11 - 5.83) | 5.26 (5.08 - 5.44) |
| OEHR | 16,231 | 68,316 | 4.4 | 33.0 | 39 | 347 | 8.82 (6.27 - 12.1) | 10.5 (9.42 - 11.7) |
| SIDIAP | 2,919 | 1,995 | 3.5 | 3.5 | 15 | 9 | 4.30 (2.40 - 7.09) | 2.54 (1.16 - 4.83) |
| Meta-analysis | 314,002 | 955,356 | 188.7 | 730.4 | 1,139 | 4,301 | 6.04 (5.69 - 6.40) | 5.89 (5.71 - 6.07) |
| Meta-analysis (w/o USOC) | 47,054 | 142,972 | 22.6 | 83.8 | 232 | 903 | 10.3 (8.99 - 11.7) | 10.8 (10.1 - 11.5) |

##### 4.3.12 Acute pancreatitis

**Supplementary Table 349:** Exposures, outcomes and risk estimates for Acute pancreatitis in SGLT2I (female) (T) vs SU (female) (C) new-user cohorts under four different propensity score (PS) adjustment / time-at-risk (TAR) designs across data sources. We report total patient exposure time in 1,000 patient-year (KYr) units, uncalibrated and calibrated hazard ratios (HRs) and their 95% confidence intervals (CIs). Greyed out entries did not pass study diagnostics and are included only for completeness.

| | Patients | | Exposure (KYr) | | Outcomes | | Incidence (per KYr) | |
|---|---|---|---|---|---|---|---|---|
| | T | C | T | C | T | C | T | C |
| **PS-matching / on-treatment TAR** | | | | | | | | |
| CCAE | 15,936 | 34,825 | 11.8 | 24.9 | 21 | 58 | 1.79 (1.11 - 2.73) | 2.33 (1.77 - 3.01) |
| DAG | 3,180 | 682 | 2.5 | 0.7 | 0 | 0 | | |
| ODOD | 11,759 | 40,592 | 7.3 | 34.0 | 16 | 101 | 2.18 (1.25 - 3.54) | 2.97 (2.42 - 3.61) |
| MDCR | 1,317 | 6,709 | 0.9 | 5.4 | <5 | 22 | < 5.73 (1.86 - 13.4) | 4.06 (2.54 - 6.14) |
| MDCD | 2,344 | 11,389 | 1.3 | 7.1 | 9 | 36 | 7.15 (3.27 - 13.6) | 5.05 (3.54 - 6.99) |
| Open Claims | 258,324 | 821,575 | 193.9 | 777.8 | 260 | 1,636 | 1.34 (1.18 - 1.51) | 2.10 (2.00 - 2.21) |
| OEHR | 15,419 | 63,891 | 4.8 | 36.6 | 9 | 76 | 1.86 (0.85 - 3.53) | 2.08 (1.64 - 2.60) |
| SIDIAP | 2,840 | 960 | 4.0 | 1.7 | <5 | 0 | < 1.26 (0.41 - 2.95) | |
| Meta-analysis | 303,782 | 972,272 | 219.1 | 880.4 | 315 | 1,907 | 1.44 (1.28 - 1.61) | 2.17 (2.07 - 2.27) |
| Meta-analysis (w/o USOC) | 45,458 | 150,697 | 25.2 | 102.5 | 55 | 271 | 2.18 (1.64 - 2.84) | 2.64 (2.34 - 2.98) |
| **PS-stratification / on-treatment TAR** | | | | | | | | |
| CCAE | 18,712 | 35,440 | 13.6 | 25.1 | 22 | 59 | 1.62 (1.02 - 2.45) | 2.35 (1.79 - 3.03) |
| DAG | 3,231 | 1,228 | 2.5 | 1.2 | 0 | 0 | | |
| ODOD | 12,741 | 41,471 | 7.8 | 34.8 | 17 | 104 | 2.18 (1.27 - 3.49) | 2.99 (2.44 - 3.62) |
| MDCR | 1,473 | 7,194 | 0.9 | 5.8 | <5 | 22 | < 5.33 (1.73 - 12.4) | 3.78 (2.37 - 5.72) |
| MDCD | 2,390 | 11,698 | 1.3 | 7.3 | 9 | 39 | 7.05 (3.22 - 13.4) | 5.31 (3.77 - 7.26) |
| Open Claims | 272,688 | 831,302 | 202.2 | 786.9 | 275 | 1,680 | 1.36 (1.20 - 1.53) | 2.13 (2.03 - 2.24) |
| OEHR | 16,582 | 70,088 | 5.2 | 40.3 | 9 | 83 | 1.73 (0.79 - 3.28) | 2.06 (1.64 - 2.55) |
| SIDIAP | 2,941 | 2,005 | 4.0 | 4.3 | <5 | <5 | < 1.25 (0.40 - 2.91) | < 1.16 (0.38 - 2.70) |
| Meta-analysis | 320,723 | 978,301 | 228.8 | 887.2 | 323 | 1,926 | 1.41 (1.26 - 1.57) | 2.17 (2.08 - 2.27) |
| Meta-analysis (w/o USOC) | 48,035 | 146,999 | 26.6 | 100.3 | 48 | 246 | 1.81 (1.33 - 2.39) | 2.45 (2.16 - 2.78) |
| **PS-matching / on-treatment TAR without escalation** | | | | | | | | |
| CCAE | 15,936 | 34,825 | 10.1 | 21.4 | 16 | 43 | 1.58 (0.90 - 2.57) | 2.01 (1.45 - 2.71) |
| DAG | 3,180 | 682 | 2.3 | 0.6 | 0 | 0 | | |
| ODOD | 11,759 | 40,592 | 6.5 | 30.3 | 12 | 84 | 1.85 (0.95 - 3.23) | 2.77 (2.21 - 3.43) |
| MDCR | 1,317 | 6,709 | 0.8 | 4.9 | <5 | 21 | < 6.49 (2.11 - 15.1) | 4.26 (2.63 - 6.50) |
| MDCD | 2,344 | 11,389 | 1.0 | 5.9 | 8 | 33 | 7.71 (3.33 - 15.2) | 5.60 (3.85 - 7.86) |
| Open Claims | 258,324 | 821,575 | 162.8 | 655.3 | 201 | 1,343 | 1.23 (1.07 - 1.42) | 2.05 (1.94 - 2.16) |
| OEHR | 15,419 | 63,891 | 4.2 | 31.0 | 7 | 62 | 1.66 (0.67 - 3.42) | 2.00 (1.53 - 2.56) |
| SIDIAP | 2,840 | 960 | 3.5 | 1.4 | <5 | 0 | < 1.44 (0.47 - 3.36) | |
| Meta-analysis | 303,782 | 972,272 | 184.7 | 743.9 | 244 | 1,565 | 1.32 (1.16 - 1.50) | 2.10 (2.00 - 2.21) |
| Meta-analysis (w/o USOC) | 45,458 | 150,697 | 21.9 | 88.6 | 43 | 222 | 1.97 (1.42 - 2.65) | 2.51 (2.19 - 2.86) |
| **PS-stratification / on-treatment TAR without escalation** | | | | | | | | |
| CCAE | 18,712 | 35,440 | 11.7 | 21.6 | 17 | 44 | 1.45 (0.85 - 2.33) | 2.04 (1.48 - 2.73) |
| DAG | 3,231 | 1,228 | 2.3 | 1.2 | 0 | 0 | | |
| ODOD | 12,741 | 41,471 | 6.9 | 31.1 | 13 | 86 | 1.88 (1.00 - 3.21) | 2.77 (2.21 - 3.42) |
| MDCR | 1,473 | 7,194 | 0.8 | 5.3 | <5 | 21 | < 6.01 (1.95 - 14.0) | 3.96 (2.45 - 6.05) |
| MDCD | 2,390 | 11,698 | 1.1 | 6.1 | 8 | 34 | 7.58 (3.27 - 14.9) | 5.59 (3.87 - 7.81) |
| Open Claims | 272,688 | 831,302 | 170.1 | 663.2 | 211 | 1,380 | 1.24 (1.08 - 1.42) | 2.08 (1.97 - 2.19) |
| OEHR | 16,582 | 70,088 | 4.5 | 34.2 | 7 | 68 | 1.54 (0.62 - 3.18) | 1.99 (1.55 - 2.52) |
| SIDIAP | 2,941 | 2,005 | 3.5 | 3.6 | <5 | <5 | < 1.42 (0.46 - 3.31) | < 1.40 (0.46 - 3.27) |
| Meta-analysis | 320,723 | 978,301 | 193.2 | 750.1 | 248 | 1,578 | 1.28 (1.13 - 1.45) | 2.10 (2.00 - 2.21) |
| Meta-analysis (w/o USOC) | 48,035 | 146,999 | 23.2 | 86.9 | 37 | 198 | 1.60 (1.12 - 2.20) | 2.28 (1.97 - 2.62) |

##### 4.3.13 Bone fracture

**Supplementary Table 350:** Exposures, outcomes and risk estimates for Bone fracture in SGLT2I (female) (T) vs SU (female) (C) new-user cohorts under four different propensity score (PS) adjustment / time-at-risk (TAR) designs across data sources. We report total patient exposure time in 1,000 patient-year (KYr) units, uncalibrated and calibrated hazard ratios (HRs) and their 95% confidence intervals (CIs). Greyed out entries did not pass study diagnostics and are included only for completeness.

| | Patients | | Exposure (KYr) | | Outcomes | | Incidence (per KYr) | |
|---|---|---|---|---|---|---|---|---|
| | T | C | T | C | T | C | T | C |
| **PS-matching / on-treatment TAR** | | | | | | | | |
| CCAE | 14,510 | 31,898 | 10.5 | 22.4 | 188 | 453 | 17.9 (15.5 - 20.7) | 20.2 (18.4 - 22.2) |
| DAG | 2,771 | 603 | 2.2 | 0.6 | 31 | 17 | 14.4 (9.78 - 20.4) | 30.2 (17.6 - 48.3) |
| ODOD | 10,623 | 36,345 | 6.5 | 29.4 | 217 | 1,069 | 33.3 (29.0 - 38.0) | 36.3 (34.2 - 38.6) |
| MDCR | 1,155 | 5,643 | 0.7 | 4.4 | 34 | 210 | 46.5 (32.2 - 65.0) | 47.3 (41.1 - 54.2) |
| MDCD | 2,052 | 10,040 | 1.1 | 6.1 | 32 | 222 | 29.6 (20.3 - 41.8) | 36.2 (31.6 - 41.3) |
| Open Claims | 225,774 | 719,920 | 168.0 | 667.2 | 2,702 | 14,367 | 16.1 (15.5 - 16.7) | 21.5 (21.2 - 21.9) |
| OEHR | 14,291 | 58,744 | 4.5 | 32.8 | 66 | 804 | 14.8 (11.4 - 18.8) | 24.5 (22.8 - 26.2) |
| SIDIAP | 2,422 | 853 | 3.4 | 1.5 | 52 | 23 | 15.3 (11.5 - 20.1) | 15.6 (9.91 - 23.5) |
| Meta-analysis | 270,827 | 863,443 | 194.7 | 763.9 | 3,291 | 17,148 | 16.9 (16.3 - 17.5) | 22.4 (22.1 - 22.8) |
| Meta-analysis (w/o USOC) | 45,053 | 143,523 | 26.7 | 96.7 | 589 | 2,781 | 22.1 (20.3 - 23.9) | 28.8 (27.7 - 29.9) |
| **PS-stratification / on-treatment TAR** | | | | | | | | |
| CCAE | 16,973 | 32,516 | 12.0 | 22.6 | 230 | 458 | 19.1 (16.7 - 21.7) | 20.2 (18.4 - 22.2) |
| DAG | 2,815 | 1,043 | 2.2 | 1.0 | 31 | 30 | 14.2 (9.67 - 20.2) | 29.2 (19.7 - 41.7) |
| ODOD | 11,478 | 37,125 | 6.9 | 30.2 | 222 | 1,114 | 32.1 (28.0 - 36.6) | 36.9 (34.7 - 39.1) |
| MDCR | 1,280 | 6,081 | 0.8 | 4.7 | 35 | 236 | 44.9 (31.3 - 62.4) | 49.9 (43.8 - 56.7) |
| MDCD | 2,102 | 10,397 | 1.1 | 6.4 | 33 | 227 | 30.0 (20.6 - 42.1) | 35.5 (31.0 - 40.4) |
| Open Claims | 237,944 | 728,813 | 175.0 | 675.5 | 2,817 | 14,731 | 16.1 (15.5 - 16.7) | 21.8 (21.5 - 22.2) |
| OEHR | 15,400 | 64,626 | 4.8 | 36.2 | 69 | 932 | 14.3 (11.2 - 18.1) | 25.7 (24.1 - 27.4) |
| SIDIAP | 2,510 | 1,740 | 3.4 | 3.6 | 55 | 62 | 16.0 (12.1 - 20.8) | 17.1 (13.1 - 21.9) |
| Meta-analysis | 281,795 | 863,080 | 198.7 | 764.5 | 3,338 | 17,235 | 16.8 (16.2 - 17.4) | 22.5 (22.2 - 22.9) |
| Meta-analysis (w/o USOC) | 43,851 | 134,267 | 23.8 | 89.0 | 521 | 2,504 | 21.9 (20.1 - 23.9) | 28.1 (27.0 - 29.2) |
| **PS-matching / on-treatment TAR without escalation** | | | | | | | | |
| CCAE | 14,510 | 31,898 | 9.0 | 19.3 | 159 | 393 | 17.6 (15.0 - 20.6) | 20.4 (18.4 - 22.5) |
| DAG | 2,771 | 603 | 2.0 | 0.5 | 28 | 17 | 14.2 (9.46 - 20.6) | 32.4 (18.9 - 51.8) |
| ODOD | 10,623 | 36,345 | 5.8 | 26.3 | 188 | 951 | 32.5 (28.0 - 37.5) | 36.1 (33.9 - 38.5) |
| MDCR | 1,155 | 5,643 | 0.6 | 4.0 | 29 | 193 | 44.8 (30.0 - 64.3) | 47.7 (41.2 - 54.9) |
| MDCD | 2,052 | 10,040 | 0.9 | 5.1 | 27 | 187 | 30.2 (19.9 - 44.0) | 36.7 (31.6 - 42.3) |
| Open Claims | 225,774 | 719,920 | 141.2 | 562.8 | 2,253 | 12,118 | 16.0 (15.3 - 16.6) | 21.5 (21.1 - 21.9) |
| OEHR | 14,291 | 58,744 | 3.9 | 28.0 | 56 | 678 | 14.4 (10.9 - 18.7) | 24.2 (22.5 - 26.1) |
| SIDIAP | 2,422 | 853 | 3.0 | 1.2 | 43 | 19 | 14.5 (10.5 - 19.5) | 15.5 (9.33 - 24.2) |
| Meta-analysis | 270,827 | 863,443 | 164.5 | 646.8 | 2,755 | 14,539 | 16.8 (16.1 - 17.4) | 22.5 (22.1 - 22.8) |
| Meta-analysis (w/o USOC) | 45,053 | 143,523 | 23.2 | 83.9 | 502 | 2,421 | 21.6 (19.8 - 23.6) | 28.8 (27.7 - 30.0) |
| **PS-stratification / on-treatment TAR without escalation** | | | | | | | | |
| CCAE | 16,973 | 32,516 | 10.4 | 19.5 | 196 | 398 | 18.9 (16.3 - 21.7) | 20.4 (18.4 - 22.5) |
| DAG | 2,815 | 1,043 | 2.0 | 1.0 | 28 | 28 | 14.1 (9.37 - 20.4) | 29.2 (19.4 - 42.2) |
| ODOD | 11,478 | 37,125 | 6.2 | 27.0 | 192 | 992 | 31.2 (27.0 - 36.0) | 36.7 (34.5 - 39.1) |
| MDCR | 1,280 | 6,081 | 0.7 | 4.3 | 30 | 218 | 43.4 (29.3 - 61.9) | 50.5 (44.0 - 57.7) |
| MDCD | 2,102 | 10,397 | 0.9 | 5.3 | 28 | 192 | 30.7 (20.4 - 44.4) | 36.2 (31.3 - 41.7) |
| Open Claims | 237,944 | 728,813 | 147.3 | 570.0 | 2,346 | 12,427 | 15.9 (15.3 - 16.6) | 21.8 (21.4 - 22.2) |
| OEHR | 15,400 | 64,626 | 4.2 | 30.8 | 59 | 786 | 14.1 (10.7 - 18.2) | 25.5 (23.7 - 27.3) |
| SIDIAP | 2,510 | 1,740 | 3.0 | 3.0 | 46 | 54 | 15.2 (11.2 - 20.3) | 18.1 (13.6 - 23.6) |
| Meta-analysis | 281,795 | 863,080 | 168.0 | 647.3 | 2,793 | 14,603 | 16.6 (16.0 - 17.2) | 22.6 (22.2 - 22.9) |
| Meta-analysis (w/o USOC) | 43,851 | 134,267 | 20.7 | 77.4 | 447 | 2,176 | 21.6 (19.6 - 23.7) | 28.1 (27.0 - 29.3) |

##### 4.3.14 Diabetic ketoacidosis

**Supplementary Table 351:** Exposures, outcomes and risk estimates for Diabetic ketoacidosis in SGLT2I (female) (T) vs SU (female) (C) new-user cohorts under four different propensity score (PS) adjustment / time-at-risk (TAR) designs across data sources. We report total patient exposure time in 1,000 patient-year (KYr) units, uncalibrated and calibrated hazard ratios (HRs) and their 95% confidence intervals (CIs). Greyed out entries did not pass study diagnostics and are included only for completeness.

| | Patients | | Exposure (KYr) | | Outcomes | | Incidence (per KYr) | |
|---|---|---|---|---|---|---|---|---|
| | T | C | T | C | T | C | T | C |
| **PS-matching / on-treatment TAR** | | | | | | | | |
| CCAE | 16,071 | 35,137 | 11.8 | 25.1 | 26 | 33 | 2.19 (1.43 - 3.22) | 1.31 (0.90 - 1.85) |
| DAG | 3,180 | 682 | 2.5 | 0.7 | 0 | 0 | | |
| ODOD | 11,833 | 40,906 | 7.4 | 34.3 | 22 | 63 | 2.98 (1.87 - 4.51) | 1.84 (1.41 - 2.35) |
| MDCR | 1,330 | 6,791 | 0.9 | 5.5 | <5 | 6 | < 5.69 (1.85 - 13.3) | 1.09 (0.40 - 2.37) |
| MDCD | 2,380 | 11,611 | 1.3 | 7.3 | 5 | 25 | 3.89 (1.26 - 9.07) | 3.44 (2.22 - 5.07) |
| Open Claims | 261,540 | 831,033 | 196.2 | 787.6 | 242 | 707 | 1.23 (1.08 - 1.40) | 0.90 (0.83 - 0.97) |
| OEHR | 15,483 | 64,194 | 4.9 | 36.8 | 11 | 58 | 2.26 (1.13 - 4.05) | 1.58 (1.20 - 2.04) |
| SIDIAP | 2,849 | 965 | 4.0 | 1.7 | 0 | 0 | | |
| Meta-analysis | 307,307 | 982,881 | 221.6 | 891.0 | 306 | 886 | 1.38 (1.23 - 1.54) | 0.99 (0.93 - 1.06) |
| Meta-analysis (w/o USOC) | 45,767 | 151,848 | 25.4 | 103.5 | 64 | 179 | 2.52 (1.94 - 3.22) | 1.73 (1.49 - 2.00) |
| **PS-stratification / on-treatment TAR** | | | | | | | | |
| CCAE | 18,865 | 35,766 | 13.7 | 25.4 | 31 | 33 | 2.27 (1.54 - 3.22) | 1.30 (0.90 - 1.83) |
| DAG | 3,231 | 1,228 | 2.5 | 1.2 | 0 | 0 | | |
| ODOD | 12,833 | 41,815 | 7.9 | 35.1 | 24 | 67 | 3.05 (1.96 - 4.54) | 1.91 (1.48 - 2.42) |
| MDCR | 1,486 | 7,279 | 0.9 | 5.9 | <5 | 7 | < 5.29 (1.72 - 12.3) | 1.18 (0.48 - 2.44) |
| MDCD | 2,427 | 11,887 | 1.3 | 7.5 | 5 | 25 | 3.83 (1.24 - 8.94) | 3.34 (2.16 - 4.94) |
| Open Claims | 276,157 | 841,119 | 204.7 | 797.0 | 254 | 720 | 1.24 (1.09 - 1.40) | 0.90 (0.84 - 0.97) |
| OEHR | 16,657 | 70,431 | 5.2 | 40.6 | 11 | 64 | 2.10 (1.05 - 3.76) | 1.58 (1.21 - 2.01) |
| SIDIAP | 2,950 | 2,013 | 4.0 | 4.3 | 0 | 0 | | |
| Meta-analysis | 324,512 | 989,131 | 231.4 | 898.0 | 320 | 884 | 1.38 (1.24 - 1.54) | 0.98 (0.92 - 1.05) |
| Meta-analysis (w/o USOC) | 48,355 | 148,012 | 26.8 | 101.1 | 66 | 164 | 2.47 (1.91 - 3.14) | 1.62 (1.38 - 1.89) |
| **PS-matching / on-treatment TAR without escalation** | | | | | | | | |
| CCAE | 16,071 | 35,137 | 10.2 | 21.6 | 20 | 25 | 1.96 (1.20 - 3.03) | 1.16 (0.75 - 1.71) |
| DAG | 3,180 | 682 | 2.3 | 0.6 | 0 | 0 | | |
| ODOD | 11,833 | 40,906 | 6.5 | 30.6 | 19 | 51 | 2.91 (1.75 - 4.54) | 1.67 (1.24 - 2.19) |
| MDCR | 1,330 | 6,791 | 0.8 | 5.0 | <5 | <5 | < 6.43 (2.09 - 15.0) | < 1.00 (0.32 - 2.33) |
| MDCD | 2,380 | 11,611 | 1.1 | 6.0 | <5 | 18 | < 4.72 (1.53 - 11.0) | 2.99 (1.77 - 4.72) |
| Open Claims | 261,540 | 831,033 | 164.7 | 663.3 | 190 | 525 | 1.15 (1.00 - 1.33) | 0.79 (0.73 - 0.86) |
| OEHR | 15,483 | 64,194 | 4.2 | 31.2 | 7 | 36 | 1.66 (0.67 - 3.41) | 1.15 (0.81 - 1.60) |
| SIDIAP | 2,849 | 965 | 3.5 | 1.4 | 0 | 0 | | |
| Meta-analysis | 304,927 | 971,270 | 185.7 | 746.7 | 236 | 637 | 1.27 (1.11 - 1.44) | 0.85 (0.79 - 0.92) |
| Meta-analysis (w/o USOC) | 43,387 | 140,237 | 21.0 | 83.4 | 46 | 112 | 2.19 (1.61 - 2.93) | 1.34 (1.11 - 1.62) |
| **PS-stratification / on-treatment TAR without escalation** | | | | | | | | |
| CCAE | 18,865 | 35,766 | 11.8 | 21.8 | 24 | 25 | 2.04 (1.31 - 3.03) | 1.15 (0.74 - 1.69) |
| DAG | 3,231 | 1,228 | 2.3 | 1.2 | 0 | 0 | | |
| ODOD | 12,833 | 41,815 | 7.0 | 31.4 | 20 | 54 | 2.87 (1.75 - 4.43) | 1.72 (1.29 - 2.24) |
| MDCR | 1,486 | 7,279 | 0.8 | 5.4 | <5 | 5 | < 5.97 (1.94 - 13.9) | 0.93 (0.30 - 2.17) |
| MDCD | 2,427 | 11,887 | 1.1 | 6.2 | <5 | 18 | < 4.64 (1.51 - 10.8) | 2.91 (1.72 - 4.59) |
| Open Claims | 276,157 | 841,119 | 172.1 | 671.5 | 200 | 536 | 1.16 (1.01 - 1.33) | 0.80 (0.73 - 0.87) |
| OEHR | 16,657 | 70,431 | 4.6 | 34.4 | 7 | 41 | 1.54 (0.62 - 3.17) | 1.19 (0.86 - 1.62) |
| SIDIAP | 2,950 | 2,013 | 3.5 | 3.6 | 0 | 0 | | |
| Meta-analysis | 324,512 | 989,131 | 195.4 | 759.1 | 251 | 656 | 1.28 (1.13 - 1.45) | 0.86 (0.80 - 0.93) |
| Meta-analysis (w/o USOC) | 48,355 | 148,012 | 23.3 | 87.5 | 51 | 120 | 2.19 (1.63 - 2.88) | 1.37 (1.14 - 1.64) |

##### 4.3.15 Diarrhea

**Supplementary Table 352:** Exposures, outcomes and risk estimates for Diarrhea in SGLT2I (female) (T) vs SU (female) (C) new-user cohorts under four different propensity score (PS) adjustment / time-at-risk (TAR) designs across data sources. We report total patient exposure time in 1,000 patient-year (KYr) units, uncalibrated and calibrated hazard ratios (HRs) and their 95% confidence intervals (CIs). Greyed out entries did not pass study diagnostics and are included only for completeness.

| | Patients | | Exposure (KYr) | | Outcomes | | Incidence (per KYr) | |
|---|---|---|---|---|---|---|---|---|
| | T | C | T | C | T | C | T | C |
| **PS-matching / on-treatment TAR** | | | | | | | | |
| CCAE | 13,530 | 29,669 | 9.9 | 20.6 | 312 | 781 | 31.5 (28.1 - 35.2) | 37.9 (35.2 - 40.6) |
| DAG | 3,105 | 653 | 2.4 | 0.6 | 7 | <5 | 2.87 (1.15 - 5.92) | < 7.95 (2.58 - 18.5) |
| ODOD | 9,768 | 32,870 | 6.0 | 26.4 | 244 | 1,368 | 40.7 (35.7 - 46.1) | 51.9 (49.2 - 54.7) |
| MDCR | 1,100 | 5,279 | 0.7 | 4.1 | 25 | 202 | 33.8 (21.9 - 49.9) | 49.0 (42.5 - 56.3) |
| MDCD | 1,761 | 8,860 | 0.9 | 5.2 | 57 | 412 | 61.3 (46.5 - 79.5) | 78.5 (71.1 - 86.5) |
| Open Claims | 218,164 | 701,722 | 162.6 | 650.6 | 3,008 | 15,651 | 18.5 (17.8 - 19.2) | 24.1 (23.7 - 24.4) |
| OEHR | 13,181 | 54,532 | 4.0 | 29.4 | 162 | 1,524 | 40.2 (34.2 - 46.9) | 51.8 (49.2 - 54.4) |
| SIDIAP | 2,679 | 892 | 3.7 | 1.6 | 56 | 20 | 15.1 (11.4 - 19.7) | 12.6 (7.72 - 19.5) |
| Meta-analysis | 260,183 | 833,824 | 187.9 | 738.0 | 3,864 | 19,958 | 20.6 (19.9 - 21.2) | 27.0 (26.7 - 27.4) |
| Meta-analysis (w/o USOC) | 42,019 | 132,102 | 25.3 | 87.4 | 856 | 4,307 | 33.9 (31.6 - 36.2) | 49.3 (47.8 - 50.8) |
| **PS-stratification / on-treatment TAR** | | | | | | | | |
| CCAE | 15,864 | 30,271 | 11.4 | 20.9 | 371 | 798 | 32.6 (29.4 - 36.1) | 38.2 (35.6 - 41.0) |
| DAG | 3,158 | 1,189 | 2.5 | 1.2 | 7 | 5 | 2.84 (1.14 - 5.86) | 4.16 (1.35 - 9.72) |
| ODOD | 10,574 | 33,671 | 6.4 | 27.1 | 262 | 1,413 | 41.1 (36.3 - 46.4) | 52.1 (49.4 - 54.9) |
| MDCR | 1,228 | 5,815 | 0.8 | 4.5 | 29 | 223 | 37.1 (24.8 - 53.2) | 49.3 (43.0 - 56.2) |
| MDCD | 1,793 | 9,021 | 0.9 | 5.4 | 57 | 420 | 60.4 (45.7 - 78.2) | 78.4 (71.1 - 86.2) |
| Open Claims | 230,071 | 711,007 | 169.4 | 659.1 | 3,165 | 16,015 | 18.7 (18.0 - 19.3) | 24.3 (23.9 - 24.7) |
| OEHR | 14,218 | 59,849 | 4.4 | 32.3 | 177 | 1,724 | 40.7 (34.9 - 47.1) | 53.3 (50.8 - 55.9) |
| SIDIAP | 2,774 | 1,870 | 3.8 | 4.0 | 58 | 50 | 15.5 (11.7 - 20.0) | 12.4 (9.22 - 16.4) |
| Meta-analysis | 270,727 | 834,798 | 191.5 | 739.4 | 3,975 | 19,950 | 20.8 (20.1 - 21.4) | 27.0 (26.6 - 27.4) |
| Meta-analysis (w/o USOC) | 40,656 | 123,791 | 22.1 | 80.3 | 810 | 3,935 | 36.7 (34.2 - 39.3) | 49.0 (47.5 - 50.5) |
| **PS-matching / on-treatment TAR without escalation** | | | | | | | | |
| CCAE | 13,530 | 29,669 | 8.6 | 17.9 | 273 | 680 | 31.9 (28.2 - 35.9) | 38.0 (35.2 - 40.9) |
| DAG | 3,105 | 653 | 2.2 | 0.6 | 7 | <5 | 3.15 (1.27 - 6.50) | < 8.56 (2.78 - 20.0) |
| ODOD | 9,768 | 32,870 | 5.3 | 23.7 | 219 | 1,247 | 41.0 (35.7 - 46.8) | 52.5 (49.7 - 55.5) |
| MDCR | 1,100 | 5,279 | 0.7 | 3.8 | 22 | 182 | 33.8 (21.2 - 51.1) | 48.3 (41.5 - 55.8) |
| MDCD | 1,761 | 8,860 | 0.8 | 4.4 | 47 | 345 | 60.9 (44.7 - 81.0) | 78.2 (70.2 - 86.9) |
| Open Claims | 218,164 | 701,722 | 137.5 | 552.1 | 2,460 | 13,269 | 17.9 (17.2 - 18.6) | 24.0 (23.6 - 24.4) |
| OEHR | 13,181 | 54,532 | 3.5 | 25.4 | 135 | 1,207 | 38.1 (32.0 - 45.1) | 47.5 (44.9 - 50.3) |
| SIDIAP | 2,679 | 892 | 3.3 | 1.3 | 49 | 18 | 15.1 (11.1 - 19.9) | 13.5 (8.01 - 21.4) |
| Meta-analysis | 260,183 | 833,824 | 159.6 | 628.7 | 3,205 | 16,948 | 20.1 (19.4 - 20.8) | 27.0 (26.6 - 27.4) |
| Meta-analysis (w/o USOC) | 42,019 | 132,102 | 22.1 | 76.6 | 745 | 3,679 | 33.7 (31.3 - 36.2) | 48.1 (46.5 - 49.6) |
| **PS-stratification / on-treatment TAR without escalation** | | | | | | | | |
| CCAE | 15,864 | 30,271 | 9.9 | 18.1 | 322 | 696 | 32.5 (29.1 - 36.3) | 38.4 (35.6 - 41.3) |
| DAG | 3,158 | 1,189 | 2.2 | 1.1 | 7 | <5 | 3.12 (1.26 - 6.44) | < 4.49 (1.46 - 10.5) |
| ODOD | 10,574 | 33,671 | 5.7 | 24.4 | 234 | 1,292 | 41.2 (36.1 - 46.8) | 52.9 (50.1 - 55.9) |
| MDCR | 1,228 | 5,815 | 0.7 | 4.1 | 26 | 200 | 37.6 (24.6 - 55.1) | 48.2 (41.8 - 55.4) |
| MDCD | 1,793 | 9,021 | 0.8 | 4.5 | 47 | 353 | 59.8 (43.9 - 79.5) | 78.4 (70.4 - 87.0) |
| Open Claims | 230,071 | 711,007 | 143.4 | 559.6 | 2,586 | 13,584 | 18.0 (17.3 - 18.7) | 24.3 (23.9 - 24.7) |
| OEHR | 14,218 | 59,849 | 3.8 | 27.9 | 147 | 1,372 | 38.4 (32.5 - 45.2) | 49.1 (46.6 - 51.8) |
| SIDIAP | 2,774 | 1,870 | 3.3 | 3.3 | 50 | 37 | 15.2 (11.2 - 20.0) | 11.1 (7.83 - 15.3) |
| Meta-analysis | 270,727 | 834,798 | 162.8 | 630.1 | 3,289 | 16,944 | 20.2 (19.5 - 20.9) | 26.9 (26.5 - 27.3) |
| Meta-analysis (w/o USOC) | 40,656 | 123,791 | 19.4 | 70.5 | 703 | 3,360 | 36.2 (33.6 - 39.0) | 47.7 (46.1 - 49.3) |

##### 4.3.16 Genitourinary infection

**Supplementary Table 353:** Exposures, outcomes and risk estimates for Genitourinary infection in SGLT2I (female) (T) vs SU (female) (C) new-user cohorts under four different propensity score (PS) adjustment / time-at-risk (TAR) designs across data sources. We report total patient exposure time in 1,000 patient-year (KYr) units, uncalibrated and calibrated hazard ratios (HRs) and their 95% confidence intervals (CIs). Greyed out entries did not pass study diagnostics and are included only for completeness.

| | Patients | | Exposure (KYr) | | Outcomes | | Incidence (per KYr) | |
|---|---|---|---|---|---|---|---|---|
| | T | C | T | C | T | C | T | C |
| **PS-matching / on-treatment TAR** | | | | | | | | |
| CCAE | 11,136 | 24,608 | 7.9 | 16.9 | 472 | 1,046 | 60.0 (54.7 - 65.7) | 62.0 (58.3 - 65.9) |
| DAG | 2,162 | 481 | 1.7 | 0.4 | 87 | 19 | 52.5 (42.1 - 64.8) | 45.4 (27.3 - 70.9) |
| ODOD | 8,195 | 27,060 | 4.9 | 20.9 | 378 | 2,001 | 76.7 (69.2 - 84.9) | 95.6 (91.4 - 99.9) |
| MDCR | 996 | 4,606 | 0.6 | 3.5 | 51 | 301 | 79.5 (59.2 - 105) | 85.7 (76.3 - 95.9) |
| MDCD | 1,575 | 7,460 | 0.8 | 4.5 | 78 | 447 | 95.4 (75.4 - 119) | 100 (91.0 - 110) |
| Open Claims | 174,087 | 563,118 | 131.3 | 518.4 | 5,119 | 25,863 | 39.0 (37.9 - 40.1) | 49.9 (49.3 - 50.5) |
| OEHR | 11,752 | 48,208 | 3.5 | 25.6 | 237 | 1,906 | 67.2 (58.9 - 76.3) | 74.3 (71.0 - 77.8) |
| SIDIAP | 1,816 | 677 | 2.6 | 1.1 | 148 | 61 | 56.8 (48.0 - 66.7) | 53.9 (41.2 - 69.2) |
| Meta-analysis | 209,557 | 675,737 | 151.6 | 590.9 | 6,483 | 31,625 | 42.8 (41.7 - 43.8) | 53.5 (52.9 - 54.1) |
| Meta-analysis (w/o USOC) | 35,470 | 112,619 | 20.4 | 72.6 | 1,364 | 5,762 | 66.9 (63.4 - 70.6) | 79.4 (77.4 - 81.5) |
| **PS-stratification / on-treatment TAR** | | | | | | | | |
| CCAE | 12,966 | 24,966 | 9.0 | 17.0 | 553 | 1,057 | 61.4 (56.4 - 66.7) | 62.2 (58.5 - 66.0) |
| DAG | 2,193 | 813 | 1.7 | 0.8 | 89 | 35 | 53.2 (42.8 - 65.5) | 45.7 (31.9 - 63.6) |
| ODOD | 8,866 | 27,519 | 5.2 | 21.3 | 402 | 2,071 | 76.9 (69.6 - 84.8) | 97.0 (92.9 - 101) |
| MDCR | 1,115 | 4,982 | 0.7 | 3.8 | 54 | 341 | 77.8 (58.4 - 102) | 90.3 (80.9 - 100) |
| MDCD | 1,602 | 7,632 | 0.8 | 4.6 | 79 | 458 | 95.5 (75.6 - 119) | 100 (91.2 - 110) |
| Open Claims | 183,262 | 571,242 | 136.5 | 525.7 | 5,341 | 26,626 | 39.1 (38.1 - 40.2) | 50.6 (50.0 - 51.3) |
| OEHR | 12,672 | 53,535 | 3.8 | 28.5 | 254 | 2,210 | 66.5 (58.5 - 75.2) | 77.5 (74.3 - 80.8) |
| SIDIAP | 1,915 | 1,459 | 2.7 | 2.9 | 154 | 169 | 57.8 (49.0 - 67.7) | 57.9 (49.5 - 67.3) |
| Meta-analysis | 217,766 | 677,262 | 154.6 | 592.5 | 6,550 | 31,964 | 42.4 (41.4 - 43.4) | 53.9 (53.4 - 54.5) |
| Meta-analysis (w/o USOC) | 34,504 | 106,020 | 18.1 | 66.9 | 1,209 | 5,338 | 67.0 (63.2 - 70.8) | 79.8 (77.7 - 82.0) |
| **PS-matching / on-treatment TAR without escalation** | | | | | | | | |
| CCAE | 11,136 | 24,608 | 6.8 | 14.7 | 412 | 886 | 60.2 (54.6 - 66.3) | 60.4 (56.5 - 64.5) |
| DAG | 2,162 | 481 | 1.5 | 0.4 | 79 | 16 | 52.4 (41.5 - 65.3) | 41.6 (23.8 - 67.6) |
| ODOD | 8,195 | 27,060 | 4.4 | 18.9 | 333 | 1,800 | 75.6 (67.7 - 84.1) | 95.1 (90.8 - 99.6) |
| MDCR | 996 | 4,606 | 0.6 | 3.2 | 46 | 271 | 81.2 (59.4 - 108) | 83.5 (73.9 - 94.1) |
| MDCD | 1,575 | 7,460 | 0.7 | 3.8 | 58 | 373 | 84.3 (64.0 - 109) | 98.7 (88.9 - 109) |
| Open Claims | 174,087 | 563,118 | 110.7 | 440.7 | 4,339 | 22,098 | 39.2 (38.0 - 40.4) | 50.1 (49.5 - 50.8) |
| OEHR | 11,752 | 48,208 | 3.1 | 22.1 | 203 | 1,556 | 66.0 (57.2 - 75.7) | 70.3 (66.9 - 73.9) |
| SIDIAP | 1,816 | 677 | 2.3 | 1.0 | 135 | 55 | 59.0 (49.4 - 69.8) | 56.8 (42.8 - 73.9) |
| Meta-analysis | 209,557 | 675,737 | 128.6 | 504.4 | 5,526 | 27,039 | 43.0 (41.9 - 44.1) | 53.6 (53.0 - 54.2) |
| Meta-analysis (w/o USOC) | 35,470 | 112,619 | 17.9 | 63.7 | 1,187 | 4,941 | 66.4 (62.7 - 70.3) | 77.6 (75.4 - 79.7) |
| **PS-stratification / on-treatment TAR without escalation** | | | | | | | | |
| CCAE | 12,966 | 24,966 | 7.8 | 14.8 | 485 | 896 | 61.8 (56.4 - 67.5) | 60.6 (56.7 - 64.7) |
| DAG | 2,193 | 813 | 1.5 | 0.7 | 81 | 31 | 53.2 (42.3 - 66.1) | 43.8 (29.8 - 62.2) |
| ODOD | 8,866 | 27,519 | 4.7 | 19.3 | 355 | 1,869 | 75.9 (68.2 - 84.2) | 96.8 (92.5 - 101) |
| MDCR | 1,115 | 4,982 | 0.6 | 3.5 | 49 | 309 | 79.4 (58.8 - 105) | 88.7 (79.1 - 99.2) |
| MDCD | 1,602 | 7,632 | 0.7 | 3.9 | 59 | 382 | 84.7 (64.5 - 109) | 98.5 (88.9 - 109) |
| Open Claims | 183,262 | 571,242 | 115.3 | 447.2 | 4,529 | 22,753 | 39.3 (38.1 - 40.4) | 50.9 (50.2 - 51.5) |
| OEHR | 12,672 | 53,535 | 3.3 | 24.6 | 218 | 1,810 | 65.4 (57.0 - 74.6) | 73.5 (70.2 - 77.0) |
| SIDIAP | 1,915 | 1,459 | 2.3 | 2.4 | 141 | 142 | 60.1 (50.6 - 70.9) | 58.6 (49.3 - 69.0) |
| Meta-analysis | 217,766 | 677,262 | 131.2 | 505.9 | 5,587 | 27,328 | 42.6 (41.5 - 43.7) | 54.0 (53.4 - 54.7) |
| Meta-analysis (w/o USOC) | 34,504 | 106,020 | 15.9 | 58.7 | 1,058 | 4,575 | 66.7 (62.7 - 70.9) | 77.9 (75.7 - 80.2) |

##### 4.3.17 Hyperkalemia

**Supplementary Table 354:** Exposures, outcomes and risk estimates for Hyperkalemia in SGLT2I (female) (T) vs SU (female) (C) new-user cohorts under four different propensity score (PS) adjustment / time-at-risk (TAR) designs across data sources. We report total patient exposure time in 1,000 patient-year (KYr) units, uncalibrated and calibrated hazard ratios (HRs) and their 95% confidence intervals (CIs). Greyed out entries did not pass study diagnostics and are included only for completeness.

| | Patients | | Exposure (KYr) | | Outcomes | | Incidence (per KYr) | |
|---|---|---|---|---|---|---|---|---|
| | T | C | T | C | T | C | T | C |
| **PS-matching / on-treatment TAR** | | | | | | | | |
| CCAE | 15,821 | 34,604 | 11.6 | 24.6 | 64 | 200 | 5.52 (4.25 - 7.05) | 8.13 (7.04 - 9.34) |
| DAG | 2,974 | 640 | 2.3 | 0.6 | 25 | 9 | 10.7 (6.90 - 15.7) | 14.8 (6.77 - 28.1) |
| ODOD | 11,422 | 38,860 | 7.1 | 31.8 | 105 | 758 | 14.8 (12.1 - 17.9) | 23.8 (22.2 - 25.6) |
| MDCR | 1,299 | 6,566 | 0.9 | 5.3 | 5 | 89 | 5.78 (1.88 - 13.5) | 16.9 (13.6 - 20.8) |
| MDCD | 2,321 | 11,258 | 1.3 | 6.9 | 7 | 121 | 5.59 (2.25 - 11.5) | 17.4 (14.5 - 20.8) |
| Open Claims | 257,321 | 815,085 | 193.0 | 765.4 | 685 | 5,838 | 3.55 (3.29 - 3.83) | 7.63 (7.43 - 7.83) |
| OEHR | 15,234 | 63,024 | 4.8 | 35.7 | 31 | 465 | 6.50 (4.41 - 9.22) | 13.0 (11.9 - 14.3) |
| SIDIAP | 2,756 | 929 | 3.8 | 1.6 | 37 | 16 | 9.65 (6.80 - 13.3) | 9.97 (5.70 - 16.2) |
| Meta-analysis | 306,174 | 970,326 | 222.4 | 871.3 | 934 | 7,487 | 4.20 (3.93 - 4.48) | 8.59 (8.40 - 8.79) |
| Meta-analysis (w/o USOC) | 48,853 | 155,241 | 29.4 | 105.9 | 249 | 1,649 | 8.46 (7.44 - 9.58) | 15.6 (14.8 - 16.3) |
| **PS-stratification / on-treatment TAR** | | | | | | | | |
| CCAE | 18,560 | 35,219 | 13.4 | 24.8 | 76 | 203 | 5.69 (4.48 - 7.12) | 8.18 (7.09 - 9.38) |
| DAG | 3,022 | 1,171 | 2.4 | 1.2 | 26 | 12 | 11.0 (7.18 - 16.1) | 10.1 (5.22 - 17.7) |
| ODOD | 12,381 | 39,679 | 7.6 | 32.6 | 109 | 790 | 14.4 (11.8 - 17.4) | 24.2 (22.6 - 26.0) |
| MDCR | 1,451 | 7,008 | 0.9 | 5.6 | 5 | 98 | 5.37 (1.75 - 12.5) | 17.4 (14.1 - 21.2) |
| MDCD | 2,363 | 11,556 | 1.3 | 7.2 | 7 | 128 | 5.52 (2.22 - 11.4) | 17.9 (14.9 - 21.3) |
| Open Claims | 271,518 | 824,713 | 201.2 | 774.2 | 717 | 6,075 | 3.56 (3.31 - 3.83) | 7.85 (7.65 - 8.05) |
| OEHR | 16,395 | 69,036 | 5.1 | 39.2 | 32 | 550 | 6.23 (4.26 - 8.80) | 14.0 (12.9 - 15.3) |
| SIDIAP | 2,851 | 1,941 | 3.9 | 4.1 | 40 | 39 | 10.3 (7.35 - 14.0) | 9.42 (6.70 - 12.9) |
| Meta-analysis | 318,854 | 968,647 | 227.2 | 870.9 | 934 | 7,618 | 4.11 (3.85 - 4.38) | 8.75 (8.55 - 8.95) |
| Meta-analysis (w/o USOC) | 47,336 | 143,934 | 26.1 | 96.6 | 217 | 1,543 | 8.32 (7.25 - 9.51) | 16.0 (15.2 - 16.8) |
| **PS-matching / on-treatment TAR without escalation** | | | | | | | | |
| CCAE | 15,821 | 34,604 | 10.0 | 21.2 | 55 | 176 | 5.50 (4.15 - 7.16) | 8.32 (7.14 - 9.65) |
| DAG | 2,974 | 640 | 2.1 | 0.6 | 22 | 8 | 10.3 (6.44 - 15.6) | 14.3 (6.16 - 28.1) |
| ODOD | 11,422 | 38,860 | 6.3 | 28.5 | 93 | 655 | 14.8 (11.9 - 18.1) | 23.0 (21.3 - 24.8) |
| MDCR | 1,299 | 6,566 | 0.8 | 4.8 | 5 | 81 | 6.54 (2.12 - 15.3) | 16.9 (13.4 - 21.0) |
| MDCD | 2,321 | 11,258 | 1.0 | 5.8 | 7 | 106 | 6.77 (2.72 - 14.0) | 18.4 (15.1 - 22.3) |
| Open Claims | 257,321 | 815,085 | 162.1 | 645.3 | 551 | 4,869 | 3.40 (3.12 - 3.70) | 7.55 (7.33 - 7.76) |
| OEHR | 15,234 | 63,024 | 4.1 | 30.3 | 26 | 352 | 6.28 (4.10 - 9.20) | 11.6 (10.4 - 12.9) |
| SIDIAP | 2,756 | 929 | 3.4 | 1.3 | 32 | 14 | 9.48 (6.49 - 13.4) | 10.4 (5.69 - 17.4) |
| Meta-analysis | 306,174 | 970,326 | 187.7 | 737.2 | 769 | 6,253 | 4.10 (3.81 - 4.40) | 8.48 (8.27 - 8.70) |
| Meta-analysis (w/o USOC) | 48,853 | 155,241 | 25.6 | 91.9 | 218 | 1,384 | 8.52 (7.42 - 9.72) | 15.1 (14.3 - 15.9) |
| **PS-stratification / on-treatment TAR without escalation** | | | | | | | | |
| CCAE | 18,560 | 35,219 | 11.5 | 21.4 | 67 | 179 | 5.81 (4.50 - 7.38) | 8.38 (7.20 - 9.70) |
| DAG | 3,022 | 1,171 | 2.2 | 1.1 | 23 | 10 | 10.6 (6.75 - 16.0) | 9.05 (4.34 - 16.7) |
| ODOD | 12,381 | 39,679 | 6.7 | 29.2 | 97 | 685 | 14.5 (11.7 - 17.6) | 23.5 (21.7 - 25.3) |
| MDCR | 1,451 | 7,008 | 0.8 | 5.1 | 5 | 90 | 6.07 (1.97 - 14.2) | 17.6 (14.1 - 21.6) |
| MDCD | 2,363 | 11,556 | 1.0 | 5.9 | 7 | 112 | 6.67 (2.68 - 13.7) | 18.9 (15.5 - 22.7) |
| Open Claims | 271,518 | 824,713 | 169.2 | 652.9 | 580 | 5,079 | 3.43 (3.15 - 3.72) | 7.78 (7.57 - 8.00) |
| OEHR | 16,395 | 69,036 | 4.5 | 33.3 | 26 | 410 | 5.82 (3.81 - 8.53) | 12.3 (11.1 - 13.5) |
| SIDIAP | 2,851 | 1,941 | 3.4 | 3.4 | 35 | 33 | 10.2 (7.12 - 14.2) | 9.68 (6.66 - 13.6) |
| Meta-analysis | 318,854 | 968,647 | 191.9 | 736.8 | 770 | 6,353 | 4.01 (3.73 - 4.31) | 8.62 (8.41 - 8.84) |
| Meta-analysis (w/o USOC) | 47,336 | 143,934 | 22.7 | 83.9 | 190 | 1,274 | 8.37 (7.22 - 9.64) | 15.2 (14.4 - 16.0) |

##### 4.3.18 Hypoglycemia

**Supplementary Table 355:** Exposures, outcomes and risk estimates for Hypoglycemia in SGLT2I (female) (T) vs SU (female) (C) new-user cohorts under four different propensity score (PS) adjustment / time-at-risk (TAR) designs across data sources. We report total patient exposure time in 1,000 patient-year (KYr) units, uncalibrated and calibrated hazard ratios (HRs) and their 95% confidence intervals (CIs). Greyed out entries did not pass study diagnostics and are included only for completeness.

| | Patients | | Exposure (KYr) | | Outcomes | | Incidence (per KYr) | |
|---|---|---|---|---|---|---|---|---|
| | T | C | T | C | T | C | T | C |
| **PS-matching / on-treatment TAR** | | | | | | | | |
| CCAE | 15,761 | 34,654 | 11.6 | 24.6 | 42 | 323 | 3.62 (2.61 - 4.89) | 13.1 (11.7 - 14.6) |
| DAG | 3,163 | 678 | 2.5 | 0.7 | <5 | 5 | < 2.00 (0.65 - 4.67) | 7.67 (2.49 - 17.9) |
| ODOD | 11,617 | 40,283 | 7.2 | 33.4 | 47 | 920 | 6.49 (4.77 - 8.63) | 27.6 (25.8 - 29.4) |
| MDCR | 1,320 | 6,722 | 0.9 | 5.4 | 0 | 143 | | 26.5 (22.4 - 31.3) |
| MDCD | 2,312 | 11,360 | 1.2 | 7.0 | 12 | 243 | 9.65 (4.99 - 16.9) | 34.7 (30.5 - 39.3) |
| Open Claims | 256,842 | 818,296 | 192.7 | 771.9 | 398 | 8,212 | 2.07 (1.87 - 2.28) | 10.6 (10.4 - 10.9) |
| OEHR | 15,300 | 63,521 | 4.8 | 36.0 | 17 | 703 | 3.55 (2.07 - 5.68) | 19.5 (18.1 - 21.0) |
| SIDIAP | 2,843 | 962 | 4.0 | 1.7 | <5 | 7 | < 1.26 (0.41 - 2.95) | 4.19 (1.68 - 8.63) |
| Meta-analysis | 301,832 | 968,114 | 217.6 | 872.9 | 516 | 10,401 | 2.37 (2.17 - 2.59) | 11.9 (11.7 - 12.1) |
| Meta-analysis (w/o USOC) | 44,990 | 149,818 | 24.9 | 101.0 | 118 | 2,189 | 4.74 (3.92 - 5.68) | 21.7 (20.8 - 22.6) |
| **PS-stratification / on-treatment TAR** | | | | | | | | |
| CCAE | 18,425 | 35,271 | 13.3 | 24.9 | 50 | 326 | 3.76 (2.79 - 4.96) | 13.1 (11.7 - 14.6) |
| DAG | 3,214 | 1,220 | 2.5 | 1.2 | <5 | 11 | < 1.98 (0.64 - 4.63) | 8.89 (4.44 - 15.9) |
| ODOD | 12,580 | 41,160 | 7.7 | 34.2 | 50 | 959 | 6.50 (4.83 - 8.57) | 28.0 (26.3 - 29.9) |
| MDCR | 1,473 | 7,208 | 0.9 | 5.8 | 0 | 162 | | 27.9 (23.8 - 32.6) |
| MDCD | 2,349 | 11,604 | 1.3 | 7.2 | 12 | 248 | 9.54 (4.93 - 16.7) | 34.5 (30.4 - 39.1) |
| Open Claims | 270,936 | 828,559 | 200.8 | 781.5 | 416 | 8,484 | 2.07 (1.88 - 2.28) | 10.9 (10.6 - 11.1) |
| OEHR | 16,444 | 69,682 | 5.2 | 39.7 | 18 | 803 | 3.49 (2.07 - 5.52) | 20.3 (18.9 - 21.7) |
| SIDIAP | 2,944 | 2,010 | 4.0 | 4.3 | <5 | 20 | < 1.25 (0.40 - 2.91) | 4.63 (2.83 - 7.14) |
| Meta-analysis | 318,385 | 974,672 | 226.9 | 880.3 | 534 | 10,572 | 2.35 (2.16 - 2.56) | 12.0 (11.8 - 12.2) |
| Meta-analysis (w/o USOC) | 47,449 | 146,113 | 26.1 | 98.7 | 118 | 2,088 | 4.51 (3.74 - 5.40) | 21.1 (20.2 - 22.1) |
| **PS-matching / on-treatment TAR without escalation** | | | | | | | | |
| CCAE | 15,761 | 34,654 | 10.0 | 21.2 | 30 | 264 | 3.00 (2.03 - 4.29) | 12.4 (11.0 - 14.0) |
| DAG | 3,163 | 678 | 2.3 | 0.6 | <5 | 5 | < 2.20 (0.71 - 5.12) | 8.28 (2.69 - 19.3) |
| ODOD | 11,617 | 40,283 | 6.4 | 29.8 | 34 | 807 | 5.30 (3.67 - 7.40) | 27.1 (25.2 - 29.0) |
| MDCR | 1,320 | 6,722 | 0.8 | 4.9 | 0 | 128 | | 26.1 (21.8 - 31.0) |
| MDCD | 2,312 | 11,360 | 1.0 | 5.8 | 9 | 197 | 8.80 (4.02 - 16.7) | 33.8 (29.2 - 38.9) |
| Open Claims | 256,842 | 818,296 | 161.8 | 650.9 | 260 | 6,809 | 1.61 (1.42 - 1.82) | 10.5 (10.2 - 10.7) |
| OEHR | 15,300 | 63,521 | 4.2 | 30.6 | 8 | 522 | 1.92 (0.83 - 3.78) | 17.0 (15.6 - 18.6) |
| SIDIAP | 2,843 | 962 | 3.5 | 1.4 | <5 | 5 | < 1.44 (0.47 - 3.37) | 3.54 (1.15 - 8.27) |
| Meta-analysis | 301,832 | 968,114 | 183.4 | 738.4 | 341 | 8,599 | 1.86 (1.67 - 2.07) | 11.6 (11.4 - 11.9) |
| Meta-analysis (w/o USOC) | 44,990 | 149,818 | 21.6 | 87.5 | 81 | 1,790 | 3.75 (2.98 - 4.66) | 20.5 (19.5 - 21.4) |
| **PS-stratification / on-treatment TAR without escalation** | | | | | | | | |
| CCAE | 18,425 | 35,271 | 11.5 | 21.4 | 37 | 264 | 3.22 (2.27 - 4.44) | 12.3 (10.9 - 13.9) |
| DAG | 3,214 | 1,220 | 2.3 | 1.1 | <5 | 11 | < 2.18 (0.71 - 5.08) | 9.60 (4.79 - 17.2) |
| ODOD | 12,580 | 41,160 | 6.8 | 30.6 | 37 | 844 | 5.42 (3.82 - 7.47) | 27.6 (25.8 - 29.5) |
| MDCR | 1,473 | 7,208 | 0.8 | 5.3 | 0 | 145 | | 27.5 (23.2 - 32.3) |
| MDCD | 2,349 | 11,604 | 1.0 | 6.0 | 9 | 202 | 8.68 (3.97 - 16.5) | 33.8 (29.3 - 38.8) |
| Open Claims | 270,936 | 828,559 | 168.9 | 659.3 | 277 | 7,050 | 1.64 (1.45 - 1.85) | 10.7 (10.4 - 10.9) |
| OEHR | 16,444 | 69,682 | 4.5 | 33.7 | 9 | 590 | 2.01 (0.92 - 3.81) | 17.5 (16.1 - 19.0) |
| SIDIAP | 2,944 | 2,010 | 3.5 | 3.6 | <5 | 15 | < 1.42 (0.46 - 3.31) | 4.20 (2.35 - 6.93) |
| Meta-analysis | 318,385 | 974,672 | 191.7 | 745.0 | 360 | 8,748 | 1.88 (1.69 - 2.08) | 11.7 (11.5 - 12.0) |
| Meta-analysis (w/o USOC) | 47,449 | 146,113 | 22.8 | 85.7 | 83 | 1,698 | 3.64 (2.90 - 4.51) | 19.8 (18.9 - 20.8) |

##### 4.3.19 Hypotension

**Supplementary Table 356:** Exposures, outcomes and risk estimates for Hypotension in SGLT2I (female) (T) vs SU (female) (C) new-user cohorts under four different propensity score (PS) adjustment / time-at-risk (TAR) designs across data sources. We report total patient exposure time in 1,000 patient-year (KYr) units, uncalibrated and calibrated hazard ratios (HRs) and their 95% confidence intervals (CIs). Greyed out entries did not pass study diagnostics and are included only for completeness.

| | Patients | | Exposure (KYr) | | Outcomes | | Incidence (per KYr) | |
|---|---|---|---|---|---|---|---|---|
| | T | C | T | C | T | C | T | C |
| **PS-matching / on-treatment TAR** | | | | | | | | |
| CCAE | 15,744 | 34,322 | 11.6 | 24.5 | 90 | 187 | 7.77 (6.25 - 9.55) | 7.65 (6.59 - 8.82) |
| DAG | 3,055 | 659 | 2.4 | 0.6 | 7 | <5 | 2.91 (1.17 - 6.00) | < 8.00 (2.60 - 18.7) |
| ODOD | 11,340 | 38,641 | 7.0 | 31.9 | 141 | 724 | 20.1 (16.9 - 23.7) | 22.7 (21.1 - 24.4) |
| MDCR | 1,279 | 6,447 | 0.8 | 5.2 | 23 | 116 | 27.3 (17.3 - 40.9) | 22.4 (18.5 - 26.9) |
| MDCD | 2,228 | 10,689 | 1.2 | 6.6 | 42 | 177 | 35.9 (25.8 - 48.5) | 26.9 (23.0 - 31.1) |
| Open Claims | 253,429 | 803,417 | 190.2 | 756.9 | 1,208 | 6,608 | 6.35 (6.00 - 6.72) | 8.73 (8.52 - 8.94) |
| OEHR | 15,130 | 62,599 | 4.7 | 35.4 | 56 | 519 | 11.8 (8.93 - 15.3) | 14.7 (13.4 - 16.0) |
| SIDIAP | 2,828 | 957 | 3.9 | 1.7 | 8 | 5 | 2.04 (0.88 - 4.01) | 3.02 (0.98 - 7.04) |
| Meta-analysis | 299,150 | 956,115 | 215.5 | 860.5 | 1,560 | 8,331 | 7.24 (6.88 - 7.61) | 9.68 (9.48 - 9.89) |
| Meta-analysis (w/o USOC) | 45,721 | 152,698 | 25.4 | 103.5 | 352 | 1,723 | 13.9 (12.5 - 15.4) | 16.6 (15.9 - 17.4) |
| **PS-stratification / on-treatment TAR** | | | | | | | | |
| CCAE | 18,471 | 35,027 | 13.3 | 24.7 | 110 | 194 | 8.25 (6.78 - 9.94) | 7.85 (6.78 - 9.03) |
| DAG | 3,105 | 1,187 | 2.4 | 1.2 | 7 | <5 | 2.88 (1.16 - 5.94) | < 4.19 (1.36 - 9.78) |
| ODOD | 12,292 | 39,562 | 7.5 | 32.8 | 148 | 771 | 19.8 (16.7 - 23.2) | 23.5 (21.9 - 25.2) |
| MDCR | 1,425 | 6,870 | 0.9 | 5.5 | 25 | 131 | 27.6 (17.9 - 40.8) | 23.8 (19.9 - 28.3) |
| MDCD | 2,263 | 11,080 | 1.2 | 6.9 | 42 | 188 | 35.4 (25.5 - 47.9) | 27.3 (23.5 - 31.5) |
| Open Claims | 267,171 | 813,252 | 198.1 | 766.1 | 1,308 | 6,875 | 6.60 (6.25 - 6.97) | 8.97 (8.76 - 9.19) |
| OEHR | 16,280 | 68,740 | 5.1 | 39.0 | 60 | 601 | 11.8 (8.97 - 15.1) | 15.4 (14.2 - 16.7) |
| SIDIAP | 2,929 | 2,001 | 4.0 | 4.3 | 8 | 8 | 2.01 (0.87 - 3.95) | 1.85 (0.80 - 3.65) |
| Meta-analysis | 314,214 | 956,581 | 224.0 | 862.6 | 1,626 | 8,441 | 7.26 (6.91 - 7.62) | 9.79 (9.58 - 10.00) |
| Meta-analysis (w/o USOC) | 47,043 | 143,329 | 25.9 | 96.5 | 318 | 1,566 | 12.3 (11.0 - 13.7) | 16.2 (15.4 - 17.1) |
| **PS-matching / on-treatment TAR without escalation** | | | | | | | | |
| CCAE | 15,744 | 34,322 | 10.0 | 21.1 | 68 | 151 | 6.82 (5.30 - 8.65) | 7.17 (6.07 - 8.41) |
| DAG | 3,055 | 659 | 2.2 | 0.6 | 7 | <5 | 3.20 (1.29 - 6.60) | < 8.64 (2.80 - 20.2) |
| ODOD | 11,340 | 38,641 | 6.2 | 28.5 | 119 | 638 | 19.1 (15.8 - 22.9) | 22.4 (20.7 - 24.2) |
| MDCR | 1,279 | 6,447 | 0.7 | 4.7 | 23 | 107 | 30.8 (19.5 - 46.2) | 22.7 (18.6 - 27.4) |
| MDCD | 2,228 | 10,689 | 1.0 | 5.5 | 32 | 136 | 32.9 (22.5 - 46.5) | 24.9 (20.9 - 29.4) |
| Open Claims | 253,429 | 803,417 | 159.8 | 638.0 | 975 | 5,490 | 6.10 (5.72 - 6.50) | 8.60 (8.38 - 8.84) |
| OEHR | 15,130 | 62,599 | 4.1 | 30.1 | 39 | 370 | 9.46 (6.73 - 12.9) | 12.3 (11.1 - 13.6) |
| SIDIAP | 2,828 | 957 | 3.5 | 1.4 | 7 | <5 | 2.03 (0.82 - 4.18) | < 3.58 (1.16 - 8.34) |
| Meta-analysis | 299,150 | 956,115 | 181.8 | 727.9 | 1,256 | 6,892 | 6.91 (6.53 - 7.30) | 9.47 (9.25 - 9.69) |
| Meta-analysis (w/o USOC) | 45,721 | 152,698 | 22.0 | 89.9 | 281 | 1,402 | 12.8 (11.3 - 14.3) | 15.6 (14.8 - 16.4) |
| **PS-stratification / on-treatment TAR without escalation** | | | | | | | | |
| CCAE | 18,471 | 35,027 | 11.5 | 21.3 | 84 | 157 | 7.30 (5.82 - 9.04) | 7.37 (6.26 - 8.62) |
| DAG | 3,105 | 1,187 | 2.2 | 1.1 | 7 | <5 | 3.17 (1.27 - 6.53) | < 4.53 (1.47 - 10.6) |
| ODOD | 12,292 | 39,562 | 6.6 | 29.3 | 126 | 680 | 19.0 (15.8 - 22.6) | 23.2 (21.5 - 25.0) |
| MDCR | 1,425 | 6,870 | 0.8 | 5.0 | 24 | 120 | 29.8 (19.1 - 44.4) | 23.9 (19.9 - 28.6) |
| MDCD | 2,263 | 11,080 | 1.0 | 5.7 | 32 | 144 | 32.5 (22.2 - 45.8) | 25.2 (21.3 - 29.7) |
| Open Claims | 267,171 | 813,252 | 166.7 | 646.0 | 1,066 | 5,725 | 6.40 (6.02 - 6.79) | 8.86 (8.63 - 9.10) |
| OEHR | 16,280 | 68,740 | 4.4 | 33.2 | 42 | 427 | 9.45 (6.81 - 12.8) | 12.9 (11.7 - 14.1) |
| SIDIAP | 2,929 | 2,001 | 3.5 | 3.6 | 7 | 7 | 2.00 (0.80 - 4.12) | 1.97 (0.79 - 4.06) |
| Meta-analysis | 314,214 | 956,581 | 189.3 | 729.8 | 1,318 | 6,989 | 6.96 (6.59 - 7.35) | 9.58 (9.35 - 9.80) |
| Meta-analysis (w/o USOC) | 47,043 | 143,329 | 22.6 | 83.8 | 252 | 1,264 | 11.2 (9.82 - 12.6) | 15.1 (14.3 - 15.9) |

##### 4.3.20 Joint pain

**Supplementary Table 357:** Exposures, outcomes and risk estimates for Joint pain in SGLT2I (female) (T) vs SU (female) (C) new-user cohorts under four different propensity score (PS) adjustment / time-at-risk (TAR) designs across data sources. We report total patient exposure time in 1,000 patient-year (KYr) units, uncalibrated and calibrated hazard ratios (HRs) and their 95% confidence intervals (CIs). Greyed out entries did not pass study diagnostics and are included only for completeness.

| | Patients | | Exposure (KYr) | | Outcomes | | Incidence (per KYr) | |
|---|---|---|---|---|---|---|---|---|
| | T | C | T | C | T | C | T | C |
| **PS-matching / on-treatment TAR** | | | | | | | | |
| CCAE | 14,820 | 32,693 | 10.9 | 23.2 | 140 | 354 | 12.9 (10.8 - 15.2) | 15.3 (13.7 - 17.0) |
| DAG | 2,618 | 573 | 2.0 | 0.5 | 45 | 16 | 22.4 (16.3 - 30.0) | 30.3 (17.3 - 49.1) |
| ODOD | 10,889 | 37,908 | 6.8 | 31.5 | 119 | 552 | 17.6 (14.6 - 21.1) | 17.5 (16.1 - 19.1) |
| MDCR | 1,265 | 6,408 | 0.8 | 5.2 | 12 | 55 | 14.4 (7.46 - 25.2) | 10.7 (8.03 - 13.9) |
| MDCD | 2,114 | 10,446 | 1.1 | 6.4 | 31 | 155 | 27.2 (18.5 - 38.6) | 24.2 (20.6 - 28.3) |
| Open Claims | 240,761 | 775,933 | 180.9 | 732.9 | 1,373 | 6,030 | 7.59 (7.19 - 8.00) | 8.23 (8.02 - 8.44) |
| OEHR | 14,340 | 60,148 | 4.4 | 33.8 | 76 | 588 | 17.1 (13.5 - 21.5) | 17.4 (16.0 - 18.9) |
| SIDIAP | 2,038 | 728 | 2.8 | 1.2 | 57 | 40 | 20.1 (15.2 - 26.1) | 32.9 (23.5 - 44.8) |
| Meta-analysis | 286,227 | 924,264 | 207.8 | 834.1 | 1,808 | 7,774 | 8.70 (8.31 - 9.11) | 9.32 (9.11 - 9.53) |
| Meta-analysis (w/o USOC) | 45,466 | 148,331 | 26.9 | 101.2 | 435 | 1,744 | 16.2 (14.7 - 17.8) | 17.2 (16.4 - 18.1) |
| **PS-stratification / on-treatment TAR** | | | | | | | | |
| CCAE | 17,271 | 33,274 | 12.5 | 23.4 | 175 | 355 | 14.0 (12.0 - 16.2) | 15.2 (13.6 - 16.8) |
| DAG | 2,660 | 1,030 | 2.0 | 1.0 | 46 | 31 | 22.7 (16.6 - 30.2) | 31.0 (21.1 - 44.0) |
| ODOD | 11,750 | 38,689 | 7.2 | 32.2 | 129 | 565 | 18.0 (15.1 - 21.4) | 17.5 (16.1 - 19.0) |
| MDCR | 1,412 | 6,861 | 0.9 | 5.5 | 14 | 60 | 15.7 (8.57 - 26.3) | 10.8 (8.26 - 13.9) |
| MDCD | 2,157 | 10,726 | 1.2 | 6.6 | 31 | 160 | 26.8 (18.2 - 38.0) | 24.3 (20.7 - 28.4) |
| Open Claims | 253,928 | 785,790 | 188.5 | 742.0 | 1,465 | 6,137 | 7.77 (7.38 - 8.18) | 8.27 (8.07 - 8.48) |
| OEHR | 15,387 | 66,105 | 4.8 | 37.3 | 80 | 643 | 16.8 (13.3 - 20.9) | 17.2 (15.9 - 18.6) |
| SIDIAP | 2,108 | 1,550 | 2.9 | 3.2 | 59 | 96 | 20.5 (15.6 - 26.5) | 29.9 (24.2 - 36.5) |
| Meta-analysis | 298,336 | 923,858 | 212.9 | 835.0 | 1,849 | 7,700 | 8.69 (8.29 - 9.09) | 9.22 (9.02 - 9.43) |
| Meta-analysis (w/o USOC) | 44,408 | 138,068 | 24.4 | 93.0 | 384 | 1,563 | 15.7 (14.2 - 17.4) | 16.8 (16.0 - 17.7) |
| **PS-matching / on-treatment TAR without escalation** | | | | | | | | |
| CCAE | 14,820 | 32,693 | 9.4 | 20.0 | 118 | 313 | 12.6 (10.4 - 15.1) | 15.7 (14.0 - 17.5) |
| DAG | 2,618 | 573 | 1.8 | 0.5 | 43 | 15 | 23.6 (17.1 - 31.8) | 30.6 (17.1 - 50.4) |
| ODOD | 10,889 | 37,908 | 6.0 | 28.2 | 103 | 494 | 17.2 (14.0 - 20.9) | 17.5 (16.0 - 19.1) |
| MDCR | 1,265 | 6,408 | 0.7 | 4.7 | 11 | 48 | 14.8 (7.41 - 26.5) | 10.2 (7.54 - 13.6) |
| MDCD | 2,114 | 10,446 | 0.9 | 5.3 | 28 | 127 | 29.7 (19.8 - 43.0) | 23.9 (19.9 - 28.4) |
| Open Claims | 240,761 | 775,933 | 152.0 | 618.5 | 1,151 | 5,053 | 7.57 (7.14 - 8.02) | 8.17 (7.95 - 8.40) |
| OEHR | 14,340 | 60,148 | 3.9 | 28.8 | 69 | 473 | 17.8 (13.9 - 22.6) | 16.4 (15.0 - 18.0) |
| SIDIAP | 2,038 | 728 | 2.5 | 1.0 | 43 | 35 | 17.2 (12.4 - 23.1) | 33.4 (23.3 - 46.5) |
| Meta-analysis | 286,227 | 924,264 | 175.4 | 706.5 | 1,523 | 6,543 | 8.68 (8.25 - 9.13) | 9.26 (9.04 - 9.49) |
| Meta-analysis (w/o USOC) | 45,466 | 148,331 | 23.4 | 88.0 | 372 | 1,490 | 15.9 (14.3 - 17.6) | 16.9 (16.1 - 17.8) |
| **PS-stratification / on-treatment TAR without escalation** | | | | | | | | |
| CCAE | 17,271 | 33,274 | 10.8 | 20.2 | 150 | 314 | 13.9 (11.8 - 16.3) | 15.6 (13.9 - 17.4) |
| DAG | 2,660 | 1,030 | 1.8 | 0.9 | 44 | 29 | 24.0 (17.4 - 32.2) | 31.1 (20.8 - 44.6) |
| ODOD | 11,750 | 38,689 | 6.3 | 28.9 | 113 | 507 | 17.8 (14.7 - 21.4) | 17.6 (16.1 - 19.2) |
| MDCR | 1,412 | 6,861 | 0.8 | 5.0 | 13 | 53 | 16.3 (8.66 - 27.8) | 10.5 (7.88 - 13.8) |
| MDCD | 2,157 | 10,726 | 1.0 | 5.5 | 28 | 131 | 29.2 (19.4 - 42.3) | 23.9 (20.0 - 28.4) |
| Open Claims | 253,928 | 785,790 | 158.6 | 626.4 | 1,233 | 5,146 | 7.77 (7.35 - 8.22) | 8.22 (7.99 - 8.44) |
| OEHR | 15,387 | 66,105 | 4.2 | 31.8 | 72 | 518 | 17.3 (13.6 - 21.8) | 16.3 (14.9 - 17.8) |
| SIDIAP | 2,108 | 1,550 | 2.5 | 2.7 | 45 | 81 | 17.7 (12.9 - 23.7) | 30.2 (24.0 - 37.5) |
| Meta-analysis | 298,336 | 923,858 | 179.9 | 707.2 | 1,568 | 6,485 | 8.72 (8.29 - 9.16) | 9.17 (8.95 - 9.40) |
| Meta-analysis (w/o USOC) | 44,408 | 138,068 | 21.3 | 80.8 | 335 | 1,339 | 15.7 (14.1 - 17.5) | 16.6 (15.7 - 17.5) |

##### 4.3.21 Lower extremity amputation

**Supplementary Table 358:** Exposures, outcomes and risk estimates for Lower extremity amputation in SGLT2I (female) (T) vs SU (female) (C) new-user cohorts under four different propensity score (PS) adjustment / time-at-risk (TAR) designs across data sources. We report total patient exposure time in 1,000 patient-year (KYr) units, uncalibrated and calibrated hazard ratios (HRs) and their 95% confidence intervals (CIs). Greyed out entries did not pass study diagnostics and are included only for completeness.

| | Patients | | Exposure (KYr) | | Outcomes | | Incidence (per KYr) | |
|---|---|---|---|---|---|---|---|---|
| | T | C | T | C | T | C | T | C |
| **PS-matching / on-treatment TAR** | | | | | | | | |
| CCAE | 16,075 | 35,099 | 11.9 | 25.1 | <5 | 5 | < 0.42 (0.14 - 0.98) | 0.20 (0.06 - 0.46) |
| DAG | 3,180 | 682 | 2.5 | 0.7 | 0 | 0 | | |
| ODOD | 11,831 | 40,935 | 7.4 | 34.3 | 5 | 22 | 0.68 (0.22 - 1.58) | 0.64 (0.40 - 0.97) |
| MDCR | 1,330 | 6,790 | 0.9 | 5.5 | <5 | <5 | < 5.72 (1.86 - 13.3) | < 0.91 (0.30 - 2.13) |
| MDCD | 2,384 | 11,649 | 1.3 | 7.3 | 0 | 7 | | 0.96 (0.39 - 1.98) |
| Open Claims | 261,706 | 831,984 | 196.3 | 788.1 | 39 | 313 | 0.20 (0.14 - 0.27) | 0.40 (0.35 - 0.44) |
| OEHR | 15,481 | 64,204 | 4.9 | 36.8 | <5 | 21 | < 1.03 (0.33 - 2.40) | 0.57 (0.35 - 0.87) |
| SIDIAP | 2,849 | 964 | 4.0 | 1.7 | 0 | 0 | | |
| Meta-analysis | 273,537 | 872,919 | 203.7 | 822.3 | 44 | 335 | 0.22 (0.16 - 0.29) | 0.41 (0.36 - 0.45) |
| Meta-analysis (w/o USOC) | 11,831 | 40,935 | 7.4 | 34.3 | 5 | 22 | 0.68 (0.22 - 1.58) | 0.64 (0.40 - 0.97) |
| **PS-stratification / on-treatment TAR** | | | | | | | | |
| CCAE | 18,874 | 35,794 | 13.7 | 25.4 | 5 | 5 | 0.37 (0.12 - 0.85) | 0.20 (0.06 - 0.46) |
| DAG | 3,231 | 1,228 | 2.5 | 1.2 | 0 | 0 | | |
| ODOD | 12,831 | 41,835 | 7.9 | 35.2 | 5 | 24 | 0.64 (0.21 - 1.48) | 0.68 (0.44 - 1.02) |
| MDCR | 1,486 | 7,280 | 0.9 | 5.9 | <5 | <5 | < 5.31 (1.73 - 12.4) | < 0.85 (0.28 - 1.98) |
| MDCD | 2,431 | 11,911 | 1.3 | 7.5 | 0 | 7 | | 0.94 (0.38 - 1.93) |
| Open Claims | 276,304 | 842,001 | 204.7 | 797.4 | 39 | 323 | 0.19 (0.14 - 0.26) | 0.41 (0.36 - 0.45) |
| OEHR | 16,654 | 70,443 | 5.2 | 40.6 | <5 | 24 | < 0.96 (0.31 - 2.23) | 0.59 (0.38 - 0.88) |
| SIDIAP | 2,950 | 2,011 | 4.0 | 4.3 | 0 | 0 | | |
| Meta-analysis | 305,789 | 954,279 | 217.8 | 873.1 | <49 | 371 | < 0.22 (0.17 - 0.30) | 0.42 (0.38 - 0.47) |
| Meta-analysis (w/o USOC) | 29,485 | 112,278 | 13.1 | 75.7 | <10 | 48 | < 0.76 (0.37 - 1.40) | 0.63 (0.47 - 0.84) |
| **PS-matching / on-treatment TAR without escalation** | | | | | | | | |
| CCAE | 16,075 | 35,099 | 10.2 | 21.6 | <5 | 5 | < 0.49 (0.16 - 1.14) | 0.23 (0.08 - 0.54) |
| DAG | 3,180 | 682 | 2.3 | 0.6 | 0 | 0 | | |
| ODOD | 11,831 | 40,935 | 6.5 | 30.6 | <5 | 18 | < 0.77 (0.25 - 1.79) | 0.59 (0.35 - 0.93) |
| MDCR | 1,330 | 6,790 | 0.8 | 5.0 | <5 | <5 | < 6.47 (2.10 - 15.1) | < 1.00 (0.32 - 2.33) |
| MDCD | 2,384 | 11,649 | 1.1 | 6.0 | 0 | <5 | | < 0.83 (0.27 - 1.93) |
| Open Claims | 261,706 | 831,984 | 164.8 | 663.6 | 27 | 239 | 0.16 (0.11 - 0.24) | 0.36 (0.32 - 0.41) |
| OEHR | 15,481 | 64,204 | 4.2 | 31.2 | <5 | 13 | < 1.18 (0.38 - 2.76) | 0.42 (0.22 - 0.71) |
| SIDIAP | 2,849 | 964 | 3.5 | 1.4 | 0 | 0 | | |
| Meta-analysis | 261,706 | 831,984 | 164.8 | 663.6 | 27 | 239 | 0.16 (0.11 - 0.24) | 0.36 (0.32 - 0.41) |
| **PS-stratification / on-treatment TAR without escalation** | | | | | | | | |
| CCAE | 18,874 | 35,794 | 11.8 | 21.8 | <5 | 5 | < 0.42 (0.14 - 0.99) | 0.23 (0.07 - 0.53) |
| DAG | 3,231 | 1,228 | 2.3 | 1.2 | 0 | 0 | | |
| ODOD | 12,831 | 41,835 | 7.0 | 31.4 | <5 | 20 | < 0.72 (0.23 - 1.67) | 0.64 (0.39 - 0.98) |
| MDCR | 1,486 | 7,280 | 0.8 | 5.4 | <5 | <5 | < 6.00 (1.95 - 14.0) | < 0.93 (0.30 - 2.17) |
| MDCD | 2,431 | 11,911 | 1.1 | 6.2 | 0 | <5 | | < 0.81 (0.26 - 1.89) |
| Open Claims | 276,304 | 842,001 | 172.1 | 671.7 | 27 | 248 | 0.16 (0.10 - 0.23) | 0.37 (0.32 - 0.42) |
| OEHR | 16,654 | 70,443 | 4.6 | 34.3 | <5 | 16 | < 1.10 (0.36 - 2.56) | 0.47 (0.27 - 0.76) |
| SIDIAP | 2,950 | 2,011 | 3.5 | 3.6 | 0 | 0 | | |
| Meta-analysis | 289,135 | 883,836 | 179.1 | 703.1 | <32 | 268 | < 0.18 (0.12 - 0.25) | 0.38 (0.34 - 0.43) |
| Meta-analysis (w/o USOC) | 12,831 | 41,835 | 7.0 | 31.4 | <5 | 20 | < 0.72 (0.23 - 1.67) | 0.64 (0.39 - 0.98) |

##### 4.3.22 Nausea

**Supplementary Table 359:** Exposures, outcomes and risk estimates for Nausea in SGLT2I (female) (T) vs SU (female) (C) new-user cohorts under four different propensity score (PS) adjustment / time-at-risk (TAR) designs across data sources. We report total patient exposure time in 1,000 patient-year (KYr) units, uncalibrated and calibrated hazard ratios (HRs) and their 95% confidence intervals (CIs). Greyed out entries did not pass study diagnostics and are included only for completeness.

| | Patients | | Exposure (KYr) | | Outcomes | | Incidence (per KYr) | |
|---|---|---|---|---|---|---|---|---|
| | T | C | T | C | T | C | T | C |
| **PS-matching / on-treatment TAR** | | | | | | | | |
| CCAE | 13,066 | 28,698 | 9.5 | 20.3 | 435 | 1,000 | 45.8 (41.6 - 50.3) | 49.3 (46.3 - 52.5) |
| DAG | 2,817 | 604 | 2.2 | 0.6 | 27 | 6 | 12.3 (8.10 - 17.9) | 10.4 (3.82 - 22.7) |
| ODOD | 9,424 | 32,387 | 5.8 | 26.5 | 296 | 1,541 | 50.8 (45.2 - 57.0) | 58.2 (55.3 - 61.1) |
| MDCR | 1,133 | 5,621 | 0.7 | 4.4 | 31 | 212 | 42.2 (28.7 - 59.9) | 48.0 (41.8 - 55.0) |
| MDCD | 1,514 | 7,446 | 0.8 | 4.6 | 78 | 483 | 96.0 (75.9 - 120) | 105 (96.2 - 115) |
| Open Claims | 210,078 | 681,648 | 158.9 | 643.8 | 3,770 | 18,261 | 23.7 (23.0 - 24.5) | 28.4 (28.0 - 28.8) |
| OEHR | 13,014 | 54,321 | 3.9 | 29.7 | 215 | 1,686 | 54.4 (47.4 - 62.2) | 56.8 (54.1 - 59.6) |
| SIDIAP | 2,664 | 897 | 3.7 | 1.6 | 20 | 12 | 5.41 (3.31 - 8.36) | 7.58 (3.92 - 13.2) |
| Meta-analysis | 250,893 | 811,018 | 183.4 | 730.9 | 4,845 | 23,195 | 26.4 (25.7 - 27.2) | 31.7 (31.3 - 32.1) |
| Meta-analysis (w/o USOC) | 40,815 | 129,370 | 24.5 | 87.0 | 1,075 | 4,934 | 43.9 (41.3 - 46.6) | 56.7 (55.1 - 58.3) |
| **PS-stratification / on-treatment TAR** | | | | | | | | |
| CCAE | 15,258 | 29,286 | 10.9 | 20.5 | 515 | 1,013 | 47.1 (43.2 - 51.4) | 49.4 (46.4 - 52.5) |
| DAG | 2,864 | 1,091 | 2.2 | 1.1 | 27 | 15 | 12.2 (8.01 - 17.7) | 13.7 (7.67 - 22.6) |
| ODOD | 10,200 | 33,096 | 6.2 | 27.2 | 321 | 1,590 | 51.9 (46.4 - 57.9) | 58.6 (55.7 - 61.5) |
| MDCR | 1,258 | 5,994 | 0.8 | 4.7 | 32 | 233 | 40.6 (27.8 - 57.4) | 49.6 (43.4 - 56.4) |
| MDCD | 1,543 | 7,617 | 0.8 | 4.7 | 80 | 494 | 97.2 (77.1 - 121) | 105 (96.1 - 115) |
| Open Claims | 221,452 | 690,768 | 165.5 | 652.4 | 3,960 | 18,680 | 23.9 (23.2 - 24.7) | 28.6 (28.2 - 29.0) |
| OEHR | 14,004 | 59,772 | 4.3 | 32.8 | 230 | 1,880 | 53.8 (47.1 - 61.2) | 57.3 (54.8 - 60.0) |
| SIDIAP | 2,761 | 1,897 | 3.7 | 4.1 | 21 | 41 | 5.60 (3.47 - 8.56) | 10.1 (7.24 - 13.7) |
| Meta-analysis | 260,914 | 812,922 | 186.8 | 732.8 | 5,026 | 23,163 | 26.9 (26.2 - 27.7) | 31.6 (31.2 - 32.0) |
| Meta-analysis (w/o USOC) | 39,462 | 122,154 | 21.4 | 80.5 | 1,066 | 4,483 | 49.9 (46.9 - 52.9) | 55.7 (54.1 - 57.4) |
| **PS-matching / on-treatment TAR without escalation** | | | | | | | | |
| CCAE | 13,066 | 28,698 | 8.2 | 17.6 | 383 | 845 | 46.4 (41.9 - 51.3) | 48.0 (44.8 - 51.3) |
| DAG | 2,817 | 604 | 2.0 | 0.5 | 23 | 6 | 11.5 (7.30 - 17.3) | 11.2 (4.12 - 24.4) |
| ODOD | 9,424 | 32,387 | 5.2 | 23.8 | 257 | 1,362 | 49.3 (43.5 - 55.8) | 57.2 (54.2 - 60.3) |
| MDCR | 1,133 | 5,621 | 0.6 | 4.0 | 28 | 192 | 43.3 (28.8 - 62.6) | 47.7 (41.2 - 54.9) |
| MDCD | 1,514 | 7,446 | 0.7 | 3.9 | 63 | 411 | 91.3 (70.2 - 117) | 106 (96.1 - 117) |
| Open Claims | 210,078 | 681,648 | 134.4 | 546.6 | 3,072 | 15,114 | 22.8 (22.0 - 23.7) | 27.6 (27.2 - 28.1) |
| OEHR | 13,014 | 54,321 | 3.5 | 25.6 | 179 | 1,333 | 51.6 (44.3 - 59.8) | 52.0 (49.2 - 54.9) |
| SIDIAP | 2,664 | 897 | 3.2 | 1.3 | 16 | 10 | 4.92 (2.81 - 8.00) | 7.54 (3.61 - 13.9) |
| Meta-analysis | 250,893 | 811,018 | 156.0 | 622.9 | 3,998 | 19,267 | 25.6 (24.8 - 26.4) | 30.9 (30.5 - 31.4) |
| Meta-analysis (w/o USOC) | 40,815 | 129,370 | 21.5 | 76.3 | 926 | 4,153 | 43.1 (40.3 - 45.9) | 54.4 (52.8 - 56.1) |
| **PS-stratification / on-treatment TAR without escalation** | | | | | | | | |
| CCAE | 15,258 | 29,286 | 9.5 | 17.8 | 450 | 858 | 47.3 (43.1 - 51.9) | 48.1 (44.9 - 51.4) |
| DAG | 2,864 | 1,091 | 2.0 | 1.0 | 23 | 14 | 11.4 (7.23 - 17.1) | 13.8 (7.53 - 23.1) |
| ODOD | 10,200 | 33,096 | 5.5 | 24.4 | 277 | 1,408 | 50.1 (44.3 - 56.3) | 57.7 (54.7 - 60.8) |
| MDCR | 1,258 | 5,994 | 0.7 | 4.3 | 29 | 210 | 41.6 (27.9 - 59.8) | 48.9 (42.5 - 55.9) |
| MDCD | 1,543 | 7,617 | 0.7 | 4.0 | 65 | 420 | 93.0 (71.8 - 118) | 106 (96.1 - 117) |
| Open Claims | 221,452 | 690,768 | 140.3 | 554.1 | 3,228 | 15,476 | 23.0 (22.2 - 23.8) | 27.9 (27.5 - 28.4) |
| OEHR | 14,004 | 59,772 | 3.8 | 28.3 | 190 | 1,496 | 50.6 (43.7 - 58.3) | 52.9 (50.2 - 55.6) |
| SIDIAP | 2,761 | 1,897 | 3.3 | 3.4 | 16 | 34 | 4.85 (2.77 - 7.87) | 10.1 (7.02 - 14.2) |
| Meta-analysis | 260,914 | 812,922 | 159.1 | 624.7 | 4,145 | 19,238 | 26.1 (25.3 - 26.9) | 30.8 (30.4 - 31.2) |
| Meta-analysis (w/o USOC) | 39,462 | 122,154 | 18.8 | 70.6 | 917 | 3,762 | 48.8 (45.7 - 52.0) | 53.3 (51.6 - 55.1) |

##### 4.3.23 Peripheral edema

**Supplementary Table 360:** Exposures, outcomes and risk estimates for Peripheral edema in SGLT2I (female) (T) vs SU (female) (C) new-user cohorts under four different propensity score (PS) adjustment / time-at-risk (TAR) designs across data sources. We report total patient exposure time in 1,000 patient-year (KYr) units, uncalibrated and calibrated hazard ratios (HRs) and their 95% confidence intervals (CIs). Greyed out entries did not pass study diagnostics and are included only for completeness.

| | Patients | | Exposure (KYr) | | Outcomes | | Incidence (per KYr) | |
|---|---|---|---|---|---|---|---|---|
| | T | C | T | C | T | C | T | C |
| **PS-matching / on-treatment TAR** | | | | | | | | |
| CCAE | 12,325 | 27,200 | 8.9 | 18.7 | 369 | 1,065 | 41.7 (37.5 - 46.1) | 57.1 (53.7 - 60.6) |
| DAG | 2,530 | 557 | 2.0 | 0.5 | 33 | 20 | 16.6 (11.4 - 23.3) | 39.7 (24.3 - 61.3) |
| ODOD | 8,641 | 29,325 | 5.3 | 22.7 | 310 | 2,178 | 59.0 (52.7 - 66.0) | 95.7 (91.8 - 99.9) |
| MDCR | 983 | 4,713 | 0.6 | 3.6 | 28 | 377 | 43.9 (29.2 - 63.4) | 106 (95.2 - 117) |
| MDCD | 1,517 | 7,521 | 0.8 | 4.3 | 69 | 454 | 89.9 (69.9 - 114) | 105 (95.3 - 115) |
| Open Claims | 192,709 | 626,018 | 144.3 | 569.4 | 3,477 | 24,047 | 24.1 (23.3 - 24.9) | 42.2 (41.7 - 42.8) |
| OEHR | 12,363 | 51,739 | 3.8 | 27.4 | 149 | 1,830 | 39.4 (33.3 - 46.2) | 66.8 (63.8 - 70.0) |
| SIDIAP | 2,386 | 847 | 3.4 | 1.5 | 47 | 35 | 14.0 (10.3 - 18.6) | 23.9 (16.6 - 33.2) |
| Meta-analysis | 230,924 | 747,363 | 167.0 | 647.5 | 4,449 | 29,986 | 26.6 (25.9 - 27.4) | 46.3 (45.8 - 46.8) |
| Meta-analysis (w/o USOC) | 38,215 | 121,345 | 22.7 | 78.2 | 972 | 5,939 | 42.9 (40.2 - 45.7) | 76.0 (74.1 - 77.9) |
| **PS-stratification / on-treatment TAR** | | | | | | | | |
| CCAE | 14,270 | 27,767 | 10.1 | 18.9 | 434 | 1,084 | 43.2 (39.2 - 47.4) | 57.4 (54.0 - 60.9) |
| DAG | 2,565 | 990 | 2.0 | 1.0 | 33 | 47 | 16.4 (11.3 - 23.1) | 48.8 (35.9 - 64.9) |
| ODOD | 9,264 | 29,843 | 5.5 | 23.2 | 325 | 2,258 | 58.9 (52.6 - 65.6) | 97.5 (93.5 - 102) |
| MDCR | 1,087 | 4,955 | 0.7 | 3.7 | 30 | 397 | 44.7 (30.1 - 63.7) | 106 (96.1 - 117) |
| MDCD | 1,536 | 7,651 | 0.8 | 4.4 | 69 | 460 | 89.1 (69.3 - 113) | 104 (95.0 - 114) |
| Open Claims | 201,879 | 633,848 | 149.5 | 576.2 | 3,614 | 24,744 | 24.2 (23.4 - 25.0) | 42.9 (42.4 - 43.5) |
| OEHR | 13,269 | 56,725 | 4.1 | 30.0 | 157 | 2,118 | 38.5 (32.7 - 45.0) | 70.6 (67.6 - 73.7) |
| SIDIAP | 2,460 | 1,790 | 3.4 | 3.7 | 48 | 84 | 14.1 (10.4 - 18.7) | 22.5 (17.9 - 27.8) |
| Meta-analysis | 238,682 | 748,183 | 169.2 | 648.3 | 4,530 | 30,204 | 26.8 (26.0 - 27.6) | 46.6 (46.1 - 47.1) |
| Meta-analysis (w/o USOC) | 36,803 | 114,335 | 19.7 | 72.1 | 916 | 5,460 | 46.6 (43.6 - 49.7) | 75.8 (73.8 - 77.8) |
| **PS-matching / on-treatment TAR without escalation** | | | | | | | | |
| CCAE | 12,325 | 27,200 | 7.7 | 16.2 | 313 | 924 | 40.8 (36.4 - 45.6) | 57.0 (53.4 - 60.8) |
| DAG | 2,530 | 557 | 1.8 | 0.5 | 31 | 17 | 17.1 (11.6 - 24.3) | 36.1 (21.0 - 57.8) |
| ODOD | 8,641 | 29,325 | 4.7 | 20.5 | 273 | 1,955 | 58.3 (51.6 - 65.6) | 95.6 (91.4 - 99.9) |
| MDCR | 983 | 4,713 | 0.6 | 3.3 | 26 | 353 | 45.1 (29.5 - 66.1) | 107 (96.2 - 119) |
| MDCD | 1,517 | 7,521 | 0.6 | 3.7 | 60 | 393 | 94.7 (72.2 - 122) | 107 (96.5 - 118) |
| Open Claims | 192,709 | 626,018 | 121.8 | 483.6 | 2,870 | 20,251 | 23.6 (22.7 - 24.4) | 41.9 (41.3 - 42.5) |
| OEHR | 12,363 | 51,739 | 3.3 | 23.7 | 125 | 1,501 | 37.8 (31.5 - 45.1) | 63.4 (60.3 - 66.7) |
| SIDIAP | 2,386 | 847 | 3.0 | 1.2 | 40 | 30 | 13.5 (9.61 - 18.3) | 24.6 (16.6 - 35.2) |
| Meta-analysis | 230,924 | 747,363 | 141.7 | 552.1 | 3,707 | 25,407 | 26.2 (25.3 - 27.0) | 46.0 (45.5 - 46.6) |
| Meta-analysis (w/o USOC) | 38,215 | 121,345 | 19.8 | 68.5 | 837 | 5,156 | 42.2 (39.4 - 45.1) | 75.2 (73.2 - 77.3) |
| **PS-stratification / on-treatment TAR without escalation** | | | | | | | | |
| CCAE | 14,270 | 27,767 | 8.7 | 16.4 | 372 | 940 | 42.7 (38.5 - 47.3) | 57.2 (53.6 - 61.0) |
| DAG | 2,565 | 990 | 1.8 | 0.9 | 31 | 44 | 17.0 (11.6 - 24.2) | 49.1 (35.7 - 66.0) |
| ODOD | 9,264 | 29,843 | 4.9 | 20.8 | 288 | 2,029 | 58.3 (51.8 - 65.5) | 97.3 (93.1 - 102) |
| MDCR | 1,087 | 4,955 | 0.6 | 3.5 | 28 | 373 | 46.1 (30.6 - 66.7) | 108 (97.3 - 120) |
| MDCD | 1,536 | 7,651 | 0.6 | 3.7 | 60 | 396 | 93.9 (71.6 - 121) | 106 (95.6 - 117) |
| Open Claims | 201,879 | 633,848 | 126.4 | 489.6 | 2,985 | 20,855 | 23.6 (22.8 - 24.5) | 42.6 (42.0 - 43.2) |
| OEHR | 13,269 | 56,725 | 3.6 | 25.9 | 131 | 1,731 | 36.8 (30.8 - 43.7) | 66.7 (63.6 - 69.9) |
| SIDIAP | 2,460 | 1,790 | 3.0 | 3.1 | 41 | 72 | 13.6 (9.76 - 18.5) | 23.3 (18.2 - 29.3) |
| Meta-analysis | 238,682 | 748,183 | 143.6 | 552.9 | 3,776 | 25,555 | 26.3 (25.5 - 27.1) | 46.2 (45.7 - 46.8) |
| Meta-analysis (w/o USOC) | 36,803 | 114,335 | 17.2 | 63.2 | 791 | 4,700 | 46.0 (42.8 - 49.3) | 74.3 (72.2 - 76.5) |

##### 4.3.24 Photosensitivity

**Supplementary Table 361:** Exposures, outcomes and risk estimates for Photosensitivity in SGLT2I (female) (T) vs SU (female) (C) new-user cohorts under four different propensity score (PS) adjustment / time-at-risk (TAR) designs across data sources. We report total patient exposure time in 1,000 patient-year (KYr) units, uncalibrated and calibrated hazard ratios (HRs) and their 95% confidence intervals (CIs). Greyed out entries did not pass study diagnostics and are included only for completeness.

| | Patients | | Exposure (KYr) | | Outcomes | | Incidence (per KYr) | |
|---|---|---|---|---|---|---|---|---|
| | T | C | T | C | T | C | T | C |
| **PS-matching / on-treatment TAR** | | | | | | | | |
| CCAE | 15,922 | 34,859 | 11.7 | 24.8 | 9 | 31 | 0.77 (0.35 - 1.46) | 1.25 (0.85 - 1.77) |
| DAG | 3,165 | 680 | 2.5 | 0.7 | <5 | 0 | < 2.00 (0.65 - 4.68) | |
| ODOD | 11,746 | 40,624 | 7.3 | 33.9 | <5 | 31 | < 0.68 (0.22 - 1.60) | 0.91 (0.62 - 1.30) |
| MDCR | 1,320 | 6,692 | 0.9 | 5.4 | <5 | 8 | < 5.75 (1.87 - 13.4) | 1.48 (0.64 - 2.91) |
| MDCD | 2,373 | 11,605 | 1.3 | 7.3 | <5 | 8 | < 3.89 (1.26 - 9.09) | 1.10 (0.48 - 2.17) |
| Open Claims | 259,076 | 825,095 | 194.3 | 780.5 | 65 | 362 | 0.33 (0.26 - 0.43) | 0.46 (0.42 - 0.51) |
| OEHR | 15,371 | 63,759 | 4.8 | 36.5 | <5 | 52 | < 1.04 (0.34 - 2.42) | 1.42 (1.06 - 1.87) |
| SIDIAP | 2,837 | 962 | 4.0 | 1.7 | 0 | <5 | | < 2.99 (0.97 - 6.97) |
| Meta-analysis | 302,115 | 964,337 | 218.1 | 875.7 | <84 | 476 | < 0.39 (0.31 - 0.48) | 0.54 (0.50 - 0.59) |
| Meta-analysis (w/o USOC) | 43,039 | 139,242 | 23.8 | 95.3 | <19 | 114 | < 0.80 (0.48 - 1.24) | 1.20 (0.99 - 1.44) |
| **PS-stratification / on-treatment TAR** | | | | | | | | |
| CCAE | 18,681 | 35,476 | 13.5 | 25.1 | 11 | 31 | 0.81 (0.41 - 1.46) | 1.24 (0.84 - 1.75) |
| DAG | 3,216 | 1,225 | 2.5 | 1.2 | <5 | <5 | < 1.98 (0.64 - 4.63) | < 4.03 (1.31 - 9.41) |
| ODOD | 12,737 | 41,516 | 7.8 | 34.8 | <5 | 33 | < 0.64 (0.21 - 1.50) | 0.95 (0.65 - 1.33) |
| MDCR | 1,473 | 7,176 | 0.9 | 5.8 | <5 | 8 | < 5.35 (1.74 - 12.5) | 1.38 (0.59 - 2.71) |
| MDCD | 2,416 | 11,868 | 1.3 | 7.5 | <5 | 8 | < 3.84 (1.25 - 8.97) | 1.07 (0.46 - 2.11) |
| Open Claims | 273,499 | 835,013 | 202.5 | 789.7 | 72 | 375 | 0.36 (0.28 - 0.45) | 0.47 (0.43 - 0.53) |
| OEHR | 16,535 | 69,939 | 5.2 | 40.2 | <5 | 61 | < 0.96 (0.31 - 2.25) | 1.52 (1.16 - 1.95) |
| SIDIAP | 2,938 | 2,004 | 4.0 | 4.3 | 0 | <5 | | < 1.15 (0.37 - 2.69) |
| Meta-analysis | 321,452 | 981,944 | 229.0 | 889.8 | <93 | 500 | < 0.41 (0.33 - 0.50) | 0.56 (0.51 - 0.61) |
| Meta-analysis (w/o USOC) | 47,953 | 146,931 | 26.5 | 100.1 | <21 | 125 | < 0.79 (0.49 - 1.21) | 1.25 (1.04 - 1.49) |
| **PS-matching / on-treatment TAR without escalation** | | | | | | | | |
| CCAE | 15,922 | 34,859 | 10.1 | 21.4 | 8 | 28 | 0.79 (0.34 - 1.56) | 1.31 (0.87 - 1.89) |
| DAG | 3,165 | 680 | 2.3 | 0.6 | <5 | 0 | < 2.20 (0.71 - 5.14) | |
| ODOD | 11,746 | 40,624 | 6.5 | 30.3 | <5 | 31 | < 0.77 (0.25 - 1.80) | 1.02 (0.70 - 1.45) |
| MDCR | 1,320 | 6,692 | 0.8 | 4.9 | <5 | 6 | < 6.51 (2.11 - 15.2) | 1.22 (0.45 - 2.65) |
| MDCD | 2,373 | 11,605 | 1.1 | 6.0 | <5 | 6 | < 4.72 (1.53 - 11.0) | 1.00 (0.37 - 2.17) |
| Open Claims | 259,076 | 825,095 | 163.0 | 657.2 | 53 | 312 | 0.33 (0.24 - 0.43) | 0.47 (0.42 - 0.53) |
| OEHR | 15,371 | 63,759 | 4.2 | 30.9 | <5 | 45 | < 1.19 (0.39 - 2.78) | 1.46 (1.06 - 1.95) |
| SIDIAP | 2,837 | 962 | 3.5 | 1.4 | 0 | <5 | | < 3.54 (1.15 - 8.27) |
| Meta-analysis | 302,115 | 964,337 | 183.8 | 739.7 | <71 | 416 | < 0.39 (0.30 - 0.49) | 0.56 (0.51 - 0.62) |
| Meta-analysis (w/o USOC) | 43,039 | 139,242 | 20.8 | 82.6 | <18 | 104 | < 0.87 (0.51 - 1.37) | 1.26 (1.03 - 1.53) |
| **PS-stratification / on-treatment TAR without escalation** | | | | | | | | |
| CCAE | 18,681 | 35,476 | 11.7 | 21.6 | 10 | 28 | 0.86 (0.41 - 1.58) | 1.30 (0.86 - 1.88) |
| DAG | 3,216 | 1,225 | 2.3 | 1.1 | <5 | <5 | < 2.18 (0.71 - 5.09) | < 4.36 (1.42 - 10.2) |
| ODOD | 12,737 | 41,516 | 6.9 | 31.1 | <5 | 32 | < 0.72 (0.23 - 1.69) | 1.03 (0.70 - 1.45) |
| MDCR | 1,473 | 7,176 | 0.8 | 5.3 | <5 | 6 | < 6.04 (1.96 - 14.1) | 1.13 (0.42 - 2.46) |
| MDCD | 2,416 | 11,868 | 1.1 | 6.2 | <5 | 6 | < 4.65 (1.51 - 10.9) | 0.97 (0.36 - 2.12) |
| Open Claims | 273,499 | 835,013 | 170.2 | 665.2 | 59 | 324 | 0.35 (0.26 - 0.45) | 0.49 (0.44 - 0.54) |
| OEHR | 16,535 | 69,939 | 4.5 | 34.0 | <5 | 51 | < 1.11 (0.36 - 2.58) | 1.50 (1.12 - 1.97) |
| SIDIAP | 2,938 | 2,004 | 3.5 | 3.6 | 0 | <5 | | < 1.40 (0.45 - 3.27) |
| Meta-analysis | 321,452 | 981,944 | 193.3 | 751.9 | <79 | 435 | < 0.41 (0.32 - 0.51) | 0.58 (0.53 - 0.64) |
| Meta-analysis (w/o USOC) | 47,953 | 146,931 | 23.1 | 86.7 | <20 | 111 | < 0.87 (0.53 - 1.34) | 1.28 (1.05 - 1.54) |

##### 4.3.25 Venous thromboembolism

**Supplementary Table 362:** Exposures, outcomes and risk estimates for Venous thromboembolism in SGLT2I (female) (T) vs SU (female) (C) new-user cohorts under four different propensity score (PS) adjustment / time-at-risk (TAR) designs across data sources. We report total patient exposure time in 1,000 patient-year (KYr) units, uncalibrated and calibrated hazard ratios (HRs) and their 95% confidence intervals (CIs). Greyed out entries did not pass study diagnostics and are included only for completeness.

| | Patients | | Exposure (KYr) | | Outcomes | | Incidence (per KYr) | |
|---|---|---|---|---|---|---|---|---|
| | T | C | T | C | T | C | T | C |
| **PS-matching / on-treatment TAR** | | | | | | | | |
| CCAE | 15,702 | 34,115 | 11.6 | 24.3 | 50 | 153 | 4.32 (3.21 - 5.70) | 6.29 (5.33 - 7.37) |
| DAG | 2,914 | 625 | 2.3 | 0.6 | 15 | 7 | 6.64 (3.71 - 10.9) | 11.8 (4.75 - 24.3) |
| ODOD | 11,476 | 39,372 | 7.1 | 32.8 | 58 | 373 | 8.12 (6.17 - 10.5) | 11.4 (10.3 - 12.6) |
| MDCR | 1,277 | 6,479 | 0.9 | 5.2 | 5 | 81 | 5.88 (1.91 - 13.7) | 15.6 (12.4 - 19.4) |
| MDCD | 2,277 | 11,099 | 1.2 | 6.9 | 10 | 76 | 8.13 (3.90 - 14.9) | 11.0 (8.67 - 13.8) |
| Open Claims | 253,292 | 804,409 | 190.1 | 758.7 | 696 | 4,734 | 3.66 (3.39 - 3.94) | 6.24 (6.06 - 6.42) |
| OEHR | 15,125 | 62,308 | 4.7 | 35.3 | 38 | 328 | 8.04 (5.69 - 11.0) | 9.28 (8.31 - 10.3) |
| SIDIAP | 2,721 | 929 | 3.8 | 1.6 | 14 | 5 | 3.70 (2.02 - 6.21) | 3.12 (1.01 - 7.29) |
| Meta-analysis | 299,149 | 957,782 | 215.6 | 863.2 | 857 | 5,745 | 3.98 (3.71 - 4.25) | 6.66 (6.48 - 6.83) |
| Meta-analysis (w/o USOC) | 45,857 | 153,373 | 25.5 | 104.5 | 161 | 1,011 | 6.31 (5.37 - 7.36) | 9.67 (9.08 - 10.3) |
| **PS-stratification / on-treatment TAR** | | | | | | | | |
| CCAE | 18,464 | 34,895 | 13.4 | 24.6 | 59 | 155 | 4.42 (3.36 - 5.70) | 6.29 (5.34 - 7.36) |
| DAG | 2,961 | 1,106 | 2.3 | 1.1 | 15 | 13 | 6.57 (3.68 - 10.8) | 11.8 (6.26 - 20.1) |
| ODOD | 12,418 | 40,278 | 7.6 | 33.7 | 60 | 393 | 7.91 (6.03 - 10.2) | 11.7 (10.5 - 12.9) |
| MDCR | 1,426 | 6,917 | 0.9 | 5.6 | 5 | 87 | 5.48 (1.78 - 12.8) | 15.7 (12.5 - 19.3) |
| MDCD | 2,324 | 11,379 | 1.2 | 7.1 | 10 | 84 | 8.00 (3.84 - 14.7) | 11.8 (9.44 - 14.6) |
| Open Claims | 267,177 | 813,929 | 198.1 | 767.6 | 730 | 4,915 | 3.69 (3.42 - 3.96) | 6.40 (6.23 - 6.58) |
| OEHR | 16,265 | 68,607 | 5.1 | 39.0 | 38 | 385 | 7.47 (5.28 - 10.2) | 9.86 (8.90 - 10.9) |
| SIDIAP | 2,816 | 1,941 | 3.8 | 4.2 | 15 | 15 | 3.91 (2.19 - 6.44) | 3.60 (2.01 - 5.93) |
| Meta-analysis | 314,324 | 957,709 | 224.1 | 864.9 | 887 | 5,848 | 3.96 (3.70 - 4.23) | 6.76 (6.59 - 6.94) |
| Meta-analysis (w/o USOC) | 47,147 | 143,780 | 26.0 | 97.3 | 157 | 933 | 6.03 (5.12 - 7.05) | 9.58 (8.98 - 10.2) |
| **PS-matching / on-treatment TAR without escalation** | | | | | | | | |
| CCAE | 15,702 | 34,115 | 9.9 | 20.9 | 41 | 137 | 4.12 (2.96 - 5.59) | 6.55 (5.50 - 7.74) |
| DAG | 2,914 | 625 | 2.1 | 0.6 | 13 | 7 | 6.31 (3.36 - 10.8) | 12.7 (5.10 - 26.1) |
| ODOD | 11,476 | 39,372 | 6.3 | 29.3 | 51 | 336 | 8.07 (6.01 - 10.6) | 11.5 (10.3 - 12.8) |
| MDCR | 1,277 | 6,479 | 0.8 | 4.7 | <5 | 75 | < 6.66 (2.16 - 15.5) | 15.9 (12.5 - 19.9) |
| MDCD | 2,277 | 11,099 | 1.0 | 5.7 | 7 | 57 | 6.92 (2.78 - 14.3) | 9.96 (7.54 - 12.9) |
| Open Claims | 253,292 | 804,409 | 159.7 | 639.2 | 569 | 4,046 | 3.56 (3.28 - 3.87) | 6.33 (6.14 - 6.53) |
| OEHR | 15,125 | 62,308 | 4.1 | 30.0 | 32 | 255 | 7.80 (5.33 - 11.0) | 8.50 (7.49 - 9.61) |
| SIDIAP | 2,721 | 929 | 3.3 | 1.3 | 13 | <5 | 3.91 (2.08 - 6.68) | < 3.71 (1.20 - 8.65) |
| Meta-analysis | 299,149 | 957,782 | 181.8 | 729.9 | <705 | 4,906 | < 3.88 (3.60 - 4.17) | 6.72 (6.53 - 6.91) |
| Meta-analysis (w/o USOC) | 45,857 | 153,373 | 22.1 | 90.7 | <136 | 860 | < 6.14 (5.15 - 7.27) | 9.49 (8.86 - 10.1) |
| **PS-stratification / on-treatment TAR without escalation** | | | | | | | | |
| CCAE | 18,464 | 34,895 | 11.5 | 21.2 | 49 | 139 | 4.25 (3.15 - 5.62) | 6.56 (5.51 - 7.74) |
| DAG | 2,961 | 1,106 | 2.1 | 1.0 | 13 | 13 | 6.25 (3.33 - 10.7) | 12.7 (6.74 - 21.6) |
| ODOD | 12,418 | 40,278 | 6.7 | 30.1 | 53 | 355 | 7.88 (5.90 - 10.3) | 11.8 (10.6 - 13.1) |
| MDCR | 1,426 | 6,917 | 0.8 | 5.1 | <5 | 80 | < 6.19 (2.01 - 14.4) | 15.8 (12.5 - 19.7) |
| MDCD | 2,324 | 11,379 | 1.0 | 5.9 | 7 | 64 | 6.80 (2.73 - 14.0) | 10.9 (8.38 - 13.9) |
| Open Claims | 267,177 | 813,929 | 166.7 | 646.9 | 597 | 4,205 | 3.58 (3.30 - 3.88) | 6.50 (6.30 - 6.70) |
| OEHR | 16,265 | 68,607 | 4.4 | 33.1 | 32 | 296 | 7.23 (4.94 - 10.2) | 8.93 (7.94 - 10.0) |
| SIDIAP | 2,816 | 1,941 | 3.4 | 3.4 | 13 | 12 | 3.85 (2.05 - 6.58) | 3.49 (1.80 - 6.09) |
| Meta-analysis | 314,324 | 957,709 | 189.3 | 731.4 | 731 | 4,995 | 3.86 (3.59 - 4.15) | 6.83 (6.64 - 7.02) |
| Meta-analysis (w/o USOC) | 47,147 | 143,780 | 22.7 | 84.4 | 134 | 790 | 5.91 (4.95 - 7.00) | 9.36 (8.72 - 10.0) |

##### 4.3.26 Vomiting

**Supplementary Table 363:** Exposures, outcomes and risk estimates for Vomiting in SGLT2I (female) (T) vs SU (female) (C) new-user cohorts under four different propensity score (PS) adjustment / time-at-risk (TAR) designs across data sources. We report total patient exposure time in 1,000 patient-year (KYr) units, uncalibrated and calibrated hazard ratios (HRs) and their 95% confidence intervals (CIs). Greyed out entries did not pass study diagnostics and are included only for completeness.

| | Patients | | Exposure (KYr) | | Outcomes | | Incidence (per KYr) | |
|---|---|---|---|---|---|---|---|---|
| | T | C | T | C | T | C | T | C |
| **PS-matching / on-treatment TAR** | | | | | | | | |
| CCAE | 14,041 | 30,666 | 10.3 | 21.7 | 300 | 694 | 29.3 (26.0 - 32.8) | 31.9 (29.6 - 34.4) |
| DAG | 2,813 | 604 | 2.2 | 0.6 | 27 | 6 | 12.3 (8.10 - 17.9) | 10.4 (3.82 - 22.7) |
| ODOD | 10,217 | 34,872 | 6.3 | 28.8 | 197 | 1,190 | 31.1 (26.9 - 35.7) | 41.4 (39.1 - 43.8) |
| MDCR | 1,211 | 5,956 | 0.8 | 4.7 | 20 | 179 | 25.2 (15.4 - 38.9) | 37.9 (32.6 - 43.9) |
| MDCD | 1,706 | 8,339 | 0.9 | 5.1 | 70 | 352 | 75.5 (58.8 - 95.3) | 68.4 (61.4 - 75.9) |
| Open Claims | 223,660 | 716,261 | 168.9 | 678.9 | 2,751 | 13,887 | 16.3 (15.7 - 16.9) | 20.5 (20.1 - 20.8) |
| OEHR | 13,955 | 57,490 | 4.3 | 32.0 | 129 | 1,172 | 30.0 (25.1 - 35.7) | 36.6 (34.5 - 38.8) |
| SIDIAP | 2,657 | 896 | 3.7 | 1.6 | 25 | 13 | 6.80 (4.40 - 10.0) | 8.24 (4.39 - 14.1) |
| Meta-analysis | 267,447 | 854,480 | 195.2 | 772.9 | 3,492 | 17,487 | 17.9 (17.3 - 18.5) | 22.6 (22.3 - 23.0) |
| Meta-analysis (w/o USOC) | 43,787 | 138,219 | 26.3 | 94.0 | 741 | 3,600 | 28.2 (26.2 - 30.3) | 38.3 (37.1 - 39.6) |
| **PS-stratification / on-treatment TAR** | | | | | | | | |
| CCAE | 16,475 | 31,156 | 11.9 | 21.9 | 353 | 699 | 29.7 (26.7 - 33.0) | 31.9 (29.6 - 34.3) |
| DAG | 2,860 | 1,091 | 2.2 | 1.1 | 27 | 15 | 12.2 (8.02 - 17.7) | 13.7 (7.67 - 22.6) |
| ODOD | 11,062 | 35,657 | 6.7 | 29.5 | 210 | 1,236 | 31.2 (27.1 - 35.7) | 41.9 (39.6 - 44.3) |
| MDCR | 1,346 | 6,340 | 0.8 | 5.0 | 21 | 196 | 24.7 (15.3 - 37.8) | 39.0 (33.7 - 44.8) |
| MDCD | 1,727 | 8,546 | 0.9 | 5.3 | 71 | 361 | 75.8 (59.2 - 95.6) | 68.2 (61.3 - 75.6) |
| Open Claims | 235,920 | 725,274 | 176.1 | 687.4 | 2,889 | 14,218 | 16.4 (15.8 - 17.0) | 20.7 (20.3 - 21.0) |
| OEHR | 15,028 | 63,337 | 4.6 | 35.4 | 137 | 1,315 | 29.5 (24.8 - 34.9) | 37.1 (35.1 - 39.2) |
| SIDIAP | 2,753 | 1,889 | 3.7 | 4.0 | 26 | 41 | 6.97 (4.55 - 10.2) | 10.2 (7.29 - 13.8) |
| Meta-analysis | 278,485 | 855,424 | 199.3 | 774.2 | 3,589 | 17,468 | 18.0 (17.4 - 18.6) | 22.6 (22.2 - 22.9) |
| Meta-analysis (w/o USOC) | 42,565 | 130,150 | 23.2 | 86.8 | 700 | 3,250 | 30.1 (27.9 - 32.4) | 37.4 (36.2 - 38.7) |
| **PS-matching / on-treatment TAR without escalation** | | | | | | | | |
| CCAE | 14,041 | 30,666 | 8.9 | 18.8 | 261 | 586 | 29.4 (26.0 - 33.2) | 31.1 (28.6 - 33.7) |
| DAG | 2,813 | 604 | 2.0 | 0.5 | 23 | 6 | 11.5 (7.30 - 17.3) | 11.2 (4.12 - 24.4) |
| ODOD | 10,217 | 34,872 | 5.6 | 25.8 | 168 | 1,057 | 29.8 (25.5 - 34.7) | 41.0 (38.6 - 43.6) |
| MDCR | 1,211 | 5,956 | 0.7 | 4.3 | 19 | 156 | 27.2 (16.4 - 42.5) | 36.2 (30.7 - 42.3) |
| MDCD | 1,706 | 8,339 | 0.8 | 4.3 | 56 | 298 | 71.9 (54.3 - 93.3) | 69.4 (61.8 - 77.8) |
| Open Claims | 223,660 | 716,261 | 142.6 | 575.1 | 2,209 | 11,482 | 15.5 (14.9 - 16.2) | 20.0 (19.6 - 20.3) |
| OEHR | 13,955 | 57,490 | 3.8 | 27.5 | 106 | 931 | 28.3 (23.1 - 34.2) | 33.9 (31.8 - 36.2) |
| SIDIAP | 2,657 | 896 | 3.2 | 1.3 | 20 | 10 | 6.17 (3.77 - 9.52) | 7.58 (3.64 - 13.9) |
| Meta-analysis | 267,447 | 854,480 | 165.6 | 657.0 | 2,839 | 14,520 | 17.1 (16.5 - 17.8) | 22.1 (21.7 - 22.5) |
| Meta-analysis (w/o USOC) | 43,787 | 138,219 | 23.0 | 82.0 | 630 | 3,038 | 27.4 (25.3 - 29.6) | 37.1 (35.8 - 38.4) |
| **PS-stratification / on-treatment TAR without escalation** | | | | | | | | |
| CCAE | 16,475 | 31,156 | 10.3 | 19.0 | 301 | 591 | 29.2 (26.0 - 32.7) | 31.1 (28.7 - 33.7) |
| DAG | 2,860 | 1,091 | 2.0 | 1.0 | 23 | 14 | 11.4 (7.23 - 17.1) | 13.8 (7.53 - 23.1) |
| ODOD | 11,062 | 35,657 | 6.0 | 26.4 | 179 | 1,101 | 29.9 (25.7 - 34.6) | 41.6 (39.2 - 44.2) |
| MDCR | 1,346 | 6,340 | 0.7 | 4.6 | 20 | 170 | 26.7 (16.3 - 41.2) | 37.0 (31.6 - 43.0) |
| MDCD | 1,727 | 8,546 | 0.8 | 4.4 | 56 | 306 | 71.2 (53.8 - 92.4) | 69.4 (61.8 - 77.6) |
| Open Claims | 235,920 | 725,274 | 148.9 | 582.5 | 2,319 | 11,766 | 15.6 (15.0 - 16.2) | 20.2 (19.8 - 20.6) |
| OEHR | 15,028 | 63,337 | 4.1 | 30.4 | 112 | 1,048 | 27.6 (22.7 - 33.2) | 34.5 (32.5 - 36.7) |
| SIDIAP | 2,753 | 1,889 | 3.3 | 3.3 | 20 | 33 | 6.07 (3.71 - 9.38) | 9.90 (6.81 - 13.9) |
| Meta-analysis | 278,485 | 855,424 | 169.2 | 658.3 | 2,911 | 14,506 | 17.2 (16.6 - 17.8) | 22.0 (21.7 - 22.4) |
| Meta-analysis (w/o USOC) | 42,565 | 130,150 | 20.3 | 75.8 | 592 | 2,740 | 29.1 (26.8 - 31.5) | 36.2 (34.8 - 37.5) |

#### 4.4 GLP1RA (female) vs DPP4I (female)

##### 4.4.1 3-point MACE

**Supplementary Table 364:** Exposures, outcomes and risk estimates for 3-point MACE in GLP1RA (female) (T) vs DPP4I (female) (C) new-user cohorts under four different propensity score (PS) adjustment / time-at-risk (TAR) designs across data sources. We report total patient exposure time in 1,000 patient-year (KYr) units, uncalibrated and calibrated hazard ratios (HRs) and their 95% confidence intervals (CIs). Greyed out entries did not pass study diagnostics and are included only for completeness.

| | Patients | | Exposure (KYr) | | Outcomes | | Incidence (per KYr) | |
|---|---|---|---|---|---|---|---|---|
| | T | C | T | C | T | C | T | C |
| **PS-matching / on-treatment TAR** | | | | | | | | |
| CCAE | 14,149 | 44,506 | 8.9 | 35.1 | 45 | 203 | 5.05 (3.69 - 6.76) | 5.78 (5.01 - 6.63) |
| ODOD | 7,703 | 27,560 | 4.5 | 21.7 | 52 | 366 | 11.6 (8.70 - 15.3) | 16.9 (15.2 - 18.7) |
| MDCR | 941 | 7,585 | 0.6 | 7.2 | 6 | 157 | 9.75 (3.58 - 21.2) | 21.9 (18.6 - 25.6) |
| MDCD | 1,771 | 7,241 | 1.0 | 5.1 | 9 | 101 | 9.21 (4.21 - 17.5) | 19.6 (16.0 - 23.8) |
| Open Claims | 139,429 | 427,400 | 101.8 | 373.9 | 532 | 3,909 | 5.23 (4.79 - 5.69) | 10.5 (10.1 - 10.8) |
| OEHR | 10,417 | 38,070 | 3.2 | 19.7 | 22 | 155 | 6.85 (4.30 - 10.4) | 7.89 (6.69 - 9.23) |
| Meta-analysis | 173,469 | 544,777 | 119.3 | 455.6 | 660 | 4,734 | 5.53 (5.12 - 5.97) | 10.4 (10.1 - 10.7) |
| Meta-analysis (w/o USOC) | 34,040 | 117,377 | 17.6 | 81.6 | 128 | 825 | 7.29 (6.08 - 8.67) | 10.1 (9.43 - 10.8) |
| **PS-stratification / on-treatment TAR** | | | | | | | | |
| CCAE | 19,084 | 44,510 | 11.3 | 35.1 | 54 | 203 | 4.79 (3.60 - 6.26) | 5.78 (5.01 - 6.63) |
| ODOD | 11,356 | 28,169 | 6.0 | 22.3 | 66 | 400 | 11.0 (8.51 - 14.0) | 18.0 (16.2 - 19.8) |
| MDCR | 1,075 | 8,124 | 0.7 | 7.6 | 6 | 182 | 9.18 (3.37 - 20.0) | 23.8 (20.5 - 27.6) |
| MDCD | 1,943 | 7,282 | 1.0 | 5.2 | 9 | 102 | 8.58 (3.92 - 16.3) | 19.7 (16.1 - 23.9) |
| Open Claims | 223,507 | 428,126 | 139.3 | 374.7 | 676 | 3,943 | 4.85 (4.49 - 5.23) | 10.5 (10.2 - 10.9) |
| OEHR | 13,452 | 38,260 | 4.0 | 19.8 | 25 | 161 | 6.32 (4.09 - 9.33) | 8.15 (6.94 - 9.51) |
| Meta-analysis | 269,342 | 546,347 | 161.6 | 457.0 | 830 | 4,809 | 5.14 (4.79 - 5.50) | 10.5 (10.2 - 10.8) |
| Meta-analysis (w/o USOC) | 45,835 | 118,221 | 22.3 | 82.4 | 154 | 866 | 6.92 (5.87 - 8.10) | 10.5 (9.83 - 11.2) |
| **PS-matching / on-treatment TAR without escalation** | | | | | | | | |
| CCAE | 14,149 | 44,506 | 8.0 | 28.9 | 36 | 158 | 4.51 (3.16 - 6.24) | 5.46 (4.64 - 6.38) |
| ODOD | 7,703 | 27,560 | 4.0 | 18.2 | 45 | 322 | 11.2 (8.17 - 15.0) | 17.7 (15.8 - 19.7) |
| MDCR | 941 | 7,585 | 0.6 | 6.2 | 6 | 124 | 10.6 (3.89 - 23.0) | 19.9 (16.5 - 23.7) |
| MDCD | 1,771 | 7,241 | 0.8 | 4.0 | 8 | 77 | 9.55 (4.12 - 18.8) | 19.2 (15.1 - 23.9) |
| Open Claims | 139,429 | 427,400 | 90.2 | 297.0 | 455 | 3,167 | 5.05 (4.59 - 5.53) | 10.7 (10.3 - 11.0) |
| OEHR | 10,417 | 38,070 | 2.8 | 15.8 | 17 | 99 | 6.15 (3.58 - 9.85) | 6.25 (5.08 - 7.61) |
| Meta-analysis | 173,469 | 544,777 | 105.8 | 364.0 | 561 | 3,823 | 5.30 (4.87 - 5.76) | 10.5 (10.2 - 10.8) |
| Meta-analysis (w/o USOC) | 34,040 | 117,377 | 15.6 | 67.0 | 106 | 656 | 6.79 (5.56 - 8.21) | 9.79 (9.06 - 10.6) |
| **PS-stratification / on-treatment TAR without escalation** | | | | | | | | |
| CCAE | 19,084 | 44,510 | 10.2 | 28.9 | 43 | 158 | 4.22 (3.05 - 5.68) | 5.46 (4.64 - 6.38) |
| ODOD | 11,356 | 28,169 | 5.5 | 18.7 | 59 | 353 | 10.8 (8.22 - 13.9) | 18.9 (17.0 - 21.0) |
| MDCR | 1,075 | 8,124 | 0.6 | 6.6 | 6 | 147 | 9.93 (3.64 - 21.6) | 22.1 (18.7 - 26.0) |
| MDCD | 1,943 | 7,282 | 0.9 | 4.0 | 8 | 78 | 8.85 (3.82 - 17.4) | 19.3 (15.2 - 24.1) |
| Open Claims | 223,507 | 428,126 | 125.7 | 297.6 | 596 | 3,196 | 4.74 (4.37 - 5.14) | 10.7 (10.4 - 11.1) |
| OEHR | 13,452 | 38,260 | 3.4 | 15.9 | 20 | 104 | 5.81 (3.55 - 8.98) | 6.53 (5.33 - 7.91) |
| Meta-analysis | 269,342 | 546,347 | 145.7 | 365.2 | 726 | 3,889 | 4.98 (4.63 - 5.36) | 10.6 (10.3 - 11.0) |
| Meta-analysis (w/o USOC) | 45,835 | 118,221 | 20.0 | 67.6 | 130 | 693 | 6.50 (5.43 - 7.72) | 10.3 (9.50 - 11.0) |

##### 4.4.2 4-point MACE

**Supplementary Table 365:** Exposures, outcomes and risk estimates for 4-point MACE in GLP1RA (female) (T) vs DPP4I (female) (C) new-user cohorts under four different propensity score (PS) adjustment / time-at-risk (TAR) designs across data sources. We report total patient exposure time in 1,000 patient-year (KYr) units, uncalibrated and calibrated hazard ratios (HRs) and their 95% confidence intervals (CIs). Greyed out entries did not pass study diagnostics and are included only for completeness.

| | Patients | | Exposure (KYr) | | Outcomes | | Incidence (per KYr) | |
|---|---|---|---|---|---|---|---|---|
| | T | C | T | C | T | C | T | C |
| **PS-matching / on-treatment TAR** | | | | | | | | |
| CCAE | 13,948 | 43,925 | 8.8 | 34.7 | 55 | 329 | 6.26 (4.72 - 8.15) | 9.48 (8.49 - 10.6) |
| ODOD | 7,470 | 26,516 | 4.3 | 20.7 | 92 | 635 | 21.3 (17.2 - 26.2) | 30.7 (28.3 - 33.1) |
| MDCR | 900 | 7,079 | 0.6 | 6.7 | 11 | 248 | 18.3 (9.15 - 32.8) | 37.0 (32.5 - 41.9) |
| MDCD | 1,655 | 6,752 | 0.9 | 4.7 | 17 | 180 | 18.7 (10.9 - 30.0) | 37.9 (32.6 - 43.9) |
| Open Claims | 135,919 | 413,784 | 99.2 | 360.7 | 820 | 6,457 | 8.26 (7.71 - 8.85) | 17.9 (17.5 - 18.3) |
| OEHR | 10,256 | 37,505 | 3.2 | 19.2 | 37 | 316 | 11.7 (8.25 - 16.1) | 16.4 (14.7 - 18.4) |
| Meta-analysis | 169,248 | 528,482 | 116.4 | 440.1 | 1,021 | 7,917 | 8.77 (8.24 - 9.33) | 18.0 (17.6 - 18.4) |
| Meta-analysis (w/o USOC) | 33,329 | 114,698 | 17.2 | 79.4 | 201 | 1,460 | 11.7 (10.2 - 13.5) | 18.4 (17.5 - 19.4) |
| **PS-stratification / on-treatment TAR** | | | | | | | | |
| CCAE | 18,823 | 43,928 | 11.1 | 34.7 | 65 | 329 | 5.85 (4.52 - 7.46) | 9.48 (8.49 - 10.6) |
| ODOD | 11,041 | 27,017 | 5.8 | 21.2 | 109 | 680 | 18.7 (15.4 - 22.6) | 32.1 (29.7 - 34.6) |
| MDCR | 1,026 | 7,581 | 0.6 | 7.1 | 11 | 286 | 17.3 (8.63 - 30.9) | 40.1 (35.6 - 45.0) |
| MDCD | 1,819 | 6,786 | 1.0 | 4.8 | 18 | 181 | 18.4 (10.9 - 29.1) | 37.9 (32.6 - 43.9) |
| Open Claims | 218,502 | 414,108 | 136.1 | 361.1 | 1,041 | 6,491 | 7.65 (7.19 - 8.13) | 18.0 (17.5 - 18.4) |
| OEHR | 13,257 | 37,672 | 3.9 | 19.3 | 41 | 325 | 10.5 (7.56 - 14.3) | 16.8 (15.1 - 18.8) |
| Meta-analysis | 263,442 | 529,511 | 157.9 | 441.0 | 1,274 | 8,006 | 8.07 (7.63 - 8.52) | 18.2 (17.8 - 18.6) |
| Meta-analysis (w/o USOC) | 44,940 | 115,403 | 21.8 | 80.0 | 233 | 1,515 | 10.7 (9.36 - 12.2) | 18.9 (18.0 - 19.9) |
| **PS-matching / on-treatment TAR without escalation** | | | | | | | | |
| CCAE | 13,948 | 43,925 | 7.9 | 28.6 | 44 | 264 | 5.58 (4.06 - 7.50) | 9.25 (8.16 - 10.4) |
| ODOD | 7,470 | 26,516 | 3.9 | 17.4 | 80 | 537 | 20.6 (16.4 - 25.7) | 30.9 (28.3 - 33.6) |
| MDCR | 900 | 7,079 | 0.6 | 5.8 | 11 | 205 | 19.9 (9.94 - 35.6) | 35.2 (30.5 - 40.3) |
| MDCD | 1,655 | 6,752 | 0.8 | 3.7 | 14 | 137 | 18.0 (9.86 - 30.3) | 36.9 (30.9 - 43.6) |
| Open Claims | 135,919 | 413,784 | 88.0 | 286.7 | 695 | 5,190 | 7.90 (7.32 - 8.51) | 18.1 (17.6 - 18.6) |
| OEHR | 10,256 | 37,505 | 2.7 | 15.5 | 29 | 215 | 10.6 (7.12 - 15.3) | 13.8 (12.0 - 15.8) |
| Meta-analysis | 169,248 | 528,482 | 103.2 | 351.9 | 862 | 6,343 | 8.35 (7.80 - 8.93) | 18.0 (17.6 - 18.5) |
| Meta-analysis (w/o USOC) | 33,329 | 114,698 | 15.3 | 65.2 | 167 | 1,153 | 10.9 (9.34 - 12.7) | 17.7 (16.7 - 18.7) |
| **PS-stratification / on-treatment TAR without escalation** | | | | | | | | |
| CCAE | 18,823 | 43,928 | 10.1 | 28.6 | 51 | 264 | 5.07 (3.78 - 6.67) | 9.24 (8.16 - 10.4) |
| ODOD | 11,041 | 27,017 | 5.3 | 17.8 | 97 | 574 | 18.3 (14.8 - 22.3) | 32.2 (29.6 - 35.0) |
| MDCR | 1,026 | 7,581 | 0.6 | 6.2 | 11 | 240 | 18.7 (9.34 - 33.5) | 38.6 (33.9 - 43.8) |
| MDCD | 1,819 | 6,786 | 0.8 | 3.7 | 15 | 138 | 17.9 (10.0 - 29.5) | 36.9 (31.0 - 43.6) |
| Open Claims | 218,502 | 414,108 | 123.0 | 287.0 | 905 | 5,220 | 7.36 (6.89 - 7.86) | 18.2 (17.7 - 18.7) |
| OEHR | 13,257 | 37,672 | 3.4 | 15.6 | 33 | 222 | 9.73 (6.70 - 13.7) | 14.2 (12.4 - 16.2) |
| Meta-analysis | 263,442 | 529,511 | 142.5 | 352.7 | 1,101 | 6,418 | 7.72 (7.27 - 8.19) | 18.2 (17.8 - 18.6) |
| Meta-analysis (w/o USOC) | 44,940 | 115,403 | 19.6 | 65.7 | 196 | 1,198 | 10.0 (8.65 - 11.5) | 18.2 (17.2 - 19.3) |

##### 4.4.3 Acute myocardial infarction

**Supplementary Table 366:** Exposures, outcomes and risk estimates for Acute myocardial infarction in GLP1RA (female) (T) vs DPP4I (female) (C) new-user cohorts under four different propensity score (PS) adjustment / time-at-risk (TAR) designs across data sources. We report total patient exposure time in 1,000 patient-year (KYr) units, uncalibrated and calibrated hazard ratios (HRs) and their 95% confidence intervals (CIs). Greyed out entries did not pass study diagnostics and are included only for completeness.

| | Patients | | Exposure (KYr) | | Outcomes | | Incidence (per KYr) | |
|---|---|---|---|---|---|---|---|---|
| | T | C | T | C | T | C | T | C |
| **PS-matching / on-treatment TAR** | | | | | | | | |
| CCAE | 14,309 | 45,022 | 9.0 | 35.5 | 22 | 85 | 2.44 (1.53 - 3.69) | 2.39 (1.91 - 2.96) |
| ODOD | 7,825 | 28,313 | 4.6 | 22.4 | 24 | 126 | 5.25 (3.37 - 7.81) | 5.63 (4.69 - 6.70) |
| MDCR | 972 | 8,213 | 0.6 | 7.8 | <5 | 67 | < 7.91 (2.57 - 18.5) | 8.60 (6.66 - 10.9) |
| MDCD | 1,822 | 7,496 | 1.0 | 5.3 | <5 | 32 | < 5.00 (1.62 - 11.7) | 5.98 (4.09 - 8.45) |
| Open Claims | 142,472 | 439,605 | 103.9 | 385.4 | 226 | 1,658 | 2.18 (1.90 - 2.48) | 4.30 (4.10 - 4.51) |
| OEHR | 10,487 | 38,386 | 3.2 | 19.9 | 12 | 69 | 3.70 (1.91 - 6.46) | 3.47 (2.70 - 4.39) |
| Meta-analysis | 176,915 | 558,822 | 121.7 | 468.6 | <289 | 1,970 | < 2.37 (2.11 - 2.66) | 4.20 (4.02 - 4.39) |
| Meta-analysis (w/o USOC) | 34,443 | 119,217 | 17.8 | 83.2 | <63 | 312 | < 3.53 (2.71 - 4.52) | 3.75 (3.35 - 4.19) |
| **PS-stratification / on-treatment TAR** | | | | | | | | |
| CCAE | 19,305 | 45,027 | 11.4 | 35.5 | 25 | 85 | 2.19 (1.42 - 3.24) | 2.39 (1.91 - 2.96) |
| ODOD | 11,516 | 28,975 | 6.1 | 23.0 | 30 | 147 | 4.90 (3.30 - 6.99) | 6.39 (5.40 - 7.51) |
| MDCR | 1,111 | 8,520 | 0.7 | 8.0 | <5 | 74 | < 7.45 (2.42 - 17.4) | 9.22 (7.24 - 11.6) |
| MDCD | 1,994 | 7,556 | 1.1 | 5.4 | <5 | 33 | < 4.66 (1.51 - 10.9) | 6.11 (4.20 - 8.58) |
| Open Claims | 227,850 | 440,479 | 142.0 | 386.3 | 281 | 1,670 | 1.98 (1.75 - 2.22) | 4.32 (4.12 - 4.54) |
| OEHR | 13,546 | 38,583 | 4.0 | 20.0 | 15 | 69 | 3.75 (2.10 - 6.19) | 3.45 (2.68 - 4.37) |
| Meta-analysis | 274,211 | 560,620 | 164.6 | 470.3 | <356 | 2,004 | < 2.16 (1.94 - 2.40) | 4.26 (4.08 - 4.45) |
| Meta-analysis (w/o USOC) | 46,361 | 120,141 | 22.6 | 84.0 | <75 | 334 | < 3.32 (2.61 - 4.16) | 3.98 (3.56 - 4.43) |
| **PS-matching / on-treatment TAR without escalation** | | | | | | | | |
| CCAE | 14,309 | 45,022 | 8.1 | 29.2 | 19 | 58 | 2.35 (1.41 - 3.67) | 1.98 (1.51 - 2.56) |
| ODOD | 7,825 | 28,313 | 4.1 | 18.8 | 20 | 113 | 4.87 (2.98 - 7.53) | 6.02 (4.96 - 7.23) |
| MDCR | 972 | 8,213 | 0.6 | 6.8 | <5 | 58 | < 8.60 (2.79 - 20.1) | 8.58 (6.51 - 11.1) |
| MDCD | 1,822 | 7,496 | 0.9 | 4.2 | <5 | 23 | < 5.82 (1.89 - 13.6) | 5.53 (3.50 - 8.29) |
| Open Claims | 142,472 | 439,605 | 92.0 | 306.1 | 196 | 1,336 | 2.13 (1.84 - 2.45) | 4.36 (4.13 - 4.60) |
| OEHR | 10,487 | 38,386 | 2.8 | 16.0 | 9 | 42 | 3.22 (1.47 - 6.12) | 2.62 (1.89 - 3.54) |
| Meta-analysis | 176,915 | 558,822 | 107.9 | 374.3 | <249 | 1,572 | < 2.31 (2.03 - 2.61) | 4.20 (3.99 - 4.41) |
| Meta-analysis (w/o USOC) | 34,443 | 119,217 | 15.8 | 68.2 | <53 | 236 | < 3.34 (2.51 - 4.37) | 3.46 (3.03 - 3.93) |
| **PS-stratification / on-treatment TAR without escalation** | | | | | | | | |
| CCAE | 19,305 | 45,027 | 10.3 | 29.2 | 22 | 58 | 2.13 (1.34 - 3.23) | 1.98 (1.51 - 2.56) |
| ODOD | 11,516 | 28,975 | 5.6 | 19.3 | 26 | 132 | 4.67 (3.05 - 6.84) | 6.84 (5.72 - 8.11) |
| MDCR | 1,111 | 8,520 | 0.6 | 7.0 | <5 | 65 | < 8.07 (2.62 - 18.8) | 9.32 (7.19 - 11.9) |
| MDCD | 1,994 | 7,556 | 0.9 | 4.2 | <5 | 23 | < 5.40 (1.75 - 12.6) | 5.47 (3.47 - 8.21) |
| Open Claims | 227,850 | 440,479 | 128.1 | 306.8 | 251 | 1,346 | 1.96 (1.72 - 2.22) | 4.39 (4.16 - 4.63) |
| OEHR | 13,546 | 38,583 | 3.5 | 16.1 | 12 | 42 | 3.45 (1.78 - 6.03) | 2.61 (1.88 - 3.52) |
| Meta-analysis | 274,211 | 560,620 | 148.4 | 375.7 | <316 | 1,601 | < 2.13 (1.90 - 2.38) | 4.26 (4.06 - 4.48) |
| Meta-analysis (w/o USOC) | 46,361 | 120,141 | 20.3 | 68.9 | <65 | 255 | < 3.20 (2.47 - 4.08) | 3.70 (3.26 - 4.19) |

##### 4.4.4 Acute renal failure

**Supplementary Table 367:** Exposures, outcomes and risk estimates for Acute renal failure in GLP1RA (female) (T) vs DPP4I (female) (C) new-user cohorts under four different propensity score (PS) adjustment / time-at-risk (TAR) designs across data sources. We report total patient exposure time in 1,000 patient-year (KYr) units, uncalibrated and calibrated hazard ratios (HRs) and their 95% confidence intervals (CIs). Greyed out entries did not pass study diagnostics and are included only for completeness.

| | Patients | | Exposure (KYr) | | Outcomes | | Incidence (per KYr) | |
|---|---|---|---|---|---|---|---|---|
| | T | C | T | C | T | C | T | C |
| **PS-matching / on-treatment TAR** | | | | | | | | |
| CCAE | 14,295 | 44,953 | 9.0 | 35.5 | 41 | 225 | 4.55 (3.27 - 6.18) | 6.34 (5.54 - 7.22) |
| ODOD | 7,660 | 27,597 | 4.5 | 21.6 | 61 | 584 | 13.7 (10.5 - 17.6) | 27.0 (24.9 - 29.3) |
| MDCR | 950 | 8,006 | 0.6 | 7.5 | 10 | 211 | 16.2 (7.77 - 29.8) | 28.0 (24.3 - 32.0) |
| MDCD | 1,767 | 7,296 | 1.0 | 5.1 | 16 | 171 | 16.7 (9.53 - 27.1) | 33.7 (28.8 - 39.1) |
| Open Claims | 140,798 | 433,196 | 102.4 | 377.3 | 735 | 5,505 | 7.18 (6.67 - 7.72) | 14.6 (14.2 - 15.0) |
| OEHR | 10,454 | 38,171 | 3.2 | 19.7 | 38 | 259 | 11.8 (8.35 - 16.2) | 13.2 (11.6 - 14.9) |
| Meta-analysis | 174,974 | 551,213 | 120.0 | 459.2 | 891 | 6,744 | 7.42 (6.94 - 7.93) | 14.7 (14.3 - 15.0) |
| Meta-analysis (w/o USOC) | 34,176 | 118,017 | 17.6 | 81.9 | 156 | 1,239 | 8.84 (7.51 - 10.3) | 15.1 (14.3 - 16.0) |
| **PS-stratification / on-treatment TAR** | | | | | | | | |
| CCAE | 19,257 | 44,957 | 11.4 | 35.5 | 59 | 225 | 5.19 (3.95 - 6.69) | 6.34 (5.54 - 7.22) |
| ODOD | 11,351 | 28,091 | 6.0 | 22.1 | 79 | 613 | 13.2 (10.4 - 16.4) | 27.8 (25.6 - 30.1) |
| MDCR | 1,089 | 8,325 | 0.7 | 7.8 | 10 | 238 | 15.2 (7.31 - 28.0) | 30.6 (26.8 - 34.7) |
| MDCD | 1,939 | 7,347 | 1.0 | 5.1 | 16 | 172 | 15.5 (8.88 - 25.2) | 33.6 (28.8 - 39.0) |
| Open Claims | 225,549 | 433,780 | 140.2 | 377.9 | 937 | 5,535 | 6.68 (6.26 - 7.12) | 14.6 (14.3 - 15.0) |
| OEHR | 13,485 | 38,353 | 4.0 | 19.8 | 45 | 267 | 11.3 (8.26 - 15.2) | 13.5 (11.9 - 15.2) |
| Meta-analysis | 271,581 | 552,528 | 162.6 | 460.4 | 1,136 | 6,812 | 6.99 (6.59 - 7.41) | 14.8 (14.4 - 15.2) |
| Meta-analysis (w/o USOC) | 46,032 | 118,748 | 22.4 | 82.4 | 199 | 1,277 | 8.89 (7.70 - 10.2) | 15.5 (14.7 - 16.4) |
| **PS-matching / on-treatment TAR without escalation** | | | | | | | | |
| CCAE | 14,295 | 44,953 | 8.1 | 29.2 | 36 | 177 | 4.46 (3.12 - 6.17) | 6.06 (5.20 - 7.03) |
| ODOD | 7,660 | 27,597 | 4.0 | 18.2 | 52 | 476 | 13.0 (9.71 - 17.0) | 26.2 (23.9 - 28.7) |
| MDCR | 950 | 8,006 | 0.6 | 6.6 | 9 | 177 | 15.8 (7.22 - 30.0) | 26.9 (23.1 - 31.1) |
| MDCD | 1,767 | 7,296 | 0.8 | 4.0 | 14 | 130 | 17.0 (9.29 - 28.5) | 32.6 (27.3 - 38.8) |
| Open Claims | 140,798 | 433,196 | 90.7 | 300.0 | 626 | 4,363 | 6.90 (6.37 - 7.46) | 14.5 (14.1 - 15.0) |
| OEHR | 10,454 | 38,171 | 2.8 | 15.9 | 23 | 178 | 8.28 (5.25 - 12.4) | 11.2 (9.63 - 13.0) |
| Meta-analysis | 174,974 | 551,213 | 106.4 | 367.3 | 751 | 5,324 | 7.06 (6.56 - 7.58) | 14.5 (14.1 - 14.9) |
| Meta-analysis (w/o USOC) | 34,176 | 118,017 | 15.7 | 67.2 | 125 | 961 | 7.97 (6.64 - 9.50) | 14.3 (13.4 - 15.2) |
| **PS-stratification / on-treatment TAR without escalation** | | | | | | | | |
| CCAE | 19,257 | 44,957 | 10.3 | 29.2 | 53 | 177 | 5.15 (3.86 - 6.74) | 6.06 (5.20 - 7.03) |
| ODOD | 11,351 | 28,091 | 5.5 | 18.6 | 69 | 501 | 12.6 (9.83 - 16.0) | 27.0 (24.7 - 29.5) |
| MDCR | 1,089 | 8,325 | 0.6 | 6.8 | 9 | 198 | 14.8 (6.77 - 28.1) | 29.1 (25.2 - 33.4) |
| MDCD | 1,939 | 7,347 | 0.9 | 4.0 | 14 | 131 | 15.8 (8.62 - 26.4) | 32.6 (27.3 - 38.7) |
| Open Claims | 225,549 | 433,780 | 126.5 | 300.5 | 810 | 4,388 | 6.40 (5.97 - 6.86) | 14.6 (14.2 - 15.0) |
| OEHR | 13,485 | 38,353 | 3.5 | 16.0 | 30 | 184 | 8.67 (5.85 - 12.4) | 11.5 (9.93 - 13.3) |
| Meta-analysis | 271,581 | 552,528 | 146.6 | 368.2 | 976 | 5,381 | 6.66 (6.24 - 7.09) | 14.6 (14.2 - 15.0) |
| Meta-analysis (w/o USOC) | 46,032 | 118,748 | 20.1 | 67.7 | 166 | 993 | 8.26 (7.05 - 9.61) | 14.7 (13.8 - 15.6) |

##### 4.4.5 Glycemic control

**Supplementary Table 368:** Exposures, outcomes and risk estimates for Glycemic control in GLP1RA (female) (T) vs DPP4I (female) (C) new-user cohorts under four different propensity score (PS) adjustment / time-at-risk (TAR) designs across data sources. We report total patient exposure time in 1,000 patient-year (KYr) units, uncalibrated and calibrated hazard ratios (HRs) and their 95% confidence intervals (CIs). Greyed out entries did not pass study diagnostics and are included only for completeness.

| | Patients | | Exposure (KYr) | | Outcomes | | Incidence (per KYr) | |
|---|---|---|---|---|---|---|---|---|
| | T | C | T | C | T | C | T | C |
| **PS-matching / on-treatment TAR** | | | | | | | | |
| CCAE | 14,306 | 44,884 | 9.0 | 35.5 | 18 | 111 | 2.00 (1.18 - 3.15) | 3.13 (2.57 - 3.77) |
| ODOD | 5,103 | 18,673 | 2.8 | 13.2 | 316 | 1,420 | 112 (100 - 126) | 108 (102 - 113) |
| MDCR | 981 | 8,392 | 0.6 | 8.0 | 0 | 15 | | 1.88 (1.05 - 3.10) |
| MDCD | 1,861 | 7,692 | 1.0 | 5.5 | 0 | 0 | | |
| Open Claims | 144,854 | 448,754 | 105.6 | 394.3 | 0 | 0 | | |
| OEHR | 10,563 | 38,693 | 3.3 | 20.1 | 0 | 0 | | |
| Meta-analysis | 19,409 | 63,557 | 11.8 | 48.7 | 334 | 1,531 | 28.2 (25.3 - 31.4) | 31.4 (29.9 - 33.1) |
| Meta-analysis (w/o USOC) | 19,409 | 63,557 | 11.8 | 48.7 | 334 | 1,531 | 28.2 (25.3 - 31.4) | 31.4 (29.9 - 33.1) |
| **PS-stratification / on-treatment TAR** | | | | | | | | |
| CCAE | 19,314 | 44,889 | 11.4 | 35.5 | 19 | 111 | 1.67 (1.00 - 2.60) | 3.13 (2.57 - 3.76) |
| ODOD | 7,236 | 19,175 | 3.6 | 13.6 | 400 | 1,468 | 110 (99.9 - 122) | 108 (103 - 114) |
| MDCR | 1,124 | 8,751 | 0.7 | 8.3 | 0 | 15 | | 1.82 (1.02 - 2.99) |
| MDCD | 2,034 | 7,771 | 1.1 | 5.6 | 0 | 0 | | |
| Open Claims | 231,281 | 449,866 | 144.1 | 395.4 | 0 | 0 | | |
| OEHR | 13,638 | 38,897 | 4.0 | 20.2 | 0 | 0 | | |
| Meta-analysis | 26,550 | 64,064 | 15.0 | 49.1 | 419 | 1,579 | 27.9 (25.3 - 30.7) | 32.2 (30.6 - 33.8) |
| Meta-analysis (w/o USOC) | 26,550 | 64,064 | 15.0 | 49.1 | 419 | 1,579 | 27.9 (25.3 - 30.7) | 32.2 (30.6 - 33.8) |
| **PS-matching / on-treatment TAR without escalation** | | | | | | | | |
| CCAE | 14,306 | 44,884 | 8.1 | 29.2 | 18 | 97 | 2.23 (1.32 - 3.52) | 3.33 (2.70 - 4.06) |
| ODOD | 5,103 | 18,673 | 2.5 | 11.1 | 288 | 1,187 | 114 (102 - 128) | 107 (101 - 114) |
| MDCR | 981 | 8,392 | 0.6 | 6.9 | 0 | 12 | | 1.73 (0.89 - 3.02) |
| MDCD | 1,861 | 7,692 | 0.9 | 4.3 | 0 | 0 | | |
| Open Claims | 144,854 | 448,754 | 93.5 | 313.0 | 0 | 0 | | |
| OEHR | 10,563 | 38,693 | 2.8 | 16.2 | 0 | 0 | | |
| Meta-analysis | 19,409 | 63,557 | 10.6 | 40.2 | 306 | 1,284 | 28.9 (25.7 - 32.3) | 31.9 (30.2 - 33.7) |
| Meta-analysis (w/o USOC) | 19,409 | 63,557 | 10.6 | 40.2 | 306 | 1,284 | 28.9 (25.7 - 32.3) | 31.9 (30.2 - 33.7) |
| **PS-stratification / on-treatment TAR without escalation** | | | | | | | | |
| CCAE | 19,314 | 44,889 | 10.3 | 29.2 | 19 | 97 | 1.84 (1.11 - 2.88) | 3.33 (2.70 - 4.06) |
| ODOD | 7,236 | 19,175 | 3.3 | 11.4 | 365 | 1,230 | 112 (100 - 124) | 108 (102 - 114) |
| MDCR | 1,124 | 8,751 | 0.6 | 7.2 | 0 | 12 | | 1.67 (0.86 - 2.92) |
| MDCD | 2,034 | 7,771 | 0.9 | 4.3 | 0 | 0 | | |
| Open Claims | 231,281 | 449,866 | 130.0 | 313.9 | 0 | 0 | | |
| OEHR | 13,638 | 38,897 | 3.5 | 16.3 | 0 | 0 | | |
| Meta-analysis | 26,550 | 64,064 | 13.6 | 40.6 | 384 | 1,327 | 28.3 (25.5 - 31.2) | 32.7 (31.0 - 34.5) |
| Meta-analysis (w/o USOC) | 26,550 | 64,064 | 13.6 | 40.6 | 384 | 1,327 | 28.3 (25.5 - 31.2) | 32.7 (31.0 - 34.5) |

##### 4.4.6 Hospitalization with heart failure

**Supplementary Table 369:** Exposures, outcomes and risk estimates for Hospitalization with heart failure in GLP1RA (female) (T) vs DPP4I (female) (C) new-user cohorts under four different propensity score (PS) adjustment / time-at-risk (TAR) designs across data sources. We report total patient exposure time in 1,000 patient-year (KYr) units, uncalibrated and calibrated hazard ratios (HRs) and their 95% confidence intervals (CIs). Greyed out entries did not pass study diagnostics and are included only for completeness.

| | Patients | | Exposure (KYr) | | Outcomes | | Incidence (per KYr) | |
|---|---|---|---|---|---|---|---|---|
| | T | C | T | C | T | C | T | C |
| **PS-matching / on-treatment TAR** | | | | | | | | |
| CCAE | 14,194 | 44,751 | 9.0 | 35.4 | 15 | 177 | 1.67 (0.94 - 2.76) | 5.00 (4.29 - 5.79) |
| ODOD | 7,628 | 27,438 | 4.4 | 21.5 | 59 | 451 | 13.3 (10.1 - 17.2) | 21.0 (19.1 - 23.0) |
| MDCR | 932 | 7,765 | 0.6 | 7.4 | 9 | 187 | 14.6 (6.70 - 27.8) | 25.3 (21.8 - 29.2) |
| MDCD | 1,715 | 7,075 | 0.9 | 5.0 | 15 | 124 | 16.0 (8.95 - 26.4) | 24.8 (20.6 - 29.6) |
| Open Claims | 140,316 | 430,165 | 102.3 | 376.1 | 479 | 4,266 | 4.68 (4.27 - 5.12) | 11.3 (11.0 - 11.7) |
| OEHR | 10,378 | 38,019 | 3.2 | 19.6 | 21 | 234 | 6.55 (4.06 - 10.0) | 12.0 (10.5 - 13.6) |
| Meta-analysis | 174,231 | 547,448 | 119.8 | 457.6 | 589 | 5,252 | 4.92 (4.53 - 5.33) | 11.5 (11.2 - 11.8) |
| Meta-analysis (w/o USOC) | 33,915 | 117,283 | 17.5 | 81.5 | 110 | 986 | 6.27 (5.15 - 7.56) | 12.1 (11.4 - 12.9) |
| **PS-stratification / on-treatment TAR** | | | | | | | | |
| CCAE | 19,133 | 44,755 | 11.3 | 35.4 | 18 | 177 | 1.59 (0.94 - 2.51) | 5.00 (4.29 - 5.79) |
| ODOD | 11,259 | 27,920 | 6.0 | 22.0 | 66 | 482 | 11.1 (8.55 - 14.1) | 21.9 (20.0 - 24.0) |
| MDCR | 1,067 | 8,032 | 0.7 | 7.6 | 9 | 203 | 13.8 (6.30 - 26.2) | 26.8 (23.2 - 30.7) |
| MDCD | 1,878 | 7,124 | 1.0 | 5.0 | 16 | 125 | 15.9 (9.07 - 25.8) | 24.8 (20.7 - 29.6) |
| Open Claims | 224,856 | 430,696 | 140.1 | 376.6 | 587 | 4,304 | 4.19 (3.86 - 4.54) | 11.4 (11.1 - 11.8) |
| OEHR | 13,410 | 38,193 | 4.0 | 19.7 | 25 | 241 | 6.33 (4.09 - 9.34) | 12.3 (10.8 - 13.9) |
| Meta-analysis | 270,536 | 548,688 | 162.3 | 458.7 | 712 | 5,329 | 4.39 (4.07 - 4.72) | 11.6 (11.3 - 11.9) |
| Meta-analysis (w/o USOC) | 45,680 | 117,992 | 22.2 | 82.1 | 125 | 1,025 | 5.62 (4.68 - 6.70) | 12.5 (11.7 - 13.3) |
| **PS-matching / on-treatment TAR without escalation** | | | | | | | | |
| CCAE | 14,194 | 44,751 | 8.0 | 29.1 | 10 | 148 | 1.24 (0.60 - 2.29) | 5.08 (4.30 - 5.97) |
| ODOD | 7,628 | 27,438 | 4.0 | 18.1 | 51 | 370 | 12.8 (9.53 - 16.8) | 20.5 (18.4 - 22.7) |
| MDCR | 932 | 7,765 | 0.6 | 6.4 | 8 | 159 | 14.1 (6.10 - 27.8) | 24.8 (21.1 - 29.0) |
| MDCD | 1,715 | 7,075 | 0.8 | 3.9 | 12 | 93 | 14.9 (7.71 - 26.1) | 23.8 (19.2 - 29.1) |
| Open Claims | 140,316 | 430,165 | 90.7 | 298.9 | 409 | 3,403 | 4.51 (4.08 - 4.97) | 11.4 (11.0 - 11.8) |
| OEHR | 10,378 | 38,019 | 2.8 | 15.8 | 16 | 166 | 5.78 (3.30 - 9.39) | 10.5 (8.96 - 12.2) |
| Meta-analysis | 174,231 | 547,448 | 106.3 | 365.8 | 498 | 4,180 | 4.69 (4.28 - 5.12) | 11.4 (11.1 - 11.8) |
| Meta-analysis (w/o USOC) | 33,915 | 117,283 | 15.6 | 66.9 | 89 | 777 | 5.70 (4.58 - 7.02) | 11.6 (10.8 - 12.5) |
| **PS-stratification / on-treatment TAR without escalation** | | | | | | | | |
| CCAE | 19,133 | 44,755 | 10.2 | 29.1 | 12 | 148 | 1.17 (0.61 - 2.05) | 5.08 (4.30 - 5.97) |
| ODOD | 11,259 | 27,920 | 5.4 | 18.5 | 58 | 394 | 10.7 (8.11 - 13.8) | 21.3 (19.3 - 23.6) |
| MDCR | 1,067 | 8,032 | 0.6 | 6.6 | 8 | 175 | 13.2 (5.72 - 26.1) | 26.6 (22.8 - 30.8) |
| MDCD | 1,878 | 7,124 | 0.9 | 3.9 | 13 | 93 | 15.0 (7.97 - 25.6) | 23.6 (19.1 - 28.9) |
| Open Claims | 224,856 | 430,696 | 126.4 | 299.3 | 507 | 3,435 | 4.01 (3.67 - 4.37) | 11.5 (11.1 - 11.9) |
| OEHR | 13,410 | 38,193 | 3.4 | 15.9 | 20 | 171 | 5.81 (3.55 - 8.97) | 10.8 (9.21 - 12.5) |
| Meta-analysis | 270,536 | 548,688 | 146.4 | 366.7 | 610 | 4,241 | 4.17 (3.84 - 4.51) | 11.6 (11.2 - 11.9) |
| Meta-analysis (w/o USOC) | 45,680 | 117,992 | 20.0 | 67.4 | 103 | 806 | 5.15 (4.21 - 6.25) | 12.0 (11.1 - 12.8) |

##### 4.4.7 Measured renal dysfunction

**Supplementary Table 370:** Exposures, outcomes and risk estimates for Measured renal dysfunction in GLP1RA (female) (T) vs DPP4I (female) (C) new-user cohorts under four different propensity score (PS) adjustment / time-at-risk (TAR) designs across data sources. We report total patient exposure time in 1,000 patient-year (KYr) units, uncalibrated and calibrated hazard ratios (HRs) and their 95% confidence intervals (CIs). Greyed out entries did not pass study diagnostics and are included only for completeness.

| | Patients | | Exposure (KYr) | | Outcomes | | Incidence (per KYr) | |
|---|---|---|---|---|---|---|---|---|
| | T | C | T | C | T | C | T | C |
| **PS-matching / on-treatment TAR** | | | | | | | | |
| CCAE | 14,416 | 45,460 | 9.1 | 36.0 | <5 | 7 | < 0.55 (0.18 - 1.28) | 0.19 (0.08 - 0.40) |
| ODOD | 7,909 | 28,772 | 4.6 | 22.8 | <5 | 46 | < 1.08 (0.35 - 2.53) | 2.02 (1.48 - 2.69) |
| MDCR | 986 | 8,446 | 0.6 | 8.0 | 0 | 0 | | |
| MDCD | 1,861 | 7,692 | 1.0 | 5.5 | 0 | 0 | | |
| Open Claims | 144,854 | 448,754 | 105.6 | 394.3 | 0 | 0 | | |
| OEHR | 10,563 | 38,693 | 3.3 | 20.1 | 0 | 0 | | |
| Meta-analysis | 7,909 | 28,772 | 4.6 | 22.8 | <5 | 46 | < 1.08 (0.35 - 2.53) | 2.02 (1.48 - 2.69) |
| Meta-analysis (w/o USOC) | 7,909 | 28,772 | 4.6 | 22.8 | <5 | 46 | < 1.08 (0.35 - 2.53) | 2.02 (1.48 - 2.69) |
| **PS-stratification / on-treatment TAR** | | | | | | | | |
| CCAE | 19,418 | 45,465 | 11.5 | 36.0 | <5 | 7 | < 0.44 (0.14 - 1.02) | 0.19 (0.08 - 0.40) |
| ODOD | 11,650 | 29,464 | 6.2 | 23.4 | <5 | 55 | < 0.81 (0.26 - 1.88) | 2.35 (1.77 - 3.06) |
| MDCR | 1,128 | 8,813 | 0.7 | 8.3 | 0 | 0 | | |
| MDCD | 2,034 | 7,771 | 1.1 | 5.6 | 0 | 0 | | |
| Open Claims | 231,281 | 449,866 | 144.1 | 395.4 | 0 | 0 | | |
| OEHR | 13,638 | 38,897 | 4.0 | 20.2 | 0 | 0 | | |
| Meta-analysis | 11,650 | 29,464 | 6.2 | 23.4 | <5 | 55 | < 0.81 (0.26 - 1.88) | 2.35 (1.77 - 3.06) |
| Meta-analysis (w/o USOC) | 11,650 | 29,464 | 6.2 | 23.4 | <5 | 55 | < 0.81 (0.26 - 1.88) | 2.35 (1.77 - 3.06) |
| **PS-matching / on-treatment TAR without escalation** | | | | | | | | |
| CCAE | 14,416 | 45,460 | 8.2 | 29.6 | <5 | 6 | < 0.61 (0.20 - 1.43) | 0.20 (0.07 - 0.44) |
| ODOD | 7,909 | 28,772 | 4.1 | 19.1 | <5 | 37 | < 1.21 (0.39 - 2.82) | 1.94 (1.36 - 2.67) |
| MDCR | 986 | 8,446 | 0.6 | 7.0 | 0 | 0 | | |
| MDCD | 1,861 | 7,692 | 0.9 | 4.3 | 0 | 0 | | |
| Open Claims | 144,854 | 448,754 | 93.5 | 313.0 | 0 | 0 | | |
| OEHR | 10,563 | 38,693 | 2.8 | 16.2 | 0 | 0 | | |
| Meta-analysis | 7,909 | 28,772 | 4.1 | 19.1 | <5 | 37 | < 1.21 (0.39 - 2.82) | 1.94 (1.36 - 2.67) |
| Meta-analysis (w/o USOC) | 7,909 | 28,772 | 4.1 | 19.1 | <5 | 37 | < 1.21 (0.39 - 2.82) | 1.94 (1.36 - 2.67) |
| **PS-stratification / on-treatment TAR without escalation** | | | | | | | | |
| CCAE | 19,418 | 45,465 | 10.4 | 29.6 | <5 | 6 | < 0.48 (0.16 - 1.12) | 0.20 (0.07 - 0.44) |
| ODOD | 11,650 | 29,464 | 5.6 | 19.6 | <5 | 46 | < 0.89 (0.29 - 2.07) | 2.34 (1.72 - 3.13) |
| MDCR | 1,128 | 8,813 | 0.6 | 7.2 | 0 | 0 | | |
| MDCD | 2,034 | 7,771 | 0.9 | 4.3 | 0 | 0 | | |
| Open Claims | 231,281 | 449,866 | 130.0 | 313.9 | 0 | 0 | | |
| OEHR | 13,638 | 38,897 | 3.5 | 16.3 | 0 | 0 | | |
| Meta-analysis | 11,650 | 29,464 | 5.6 | 19.6 | <5 | 46 | < 0.89 (0.29 - 2.07) | 2.34 (1.72 - 3.13) |
| Meta-analysis (w/o USOC) | 11,650 | 29,464 | 5.6 | 19.6 | <5 | 46 | < 0.89 (0.29 - 2.07) | 2.34 (1.72 - 3.13) |

##### 4.4.8 Stroke

**Supplementary Table 371:** Exposures, outcomes and risk estimates for Stroke in GLP1RA (female) (T) vs DPP4I (female) (C) new-user cohorts under four different propensity score (PS) adjustment / time-at-risk (TAR) designs across data sources. We report total patient exposure time in 1,000 patient-year (KYr) units, uncalibrated and calibrated hazard ratios (HRs) and their 95% confidence intervals (CIs). Greyed out entries did not pass study diagnostics and are included only for completeness.

| | Patients | | Exposure (KYr) | | Outcomes | | Incidence (per KYr) | |
|---|---|---|---|---|---|---|---|---|
| | T | C | T | C | T | C | T | C |
| **PS-matching / on-treatment TAR** | | | | | | | | |
| CCAE | 14,268 | 44,964 | 9.0 | 35.6 | 22 | 123 | 2.44 (1.53 - 3.70) | 3.46 (2.87 - 4.12) |
| ODOD | 7,799 | 28,030 | 4.5 | 22.1 | 27 | 234 | 5.98 (3.94 - 8.70) | 10.6 (9.28 - 12.0) |
| MDCR | 957 | 7,797 | 0.6 | 7.4 | <5 | 108 | < 8.01 (2.60 - 18.7) | 14.6 (12.0 - 17.6) |
| MDCD | 1,805 | 7,419 | 1.0 | 5.3 | 8 | 69 | 8.06 (3.48 - 15.9) | 13.1 (10.2 - 16.6) |
| Open Claims | 141,653 | 435,868 | 103.4 | 382.2 | 309 | 2,323 | 2.99 (2.67 - 3.34) | 6.08 (5.83 - 6.33) |
| OEHR | 10,486 | 38,365 | 3.2 | 19.9 | 9 | 80 | 2.79 (1.27 - 5.29) | 4.03 (3.19 - 5.01) |
| Meta-analysis | 176,011 | 554,646 | 121.1 | 465.0 | 375 | 2,829 | 3.10 (2.79 - 3.43) | 6.08 (5.86 - 6.31) |
| Meta-analysis (w/o USOC) | 34,358 | 118,778 | 17.7 | 82.8 | 66 | 506 | 3.72 (2.88 - 4.73) | 6.11 (5.59 - 6.67) |
| **PS-stratification / on-treatment TAR** | | | | | | | | |
| CCAE | 19,232 | 44,968 | 11.4 | 35.6 | 29 | 123 | 2.55 (1.71 - 3.66) | 3.46 (2.87 - 4.12) |
| ODOD | 11,498 | 28,658 | 6.1 | 22.7 | 35 | 254 | 5.77 (4.02 - 8.02) | 11.2 (9.85 - 12.6) |
| MDCR | 1,094 | 8,390 | 0.7 | 7.9 | <5 | 120 | < 7.54 (2.45 - 17.6) | 15.2 (12.6 - 18.2) |
| MDCD | 1,977 | 7,475 | 1.1 | 5.3 | 8 | 69 | 7.52 (3.25 - 14.8) | 13.0 (10.1 - 16.4) |
| Open Claims | 226,789 | 436,800 | 141.4 | 383.1 | 396 | 2,350 | 2.80 (2.53 - 3.09) | 6.13 (5.89 - 6.39) |
| OEHR | 13,541 | 38,563 | 4.0 | 20.0 | 9 | 86 | 2.26 (1.03 - 4.29) | 4.31 (3.44 - 5.32) |
| Meta-analysis | 273,037 | 556,464 | 163.9 | 466.7 | 477 | 2,882 | 2.91 (2.66 - 3.18) | 6.17 (5.95 - 6.40) |
| Meta-analysis (w/o USOC) | 46,248 | 119,664 | 22.5 | 83.6 | 81 | 532 | 3.60 (2.86 - 4.48) | 6.36 (5.83 - 6.93) |
| **PS-matching / on-treatment TAR without escalation** | | | | | | | | |
| CCAE | 14,268 | 44,964 | 8.1 | 29.3 | 16 | 103 | 1.98 (1.13 - 3.22) | 3.52 (2.87 - 4.27) |
| ODOD | 7,799 | 28,030 | 4.1 | 18.5 | 23 | 207 | 5.66 (3.59 - 8.50) | 11.2 (9.69 - 12.8) |
| MDCR | 957 | 7,797 | 0.6 | 6.4 | <5 | 86 | < 8.69 (2.82 - 20.3) | 13.4 (10.7 - 16.5) |
| MDCD | 1,805 | 7,419 | 0.9 | 4.1 | 7 | 57 | 8.22 (3.30 - 16.9) | 13.9 (10.5 - 18.0) |
| Open Claims | 141,653 | 435,868 | 91.6 | 303.4 | 263 | 1,877 | 2.87 (2.54 - 3.24) | 6.19 (5.91 - 6.47) |
| OEHR | 10,486 | 38,365 | 2.8 | 16.0 | 7 | 55 | 2.52 (1.01 - 5.19) | 3.44 (2.59 - 4.48) |
| Meta-analysis | 176,011 | 554,646 | 107.3 | 371.3 | 316 | 2,299 | 2.94 (2.63 - 3.29) | 6.19 (5.94 - 6.45) |
| Meta-analysis (w/o USOC) | 34,358 | 118,778 | 15.8 | 67.9 | 53 | 422 | 3.36 (2.52 - 4.40) | 6.21 (5.63 - 6.84) |
| **PS-stratification / on-treatment TAR without escalation** | | | | | | | | |
| CCAE | 19,232 | 44,968 | 10.3 | 29.3 | 21 | 103 | 2.04 (1.26 - 3.12) | 3.52 (2.87 - 4.27) |
| ODOD | 11,498 | 28,658 | 5.5 | 19.1 | 31 | 227 | 5.61 (3.81 - 7.96) | 11.9 (10.4 - 13.6) |
| MDCR | 1,094 | 8,390 | 0.6 | 6.9 | <5 | 96 | < 8.16 (2.65 - 19.0) | 13.9 (11.3 - 17.0) |
| MDCD | 1,977 | 7,475 | 0.9 | 4.2 | 7 | 57 | 7.64 (3.07 - 15.7) | 13.7 (10.4 - 17.8) |
| Open Claims | 226,789 | 436,800 | 127.5 | 304.2 | 348 | 1,900 | 2.73 (2.45 - 3.03) | 6.25 (5.97 - 6.53) |
| OEHR | 13,541 | 38,563 | 3.5 | 16.1 | 7 | 60 | 2.02 (0.81 - 4.16) | 3.73 (2.85 - 4.80) |
| Meta-analysis | 273,037 | 556,464 | 147.7 | 372.8 | 414 | 2,347 | 2.80 (2.54 - 3.09) | 6.30 (6.04 - 6.56) |
| Meta-analysis (w/o USOC) | 46,248 | 119,664 | 20.2 | 68.6 | 66 | 447 | 3.27 (2.53 - 4.16) | 6.52 (5.93 - 7.15) |

##### 4.4.9 Sudden cardiac death

**Supplementary Table 372:** Exposures, outcomes and risk estimates for Sudden cardiac death in GLP1RA (female) (T) vs DPP4I (female) (C) new-user cohorts under four different propensity score (PS) adjustment / time-at-risk (TAR) designs across data sources. We report total patient exposure time in 1,000 patient-year (KYr) units, uncalibrated and calibrated hazard ratios (HRs) and their 95% confidence intervals (CIs). Greyed out entries did not pass study diagnostics and are included only for completeness.

| | Patients | | Exposure (KYr) | | Outcomes | | Incidence (per KYr) | |
|---|---|---|---|---|---|---|---|---|
| | T | C | T | C | T | C | T | C |
| **PS-matching / on-treatment TAR** | | | | | | | | |
| CCAE | 14,422 | 45,475 | 9.1 | 36.0 | 6 | 16 | 0.66 (0.24 - 1.43) | 0.44 (0.25 - 0.72) |
| ODOD | 7,916 | 28,775 | 4.6 | 22.8 | 7 | 61 | 1.51 (0.61 - 3.12) | 2.67 (2.04 - 3.43) |
| MDCR | 983 | 8,427 | 0.6 | 8.0 | 0 | 11 | | 1.37 (0.69 - 2.46) |
| MDCD | 1,860 | 7,678 | 1.0 | 5.5 | 0 | 18 | | 3.29 (1.95 - 5.20) |
| Open Claims | 144,684 | 448,094 | 105.4 | 393.7 | 54 | 470 | 0.51 (0.38 - 0.67) | 1.19 (1.09 - 1.31) |
| OEHR | 10,558 | 38,677 | 3.3 | 20.1 | <5 | 19 | < 1.53 (0.50 - 3.58) | 0.95 (0.57 - 1.48) |
| Meta-analysis | 152,600 | 476,869 | 110.1 | 416.5 | 61 | 531 | 0.55 (0.42 - 0.71) | 1.27 (1.17 - 1.39) |
| Meta-analysis (w/o USOC) | 7,916 | 28,775 | 4.6 | 22.8 | 7 | 61 | 1.51 (0.61 - 3.12) | 2.67 (2.04 - 3.43) |
| **PS-stratification / on-treatment TAR** | | | | | | | | |
| CCAE | 19,424 | 45,480 | 11.5 | 36.0 | 6 | 16 | 0.52 (0.19 - 1.14) | 0.44 (0.25 - 0.72) |
| ODOD | 11,656 | 29,466 | 6.2 | 23.5 | 8 | 66 | 1.29 (0.56 - 2.55) | 2.81 (2.18 - 3.58) |
| MDCR | 1,124 | 8,792 | 0.7 | 8.3 | 0 | 14 | | 1.69 (0.92 - 2.83) |
| MDCD | 2,033 | 7,756 | 1.1 | 5.5 | 0 | 20 | | 3.61 (2.21 - 5.58) |
| Open Claims | 230,968 | 449,183 | 143.9 | 394.7 | 66 | 479 | 0.46 (0.35 - 0.58) | 1.21 (1.11 - 1.33) |
| OEHR | 13,631 | 38,881 | 4.0 | 20.2 | <5 | 20 | < 1.24 (0.40 - 2.90) | 0.99 (0.60 - 1.53) |
| Meta-analysis | 275,679 | 563,010 | 165.6 | 474.4 | <85 | 581 | < 0.51 (0.41 - 0.63) | 1.22 (1.13 - 1.33) |
| Meta-analysis (w/o USOC) | 44,711 | 113,827 | 21.7 | 79.7 | <19 | 102 | < 0.88 (0.53 - 1.37) | 1.28 (1.04 - 1.55) |
| **PS-matching / on-treatment TAR without escalation** | | | | | | | | |
| CCAE | 14,422 | 45,475 | 8.2 | 29.6 | 5 | 11 | 0.61 (0.20 - 1.43) | 0.37 (0.19 - 0.67) |
| ODOD | 7,916 | 28,775 | 4.1 | 19.1 | 7 | 51 | 1.69 (0.68 - 3.48) | 2.67 (1.98 - 3.50) |
| MDCR | 983 | 8,427 | 0.6 | 7.0 | 0 | 7 | | 1.01 (0.40 - 2.07) |
| MDCD | 1,860 | 7,678 | 0.9 | 4.3 | 0 | 11 | | 2.58 (1.29 - 4.62) |
| Open Claims | 144,684 | 448,094 | 93.4 | 312.5 | 47 | 381 | 0.50 (0.37 - 0.67) | 1.22 (1.10 - 1.35) |
| OEHR | 10,558 | 38,677 | 2.8 | 16.2 | <5 | 10 | < 1.78 (0.58 - 4.15) | 0.62 (0.30 - 1.14) |
| Meta-analysis | 152,600 | 476,869 | 97.5 | 331.6 | 54 | 432 | 0.55 (0.42 - 0.72) | 1.30 (1.18 - 1.43) |
| Meta-analysis (w/o USOC) | 7,916 | 28,775 | 4.1 | 19.1 | 7 | 51 | 1.69 (0.68 - 3.48) | 2.67 (1.98 - 3.50) |
| **PS-stratification / on-treatment TAR without escalation** | | | | | | | | |
| CCAE | 19,424 | 45,480 | 10.4 | 29.6 | 5 | 11 | 0.48 (0.16 - 1.12) | 0.37 (0.19 - 0.67) |
| ODOD | 11,656 | 29,466 | 5.6 | 19.7 | 8 | 53 | 1.42 (0.61 - 2.80) | 2.69 (2.02 - 3.52) |
| MDCR | 1,124 | 8,792 | 0.6 | 7.2 | 0 | 10 | | 1.39 (0.67 - 2.55) |
| MDCD | 2,033 | 7,756 | 0.9 | 4.3 | 0 | 12 | | 2.78 (1.44 - 4.86) |
| Open Claims | 230,968 | 449,183 | 129.8 | 313.4 | 57 | 389 | 0.44 (0.33 - 0.57) | 1.24 (1.12 - 1.37) |
| OEHR | 13,631 | 38,881 | 3.5 | 16.3 | <5 | 11 | < 1.43 (0.46 - 3.33) | 0.68 (0.34 - 1.21) |
| Meta-analysis | 242,624 | 478,649 | 135.4 | 333.1 | 65 | 442 | 0.48 (0.37 - 0.61) | 1.33 (1.21 - 1.46) |
| Meta-analysis (w/o USOC) | 11,656 | 29,466 | 5.6 | 19.7 | 8 | 53 | 1.42 (0.61 - 2.80) | 2.69 (2.02 - 3.52) |

##### 4.4.10 Abnormal weight gain

**Supplementary Table 373:** Exposures, outcomes and risk estimates for Abnormal weight gain in GLP1RA (female) (T) vs DPP4I (female) (C) new-user cohorts under four different propensity score (PS) adjustment / time-at-risk (TAR) designs across data sources. We report total patient exposure time in 1,000 patient-year (KYr) units, uncalibrated and calibrated hazard ratios (HRs) and their 95% confidence intervals (CIs). Greyed out entries did not pass study diagnostics and are included only for completeness.

| | Patients | | Exposure (KYr) | | Outcomes | | Incidence (per KYr) | |
|---|---|---|---|---|---|---|---|---|
| | T | C | T | C | T | C | T | C |
| **PS-matching / on-treatment TAR** | | | | | | | | |
| CCAE | 12,839 | 42,506 | 8.0 | 33.6 | 85 | 255 | 10.6 (8.45 - 13.1) | 7.59 (6.68 - 8.58) |
| ODOD | 7,201 | 27,139 | 4.2 | 21.4 | 44 | 172 | 10.5 (7.63 - 14.1) | 8.03 (6.87 - 9.32) |
| MDCR | 950 | 8,313 | 0.6 | 7.9 | <5 | 26 | < 8.13 (2.64 - 19.0) | 3.30 (2.15 - 4.83) |
| MDCD | 1,607 | 6,993 | 0.9 | 4.9 | 11 | 49 | 12.5 (6.22 - 22.3) | 9.92 (7.34 - 13.1) |
| Open Claims | 132,024 | 424,771 | 95.7 | 373.4 | 375 | 1,127 | 3.92 (3.53 - 4.34) | 3.02 (2.84 - 3.20) |
| OEHR | 9,609 | 36,545 | 2.9 | 18.8 | 36 | 166 | 12.3 (8.59 - 17.0) | 8.83 (7.54 - 10.3) |
| Meta-analysis | 163,280 | 537,954 | 111.7 | 452.2 | 551 | 1,769 | 4.93 (4.53 - 5.36) | 3.91 (3.73 - 4.10) |
| Meta-analysis (w/o USOC) | 31,256 | 113,183 | 16.0 | 78.8 | 176 | 642 | 11.0 (9.41 - 12.7) | 8.15 (7.53 - 8.80) |
| **PS-stratification / on-treatment TAR** | | | | | | | | |
| CCAE | 16,752 | 42,511 | 9.9 | 33.6 | 110 | 255 | 11.1 (9.15 - 13.4) | 7.58 (6.68 - 8.57) |
| ODOD | 10,178 | 27,829 | 5.4 | 22.1 | 61 | 179 | 11.4 (8.69 - 14.6) | 8.11 (6.96 - 9.39) |
| MDCR | 1,069 | 8,672 | 0.6 | 8.2 | <5 | 26 | < 7.73 (2.51 - 18.0) | 3.19 (2.08 - 4.67) |
| MDCD | 1,722 | 7,067 | 0.9 | 5.0 | 13 | 50 | 14.1 (7.50 - 24.1) | 10.00 (7.42 - 13.2) |
| Open Claims | 202,244 | 425,856 | 126.4 | 374.5 | 549 | 1,130 | 4.34 (3.99 - 4.72) | 3.02 (2.84 - 3.20) |
| OEHR | 12,038 | 36,753 | 3.5 | 18.9 | 53 | 168 | 15.0 (11.3 - 19.7) | 8.88 (7.59 - 10.3) |
| Meta-analysis | 242,934 | 540,016 | 146.1 | 454.1 | 786 | 1,782 | 5.38 (5.01 - 5.77) | 3.92 (3.74 - 4.11) |
| Meta-analysis (w/o USOC) | 40,690 | 114,160 | 19.7 | 79.6 | 237 | 652 | 12.0 (10.5 - 13.7) | 8.19 (7.57 - 8.84) |
| **PS-matching / on-treatment TAR without escalation** | | | | | | | | |
| CCAE | 12,839 | 42,506 | 7.2 | 27.7 | 78 | 202 | 10.8 (8.56 - 13.5) | 7.30 (6.33 - 8.38) |
| ODOD | 7,201 | 27,139 | 3.8 | 18.0 | 40 | 139 | 10.6 (7.58 - 14.5) | 7.71 (6.48 - 9.10) |
| MDCR | 950 | 8,313 | 0.6 | 6.9 | <5 | 20 | < 8.85 (2.87 - 20.6) | 2.92 (1.78 - 4.50) |
| MDCD | 1,607 | 6,993 | 0.8 | 3.8 | 9 | 35 | 11.9 (5.43 - 22.5) | 9.12 (6.35 - 12.7) |
| Open Claims | 132,024 | 424,771 | 84.7 | 296.9 | 331 | 857 | 3.91 (3.50 - 4.35) | 2.89 (2.70 - 3.09) |
| OEHR | 9,609 | 36,545 | 2.5 | 15.2 | 30 | 113 | 11.8 (7.97 - 16.9) | 7.46 (6.14 - 8.96) |
| Meta-analysis | 163,280 | 537,954 | 99.0 | 361.6 | 488 | 1,346 | 4.93 (4.50 - 5.39) | 3.72 (3.53 - 3.93) |
| Meta-analysis (w/o USOC) | 31,256 | 113,183 | 14.3 | 64.7 | 157 | 489 | 11.0 (9.35 - 12.9) | 7.56 (6.91 - 8.26) |
| **PS-stratification / on-treatment TAR without escalation** | | | | | | | | |
| CCAE | 16,752 | 42,511 | 8.9 | 27.7 | 102 | 202 | 11.4 (9.30 - 13.8) | 7.30 (6.33 - 8.38) |
| ODOD | 10,178 | 27,829 | 4.9 | 18.6 | 56 | 145 | 11.5 (8.67 - 14.9) | 7.81 (6.59 - 9.19) |
| MDCR | 1,069 | 8,672 | 0.6 | 7.1 | <5 | 20 | < 8.39 (2.72 - 19.6) | 2.82 (1.72 - 4.35) |
| MDCD | 1,722 | 7,067 | 0.8 | 3.9 | 11 | 36 | 13.8 (6.90 - 24.7) | 9.27 (6.49 - 12.8) |
| Open Claims | 202,244 | 425,856 | 113.9 | 297.8 | 499 | 860 | 4.38 (4.01 - 4.78) | 2.89 (2.70 - 3.09) |
| OEHR | 12,038 | 36,753 | 3.1 | 15.3 | 46 | 115 | 15.0 (11.0 - 20.0) | 7.54 (6.22 - 9.05) |
| Meta-analysis | 242,934 | 540,016 | 131.6 | 363.2 | 714 | 1,358 | 5.43 (5.04 - 5.84) | 3.74 (3.54 - 3.94) |
| Meta-analysis (w/o USOC) | 40,690 | 114,160 | 17.7 | 65.4 | 215 | 498 | 12.1 (10.6 - 13.9) | 7.62 (6.96 - 8.32) |

##### 4.4.11 Abnormal weight loss

**Supplementary Table 374:** Exposures, outcomes and risk estimates for Abnormal weight loss in GLP1RA (female) (T) vs DPP4I (female) (C) new-user cohorts under four different propensity score (PS) adjustment / time-at-risk (TAR) designs across data sources. We report total patient exposure time in 1,000 patient-year (KYr) units, uncalibrated and calibrated hazard ratios (HRs) and their 95% confidence intervals (CIs). Greyed out entries did not pass study diagnostics and are included only for completeness.

| | Patients | | Exposure (KYr) | | Outcomes | | Incidence (per KYr) | |
|---|---|---|---|---|---|---|---|---|
| | T | C | T | C | T | C | T | C |
| **PS-matching / on-treatment TAR** | | | | | | | | |
| CCAE | 14,105 | 44,496 | 8.9 | 35.0 | 81 | 243 | 9.14 (7.26 - 11.4) | 6.93 (6.09 - 7.86) |
| ODOD | 7,697 | 27,623 | 4.5 | 21.7 | 73 | 313 | 16.4 (12.8 - 20.6) | 14.4 (12.9 - 16.1) |
| MDCR | 964 | 8,114 | 0.6 | 7.7 | 11 | 109 | 17.6 (8.77 - 31.4) | 14.2 (11.7 - 17.2) |
| MDCD | 1,790 | 7,318 | 1.0 | 5.2 | 20 | 84 | 20.7 (12.6 - 31.9) | 16.3 (13.0 - 20.2) |
| Open Claims | 139,999 | 430,739 | 101.8 | 376.4 | 539 | 2,632 | 5.29 (4.86 - 5.76) | 6.99 (6.73 - 7.27) |
| OEHR | 10,300 | 37,586 | 3.2 | 19.3 | 27 | 220 | 8.51 (5.61 - 12.4) | 11.4 (9.92 - 13.0) |
| Meta-analysis | 173,891 | 547,762 | 119.3 | 457.6 | 740 | 3,492 | 6.20 (5.76 - 6.67) | 7.63 (7.38 - 7.89) |
| Meta-analysis (w/o USOC) | 33,892 | 117,023 | 17.5 | 81.3 | 201 | 860 | 11.5 (9.97 - 13.2) | 10.6 (9.89 - 11.3) |
| **PS-stratification / on-treatment TAR** | | | | | | | | |
| CCAE | 18,985 | 44,501 | 11.2 | 35.0 | 106 | 243 | 9.49 (7.77 - 11.5) | 6.93 (6.09 - 7.86) |
| ODOD | 11,296 | 28,127 | 6.0 | 22.2 | 88 | 328 | 14.7 (11.8 - 18.2) | 14.8 (13.2 - 16.5) |
| MDCR | 1,107 | 8,411 | 0.7 | 7.9 | 12 | 120 | 18.0 (9.32 - 31.5) | 15.2 (12.6 - 18.2) |
| MDCD | 1,955 | 7,383 | 1.0 | 5.2 | 20 | 88 | 19.3 (11.8 - 29.8) | 16.9 (13.6 - 20.8) |
| Open Claims | 223,395 | 431,515 | 138.9 | 377.1 | 739 | 2,654 | 5.32 (4.94 - 5.72) | 7.04 (6.77 - 7.31) |
| OEHR | 13,291 | 37,764 | 3.9 | 19.4 | 34 | 223 | 8.71 (6.03 - 12.2) | 11.5 (10.0 - 13.1) |
| Meta-analysis | 268,922 | 549,290 | 161.0 | 459.0 | 987 | 3,536 | 6.13 (5.75 - 6.52) | 7.70 (7.45 - 7.96) |
| Meta-analysis (w/o USOC) | 45,527 | 117,775 | 22.1 | 81.9 | 248 | 882 | 11.2 (9.87 - 12.7) | 10.8 (10.1 - 11.5) |
| **PS-matching / on-treatment TAR without escalation** | | | | | | | | |
| CCAE | 14,105 | 44,496 | 8.0 | 28.8 | 69 | 208 | 8.67 (6.74 - 11.0) | 7.21 (6.26 - 8.26) |
| ODOD | 7,697 | 27,623 | 4.0 | 18.2 | 65 | 263 | 16.2 (12.5 - 20.7) | 14.4 (12.7 - 16.3) |
| MDCR | 964 | 8,114 | 0.6 | 6.7 | 10 | 98 | 17.4 (8.33 - 32.0) | 14.7 (12.0 - 18.0) |
| MDCD | 1,790 | 7,318 | 0.8 | 4.0 | 13 | 59 | 15.7 (8.33 - 26.8) | 14.6 (11.1 - 18.8) |
| Open Claims | 139,999 | 430,739 | 90.2 | 298.7 | 475 | 2,139 | 5.26 (4.80 - 5.76) | 7.16 (6.86 - 7.47) |
| OEHR | 10,300 | 37,586 | 2.7 | 15.6 | 21 | 172 | 7.67 (4.75 - 11.7) | 11.0 (9.45 - 12.8) |
| Meta-analysis | 173,891 | 547,762 | 105.8 | 365.5 | 643 | 2,841 | 6.08 (5.62 - 6.57) | 7.77 (7.49 - 8.07) |
| Meta-analysis (w/o USOC) | 33,892 | 117,023 | 15.5 | 66.7 | 168 | 702 | 10.8 (9.24 - 12.6) | 10.5 (9.76 - 11.3) |
| **PS-stratification / on-treatment TAR without escalation** | | | | | | | | |
| CCAE | 18,985 | 44,501 | 10.1 | 28.8 | 88 | 208 | 8.69 (6.97 - 10.7) | 7.21 (6.26 - 8.26) |
| ODOD | 11,296 | 28,127 | 5.4 | 18.6 | 79 | 275 | 14.6 (11.5 - 18.1) | 14.8 (13.1 - 16.6) |
| MDCR | 1,107 | 8,411 | 0.6 | 6.8 | 11 | 109 | 17.9 (8.95 - 32.1) | 15.9 (13.1 - 19.2) |
| MDCD | 1,955 | 7,383 | 0.9 | 4.1 | 13 | 63 | 14.5 (7.74 - 24.8) | 15.5 (11.9 - 19.8) |
| Open Claims | 223,395 | 431,515 | 125.4 | 299.3 | 666 | 2,158 | 5.31 (4.92 - 5.73) | 7.21 (6.91 - 7.52) |
| OEHR | 13,291 | 37,764 | 3.4 | 15.7 | 28 | 175 | 8.23 (5.47 - 11.9) | 11.2 (9.58 - 13.0) |
| Meta-analysis | 268,922 | 549,290 | 145.2 | 366.6 | 874 | 2,879 | 6.02 (5.63 - 6.43) | 7.85 (7.57 - 8.15) |
| Meta-analysis (w/o USOC) | 45,527 | 117,775 | 19.8 | 67.2 | 208 | 721 | 10.5 (9.10 - 12.0) | 10.7 (9.96 - 11.5) |

##### 4.4.12 Acute pancreatitis

**Supplementary Table 375:** Exposures, outcomes and risk estimates for Acute pancreatitis in GLP1RA (female) (T) vs DPP4I (female) (C) new-user cohorts under four different propensity score (PS) adjustment / time-at-risk (TAR) designs across data sources. We report total patient exposure time in 1,000 patient-year (KYr) units, uncalibrated and calibrated hazard ratios (HRs) and their 95% confidence intervals (CIs). Greyed out entries did not pass study diagnostics and are included only for completeness.

| | Patients | | Exposure (KYr) | | Outcomes | | Incidence (per KYr) | |
|---|---|---|---|---|---|---|---|---|
| | T | C | T | C | T | C | T | C |
| **PS-matching / on-treatment TAR** | | | | | | | | |
| CCAE | 14,343 | 45,207 | 9.1 | 35.7 | 15 | 76 | 1.65 (0.93 - 2.73) | 2.13 (1.68 - 2.66) |
| ODOD | 7,876 | 28,636 | 4.6 | 22.7 | 13 | 62 | 2.83 (1.51 - 4.83) | 2.73 (2.09 - 3.50) |
| MDCR | 982 | 8,389 | 0.6 | 8.0 | <5 | 16 | < 7.85 (2.55 - 18.3) | 2.01 (1.15 - 3.26) |
| MDCD | 1,840 | 7,561 | 1.0 | 5.4 | 5 | 25 | 4.96 (1.61 - 11.6) | 4.64 (3.00 - 6.84) |
| Open Claims | 143,294 | 443,745 | 104.5 | 389.7 | 169 | 838 | 1.62 (1.38 - 1.88) | 2.15 (2.01 - 2.30) |
| OEHR | 10,522 | 38,526 | 3.2 | 20.0 | 9 | 39 | 2.77 (1.27 - 5.26) | 1.95 (1.39 - 2.67) |
| Meta-analysis | 177,875 | 563,675 | 122.5 | 473.6 | 211 | 1,040 | 1.72 (1.50 - 1.97) | 2.20 (2.06 - 2.33) |
| Meta-analysis (w/o USOC) | 34,581 | 119,930 | 17.9 | 83.8 | 42 | 202 | 2.34 (1.69 - 3.17) | 2.41 (2.09 - 2.77) |
| **PS-stratification / on-treatment TAR** | | | | | | | | |
| CCAE | 19,337 | 45,212 | 11.4 | 35.7 | 16 | 77 | 1.40 (0.80 - 2.27) | 2.16 (1.70 - 2.69) |
| ODOD | 11,600 | 29,302 | 6.2 | 23.3 | 16 | 67 | 2.60 (1.48 - 4.22) | 2.87 (2.23 - 3.65) |
| MDCR | 1,124 | 8,746 | 0.7 | 8.3 | <5 | 18 | < 7.39 (2.40 - 17.3) | 2.18 (1.29 - 3.45) |
| MDCD | 2,010 | 7,636 | 1.1 | 5.5 | 5 | 25 | 4.63 (1.50 - 10.8) | 4.58 (2.96 - 6.76) |
| Open Claims | 228,988 | 444,912 | 142.8 | 390.9 | 217 | 842 | 1.52 (1.32 - 1.74) | 2.15 (2.01 - 2.30) |
| OEHR | 13,579 | 38,728 | 4.0 | 20.1 | 10 | 39 | 2.50 (1.20 - 4.59) | 1.94 (1.38 - 2.65) |
| Meta-analysis | 275,514 | 565,790 | 165.5 | 475.5 | 264 | 1,050 | 1.60 (1.41 - 1.80) | 2.21 (2.08 - 2.35) |
| Meta-analysis (w/o USOC) | 46,526 | 120,878 | 22.7 | 84.6 | 47 | 208 | 2.07 (1.52 - 2.75) | 2.46 (2.14 - 2.82) |
| **PS-matching / on-treatment TAR without escalation** | | | | | | | | |
| CCAE | 14,343 | 45,207 | 8.1 | 29.4 | 12 | 59 | 1.48 (0.76 - 2.58) | 2.01 (1.53 - 2.59) |
| ODOD | 7,876 | 28,636 | 4.1 | 19.0 | 11 | 52 | 2.66 (1.33 - 4.77) | 2.73 (2.04 - 3.58) |
| MDCR | 982 | 8,389 | 0.6 | 6.9 | <5 | 12 | < 8.53 (2.77 - 19.9) | 1.73 (0.89 - 3.02) |
| MDCD | 1,840 | 7,561 | 0.9 | 4.2 | <5 | 16 | < 5.75 (1.87 - 13.4) | 3.81 (2.18 - 6.19) |
| Open Claims | 143,294 | 443,745 | 92.6 | 309.4 | 149 | 649 | 1.61 (1.36 - 1.89) | 2.10 (1.94 - 2.27) |
| OEHR | 10,522 | 38,526 | 2.8 | 16.1 | 8 | 29 | 2.86 (1.23 - 5.63) | 1.80 (1.21 - 2.59) |
| Meta-analysis | 176,035 | 556,114 | 107.6 | 373.9 | 180 | 789 | 1.67 (1.44 - 1.94) | 2.11 (1.97 - 2.26) |
| Meta-analysis (w/o USOC) | 32,741 | 112,369 | 15.1 | 64.5 | 31 | 140 | 2.06 (1.40 - 2.92) | 2.17 (1.83 - 2.56) |
| **PS-stratification / on-treatment TAR without escalation** | | | | | | | | |
| CCAE | 19,337 | 45,212 | 10.4 | 29.4 | 13 | 60 | 1.26 (0.67 - 2.15) | 2.04 (1.56 - 2.63) |
| ODOD | 11,600 | 29,302 | 5.6 | 19.6 | 14 | 56 | 2.50 (1.37 - 4.19) | 2.86 (2.16 - 3.72) |
| MDCR | 1,124 | 8,746 | 0.6 | 7.2 | <5 | 14 | < 8.01 (2.60 - 18.7) | 1.95 (1.07 - 3.27) |
| MDCD | 2,010 | 7,636 | 0.9 | 4.2 | <5 | 16 | < 5.34 (1.73 - 12.5) | 3.77 (2.15 - 6.12) |
| Open Claims | 228,988 | 444,912 | 128.8 | 310.4 | 193 | 653 | 1.50 (1.29 - 1.73) | 2.10 (1.95 - 2.27) |
| OEHR | 13,579 | 38,728 | 3.5 | 16.2 | 9 | 29 | 2.58 (1.18 - 4.90) | 1.79 (1.20 - 2.57) |
| Meta-analysis | 273,504 | 558,154 | 148.2 | 375.5 | 229 | 798 | 1.54 (1.35 - 1.76) | 2.13 (1.98 - 2.28) |
| Meta-analysis (w/o USOC) | 44,516 | 113,242 | 19.4 | 65.1 | 36 | 145 | 1.85 (1.30 - 2.56) | 2.23 (1.88 - 2.62) |

##### 4.4.13 Bone fracture

**Supplementary Table 376:** Exposures, outcomes and risk estimates for Bone fracture in GLP1RA (female) (T) vs DPP4I (female) (C) new-user cohorts under four different propensity score (PS) adjustment / time-at-risk (TAR) designs across data sources. We report total patient exposure time in 1,000 patient-year (KYr) units, uncalibrated and calibrated hazard ratios (HRs) and their 95% confidence intervals (CIs). Greyed out entries did not pass study diagnostics and are included only for completeness.

| | Patients | | Exposure (KYr) | | Outcomes | | Incidence (per KYr) | |
|---|---|---|---|---|---|---|---|---|
| | T | C | T | C | T | C | T | C |
| **PS-matching / on-treatment TAR** | | | | | | | | |
| CCAE | 12,951 | 41,233 | 8.0 | 31.8 | 163 | 694 | 20.4 (17.4 - 23.8) | 21.8 (20.2 - 23.5) |
| ODOD | 7,110 | 25,541 | 4.1 | 19.7 | 123 | 672 | 30.2 (25.1 - 36.0) | 34.2 (31.6 - 36.9) |
| MDCR | 835 | 7,107 | 0.5 | 6.6 | 18 | 301 | 33.3 (19.7 - 52.6) | 45.7 (40.7 - 51.1) |
| MDCD | 1,600 | 6,648 | 0.9 | 4.6 | 32 | 182 | 37.3 (25.5 - 52.6) | 39.4 (33.9 - 45.6) |
| Open Claims | 122,716 | 384,052 | 88.4 | 330.6 | 1,321 | 7,197 | 14.9 (14.1 - 15.8) | 21.8 (21.3 - 22.3) |
| OEHR | 9,754 | 35,795 | 3.0 | 18.3 | 36 | 409 | 12.0 (8.40 - 16.6) | 22.4 (20.3 - 24.6) |
| Meta-analysis | 154,131 | 493,269 | 104.3 | 405.0 | 1,675 | 9,154 | 16.1 (15.3 - 16.8) | 22.6 (22.1 - 23.1) |
| Meta-analysis (w/o USOC) | 31,415 | 109,217 | 15.9 | 74.4 | 354 | 1,957 | 22.2 (20.0 - 24.7) | 26.3 (25.2 - 27.5) |
| **PS-stratification / on-treatment TAR** | | | | | | | | |
| CCAE | 17,366 | 41,238 | 10.1 | 31.8 | 203 | 694 | 20.2 (17.5 - 23.1) | 21.8 (20.2 - 23.5) |
| ODOD | 10,452 | 26,228 | 5.4 | 20.3 | 161 | 709 | 29.6 (25.2 - 34.6) | 35.0 (32.4 - 37.6) |
| MDCR | 957 | 7,400 | 0.6 | 6.8 | 18 | 311 | 31.3 (18.6 - 49.5) | 45.7 (40.7 - 51.0) |
| MDCD | 1,744 | 6,705 | 0.9 | 4.7 | 33 | 185 | 36.0 (24.8 - 50.6) | 39.7 (34.2 - 45.8) |
| Open Claims | 194,843 | 385,020 | 120.3 | 331.6 | 1,758 | 7,243 | 14.6 (13.9 - 15.3) | 21.8 (21.3 - 22.4) |
| OEHR | 12,509 | 35,951 | 3.7 | 18.4 | 48 | 416 | 13.1 (9.67 - 17.4) | 22.7 (20.5 - 24.9) |
| Meta-analysis | 236,914 | 495,142 | 140.4 | 406.7 | 2,203 | 9,247 | 15.7 (15.0 - 16.4) | 22.7 (22.3 - 23.2) |
| Meta-analysis (w/o USOC) | 42,071 | 110,122 | 20.1 | 75.1 | 445 | 2,004 | 22.2 (20.2 - 24.3) | 26.7 (25.5 - 27.9) |
| **PS-matching / on-treatment TAR without escalation** | | | | | | | | |
| CCAE | 12,951 | 41,233 | 7.2 | 26.3 | 145 | 558 | 20.2 (17.1 - 23.8) | 21.2 (19.5 - 23.1) |
| ODOD | 7,110 | 25,541 | 3.7 | 16.6 | 106 | 551 | 28.9 (23.7 - 35.0) | 33.3 (30.5 - 36.1) |
| MDCR | 835 | 7,107 | 0.5 | 5.8 | 17 | 250 | 34.1 (19.9 - 54.6) | 43.2 (38.0 - 48.9) |
| MDCD | 1,600 | 6,648 | 0.7 | 3.6 | 29 | 136 | 39.2 (26.2 - 56.3) | 37.7 (31.6 - 44.6) |
| Open Claims | 122,716 | 384,052 | 78.4 | 263.2 | 1,134 | 5,706 | 14.5 (13.6 - 15.3) | 21.7 (21.1 - 22.3) |
| OEHR | 9,754 | 35,795 | 2.6 | 14.8 | 31 | 306 | 12.0 (8.16 - 17.0) | 20.7 (18.4 - 23.2) |
| Meta-analysis | 154,131 | 493,269 | 92.5 | 324.4 | 1,445 | 7,257 | 15.6 (14.8 - 16.4) | 22.4 (21.9 - 22.9) |
| Meta-analysis (w/o USOC) | 31,415 | 109,217 | 14.2 | 61.3 | 311 | 1,551 | 22.0 (19.6 - 24.6) | 25.3 (24.1 - 26.6) |
| **PS-stratification / on-treatment TAR without escalation** | | | | | | | | |
| CCAE | 17,366 | 41,238 | 9.1 | 26.3 | 182 | 558 | 19.9 (17.1 - 23.1) | 21.2 (19.5 - 23.1) |
| ODOD | 10,452 | 26,228 | 5.0 | 17.1 | 140 | 582 | 28.3 (23.8 - 33.4) | 34.0 (31.3 - 36.9) |
| MDCR | 957 | 7,400 | 0.5 | 6.0 | 17 | 260 | 31.9 (18.6 - 51.1) | 43.5 (38.3 - 49.1) |
| MDCD | 1,744 | 6,705 | 0.8 | 3.6 | 30 | 138 | 37.7 (25.5 - 53.9) | 37.9 (31.9 - 44.8) |
| Open Claims | 194,843 | 385,020 | 108.6 | 263.9 | 1,548 | 5,747 | 14.3 (13.6 - 15.0) | 21.8 (21.2 - 22.3) |
| OEHR | 12,509 | 35,951 | 3.2 | 14.8 | 43 | 310 | 13.5 (9.79 - 18.2) | 20.9 (18.6 - 23.3) |
| Meta-analysis | 236,914 | 495,142 | 126.7 | 325.8 | 1,943 | 7,335 | 15.3 (14.7 - 16.0) | 22.5 (22.0 - 23.0) |
| Meta-analysis (w/o USOC) | 42,071 | 110,122 | 18.1 | 61.9 | 395 | 1,588 | 21.9 (19.8 - 24.1) | 25.7 (24.4 - 27.0) |

##### 4.4.14 Diabetic ketoacidosis

**Supplementary Table 377:** Exposures, outcomes and risk estimates for Diabetic ketoacidosis in GLP1RA (female) (T) vs DPP4I (female) (C) new-user cohorts under four different propensity score (PS) adjustment / time-at-risk (TAR) designs across data sources. We report total patient exposure time in 1,000 patient-year (KYr) units, uncalibrated and calibrated hazard ratios (HRs) and their 95% confidence intervals (CIs). Greyed out entries did not pass study diagnostics and are included only for completeness.

| | Patients | | Exposure (KYr) | | Outcomes | | Incidence (per KYr) | |
|---|---|---|---|---|---|---|---|---|
| | T | C | T | C | T | C | T | C |
| **PS-matching / on-treatment TAR** | | | | | | | | |
| CCAE | 14,417 | 45,448 | 9.1 | 36.0 | 6 | 29 | 0.66 (0.24 - 1.43) | 0.81 (0.54 - 1.16) |
| ODOD | 7,919 | 28,782 | 4.6 | 22.8 | 5 | 36 | 1.08 (0.35 - 2.53) | 1.58 (1.11 - 2.18) |
| MDCR | 986 | 8,431 | 0.6 | 8.0 | 0 | 10 | | 1.25 (0.60 - 2.29) |
| MDCD | 1,854 | 7,669 | 1.0 | 5.5 | <5 | 17 | < 4.94 (1.61 - 11.5) | 3.11 (1.81 - 4.98) |
| Open Claims | 144,634 | 448,054 | 105.4 | 393.6 | 59 | 362 | 0.56 (0.43 - 0.72) | 0.92 (0.83 - 1.02) |
| OEHR | 10,556 | 38,675 | 3.3 | 20.1 | 5 | 24 | 1.53 (0.50 - 3.58) | 1.20 (0.77 - 1.78) |
| Meta-analysis | 177,526 | 560,959 | 122.4 | 472.4 | 75 | 451 | 0.61 (0.48 - 0.77) | 0.95 (0.87 - 1.05) |
| Meta-analysis (w/o USOC) | 32,892 | 112,905 | 17.0 | 78.9 | 16 | 89 | 0.94 (0.54 - 1.53) | 1.13 (0.91 - 1.39) |
| **PS-stratification / on-treatment TAR** | | | | | | | | |
| CCAE | 19,428 | 45,453 | 11.5 | 36.0 | 7 | 29 | 0.61 (0.24 - 1.25) | 0.81 (0.54 - 1.16) |
| ODOD | 11,652 | 29,474 | 6.2 | 23.5 | 7 | 37 | 1.13 (0.45 - 2.33) | 1.58 (1.11 - 2.17) |
| MDCR | 1,128 | 8,794 | 0.7 | 8.3 | 0 | 10 | | 1.20 (0.58 - 2.22) |
| MDCD | 2,026 | 7,748 | 1.1 | 5.5 | <5 | 17 | < 4.61 (1.50 - 10.8) | 3.07 (1.79 - 4.92) |
| Open Claims | 230,974 | 449,158 | 143.9 | 394.7 | 78 | 362 | 0.54 (0.43 - 0.68) | 0.92 (0.83 - 1.02) |
| OEHR | 13,629 | 38,879 | 4.0 | 20.2 | 8 | 25 | 1.99 (0.86 - 3.92) | 1.24 (0.80 - 1.83) |
| Meta-analysis | 275,683 | 562,964 | 165.7 | 474.3 | 100 | 453 | 0.60 (0.49 - 0.73) | 0.96 (0.87 - 1.05) |
| Meta-analysis (w/o USOC) | 44,709 | 113,806 | 21.7 | 79.6 | 22 | 91 | 1.01 (0.64 - 1.53) | 1.14 (0.92 - 1.40) |
| **PS-matching / on-treatment TAR without escalation** | | | | | | | | |
| CCAE | 14,417 | 45,448 | 8.2 | 29.6 | 5 | 18 | 0.61 (0.20 - 1.43) | 0.61 (0.36 - 0.96) |
| ODOD | 7,919 | 28,782 | 4.1 | 19.1 | <5 | 29 | < 1.21 (0.39 - 2.81) | 1.52 (1.02 - 2.18) |
| MDCR | 986 | 8,431 | 0.6 | 7.0 | 0 | 8 | | 1.15 (0.50 - 2.26) |
| MDCD | 1,854 | 7,669 | 0.9 | 4.3 | <5 | 12 | < 5.76 (1.87 - 13.4) | 2.82 (1.46 - 4.92) |
| Open Claims | 144,634 | 448,054 | 93.4 | 312.6 | 48 | 252 | 0.51 (0.38 - 0.68) | 0.81 (0.71 - 0.91) |
| OEHR | 10,556 | 38,675 | 2.8 | 16.2 | <5 | 14 | < 1.78 (0.58 - 4.15) | 0.87 (0.47 - 1.45) |
| Meta-analysis | 166,970 | 522,284 | 105.7 | 361.3 | <58 | 299 | < 0.55 (0.42 - 0.71) | 0.83 (0.74 - 0.93) |
| Meta-analysis (w/o USOC) | 22,336 | 74,230 | 12.3 | 48.7 | <10 | 47 | < 0.81 (0.39 - 1.49) | 0.96 (0.71 - 1.28) |
| **PS-stratification / on-treatment TAR without escalation** | | | | | | | | |
| CCAE | 19,428 | 45,453 | 10.4 | 29.6 | 5 | 18 | 0.48 (0.16 - 1.12) | 0.61 (0.36 - 0.96) |
| ODOD | 11,652 | 29,474 | 5.6 | 19.7 | 6 | 30 | 1.07 (0.39 - 2.32) | 1.53 (1.03 - 2.18) |
| MDCR | 1,128 | 8,794 | 0.6 | 7.2 | 0 | 8 | | 1.11 (0.48 - 2.18) |
| MDCD | 2,026 | 7,748 | 0.9 | 4.3 | <5 | 12 | < 5.34 (1.73 - 12.5) | 2.78 (1.44 - 4.86) |
| Open Claims | 230,974 | 449,158 | 129.9 | 313.5 | 67 | 252 | 0.52 (0.40 - 0.66) | 0.80 (0.71 - 0.91) |
| OEHR | 13,629 | 38,879 | 3.5 | 16.3 | <5 | 14 | < 1.43 (0.46 - 3.33) | 0.86 (0.47 - 1.44) |
| Meta-analysis | 262,054 | 524,085 | 145.9 | 362.7 | 78 | 300 | 0.53 (0.42 - 0.67) | 0.83 (0.74 - 0.93) |
| Meta-analysis (w/o USOC) | 31,080 | 74,927 | 16.0 | 49.2 | 11 | 48 | 0.69 (0.34 - 1.23) | 0.97 (0.72 - 1.29) |

##### 4.4.15 Diarrhea

**Supplementary Table 378:** Exposures, outcomes and risk estimates for Diarrhea in GLP1RA (female) (T) vs DPP4I (female) (C) new-user cohorts under four different propensity score (PS) adjustment / time-at-risk (TAR) designs across data sources. We report total patient exposure time in 1,000 patient-year (KYr) units, uncalibrated and calibrated hazard ratios (HRs) and their 95% confidence intervals (CIs). Greyed out entries did not pass study diagnostics and are included only for completeness.

| | Patients | | Exposure (KYr) | | Outcomes | | Incidence (per KYr) | |
|---|---|---|---|---|---|---|---|---|
| | T | C | T | C | T | C | T | C |
| **PS-matching / on-treatment TAR** | | | | | | | | |
| CCAE | 12,135 | 38,768 | 7.4 | 29.7 | 335 | 1,050 | 45.4 (40.7 - 50.6) | 35.3 (33.2 - 37.5) |
| ODOD | 6,507 | 23,550 | 3.7 | 18.0 | 185 | 867 | 50.1 (43.1 - 57.8) | 48.2 (45.0 - 51.5) |
| MDCR | 840 | 7,035 | 0.5 | 6.4 | 24 | 275 | 44.9 (28.8 - 66.9) | 42.7 (37.8 - 48.1) |
| MDCD | 1,404 | 5,749 | 0.7 | 3.9 | 64 | 272 | 86.3 (66.4 - 110) | 70.6 (62.5 - 79.5) |
| Open Claims | 118,300 | 373,356 | 85.1 | 320.7 | 2,145 | 8,088 | 25.2 (24.1 - 26.3) | 25.2 (24.7 - 25.8) |
| OEHR | 8,981 | 33,026 | 2.7 | 16.2 | 168 | 800 | 62.5 (53.4 - 72.7) | 49.2 (45.9 - 52.8) |
| Meta-analysis | 147,327 | 474,449 | 99.6 | 388.6 | 2,897 | 11,077 | 29.1 (28.0 - 30.2) | 28.5 (28.0 - 29.0) |
| Meta-analysis (w/o USOC) | 29,027 | 101,093 | 14.5 | 67.8 | 752 | 2,989 | 51.9 (48.2 - 55.7) | 44.1 (42.5 - 45.7) |
| **PS-stratification / on-treatment TAR** | | | | | | | | |
| CCAE | 16,274 | 38,768 | 9.3 | 29.7 | 457 | 1,050 | 49.2 (44.8 - 53.9) | 35.3 (33.2 - 37.5) |
| ODOD | 9,491 | 24,049 | 4.9 | 18.4 | 264 | 897 | 54.0 (47.7 - 60.9) | 48.7 (45.6 - 52.0) |
| MDCR | 960 | 7,235 | 0.6 | 6.6 | 25 | 285 | 44.2 (28.6 - 65.3) | 43.4 (38.5 - 48.7) |
| MDCD | 1,534 | 5,785 | 0.8 | 3.9 | 73 | 273 | 92.3 (72.3 - 116) | 70.5 (62.4 - 79.4) |
| Open Claims | 186,885 | 374,449 | 115.2 | 321.8 | 3,059 | 8,141 | 26.6 (25.6 - 27.5) | 25.3 (24.8 - 25.9) |
| OEHR | 11,445 | 33,203 | 3.3 | 16.4 | 199 | 804 | 60.8 (52.7 - 69.9) | 49.2 (45.8 - 52.7) |
| Meta-analysis | 225,629 | 476,254 | 133.4 | 390.2 | 4,052 | 11,165 | 30.4 (29.4 - 31.3) | 28.6 (28.1 - 29.2) |
| Meta-analysis (w/o USOC) | 38,744 | 101,805 | 18.2 | 68.4 | 993 | 3,024 | 54.4 (51.1 - 57.9) | 44.2 (42.7 - 45.8) |
| **PS-matching / on-treatment TAR without escalation** | | | | | | | | |
| CCAE | 12,135 | 38,768 | 6.7 | 24.8 | 303 | 865 | 45.5 (40.6 - 51.0) | 34.9 (32.6 - 37.3) |
| ODOD | 6,507 | 23,550 | 3.3 | 15.2 | 174 | 723 | 52.2 (44.7 - 60.6) | 47.5 (44.1 - 51.0) |
| MDCR | 840 | 7,035 | 0.5 | 5.7 | 21 | 244 | 42.7 (26.4 - 65.2) | 43.2 (37.9 - 48.9) |
| MDCD | 1,404 | 5,749 | 0.7 | 3.1 | 55 | 224 | 83.8 (63.2 - 109) | 72.9 (63.6 - 83.1) |
| Open Claims | 118,300 | 373,356 | 75.7 | 256.9 | 1,904 | 6,481 | 25.2 (24.0 - 26.3) | 25.2 (24.6 - 25.8) |
| OEHR | 8,981 | 33,026 | 2.3 | 13.2 | 152 | 638 | 65.2 (55.2 - 76.4) | 48.2 (44.5 - 52.0) |
| Meta-analysis | 147,327 | 474,449 | 88.6 | 313.3 | 2,588 | 8,931 | 29.2 (28.1 - 30.3) | 28.5 (27.9 - 29.1) |
| Meta-analysis (w/o USOC) | 29,027 | 101,093 | 13.0 | 56.4 | 684 | 2,450 | 52.7 (48.8 - 56.8) | 43.5 (41.8 - 45.2) |
| **PS-stratification / on-treatment TAR without escalation** | | | | | | | | |
| CCAE | 16,274 | 38,768 | 8.5 | 24.8 | 409 | 865 | 48.3 (43.8 - 53.3) | 34.9 (32.6 - 37.3) |
| ODOD | 9,491 | 24,049 | 4.5 | 15.6 | 249 | 752 | 55.8 (49.1 - 63.2) | 48.1 (44.8 - 51.7) |
| MDCR | 960 | 7,235 | 0.5 | 5.8 | 22 | 252 | 42.1 (26.4 - 63.7) | 43.7 (38.5 - 49.4) |
| MDCD | 1,534 | 5,785 | 0.7 | 3.1 | 64 | 225 | 91.2 (70.2 - 116) | 72.9 (63.7 - 83.1) |
| Open Claims | 186,885 | 374,449 | 104.1 | 257.8 | 2,767 | 6,522 | 26.6 (25.6 - 27.6) | 25.3 (24.7 - 25.9) |
| OEHR | 11,445 | 33,203 | 2.9 | 13.3 | 180 | 641 | 62.7 (53.9 - 72.6) | 48.1 (44.4 - 51.9) |
| Meta-analysis | 225,629 | 476,254 | 120.6 | 314.7 | 3,669 | 9,005 | 30.4 (29.4 - 31.4) | 28.6 (28.0 - 29.2) |
| Meta-analysis (w/o USOC) | 38,744 | 101,805 | 16.5 | 56.9 | 902 | 2,483 | 54.7 (51.2 - 58.4) | 43.7 (42.0 - 45.4) |

##### 4.4.16 Genitourinary infection

**Supplementary Table 379:** Exposures, outcomes and risk estimates for Genitourinary infection in GLP1RA (female) (T) vs DPP4I (female) (C) new-user cohorts under four different propensity score (PS) adjustment / time-at-risk (TAR) designs across data sources. We report total patient exposure time in 1,000 patient-year (KYr) units, uncalibrated and calibrated hazard ratios (HRs) and their 95% confidence intervals (CIs). Greyed out entries did not pass study diagnostics and are included only for completeness.

| | Patients | | Exposure (KYr) | | Outcomes | | Incidence (per KYr) | |
|---|---|---|---|---|---|---|---|---|
| | T | C | T | C | T | C | T | C |
| **PS-matching / on-treatment TAR** | | | | | | | | |
| CCAE | 9,865 | 31,463 | 5.8 | 23.4 | 425 | 1,750 | 73.2 (66.4 - 80.6) | 74.8 (71.3 - 78.4) |
| ODOD | 5,459 | 18,987 | 3.0 | 13.9 | 271 | 1,382 | 90.1 (79.7 - 102) | 99.2 (94.1 - 105) |
| MDCR | 672 | 5,842 | 0.4 | 5.2 | 32 | 489 | 79.0 (54.0 - 111) | 94.5 (86.3 - 103) |
| MDCD | 1,192 | 4,948 | 0.6 | 3.3 | 73 | 344 | 113 (88.8 - 142) | 105 (94.6 - 117) |
| Open Claims | 92,305 | 294,588 | 66.2 | 248.8 | 2,742 | 13,292 | 41.4 (39.9 - 43.0) | 53.4 (52.5 - 54.3) |
| OEHR | 8,089 | 29,595 | 2.4 | 14.3 | 192 | 1,107 | 81.0 (70.0 - 93.3) | 77.4 (72.9 - 82.1) |
| Meta-analysis | 116,910 | 379,581 | 78.0 | 303.7 | 3,703 | 17,875 | 47.5 (46.0 - 49.0) | 58.9 (58.0 - 59.7) |
| Meta-analysis (w/o USOC) | 24,605 | 84,993 | 11.8 | 54.9 | 961 | 4,583 | 81.3 (76.2 - 86.6) | 83.5 (81.1 - 85.9) |
| **PS-stratification / on-treatment TAR** | | | | | | | | |
| CCAE | 13,187 | 31,471 | 7.3 | 23.4 | 555 | 1,751 | 75.7 (69.5 - 82.2) | 74.8 (71.3 - 78.4) |
| ODOD | 7,974 | 19,378 | 4.0 | 14.3 | 361 | 1,439 | 89.8 (80.8 - 99.6) | 101 (95.7 - 106) |
| MDCR | 772 | 6,012 | 0.4 | 5.3 | 35 | 505 | 81.5 (56.7 - 113) | 95.5 (87.3 - 104) |
| MDCD | 1,310 | 4,991 | 0.7 | 3.3 | 78 | 346 | 113 (89.1 - 141) | 105 (94.2 - 117) |
| Open Claims | 144,467 | 295,210 | 89.2 | 249.4 | 3,719 | 13,368 | 41.7 (40.4 - 43.0) | 53.6 (52.7 - 54.5) |
| OEHR | 10,291 | 29,709 | 2.9 | 14.4 | 243 | 1,118 | 84.3 (74.1 - 95.6) | 77.8 (73.3 - 82.5) |
| Meta-analysis | 177,229 | 380,759 | 104.1 | 304.7 | 4,956 | 18,022 | 47.6 (46.3 - 48.9) | 59.1 (58.3 - 60.0) |
| Meta-analysis (w/o USOC) | 32,762 | 85,549 | 14.9 | 55.3 | 1,237 | 4,654 | 82.9 (78.3 - 87.6) | 84.1 (81.7 - 86.6) |
| **PS-matching / on-treatment TAR without escalation** | | | | | | | | |
| CCAE | 9,865 | 31,463 | 5.3 | 19.5 | 379 | 1,467 | 72.1 (65.0 - 79.7) | 75.4 (71.6 - 79.4) |
| ODOD | 5,459 | 18,987 | 2.7 | 11.9 | 240 | 1,169 | 87.9 (77.1 - 99.7) | 98.1 (92.6 - 104) |
| MDCR | 672 | 5,842 | 0.4 | 4.5 | 25 | 418 | 66.1 (42.8 - 97.6) | 92.2 (83.6 - 101) |
| MDCD | 1,192 | 4,948 | 0.6 | 2.6 | 59 | 254 | 105 (80.0 - 136) | 97.5 (85.9 - 110) |
| Open Claims | 92,305 | 294,588 | 58.7 | 199.1 | 2,385 | 10,685 | 40.6 (39.0 - 42.3) | 53.7 (52.7 - 54.7) |
| OEHR | 8,089 | 29,595 | 2.1 | 11.7 | 160 | 863 | 77.2 (65.7 - 90.1) | 74.0 (69.1 - 79.1) |
| Meta-analysis | 116,910 | 379,581 | 69.3 | 244.7 | 3,223 | 14,438 | 46.5 (44.9 - 48.1) | 59.0 (58.0 - 60.0) |
| Meta-analysis (w/o USOC) | 24,605 | 84,993 | 10.6 | 45.6 | 838 | 3,753 | 78.9 (73.6 - 84.4) | 82.2 (79.6 - 84.9) |
| **PS-stratification / on-treatment TAR without escalation** | | | | | | | | |
| CCAE | 13,187 | 31,471 | 6.7 | 19.5 | 505 | 1,468 | 75.4 (68.9 - 82.2) | 75.4 (71.6 - 79.4) |
| ODOD | 7,974 | 19,378 | 3.7 | 12.2 | 324 | 1,220 | 87.8 (78.5 - 97.9) | 100.0 (94.4 - 106) |
| MDCR | 772 | 6,012 | 0.4 | 4.6 | 27 | 431 | 67.2 (44.3 - 97.8) | 93.0 (84.4 - 102) |
| MDCD | 1,310 | 4,991 | 0.6 | 2.6 | 63 | 255 | 104 (79.9 - 133) | 96.9 (85.4 - 110) |
| Open Claims | 144,467 | 295,210 | 80.5 | 199.6 | 3,307 | 10,757 | 41.1 (39.7 - 42.5) | 53.9 (52.9 - 54.9) |
| OEHR | 10,291 | 29,709 | 2.5 | 11.7 | 206 | 874 | 81.1 (70.4 - 92.9) | 74.6 (69.7 - 79.7) |
| Meta-analysis | 177,229 | 380,759 | 94.1 | 245.6 | 4,405 | 14,574 | 46.8 (45.5 - 48.2) | 59.3 (58.4 - 60.3) |
| Meta-analysis (w/o USOC) | 32,762 | 85,549 | 13.5 | 46.0 | 1,098 | 3,817 | 81.1 (76.4 - 86.0) | 83.0 (80.3 - 85.6) |

##### 4.4.17 Hyperkalemia

**Supplementary Table 380:** Exposures, outcomes and risk estimates for Hyperkalemia in GLP1RA (female) (T) vs DPP4I (female) (C) new-user cohorts under four different propensity score (PS) adjustment / time-at-risk (TAR) designs across data sources. We report total patient exposure time in 1,000 patient-year (KYr) units, uncalibrated and calibrated hazard ratios (HRs) and their 95% confidence intervals (CIs). Greyed out entries did not pass study diagnostics and are included only for completeness.

| | Patients | | Exposure (KYr) | | Outcomes | | Incidence (per KYr) | |
|---|---|---|---|---|---|---|---|---|
| | T | C | T | C | T | C | T | C |
| **PS-matching / on-treatment TAR** | | | | | | | | |
| CCAE | 14,235 | 44,801 | 8.9 | 35.3 | 37 | 205 | 4.13 (2.91 - 5.70) | 5.81 (5.04 - 6.67) |
| ODOD | 7,683 | 27,663 | 4.5 | 21.5 | 51 | 463 | 11.4 (8.52 - 15.1) | 21.5 (19.6 - 23.6) |
| MDCR | 966 | 8,221 | 0.6 | 7.8 | 5 | 104 | 8.08 (2.62 - 18.8) | 13.4 (10.9 - 16.2) |
| MDCD | 1,807 | 7,452 | 1.0 | 5.2 | 8 | 92 | 8.10 (3.50 - 16.0) | 17.7 (14.3 - 21.7) |
| Open Claims | 142,284 | 438,407 | 103.5 | 382.1 | 341 | 2,955 | 3.29 (2.95 - 3.66) | 7.73 (7.46 - 8.02) |
| OEHR | 10,414 | 37,959 | 3.2 | 19.5 | 25 | 255 | 7.78 (5.04 - 11.5) | 13.1 (11.5 - 14.8) |
| Meta-analysis | 176,423 | 556,282 | 121.1 | 463.6 | 462 | 3,970 | 3.81 (3.47 - 4.18) | 8.56 (8.30 - 8.83) |
| Meta-analysis (w/o USOC) | 34,139 | 117,875 | 17.6 | 81.4 | 121 | 1,015 | 6.87 (5.70 - 8.21) | 12.5 (11.7 - 13.3) |
| **PS-stratification / on-treatment TAR** | | | | | | | | |
| CCAE | 19,147 | 44,818 | 11.3 | 35.3 | 47 | 206 | 4.17 (3.06 - 5.54) | 5.84 (5.07 - 6.69) |
| ODOD | 11,337 | 28,155 | 6.0 | 22.0 | 67 | 480 | 11.2 (8.67 - 14.2) | 21.9 (19.9 - 23.9) |
| MDCR | 1,105 | 8,538 | 0.7 | 8.0 | 6 | 119 | 9.13 (3.35 - 19.9) | 14.8 (12.3 - 17.8) |
| MDCD | 1,985 | 7,515 | 1.1 | 5.3 | 8 | 94 | 7.52 (3.25 - 14.8) | 17.9 (14.5 - 21.9) |
| Open Claims | 227,579 | 439,399 | 141.5 | 383.1 | 427 | 2,989 | 3.02 (2.74 - 3.32) | 7.80 (7.53 - 8.09) |
| OEHR | 13,435 | 38,147 | 4.0 | 19.6 | 30 | 261 | 7.57 (5.11 - 10.8) | 13.3 (11.8 - 15.1) |
| Meta-analysis | 273,483 | 558,034 | 163.8 | 465.1 | 579 | 4,030 | 3.53 (3.25 - 3.83) | 8.66 (8.40 - 8.94) |
| Meta-analysis (w/o USOC) | 45,904 | 118,635 | 22.3 | 82.1 | 152 | 1,041 | 6.82 (5.78 - 7.99) | 12.7 (11.9 - 13.5) |
| **PS-matching / on-treatment TAR without escalation** | | | | | | | | |
| CCAE | 14,235 | 44,801 | 8.0 | 29.0 | 33 | 166 | 4.11 (2.83 - 5.78) | 5.72 (4.88 - 6.66) |
| ODOD | 7,683 | 27,663 | 4.0 | 18.1 | 50 | 379 | 12.5 (9.28 - 16.5) | 21.0 (18.9 - 23.2) |
| MDCR | 966 | 8,221 | 0.6 | 6.8 | 5 | 91 | 8.75 (2.84 - 20.4) | 13.5 (10.8 - 16.5) |
| MDCD | 1,807 | 7,452 | 0.8 | 4.1 | 8 | 71 | 9.43 (4.07 - 18.6) | 17.5 (13.6 - 22.0) |
| Open Claims | 142,284 | 438,407 | 91.7 | 303.6 | 294 | 2,339 | 3.21 (2.85 - 3.59) | 7.71 (7.40 - 8.02) |
| OEHR | 10,414 | 37,959 | 2.8 | 15.7 | 23 | 190 | 8.32 (5.27 - 12.5) | 12.1 (10.4 - 14.0) |
| Meta-analysis | 176,423 | 556,282 | 107.4 | 370.4 | 408 | 3,145 | 3.80 (3.44 - 4.19) | 8.49 (8.20 - 8.79) |
| Meta-analysis (w/o USOC) | 34,139 | 117,875 | 15.6 | 66.9 | 114 | 806 | 7.29 (6.01 - 8.76) | 12.1 (11.2 - 12.9) |
| **PS-stratification / on-treatment TAR without escalation** | | | | | | | | |
| CCAE | 19,147 | 44,818 | 10.2 | 29.0 | 43 | 167 | 4.21 (3.05 - 5.67) | 5.75 (4.91 - 6.70) |
| ODOD | 11,337 | 28,155 | 5.4 | 18.4 | 64 | 395 | 11.8 (9.05 - 15.0) | 21.4 (19.4 - 23.6) |
| MDCR | 1,105 | 8,538 | 0.6 | 7.0 | 6 | 104 | 9.89 (3.63 - 21.5) | 14.9 (12.2 - 18.1) |
| MDCD | 1,985 | 7,515 | 0.9 | 4.1 | 8 | 73 | 8.71 (3.76 - 17.2) | 17.8 (13.9 - 22.3) |
| Open Claims | 227,579 | 439,399 | 127.7 | 304.3 | 374 | 2,370 | 2.93 (2.64 - 3.24) | 7.79 (7.48 - 8.11) |
| OEHR | 13,435 | 38,147 | 3.4 | 15.8 | 28 | 193 | 8.13 (5.40 - 11.7) | 12.2 (10.6 - 14.1) |
| Meta-analysis | 273,483 | 558,034 | 147.7 | 371.7 | 517 | 3,198 | 3.50 (3.20 - 3.81) | 8.60 (8.31 - 8.91) |
| Meta-analysis (w/o USOC) | 45,904 | 118,635 | 20.0 | 67.4 | 143 | 828 | 7.14 (6.02 - 8.42) | 12.3 (11.5 - 13.2) |

##### 4.4.18 Hypoglycemia

**Supplementary Table 381:** Exposures, outcomes and risk estimates for Hypoglycemia in GLP1RA (female) (T) vs DPP4I (female) (C) new-user cohorts under four different propensity score (PS) adjustment / time-at-risk (TAR) designs across data sources. We report total patient exposure time in 1,000 patient-year (KYr) units, uncalibrated and calibrated hazard ratios (HRs) and their 95% confidence intervals (CIs). Greyed out entries did not pass study diagnostics and are included only for completeness.

| | Patients | | Exposure (KYr) | | Outcomes | | Incidence (per KYr) | |
|---|---|---|---|---|---|---|---|---|
| | T | C | T | C | T | C | T | C |
| **PS-matching / on-treatment TAR** | | | | | | | | |
| CCAE | 14,052 | 44,595 | 8.9 | 35.3 | 41 | 120 | 4.62 (3.32 - 6.27) | 3.40 (2.82 - 4.07) |
| ODOD | 7,741 | 28,286 | 4.5 | 22.4 | 32 | 175 | 7.08 (4.84 - 9.99) | 7.82 (6.70 - 9.06) |
| MDCR | 972 | 8,349 | 0.6 | 7.9 | <5 | 42 | < 7.93 (2.58 - 18.5) | 5.31 (3.83 - 7.18) |
| MDCD | 1,787 | 7,432 | 1.0 | 5.3 | 9 | 58 | 9.16 (4.19 - 17.4) | 11.0 (8.36 - 14.2) |
| Open Claims | 141,638 | 440,239 | 103.2 | 386.3 | 284 | 1,258 | 2.75 (2.44 - 3.09) | 3.26 (3.08 - 3.44) |
| OEHR | 10,394 | 38,200 | 3.2 | 19.7 | 27 | 137 | 8.41 (5.54 - 12.2) | 6.95 (5.83 - 8.21) |
| Meta-analysis | 175,612 | 558,752 | 120.7 | 468.9 | 393 | 1,748 | 3.25 (2.94 - 3.59) | 3.73 (3.55 - 3.91) |
| Meta-analysis (w/o USOC) | 33,974 | 118,513 | 17.6 | 82.6 | 109 | 490 | 6.20 (5.09 - 7.48) | 5.93 (5.42 - 6.48) |
| **PS-stratification / on-treatment TAR** | | | | | | | | |
| CCAE | 18,798 | 44,599 | 11.1 | 35.3 | 60 | 120 | 5.39 (4.11 - 6.94) | 3.40 (2.82 - 4.07) |
| ODOD | 11,351 | 28,947 | 6.0 | 23.0 | 56 | 182 | 9.30 (7.02 - 12.1) | 7.91 (6.80 - 9.15) |
| MDCR | 1,110 | 8,701 | 0.7 | 8.2 | <5 | 45 | < 7.48 (2.43 - 17.5) | 5.50 (4.01 - 7.36) |
| MDCD | 1,953 | 7,505 | 1.1 | 5.3 | 9 | 60 | 8.56 (3.92 - 16.3) | 11.3 (8.60 - 14.5) |
| Open Claims | 225,048 | 441,240 | 140.3 | 387.3 | 401 | 1,269 | 2.86 (2.59 - 3.15) | 3.28 (3.10 - 3.46) |
| OEHR | 13,387 | 38,401 | 3.9 | 19.8 | 31 | 139 | 7.86 (5.34 - 11.2) | 7.01 (5.89 - 8.27) |
| Meta-analysis | 270,537 | 560,692 | 162.5 | 470.7 | 557 | 1,770 | 3.43 (3.15 - 3.73) | 3.76 (3.59 - 3.94) |
| Meta-analysis (w/o USOC) | 45,489 | 119,452 | 22.1 | 83.4 | 156 | 501 | 7.04 (5.98 - 8.24) | 6.00 (5.49 - 6.55) |
| **PS-matching / on-treatment TAR without escalation** | | | | | | | | |
| CCAE | 14,052 | 44,595 | 8.0 | 29.0 | 34 | 70 | 4.28 (2.96 - 5.98) | 2.41 (1.88 - 3.05) |
| ODOD | 7,741 | 28,286 | 4.1 | 18.8 | 23 | 96 | 5.67 (3.59 - 8.50) | 5.10 (4.13 - 6.23) |
| MDCR | 972 | 8,349 | 0.6 | 6.9 | <5 | 26 | < 8.63 (2.80 - 20.1) | 3.78 (2.47 - 5.54) |
| MDCD | 1,787 | 7,432 | 0.8 | 4.1 | 7 | 32 | 8.24 (3.31 - 17.0) | 7.80 (5.33 - 11.0) |
| Open Claims | 141,638 | 440,239 | 91.4 | 307.1 | 233 | 685 | 2.55 (2.23 - 2.90) | 2.23 (2.07 - 2.40) |
| OEHR | 10,394 | 38,200 | 2.8 | 15.9 | 20 | 66 | 7.23 (4.41 - 11.2) | 4.15 (3.21 - 5.28) |
| Meta-analysis | 175,612 | 558,752 | 107.1 | 374.9 | 317 | 949 | 2.96 (2.64 - 3.31) | 2.53 (2.37 - 2.70) |
| Meta-analysis (w/o USOC) | 33,974 | 118,513 | 15.6 | 67.8 | 84 | 264 | 5.38 (4.29 - 6.66) | 3.89 (3.44 - 4.39) |
| **PS-stratification / on-treatment TAR without escalation** | | | | | | | | |
| CCAE | 18,798 | 44,599 | 10.1 | 29.0 | 52 | 70 | 5.17 (3.86 - 6.77) | 2.41 (1.88 - 3.05) |
| ODOD | 11,351 | 28,947 | 5.5 | 19.3 | 46 | 101 | 8.40 (6.15 - 11.2) | 5.22 (4.25 - 6.35) |
| MDCR | 1,110 | 8,701 | 0.6 | 7.1 | <5 | 28 | < 8.11 (2.63 - 18.9) | 3.94 (2.62 - 5.69) |
| MDCD | 1,953 | 7,505 | 0.9 | 4.2 | 7 | 33 | 7.68 (3.09 - 15.8) | 7.94 (5.47 - 11.2) |
| Open Claims | 225,048 | 441,240 | 126.6 | 307.9 | 337 | 690 | 2.66 (2.39 - 2.96) | 2.24 (2.08 - 2.41) |
| OEHR | 13,387 | 38,401 | 3.4 | 16.0 | 24 | 66 | 6.99 (4.48 - 10.4) | 4.13 (3.19 - 5.25) |
| Meta-analysis | 270,537 | 560,692 | 146.5 | 376.4 | 466 | 960 | 3.18 (2.90 - 3.48) | 2.55 (2.39 - 2.72) |
| Meta-analysis (w/o USOC) | 45,489 | 119,452 | 19.9 | 68.5 | 129 | 270 | 6.49 (5.42 - 7.71) | 3.94 (3.49 - 4.44) |

##### 4.4.19 Hypotension

**Supplementary Table 382:** Exposures, outcomes and risk estimates for Hypotension in GLP1RA (female) (T) vs DPP4I (female) (C) new-user cohorts under four different propensity score (PS) adjustment / time-at-risk (TAR) designs across data sources. We report total patient exposure time in 1,000 patient-year (KYr) units, uncalibrated and calibrated hazard ratios (HRs) and their 95% confidence intervals (CIs). Greyed out entries did not pass study diagnostics and are included only for completeness.

| | Patients | | Exposure (KYr) | | Outcomes | | Incidence (per KYr) | |
|---|---|---|---|---|---|---|---|---|
| | T | C | T | C | T | C | T | C |
| **PS-matching / on-treatment TAR** | | | | | | | | |
| CCAE | 14,152 | 44,621 | 8.9 | 35.2 | 72 | 257 | 8.11 (6.34 - 10.2) | 7.31 (6.44 - 8.26) |
| ODOD | 7,602 | 27,545 | 4.4 | 21.6 | 74 | 461 | 16.8 (13.2 - 21.1) | 21.4 (19.5 - 23.4) |
| MDCR | 934 | 8,043 | 0.6 | 7.6 | 9 | 155 | 14.7 (6.72 - 27.9) | 20.3 (17.3 - 23.8) |
| MDCD | 1,753 | 7,149 | 0.9 | 5.0 | 28 | 151 | 29.9 (19.8 - 43.2) | 30.2 (25.6 - 35.5) |
| Open Claims | 139,697 | 431,616 | 101.4 | 376.8 | 697 | 3,615 | 6.87 (6.37 - 7.40) | 9.59 (9.28 - 9.91) |
| OEHR | 10,378 | 37,870 | 3.2 | 19.4 | 52 | 281 | 16.3 (12.2 - 21.4) | 14.5 (12.8 - 16.3) |
| Meta-analysis | 173,582 | 548,801 | 118.8 | 457.9 | 923 | 4,765 | 7.77 (7.27 - 8.28) | 10.4 (10.1 - 10.7) |
| Meta-analysis (w/o USOC) | 33,885 | 117,185 | 17.4 | 81.1 | 226 | 1,150 | 13.0 (11.4 - 14.8) | 14.2 (13.4 - 15.0) |
| **PS-stratification / on-treatment TAR** | | | | | | | | |
| CCAE | 19,037 | 44,624 | 11.2 | 35.2 | 98 | 257 | 8.75 (7.11 - 10.7) | 7.30 (6.44 - 8.25) |
| ODOD | 11,208 | 28,041 | 5.9 | 22.0 | 103 | 486 | 17.4 (14.2 - 21.1) | 22.1 (20.1 - 24.1) |
| MDCR | 1,073 | 8,338 | 0.6 | 7.8 | 9 | 172 | 13.9 (6.34 - 26.3) | 21.9 (18.8 - 25.5) |
| MDCD | 1,922 | 7,199 | 1.0 | 5.0 | 28 | 153 | 27.7 (18.4 - 40.0) | 30.4 (25.7 - 35.6) |
| Open Claims | 223,218 | 432,712 | 138.6 | 377.9 | 939 | 3,663 | 6.77 (6.35 - 7.22) | 9.69 (9.38 - 10.0) |
| OEHR | 13,380 | 38,051 | 3.9 | 19.5 | 60 | 287 | 15.3 (11.7 - 19.7) | 14.7 (13.1 - 16.5) |
| Meta-analysis | 268,765 | 550,627 | 160.7 | 459.6 | 1,228 | 4,846 | 7.64 (7.22 - 8.08) | 10.5 (10.2 - 10.8) |
| Meta-analysis (w/o USOC) | 45,547 | 117,915 | 22.0 | 81.7 | 289 | 1,183 | 13.1 (11.6 - 14.7) | 14.5 (13.7 - 15.3) |
| **PS-matching / on-treatment TAR without escalation** | | | | | | | | |
| CCAE | 14,152 | 44,621 | 8.0 | 29.0 | 62 | 202 | 7.79 (5.97 - 9.98) | 6.98 (6.05 - 8.01) |
| ODOD | 7,602 | 27,545 | 4.0 | 18.1 | 65 | 386 | 16.4 (12.7 - 20.9) | 21.3 (19.2 - 23.5) |
| MDCR | 934 | 8,043 | 0.6 | 6.6 | 9 | 136 | 16.0 (7.30 - 30.3) | 20.5 (17.2 - 24.2) |
| MDCD | 1,753 | 7,149 | 0.8 | 3.9 | 23 | 103 | 28.5 (18.1 - 42.7) | 26.2 (21.4 - 31.8) |
| Open Claims | 139,697 | 431,616 | 89.9 | 299.5 | 584 | 2,874 | 6.49 (5.98 - 7.04) | 9.60 (9.25 - 9.95) |
| OEHR | 10,378 | 37,870 | 2.7 | 15.7 | 41 | 194 | 14.9 (10.7 - 20.2) | 12.4 (10.7 - 14.3) |
| Meta-analysis | 173,582 | 548,801 | 105.4 | 366.2 | 775 | 3,759 | 7.35 (6.84 - 7.89) | 10.3 (9.94 - 10.6) |
| Meta-analysis (w/o USOC) | 33,885 | 117,185 | 15.5 | 66.7 | 191 | 885 | 12.3 (10.7 - 14.2) | 13.3 (12.4 - 14.2) |
| **PS-stratification / on-treatment TAR without escalation** | | | | | | | | |
| CCAE | 19,037 | 44,624 | 10.1 | 29.0 | 87 | 202 | 8.59 (6.88 - 10.6) | 6.98 (6.05 - 8.01) |
| ODOD | 11,208 | 28,041 | 5.4 | 18.5 | 93 | 410 | 17.3 (13.9 - 21.2) | 22.1 (20.0 - 24.4) |
| MDCR | 1,073 | 8,338 | 0.6 | 6.8 | 9 | 151 | 15.0 (6.86 - 28.5) | 22.1 (18.7 - 25.9) |
| MDCD | 1,922 | 7,199 | 0.9 | 4.0 | 23 | 104 | 26.2 (16.6 - 39.4) | 26.2 (21.4 - 31.8) |
| Open Claims | 223,218 | 432,712 | 125.1 | 300.4 | 809 | 2,916 | 6.47 (6.03 - 6.93) | 9.71 (9.36 - 10.1) |
| OEHR | 13,380 | 38,051 | 3.4 | 15.8 | 47 | 200 | 13.8 (10.1 - 18.3) | 12.7 (11.0 - 14.6) |
| Meta-analysis | 268,765 | 550,627 | 144.9 | 367.6 | 1,059 | 3,832 | 7.31 (6.87 - 7.76) | 10.4 (10.1 - 10.8) |
| Meta-analysis (w/o USOC) | 45,547 | 117,915 | 19.8 | 67.2 | 250 | 916 | 12.6 (11.1 - 14.3) | 13.6 (12.8 - 14.5) |

##### 4.4.20 Joint pain

**Supplementary Table 383:** Exposures, outcomes and risk estimates for Joint pain in GLP1RA (female) (T) vs DPP4I (female) (C) new-user cohorts under four different propensity score (PS) adjustment / time-at-risk (TAR) designs across data sources. We report total patient exposure time in 1,000 patient-year (KYr) units, uncalibrated and calibrated hazard ratios (HRs) and their 95% confidence intervals (CIs). Greyed out entries did not pass study diagnostics and are included only for completeness.

| | Patients | | Exposure (KYr) | | Outcomes | | Incidence (per KYr) | |
|---|---|---|---|---|---|---|---|---|
| | T | C | T | C | T | C | T | C |
| **PS-matching / on-treatment TAR** | | | | | | | | |
| CCAE | 13,190 | 42,276 | 8.2 | 33.2 | 128 | 487 | 15.6 (13.0 - 18.6) | 14.7 (13.4 - 16.0) |
| ODOD | 7,217 | 26,640 | 4.2 | 20.9 | 83 | 396 | 19.9 (15.9 - 24.7) | 19.0 (17.1 - 20.9) |
| MDCR | 918 | 8,004 | 0.6 | 7.6 | 5 | 97 | 8.31 (2.70 - 19.4) | 12.8 (10.4 - 15.6) |
| MDCD | 1,623 | 6,807 | 0.9 | 4.7 | 24 | 145 | 27.7 (17.8 - 41.2) | 30.9 (26.1 - 36.4) |
| Open Claims | 131,575 | 413,816 | 95.6 | 361.2 | 780 | 3,517 | 8.16 (7.60 - 8.75) | 9.74 (9.42 - 10.1) |
| OEHR | 9,737 | 36,169 | 3.0 | 18.4 | 51 | 310 | 17.3 (12.9 - 22.7) | 16.9 (15.0 - 18.8) |
| Meta-analysis | 163,342 | 525,708 | 111.8 | 438.3 | 1,066 | 4,855 | 9.54 (8.97 - 10.1) | 11.1 (10.8 - 11.4) |
| Meta-analysis (w/o USOC) | 31,767 | 111,892 | 16.2 | 77.2 | 286 | 1,338 | 17.7 (15.7 - 19.8) | 17.3 (16.4 - 18.3) |
| **PS-stratification / on-treatment TAR** | | | | | | | | |
| CCAE | 17,578 | 42,281 | 10.3 | 33.2 | 163 | 487 | 15.9 (13.5 - 18.5) | 14.7 (13.4 - 16.0) |
| ODOD | 10,448 | 27,243 | 5.5 | 21.4 | 109 | 412 | 19.8 (16.3 - 23.9) | 19.2 (17.4 - 21.2) |
| MDCR | 1,038 | 8,335 | 0.6 | 7.8 | 5 | 100 | 7.89 (2.56 - 18.4) | 12.8 (10.4 - 15.5) |
| MDCD | 1,769 | 6,865 | 0.9 | 4.7 | 28 | 148 | 30.1 (20.0 - 43.6) | 31.3 (26.4 - 36.7) |
| Open Claims | 207,745 | 415,056 | 129.4 | 362.4 | 1,119 | 3,534 | 8.65 (8.15 - 9.17) | 9.75 (9.43 - 10.1) |
| OEHR | 12,431 | 36,357 | 3.6 | 18.5 | 66 | 310 | 18.3 (14.2 - 23.3) | 16.8 (14.9 - 18.7) |
| Meta-analysis | 249,971 | 527,802 | 149.7 | 440.3 | 1,485 | 4,891 | 9.92 (9.42 - 10.4) | 11.1 (10.8 - 11.4) |
| Meta-analysis (w/o USOC) | 42,226 | 112,746 | 20.3 | 77.9 | 366 | 1,357 | 18.0 (16.2 - 20.0) | 17.4 (16.5 - 18.4) |
| **PS-matching / on-treatment TAR without escalation** | | | | | | | | |
| CCAE | 13,190 | 42,276 | 7.4 | 27.4 | 112 | 402 | 15.2 (12.5 - 18.3) | 14.7 (13.3 - 16.2) |
| ODOD | 7,217 | 26,640 | 3.7 | 17.6 | 70 | 330 | 18.7 (14.6 - 23.6) | 18.8 (16.8 - 20.9) |
| MDCR | 918 | 8,004 | 0.6 | 6.6 | 5 | 84 | 9.04 (2.94 - 21.1) | 12.7 (10.2 - 15.8) |
| MDCD | 1,623 | 6,807 | 0.7 | 3.7 | 22 | 113 | 29.5 (18.5 - 44.7) | 30.9 (25.4 - 37.1) |
| Open Claims | 131,575 | 413,816 | 84.6 | 287.2 | 689 | 2,792 | 8.14 (7.55 - 8.77) | 9.72 (9.36 - 10.1) |
| OEHR | 9,737 | 36,169 | 2.6 | 14.9 | 42 | 241 | 16.4 (11.8 - 22.2) | 16.2 (14.2 - 18.4) |
| Meta-analysis | 163,342 | 525,708 | 99.0 | 350.6 | 935 | 3,878 | 9.44 (8.85 - 10.1) | 11.1 (10.7 - 11.4) |
| Meta-analysis (w/o USOC) | 31,767 | 111,892 | 14.4 | 63.4 | 246 | 1,086 | 17.1 (15.0 - 19.3) | 17.1 (16.1 - 18.2) |
| **PS-stratification / on-treatment TAR without escalation** | | | | | | | | |
| CCAE | 17,578 | 42,281 | 9.3 | 27.4 | 143 | 402 | 15.4 (13.0 - 18.1) | 14.7 (13.3 - 16.2) |
| ODOD | 10,448 | 27,243 | 5.0 | 18.0 | 94 | 345 | 18.8 (15.2 - 23.0) | 19.2 (17.2 - 21.3) |
| MDCR | 1,038 | 8,335 | 0.6 | 6.8 | 5 | 87 | 8.56 (2.78 - 20.0) | 12.8 (10.2 - 15.7) |
| MDCD | 1,769 | 6,865 | 0.8 | 3.7 | 26 | 115 | 32.3 (21.1 - 47.4) | 31.1 (25.7 - 37.4) |
| Open Claims | 207,745 | 415,056 | 116.6 | 288.2 | 1,006 | 2,807 | 8.63 (8.11 - 9.18) | 9.74 (9.38 - 10.1) |
| OEHR | 12,431 | 36,357 | 3.2 | 14.9 | 57 | 241 | 18.1 (13.7 - 23.4) | 16.1 (14.2 - 18.3) |
| Meta-analysis | 249,971 | 527,802 | 134.8 | 352.2 | 1,326 | 3,910 | 9.83 (9.31 - 10.4) | 11.1 (10.8 - 11.5) |
| Meta-analysis (w/o USOC) | 42,226 | 112,746 | 18.3 | 64.0 | 320 | 1,103 | 17.5 (15.7 - 19.5) | 17.2 (16.2 - 18.3) |

##### 4.4.21 Lower extremity amputation

**Supplementary Table 384:** Exposures, outcomes and risk estimates for Lower extremity amputation in GLP1RA (female) (T) vs DPP4I (female) (C) new-user cohorts under four different propensity score (PS) adjustment / time-at-risk (TAR) designs across data sources. We report total patient exposure time in 1,000 patient-year (KYr) units, uncalibrated and calibrated hazard ratios (HRs) and their 95% confidence intervals (CIs). Greyed out entries did not pass study diagnostics and are included only for completeness.

| | Patients | | Exposure (KYr) | | Outcomes | | Incidence (per KYr) | |
|---|---|---|---|---|---|---|---|---|
| | T | C | T | C | T | C | T | C |
| **PS-matching / on-treatment TAR** | | | | | | | | |
| CCAE | 14,428 | 45,476 | 9.1 | 36.0 | <5 | 10 | < 0.55 (0.18 - 1.28) | 0.28 (0.13 - 0.51) |
| ODOD | 7,911 | 28,787 | 4.6 | 22.8 | <5 | 10 | < 1.08 (0.35 - 2.53) | 0.44 (0.21 - 0.81) |
| MDCR | 985 | 8,426 | 0.6 | 8.0 | 0 | 5 | | 0.62 (0.20 - 1.46) |
| MDCD | 1,858 | 7,679 | 1.0 | 5.5 | <5 | 5 | < 4.92 (1.60 - 11.5) | 0.91 (0.30 - 2.13) |
| Open Claims | 144,729 | 448,280 | 105.5 | 393.9 | 19 | 107 | 0.18 (0.11 - 0.28) | 0.27 (0.22 - 0.33) |
| OEHR | 10,560 | 38,681 | 3.3 | 20.1 | 0 | 7 | | 0.35 (0.14 - 0.72) |
| Meta-analysis | 144,729 | 448,280 | 105.5 | 393.9 | 19 | 107 | 0.18 (0.11 - 0.28) | 0.27 (0.22 - 0.33) |
| **PS-stratification / on-treatment TAR** | | | | | | | | |
| CCAE | 19,436 | 45,481 | 11.5 | 36.0 | <5 | 10 | < 0.44 (0.14 - 1.02) | 0.28 (0.13 - 0.51) |
| ODOD | 11,652 | 29,479 | 6.2 | 23.5 | <5 | 10 | < 0.81 (0.26 - 1.88) | 0.43 (0.20 - 0.78) |
| MDCR | 1,127 | 8,789 | 0.7 | 8.3 | 0 | 6 | | 0.72 (0.27 - 1.57) |
| MDCD | 2,031 | 7,756 | 1.1 | 5.5 | <5 | 5 | < 4.59 (1.49 - 10.7) | 0.90 (0.29 - 2.11) |
| Open Claims | 231,103 | 449,385 | 144.0 | 395.0 | 22 | 110 | 0.15 (0.10 - 0.23) | 0.28 (0.23 - 0.34) |
| OEHR | 13,629 | 38,885 | 4.0 | 20.2 | 0 | 7 | | 0.35 (0.14 - 0.71) |
| Meta-analysis | 231,103 | 449,385 | 144.0 | 395.0 | 22 | 110 | 0.15 (0.10 - 0.23) | 0.28 (0.23 - 0.34) |
| **PS-matching / on-treatment TAR without escalation** | | | | | | | | |
| CCAE | 14,428 | 45,476 | 8.2 | 29.6 | <5 | 9 | < 0.61 (0.20 - 1.43) | 0.30 (0.14 - 0.58) |
| ODOD | 7,911 | 28,787 | 4.1 | 19.1 | <5 | 8 | < 1.21 (0.39 - 2.82) | 0.42 (0.18 - 0.82) |
| MDCR | 985 | 8,426 | 0.6 | 7.0 | 0 | 5 | | 0.72 (0.23 - 1.68) |
| MDCD | 1,858 | 7,679 | 0.9 | 4.3 | <5 | <5 | < 5.73 (1.86 - 13.4) | < 1.17 (0.38 - 2.74) |
| Open Claims | 144,729 | 448,280 | 93.4 | 312.7 | 13 | 85 | 0.14 (0.07 - 0.24) | 0.27 (0.22 - 0.34) |
| OEHR | 10,560 | 38,681 | 2.8 | 16.2 | 0 | <5 | | < 0.31 (0.10 - 0.72) |
| Meta-analysis | 144,729 | 448,280 | 93.4 | 312.7 | 13 | 85 | 0.14 (0.07 - 0.24) | 0.27 (0.22 - 0.34) |
| **PS-stratification / on-treatment TAR without escalation** | | | | | | | | |
| CCAE | 19,436 | 45,481 | 10.4 | 29.6 | <5 | 9 | < 0.48 (0.16 - 1.12) | 0.30 (0.14 - 0.58) |
| ODOD | 11,652 | 29,479 | 5.6 | 19.7 | <5 | 8 | < 0.89 (0.29 - 2.07) | 0.41 (0.18 - 0.80) |
| MDCR | 1,127 | 8,789 | 0.6 | 7.2 | 0 | 6 | | 0.83 (0.31 - 1.81) |
| MDCD | 2,031 | 7,756 | 0.9 | 4.3 | <5 | <5 | < 5.32 (1.73 - 12.4) | < 1.16 (0.38 - 2.71) |
| Open Claims | 231,103 | 449,385 | 129.9 | 313.6 | 16 | 87 | 0.12 (0.07 - 0.20) | 0.28 (0.22 - 0.34) |
| OEHR | 13,629 | 38,885 | 3.5 | 16.3 | 0 | <5 | | < 0.31 (0.10 - 0.72) |
| Meta-analysis | 231,103 | 449,385 | 129.9 | 313.6 | 16 | 87 | 0.12 (0.07 - 0.20) | 0.28 (0.22 - 0.34) |

##### 4.4.22 Nausea

**Supplementary Table 385:** Exposures, outcomes and risk estimates for Nausea in GLP1RA (female) (T) vs DPP4I (female) (C) new-user cohorts under four different propensity score (PS) adjustment / time-at-risk (TAR) designs across data sources. We report total patient exposure time in 1,000 patient-year (KYr) units, uncalibrated and calibrated hazard ratios (HRs) and their 95% confidence intervals (CIs). Greyed out entries did not pass study diagnostics and are included only for completeness.

| | Patients | | Exposure (KYr) | | Outcomes | | Incidence (per KYr) | |
|---|---|---|---|---|---|---|---|---|
| | T | C | T | C | T | C | T | C |
| **PS-matching / on-treatment TAR** | | | | | | | | |
| CCAE | 11,842 | 38,596 | 7.2 | 30.0 | 441 | 1,285 | 61.1 (55.6 - 67.1) | 42.8 (40.5 - 45.2) |
| ODOD | 6,280 | 23,313 | 3.5 | 18.0 | 306 | 972 | 86.4 (76.9 - 96.6) | 54.1 (50.7 - 57.6) |
| MDCR | 882 | 7,237 | 0.5 | 6.8 | 29 | 306 | 52.7 (35.3 - 75.7) | 45.3 (40.3 - 50.6) |
| MDCD | 1,189 | 5,111 | 0.6 | 3.4 | 98 | 403 | 155 (126 - 189) | 117 (106 - 130) |
| Open Claims | 112,939 | 366,334 | 81.8 | 319.0 | 3,158 | 9,386 | 38.6 (37.3 - 40.0) | 29.4 (28.8 - 30.0) |
| OEHR | 8,753 | 33,160 | 2.6 | 16.5 | 226 | 833 | 85.5 (74.7 - 97.4) | 50.3 (47.0 - 53.9) |
| Meta-analysis | 141,003 | 466,514 | 95.8 | 386.9 | 4,229 | 12,879 | 44.1 (42.8 - 45.5) | 33.3 (32.7 - 33.9) |
| Meta-analysis (w/o USOC) | 28,064 | 100,180 | 14.0 | 68.0 | 1,071 | 3,493 | 76.3 (71.8 - 81.0) | 51.4 (49.7 - 53.1) |
| **PS-stratification / on-treatment TAR** | | | | | | | | |
| CCAE | 15,622 | 38,599 | 8.9 | 30.0 | 606 | 1,285 | 67.7 (62.5 - 73.4) | 42.8 (40.5 - 45.2) |
| ODOD | 9,065 | 23,886 | 4.7 | 18.5 | 450 | 1,003 | 96.5 (87.8 - 106) | 54.3 (51.0 - 57.7) |
| MDCR | 1,005 | 7,552 | 0.6 | 7.0 | 33 | 317 | 56.4 (38.8 - 79.1) | 45.2 (40.4 - 50.5) |
| MDCD | 1,275 | 5,142 | 0.7 | 3.4 | 106 | 404 | 161 (132 - 195) | 117 (106 - 129) |
| Open Claims | 175,888 | 367,383 | 109.5 | 320.0 | 4,651 | 9,435 | 42.5 (41.2 - 43.7) | 29.5 (28.9 - 30.1) |
| OEHR | 11,092 | 33,348 | 3.2 | 16.7 | 290 | 842 | 91.3 (81.1 - 102) | 50.6 (47.2 - 54.1) |
| Meta-analysis | 212,942 | 468,358 | 127.0 | 388.6 | 6,103 | 12,969 | 48.1 (46.9 - 49.3) | 33.4 (32.8 - 34.0) |
| Meta-analysis (w/o USOC) | 37,054 | 100,975 | 17.4 | 68.6 | 1,452 | 3,534 | 83.2 (79.0 - 87.6) | 51.5 (49.8 - 53.3) |
| **PS-matching / on-treatment TAR without escalation** | | | | | | | | |
| CCAE | 11,842 | 38,596 | 6.5 | 24.9 | 403 | 1,058 | 61.9 (56.0 - 68.2) | 42.5 (40.0 - 45.1) |
| ODOD | 6,280 | 23,313 | 3.2 | 15.2 | 280 | 821 | 87.7 (77.7 - 98.6) | 53.9 (50.3 - 57.7) |
| MDCR | 882 | 7,237 | 0.5 | 5.9 | 27 | 261 | 52.8 (34.8 - 76.8) | 44.3 (39.1 - 50.1) |
| MDCD | 1,189 | 5,111 | 0.6 | 2.8 | 89 | 316 | 162 (130 - 199) | 115 (102 - 128) |
| Open Claims | 112,939 | 366,334 | 72.7 | 255.3 | 2,808 | 7,440 | 38.6 (37.2 - 40.1) | 29.1 (28.5 - 29.8) |
| OEHR | 8,753 | 33,160 | 2.3 | 13.5 | 196 | 630 | 85.5 (74.0 - 98.4) | 46.8 (43.2 - 50.6) |
| Meta-analysis | 141,003 | 466,514 | 85.2 | 311.6 | 3,776 | 10,265 | 44.3 (42.9 - 45.7) | 32.9 (32.3 - 33.6) |
| Meta-analysis (w/o USOC) | 28,064 | 100,180 | 12.5 | 56.3 | 968 | 2,825 | 77.1 (72.4 - 82.2) | 50.1 (48.3 - 52.0) |
| **PS-stratification / on-treatment TAR without escalation** | | | | | | | | |
| CCAE | 15,622 | 38,599 | 8.1 | 24.9 | 557 | 1,058 | 68.4 (62.8 - 74.3) | 42.5 (40.0 - 45.1) |
| ODOD | 9,065 | 23,886 | 4.3 | 15.7 | 420 | 846 | 98.7 (89.5 - 109) | 54.0 (50.4 - 57.7) |
| MDCR | 1,005 | 7,552 | 0.5 | 6.1 | 30 | 271 | 54.8 (37.0 - 78.3) | 44.3 (39.2 - 49.9) |
| MDCD | 1,275 | 5,142 | 0.6 | 2.8 | 97 | 316 | 169 (137 - 206) | 114 (102 - 128) |
| Open Claims | 175,888 | 367,383 | 99.0 | 256.1 | 4,235 | 7,482 | 42.8 (41.5 - 44.1) | 29.2 (28.6 - 29.9) |
| OEHR | 11,092 | 33,348 | 2.8 | 13.6 | 253 | 638 | 91.2 (80.3 - 103) | 47.1 (43.5 - 50.9) |
| Meta-analysis | 212,942 | 468,358 | 114.8 | 313.0 | 5,562 | 10,340 | 48.5 (47.2 - 49.8) | 33.0 (32.4 - 33.7) |
| Meta-analysis (w/o USOC) | 37,054 | 100,975 | 15.7 | 56.9 | 1,327 | 2,858 | 84.3 (79.8 - 88.9) | 50.2 (48.4 - 52.1) |

##### 4.4.23 Peripheral edema

**Supplementary Table 386:** Exposures, outcomes and risk estimates for Peripheral edema in GLP1RA (female) (T) vs DPP4I (female) (C) new-user cohorts under four different propensity score (PS) adjustment / time-at-risk (TAR) designs across data sources. We report total patient exposure time in 1,000 patient-year (KYr) units, uncalibrated and calibrated hazard ratios (HRs) and their 95% confidence intervals (CIs). Greyed out entries did not pass study diagnostics and are included only for completeness.

| | Patients | | Exposure (KYr) | | Outcomes | | Incidence (per KYr) | |
|---|---|---|---|---|---|---|---|---|
| | T | C | T | C | T | C | T | C |
| **PS-matching / on-treatment TAR** | | | | | | | | |
| CCAE | 10,948 | 35,719 | 6.7 | 27.2 | 331 | 1,396 | 49.3 (44.1 - 54.9) | 51.4 (48.7 - 54.1) |
| ODOD | 5,713 | 20,985 | 3.2 | 15.4 | 189 | 1,413 | 58.8 (50.7 - 67.9) | 92.0 (87.3 - 96.9) |
| MDCR | 710 | 6,031 | 0.4 | 5.4 | 29 | 487 | 65.2 (43.7 - 93.7) | 90.0 (82.2 - 98.3) |
| MDCD | 1,168 | 4,889 | 0.6 | 3.1 | 56 | 369 | 88.6 (66.9 - 115) | 120 (108 - 133) |
| Open Claims | 102,964 | 329,300 | 73.9 | 279.2 | 1,937 | 11,266 | 26.2 (25.0 - 27.4) | 40.3 (39.6 - 41.1) |
| OEHR | 8,443 | 31,545 | 2.5 | 15.4 | 126 | 849 | 49.9 (41.6 - 59.4) | 55.2 (51.5 - 59.0) |
| Meta-analysis | 129,236 | 422,438 | 87.0 | 340.2 | 2,639 | 15,293 | 30.3 (29.2 - 31.5) | 44.9 (44.2 - 45.7) |
| Meta-analysis (w/o USOC) | 26,272 | 93,138 | 13.1 | 61.0 | 702 | 4,027 | 53.6 (49.7 - 57.8) | 66.0 (64.0 - 68.1) |
| **PS-stratification / on-treatment TAR** | | | | | | | | |
| CCAE | 14,476 | 35,723 | 8.3 | 27.2 | 411 | 1,396 | 49.4 (44.7 - 54.4) | 51.4 (48.7 - 54.1) |
| ODOD | 8,358 | 21,408 | 4.3 | 15.7 | 261 | 1,486 | 61.0 (53.8 - 68.9) | 94.7 (89.9 - 99.6) |
| MDCR | 804 | 6,251 | 0.5 | 5.6 | 32 | 516 | 68.1 (46.6 - 96.2) | 92.8 (84.9 - 101) |
| MDCD | 1,281 | 4,930 | 0.7 | 3.1 | 60 | 377 | 88.9 (67.9 - 114) | 121 (109 - 134) |
| Open Claims | 160,813 | 329,848 | 99.2 | 279.7 | 2,532 | 11,328 | 25.5 (24.5 - 26.5) | 40.5 (39.8 - 41.3) |
| OEHR | 10,620 | 31,683 | 3.0 | 15.5 | 147 | 858 | 48.6 (41.1 - 57.1) | 55.5 (51.8 - 59.3) |
| Meta-analysis | 195,548 | 423,592 | 115.5 | 341.2 | 3,411 | 15,445 | 29.5 (28.5 - 30.5) | 45.3 (44.6 - 46.0) |
| Meta-analysis (w/o USOC) | 34,735 | 93,744 | 16.3 | 61.4 | 879 | 4,117 | 53.9 (50.4 - 57.6) | 67.0 (65.0 - 69.1) |
| **PS-matching / on-treatment TAR without escalation** | | | | | | | | |
| CCAE | 10,948 | 35,719 | 6.0 | 22.6 | 299 | 1,160 | 49.5 (44.1 - 55.5) | 51.4 (48.5 - 54.4) |
| ODOD | 5,713 | 20,985 | 2.9 | 13.0 | 175 | 1,167 | 60.1 (51.5 - 69.6) | 89.8 (84.8 - 95.1) |
| MDCR | 710 | 6,031 | 0.4 | 4.8 | 28 | 425 | 68.7 (45.7 - 99.4) | 89.4 (81.1 - 98.3) |
| MDCD | 1,168 | 4,889 | 0.5 | 2.5 | 45 | 289 | 82.4 (60.1 - 110) | 117 (104 - 131) |
| Open Claims | 102,964 | 329,300 | 65.5 | 223.6 | 1,655 | 8,960 | 25.3 (24.1 - 26.5) | 40.1 (39.2 - 40.9) |
| OEHR | 8,443 | 31,545 | 2.2 | 12.6 | 115 | 649 | 52.6 (43.5 - 63.2) | 51.7 (47.8 - 55.8) |
| Meta-analysis | 129,236 | 422,438 | 77.2 | 274.2 | 2,289 | 12,225 | 29.6 (28.4 - 30.9) | 44.6 (43.8 - 45.4) |
| Meta-analysis (w/o USOC) | 26,272 | 93,138 | 11.7 | 50.6 | 634 | 3,265 | 54.3 (50.1 - 58.7) | 64.5 (62.3 - 66.8) |
| **PS-stratification / on-treatment TAR without escalation** | | | | | | | | |
| CCAE | 14,476 | 35,723 | 7.6 | 22.6 | 368 | 1,160 | 48.7 (43.9 - 54.0) | 51.4 (48.5 - 54.4) |
| ODOD | 8,358 | 21,408 | 3.9 | 13.3 | 239 | 1,231 | 60.9 (53.5 - 69.2) | 92.7 (87.6 - 98.0) |
| MDCR | 804 | 6,251 | 0.4 | 4.9 | 31 | 449 | 71.8 (48.8 - 102) | 91.9 (83.6 - 101) |
| MDCD | 1,281 | 4,930 | 0.6 | 2.5 | 49 | 295 | 83.6 (61.8 - 111) | 118 (105 - 133) |
| Open Claims | 160,813 | 329,848 | 89.5 | 224.0 | 2,215 | 9,012 | 24.7 (23.7 - 25.8) | 40.2 (39.4 - 41.1) |
| OEHR | 10,620 | 31,683 | 2.6 | 12.6 | 135 | 658 | 51.1 (42.8 - 60.5) | 52.1 (48.2 - 56.3) |
| Meta-analysis | 195,548 | 423,592 | 104.2 | 274.9 | 3,006 | 12,356 | 28.8 (27.8 - 29.9) | 44.9 (44.2 - 45.7) |
| Meta-analysis (w/o USOC) | 34,735 | 93,744 | 14.7 | 51.0 | 791 | 3,344 | 53.8 (50.1 - 57.7) | 65.6 (63.4 - 67.9) |

##### 4.4.24 Photosensitivity

**Supplementary Table 387:** Exposures, outcomes and risk estimates for Photosensitivity in GLP1RA (female) (T) vs DPP4I (female) (C) new-user cohorts under four different propensity score (PS) adjustment / time-at-risk (TAR) designs across data sources. We report total patient exposure time in 1,000 patient-year (KYr) units, uncalibrated and calibrated hazard ratios (HRs) and their 95% confidence intervals (CIs). Greyed out entries did not pass study diagnostics and are included only for completeness.

| | Patients | | Exposure (KYr) | | Outcomes | | Incidence (per KYr) | |
|---|---|---|---|---|---|---|---|---|
| | T | C | T | C | T | C | T | C |
| **PS-matching / on-treatment TAR** | | | | | | | | |
| CCAE | 14,289 | 45,011 | 9.0 | 35.5 | 13 | 50 | 1.44 (0.77 - 2.46) | 1.41 (1.05 - 1.86) |
| ODOD | 7,863 | 28,515 | 4.6 | 22.5 | 10 | 45 | 2.18 (1.05 - 4.02) | 2.00 (1.46 - 2.67) |
| MDCR | 961 | 8,205 | 0.6 | 7.8 | <5 | 25 | < 8.04 (2.61 - 18.8) | 3.21 (2.08 - 4.74) |
| MDCD | 1,850 | 7,629 | 1.0 | 5.4 | <5 | 11 | < 4.93 (1.60 - 11.5) | 2.03 (1.01 - 3.63) |
| Open Claims | 143,120 | 443,736 | 104.1 | 389.4 | 39 | 202 | 0.37 (0.27 - 0.51) | 0.52 (0.45 - 0.60) |
| OEHR | 10,498 | 38,415 | 3.2 | 19.9 | 9 | 37 | 2.78 (1.27 - 5.28) | 1.86 (1.31 - 2.56) |
| Meta-analysis | 175,770 | 555,677 | 121.0 | 467.3 | 71 | 334 | 0.59 (0.46 - 0.74) | 0.71 (0.64 - 0.80) |
| Meta-analysis (w/o USOC) | 32,650 | 111,941 | 16.8 | 77.9 | 32 | 132 | 1.90 (1.30 - 2.68) | 1.69 (1.42 - 2.01) |
| **PS-stratification / on-treatment TAR** | | | | | | | | |
| CCAE | 19,239 | 45,015 | 11.4 | 35.5 | 15 | 50 | 1.32 (0.74 - 2.18) | 1.41 (1.05 - 1.86) |
| ODOD | 11,575 | 29,190 | 6.1 | 23.2 | 11 | 45 | 1.79 (0.89 - 3.21) | 1.94 (1.42 - 2.60) |
| MDCR | 1,102 | 8,668 | 0.7 | 8.2 | <5 | 25 | < 7.56 (2.45 - 17.6) | 3.06 (1.98 - 4.52) |
| MDCD | 2,013 | 7,706 | 1.1 | 5.5 | <5 | 11 | < 4.62 (1.50 - 10.8) | 2.00 (1.00 - 3.59) |
| Open Claims | 228,366 | 444,856 | 142.1 | 390.5 | 52 | 204 | 0.37 (0.27 - 0.48) | 0.52 (0.45 - 0.60) |
| OEHR | 13,527 | 38,618 | 4.0 | 20.0 | 9 | 37 | 2.26 (1.03 - 4.29) | 1.85 (1.30 - 2.55) |
| Meta-analysis | 272,707 | 557,679 | 163.6 | 469.2 | 87 | 336 | 0.53 (0.43 - 0.66) | 0.72 (0.64 - 0.80) |
| Meta-analysis (w/o USOC) | 44,341 | 112,823 | 21.5 | 78.7 | 35 | 132 | 1.63 (1.13 - 2.26) | 1.68 (1.40 - 1.99) |
| **PS-matching / on-treatment TAR without escalation** | | | | | | | | |
| CCAE | 14,289 | 45,011 | 8.1 | 29.2 | 11 | 45 | 1.36 (0.68 - 2.43) | 1.54 (1.12 - 2.06) |
| ODOD | 7,863 | 28,515 | 4.1 | 18.9 | 10 | 37 | 2.43 (1.17 - 4.47) | 1.96 (1.38 - 2.70) |
| MDCR | 961 | 8,205 | 0.6 | 6.8 | <5 | 23 | < 8.72 (2.83 - 20.4) | 3.40 (2.15 - 5.10) |
| MDCD | 1,850 | 7,629 | 0.9 | 4.2 | <5 | 9 | < 5.72 (1.86 - 13.3) | 2.13 (0.97 - 4.04) |
| Open Claims | 143,120 | 443,736 | 92.3 | 309.1 | 27 | 175 | 0.29 (0.19 - 0.43) | 0.57 (0.49 - 0.66) |
| OEHR | 10,498 | 38,415 | 2.8 | 16.0 | 6 | 31 | 2.15 (0.79 - 4.69) | 1.93 (1.31 - 2.74) |
| Meta-analysis | 175,770 | 555,677 | 107.2 | 373.2 | 54 | 288 | 0.50 (0.38 - 0.66) | 0.77 (0.69 - 0.87) |
| Meta-analysis (w/o USOC) | 32,650 | 111,941 | 15.0 | 64.1 | 27 | 113 | 1.80 (1.19 - 2.62) | 1.76 (1.45 - 2.12) |
| **PS-stratification / on-treatment TAR without escalation** | | | | | | | | |
| CCAE | 19,239 | 45,015 | 10.3 | 29.2 | 13 | 45 | 1.26 (0.67 - 2.16) | 1.54 (1.12 - 2.06) |
| ODOD | 11,575 | 29,190 | 5.6 | 19.4 | 11 | 37 | 1.97 (0.98 - 3.53) | 1.91 (1.34 - 2.63) |
| MDCR | 1,102 | 8,668 | 0.6 | 7.1 | <5 | 23 | < 8.17 (2.65 - 19.1) | 3.24 (2.06 - 4.86) |
| MDCD | 2,013 | 7,706 | 0.9 | 4.3 | <5 | 9 | < 5.35 (1.74 - 12.5) | 2.10 (0.96 - 3.99) |
| Open Claims | 228,366 | 444,856 | 128.2 | 310.0 | 39 | 176 | 0.30 (0.22 - 0.42) | 0.57 (0.49 - 0.66) |
| OEHR | 13,527 | 38,618 | 3.5 | 16.1 | 6 | 31 | 1.73 (0.64 - 3.77) | 1.92 (1.31 - 2.73) |
| Meta-analysis | 272,707 | 557,679 | 147.5 | 374.8 | 69 | 289 | 0.47 (0.36 - 0.59) | 0.77 (0.68 - 0.87) |
| Meta-analysis (w/o USOC) | 44,341 | 112,823 | 19.3 | 64.7 | 30 | 113 | 1.55 (1.05 - 2.22) | 1.75 (1.44 - 2.10) |

##### 4.4.25 Venous thromboembolism

**Supplementary Table 388:** Exposures, outcomes and risk estimates for Venous thromboembolism in GLP1RA (female) (T) vs DPP4I (female) (C) new-user cohorts under four different propensity score (PS) adjustment / time-at-risk (TAR) designs across data sources. We report total patient exposure time in 1,000 patient-year (KYr) units, uncalibrated and calibrated hazard ratios (HRs) and their 95% confidence intervals (CIs). Greyed out entries did not pass study diagnostics and are included only for completeness.

| | Patients | | Exposure (KYr) | | Outcomes | | Incidence (per KYr) | |
|---|---|---|---|---|---|---|---|---|
| | T | C | T | C | T | C | T | C |
| **PS-matching / on-treatment TAR** | | | | | | | | |
| CCAE | 14,066 | 44,438 | 8.8 | 35.0 | 40 | 185 | 4.54 (3.24 - 6.18) | 5.28 (4.55 - 6.10) |
| ODOD | 7,637 | 27,903 | 4.5 | 21.9 | 40 | 230 | 8.97 (6.41 - 12.2) | 10.5 (9.18 - 11.9) |
| MDCR | 947 | 8,008 | 0.6 | 7.6 | 6 | 103 | 9.88 (3.62 - 21.5) | 13.5 (11.0 - 16.4) |
| MDCD | 1,771 | 7,342 | 1.0 | 5.2 | 10 | 61 | 10.4 (4.97 - 19.1) | 11.8 (9.04 - 15.2) |
| Open Claims | 139,255 | 432,685 | 101.4 | 379.1 | 397 | 2,005 | 3.91 (3.54 - 4.32) | 5.29 (5.06 - 5.53) |
| OEHR | 10,290 | 37,693 | 3.2 | 19.4 | 25 | 135 | 7.85 (5.08 - 11.6) | 6.96 (5.83 - 8.24) |
| Meta-analysis | 173,019 | 550,061 | 118.9 | 460.6 | 512 | 2,616 | 4.31 (3.94 - 4.70) | 5.68 (5.46 - 5.90) |
| Meta-analysis (w/o USOC) | 33,764 | 117,376 | 17.4 | 81.5 | 115 | 611 | 6.60 (5.45 - 7.92) | 7.50 (6.91 - 8.11) |
| **PS-stratification / on-treatment TAR** | | | | | | | | |
| CCAE | 18,912 | 44,442 | 11.1 | 35.0 | 50 | 185 | 4.50 (3.34 - 5.94) | 5.28 (4.55 - 6.10) |
| ODOD | 11,250 | 28,540 | 6.0 | 22.6 | 52 | 240 | 8.71 (6.50 - 11.4) | 10.6 (9.34 - 12.1) |
| MDCR | 1,082 | 8,418 | 0.6 | 7.9 | 6 | 108 | 9.32 (3.42 - 20.3) | 13.6 (11.1 - 16.4) |
| MDCD | 1,938 | 7,411 | 1.0 | 5.2 | 10 | 61 | 9.66 (4.63 - 17.8) | 11.7 (8.93 - 15.0) |
| Open Claims | 221,964 | 433,661 | 138.3 | 380.1 | 508 | 2,031 | 3.67 (3.36 - 4.01) | 5.34 (5.11 - 5.58) |
| OEHR | 13,249 | 37,874 | 3.9 | 19.5 | 30 | 139 | 7.68 (5.18 - 11.0) | 7.13 (5.99 - 8.42) |
| Meta-analysis | 267,313 | 551,928 | 160.3 | 462.4 | 650 | 2,656 | 4.06 (3.75 - 4.38) | 5.74 (5.53 - 5.97) |
| Meta-analysis (w/o USOC) | 45,349 | 118,267 | 22.0 | 82.3 | 142 | 625 | 6.45 (5.43 - 7.60) | 7.59 (7.01 - 8.21) |
| **PS-matching / on-treatment TAR without escalation** | | | | | | | | |
| CCAE | 14,066 | 44,438 | 7.9 | 28.8 | 35 | 147 | 4.43 (3.08 - 6.16) | 5.10 (4.31 - 6.00) |
| ODOD | 7,637 | 27,903 | 4.0 | 18.4 | 34 | 203 | 8.48 (5.87 - 11.9) | 11.0 (9.57 - 12.7) |
| MDCR | 947 | 8,008 | 0.6 | 6.6 | <5 | 87 | < 8.88 (2.88 - 20.7) | 13.2 (10.5 - 16.2) |
| MDCD | 1,771 | 7,342 | 0.8 | 4.0 | <5 | 44 | < 5.98 (1.94 - 14.0) | 10.9 (7.92 - 14.6) |
| Open Claims | 139,255 | 432,685 | 89.9 | 301.2 | 343 | 1,613 | 3.82 (3.42 - 4.24) | 5.36 (5.10 - 5.62) |
| OEHR | 10,290 | 37,693 | 2.7 | 15.7 | 20 | 91 | 7.30 (4.46 - 11.3) | 5.81 (4.68 - 7.13) |
| Meta-analysis | 173,019 | 550,061 | 105.4 | 368.1 | <437 | 2,098 | < 4.15 (3.77 - 4.55) | 5.70 (5.46 - 5.95) |
| Meta-analysis (w/o USOC) | 33,764 | 117,376 | 15.5 | 66.9 | <94 | 485 | < 6.07 (4.90 - 7.43) | 7.25 (6.62 - 7.92) |
| **PS-stratification / on-treatment TAR without escalation** | | | | | | | | |
| CCAE | 18,912 | 44,442 | 10.0 | 28.8 | 45 | 147 | 4.48 (3.27 - 5.99) | 5.10 (4.31 - 6.00) |
| ODOD | 11,250 | 28,540 | 5.4 | 18.9 | 46 | 211 | 8.48 (6.21 - 11.3) | 11.2 (9.70 - 12.8) |
| MDCR | 1,082 | 8,418 | 0.6 | 6.9 | <5 | 89 | < 8.36 (2.71 - 19.5) | 12.9 (10.3 - 15.9) |
| MDCD | 1,938 | 7,411 | 0.9 | 4.1 | <5 | 44 | < 5.56 (1.80 - 13.0) | 10.8 (7.83 - 14.5) |
| Open Claims | 221,964 | 433,661 | 124.8 | 302.0 | 445 | 1,637 | 3.57 (3.24 - 3.91) | 5.42 (5.16 - 5.69) |
| OEHR | 13,249 | 37,874 | 3.4 | 15.7 | 25 | 95 | 7.36 (4.76 - 10.9) | 6.04 (4.88 - 7.38) |
| Meta-analysis | 267,313 | 551,928 | 144.5 | 369.5 | <566 | 2,134 | < 3.92 (3.60 - 4.25) | 5.78 (5.53 - 6.03) |
| Meta-analysis (w/o USOC) | 45,349 | 118,267 | 19.8 | 67.6 | <121 | 497 | < 6.12 (5.08 - 7.31) | 7.36 (6.72 - 8.03) |

##### 4.4.26 Vomiting

**Supplementary Table 389:** Exposures, outcomes and risk estimates for Vomiting in GLP1RA (female) (T) vs DPP4I (female) (C) new-user cohorts under four different propensity score (PS) adjustment / time-at-risk (TAR) designs across data sources. We report total patient exposure time in 1,000 patient-year (KYr) units, uncalibrated and calibrated hazard ratios (HRs) and their 95% confidence intervals (CIs). Greyed out entries did not pass study diagnostics and are included only for completeness.

| | Patients | | Exposure (KYr) | | Outcomes | | Incidence (per KYr) | |
|---|---|---|---|---|---|---|---|---|
| | T | C | T | C | T | C | T | C |
| **PS-matching / on-treatment TAR** | | | | | | | | |
| CCAE | 12,634 | 40,547 | 7.8 | 31.7 | 348 | 924 | 44.4 (39.9 - 49.4) | 29.1 (27.3 - 31.1) |
| ODOD | 6,827 | 25,080 | 3.9 | 19.5 | 218 | 708 | 55.5 (48.4 - 63.4) | 36.3 (33.6 - 39.0) |
| MDCR | 907 | 7,519 | 0.6 | 7.0 | 22 | 230 | 38.1 (23.9 - 57.7) | 32.7 (28.6 - 37.2) |
| MDCD | 1,350 | 5,634 | 0.7 | 3.9 | 80 | 297 | 112 (88.7 - 139) | 76.7 (68.2 - 85.9) |
| Open Claims | 121,312 | 385,568 | 88.1 | 337.2 | 2,315 | 7,187 | 26.3 (25.2 - 27.4) | 21.3 (20.8 - 21.8) |
| OEHR | 9,413 | 35,152 | 2.9 | 17.8 | 136 | 573 | 47.3 (39.7 - 56.0) | 32.2 (29.7 - 35.0) |
| Meta-analysis | 151,536 | 491,981 | 103.4 | 410.1 | 3,097 | 9,689 | 29.9 (28.9 - 31.0) | 23.6 (23.2 - 24.1) |
| Meta-analysis (w/o USOC) | 30,224 | 106,413 | 15.3 | 72.9 | 782 | 2,502 | 51.0 (47.5 - 54.7) | 34.3 (33.0 - 35.7) |
| **PS-stratification / on-treatment TAR** | | | | | | | | |
| CCAE | 16,862 | 40,551 | 9.8 | 31.7 | 469 | 925 | 47.8 (43.6 - 52.3) | 29.1 (27.3 - 31.1) |
| ODOD | 9,989 | 25,529 | 5.2 | 19.9 | 323 | 732 | 62.0 (55.4 - 69.1) | 36.7 (34.1 - 39.5) |
| MDCR | 1,032 | 7,844 | 0.6 | 7.3 | 24 | 245 | 39.2 (25.1 - 58.4) | 33.6 (29.5 - 38.1) |
| MDCD | 1,441 | 5,668 | 0.7 | 3.9 | 87 | 300 | 116 (93.2 - 144) | 77.0 (68.5 - 86.2) |
| Open Claims | 191,403 | 386,745 | 119.3 | 338.3 | 3,339 | 7,240 | 28.0 (27.1 - 29.0) | 21.4 (20.9 - 21.9) |
| OEHR | 12,003 | 35,368 | 3.5 | 17.9 | 178 | 580 | 51.0 (43.8 - 59.1) | 32.4 (29.8 - 35.1) |
| Meta-analysis | 231,698 | 493,861 | 138.5 | 411.7 | 4,396 | 9,777 | 31.7 (30.8 - 32.7) | 23.7 (23.3 - 24.2) |
| Meta-analysis (w/o USOC) | 40,295 | 107,116 | 19.3 | 73.5 | 1,057 | 2,537 | 54.9 (51.6 - 58.3) | 34.5 (33.2 - 35.9) |
| **PS-matching / on-treatment TAR without escalation** | | | | | | | | |
| CCAE | 12,634 | 40,547 | 7.1 | 26.3 | 321 | 760 | 45.4 (40.6 - 50.7) | 28.9 (26.9 - 31.1) |
| ODOD | 6,827 | 25,080 | 3.5 | 16.5 | 196 | 590 | 55.5 (48.0 - 63.8) | 35.7 (32.9 - 38.7) |
| MDCR | 907 | 7,519 | 0.5 | 6.1 | 21 | 191 | 39.4 (24.4 - 60.3) | 31.2 (26.9 - 36.0) |
| MDCD | 1,350 | 5,634 | 0.6 | 3.1 | 71 | 224 | 115 (89.7 - 145) | 72.5 (63.3 - 82.6) |
| Open Claims | 121,312 | 385,568 | 78.2 | 269.2 | 2,052 | 5,665 | 26.2 (25.1 - 27.4) | 21.0 (20.5 - 21.6) |
| OEHR | 9,413 | 35,152 | 2.5 | 14.4 | 116 | 437 | 46.7 (38.6 - 56.0) | 30.3 (27.5 - 33.3) |
| Meta-analysis | 151,536 | 491,981 | 91.9 | 329.5 | 2,756 | 7,676 | 30.0 (28.9 - 31.1) | 23.3 (22.8 - 23.8) |
| Meta-analysis (w/o USOC) | 30,224 | 106,413 | 13.7 | 60.3 | 704 | 2,011 | 51.4 (47.6 - 55.3) | 33.4 (31.9 - 34.8) |
| **PS-stratification / on-treatment TAR without escalation** | | | | | | | | |
| CCAE | 16,862 | 40,551 | 8.9 | 26.3 | 430 | 761 | 48.2 (43.8 - 53.0) | 29.0 (27.0 - 31.1) |
| ODOD | 9,989 | 25,529 | 4.7 | 16.9 | 294 | 609 | 62.0 (55.1 - 69.5) | 36.1 (33.3 - 39.1) |
| MDCR | 1,032 | 7,844 | 0.6 | 6.3 | 22 | 204 | 38.8 (24.3 - 58.8) | 32.1 (27.9 - 36.9) |
| MDCD | 1,441 | 5,668 | 0.6 | 3.1 | 78 | 227 | 120 (95.1 - 150) | 73.2 (64.0 - 83.4) |
| Open Claims | 191,403 | 386,745 | 107.8 | 270.1 | 3,025 | 5,707 | 28.1 (27.1 - 29.1) | 21.1 (20.6 - 21.7) |
| OEHR | 12,003 | 35,368 | 3.0 | 14.5 | 154 | 442 | 50.5 (42.9 - 59.2) | 30.4 (27.6 - 33.4) |
| Meta-analysis | 231,698 | 493,861 | 125.1 | 330.9 | 3,981 | 7,746 | 31.8 (30.8 - 32.8) | 23.4 (22.9 - 23.9) |
| Meta-analysis (w/o USOC) | 40,295 | 107,116 | 17.4 | 60.8 | 956 | 2,039 | 55.1 (51.6 - 58.7) | 33.6 (32.1 - 35.0) |

#### 4.5 GLP1RA (female) vs SU (female)

##### 4.5.1 3-point MACE

**Supplementary Table 390:** Exposures, outcomes and risk estimates for 3-point MACE in GLP1RA (female) (T) vs SU (female) (C) new-user cohorts under four different propensity score (PS) adjustment / time-at-risk (TAR) designs across data sources. We report total patient exposure time in 1,000 patient-year (KYr) units, uncalibrated and calibrated hazard ratios (HRs) and their 95% confidence intervals (CIs). Greyed out entries did not pass study diagnostics and are included only for completeness.

| | Patients | | Exposure (KYr) | | Outcomes | | Incidence (per KYr) | |
|---|---|---|---|---|---|---|---|---|
| | T | C | T | C | T | C | T | C |
| **PS-matching / on-treatment TAR** | | | | | | | | |
| CCAE | 15,035 | 68,011 | 9.3 | 50.2 | 45 | 363 | 4.83 (3.52 - 6.46) | 7.23 (6.50 - 8.01) |
| ODOD | 8,688 | 53,222 | 4.9 | 44.8 | 58 | 889 | 11.9 (9.05 - 15.4) | 19.9 (18.6 - 21.2) |
| MDCR | 1,051 | 12,167 | 0.7 | 11.0 | 8 | 300 | 11.6 (5.00 - 22.8) | 27.2 (24.2 - 30.4) |
| MDCD | 1,801 | 14,292 | 1.0 | 9.1 | 9 | 178 | 9.16 (4.19 - 17.4) | 19.5 (16.8 - 22.6) |
| Open Claims | 171,822 | 796,792 | 115.8 | 754.6 | 584 | 8,693 | 5.04 (4.64 - 5.47) | 11.5 (11.3 - 11.8) |
| OEHR | 10,976 | 76,984 | 3.3 | 43.2 | 21 | 483 | 6.33 (3.92 - 9.68) | 11.2 (10.2 - 12.2) |
| Meta-analysis | 209,373 | 1,021,468 | 135.0 | 912.9 | 725 | 10,906 | 5.37 (4.99 - 5.78) | 11.9 (11.7 - 12.2) |
| Meta-analysis (w/o USOC) | 37,551 | 224,676 | 19.2 | 158.3 | 141 | 2,213 | 7.35 (6.19 - 8.67) | 14.0 (13.4 - 14.6) |
| **PS-stratification / on-treatment TAR** | | | | | | | | |
| CCAE | 20,219 | 70,165 | 12.0 | 51.5 | 56 | 372 | 4.67 (3.52 - 6.06) | 7.23 (6.51 - 8.00) |
| ODOD | 11,357 | 57,249 | 6.0 | 48.9 | 66 | 1,113 | 11.0 (8.50 - 14.0) | 22.7 (21.4 - 24.1) |
| MDCR | 1,175 | 16,035 | 0.7 | 14.3 | 8 | 454 | 10.8 (4.67 - 21.3) | 31.7 (28.9 - 34.8) |
| MDCD | 1,943 | 15,757 | 1.0 | 10.2 | 9 | 208 | 8.58 (3.92 - 16.3) | 20.3 (17.6 - 23.2) |
| Open Claims | 223,507 | 810,783 | 139.3 | 767.8 | 676 | 9,194 | 4.85 (4.49 - 5.23) | 12.0 (11.7 - 12.2) |
| OEHR | 13,453 | 81,779 | 4.0 | 46.2 | 25 | 586 | 6.32 (4.09 - 9.33) | 12.7 (11.7 - 13.8) |
| Meta-analysis | 268,536 | 1,019,976 | 161.3 | 914.4 | 823 | 11,265 | 5.10 (4.76 - 5.46) | 12.3 (12.1 - 12.5) |
| Meta-analysis (w/o USOC) | 45,029 | 209,193 | 22.0 | 146.6 | 147 | 2,071 | 6.69 (5.65 - 7.87) | 14.1 (13.5 - 14.8) |
| **PS-matching / on-treatment TAR without escalation** | | | | | | | | |
| CCAE | 15,035 | 68,011 | 8.4 | 43.1 | 37 | 303 | 4.42 (3.11 - 6.10) | 7.03 (6.26 - 7.87) |
| ODOD | 8,688 | 53,222 | 4.4 | 39.8 | 51 | 784 | 11.6 (8.62 - 15.2) | 19.7 (18.4 - 21.2) |
| MDCR | 1,051 | 12,167 | 0.6 | 10.1 | 8 | 267 | 12.7 (5.50 - 25.1) | 26.4 (23.4 - 29.8) |
| MDCD | 1,801 | 14,292 | 0.8 | 7.5 | 8 | 144 | 9.47 (4.09 - 18.7) | 19.1 (16.1 - 22.5) |
| Open Claims | 171,822 | 796,792 | 103.4 | 634.7 | 507 | 7,373 | 4.90 (4.49 - 5.35) | 11.6 (11.4 - 11.9) |
| OEHR | 10,976 | 76,984 | 2.8 | 36.7 | 16 | 354 | 5.62 (3.21 - 9.12) | 9.65 (8.67 - 10.7) |
| Meta-analysis | 209,373 | 1,021,468 | 120.5 | 771.9 | 627 | 9,225 | 5.20 (4.80 - 5.63) | 12.0 (11.7 - 12.2) |
| Meta-analysis (w/o USOC) | 37,551 | 224,676 | 17.1 | 137.2 | 120 | 1,852 | 7.02 (5.82 - 8.40) | 13.5 (12.9 - 14.1) |
| **PS-stratification / on-treatment TAR without escalation** | | | | | | | | |
| CCAE | 20,219 | 70,165 | 10.9 | 44.3 | 45 | 310 | 4.14 (3.02 - 5.54) | 7.01 (6.25 - 7.83) |
| ODOD | 11,357 | 57,249 | 5.5 | 43.6 | 59 | 989 | 10.8 (8.22 - 13.9) | 22.7 (21.3 - 24.2) |
| MDCR | 1,175 | 16,035 | 0.7 | 13.1 | 8 | 407 | 11.8 (5.10 - 23.3) | 31.0 (28.1 - 34.2) |
| MDCD | 1,943 | 15,757 | 0.9 | 8.5 | 8 | 170 | 8.85 (3.82 - 17.4) | 20.0 (17.1 - 23.2) |
| Open Claims | 223,507 | 810,783 | 125.7 | 646.3 | 596 | 7,802 | 4.74 (4.37 - 5.14) | 12.1 (11.8 - 12.3) |
| OEHR | 13,453 | 81,779 | 3.4 | 39.3 | 20 | 426 | 5.81 (3.55 - 8.98) | 10.8 (9.83 - 11.9) |
| Meta-analysis | 268,536 | 1,019,976 | 145.5 | 773.4 | 720 | 9,527 | 4.95 (4.59 - 5.32) | 12.3 (12.1 - 12.6) |
| Meta-analysis (w/o USOC) | 45,029 | 209,193 | 19.8 | 127.1 | 124 | 1,725 | 6.27 (5.21 - 7.47) | 13.6 (12.9 - 14.2) |

##### 4.5.2 4-point MACE

**Supplementary Table 391:** Exposures, outcomes and risk estimates for 4-point MACE in GLP1RA (female) (T) vs SU (female) (C) new-user cohorts under four different propensity score (PS) adjustment / time-at-risk (TAR) designs across data sources. We report total patient exposure time in 1,000 patient-year (KYr) units, uncalibrated and calibrated hazard ratios (HRs) and their 95% confidence intervals (CIs). Greyed out entries did not pass study diagnostics and are included only for completeness.

| | Patients | | Exposure (KYr) | | Outcomes | | Incidence (per KYr) | |
|---|---|---|---|---|---|---|---|---|
| | T | C | T | C | T | C | T | C |
| **PS-matching / on-treatment TAR** | | | | | | | | |
| CCAE | 14,823 | 67,020 | 9.2 | 49.4 | 57 | 584 | 6.20 (4.69 - 8.03) | 11.8 (10.9 - 12.8) |
| ODOD | 8,427 | 51,180 | 4.7 | 42.6 | 100 | 1,511 | 21.3 (17.3 - 25.9) | 35.4 (33.7 - 37.3) |
| MDCR | 1,011 | 11,143 | 0.7 | 10.0 | 13 | 445 | 19.2 (10.2 - 32.8) | 44.7 (40.6 - 49.1) |
| MDCD | 1,689 | 13,397 | 0.9 | 8.4 | 17 | 308 | 18.6 (10.8 - 29.7) | 36.5 (32.5 - 40.8) |
| Open Claims | 167,622 | 771,419 | 113.0 | 726.5 | 900 | 14,557 | 7.96 (7.45 - 8.50) | 20.0 (19.7 - 20.4) |
| OEHR | 10,817 | 75,708 | 3.3 | 42.0 | 36 | 916 | 11.0 (7.74 - 15.3) | 21.8 (20.4 - 23.3) |
| Meta-analysis | 204,389 | 989,867 | 131.8 | 878.9 | 1,123 | 18,321 | 8.52 (8.03 - 9.03) | 20.8 (20.5 - 21.1) |
| Meta-analysis (w/o USOC) | 36,767 | 218,448 | 18.8 | 152.4 | 223 | 3,764 | 11.9 (10.4 - 13.6) | 24.7 (23.9 - 25.5) |
| **PS-stratification / on-treatment TAR** | | | | | | | | |
| CCAE | 19,944 | 69,057 | 11.8 | 50.6 | 70 | 599 | 5.91 (4.61 - 7.47) | 11.8 (10.9 - 12.8) |
| ODOD | 11,042 | 54,541 | 5.8 | 46.2 | 109 | 1,805 | 18.7 (15.4 - 22.6) | 39.1 (37.3 - 40.9) |
| MDCR | 1,125 | 14,786 | 0.7 | 13.1 | 13 | 731 | 18.0 (9.59 - 30.8) | 56.0 (52.0 - 60.2) |
| MDCD | 1,819 | 14,676 | 1.0 | 9.4 | 18 | 355 | 18.4 (10.9 - 29.1) | 37.7 (33.8 - 41.8) |
| Open Claims | 218,502 | 783,706 | 136.1 | 738.0 | 1,041 | 15,361 | 7.65 (7.19 - 8.13) | 20.8 (20.5 - 21.1) |
| OEHR | 13,258 | 80,292 | 3.9 | 44.8 | 41 | 1,088 | 10.5 (7.56 - 14.3) | 24.3 (22.9 - 25.8) |
| Meta-analysis | 262,746 | 987,596 | 157.7 | 879.5 | 1,261 | 18,853 | 8.00 (7.56 - 8.45) | 21.4 (21.1 - 21.7) |
| Meta-analysis (w/o USOC) | 44,244 | 203,890 | 21.6 | 141.5 | 220 | 3,492 | 10.2 (8.90 - 11.6) | 24.7 (23.9 - 25.5) |
| **PS-matching / on-treatment TAR without escalation** | | | | | | | | |
| CCAE | 14,823 | 67,020 | 8.3 | 42.4 | 47 | 503 | 5.69 (4.18 - 7.57) | 11.9 (10.8 - 12.9) |
| ODOD | 8,427 | 51,180 | 4.3 | 37.9 | 89 | 1,325 | 20.9 (16.8 - 25.7) | 34.9 (33.1 - 36.9) |
| MDCR | 1,011 | 11,143 | 0.6 | 9.1 | 13 | 394 | 21.1 (11.2 - 36.0) | 43.1 (38.9 - 47.6) |
| MDCD | 1,689 | 13,397 | 0.8 | 7.0 | 15 | 248 | 19.1 (10.7 - 31.5) | 35.3 (31.1 - 40.0) |
| Open Claims | 167,622 | 771,419 | 100.9 | 611.6 | 772 | 12,285 | 7.65 (7.12 - 8.21) | 20.1 (19.7 - 20.4) |
| OEHR | 10,817 | 75,708 | 2.8 | 35.9 | 28 | 689 | 9.97 (6.63 - 14.4) | 19.2 (17.8 - 20.7) |
| Meta-analysis | 204,389 | 989,867 | 117.7 | 743.9 | 964 | 15,444 | 8.19 (7.68 - 8.73) | 20.8 (20.4 - 21.1) |
| Meta-analysis (w/o USOC) | 36,767 | 218,448 | 16.7 | 132.4 | 192 | 3,159 | 11.5 (9.91 - 13.2) | 23.9 (23.0 - 24.7) |
| **PS-stratification / on-treatment TAR without escalation** | | | | | | | | |
| CCAE | 19,944 | 69,057 | 10.7 | 43.5 | 56 | 513 | 5.22 (3.94 - 6.78) | 11.8 (10.8 - 12.9) |
| ODOD | 11,042 | 54,541 | 5.3 | 41.1 | 97 | 1,590 | 18.3 (14.8 - 22.3) | 38.6 (36.8 - 40.6) |
| MDCR | 1,125 | 14,786 | 0.7 | 12.0 | 13 | 659 | 19.7 (10.5 - 33.7) | 54.9 (50.8 - 59.3) |
| MDCD | 1,819 | 14,676 | 0.8 | 7.8 | 15 | 289 | 17.9 (10.0 - 29.5) | 36.9 (32.7 - 41.4) |
| Open Claims | 218,502 | 783,706 | 123.0 | 621.7 | 905 | 12,981 | 7.36 (6.89 - 7.86) | 20.9 (20.5 - 21.2) |
| OEHR | 13,258 | 80,292 | 3.4 | 38.3 | 33 | 827 | 9.73 (6.69 - 13.7) | 21.6 (20.1 - 23.1) |
| Meta-analysis | 262,746 | 987,596 | 142.4 | 744.7 | 1,091 | 15,911 | 7.66 (7.21 - 8.13) | 21.4 (21.0 - 21.7) |
| Meta-analysis (w/o USOC) | 44,244 | 203,890 | 19.4 | 123.0 | 186 | 2,930 | 9.57 (8.25 - 11.1) | 23.8 (23.0 - 24.7) |

##### 4.5.3 Acute myocardial infarction

**Supplementary Table 392:** Exposures, outcomes and risk estimates for Acute myocardial infarction in GLP1RA (female) (T) vs SU (female) (C) new-user cohorts under four different propensity score (PS) adjustment / time-at-risk (TAR) designs across data sources. We report total patient exposure time in 1,000 patient-year (KYr) units, uncalibrated and calibrated hazard ratios (HRs) and their 95% confidence intervals (CIs). Greyed out entries did not pass study diagnostics and are included only for completeness.

| | Patients | | Exposure (KYr) | | Outcomes | | Incidence (per KYr) | |
|---|---|---|---|---|---|---|---|---|
| | T | C | T | C | T | C | T | C |
| **PS-matching / on-treatment TAR** | | | | | | | | |
| CCAE | 15,204 | 68,858 | 9.4 | 51.0 | 22 | 160 | 2.33 (1.46 - 3.53) | 3.14 (2.67 - 3.66) |
| ODOD | 8,837 | 54,488 | 5.0 | 46.2 | 28 | 385 | 5.62 (3.74 - 8.13) | 8.34 (7.53 - 9.22) |
| MDCR | 1,084 | 13,225 | 0.7 | 12.1 | <5 | 124 | < 7.06 (2.29 - 16.5) | 10.3 (8.55 - 12.3) |
| MDCD | 1,847 | 14,776 | 1.0 | 9.5 | <5 | 67 | < 4.97 (1.61 - 11.6) | 7.08 (5.48 - 8.99) |
| Open Claims | 175,427 | 818,268 | 118.2 | 777.2 | 251 | 3,750 | 2.12 (1.87 - 2.40) | 4.82 (4.67 - 4.98) |
| OEHR | 11,063 | 77,588 | 3.4 | 43.7 | 12 | 234 | 3.58 (1.85 - 6.26) | 5.36 (4.70 - 6.09) |
| Meta-analysis | 213,462 | 1,047,203 | 137.7 | 939.5 | <323 | 4,720 | < 2.35 (2.10 - 2.62) | 5.02 (4.88 - 5.17) |
| Meta-analysis (w/o USOC) | 38,035 | 228,935 | 19.5 | 162.3 | <72 | 970 | < 3.69 (2.89 - 4.65) | 5.98 (5.61 - 6.36) |
| **PS-stratification / on-treatment TAR** | | | | | | | | |
| CCAE | 20,444 | 71,108 | 12.1 | 52.3 | 25 | 164 | 2.06 (1.33 - 3.04) | 3.14 (2.68 - 3.66) |
| ODOD | 11,517 | 59,011 | 6.1 | 50.9 | 30 | 481 | 4.90 (3.30 - 6.99) | 9.45 (8.62 - 10.3) |
| MDCR | 1,213 | 16,841 | 0.8 | 15.1 | <5 | 183 | < 6.59 (2.14 - 15.4) | 12.1 (10.4 - 14.0) |
| MDCD | 1,994 | 16,407 | 1.1 | 10.7 | <5 | 83 | < 4.66 (1.51 - 10.9) | 7.75 (6.17 - 9.60) |
| Open Claims | 227,850 | 833,873 | 142.0 | 792.0 | 281 | 3,979 | 1.98 (1.75 - 2.22) | 5.02 (4.87 - 5.18) |
| OEHR | 13,547 | 82,582 | 4.0 | 46.8 | 15 | 277 | 3.75 (2.10 - 6.19) | 5.92 (5.24 - 6.66) |
| Meta-analysis | 273,358 | 1,046,574 | 164.3 | 942.0 | 351 | 4,901 | 2.14 (1.92 - 2.37) | 5.20 (5.06 - 5.35) |
| Meta-analysis (w/o USOC) | 45,508 | 212,701 | 22.3 | 150.0 | 70 | 922 | 3.14 (2.45 - 3.97) | 6.15 (5.76 - 6.56) |
| **PS-matching / on-treatment TAR without escalation** | | | | | | | | |
| CCAE | 15,204 | 68,858 | 8.5 | 43.7 | 19 | 136 | 2.24 (1.35 - 3.50) | 3.11 (2.61 - 3.68) |
| ODOD | 8,837 | 54,488 | 4.5 | 41.0 | 24 | 335 | 5.33 (3.42 - 7.94) | 8.18 (7.33 - 9.10) |
| MDCR | 1,084 | 13,225 | 0.6 | 11.0 | <5 | 109 | < 7.76 (2.52 - 18.1) | 9.89 (8.12 - 11.9) |
| MDCD | 1,847 | 14,776 | 0.9 | 7.8 | <5 | 51 | < 5.77 (1.87 - 13.5) | 6.50 (4.84 - 8.55) |
| Open Claims | 175,427 | 818,268 | 105.5 | 653.4 | 223 | 3,192 | 2.11 (1.85 - 2.41) | 4.88 (4.72 - 5.06) |
| OEHR | 11,063 | 77,588 | 2.9 | 37.0 | 9 | 166 | 3.13 (1.43 - 5.93) | 4.48 (3.82 - 5.22) |
| Meta-analysis | 213,462 | 1,047,203 | 122.8 | 794.0 | <285 | 3,989 | < 2.32 (2.06 - 2.61) | 5.02 (4.87 - 5.18) |
| Meta-analysis (w/o USOC) | 38,035 | 228,935 | 17.4 | 140.6 | <62 | 797 | < 3.57 (2.74 - 4.58) | 5.67 (5.28 - 6.08) |
| **PS-stratification / on-treatment TAR without escalation** | | | | | | | | |
| CCAE | 20,444 | 71,108 | 11.0 | 44.9 | 22 | 139 | 2.00 (1.25 - 3.03) | 3.10 (2.60 - 3.66) |
| ODOD | 11,517 | 59,011 | 5.6 | 45.3 | 26 | 423 | 4.67 (3.05 - 6.84) | 9.34 (8.47 - 10.3) |
| MDCR | 1,213 | 16,841 | 0.7 | 13.8 | <5 | 163 | < 7.21 (2.34 - 16.8) | 11.8 (10.0 - 13.7) |
| MDCD | 1,994 | 16,407 | 0.9 | 8.9 | <5 | 66 | < 5.40 (1.75 - 12.6) | 7.42 (5.74 - 9.44) |
| Open Claims | 227,850 | 833,873 | 128.1 | 666.3 | 251 | 3,391 | 1.96 (1.72 - 2.22) | 5.09 (4.92 - 5.26) |
| OEHR | 13,547 | 82,582 | 3.5 | 39.8 | 12 | 201 | 3.45 (1.78 - 6.03) | 5.05 (4.38 - 5.80) |
| Meta-analysis | 273,358 | 1,046,574 | 148.2 | 796.3 | 311 | 4,154 | 2.10 (1.87 - 2.35) | 5.22 (5.06 - 5.38) |
| Meta-analysis (w/o USOC) | 45,508 | 212,701 | 20.1 | 130.0 | 60 | 763 | 2.99 (2.28 - 3.85) | 5.87 (5.46 - 6.30) |

##### 4.5.4 Acute renal failure

**Supplementary Table 393:** Exposures, outcomes and risk estimates for Acute renal failure in GLP1RA (female) (T) vs SU (female) (C) new-user cohorts under four different propensity score (PS) adjustment / time-at-risk (TAR) designs across data sources. We report total patient exposure time in 1,000 patient-year (KYr) units, uncalibrated and calibrated hazard ratios (HRs) and their 95% confidence intervals (CIs). Greyed out entries did not pass study diagnostics and are included only for completeness.

| | Patients | | Exposure (KYr) | | Outcomes | | Incidence (per KYr) | |
|---|---|---|---|---|---|---|---|---|
| | T | C | T | C | T | C | T | C |
| **PS-matching / on-treatment TAR** | | | | | | | | |
| CCAE | 15,192 | 68,720 | 9.4 | 50.8 | 44 | 421 | 4.67 (3.39 - 6.27) | 8.29 (7.51 - 9.12) |
| ODOD | 8,692 | 53,334 | 4.9 | 44.7 | 65 | 1,305 | 13.3 (10.3 - 17.0) | 29.2 (27.6 - 30.8) |
| MDCR | 1,061 | 12,944 | 0.7 | 11.7 | 10 | 339 | 14.3 (6.87 - 26.4) | 28.9 (25.9 - 32.2) |
| MDCD | 1,797 | 14,412 | 1.0 | 9.1 | 16 | 268 | 16.6 (9.48 - 26.9) | 29.5 (26.0 - 33.2) |
| Open Claims | 173,476 | 805,988 | 116.6 | 761.0 | 802 | 11,625 | 6.88 (6.41 - 7.37) | 15.3 (15.0 - 15.6) |
| OEHR | 11,015 | 77,132 | 3.3 | 43.1 | 41 | 629 | 12.3 (8.84 - 16.7) | 14.6 (13.5 - 15.8) |
| Meta-analysis | 211,233 | 1,032,530 | 135.9 | 920.4 | 978 | 14,587 | 7.20 (6.75 - 7.66) | 15.8 (15.6 - 16.1) |
| Meta-analysis (w/o USOC) | 37,757 | 226,542 | 19.3 | 159.4 | 176 | 2,962 | 9.13 (7.83 - 10.6) | 18.6 (17.9 - 19.3) |
| **PS-stratification / on-treatment TAR** | | | | | | | | |
| CCAE | 20,403 | 70,859 | 12.1 | 52.0 | 60 | 434 | 4.95 (3.78 - 6.38) | 8.34 (7.57 - 9.16) |
| ODOD | 11,352 | 57,109 | 6.0 | 48.7 | 79 | 1,557 | 13.2 (10.4 - 16.4) | 32.0 (30.4 - 33.6) |
| MDCR | 1,192 | 16,436 | 0.7 | 14.6 | 11 | 489 | 14.8 (7.37 - 26.4) | 33.4 (30.5 - 36.5) |
| MDCD | 1,939 | 15,879 | 1.0 | 10.2 | 16 | 323 | 15.5 (8.88 - 25.2) | 31.7 (28.3 - 35.4) |
| Open Claims | 225,549 | 821,391 | 140.2 | 775.7 | 937 | 12,428 | 6.68 (6.26 - 7.12) | 16.0 (15.7 - 16.3) |
| OEHR | 13,486 | 82,060 | 4.0 | 46.2 | 45 | 763 | 11.3 (8.26 - 15.2) | 16.5 (15.4 - 17.7) |
| Meta-analysis | 270,790 | 1,031,419 | 162.3 | 922.6 | 1,121 | 15,182 | 6.91 (6.51 - 7.32) | 16.5 (16.2 - 16.7) |
| Meta-analysis (w/o USOC) | 45,241 | 210,028 | 22.1 | 146.9 | 184 | 2,754 | 8.33 (7.17 - 9.63) | 18.7 (18.1 - 19.5) |
| **PS-matching / on-treatment TAR without escalation** | | | | | | | | |
| CCAE | 15,192 | 68,720 | 8.5 | 43.6 | 40 | 340 | 4.73 (3.38 - 6.44) | 7.79 (6.99 - 8.67) |
| ODOD | 8,692 | 53,334 | 4.4 | 39.7 | 55 | 1,128 | 12.5 (9.40 - 16.2) | 28.4 (26.7 - 30.1) |
| MDCR | 1,061 | 12,944 | 0.6 | 10.7 | 9 | 301 | 14.2 (6.47 - 26.9) | 28.1 (25.0 - 31.5) |
| MDCD | 1,797 | 14,412 | 0.8 | 7.6 | 14 | 208 | 16.9 (9.22 - 28.3) | 27.4 (23.8 - 31.4) |
| Open Claims | 173,476 | 805,988 | 104.1 | 640.4 | 684 | 9,683 | 6.57 (6.09 - 7.08) | 15.1 (14.8 - 15.4) |
| OEHR | 11,015 | 77,132 | 2.9 | 36.7 | 26 | 430 | 9.07 (5.92 - 13.3) | 11.7 (10.6 - 12.9) |
| Meta-analysis | 211,233 | 1,032,530 | 121.3 | 778.8 | 828 | 12,090 | 6.83 (6.37 - 7.31) | 15.5 (15.2 - 15.8) |
| Meta-analysis (w/o USOC) | 37,757 | 226,542 | 17.2 | 138.4 | 144 | 2,407 | 8.37 (7.06 - 9.86) | 17.4 (16.7 - 18.1) |
| **PS-stratification / on-treatment TAR without escalation** | | | | | | | | |
| CCAE | 20,403 | 70,859 | 11.0 | 44.8 | 54 | 351 | 4.92 (3.70 - 6.42) | 7.84 (7.04 - 8.71) |
| ODOD | 11,352 | 57,109 | 5.5 | 43.4 | 69 | 1,359 | 12.6 (9.82 - 16.0) | 31.3 (29.7 - 33.0) |
| MDCR | 1,192 | 16,436 | 0.7 | 13.4 | 10 | 430 | 14.6 (7.02 - 26.9) | 32.1 (29.2 - 35.3) |
| MDCD | 1,939 | 15,879 | 0.9 | 8.5 | 14 | 256 | 15.8 (8.62 - 26.4) | 30.1 (26.6 - 34.1) |
| Open Claims | 225,549 | 821,391 | 126.5 | 653.4 | 810 | 10,377 | 6.40 (5.97 - 6.86) | 15.9 (15.6 - 16.2) |
| OEHR | 13,486 | 82,060 | 3.5 | 39.4 | 30 | 527 | 8.67 (5.85 - 12.4) | 13.4 (12.3 - 14.6) |
| Meta-analysis | 270,790 | 1,031,419 | 146.4 | 781.0 | 963 | 12,614 | 6.58 (6.17 - 7.00) | 16.2 (15.9 - 16.4) |
| Meta-analysis (w/o USOC) | 45,241 | 210,028 | 19.9 | 127.6 | 153 | 2,237 | 7.69 (6.52 - 9.01) | 17.5 (16.8 - 18.3) |

##### 4.5.5 Glycemic control

**Supplementary Table 394:** Exposures, outcomes and risk estimates for Glycemic control in GLP1RA (female) (T) vs SU (female) (C) new-user cohorts under four different propensity score (PS) adjustment / time-at-risk (TAR) designs across data sources. We report total patient exposure time in 1,000 patient-year (KYr) units, uncalibrated and calibrated hazard ratios (HRs) and their 95% confidence intervals (CIs). Greyed out entries did not pass study diagnostics and are included only for completeness.

| | Patients | | Exposure (KYr) | | Outcomes | | Incidence (per KYr) | |
|---|---|---|---|---|---|---|---|---|
| | T | C | T | C | T | C | T | C |
| **PS-matching / on-treatment TAR** | | | | | | | | |
| CCAE | 15,234 | 68,978 | 9.5 | 51.0 | 20 | 285 | 2.11 (1.29 - 3.26) | 5.59 (4.96 - 6.28) |
| ODOD | 5,847 | 38,821 | 3.1 | 29.5 | 336 | 2,844 | 109 (97.8 - 121) | 96.4 (92.8 - 100.0) |
| MDCR | 1,094 | 13,448 | 0.7 | 12.3 | 0 | 41 | | 3.34 (2.39 - 4.53) |
| MDCD | 1,887 | 15,102 | 1.0 | 9.7 | 0 | 0 | | |
| Open Claims | 178,376 | 835,401 | 120.1 | 795.9 | 0 | 0 | | |
| OEHR | 11,145 | 78,232 | 3.4 | 44.2 | 0 | 0 | | |
| Meta-analysis | 21,081 | 107,799 | 12.5 | 80.5 | 356 | 3,129 | 28.4 (25.5 - 31.5) | 38.9 (37.5 - 40.3) |
| Meta-analysis (w/o USOC) | 21,081 | 107,799 | 12.5 | 80.5 | 356 | 3,129 | 28.4 (25.5 - 31.5) | 38.9 (37.5 - 40.3) |
| **PS-stratification / on-treatment TAR** | | | | | | | | |
| CCAE | 20,459 | 70,856 | 12.1 | 51.9 | 24 | 393 | 1.98 (1.27 - 2.94) | 7.57 (6.84 - 8.36) |
| ODOD | 7,237 | 42,368 | 3.6 | 32.7 | 400 | 3,230 | 110 (99.8 - 122) | 98.8 (95.4 - 102) |
| MDCR | 1,227 | 17,202 | 0.8 | 15.5 | 0 | 101 | | 6.53 (5.31 - 7.93) |
| MDCD | 2,034 | 16,863 | 1.1 | 11.0 | 0 | 0 | | |
| Open Claims | 231,281 | 852,348 | 144.1 | 811.8 | 0 | 0 | | |
| OEHR | 13,639 | 83,370 | 4.0 | 47.5 | 0 | 0 | | |
| Meta-analysis | 27,696 | 113,224 | 15.8 | 84.6 | 424 | 3,623 | 26.9 (24.4 - 29.6) | 42.8 (41.4 - 44.2) |
| Meta-analysis (w/o USOC) | 27,696 | 113,224 | 15.8 | 84.6 | 424 | 3,623 | 26.9 (24.4 - 29.6) | 42.8 (41.4 - 44.2) |
| **PS-matching / on-treatment TAR without escalation** | | | | | | | | |
| CCAE | 15,234 | 68,978 | 8.5 | 43.6 | 20 | 254 | 2.36 (1.44 - 3.64) | 5.82 (5.13 - 6.58) |
| ODOD | 5,847 | 38,821 | 2.8 | 26.2 | 305 | 2,559 | 110 (98.0 - 123) | 97.6 (93.8 - 101) |
| MDCR | 1,094 | 13,448 | 0.7 | 11.2 | 0 | 37 | | 3.30 (2.33 - 4.55) |
| MDCD | 1,887 | 15,102 | 0.9 | 8.1 | 0 | 0 | | |
| Open Claims | 178,376 | 835,401 | 107.1 | 668.8 | 0 | 0 | | |
| OEHR | 11,145 | 78,232 | 2.9 | 37.4 | 0 | 0 | | |
| Meta-analysis | 21,081 | 107,799 | 11.3 | 69.9 | 325 | 2,813 | 28.9 (25.8 - 32.2) | 40.3 (38.8 - 41.8) |
| Meta-analysis (w/o USOC) | 21,081 | 107,799 | 11.3 | 69.9 | 325 | 2,813 | 28.9 (25.8 - 32.2) | 40.3 (38.8 - 41.8) |
| **PS-stratification / on-treatment TAR without escalation** | | | | | | | | |
| CCAE | 20,459 | 70,856 | 11.0 | 44.5 | 24 | 358 | 2.18 (1.40 - 3.25) | 8.04 (7.23 - 8.92) |
| ODOD | 7,237 | 42,368 | 3.3 | 29.2 | 365 | 2,923 | 112 (100 - 124) | 100 (96.6 - 104) |
| MDCR | 1,227 | 17,202 | 0.7 | 14.1 | 0 | 92 | | 6.53 (5.26 - 8.01) |
| MDCD | 2,034 | 16,863 | 0.9 | 9.2 | 0 | 0 | | |
| Open Claims | 231,281 | 852,348 | 130.0 | 682.7 | 0 | 0 | | |
| OEHR | 13,639 | 83,370 | 3.5 | 40.3 | 0 | 0 | | |
| Meta-analysis | 27,696 | 113,224 | 14.3 | 73.7 | 389 | 3,281 | 27.3 (24.6 - 30.1) | 44.5 (43.0 - 46.1) |
| Meta-analysis (w/o USOC) | 27,696 | 113,224 | 14.3 | 73.7 | 389 | 3,281 | 27.3 (24.6 - 30.1) | 44.5 (43.0 - 46.1) |

##### 4.5.6 Hospitalization with heart failure

**Supplementary Table 395:** Exposures, outcomes and risk estimates for Hospitalization with heart failure in GLP1RA (female) (T) vs SU (female) (C) new-user cohorts under four different propensity score (PS) adjustment / time-at-risk (TAR) designs across data sources. We report total patient exposure time in 1,000 patient-year (KYr) units, uncalibrated and calibrated hazard ratios (HRs) and their 95% confidence intervals (CIs). Greyed out entries did not pass study diagnostics and are included only for completeness.

| | Patients | | Exposure (KYr) | | Outcomes | | Incidence (per KYr) | |
|---|---|---|---|---|---|---|---|---|
| | T | C | T | C | T | C | T | C |
| **PS-matching / on-treatment TAR** | | | | | | | | |
| CCAE | 15,081 | 68,367 | 9.4 | 50.6 | 17 | 321 | 1.81 (1.05 - 2.90) | 6.35 (5.67 - 7.08) |
| ODOD | 8,619 | 52,820 | 4.8 | 44.4 | 63 | 1,049 | 13.0 (10.0 - 16.7) | 23.6 (22.2 - 25.1) |
| MDCR | 1,050 | 12,178 | 0.7 | 11.0 | 10 | 333 | 14.3 (6.88 - 26.4) | 30.4 (27.2 - 33.9) |
| MDCD | 1,745 | 13,943 | 0.9 | 8.8 | 15 | 215 | 15.9 (8.87 - 26.1) | 24.4 (21.2 - 27.9) |
| Open Claims | 172,903 | 801,531 | 116.4 | 758.1 | 512 | 9,369 | 4.40 (4.02 - 4.79) | 12.4 (12.1 - 12.6) |
| OEHR | 10,956 | 76,773 | 3.3 | 42.8 | 23 | 618 | 6.94 (4.40 - 10.4) | 14.4 (13.3 - 15.6) |
| Meta-analysis | 210,354 | 1,025,612 | 135.6 | 915.7 | 640 | 11,905 | 4.72 (4.36 - 5.10) | 13.0 (12.8 - 13.2) |
| Meta-analysis (w/o USOC) | 37,451 | 224,081 | 19.2 | 157.5 | 128 | 2,536 | 6.67 (5.57 - 7.93) | 16.1 (15.5 - 16.7) |
| **PS-stratification / on-treatment TAR** | | | | | | | | |
| CCAE | 20,267 | 70,548 | 12.1 | 51.8 | 21 | 330 | 1.74 (1.08 - 2.66) | 6.36 (5.70 - 7.09) |
| ODOD | 11,260 | 56,641 | 6.0 | 48.4 | 66 | 1,270 | 11.1 (8.55 - 14.1) | 26.2 (24.8 - 27.7) |
| MDCR | 1,169 | 15,734 | 0.7 | 14.0 | 10 | 530 | 13.5 (6.46 - 24.8) | 37.8 (34.7 - 41.2) |
| MDCD | 1,878 | 15,407 | 1.0 | 9.9 | 16 | 250 | 15.9 (9.07 - 25.8) | 25.2 (22.2 - 28.6) |
| Open Claims | 224,856 | 815,621 | 140.1 | 771.4 | 587 | 10,030 | 4.19 (3.86 - 4.54) | 13.0 (12.7 - 13.3) |
| OEHR | 13,411 | 81,589 | 4.0 | 45.8 | 25 | 751 | 6.33 (4.09 - 9.34) | 16.4 (15.2 - 17.6) |
| Meta-analysis | 269,794 | 1,024,399 | 162.0 | 917.5 | 699 | 12,381 | 4.31 (4.00 - 4.65) | 13.5 (13.3 - 13.7) |
| Meta-analysis (w/o USOC) | 44,938 | 208,778 | 22.0 | 146.1 | 112 | 2,351 | 5.10 (4.20 - 6.13) | 16.1 (15.5 - 16.8) |
| **PS-matching / on-treatment TAR without escalation** | | | | | | | | |
| CCAE | 15,081 | 68,367 | 8.4 | 43.4 | 13 | 281 | 1.54 (0.82 - 2.64) | 6.48 (5.74 - 7.28) |
| ODOD | 8,619 | 52,820 | 4.4 | 39.4 | 56 | 913 | 12.8 (9.66 - 16.6) | 23.2 (21.7 - 24.7) |
| MDCR | 1,050 | 12,178 | 0.6 | 10.0 | 9 | 293 | 14.2 (6.48 - 26.9) | 29.2 (26.0 - 32.8) |
| MDCD | 1,745 | 13,943 | 0.8 | 7.3 | 13 | 172 | 16.0 (8.51 - 27.3) | 23.5 (20.1 - 27.3) |
| Open Claims | 172,903 | 801,531 | 104.0 | 637.8 | 438 | 7,864 | 4.21 (3.83 - 4.63) | 12.3 (12.1 - 12.6) |
| OEHR | 10,956 | 76,773 | 2.9 | 36.5 | 18 | 449 | 6.30 (3.74 - 9.96) | 12.3 (11.2 - 13.5) |
| Meta-analysis | 210,354 | 1,025,612 | 121.1 | 774.4 | 547 | 9,972 | 4.52 (4.15 - 4.91) | 12.9 (12.6 - 13.1) |
| Meta-analysis (w/o USOC) | 37,451 | 224,081 | 17.1 | 136.6 | 109 | 2,108 | 6.37 (5.23 - 7.68) | 15.4 (14.8 - 16.1) |
| **PS-stratification / on-treatment TAR without escalation** | | | | | | | | |
| CCAE | 20,267 | 70,548 | 10.9 | 44.6 | 15 | 287 | 1.37 (0.77 - 2.26) | 6.44 (5.72 - 7.23) |
| ODOD | 11,260 | 56,641 | 5.4 | 43.1 | 58 | 1,110 | 10.7 (8.11 - 13.8) | 25.7 (24.2 - 27.3) |
| MDCR | 1,169 | 15,734 | 0.7 | 12.8 | 9 | 475 | 13.2 (6.06 - 25.1) | 37.0 (33.8 - 40.5) |
| MDCD | 1,878 | 15,407 | 0.9 | 8.2 | 13 | 201 | 15.0 (7.97 - 25.6) | 24.4 (21.1 - 28.0) |
| Open Claims | 224,856 | 815,621 | 126.4 | 649.4 | 507 | 8,427 | 4.01 (3.67 - 4.37) | 13.0 (12.7 - 13.3) |
| OEHR | 13,411 | 81,589 | 3.4 | 39.1 | 20 | 561 | 5.81 (3.55 - 8.97) | 14.3 (13.2 - 15.6) |
| Meta-analysis | 269,794 | 1,024,399 | 146.2 | 776.2 | 600 | 10,385 | 4.10 (3.78 - 4.44) | 13.4 (13.1 - 13.6) |
| Meta-analysis (w/o USOC) | 44,938 | 208,778 | 19.8 | 126.8 | 93 | 1,958 | 4.70 (3.79 - 5.75) | 15.4 (14.8 - 16.1) |

##### 4.5.7 Measured renal dysfunction

**Supplementary Table 396:** Exposures, outcomes and risk estimates for Measured renal dysfunction in GLP1RA (female) (T) vs SU (female) (C) new-user cohorts under four different propensity score (PS) adjustment / time-at-risk (TAR) designs across data sources. We report total patient exposure time in 1,000 patient-year (KYr) units, uncalibrated and calibrated hazard ratios (HRs) and their 95% confidence intervals (CIs). Greyed out entries did not pass study diagnostics and are included only for completeness.

| | Patients | | Exposure (KYr) | | Outcomes | | Incidence (per KYr) | |
|---|---|---|---|---|---|---|---|---|
| | T | C | T | C | T | C | T | C |
| **PS-matching / on-treatment TAR** | | | | | | | | |
| CCAE | 15,319 | 69,514 | 9.5 | 51.6 | <5 | 17 | < 0.52 (0.17 - 1.22) | 0.33 (0.19 - 0.53) |
| ODOD | 8,942 | 55,439 | 5.0 | 47.1 | <5 | 55 | < 0.99 (0.32 - 2.32) | 1.17 (0.88 - 1.52) |
| MDCR | 1,098 | 13,508 | 0.7 | 12.4 | 0 | <5 | | < 0.40 (0.13 - 0.94) |
| MDCD | 1,887 | 15,102 | 1.0 | 9.7 | 0 | 0 | | |
| Open Claims | 178,376 | 835,401 | 120.1 | 795.9 | 0 | 0 | | |
| OEHR | 11,145 | 78,232 | 3.4 | 44.2 | 0 | 0 | | |
| Meta-analysis | 8,942 | 55,439 | 5.0 | 47.1 | <5 | 55 | < 0.99 (0.32 - 2.32) | 1.17 (0.88 - 1.52) |
| Meta-analysis (w/o USOC) | 8,942 | 55,439 | 5.0 | 47.1 | <5 | 55 | < 0.99 (0.32 - 2.32) | 1.17 (0.88 - 1.52) |
| **PS-stratification / on-treatment TAR** | | | | | | | | |
| CCAE | 20,567 | 71,860 | 12.2 | 52.9 | <5 | 17 | < 0.41 (0.13 - 0.95) | 0.32 (0.19 - 0.51) |
| ODOD | 11,651 | 60,230 | 6.2 | 52.2 | <5 | 82 | < 0.81 (0.26 - 1.88) | 1.57 (1.25 - 1.95) |
| MDCR | 1,231 | 17,494 | 0.8 | 15.8 | 0 | <5 | | < 0.32 (0.10 - 0.74) |
| MDCD | 2,034 | 16,863 | 1.1 | 11.0 | 0 | 0 | | |
| Open Claims | 231,281 | 852,348 | 144.1 | 811.8 | 0 | 0 | | |
| OEHR | 13,639 | 83,370 | 4.0 | 47.5 | 0 | 0 | | |
| Meta-analysis | 11,651 | 60,230 | 6.2 | 52.2 | <5 | 82 | < 0.81 (0.26 - 1.88) | 1.57 (1.25 - 1.95) |
| Meta-analysis (w/o USOC) | 11,651 | 60,230 | 6.2 | 52.2 | <5 | 82 | < 0.81 (0.26 - 1.88) | 1.57 (1.25 - 1.95) |
| **PS-matching / on-treatment TAR without escalation** | | | | | | | | |
| CCAE | 15,319 | 69,514 | 8.6 | 44.2 | <5 | 14 | < 0.58 (0.19 - 1.36) | 0.32 (0.17 - 0.53) |
| ODOD | 8,942 | 55,439 | 4.5 | 41.8 | <5 | 44 | < 1.10 (0.36 - 2.57) | 1.05 (0.76 - 1.41) |
| MDCR | 1,098 | 13,508 | 0.7 | 11.3 | 0 | <5 | | < 0.44 (0.14 - 1.03) |
| MDCD | 1,887 | 15,102 | 0.9 | 8.1 | 0 | 0 | | |
| Open Claims | 178,376 | 835,401 | 107.1 | 668.8 | 0 | 0 | | |
| OEHR | 11,145 | 78,232 | 2.9 | 37.4 | 0 | 0 | | |
| Meta-analysis | 8,942 | 55,439 | 4.5 | 41.8 | <5 | 44 | < 1.10 (0.36 - 2.57) | 1.05 (0.76 - 1.41) |
| Meta-analysis (w/o USOC) | 8,942 | 55,439 | 4.5 | 41.8 | <5 | 44 | < 1.10 (0.36 - 2.57) | 1.05 (0.76 - 1.41) |
| **PS-stratification / on-treatment TAR without escalation** | | | | | | | | |
| CCAE | 20,567 | 71,860 | 11.1 | 45.4 | <5 | 14 | < 0.45 (0.15 - 1.05) | 0.31 (0.17 - 0.52) |
| ODOD | 11,651 | 60,230 | 5.6 | 46.4 | <5 | 70 | < 0.89 (0.29 - 2.07) | 1.51 (1.18 - 1.91) |
| MDCR | 1,231 | 17,494 | 0.7 | 14.4 | 0 | <5 | | < 0.35 (0.11 - 0.81) |
| MDCD | 2,034 | 16,863 | 0.9 | 9.2 | 0 | 0 | | |
| Open Claims | 231,281 | 852,348 | 130.0 | 682.7 | 0 | 0 | | |
| OEHR | 13,639 | 83,370 | 3.5 | 40.3 | 0 | 0 | | |
| Meta-analysis | 11,651 | 60,230 | 5.6 | 46.4 | <5 | 70 | < 0.89 (0.29 - 2.07) | 1.51 (1.18 - 1.91) |
| Meta-analysis (w/o USOC) | 11,651 | 60,230 | 5.6 | 46.4 | <5 | 70 | < 0.89 (0.29 - 2.07) | 1.51 (1.18 - 1.91) |

##### 4.5.8 Stroke

**Supplementary Table 397:** Exposures, outcomes and risk estimates for Stroke in GLP1RA (female) (T) vs SU (female) (C) new-user cohorts under four different propensity score (PS) adjustment / time-at-risk (TAR) designs across data sources. We report total patient exposure time in 1,000 patient-year (KYr) units, uncalibrated and calibrated hazard ratios (HRs) and their 95% confidence intervals (CIs). Greyed out entries did not pass study diagnostics and are included only for completeness.

| | Patients | | Exposure (KYr) | | Outcomes | | Incidence (per KYr) | |
|---|---|---|---|---|---|---|---|---|
| | T | C | T | C | T | C | T | C |
| **PS-matching / on-treatment TAR** | | | | | | | | |
| CCAE | 15,170 | 68,693 | 9.4 | 50.9 | 24 | 188 | 2.55 (1.63 - 3.79) | 3.70 (3.19 - 4.26) |
| ODOD | 8,800 | 54,222 | 4.9 | 45.8 | 28 | 491 | 5.68 (3.78 - 8.21) | 10.7 (9.79 - 11.7) |
| MDCR | 1,067 | 12,540 | 0.7 | 11.4 | <5 | 197 | < 7.13 (2.32 - 16.6) | 17.2 (14.9 - 19.8) |
| MDCD | 1,835 | 14,600 | 1.0 | 9.3 | 8 | 111 | 8.01 (3.46 - 15.8) | 11.9 (9.78 - 14.3) |
| Open Claims | 174,625 | 812,865 | 117.7 | 772.1 | 338 | 5,028 | 2.87 (2.57 - 3.20) | 6.51 (6.33 - 6.69) |
| OEHR | 11,054 | 77,589 | 3.3 | 43.7 | 8 | 248 | 2.40 (1.03 - 4.72) | 5.68 (4.99 - 6.43) |
| Meta-analysis | 212,551 | 1,040,509 | 137.1 | 933.2 | <411 | 6,263 | < 3.00 (2.72 - 3.30) | 6.71 (6.55 - 6.88) |
| Meta-analysis (w/o USOC) | 37,926 | 227,644 | 19.4 | 161.1 | <73 | 1,235 | < 3.76 (2.95 - 4.73) | 7.66 (7.24 - 8.10) |
| **PS-stratification / on-treatment TAR** | | | | | | | | |
| CCAE | 20,377 | 70,931 | 12.1 | 52.2 | 32 | 192 | 2.64 (1.81 - 3.73) | 3.68 (3.18 - 4.24) |
| ODOD | 11,499 | 58,487 | 6.1 | 50.3 | 35 | 633 | 5.76 (4.02 - 8.02) | 12.6 (11.6 - 13.6) |
| MDCR | 1,195 | 16,620 | 0.8 | 14.9 | <5 | 303 | < 6.66 (2.16 - 15.5) | 20.3 (18.1 - 22.8) |
| MDCD | 1,977 | 16,180 | 1.1 | 10.5 | 8 | 129 | 7.52 (3.25 - 14.8) | 12.2 (10.2 - 14.5) |
| Open Claims | 226,789 | 828,014 | 141.4 | 786.3 | 396 | 5,350 | 2.80 (2.53 - 3.09) | 6.80 (6.62 - 6.99) |
| OEHR | 13,542 | 82,512 | 4.0 | 46.8 | 9 | 295 | 2.26 (1.03 - 4.29) | 6.30 (5.60 - 7.07) |
| Meta-analysis | 272,207 | 1,039,944 | 163.5 | 935.6 | 472 | 6,470 | 2.89 (2.63 - 3.16) | 6.92 (6.75 - 7.09) |
| Meta-analysis (w/o USOC) | 45,418 | 211,930 | 22.2 | 149.3 | 76 | 1,120 | 3.43 (2.70 - 4.29) | 7.50 (7.07 - 7.96) |
| **PS-matching / on-treatment TAR without escalation** | | | | | | | | |
| CCAE | 15,170 | 68,693 | 8.5 | 43.6 | 19 | 158 | 2.25 (1.35 - 3.51) | 3.62 (3.08 - 4.24) |
| ODOD | 8,800 | 54,222 | 4.5 | 40.7 | 24 | 436 | 5.38 (3.45 - 8.01) | 10.7 (9.73 - 11.8) |
| MDCR | 1,067 | 12,540 | 0.6 | 10.4 | <5 | 175 | < 7.84 (2.55 - 18.3) | 16.8 (14.4 - 19.5) |
| MDCD | 1,835 | 14,600 | 0.9 | 7.7 | 7 | 94 | 8.16 (3.28 - 16.8) | 12.1 (9.81 - 14.9) |
| Open Claims | 174,625 | 812,865 | 105.0 | 649.1 | 291 | 4,259 | 2.77 (2.46 - 3.11) | 6.56 (6.37 - 6.76) |
| OEHR | 11,054 | 77,589 | 2.9 | 37.0 | 6 | 190 | 2.09 (0.77 - 4.55) | 5.13 (4.43 - 5.91) |
| Meta-analysis | 212,551 | 1,040,509 | 122.2 | 788.6 | <352 | 5,312 | < 2.88 (2.59 - 3.20) | 6.74 (6.56 - 6.92) |
| Meta-analysis (w/o USOC) | 37,926 | 227,644 | 17.3 | 139.5 | <61 | 1,053 | < 3.53 (2.70 - 4.53) | 7.55 (7.10 - 8.02) |
| **PS-stratification / on-treatment TAR without escalation** | | | | | | | | |
| CCAE | 20,377 | 70,931 | 11.0 | 44.8 | 24 | 161 | 2.19 (1.40 - 3.25) | 3.59 (3.06 - 4.19) |
| ODOD | 11,499 | 58,487 | 5.5 | 44.8 | 31 | 564 | 5.61 (3.81 - 7.96) | 12.6 (11.6 - 13.7) |
| MDCR | 1,195 | 16,620 | 0.7 | 13.6 | <5 | 272 | < 7.27 (2.36 - 17.0) | 20.0 (17.7 - 22.5) |
| MDCD | 1,977 | 16,180 | 0.9 | 8.8 | 7 | 107 | 7.64 (3.07 - 15.7) | 12.2 (10.0 - 14.8) |
| Open Claims | 226,789 | 828,014 | 127.5 | 661.6 | 348 | 4,533 | 2.73 (2.45 - 3.03) | 6.85 (6.65 - 7.05) |
| OEHR | 13,542 | 82,512 | 3.5 | 39.7 | 7 | 223 | 2.02 (0.81 - 4.16) | 5.61 (4.90 - 6.40) |
| Meta-analysis | 272,207 | 1,039,944 | 147.5 | 790.9 | 410 | 5,481 | 2.78 (2.52 - 3.06) | 6.93 (6.75 - 7.12) |
| Meta-analysis (w/o USOC) | 45,418 | 211,930 | 20.0 | 129.3 | 62 | 948 | 3.11 (2.38 - 3.98) | 7.33 (6.87 - 7.81) |

##### 4.5.9 Sudden cardiac death

**Supplementary Table 398:** Exposures, outcomes and risk estimates for Sudden cardiac death in GLP1RA (female) (T) vs SU (female) (C) new-user cohorts under four different propensity score (PS) adjustment / time-at-risk (TAR) designs across data sources. We report total patient exposure time in 1,000 patient-year (KYr) units, uncalibrated and calibrated hazard ratios (HRs) and their 95% confidence intervals (CIs). Greyed out entries did not pass study diagnostics and are included only for completeness.

| | Patients | | Exposure (KYr) | | Outcomes | | Incidence (per KYr) | |
|---|---|---|---|---|---|---|---|---|
| | T | C | T | C | T | C | T | C |
| **PS-matching / on-treatment TAR** | | | | | | | | |
| CCAE | 15,327 | 69,542 | 9.5 | 51.6 | 6 | 52 | 0.63 (0.23 - 1.37) | 1.01 (0.75 - 1.32) |
| ODOD | 8,947 | 55,512 | 5.0 | 47.2 | 8 | 122 | 1.59 (0.69 - 3.13) | 2.58 (2.15 - 3.09) |
| MDCR | 1,095 | 13,395 | 0.7 | 12.3 | 0 | 32 | | 2.61 (1.79 - 3.69) |
| MDCD | 1,886 | 15,066 | 1.0 | 9.7 | 0 | 29 | | 2.99 (2.00 - 4.29) |
| Open Claims | 178,120 | 834,180 | 119.9 | 794.5 | 58 | 1,004 | 0.48 (0.37 - 0.63) | 1.26 (1.19 - 1.34) |
| OEHR | 11,139 | 78,206 | 3.4 | 44.2 | <5 | 55 | < 1.48 (0.48 - 3.46) | 1.24 (0.94 - 1.62) |
| Meta-analysis | 213,533 | 1,037,440 | 137.9 | 937.5 | <77 | 1,233 | < 0.56 (0.44 - 0.70) | 1.32 (1.24 - 1.39) |
| Meta-analysis (w/o USOC) | 35,413 | 203,260 | 17.9 | 143.0 | <19 | 229 | < 1.06 (0.64 - 1.65) | 1.60 (1.40 - 1.82) |
| **PS-stratification / on-treatment TAR** | | | | | | | | |
| CCAE | 20,573 | 71,879 | 12.2 | 52.9 | 6 | 53 | 0.49 (0.18 - 1.07) | 1.00 (0.75 - 1.31) |
| ODOD | 11,657 | 60,285 | 6.2 | 52.3 | 8 | 163 | 1.29 (0.56 - 2.54) | 3.12 (2.66 - 3.64) |
| MDCR | 1,227 | 17,461 | 0.8 | 15.8 | 0 | 54 | | 3.42 (2.57 - 4.47) |
| MDCD | 2,033 | 16,806 | 1.1 | 11.0 | 0 | 35 | | 3.18 (2.21 - 4.42) |
| Open Claims | 230,968 | 850,946 | 143.9 | 810.3 | 66 | 1,077 | 0.46 (0.35 - 0.58) | 1.33 (1.25 - 1.41) |
| OEHR | 13,632 | 83,333 | 4.0 | 47.5 | <5 | 81 | < 1.24 (0.40 - 2.90) | 1.71 (1.36 - 2.12) |
| Meta-analysis | 276,830 | 1,066,443 | 166.4 | 963.0 | <85 | 1,374 | < 0.51 (0.41 - 0.63) | 1.43 (1.35 - 1.50) |
| Meta-analysis (w/o USOC) | 45,862 | 215,497 | 22.5 | 152.7 | <19 | 297 | < 0.85 (0.51 - 1.32) | 1.95 (1.73 - 2.18) |
| **PS-matching / on-treatment TAR without escalation** | | | | | | | | |
| CCAE | 15,327 | 69,542 | 8.6 | 44.2 | 5 | 38 | 0.58 (0.19 - 1.36) | 0.86 (0.61 - 1.18) |
| ODOD | 8,947 | 55,512 | 4.5 | 41.9 | 8 | 104 | 1.76 (0.76 - 3.47) | 2.48 (2.03 - 3.01) |
| MDCR | 1,095 | 13,395 | 0.6 | 11.2 | 0 | 29 | | 2.59 (1.74 - 3.72) |
| MDCD | 1,886 | 15,066 | 0.9 | 8.0 | 0 | 25 | | 3.11 (2.01 - 4.59) |
| Open Claims | 178,120 | 834,180 | 107.0 | 667.7 | 49 | 841 | 0.46 (0.34 - 0.61) | 1.26 (1.18 - 1.35) |
| OEHR | 11,139 | 78,206 | 2.9 | 37.4 | <5 | 36 | < 1.72 (0.56 - 4.02) | 0.96 (0.67 - 1.33) |
| Meta-analysis | 213,533 | 1,037,440 | 123.0 | 791.2 | <67 | 1,019 | < 0.54 (0.42 - 0.69) | 1.29 (1.21 - 1.37) |
| Meta-analysis (w/o USOC) | 35,413 | 203,260 | 16.0 | 123.5 | <18 | 178 | < 1.12 (0.67 - 1.78) | 1.44 (1.24 - 1.67) |
| **PS-stratification / on-treatment TAR without escalation** | | | | | | | | |
| CCAE | 20,573 | 71,879 | 11.1 | 45.4 | 5 | 39 | 0.45 (0.15 - 1.05) | 0.86 (0.61 - 1.17) |
| ODOD | 11,657 | 60,285 | 5.6 | 46.5 | 8 | 141 | 1.42 (0.61 - 2.80) | 3.03 (2.55 - 3.58) |
| MDCR | 1,227 | 17,461 | 0.7 | 14.4 | 0 | 50 | | 3.48 (2.58 - 4.59) |
| MDCD | 2,033 | 16,806 | 0.9 | 9.1 | 0 | 31 | | 3.39 (2.30 - 4.81) |
| Open Claims | 230,968 | 850,946 | 129.8 | 681.5 | 57 | 904 | 0.44 (0.33 - 0.57) | 1.33 (1.24 - 1.42) |
| OEHR | 13,632 | 83,333 | 3.5 | 40.3 | <5 | 49 | < 1.43 (0.46 - 3.33) | 1.22 (0.90 - 1.61) |
| Meta-analysis | 276,830 | 1,066,443 | 150.0 | 813.7 | <75 | 1,133 | < 0.50 (0.39 - 0.63) | 1.39 (1.31 - 1.48) |
| Meta-analysis (w/o USOC) | 45,862 | 215,497 | 20.2 | 132.2 | <18 | 229 | < 0.89 (0.53 - 1.41) | 1.73 (1.52 - 1.97) |

##### 4.5.10 Abnormal weight gain

**Supplementary Table 399:** Exposures, outcomes and risk estimates for Abnormal weight gain in GLP1RA (female) (T) vs SU (female) (C) new-user cohorts under four different propensity score (PS) adjustment / time-at-risk (TAR) designs across data sources. We report total patient exposure time in 1,000 patient-year (KYr) units, uncalibrated and calibrated hazard ratios (HRs) and their 95% confidence intervals (CIs). Greyed out entries did not pass study diagnostics and are included only for completeness.

| | Patients | | Exposure (KYr) | | Outcomes | | Incidence (per KYr) | |
|---|---|---|---|---|---|---|---|---|
| | T | C | T | C | T | C | T | C |
| **PS-matching / on-treatment TAR** | | | | | | | | |
| CCAE | 13,857 | 66,170 | 8.5 | 49.0 | 83 | 394 | 9.72 (7.75 - 12.1) | 8.04 (7.26 - 8.87) |
| ODOD | 8,189 | 52,870 | 4.6 | 44.7 | 50 | 409 | 11.0 (8.15 - 14.5) | 9.14 (8.28 - 10.1) |
| MDCR | 1,061 | 13,346 | 0.7 | 12.2 | <5 | 35 | < 7.17 (2.33 - 16.7) | 2.87 (2.00 - 4.00) |
| MDCD | 1,632 | 13,899 | 0.9 | 8.9 | 13 | 118 | 14.7 (7.82 - 25.1) | 13.3 (11.0 - 15.9) |
| Open Claims | 161,505 | 795,705 | 108.4 | 758.2 | 413 | 2,381 | 3.81 (3.45 - 4.20) | 3.14 (3.02 - 3.27) |
| OEHR | 10,162 | 74,811 | 3.1 | 41.9 | 34 | 397 | 11.1 (7.69 - 15.5) | 9.47 (8.57 - 10.5) |
| Meta-analysis | 195,345 | 1,003,455 | 125.4 | 902.7 | 593 | 3,699 | 4.73 (4.35 - 5.12) | 4.10 (3.97 - 4.23) |
| Meta-analysis (w/o USOC) | 33,840 | 207,750 | 17.0 | 144.5 | 180 | 1,318 | 10.6 (9.08 - 12.2) | 9.12 (8.63 - 9.62) |
| **PS-stratification / on-treatment TAR** | | | | | | | | |
| CCAE | 17,829 | 68,489 | 10.6 | 50.4 | 116 | 399 | 11.0 (9.06 - 13.2) | 7.92 (7.16 - 8.74) |
| ODOD | 10,179 | 57,785 | 5.4 | 49.9 | 61 | 452 | 11.4 (8.69 - 14.6) | 9.06 (8.25 - 9.94) |
| MDCR | 1,171 | 17,288 | 0.7 | 15.6 | <5 | 55 | < 6.79 (2.21 - 15.9) | 3.53 (2.66 - 4.59) |
| MDCD | 1,722 | 15,658 | 0.9 | 10.2 | 13 | 130 | 14.1 (7.50 - 24.1) | 12.7 (10.6 - 15.1) |
| Open Claims | 202,244 | 813,475 | 126.4 | 775.1 | 549 | 2,456 | 4.34 (3.99 - 4.72) | 3.17 (3.04 - 3.30) |
| OEHR | 12,039 | 80,074 | 3.5 | 45.2 | 53 | 432 | 15.0 (11.3 - 19.7) | 9.56 (8.68 - 10.5) |
| Meta-analysis | 242,291 | 1,019,823 | 145.9 | 920.6 | 779 | 3,739 | 5.34 (4.97 - 5.73) | 4.06 (3.93 - 4.19) |
| Meta-analysis (w/o USOC) | 40,047 | 206,348 | 19.5 | 145.4 | 230 | 1,283 | 11.8 (10.3 - 13.4) | 8.82 (8.35 - 9.32) |
| **PS-matching / on-treatment TAR without escalation** | | | | | | | | |
| CCAE | 13,857 | 66,170 | 7.7 | 42.1 | 75 | 325 | 9.76 (7.68 - 12.2) | 7.72 (6.90 - 8.61) |
| ODOD | 8,189 | 52,870 | 4.1 | 39.8 | 45 | 346 | 10.9 (7.98 - 14.6) | 8.70 (7.80 - 9.66) |
| MDCR | 1,061 | 13,346 | 0.6 | 11.1 | <5 | 30 | < 7.90 (2.56 - 18.4) | 2.70 (1.82 - 3.85) |
| MDCD | 1,632 | 13,899 | 0.8 | 7.4 | 11 | 99 | 14.4 (7.21 - 25.8) | 13.4 (10.9 - 16.3) |
| Open Claims | 161,505 | 795,705 | 96.7 | 638.6 | 369 | 1,959 | 3.82 (3.44 - 4.23) | 3.07 (2.93 - 3.21) |
| OEHR | 10,162 | 74,811 | 2.6 | 35.6 | 29 | 310 | 10.9 (7.33 - 15.7) | 8.71 (7.77 - 9.74) |
| Meta-analysis | 195,345 | 1,003,455 | 111.9 | 763.5 | 529 | 3,039 | 4.73 (4.33 - 5.15) | 3.98 (3.84 - 4.12) |
| Meta-analysis (w/o USOC) | 33,840 | 207,750 | 15.2 | 124.9 | 160 | 1,080 | 10.5 (8.95 - 12.3) | 8.65 (8.14 - 9.18) |
| **PS-stratification / on-treatment TAR without escalation** | | | | | | | | |
| CCAE | 17,829 | 68,489 | 9.6 | 43.4 | 107 | 330 | 11.2 (9.15 - 13.5) | 7.61 (6.81 - 8.48) |
| ODOD | 10,179 | 57,785 | 4.9 | 44.5 | 56 | 386 | 11.5 (8.67 - 14.9) | 8.68 (7.83 - 9.59) |
| MDCR | 1,171 | 17,288 | 0.7 | 14.2 | <5 | 48 | < 7.44 (2.42 - 17.4) | 3.37 (2.49 - 4.47) |
| MDCD | 1,722 | 15,658 | 0.8 | 8.5 | 11 | 110 | 13.8 (6.90 - 24.7) | 12.9 (10.6 - 15.6) |
| Open Claims | 202,244 | 813,475 | 113.9 | 653.4 | 499 | 2,028 | 4.38 (4.01 - 4.78) | 3.10 (2.97 - 3.24) |
| OEHR | 12,039 | 80,074 | 3.1 | 38.5 | 46 | 337 | 15.0 (11.0 - 20.0) | 8.76 (7.85 - 9.75) |
| Meta-analysis | 242,291 | 1,019,823 | 131.4 | 779.7 | 708 | 3,081 | 5.39 (5.00 - 5.80) | 3.95 (3.81 - 4.09) |
| Meta-analysis (w/o USOC) | 40,047 | 206,348 | 17.5 | 126.3 | 209 | 1,053 | 11.9 (10.4 - 13.6) | 8.34 (7.84 - 8.86) |

##### 4.5.11 Abnormal weight loss

**Supplementary Table 400:** Exposures, outcomes and risk estimates for Abnormal weight loss in GLP1RA (female) (T) vs SU (female) (C) new-user cohorts under four different propensity score (PS) adjustment / time-at-risk (TAR) designs across data sources. We report total patient exposure time in 1,000 patient-year (KYr) units, uncalibrated and calibrated hazard ratios (HRs) and their 95% confidence intervals (CIs). Greyed out entries did not pass study diagnostics and are included only for completeness.

| | Patients | | Exposure (KYr) | | Outcomes | | Incidence (per KYr) | |
|---|---|---|---|---|---|---|---|---|
| | T | C | T | C | T | C | T | C |
| **PS-matching / on-treatment TAR** | | | | | | | | |
| CCAE | 15,023 | 68,111 | 9.3 | 50.4 | 77 | 255 | 8.28 (6.54 - 10.4) | 5.05 (4.45 - 5.71) |
| ODOD | 8,688 | 53,392 | 4.9 | 45.0 | 74 | 599 | 15.2 (11.9 - 19.1) | 13.3 (12.3 - 14.4) |
| MDCR | 1,076 | 13,152 | 0.7 | 12.0 | 12 | 127 | 17.1 (8.81 - 29.8) | 10.6 (8.85 - 12.6) |
| MDCD | 1,813 | 14,392 | 1.0 | 9.1 | 19 | 141 | 19.5 (11.7 - 30.4) | 15.4 (13.0 - 18.2) |
| Open Claims | 172,652 | 806,459 | 115.9 | 765.8 | 591 | 3,941 | 5.10 (4.70 - 5.53) | 5.15 (4.99 - 5.31) |
| OEHR | 10,878 | 76,256 | 3.3 | 42.7 | 27 | 458 | 8.24 (5.43 - 12.0) | 10.7 (9.77 - 11.8) |
| Meta-analysis | 210,130 | 1,031,762 | 135.0 | 925.0 | 800 | 5,521 | 5.92 (5.52 - 6.35) | 5.97 (5.81 - 6.13) |
| Meta-analysis (w/o USOC) | 37,478 | 225,303 | 19.1 | 159.3 | 209 | 1,580 | 10.9 (9.50 - 12.5) | 9.92 (9.44 - 10.4) |
| **PS-stratification / on-treatment TAR** | | | | | | | | |
| CCAE | 20,123 | 70,373 | 11.9 | 51.8 | 107 | 264 | 8.98 (7.36 - 10.9) | 5.10 (4.50 - 5.75) |
| ODOD | 11,297 | 57,469 | 6.0 | 49.3 | 88 | 704 | 14.7 (11.8 - 18.1) | 14.3 (13.2 - 15.4) |
| MDCR | 1,208 | 16,711 | 0.8 | 15.0 | 12 | 183 | 15.9 (8.22 - 27.8) | 12.2 (10.5 - 14.1) |
| MDCD | 1,955 | 16,069 | 1.0 | 10.4 | 20 | 167 | 19.3 (11.8 - 29.8) | 16.1 (13.7 - 18.7) |
| Open Claims | 223,395 | 821,471 | 138.9 | 779.9 | 739 | 4,151 | 5.32 (4.94 - 5.72) | 5.32 (5.16 - 5.49) |
| OEHR | 13,292 | 80,965 | 3.9 | 45.7 | 34 | 527 | 8.71 (6.03 - 12.2) | 11.5 (10.6 - 12.6) |
| Meta-analysis | 268,107 | 1,030,278 | 160.7 | 926.6 | 968 | 5,646 | 6.02 (5.65 - 6.41) | 6.09 (5.94 - 6.25) |
| Meta-analysis (w/o USOC) | 44,712 | 208,807 | 21.8 | 146.7 | 229 | 1,495 | 10.5 (9.19 - 12.0) | 10.2 (9.68 - 10.7) |
| **PS-matching / on-treatment TAR without escalation** | | | | | | | | |
| CCAE | 15,023 | 68,111 | 8.4 | 43.2 | 63 | 210 | 7.54 (5.80 - 9.65) | 4.86 (4.22 - 5.56) |
| ODOD | 8,688 | 53,392 | 4.4 | 40.0 | 67 | 533 | 15.2 (11.8 - 19.3) | 13.3 (12.2 - 14.5) |
| MDCR | 1,076 | 13,152 | 0.6 | 10.9 | 11 | 114 | 17.2 (8.60 - 30.8) | 10.4 (8.62 - 12.6) |
| MDCD | 1,813 | 14,392 | 0.8 | 7.6 | 13 | 123 | 15.5 (8.25 - 26.5) | 16.2 (13.5 - 19.3) |
| Open Claims | 172,652 | 806,459 | 103.5 | 643.9 | 528 | 3,309 | 5.10 (4.68 - 5.56) | 5.14 (4.97 - 5.32) |
| OEHR | 10,878 | 76,256 | 2.8 | 36.2 | 21 | 355 | 7.44 (4.61 - 11.4) | 9.80 (8.81 - 10.9) |
| Meta-analysis | 210,130 | 1,031,762 | 120.5 | 781.9 | 703 | 4,644 | 5.83 (5.41 - 6.28) | 5.94 (5.77 - 6.11) |
| Meta-analysis (w/o USOC) | 37,478 | 225,303 | 17.0 | 137.9 | 175 | 1,335 | 10.3 (8.80 - 11.9) | 9.68 (9.17 - 10.2) |
| **PS-stratification / on-treatment TAR without escalation** | | | | | | | | |
| CCAE | 20,123 | 70,373 | 10.8 | 44.5 | 89 | 219 | 8.24 (6.61 - 10.1) | 4.93 (4.29 - 5.62) |
| ODOD | 11,297 | 57,469 | 5.4 | 43.9 | 79 | 628 | 14.6 (11.5 - 18.1) | 14.3 (13.2 - 15.5) |
| MDCR | 1,208 | 16,711 | 0.7 | 13.7 | 11 | 167 | 16.0 (7.97 - 28.6) | 12.2 (10.4 - 14.2) |
| MDCD | 1,955 | 16,069 | 0.9 | 8.6 | 13 | 146 | 14.5 (7.74 - 24.8) | 16.9 (14.3 - 19.9) |
| Open Claims | 223,395 | 821,471 | 125.4 | 656.3 | 666 | 3,496 | 5.31 (4.92 - 5.73) | 5.33 (5.15 - 5.51) |
| OEHR | 13,292 | 80,965 | 3.4 | 38.8 | 28 | 413 | 8.23 (5.47 - 11.9) | 10.6 (9.64 - 11.7) |
| Meta-analysis | 268,107 | 1,030,278 | 145.0 | 783.4 | 862 | 4,756 | 5.94 (5.55 - 6.36) | 6.07 (5.90 - 6.25) |
| Meta-analysis (w/o USOC) | 44,712 | 208,807 | 19.6 | 127.1 | 196 | 1,260 | 9.98 (8.63 - 11.5) | 9.91 (9.37 - 10.5) |

##### 4.5.12 Acute pancreatitis

**Supplementary Table 401:** Exposures, outcomes and risk estimates for Acute pancreatitis in GLP1RA (female) (T) vs SU (female) (C) new-user cohorts under four different propensity score (PS) adjustment / time-at-risk (TAR) designs across data sources. We report total patient exposure time in 1,000 patient-year (KYr) units, uncalibrated and calibrated hazard ratios (HRs) and their 95% confidence intervals (CIs). Greyed out entries did not pass study diagnostics and are included only for completeness.

| | Patients | | Exposure (KYr) | | Outcomes | | Incidence (per KYr) | |
|---|---|---|---|---|---|---|---|---|
| | T | C | T | C | T | C | T | C |
| **PS-matching / on-treatment TAR** | | | | | | | | |
| CCAE | 15,250 | 68,961 | 9.5 | 51.1 | 15 | 119 | 1.58 (0.88 - 2.60) | 2.33 (1.93 - 2.79) |
| ODOD | 8,897 | 55,154 | 5.0 | 46.9 | 12 | 130 | 2.40 (1.24 - 4.19) | 2.77 (2.32 - 3.29) |
| MDCR | 1,093 | 13,410 | 0.7 | 12.2 | <5 | 44 | < 7.00 (2.27 - 16.3) | 3.60 (2.61 - 4.83) |
| MDCD | 1,866 | 14,860 | 1.0 | 9.6 | 5 | 46 | 4.93 (1.60 - 11.5) | 4.81 (3.52 - 6.42) |
| Open Claims | 176,408 | 823,569 | 118.9 | 783.8 | 186 | 1,639 | 1.56 (1.35 - 1.81) | 2.09 (1.99 - 2.19) |
| OEHR | 11,103 | 77,881 | 3.4 | 44.0 | 9 | 94 | 2.68 (1.23 - 5.09) | 2.14 (1.73 - 2.62) |
| Meta-analysis | 213,524 | 1,040,425 | 137.8 | 935.3 | 227 | 2,028 | 1.65 (1.44 - 1.88) | 2.17 (2.07 - 2.26) |
| Meta-analysis (w/o USOC) | 37,116 | 216,856 | 18.9 | 151.5 | 41 | 389 | 2.17 (1.56 - 2.95) | 2.57 (2.32 - 2.84) |
| **PS-stratification / on-treatment TAR** | | | | | | | | |
| CCAE | 20,479 | 71,257 | 12.2 | 52.4 | 18 | 124 | 1.48 (0.88 - 2.33) | 2.36 (1.97 - 2.82) |
| ODOD | 11,601 | 59,795 | 6.2 | 51.8 | 16 | 155 | 2.60 (1.48 - 4.21) | 2.99 (2.54 - 3.50) |
| MDCR | 1,226 | 17,293 | 0.8 | 15.6 | <5 | 63 | < 6.53 (2.12 - 15.2) | 4.04 (3.11 - 5.17) |
| MDCD | 2,010 | 16,533 | 1.1 | 10.8 | 5 | 59 | 4.63 (1.50 - 10.8) | 5.45 (4.15 - 7.03) |
| Open Claims | 228,988 | 840,593 | 142.8 | 800.1 | 217 | 1,714 | 1.52 (1.32 - 1.74) | 2.14 (2.04 - 2.25) |
| OEHR | 13,580 | 82,939 | 4.0 | 47.2 | 10 | 105 | 2.50 (1.20 - 4.59) | 2.23 (1.82 - 2.69) |
| Meta-analysis | 274,648 | 1,054,584 | 165.1 | 951.5 | 261 | 2,098 | 1.58 (1.39 - 1.78) | 2.20 (2.11 - 2.30) |
| Meta-analysis (w/o USOC) | 45,660 | 213,991 | 22.4 | 151.4 | 44 | 384 | 1.97 (1.43 - 2.64) | 2.54 (2.29 - 2.80) |
| **PS-matching / on-treatment TAR without escalation** | | | | | | | | |
| CCAE | 15,250 | 68,961 | 8.5 | 43.8 | 12 | 91 | 1.41 (0.73 - 2.46) | 2.08 (1.67 - 2.55) |
| ODOD | 8,897 | 55,154 | 4.5 | 41.6 | 11 | 107 | 2.43 (1.21 - 4.35) | 2.57 (2.11 - 3.11) |
| MDCR | 1,093 | 13,410 | 0.6 | 11.2 | <5 | 37 | < 7.70 (2.50 - 18.0) | 3.32 (2.33 - 4.57) |
| MDCD | 1,866 | 14,860 | 0.9 | 7.9 | <5 | 40 | < 5.70 (1.85 - 13.3) | 5.05 (3.61 - 6.87) |
| Open Claims | 176,408 | 823,569 | 106.1 | 659.0 | 164 | 1,337 | 1.55 (1.32 - 1.80) | 2.03 (1.92 - 2.14) |
| OEHR | 11,103 | 77,881 | 2.9 | 37.2 | 8 | 79 | 2.77 (1.20 - 5.46) | 2.12 (1.68 - 2.64) |
| Meta-analysis | 213,524 | 1,040,425 | 122.9 | 789.6 | <200 | 1,654 | < 1.63 (1.41 - 1.87) | 2.09 (1.99 - 2.20) |
| Meta-analysis (w/o USOC) | 37,116 | 216,856 | 16.8 | 130.6 | <36 | 317 | < 2.14 (1.50 - 2.96) | 2.43 (2.17 - 2.71) |
| **PS-stratification / on-treatment TAR without escalation** | | | | | | | | |
| CCAE | 20,479 | 71,257 | 11.0 | 45.1 | 15 | 96 | 1.36 (0.76 - 2.24) | 2.13 (1.73 - 2.60) |
| ODOD | 11,601 | 59,795 | 5.6 | 46.1 | 14 | 131 | 2.50 (1.37 - 4.19) | 2.84 (2.38 - 3.37) |
| MDCR | 1,226 | 17,293 | 0.7 | 14.2 | <5 | 54 | < 7.14 (2.32 - 16.7) | 3.80 (2.86 - 4.96) |
| MDCD | 2,010 | 16,533 | 0.9 | 9.0 | <5 | 50 | < 5.34 (1.73 - 12.5) | 5.56 (4.13 - 7.33) |
| Open Claims | 228,988 | 840,593 | 128.8 | 673.2 | 193 | 1,402 | 1.50 (1.29 - 1.73) | 2.08 (1.97 - 2.19) |
| OEHR | 13,580 | 82,939 | 3.5 | 40.0 | 9 | 88 | 2.58 (1.18 - 4.90) | 2.20 (1.76 - 2.71) |
| Meta-analysis | 274,648 | 1,054,584 | 148.9 | 804.4 | 231 | 1,717 | 1.55 (1.36 - 1.76) | 2.13 (2.03 - 2.24) |
| Meta-analysis (w/o USOC) | 45,660 | 213,991 | 20.1 | 131.2 | 38 | 315 | 1.89 (1.34 - 2.59) | 2.40 (2.14 - 2.68) |

##### 4.5.13 Bone fracture

**Supplementary Table 402:** Exposures, outcomes and risk estimates for Bone fracture in GLP1RA (female) (T) vs SU (female) (C) new-user cohorts under four different propensity score (PS) adjustment / time-at-risk (TAR) designs across data sources. We report total patient exposure time in 1,000 patient-year (KYr) units, uncalibrated and calibrated hazard ratios (HRs) and their 95% confidence intervals (CIs). Greyed out entries did not pass study diagnostics and are included only for completeness.

| | Patients | | Exposure (KYr) | | Outcomes | | Incidence (per KYr) | |
|---|---|---|---|---|---|---|---|---|
| | T | C | T | C | T | C | T | C |
| **PS-matching / on-treatment TAR** | | | | | | | | |
| CCAE | 13,821 | 63,626 | 8.4 | 46.3 | 169 | 968 | 20.1 (17.1 - 23.3) | 20.9 (19.6 - 22.3) |
| ODOD | 8,076 | 50,040 | 4.4 | 41.2 | 133 | 1,432 | 30.1 (25.2 - 35.7) | 34.8 (33.0 - 36.6) |
| MDCR | 941 | 11,539 | 0.6 | 10.2 | 22 | 479 | 36.1 (22.6 - 54.7) | 47.0 (42.9 - 51.4) |
| MDCD | 1,622 | 13,355 | 0.9 | 8.4 | 30 | 296 | 34.6 (23.4 - 49.5) | 35.4 (31.5 - 39.7) |
| Open Claims | 151,263 | 722,142 | 100.8 | 673.0 | 1,461 | 14,383 | 14.5 (13.8 - 15.3) | 21.4 (21.0 - 21.7) |
| OEHR | 10,253 | 72,315 | 3.1 | 39.8 | 42 | 982 | 13.6 (9.80 - 18.4) | 24.7 (23.2 - 26.3) |
| Meta-analysis | 185,976 | 933,017 | 118.2 | 818.8 | 1,857 | 18,540 | 15.7 (15.0 - 16.4) | 22.6 (22.3 - 23.0) |
| Meta-analysis (w/o USOC) | 34,713 | 210,875 | 17.4 | 145.8 | 396 | 4,157 | 22.8 (20.6 - 25.1) | 28.5 (27.6 - 29.4) |
| **PS-stratification / on-treatment TAR** | | | | | | | | |
| CCAE | 18,418 | 65,775 | 10.7 | 47.5 | 219 | 987 | 20.4 (17.8 - 23.3) | 20.8 (19.5 - 22.1) |
| ODOD | 10,453 | 53,994 | 5.4 | 45.3 | 161 | 1,660 | 29.6 (25.2 - 34.6) | 36.6 (34.9 - 38.4) |
| MDCR | 1,049 | 14,758 | 0.6 | 12.9 | 23 | 626 | 35.4 (22.5 - 53.1) | 48.6 (44.9 - 52.6) |
| MDCD | 1,744 | 14,781 | 0.9 | 9.4 | 33 | 331 | 36.0 (24.8 - 50.6) | 35.2 (31.5 - 39.2) |
| Open Claims | 194,843 | 737,097 | 120.3 | 686.7 | 1,758 | 15,021 | 14.6 (13.9 - 15.3) | 21.9 (21.5 - 22.2) |
| OEHR | 12,510 | 76,889 | 3.7 | 42.6 | 48 | 1,112 | 13.1 (9.67 - 17.4) | 26.1 (24.6 - 27.7) |
| Meta-analysis | 236,224 | 933,755 | 140.1 | 822.2 | 2,186 | 18,780 | 15.6 (15.0 - 16.3) | 22.8 (22.5 - 23.2) |
| Meta-analysis (w/o USOC) | 41,381 | 196,658 | 19.8 | 135.4 | 428 | 3,759 | 21.6 (19.6 - 23.7) | 27.8 (26.9 - 28.7) |
| **PS-matching / on-treatment TAR without escalation** | | | | | | | | |
| CCAE | 13,821 | 63,626 | 7.6 | 39.8 | 150 | 815 | 19.8 (16.8 - 23.2) | 20.5 (19.1 - 21.9) |
| ODOD | 8,076 | 50,040 | 4.0 | 36.7 | 114 | 1,268 | 28.5 (23.5 - 34.3) | 34.6 (32.7 - 36.5) |
| MDCR | 941 | 11,539 | 0.6 | 9.3 | 21 | 442 | 37.9 (23.5 - 58.0) | 47.3 (43.0 - 51.9) |
| MDCD | 1,622 | 13,355 | 0.7 | 7.0 | 27 | 247 | 36.1 (23.8 - 52.5) | 35.4 (31.1 - 40.1) |
| Open Claims | 151,263 | 722,142 | 90.0 | 566.6 | 1,266 | 12,096 | 14.1 (13.3 - 14.9) | 21.3 (21.0 - 21.7) |
| OEHR | 10,253 | 72,315 | 2.6 | 33.9 | 37 | 814 | 14.0 (9.84 - 19.3) | 24.0 (22.4 - 25.7) |
| Meta-analysis | 185,976 | 933,017 | 105.5 | 693.4 | 1,615 | 15,682 | 15.3 (14.6 - 16.1) | 22.6 (22.3 - 23.0) |
| Meta-analysis (w/o USOC) | 34,713 | 210,875 | 15.5 | 126.7 | 349 | 3,586 | 22.5 (20.2 - 25.0) | 28.3 (27.4 - 29.2) |
| **PS-stratification / on-treatment TAR without escalation** | | | | | | | | |
| CCAE | 18,418 | 65,775 | 9.7 | 41.0 | 196 | 832 | 20.1 (17.4 - 23.1) | 20.3 (18.9 - 21.7) |
| ODOD | 10,453 | 53,994 | 5.0 | 40.5 | 140 | 1,479 | 28.3 (23.8 - 33.3) | 36.6 (34.7 - 38.5) |
| MDCR | 1,049 | 14,758 | 0.6 | 11.8 | 22 | 574 | 37.0 (23.2 - 56.0) | 48.7 (44.8 - 52.8) |
| MDCD | 1,744 | 14,781 | 0.8 | 7.9 | 30 | 277 | 37.7 (25.5 - 53.9) | 35.3 (31.3 - 39.7) |
| Open Claims | 194,843 | 737,097 | 108.6 | 578.6 | 1,548 | 12,654 | 14.3 (13.6 - 15.0) | 21.9 (21.5 - 22.3) |
| OEHR | 12,510 | 76,889 | 3.2 | 36.3 | 43 | 927 | 13.5 (9.79 - 18.2) | 25.5 (23.9 - 27.2) |
| Meta-analysis | 236,224 | 933,755 | 126.5 | 696.4 | 1,927 | 15,892 | 15.2 (14.6 - 15.9) | 22.8 (22.5 - 23.2) |
| Meta-analysis (w/o USOC) | 41,381 | 196,658 | 17.9 | 117.8 | 379 | 3,238 | 21.2 (19.1 - 23.5) | 27.5 (26.6 - 28.5) |

##### 4.5.14 Diabetic ketoacidosis

**Supplementary Table 403:** Exposures, outcomes and risk estimates for Diabetic ketoacidosis in GLP1RA (female) (T) vs SU (female) (C) new-user cohorts under four different propensity score (PS) adjustment / time-at-risk (TAR) designs across data sources. We report total patient exposure time in 1,000 patient-year (KYr) units, uncalibrated and calibrated hazard ratios (HRs) and their 95% confidence intervals (CIs). Greyed out entries did not pass study diagnostics and are included only for completeness.

| | Patients | | Exposure (KYr) | | Outcomes | | Incidence (per KYr) | |
|---|---|---|---|---|---|---|---|---|
| | T | C | T | C | T | C | T | C |
| **PS-matching / on-treatment TAR** | | | | | | | | |
| CCAE | 15,323 | 69,477 | 9.5 | 51.5 | 5 | 59 | 0.52 (0.17 - 1.22) | 1.15 (0.87 - 1.48) |
| ODOD | 8,942 | 55,476 | 5.0 | 47.2 | 6 | 83 | 1.19 (0.44 - 2.60) | 1.76 (1.40 - 2.18) |
| MDCR | 1,098 | 13,499 | 0.7 | 12.4 | 0 | 11 | | 0.89 (0.44 - 1.59) |
| MDCD | 1,881 | 15,037 | 1.0 | 9.7 | <5 | 35 | < 4.91 (1.59 - 11.5) | 3.62 (2.52 - 5.03) |
| Open Claims | 178,113 | 833,753 | 119.9 | 794.5 | 67 | 709 | 0.56 (0.43 - 0.71) | 0.89 (0.83 - 0.96) |
| OEHR | 11,136 | 78,175 | 3.4 | 44.2 | 5 | 60 | 1.48 (0.48 - 3.46) | 1.36 (1.04 - 1.75) |
| Meta-analysis | 213,514 | 1,036,881 | 137.9 | 937.4 | 83 | 911 | 0.60 (0.48 - 0.75) | 0.97 (0.91 - 1.04) |
| Meta-analysis (w/o USOC) | 35,401 | 203,128 | 17.9 | 142.9 | 16 | 202 | 0.89 (0.51 - 1.45) | 1.41 (1.23 - 1.62) |
| **PS-stratification / on-treatment TAR** | | | | | | | | |
| CCAE | 20,575 | 71,808 | 12.2 | 52.9 | 7 | 64 | 0.57 (0.23 - 1.18) | 1.21 (0.93 - 1.55) |
| ODOD | 11,653 | 60,263 | 6.2 | 52.3 | 7 | 98 | 1.13 (0.45 - 2.33) | 1.87 (1.52 - 2.28) |
| MDCR | 1,231 | 17,473 | 0.8 | 15.8 | 0 | 19 | | 1.20 (0.72 - 1.88) |
| MDCD | 2,026 | 16,782 | 1.1 | 11.0 | <5 | 39 | < 4.61 (1.50 - 10.8) | 3.55 (2.52 - 4.85) |
| Open Claims | 230,974 | 850,481 | 143.9 | 810.3 | 78 | 732 | 0.54 (0.43 - 0.68) | 0.90 (0.84 - 0.97) |
| OEHR | 13,630 | 83,305 | 4.0 | 47.5 | 8 | 68 | 1.99 (0.86 - 3.92) | 1.43 (1.11 - 1.82) |
| Meta-analysis | 276,832 | 1,065,857 | 166.4 | 962.9 | 100 | 962 | 0.60 (0.49 - 0.73) | 1.00 (0.94 - 1.06) |
| Meta-analysis (w/o USOC) | 45,858 | 215,376 | 22.5 | 152.6 | 22 | 230 | 0.98 (0.61 - 1.48) | 1.51 (1.32 - 1.71) |
| **PS-matching / on-treatment TAR without escalation** | | | | | | | | |
| CCAE | 15,323 | 69,477 | 8.6 | 44.1 | <5 | 45 | < 0.58 (0.19 - 1.36) | 1.02 (0.74 - 1.36) |
| ODOD | 8,942 | 55,476 | 4.5 | 41.9 | 5 | 64 | 1.10 (0.36 - 2.57) | 1.53 (1.18 - 1.95) |
| MDCR | 1,098 | 13,499 | 0.7 | 11.3 | 0 | 9 | | 0.80 (0.37 - 1.52) |
| MDCD | 1,881 | 15,037 | 0.9 | 8.0 | <5 | 23 | < 5.70 (1.85 - 13.3) | 2.87 (1.82 - 4.30) |
| Open Claims | 178,113 | 833,753 | 107.0 | 667.9 | 59 | 525 | 0.55 (0.42 - 0.71) | 0.79 (0.72 - 0.86) |
| OEHR | 11,136 | 78,175 | 2.9 | 37.4 | <5 | 38 | < 1.72 (0.56 - 4.02) | 1.02 (0.72 - 1.39) |
| Meta-analysis | 213,514 | 1,036,881 | 123.0 | 791.3 | <74 | 672 | < 0.60 (0.47 - 0.76) | 0.85 (0.79 - 0.92) |
| Meta-analysis (w/o USOC) | 35,401 | 203,128 | 16.0 | 123.5 | <15 | 147 | < 0.94 (0.52 - 1.55) | 1.19 (1.01 - 1.40) |
| **PS-stratification / on-treatment TAR without escalation** | | | | | | | | |
| CCAE | 20,575 | 71,808 | 11.1 | 45.4 | 5 | 50 | 0.45 (0.15 - 1.05) | 1.10 (0.82 - 1.45) |
| ODOD | 11,653 | 60,263 | 5.6 | 46.5 | 6 | 78 | 1.07 (0.39 - 2.32) | 1.68 (1.33 - 2.09) |
| MDCR | 1,231 | 17,473 | 0.7 | 14.4 | 0 | 16 | | 1.11 (0.64 - 1.81) |
| MDCD | 2,026 | 16,782 | 0.9 | 9.1 | <5 | 26 | < 5.34 (1.73 - 12.5) | 2.85 (1.86 - 4.17) |
| Open Claims | 230,974 | 850,481 | 129.9 | 681.6 | 67 | 542 | 0.52 (0.40 - 0.66) | 0.80 (0.73 - 0.86) |
| OEHR | 13,630 | 83,305 | 3.5 | 40.2 | <5 | 44 | < 1.43 (0.46 - 3.33) | 1.09 (0.79 - 1.47) |
| Meta-analysis | 276,832 | 1,065,857 | 150.1 | 813.8 | <83 | 714 | < 0.55 (0.44 - 0.69) | 0.88 (0.81 - 0.94) |
| Meta-analysis (w/o USOC) | 45,858 | 215,376 | 20.2 | 132.2 | <16 | 172 | < 0.79 (0.45 - 1.29) | 1.30 (1.11 - 1.51) |

##### 4.5.15 Diarrhea

**Supplementary Table 404:** Exposures, outcomes and risk estimates for Diarrhea in GLP1RA (female) (T) vs SU (female) (C) new-user cohorts under four different propensity score (PS) adjustment / time-at-risk (TAR) designs across data sources. We report total patient exposure time in 1,000 patient-year (KYr) units, uncalibrated and calibrated hazard ratios (HRs) and their 95% confidence intervals (CIs). Greyed out entries did not pass study diagnostics and are included only for completeness.

| | Patients | | Exposure (KYr) | | Outcomes | | Incidence (per KYr) | |
|---|---|---|---|---|---|---|---|---|
| | T | C | T | C | T | C | T | C |
| **PS-matching / on-treatment TAR** | | | | | | | | |
| CCAE | 12,969 | 59,928 | 7.8 | 43.4 | 354 | 1,537 | 45.7 (41.0 - 50.7) | 35.4 (33.7 - 37.2) |
| ODOD | 7,324 | 45,848 | 4.0 | 37.2 | 210 | 1,882 | 52.5 (45.6 - 60.1) | 50.6 (48.3 - 52.9) |
| MDCR | 949 | 11,402 | 0.6 | 10.1 | 27 | 440 | 45.0 (29.6 - 65.4) | 43.7 (39.7 - 47.9) |
| MDCD | 1,430 | 11,794 | 0.7 | 7.2 | 65 | 533 | 87.1 (67.2 - 111) | 73.9 (67.8 - 80.5) |
| Open Claims | 145,604 | 703,313 | 96.9 | 655.7 | 2,408 | 15,718 | 24.9 (23.9 - 25.9) | 24.0 (23.6 - 24.4) |
| OEHR | 9,403 | 67,296 | 2.8 | 35.7 | 162 | 1,892 | 58.4 (49.8 - 68.1) | 53.0 (50.7 - 55.5) |
| Meta-analysis | 177,679 | 899,581 | 112.8 | 789.2 | 3,226 | 22,002 | 28.6 (27.6 - 29.6) | 27.9 (27.5 - 28.2) |
| Meta-analysis (w/o USOC) | 32,075 | 196,268 | 15.9 | 133.6 | 818 | 6,284 | 51.5 (48.1 - 55.2) | 47.0 (45.9 - 48.2) |
| **PS-stratification / on-treatment TAR** | | | | | | | | |
| CCAE | 17,309 | 62,314 | 10.0 | 44.8 | 474 | 1,588 | 47.6 (43.4 - 52.1) | 35.5 (33.7 - 37.2) |
| ODOD | 9,492 | 49,358 | 4.9 | 40.7 | 264 | 2,104 | 54.0 (47.7 - 60.9) | 51.7 (49.5 - 54.0) |
| MDCR | 1,049 | 14,631 | 0.6 | 12.8 | 29 | 580 | 45.7 (30.6 - 65.6) | 45.2 (41.6 - 49.1) |
| MDCD | 1,534 | 13,053 | 0.8 | 8.1 | 73 | 591 | 92.3 (72.3 - 116) | 73.2 (67.4 - 79.4) |
| Open Claims | 186,885 | 719,371 | 115.2 | 670.5 | 3,059 | 16,351 | 26.6 (25.6 - 27.5) | 24.4 (24.0 - 24.8) |
| OEHR | 11,446 | 71,651 | 3.3 | 38.2 | 199 | 2,051 | 60.8 (52.6 - 69.9) | 53.6 (51.3 - 56.0) |
| Meta-analysis | 225,132 | 902,694 | 133.3 | 794.2 | 3,996 | 22,094 | 30.0 (29.1 - 30.9) | 27.8 (27.5 - 28.2) |
| Meta-analysis (w/o USOC) | 38,247 | 183,323 | 18.1 | 123.7 | 937 | 5,743 | 51.7 (48.5 - 55.1) | 46.4 (45.2 - 47.6) |
| **PS-matching / on-treatment TAR without escalation** | | | | | | | | |
| CCAE | 12,969 | 59,928 | 7.0 | 37.5 | 318 | 1,344 | 45.4 (40.6 - 50.7) | 35.8 (33.9 - 37.8) |
| ODOD | 7,324 | 45,848 | 3.6 | 33.4 | 199 | 1,698 | 54.9 (47.5 - 63.0) | 50.8 (48.4 - 53.3) |
| MDCR | 949 | 11,402 | 0.6 | 9.3 | 23 | 394 | 41.8 (26.5 - 62.7) | 42.5 (38.4 - 47.0) |
| MDCD | 1,430 | 11,794 | 0.7 | 6.0 | 56 | 449 | 84.8 (64.1 - 110) | 74.2 (67.5 - 81.4) |
| Open Claims | 145,604 | 703,313 | 86.7 | 555.3 | 2,143 | 13,288 | 24.7 (23.7 - 25.8) | 23.9 (23.5 - 24.3) |
| OEHR | 9,403 | 67,296 | 2.4 | 30.8 | 145 | 1,518 | 60.3 (50.9 - 71.0) | 49.3 (46.8 - 51.8) |
| Meta-analysis | 177,679 | 899,581 | 100.9 | 672.3 | 2,884 | 18,691 | 28.6 (27.5 - 29.6) | 27.8 (27.4 - 28.2) |
| Meta-analysis (w/o USOC) | 32,075 | 196,268 | 14.2 | 117.0 | 741 | 5,403 | 52.0 (48.3 - 55.9) | 46.2 (44.9 - 47.4) |
| **PS-stratification / on-treatment TAR without escalation** | | | | | | | | |
| CCAE | 17,309 | 62,314 | 9.1 | 38.8 | 426 | 1,391 | 46.9 (42.6 - 51.6) | 35.9 (34.0 - 37.8) |
| ODOD | 9,492 | 49,358 | 4.5 | 36.6 | 249 | 1,909 | 55.8 (49.1 - 63.2) | 52.2 (49.9 - 54.6) |
| MDCR | 1,049 | 14,631 | 0.6 | 11.8 | 25 | 520 | 42.8 (27.7 - 63.1) | 44.3 (40.5 - 48.2) |
| MDCD | 1,534 | 13,053 | 0.7 | 6.8 | 64 | 500 | 91.2 (70.2 - 116) | 73.6 (67.3 - 80.3) |
| Open Claims | 186,885 | 719,371 | 104.1 | 568.4 | 2,767 | 13,842 | 26.6 (25.6 - 27.6) | 24.4 (23.9 - 24.8) |
| OEHR | 11,446 | 71,651 | 2.9 | 33.1 | 180 | 1,645 | 62.7 (53.9 - 72.6) | 49.7 (47.3 - 52.2) |
| Meta-analysis | 225,132 | 902,694 | 120.6 | 676.9 | 3,622 | 18,787 | 30.0 (29.1 - 31.0) | 27.8 (27.4 - 28.2) |
| Meta-analysis (w/o USOC) | 38,247 | 183,323 | 16.4 | 108.5 | 855 | 4,945 | 52.1 (48.7 - 55.7) | 45.6 (44.3 - 46.9) |

##### 4.5.16 Genitourinary infection

**Supplementary Table 405:** Exposures, outcomes and risk estimates for Genitourinary infection in GLP1RA (female) (T) vs SU (female) (C) new-user cohorts under four different propensity score (PS) adjustment / time-at-risk (TAR) designs across data sources. We report total patient exposure time in 1,000 patient-year (KYr) units, uncalibrated and calibrated hazard ratios (HRs) and their 95% confidence intervals (CIs). Greyed out entries did not pass study diagnostics and are included only for completeness.

| | Patients | | Exposure (KYr) | | Outcomes | | Incidence (per KYr) | |
|---|---|---|---|---|---|---|---|---|
| | T | C | T | C | T | C | T | C |
| **PS-matching / on-treatment TAR** | | | | | | | | |
| CCAE | 10,631 | 49,762 | 6.2 | 35.3 | 451 | 2,355 | 72.4 (65.8 - 79.4) | 66.7 (64.0 - 69.4) |
| ODOD | 6,206 | 37,882 | 3.3 | 29.5 | 302 | 2,889 | 91.8 (81.7 - 103) | 97.8 (94.3 - 101) |
| MDCR | 773 | 9,213 | 0.5 | 8.0 | 39 | 711 | 84.5 (60.1 - 115) | 89.2 (82.8 - 96.0) |
| MDCD | 1,206 | 9,894 | 0.6 | 6.0 | 70 | 615 | 109 (84.7 - 137) | 103 (95.1 - 112) |
| Open Claims | 113,858 | 564,876 | 75.7 | 522.7 | 3,094 | 25,854 | 40.9 (39.4 - 42.3) | 49.5 (48.9 - 50.1) |
| OEHR | 8,488 | 60,875 | 2.4 | 31.8 | 208 | 2,409 | 85.0 (73.8 - 97.4) | 75.8 (72.8 - 78.8) |
| Meta-analysis | 141,162 | 732,502 | 88.8 | 633.3 | 4,164 | 34,833 | 46.9 (45.5 - 48.3) | 55.0 (54.4 - 55.6) |
| Meta-analysis (w/o USOC) | 27,304 | 167,626 | 13.1 | 110.6 | 1,070 | 8,979 | 81.8 (77.0 - 86.9) | 81.2 (79.5 - 82.9) |
| **PS-stratification / on-treatment TAR** | | | | | | | | |
| CCAE | 14,042 | 51,428 | 7.9 | 36.3 | 598 | 2,428 | 76.1 (70.1 - 82.4) | 66.9 (64.3 - 69.6) |
| ODOD | 7,975 | 40,393 | 4.0 | 32.0 | 361 | 3,269 | 89.8 (80.7 - 99.5) | 102 (98.8 - 106) |
| MDCR | 849 | 12,248 | 0.5 | 10.5 | 41 | 1,003 | 84.5 (60.7 - 115) | 95.9 (90.1 - 102) |
| MDCD | 1,310 | 10,858 | 0.7 | 6.6 | 78 | 707 | 113 (89.1 - 141) | 107 (99.0 - 115) |
| Open Claims | 144,467 | 578,203 | 89.2 | 534.8 | 3,719 | 27,149 | 41.7 (40.4 - 43.0) | 50.8 (50.2 - 51.4) |
| OEHR | 10,291 | 64,267 | 2.9 | 33.7 | 243 | 2,686 | 84.3 (74.1 - 95.6) | 79.7 (76.7 - 82.8) |
| Meta-analysis | 176,775 | 734,291 | 104.0 | 636.8 | 4,921 | 35,532 | 47.3 (46.0 - 48.7) | 55.8 (55.2 - 56.4) |
| Meta-analysis (w/o USOC) | 32,308 | 156,088 | 14.8 | 102.0 | 1,202 | 8,383 | 81.4 (76.9 - 86.2) | 82.2 (80.5 - 84.0) |
| **PS-matching / on-treatment TAR without escalation** | | | | | | | | |
| CCAE | 10,631 | 49,762 | 5.6 | 30.7 | 405 | 2,027 | 71.8 (65.0 - 79.1) | 66.1 (63.3 - 69.1) |
| ODOD | 6,206 | 37,882 | 3.0 | 26.6 | 269 | 2,603 | 89.5 (79.1 - 101) | 97.8 (94.1 - 102) |
| MDCR | 773 | 9,213 | 0.4 | 7.3 | 30 | 638 | 69.8 (47.1 - 99.7) | 87.1 (80.5 - 94.1) |
| MDCD | 1,206 | 9,894 | 0.6 | 5.1 | 55 | 512 | 98.0 (73.8 - 128) | 101 (92.4 - 110) |
| Open Claims | 113,858 | 564,876 | 67.7 | 443.5 | 2,711 | 22,003 | 40.1 (38.6 - 41.6) | 49.6 (49.0 - 50.3) |
| OEHR | 8,488 | 60,875 | 2.1 | 27.5 | 174 | 1,969 | 81.6 (70.0 - 94.7) | 71.5 (68.4 - 74.7) |
| Meta-analysis | 141,162 | 732,502 | 79.4 | 540.7 | 3,644 | 29,752 | 45.9 (44.4 - 47.4) | 55.0 (54.4 - 55.7) |
| Meta-analysis (w/o USOC) | 27,304 | 167,626 | 11.8 | 97.2 | 933 | 7,749 | 79.3 (74.3 - 84.5) | 79.7 (78.0 - 81.5) |
| **PS-stratification / on-treatment TAR without escalation** | | | | | | | | |
| CCAE | 14,042 | 51,428 | 7.2 | 31.6 | 547 | 2,092 | 76.1 (69.9 - 82.8) | 66.3 (63.5 - 69.2) |
| ODOD | 7,975 | 40,393 | 3.7 | 28.9 | 324 | 2,959 | 87.7 (78.4 - 97.8) | 103 (98.9 - 106) |
| MDCR | 849 | 12,248 | 0.5 | 9.6 | 32 | 901 | 70.6 (48.3 - 99.7) | 93.6 (87.6 - 100.0) |
| MDCD | 1,310 | 10,858 | 0.6 | 5.6 | 63 | 595 | 104 (79.9 - 133) | 105 (97.0 - 114) |
| Open Claims | 144,467 | 578,203 | 80.5 | 454.3 | 3,307 | 23,155 | 41.1 (39.7 - 42.5) | 51.0 (50.3 - 51.6) |
| OEHR | 10,291 | 64,267 | 2.5 | 29.2 | 206 | 2,200 | 81.1 (70.4 - 92.9) | 75.3 (72.2 - 78.5) |
| Meta-analysis | 176,775 | 734,291 | 94.0 | 543.9 | 4,384 | 30,406 | 46.7 (45.3 - 48.1) | 55.9 (55.3 - 56.5) |
| Meta-analysis (w/o USOC) | 32,308 | 156,088 | 13.4 | 89.6 | 1,077 | 7,251 | 80.2 (75.5 - 85.2) | 80.9 (79.1 - 82.8) |

##### 4.5.17 Hyperkalemia

**Supplementary Table 406:** Exposures, outcomes and risk estimates for Hyperkalemia in GLP1RA (female) (T) vs SU (female) (C) new-user cohorts under four different propensity score (PS) adjustment / time-at-risk (TAR) designs across data sources. We report total patient exposure time in 1,000 patient-year (KYr) units, uncalibrated and calibrated hazard ratios (HRs) and their 95% confidence intervals (CIs). Greyed out entries did not pass study diagnostics and are included only for completeness.

| | Patients | | Exposure (KYr) | | Outcomes | | Incidence (per KYr) | |
|---|---|---|---|---|---|---|---|---|
| | T | C | T | C | T | C | T | C |
| **PS-matching / on-treatment TAR** | | | | | | | | |
| CCAE | 15,116 | 68,606 | 9.4 | 50.6 | 41 | 379 | 4.38 (3.14 - 5.94) | 7.49 (6.76 - 8.29) |
| ODOD | 8,696 | 53,210 | 4.9 | 44.3 | 60 | 937 | 12.3 (9.40 - 15.8) | 21.2 (19.8 - 22.6) |
| MDCR | 1,084 | 13,259 | 0.7 | 12.0 | 5 | 155 | 7.13 (2.31 - 16.6) | 12.9 (10.9 - 15.1) |
| MDCD | 1,842 | 14,702 | 1.0 | 9.3 | 8 | 148 | 8.02 (3.46 - 15.8) | 15.9 (13.4 - 18.7) |
| Open Claims | 175,343 | 817,461 | 117.8 | 771.8 | 368 | 5,803 | 3.12 (2.81 - 3.46) | 7.52 (7.33 - 7.71) |
| OEHR | 10,986 | 76,869 | 3.3 | 42.9 | 26 | 530 | 7.83 (5.11 - 11.5) | 12.4 (11.3 - 13.5) |
| Meta-analysis | 213,067 | 1,044,107 | 137.0 | 930.9 | 508 | 7,952 | 3.71 (3.39 - 4.04) | 8.54 (8.36 - 8.73) |
| Meta-analysis (w/o USOC) | 37,724 | 226,646 | 19.3 | 159.1 | 140 | 2,149 | 7.27 (6.12 - 8.58) | 13.5 (12.9 - 14.1) |
| **PS-stratification / on-treatment TAR** | | | | | | | | |
| CCAE | 20,285 | 70,883 | 12.0 | 51.9 | 49 | 390 | 4.08 (3.02 - 5.39) | 7.51 (6.79 - 8.30) |
| ODOD | 11,338 | 57,592 | 6.0 | 48.8 | 67 | 1,125 | 11.2 (8.67 - 14.2) | 23.1 (21.7 - 24.5) |
| MDCR | 1,208 | 16,970 | 0.7 | 15.2 | 6 | 220 | 8.04 (2.95 - 17.5) | 14.5 (12.6 - 16.5) |
| MDCD | 1,985 | 16,325 | 1.1 | 10.5 | 8 | 179 | 7.52 (3.25 - 14.8) | 17.0 (14.6 - 19.7) |
| Open Claims | 227,579 | 833,943 | 141.5 | 787.1 | 427 | 6,214 | 3.02 (2.74 - 3.32) | 7.89 (7.70 - 8.09) |
| OEHR | 13,436 | 81,722 | 4.0 | 45.9 | 30 | 642 | 7.57 (5.11 - 10.8) | 14.0 (12.9 - 15.1) |
| Meta-analysis | 272,638 | 1,044,140 | 163.5 | 933.6 | 573 | 8,371 | 3.50 (3.22 - 3.80) | 8.97 (8.77 - 9.16) |
| Meta-analysis (w/o USOC) | 45,059 | 210,197 | 22.0 | 146.5 | 146 | 2,157 | 6.65 (5.61 - 7.82) | 14.7 (14.1 - 15.4) |
| **PS-matching / on-treatment TAR without escalation** | | | | | | | | |
| CCAE | 15,116 | 68,606 | 8.4 | 43.4 | 36 | 335 | 4.28 (3.00 - 5.93) | 7.73 (6.92 - 8.60) |
| ODOD | 8,696 | 53,210 | 4.4 | 39.4 | 57 | 810 | 12.9 (9.80 - 16.8) | 20.6 (19.2 - 22.0) |
| MDCR | 1,084 | 13,259 | 0.6 | 11.0 | 5 | 140 | 7.84 (2.55 - 18.3) | 12.7 (10.7 - 15.0) |
| MDCD | 1,842 | 14,702 | 0.9 | 7.7 | 8 | 128 | 9.30 (4.01 - 18.3) | 16.5 (13.8 - 19.6) |
| Open Claims | 175,343 | 817,461 | 105.1 | 649.4 | 318 | 4,817 | 3.02 (2.70 - 3.38) | 7.42 (7.21 - 7.63) |
| OEHR | 10,986 | 76,869 | 2.9 | 36.5 | 24 | 389 | 8.40 (5.38 - 12.5) | 10.7 (9.63 - 11.8) |
| Meta-analysis | 213,067 | 1,044,107 | 122.3 | 787.4 | 448 | 6,619 | 3.66 (3.33 - 4.02) | 8.41 (8.21 - 8.61) |
| Meta-analysis (w/o USOC) | 37,724 | 226,646 | 17.2 | 138.0 | 130 | 1,802 | 7.57 (6.33 - 8.99) | 13.1 (12.5 - 13.7) |
| **PS-stratification / on-treatment TAR without escalation** | | | | | | | | |
| CCAE | 20,285 | 70,883 | 10.9 | 44.6 | 44 | 345 | 4.04 (2.94 - 5.42) | 7.74 (6.94 - 8.60) |
| ODOD | 11,338 | 57,592 | 5.4 | 43.5 | 64 | 982 | 11.8 (9.05 - 15.0) | 22.6 (21.2 - 24.0) |
| MDCR | 1,208 | 16,970 | 0.7 | 13.9 | 6 | 192 | 8.80 (3.23 - 19.1) | 13.8 (11.9 - 15.9) |
| MDCD | 1,985 | 16,325 | 0.9 | 8.8 | 8 | 156 | 8.71 (3.76 - 17.2) | 17.8 (15.1 - 20.8) |
| Open Claims | 227,579 | 833,943 | 127.7 | 662.8 | 374 | 5,176 | 2.93 (2.64 - 3.24) | 7.81 (7.60 - 8.02) |
| OEHR | 13,436 | 81,722 | 3.4 | 39.1 | 28 | 474 | 8.12 (5.40 - 11.7) | 12.1 (11.1 - 13.3) |
| Meta-analysis | 272,638 | 1,044,140 | 147.5 | 790.0 | 510 | 6,977 | 3.46 (3.16 - 3.77) | 8.83 (8.63 - 9.04) |
| Meta-analysis (w/o USOC) | 45,059 | 210,197 | 19.8 | 127.2 | 136 | 1,801 | 6.88 (5.77 - 8.13) | 14.2 (13.5 - 14.8) |

##### 4.5.18 Hypoglycemia

**Supplementary Table 407:** Exposures, outcomes and risk estimates for Hypoglycemia in GLP1RA (female) (T) vs SU (female) (C) new-user cohorts under four different propensity score (PS) adjustment / time-at-risk (TAR) designs across data sources. We report total patient exposure time in 1,000 patient-year (KYr) units, uncalibrated and calibrated hazard ratios (HRs) and their 95% confidence intervals (CIs). Greyed out entries did not pass study diagnostics and are included only for completeness.

| | Patients | | Exposure (KYr) | | Outcomes | | Incidence (per KYr) | |
|---|---|---|---|---|---|---|---|---|
| | T | C | T | C | T | C | T | C |
| **PS-matching / on-treatment TAR** | | | | | | | | |
| CCAE | 14,974 | 68,551 | 9.3 | 50.7 | 46 | 535 | 4.94 (3.62 - 6.59) | 10.6 (9.68 - 11.5) |
| ODOD | 8,791 | 54,693 | 4.9 | 46.0 | 37 | 1,105 | 7.48 (5.27 - 10.3) | 24.0 (22.6 - 25.5) |
| MDCR | 1,083 | 13,413 | 0.7 | 12.2 | <5 | 238 | < 7.05 (2.29 - 16.5) | 19.5 (17.1 - 22.2) |
| MDCD | 1,823 | 14,730 | 1.0 | 9.3 | 9 | 288 | 9.05 (4.14 - 17.2) | 30.9 (27.4 - 34.7) |
| Open Claims | 174,559 | 821,394 | 117.4 | 779.2 | 326 | 8,103 | 2.78 (2.48 - 3.10) | 10.4 (10.2 - 10.6) |
| OEHR | 10,983 | 77,456 | 3.3 | 43.3 | 26 | 828 | 7.84 (5.12 - 11.5) | 19.1 (17.9 - 20.5) |
| Meta-analysis | 212,213 | 1,050,237 | 136.7 | 940.7 | <449 | 11,097 | < 3.29 (2.99 - 3.60) | 11.8 (11.6 - 12.0) |
| Meta-analysis (w/o USOC) | 37,654 | 228,843 | 19.3 | 161.5 | <123 | 2,994 | < 6.38 (5.30 - 7.61) | 18.5 (17.9 - 19.2) |
| **PS-stratification / on-treatment TAR** | | | | | | | | |
| CCAE | 19,900 | 70,862 | 11.8 | 52.0 | 60 | 560 | 5.07 (3.87 - 6.52) | 10.8 (9.90 - 11.7) |
| ODOD | 11,352 | 59,401 | 6.0 | 50.9 | 56 | 1,322 | 9.29 (7.02 - 12.1) | 25.9 (24.6 - 27.4) |
| MDCR | 1,212 | 17,322 | 0.8 | 15.5 | <5 | 352 | < 6.60 (2.14 - 15.4) | 22.7 (20.4 - 25.2) |
| MDCD | 1,953 | 16,411 | 1.1 | 10.6 | 9 | 341 | 8.56 (3.92 - 16.3) | 32.2 (28.9 - 35.8) |
| Open Claims | 225,048 | 837,735 | 140.3 | 794.4 | 401 | 8,599 | 2.86 (2.59 - 3.15) | 10.8 (10.6 - 11.1) |
| OEHR | 13,388 | 82,491 | 3.9 | 46.4 | 31 | 934 | 7.86 (5.34 - 11.2) | 20.1 (18.9 - 21.5) |
| Meta-analysis | 269,688 | 1,050,489 | 162.1 | 943.8 | 548 | 11,415 | 3.38 (3.10 - 3.68) | 12.1 (11.9 - 12.3) |
| Meta-analysis (w/o USOC) | 44,640 | 212,754 | 21.8 | 149.4 | 147 | 2,816 | 6.74 (5.70 - 7.92) | 18.9 (18.2 - 19.6) |
| **PS-matching / on-treatment TAR without escalation** | | | | | | | | |
| CCAE | 14,974 | 68,551 | 8.4 | 43.5 | 40 | 444 | 4.79 (3.42 - 6.52) | 10.2 (9.29 - 11.2) |
| ODOD | 8,791 | 54,693 | 4.5 | 40.9 | 28 | 962 | 6.27 (4.16 - 9.06) | 23.5 (22.0 - 25.0) |
| MDCR | 1,083 | 13,413 | 0.6 | 11.1 | <5 | 212 | < 7.77 (2.52 - 18.1) | 19.1 (16.6 - 21.8) |
| MDCD | 1,823 | 14,730 | 0.9 | 7.8 | 7 | 231 | 8.12 (3.26 - 16.7) | 29.7 (26.0 - 33.8) |
| Open Claims | 174,559 | 821,394 | 104.8 | 655.8 | 268 | 6,678 | 2.56 (2.26 - 2.88) | 10.2 (9.94 - 10.4) |
| OEHR | 10,983 | 77,456 | 2.9 | 36.8 | 19 | 610 | 6.65 (4.01 - 10.4) | 16.6 (15.3 - 17.9) |
| Meta-analysis | 212,213 | 1,050,237 | 122.0 | 795.9 | <367 | 9,137 | < 3.01 (2.71 - 3.33) | 11.5 (11.2 - 11.7) |
| Meta-analysis (w/o USOC) | 37,654 | 228,843 | 17.2 | 140.1 | <99 | 2,459 | < 5.76 (4.68 - 7.01) | 17.6 (16.9 - 18.3) |
| **PS-stratification / on-treatment TAR without escalation** | | | | | | | | |
| CCAE | 19,900 | 70,862 | 10.7 | 44.7 | 52 | 468 | 4.85 (3.62 - 6.36) | 10.5 (9.54 - 11.5) |
| ODOD | 11,352 | 59,401 | 5.5 | 45.4 | 46 | 1,161 | 8.40 (6.15 - 11.2) | 25.6 (24.1 - 27.1) |
| MDCR | 1,212 | 17,322 | 0.7 | 14.2 | <5 | 315 | < 7.22 (2.35 - 16.9) | 22.2 (19.8 - 24.8) |
| MDCD | 1,953 | 16,411 | 0.9 | 8.8 | 7 | 276 | 7.68 (3.09 - 15.8) | 31.2 (27.6 - 35.1) |
| Open Claims | 225,048 | 837,735 | 126.6 | 669.1 | 337 | 7,127 | 2.66 (2.39 - 2.96) | 10.7 (10.4 - 10.9) |
| OEHR | 13,388 | 82,491 | 3.4 | 39.5 | 24 | 687 | 6.99 (4.48 - 10.4) | 17.4 (16.1 - 18.7) |
| Meta-analysis | 269,688 | 1,050,489 | 146.2 | 798.8 | 459 | 9,443 | 3.14 (2.86 - 3.44) | 11.8 (11.6 - 12.1) |
| Meta-analysis (w/o USOC) | 44,640 | 212,754 | 19.6 | 129.6 | 122 | 2,316 | 6.21 (5.16 - 7.42) | 17.9 (17.1 - 18.6) |

##### 4.5.19 Hypotension

**Supplementary Table 408:** Exposures, outcomes and risk estimates for Hypotension in GLP1RA (female) (T) vs SU (female) (C) new-user cohorts under four different propensity score (PS) adjustment / time-at-risk (TAR) designs across data sources. We report total patient exposure time in 1,000 patient-year (KYr) units, uncalibrated and calibrated hazard ratios (HRs) and their 95% confidence intervals (CIs). Greyed out entries did not pass study diagnostics and are included only for completeness.

| | Patients | | Exposure (KYr) | | Outcomes | | Incidence (per KYr) | |
|---|---|---|---|---|---|---|---|---|
| | T | C | T | C | T | C | T | C |
| **PS-matching / on-treatment TAR** | | | | | | | | |
| CCAE | 15,039 | 68,260 | 9.3 | 50.5 | 78 | 356 | 8.38 (6.63 - 10.5) | 7.05 (6.34 - 7.82) |
| ODOD | 8,607 | 53,283 | 4.8 | 44.7 | 81 | 915 | 16.8 (13.4 - 20.9) | 20.5 (19.1 - 21.8) |
| MDCR | 1,052 | 12,843 | 0.7 | 11.6 | 10 | 216 | 14.5 (6.94 - 26.6) | 18.6 (16.2 - 21.2) |
| MDCD | 1,780 | 14,216 | 0.9 | 9.0 | 28 | 235 | 29.6 (19.6 - 42.7) | 26.0 (22.8 - 29.6) |
| Open Claims | 172,226 | 805,415 | 115.6 | 762.8 | 761 | 6,533 | 6.58 (6.13 - 7.07) | 8.56 (8.36 - 8.77) |
| OEHR | 10,930 | 76,582 | 3.3 | 42.7 | 56 | 612 | 17.0 (12.9 - 22.1) | 14.3 (13.2 - 15.5) |
| Meta-analysis | 209,634 | 1,030,599 | 134.6 | 921.4 | 1,014 | 8,867 | 7.53 (7.08 - 8.01) | 9.62 (9.42 - 9.83) |
| Meta-analysis (w/o USOC) | 37,408 | 225,184 | 19.0 | 158.6 | 253 | 2,334 | 13.3 (11.7 - 15.0) | 14.7 (14.1 - 15.3) |
| **PS-stratification / on-treatment TAR** | | | | | | | | |
| CCAE | 20,175 | 70,624 | 11.9 | 51.9 | 104 | 375 | 8.72 (7.12 - 10.6) | 7.23 (6.51 - 8.00) |
| ODOD | 11,209 | 57,318 | 5.9 | 49.0 | 103 | 1,103 | 17.4 (14.2 - 21.1) | 22.5 (21.2 - 23.9) |
| MDCR | 1,176 | 16,652 | 0.7 | 14.9 | 10 | 323 | 13.6 (6.50 - 24.9) | 21.7 (19.4 - 24.2) |
| MDCD | 1,922 | 15,691 | 1.0 | 10.1 | 28 | 287 | 27.7 (18.4 - 40.0) | 28.4 (25.2 - 31.8) |
| Open Claims | 223,218 | 822,404 | 138.6 | 778.9 | 939 | 6,995 | 6.77 (6.35 - 7.22) | 8.98 (8.77 - 9.19) |
| OEHR | 13,381 | 81,467 | 3.9 | 45.7 | 60 | 717 | 15.3 (11.7 - 19.7) | 15.7 (14.6 - 16.9) |
| Meta-analysis | 267,983 | 1,031,813 | 160.4 | 925.5 | 1,206 | 9,190 | 7.52 (7.10 - 7.96) | 9.93 (9.73 - 10.1) |
| Meta-analysis (w/o USOC) | 44,765 | 209,409 | 21.8 | 146.6 | 267 | 2,195 | 12.3 (10.8 - 13.8) | 15.0 (14.4 - 15.6) |
| **PS-matching / on-treatment TAR without escalation** | | | | | | | | |
| CCAE | 15,039 | 68,260 | 8.4 | 43.3 | 67 | 293 | 8.02 (6.22 - 10.2) | 6.76 (6.01 - 7.58) |
| ODOD | 8,607 | 53,283 | 4.4 | 39.8 | 71 | 802 | 16.3 (12.7 - 20.6) | 20.2 (18.8 - 21.6) |
| MDCR | 1,052 | 12,843 | 0.6 | 10.6 | 10 | 192 | 15.9 (7.62 - 29.2) | 18.0 (15.6 - 20.8) |
| MDCD | 1,780 | 14,216 | 0.8 | 7.5 | 23 | 178 | 28.1 (17.8 - 42.1) | 23.6 (20.3 - 27.3) |
| Open Claims | 172,226 | 805,415 | 103.2 | 641.6 | 646 | 5,405 | 6.26 (5.79 - 6.76) | 8.42 (8.20 - 8.65) |
| OEHR | 10,930 | 76,582 | 2.8 | 36.3 | 44 | 423 | 15.5 (11.3 - 20.8) | 11.6 (10.6 - 12.8) |
| Meta-analysis | 209,634 | 1,030,599 | 120.2 | 779.2 | 861 | 7,293 | 7.17 (6.69 - 7.66) | 9.36 (9.15 - 9.58) |
| Meta-analysis (w/o USOC) | 37,408 | 225,184 | 17.0 | 137.6 | 215 | 1,888 | 12.7 (11.0 - 14.5) | 13.7 (13.1 - 14.4) |
| **PS-stratification / on-treatment TAR without escalation** | | | | | | | | |
| CCAE | 20,175 | 70,624 | 10.8 | 44.6 | 93 | 311 | 8.60 (6.94 - 10.5) | 6.97 (6.22 - 7.79) |
| ODOD | 11,209 | 57,318 | 5.4 | 43.6 | 93 | 972 | 17.3 (13.9 - 21.1) | 22.3 (20.9 - 23.7) |
| MDCR | 1,176 | 16,652 | 0.7 | 13.6 | 10 | 285 | 14.8 (7.10 - 27.2) | 20.9 (18.6 - 23.5) |
| MDCD | 1,922 | 15,691 | 0.9 | 8.4 | 23 | 226 | 26.2 (16.6 - 39.4) | 26.8 (23.4 - 30.5) |
| Open Claims | 223,218 | 822,404 | 125.1 | 655.8 | 809 | 5,814 | 6.47 (6.03 - 6.93) | 8.87 (8.64 - 9.10) |
| OEHR | 13,381 | 81,467 | 3.4 | 39.0 | 47 | 501 | 13.7 (10.1 - 18.3) | 12.9 (11.8 - 14.0) |
| Meta-analysis | 267,983 | 1,031,813 | 144.7 | 783.0 | 1,042 | 7,598 | 7.20 (6.77 - 7.65) | 9.70 (9.49 - 9.92) |
| Meta-analysis (w/o USOC) | 44,765 | 209,409 | 19.6 | 127.2 | 233 | 1,784 | 11.9 (10.4 - 13.5) | 14.0 (13.4 - 14.7) |

##### 4.5.20 Joint pain

**Supplementary Table 409:** Exposures, outcomes and risk estimates for Joint pain in GLP1RA (female) (T) vs SU (female) (C) new-user cohorts under four different propensity score (PS) adjustment / time-at-risk (TAR) designs across data sources. We report total patient exposure time in 1,000 patient-year (KYr) units, uncalibrated and calibrated hazard ratios (HRs) and their 95% confidence intervals (CIs). Greyed out entries did not pass study diagnostics and are included only for completeness.

| | Patients | | Exposure (KYr) | | Outcomes | | Incidence (per KYr) | |
|---|---|---|---|---|---|---|---|---|
| | T | C | T | C | T | C | T | C |
| **PS-matching / on-treatment TAR** | | | | | | | | |
| CCAE | 14,119 | 65,351 | 8.7 | 48.1 | 126 | 624 | 14.5 (12.1 - 17.3) | 13.0 (12.0 - 14.0) |
| ODOD | 8,139 | 51,604 | 4.5 | 43.4 | 76 | 737 | 16.7 (13.2 - 21.0) | 17.0 (15.8 - 18.2) |
| MDCR | 1,016 | 12,901 | 0.7 | 11.8 | <5 | 102 | < 7.42 (2.41 - 17.3) | 8.68 (7.08 - 10.5) |
| MDCD | 1,652 | 13,655 | 0.9 | 8.6 | 26 | 207 | 29.8 (19.5 - 43.7) | 24.0 (20.8 - 27.5) |
| Open Claims | 161,845 | 777,235 | 108.7 | 738.2 | 878 | 6,030 | 8.08 (7.55 - 8.63) | 8.17 (7.96 - 8.38) |
| OEHR | 10,237 | 73,432 | 3.1 | 40.6 | 50 | 700 | 16.4 (12.2 - 21.6) | 17.2 (16.0 - 18.5) |
| Meta-analysis | 197,008 | 994,178 | 126.5 | 890.8 | <1,161 | 8,400 | < 9.18 (8.66 - 9.72) | 9.43 (9.23 - 9.63) |
| Meta-analysis (w/o USOC) | 35,163 | 216,943 | 17.8 | 152.6 | <283 | 2,370 | < 15.9 (14.1 - 17.9) | 15.5 (14.9 - 16.2) |
| **PS-stratification / on-treatment TAR** | | | | | | | | |
| CCAE | 18,670 | 67,542 | 11.0 | 49.4 | 172 | 634 | 15.7 (13.4 - 18.2) | 12.8 (11.9 - 13.9) |
| ODOD | 10,449 | 56,251 | 5.5 | 48.3 | 109 | 815 | 19.8 (16.3 - 23.9) | 16.9 (15.7 - 18.1) |
| MDCR | 1,137 | 16,702 | 0.7 | 15.0 | 5 | 144 | 6.95 (2.26 - 16.2) | 9.60 (8.09 - 11.3) |
| MDCD | 1,769 | 15,272 | 0.9 | 9.8 | 28 | 230 | 30.1 (20.0 - 43.6) | 23.4 (20.5 - 26.6) |
| Open Claims | 207,745 | 794,699 | 129.4 | 754.5 | 1,119 | 6,218 | 8.65 (8.15 - 9.17) | 8.24 (8.04 - 8.45) |
| OEHR | 12,432 | 78,546 | 3.6 | 43.8 | 66 | 742 | 18.3 (14.2 - 23.3) | 16.9 (15.7 - 18.2) |
| Meta-analysis | 249,296 | 997,038 | 149.4 | 896.1 | 1,466 | 8,409 | 9.81 (9.31 - 10.3) | 9.38 (9.18 - 9.59) |
| Meta-analysis (w/o USOC) | 41,551 | 202,339 | 20.1 | 141.6 | 347 | 2,191 | 17.3 (15.5 - 19.2) | 15.5 (14.8 - 16.1) |
| **PS-matching / on-treatment TAR without escalation** | | | | | | | | |
| CCAE | 14,119 | 65,351 | 7.8 | 41.3 | 111 | 536 | 14.2 (11.7 - 17.2) | 13.0 (11.9 - 14.1) |
| ODOD | 8,139 | 51,604 | 4.1 | 38.6 | 63 | 660 | 15.3 (11.8 - 19.6) | 17.1 (15.8 - 18.4) |
| MDCR | 1,016 | 12,901 | 0.6 | 10.7 | <5 | 92 | < 8.19 (2.66 - 19.1) | 8.57 (6.91 - 10.5) |
| MDCD | 1,652 | 13,655 | 0.8 | 7.2 | 24 | 174 | 31.9 (20.5 - 47.5) | 24.2 (20.7 - 28.1) |
| Open Claims | 161,845 | 777,235 | 96.9 | 621.6 | 776 | 5,043 | 8.00 (7.45 - 8.59) | 8.11 (7.89 - 8.34) |
| OEHR | 10,237 | 73,432 | 2.6 | 34.6 | 41 | 564 | 15.5 (11.1 - 21.1) | 16.3 (15.0 - 17.7) |
| Meta-analysis | 197,008 | 994,178 | 112.8 | 754.2 | <1,020 | 7,069 | < 9.04 (8.49 - 9.61) | 9.37 (9.16 - 9.59) |
| Meta-analysis (w/o USOC) | 35,163 | 216,943 | 15.9 | 132.5 | <244 | 2,026 | < 15.3 (13.5 - 17.4) | 15.3 (14.6 - 16.0) |
| **PS-stratification / on-treatment TAR without escalation** | | | | | | | | |
| CCAE | 18,670 | 67,542 | 10.0 | 42.5 | 150 | 545 | 15.1 (12.8 - 17.7) | 12.8 (11.8 - 13.9) |
| ODOD | 10,449 | 56,251 | 5.0 | 43.1 | 94 | 734 | 18.8 (15.2 - 23.0) | 17.0 (15.8 - 18.3) |
| MDCR | 1,137 | 16,702 | 0.7 | 13.7 | 5 | 130 | 7.62 (2.47 - 17.8) | 9.50 (7.93 - 11.3) |
| MDCD | 1,769 | 15,272 | 0.8 | 8.2 | 26 | 193 | 32.3 (21.1 - 47.4) | 23.5 (20.3 - 27.0) |
| Open Claims | 207,745 | 794,699 | 116.6 | 636.0 | 1,006 | 5,207 | 8.63 (8.11 - 9.18) | 8.19 (7.97 - 8.41) |
| OEHR | 12,432 | 78,546 | 3.2 | 37.4 | 57 | 602 | 18.1 (13.7 - 23.4) | 16.1 (14.8 - 17.4) |
| Meta-analysis | 249,296 | 997,038 | 134.7 | 758.9 | 1,307 | 7,088 | 9.70 (9.19 - 10.2) | 9.34 (9.12 - 9.56) |
| Meta-analysis (w/o USOC) | 41,551 | 202,339 | 18.1 | 123.0 | 301 | 1,881 | 16.6 (14.8 - 18.6) | 15.3 (14.6 - 16.0) |

##### 4.5.21 Lower extremity amputation

**Supplementary Table 410:** Exposures, outcomes and risk estimates for Lower extremity amputation in GLP1RA (female) (T) vs SU (female) (C) new-user cohorts under four different propensity score (PS) adjustment / time-at-risk (TAR) designs across data sources. We report total patient exposure time in 1,000 patient-year (KYr) units, uncalibrated and calibrated hazard ratios (HRs) and their 95% confidence intervals (CIs). Greyed out entries did not pass study diagnostics and are included only for completeness.

| | Patients | | Exposure (KYr) | | Outcomes | | Incidence (per KYr) | |
|---|---|---|---|---|---|---|---|---|
| | T | C | T | C | T | C | T | C |
| **PS-matching / on-treatment TAR** | | | | | | | | |
| CCAE | 15,332 | 69,524 | 9.5 | 51.6 | <5 | 16 | < 0.52 (0.17 - 1.22) | 0.31 (0.18 - 0.50) |
| ODOD | 8,941 | 55,511 | 5.0 | 47.2 | <5 | 27 | < 0.99 (0.32 - 2.32) | 0.57 (0.38 - 0.83) |
| MDCR | 1,097 | 13,489 | 0.7 | 12.3 | 0 | 10 | | 0.81 (0.39 - 1.49) |
| MDCD | 1,884 | 15,079 | 1.0 | 9.7 | <5 | 7 | < 4.89 (1.59 - 11.4) | 0.72 (0.29 - 1.49) |
| Open Claims | 178,234 | 834,411 | 120.0 | 794.8 | 21 | 310 | 0.18 (0.11 - 0.27) | 0.39 (0.35 - 0.44) |
| OEHR | 11,141 | 78,091 | 3.4 | 44.1 | 0 | 24 | | 0.54 (0.35 - 0.81) |
| Meta-analysis | 178,234 | 834,411 | 120.0 | 794.8 | 21 | 310 | 0.18 (0.11 - 0.27) | 0.39 (0.35 - 0.44) |
| **PS-stratification / on-treatment TAR** | | | | | | | | |
| CCAE | 20,584 | 71,863 | 12.2 | 52.9 | <5 | 16 | < 0.41 (0.13 - 0.95) | 0.30 (0.17 - 0.49) |
| ODOD | 11,653 | 60,303 | 6.2 | 52.3 | <5 | 39 | < 0.81 (0.26 - 1.88) | 0.75 (0.53 - 1.02) |
| MDCR | 1,230 | 17,455 | 0.8 | 15.7 | 0 | 12 | | 0.76 (0.39 - 1.33) |
| MDCD | 2,031 | 16,825 | 1.1 | 11.0 | <5 | 8 | < 4.59 (1.49 - 10.7) | 0.73 (0.31 - 1.43) |
| Open Claims | 231,103 | 851,361 | 144.0 | 810.7 | 22 | 331 | 0.15 (0.10 - 0.23) | 0.41 (0.37 - 0.45) |
| OEHR | 13,630 | 83,320 | 4.0 | 47.4 | 0 | 28 | | 0.59 (0.39 - 0.85) |
| Meta-analysis | 242,756 | 911,664 | 150.2 | 863.0 | <27 | 370 | < 0.18 (0.12 - 0.26) | 0.43 (0.39 - 0.47) |
| Meta-analysis (w/o USOC) | 11,653 | 60,303 | 6.2 | 52.3 | <5 | 39 | < 0.81 (0.26 - 1.88) | 0.75 (0.53 - 1.02) |
| **PS-matching / on-treatment TAR without escalation** | | | | | | | | |
| CCAE | 15,332 | 69,524 | 8.6 | 44.2 | <5 | 13 | < 0.58 (0.19 - 1.36) | 0.29 (0.16 - 0.50) |
| ODOD | 8,941 | 55,511 | 4.5 | 41.9 | <5 | 20 | < 1.10 (0.36 - 2.57) | 0.48 (0.29 - 0.74) |
| MDCR | 1,097 | 13,489 | 0.7 | 11.2 | 0 | 6 | | 0.53 (0.20 - 1.16) |
| MDCD | 1,884 | 15,079 | 0.9 | 8.0 | <5 | <5 | < 5.68 (1.84 - 13.2) | < 0.62 (0.20 - 1.45) |
| Open Claims | 178,234 | 834,411 | 107.1 | 667.9 | 15 | 233 | 0.14 (0.08 - 0.23) | 0.35 (0.31 - 0.40) |
| OEHR | 11,141 | 78,091 | 2.9 | 37.4 | 0 | 13 | | 0.35 (0.19 - 0.59) |
| Meta-analysis | 178,234 | 834,411 | 107.1 | 667.9 | 15 | 233 | 0.14 (0.08 - 0.23) | 0.35 (0.31 - 0.40) |
| **PS-stratification / on-treatment TAR without escalation** | | | | | | | | |
| CCAE | 20,584 | 71,863 | 11.1 | 45.4 | <5 | 13 | < 0.45 (0.15 - 1.05) | 0.29 (0.15 - 0.49) |
| ODOD | 11,653 | 60,303 | 5.6 | 46.5 | <5 | 32 | < 0.89 (0.29 - 2.07) | 0.69 (0.47 - 0.97) |
| MDCR | 1,230 | 17,455 | 0.7 | 14.4 | 0 | 8 | | 0.56 (0.24 - 1.10) |
| MDCD | 2,031 | 16,825 | 0.9 | 9.1 | <5 | 5 | < 5.32 (1.73 - 12.4) | 0.55 (0.18 - 1.28) |
| Open Claims | 231,103 | 851,361 | 129.9 | 681.8 | 16 | 252 | 0.12 (0.07 - 0.20) | 0.37 (0.33 - 0.42) |
| OEHR | 13,630 | 83,320 | 3.5 | 40.2 | 0 | 16 | | 0.40 (0.23 - 0.65) |
| Meta-analysis | 242,756 | 911,664 | 135.5 | 728.3 | <21 | 284 | < 0.15 (0.10 - 0.24) | 0.39 (0.35 - 0.44) |
| Meta-analysis (w/o USOC) | 11,653 | 60,303 | 5.6 | 46.5 | <5 | 32 | < 0.89 (0.29 - 2.07) | 0.69 (0.47 - 0.97) |

##### 4.5.22 Nausea

**Supplementary Table 411:** Exposures, outcomes and risk estimates for Nausea in GLP1RA (female) (T) vs SU (female) (C) new-user cohorts under four different propensity score (PS) adjustment / time-at-risk (TAR) designs across data sources. We report total patient exposure time in 1,000 patient-year (KYr) units, uncalibrated and calibrated hazard ratios (HRs) and their 95% confidence intervals (CIs). Greyed out entries did not pass study diagnostics and are included only for completeness.

| | Patients | | Exposure (KYr) | | Outcomes | | Incidence (per KYr) | |
|---|---|---|---|---|---|---|---|---|
| | T | C | T | C | T | C | T | C |
| **PS-matching / on-treatment TAR** | | | | | | | | |
| CCAE | 12,637 | 59,138 | 7.6 | 43.5 | 449 | 1,793 | 59.0 (53.7 - 64.7) | 41.2 (39.3 - 43.2) |
| ODOD | 7,071 | 45,139 | 3.8 | 37.3 | 346 | 2,104 | 89.9 (80.7 - 99.9) | 56.4 (54.0 - 58.8) |
| MDCR | 990 | 11,463 | 0.6 | 10.3 | 36 | 433 | 58.5 (41.0 - 81.0) | 42.2 (38.3 - 46.3) |
| MDCD | 1,197 | 10,014 | 0.6 | 6.3 | 96 | 628 | 153 (124 - 187) | 100 (92.7 - 109) |
| Open Claims | 138,061 | 683,664 | 92.6 | 649.2 | 3,666 | 18,327 | 39.6 (38.3 - 40.9) | 28.2 (27.8 - 28.6) |
| OEHR | 9,169 | 67,499 | 2.7 | 36.3 | 225 | 2,024 | 82.7 (72.2 - 94.2) | 55.8 (53.4 - 58.3) |
| Meta-analysis | 169,125 | 876,917 | 108.0 | 782.8 | 4,818 | 25,309 | 44.6 (43.3 - 45.9) | 32.3 (31.9 - 32.7) |
| Meta-analysis (w/o USOC) | 31,064 | 193,253 | 15.4 | 133.6 | 1,152 | 6,982 | 74.7 (70.4 - 79.1) | 52.3 (51.0 - 53.5) |
| **PS-stratification / on-treatment TAR** | | | | | | | | |
| CCAE | 16,677 | 61,481 | 9.6 | 44.9 | 634 | 1,853 | 66.0 (61.0 - 71.4) | 41.3 (39.4 - 43.2) |
| ODOD | 9,066 | 48,730 | 4.7 | 41.0 | 450 | 2,384 | 96.5 (87.8 - 106) | 58.1 (55.8 - 60.5) |
| MDCR | 1,099 | 14,996 | 0.7 | 13.2 | 37 | 606 | 56.3 (39.6 - 77.6) | 45.8 (42.2 - 49.6) |
| MDCD | 1,275 | 11,261 | 0.7 | 7.2 | 106 | 727 | 161 (132 - 195) | 102 (94.3 - 109) |
| Open Claims | 175,888 | 699,022 | 109.5 | 663.8 | 4,651 | 19,013 | 42.5 (41.2 - 43.7) | 28.6 (28.2 - 29.1) |
| OEHR | 11,092 | 71,754 | 3.2 | 38.8 | 290 | 2,206 | 91.3 (81.1 - 102) | 56.8 (54.5 - 59.2) |
| Meta-analysis | 212,723 | 880,987 | 127.0 | 788.6 | 6,025 | 25,456 | 47.4 (46.3 - 48.7) | 32.3 (31.9 - 32.7) |
| Meta-analysis (w/o USOC) | 36,835 | 181,965 | 17.4 | 124.8 | 1,374 | 6,443 | 78.8 (74.7 - 83.1) | 51.6 (50.4 - 52.9) |
| **PS-matching / on-treatment TAR without escalation** | | | | | | | | |
| CCAE | 12,637 | 59,138 | 6.9 | 37.6 | 410 | 1,506 | 59.6 (54.0 - 65.7) | 40.1 (38.1 - 42.2) |
| ODOD | 7,071 | 45,139 | 3.5 | 33.4 | 322 | 1,866 | 92.4 (82.5 - 103) | 55.9 (53.4 - 58.5) |
| MDCR | 990 | 11,463 | 0.6 | 9.4 | 31 | 386 | 54.5 (37.0 - 77.4) | 41.1 (37.1 - 45.4) |
| MDCD | 1,197 | 10,014 | 0.5 | 5.3 | 88 | 528 | 162 (130 - 199) | 99.9 (91.6 - 109) |
| Open Claims | 138,061 | 683,664 | 82.9 | 550.0 | 3,298 | 15,134 | 39.8 (38.4 - 41.2) | 27.5 (27.1 - 28.0) |
| OEHR | 9,169 | 67,499 | 2.4 | 31.3 | 196 | 1,608 | 83.4 (72.1 - 95.9) | 51.3 (48.9 - 53.9) |
| Meta-analysis | 169,125 | 876,917 | 96.7 | 667.0 | 4,345 | 21,028 | 44.9 (43.6 - 46.3) | 31.5 (31.1 - 32.0) |
| Meta-analysis (w/o USOC) | 31,064 | 193,253 | 13.8 | 117.0 | 1,047 | 5,894 | 75.7 (71.2 - 80.4) | 50.4 (49.1 - 51.7) |
| **PS-stratification / on-treatment TAR without escalation** | | | | | | | | |
| CCAE | 16,677 | 61,481 | 8.7 | 38.9 | 584 | 1,562 | 66.7 (61.4 - 72.4) | 40.2 (38.2 - 42.2) |
| ODOD | 9,066 | 48,730 | 4.3 | 36.8 | 420 | 2,124 | 98.7 (89.5 - 109) | 57.8 (55.3 - 60.3) |
| MDCR | 1,099 | 14,996 | 0.6 | 12.1 | 32 | 539 | 52.4 (35.8 - 73.9) | 44.5 (40.8 - 48.4) |
| MDCD | 1,275 | 11,261 | 0.6 | 6.1 | 97 | 621 | 169 (137 - 206) | 102 (94.5 - 111) |
| Open Claims | 175,888 | 699,022 | 99.0 | 563.0 | 4,235 | 15,739 | 42.8 (41.5 - 44.1) | 28.0 (27.5 - 28.4) |
| OEHR | 11,092 | 71,754 | 2.8 | 33.6 | 253 | 1,758 | 91.2 (80.3 - 103) | 52.3 (49.9 - 54.8) |
| Meta-analysis | 212,723 | 880,987 | 114.8 | 672.2 | 5,492 | 21,183 | 47.8 (46.6 - 49.1) | 31.5 (31.1 - 31.9) |
| Meta-analysis (w/o USOC) | 36,835 | 181,965 | 15.8 | 109.3 | 1,257 | 5,444 | 79.7 (75.3 - 84.2) | 49.8 (48.5 - 51.2) |

##### 4.5.23 Peripheral edema

**Supplementary Table 412:** Exposures, outcomes and risk estimates for Peripheral edema in GLP1RA (female) (T) vs SU (female) (C) new-user cohorts under four different propensity score (PS) adjustment / time-at-risk (TAR) designs across data sources. We report total patient exposure time in 1,000 patient-year (KYr) units, uncalibrated and calibrated hazard ratios (HRs) and their 95% confidence intervals (CIs). Greyed out entries did not pass study diagnostics and are included only for completeness.

| | Patients | | Exposure (KYr) | | Outcomes | | Incidence (per KYr) | |
|---|---|---|---|---|---|---|---|---|
| | T | C | T | C | T | C | T | C |
| **PS-matching / on-treatment TAR** | | | | | | | | |
| CCAE | 11,776 | 55,745 | 7.1 | 39.7 | 350 | 2,266 | 49.2 (44.2 - 54.7) | 57.1 (54.8 - 59.5) |
| ODOD | 6,523 | 41,013 | 3.5 | 32.0 | 208 | 3,009 | 59.0 (51.3 - 67.6) | 94.0 (90.7 - 97.5) |
| MDCR | 816 | 9,763 | 0.5 | 8.4 | 27 | 795 | 52.8 (34.8 - 76.8) | 95.1 (88.6 - 102) |
| MDCD | 1,192 | 9,639 | 0.6 | 5.6 | 57 | 597 | 89.9 (68.1 - 116) | 107 (98.1 - 115) |
| Open Claims | 126,791 | 627,042 | 84.2 | 573.5 | 2,132 | 23,997 | 25.3 (24.3 - 26.4) | 41.8 (41.3 - 42.4) |
| OEHR | 8,807 | 64,056 | 2.6 | 33.2 | 126 | 2,285 | 48.9 (40.7 - 58.2) | 68.8 (66.0 - 71.6) |
| Meta-analysis | 155,905 | 807,258 | 98.6 | 692.4 | 2,900 | 32,949 | 29.4 (28.4 - 30.5) | 47.6 (47.1 - 48.1) |
| Meta-analysis (w/o USOC) | 29,114 | 180,216 | 14.4 | 118.9 | 768 | 8,952 | 53.5 (49.8 - 57.4) | 75.3 (73.8 - 76.9) |
| **PS-stratification / on-treatment TAR** | | | | | | | | |
| CCAE | 15,438 | 57,760 | 8.9 | 40.8 | 445 | 2,347 | 49.9 (45.3 - 54.7) | 57.5 (55.2 - 59.9) |
| ODOD | 8,359 | 44,279 | 4.3 | 35.1 | 261 | 3,509 | 61.0 (53.8 - 68.9) | 100 (96.8 - 103) |
| MDCR | 890 | 12,728 | 0.5 | 10.7 | 33 | 1,107 | 61.6 (42.4 - 86.5) | 103 (97.2 - 109) |
| MDCD | 1,281 | 11,097 | 0.7 | 6.6 | 60 | 726 | 88.9 (67.9 - 114) | 111 (103 - 119) |
| Open Claims | 160,813 | 641,277 | 99.2 | 586.1 | 2,532 | 25,278 | 25.5 (24.5 - 26.5) | 43.1 (42.6 - 43.7) |
| OEHR | 10,621 | 68,077 | 3.0 | 35.5 | 147 | 2,577 | 48.6 (41.1 - 57.1) | 72.6 (69.8 - 75.5) |
| Meta-analysis | 195,231 | 811,393 | 115.5 | 697.5 | 3,385 | 33,711 | 29.3 (28.3 - 30.3) | 48.3 (47.8 - 48.9) |
| Meta-analysis (w/o USOC) | 34,418 | 170,116 | 16.2 | 111.4 | 853 | 8,433 | 52.6 (49.1 - 56.2) | 75.7 (74.1 - 77.3) |
| **PS-matching / on-treatment TAR without escalation** | | | | | | | | |
| CCAE | 11,776 | 55,745 | 6.4 | 34.4 | 312 | 1,949 | 48.7 (43.5 - 54.5) | 56.6 (54.1 - 59.2) |
| ODOD | 6,523 | 41,013 | 3.2 | 28.7 | 189 | 2,667 | 58.8 (50.7 - 67.8) | 92.8 (89.3 - 96.4) |
| MDCR | 816 | 9,763 | 0.5 | 7.7 | 26 | 725 | 56.0 (36.6 - 82.1) | 94.2 (87.5 - 101) |
| MDCD | 1,192 | 9,639 | 0.6 | 4.8 | 46 | 508 | 83.6 (61.2 - 112) | 107 (97.5 - 116) |
| Open Claims | 126,791 | 627,042 | 75.2 | 486.2 | 1,837 | 20,118 | 24.4 (23.3 - 25.6) | 41.4 (40.8 - 42.0) |
| OEHR | 8,807 | 64,056 | 2.2 | 28.8 | 115 | 1,862 | 51.6 (42.6 - 61.9) | 64.6 (61.7 - 67.6) |
| Meta-analysis | 155,905 | 807,258 | 88.0 | 590.6 | 2,525 | 27,829 | 28.7 (27.6 - 29.8) | 47.1 (46.6 - 47.7) |
| Meta-analysis (w/o USOC) | 29,114 | 180,216 | 12.9 | 104.5 | 688 | 7,711 | 53.5 (49.6 - 57.7) | 73.8 (72.2 - 75.5) |
| **PS-stratification / on-treatment TAR without escalation** | | | | | | | | |
| CCAE | 15,438 | 57,760 | 8.1 | 35.5 | 398 | 2,024 | 49.1 (44.4 - 54.2) | 57.0 (54.5 - 59.5) |
| ODOD | 8,359 | 44,279 | 3.9 | 31.6 | 239 | 3,128 | 60.9 (53.4 - 69.1) | 99.1 (95.7 - 103) |
| MDCR | 890 | 12,728 | 0.5 | 9.9 | 32 | 1,016 | 65.5 (44.8 - 92.4) | 103 (96.4 - 109) |
| MDCD | 1,281 | 11,097 | 0.6 | 5.6 | 49 | 621 | 83.6 (61.8 - 111) | 111 (102 - 120) |
| Open Claims | 160,813 | 641,277 | 89.5 | 497.3 | 2,215 | 21,268 | 24.7 (23.7 - 25.8) | 42.8 (42.2 - 43.3) |
| OEHR | 10,621 | 68,077 | 2.6 | 30.8 | 135 | 2,106 | 51.1 (42.8 - 60.5) | 68.4 (65.5 - 71.4) |
| Meta-analysis | 195,231 | 811,393 | 104.2 | 595.2 | 2,987 | 28,526 | 28.7 (27.7 - 29.7) | 47.9 (47.4 - 48.5) |
| Meta-analysis (w/o USOC) | 34,418 | 170,116 | 14.7 | 97.9 | 772 | 7,258 | 52.6 (49.0 - 56.5) | 74.2 (72.5 - 75.9) |

##### 4.5.24 Photosensitivity

**Supplementary Table 413:** Exposures, outcomes and risk estimates for Photosensitivity in GLP1RA (female) (T) vs SU (female) (C) new-user cohorts under four different propensity score (PS) adjustment / time-at-risk (TAR) designs across data sources. We report total patient exposure time in 1,000 patient-year (KYr) units, uncalibrated and calibrated hazard ratios (HRs) and their 95% confidence intervals (CIs). Greyed out entries did not pass study diagnostics and are included only for completeness.

| | Patients | | Exposure (KYr) | | Outcomes | | Incidence (per KYr) | |
|---|---|---|---|---|---|---|---|---|
| | T | C | T | C | T | C | T | C |
| **PS-matching / on-treatment TAR** | | | | | | | | |
| CCAE | 15,194 | 69,000 | 9.5 | 51.0 | 14 | 85 | 1.48 (0.81 - 2.49) | 1.67 (1.33 - 2.06) |
| ODOD | 8,890 | 55,046 | 5.0 | 46.7 | 10 | 71 | 2.00 (0.96 - 3.68) | 1.52 (1.19 - 1.92) |
| MDCR | 1,072 | 13,148 | 0.7 | 12.0 | <5 | 27 | < 7.15 (2.32 - 16.7) | 2.25 (1.49 - 3.28) |
| MDCD | 1,874 | 15,015 | 1.0 | 9.7 | <5 | 10 | < 4.90 (1.59 - 11.4) | 1.04 (0.50 - 1.90) |
| Open Claims | 176,301 | 827,290 | 118.5 | 786.8 | 43 | 373 | 0.36 (0.26 - 0.49) | 0.47 (0.43 - 0.52) |
| OEHR | 11,073 | 77,729 | 3.3 | 43.9 | 9 | 58 | 2.69 (1.23 - 5.10) | 1.32 (1.00 - 1.71) |
| Meta-analysis | 211,458 | 1,029,065 | 136.3 | 928.3 | 76 | 587 | 0.56 (0.44 - 0.70) | 0.63 (0.58 - 0.69) |
| Meta-analysis (w/o USOC) | 35,157 | 201,775 | 17.8 | 141.6 | 33 | 214 | 1.86 (1.28 - 2.61) | 1.51 (1.32 - 1.73) |
| **PS-stratification / on-treatment TAR** | | | | | | | | |
| CCAE | 20,378 | 71,317 | 12.1 | 52.4 | 18 | 87 | 1.49 (0.88 - 2.35) | 1.66 (1.33 - 2.05) |
| ODOD | 11,576 | 59,797 | 6.1 | 51.7 | 11 | 81 | 1.79 (0.89 - 3.21) | 1.57 (1.24 - 1.95) |
| MDCR | 1,204 | 17,247 | 0.8 | 15.5 | <5 | 35 | < 6.66 (2.16 - 15.6) | 2.25 (1.57 - 3.14) |
| MDCD | 2,013 | 16,755 | 1.1 | 11.0 | <5 | 10 | < 4.62 (1.50 - 10.8) | 0.91 (0.44 - 1.68) |
| Open Claims | 228,366 | 844,310 | 142.1 | 802.8 | 52 | 387 | 0.37 (0.27 - 0.48) | 0.48 (0.44 - 0.53) |
| OEHR | 13,528 | 82,778 | 4.0 | 47.1 | 9 | 67 | 2.26 (1.03 - 4.29) | 1.42 (1.10 - 1.81) |
| Meta-analysis | 273,848 | 1,058,202 | 164.3 | 953.9 | 90 | 622 | 0.55 (0.44 - 0.67) | 0.65 (0.60 - 0.71) |
| Meta-analysis (w/o USOC) | 45,482 | 213,892 | 22.2 | 151.1 | 38 | 235 | 1.71 (1.21 - 2.35) | 1.55 (1.36 - 1.77) |
| **PS-matching / on-treatment TAR without escalation** | | | | | | | | |
| CCAE | 15,194 | 69,000 | 8.5 | 43.7 | 12 | 74 | 1.41 (0.73 - 2.47) | 1.69 (1.33 - 2.12) |
| ODOD | 8,890 | 55,046 | 4.5 | 41.4 | 10 | 67 | 2.22 (1.06 - 4.08) | 1.62 (1.25 - 2.05) |
| MDCR | 1,072 | 13,148 | 0.6 | 10.9 | <5 | 24 | < 7.86 (2.55 - 18.3) | 2.20 (1.41 - 3.27) |
| MDCD | 1,874 | 15,015 | 0.9 | 8.0 | <5 | 8 | < 5.69 (1.85 - 13.3) | 1.00 (0.43 - 1.97) |
| Open Claims | 176,301 | 827,290 | 105.8 | 661.2 | 30 | 322 | 0.28 (0.19 - 0.40) | 0.49 (0.44 - 0.54) |
| OEHR | 11,073 | 77,729 | 2.9 | 37.1 | 6 | 50 | 2.09 (0.77 - 4.54) | 1.35 (1.00 - 1.78) |
| Meta-analysis | 211,458 | 1,029,065 | 121.6 | 783.5 | 58 | 513 | 0.48 (0.36 - 0.62) | 0.65 (0.60 - 0.71) |
| Meta-analysis (w/o USOC) | 35,157 | 201,775 | 15.9 | 122.3 | 28 | 191 | 1.76 (1.17 - 2.55) | 1.56 (1.35 - 1.80) |
| **PS-stratification / on-treatment TAR without escalation** | | | | | | | | |
| CCAE | 20,378 | 71,317 | 11.0 | 45.0 | 16 | 76 | 1.46 (0.83 - 2.37) | 1.69 (1.33 - 2.11) |
| ODOD | 11,576 | 59,797 | 5.6 | 46.0 | 11 | 76 | 1.97 (0.98 - 3.53) | 1.65 (1.30 - 2.07) |
| MDCR | 1,204 | 17,247 | 0.7 | 14.2 | <5 | 31 | < 7.28 (2.36 - 17.0) | 2.19 (1.49 - 3.11) |
| MDCD | 2,013 | 16,755 | 0.9 | 9.1 | <5 | 8 | < 5.35 (1.74 - 12.5) | 0.88 (0.38 - 1.73) |
| Open Claims | 228,366 | 844,310 | 128.2 | 675.2 | 39 | 334 | 0.30 (0.22 - 0.42) | 0.49 (0.44 - 0.55) |
| OEHR | 13,528 | 82,778 | 3.5 | 39.9 | 6 | 57 | 1.73 (0.64 - 3.77) | 1.43 (1.08 - 1.85) |
| Meta-analysis | 273,848 | 1,058,202 | 148.2 | 806.1 | 72 | 543 | 0.49 (0.38 - 0.61) | 0.67 (0.62 - 0.73) |
| Meta-analysis (w/o USOC) | 45,482 | 213,892 | 20.0 | 130.9 | 33 | 209 | 1.65 (1.14 - 2.32) | 1.60 (1.39 - 1.83) |

##### 4.5.25 Venous thromboembolism

**Supplementary Table 414:** Exposures, outcomes and risk estimates for Venous thromboembolism in GLP1RA (female) (T) vs SU (female) (C) new-user cohorts under four different propensity score (PS) adjustment / time-at-risk (TAR) designs across data sources. We report total patient exposure time in 1,000 patient-year (KYr) units, uncalibrated and calibrated hazard ratios (HRs) and their 95% confidence intervals (CIs). Greyed out entries did not pass study diagnostics and are included only for completeness.

| | Patients | | Exposure (KYr) | | Outcomes | | Incidence (per KYr) | |
|---|---|---|---|---|---|---|---|---|
| | T | C | T | C | T | C | T | C |
| **PS-matching / on-treatment TAR** | | | | | | | | |
| CCAE | 14,954 | 68,098 | 9.3 | 50.3 | 43 | 322 | 4.64 (3.36 - 6.25) | 6.40 (5.72 - 7.14) |
| ODOD | 8,642 | 53,731 | 4.9 | 45.3 | 43 | 508 | 8.86 (6.41 - 11.9) | 11.2 (10.3 - 12.2) |
| MDCR | 1,060 | 12,877 | 0.7 | 11.7 | 6 | 180 | 8.76 (3.22 - 19.1) | 15.4 (13.2 - 17.8) |
| MDCD | 1,803 | 14,367 | 1.0 | 9.1 | 10 | 106 | 10.3 (4.93 - 18.9) | 11.6 (9.50 - 14.0) |
| Open Claims | 171,467 | 806,584 | 115.4 | 764.8 | 420 | 4,709 | 3.64 (3.30 - 4.01) | 6.16 (5.98 - 6.34) |
| OEHR | 10,853 | 76,373 | 3.3 | 42.7 | 26 | 403 | 7.92 (5.17 - 11.6) | 9.45 (8.55 - 10.4) |
| Meta-analysis | 208,779 | 1,032,030 | 134.4 | 923.9 | 548 | 6,228 | 4.08 (3.74 - 4.43) | 6.74 (6.57 - 6.91) |
| Meta-analysis (w/o USOC) | 37,312 | 225,446 | 19.1 | 159.1 | 128 | 1,519 | 6.72 (5.60 - 7.98) | 9.55 (9.07 - 10.0) |
| **PS-stratification / on-treatment TAR** | | | | | | | | |
| CCAE | 20,049 | 70,354 | 11.8 | 51.6 | 54 | 332 | 4.56 (3.43 - 5.95) | 6.43 (5.76 - 7.16) |
| ODOD | 11,251 | 58,278 | 6.0 | 50.2 | 52 | 622 | 8.71 (6.50 - 11.4) | 12.4 (11.4 - 13.4) |
| MDCR | 1,185 | 16,713 | 0.7 | 15.0 | 7 | 248 | 9.57 (3.85 - 19.7) | 16.5 (14.6 - 18.7) |
| MDCD | 1,938 | 16,115 | 1.0 | 10.5 | 10 | 131 | 9.66 (4.63 - 17.8) | 12.5 (10.5 - 14.9) |
| Open Claims | 221,964 | 823,043 | 138.3 | 780.4 | 508 | 4,999 | 3.67 (3.36 - 4.01) | 6.41 (6.23 - 6.59) |
| OEHR | 13,250 | 81,268 | 3.9 | 45.7 | 30 | 472 | 7.68 (5.18 - 11.0) | 10.3 (9.42 - 11.3) |
| Meta-analysis | 266,514 | 1,032,943 | 160.0 | 927.9 | 644 | 6,425 | 4.03 (3.72 - 4.35) | 6.92 (6.76 - 7.10) |
| Meta-analysis (w/o USOC) | 44,550 | 209,900 | 21.7 | 147.5 | 136 | 1,426 | 6.26 (5.25 - 7.41) | 9.67 (9.17 - 10.2) |
| **PS-matching / on-treatment TAR without escalation** | | | | | | | | |
| CCAE | 14,954 | 68,098 | 8.3 | 43.2 | 38 | 279 | 4.57 (3.24 - 6.28) | 6.47 (5.73 - 7.27) |
| ODOD | 8,642 | 53,731 | 4.4 | 40.2 | 38 | 458 | 8.67 (6.13 - 11.9) | 11.4 (10.4 - 12.5) |
| MDCR | 1,060 | 12,877 | 0.6 | 10.7 | <5 | 162 | < 7.99 (2.59 - 18.6) | 15.2 (12.9 - 17.7) |
| MDCD | 1,803 | 14,367 | 0.8 | 7.6 | <5 | 89 | < 5.93 (1.92 - 13.8) | 11.7 (9.42 - 14.4) |
| Open Claims | 171,467 | 806,584 | 103.0 | 643.1 | 365 | 4,011 | 3.54 (3.19 - 3.93) | 6.24 (6.05 - 6.43) |
| OEHR | 10,853 | 76,373 | 2.8 | 36.2 | 21 | 317 | 7.44 (4.61 - 11.4) | 8.75 (7.81 - 9.77) |
| Meta-analysis | 208,779 | 1,032,030 | 119.9 | 781.0 | <472 | 5,316 | < 3.94 (3.59 - 4.31) | 6.81 (6.63 - 6.99) |
| Meta-analysis (w/o USOC) | 37,312 | 225,446 | 17.0 | 137.8 | <107 | 1,305 | < 6.30 (5.16 - 7.61) | 9.47 (8.96 - 10.00) |
| **PS-stratification / on-treatment TAR without escalation** | | | | | | | | |
| CCAE | 20,049 | 70,354 | 10.7 | 44.4 | 49 | 286 | 4.57 (3.38 - 6.04) | 6.45 (5.72 - 7.24) |
| ODOD | 11,251 | 58,278 | 5.4 | 44.6 | 46 | 564 | 8.48 (6.21 - 11.3) | 12.6 (11.6 - 13.7) |
| MDCR | 1,185 | 16,713 | 0.7 | 13.7 | 5 | 225 | 7.43 (2.41 - 17.3) | 16.4 (14.4 - 18.7) |
| MDCD | 1,938 | 16,115 | 0.9 | 8.7 | <5 | 108 | < 5.56 (1.80 - 13.0) | 12.4 (10.2 - 15.0) |
| Open Claims | 221,964 | 823,043 | 124.8 | 656.7 | 445 | 4,270 | 3.57 (3.24 - 3.91) | 6.50 (6.31 - 6.70) |
| OEHR | 13,250 | 81,268 | 3.4 | 38.9 | 25 | 366 | 7.36 (4.76 - 10.9) | 9.42 (8.48 - 10.4) |
| Meta-analysis | 266,514 | 1,032,943 | 144.3 | 784.6 | 565 | 5,486 | 3.92 (3.60 - 4.25) | 6.99 (6.81 - 7.18) |
| Meta-analysis (w/o USOC) | 44,550 | 209,900 | 19.6 | 127.9 | 120 | 1,216 | 6.14 (5.09 - 7.34) | 9.51 (8.98 - 10.1) |

##### 4.5.26 Vomiting

**Supplementary Table 415:** Exposures, outcomes and risk estimates for Vomiting in GLP1RA (female) (T) vs SU (female) (C) new-user cohorts under four different propensity score (PS) adjustment / time-at-risk (TAR) designs across data sources. We report total patient exposure time in 1,000 patient-year (KYr) units, uncalibrated and calibrated hazard ratios (HRs) and their 95% confidence intervals (CIs). Greyed out entries did not pass study diagnostics and are included only for completeness.

| | Patients | | Exposure (KYr) | | Outcomes | | Incidence (per KYr) | |
|---|---|---|---|---|---|---|---|---|
| | T | C | T | C | T | C | T | C |
| **PS-matching / on-treatment TAR** | | | | | | | | |
| CCAE | 13,485 | 61,930 | 8.2 | 45.6 | 348 | 1,327 | 42.3 (38.0 - 47.0) | 29.1 (27.6 - 30.7) |
| ODOD | 7,691 | 48,204 | 4.3 | 40.2 | 248 | 1,639 | 58.3 (51.2 - 66.0) | 40.7 (38.8 - 42.8) |
| MDCR | 1,024 | 12,009 | 0.6 | 10.8 | 28 | 382 | 43.7 (29.1 - 63.2) | 35.4 (32.0 - 39.2) |
| MDCD | 1,353 | 11,126 | 0.7 | 7.0 | 82 | 488 | 115 (91.7 - 143) | 69.8 (63.7 - 76.3) |
| Open Claims | 148,913 | 718,518 | 100.1 | 684.7 | 2,639 | 13,928 | 26.4 (25.4 - 27.4) | 20.3 (20.0 - 20.7) |
| OEHR | 9,884 | 71,116 | 3.0 | 39.0 | 139 | 1,408 | 46.9 (39.4 - 55.3) | 36.1 (34.2 - 38.0) |
| Meta-analysis | 182,350 | 922,903 | 116.9 | 827.3 | 3,484 | 19,172 | 29.8 (28.8 - 30.8) | 23.2 (22.8 - 23.5) |
| Meta-analysis (w/o USOC) | 33,437 | 204,385 | 16.8 | 142.6 | 845 | 5,244 | 50.3 (46.9 - 53.8) | 36.8 (35.8 - 37.8) |
| **PS-stratification / on-treatment TAR** | | | | | | | | |
| CCAE | 17,941 | 64,184 | 10.5 | 47.0 | 495 | 1,369 | 47.2 (43.1 - 51.6) | 29.2 (27.6 - 30.7) |
| ODOD | 9,990 | 52,035 | 5.2 | 44.2 | 323 | 1,857 | 62.0 (55.4 - 69.1) | 42.0 (40.1 - 44.0) |
| MDCR | 1,132 | 15,584 | 0.7 | 13.8 | 28 | 515 | 41.2 (27.4 - 59.5) | 37.3 (34.1 - 40.7) |
| MDCD | 1,441 | 12,420 | 0.7 | 7.9 | 87 | 565 | 116 (93.2 - 144) | 71.3 (65.5 - 77.4) |
| Open Claims | 191,403 | 733,774 | 119.3 | 699.2 | 3,339 | 14,509 | 28.0 (27.1 - 29.0) | 20.8 (20.4 - 21.1) |
| OEHR | 12,004 | 75,585 | 3.5 | 41.7 | 178 | 1,545 | 51.0 (43.8 - 59.1) | 37.0 (35.2 - 38.9) |
| Meta-analysis | 231,338 | 925,578 | 138.5 | 832.1 | 4,335 | 19,280 | 31.3 (30.4 - 32.3) | 23.2 (22.8 - 23.5) |
| Meta-analysis (w/o USOC) | 39,935 | 191,804 | 19.2 | 132.9 | 996 | 4,771 | 51.9 (48.7 - 55.2) | 35.9 (34.9 - 36.9) |
| **PS-matching / on-treatment TAR without escalation** | | | | | | | | |
| CCAE | 13,485 | 61,930 | 7.4 | 39.3 | 318 | 1,108 | 42.8 (38.2 - 47.8) | 28.2 (26.5 - 29.9) |
| ODOD | 7,691 | 48,204 | 3.9 | 35.9 | 226 | 1,463 | 58.6 (51.2 - 66.8) | 40.7 (38.7 - 42.9) |
| MDCR | 1,024 | 12,009 | 0.6 | 9.9 | 25 | 334 | 42.4 (27.4 - 62.5) | 33.9 (30.4 - 37.7) |
| MDCD | 1,353 | 11,126 | 0.6 | 5.9 | 73 | 414 | 119 (93.3 - 150) | 70.8 (64.1 - 77.9) |
| Open Claims | 148,913 | 718,518 | 89.5 | 578.7 | 2,366 | 11,482 | 26.4 (25.4 - 27.5) | 19.8 (19.5 - 20.2) |
| OEHR | 9,884 | 71,116 | 2.6 | 33.4 | 120 | 1,114 | 46.9 (38.9 - 56.1) | 33.3 (31.4 - 35.3) |
| Meta-analysis | 182,350 | 922,903 | 104.6 | 703.2 | 3,128 | 15,915 | 29.9 (28.9 - 31.0) | 22.6 (22.3 - 23.0) |
| Meta-analysis (w/o USOC) | 33,437 | 204,385 | 15.0 | 124.4 | 762 | 4,433 | 50.7 (47.1 - 54.4) | 35.6 (34.6 - 36.7) |
| **PS-stratification / on-treatment TAR without escalation** | | | | | | | | |
| CCAE | 17,941 | 64,184 | 9.5 | 40.6 | 456 | 1,148 | 47.8 (43.5 - 52.4) | 28.3 (26.7 - 30.0) |
| ODOD | 9,990 | 52,035 | 4.7 | 39.5 | 294 | 1,667 | 61.9 (55.1 - 69.4) | 42.2 (40.2 - 44.2) |
| MDCR | 1,132 | 15,584 | 0.6 | 12.6 | 25 | 449 | 39.7 (25.7 - 58.6) | 35.6 (32.4 - 39.0) |
| MDCD | 1,441 | 12,420 | 0.6 | 6.7 | 78 | 487 | 120 (95.1 - 150) | 73.2 (66.9 - 80.0) |
| Open Claims | 191,403 | 733,774 | 107.8 | 591.6 | 3,025 | 11,994 | 28.1 (27.1 - 29.1) | 20.3 (19.9 - 20.6) |
| OEHR | 12,004 | 75,585 | 3.0 | 35.9 | 154 | 1,228 | 50.5 (42.9 - 59.2) | 34.2 (32.4 - 36.2) |
| Meta-analysis | 231,338 | 925,578 | 125.1 | 707.5 | 3,929 | 16,037 | 31.4 (30.4 - 32.4) | 22.7 (22.3 - 23.0) |
| Meta-analysis (w/o USOC) | 39,935 | 191,804 | 17.3 | 116.0 | 904 | 4,043 | 52.2 (48.8 - 55.7) | 34.9 (33.8 - 36.0) |

#### 4.6 DPP4I (female) vs SU (female)

##### 4.6.1 3-point MACE

**Supplementary Table 416:** Exposures, outcomes and risk estimates for 3-point MACE in DPP4I (female) (T) vs SU (female) (C) new-user cohorts under four different propensity score (PS) adjustment / time-at-risk (TAR) designs across data sources. We report total patient exposure time in 1,000 patient-year (KYr) units, uncalibrated and calibrated hazard ratios (HRs) and their 95% confidence intervals (CIs). Greyed out entries did not pass study diagnostics and are included only for completeness.

| | Patients | | Exposure (KYr) | | Outcomes | | Incidence (per KYr) | |
|---|---|---|---|---|---|---|---|---|
| | T | C | T | C | T | C | T | C |
| **PS-matching / on-treatment TAR** | | | | | | | | |
| CCAE | 38,239 | 64,329 | 29.9 | 46.8 | 175 | 336 | 5.85 (5.02 - 6.78) | 7.18 (6.43 - 7.99) |
| DAG | 10,485 | 2,040 | 11.4 | 2.1 | 0 | 0 | | |
| ODOD | 26,182 | 57,090 | 20.6 | 48.9 | 376 | 1,113 | 18.2 (16.4 - 20.2) | 22.8 (21.4 - 24.1) |
| MDCR | 7,532 | 14,216 | 7.1 | 12.6 | 169 | 392 | 23.9 (20.5 - 27.8) | 31.2 (28.2 - 34.4) |
| MDCD | 6,936 | 15,653 | 4.9 | 10.2 | 96 | 208 | 19.6 (15.9 - 24.0) | 20.3 (17.7 - 23.3) |
| Open Claims | 418,312 | 810,705 | 363.9 | 767.8 | 3,791 | 9,194 | 10.4 (10.1 - 10.8) | 12.0 (11.7 - 12.2) |
| OEHR | 35,813 | 81,690 | 18.3 | 46.1 | 153 | 586 | 8.38 (7.11 - 9.82) | 12.7 (11.7 - 13.8) |
| VA | 1,770 | 9,474 | 1.3 | 7.3 | <10 | 31 | < 7.43 (3.56 - 13.7) | 4.24 (2.88 - 6.02) |
| SIDIAP | 19,287 | 5,403 | 53.0 | 14.1 | 144 | 44 | 2.72 (2.29 - 3.20) | 3.12 (2.27 - 4.19) |
| Meta-analysis | 554,071 | 1,058,560 | 499.0 | 953.8 | <4,914 | 11,904 | < 9.85 (9.57 - 10.1) | 12.5 (12.3 - 12.7) |
| Meta-analysis (w/o USOC) | 135,759 | 247,855 | 135.1 | 186.0 | <1,123 | 2,710 | < 8.31 (7.83 - 8.81) | 14.6 (14.0 - 15.1) |
| **PS-stratification / on-treatment TAR** | | | | | | | | |
| CCAE | 44,510 | 66,329 | 35.1 | 48.0 | 203 | 341 | 5.78 (5.01 - 6.63) | 7.10 (6.37 - 7.90) |
| DAG | 10,516 | 2,367 | 11.4 | 2.5 | 0 | 0 | | |
| ODOD | 28,169 | 57,247 | 22.3 | 48.9 | 400 | 1,113 | 18.0 (16.2 - 19.8) | 22.7 (21.4 - 24.1) |
| MDCR | 8,124 | 14,511 | 7.6 | 12.8 | 182 | 401 | 23.8 (20.5 - 27.6) | 31.2 (28.3 - 34.5) |
| MDCD | 7,282 | 15,757 | 5.2 | 10.2 | 102 | 208 | 19.7 (16.1 - 23.9) | 20.3 (17.6 - 23.2) |
| Open Claims | 428,138 | 810,802 | 374.7 | 767.8 | 3,943 | 9,195 | 10.5 (10.2 - 10.9) | 12.0 (11.7 - 12.2) |
| OEHR | 38,260 | 81,775 | 19.8 | 46.2 | 161 | 586 | 8.15 (6.94 - 9.51) | 12.7 (11.7 - 13.8) |
| VA | 2,150 | 12,928 | 1.6 | 10.4 | <10 | 50 | < 6.33 (3.03 - 11.6) | 4.82 (3.58 - 6.35) |
| SIDIAP | 19,389 | 7,031 | 53.1 | 19.3 | 144 | 55 | 2.71 (2.29 - 3.19) | 2.86 (2.15 - 3.72) |
| Meta-analysis | 566,590 | 1,037,695 | 512.6 | 943.0 | 5,033 | 11,691 | 9.82 (9.55 - 10.1) | 12.4 (12.2 - 12.6) |
| Meta-analysis (w/o USOC) | 138,452 | 226,893 | 137.9 | 175.2 | 1,090 | 2,496 | 7.90 (7.44 - 8.39) | 14.2 (13.7 - 14.8) |
| **PS-matching / on-treatment TAR without escalation** | | | | | | | | |
| CCAE | 38,239 | 64,329 | 24.6 | 40.3 | 135 | 282 | 5.49 (4.60 - 6.50) | 6.99 (6.20 - 7.86) |
| DAG | 10,485 | 2,040 | 10.3 | 2.0 | 0 | 0 | | |
| ODOD | 26,182 | 57,090 | 17.3 | 43.5 | 331 | 989 | 19.1 (17.1 - 21.3) | 22.7 (21.3 - 24.2) |
| MDCR | 7,532 | 14,216 | 6.1 | 11.6 | 136 | 352 | 22.1 (18.6 - 26.2) | 30.4 (27.3 - 33.8) |
| MDCD | 6,936 | 15,653 | 3.8 | 8.5 | 72 | 170 | 18.8 (14.7 - 23.7) | 20.1 (17.2 - 23.3) |
| Open Claims | 418,312 | 810,705 | 288.9 | 646.3 | 3,075 | 7,802 | 10.6 (10.3 - 11.0) | 12.1 (11.8 - 12.3) |
| OEHR | 35,813 | 81,690 | 14.7 | 39.3 | 98 | 426 | 6.66 (5.41 - 8.12) | 10.9 (9.85 - 11.9) |
| VA | 1,770 | 9,474 | 1.1 | 6.3 | <10 | 24 | < 9.34 (4.48 - 17.2) | 3.80 (2.43 - 5.65) |
| SIDIAP | 19,287 | 5,403 | 39.4 | 11.5 | 114 | 39 | 2.89 (2.39 - 3.47) | 3.40 (2.42 - 4.65) |
| Meta-analysis | 554,071 | 1,058,560 | 396.0 | 807.2 | <3,971 | 10,084 | < 10.0 (9.72 - 10.3) | 12.5 (12.2 - 12.7) |
| Meta-analysis (w/o USOC) | 135,759 | 247,855 | 107.1 | 160.9 | <896 | 2,282 | < 8.37 (7.83 - 8.93) | 14.2 (13.6 - 14.8) |
| **PS-stratification / on-treatment TAR without escalation** | | | | | | | | |
| CCAE | 44,510 | 66,329 | 28.9 | 41.4 | 158 | 286 | 5.46 (4.64 - 6.38) | 6.90 (6.13 - 7.75) |
| DAG | 10,516 | 2,367 | 10.3 | 2.3 | 0 | 0 | | |
| ODOD | 28,169 | 57,247 | 18.7 | 43.6 | 353 | 989 | 18.9 (17.0 - 21.0) | 22.7 (21.3 - 24.2) |
| MDCR | 8,124 | 14,511 | 6.6 | 11.8 | 147 | 359 | 22.1 (18.7 - 26.0) | 30.4 (27.3 - 33.7) |
| MDCD | 7,282 | 15,757 | 4.0 | 8.5 | 78 | 170 | 19.3 (15.2 - 24.1) | 20.0 (17.1 - 23.2) |
| Open Claims | 428,138 | 810,802 | 297.6 | 646.3 | 3,196 | 7,803 | 10.7 (10.4 - 11.1) | 12.1 (11.8 - 12.3) |
| OEHR | 38,260 | 81,775 | 15.9 | 39.3 | 104 | 426 | 6.53 (5.33 - 7.91) | 10.8 (9.83 - 11.9) |
| VA | 2,150 | 12,928 | 1.3 | 9.0 | <10 | 41 | < 7.94 (3.81 - 14.6) | 4.53 (3.25 - 6.15) |
| SIDIAP | 19,389 | 7,031 | 39.5 | 15.6 | 114 | 48 | 2.89 (2.38 - 3.47) | 3.08 (2.27 - 4.09) |
| Meta-analysis | 566,590 | 1,037,695 | 407.3 | 798.0 | 4,072 | 9,911 | 10.00 (9.69 - 10.3) | 12.4 (12.2 - 12.7) |
| Meta-analysis (w/o USOC) | 138,452 | 226,893 | 109.7 | 151.7 | 876 | 2,108 | 7.99 (7.47 - 8.53) | 13.9 (13.3 - 14.5) |

##### 4.6.2 4-point MACE

**Supplementary Table 417:** Exposures, outcomes and risk estimates for 4-point MACE in DPP4I (female) (T) vs SU (female) (C) new-user cohorts under four different propensity score (PS) adjustment / time-at-risk (TAR) designs across data sources. We report total patient exposure time in 1,000 patient-year (KYr) units, uncalibrated and calibrated hazard ratios (HRs) and their 95% confidence intervals (CIs). Greyed out entries did not pass study diagnostics and are included only for completeness.

| | Patients | | Exposure (KYr) | | Outcomes | | Incidence (per KYr) | |
|---|---|---|---|---|---|---|---|---|
| | T | C | T | C | T | C | T | C |
| **PS-matching / on-treatment TAR** | | | | | | | | |
| CCAE | 37,689 | 63,317 | 29.5 | 46.0 | 285 | 546 | 9.66 (8.57 - 10.9) | 11.9 (10.9 - 12.9) |
| DAG | 10,485 | 2,040 | 11.4 | 2.1 | 0 | 0 | | |
| ODOD | 25,033 | 54,385 | 19.6 | 46.1 | 642 | 1,803 | 32.8 (30.3 - 35.5) | 39.1 (37.3 - 40.9) |
| MDCR | 7,009 | 13,031 | 6.6 | 11.4 | 258 | 622 | 39.1 (34.4 - 44.1) | 54.7 (50.4 - 59.1) |
| MDCD | 6,470 | 14,576 | 4.5 | 9.4 | 170 | 355 | 37.8 (32.3 - 43.9) | 37.7 (33.9 - 41.9) |
| Open Claims | 404,455 | 783,629 | 350.4 | 738.0 | 6,256 | 15,360 | 17.9 (17.4 - 18.3) | 20.8 (20.5 - 21.1) |
| OEHR | 35,226 | 80,207 | 17.8 | 44.7 | 310 | 1,088 | 17.4 (15.5 - 19.5) | 24.3 (22.9 - 25.8) |
| VA | 1,738 | 9,206 | 1.3 | 7.1 | 12 | 58 | 9.10 (4.70 - 15.9) | 8.20 (6.23 - 10.6) |
| SIDIAP | 19,149 | 5,388 | 52.8 | 14.0 | 195 | 48 | 3.69 (3.19 - 4.25) | 3.42 (2.52 - 4.53) |
| Meta-analysis | 536,769 | 1,023,739 | 482.5 | 916.7 | 8,128 | 19,880 | 16.8 (16.5 - 17.2) | 21.7 (21.4 - 22.0) |
| Meta-analysis (w/o USOC) | 132,314 | 240,110 | 132.1 | 178.7 | 1,872 | 4,520 | 14.2 (13.5 - 14.8) | 25.3 (24.6 - 26.0) |
| **PS-stratification / on-treatment TAR** | | | | | | | | |
| CCAE | 43,928 | 65,293 | 34.7 | 47.2 | 329 | 558 | 9.48 (8.49 - 10.6) | 11.8 (10.9 - 12.9) |
| DAG | 10,516 | 2,367 | 11.4 | 2.5 | 0 | 0 | | |
| ODOD | 27,017 | 54,539 | 21.2 | 46.2 | 680 | 1,805 | 32.1 (29.7 - 34.6) | 39.1 (37.3 - 41.0) |
| MDCR | 7,581 | 13,379 | 7.1 | 11.7 | 286 | 641 | 40.1 (35.6 - 45.0) | 54.7 (50.6 - 59.1) |
| MDCD | 6,786 | 14,676 | 4.8 | 9.4 | 181 | 355 | 37.9 (32.6 - 43.9) | 37.7 (33.8 - 41.8) |
| Open Claims | 414,120 | 783,725 | 361.1 | 738.0 | 6,491 | 15,362 | 18.0 (17.5 - 18.4) | 20.8 (20.5 - 21.1) |
| OEHR | 37,672 | 80,288 | 19.3 | 44.8 | 325 | 1,088 | 16.8 (15.1 - 18.8) | 24.3 (22.9 - 25.8) |
| VA | 2,111 | 12,720 | 1.5 | 10.2 | 15 | 101 | 9.68 (5.42 - 16.0) | 9.91 (8.07 - 12.0) |
| SIDIAP | 19,309 | 7,015 | 52.9 | 19.2 | 195 | 65 | 3.69 (3.19 - 4.24) | 3.39 (2.61 - 4.31) |
| Meta-analysis | 549,627 | 1,004,239 | 496.3 | 907.0 | 8,306 | 19,519 | 16.7 (16.4 - 17.1) | 21.5 (21.2 - 21.8) |
| Meta-analysis (w/o USOC) | 135,507 | 220,514 | 135.2 | 169.0 | 1,815 | 4,157 | 13.4 (12.8 - 14.1) | 24.6 (23.9 - 25.4) |
| **PS-matching / on-treatment TAR without escalation** | | | | | | | | |
| CCAE | 37,689 | 63,317 | 24.2 | 39.7 | 227 | 471 | 9.36 (8.18 - 10.7) | 11.9 (10.8 - 13.0) |
| DAG | 10,485 | 2,040 | 10.3 | 2.0 | 0 | 0 | | |
| ODOD | 25,033 | 54,385 | 16.4 | 41.1 | 543 | 1,588 | 33.0 (30.3 - 35.9) | 38.6 (36.7 - 40.6) |
| MDCR | 7,009 | 13,031 | 5.8 | 10.5 | 216 | 566 | 37.6 (32.7 - 42.9) | 53.9 (49.5 - 58.5) |
| MDCD | 6,470 | 14,576 | 3.5 | 7.8 | 129 | 289 | 36.7 (30.6 - 43.6) | 37.0 (32.8 - 41.5) |
| Open Claims | 404,455 | 783,629 | 278.5 | 621.7 | 5,035 | 12,980 | 18.1 (17.6 - 18.6) | 20.9 (20.5 - 21.2) |
| OEHR | 35,226 | 80,207 | 14.4 | 38.3 | 210 | 827 | 14.6 (12.7 - 16.7) | 21.6 (20.2 - 23.1) |
| VA | 1,738 | 9,206 | 1.0 | 6.1 | <10 | 48 | < 9.54 (4.57 - 17.5) | 7.84 (5.78 - 10.4) |
| SIDIAP | 19,149 | 5,388 | 39.2 | 11.4 | 145 | 43 | 3.70 (3.12 - 4.35) | 3.77 (2.73 - 5.07) |
| Meta-analysis | 536,769 | 1,023,739 | 383.1 | 776.6 | <6,515 | 16,812 | < 17.0 (16.6 - 17.4) | 21.6 (21.3 - 22.0) |
| Meta-analysis (w/o USOC) | 132,314 | 240,110 | 104.6 | 154.9 | <1,480 | 3,832 | < 14.1 (13.4 - 14.9) | 24.7 (24.0 - 25.5) |
| **PS-stratification / on-treatment TAR without escalation** | | | | | | | | |
| CCAE | 43,928 | 65,293 | 28.6 | 40.7 | 264 | 480 | 9.24 (8.16 - 10.4) | 11.8 (10.8 - 12.9) |
| DAG | 10,516 | 2,367 | 10.3 | 2.3 | 0 | 0 | | |
| ODOD | 27,017 | 54,539 | 17.8 | 41.1 | 574 | 1,590 | 32.2 (29.6 - 35.0) | 38.6 (36.8 - 40.6) |
| MDCR | 7,581 | 13,379 | 6.2 | 10.8 | 240 | 583 | 38.6 (33.9 - 43.8) | 54.0 (49.7 - 58.5) |
| MDCD | 6,786 | 14,676 | 3.7 | 7.8 | 138 | 289 | 36.9 (31.0 - 43.6) | 36.9 (32.7 - 41.4) |
| Open Claims | 414,120 | 783,725 | 287.0 | 621.7 | 5,220 | 12,982 | 18.2 (17.7 - 18.7) | 20.9 (20.5 - 21.2) |
| OEHR | 37,672 | 80,288 | 15.6 | 38.3 | 222 | 827 | 14.2 (12.4 - 16.2) | 21.6 (20.1 - 23.1) |
| VA | 2,111 | 12,720 | 1.2 | 8.9 | <10 | 83 | < 8.11 (3.89 - 14.9) | 9.33 (7.43 - 11.6) |
| SIDIAP | 19,309 | 7,015 | 39.4 | 15.5 | 145 | 58 | 3.68 (3.11 - 4.34) | 3.74 (2.84 - 4.83) |
| Meta-analysis | 549,627 | 1,004,239 | 394.6 | 768.3 | 6,665 | 16,520 | 16.9 (16.5 - 17.3) | 21.5 (21.2 - 21.8) |
| Meta-analysis (w/o USOC) | 135,507 | 220,514 | 107.6 | 146.5 | 1,445 | 3,538 | 13.4 (12.8 - 14.1) | 24.1 (23.4 - 25.0) |

##### 4.6.3 Acute myocardial infarction

**Supplementary Table 418:** Exposures, outcomes and risk estimates for Acute myocardial infarction in DPP4I (female) (T) vs SU (female) (C) new-user cohorts under four different propensity score (PS) adjustment / time-at-risk (TAR) designs across data sources. We report total patient exposure time in 1,000 patient-year (KYr) units, uncalibrated and calibrated hazard ratios (HRs) and their 95% confidence intervals (CIs). Greyed out entries did not pass study diagnostics and are included only for completeness.

| | Patients | | Exposure (KYr) | | Outcomes | | Incidence (per KYr) | |
|---|---|---|---|---|---|---|---|---|
| | T | C | T | C | T | C | T | C |
| **PS-matching / on-treatment TAR** | | | | | | | | |
| CCAE | 38,717 | 65,227 | 30.3 | 47.6 | 75 | 145 | 2.48 (1.95 - 3.11) | 3.05 (2.57 - 3.59) |
| DAG | 10,485 | 2,040 | 11.4 | 2.1 | 0 | 0 | | |
| ODOD | 26,968 | 58,852 | 21.3 | 50.9 | 141 | 481 | 6.61 (5.56 - 7.79) | 9.45 (8.63 - 10.3) |
| MDCR | 7,901 | 14,944 | 7.4 | 13.3 | 71 | 160 | 9.57 (7.48 - 12.1) | 12.0 (10.2 - 14.0) |
| MDCD | 7,213 | 16,301 | 5.1 | 10.7 | 31 | 83 | 6.07 (4.12 - 8.61) | 7.77 (6.19 - 9.63) |
| Open Claims | 430,562 | 833,795 | 375.3 | 792.0 | 1,609 | 3,979 | 4.29 (4.08 - 4.50) | 5.02 (4.87 - 5.18) |
| OEHR | 36,117 | 82,493 | 18.5 | 46.7 | 64 | 277 | 3.46 (2.67 - 4.42) | 5.93 (5.25 - 6.67) |
| VA | 1,796 | 9,581 | 1.4 | 7.4 | <10 | 16 | < 7.31 (3.51 - 13.4) | 2.16 (1.24 - 3.51) |
| SIDIAP | 19,424 | 5,426 | 53.5 | 14.2 | 45 | 22 | 0.84 (0.61 - 1.13) | 1.55 (0.97 - 2.35) |
| Meta-analysis | 566,902 | 1,077,038 | 511.4 | 975.3 | 2,036 | 5,147 | 3.98 (3.81 - 4.16) | 5.28 (5.13 - 5.42) |
| Meta-analysis (w/o USOC) | 136,340 | 243,243 | 136.1 | 183.4 | 427 | 1,168 | 3.14 (2.85 - 3.45) | 6.37 (6.01 - 6.75) |
| **PS-stratification / on-treatment TAR** | | | | | | | | |
| CCAE | 45,027 | 67,241 | 35.5 | 48.8 | 85 | 146 | 2.39 (1.91 - 2.96) | 2.99 (2.53 - 3.52) |
| DAG | 10,516 | 2,367 | 11.4 | 2.5 | 0 | 0 | | |
| ODOD | 28,975 | 59,009 | 23.0 | 50.9 | 147 | 481 | 6.39 (5.40 - 7.51) | 9.45 (8.62 - 10.3) |
| MDCR | 8,520 | 15,256 | 8.0 | 13.6 | 74 | 163 | 9.22 (7.24 - 11.6) | 12.0 (10.2 - 14.0) |
| MDCD | 7,556 | 16,407 | 5.4 | 10.7 | 33 | 83 | 6.11 (4.20 - 8.58) | 7.75 (6.17 - 9.60) |
| Open Claims | 440,491 | 833,892 | 386.3 | 792.0 | 1,670 | 3,979 | 4.32 (4.12 - 4.54) | 5.02 (4.87 - 5.18) |
| OEHR | 38,583 | 82,578 | 20.0 | 46.8 | 69 | 277 | 3.45 (2.68 - 4.37) | 5.92 (5.24 - 6.66) |
| VA | 2,188 | 13,082 | 1.6 | 10.5 | <10 | 29 | < 6.22 (2.98 - 11.4) | 2.76 (1.85 - 3.96) |
| SIDIAP | 19,530 | 7,058 | 53.5 | 19.3 | 45 | 26 | 0.84 (0.61 - 1.12) | 1.34 (0.88 - 1.97) |
| Meta-analysis | 581,126 | 1,065,034 | 526.5 | 971.4 | 2,090 | 5,072 | 3.97 (3.80 - 4.14) | 5.22 (5.08 - 5.37) |
| Meta-analysis (w/o USOC) | 140,635 | 231,142 | 140.1 | 179.4 | 420 | 1,093 | 3.00 (2.72 - 3.30) | 6.09 (5.74 - 6.46) |
| **PS-matching / on-treatment TAR without escalation** | | | | | | | | |
| CCAE | 38,717 | 65,227 | 24.9 | 41.0 | 52 | 123 | 2.09 (1.56 - 2.74) | 3.00 (2.50 - 3.58) |
| DAG | 10,485 | 2,040 | 10.3 | 2.0 | 0 | 0 | | |
| ODOD | 26,968 | 58,852 | 17.9 | 45.2 | 126 | 423 | 7.04 (5.87 - 8.38) | 9.35 (8.48 - 10.3) |
| MDCR | 7,901 | 14,944 | 6.4 | 12.2 | 63 | 143 | 9.78 (7.52 - 12.5) | 11.7 (9.87 - 13.8) |
| MDCD | 7,213 | 16,301 | 4.0 | 8.9 | 21 | 66 | 5.28 (3.27 - 8.07) | 7.44 (5.76 - 9.47) |
| Open Claims | 430,562 | 833,795 | 298.0 | 666.3 | 1,301 | 3,391 | 4.37 (4.13 - 4.61) | 5.09 (4.92 - 5.26) |
| OEHR | 36,117 | 82,493 | 14.9 | 39.7 | 39 | 201 | 2.62 (1.87 - 3.59) | 5.06 (4.38 - 5.81) |
| VA | 1,796 | 9,581 | 1.1 | 6.4 | <10 | 14 | < 9.19 (4.41 - 16.9) | 2.19 (1.20 - 3.67) |
| SIDIAP | 19,424 | 5,426 | 39.8 | 11.5 | 34 | 19 | 0.86 (0.59 - 1.19) | 1.65 (0.99 - 2.58) |
| Meta-analysis | 566,902 | 1,077,038 | 405.8 | 824.9 | 1,636 | 4,366 | 4.03 (3.84 - 4.23) | 5.29 (5.14 - 5.45) |
| Meta-analysis (w/o USOC) | 136,340 | 243,243 | 107.8 | 158.6 | 335 | 975 | 3.11 (2.78 - 3.46) | 6.15 (5.77 - 6.55) |
| **PS-stratification / on-treatment TAR without escalation** | | | | | | | | |
| CCAE | 45,027 | 67,241 | 29.2 | 42.1 | 58 | 124 | 1.98 (1.51 - 2.56) | 2.95 (2.45 - 3.52) |
| DAG | 10,516 | 2,367 | 10.3 | 2.3 | 0 | 0 | | |
| ODOD | 28,975 | 59,009 | 19.3 | 45.3 | 132 | 423 | 6.84 (5.72 - 8.11) | 9.34 (8.47 - 10.3) |
| MDCR | 8,520 | 15,256 | 7.0 | 12.5 | 65 | 145 | 9.32 (7.19 - 11.9) | 11.6 (9.81 - 13.7) |
| MDCD | 7,556 | 16,407 | 4.2 | 8.9 | 23 | 66 | 5.47 (3.47 - 8.21) | 7.42 (5.74 - 9.44) |
| Open Claims | 440,491 | 833,892 | 306.8 | 666.4 | 1,346 | 3,391 | 4.39 (4.16 - 4.63) | 5.09 (4.92 - 5.26) |
| OEHR | 38,583 | 82,578 | 16.1 | 39.8 | 42 | 201 | 2.61 (1.88 - 3.52) | 5.05 (4.38 - 5.80) |
| VA | 2,188 | 13,082 | 1.3 | 9.2 | <10 | 26 | < 7.79 (3.74 - 14.3) | 2.84 (1.85 - 4.16) |
| SIDIAP | 19,530 | 7,058 | 39.8 | 15.7 | 34 | 22 | 0.85 (0.59 - 1.19) | 1.41 (0.88 - 2.13) |
| Meta-analysis | 581,126 | 1,065,034 | 418.3 | 821.6 | 1,677 | 4,306 | 4.01 (3.82 - 4.21) | 5.24 (5.09 - 5.40) |
| Meta-analysis (w/o USOC) | 140,635 | 231,142 | 111.5 | 155.3 | 331 | 915 | 2.97 (2.66 - 3.31) | 5.89 (5.52 - 6.29) |

##### 4.6.4 Acute renal failure

**Supplementary Table 419:** Exposures, outcomes and risk estimates for Acute renal failure in DPP4I (female) (T) vs SU (female) (C) new-user cohorts under four different propensity score (PS) adjustment / time-at-risk (TAR) designs across data sources. We report total patient exposure time in 1,000 patient-year (KYr) units, uncalibrated and calibrated hazard ratios (HRs) and their 95% confidence intervals (CIs). Greyed out entries did not pass study diagnostics and are included only for completeness.

| | Patients | | Exposure (KYr) | | Outcomes | | Incidence (per KYr) | |
|---|---|---|---|---|---|---|---|---|
| | T | C | T | C | T | C | T | C |
| **PS-matching / on-treatment TAR** | | | | | | | | |
| CCAE | 38,636 | 64,976 | 30.2 | 47.3 | 194 | 406 | 6.41 (5.54 - 7.38) | 8.58 (7.77 - 9.46) |
| DAG | 10,485 | 2,040 | 11.4 | 2.1 | 0 | 0 | | |
| ODOD | 26,125 | 56,950 | 20.4 | 48.6 | 577 | 1,557 | 28.2 (26.0 - 30.6) | 32.0 (30.4 - 33.6) |
| MDCR | 7,704 | 14,462 | 7.2 | 12.7 | 218 | 458 | 30.4 (26.5 - 34.7) | 36.0 (32.8 - 39.4) |
| MDCD | 7,000 | 15,775 | 4.8 | 10.2 | 157 | 322 | 32.4 (27.6 - 37.9) | 31.7 (28.3 - 35.3) |
| Open Claims | 423,871 | 821,287 | 366.9 | 775.7 | 5,336 | 12,429 | 14.5 (14.2 - 14.9) | 16.0 (15.7 - 16.3) |
| OEHR | 35,910 | 81,969 | 18.3 | 46.1 | 259 | 763 | 14.2 (12.5 - 16.0) | 16.5 (15.4 - 17.8) |
| VA | 1,766 | 9,451 | 1.3 | 7.2 | 16 | 78 | 12.1 (6.89 - 19.6) | 10.8 (8.51 - 13.4) |
| SIDIAP | 19,487 | 5,438 | 53.7 | 14.2 | 16 | <5 | 0.30 (0.17 - 0.48) | < 0.35 (0.11 - 0.82) |
| Meta-analysis | 541,012 | 1,064,870 | 449.2 | 947.9 | 6,757 | 16,013 | 15.0 (14.7 - 15.4) | 16.9 (16.6 - 17.2) |
| Meta-analysis (w/o USOC) | 117,141 | 243,583 | 82.3 | 172.2 | 1,421 | 3,584 | 17.3 (16.4 - 18.2) | 20.8 (20.1 - 21.5) |
| **PS-stratification / on-treatment TAR** | | | | | | | | |
| CCAE | 44,957 | 66,985 | 35.5 | 48.5 | 225 | 413 | 6.34 (5.54 - 7.22) | 8.51 (7.71 - 9.38) |
| DAG | 10,516 | 2,367 | 11.4 | 2.5 | 0 | 0 | | |
| ODOD | 28,091 | 57,107 | 22.1 | 48.7 | 613 | 1,557 | 27.8 (25.6 - 30.1) | 32.0 (30.4 - 33.6) |
| MDCR | 8,325 | 14,834 | 7.8 | 13.1 | 238 | 466 | 30.6 (26.8 - 34.7) | 35.6 (32.5 - 39.0) |
| MDCD | 7,347 | 15,879 | 5.1 | 10.2 | 172 | 323 | 33.6 (28.8 - 39.0) | 31.7 (28.3 - 35.4) |
| Open Claims | 433,792 | 821,409 | 377.9 | 775.7 | 5,536 | 12,429 | 14.6 (14.3 - 15.0) | 16.0 (15.7 - 16.3) |
| OEHR | 38,353 | 82,056 | 19.8 | 46.2 | 267 | 763 | 13.5 (11.9 - 15.2) | 16.5 (15.4 - 17.7) |
| VA | 2,163 | 12,941 | 1.6 | 10.3 | 17 | 116 | 10.8 (6.30 - 17.3) | 11.2 (9.27 - 13.5) |
| SIDIAP | 19,596 | 7,072 | 53.8 | 19.4 | 16 | <5 | 0.30 (0.17 - 0.48) | < 0.26 (0.08 - 0.60) |
| Meta-analysis | 573,114 | 1,049,463 | 516.8 | 951.5 | 6,895 | <15,633 | 13.3 (13.0 - 13.7) | < 16.4 (16.2 - 16.7) |
| Meta-analysis (w/o USOC) | 139,322 | 228,054 | 138.9 | 175.8 | 1,359 | <3,204 | 9.79 (9.27 - 10.3) | < 18.2 (17.6 - 18.9) |
| **PS-matching / on-treatment TAR without escalation** | | | | | | | | |
| CCAE | 38,636 | 64,976 | 24.8 | 40.8 | 153 | 334 | 6.16 (5.22 - 7.22) | 8.19 (7.33 - 9.11) |
| DAG | 10,485 | 2,040 | 10.3 | 2.0 | 0 | 0 | | |
| ODOD | 26,125 | 56,950 | 17.2 | 43.4 | 471 | 1,359 | 27.4 (25.0 - 30.0) | 31.3 (29.7 - 33.1) |
| MDCR | 7,704 | 14,462 | 6.3 | 11.7 | 180 | 405 | 28.7 (24.6 - 33.2) | 34.6 (31.3 - 38.2) |
| MDCD | 7,000 | 15,775 | 3.8 | 8.5 | 117 | 255 | 30.8 (25.5 - 37.0) | 30.1 (26.5 - 34.0) |
| Open Claims | 423,871 | 821,287 | 291.6 | 653.4 | 4,240 | 10,378 | 14.5 (14.1 - 15.0) | 15.9 (15.6 - 16.2) |
| OEHR | 35,910 | 81,969 | 14.7 | 39.4 | 177 | 527 | 12.0 (10.3 - 13.9) | 13.4 (12.3 - 14.6) |
| VA | 1,766 | 9,451 | 1.1 | 6.3 | 13 | 59 | 12.3 (6.55 - 21.0) | 9.40 (7.16 - 12.1) |
| SIDIAP | 19,487 | 5,438 | 39.9 | 11.6 | 15 | <5 | 0.38 (0.21 - 0.62) | < 0.43 (0.14 - 1.01) |
| Meta-analysis | 541,012 | 1,064,870 | 359.5 | 803.3 | 5,351 | 13,317 | 14.9 (14.5 - 15.3) | 16.6 (16.3 - 16.9) |
| Meta-analysis (w/o USOC) | 117,141 | 243,583 | 67.9 | 150.0 | 1,111 | 2,939 | 16.4 (15.4 - 17.4) | 19.6 (18.9 - 20.3) |
| **PS-stratification / on-treatment TAR without escalation** | | | | | | | | |
| CCAE | 44,957 | 66,985 | 29.2 | 41.9 | 177 | 339 | 6.06 (5.20 - 7.03) | 8.09 (7.25 - 9.00) |
| DAG | 10,516 | 2,367 | 10.3 | 2.3 | 0 | 0 | | |
| ODOD | 28,091 | 57,107 | 18.6 | 43.4 | 501 | 1,359 | 27.0 (24.7 - 29.5) | 31.3 (29.7 - 33.0) |
| MDCR | 8,325 | 14,834 | 6.8 | 12.0 | 198 | 413 | 29.1 (25.2 - 33.4) | 34.4 (31.1 - 37.9) |
| MDCD | 7,347 | 15,879 | 4.0 | 8.5 | 131 | 256 | 32.6 (27.3 - 38.7) | 30.1 (26.6 - 34.1) |
| Open Claims | 433,792 | 821,409 | 300.5 | 653.4 | 4,388 | 10,378 | 14.6 (14.2 - 15.0) | 15.9 (15.6 - 16.2) |
| OEHR | 38,353 | 82,056 | 16.0 | 39.4 | 184 | 527 | 11.5 (9.93 - 13.3) | 13.4 (12.3 - 14.6) |
| VA | 2,163 | 12,941 | 1.3 | 9.0 | 14 | 92 | 11.2 (6.12 - 18.8) | 10.2 (8.23 - 12.5) |
| SIDIAP | 19,596 | 7,072 | 40.0 | 15.7 | 15 | <5 | 0.38 (0.21 - 0.62) | < 0.32 (0.10 - 0.74) |
| Meta-analysis | 553,518 | 1,042,391 | 371.0 | 790.1 | 5,448 | 13,016 | 14.7 (14.3 - 15.1) | 16.5 (16.2 - 16.8) |
| Meta-analysis (w/o USOC) | 119,726 | 220,982 | 70.5 | 136.7 | 1,060 | 2,638 | 15.0 (14.1 - 16.0) | 19.3 (18.6 - 20.0) |

##### 4.6.5 Glycemic control

**Supplementary Table 420:** Exposures, outcomes and risk estimates for Glycemic control in DPP4I (female) (T) vs SU (female) (C) new-user cohorts under four different propensity score (PS) adjustment / time-at-risk (TAR) designs across data sources. We report total patient exposure time in 1,000 patient-year (KYr) units, uncalibrated and calibrated hazard ratios (HRs) and their 95% confidence intervals (CIs). Greyed out entries did not pass study diagnostics and are included only for completeness.

| | Patients | | Exposure (KYr) | | Outcomes | | Incidence (per KYr) | |
|---|---|---|---|---|---|---|---|---|
| | T | C | T | C | T | C | T | C |
| **PS-matching / on-treatment TAR** | | | | | | | | |
| CCAE | 38,694 | 65,531 | 30.3 | 47.7 | 81 | 255 | 2.67 (2.12 - 3.32) | 5.35 (4.71 - 6.05) |
| DAG | 3,753 | 846 | 3.3 | 0.8 | 465 | 95 | 140 (128 - 153) | 125 (101 - 153) |
| ODOD | 18,261 | 42,267 | 13.0 | 32.7 | 1,376 | 3,227 | 106 (101 - 112) | 98.7 (95.4 - 102) |
| MDCR | 8,123 | 15,281 | 7.7 | 13.6 | 12 | 44 | 1.57 (0.81 - 2.74) | 3.23 (2.34 - 4.33) |
| MDCD | 7,427 | 16,757 | 5.3 | 11.0 | 0 | 0 | | |
| Open Claims | 439,873 | 852,269 | 384.3 | 811.8 | 0 | 0 | | |
| OEHR | 36,419 | 83,277 | 18.7 | 47.4 | 0 | 0 | | |
| VA | 1,766 | 9,359 | 1.3 | 7.2 | <10 | <10 | < 7.50 (3.60 - 13.8) | < 1.39 (0.67 - 2.56) |
| SIDIAP | 2,096 | 1,017 | 3.4 | 1.7 | 881 | 348 | 261 (244 - 279) | 209 (187 - 232) |
| Meta-analysis | 70,927 | 124,942 | 57.6 | 96.4 | 2,815 | 3,969 | 48.9 (47.1 - 50.7) | 41.2 (39.9 - 42.5) |
| Meta-analysis (w/o USOC) | 70,927 | 124,942 | 57.6 | 96.4 | 2,815 | 3,969 | 48.9 (47.1 - 50.7) | 41.2 (39.9 - 42.5) |
| **PS-stratification / on-treatment TAR** | | | | | | | | |
| CCAE | 44,889 | 66,951 | 35.5 | 48.4 | 111 | 371 | 3.13 (2.57 - 3.76) | 7.67 (6.91 - 8.49) |
| DAG | 3,759 | 1,004 | 3.3 | 0.9 | 465 | 124 | 140 (128 - 153) | 138 (115 - 165) |
| ODOD | 19,175 | 42,367 | 13.6 | 32.7 | 1,468 | 3,230 | 108 (103 - 114) | 98.8 (95.4 - 102) |
| MDCR | 8,751 | 15,548 | 8.3 | 13.9 | 15 | 98 | 1.82 (1.02 - 2.99) | 7.07 (5.74 - 8.62) |
| MDCD | 7,771 | 16,863 | 5.6 | 11.0 | 0 | 0 | | |
| Open Claims | 449,878 | 852,367 | 395.5 | 811.8 | 0 | 0 | | |
| OEHR | 38,897 | 83,366 | 20.2 | 47.5 | 0 | 0 | | |
| VA | 2,158 | 12,709 | 1.6 | 10.1 | <10 | 16 | < 6.35 (3.05 - 11.7) | 1.58 (0.90 - 2.56) |
| SIDIAP | 2,111 | 1,580 | 3.4 | 2.8 | 885 | 545 | 261 (244 - 279) | 195 (179 - 212) |
| Meta-analysis | 78,685 | 127,450 | 64.1 | 98.6 | 2,944 | 4,368 | 45.9 (44.3 - 47.6) | 44.3 (43.0 - 45.6) |
| Meta-analysis (w/o USOC) | 78,685 | 127,450 | 64.1 | 98.6 | 2,944 | 4,368 | 45.9 (44.3 - 47.6) | 44.3 (43.0 - 45.6) |
| **PS-matching / on-treatment TAR without escalation** | | | | | | | | |
| CCAE | 38,694 | 65,531 | 24.9 | 41.0 | 72 | 232 | 2.89 (2.27 - 3.65) | 5.66 (4.95 - 6.43) |
| DAG | 3,753 | 846 | 3.0 | 0.7 | 415 | 94 | 138 (125 - 152) | 133 (107 - 162) |
| ODOD | 18,261 | 42,267 | 10.9 | 29.1 | 1,157 | 2,920 | 106 (100 - 113) | 100 (96.6 - 104) |
| MDCR | 8,123 | 15,281 | 6.7 | 12.5 | 9 | 42 | 1.35 (0.62 - 2.56) | 3.36 (2.43 - 4.55) |
| MDCD | 7,427 | 16,757 | 4.1 | 9.1 | 0 | 0 | | |
| Open Claims | 439,873 | 852,269 | 305.0 | 682.7 | 0 | 0 | | |
| OEHR | 36,419 | 83,277 | 15.0 | 40.2 | 0 | 0 | | |
| VA | 1,766 | 9,359 | 1.1 | 6.2 | <10 | <10 | < 9.46 (4.54 - 17.4) | < 1.61 (0.77 - 2.96) |
| SIDIAP | 2,096 | 1,017 | 2.3 | 1.3 | 684 | 287 | 298 (276 - 321) | 214 (190 - 240) |
| Meta-analysis | 70,927 | 124,942 | 47.7 | 84.7 | 2,337 | 3,575 | 49.0 (47.0 - 51.0) | 42.2 (40.8 - 43.6) |
| Meta-analysis (w/o USOC) | 70,927 | 124,942 | 47.7 | 84.7 | 2,337 | 3,575 | 49.0 (47.0 - 51.0) | 42.2 (40.8 - 43.6) |
| **PS-stratification / on-treatment TAR without escalation** | | | | | | | | |
| CCAE | 44,889 | 66,951 | 29.2 | 41.6 | 97 | 344 | 3.33 (2.70 - 4.06) | 8.26 (7.41 - 9.18) |
| DAG | 3,759 | 1,004 | 3.0 | 0.8 | 415 | 120 | 138 (125 - 151) | 143 (119 - 171) |
| ODOD | 19,175 | 42,367 | 11.4 | 29.2 | 1,230 | 2,923 | 108 (102 - 114) | 100 (96.6 - 104) |
| MDCR | 8,751 | 15,548 | 7.2 | 12.7 | 12 | 90 | 1.67 (0.86 - 2.92) | 7.10 (5.71 - 8.73) |
| MDCD | 7,771 | 16,863 | 4.3 | 9.2 | 0 | 0 | | |
| Open Claims | 449,878 | 852,367 | 314.0 | 682.7 | 0 | 0 | | |
| OEHR | 38,897 | 83,366 | 16.3 | 40.3 | 0 | 0 | | |
| VA | 2,158 | 12,709 | 1.3 | 8.8 | <10 | 15 | < 7.99 (3.83 - 14.7) | 1.70 (0.95 - 2.80) |
| SIDIAP | 2,111 | 1,580 | 2.3 | 2.2 | 688 | 456 | 298 (276 - 321) | 204 (186 - 224) |
| Meta-analysis | 78,685 | 127,450 | 53.1 | 86.5 | 2,442 | 3,933 | 46.0 (44.2 - 47.9) | 45.4 (44.0 - 46.9) |
| Meta-analysis (w/o USOC) | 78,685 | 127,450 | 53.1 | 86.5 | 2,442 | 3,933 | 46.0 (44.2 - 47.9) | 45.4 (44.0 - 46.9) |

##### 4.6.6 Hospitalization with heart failure

**Supplementary Table 421:** Exposures, outcomes and risk estimates for Hospitalization with heart failure in DPP4I (female) (T) vs SU (female) (C) new-user cohorts under four different propensity score (PS) adjustment / time-at-risk (TAR) designs across data sources. We report total patient exposure time in 1,000 patient-year (KYr) units, uncalibrated and calibrated hazard ratios (HRs) and their 95% confidence intervals (CIs). Greyed out entries did not pass study diagnostics and are included only for completeness.

| | Patients | | Exposure (KYr) | | Outcomes | | Incidence (per KYr) | |
|---|---|---|---|---|---|---|---|---|
| | T | C | T | C | T | C | T | C |
| **PS-matching / on-treatment TAR** | | | | | | | | |
| CCAE | 38,432 | 64,723 | 30.1 | 47.2 | 155 | 302 | 5.15 (4.37 - 6.02) | 6.40 (5.70 - 7.17) |
| DAG | 10,485 | 2,040 | 11.4 | 2.1 | 0 | 0 | | |
| ODOD | 25,936 | 56,484 | 20.3 | 48.4 | 452 | 1,268 | 22.2 (20.2 - 24.4) | 26.2 (24.8 - 27.7) |
| MDCR | 7,431 | 13,869 | 7.0 | 12.2 | 186 | 451 | 26.5 (22.8 - 30.6) | 37.0 (33.6 - 40.5) |
| MDCD | 6,798 | 15,306 | 4.8 | 9.9 | 114 | 250 | 24.0 (19.8 - 28.8) | 25.3 (22.3 - 28.6) |
| Open Claims | 420,971 | 815,544 | 365.9 | 771.4 | 4,146 | 10,029 | 11.3 (11.0 - 11.7) | 13.0 (12.7 - 13.3) |
| OEHR | 35,723 | 81,503 | 18.1 | 45.7 | 233 | 751 | 12.8 (11.2 - 14.6) | 16.4 (15.3 - 17.6) |
| VA | 1,778 | 9,366 | 1.3 | 7.2 | <10 | 39 | < 7.41 (3.55 - 13.6) | 5.41 (3.84 - 7.39) |
| SIDIAP | 19,369 | 5,425 | 53.5 | 14.2 | 58 | <5 | 1.08 (0.82 - 1.40) | < 0.35 (0.11 - 0.82) |
| Meta-analysis | 556,438 | 1,062,220 | 501.1 | 956.2 | <5,354 | <13,095 | < 10.7 (10.4 - 11.0) | < 13.7 (13.5 - 13.9) |
| Meta-analysis (w/o USOC) | 135,467 | 246,676 | 135.2 | 184.8 | <1,208 | <3,066 | < 8.93 (8.44 - 9.45) | < 16.6 (16.0 - 17.2) |
| **PS-stratification / on-treatment TAR** | | | | | | | | |
| CCAE | 44,755 | 66,729 | 35.4 | 48.4 | 177 | 311 | 5.00 (4.29 - 5.79) | 6.43 (5.74 - 7.19) |
| DAG | 10,516 | 2,367 | 11.4 | 2.5 | 0 | 0 | | |
| ODOD | 27,920 | 56,639 | 22.0 | 48.4 | 482 | 1,270 | 21.9 (20.0 - 24.0) | 26.2 (24.8 - 27.7) |
| MDCR | 8,032 | 14,238 | 7.6 | 12.6 | 203 | 466 | 26.8 (23.2 - 30.7) | 37.1 (33.8 - 40.6) |
| MDCD | 7,124 | 15,407 | 5.0 | 9.9 | 125 | 250 | 24.8 (20.7 - 29.6) | 25.2 (22.2 - 28.6) |
| Open Claims | 430,708 | 815,640 | 376.7 | 771.4 | 4,304 | 10,030 | 11.4 (11.1 - 11.8) | 13.0 (12.7 - 13.3) |
| OEHR | 38,193 | 81,585 | 19.7 | 45.8 | 241 | 751 | 12.3 (10.8 - 13.9) | 16.4 (15.2 - 17.6) |
| VA | 2,167 | 12,966 | 1.6 | 10.4 | <10 | 71 | < 6.30 (3.02 - 11.6) | 6.82 (5.33 - 8.60) |
| SIDIAP | 19,541 | 7,057 | 53.6 | 19.4 | 58 | 10 | 1.08 (0.82 - 1.40) | 0.52 (0.25 - 0.95) |
| Meta-analysis | 569,149 | 1,041,888 | 514.9 | 945.9 | 5,465 | 12,838 | 10.6 (10.3 - 10.9) | 13.6 (13.3 - 13.8) |
| Meta-analysis (w/o USOC) | 138,441 | 226,248 | 138.3 | 174.5 | 1,161 | 2,808 | 8.40 (7.92 - 8.89) | 16.1 (15.5 - 16.7) |
| **PS-matching / on-treatment TAR without escalation** | | | | | | | | |
| CCAE | 38,432 | 64,723 | 24.7 | 40.6 | 131 | 263 | 5.30 (4.43 - 6.28) | 6.47 (5.71 - 7.30) |
| DAG | 10,485 | 2,040 | 10.3 | 2.0 | 0 | 0 | | |
| ODOD | 25,936 | 56,484 | 17.1 | 43.1 | 369 | 1,108 | 21.6 (19.5 - 23.9) | 25.7 (24.2 - 27.3) |
| MDCR | 7,431 | 13,869 | 6.1 | 11.2 | 159 | 410 | 26.1 (22.2 - 30.4) | 36.5 (33.1 - 40.2) |
| MDCD | 6,798 | 15,306 | 3.7 | 8.2 | 86 | 201 | 23.2 (18.5 - 28.6) | 24.5 (21.2 - 28.1) |
| Open Claims | 420,971 | 815,544 | 290.7 | 649.4 | 3,313 | 8,426 | 11.4 (11.0 - 11.8) | 13.0 (12.7 - 13.3) |
| OEHR | 35,723 | 81,503 | 14.6 | 39.1 | 164 | 561 | 11.2 (9.55 - 13.1) | 14.4 (13.2 - 15.6) |
| VA | 1,778 | 9,366 | 1.1 | 6.2 | <10 | 35 | < 9.32 (4.47 - 17.1) | 5.61 (3.91 - 7.80) |
| SIDIAP | 19,369 | 5,425 | 39.8 | 11.5 | 37 | <5 | 0.93 (0.66 - 1.28) | < 0.43 (0.14 - 1.01) |
| Meta-analysis | 556,438 | 1,062,220 | 397.8 | 809.4 | <4,269 | <11,009 | < 10.7 (10.4 - 11.1) | < 13.6 (13.3 - 13.9) |
| Meta-analysis (w/o USOC) | 135,467 | 246,676 | 107.1 | 160.0 | <956 | <2,583 | < 8.92 (8.37 - 9.51) | < 16.1 (15.5 - 16.8) |
| **PS-stratification / on-treatment TAR without escalation** | | | | | | | | |
| CCAE | 44,755 | 66,729 | 29.1 | 41.7 | 148 | 270 | 5.08 (4.30 - 5.97) | 6.47 (5.72 - 7.29) |
| DAG | 10,516 | 2,367 | 10.3 | 2.3 | 0 | 0 | | |
| ODOD | 27,920 | 56,639 | 18.5 | 43.1 | 394 | 1,110 | 21.3 (19.3 - 23.6) | 25.7 (24.2 - 27.3) |
| MDCR | 8,032 | 14,238 | 6.6 | 11.5 | 175 | 424 | 26.6 (22.8 - 30.8) | 36.7 (33.3 - 40.4) |
| MDCD | 7,124 | 15,407 | 3.9 | 8.2 | 93 | 201 | 23.6 (19.1 - 28.9) | 24.4 (21.1 - 28.0) |
| Open Claims | 430,708 | 815,640 | 299.3 | 649.4 | 3,435 | 8,427 | 11.5 (11.1 - 11.9) | 13.0 (12.7 - 13.3) |
| OEHR | 38,193 | 81,585 | 15.9 | 39.1 | 171 | 561 | 10.8 (9.21 - 12.5) | 14.3 (13.2 - 15.6) |
| VA | 2,167 | 12,966 | 1.3 | 9.1 | <10 | 61 | < 7.91 (3.79 - 14.5) | 6.72 (5.14 - 8.63) |
| SIDIAP | 19,541 | 7,057 | 39.9 | 15.7 | 37 | 10 | 0.93 (0.65 - 1.28) | 0.64 (0.31 - 1.17) |
| Meta-analysis | 569,149 | 1,041,888 | 409.2 | 800.6 | 4,360 | 10,802 | 10.7 (10.3 - 11.0) | 13.5 (13.2 - 13.7) |
| Meta-analysis (w/o USOC) | 138,441 | 226,248 | 109.9 | 151.2 | 925 | 2,375 | 8.41 (7.88 - 8.97) | 15.7 (15.1 - 16.4) |

##### 4.6.7 Measured renal dysfunction

**Supplementary Table 422:** Exposures, outcomes and risk estimates for Measured renal dysfunction in DPP4I (female) (T) vs SU (female) (C) new-user cohorts under four different propensity score (PS) adjustment / time-at-risk (TAR) designs across data sources. We report total patient exposure time in 1,000 patient-year (KYr) units, uncalibrated and calibrated hazard ratios (HRs) and their 95% confidence intervals (CIs). Greyed out entries did not pass study diagnostics and are included only for completeness.

| | Patients | | Exposure (KYr) | | Outcomes | | Incidence (per KYr) | |
|---|---|---|---|---|---|---|---|---|
| | T | C | T | C | T | C | T | C |
| **PS-matching / on-treatment TAR** | | | | | | | | |
| CCAE | 39,095 | 66,019 | 30.6 | 48.2 | 5 | 16 | 0.16 (0.05 - 0.38) | 0.33 (0.19 - 0.54) |
| DAG | 10,485 | 2,040 | 11.4 | 2.1 | 0 | 0 | | |
| ODOD | 27,459 | 60,070 | 21.7 | 52.2 | 52 | 82 | 2.40 (1.79 - 3.14) | 1.57 (1.25 - 1.95) |
| MDCR | 8,176 | 15,519 | 7.7 | 13.9 | 0 | <5 | | < 0.36 (0.12 - 0.84) |
| MDCD | 7,427 | 16,757 | 5.3 | 11.0 | 0 | 0 | | |
| Open Claims | 439,873 | 852,269 | 384.3 | 811.8 | 0 | 0 | | |
| OEHR | 36,419 | 83,277 | 18.7 | 47.4 | 0 | 0 | | |
| VA | 1,805 | 9,455 | 1.4 | 7.3 | <10 | 20 | < 7.36 (3.53 - 13.5) | 2.75 (1.68 - 4.25) |
| SIDIAP | 19,439 | 5,430 | 53.4 | 14.2 | 141 | 16 | 2.64 (2.22 - 3.11) | 1.13 (0.64 - 1.83) |
| Meta-analysis | 46,898 | 65,500 | 75.1 | 66.3 | 193 | 98 | 2.57 (2.22 - 2.96) | 1.48 (1.20 - 1.80) |
| Meta-analysis (w/o USOC) | 46,898 | 65,500 | 75.1 | 66.3 | 193 | 98 | 2.57 (2.22 - 2.96) | 1.48 (1.20 - 1.80) |
| **PS-stratification / on-treatment TAR** | | | | | | | | |
| CCAE | 45,465 | 67,954 | 36.0 | 49.3 | 7 | 16 | 0.19 (0.08 - 0.40) | 0.32 (0.19 - 0.53) |
| DAG | 10,516 | 2,367 | 11.4 | 2.5 | 0 | 0 | | |
| ODOD | 29,464 | 60,228 | 23.4 | 52.2 | 55 | 82 | 2.35 (1.77 - 3.06) | 1.57 (1.25 - 1.95) |
| MDCR | 8,813 | 15,840 | 8.3 | 14.2 | 0 | <5 | | < 0.35 (0.11 - 0.82) |
| MDCD | 7,771 | 16,863 | 5.6 | 11.0 | 0 | 0 | | |
| Open Claims | 449,878 | 852,367 | 395.5 | 811.8 | 0 | 0 | | |
| OEHR | 38,897 | 83,366 | 20.2 | 47.5 | 0 | 0 | | |
| VA | 2,193 | 13,010 | 1.6 | 10.4 | <10 | 32 | < 6.27 (3.01 - 11.5) | 3.07 (2.10 - 4.33) |
| SIDIAP | 19,548 | 7,062 | 53.5 | 19.4 | 143 | 19 | 2.67 (2.25 - 3.15) | 0.98 (0.59 - 1.53) |
| Meta-analysis | 94,477 | 135,244 | 112.9 | 120.9 | 205 | 117 | 1.82 (1.58 - 2.08) | 0.97 (0.80 - 1.16) |
| Meta-analysis (w/o USOC) | 94,477 | 135,244 | 112.9 | 120.9 | 205 | 117 | 1.82 (1.58 - 2.08) | 0.97 (0.80 - 1.16) |
| **PS-matching / on-treatment TAR without escalation** | | | | | | | | |
| CCAE | 39,095 | 66,019 | 25.2 | 41.5 | 5 | 13 | 0.20 (0.06 - 0.46) | 0.31 (0.17 - 0.54) |
| DAG | 10,485 | 2,040 | 10.3 | 2.0 | 0 | 0 | | |
| ODOD | 27,459 | 60,070 | 18.2 | 46.4 | 44 | 70 | 2.42 (1.76 - 3.25) | 1.51 (1.18 - 1.91) |
| MDCR | 8,176 | 15,519 | 6.7 | 12.7 | 0 | <5 | | < 0.39 (0.13 - 0.92) |
| MDCD | 7,427 | 16,757 | 4.1 | 9.1 | 0 | 0 | | |
| Open Claims | 439,873 | 852,269 | 305.0 | 682.7 | 0 | 0 | | |
| OEHR | 36,419 | 83,277 | 15.0 | 40.2 | 0 | 0 | | |
| VA | 1,805 | 9,455 | 1.1 | 6.3 | <10 | 19 | < 9.24 (4.43 - 17.0) | 3.02 (1.82 - 4.71) |
| SIDIAP | 19,439 | 5,430 | 39.7 | 11.6 | 108 | 14 | 2.72 (2.23 - 3.28) | 1.21 (0.66 - 2.03) |
| Meta-analysis | 46,898 | 65,500 | 57.9 | 57.9 | 152 | 84 | 2.62 (2.22 - 3.08) | 1.45 (1.16 - 1.80) |
| Meta-analysis (w/o USOC) | 46,898 | 65,500 | 57.9 | 57.9 | 152 | 84 | 2.62 (2.22 - 3.08) | 1.45 (1.16 - 1.80) |
| **PS-stratification / on-treatment TAR without escalation** | | | | | | | | |
| CCAE | 45,465 | 67,954 | 29.6 | 42.5 | 6 | 13 | 0.20 (0.07 - 0.44) | 0.31 (0.16 - 0.52) |
| DAG | 10,516 | 2,367 | 10.3 | 2.3 | 0 | 0 | | |
| ODOD | 29,464 | 60,228 | 19.6 | 46.4 | 46 | 70 | 2.34 (1.72 - 3.13) | 1.51 (1.18 - 1.91) |
| MDCR | 8,813 | 15,840 | 7.2 | 13.0 | 0 | <5 | | < 0.39 (0.13 - 0.90) |
| MDCD | 7,771 | 16,863 | 4.3 | 9.2 | 0 | 0 | | |
| Open Claims | 449,878 | 852,367 | 314.0 | 682.7 | 0 | 0 | | |
| OEHR | 38,897 | 83,366 | 16.3 | 40.3 | 0 | 0 | | |
| VA | 2,193 | 13,010 | 1.3 | 9.1 | <10 | 29 | < 7.87 (3.77 - 14.5) | 3.19 (2.14 - 4.59) |
| SIDIAP | 19,548 | 7,062 | 39.8 | 15.7 | 110 | 17 | 2.76 (2.27 - 3.33) | 1.08 (0.63 - 1.74) |
| Meta-analysis | 94,477 | 135,244 | 89.0 | 104.6 | 162 | 100 | 1.82 (1.55 - 2.12) | 0.96 (0.78 - 1.16) |
| Meta-analysis (w/o USOC) | 94,477 | 135,244 | 89.0 | 104.6 | 162 | 100 | 1.82 (1.55 - 2.12) | 0.96 (0.78 - 1.16) |

##### 4.6.8 Stroke

**Supplementary Table 423:** Exposures, outcomes and risk estimates for Stroke in DPP4I (female) (T) vs SU (female) (C) new-user cohorts under four different propensity score (PS) adjustment / time-at-risk (TAR) designs across data sources. We report total patient exposure time in 1,000 patient-year (KYr) units, uncalibrated and calibrated hazard ratios (HRs) and their 95% confidence intervals (CIs). Greyed out entries did not pass study diagnostics and are included only for completeness.

| | Patients | | Exposure (KYr) | | Outcomes | | Incidence (per KYr) | |
|---|---|---|---|---|---|---|---|---|
| | T | C | T | C | T | C | T | C |
| **PS-matching / on-treatment TAR** | | | | | | | | |
| CCAE | 38,640 | 65,140 | 30.3 | 47.5 | 108 | 178 | 3.56 (2.92 - 4.30) | 3.75 (3.22 - 4.34) |
| DAG | 10,485 | 2,040 | 11.4 | 2.1 | 0 | 0 | | |
| ODOD | 26,684 | 58,327 | 21.1 | 50.3 | 244 | 633 | 11.6 (10.2 - 13.1) | 12.6 (11.6 - 13.6) |
| MDCR | 7,786 | 14,724 | 7.3 | 13.1 | 111 | 264 | 15.2 (12.5 - 18.3) | 20.2 (17.9 - 22.8) |
| MDCD | 7,135 | 16,076 | 5.0 | 10.5 | 64 | 129 | 12.7 (9.79 - 16.2) | 12.3 (10.2 - 14.6) |
| Open Claims | 426,881 | 827,934 | 372.1 | 786.3 | 2,257 | 5,349 | 6.07 (5.82 - 6.32) | 6.80 (6.62 - 6.99) |
| OEHR | 36,097 | 82,419 | 18.5 | 46.7 | 82 | 295 | 4.44 (3.53 - 5.52) | 6.31 (5.61 - 7.08) |
| VA | 1,793 | 9,562 | 1.4 | 7.4 | <10 | 16 | < 7.37 (3.53 - 13.6) | 2.16 (1.24 - 3.51) |
| SIDIAP | 19,363 | 5,414 | 53.3 | 14.1 | 112 | 23 | 2.10 (1.73 - 2.53) | 1.63 (1.03 - 2.44) |
| Meta-analysis | 562,586 | 1,070,034 | 507.5 | 968.5 | 2,978 | 6,871 | 5.87 (5.66 - 6.08) | 7.09 (6.93 - 7.26) |
| Meta-analysis (w/o USOC) | 135,705 | 242,100 | 135.4 | 182.2 | 721 | 1,522 | 5.32 (4.94 - 5.73) | 8.35 (7.94 - 8.78) |
| **PS-stratification / on-treatment TAR** | | | | | | | | |
| CCAE | 44,968 | 67,059 | 35.6 | 48.6 | 123 | 181 | 3.46 (2.87 - 4.12) | 3.72 (3.20 - 4.31) |
| DAG | 10,516 | 2,367 | 11.4 | 2.5 | 0 | 0 | | |
| ODOD | 28,658 | 58,485 | 22.7 | 50.3 | 254 | 633 | 11.2 (9.85 - 12.6) | 12.6 (11.6 - 13.6) |
| MDCR | 8,390 | 15,031 | 7.9 | 13.3 | 120 | 268 | 15.2 (12.6 - 18.2) | 20.1 (17.8 - 22.7) |
| MDCD | 7,475 | 16,180 | 5.3 | 10.5 | 69 | 129 | 13.0 (10.1 - 16.4) | 12.2 (10.2 - 14.5) |
| Open Claims | 436,812 | 828,032 | 383.1 | 786.3 | 2,350 | 5,350 | 6.13 (5.89 - 6.39) | 6.80 (6.62 - 6.99) |
| OEHR | 38,563 | 82,508 | 20.0 | 46.8 | 86 | 295 | 4.31 (3.44 - 5.32) | 6.30 (5.61 - 7.07) |
| VA | 2,180 | 13,066 | 1.6 | 10.5 | 10 | 25 | 6.27 (3.01 - 11.5) | 2.38 (1.54 - 3.51) |
| SIDIAP | 19,471 | 7,043 | 53.3 | 19.3 | 112 | 30 | 2.10 (1.73 - 2.53) | 1.55 (1.05 - 2.22) |
| Meta-analysis | 576,862 | 1,058,158 | 522.7 | 964.7 | 3,045 | 6,757 | 5.83 (5.62 - 6.04) | 7.00 (6.84 - 7.17) |
| Meta-analysis (w/o USOC) | 140,050 | 230,126 | 139.5 | 178.4 | 695 | 1,407 | 4.98 (4.62 - 5.37) | 7.89 (7.48 - 8.31) |
| **PS-matching / on-treatment TAR without escalation** | | | | | | | | |
| CCAE | 38,640 | 65,140 | 24.9 | 40.9 | 89 | 151 | 3.57 (2.87 - 4.40) | 3.69 (3.13 - 4.33) |
| DAG | 10,485 | 2,040 | 10.3 | 2.0 | 0 | 0 | | |
| ODOD | 26,684 | 58,327 | 17.7 | 44.7 | 217 | 564 | 12.3 (10.7 - 14.0) | 12.6 (11.6 - 13.7) |
| MDCR | 7,786 | 14,724 | 6.4 | 12.0 | 89 | 238 | 14.0 (11.2 - 17.2) | 19.9 (17.4 - 22.6) |
| MDCD | 7,135 | 16,076 | 3.9 | 8.7 | 52 | 107 | 13.2 (9.88 - 17.3) | 12.3 (10.0 - 14.8) |
| Open Claims | 426,881 | 827,934 | 295.3 | 661.6 | 1,827 | 4,532 | 6.19 (5.91 - 6.48) | 6.85 (6.65 - 7.05) |
| OEHR | 36,097 | 82,419 | 14.8 | 39.7 | 57 | 223 | 3.84 (2.91 - 4.98) | 5.62 (4.91 - 6.41) |
| VA | 1,793 | 9,562 | 1.1 | 6.4 | <10 | 11 | < 9.25 (4.44 - 17.0) | 1.72 (0.86 - 3.08) |
| SIDIAP | 19,363 | 5,414 | 39.6 | 11.5 | 90 | 20 | 2.27 (1.83 - 2.80) | 1.74 (1.06 - 2.69) |
| Meta-analysis | 562,586 | 1,070,034 | 402.7 | 819.1 | 2,421 | 5,835 | 6.01 (5.78 - 6.26) | 7.12 (6.94 - 7.31) |
| Meta-analysis (w/o USOC) | 135,705 | 242,100 | 107.3 | 157.5 | 594 | 1,303 | 5.53 (5.10 - 6.00) | 8.27 (7.83 - 8.73) |
| **PS-stratification / on-treatment TAR without escalation** | | | | | | | | |
| CCAE | 44,968 | 67,059 | 29.3 | 41.9 | 103 | 153 | 3.52 (2.87 - 4.27) | 3.65 (3.09 - 4.27) |
| DAG | 10,516 | 2,367 | 10.3 | 2.3 | 0 | 0 | | |
| ODOD | 28,658 | 58,485 | 19.1 | 44.8 | 227 | 564 | 11.9 (10.4 - 13.6) | 12.6 (11.6 - 13.7) |
| MDCR | 8,390 | 15,031 | 6.9 | 12.2 | 96 | 242 | 13.9 (11.3 - 17.0) | 19.8 (17.4 - 22.4) |
| MDCD | 7,475 | 16,180 | 4.2 | 8.8 | 57 | 107 | 13.7 (10.4 - 17.8) | 12.2 (10.0 - 14.8) |
| Open Claims | 436,812 | 828,032 | 304.2 | 661.6 | 1,900 | 4,533 | 6.25 (5.97 - 6.53) | 6.85 (6.65 - 7.05) |
| OEHR | 38,563 | 82,508 | 16.1 | 39.7 | 60 | 223 | 3.73 (2.85 - 4.80) | 5.61 (4.90 - 6.40) |
| VA | 2,180 | 13,066 | 1.3 | 9.2 | <10 | 19 | < 7.86 (3.77 - 14.4) | 2.08 (1.25 - 3.24) |
| SIDIAP | 19,471 | 7,043 | 39.7 | 15.6 | 90 | 26 | 2.27 (1.82 - 2.79) | 1.67 (1.09 - 2.44) |
| Meta-analysis | 576,862 | 1,058,158 | 415.2 | 815.9 | 2,476 | 5,741 | 5.96 (5.73 - 6.20) | 7.04 (6.86 - 7.22) |
| Meta-analysis (w/o USOC) | 140,050 | 230,126 | 111.0 | 154.3 | 576 | 1,208 | 5.19 (4.78 - 5.63) | 7.83 (7.39 - 8.28) |

##### 4.6.9 Sudden cardiac death

**Supplementary Table 424:** Exposures, outcomes and risk estimates for Sudden cardiac death in DPP4I (female) (T) vs SU (female) (C) new-user cohorts under four different propensity score (PS) adjustment / time-at-risk (TAR) designs across data sources. We report total patient exposure time in 1,000 patient-year (KYr) units, uncalibrated and calibrated hazard ratios (HRs) and their 95% confidence intervals (CIs). Greyed out entries did not pass study diagnostics and are included only for completeness.

| | Patients | | Exposure (KYr) | | Outcomes | | Incidence (per KYr) | |
|---|---|---|---|---|---|---|---|---|
| | T | C | T | C | T | C | T | C |
| **PS-matching / on-treatment TAR** | | | | | | | | |
| CCAE | 39,112 | 66,040 | 30.7 | 48.2 | 11 | 48 | 0.36 (0.18 - 0.64) | 1.00 (0.73 - 1.32) |
| DAG | 10,485 | 2,040 | 11.4 | 2.1 | 0 | 0 | | |
| ODOD | 27,462 | 60,125 | 21.8 | 52.2 | 61 | 163 | 2.80 (2.14 - 3.60) | 3.12 (2.66 - 3.64) |
| MDCR | 8,154 | 15,491 | 7.7 | 13.9 | 14 | 44 | 1.82 (1.00 - 3.06) | 3.18 (2.31 - 4.26) |
| MDCD | 7,412 | 16,700 | 5.3 | 11.0 | 19 | 35 | 3.62 (2.18 - 5.65) | 3.19 (2.22 - 4.43) |
| Open Claims | 439,226 | 850,867 | 383.6 | 810.3 | 462 | 1,077 | 1.20 (1.10 - 1.32) | 1.33 (1.25 - 1.41) |
| OEHR | 36,404 | 83,240 | 18.7 | 47.4 | 20 | 81 | 1.07 (0.65 - 1.65) | 1.71 (1.36 - 2.12) |
| VA | 1,820 | 9,662 | 1.4 | 7.5 | 0 | <10 | | < 1.34 (0.64 - 2.46) |
| SIDIAP | 19,513 | 5,440 | 53.7 | 14.2 | 0 | 0 | | |
| Meta-analysis | 557,770 | 1,092,463 | 467.6 | 983.0 | 587 | 1,448 | 1.26 (1.16 - 1.36) | 1.47 (1.40 - 1.55) |
| Meta-analysis (w/o USOC) | 118,544 | 241,596 | 84.0 | 172.7 | 125 | 371 | 1.49 (1.24 - 1.77) | 2.15 (1.94 - 2.38) |
| **PS-stratification / on-treatment TAR** | | | | | | | | |
| CCAE | 45,480 | 67,974 | 36.0 | 49.4 | 16 | 49 | 0.44 (0.25 - 0.72) | 0.99 (0.73 - 1.31) |
| DAG | 10,516 | 2,367 | 11.4 | 2.5 | 0 | 0 | | |
| ODOD | 29,466 | 60,283 | 23.5 | 52.3 | 66 | 163 | 2.81 (2.18 - 3.58) | 3.12 (2.66 - 3.64) |
| MDCR | 8,792 | 15,812 | 8.3 | 14.1 | 14 | 47 | 1.69 (0.92 - 2.83) | 3.32 (2.44 - 4.42) |
| MDCD | 7,756 | 16,806 | 5.5 | 11.0 | 20 | 35 | 3.61 (2.21 - 5.58) | 3.18 (2.21 - 4.42) |
| Open Claims | 449,195 | 850,965 | 394.8 | 810.4 | 479 | 1,077 | 1.21 (1.11 - 1.33) | 1.33 (1.25 - 1.41) |
| OEHR | 38,881 | 83,329 | 20.2 | 47.5 | 20 | 81 | 0.99 (0.60 - 1.53) | 1.71 (1.36 - 2.12) |
| VA | 2,220 | 13,214 | 1.6 | 10.7 | 0 | <10 | | < 0.94 (0.45 - 1.73) |
| SIDIAP | 19,626 | 7,073 | 53.8 | 19.4 | 0 | 0 | | |
| Meta-analysis | 571,814 | 1,078,363 | 482.7 | 973.6 | 595 | 1,417 | 1.23 (1.14 - 1.34) | 1.46 (1.38 - 1.53) |
| Meta-analysis (w/o USOC) | 122,619 | 227,398 | 88.0 | 163.2 | 116 | 340 | 1.32 (1.09 - 1.58) | 2.08 (1.87 - 2.32) |
| **PS-matching / on-treatment TAR without escalation** | | | | | | | | |
| CCAE | 39,112 | 66,040 | 25.2 | 41.5 | 8 | 35 | 0.32 (0.14 - 0.63) | 0.84 (0.59 - 1.17) |
| DAG | 10,485 | 2,040 | 10.3 | 2.0 | 0 | 0 | | |
| ODOD | 27,462 | 60,125 | 18.3 | 46.5 | 49 | 141 | 2.68 (1.99 - 3.55) | 3.04 (2.56 - 3.58) |
| MDCR | 8,154 | 15,491 | 6.7 | 12.7 | 10 | 41 | 1.50 (0.72 - 2.76) | 3.23 (2.32 - 4.39) |
| MDCD | 7,412 | 16,700 | 4.1 | 9.1 | 11 | 31 | 2.69 (1.34 - 4.81) | 3.40 (2.31 - 4.83) |
| Open Claims | 439,226 | 850,867 | 304.5 | 681.5 | 376 | 904 | 1.23 (1.11 - 1.37) | 1.33 (1.24 - 1.42) |
| OEHR | 36,404 | 83,240 | 15.0 | 40.2 | 11 | 49 | 0.73 (0.37 - 1.31) | 1.22 (0.90 - 1.61) |
| VA | 1,820 | 9,662 | 1.1 | 6.5 | 0 | <10 | | < 1.55 (0.74 - 2.84) |
| SIDIAP | 19,513 | 5,440 | 40.0 | 11.6 | 0 | 0 | | |
| Meta-analysis | 557,770 | 1,092,463 | 373.7 | 831.4 | 465 | 1,201 | 1.24 (1.13 - 1.36) | 1.44 (1.36 - 1.53) |
| Meta-analysis (w/o USOC) | 118,544 | 241,596 | 69.2 | 149.9 | 89 | 297 | 1.29 (1.03 - 1.58) | 1.98 (1.76 - 2.22) |
| **PS-stratification / on-treatment TAR without escalation** | | | | | | | | |
| CCAE | 45,480 | 67,974 | 29.6 | 42.6 | 11 | 36 | 0.37 (0.19 - 0.67) | 0.85 (0.59 - 1.17) |
| DAG | 10,516 | 2,367 | 10.3 | 2.3 | 0 | 0 | | |
| ODOD | 29,466 | 60,283 | 19.7 | 46.5 | 53 | 141 | 2.69 (2.02 - 3.52) | 3.03 (2.55 - 3.58) |
| MDCR | 8,792 | 15,812 | 7.2 | 12.9 | 10 | 43 | 1.39 (0.67 - 2.55) | 3.32 (2.40 - 4.47) |
| MDCD | 7,756 | 16,806 | 4.3 | 9.1 | 12 | 31 | 2.78 (1.44 - 4.86) | 3.39 (2.30 - 4.81) |
| Open Claims | 449,195 | 850,965 | 313.4 | 681.5 | 389 | 904 | 1.24 (1.12 - 1.37) | 1.33 (1.24 - 1.42) |
| OEHR | 38,881 | 83,329 | 16.3 | 40.2 | 11 | 49 | 0.68 (0.34 - 1.21) | 1.22 (0.90 - 1.61) |
| VA | 2,220 | 13,214 | 1.3 | 9.3 | 0 | <10 | | < 1.08 (0.52 - 1.98) |
| SIDIAP | 19,626 | 7,073 | 40.0 | 15.7 | 0 | 0 | | |
| Meta-analysis | 571,814 | 1,078,363 | 386.1 | 823.7 | 474 | 1,173 | 1.23 (1.12 - 1.34) | 1.42 (1.34 - 1.51) |
| Meta-analysis (w/o USOC) | 122,619 | 227,398 | 72.7 | 142.2 | 85 | 269 | 1.17 (0.93 - 1.45) | 1.89 (1.67 - 2.13) |

##### 4.6.10 Abnormal weight gain

**Supplementary Table 425:** Exposures, outcomes and risk estimates for Abnormal weight gain in DPP4I (female) (T) vs SU (female) (C) new-user cohorts under four different propensity score (PS) adjustment / time-at-risk (TAR) designs across data sources. We report total patient exposure time in 1,000 patient-year (KYr) units, uncalibrated and calibrated hazard ratios (HRs) and their 95% confidence intervals (CIs). Greyed out entries did not pass study diagnostics and are included only for completeness.

| | Patients | | Exposure (KYr) | | Outcomes | | Incidence (per KYr) | |
|---|---|---|---|---|---|---|---|---|
| | T | C | T | C | T | C | T | C |
| **PS-matching / on-treatment TAR** | | | | | | | | |
| CCAE | 36,796 | 62,846 | 28.8 | 45.8 | 205 | 377 | 7.12 (6.18 - 8.16) | 8.24 (7.43 - 9.11) |
| DAG | 10,380 | 2,020 | 11.2 | 2.1 | 15 | <5 | 1.34 (0.75 - 2.20) | < 2.36 (0.77 - 5.51) |
| ODOD | 26,061 | 57,645 | 20.6 | 49.8 | 157 | 452 | 7.63 (6.48 - 8.92) | 9.07 (8.25 - 9.95) |
| MDCR | 8,073 | 15,327 | 7.6 | 13.7 | 24 | 54 | 3.16 (2.03 - 4.71) | 3.95 (2.96 - 5.15) |
| MDCD | 6,776 | 15,565 | 4.8 | 10.2 | 44 | 130 | 9.24 (6.72 - 12.4) | 12.7 (10.6 - 15.1) |
| Open Claims | 416,823 | 813,408 | 364.2 | 775.1 | 1,087 | 2,456 | 2.98 (2.81 - 3.17) | 3.17 (3.04 - 3.30) |
| OEHR | 34,545 | 79,985 | 17.6 | 45.1 | 153 | 432 | 8.69 (7.37 - 10.2) | 9.57 (8.69 - 10.5) |
| VA | 1,733 | 9,121 | 1.3 | 7.0 | <10 | 22 | < 7.72 (3.70 - 14.2) | 3.13 (1.96 - 4.74) |
| SIDIAP | 19,342 | 5,409 | 53.2 | 14.1 | 63 | 18 | 1.18 (0.91 - 1.51) | 1.28 (0.76 - 2.02) |
| Meta-analysis | 550,149 | 1,059,306 | 498.0 | 960.9 | <1,743 | 3,941 | < 3.50 (3.34 - 3.67) | 4.10 (3.97 - 4.23) |
| Meta-analysis (w/o USOC) | 133,326 | 245,898 | 133.8 | 185.8 | <656 | 1,485 | < 4.90 (4.53 - 5.29) | 7.99 (7.59 - 8.41) |
| **PS-stratification / on-treatment TAR** | | | | | | | | |
| CCAE | 42,511 | 64,683 | 33.6 | 46.9 | 255 | 380 | 7.58 (6.68 - 8.57) | 8.10 (7.31 - 8.96) |
| DAG | 10,411 | 2,349 | 11.3 | 2.5 | 15 | 5 | 1.33 (0.75 - 2.20) | 2.00 (0.65 - 4.66) |
| ODOD | 27,829 | 57,783 | 22.1 | 49.9 | 179 | 452 | 8.11 (6.96 - 9.39) | 9.06 (8.25 - 9.94) |
| MDCR | 8,672 | 15,644 | 8.2 | 14.0 | 26 | 55 | 3.19 (2.08 - 4.67) | 3.94 (2.97 - 5.12) |
| MDCD | 7,067 | 15,658 | 5.0 | 10.2 | 50 | 130 | 10.00 (7.42 - 13.2) | 12.7 (10.6 - 15.1) |
| Open Claims | 425,868 | 813,494 | 374.5 | 775.2 | 1,130 | 2,456 | 3.02 (2.84 - 3.20) | 3.17 (3.04 - 3.30) |
| OEHR | 36,753 | 80,070 | 18.9 | 45.2 | 168 | 432 | 8.88 (7.59 - 10.3) | 9.56 (8.68 - 10.5) |
| VA | 2,099 | 12,636 | 1.5 | 10.1 | <10 | 38 | < 6.60 (3.17 - 12.1) | 3.75 (2.66 - 5.15) |
| SIDIAP | 19,454 | 7,041 | 53.3 | 19.3 | 63 | 24 | 1.18 (0.91 - 1.51) | 1.24 (0.80 - 1.85) |
| Meta-analysis | 561,087 | 1,038,715 | 510.6 | 950.3 | 1,821 | 3,799 | 3.57 (3.40 - 3.73) | 4.00 (3.87 - 4.13) |
| Meta-analysis (w/o USOC) | 135,219 | 225,221 | 136.1 | 175.2 | 691 | 1,343 | 5.08 (4.71 - 5.47) | 7.67 (7.26 - 8.09) |
| **PS-matching / on-treatment TAR without escalation** | | | | | | | | |
| CCAE | 36,796 | 62,846 | 23.7 | 39.5 | 163 | 310 | 6.88 (5.87 - 8.02) | 7.85 (7.00 - 8.77) |
| DAG | 10,380 | 2,020 | 10.1 | 2.0 | 12 | <5 | 1.18 (0.61 - 2.07) | < 2.56 (0.83 - 5.97) |
| ODOD | 26,061 | 57,645 | 17.3 | 44.5 | 124 | 386 | 7.17 (5.96 - 8.55) | 8.68 (7.84 - 9.59) |
| MDCR | 8,073 | 15,327 | 6.6 | 12.5 | 18 | 47 | 2.73 (1.62 - 4.31) | 3.75 (2.75 - 4.98) |
| MDCD | 6,776 | 15,565 | 3.7 | 8.5 | 30 | 110 | 8.10 (5.46 - 11.6) | 12.9 (10.6 - 15.6) |
| Open Claims | 416,823 | 813,408 | 289.7 | 653.4 | 829 | 2,028 | 2.86 (2.67 - 3.06) | 3.10 (2.97 - 3.24) |
| OEHR | 34,545 | 79,985 | 14.2 | 38.4 | 106 | 337 | 7.48 (6.12 - 9.04) | 8.78 (7.86 - 9.77) |
| VA | 1,733 | 9,121 | 1.0 | 6.1 | <10 | 17 | < 9.71 (4.66 - 17.9) | 2.79 (1.63 - 4.47) |
| SIDIAP | 19,342 | 5,409 | 39.6 | 11.5 | 49 | 18 | 1.24 (0.92 - 1.64) | 1.57 (0.93 - 2.47) |
| Meta-analysis | 548,416 | 1,050,185 | 394.7 | 808.3 | 1,319 | 3,236 | 3.34 (3.16 - 3.53) | 4.00 (3.87 - 4.14) |
| Meta-analysis (w/o USOC) | 131,593 | 236,777 | 105.0 | 154.9 | 490 | 1,208 | 4.67 (4.26 - 5.10) | 7.80 (7.37 - 8.25) |
| **PS-stratification / on-treatment TAR without escalation** | | | | | | | | |
| CCAE | 42,511 | 64,683 | 27.7 | 40.5 | 202 | 313 | 7.30 (6.33 - 8.38) | 7.72 (6.89 - 8.63) |
| DAG | 10,411 | 2,349 | 10.2 | 2.3 | 12 | 5 | 1.18 (0.61 - 2.06) | 2.18 (0.71 - 5.08) |
| ODOD | 27,829 | 57,783 | 18.6 | 44.5 | 145 | 386 | 7.81 (6.59 - 9.19) | 8.68 (7.83 - 9.59) |
| MDCR | 8,672 | 15,644 | 7.1 | 12.8 | 20 | 48 | 2.82 (1.72 - 4.35) | 3.75 (2.76 - 4.97) |
| MDCD | 7,067 | 15,658 | 3.9 | 8.5 | 36 | 110 | 9.27 (6.49 - 12.8) | 12.9 (10.6 - 15.6) |
| Open Claims | 425,868 | 813,494 | 297.8 | 653.4 | 860 | 2,028 | 2.89 (2.70 - 3.09) | 3.10 (2.97 - 3.24) |
| OEHR | 36,753 | 80,070 | 15.3 | 38.4 | 115 | 337 | 7.54 (6.22 - 9.05) | 8.77 (7.85 - 9.75) |
| VA | 2,099 | 12,636 | 1.2 | 8.8 | <10 | 30 | < 8.29 (3.97 - 15.2) | 3.40 (2.29 - 4.85) |
| SIDIAP | 19,454 | 7,041 | 39.7 | 15.6 | 49 | 22 | 1.24 (0.91 - 1.63) | 1.41 (0.88 - 2.13) |
| Meta-analysis | 561,087 | 1,038,715 | 406.1 | 805.3 | 1,391 | 3,134 | 3.43 (3.25 - 3.61) | 3.89 (3.76 - 4.03) |
| Meta-analysis (w/o USOC) | 135,219 | 225,221 | 108.2 | 151.9 | 531 | 1,106 | 4.91 (4.50 - 5.34) | 7.28 (6.86 - 7.72) |

##### 4.6.11 Abnormal weight loss

**Supplementary Table 426:** Exposures, outcomes and risk estimates for Abnormal weight loss in DPP4I (female) (T) vs SU (female) (C) new-user cohorts under four different propensity score (PS) adjustment / time-at-risk (TAR) designs across data sources. We report total patient exposure time in 1,000 patient-year (KYr) units, uncalibrated and calibrated hazard ratios (HRs) and their 95% confidence intervals (CIs). Greyed out entries did not pass study diagnostics and are included only for completeness.

| | Patients | | Exposure (KYr) | | Outcomes | | Incidence (per KYr) | |
|---|---|---|---|---|---|---|---|---|
| | T | C | T | C | T | C | T | C |
| **PS-matching / on-treatment TAR** | | | | | | | | |
| CCAE | 38,258 | 64,600 | 29.8 | 47.1 | 209 | 251 | 7.00 (6.09 - 8.02) | 5.33 (4.69 - 6.03) |
| DAG | 10,197 | 1,981 | 11.0 | 2.1 | 79 | 15 | 7.21 (5.71 - 8.98) | 7.28 (4.07 - 12.0) |
| ODOD | 26,218 | 57,313 | 20.6 | 49.3 | 291 | 704 | 14.1 (12.5 - 15.8) | 14.3 (13.3 - 15.4) |
| MDCR | 7,809 | 14,790 | 7.3 | 13.1 | 105 | 167 | 14.4 (11.7 - 17.4) | 12.7 (10.9 - 14.8) |
| MDCD | 7,062 | 15,969 | 4.9 | 10.4 | 82 | 167 | 16.6 (13.2 - 20.6) | 16.1 (13.8 - 18.8) |
| Open Claims | 422,034 | 821,394 | 366.6 | 779.9 | 2,548 | 4,151 | 6.95 (6.68 - 7.23) | 5.32 (5.16 - 5.49) |
| OEHR | 35,345 | 80,877 | 18.0 | 45.6 | 204 | 525 | 11.4 (9.85 - 13.0) | 11.5 (10.5 - 12.5) |
| VA | 1,758 | 9,356 | 1.3 | 7.2 | <10 | 37 | < 7.51 (3.60 - 13.8) | 5.12 (3.61 - 7.06) |
| SIDIAP | 19,270 | 5,415 | 53.0 | 14.1 | 184 | 34 | 3.47 (2.99 - 4.01) | 2.41 (1.67 - 3.37) |
| Meta-analysis | 567,951 | 1,071,695 | 512.5 | 968.7 | <3,712 | 6,051 | < 7.24 (7.01 - 7.48) | 6.25 (6.09 - 6.41) |
| Meta-analysis (w/o USOC) | 145,917 | 250,301 | 145.9 | 188.8 | <1,164 | 1,900 | < 7.98 (7.53 - 8.45) | 10.1 (9.61 - 10.5) |
| **PS-stratification / on-treatment TAR** | | | | | | | | |
| CCAE | 44,501 | 66,506 | 35.0 | 48.2 | 243 | 255 | 6.93 (6.09 - 7.86) | 5.29 (4.66 - 5.98) |
| DAG | 10,227 | 2,299 | 11.0 | 2.4 | 79 | 17 | 7.19 (5.70 - 8.97) | 7.01 (4.08 - 11.2) |
| ODOD | 28,127 | 57,467 | 22.2 | 49.3 | 328 | 704 | 14.8 (13.2 - 16.5) | 14.3 (13.2 - 15.4) |
| MDCR | 8,411 | 15,103 | 7.9 | 13.4 | 120 | 167 | 15.2 (12.6 - 18.2) | 12.5 (10.6 - 14.5) |
| MDCD | 7,383 | 16,069 | 5.2 | 10.4 | 88 | 167 | 16.9 (13.6 - 20.8) | 16.1 (13.7 - 18.7) |
| Open Claims | 431,527 | 821,490 | 377.1 | 779.9 | 2,654 | 4,151 | 7.04 (6.77 - 7.31) | 5.32 (5.16 - 5.49) |
| OEHR | 37,764 | 80,961 | 19.4 | 45.7 | 223 | 527 | 11.5 (10.0 - 13.1) | 11.5 (10.6 - 12.6) |
| VA | 2,135 | 12,725 | 1.6 | 10.2 | 12 | 55 | 7.71 (3.98 - 13.5) | 5.41 (4.07 - 7.04) |
| SIDIAP | 19,387 | 7,046 | 53.0 | 19.3 | 186 | 44 | 3.51 (3.02 - 4.05) | 2.28 (1.66 - 3.06) |
| Meta-analysis | 579,944 | 1,050,872 | 525.7 | 958.2 | 3,833 | 5,865 | 7.29 (7.06 - 7.53) | 6.12 (5.97 - 6.28) |
| Meta-analysis (w/o USOC) | 148,417 | 229,382 | 148.6 | 178.3 | 1,179 | 1,714 | 7.93 (7.49 - 8.40) | 9.61 (9.16 - 10.1) |
| **PS-matching / on-treatment TAR without escalation** | | | | | | | | |
| CCAE | 38,258 | 64,600 | 24.6 | 40.6 | 178 | 209 | 7.25 (6.22 - 8.40) | 5.15 (4.48 - 5.90) |
| DAG | 10,197 | 1,981 | 9.9 | 1.9 | 69 | 13 | 6.97 (5.42 - 8.82) | 6.83 (3.64 - 11.7) |
| ODOD | 26,218 | 57,313 | 17.3 | 43.8 | 241 | 628 | 13.9 (12.2 - 15.8) | 14.3 (13.2 - 15.5) |
| MDCR | 7,809 | 14,790 | 6.3 | 12.0 | 98 | 153 | 15.4 (12.5 - 18.8) | 12.7 (10.8 - 14.9) |
| MDCD | 7,062 | 15,969 | 3.9 | 8.6 | 58 | 146 | 15.0 (11.4 - 19.4) | 16.9 (14.3 - 19.9) |
| Open Claims | 422,034 | 821,394 | 290.9 | 656.3 | 2,071 | 3,496 | 7.12 (6.82 - 7.43) | 5.33 (5.15 - 5.51) |
| OEHR | 35,345 | 80,877 | 14.5 | 38.7 | 160 | 412 | 11.1 (9.42 - 12.9) | 10.6 (9.63 - 11.7) |
| VA | 1,758 | 9,356 | 1.1 | 6.3 | <10 | 32 | < 9.47 (4.54 - 17.4) | 5.12 (3.50 - 7.22) |
| SIDIAP | 19,270 | 5,415 | 39.4 | 11.5 | 143 | 28 | 3.63 (3.06 - 4.28) | 2.44 (1.62 - 3.52) |
| Meta-analysis | 567,951 | 1,071,695 | 407.8 | 819.7 | <3,028 | 5,117 | < 7.43 (7.16 - 7.69) | 6.24 (6.07 - 6.42) |
| Meta-analysis (w/o USOC) | 145,917 | 250,301 | 116.9 | 163.4 | <957 | 1,621 | < 8.19 (7.68 - 8.73) | 9.92 (9.44 - 10.4) |
| **PS-stratification / on-treatment TAR without escalation** | | | | | | | | |
| CCAE | 44,501 | 66,506 | 28.8 | 41.6 | 208 | 213 | 7.21 (6.26 - 8.26) | 5.12 (4.46 - 5.86) |
| DAG | 10,227 | 2,299 | 9.9 | 2.2 | 69 | 15 | 6.96 (5.41 - 8.80) | 6.72 (3.76 - 11.1) |
| ODOD | 28,127 | 57,467 | 18.6 | 43.9 | 275 | 628 | 14.8 (13.1 - 16.6) | 14.3 (13.2 - 15.5) |
| MDCR | 8,411 | 15,103 | 6.8 | 12.3 | 109 | 153 | 15.9 (13.1 - 19.2) | 12.5 (10.6 - 14.6) |
| MDCD | 7,383 | 16,069 | 4.1 | 8.6 | 63 | 146 | 15.5 (11.9 - 19.8) | 16.9 (14.3 - 19.9) |
| Open Claims | 431,527 | 821,490 | 299.4 | 656.3 | 2,158 | 3,496 | 7.21 (6.91 - 7.52) | 5.33 (5.15 - 5.51) |
| OEHR | 37,764 | 80,961 | 15.7 | 38.8 | 175 | 413 | 11.2 (9.58 - 13.0) | 10.6 (9.65 - 11.7) |
| VA | 2,135 | 12,725 | 1.2 | 8.9 | <10 | 44 | < 8.05 (3.86 - 14.8) | 4.96 (3.60 - 6.66) |
| SIDIAP | 19,387 | 7,046 | 39.5 | 15.6 | 145 | 36 | 3.67 (3.10 - 4.32) | 2.31 (1.62 - 3.19) |
| Meta-analysis | 579,944 | 1,050,872 | 418.7 | 810.6 | 3,139 | 4,954 | 7.50 (7.24 - 7.76) | 6.11 (5.94 - 6.28) |
| Meta-analysis (w/o USOC) | 148,417 | 229,382 | 119.4 | 154.4 | 981 | 1,458 | 8.22 (7.71 - 8.75) | 9.45 (8.97 - 9.94) |

##### 4.6.12 Acute pancreatitis

**Supplementary Table 427:** Exposures, outcomes and risk estimates for Acute pancreatitis in DPP4I (female) (T) vs SU (female) (C) new-user cohorts under four different propensity score (PS) adjustment / time-at-risk (TAR) designs across data sources. We report total patient exposure time in 1,000 patient-year (KYr) units, uncalibrated and calibrated hazard ratios (HRs) and their 95% confidence intervals (CIs). Greyed out entries did not pass study diagnostics and are included only for completeness.

| | Patients | | Exposure (KYr) | | Outcomes | | Incidence (per KYr) | |
|---|---|---|---|---|---|---|---|---|
| | T | C | T | C | T | C | T | C |
| **PS-matching / on-treatment TAR** | | | | | | | | |
| CCAE | 38,847 | 65,460 | 30.4 | 47.7 | 71 | 112 | 2.33 (1.82 - 2.94) | 2.35 (1.93 - 2.82) |
| DAG | 10,485 | 2,040 | 11.4 | 2.1 | 0 | 0 | | |
| ODOD | 27,307 | 59,635 | 21.6 | 51.8 | 62 | 155 | 2.87 (2.20 - 3.67) | 2.99 (2.54 - 3.50) |
| MDCR | 8,109 | 15,336 | 7.6 | 13.7 | 18 | 53 | 2.35 (1.40 - 3.72) | 3.87 (2.90 - 5.06) |
| MDCD | 7,291 | 16,429 | 5.2 | 10.8 | 24 | 58 | 4.64 (2.97 - 6.90) | 5.37 (4.08 - 6.95) |
| Open Claims | 434,881 | 840,518 | 379.6 | 800.1 | 817 | 1,714 | 2.15 (2.01 - 2.30) | 2.14 (2.04 - 2.25) |
| OEHR | 36,246 | 82,846 | 18.6 | 47.1 | 36 | 104 | 1.94 (1.36 - 2.68) | 2.21 (1.80 - 2.67) |
| VA | 1,802 | 9,564 | 1.4 | 7.4 | <10 | 11 | < 7.31 (3.50 - 13.4) | 1.48 (0.74 - 2.66) |
| SIDIAP | 19,480 | 5,431 | 53.6 | 14.2 | 18 | 6 | 0.34 (0.20 - 0.53) | 0.42 (0.16 - 0.92) |
| Meta-analysis | 572,161 | 1,085,655 | 516.8 | 985.4 | 1,046 | 2,202 | 2.02 (1.90 - 2.15) | 2.23 (2.14 - 2.33) |
| Meta-analysis (w/o USOC) | 137,280 | 245,137 | 137.1 | 185.3 | 229 | 488 | 1.67 (1.46 - 1.90) | 2.63 (2.40 - 2.88) |
| **PS-stratification / on-treatment TAR** | | | | | | | | |
| CCAE | 45,212 | 67,373 | 35.7 | 48.9 | 77 | 115 | 2.16 (1.70 - 2.69) | 2.35 (1.94 - 2.82) |
| DAG | 10,516 | 2,367 | 11.4 | 2.5 | 0 | 0 | | |
| ODOD | 29,302 | 59,793 | 23.3 | 51.8 | 67 | 155 | 2.87 (2.23 - 3.65) | 2.99 (2.54 - 3.50) |
| MDCR | 8,746 | 15,653 | 8.3 | 14.0 | 18 | 54 | 2.18 (1.29 - 3.45) | 3.86 (2.90 - 5.04) |
| MDCD | 7,636 | 16,533 | 5.5 | 10.8 | 25 | 59 | 4.58 (2.96 - 6.76) | 5.45 (4.15 - 7.03) |
| Open Claims | 444,924 | 840,612 | 390.9 | 800.1 | 842 | 1,714 | 2.15 (2.01 - 2.30) | 2.14 (2.04 - 2.25) |
| OEHR | 38,728 | 82,935 | 20.1 | 47.2 | 39 | 105 | 1.94 (1.38 - 2.65) | 2.23 (1.82 - 2.69) |
| VA | 2,200 | 13,062 | 1.6 | 10.5 | <10 | 19 | < 6.21 (2.98 - 11.4) | 1.81 (1.09 - 2.82) |
| SIDIAP | 19,591 | 7,065 | 53.7 | 19.4 | 18 | 6 | 0.33 (0.20 - 0.53) | 0.31 (0.11 - 0.67) |
| Meta-analysis | 586,503 | 1,073,431 | 532.1 | 981.3 | 1,061 | 2,149 | 1.99 (1.88 - 2.12) | 2.19 (2.10 - 2.28) |
| Meta-analysis (w/o USOC) | 141,579 | 232,819 | 141.2 | 181.2 | 219 | 435 | 1.55 (1.35 - 1.77) | 2.40 (2.18 - 2.64) |
| **PS-matching / on-treatment TAR without escalation** | | | | | | | | |
| CCAE | 38,847 | 65,460 | 25.0 | 41.1 | 55 | 86 | 2.20 (1.66 - 2.87) | 2.09 (1.67 - 2.58) |
| DAG | 10,485 | 2,040 | 10.3 | 2.0 | 0 | 0 | | |
| ODOD | 27,307 | 59,635 | 18.2 | 46.1 | 51 | 131 | 2.81 (2.09 - 3.69) | 2.84 (2.38 - 3.37) |
| MDCR | 8,109 | 15,336 | 6.7 | 12.5 | 14 | 46 | 2.10 (1.15 - 3.53) | 3.67 (2.69 - 4.90) |
| MDCD | 7,291 | 16,429 | 4.0 | 9.0 | 16 | 49 | 3.97 (2.27 - 6.45) | 5.46 (4.04 - 7.22) |
| Open Claims | 434,881 | 840,518 | 301.4 | 673.2 | 635 | 1,402 | 2.11 (1.95 - 2.28) | 2.08 (1.97 - 2.19) |
| OEHR | 36,246 | 82,846 | 14.9 | 40.0 | 27 | 87 | 1.81 (1.19 - 2.63) | 2.18 (1.74 - 2.68) |
| VA | 1,802 | 9,564 | 1.1 | 6.4 | <10 | <10 | < 9.17 (4.40 - 16.9) | < 1.56 (0.75 - 2.87) |
| SIDIAP | 19,480 | 5,431 | 39.9 | 11.6 | 13 | <5 | 0.33 (0.17 - 0.56) | < 0.43 (0.14 - 1.01) |
| Meta-analysis | 552,681 | 1,080,224 | 370.1 | 821.9 | 798 | 1,801 | 2.16 (2.01 - 2.31) | 2.19 (2.09 - 2.29) |
| Meta-analysis (w/o USOC) | 117,800 | 239,706 | 68.8 | 148.7 | 163 | 399 | 2.37 (2.02 - 2.76) | 2.68 (2.43 - 2.96) |
| **PS-stratification / on-treatment TAR without escalation** | | | | | | | | |
| CCAE | 45,212 | 67,373 | 29.4 | 42.2 | 60 | 89 | 2.04 (1.56 - 2.63) | 2.11 (1.69 - 2.60) |
| DAG | 10,516 | 2,367 | 10.3 | 2.3 | 0 | 0 | | |
| ODOD | 29,302 | 59,793 | 19.6 | 46.1 | 56 | 131 | 2.86 (2.16 - 3.72) | 2.84 (2.38 - 3.37) |
| MDCR | 8,746 | 15,653 | 7.2 | 12.8 | 14 | 47 | 1.95 (1.07 - 3.27) | 3.67 (2.70 - 4.89) |
| MDCD | 7,636 | 16,533 | 4.2 | 9.0 | 16 | 50 | 3.77 (2.15 - 6.12) | 5.56 (4.13 - 7.33) |
| Open Claims | 444,924 | 840,612 | 310.4 | 673.3 | 653 | 1,402 | 2.10 (1.95 - 2.27) | 2.08 (1.97 - 2.19) |
| OEHR | 38,728 | 82,935 | 16.2 | 40.0 | 29 | 88 | 1.79 (1.20 - 2.57) | 2.20 (1.76 - 2.71) |
| VA | 2,200 | 13,062 | 1.3 | 9.2 | <10 | 16 | < 7.78 (3.73 - 14.3) | 1.75 (1.00 - 2.84) |
| SIDIAP | 19,591 | 7,065 | 40.0 | 15.7 | 13 | <5 | 0.33 (0.17 - 0.56) | < 0.32 (0.10 - 0.74) |
| Meta-analysis | 566,912 | 1,066,366 | 382.7 | 814.4 | 812 | 1,757 | 2.12 (1.98 - 2.27) | 2.16 (2.06 - 2.26) |
| Meta-analysis (w/o USOC) | 121,988 | 225,754 | 72.3 | 141.1 | 159 | 355 | 2.20 (1.87 - 2.57) | 2.52 (2.26 - 2.79) |

##### 4.6.13 Bone fracture

**Supplementary Table 428:** Exposures, outcomes and risk estimates for Bone fracture in DPP4I (female) (T) vs SU (female) (C) new-user cohorts under four different propensity score (PS) adjustment / time-at-risk (TAR) designs across data sources. We report total patient exposure time in 1,000 patient-year (KYr) units, uncalibrated and calibrated hazard ratios (HRs) and their 95% confidence intervals (CIs). Greyed out entries did not pass study diagnostics and are included only for completeness.

| | Patients | | Exposure (KYr) | | Outcomes | | Incidence (per KYr) | |
|---|---|---|---|---|---|---|---|---|
| | T | C | T | C | T | C | T | C |
| **PS-matching / on-treatment TAR** | | | | | | | | |
| CCAE | 35,539 | 60,320 | 27.2 | 43.2 | 573 | 892 | 21.1 (19.4 - 22.9) | 20.7 (19.3 - 22.1) |
| DAG | 9,003 | 1,776 | 9.5 | 1.8 | 236 | 55 | 24.8 (21.7 - 28.1) | 30.7 (23.2 - 40.0) |
| ODOD | 24,473 | 53,839 | 18.9 | 45.3 | 656 | 1,658 | 34.7 (32.1 - 37.5) | 36.6 (34.9 - 38.4) |
| MDCR | 6,884 | 13,051 | 6.3 | 11.3 | 286 | 551 | 45.1 (40.0 - 50.6) | 48.9 (44.9 - 53.2) |
| MDCD | 6,435 | 14,654 | 4.4 | 9.4 | 176 | 329 | 39.6 (34.0 - 45.9) | 35.1 (31.4 - 39.1) |
| Open Claims | 376,410 | 737,017 | 322.0 | 686.7 | 6,970 | 15,022 | 21.6 (21.1 - 22.2) | 21.9 (21.5 - 22.2) |
| OEHR | 33,656 | 76,804 | 17.0 | 42.5 | 380 | 1,111 | 22.4 (20.2 - 24.8) | 26.1 (24.6 - 27.7) |
| VA | 1,570 | 8,364 | 1.2 | 6.3 | 24 | 99 | 20.8 (13.3 - 31.0) | 15.7 (12.8 - 19.1) |
| SIDIAP | 16,703 | 4,925 | 44.6 | 12.4 | 1,013 | 252 | 22.7 (21.3 - 24.1) | 20.3 (17.9 - 23.0) |
| Meta-analysis | 510,673 | 970,750 | 451.2 | 858.8 | 10,314 | 19,969 | 22.9 (22.4 - 23.3) | 23.3 (22.9 - 23.6) |
| Meta-analysis (w/o USOC) | 134,263 | 233,733 | 129.1 | 172.1 | 3,344 | 4,947 | 25.9 (25.0 - 26.8) | 28.7 (27.9 - 29.6) |
| **PS-stratification / on-treatment TAR** | | | | | | | | |
| CCAE | 41,238 | 62,146 | 31.8 | 44.3 | 694 | 903 | 21.8 (20.2 - 23.5) | 20.4 (19.1 - 21.8) |
| DAG | 9,043 | 2,056 | 9.6 | 2.1 | 237 | 64 | 24.8 (21.7 - 28.2) | 30.4 (23.4 - 38.9) |
| ODOD | 26,228 | 53,992 | 20.3 | 45.3 | 709 | 1,659 | 35.0 (32.4 - 37.6) | 36.6 (34.9 - 38.4) |
| MDCR | 7,400 | 13,337 | 6.8 | 11.5 | 311 | 559 | 45.7 (40.7 - 51.0) | 48.5 (44.6 - 52.7) |
| MDCD | 6,705 | 14,781 | 4.7 | 9.4 | 185 | 331 | 39.7 (34.2 - 45.8) | 35.2 (31.5 - 39.2) |
| Open Claims | 385,029 | 737,112 | 331.6 | 686.7 | 7,245 | 15,022 | 21.9 (21.4 - 22.4) | 21.9 (21.5 - 22.2) |
| OEHR | 35,951 | 76,885 | 18.4 | 42.6 | 416 | 1,112 | 22.7 (20.5 - 24.9) | 26.1 (24.6 - 27.7) |
| VA | 1,908 | 11,555 | 1.4 | 9.0 | 29 | 146 | 21.5 (14.4 - 30.8) | 16.2 (13.7 - 19.0) |
| SIDIAP | 16,780 | 6,472 | 44.7 | 17.1 | 1,013 | 350 | 22.7 (21.3 - 24.1) | 20.5 (18.4 - 22.8) |
| Meta-analysis | 521,669 | 952,000 | 463.1 | 849.6 | 10,625 | 19,669 | 22.9 (22.5 - 23.4) | 23.2 (22.8 - 23.5) |
| Meta-analysis (w/o USOC) | 136,640 | 214,888 | 131.5 | 162.8 | 3,380 | 4,647 | 25.7 (24.8 - 26.6) | 28.5 (27.7 - 29.4) |
| **PS-matching / on-treatment TAR without escalation** | | | | | | | | |
| CCAE | 35,539 | 60,320 | 22.4 | 37.3 | 460 | 759 | 20.5 (18.7 - 22.5) | 20.3 (18.9 - 21.8) |
| DAG | 9,003 | 1,776 | 8.6 | 1.7 | 217 | 52 | 25.2 (22.0 - 28.8) | 31.3 (23.4 - 41.0) |
| ODOD | 24,473 | 53,839 | 15.9 | 40.4 | 533 | 1,478 | 33.5 (30.7 - 36.4) | 36.6 (34.7 - 38.5) |
| MDCR | 6,884 | 13,051 | 5.6 | 10.3 | 236 | 509 | 42.3 (37.1 - 48.1) | 49.2 (45.0 - 53.6) |
| MDCD | 6,435 | 14,654 | 3.5 | 7.8 | 131 | 276 | 37.8 (31.6 - 44.8) | 35.3 (31.3 - 39.8) |
| Open Claims | 376,410 | 737,017 | 256.3 | 578.6 | 5,528 | 12,655 | 21.6 (21.0 - 22.1) | 21.9 (21.5 - 22.3) |
| OEHR | 33,656 | 76,804 | 13.7 | 36.3 | 283 | 926 | 20.6 (18.3 - 23.2) | 25.5 (23.9 - 27.2) |
| VA | 1,570 | 8,364 | 0.9 | 5.5 | 18 | 84 | 19.5 (11.6 - 30.9) | 15.3 (12.2 - 18.9) |
| SIDIAP | 16,703 | 4,925 | 33.1 | 10.2 | 783 | 214 | 23.6 (22.0 - 25.3) | 21.1 (18.3 - 24.1) |
| Meta-analysis | 510,673 | 970,750 | 360.1 | 728.1 | 8,189 | 16,953 | 22.7 (22.3 - 23.2) | 23.3 (22.9 - 23.6) |
| Meta-analysis (w/o USOC) | 134,263 | 233,733 | 103.8 | 149.5 | 2,661 | 4,298 | 25.6 (24.7 - 26.6) | 28.7 (27.9 - 29.6) |
| **PS-stratification / on-treatment TAR without escalation** | | | | | | | | |
| CCAE | 41,238 | 62,146 | 26.3 | 38.3 | 558 | 768 | 21.2 (19.5 - 23.1) | 20.0 (18.6 - 21.5) |
| DAG | 9,043 | 2,056 | 8.6 | 1.9 | 218 | 60 | 25.2 (22.0 - 28.8) | 30.8 (23.5 - 39.7) |
| ODOD | 26,228 | 53,992 | 17.1 | 40.5 | 582 | 1,478 | 34.0 (31.3 - 36.9) | 36.5 (34.7 - 38.4) |
| MDCR | 7,400 | 13,337 | 6.0 | 10.6 | 260 | 515 | 43.5 (38.3 - 49.1) | 48.6 (44.5 - 53.0) |
| MDCD | 6,705 | 14,781 | 3.6 | 7.9 | 138 | 277 | 37.9 (31.9 - 44.8) | 35.3 (31.3 - 39.7) |
| Open Claims | 385,029 | 737,112 | 264.0 | 578.6 | 5,749 | 12,655 | 21.8 (21.2 - 22.4) | 21.9 (21.5 - 22.3) |
| OEHR | 35,951 | 76,885 | 14.8 | 36.3 | 310 | 927 | 20.9 (18.6 - 23.3) | 25.5 (23.9 - 27.2) |
| VA | 1,908 | 11,555 | 1.1 | 7.9 | 22 | 124 | 20.3 (12.7 - 30.8) | 15.7 (13.0 - 18.7) |
| SIDIAP | 16,780 | 6,472 | 33.2 | 13.9 | 783 | 289 | 23.6 (22.0 - 25.3) | 20.8 (18.5 - 23.4) |
| Meta-analysis | 521,669 | 952,000 | 370.0 | 720.2 | 8,460 | 16,692 | 22.9 (22.4 - 23.4) | 23.2 (22.8 - 23.5) |
| Meta-analysis (w/o USOC) | 136,640 | 214,888 | 106.1 | 141.5 | 2,711 | 4,037 | 25.6 (24.6 - 26.5) | 28.5 (27.7 - 29.4) |

###### 4.6.14 Diabetic ketoacidosis

**Supplementary Table 429:** Exposures, outcomes and risk estimates for Diabetic ketoacidosis in DPP4I (female) (T) vs SU (female) (C) new-user cohorts under four different propensity score (PS) adjustment / time-at-risk (TAR) designs across data sources. We report total patient exposure time in 1,000 patient-year (KYr) units, uncalibrated and calibrated hazard ratios (HRs) and their 95% confidence intervals (CIs). Greyed out entries did not pass study diagnostics and are included only for completeness.

| | Patients | | Exposure (KYr) | | Outcomes | | Incidence (per KYr) | |
|---|---|---|---|---|---|---|---|---|
| | T | C | T | C | T | C | T | C |
| **PS-matching / on-treatment TAR** | | | | | | | | |
| CCAE | 39,091 | 65,975 | 30.7 | 48.2 | 26 | 60 | 0.85 (0.55 - 1.24) | 1.25 (0.95 - 1.60) |
| DAG | 10,485 | 2,040 | 11.4 | 2.1 | 0 | 0 | | |
| ODOD | 27,468 | 60,103 | 21.8 | 52.3 | 36 | 98 | 1.66 (1.16 - 2.29) | 1.88 (1.52 - 2.29) |
| MDCR | 8,158 | 15,498 | 7.7 | 13.9 | 10 | 19 | 1.30 (0.62 - 2.39) | 1.37 (0.82 - 2.14) |
| MDCD | 7,403 | 16,677 | 5.2 | 11.0 | 17 | 39 | 3.24 (1.89 - 5.19) | 3.56 (2.53 - 4.86) |
| Open Claims | 439,162 | 850,402 | 383.5 | 810.3 | 354 | 732 | 0.92 (0.83 - 1.02) | 0.90 (0.84 - 0.97) |
| OEHR | 36,401 | 83,212 | 18.7 | 47.4 | 24 | 68 | 1.29 (0.82 - 1.91) | 1.43 (1.11 - 1.82) |
| VA | 1,820 | 9,661 | 1.4 | 7.5 | <10 | <10 | < 7.24 (3.47 - 13.3) | < 1.34 (0.64 - 2.46) |
| SIDIAP | 19,514 | 5,440 | 53.7 | 14.2 | 0 | 0 | | |
| Meta-analysis | 557,683 | 1,091,867 | 467.5 | 982.9 | 467 | 1,016 | 1.00 (0.91 - 1.09) | 1.03 (0.97 - 1.10) |
| Meta-analysis (w/o USOC) | 118,521 | 241,465 | 84.0 | 172.6 | 113 | 284 | 1.34 (1.11 - 1.62) | 1.65 (1.46 - 1.85) |
| **PS-stratification / on-treatment TAR** | | | | | | | | |
| CCAE | 45,453 | 67,906 | 36.0 | 49.3 | 29 | 62 | 0.81 (0.54 - 1.16) | 1.26 (0.96 - 1.61) |
| DAG | 10,516 | 2,367 | 11.4 | 2.5 | 0 | 0 | | |
| ODOD | 29,474 | 60,261 | 23.5 | 52.3 | 37 | 98 | 1.58 (1.11 - 2.17) | 1.87 (1.52 - 2.28) |
| MDCR | 8,794 | 15,819 | 8.3 | 14.2 | 10 | 19 | 1.20 (0.58 - 2.22) | 1.34 (0.81 - 2.10) |
| MDCD | 7,748 | 16,782 | 5.5 | 11.0 | 17 | 39 | 3.07 (1.79 - 4.92) | 3.55 (2.52 - 4.85) |
| Open Claims | 449,170 | 850,500 | 394.7 | 810.3 | 362 | 732 | 0.92 (0.83 - 1.02) | 0.90 (0.84 - 0.97) |
| OEHR | 38,879 | 83,301 | 20.2 | 47.4 | 25 | 68 | 1.24 (0.80 - 1.83) | 1.43 (1.11 - 1.82) |
| VA | 2,219 | 13,213 | 1.6 | 10.6 | <10 | 12 | < 6.15 (2.95 - 11.3) | 1.13 (0.58 - 1.97) |
| SIDIAP | 19,627 | 7,073 | 53.8 | 19.4 | 0 | 0 | | |
| Meta-analysis | 571,770 | 1,077,787 | 482.6 | 973.5 | 463 | 979 | 0.96 (0.87 - 1.05) | 1.01 (0.94 - 1.07) |
| Meta-analysis (w/o USOC) | 122,600 | 227,287 | 87.9 | 163.2 | 101 | 247 | 1.15 (0.94 - 1.40) | 1.51 (1.33 - 1.71) |
| **PS-matching / on-treatment TAR without escalation** | | | | | | | | |
| CCAE | 39,091 | 65,975 | 25.2 | 41.5 | 16 | 46 | 0.64 (0.36 - 1.03) | 1.11 (0.81 - 1.48) |
| DAG | 10,485 | 2,040 | 10.3 | 2.0 | 0 | 0 | | |
| ODOD | 27,468 | 60,103 | 18.2 | 46.5 | 29 | 78 | 1.59 (1.06 - 2.28) | 1.68 (1.33 - 2.09) |
| MDCR | 8,158 | 15,498 | 6.7 | 12.7 | 8 | 16 | 1.20 (0.52 - 2.36) | 1.26 (0.72 - 2.05) |
| MDCD | 7,403 | 16,677 | 4.1 | 9.1 | 12 | 26 | 2.94 (1.52 - 5.13) | 2.85 (1.86 - 4.18) |
| Open Claims | 439,162 | 850,402 | 304.5 | 681.6 | 248 | 542 | 0.81 (0.72 - 0.92) | 0.80 (0.73 - 0.86) |
| OEHR | 36,401 | 83,212 | 15.0 | 40.2 | 13 | 44 | 0.87 (0.46 - 1.48) | 1.09 (0.80 - 1.47) |
| VA | 1,820 | 9,661 | 1.1 | 6.5 | <10 | <10 | < 9.09 (4.36 - 16.7) | < 1.55 (0.74 - 2.84) |
| SIDIAP | 19,514 | 5,440 | 40.0 | 11.6 | 0 | 0 | | |
| Meta-analysis | 557,683 | 1,091,867 | 373.7 | 831.6 | 326 | 752 | 0.87 (0.78 - 0.97) | 0.90 (0.84 - 0.97) |
| Meta-analysis (w/o USOC) | 118,521 | 241,465 | 69.2 | 149.9 | 78 | 210 | 1.13 (0.89 - 1.41) | 1.40 (1.22 - 1.60) |
| **PS-stratification / on-treatment TAR without escalation** | | | | | | | | |
| CCAE | 45,453 | 67,906 | 29.6 | 42.5 | 18 | 48 | 0.61 (0.36 - 0.96) | 1.13 (0.83 - 1.50) |
| DAG | 10,516 | 2,367 | 10.3 | 2.3 | 0 | 0 | | |
| ODOD | 29,474 | 60,261 | 19.7 | 46.5 | 30 | 78 | 1.53 (1.03 - 2.18) | 1.68 (1.33 - 2.09) |
| MDCR | 8,794 | 15,819 | 7.2 | 13.0 | 8 | 16 | 1.11 (0.48 - 2.18) | 1.23 (0.71 - 2.00) |
| MDCD | 7,748 | 16,782 | 4.3 | 9.1 | 12 | 26 | 2.78 (1.44 - 4.86) | 2.85 (1.86 - 4.17) |
| Open Claims | 449,170 | 850,500 | 313.5 | 681.7 | 252 | 542 | 0.80 (0.71 - 0.91) | 0.80 (0.73 - 0.86) |
| OEHR | 38,879 | 83,301 | 16.3 | 40.2 | 14 | 44 | 0.86 (0.47 - 1.44) | 1.09 (0.79 - 1.47) |
| VA | 2,219 | 13,213 | 1.3 | 9.3 | <10 | <10 | < 7.71 (3.70 - 14.2) | < 1.08 (0.52 - 1.99) |
| SIDIAP | 19,627 | 7,073 | 40.0 | 15.7 | 0 | 0 | | |
| Meta-analysis | 571,770 | 1,077,787 | 386.2 | 823.9 | 322 | 728 | 0.83 (0.75 - 0.93) | 0.88 (0.82 - 0.95) |
| Meta-analysis (w/o USOC) | 122,600 | 227,287 | 72.7 | 142.2 | 70 | 186 | 0.96 (0.75 - 1.22) | 1.31 (1.13 - 1.51) |

##### 4.6.15 Diarrhea

**Supplementary Table 430:** Exposures, outcomes and risk estimates for Diarrhea in DPP4I (female) (T) vs SU (female) (C) new-user cohorts under four different propensity score (PS) adjustment / time-at-risk (TAR) designs across data sources. We report total patient exposure time in 1,000 patient-year (KYr) units, uncalibrated and calibrated hazard ratios (HRs) and their 95% confidence intervals (CIs). Greyed out entries did not pass study diagnostics and are included only for completeness.

| | Patients | | Exposure (KYr) | | Outcomes | | Incidence (per KYr) | |
|---|---|---|---|---|---|---|---|---|
| | T | C | T | C | T | C | T | C |
| **PS-matching / on-treatment TAR** | | | | | | | | |
| CCAE | 33,476 | 56,954 | 25.5 | 40.5 | 871 | 1,460 | 34.2 (32.0 - 36.5) | 36.0 (34.2 - 37.9) |
| DAG | 10,305 | 1,988 | 11.1 | 2.1 | 29 | 7 | 2.61 (1.75 - 3.75) | 3.38 (1.36 - 6.96) |
| ODOD | 22,398 | 49,215 | 17.1 | 40.6 | 830 | 2,101 | 48.6 (45.3 - 52.0) | 51.7 (49.5 - 54.0) |
| MDCR | 6,671 | 12,831 | 6.1 | 11.1 | 251 | 514 | 41.5 (36.5 - 47.0) | 46.2 (42.3 - 50.4) |
| MDCD | 5,573 | 12,971 | 3.7 | 8.1 | 256 | 591 | 69.0 (60.8 - 78.0) | 73.4 (67.6 - 79.5) |
| Open Claims | 366,241 | 719,300 | 312.9 | 670.5 | 7,824 | 16,350 | 25.0 (24.5 - 25.6) | 24.4 (24.0 - 24.8) |
| OEHR | 31,061 | 71,577 | 15.1 | 38.2 | 744 | 2,051 | 49.2 (45.7 - 52.9) | 53.7 (51.4 - 56.1) |
| VA | 1,386 | 7,321 | 1.0 | 5.3 | 37 | 187 | 37.1 (26.1 - 51.1) | 35.1 (30.2 - 40.5) |
| SIDIAP | 18,349 | 5,245 | 50.2 | 13.6 | 621 | 121 | 12.4 (11.4 - 13.4) | 8.92 (7.40 - 10.7) |
| Meta-analysis | 495,460 | 937,402 | 442.6 | 830.0 | 11,463 | 23,382 | 25.9 (25.4 - 26.4) | 28.2 (27.8 - 28.5) |
| Meta-analysis (w/o USOC) | 129,219 | 218,102 | 129.7 | 159.5 | 3,639 | 7,032 | 28.1 (27.1 - 29.0) | 44.1 (43.1 - 45.1) |
| **PS-stratification / on-treatment TAR** | | | | | | | | |
| CCAE | 38,768 | 58,736 | 29.7 | 41.6 | 1,050 | 1,493 | 35.3 (33.2 - 37.5) | 35.9 (34.1 - 37.7) |
| DAG | 10,336 | 2,309 | 11.1 | 2.4 | 29 | 8 | 2.60 (1.74 - 3.74) | 3.27 (1.41 - 6.44) |
| ODOD | 24,049 | 49,356 | 18.4 | 40.7 | 897 | 2,104 | 48.7 (45.6 - 52.0) | 51.7 (49.5 - 54.0) |
| MDCR | 7,235 | 13,140 | 6.6 | 11.4 | 285 | 523 | 43.4 (38.5 - 48.7) | 46.0 (42.1 - 50.1) |
| MDCD | 5,785 | 13,053 | 3.9 | 8.1 | 273 | 591 | 70.5 (62.4 - 79.4) | 73.2 (67.4 - 79.4) |
| Open Claims | 374,458 | 719,384 | 321.8 | 670.5 | 8,141 | 16,352 | 25.3 (24.8 - 25.9) | 24.4 (24.0 - 24.8) |
| OEHR | 33,203 | 71,648 | 16.4 | 38.2 | 804 | 2,051 | 49.2 (45.8 - 52.7) | 53.6 (51.3 - 56.0) |
| VA | 1,662 | 10,429 | 1.2 | 7.9 | 47 | 296 | 40.2 (29.5 - 53.5) | 37.3 (33.1 - 41.8) |
| SIDIAP | 18,475 | 6,861 | 50.3 | 18.6 | 627 | 142 | 12.5 (11.5 - 13.5) | 7.62 (6.42 - 8.99) |
| Meta-analysis | 506,524 | 921,434 | 454.3 | 823.5 | 11,833 | 22,673 | 26.0 (25.6 - 26.5) | 27.5 (27.2 - 27.9) |
| Meta-analysis (w/o USOC) | 132,066 | 202,050 | 132.5 | 153.0 | 3,692 | 6,321 | 27.9 (27.0 - 28.8) | 41.3 (40.3 - 42.4) |
| **PS-matching / on-treatment TAR without escalation** | | | | | | | | |
| CCAE | 33,476 | 56,954 | 21.2 | 35.2 | 710 | 1,275 | 33.5 (31.0 - 36.0) | 36.2 (34.2 - 38.2) |
| DAG | 10,305 | 1,988 | 10.1 | 1.9 | 28 | 6 | 2.78 (1.85 - 4.02) | 3.13 (1.15 - 6.81) |
| ODOD | 22,398 | 49,215 | 14.5 | 36.6 | 700 | 1,906 | 48.3 (44.8 - 52.0) | 52.1 (49.8 - 54.5) |
| MDCR | 6,671 | 12,831 | 5.3 | 10.2 | 221 | 464 | 41.7 (36.3 - 47.5) | 45.4 (41.3 - 49.7) |
| MDCD | 5,573 | 12,971 | 3.0 | 6.8 | 211 | 500 | 71.2 (61.9 - 81.5) | 73.8 (67.4 - 80.5) |
| Open Claims | 366,241 | 719,300 | 250.6 | 568.4 | 6,265 | 13,841 | 25.0 (24.4 - 25.6) | 24.4 (23.9 - 24.8) |
| OEHR | 31,061 | 71,577 | 12.3 | 33.1 | 590 | 1,645 | 47.9 (44.1 - 51.9) | 49.8 (47.4 - 52.2) |
| VA | 1,386 | 7,321 | 0.8 | 4.7 | 31 | 160 | 38.8 (26.3 - 55.0) | 34.3 (29.2 - 40.0) |
| SIDIAP | 18,349 | 5,245 | 37.4 | 11.1 | 462 | 92 | 12.3 (11.2 - 13.5) | 8.29 (6.69 - 10.2) |
| Meta-analysis | 495,460 | 937,402 | 355.2 | 707.9 | 9,218 | 19,889 | 26.0 (25.4 - 26.5) | 28.1 (27.7 - 28.5) |
| Meta-analysis (w/o USOC) | 129,219 | 218,102 | 104.6 | 139.5 | 2,953 | 6,048 | 28.2 (27.2 - 29.3) | 43.4 (42.3 - 44.5) |
| **PS-stratification / on-treatment TAR without escalation** | | | | | | | | |
| CCAE | 38,768 | 58,736 | 24.8 | 36.2 | 865 | 1,306 | 34.9 (32.6 - 37.3) | 36.1 (34.1 - 38.1) |
| DAG | 10,336 | 2,309 | 10.1 | 2.3 | 28 | 7 | 2.78 (1.84 - 4.01) | 3.11 (1.25 - 6.41) |
| ODOD | 24,049 | 49,356 | 15.6 | 36.6 | 752 | 1,909 | 48.1 (44.8 - 51.7) | 52.2 (49.9 - 54.6) |
| MDCR | 7,235 | 13,140 | 5.8 | 10.5 | 252 | 472 | 43.7 (38.5 - 49.4) | 45.1 (41.1 - 49.3) |
| MDCD | 5,785 | 13,053 | 3.1 | 6.8 | 225 | 500 | 72.9 (63.7 - 83.1) | 73.6 (67.3 - 80.3) |
| Open Claims | 374,458 | 719,384 | 257.8 | 568.4 | 6,522 | 13,843 | 25.3 (24.7 - 25.9) | 24.4 (24.0 - 24.8) |
| OEHR | 33,203 | 71,648 | 13.3 | 33.1 | 641 | 1,645 | 48.1 (44.4 - 51.9) | 49.7 (47.3 - 52.2) |
| VA | 1,662 | 10,429 | 0.9 | 7.0 | 38 | 252 | 40.4 (28.6 - 55.4) | 36.0 (31.7 - 40.8) |
| SIDIAP | 18,475 | 6,861 | 37.5 | 15.1 | 466 | 112 | 12.4 (11.3 - 13.6) | 7.40 (6.10 - 8.91) |
| Meta-analysis | 506,524 | 921,434 | 365.0 | 702.1 | 9,526 | 19,294 | 26.1 (25.6 - 26.6) | 27.5 (27.1 - 27.9) |
| Meta-analysis (w/o USOC) | 132,066 | 202,050 | 107.1 | 133.7 | 3,004 | 5,451 | 28.0 (27.0 - 29.1) | 40.8 (39.7 - 41.9) |

##### 4.6.16 Genitourinary infection

**Supplementary Table 431:** Exposures, outcomes and risk estimates for Genitourinary infection in DPP4I (female) (T) vs SU (female) (C) new-user cohorts under four different propensity score (PS) adjustment / time-at-risk (TAR) designs across data sources. We report total patient exposure time in 1,000 patient-year (KYr) units, uncalibrated and calibrated hazard ratios (HRs) and their 95% confidence intervals (CIs). Greyed out entries did not pass study diagnostics and are included only for completeness.

| | Patients | | Exposure (KYr) | | Outcomes | | Incidence (per KYr) | |
|---|---|---|---|---|---|---|---|---|
| | T | C | T | C | T | C | T | C |
| **PS-matching / on-treatment TAR** | | | | | | | | |
| CCAE | 27,245 | 46,972 | 20.2 | 32.8 | 1,460 | 2,153 | 72.3 (68.6 - 76.1) | 65.6 (62.9 - 68.4) |
| DAG | 7,047 | 1,394 | 7.1 | 1.4 | 384 | 67 | 53.8 (48.5 - 59.4) | 49.4 (38.3 - 62.7) |
| ODOD | 18,104 | 40,283 | 13.3 | 31.9 | 1,331 | 3,262 | 99.8 (94.5 - 105) | 102 (98.7 - 106) |
| MDCR | 5,639 | 10,677 | 4.9 | 9.0 | 479 | 876 | 96.8 (88.3 - 106) | 97.1 (90.8 - 104) |
| MDCD | 4,784 | 10,812 | 3.2 | 6.6 | 328 | 700 | 104 (93.0 - 116) | 106 (98.0 - 114) |
| Open Claims | 289,230 | 578,011 | 243.1 | 534.8 | 12,925 | 27,146 | 53.2 (52.3 - 54.1) | 50.8 (50.2 - 51.4) |
| OEHR | 27,853 | 64,207 | 13.3 | 33.7 | 1,032 | 2,683 | 77.4 (72.8 - 82.3) | 79.7 (76.7 - 82.8) |
| VA | 1,178 | 6,123 | 0.8 | 4.4 | 45 | 202 | 53.2 (38.8 - 71.2) | 45.4 (39.4 - 52.1) |
| SIDIAP | 13,675 | 4,493 | 35.6 | 10.8 | 2,053 | 539 | 57.6 (55.2 - 60.2) | 50.1 (46.0 - 54.5) |
| Meta-analysis | 394,755 | 762,972 | 341.7 | 665.4 | 20,037 | 37,628 | 58.6 (57.8 - 59.5) | 56.5 (56.0 - 57.1) |
| Meta-analysis (w/o USOC) | 105,525 | 184,961 | 98.6 | 130.6 | 7,112 | 10,482 | 72.1 (70.5 - 73.8) | 80.2 (78.7 - 81.8) |
| **PS-stratification / on-treatment TAR** | | | | | | | | |
| CCAE | 31,471 | 48,414 | 23.4 | 33.7 | 1,751 | 2,226 | 74.8 (71.3 - 78.4) | 66.0 (63.3 - 68.8) |
| DAG | 7,079 | 1,619 | 7.2 | 1.6 | 386 | 82 | 53.8 (48.6 - 59.5) | 52.1 (41.4 - 64.7) |
| ODOD | 19,378 | 40,392 | 14.3 | 32.0 | 1,439 | 3,269 | 101 (95.7 - 106) | 102 (98.8 - 106) |
| MDCR | 6,012 | 11,044 | 5.3 | 9.3 | 505 | 900 | 95.5 (87.3 - 104) | 96.3 (90.1 - 103) |
| MDCD | 4,991 | 10,858 | 3.3 | 6.6 | 346 | 707 | 105 (94.2 - 117) | 107 (99.0 - 115) |
| Open Claims | 295,218 | 578,219 | 249.4 | 534.8 | 13,368 | 27,151 | 53.6 (52.7 - 54.5) | 50.8 (50.2 - 51.4) |
| OEHR | 29,709 | 64,264 | 14.4 | 33.7 | 1,118 | 2,686 | 77.8 (73.3 - 82.5) | 79.7 (76.7 - 82.8) |
| VA | 1,400 | 8,809 | 1.0 | 6.7 | 51 | 340 | 52.3 (39.0 - 68.8) | 50.8 (45.5 - 56.5) |
| SIDIAP | 13,757 | 5,991 | 35.7 | 15.0 | 2,058 | 737 | 57.7 (55.2 - 60.2) | 49.2 (45.7 - 52.9) |
| Meta-analysis | 402,624 | 749,943 | 349.6 | 660.1 | 20,625 | 37,051 | 59.0 (58.2 - 59.8) | 56.1 (55.6 - 56.7) |
| Meta-analysis (w/o USOC) | 107,406 | 171,724 | 100.2 | 125.3 | 7,257 | 9,900 | 72.4 (70.8 - 74.1) | 79.0 (77.5 - 80.6) |
| **PS-matching / on-treatment TAR without escalation** | | | | | | | | |
| CCAE | 27,245 | 46,972 | 16.7 | 28.6 | 1,222 | 1,852 | 73.0 (69.0 - 77.2) | 64.8 (61.8 - 67.8) |
| DAG | 7,047 | 1,394 | 6.5 | 1.3 | 352 | 60 | 54.2 (48.7 - 60.2) | 47.8 (36.5 - 61.5) |
| ODOD | 18,104 | 40,283 | 11.4 | 28.8 | 1,126 | 2,953 | 98.7 (93.0 - 105) | 102 (98.8 - 106) |
| MDCR | 5,639 | 10,677 | 4.3 | 8.3 | 407 | 787 | 93.8 (84.9 - 103) | 94.4 (87.9 - 101) |
| MDCD | 4,784 | 10,812 | 2.5 | 5.6 | 241 | 588 | 95.6 (83.9 - 108) | 104 (96.0 - 113) |
| Open Claims | 289,230 | 578,011 | 194.5 | 454.2 | 10,398 | 23,153 | 53.4 (52.4 - 54.5) | 51.0 (50.3 - 51.6) |
| OEHR | 27,853 | 64,207 | 10.9 | 29.2 | 804 | 2,197 | 74.0 (69.0 - 79.3) | 75.3 (72.2 - 78.5) |
| VA | 1,178 | 6,123 | 0.7 | 3.9 | 35 | 175 | 52.9 (36.9 - 73.6) | 44.9 (38.5 - 52.1) |
| SIDIAP | 13,675 | 4,493 | 27.0 | 8.9 | 1,502 | 441 | 55.6 (52.9 - 58.5) | 49.7 (45.1 - 54.5) |
| Meta-analysis | 394,755 | 762,972 | 274.6 | 568.8 | 16,087 | 32,206 | 58.6 (57.7 - 59.5) | 56.6 (56.0 - 57.2) |
| Meta-analysis (w/o USOC) | 105,525 | 184,961 | 80.0 | 114.6 | 5,689 | 9,053 | 71.1 (69.3 - 73.0) | 79.0 (77.4 - 80.6) |
| **PS-stratification / on-treatment TAR without escalation** | | | | | | | | |
| CCAE | 31,471 | 48,414 | 19.5 | 29.4 | 1,468 | 1,918 | 75.4 (71.6 - 79.4) | 65.2 (62.3 - 68.2) |
| DAG | 7,079 | 1,619 | 6.5 | 1.5 | 354 | 73 | 54.3 (48.8 - 60.3) | 50.1 (39.3 - 63.0) |
| ODOD | 19,378 | 40,392 | 12.2 | 28.9 | 1,220 | 2,959 | 100.0 (94.4 - 106) | 103 (98.9 - 106) |
| MDCR | 6,012 | 11,044 | 4.6 | 8.6 | 431 | 809 | 93.0 (84.4 - 102) | 93.7 (87.3 - 100) |
| MDCD | 4,991 | 10,858 | 2.6 | 5.6 | 255 | 595 | 96.9 (85.4 - 110) | 105 (97.0 - 114) |
| Open Claims | 295,218 | 578,219 | 199.6 | 454.3 | 10,757 | 23,157 | 53.9 (52.9 - 54.9) | 51.0 (50.3 - 51.6) |
| OEHR | 29,709 | 64,264 | 11.7 | 29.2 | 874 | 2,200 | 74.6 (69.7 - 79.7) | 75.4 (72.2 - 78.6) |
| VA | 1,400 | 8,809 | 0.8 | 5.9 | 40 | 297 | 52.2 (37.3 - 71.0) | 50.5 (44.9 - 56.6) |
| SIDIAP | 13,757 | 5,991 | 27.1 | 12.3 | 1,507 | 581 | 55.7 (52.9 - 58.6) | 47.2 (43.4 - 51.2) |
| Meta-analysis | 402,624 | 749,943 | 281.2 | 564.2 | 16,611 | 31,697 | 59.1 (58.2 - 60.0) | 56.2 (55.6 - 56.8) |
| Meta-analysis (w/o USOC) | 107,406 | 171,724 | 81.6 | 109.9 | 5,854 | 8,540 | 71.8 (69.9 - 73.6) | 77.7 (76.1 - 79.4) |

##### 4.6.17 Hyperkalemia

**Supplementary Table 432:** Exposures, outcomes and risk estimates for Hyperkalemia in DPP4I (female) (T) vs SU (female) (C) new-user cohorts under four different propensity score (PS) adjustment / time-at-risk (TAR) designs across data sources. We report total patient exposure time in 1,000 patient-year (KYr) units, uncalibrated and calibrated hazard ratios (HRs) and their 95% confidence intervals (CIs). Greyed out entries did not pass study diagnostics and are included only for completeness.

| | Patients | | Exposure (KYr) | | Outcomes | | Incidence (per KYr) | |
|---|---|---|---|---|---|---|---|---|
| | T | C | T | C | T | C | T | C |
| **PS-matching / on-treatment TAR** | | | | | | | | |
| CCAE | 38,521 | 65,087 | 30.0 | 47.2 | 172 | 368 | 5.73 (4.90 - 6.65) | 7.79 (7.02 - 8.63) |
| DAG | 9,775 | 1,951 | 10.4 | 2.0 | 176 | 22 | 16.9 (14.5 - 19.6) | 10.9 (6.85 - 16.5) |
| ODOD | 26,208 | 57,432 | 20.4 | 48.7 | 448 | 1,124 | 22.0 (20.0 - 24.1) | 23.1 (21.7 - 24.5) |
| MDCR | 7,925 | 14,952 | 7.4 | 13.2 | 111 | 203 | 15.0 (12.3 - 18.0) | 15.3 (13.3 - 17.6) |
| MDCD | 7,174 | 16,221 | 5.0 | 10.5 | 86 | 178 | 17.3 (13.8 - 21.3) | 17.0 (14.6 - 19.6) |
| Open Claims | 429,599 | 833,862 | 372.3 | 787.1 | 2,865 | 6,215 | 7.70 (7.42 - 7.98) | 7.90 (7.70 - 8.09) |
| OEHR | 35,700 | 81,633 | 18.1 | 45.8 | 242 | 642 | 13.4 (11.7 - 15.2) | 14.0 (12.9 - 15.1) |
| VA | 1,735 | 9,218 | 1.3 | 7.0 | 14 | 88 | 10.8 (5.90 - 18.1) | 12.5 (10.0 - 15.4) |
| SIDIAP | 18,570 | 5,270 | 50.4 | 13.6 | 453 | 128 | 8.99 (8.18 - 9.86) | 9.41 (7.85 - 11.2) |
| Meta-analysis | 575,207 | 1,085,626 | 515.3 | 975.3 | 4,567 | 8,968 | 8.86 (8.61 - 9.12) | 9.20 (9.01 - 9.39) |
| Meta-analysis (w/o USOC) | 145,608 | 251,764 | 143.0 | 188.1 | 1,702 | 2,753 | 11.9 (11.3 - 12.5) | 14.6 (14.1 - 15.2) |
| **PS-stratification / on-treatment TAR** | | | | | | | | |
| CCAE | 44,818 | 67,007 | 35.3 | 48.4 | 206 | 371 | 5.84 (5.07 - 6.69) | 7.67 (6.91 - 8.49) |
| DAG | 9,799 | 2,266 | 10.4 | 2.4 | 176 | 29 | 16.9 (14.5 - 19.6) | 12.2 (8.18 - 17.5) |
| ODOD | 28,155 | 57,590 | 22.0 | 48.8 | 480 | 1,125 | 21.9 (19.9 - 23.9) | 23.1 (21.7 - 24.5) |
| MDCR | 8,538 | 15,344 | 8.0 | 13.6 | 119 | 209 | 14.8 (12.3 - 17.8) | 15.4 (13.3 - 17.6) |
| MDCD | 7,515 | 16,325 | 5.3 | 10.5 | 94 | 179 | 17.9 (14.5 - 21.9) | 17.0 (14.6 - 19.7) |
| Open Claims | 439,411 | 833,960 | 383.1 | 787.1 | 2,989 | 6,215 | 7.80 (7.52 - 8.09) | 7.90 (7.70 - 8.09) |
| OEHR | 38,147 | 81,718 | 19.6 | 45.9 | 261 | 642 | 13.3 (11.8 - 15.1) | 14.0 (12.9 - 15.1) |
| VA | 2,107 | 12,693 | 1.5 | 10.0 | 15 | 139 | 9.83 (5.50 - 16.2) | 13.8 (11.6 - 16.3) |
| SIDIAP | 18,669 | 6,864 | 50.5 | 18.6 | 455 | 176 | 9.01 (8.21 - 9.88) | 9.48 (8.13 - 11.0) |
| Meta-analysis | 587,537 | 1,064,749 | 528.8 | 964.7 | 4,686 | 8,767 | 8.86 (8.61 - 9.12) | 9.09 (8.90 - 9.28) |
| Meta-analysis (w/o USOC) | 148,126 | 230,789 | 145.7 | 177.5 | 1,697 | 2,552 | 11.6 (11.1 - 12.2) | 14.4 (13.8 - 14.9) |
| **PS-matching / on-treatment TAR without escalation** | | | | | | | | |
| CCAE | 38,521 | 65,087 | 24.7 | 40.7 | 140 | 325 | 5.67 (4.77 - 6.69) | 7.99 (7.14 - 8.91) |
| DAG | 9,775 | 1,951 | 9.4 | 1.9 | 159 | 20 | 16.9 (14.3 - 19.7) | 10.7 (6.56 - 16.6) |
| ODOD | 26,208 | 57,432 | 17.1 | 43.5 | 367 | 981 | 21.5 (19.3 - 23.8) | 22.6 (21.2 - 24.0) |
| MDCR | 7,925 | 14,952 | 6.5 | 12.1 | 97 | 176 | 15.0 (12.2 - 18.3) | 14.5 (12.4 - 16.8) |
| MDCD | 7,174 | 16,221 | 3.9 | 8.7 | 65 | 155 | 16.7 (12.9 - 21.3) | 17.7 (15.0 - 20.7) |
| Open Claims | 429,599 | 833,862 | 295.7 | 662.8 | 2,271 | 5,177 | 7.68 (7.37 - 8.00) | 7.81 (7.60 - 8.03) |
| OEHR | 35,700 | 81,633 | 14.6 | 39.0 | 177 | 474 | 12.2 (10.4 - 14.1) | 12.1 (11.1 - 13.3) |
| VA | 1,735 | 9,218 | 1.0 | 6.1 | 12 | 73 | 11.5 (5.95 - 20.1) | 12.0 (9.41 - 15.1) |
| SIDIAP | 18,570 | 5,270 | 37.6 | 11.1 | 339 | 109 | 9.02 (8.08 - 10.0) | 9.86 (8.09 - 11.9) |
| Meta-analysis | 575,207 | 1,085,626 | 410.4 | 825.9 | 3,627 | 7,490 | 8.84 (8.55 - 9.13) | 9.07 (8.86 - 9.28) |
| Meta-analysis (w/o USOC) | 145,608 | 251,764 | 114.7 | 163.1 | 1,356 | 2,313 | 11.8 (11.2 - 12.5) | 14.2 (13.6 - 14.8) |
| **PS-stratification / on-treatment TAR without escalation** | | | | | | | | |
| CCAE | 44,818 | 67,007 | 29.0 | 41.7 | 167 | 328 | 5.75 (4.91 - 6.70) | 7.86 (7.03 - 8.76) |
| DAG | 9,799 | 2,266 | 9.4 | 2.2 | 159 | 26 | 16.8 (14.3 - 19.7) | 11.9 (7.76 - 17.4) |
| ODOD | 28,155 | 57,590 | 18.4 | 43.5 | 395 | 982 | 21.4 (19.4 - 23.6) | 22.6 (21.2 - 24.0) |
| MDCR | 8,538 | 15,344 | 7.0 | 12.5 | 104 | 181 | 14.9 (12.2 - 18.1) | 14.5 (12.5 - 16.8) |
| MDCD | 7,515 | 16,325 | 4.1 | 8.8 | 73 | 156 | 17.8 (13.9 - 22.3) | 17.8 (15.1 - 20.8) |
| Open Claims | 439,411 | 833,960 | 304.3 | 662.8 | 2,370 | 5,177 | 7.79 (7.48 - 8.11) | 7.81 (7.60 - 8.03) |
| OEHR | 38,147 | 81,718 | 15.8 | 39.1 | 193 | 474 | 12.2 (10.6 - 14.1) | 12.1 (11.1 - 13.3) |
| VA | 2,107 | 12,693 | 1.2 | 8.8 | 13 | 109 | 10.6 (5.66 - 18.2) | 12.4 (10.2 - 15.0) |
| SIDIAP | 18,669 | 6,864 | 37.7 | 15.0 | 341 | 147 | 9.05 (8.12 - 10.1) | 9.78 (8.26 - 11.5) |
| Meta-analysis | 587,537 | 1,064,749 | 421.7 | 816.9 | 3,729 | 7,315 | 8.84 (8.56 - 9.13) | 8.96 (8.75 - 9.16) |
| Meta-analysis (w/o USOC) | 148,126 | 230,789 | 117.3 | 154.0 | 1,359 | 2,138 | 11.6 (11.0 - 12.2) | 13.9 (13.3 - 14.5) |

##### 4.6.18 Hypoglycemia

**Supplementary Table 433:** Exposures, outcomes and risk estimates for Hypoglycemia in DPP4I (female) (T) vs SU (female) (C) new-user cohorts under four different propensity score (PS) adjustment / time-at-risk (TAR) designs across data sources. We report total patient exposure time in 1,000 patient-year (KYr) units, uncalibrated and calibrated hazard ratios (HRs) and their 95% confidence intervals (CIs). Greyed out entries did not pass study diagnostics and are included only for completeness.

| | Patients | | Exposure (KYr) | | Outcomes | | Incidence (per KYr) | |
|---|---|---|---|---|---|---|---|---|
| | T | C | T | C | T | C | T | C |
| **PS-matching / on-treatment TAR** | | | | | | | | |
| CCAE | 38,440 | 65,118 | 30.1 | 47.3 | 107 | 515 | 3.55 (2.91 - 4.29) | 10.9 (9.96 - 11.9) |
| DAG | 10,390 | 2,025 | 11.2 | 2.1 | 30 | 20 | 2.67 (1.80 - 3.81) | 9.43 (5.76 - 14.6) |
| ODOD | 27,017 | 59,247 | 21.4 | 50.9 | 165 | 1,320 | 7.72 (6.58 - 8.99) | 25.9 (24.5 - 27.4) |
| MDCR | 8,089 | 15,363 | 7.6 | 13.6 | 42 | 305 | 5.53 (3.99 - 7.47) | 22.3 (19.9 - 25.0) |
| MDCD | 7,197 | 16,312 | 5.1 | 10.6 | 60 | 340 | 11.8 (9.04 - 15.2) | 32.2 (28.8 - 35.8) |
| Open Claims | 431,606 | 837,656 | 376.5 | 794.4 | 1,208 | 8,598 | 3.21 (3.03 - 3.39) | 10.8 (10.6 - 11.1) |
| OEHR | 35,994 | 82,400 | 18.4 | 46.4 | 134 | 932 | 7.29 (6.11 - 8.64) | 20.1 (18.8 - 21.4) |
| VA | 1,785 | 9,513 | 1.3 | 7.3 | <10 | 92 | < 7.43 (3.56 - 13.7) | 12.6 (10.1 - 15.4) |
| SIDIAP | 19,478 | 5,435 | 53.6 | 14.1 | 50 | 66 | 0.93 (0.69 - 1.23) | 4.68 (3.62 - 5.95) |
| Meta-analysis | 579,996 | 1,093,069 | 525.2 | 986.8 | <1,806 | 12,188 | < 3.44 (3.28 - 3.60) | 12.4 (12.1 - 12.6) |
| Meta-analysis (w/o USOC) | 148,390 | 255,413 | 148.7 | 192.4 | <598 | 3,590 | < 4.02 (3.71 - 4.36) | 18.7 (18.1 - 19.3) |
| **PS-stratification / on-treatment TAR** | | | | | | | | |
| CCAE | 44,599 | 67,013 | 35.3 | 48.5 | 120 | 533 | 3.40 (2.82 - 4.07) | 11.0 (10.1 - 12.0) |
| DAG | 10,421 | 2,349 | 11.3 | 2.5 | 30 | 24 | 2.67 (1.80 - 3.80) | 9.58 (6.14 - 14.2) |
| ODOD | 28,947 | 59,399 | 23.0 | 50.9 | 182 | 1,322 | 7.91 (6.80 - 9.15) | 25.9 (24.6 - 27.4) |
| MDCR | 8,701 | 15,683 | 8.2 | 13.9 | 45 | 312 | 5.50 (4.01 - 7.36) | 22.4 (20.0 - 25.0) |
| MDCD | 7,505 | 16,411 | 5.3 | 10.6 | 60 | 341 | 11.3 (8.60 - 14.5) | 32.2 (28.9 - 35.8) |
| Open Claims | 441,252 | 837,752 | 387.3 | 794.5 | 1,271 | 8,599 | 3.28 (3.10 - 3.47) | 10.8 (10.6 - 11.1) |
| OEHR | 38,401 | 82,487 | 19.8 | 46.4 | 139 | 934 | 7.01 (5.89 - 8.27) | 20.1 (18.9 - 21.5) |
| VA | 2,176 | 12,984 | 1.6 | 10.4 | <10 | 138 | < 6.27 (3.01 - 11.5) | 13.3 (11.2 - 15.7) |
| SIDIAP | 19,590 | 7,064 | 53.7 | 19.3 | 50 | 84 | 0.93 (0.69 - 1.23) | 4.36 (3.47 - 5.39) |
| Meta-analysis | 591,911 | 1,071,747 | 538.5 | 976.0 | 1,837 | 11,808 | 3.41 (3.26 - 3.57) | 12.1 (11.9 - 12.3) |
| Meta-analysis (w/o USOC) | 150,659 | 233,995 | 151.2 | 181.6 | 566 | 3,209 | 3.74 (3.44 - 4.06) | 17.7 (17.1 - 18.3) |
| **PS-matching / on-treatment TAR without escalation** | | | | | | | | |
| CCAE | 38,440 | 65,118 | 24.7 | 40.8 | 60 | 430 | 2.43 (1.85 - 3.12) | 10.5 (9.56 - 11.6) |
| DAG | 10,390 | 2,025 | 10.1 | 2.0 | 26 | 20 | 2.56 (1.67 - 3.75) | 10.2 (6.23 - 15.8) |
| ODOD | 27,017 | 59,247 | 18.0 | 45.4 | 89 | 1,159 | 4.95 (3.98 - 6.09) | 25.6 (24.1 - 27.1) |
| MDCR | 8,089 | 15,363 | 6.6 | 12.5 | 26 | 276 | 3.93 (2.57 - 5.76) | 22.1 (19.6 - 24.8) |
| MDCD | 7,197 | 16,312 | 4.0 | 8.8 | 33 | 275 | 8.34 (5.74 - 11.7) | 31.2 (27.6 - 35.1) |
| Open Claims | 431,606 | 837,656 | 299.2 | 669.1 | 657 | 7,126 | 2.20 (2.03 - 2.37) | 10.6 (10.4 - 10.9) |
| OEHR | 35,994 | 82,400 | 14.8 | 39.5 | 63 | 686 | 4.26 (3.27 - 5.45) | 17.4 (16.1 - 18.7) |
| VA | 1,785 | 9,513 | 1.1 | 6.3 | <10 | 67 | < 9.28 (4.45 - 17.1) | 10.6 (8.19 - 13.4) |
| SIDIAP | 19,478 | 5,435 | 39.9 | 11.5 | 14 | 51 | 0.35 (0.19 - 0.59) | 4.43 (3.30 - 5.82) |
| Meta-analysis | 579,996 | 1,093,069 | 418.4 | 835.9 | <978 | 10,090 | < 2.34 (2.19 - 2.49) | 12.1 (11.8 - 12.3) |
| Meta-analysis (w/o USOC) | 148,390 | 255,413 | 119.2 | 166.8 | <321 | 2,964 | < 2.69 (2.41 - 3.01) | 17.8 (17.1 - 18.4) |
| **PS-stratification / on-treatment TAR without escalation** | | | | | | | | |
| CCAE | 44,599 | 67,013 | 29.0 | 41.9 | 70 | 447 | 2.41 (1.88 - 3.05) | 10.7 (9.71 - 11.7) |
| DAG | 10,421 | 2,349 | 10.2 | 2.3 | 26 | 24 | 2.56 (1.67 - 3.75) | 10.4 (6.68 - 15.5) |
| ODOD | 28,947 | 59,399 | 19.3 | 45.4 | 101 | 1,161 | 5.22 (4.25 - 6.35) | 25.6 (24.1 - 27.1) |
| MDCR | 8,701 | 15,683 | 7.1 | 12.8 | 28 | 280 | 3.94 (2.62 - 5.69) | 21.9 (19.4 - 24.7) |
| MDCD | 7,505 | 16,411 | 4.2 | 8.8 | 33 | 276 | 7.94 (5.47 - 11.2) | 31.2 (27.6 - 35.1) |
| Open Claims | 441,252 | 837,752 | 307.9 | 669.2 | 691 | 7,127 | 2.24 (2.08 - 2.42) | 10.7 (10.4 - 10.9) |
| OEHR | 38,401 | 82,487 | 16.0 | 39.5 | 66 | 687 | 4.13 (3.19 - 5.25) | 17.4 (16.1 - 18.7) |
| VA | 2,176 | 12,984 | 1.3 | 9.1 | <10 | 104 | < 7.80 (3.74 - 14.4) | 11.5 (9.38 - 13.9) |
| SIDIAP | 19,590 | 7,064 | 40.0 | 15.6 | 14 | 63 | 0.35 (0.19 - 0.59) | 4.03 (3.10 - 5.16) |
| Meta-analysis | 591,911 | 1,071,747 | 429.5 | 826.6 | 996 | 9,789 | 2.32 (2.18 - 2.47) | 11.8 (11.6 - 12.1) |
| Meta-analysis (w/o USOC) | 150,659 | 233,995 | 121.6 | 157.5 | 305 | 2,662 | 2.51 (2.23 - 2.81) | 16.9 (16.3 - 17.6) |

##### 4.6.19 Hypotension

**Supplementary Table 434:** Exposures, outcomes and risk estimates for Hypotension in DPP4I (female) (T) vs SU (female) (C) new-user cohorts under four different propensity score (PS) adjustment / time-at-risk (TAR) designs across data sources. We report total patient exposure time in 1,000 patient-year (KYr) units, uncalibrated and calibrated hazard ratios (HRs) and their 95% confidence intervals (CIs). Greyed out entries did not pass study diagnostics and are included only for completeness.

| | Patients | | Exposure (KYr) | | Outcomes | | Incidence (per KYr) | |
|---|---|---|---|---|---|---|---|---|
| | T | C | T | C | T | C | T | C |
| **PS-matching / on-treatment TAR** | | | | | | | | |
| CCAE | 38,381 | 64,846 | 30.0 | 47.2 | 211 | 343 | 7.03 (6.11 - 8.04) | 7.27 (6.52 - 8.08) |
| DAG | 10,116 | 1,972 | 10.9 | 2.0 | 69 | 6 | 6.32 (4.92 - 8.00) | 2.93 (1.08 - 6.38) |
| ODOD | 26,107 | 57,157 | 20.4 | 48.9 | 448 | 1,102 | 21.9 (20.0 - 24.1) | 22.5 (21.2 - 23.9) |
| MDCR | 7,752 | 14,741 | 7.3 | 13.0 | 159 | 297 | 21.9 (18.6 - 25.6) | 22.8 (20.3 - 25.5) |
| MDCD | 6,891 | 15,590 | 4.8 | 10.1 | 138 | 285 | 28.8 (24.2 - 34.0) | 28.2 (25.1 - 31.7) |
| Open Claims | 423,139 | 822,249 | 367.3 | 778.9 | 3,518 | 6,996 | 9.58 (9.26 - 9.90) | 8.98 (8.77 - 9.19) |
| OEHR | 35,607 | 81,377 | 18.0 | 45.6 | 262 | 715 | 14.5 (12.8 - 16.4) | 15.7 (14.5 - 16.9) |
| VA | 1,711 | 9,192 | 1.3 | 7.0 | 12 | 65 | 9.33 (4.82 - 16.3) | 9.24 (7.13 - 11.8) |
| SIDIAP | 19,382 | 5,423 | 53.3 | 14.2 | 95 | 18 | 1.78 (1.44 - 2.18) | 1.27 (0.75 - 2.01) |
| Meta-analysis | 569,086 | 1,072,547 | 513.3 | 967.1 | 4,912 | 9,827 | 9.57 (9.30 - 9.84) | 10.2 (9.96 - 10.4) |
| Meta-analysis (w/o USOC) | 145,947 | 250,298 | 146.0 | 188.2 | 1,394 | 2,831 | 9.55 (9.05 - 10.1) | 15.0 (14.5 - 15.6) |
| **PS-stratification / on-treatment TAR** | | | | | | | | |
| CCAE | 44,624 | 66,758 | 35.2 | 48.4 | 257 | 351 | 7.30 (6.44 - 8.25) | 7.26 (6.52 - 8.06) |
| DAG | 10,145 | 2,280 | 10.9 | 2.4 | 69 | 9 | 6.31 (4.91 - 7.98) | 3.72 (1.70 - 7.07) |
| ODOD | 28,041 | 57,316 | 22.0 | 49.0 | 486 | 1,103 | 22.1 (20.1 - 24.1) | 22.5 (21.2 - 23.9) |
| MDCR | 8,338 | 15,049 | 7.8 | 13.3 | 172 | 301 | 21.9 (18.8 - 25.5) | 22.6 (20.1 - 25.3) |
| MDCD | 7,199 | 15,691 | 5.0 | 10.1 | 153 | 287 | 30.4 (25.7 - 35.6) | 28.4 (25.2 - 31.8) |
| Open Claims | 432,724 | 822,422 | 377.9 | 778.9 | 3,664 | 6,996 | 9.70 (9.38 - 10.0) | 8.98 (8.77 - 9.19) |
| OEHR | 38,051 | 81,463 | 19.5 | 45.7 | 287 | 717 | 14.7 (13.1 - 16.5) | 15.7 (14.6 - 16.9) |
| VA | 2,102 | 12,553 | 1.5 | 10.0 | 15 | 100 | 9.79 (5.48 - 16.1) | 10.0 (8.15 - 12.2) |
| SIDIAP | 19,493 | 7,053 | 53.4 | 19.3 | 95 | 20 | 1.78 (1.44 - 2.18) | 1.03 (0.63 - 1.60) |
| Meta-analysis | 581,416 | 1,052,341 | 526.7 | 957.0 | 5,030 | 9,497 | 9.55 (9.29 - 9.82) | 9.92 (9.72 - 10.1) |
| Meta-analysis (w/o USOC) | 148,692 | 229,919 | 148.9 | 178.1 | 1,366 | 2,501 | 9.18 (8.70 - 9.68) | 14.0 (13.5 - 14.6) |
| **PS-matching / on-treatment TAR without escalation** | | | | | | | | |
| CCAE | 38,381 | 64,846 | 24.7 | 40.7 | 164 | 282 | 6.65 (5.67 - 7.75) | 6.93 (6.15 - 7.79) |
| DAG | 10,116 | 1,972 | 9.9 | 1.9 | 56 | 6 | 5.67 (4.29 - 7.37) | 3.16 (1.16 - 6.89) |
| ODOD | 26,107 | 57,157 | 17.2 | 43.6 | 383 | 971 | 22.3 (20.1 - 24.6) | 22.3 (20.9 - 23.7) |
| MDCR | 7,752 | 14,741 | 6.3 | 12.0 | 139 | 266 | 22.0 (18.5 - 25.9) | 22.2 (19.6 - 25.1) |
| MDCD | 6,891 | 15,590 | 3.8 | 8.4 | 91 | 224 | 24.2 (19.5 - 29.7) | 26.6 (23.2 - 30.3) |
| Open Claims | 423,139 | 822,249 | 291.9 | 655.8 | 2,798 | 5,815 | 9.59 (9.23 - 9.95) | 8.87 (8.64 - 9.10) |
| OEHR | 35,607 | 81,377 | 14.5 | 38.9 | 178 | 499 | 12.2 (10.5 - 14.2) | 12.8 (11.7 - 14.0) |
| VA | 1,711 | 9,192 | 1.0 | 6.1 | <10 | 57 | < 9.80 (4.70 - 18.0) | 9.35 (7.08 - 12.1) |
| SIDIAP | 19,382 | 5,423 | 39.6 | 11.5 | 72 | 13 | 1.82 (1.42 - 2.29) | 1.13 (0.60 - 1.93) |
| Meta-analysis | 569,086 | 1,072,547 | 408.9 | 818.9 | <3,891 | 8,133 | < 9.52 (9.22 - 9.82) | 9.93 (9.72 - 10.1) |
| Meta-analysis (w/o USOC) | 145,947 | 250,298 | 117.0 | 163.1 | <1,093 | 2,318 | < 9.34 (8.80 - 9.91) | 14.2 (13.6 - 14.8) |
| **PS-stratification / on-treatment TAR without escalation** | | | | | | | | |
| CCAE | 44,624 | 66,758 | 29.0 | 41.7 | 202 | 289 | 6.98 (6.05 - 8.01) | 6.92 (6.15 - 7.77) |
| DAG | 10,145 | 2,280 | 9.9 | 2.2 | 56 | 9 | 5.66 (4.28 - 7.35) | 4.05 (1.85 - 7.68) |
| ODOD | 28,041 | 57,316 | 18.5 | 43.6 | 410 | 972 | 22.1 (20.0 - 24.4) | 22.3 (20.9 - 23.7) |
| MDCR | 8,338 | 15,049 | 6.8 | 12.2 | 151 | 269 | 22.1 (18.7 - 25.9) | 22.0 (19.5 - 24.8) |
| MDCD | 7,199 | 15,691 | 4.0 | 8.4 | 104 | 226 | 26.2 (21.4 - 31.8) | 26.8 (23.4 - 30.5) |
| Open Claims | 432,724 | 822,422 | 300.4 | 655.8 | 2,917 | 5,815 | 9.71 (9.36 - 10.1) | 8.87 (8.64 - 9.10) |
| OEHR | 38,051 | 81,463 | 15.8 | 38.9 | 200 | 501 | 12.7 (11.0 - 14.6) | 12.9 (11.8 - 14.0) |
| VA | 2,102 | 12,553 | 1.2 | 8.7 | 12 | 85 | 9.84 (5.08 - 17.2) | 9.75 (7.79 - 12.1) |
| SIDIAP | 19,493 | 7,053 | 39.7 | 15.7 | 72 | 13 | 1.81 (1.42 - 2.28) | 0.83 (0.44 - 1.42) |
| Meta-analysis | 581,416 | 1,052,341 | 420.0 | 810.2 | 4,008 | 7,868 | 9.54 (9.25 - 9.84) | 9.71 (9.50 - 9.93) |
| Meta-analysis (w/o USOC) | 148,692 | 229,919 | 119.7 | 154.4 | 1,091 | 2,053 | 9.12 (8.58 - 9.67) | 13.3 (12.7 - 13.9) |

##### 4.6.20 Joint pain

**Supplementary Table 435:** Exposures, outcomes and risk estimates for Joint pain in DPP4I (female) (T) vs SU (female) (C) new-user cohorts under four different propensity score (PS) adjustment / time-at-risk (TAR) designs across data sources. We report total patient exposure time in 1,000 patient-year (KYr) units, uncalibrated and calibrated hazard ratios (HRs) and their 95% confidence intervals (CIs). Greyed out entries did not pass study diagnostics and are included only for completeness.

| | Patients | | Exposure (KYr) | | Outcomes | | Incidence (per KYr) | |
|---|---|---|---|---|---|---|---|---|
| | T | C | T | C | T | C | T | C |
| **PS-matching / on-treatment TAR** | | | | | | | | |
| CCAE | 36,500 | 61,985 | 28.4 | 44.9 | 396 | 596 | 13.9 (12.6 - 15.4) | 13.3 (12.2 - 14.4) |
| DAG | 8,831 | 1,761 | 9.3 | 1.8 | 212 | 50 | 22.8 (19.8 - 26.1) | 28.0 (20.8 - 36.9) |
| ODOD | 25,431 | 56,095 | 19.9 | 48.3 | 385 | 814 | 19.3 (17.4 - 21.3) | 16.9 (15.7 - 18.1) |
| MDCR | 7,747 | 14,779 | 7.2 | 13.1 | 91 | 134 | 12.6 (10.1 - 15.4) | 10.2 (8.54 - 12.1) |
| MDCD | 6,596 | 15,176 | 4.5 | 9.8 | 143 | 230 | 31.6 (26.6 - 37.2) | 23.4 (20.5 - 26.7) |
| Open Claims | 406,341 | 794,621 | 352.7 | 754.5 | 3,374 | 6,218 | 9.57 (9.25 - 9.90) | 8.24 (8.04 - 8.45) |
| OEHR | 34,136 | 78,457 | 17.2 | 43.8 | 279 | 742 | 16.3 (14.4 - 18.3) | 16.9 (15.7 - 18.2) |
| VA | 1,424 | 7,508 | 1.1 | 5.7 | 10 | 79 | 9.15 (4.39 - 16.8) | 13.9 (11.0 - 17.3) |
| SIDIAP | 15,176 | 4,670 | 40.4 | 11.6 | 1,166 | 344 | 28.8 (27.2 - 30.5) | 29.7 (26.7 - 33.0) |
| Meta-analysis | 542,182 | 1,035,052 | 480.8 | 933.5 | 6,056 | 9,207 | 12.6 (12.3 - 12.9) | 9.86 (9.66 - 10.1) |
| Meta-analysis (w/o USOC) | 135,841 | 240,431 | 128.1 | 179.0 | 2,682 | 2,989 | 20.9 (20.1 - 21.7) | 16.7 (16.1 - 17.3) |
| **PS-stratification / on-treatment TAR** | | | | | | | | |
| CCAE | 42,281 | 63,796 | 33.2 | 46.0 | 487 | 601 | 14.7 (13.4 - 16.0) | 13.1 (12.0 - 14.2) |
| DAG | 8,855 | 2,055 | 9.3 | 2.1 | 213 | 61 | 22.8 (19.9 - 26.1) | 28.8 (22.0 - 37.0) |
| ODOD | 27,243 | 56,249 | 21.4 | 48.3 | 412 | 815 | 19.2 (17.4 - 21.2) | 16.9 (15.7 - 18.1) |
| MDCR | 8,335 | 15,094 | 7.8 | 13.4 | 100 | 135 | 12.8 (10.4 - 15.5) | 10.1 (8.43 - 11.9) |
| MDCD | 6,865 | 15,272 | 4.7 | 9.8 | 148 | 230 | 31.3 (26.4 - 36.7) | 23.4 (20.5 - 26.6) |
| Open Claims | 415,068 | 794,718 | 362.4 | 754.6 | 3,534 | 6,218 | 9.75 (9.43 - 10.1) | 8.24 (8.04 - 8.45) |
| OEHR | 36,357 | 78,542 | 18.5 | 43.8 | 310 | 742 | 16.8 (14.9 - 18.7) | 16.9 (15.7 - 18.2) |
| VA | 1,694 | 10,443 | 1.3 | 8.3 | 10 | 140 | 7.95 (3.81 - 14.6) | 16.9 (14.2 - 20.0) |
| SIDIAP | 15,301 | 6,185 | 40.5 | 16.0 | 1,167 | 465 | 28.8 (27.2 - 30.5) | 29.1 (26.5 - 31.8) |
| Meta-analysis | 553,440 | 1,016,639 | 493.2 | 924.3 | 6,223 | 9,037 | 12.6 (12.3 - 12.9) | 9.78 (9.58 - 9.98) |
| Meta-analysis (w/o USOC) | 138,372 | 221,921 | 130.8 | 169.7 | 2,689 | 2,819 | 20.6 (19.8 - 21.3) | 16.6 (16.0 - 17.2) |
| **PS-matching / on-treatment TAR without escalation** | | | | | | | | |
| CCAE | 36,500 | 61,985 | 23.4 | 38.7 | 328 | 512 | 14.0 (12.5 - 15.6) | 13.2 (12.1 - 14.4) |
| DAG | 8,831 | 1,761 | 8.4 | 1.7 | 198 | 47 | 23.6 (20.4 - 27.1) | 28.4 (20.9 - 37.8) |
| ODOD | 25,431 | 56,095 | 16.8 | 43.0 | 322 | 734 | 19.2 (17.2 - 21.4) | 17.1 (15.8 - 18.3) |
| MDCR | 7,747 | 14,779 | 6.3 | 12.0 | 79 | 122 | 12.5 (9.93 - 15.6) | 10.1 (8.42 - 12.1) |
| MDCD | 6,596 | 15,176 | 3.5 | 8.2 | 110 | 193 | 31.0 (25.5 - 37.4) | 23.6 (20.3 - 27.1) |
| Open Claims | 406,341 | 794,621 | 280.4 | 636.0 | 2,679 | 5,207 | 9.55 (9.20 - 9.92) | 8.19 (7.97 - 8.41) |
| OEHR | 34,136 | 78,457 | 13.8 | 37.3 | 220 | 602 | 15.9 (13.9 - 18.1) | 16.1 (14.9 - 17.5) |
| VA | 1,424 | 7,508 | 0.9 | 4.9 | <10 | 67 | < 11.5 (5.50 - 21.1) | 13.5 (10.5 - 17.2) |
| SIDIAP | 15,176 | 4,670 | 30.1 | 9.6 | 859 | 278 | 28.5 (26.6 - 30.5) | 29.1 (25.8 - 32.7) |
| Meta-analysis | 542,182 | 1,035,052 | 383.7 | 791.5 | <4,805 | 7,762 | < 12.5 (12.2 - 12.9) | 9.81 (9.59 - 10.0) |
| Meta-analysis (w/o USOC) | 135,841 | 240,431 | 103.3 | 155.5 | <2,126 | 2,555 | < 20.6 (19.7 - 21.5) | 16.4 (15.8 - 17.1) |
| **PS-stratification / on-treatment TAR without escalation** | | | | | | | | |
| CCAE | 42,281 | 63,796 | 27.4 | 39.7 | 402 | 517 | 14.7 (13.3 - 16.2) | 13.0 (11.9 - 14.2) |
| DAG | 8,855 | 2,055 | 8.4 | 2.0 | 199 | 58 | 23.6 (20.5 - 27.1) | 29.7 (22.6 - 38.4) |
| ODOD | 27,243 | 56,249 | 18.0 | 43.1 | 345 | 734 | 19.2 (17.2 - 21.3) | 17.0 (15.8 - 18.3) |
| MDCR | 8,335 | 15,094 | 6.8 | 12.3 | 87 | 123 | 12.8 (10.2 - 15.7) | 10.00 (8.31 - 11.9) |
| MDCD | 6,865 | 15,272 | 3.7 | 8.2 | 115 | 193 | 31.1 (25.7 - 37.4) | 23.5 (20.3 - 27.0) |
| Open Claims | 415,068 | 794,718 | 288.2 | 636.0 | 2,807 | 5,207 | 9.74 (9.38 - 10.1) | 8.19 (7.97 - 8.41) |
| OEHR | 36,357 | 78,542 | 14.9 | 37.4 | 241 | 602 | 16.1 (14.2 - 18.3) | 16.1 (14.8 - 17.4) |
| VA | 1,694 | 10,443 | 1.0 | 7.3 | <10 | 121 | < 9.94 (4.76 - 18.3) | 16.7 (13.8 - 19.9) |
| SIDIAP | 15,301 | 6,185 | 30.2 | 13.1 | 860 | 376 | 28.5 (26.6 - 30.4) | 28.7 (25.9 - 31.8) |
| Meta-analysis | 553,440 | 1,016,639 | 394.0 | 783.5 | 4,941 | 7,617 | 12.5 (12.2 - 12.9) | 9.72 (9.50 - 9.94) |
| Meta-analysis (w/o USOC) | 138,372 | 221,921 | 105.8 | 147.5 | 2,134 | 2,410 | 20.2 (19.3 - 21.0) | 16.3 (15.7 - 17.0) |

##### 4.6.21 Lower extremity amputation

**Supplementary Table 436:** Exposures, outcomes and risk estimates for Lower extremity amputation in DPP4I (female) (T) vs SU (female) (C) new-user cohorts under four different propensity score (PS) adjustment / time-at-risk (TAR) designs across data sources. We report total patient exposure time in 1,000 patient-year (KYr) units, uncalibrated and calibrated hazard ratios (HRs) and their 95% confidence intervals (CIs). Greyed out entries did not pass study diagnostics and are included only for completeness.

| | Patients | | Exposure (KYr) | | Outcomes | | Incidence (per KYr) | |
|---|---|---|---|---|---|---|---|---|
| | T | C | T | C | T | C | T | C |
| **PS-matching / on-treatment TAR** | | | | | | | | |
| CCAE | 39,120 | 66,027 | 30.7 | 48.2 | 9 | 15 | 0.29 (0.13 - 0.56) | 0.31 (0.17 - 0.51) |
| DAG | 10,485 | 2,040 | 11.4 | 2.1 | 0 | 0 | | |
| ODOD | 27,477 | 60,143 | 21.8 | 52.3 | 10 | 39 | 0.46 (0.22 - 0.84) | 0.75 (0.53 - 1.02) |
| MDCR | 8,154 | 15,485 | 7.7 | 13.8 | 6 | 9 | 0.78 (0.29 - 1.70) | 0.65 (0.30 - 1.24) |
| MDCD | 7,409 | 16,719 | 5.2 | 11.0 | <5 | 8 | < 0.95 (0.31 - 2.22) | 0.73 (0.31 - 1.44) |
| Open Claims | 439,414 | 851,282 | 383.8 | 810.7 | 106 | 331 | 0.28 (0.23 - 0.33) | 0.41 (0.37 - 0.45) |
| OEHR | 36,406 | 83,227 | 18.7 | 47.4 | 7 | 28 | 0.37 (0.15 - 0.77) | 0.59 (0.39 - 0.85) |
| VA | 1,820 | 9,660 | 1.4 | 7.5 | 0 | <10 | | < 1.34 (0.64 - 2.46) |
| SIDIAP | 19,504 | 5,435 | 53.7 | 14.2 | 8 | <5 | 0.15 (0.06 - 0.29) | < 0.35 (0.11 - 0.82) |
| Meta-analysis | 542,417 | 1,060,679 | 455.0 | 958.5 | 132 | 413 | 0.29 (0.24 - 0.34) | 0.43 (0.39 - 0.47) |
| Meta-analysis (w/o USOC) | 103,003 | 209,397 | 71.1 | 147.9 | 26 | 82 | 0.37 (0.24 - 0.54) | 0.55 (0.44 - 0.69) |
| **PS-stratification / on-treatment TAR** | | | | | | | | |
| CCAE | 45,481 | 67,959 | 36.0 | 49.4 | 10 | 15 | 0.28 (0.13 - 0.51) | 0.30 (0.17 - 0.50) |
| DAG | 10,516 | 2,367 | 11.4 | 2.5 | 0 | 0 | | |
| ODOD | 29,479 | 60,301 | 23.5 | 52.3 | 10 | 39 | 0.43 (0.20 - 0.78) | 0.75 (0.53 - 1.02) |
| MDCR | 8,789 | 15,805 | 8.3 | 14.1 | 6 | 9 | 0.72 (0.27 - 1.57) | 0.64 (0.29 - 1.21) |
| MDCD | 7,756 | 16,825 | 5.5 | 11.0 | 5 | 8 | 0.90 (0.29 - 2.11) | 0.73 (0.31 - 1.43) |
| Open Claims | 449,397 | 851,380 | 395.0 | 810.7 | 110 | 331 | 0.28 (0.23 - 0.34) | 0.41 (0.37 - 0.45) |
| OEHR | 38,885 | 83,316 | 20.2 | 47.4 | 7 | 28 | 0.35 (0.14 - 0.71) | 0.59 (0.39 - 0.85) |
| VA | 2,218 | 13,208 | 1.6 | 10.6 | 0 | <10 | | < 0.94 (0.45 - 1.73) |
| SIDIAP | 19,617 | 7,063 | 53.8 | 19.4 | 8 | <5 | 0.15 (0.06 - 0.29) | < 0.26 (0.08 - 0.60) |
| Meta-analysis | 563,242 | 1,062,956 | 474.7 | 959.8 | 137 | 413 | 0.29 (0.24 - 0.34) | 0.43 (0.39 - 0.47) |
| Meta-analysis (w/o USOC) | 113,845 | 211,576 | 79.7 | 149.1 | 27 | 82 | 0.34 (0.22 - 0.49) | 0.55 (0.44 - 0.68) |
| **PS-matching / on-treatment TAR without escalation** | | | | | | | | |
| CCAE | 39,120 | 66,027 | 25.2 | 41.5 | 8 | 13 | 0.32 (0.14 - 0.63) | 0.31 (0.17 - 0.54) |
| DAG | 10,485 | 2,040 | 10.3 | 2.0 | 0 | 0 | | |
| ODOD | 27,477 | 60,143 | 18.3 | 46.5 | 8 | 32 | 0.44 (0.19 - 0.86) | 0.69 (0.47 - 0.97) |
| MDCR | 8,154 | 15,485 | 6.7 | 12.7 | 6 | 6 | 0.90 (0.33 - 1.96) | 0.47 (0.17 - 1.03) |
| MDCD | 7,409 | 16,719 | 4.1 | 9.1 | <5 | 5 | < 1.22 (0.40 - 2.85) | 0.55 (0.18 - 1.28) |
| Open Claims | 439,414 | 851,282 | 304.7 | 681.8 | 84 | 252 | 0.28 (0.22 - 0.34) | 0.37 (0.33 - 0.42) |
| OEHR | 36,406 | 83,227 | 15.0 | 40.2 | <5 | 16 | < 0.33 (0.11 - 0.78) | 0.40 (0.23 - 0.65) |
| VA | 1,820 | 9,660 | 1.1 | 6.5 | 0 | <10 | | < 1.55 (0.74 - 2.85) |
| SIDIAP | 19,504 | 5,435 | 39.9 | 11.6 | 6 | <5 | 0.15 (0.06 - 0.33) | < 0.43 (0.14 - 1.01) |
| Meta-analysis | 542,417 | 1,060,679 | 363.1 | 810.0 | <105 | 313 | < 0.29 (0.24 - 0.35) | 0.39 (0.34 - 0.43) |
| Meta-analysis (w/o USOC) | 103,003 | 209,397 | 58.5 | 128.2 | <21 | 61 | < 0.36 (0.22 - 0.55) | 0.48 (0.36 - 0.61) |
| **PS-stratification / on-treatment TAR without escalation** | | | | | | | | |
| CCAE | 45,481 | 67,959 | 29.6 | 42.6 | 9 | 13 | 0.30 (0.14 - 0.58) | 0.31 (0.16 - 0.52) |
| DAG | 10,516 | 2,367 | 10.3 | 2.3 | 0 | 0 | | |
| ODOD | 29,479 | 60,301 | 19.7 | 46.5 | 8 | 32 | 0.41 (0.18 - 0.80) | 0.69 (0.47 - 0.97) |
| MDCR | 8,789 | 15,805 | 7.2 | 12.9 | 6 | 6 | 0.83 (0.31 - 1.81) | 0.46 (0.17 - 1.01) |
| MDCD | 7,756 | 16,825 | 4.3 | 9.1 | <5 | 5 | < 1.16 (0.38 - 2.71) | 0.55 (0.18 - 1.28) |
| Open Claims | 449,397 | 851,380 | 313.6 | 681.8 | 87 | 252 | 0.28 (0.22 - 0.34) | 0.37 (0.33 - 0.42) |
| OEHR | 38,885 | 83,316 | 16.3 | 40.2 | <5 | 16 | < 0.31 (0.10 - 0.72) | 0.40 (0.23 - 0.65) |
| VA | 2,218 | 13,208 | 1.3 | 9.3 | 0 | <10 | | < 1.08 (0.52 - 1.99) |
| SIDIAP | 19,617 | 7,063 | 40.0 | 15.7 | 6 | <5 | 0.15 (0.06 - 0.33) | < 0.32 (0.10 - 0.74) |
| Meta-analysis | 563,242 | 1,062,956 | 379.1 | 811.1 | <109 | 313 | < 0.29 (0.24 - 0.35) | 0.39 (0.34 - 0.43) |
| Meta-analysis (w/o USOC) | 113,845 | 211,576 | 65.5 | 129.3 | <22 | 61 | < 0.34 (0.21 - 0.51) | 0.47 (0.36 - 0.61) |

##### 4.6.22 Nausea

**Supplementary Table 437:** Exposures, outcomes and risk estimates for Nausea in DPP4I (female) (T) vs SU (female) (C) new-user cohorts under four different propensity score (PS) adjustment / time-at-risk (TAR) designs across data sources. We report total patient exposure time in 1,000 patient-year (KYr) units, uncalibrated and calibrated hazard ratios (HRs) and their 95% confidence intervals (CIs). Greyed out entries did not pass study diagnostics and are included only for completeness.

| | Patients | | Exposure (KYr) | | Outcomes | | Incidence (per KYr) | |
|---|---|---|---|---|---|---|---|---|
| | T | C | T | C | T | C | T | C |
| **PS-matching / on-treatment TAR** | | | | | | | | |
| CCAE | 33,169 | 56,168 | 25.6 | 40.6 | 1,088 | 1,724 | 42.4 (40.0 - 45.0) | 42.5 (40.5 - 44.5) |
| DAG | 9,491 | 1,839 | 10.2 | 1.9 | 150 | 24 | 14.8 (12.5 - 17.3) | 12.5 (8.02 - 18.6) |
| ODOD | 22,223 | 48,605 | 17.1 | 41.0 | 924 | 2,379 | 54.0 (50.6 - 57.6) | 58.0 (55.7 - 60.4) |
| MDCR | 7,015 | 13,103 | 6.5 | 11.4 | 289 | 541 | 44.3 (39.3 - 49.7) | 47.4 (43.5 - 51.5) |
| MDCD | 4,961 | 11,206 | 3.3 | 7.1 | 387 | 724 | 117 (106 - 129) | 101 (94.1 - 109) |
| Open Claims | 358,720 | 698,954 | 310.6 | 663.8 | 9,048 | 19,011 | 29.1 (28.5 - 29.7) | 28.6 (28.2 - 29.0) |
| OEHR | 31,233 | 71,686 | 15.4 | 38.8 | 774 | 2,205 | 50.3 (46.8 - 53.9) | 56.8 (54.5 - 59.3) |
| VA | 1,492 | 7,880 | 1.1 | 6.0 | 29 | 141 | 26.3 (17.6 - 37.7) | 23.7 (19.9 - 27.9) |
| SIDIAP | 18,524 | 5,273 | 50.9 | 13.7 | 448 | 116 | 8.80 (8.01 - 9.66) | 8.49 (7.02 - 10.2) |
| Meta-analysis | 486,828 | 914,714 | 440.7 | 824.3 | 13,137 | 26,865 | 29.8 (29.3 - 30.3) | 32.6 (32.2 - 33.0) |
| Meta-analysis (w/o USOC) | 128,108 | 215,760 | 130.1 | 160.5 | 4,089 | 7,854 | 31.4 (30.5 - 32.4) | 48.9 (47.9 - 50.0) |
| **PS-stratification / on-treatment TAR** | | | | | | | | |
| CCAE | 38,599 | 57,863 | 30.0 | 41.6 | 1,285 | 1,755 | 42.8 (40.5 - 45.2) | 42.2 (40.2 - 44.2) |
| DAG | 9,527 | 2,130 | 10.2 | 2.3 | 150 | 30 | 14.7 (12.5 - 17.3) | 13.3 (8.94 - 18.9) |
| ODOD | 23,886 | 48,728 | 18.5 | 41.0 | 1,003 | 2,384 | 54.3 (51.0 - 57.7) | 58.1 (55.8 - 60.5) |
| MDCR | 7,552 | 13,481 | 7.0 | 11.8 | 317 | 552 | 45.2 (40.4 - 50.5) | 47.0 (43.1 - 51.1) |
| MDCD | 5,142 | 11,261 | 3.4 | 7.2 | 404 | 727 | 117 (106 - 129) | 102 (94.3 - 109) |
| Open Claims | 367,392 | 699,041 | 320.0 | 663.8 | 9,435 | 19,015 | 29.5 (28.9 - 30.1) | 28.6 (28.2 - 29.1) |
| OEHR | 33,348 | 71,751 | 16.7 | 38.8 | 842 | 2,206 | 50.6 (47.2 - 54.1) | 56.8 (54.5 - 59.2) |
| VA | 1,778 | 11,035 | 1.3 | 8.7 | 33 | 223 | 25.8 (17.7 - 36.2) | 25.6 (22.4 - 29.2) |
| SIDIAP | 18,621 | 6,878 | 51.0 | 18.7 | 449 | 152 | 8.81 (8.01 - 9.66) | 8.13 (6.89 - 9.53) |
| Meta-analysis | 498,925 | 899,872 | 453.3 | 818.1 | 13,481 | 26,094 | 29.7 (29.2 - 30.2) | 31.9 (31.5 - 32.3) |
| Meta-analysis (w/o USOC) | 131,533 | 200,831 | 133.3 | 154.2 | 4,046 | 7,079 | 30.3 (29.4 - 31.3) | 45.9 (44.8 - 47.0) |
| **PS-matching / on-treatment TAR without escalation** | | | | | | | | |
| CCAE | 33,169 | 56,168 | 21.2 | 35.3 | 892 | 1,457 | 42.0 (39.3 - 44.9) | 41.3 (39.2 - 43.5) |
| DAG | 9,491 | 1,839 | 9.2 | 1.8 | 135 | 21 | 14.7 (12.3 - 17.4) | 11.8 (7.33 - 18.1) |
| ODOD | 22,223 | 48,605 | 14.5 | 36.7 | 780 | 2,119 | 53.7 (50.0 - 57.6) | 57.7 (55.2 - 60.2) |
| MDCR | 7,015 | 13,103 | 5.7 | 10.5 | 244 | 486 | 43.0 (37.8 - 48.7) | 46.3 (42.2 - 50.6) |
| MDCD | 4,961 | 11,206 | 2.7 | 6.0 | 303 | 618 | 114 (101 - 127) | 102 (94.3 - 111) |
| Open Claims | 358,720 | 698,954 | 248.4 | 563.0 | 7,180 | 15,737 | 28.9 (28.2 - 29.6) | 28.0 (27.5 - 28.4) |
| OEHR | 31,233 | 71,686 | 12.5 | 33.6 | 584 | 1,757 | 46.6 (42.9 - 50.5) | 52.3 (49.9 - 54.8) |
| VA | 1,492 | 7,880 | 0.9 | 5.2 | 24 | 121 | 27.4 (17.5 - 40.7) | 23.3 (19.3 - 27.8) |
| SIDIAP | 18,524 | 5,273 | 37.9 | 11.2 | 331 | 93 | 8.72 (7.81 - 9.72) | 8.34 (6.73 - 10.2) |
| Meta-analysis | 486,828 | 914,714 | 353.0 | 703.2 | 10,473 | 22,409 | 29.7 (29.1 - 30.2) | 31.9 (31.5 - 32.3) |
| Meta-analysis (w/o USOC) | 128,108 | 215,760 | 104.6 | 140.2 | 3,293 | 6,672 | 31.5 (30.4 - 32.6) | 47.6 (46.4 - 48.7) |
| **PS-stratification / on-treatment TAR without escalation** | | | | | | | | |
| CCAE | 38,599 | 57,863 | 24.9 | 36.2 | 1,058 | 1,486 | 42.5 (40.0 - 45.1) | 41.0 (39.0 - 43.2) |
| DAG | 9,527 | 2,130 | 9.2 | 2.1 | 135 | 27 | 14.7 (12.3 - 17.4) | 13.0 (8.55 - 18.9) |
| ODOD | 23,886 | 48,728 | 15.7 | 36.8 | 846 | 2,124 | 54.0 (50.4 - 57.7) | 57.8 (55.3 - 60.3) |
| MDCR | 7,552 | 13,481 | 6.1 | 10.8 | 271 | 495 | 44.3 (39.2 - 49.9) | 45.8 (41.8 - 50.0) |
| MDCD | 5,142 | 11,261 | 2.8 | 6.1 | 316 | 621 | 114 (102 - 128) | 102 (94.5 - 111) |
| Open Claims | 367,392 | 699,041 | 256.1 | 563.0 | 7,482 | 15,741 | 29.2 (28.6 - 29.9) | 28.0 (27.5 - 28.4) |
| OEHR | 33,348 | 71,751 | 13.6 | 33.6 | 638 | 1,758 | 47.1 (43.5 - 50.9) | 52.3 (49.9 - 54.8) |
| VA | 1,778 | 11,035 | 1.0 | 7.6 | 26 | 189 | 25.4 (16.6 - 37.2) | 24.8 (21.4 - 28.6) |
| SIDIAP | 18,621 | 6,878 | 38.0 | 15.2 | 332 | 122 | 8.73 (7.82 - 9.73) | 8.04 (6.68 - 9.60) |
| Meta-analysis | 498,925 | 899,872 | 363.5 | 697.6 | 10,762 | 21,753 | 29.6 (29.0 - 30.2) | 31.2 (30.8 - 31.6) |
| Meta-analysis (w/o USOC) | 131,533 | 200,831 | 107.4 | 134.7 | 3,280 | 6,012 | 30.5 (29.5 - 31.6) | 44.6 (43.5 - 45.8) |

##### 4.6.23 Peripheral edema

**Supplementary Table 438:** Exposures, outcomes and risk estimates for Peripheral edema in DPP4I (female) (T) vs SU (female) (C) new-user cohorts under four different propensity score (PS) adjustment / time-at-risk (TAR) designs across data sources. We report total patient exposure time in 1,000 patient-year (KYr) units, uncalibrated and calibrated hazard ratios (HRs) and their 95% confidence intervals (CIs). Greyed out entries did not pass study diagnostics and are included only for completeness.

| | Patients | | Exposure (KYr) | | Outcomes | | Incidence (per KYr) | |
|---|---|---|---|---|---|---|---|---|
| | T | C | T | C | T | C | T | C |
| **PS-matching / on-treatment TAR** | | | | | | | | |
| CCAE | 30,818 | 52,614 | 23.2 | 36.9 | 1,179 | 2,139 | 50.7 (47.9 - 53.7) | 58.0 (55.6 - 60.6) |
| DAG | 8,516 | 1,689 | 9.0 | 1.7 | 312 | 71 | 34.7 (30.9 - 38.7) | 41.8 (32.6 - 52.7) |
| ODOD | 19,974 | 44,143 | 14.6 | 35.0 | 1,377 | 3,506 | 94.5 (89.6 - 99.7) | 100 (96.8 - 103) |
| MDCR | 5,797 | 11,111 | 5.1 | 9.3 | 468 | 998 | 90.9 (82.8 - 99.5) | 108 (101 - 115) |
| MDCD | 4,727 | 11,019 | 3.0 | 6.5 | 352 | 725 | 119 (107 - 132) | 111 (103 - 119) |
| Open Claims | 322,999 | 641,166 | 272.3 | 586.1 | 10,940 | 25,277 | 40.2 (39.4 - 40.9) | 43.1 (42.6 - 43.7) |
| OEHR | 29,650 | 68,002 | 14.3 | 35.5 | 806 | 2,574 | 56.3 (52.5 - 60.3) | 72.6 (69.8 - 75.4) |
| VA | 1,246 | 6,497 | 0.9 | 4.7 | 49 | 207 | 55.5 (41.1 - 73.4) | 43.9 (38.1 - 50.3) |
| SIDIAP | 17,390 | 5,086 | 47.0 | 12.9 | 926 | 235 | 19.7 (18.4 - 21.0) | 18.2 (15.9 - 20.6) |
| Meta-analysis | 441,117 | 841,327 | 389.5 | 728.6 | 16,409 | 35,732 | 42.1 (41.5 - 42.8) | 49.0 (48.5 - 49.6) |
| Meta-analysis (w/o USOC) | 118,118 | 200,161 | 117.1 | 142.5 | 5,469 | 10,455 | 46.7 (45.5 - 47.9) | 73.4 (72.0 - 74.8) |
| **PS-stratification / on-treatment TAR** | | | | | | | | |
| CCAE | 35,723 | 54,344 | 27.2 | 37.9 | 1,396 | 2,189 | 51.4 (48.7 - 54.1) | 57.8 (55.4 - 60.3) |
| DAG | 8,543 | 1,952 | 9.0 | 2.0 | 315 | 81 | 34.9 (31.2 - 39.0) | 40.7 (32.4 - 50.6) |
| ODOD | 21,408 | 44,278 | 15.7 | 35.1 | 1,486 | 3,509 | 94.7 (89.9 - 99.6) | 100 (96.8 - 103) |
| MDCR | 6,251 | 11,373 | 5.6 | 9.5 | 516 | 1,017 | 92.8 (84.9 - 101) | 107 (101 - 114) |
| MDCD | 4,930 | 11,097 | 3.1 | 6.6 | 377 | 726 | 121 (109 - 134) | 111 (103 - 119) |
| Open Claims | 329,855 | 641,293 | 279.7 | 586.1 | 11,328 | 25,279 | 40.5 (39.8 - 41.3) | 43.1 (42.6 - 43.7) |
| OEHR | 31,683 | 68,073 | 15.5 | 35.5 | 858 | 2,577 | 55.5 (51.8 - 59.3) | 72.6 (69.8 - 75.5) |
| VA | 1,478 | 9,426 | 1.0 | 7.1 | 55 | 346 | 54.0 (40.7 - 70.2) | 48.5 (43.5 - 53.9) |
| SIDIAP | 17,464 | 6,675 | 47.1 | 17.7 | 930 | 301 | 19.8 (18.5 - 21.1) | 17.0 (15.1 - 19.0) |
| Meta-analysis | 450,927 | 827,988 | 399.7 | 723.7 | 16,829 | 34,953 | 42.1 (41.5 - 42.7) | 48.3 (47.8 - 48.8) |
| Meta-analysis (w/o USOC) | 121,072 | 186,695 | 120.0 | 137.6 | 5,501 | 9,674 | 45.8 (44.6 - 47.1) | 70.3 (68.9 - 71.7) |
| **PS-matching / on-treatment TAR without escalation** | | | | | | | | |
| CCAE | 30,818 | 52,614 | 19.3 | 32.1 | 986 | 1,857 | 51.2 (48.0 - 54.5) | 57.8 (55.2 - 60.5) |
| DAG | 8,516 | 1,689 | 8.1 | 1.6 | 284 | 65 | 34.9 (30.9 - 39.2) | 41.3 (31.9 - 52.7) |
| ODOD | 19,974 | 44,143 | 12.3 | 31.5 | 1,144 | 3,125 | 92.9 (87.6 - 98.5) | 99.1 (95.7 - 103) |
| MDCR | 5,797 | 11,111 | 4.5 | 8.6 | 404 | 929 | 89.4 (80.9 - 98.6) | 108 (101 - 115) |
| MDCD | 4,727 | 11,019 | 2.4 | 5.6 | 275 | 620 | 115 (102 - 130) | 111 (102 - 120) |
| Open Claims | 322,999 | 641,166 | 218.1 | 497.3 | 8,695 | 21,267 | 39.9 (39.0 - 40.7) | 42.8 (42.2 - 43.3) |
| OEHR | 29,650 | 68,002 | 11.7 | 30.8 | 613 | 2,105 | 52.5 (48.4 - 56.8) | 68.4 (65.5 - 71.4) |
| VA | 1,246 | 6,497 | 0.7 | 4.1 | 41 | 176 | 58.1 (41.7 - 78.8) | 42.5 (36.4 - 49.2) |
| SIDIAP | 17,390 | 5,086 | 35.0 | 10.6 | 690 | 194 | 19.7 (18.3 - 21.3) | 18.3 (15.8 - 21.1) |
| Meta-analysis | 441,117 | 841,327 | 312.0 | 622.2 | 13,132 | 30,338 | 42.1 (41.4 - 42.8) | 48.8 (48.2 - 49.3) |
| Meta-analysis (w/o USOC) | 118,118 | 200,161 | 94.0 | 124.9 | 4,437 | 9,071 | 47.2 (45.8 - 48.6) | 72.6 (71.1 - 74.1) |
| **PS-stratification / on-treatment TAR without escalation** | | | | | | | | |
| CCAE | 35,723 | 54,344 | 22.6 | 33.1 | 1,160 | 1,900 | 51.4 (48.5 - 54.4) | 57.5 (54.9 - 60.1) |
| DAG | 8,543 | 1,952 | 8.2 | 1.8 | 287 | 73 | 35.2 (31.2 - 39.5) | 39.9 (31.3 - 50.1) |
| ODOD | 21,408 | 44,278 | 13.3 | 31.6 | 1,231 | 3,128 | 92.7 (87.6 - 98.0) | 99.1 (95.7 - 103) |
| MDCR | 6,251 | 11,373 | 4.9 | 8.8 | 449 | 944 | 91.9 (83.6 - 101) | 107 (101 - 114) |
| MDCD | 4,930 | 11,097 | 2.5 | 5.6 | 295 | 621 | 118 (105 - 133) | 111 (102 - 120) |
| Open Claims | 329,855 | 641,293 | 224.0 | 497.3 | 9,012 | 21,269 | 40.2 (39.4 - 41.1) | 42.8 (42.2 - 43.3) |
| OEHR | 31,683 | 68,073 | 12.6 | 30.8 | 658 | 2,106 | 52.1 (48.2 - 56.3) | 68.4 (65.5 - 71.4) |
| VA | 1,478 | 9,426 | 0.8 | 6.3 | 46 | 293 | 56.0 (41.0 - 74.7) | 46.5 (41.4 - 52.2) |
| SIDIAP | 17,464 | 6,675 | 35.0 | 14.4 | 694 | 243 | 19.8 (18.4 - 21.4) | 16.8 (14.8 - 19.1) |
| Meta-analysis | 450,927 | 827,988 | 320.5 | 617.8 | 13,491 | 29,663 | 42.1 (41.4 - 42.8) | 48.0 (47.5 - 48.6) |
| Meta-analysis (w/o USOC) | 121,072 | 186,695 | 96.6 | 120.5 | 4,479 | 8,394 | 46.4 (45.0 - 47.8) | 69.7 (68.2 - 71.2) |

###### 4.6.24 Photosensitivity

**Supplementary Table 439:** Exposures, outcomes and risk estimates for Photosensitivity in DPP4I (female) (T) vs SU (female) (C) new-user cohorts under four different propensity score (PS) adjustment / time-at-risk (TAR) designs across data sources. We report total patient exposure time in 1,000 patient-year (KYr) units, uncalibrated and calibrated hazard ratios (HRs) and their 95% confidence intervals (CIs). Greyed out entries did not pass study diagnostics and are included only for completeness.

| | Patients | | Exposure (KYr) | | Outcomes | | Incidence (per KYr) | |
|---|---|---|---|---|---|---|---|---|
| | T | C | T | C | T | C | T | C |
| **PS-matching / on-treatment TAR** | | | | | | | | |
| CCAE | 38,750 | 65,513 | 30.3 | 47.7 | 42 | 76 | 1.39 (1.00 - 1.88) | 1.59 (1.26 - 1.99) |
| DAG | 10,440 | 2,033 | 11.3 | 2.1 | <5 | <5 | < 0.44 (0.14 - 1.03) | < 2.35 (0.76 - 5.49) |
| ODOD | 27,224 | 59,640 | 21.5 | 51.7 | 42 | 81 | 1.95 (1.41 - 2.64) | 1.57 (1.25 - 1.95) |
| MDCR | 8,046 | 15,282 | 7.6 | 13.6 | 24 | 30 | 3.17 (2.03 - 4.72) | 2.20 (1.49 - 3.14) |
| MDCD | 7,363 | 16,649 | 5.2 | 10.9 | 10 | 10 | 1.92 (0.92 - 3.53) | 0.91 (0.44 - 1.68) |
| Open Claims | 435,042 | 844,230 | 379.6 | 802.8 | 199 | 387 | 0.52 (0.45 - 0.60) | 0.48 (0.44 - 0.53) |
| OEHR | 36,161 | 82,685 | 18.5 | 47.0 | 35 | 67 | 1.89 (1.32 - 2.63) | 1.43 (1.10 - 1.81) |
| VA | 1,802 | 9,559 | 1.4 | 7.4 | 0 | <10 | | < 1.35 (0.65 - 2.49) |
| SIDIAP | 19,472 | 5,426 | 53.6 | 14.2 | 10 | <5 | 0.19 (0.09 - 0.34) | < 0.35 (0.11 - 0.82) |
| Meta-analysis | 552,586 | 1,083,999 | 462.7 | 973.7 | 352 | 651 | 0.76 (0.68 - 0.84) | 0.67 (0.62 - 0.72) |
| Meta-analysis (w/o USOC) | 117,544 | 239,769 | 83.1 | 170.9 | 153 | 264 | 1.84 (1.56 - 2.16) | 1.54 (1.36 - 1.74) |
| **PS-stratification / on-treatment TAR** | | | | | | | | |
| CCAE | 45,015 | 67,427 | 35.5 | 48.8 | 50 | 77 | 1.41 (1.05 - 1.86) | 1.58 (1.24 - 1.97) |
| DAG | 10,471 | 2,360 | 11.3 | 2.5 | <5 | <5 | < 0.44 (0.14 - 1.03) | < 1.99 (0.65 - 4.64) |
| ODOD | 29,190 | 59,795 | 23.2 | 51.7 | 45 | 81 | 1.94 (1.42 - 2.60) | 1.57 (1.24 - 1.95) |
| MDCR | 8,668 | 15,603 | 8.2 | 13.9 | 25 | 30 | 3.06 (1.98 - 4.52) | 2.16 (1.46 - 3.08) |
| MDCD | 7,706 | 16,755 | 5.5 | 11.0 | 11 | 10 | 2.00 (1.00 - 3.59) | 0.91 (0.44 - 1.68) |
| Open Claims | 444,867 | 844,327 | 390.5 | 802.8 | 204 | 387 | 0.52 (0.45 - 0.60) | 0.48 (0.44 - 0.53) |
| OEHR | 38,618 | 82,774 | 20.0 | 47.0 | 37 | 67 | 1.85 (1.30 - 2.55) | 1.42 (1.10 - 1.81) |
| VA | 2,194 | 13,060 | 1.6 | 10.5 | 0 | <10 | | < 0.95 (0.46 - 1.75) |
| SIDIAP | 19,584 | 7,060 | 53.7 | 19.4 | 10 | <5 | 0.19 (0.09 - 0.34) | < 0.26 (0.08 - 0.60) |
| Meta-analysis | 566,358 | 1,069,926 | 477.4 | 964.3 | 361 | 642 | 0.76 (0.68 - 0.84) | 0.67 (0.62 - 0.72) |
| Meta-analysis (w/o USOC) | 121,491 | 225,599 | 86.8 | 161.5 | 157 | 255 | 1.81 (1.54 - 2.11) | 1.58 (1.39 - 1.79) |
| **PS-matching / on-treatment TAR without escalation** | | | | | | | | |
| CCAE | 38,750 | 65,513 | 24.9 | 41.1 | 39 | 68 | 1.57 (1.11 - 2.14) | 1.66 (1.29 - 2.10) |
| DAG | 10,440 | 2,033 | 10.2 | 2.0 | <5 | <5 | < 0.49 (0.16 - 1.14) | < 2.55 (0.83 - 5.94) |
| ODOD | 27,224 | 59,640 | 18.0 | 46.0 | 36 | 76 | 2.00 (1.40 - 2.76) | 1.65 (1.30 - 2.07) |
| MDCR | 8,046 | 15,282 | 6.6 | 12.5 | 22 | 27 | 3.35 (2.10 - 5.07) | 2.17 (1.43 - 3.15) |
| MDCD | 7,363 | 16,649 | 4.1 | 9.1 | 8 | 8 | 1.97 (0.85 - 3.88) | 0.88 (0.38 - 1.73) |
| Open Claims | 435,042 | 844,230 | 301.3 | 675.2 | 172 | 334 | 0.57 (0.49 - 0.66) | 0.49 (0.44 - 0.55) |
| OEHR | 36,161 | 82,685 | 14.9 | 39.8 | 30 | 57 | 2.02 (1.36 - 2.88) | 1.43 (1.08 - 1.85) |
| VA | 1,802 | 9,559 | 1.1 | 6.4 | 0 | <10 | | < 1.56 (0.75 - 2.88) |
| SIDIAP | 19,472 | 5,426 | 39.9 | 11.5 | 9 | <5 | 0.23 (0.10 - 0.43) | < 0.43 (0.14 - 1.01) |
| Meta-analysis | 545,223 | 1,067,350 | 365.7 | 814.5 | 299 | 562 | 0.82 (0.73 - 0.92) | 0.69 (0.63 - 0.75) |
| Meta-analysis (w/o USOC) | 110,181 | 223,120 | 64.4 | 139.3 | 127 | 228 | 1.97 (1.64 - 2.35) | 1.64 (1.43 - 1.86) |
| **PS-stratification / on-treatment TAR without escalation** | | | | | | | | |
| CCAE | 45,015 | 67,427 | 29.2 | 42.1 | 45 | 69 | 1.54 (1.12 - 2.06) | 1.64 (1.27 - 2.07) |
| DAG | 10,471 | 2,360 | 10.2 | 2.3 | <5 | <5 | < 0.49 (0.16 - 1.14) | < 2.16 (0.70 - 5.05) |
| ODOD | 29,190 | 59,795 | 19.4 | 46.0 | 37 | 76 | 1.91 (1.34 - 2.63) | 1.65 (1.30 - 2.07) |
| MDCR | 8,668 | 15,603 | 7.1 | 12.7 | 23 | 27 | 3.24 (2.06 - 4.86) | 2.12 (1.40 - 3.08) |
| MDCD | 7,706 | 16,755 | 4.3 | 9.1 | 9 | 8 | 2.10 (0.96 - 3.99) | 0.88 (0.38 - 1.73) |
| Open Claims | 444,867 | 844,327 | 310.0 | 675.2 | 176 | 334 | 0.57 (0.49 - 0.66) | 0.49 (0.44 - 0.55) |
| OEHR | 38,618 | 82,774 | 16.1 | 39.9 | 31 | 57 | 1.92 (1.31 - 2.73) | 1.43 (1.08 - 1.85) |
| VA | 2,194 | 13,060 | 1.3 | 9.2 | 0 | <10 | | < 1.09 (0.52 - 2.01) |
| SIDIAP | 19,584 | 7,060 | 39.9 | 15.7 | 9 | <5 | 0.23 (0.10 - 0.43) | < 0.32 (0.10 - 0.74) |
| Meta-analysis | 566,358 | 1,069,926 | 381.9 | 816.0 | 312 | 563 | 0.82 (0.73 - 0.91) | 0.69 (0.63 - 0.75) |
| Meta-analysis (w/o USOC) | 121,491 | 225,599 | 71.8 | 140.7 | 136 | 229 | 1.89 (1.59 - 2.24) | 1.63 (1.42 - 1.85) |

##### 4.6.25 Venous thromboembolism

**Supplementary Table 440:** Exposures, outcomes and risk estimates for Venous thromboembolism in DPP4I (female) (T) vs SU (female) (C) new-user cohorts under four different propensity score (PS) adjustment / time-at-risk (TAR) designs across data sources. We report total patient exposure time in 1,000 patient-year (KYr) units, uncalibrated and calibrated hazard ratios (HRs) and their 95% confidence intervals (CIs). Greyed out entries did not pass study diagnostics and are included only for completeness.

| | Patients | | Exposure (KYr) | | Outcomes | | Incidence (per KYr) | |
|---|---|---|---|---|---|---|---|---|
| | T | C | T | C | T | C | T | C |
| **PS-matching / on-treatment TAR** | | | | | | | | |
| CCAE | 38,199 | 64,583 | 29.8 | 47.0 | 160 | 302 | 5.36 (4.56 - 6.26) | 6.43 (5.72 - 7.19) |
| DAG | 9,609 | 1,856 | 10.3 | 1.9 | 113 | 22 | 11.0 (9.06 - 13.2) | 11.5 (7.19 - 17.4) |
| ODOD | 26,631 | 58,121 | 21.0 | 50.1 | 231 | 621 | 11.0 (9.65 - 12.5) | 12.4 (11.4 - 13.4) |
| MDCR | 7,810 | 14,717 | 7.4 | 13.0 | 101 | 227 | 13.7 (11.1 - 16.6) | 17.4 (15.2 - 19.8) |
| MDCD | 7,076 | 16,013 | 5.0 | 10.4 | 56 | 131 | 11.3 (8.54 - 14.7) | 12.6 (10.5 - 14.9) |
| Open Claims | 423,945 | 822,965 | 369.2 | 780.4 | 1,956 | 4,999 | 5.30 (5.07 - 5.54) | 6.41 (6.23 - 6.59) |
| OEHR | 35,455 | 81,178 | 18.0 | 45.6 | 131 | 472 | 7.28 (6.09 - 8.64) | 10.3 (9.43 - 11.3) |
| VA | 1,767 | 9,456 | 1.3 | 7.3 | <10 | 28 | < 7.52 (3.60 - 13.8) | 3.85 (2.56 - 5.56) |
| SIDIAP | 18,840 | 5,317 | 51.8 | 13.8 | 264 | 63 | 5.10 (4.50 - 5.75) | 4.56 (3.50 - 5.83) |
| Meta-analysis | 569,332 | 1,074,206 | 513.7 | 969.6 | <3,022 | 6,865 | < 5.88 (5.67 - 6.10) | 7.08 (6.91 - 7.25) |
| Meta-analysis (w/o USOC) | 145,387 | 251,241 | 144.5 | 189.2 | <1,066 | 1,866 | < 7.38 (6.94 - 7.83) | 9.86 (9.42 - 10.3) |
| **PS-stratification / on-treatment TAR** | | | | | | | | |
| CCAE | 44,442 | 66,492 | 35.0 | 48.1 | 185 | 310 | 5.28 (4.55 - 6.10) | 6.44 (5.74 - 7.20) |
| DAG | 9,629 | 2,158 | 10.3 | 2.3 | 115 | 22 | 11.2 (9.22 - 13.4) | 9.66 (6.05 - 14.6) |
| ODOD | 28,540 | 58,276 | 22.6 | 50.2 | 240 | 622 | 10.6 (9.34 - 12.1) | 12.4 (11.4 - 13.4) |
| MDCR | 8,418 | 15,107 | 7.9 | 13.4 | 108 | 229 | 13.6 (11.1 - 16.4) | 17.1 (14.9 - 19.4) |
| MDCD | 7,411 | 16,115 | 5.2 | 10.5 | 61 | 131 | 11.7 (8.93 - 15.0) | 12.5 (10.5 - 14.9) |
| Open Claims | 433,673 | 823,062 | 380.1 | 780.4 | 2,031 | 4,999 | 5.34 (5.11 - 5.58) | 6.41 (6.23 - 6.59) |
| OEHR | 37,874 | 81,264 | 19.5 | 45.7 | 139 | 472 | 7.13 (5.99 - 8.42) | 10.3 (9.42 - 11.3) |
| VA | 2,154 | 12,911 | 1.6 | 10.4 | <10 | 42 | < 6.38 (3.06 - 11.7) | 4.06 (2.92 - 5.49) |
| SIDIAP | 18,952 | 6,941 | 51.9 | 18.9 | 265 | 84 | 5.11 (4.51 - 5.76) | 4.43 (3.54 - 5.49) |
| Meta-analysis | 581,528 | 1,053,300 | 527.3 | 959.0 | 3,083 | 6,738 | 5.85 (5.64 - 6.06) | 7.03 (6.86 - 7.20) |
| Meta-analysis (w/o USOC) | 147,855 | 230,238 | 147.2 | 178.6 | 1,052 | 1,739 | 7.15 (6.72 - 7.59) | 9.74 (9.29 - 10.2) |
| **PS-matching / on-treatment TAR without escalation** | | | | | | | | |
| CCAE | 38,199 | 64,583 | 24.5 | 40.5 | 128 | 260 | 5.22 (4.36 - 6.21) | 6.42 (5.67 - 7.26) |
| DAG | 9,609 | 1,856 | 9.3 | 1.8 | 105 | 21 | 11.3 (9.23 - 13.7) | 11.9 (7.34 - 18.1) |
| ODOD | 26,631 | 58,121 | 17.6 | 44.6 | 204 | 563 | 11.6 (10.1 - 13.3) | 12.6 (11.6 - 13.7) |
| MDCR | 7,810 | 14,717 | 6.4 | 12.0 | 85 | 207 | 13.3 (10.6 - 16.4) | 17.3 (15.0 - 19.8) |
| MDCD | 7,076 | 16,013 | 3.9 | 8.7 | 39 | 108 | 10.1 (7.16 - 13.8) | 12.4 (10.2 - 15.0) |
| Open Claims | 423,945 | 822,965 | 293.2 | 656.7 | 1,576 | 4,270 | 5.37 (5.11 - 5.65) | 6.50 (6.31 - 6.70) |
| OEHR | 35,455 | 81,178 | 14.5 | 38.8 | 87 | 366 | 6.00 (4.80 - 7.40) | 9.43 (8.49 - 10.4) |
| VA | 1,767 | 9,456 | 1.1 | 6.3 | <10 | 21 | < 9.41 (4.51 - 17.3) | 3.33 (2.06 - 5.09) |
| SIDIAP | 18,840 | 5,317 | 38.5 | 11.3 | 191 | 48 | 4.96 (4.28 - 5.72) | 4.26 (3.14 - 5.65) |
| Meta-analysis | 569,332 | 1,074,206 | 409.0 | 820.6 | <2,425 | 5,864 | < 5.93 (5.70 - 6.17) | 7.15 (6.96 - 7.33) |
| Meta-analysis (w/o USOC) | 145,387 | 251,241 | 115.8 | 163.9 | <849 | 1,594 | < 7.33 (6.85 - 7.84) | 9.73 (9.26 - 10.2) |
| **PS-stratification / on-treatment TAR without escalation** | | | | | | | | |
| CCAE | 44,442 | 66,492 | 28.8 | 41.5 | 147 | 266 | 5.10 (4.31 - 6.00) | 6.41 (5.66 - 7.23) |
| DAG | 9,629 | 2,158 | 9.3 | 2.1 | 107 | 21 | 11.5 (9.41 - 13.9) | 10.0 (6.22 - 15.3) |
| ODOD | 28,540 | 58,276 | 18.9 | 44.6 | 211 | 564 | 11.2 (9.70 - 12.8) | 12.6 (11.6 - 13.7) |
| MDCR | 8,418 | 15,107 | 6.9 | 12.3 | 89 | 209 | 12.9 (10.3 - 15.9) | 17.0 (14.8 - 19.5) |
| MDCD | 7,411 | 16,115 | 4.1 | 8.7 | 44 | 108 | 10.8 (7.83 - 14.5) | 12.4 (10.2 - 15.0) |
| Open Claims | 433,673 | 823,062 | 302.0 | 656.7 | 1,637 | 4,270 | 5.42 (5.16 - 5.69) | 6.50 (6.31 - 6.70) |
| OEHR | 37,874 | 81,264 | 15.7 | 38.9 | 95 | 366 | 6.04 (4.88 - 7.38) | 9.42 (8.48 - 10.4) |
| VA | 2,154 | 12,911 | 1.3 | 9.0 | <10 | 33 | < 7.96 (3.82 - 14.6) | 3.66 (2.52 - 5.14) |
| SIDIAP | 18,952 | 6,941 | 38.6 | 15.4 | 192 | 61 | 4.98 (4.30 - 5.73) | 3.97 (3.04 - 5.10) |
| Meta-analysis | 581,528 | 1,053,300 | 420.3 | 811.5 | 2,478 | 5,757 | 5.90 (5.67 - 6.13) | 7.09 (6.91 - 7.28) |
| Meta-analysis (w/o USOC) | 147,855 | 230,238 | 118.3 | 154.7 | 841 | 1,487 | 7.11 (6.64 - 7.61) | 9.61 (9.13 - 10.1) |

##### 4.6.26 Vomiting

**Supplementary Table 441:** Exposures, outcomes and risk estimates for Vomiting in DPP4I (female) (T) vs SU (female) (C) new-user cohorts under four different propensity score (PS) adjustment / time-at-risk (TAR) designs across data sources. We report total patient exposure time in 1,000 patient-year (KYr) units, uncalibrated and calibrated hazard ratios (HRs) and their 95% confidence intervals (CIs). Greyed out entries did not pass study diagnostics and are included only for completeness.

| | Patients | | Exposure (KYr) | | Outcomes | | Incidence (per KYr) | |
|---|---|---|---|---|---|---|---|---|
| | T | C | T | C | T | C | T | C |
| **PS-matching / on-treatment TAR** | | | | | | | | |
| CCAE | 34,815 | 58,725 | 27.1 | 42.5 | 791 | 1,266 | 29.2 (27.2 - 31.3) | 29.8 (28.2 - 31.5) |
| DAG | 9,486 | 1,839 | 10.2 | 1.9 | 150 | 24 | 14.8 (12.5 - 17.3) | 12.5 (8.02 - 18.6) |
| ODOD | 23,788 | 51,899 | 18.5 | 44.2 | 672 | 1,854 | 36.4 (33.7 - 39.3) | 42.0 (40.1 - 43.9) |
| MDCR | 7,281 | 13,676 | 6.7 | 12.0 | 227 | 453 | 33.6 (29.4 - 38.3) | 37.8 (34.4 - 41.4) |
| MDCD | 5,459 | 12,359 | 3.7 | 7.9 | 288 | 562 | 77.0 (68.4 - 86.5) | 71.0 (65.3 - 77.1) |
| Open Claims | 377,793 | 733,701 | 328.5 | 699.2 | 6,955 | 14,507 | 21.2 (20.7 - 21.7) | 20.7 (20.4 - 21.1) |
| OEHR | 33,099 | 75,511 | 16.5 | 41.7 | 535 | 1,545 | 32.3 (29.7 - 35.2) | 37.1 (35.2 - 38.9) |
| VA | 1,579 | 8,275 | 1.2 | 6.3 | 18 | 106 | 15.3 (9.10 - 24.3) | 16.9 (13.8 - 20.4) |
| SIDIAP | 18,478 | 5,262 | 50.8 | 13.6 | 476 | 117 | 9.37 (8.55 - 10.3) | 8.59 (7.10 - 10.3) |
| Meta-analysis | 511,778 | 961,247 | 463.3 | 869.3 | 10,112 | 20,434 | 21.8 (21.4 - 22.3) | 23.5 (23.2 - 23.8) |
| Meta-analysis (w/o USOC) | 133,985 | 227,546 | 134.7 | 170.1 | 3,157 | 5,927 | 23.4 (22.6 - 24.3) | 34.8 (34.0 - 35.7) |
| **PS-stratification / on-treatment TAR** | | | | | | | | |
| CCAE | 40,551 | 60,512 | 31.7 | 43.6 | 925 | 1,294 | 29.1 (27.3 - 31.1) | 29.7 (28.1 - 31.3) |
| DAG | 9,522 | 2,129 | 10.2 | 2.3 | 150 | 31 | 14.7 (12.5 - 17.3) | 13.7 (9.32 - 19.5) |
| ODOD | 25,529 | 52,033 | 19.9 | 44.2 | 732 | 1,857 | 36.7 (34.1 - 39.5) | 42.0 (40.1 - 44.0) |
| MDCR | 7,844 | 14,058 | 7.3 | 12.3 | 245 | 461 | 33.6 (29.5 - 38.1) | 37.4 (34.1 - 41.0) |
| MDCD | 5,668 | 12,420 | 3.9 | 7.9 | 300 | 565 | 77.0 (68.5 - 86.2) | 71.3 (65.5 - 77.4) |
| Open Claims | 386,754 | 733,792 | 338.3 | 699.2 | 7,240 | 14,509 | 21.4 (20.9 - 21.9) | 20.8 (20.4 - 21.1) |
| OEHR | 35,368 | 75,582 | 17.9 | 41.7 | 580 | 1,545 | 32.4 (29.8 - 35.1) | 37.0 (35.2 - 38.9) |
| VA | 1,911 | 11,600 | 1.4 | 9.2 | 20 | 161 | 14.6 (8.90 - 22.5) | 17.5 (14.9 - 20.4) |
| SIDIAP | 18,575 | 6,868 | 50.9 | 18.7 | 477 | 154 | 9.38 (8.56 - 10.3) | 8.25 (7.00 - 9.66) |
| Meta-analysis | 524,143 | 944,974 | 476.2 | 862.0 | 10,349 | 19,851 | 21.7 (21.3 - 22.2) | 23.0 (22.7 - 23.4) |
| Meta-analysis (w/o USOC) | 137,389 | 211,182 | 137.9 | 162.8 | 3,109 | 5,342 | 22.5 (21.8 - 23.4) | 32.8 (31.9 - 33.7) |
| **PS-matching / on-treatment TAR without escalation** | | | | | | | | |
| CCAE | 34,815 | 58,725 | 22.4 | 36.9 | 648 | 1,064 | 28.9 (26.7 - 31.2) | 28.9 (27.2 - 30.7) |
| DAG | 9,486 | 1,839 | 9.2 | 1.8 | 135 | 21 | 14.7 (12.3 - 17.4) | 11.8 (7.33 - 18.1) |
| ODOD | 23,788 | 51,899 | 15.6 | 39.5 | 560 | 1,664 | 35.8 (32.9 - 38.9) | 42.1 (40.1 - 44.2) |
| MDCR | 7,281 | 13,676 | 5.9 | 11.0 | 187 | 398 | 31.9 (27.5 - 36.8) | 36.2 (32.7 - 39.9) |
| MDCD | 5,459 | 12,359 | 3.0 | 6.6 | 217 | 484 | 72.7 (63.4 - 83.0) | 72.9 (66.6 - 79.7) |
| Open Claims | 377,793 | 733,701 | 262.3 | 591.6 | 5,476 | 11,992 | 20.9 (20.3 - 21.4) | 20.3 (19.9 - 20.6) |
| OEHR | 33,099 | 75,511 | 13.4 | 35.8 | 406 | 1,228 | 30.3 (27.4 - 33.3) | 34.3 (32.4 - 36.3) |
| VA | 1,579 | 8,275 | 0.9 | 5.5 | 14 | 85 | 14.9 (8.17 - 25.1) | 15.5 (12.4 - 19.2) |
| SIDIAP | 18,478 | 5,262 | 37.8 | 11.1 | 345 | 92 | 9.12 (8.18 - 10.1) | 8.27 (6.67 - 10.1) |
| Meta-analysis | 511,778 | 961,247 | 370.6 | 739.7 | 7,988 | 17,028 | 21.6 (21.1 - 22.0) | 23.0 (22.7 - 23.4) |
| Meta-analysis (w/o USOC) | 133,985 | 227,546 | 108.3 | 148.2 | 2,512 | 5,036 | 23.2 (22.3 - 24.1) | 34.0 (33.1 - 34.9) |
| **PS-stratification / on-treatment TAR without escalation** | | | | | | | | |
| CCAE | 40,551 | 60,512 | 26.3 | 37.9 | 761 | 1,092 | 29.0 (27.0 - 31.1) | 28.8 (27.2 - 30.6) |
| DAG | 9,522 | 2,129 | 9.2 | 2.1 | 135 | 28 | 14.7 (12.3 - 17.4) | 13.5 (8.96 - 19.5) |
| ODOD | 25,529 | 52,033 | 16.9 | 39.5 | 609 | 1,667 | 36.1 (33.3 - 39.1) | 42.2 (40.2 - 44.2) |
| MDCR | 7,844 | 14,058 | 6.3 | 11.3 | 204 | 404 | 32.1 (27.9 - 36.9) | 35.7 (32.3 - 39.3) |
| MDCD | 5,668 | 12,420 | 3.1 | 6.7 | 227 | 487 | 73.2 (64.0 - 83.4) | 73.2 (66.9 - 80.0) |
| Open Claims | 386,754 | 733,792 | 270.1 | 591.6 | 5,707 | 11,994 | 21.1 (20.6 - 21.7) | 20.3 (19.9 - 20.6) |
| OEHR | 35,368 | 75,582 | 14.5 | 35.9 | 442 | 1,228 | 30.4 (27.6 - 33.4) | 34.3 (32.4 - 36.2) |
| VA | 1,911 | 11,600 | 1.1 | 8.1 | 15 | 131 | 13.6 (7.62 - 22.4) | 16.2 (13.6 - 19.3) |
| SIDIAP | 18,575 | 6,868 | 37.9 | 15.1 | 346 | 122 | 9.12 (8.19 - 10.1) | 8.06 (6.69 - 9.62) |
| Meta-analysis | 524,143 | 944,974 | 381.3 | 733.4 | 8,204 | 16,535 | 21.5 (21.1 - 22.0) | 22.5 (22.2 - 22.9) |
| Meta-analysis (w/o USOC) | 137,389 | 211,182 | 111.1 | 141.8 | 2,497 | 4,541 | 22.5 (21.6 - 23.4) | 32.0 (31.1 - 33.0) |
