## Supplement B and C for "Sex Differences in Comparative Effectiveness and Safety of Second-line Antidiabetic Agents: Real-world Evidence from Large-scale Multinational Study"

### Supplementary Material C

for

### MALE SUBJECTS

in

### Sex Differences in Comparative Effectiveness and Safety of Second-line Antidiabetic Agents: Real-world Evidence from Large-scale Multinational Study

### 1 Exposures, outcomes and risk across data sources and outcomes

#### 1.1 GLP1RA (male) vs SGLT2I (male)

##### 1.1.1 3-point MACE

**Supplementary Table 1:** Exposures, outcomes and risk estimates for 3-point MACE in GLP1RA (male) (T) vs SGLT2I (male) (C) new-user cohorts under four different propensity score (PS) adjustment / time-at-risk (TAR) designs across data sources. We report total patient exposure time in 1,000 patient-year (KYr) units, uncalibrated and calibrated hazard ratios (HRs) and their 95% confidence intervals (CIs). Greyed out entries did not pass study diagnostics and are included only for completeness.

| | Patients | | Exposure (KYr) | | Outcomes | | Uncalibrated | | Calibrated | |
|---|---|---|---|---|---|---|---|---|---|---|
| | T | C | T | C | T | C | HR (95% CI) | $p$ | HR (95% CI) | $p$ |
| **PS-matching / on-treatment TAR** | | | | | | | | | | |
| CCAE | 9,303 | 23,130 | 6.3 | 19.3 | 50 | 140 | 1.27 (0.81 - 1.97) | 0.29 | 1.18 (0.76 - 1.84) | 0.47 |
| ODOD | 6,825 | 16,405 | 4.0 | 11.4 | 34 | 170 | 0.57 (0.35 - 0.91) | 0.02 | 0.60 (0.37 - 0.97) | 0.04 |
| MDCR | 653 | 1,972 | 0.4 | 1.6 | 9 | 35 | 1.20 (0.39 - 3.42) | 0.74 | 0.92 (0.31 - 2.72) | 0.86 |
| Open Claims | 137,357 | 308,574 | 96.7 | 270.1 | 615 | 2,092 | 0.85 (0.75 - 0.96) | <0.01 | 0.80 (0.71 - 0.91) | <0.01 |
| OEHR | 7,754 | 19,529 | 2.2 | 6.2 | 22 | 48 | 1.00 (0.47 - 1.99) | 0.99 | 0.94 (0.46 - 1.93) | 0.85 |
| VA | 1,119 | 10,349 | 0.8 | 6.3 | <10 | 38 | 0.88 (0.18 - 3.10) | 0.86 | 0.59 (0.15 - 2.44) | 0.47 |
| Meta-analysis | 162,358 | 377,987 | 109.9 | 313.2 | <731 | 2,488 | 0.87 (0.67 - 1.14) | 0.32 | 0.83 (0.63 - 1.09) | 0.18 |
| Meta-analysis (w/o USOC) | 25,001 | 69,413 | 13.2 | 43.1 | <116 | 396 | 0.89 (0.57 - 1.39) | 0.62 | 0.85 (0.54 - 1.33) | 0.49 |
| **PS-stratification / on-treatment TAR** | | | | | | | | | | |
| CCAE | 9,931 | 23,133 | 6.6 | 19.3 | 52 | 140 | 1.22 (0.87 - 1.69) | 0.25 | 1.13 (0.81 - 1.59) | 0.47 |
| ODOD | 7,409 | 16,422 | 4.2 | 11.4 | 37 | 170 | 0.58 (0.40 - 0.83) | <0.01 | 0.56 (0.39 - 0.81) | <0.01 |
| MDCR | 685 | 1,977 | 0.4 | 1.6 | 9 | 36 | 1.15 (0.51 - 2.35) | 0.72 | 0.95 (0.44 - 2.04) | 0.87 |
| Open Claims | 147,406 | 308,612 | 101.6 | 270.1 | 644 | 2,093 | 0.91 (0.83 - 0.99) | 0.04 | 0.87 (0.79 - 0.95) | <0.01 |
| OEHR | 8,460 | 19,539 | 2.3 | 6.2 | 25 | 48 | 1.22 (0.71 - 2.03) | 0.46 | 1.09 (0.65 - 1.84) | 0.74 |
| VA | 1,281 | 11,375 | 0.9 | 6.9 | <10 | 50 | 1.68 (0.46 - 4.83) | 0.39 | 1.22 (0.37 - 3.96) | 0.75 |
| Meta-analysis | 173,206 | 367,706 | 114.8 | 307.0 | 758 | 2,451 | 0.93 (0.68 - 1.27) | 0.64 | 0.88 (0.64 - 1.21) | 0.43 |
| Meta-analysis (w/o USOC) | 25,800 | 59,094 | 13.2 | 36.9 | 114 | 358 | 0.94 (0.57 - 1.57) | 0.82 | 0.88 (0.53 - 1.47) | 0.64 |
| **PS-matching / on-treatment TAR without escalation** | | | | | | | | | | |
| CCAE | 9,303 | 23,130 | 5.5 | 16.8 | 43 | 116 | 1.30 (0.81 - 2.07) | 0.27 | 1.20 (0.75 - 1.93) | 0.45 |
| ODOD | 6,825 | 16,405 | 3.5 | 10.1 | 28 | 150 | 0.50 (0.28 - 0.83) | 0.01 | 0.51 (0.30 - 0.88) | 0.02 |
| MDCR | 653 | 1,972 | 0.4 | 1.4 | 6 | 32 | 0.81 (0.21 - 2.56) | 0.74 | 0.61 (0.17 - 2.10) | 0.43 |
| Open Claims | 137,357 | 308,574 | 84.4 | 229.5 | 510 | 1,785 | 0.86 (0.75 - 0.98) | 0.03 | 0.80 (0.70 - 0.92) | <0.01 |
| OEHR | 7,754 | 19,529 | 1.9 | 5.5 | 18 | 38 | 1.19 (0.55 - 2.44) | 0.65 | 1.08 (0.51 - 2.27) | 0.84 |
| VA | 1,119 | 10,349 | 0.6 | 5.4 | <10 | 32 | 0.75 (0.11 - 3.17) | 0.74 | 0.48 (0.09 - 2.63) | 0.40 |
| Meta-analysis | 161,239 | 367,638 | 95.3 | 261.9 | 599 | 2,089 | 0.89 (0.61 - 1.30) | 0.55 | 0.83 (0.57 - 1.22) | 0.35 |
| Meta-analysis (w/o USOC) | 23,882 | 59,064 | 10.9 | 32.4 | 89 | 304 | 0.91 (0.48 - 1.73) | 0.77 | 0.85 (0.44 - 1.61) | 0.62 |
| **PS-stratification / on-treatment TAR without escalation** | | | | | | | | | | |
| CCAE | 9,931 | 23,133 | 5.8 | 16.8 | 45 | 116 | 1.27 (0.88 - 1.81) | 0.19 | 1.16 (0.81 - 1.67) | 0.44 |
| ODOD | 7,409 | 16,422 | 3.8 | 10.1 | 31 | 150 | 0.57 (0.37 - 0.84) | <0.01 | 0.53 (0.36 - 0.80) | <0.01 |
| MDCR | 685 | 1,977 | 0.4 | 1.4 | 6 | 32 | 0.89 (0.32 - 2.05) | 0.80 | 0.71 (0.28 - 1.79) | 0.47 |
| Open Claims | 147,406 | 308,612 | 88.9 | 229.5 | 537 | 1,786 | 0.87 (0.78 - 0.96) | <0.01 | 0.82 (0.74 - 0.91) | <0.01 |
| OEHR | 8,460 | 19,539 | 2.0 | 5.5 | 21 | 38 | 1.29 (0.72 - 2.29) | 0.38 | 1.16 (0.65 - 2.07) | 0.62 |
| VA | 1,281 | 11,375 | 0.7 | 6.0 | <10 | 42 | 2.31 (0.64 - 6.54) | 0.16 | 1.67 (0.52 - 5.35) | 0.40 |
| Meta-analysis | 173,206 | 367,706 | 100.5 | 261.9 | 634 | 2,090 | 0.93 (0.65 - 1.32) | 0.68 | 0.87 (0.61 - 1.24) | 0.45 |
| Meta-analysis (w/o USOC) | 25,800 | 59,094 | 11.6 | 32.4 | 97 | 304 | 0.97 (0.55 - 1.70) | 0.90 | 0.89 (0.51 - 1.56) | 0.69 |

##### 1.1.2 4-point MACE

**Supplementary Table 2:** Exposures, outcomes and risk estimates for 4-point MACE in GLP1RA (male) (T) vs SGLT2I (male) (C) new-user cohorts under four different propensity score (PS) adjustment / time-at-risk (TAR) designs across data sources. We report total patient exposure time in 1,000 patient-year (KYr) units, uncalibrated and calibrated hazard ratios (HRs) and their 95% confidence intervals (CIs). Greyed out entries did not pass study diagnostics and are included only for completeness.

| | Patients | | Exposure (KYr) | | Outcomes | | Uncalibrated | | Calibrated | |
|---|---|---|---|---|---|---|---|---|---|---|
| | T | C | T | C | T | C | HR (95% CI) | $p$ | HR (95% CI) | $p$ |
| **PS-matching / on-treatment TAR** | | | | | | | | | | |
| CCAE | 9,140 | 22,776 | 6.2 | 19.0 | 81 | 211 | 1.27 (0.88 - 1.83) | 0.20 | 1.18 (0.82 - 1.71) | 0.39 |
| ODOD | 6,578 | 15,701 | 3.8 | 10.9 | 66 | 257 | 0.81 (0.56 - 1.16) | 0.26 | 0.85 (0.59 - 1.21) | 0.38 |
| MDCR | 619 | 1,857 | 0.4 | 1.5 | 15 | 51 | 1.01 (0.40 - 2.38) | 0.99 | 0.77 (0.31 - 1.89) | 0.57 |
| Open Claims | 133,612 | 297,193 | 94.2 | 261.9 | 927 | 2,979 | 0.95 (0.86 - 1.05) | 0.28 | 0.90 (0.81 - 1.00) | 0.07 |
| OEHR | 7,624 | 19,116 | 2.1 | 6.1 | 27 | 92 | 0.49 (0.25 - 0.89) | 0.03 | 0.47 (0.25 - 0.87) | 0.02 |
| VA | 1,072 | 9,618 | 0.7 | 5.8 | <10 | 67 | 0.83 (0.19 - 2.51) | 0.78 | 0.56 (0.15 - 2.05) | 0.38 |
| Meta-analysis | 158,026 | 364,404 | 107.1 | 303.8 | <1,111 | 3,606 | 0.90 (0.71 - 1.15) | 0.42 | 0.86 (0.67 - 1.10) | 0.23 |
| Meta-analysis (w/o USOC) | 24,414 | 67,211 | 12.9 | 41.9 | <184 | 627 | 0.85 (0.56 - 1.28) | 0.44 | 0.81 (0.53 - 1.22) | 0.32 |
| **PS-stratification / on-treatment TAR** | | | | | | | | | | |
| CCAE | 9,749 | 22,780 | 6.5 | 19.0 | 85 | 211 | 1.19 (0.91 - 1.55) | 0.20 | 1.11 (0.85 - 1.45) | 0.46 |
| ODOD | 7,135 | 15,713 | 4.1 | 10.9 | 70 | 257 | 0.76 (0.57 - 1.00) | 0.05 | 0.73 (0.55 - 0.97) | 0.03 |
| MDCR | 650 | 1,862 | 0.4 | 1.5 | 15 | 52 | 1.26 (0.67 - 2.24) | 0.46 | 1.04 (0.57 - 1.90) | 0.85 |
| Open Claims | 143,483 | 297,226 | 99.1 | 262.0 | 973 | 2,981 | 0.96 (0.89 - 1.03) | 0.25 | 0.91 (0.84 - 0.99) | 0.06 |
| OEHR | 8,332 | 19,146 | 2.3 | 6.1 | 31 | 93 | 0.75 (0.48 - 1.14) | 0.19 | 0.67 (0.43 - 1.04) | 0.08 |
| VA | 1,229 | 10,518 | 0.9 | 6.4 | <10 | 86 | 0.93 (0.27 - 2.44) | 0.89 | 0.67 (0.22 - 2.03) | 0.48 |
| Meta-analysis | 168,699 | 354,865 | 111.9 | 298.0 | 1,159 | 3,542 | 0.93 (0.77 - 1.11) | 0.42 | 0.88 (0.72 - 1.07) | 0.20 |
| Meta-analysis (w/o USOC) | 25,216 | 57,639 | 12.9 | 36.0 | 186 | 561 | 0.89 (0.65 - 1.24) | 0.50 | 0.84 (0.60 - 1.16) | 0.31 |
| **PS-matching / on-treatment TAR without escalation** | | | | | | | | | | |
| CCAE | 9,140 | 22,776 | 5.4 | 16.6 | 70 | 176 | 1.39 (0.94 - 2.04) | 0.09 | 1.28 (0.87 - 1.89) | 0.22 |
| ODOD | 6,578 | 15,701 | 3.4 | 9.7 | 58 | 226 | 0.75 (0.50 - 1.10) | 0.15 | 0.77 (0.52 - 1.14) | 0.21 |
| MDCR | 619 | 1,857 | 0.4 | 1.3 | 12 | 46 | 0.85 (0.29 - 2.17) | 0.75 | 0.64 (0.23 - 1.73) | 0.38 |
| Open Claims | 133,612 | 297,193 | 82.2 | 222.6 | 784 | 2,506 | 0.98 (0.88 - 1.09) | 0.73 | 0.91 (0.81 - 1.02) | 0.15 |
| OEHR | 7,624 | 19,116 | 1.9 | 5.4 | 22 | 74 | 0.55 (0.27 - 1.03) | 0.08 | 0.50 (0.26 - 0.97) | 0.04 |
| VA | 1,072 | 9,618 | 0.6 | 5.0 | <10 | 55 | 0.80 (0.13 - 2.87) | 0.79 | 0.52 (0.11 - 2.48) | 0.41 |
| Meta-analysis | 158,026 | 364,404 | 93.5 | 259.3 | <944 | 3,037 | 0.92 (0.68 - 1.24) | 0.59 | 0.86 (0.64 - 1.16) | 0.33 |
| Meta-analysis (w/o USOC) | 24,414 | 67,211 | 11.3 | 36.7 | <160 | 531 | 0.87 (0.54 - 1.38) | 0.55 | 0.81 (0.51 - 1.29) | 0.38 |
| **PS-stratification / on-treatment TAR without escalation** | | | | | | | | | | |
| CCAE | 9,749 | 22,780 | 5.7 | 16.6 | 74 | 176 | 1.27 (0.95 - 1.69) | 0.11 | 1.16 (0.86 - 1.54) | 0.34 |
| ODOD | 7,135 | 15,713 | 3.6 | 9.7 | 62 | 226 | 0.77 (0.57 - 1.03) | 0.08 | 0.73 (0.54 - 0.98) | 0.04 |
| MDCR | 650 | 1,862 | 0.4 | 1.3 | 12 | 46 | 1.18 (0.58 - 2.24) | 0.63 | 0.95 (0.48 - 1.87) | 0.84 |
| Open Claims | 143,483 | 297,226 | 86.7 | 222.7 | 828 | 2,508 | 0.95 (0.87 - 1.03) | 0.20 | 0.90 (0.82 - 0.98) | 0.03 |
| OEHR | 8,332 | 19,146 | 2.0 | 5.4 | 26 | 75 | 0.80 (0.49 - 1.28) | 0.36 | 0.72 (0.44 - 1.16) | 0.18 |
| VA | 1,229 | 10,518 | 0.7 | 5.5 | <10 | 71 | 1.39 (0.40 - 3.66) | 0.56 | 1.00 (0.33 - 3.04) | 0.92 |
| Meta-analysis | 168,699 | 354,865 | 98.0 | 254.4 | 990 | 2,985 | 0.95 (0.78 - 1.15) | 0.59 | 0.89 (0.73 - 1.09) | 0.26 |
| Meta-analysis (w/o USOC) | 25,216 | 57,639 | 11.4 | 31.7 | 162 | 477 | 0.94 (0.66 - 1.33) | 0.72 | 0.86 (0.61 - 1.23) | 0.42 |

##### 1.1.3 Acute myocardial infarction

**Supplementary Table 3:** Exposures, outcomes and risk estimates for Acute myocardial infarction in GLP1RA (male) (T) vs SGLT2I (male) (C) new-user cohorts under four different propensity score (PS) adjustment / time-at-risk (TAR) designs across data sources. We report total patient exposure time in 1,000 patient-year (KYr) units, uncalibrated and calibrated hazard ratios (HRs) and their 95% confidence intervals (CIs). Greyed out entries did not pass study diagnostics and are included only for completeness.

| | Patients | | Exposure (KYr) | | Outcomes | | Uncalibrated | | Calibrated | |
|---|---|---|---|---|---|---|---|---|---|---|
| | T | C | T | C | T | C | HR (95% CI) | $p$ | HR (95% CI) | $p$ |
| **PS-matching / on-treatment TAR** | | | | | | | | | | |
| CCAE | 9,438 | 23,422 | 6.4 | 19.5 | 32 | 82 | 1.33 (0.72 - 2.38) | 0.35 | 1.23 (0.68 - 2.25) | 0.50 |
| ODOD | 6,953 | 16,763 | 4.0 | 11.7 | 16 | 101 | 0.49 (0.24 - 0.91) | 0.03 | 0.51 (0.26 - 0.99) | 0.05 |
| MDCR | 679 | 2,027 | 0.4 | 1.6 | 5 | 15 | 0.83 (0.17 - 3.20) | 0.80 | 0.63 (0.15 - 2.75) | 0.54 |
| Open Claims | 140,293 | 316,161 | 98.7 | 276.1 | 328 | 1,186 | 0.84 (0.72 - 0.99) | 0.04 | 0.80 (0.68 - 0.94) | 0.01 |
| OEHR | 7,822 | 19,712 | 2.2 | 6.3 | 9 | 26 | 0.98 (0.36 - 2.37) | 0.96 | 0.92 (0.36 - 2.34) | 0.86 |
| VA | 1,143 | 10,556 | 0.8 | 6.4 | <10 | 27 | 2.90 (0.69 - 9.70) | 0.11 | 1.97 (0.53 - 7.35) | 0.32 |
| Meta-analysis | 164,506 | 376,058 | 111.3 | 313.5 | 385 | 1,395 | 0.85 (0.63 - 1.16) | 0.31 | 0.81 (0.59 - 1.11) | 0.19 |
| Meta-analysis (w/o USOC) | 24,213 | 59,897 | 12.6 | 37.5 | 57 | 209 | 0.86 (0.45 - 1.64) | 0.64 | 0.82 (0.43 - 1.56) | 0.55 |
| **PS-stratification / on-treatment TAR** | | | | | | | | | | |
| CCAE | 10,071 | 23,425 | 6.7 | 19.5 | 33 | 82 | 1.38 (0.89 - 2.10) | 0.14 | 1.29 (0.84 - 1.97) | 0.25 |
| ODOD | 7,546 | 16,779 | 4.3 | 11.7 | 17 | 101 | 0.47 (0.27 - 0.78) | <0.01 | 0.45 (0.26 - 0.77) | <0.01 |
| MDCR | 709 | 2,032 | 0.5 | 1.6 | 5 | 15 | 1.12 (0.36 - 3.01) | 0.83 | 0.93 (0.32 - 2.70) | 0.88 |
| Open Claims | 150,437 | 316,207 | 103.6 | 276.1 | 344 | 1,187 | 0.85 (0.75 - 0.97) | 0.01 | 0.81 (0.72 - 0.93) | <0.01 |
| OEHR | 8,530 | 19,723 | 2.4 | 6.3 | 10 | 26 | 0.98 (0.43 - 2.07) | 0.95 | 0.88 (0.40 - 1.93) | 0.75 |
| VA | 1,307 | 11,651 | 0.9 | 7.0 | <10 | 35 | 3.66 (0.98 - 10.8) | 0.03 | 2.65 (0.80 - 8.79) | 0.11 |
| Meta-analysis | 176,584 | 376,134 | 117.0 | 313.6 | 404 | 1,396 | 0.86 (0.56 - 1.33) | 0.50 | 0.82 (0.53 - 1.27) | 0.37 |
| Meta-analysis (w/o USOC) | 26,147 | 59,927 | 13.4 | 37.5 | 60 | 209 | 0.86 (0.42 - 1.77) | 0.69 | 0.81 (0.39 - 1.66) | 0.57 |
| **PS-matching / on-treatment TAR without escalation** | | | | | | | | | | |
| CCAE | 9,438 | 23,422 | 5.6 | 17.0 | 29 | 70 | 1.53 (0.82 - 2.81) | 0.17 | 1.41 (0.76 - 2.62) | 0.28 |
| ODOD | 6,953 | 16,763 | 3.6 | 10.3 | 13 | 90 | 0.43 (0.20 - 0.85) | 0.02 | 0.45 (0.22 - 0.91) | 0.03 |
| MDCR | 679 | 2,027 | 0.4 | 1.4 | <5 | 14 | 0.31 (0.02 - 1.85) | 0.33 | 0.23 (0.02 - 2.47) | 0.23 |
| Open Claims | 140,293 | 316,161 | 86.1 | 234.6 | 272 | 999 | 0.86 (0.72 - 1.02) | 0.09 | 0.80 (0.66 - 0.95) | 0.02 |
| OEHR | 7,822 | 19,712 | 1.9 | 5.5 | 8 | 25 | 0.88 (0.30 - 2.23) | 0.79 | 0.80 (0.29 - 2.16) | 0.66 |
| VA | 1,143 | 10,556 | 0.6 | 5.5 | <10 | 23 | 2.82 (0.56 - 10.4) | 0.17 | 1.82 (0.42 - 7.87) | 0.43 |
| Meta-analysis | 164,506 | 376,058 | 97.2 | 267.5 | 322 | 1,184 | 0.86 (0.54 - 1.37) | 0.52 | 0.80 (0.50 - 1.29) | 0.36 |
| Meta-analysis (w/o USOC) | 24,213 | 59,897 | 11.1 | 32.9 | 50 | 185 | 0.85 (0.38 - 1.91) | 0.69 | 0.79 (0.35 - 1.78) | 0.57 |
| **PS-stratification / on-treatment TAR without escalation** | | | | | | | | | | |
| CCAE | 10,071 | 23,425 | 5.9 | 17.0 | 30 | 70 | 1.48 (0.93 - 2.30) | 0.09 | 1.34 (0.85 - 2.12) | 0.21 |
| ODOD | 7,546 | 16,779 | 3.8 | 10.3 | 14 | 90 | 0.43 (0.23 - 0.75) | <0.01 | 0.41 (0.23 - 0.74) | <0.01 |
| MDCR | 709 | 2,032 | 0.4 | 1.5 | <5 | 14 | 0.76 (0.17 - 2.41) | 0.68 | 0.61 (0.16 - 2.29) | 0.47 |
| Open Claims | 150,437 | 316,207 | 90.7 | 234.6 | 288 | 1,000 | 0.83 (0.72 - 0.95) | <0.01 | 0.79 (0.68 - 0.90) | <0.01 |
| OEHR | 8,530 | 19,723 | 2.1 | 5.5 | 9 | 25 | 0.90 (0.38 - 1.98) | 0.80 | 0.81 (0.35 - 1.85) | 0.61 |
| VA | 1,307 | 11,651 | 0.7 | 6.1 | <10 | 31 | 4.18 (1.11 - 12.5) | 0.02 | 3.02 (0.90 - 10.1) | 0.08 |
| Meta-analysis | 176,584 | 376,134 | 102.5 | 267.5 | 341 | 1,185 | 0.85 (0.52 - 1.37) | 0.50 | 0.79 (0.49 - 1.29) | 0.36 |
| Meta-analysis (w/o USOC) | 26,147 | 59,927 | 11.8 | 32.9 | 53 | 185 | 0.84 (0.38 - 1.88) | 0.68 | 0.78 (0.35 - 1.73) | 0.54 |

##### 1.1.4 Acute renal failure

**Supplementary Table 4:** Exposures, outcomes and risk estimates for Acute renal failure in GLP1RA (male) (T) vs SGLT2I (male) (C) new-user cohorts under four different propensity score (PS) adjustment / time-at-risk (TAR) designs across data sources. We report total patient exposure time in 1,000 patient-year (KYr) units, uncalibrated and calibrated hazard ratios (HRs) and their 95% confidence intervals (CIs). Greyed out entries did not pass study diagnostics and are included only for completeness.

| | Patients | | Exposure (KYr) | | Outcomes | | Uncalibrated | | Calibrated | |
|---|---|---|---|---|---|---|---|---|---|---|
| | T | C | T | C | T | C | HR (95% CI) | $p$ | HR (95% CI) | $p$ |
| **PS-matching / on-treatment TAR** | | | | | | | | | | |
| CCAE | 9,463 | 23,693 | 6.4 | 19.7 | 57 | 127 | 1.40 (0.91 - 2.14) | 0.13 | 1.30 (0.85 - 2.00) | 0.24 |
| ODOD | 6,861 | 16,732 | 4.0 | 11.7 | 79 | 198 | 1.25 (0.88 - 1.76) | 0.21 | 1.30 (0.92 - 1.84) | 0.15 |
| MDCR | 661 | 2,014 | 0.4 | 1.6 | 18 | 36 | 2.39 (1.03 - 5.62) | 0.04 | 1.83 (0.78 - 4.27) | 0.17 |
| Open Claims | 140,078 | 320,717 | 98.4 | 280.5 | 846 | 2,328 | 1.03 (0.92 - 1.14) | 0.62 | 0.97 (0.87 - 1.09) | 0.65 |
| OEHR | 7,820 | 19,901 | 2.2 | 6.4 | 28 | 66 | 0.98 (0.53 - 1.74) | 0.94 | 0.92 (0.51 - 1.67) | 0.79 |
| VA | 1,126 | 10,563 | 0.8 | 6.4 | <10 | 72 | 0.93 (0.30 - 2.43) | 0.89 | 0.63 (0.22 - 1.81) | 0.40 |
| Meta-analysis | 165,348 | 391,606 | 111.8 | 324.6 | <1,020 | 2,791 | 1.08 (0.94 - 1.24) | 0.26 | 1.03 (0.89 - 1.19) | 0.70 |
| Meta-analysis (w/o USOC) | 25,270 | 70,889 | 13.4 | 44.1 | <174 | 463 | 1.23 (0.96 - 1.56) | 0.10 | 1.17 (0.91 - 1.49) | 0.24 |
| **PS-stratification / on-treatment TAR** | | | | | | | | | | |
| CCAE | 10,081 | 23,699 | 6.7 | 19.7 | 58 | 127 | 1.22 (0.87 - 1.69) | 0.24 | 1.13 (0.81 - 1.58) | 0.46 |
| ODOD | 7,422 | 16,747 | 4.2 | 11.7 | 87 | 199 | 1.12 (0.85 - 1.46) | 0.40 | 1.08 (0.82 - 1.41) | 0.60 |
| MDCR | 692 | 2,014 | 0.4 | 1.6 | 19 | 36 | 1.97 (1.07 - 3.53) | 0.03 | 1.63 (0.90 - 2.96) | 0.12 |
| Open Claims | 149,976 | 320,733 | 103.3 | 280.5 | 897 | 2,329 | 1.10 (1.01 - 1.19) | 0.02 | 1.05 (0.97 - 1.15) | 0.33 |
| OEHR | 8,532 | 19,913 | 2.4 | 6.4 | 32 | 66 | 1.01 (0.64 - 1.58) | 0.95 | 0.91 (0.58 - 1.43) | 0.69 |
| VA | 1,290 | 11,731 | 0.9 | 7.1 | <10 | 93 | 1.02 (0.42 - 2.18) | 0.96 | 0.74 (0.32 - 1.68) | 0.47 |
| Meta-analysis | 176,011 | 381,092 | 116.6 | 318.2 | 1,074 | 2,721 | 1.10 (1.03 - 1.19) | <0.01 | 1.05 (0.96 - 1.15) | 0.32 |
| Meta-analysis (w/o USOC) | 26,035 | 60,359 | 13.3 | 37.8 | 177 | 392 | 1.13 (0.94 - 1.37) | 0.19 | 1.06 (0.88 - 1.29) | 0.56 |
| **PS-matching / on-treatment TAR without escalation** | | | | | | | | | | |
| CCAE | 9,463 | 23,693 | 5.6 | 17.2 | 49 | 102 | 1.47 (0.93 - 2.31) | 0.10 | 1.36 (0.86 - 2.14) | 0.20 |
| ODOD | 6,861 | 16,732 | 3.5 | 10.4 | 69 | 166 | 1.26 (0.87 - 1.81) | 0.22 | 1.29 (0.89 - 1.87) | 0.19 |
| MDCR | 661 | 2,014 | 0.4 | 1.5 | 14 | 32 | 2.28 (0.92 - 5.81) | 0.08 | 1.71 (0.68 - 4.30) | 0.27 |
| Open Claims | 140,078 | 320,717 | 85.9 | 238.6 | 707 | 1,919 | 1.01 (0.90 - 1.13) | 0.90 | 0.94 (0.83 - 1.06) | 0.33 |
| OEHR | 7,820 | 19,901 | 1.9 | 5.6 | 24 | 46 | 1.23 (0.62 - 2.34) | 0.54 | 1.12 (0.58 - 2.17) | 0.75 |
| VA | 1,126 | 10,563 | 0.6 | 5.5 | <10 | 61 | 0.88 (0.24 - 2.55) | 0.84 | 0.57 (0.18 - 1.85) | 0.36 |
| Meta-analysis | 165,348 | 391,606 | 97.6 | 277.4 | <859 | 2,294 | 1.12 (0.93 - 1.36) | 0.24 | 1.05 (0.87 - 1.27) | 0.62 |
| Meta-analysis (w/o USOC) | 25,270 | 70,889 | 11.7 | 38.7 | <152 | 375 | 1.30 (1.00 - 1.68) | 0.05 | 1.21 (0.93 - 1.57) | 0.19 |
| **PS-stratification / on-treatment TAR without escalation** | | | | | | | | | | |
| CCAE | 10,081 | 23,699 | 5.9 | 17.3 | 50 | 102 | 1.26 (0.87 - 1.79) | 0.21 | 1.14 (0.80 - 1.64) | 0.47 |
| ODOD | 7,422 | 16,747 | 3.8 | 10.4 | 77 | 167 | 1.24 (0.92 - 1.65) | 0.15 | 1.16 (0.87 - 1.56) | 0.32 |
| MDCR | 692 | 2,014 | 0.4 | 1.5 | 15 | 32 | 1.60 (0.82 - 3.03) | 0.16 | 1.29 (0.67 - 2.48) | 0.47 |
| Open Claims | 149,976 | 320,733 | 90.3 | 238.7 | 752 | 1,920 | 1.10 (1.01 - 1.20) | 0.03 | 1.04 (0.95 - 1.14) | 0.44 |
| OEHR | 8,532 | 19,913 | 2.1 | 5.6 | 27 | 46 | 1.34 (0.79 - 2.21) | 0.27 | 1.20 (0.72 - 1.99) | 0.50 |
| VA | 1,290 | 11,731 | 0.7 | 6.2 | <10 | 80 | 1.12 (0.43 - 2.54) | 0.80 | 0.81 (0.33 - 1.97) | 0.64 |
| Meta-analysis | 176,011 | 381,092 | 102.1 | 271.9 | 906 | 2,235 | 1.12 (1.04 - 1.22) | <0.01 | 1.06 (0.96 - 1.16) | 0.27 |
| Meta-analysis (w/o USOC) | 26,035 | 60,359 | 11.7 | 33.2 | 154 | 315 | 1.26 (1.02 - 1.55) | 0.03 | 1.16 (0.94 - 1.43) | 0.18 |

##### 1.1.5 Glycemic control

**Supplementary Table 5:** Exposures, outcomes and risk estimates for Glycemic control in GLP1RA (male) (T) vs SGLT2I (male) (C) new-user cohorts under four different propensity score (PS) adjustment / time-at-risk (TAR) designs across data sources. We report total patient exposure time in 1,000 patient-year (KYr) units, uncalibrated and calibrated hazard ratios (HRs) and their 95% confidence intervals (CIs). Greyed out entries did not pass study diagnostics and are included only for completeness.

| | Patients | | Exposure (KYr) | | Outcomes | | Uncalibrated | | Calibrated | |
|---|---|---|---|---|---|---|---|---|---|---|
| | T | C | T | C | T | C | HR (95% CI) | $p$ | HR (95% CI) | $p$ |
| **PS-matching / on-treatment TAR** | | | | | | | | | | |
| CCAE | 9,646 | 23,972 | 6.6 | 20.0 | <5 | 26 | 0.33 (0.05 - 1.26) | 0.18 | 0.31 (0.06 - 1.54) | 0.15 |
| ODOD | 4,723 | 11,454 | 2.5 | 7.3 | 280 | 760 | 1.11 (0.91 - 1.33) | 0.30 | 1.15 (0.95 - 1.39) | 0.19 |
| MDCR | 700 | 2,128 | 0.4 | 1.7 | <5 | 0 | | | | |
| Open Claims | 145,214 | 332,772 | 102.0 | 289.6 | 0 | 0 | | | | |
| OEHR | 7,951 | 20,219 | 2.2 | 6.5 | 0 | 0 | | | | |
| VA | 1,123 | 10,656 | 0.8 | 6.4 | 0 | 10 | | | | |
| Meta-analysis | 14,369 | 35,426 | 9.1 | 27.2 | <285 | 786 | 0.80 (0.28 - 2.27) | 0.67 | 0.76 (0.27 - 2.16) | 0.61 |
| Meta-analysis (w/o USOC) | 14,369 | 35,426 | 9.1 | 27.2 | <285 | 786 | 0.80 (0.28 - 2.27) | 0.67 | 0.76 (0.27 - 2.16) | 0.61 |
| **PS-stratification / on-treatment TAR** | | | | | | | | | | |
| CCAE | 10,271 | 23,976 | 6.9 | 20.0 | <5 | 26 | 0.57 (0.16 - 1.51) | 0.32 | 0.53 (0.17 - 1.60) | 0.26 |
| ODOD | 5,040 | 11,471 | 2.7 | 7.3 | 299 | 761 | 1.14 (0.99 - 1.31) | 0.08 | 1.09 (0.95 - 1.26) | 0.28 |
| MDCR | 730 | 2,128 | 0.5 | 1.7 | <5 | 0 | | | | |
| Open Claims | 155,418 | 332,801 | 107.0 | 289.7 | 0 | 0 | | | | |
| OEHR | 8,668 | 20,231 | 2.4 | 6.5 | 0 | 0 | | | | |
| VA | 1,288 | 11,946 | 0.9 | 7.2 | 0 | 12 | | | | |
| Meta-analysis | 15,311 | 35,447 | 9.5 | 27.2 | <304 | 787 | 1.01 (0.61 - 1.68) | 0.97 | 0.96 (0.57 - 1.60) | 0.87 |
| Meta-analysis (w/o USOC) | 15,311 | 35,447 | 9.5 | 27.2 | <304 | 787 | 1.01 (0.61 - 1.68) | 0.97 | 0.95 (0.57 - 1.58) | 0.84 |
| **PS-matching / on-treatment TAR without escalation** | | | | | | | | | | |
| CCAE | 9,646 | 23,972 | 5.7 | 17.4 | <5 | 25 | 0.39 (0.06 - 1.50) | 0.26 | 0.36 (0.07 - 1.81) | 0.22 |
| ODOD | 4,723 | 11,454 | 2.2 | 6.5 | 251 | 668 | 1.16 (0.95 - 1.41) | 0.15 | 1.19 (0.97 - 1.45) | 0.14 |
| MDCR | 700 | 2,128 | 0.4 | 1.5 | <5 | 0 | | | | |
| Open Claims | 145,214 | 332,772 | 88.9 | 246.2 | 0 | 0 | | | | |
| OEHR | 7,951 | 20,219 | 1.9 | 5.7 | 0 | 0 | | | | |
| VA | 1,123 | 10,656 | 0.6 | 5.5 | 0 | <10 | | | | |
| Meta-analysis | 14,369 | 35,426 | 8.0 | 23.9 | <256 | 693 | 0.92 (0.38 - 2.19) | 0.85 | 0.86 (0.36 - 2.05) | 0.73 |
| Meta-analysis (w/o USOC) | 14,369 | 35,426 | 8.0 | 23.9 | <256 | 693 | 0.92 (0.38 - 2.19) | 0.85 | 0.85 (0.36 - 2.04) | 0.72 |
| **PS-stratification / on-treatment TAR without escalation** | | | | | | | | | | |
| CCAE | 10,271 | 23,976 | 6.0 | 17.4 | <5 | 25 | 0.50 (0.12 - 1.46) | 0.28 | 0.46 (0.13 - 1.60) | 0.22 |
| ODOD | 5,040 | 11,471 | 2.4 | 6.5 | 269 | 669 | 1.18 (1.02 - 1.37) | 0.03 | 1.11 (0.96 - 1.30) | 0.20 |
| MDCR | 730 | 2,128 | 0.4 | 1.5 | <5 | 0 | | | | |
| Open Claims | 155,418 | 332,801 | 93.5 | 246.2 | 0 | 0 | | | | |
| OEHR | 8,668 | 20,231 | 2.1 | 5.7 | 0 | 0 | | | | |
| VA | 1,288 | 11,946 | 0.7 | 6.2 | 0 | <10 | | | | |
| Meta-analysis | 15,311 | 35,447 | 8.4 | 23.9 | <274 | 694 | 0.97 (0.48 - 1.97) | 0.94 | 0.91 (0.45 - 1.86) | 0.81 |
| Meta-analysis (w/o USOC) | 15,311 | 35,447 | 8.4 | 23.9 | <274 | 694 | 0.97 (0.48 - 1.97) | 0.94 | 0.90 (0.44 - 1.82) | 0.76 |

##### 1.1.6 Hospitalization with heart failure

**Supplementary Table 6:** Exposures, outcomes and risk estimates for Hospitalization with heart failure in GLP1RA (male) (T) vs SGLT2I (male) (C) new-user cohorts under four different propensity score (PS) adjustment / time-at-risk (TAR) designs across data sources. We report total patient exposure time in 1,000 patient-year (KYr) units, uncalibrated and calibrated hazard ratios (HRs) and their 95% confidence intervals (CIs). Greyed out entries did not pass study diagnostics and are included only for completeness.

| | Patients | | Exposure (KYr) | | Outcomes | | Uncalibrated | | Calibrated | |
|---|---|---|---|---|---|---|---|---|---|---|
| | T | C | T | C | T | C | HR (95% CI) | $p$ | HR (95% CI) | $p$ |
| **PS-matching / on-treatment TAR** | | | | | | | | | | |
| CCAE | 9,478 | 23,592 | 6.4 | 19.6 | 44 | 113 | 1.16 (0.69 - 1.93) | 0.56 | 1.08 (0.65 - 1.82) | 0.77 |
| ODOD | 6,804 | 16,405 | 3.9 | 11.5 | 47 | 163 | 1.01 (0.64 - 1.57) | 0.96 | 1.05 (0.67 - 1.65) | 0.82 |
| MDCR | 648 | 1,957 | 0.4 | 1.6 | 11 | 32 | 0.78 (0.24 - 2.18) | 0.66 | 0.60 (0.20 - 1.80) | 0.36 |
| Open Claims | 139,941 | 315,167 | 98.5 | 277.3 | 507 | 1,629 | 1.00 (0.88 - 1.15) | 0.94 | 0.95 (0.83 - 1.09) | 0.50 |
| OEHR | 7,782 | 19,667 | 2.2 | 6.3 | 12 | 63 | 0.29 (0.11 - 0.64) | <0.01 | 0.27 (0.11 - 0.66) | <0.01 |
| VA | 1,125 | 10,173 | 0.8 | 6.2 | <10 | 56 | 0.52 (0.08 - 1.89) | 0.42 | 0.35 (0.07 - 1.72) | 0.20 |
| Meta-analysis | 165,130 | 385,004 | 111.8 | 320.9 | <620 | 2,024 | 0.99 (0.87 - 1.11) | 0.82 | 0.94 (0.82 - 1.07) | 0.35 |
| Meta-analysis (w/o USOC) | 25,189 | 69,837 | 13.3 | 43.6 | <113 | 395 | 0.75 (0.42 - 1.35) | 0.34 | 0.72 (0.40 - 1.28) | 0.27 |
| **PS-stratification / on-treatment TAR** | | | | | | | | | | |
| CCAE | 10,091 | 23,598 | 6.7 | 19.7 | 47 | 113 | 1.13 (0.78 - 1.61) | 0.51 | 1.05 (0.73 - 1.51) | 0.79 |
| ODOD | 7,372 | 16,421 | 4.2 | 11.5 | 51 | 163 | 0.93 (0.66 - 1.29) | 0.68 | 0.90 (0.64 - 1.25) | 0.53 |
| MDCR | 679 | 1,962 | 0.4 | 1.6 | 11 | 33 | 1.26 (0.59 - 2.52) | 0.53 | 1.04 (0.50 - 2.16) | 0.87 |
| Open Claims | 150,000 | 315,211 | 103.5 | 277.3 | 537 | 1,631 | 1.01 (0.91 - 1.12) | 0.87 | 0.96 (0.87 - 1.07) | 0.55 |
| OEHR | 8,499 | 19,700 | 2.4 | 6.3 | 16 | 64 | 0.56 (0.30 - 0.97) | 0.05 | 0.50 (0.28 - 0.90) | 0.02 |
| VA | 1,287 | 11,226 | 0.9 | 6.8 | <10 | 73 | 0.49 (0.08 - 1.68) | 0.36 | 0.35 (0.07 - 1.64) | 0.19 |
| Meta-analysis | 175,962 | 374,930 | 116.8 | 314.8 | 651 | 1,971 | 0.99 (0.91 - 1.09) | 0.91 | 0.94 (0.85 - 1.05) | 0.32 |
| Meta-analysis (w/o USOC) | 25,962 | 59,719 | 13.3 | 37.5 | 114 | 340 | 0.89 (0.64 - 1.25) | 0.51 | 0.84 (0.60 - 1.18) | 0.32 |
| **PS-matching / on-treatment TAR without escalation** | | | | | | | | | | |
| CCAE | 9,478 | 23,592 | 5.6 | 17.2 | 39 | 93 | 1.33 (0.77 - 2.28) | 0.30 | 1.23 (0.71 - 2.12) | 0.47 |
| ODOD | 6,804 | 16,405 | 3.5 | 10.2 | 44 | 144 | 1.02 (0.63 - 1.61) | 0.92 | 1.05 (0.66 - 1.67) | 0.84 |
| MDCR | 648 | 1,957 | 0.4 | 1.4 | 9 | 28 | 0.59 (0.13 - 1.95) | 0.44 | 0.44 (0.11 - 1.71) | 0.24 |
| Open Claims | 139,941 | 315,167 | 85.9 | 235.8 | 434 | 1,367 | 1.05 (0.91 - 1.22) | 0.47 | 0.98 (0.85 - 1.14) | 0.80 |
| OEHR | 7,782 | 19,667 | 1.9 | 5.5 | 11 | 49 | 0.32 (0.11 - 0.77) | 0.02 | 0.29 (0.11 - 0.78) | 0.01 |
| VA | 1,125 | 10,173 | 0.6 | 5.3 | <10 | 51 | 0.67 (0.10 - 2.57) | 0.63 | 0.43 (0.08 - 2.18) | 0.31 |
| Meta-analysis | 165,130 | 385,004 | 97.5 | 274.0 | <538 | 1,704 | 1.04 (0.91 - 1.19) | 0.55 | 0.98 (0.85 - 1.12) | 0.73 |
| Meta-analysis (w/o USOC) | 25,189 | 69,837 | 11.7 | 38.2 | <104 | 337 | 0.86 (0.49 - 1.49) | 0.59 | 0.80 (0.46 - 1.39) | 0.43 |
| **PS-stratification / on-treatment TAR without escalation** | | | | | | | | | | |
| CCAE | 10,091 | 23,598 | 5.9 | 17.2 | 42 | 93 | 1.27 (0.85 - 1.87) | 0.23 | 1.16 (0.78 - 1.71) | 0.47 |
| ODOD | 7,372 | 16,421 | 3.7 | 10.2 | 48 | 144 | 0.99 (0.70 - 1.39) | 0.97 | 0.93 (0.66 - 1.32) | 0.71 |
| MDCR | 679 | 1,962 | 0.4 | 1.4 | 9 | 28 | 1.20 (0.51 - 2.58) | 0.66 | 0.96 (0.43 - 2.17) | 0.88 |
| Open Claims | 150,000 | 315,211 | 90.4 | 235.8 | 462 | 1,369 | 1.02 (0.91 - 1.13) | 0.79 | 0.96 (0.86 - 1.08) | 0.55 |
| OEHR | 8,499 | 19,700 | 2.1 | 5.5 | 15 | 50 | 0.72 (0.37 - 1.30) | 0.29 | 0.64 (0.34 - 1.20) | 0.17 |
| VA | 1,287 | 11,226 | 0.7 | 5.9 | <10 | 66 | 0.54 (0.08 - 1.91) | 0.44 | 0.39 (0.08 - 1.86) | 0.24 |
| Meta-analysis | 175,962 | 374,930 | 102.2 | 268.7 | 567 | 1,656 | 1.02 (0.92 - 1.13) | 0.71 | 0.96 (0.86 - 1.07) | 0.46 |
| Meta-analysis (w/o USOC) | 25,962 | 59,719 | 11.7 | 32.9 | 105 | 287 | 1.03 (0.78 - 1.35) | 0.83 | 0.95 (0.72 - 1.25) | 0.72 |

##### 1.1.7 Measured renal dysfunction

**Supplementary Table 7:** Exposures, outcomes and risk estimates for Measured renal dysfunction in GLP1RA (male) (T) vs SGLT2I (male) (C) new-user cohorts under four different propensity score (PS) adjustment / time-at-risk (TAR) designs across data sources. We report total patient exposure time in 1,000 patient-year (KYr) units, uncalibrated and calibrated hazard ratios (HRs) and their 95% confidence intervals (CIs). Greyed out entries did not pass study diagnostics and are included only for completeness.

| | Patients | | Exposure (KYr) | | Outcomes | | Uncalibrated | | Calibrated | |
|---|---|---|---|---|---|---|---|---|---|---|
| | T | C | T | C | T | C | HR (95% CI) | $p$ | HR (95% CI) | $p$ |
| **PS-matching / on-treatment TAR** | | | | | | | | | | |
| CCAE | 9,689 | 24,078 | 6.6 | 20.0 | 0 | 5 | | | | |
| ODOD | 7,134 | 17,452 | 4.1 | 12.1 | 6 | 6 | 2.54 (0.54 - 12.1) | 0.24 | 2.64 (0.56 - 12.5) | 0.22 |
| MDCR | 701 | 2,136 | 0.4 | 1.7 | 0 | 0 | | | | |
| Open Claims | 145,214 | 332,772 | 102.0 | 289.6 | 0 | 0 | | | | |
| OEHR | 7,951 | 20,219 | 2.2 | 6.5 | 0 | 0 | | | | |
| VA | 1,166 | 11,040 | 0.8 | 6.7 | <10 | 15 | 3.67 (0.93 - 13.7) | 0.06 | 2.49 (0.65 - 9.53) | 0.19 |
| **PS-stratification / on-treatment TAR** | | | | | | | | | | |
| CCAE | 10,321 | 24,082 | 6.9 | 20.0 | 0 | 5 | | | | |
| ODOD | 7,725 | 17,471 | 4.4 | 12.1 | 6 | 7 | 2.98 (0.86 - 9.91) | 0.08 | 2.86 (0.85 - 9.70) | 0.09 |
| MDCR | 731 | 2,136 | 0.5 | 1.7 | 0 | 0 | | | | |
| Open Claims | 155,418 | 332,801 | 107.0 | 289.7 | 0 | 0 | | | | |
| OEHR | 8,668 | 20,231 | 2.4 | 6.5 | 0 | 0 | | | | |
| VA | 1,336 | 12,350 | 0.9 | 7.4 | <10 | 23 | 3.12 (0.87 - 9.80) | 0.07 | 2.25 (0.67 - 7.58) | 0.19 |
| **PS-matching / on-treatment TAR without escalation** | | | | | | | | | | |
| CCAE | 9,689 | 24,078 | 5.8 | 17.5 | 0 | 5 | | | | |
| ODOD | 7,134 | 17,452 | 3.7 | 10.7 | <5 | 6 | 1.44 (0.19 - 8.24) | 0.71 | 1.47 (0.22 - 9.84) | 0.69 |
| MDCR | 701 | 2,136 | 0.4 | 1.5 | 0 | 0 | | | | |
| Open Claims | 145,214 | 332,772 | 88.9 | 246.2 | 0 | 0 | | | | |
| OEHR | 7,951 | 20,219 | 1.9 | 5.7 | 0 | 0 | | | | |
| VA | 1,166 | 11,040 | 0.6 | 5.8 | <10 | 13 | 3.21 (0.67 - 14.1) | 0.13 | 2.07 (0.45 - 9.52) | 0.35 |
| **PS-stratification / on-treatment TAR without escalation** | | | | | | | | | | |
| CCAE | 10,321 | 24,082 | 6.0 | 17.5 | 0 | 5 | | | | |
| ODOD | 7,725 | 17,471 | 3.9 | 10.8 | <5 | 7 | 1.99 (0.47 - 7.50) | 0.33 | 1.88 (0.47 - 7.54) | 0.38 |
| MDCR | 731 | 2,136 | 0.4 | 1.5 | 0 | 0 | | | | |
| Open Claims | 155,418 | 332,801 | 93.5 | 246.2 | 0 | 0 | | | | |
| OEHR | 8,668 | 20,231 | 2.1 | 5.7 | 0 | 0 | | | | |
| VA | 1,336 | 12,350 | 0.7 | 6.4 | <10 | 18 | 3.86 (0.93 - 13.4) | 0.05 | 2.79 (0.73 - 10.6) | 0.14 |

##### 1.1.8 Stroke

**Supplementary Table 8:** Exposures, outcomes and risk estimates for Stroke in GLP1RA (male) (T) vs SGLT2I (male) (C) new-user cohorts under four different propensity score (PS) adjustment / time-at-risk (TAR) designs across data sources. We report total patient exposure time in 1,000 patient-year (KYr) units, uncalibrated and calibrated hazard ratios (HRs) and their 95% confidence intervals (CIs). Greyed out entries did not pass study diagnostics and are included only for completeness.

| | Patients | | Exposure (KYr) | | Outcomes | | Uncalibrated | | Calibrated | |
|---|---|---|---|---|---|---|---|---|---|---|
| | T | C | T | C | T | C | HR (95% CI) | $p$ | HR (95% CI) | $p$ |
| **PS-matching / on-treatment TAR** | | | | | | | | | | |
| CCAE | 9,572 | 23,839 | 6.5 | 19.9 | 20 | 58 | 1.02 (0.51 - 1.94) | 0.96 | 0.94 (0.48 - 1.85) | 0.87 |
| ODOD | 7,003 | 17,071 | 4.1 | 11.8 | 19 | 81 | 0.72 (0.37 - 1.32) | 0.31 | 0.75 (0.40 - 1.41) | 0.38 |
| MDCR | 675 | 2,074 | 0.4 | 1.7 | <5 | 18 | | 0.14 | | 0.21 |
| Open Claims | 142,136 | 324,458 | 99.9 | 283.2 | 277 | 934 | 0.85 (0.71 - 1.02) | 0.09 | 0.81 (0.67 - 0.97) | 0.03 |
| OEHR | 7,882 | 20,026 | 2.2 | 6.4 | 11 | 19 | 1.35 (0.43 - 4.02) | 0.60 | 1.27 (0.42 - 3.88) | 0.68 |
| VA | 1,158 | 10,944 | 0.8 | 6.6 | <10 | 12 | 1.47 (0.07 - 13.6) | 0.78 | 0.99 (0.07 - 14.3) | 0.96 |
| Meta-analysis | 166,593 | 385,394 | 112.7 | 321.3 | 327 | 1,092 | 0.86 (0.73 - 1.02) | 0.08 | 0.82 (0.69 - 0.98) | 0.03 |
| Meta-analysis (w/o USOC) | 24,457 | 60,936 | 12.7 | 38.1 | 50 | 158 | 0.91 (0.59 - 1.39) | 0.65 | 0.86 (0.56 - 1.32) | 0.51 |
| **PS-stratification / on-treatment TAR** | | | | | | | | | | |
| CCAE | 10,198 | 23,843 | 6.8 | 19.9 | 21 | 58 | 1.12 (0.65 - 1.85) | 0.67 | 1.04 (0.62 - 1.75) | 0.87 |
| ODOD | 7,593 | 17,093 | 4.3 | 11.8 | 21 | 81 | 0.78 (0.46 - 1.27) | 0.34 | 0.75 (0.45 - 1.25) | 0.28 |
| MDCR | 707 | 2,074 | 0.4 | 1.7 | <5 | 18 | 1.69 (0.47 - 4.83) | 0.38 | 1.40 (0.44 - 4.48) | 0.57 |
| Open Claims | 152,244 | 324,481 | 104.9 | 283.2 | 291 | 934 | 0.95 (0.82 - 1.09) | 0.44 | 0.91 (0.79 - 1.04) | 0.20 |
| OEHR | 8,594 | 20,037 | 2.4 | 6.4 | 11 | 19 | 1.38 (0.61 - 2.98) | 0.42 | 1.24 (0.56 - 2.74) | 0.59 |
| VA | 1,329 | 12,216 | 0.9 | 7.4 | <10 | 19 | 0.84 (0.04 - 5.67) | 0.89 | 0.61 (0.05 - 7.17) | 0.69 |
| Meta-analysis | 178,629 | 385,454 | 118.4 | 321.3 | 344 | 1,092 | 0.95 (0.84 - 1.08) | 0.47 | 0.91 (0.79 - 1.04) | 0.17 |
| Meta-analysis (w/o USOC) | 26,385 | 60,973 | 13.5 | 38.1 | 53 | 158 | 1.00 (0.72 - 1.39) | 0.99 | 0.94 (0.67 - 1.31) | 0.71 |
| **PS-matching / on-treatment TAR without escalation** | | | | | | | | | | |
| CCAE | 9,572 | 23,839 | 5.7 | 17.3 | 17 | 46 | 1.04 (0.49 - 2.09) | 0.93 | 0.95 (0.46 - 1.98) | 0.90 |
| ODOD | 7,003 | 17,071 | 3.6 | 10.5 | 17 | 72 | 0.74 (0.36 - 1.41) | 0.38 | 0.75 (0.38 - 1.49) | 0.43 |
| MDCR | 675 | 2,074 | 0.4 | 1.5 | <5 | 15 | | 0.22 | | 0.31 |
| Open Claims | 142,136 | 324,458 | 87.1 | 240.8 | 233 | 806 | 0.87 (0.71 - 1.05) | 0.16 | 0.80 (0.66 - 0.98) | 0.04 |
| OEHR | 7,882 | 20,026 | 1.9 | 5.6 | 9 | 12 | 1.81 (0.54 - 6.03) | 0.34 | 1.64 (0.49 - 5.48) | 0.42 |
| VA | 1,158 | 10,944 | 0.6 | 5.7 | <10 | 10 | 2.74 (0.09 - 32.6) | 0.50 | 1.77 (0.09 - 33.8) | 0.71 |
| Meta-analysis | 166,593 | 385,394 | 98.3 | 274.2 | 276 | 936 | 0.88 (0.73 - 1.06) | 0.17 | 0.82 (0.68 - 0.99) | 0.04 |
| Meta-analysis (w/o USOC) | 24,457 | 60,936 | 11.2 | 33.5 | 43 | 130 | 0.96 (0.61 - 1.52) | 0.87 | 0.90 (0.56 - 1.42) | 0.65 |
| **PS-stratification / on-treatment TAR without escalation** | | | | | | | | | | |
| CCAE | 10,198 | 23,843 | 6.0 | 17.3 | 18 | 46 | 1.14 (0.63 - 1.96) | 0.66 | 1.03 (0.59 - 1.82) | 0.91 |
| ODOD | 7,593 | 17,093 | 3.9 | 10.5 | 19 | 72 | 0.81 (0.46 - 1.35) | 0.43 | 0.76 (0.45 - 1.30) | 0.32 |
| MDCR | 707 | 2,074 | 0.4 | 1.5 | <5 | 15 | 1.38 (0.31 - 4.54) | 0.64 | 1.11 (0.29 - 4.27) | 0.88 |
| Open Claims | 152,244 | 324,481 | 91.7 | 240.8 | 245 | 806 | 0.91 (0.78 - 1.05) | 0.19 | 0.86 (0.74 - 1.00) | 0.06 |
| OEHR | 8,594 | 20,037 | 2.1 | 5.6 | 9 | 12 | 1.71 (0.66 - 4.28) | 0.26 | 1.53 (0.60 - 3.89) | 0.38 |
| VA | 1,329 | 12,216 | 0.7 | 6.4 | <10 | 14 | 1.69 (0.09 - 10.5) | 0.67 | 1.22 (0.11 - 13.5) | 0.87 |
| Meta-analysis | 178,629 | 385,454 | 103.6 | 274.3 | 291 | 936 | 0.92 (0.80 - 1.06) | 0.26 | 0.87 (0.75 - 1.00) | 0.06 |
| Meta-analysis (w/o USOC) | 26,385 | 60,973 | 11.9 | 33.5 | 46 | 130 | 1.03 (0.72 - 1.48) | 0.85 | 0.95 (0.66 - 1.37) | 0.80 |

##### 1.1.9 Sudden cardiac death

**Supplementary Table 9:** Exposures, outcomes and risk estimates for Sudden cardiac death in GLP1RA (male) (T) vs SGLT2I (male) (C) new-user cohorts under four different propensity score (PS) adjustment / time-at-risk (TAR) designs across data sources. We report total patient exposure time in 1,000 patient-year (KYr) units, uncalibrated and calibrated hazard ratios (HRs) and their 95% confidence intervals (CIs). Greyed out entries did not pass study diagnostics and are included only for completeness.

| | Patients | | Exposure (KYr) | | Outcomes | | Uncalibrated | | Calibrated | |
|---|---|---|---|---|---|---|---|---|---|---|
| | T | C | T | C | T | C | HR (95% CI) | $p$ | HR (95% CI) | $p$ |
| **PS-matching / on-treatment TAR** | | | | | | | | | | |
| CCAE | 9,696 | 24,107 | 6.6 | 20.1 | <5 | 15 | 0.00 (0.00 - 0.84) | 0.35 | 0.00 (0.00 - 1647995077) | 0.35 |
| ODOD | 7,129 | 17,424 | 4.1 | 12.1 | 8 | 19 | 0.90 (0.27 - 2.66) | 0.86 | 0.94 (0.30 - 2.94) | 0.91 |
| MDCR | 698 | 2,130 | 0.4 | 1.7 | <5 | 11 | | | | |
| Open Claims | 144,881 | 331,659 | 101.8 | 288.8 | 85 | 258 | 1.01 (0.72 - 1.41) | 0.96 | 0.95 (0.68 - 1.34) | 0.79 |
| OEHR | 7,945 | 20,193 | 2.2 | 6.5 | <5 | 5 | 1.50 (0.17 - 14.3) | 0.72 | 1.41 (0.15 - 13.1) | 0.76 |
| VA | 1,181 | 11,164 | 0.8 | 6.7 | 0 | <10 | | | | |
| Meta-analysis | 161,706 | 373,190 | 112.5 | 321.0 | <98 | 292 | 1.00 (0.72 - 1.38) | 1.00 | 0.95 (0.68 - 1.32) | 0.76 |
| Meta-analysis (w/o USOC) | 16,825 | 41,531 | 10.7 | 32.2 | <13 | 34 | 0.89 (0.29 - 2.78) | 0.84 | 0.85 (0.27 - 2.65) | 0.78 |
| **PS-stratification / on-treatment TAR** | | | | | | | | | | |
| CCAE | 10,325 | 24,111 | 6.9 | 20.1 | <5 | 15 | 0.54 (0.12 - 1.71) | 0.37 | 0.51 (0.14 - 1.89) | 0.31 |
| ODOD | 7,720 | 17,442 | 4.4 | 12.1 | 9 | 19 | 1.23 (0.51 - 2.79) | 0.63 | 1.19 (0.51 - 2.78) | 0.69 |
| MDCR | 728 | 2,130 | 0.5 | 1.7 | <5 | 11 | 0.34 (0.02 - 1.79) | 0.36 | 0.28 (0.03 - 2.77) | 0.28 |
| Open Claims | 155,084 | 331,685 | 106.8 | 288.8 | 87 | 259 | 1.03 (0.80 - 1.33) | 0.80 | 0.99 (0.76 - 1.28) | 0.93 |
| OEHR | 8,664 | 20,205 | 2.4 | 6.5 | 5 | 5 | 1.93 (0.45 - 8.51) | 0.38 | 1.74 (0.40 - 7.53) | 0.46 |
| VA | 1,352 | 12,496 | 0.9 | 7.5 | 0 | <10 | | | | |
| Meta-analysis | 173,129 | 373,238 | 118.1 | 321.0 | <101 | 293 | 1.03 (0.81 - 1.31) | 0.83 | 0.98 (0.76 - 1.25) | 0.84 |
| Meta-analysis (w/o USOC) | 18,045 | 41,553 | 11.3 | 32.2 | <14 | 34 | 0.96 (0.46 - 2.01) | 0.92 | 0.90 (0.43 - 1.89) | 0.79 |
| **PS-matching / on-treatment TAR without escalation** | | | | | | | | | | |
| CCAE | 9,696 | 24,107 | 5.8 | 17.5 | <5 | 11 | | | | |
| ODOD | 7,129 | 17,424 | 3.7 | 10.7 | 7 | 16 | 0.94 (0.24 - 3.12) | 0.93 | 0.97 (0.27 - 3.46) | 0.95 |
| MDCR | 698 | 2,130 | 0.4 | 1.5 | <5 | 10 | | | | |
| Open Claims | 144,881 | 331,659 | 88.7 | 245.4 | 70 | 227 | 1.05 (0.73 - 1.49) | 0.81 | 0.97 (0.68 - 1.39) | 0.88 |
| OEHR | 7,945 | 20,193 | 1.9 | 5.7 | <5 | <5 | 3.70 (0.31 - 88.7) | 0.37 | 3.37 (0.20 - 57.2) | 0.40 |
| VA | 1,181 | 11,164 | 0.6 | 5.8 | 0 | <10 | | | | |
| Meta-analysis | 152,010 | 349,083 | 92.4 | 256.2 | 77 | 243 | 1.04 (0.73 - 1.47) | 0.84 | 0.97 (0.69 - 1.38) | 0.87 |
| Meta-analysis (w/o USOC) | 7,129 | 17,424 | 3.7 | 10.7 | 7 | 16 | 0.94 (0.24 - 3.12) | 0.93 | 0.88 (0.24 - 3.15) | 0.84 |
| **PS-stratification / on-treatment TAR without escalation** | | | | | | | | | | |
| CCAE | 10,325 | 24,111 | 6.0 | 17.5 | <5 | 11 | 0.29 (0.02 - 1.55) | 0.29 | 0.27 (0.03 - 2.62) | 0.26 |
| ODOD | 7,720 | 17,442 | 3.9 | 10.7 | 8 | 16 | 1.39 (0.54 - 3.31) | 0.48 | 1.30 (0.53 - 3.24) | 0.57 |
| MDCR | 728 | 2,130 | 0.4 | 1.5 | <5 | 10 | 0.44 (0.02 - 2.45) | 0.49 | 0.35 (0.03 - 3.60) | 0.38 |
| Open Claims | 155,084 | 331,685 | 93.3 | 245.5 | 72 | 228 | 0.99 (0.74 - 1.30) | 0.94 | 0.94 (0.71 - 1.24) | 0.66 |
| OEHR | 8,664 | 20,205 | 2.1 | 5.7 | <5 | <5 | 3.08 (0.55 - 18.9) | 0.21 | 2.75 (0.47 - 16.2) | 0.26 |
| VA | 1,352 | 12,496 | 0.7 | 6.5 | 0 | <10 | | | | |
| Meta-analysis | 162,804 | 349,127 | 97.2 | 256.2 | 80 | 244 | 1.02 (0.78 - 1.33) | 0.89 | 0.96 (0.73 - 1.26) | 0.75 |
| Meta-analysis (w/o USOC) | 7,720 | 17,442 | 3.9 | 10.7 | 8 | 16 | 1.39 (0.54 - 3.31) | 0.48 | 1.28 (0.51 - 3.17) | 0.60 |

##### 1.1.10 Abnormal weight gain

**Supplementary Table 10:** Exposures, outcomes and risk estimates for Abnormal weight gain in GLP1RA (male) (T) vs SGLT2I (male) (C) new-user cohorts under four different propensity score (PS) adjustment / time-at-risk (TAR) designs across data sources. We report total patient exposure time in 1,000 patient-year (KYr) units, uncalibrated and calibrated hazard ratios (HRs) and their 95% confidence intervals (CIs). Greyed out entries did not pass study diagnostics and are included only for completeness.

| | Patients | | Exposure (KYr) | | Outcomes | | Uncalibrated | | Calibrated | |
|---|---|---|---|---|---|---|---|---|---|---|
| | T | C | T | C | T | C | HR (95% CI) | $p$ | HR (95% CI) | $p$ |
| **PS-matching / on-treatment TAR** | | | | | | | | | | |
| CCAE | 9,199 | 23,296 | 6.3 | 19.3 | 30 | 68 | 0.98 (0.51 - 1.82) | 0.96 | 0.91 (0.48 - 1.72) | 0.78 |
| ODOD | 6,773 | 16,848 | 3.9 | 11.6 | 30 | 31 | 1.58 (0.75 - 3.34) | 0.23 | 1.64 (0.78 - 3.47) | 0.20 |
| MDCR | 682 | 2,099 | 0.4 | 1.7 | <5 | <5 | | | | |
| Open Claims | 138,102 | 321,351 | 96.9 | 279.0 | 209 | 317 | 1.51 (1.18 - 1.94) | <0.01 | 1.43 (1.12 - 1.84) | <0.01 |
| OEHR | 7,589 | 19,544 | 2.1 | 6.2 | 15 | 30 | 1.09 (0.36 - 3.09) | 0.87 | 1.03 (0.35 - 3.03) | 0.92 |
| VA | 1,144 | 10,914 | 0.8 | 6.6 | <10 | <10 | 1.49 (0.18 - 10.1) | 0.70 | 1.01 (0.13 - 7.64) | 0.95 |
| Meta-analysis | 161,663 | 381,039 | 109.2 | 316.2 | 284 | 446 | 1.43 (1.15 - 1.77) | <0.01 | 1.36 (1.09 - 1.69) | <0.01 |
| Meta-analysis (w/o USOC) | 23,561 | 59,688 | 12.3 | 37.2 | 75 | 129 | 1.18 (0.76 - 1.83) | 0.46 | 1.12 (0.72 - 1.75) | 0.61 |
| **PS-stratification / on-treatment TAR** | | | | | | | | | | |
| CCAE | 9,729 | 23,300 | 6.5 | 19.3 | 34 | 68 | 1.16 (0.74 - 1.78) | 0.51 | 1.08 (0.69 - 1.67) | 0.74 |
| ODOD | 7,282 | 16,867 | 4.2 | 11.7 | 38 | 32 | 2.53 (1.53 - 4.22) | <0.01 | 2.43 (1.46 - 4.05) | <0.01 |
| MDCR | 709 | 2,099 | 0.4 | 1.7 | <5 | <5 | 4.65 (0.75 - 36.1) | 0.12 | 3.84 (0.55 - 26.6) | 0.17 |
| Open Claims | 146,733 | 321,381 | 101.1 | 279.0 | 235 | 317 | 1.76 (1.46 - 2.10) | <0.01 | 1.68 (1.40 - 2.02) | <0.01 |
| OEHR | 8,164 | 19,556 | 2.3 | 6.2 | 20 | 30 | 1.34 (0.72 - 2.43) | 0.35 | 1.20 (0.65 - 2.21) | 0.56 |
| VA | 1,302 | 12,226 | 0.9 | 7.4 | <10 | <10 | 1.38 (0.26 - 6.40) | 0.69 | 1.00 (0.20 - 4.98) | 0.95 |
| Meta-analysis | 171,908 | 381,104 | 114.1 | 316.2 | 327 | 447 | 1.64 (1.23 - 2.20) | <0.01 | 1.56 (1.16 - 2.10) | <0.01 |
| Meta-analysis (w/o USOC) | 25,175 | 59,723 | 12.9 | 37.2 | 92 | 130 | 1.57 (0.96 - 2.57) | 0.07 | 1.47 (0.90 - 2.42) | 0.13 |
| **PS-matching / on-treatment TAR without escalation** | | | | | | | | | | |
| CCAE | 9,199 | 23,296 | 5.5 | 16.9 | 23 | 60 | 0.88 (0.43 - 1.71) | 0.71 | 0.81 (0.40 - 1.61) | 0.54 |
| ODOD | 6,773 | 16,848 | 3.5 | 10.4 | 23 | 24 | 1.78 (0.80 - 4.01) | 0.16 | 1.83 (0.82 - 4.08) | 0.15 |
| MDCR | 682 | 2,099 | 0.4 | 1.5 | <5 | <5 | | | | |
| Open Claims | 138,102 | 321,351 | 84.5 | 237.4 | 182 | 254 | 1.58 (1.21 - 2.07) | <0.01 | 1.47 (1.12 - 1.92) | <0.01 |
| OEHR | 7,589 | 19,544 | 1.8 | 5.5 | 14 | 24 | 1.53 (0.47 - 4.83) | 0.47 | 1.39 (0.44 - 4.46) | 0.58 |
| VA | 1,144 | 10,914 | 0.6 | 5.7 | <10 | <10 | 1.49 (0.18 - 10.1) | 0.70 | 0.96 (0.13 - 7.26) | 0.94 |
| Meta-analysis | 161,663 | 381,039 | 95.3 | 270.1 | 242 | 362 | 1.48 (1.16 - 1.90) | <0.01 | 1.39 (1.08 - 1.79) | 0.01 |
| Meta-analysis (w/o USOC) | 23,561 | 59,688 | 10.8 | 32.7 | 60 | 108 | 1.24 (0.77 - 2.00) | 0.38 | 1.15 (0.71 - 1.86) | 0.57 |
| **PS-stratification / on-treatment TAR without escalation** | | | | | | | | | | |
| CCAE | 9,729 | 23,300 | 5.7 | 16.9 | 27 | 60 | 0.99 (0.60 - 1.60) | 0.98 | 0.90 (0.55 - 1.47) | 0.69 |
| ODOD | 7,282 | 16,867 | 3.7 | 10.4 | 31 | 25 | 2.63 (1.48 - 4.71) | <0.01 | 2.48 (1.39 - 4.42) | <0.01 |
| MDCR | 709 | 2,099 | 0.4 | 1.5 | <5 | <5 | 4.31 (0.69 - 33.7) | 0.14 | 3.46 (0.50 - 24.2) | 0.21 |
| Open Claims | 146,733 | 321,381 | 88.3 | 237.4 | 206 | 254 | 1.84 (1.51 - 2.24) | <0.01 | 1.74 (1.43 - 2.13) | <0.01 |
| OEHR | 8,164 | 19,556 | 2.0 | 5.5 | 17 | 24 | 1.39 (0.70 - 2.69) | 0.34 | 1.24 (0.63 - 2.43) | 0.53 |
| VA | 1,302 | 12,226 | 0.7 | 6.4 | <10 | <10 | 1.04 (0.14 - 5.74) | 0.97 | 0.75 (0.12 - 4.89) | 0.76 |
| Meta-analysis | 171,908 | 381,104 | 99.7 | 270.1 | 281 | 363 | 1.62 (1.11 - 2.38) | 0.01 | 1.52 (1.04 - 2.24) | 0.03 |
| Meta-analysis (w/o USOC) | 25,175 | 59,723 | 11.4 | 32.7 | 75 | 109 | 1.52 (0.84 - 2.75) | 0.17 | 1.40 (0.77 - 2.54) | 0.27 |

##### 1.1.11 Abnormal weight loss

**Supplementary Table 11:** Exposures, outcomes and risk estimates for Abnormal weight loss in GLP1RA (male) (T) vs SGLT2I (male) (C) new-user cohorts under four different propensity score (PS) adjustment / time-at-risk (TAR) designs across data sources. We report total patient exposure time in 1,000 patient-year (KYr) units, uncalibrated and calibrated hazard ratios (HRs) and their 95% confidence intervals (CIs). Greyed out entries did not pass study diagnostics and are included only for completeness.

| | Patients | | Exposure (KYr) | | Outcomes | | Uncalibrated | | Calibrated | |
|---|---|---|---|---|---|---|---|---|---|---|
| | T | C | T | C | T | C | HR (95% CI) | $p$ | HR (95% CI) | $p$ |
| **PS-matching / on-treatment TAR** | | | | | | | | | | |
| CCAE | 9,478 | 23,564 | 6.4 | 19.6 | 58 | 145 | 1.28 (0.86 - 1.87) | 0.21 | 1.19 (0.81 - 1.75) | 0.40 |
| ODOD | 6,937 | 16,871 | 4.0 | 11.6 | 54 | 184 | 1.02 (0.68 - 1.51) | 0.93 | 1.06 (0.71 - 1.58) | 0.79 |
| MDCR | 684 | 2,062 | 0.4 | 1.6 | 5 | 35 | 0.82 (0.26 - 2.19) | 0.71 | 0.63 (0.21 - 1.82) | 0.39 |
| Open Claims | 141,191 | 322,477 | 99.1 | 280.1 | 435 | 1,594 | 0.85 (0.74 - 0.97) | 0.02 | 0.80 (0.70 - 0.93) | <0.01 |
| OEHR | 7,738 | 19,632 | 2.2 | 6.3 | 21 | 79 | 0.71 (0.36 - 1.35) | 0.32 | 0.67 (0.34 - 1.31) | 0.25 |
| VA | 1,144 | 10,816 | 0.8 | 6.5 | <10 | 82 | 1.34 (0.50 - 3.21) | 0.54 | 0.91 (0.36 - 2.31) | 0.83 |
| Meta-analysis | 166,488 | 393,360 | 112.5 | 324.0 | <578 | 2,084 | 0.96 (0.78 - 1.18) | 0.68 | 0.91 (0.74 - 1.12) | 0.39 |
| Meta-analysis (w/o USOC) | 25,297 | 70,883 | 13.3 | 43.9 | <143 | 490 | 1.08 (0.85 - 1.39) | 0.52 | 1.03 (0.80 - 1.33) | 0.81 |
| **PS-stratification / on-treatment TAR** | | | | | | | | | | |
| CCAE | 10,096 | 23,568 | 6.7 | 19.6 | 63 | 145 | 1.21 (0.88 - 1.64) | 0.24 | 1.12 (0.82 - 1.53) | 0.48 |
| ODOD | 7,505 | 16,888 | 4.3 | 11.6 | 58 | 184 | 0.85 (0.62 - 1.16) | 0.32 | 0.82 (0.60 - 1.12) | 0.23 |
| MDCR | 714 | 2,062 | 0.5 | 1.6 | 5 | 35 | 0.58 (0.19 - 1.37) | 0.27 | 0.48 (0.18 - 1.26) | 0.14 |
| Open Claims | 151,068 | 322,505 | 104.0 | 280.1 | 462 | 1,595 | 0.82 (0.73 - 0.91) | <0.01 | 0.78 (0.70 - 0.88) | <0.01 |
| OEHR | 8,433 | 19,644 | 2.3 | 6.3 | 23 | 79 | 0.78 (0.47 - 1.25) | 0.33 | 0.70 (0.43 - 1.15) | 0.17 |
| VA | 1,315 | 12,060 | 0.9 | 7.2 | 11 | 100 | 1.01 (0.47 - 2.00) | 0.98 | 0.73 (0.35 - 1.51) | 0.40 |
| Meta-analysis | 177,102 | 382,605 | 117.3 | 317.5 | 606 | 2,003 | 0.89 (0.74 - 1.07) | 0.23 | 0.85 (0.70 - 1.03) | 0.10 |
| Meta-analysis (w/o USOC) | 26,034 | 60,100 | 13.3 | 37.5 | 144 | 408 | 0.96 (0.74 - 1.25) | 0.76 | 0.90 (0.69 - 1.18) | 0.45 |
| **PS-matching / on-treatment TAR without escalation** | | | | | | | | | | |
| CCAE | 9,478 | 23,564 | 5.6 | 17.1 | 52 | 126 | 1.25 (0.82 - 1.87) | 0.29 | 1.15 (0.76 - 1.73) | 0.52 |
| ODOD | 6,937 | 16,871 | 3.6 | 10.3 | 47 | 160 | 0.91 (0.59 - 1.37) | 0.65 | 0.93 (0.61 - 1.42) | 0.74 |
| MDCR | 684 | 2,062 | 0.4 | 1.5 | 5 | 31 | 1.02 (0.32 - 2.81) | 0.98 | 0.76 (0.26 - 2.27) | 0.63 |
| Open Claims | 141,191 | 322,477 | 86.4 | 238.3 | 368 | 1,385 | 0.81 (0.70 - 0.94) | <0.01 | 0.76 (0.65 - 0.88) | <0.01 |
| OEHR | 7,738 | 19,632 | 1.9 | 5.5 | 18 | 70 | 0.71 (0.34 - 1.38) | 0.33 | 0.64 (0.32 - 1.30) | 0.22 |
| VA | 1,144 | 10,816 | 0.6 | 5.6 | <10 | 73 | 1.28 (0.40 - 3.39) | 0.65 | 0.82 (0.28 - 2.39) | 0.72 |
| Meta-analysis | 166,488 | 393,360 | 98.1 | 276.8 | <495 | 1,814 | 0.90 (0.74 - 1.11) | 0.33 | 0.85 (0.69 - 1.04) | 0.12 |
| Meta-analysis (w/o USOC) | 25,297 | 70,883 | 11.7 | 38.5 | <127 | 429 | 1.02 (0.78 - 1.33) | 0.89 | 0.95 (0.72 - 1.24) | 0.72 |
| **PS-stratification / on-treatment TAR without escalation** | | | | | | | | | | |
| CCAE | 10,096 | 23,568 | 5.9 | 17.1 | 57 | 126 | 1.26 (0.90 - 1.74) | 0.17 | 1.14 (0.82 - 1.59) | 0.44 |
| ODOD | 7,505 | 16,888 | 3.8 | 10.3 | 51 | 160 | 0.89 (0.63 - 1.23) | 0.47 | 0.83 (0.60 - 1.16) | 0.30 |
| MDCR | 714 | 2,062 | 0.4 | 1.5 | 5 | 31 | 0.66 (0.22 - 1.58) | 0.40 | 0.53 (0.20 - 1.41) | 0.21 |
| Open Claims | 151,068 | 322,505 | 90.9 | 238.3 | 394 | 1,386 | 0.79 (0.70 - 0.89) | <0.01 | 0.75 (0.66 - 0.84) | <0.01 |
| OEHR | 8,433 | 19,644 | 2.0 | 5.5 | 19 | 70 | 0.75 (0.43 - 1.25) | 0.30 | 0.67 (0.40 - 1.15) | 0.15 |
| VA | 1,315 | 12,060 | 0.7 | 6.3 | <10 | 87 | 0.84 (0.32 - 1.87) | 0.69 | 0.60 (0.25 - 1.45) | 0.27 |
| Meta-analysis | 177,102 | 382,605 | 102.6 | 271.2 | 521 | 1,742 | 0.90 (0.72 - 1.12) | 0.35 | 0.84 (0.67 - 1.06) | 0.15 |
| Meta-analysis (w/o USOC) | 26,034 | 60,100 | 11.7 | 32.9 | 127 | 356 | 0.98 (0.73 - 1.31) | 0.90 | 0.90 (0.67 - 1.21) | 0.51 |

##### 1.1.12 Acute pancreatitis

**Supplementary Table 12:** Exposures, outcomes and risk estimates for Acute pancreatitis in GLP1RA (male) (T) vs SGLT2I (male) (C) new-user cohorts under four different propensity score (PS) adjustment / time-at-risk (TAR) designs across data sources. We report total patient exposure time in 1,000 patient-year (KYr) units, uncalibrated and calibrated hazard ratios (HRs) and their 95% confidence intervals (CIs). Greyed out entries did not pass study diagnostics and are included only for completeness.

| | Patients | | Exposure (KYr) | | Outcomes | | Uncalibrated | | Calibrated | |
|---|---|---|---|---|---|---|---|---|---|---|
| | T | C | T | C | T | C | HR (95% CI) | $p$ | HR (95% CI) | $p$ |
| **PS-matching / on-treatment TAR** | | | | | | | | | | |
| CCAE | 9,638 | 23,886 | 6.6 | 19.9 | 16 | 42 | 0.64 (0.29 - 1.32) | 0.25 | 0.60 (0.28 - 1.28) | 0.19 |
| ODOD | 7,100 | 17,288 | 4.1 | 12.0 | 8 | 29 | 1.08 (0.37 - 2.86) | 0.88 | 1.13 (0.40 - 3.15) | 0.82 |
| MDCR | 696 | 2,119 | 0.4 | 1.7 | 0 | <5 | | | | |
| Open Claims | 143,625 | 327,916 | 101.0 | 285.4 | 138 | 377 | 0.92 (0.70 - 1.19) | 0.53 | 0.87 (0.67 - 1.13) | 0.31 |
| OEHR | 7,909 | 20,096 | 2.2 | 6.4 | 7 | 13 | 1.67 (0.44 - 6.44) | 0.46 | 1.57 (0.41 - 6.03) | 0.51 |
| VA | 1,172 | 11,059 | 0.8 | 6.7 | 0 | <10 | | | | |
| Meta-analysis | 160,363 | 369,090 | 111.7 | 317.3 | 162 | 448 | 0.89 (0.70 - 1.14) | 0.36 | 0.85 (0.66 - 1.09) | 0.20 |
| Meta-analysis (w/o USOC) | 16,738 | 41,174 | 10.7 | 31.9 | 24 | 71 | 0.77 (0.42 - 1.42) | 0.41 | 0.73 (0.40 - 1.35) | 0.33 |
| **PS-stratification / on-treatment TAR** | | | | | | | | | | |
| CCAE | 10,266 | 23,890 | 6.9 | 19.9 | 17 | 42 | 0.97 (0.52 - 1.73) | 0.92 | 0.90 (0.50 - 1.65) | 0.74 |
| ODOD | 7,691 | 17,307 | 4.4 | 12.0 | 8 | 29 | 0.76 (0.31 - 1.67) | 0.52 | 0.73 (0.32 - 1.70) | 0.47 |
| MDCR | 727 | 2,119 | 0.5 | 1.7 | 0 | <5 | | | | |
| Open Claims | 153,856 | 327,945 | 106.0 | 285.4 | 149 | 377 | 1.01 (0.83 - 1.24) | 0.89 | 0.97 (0.79 - 1.19) | 0.77 |
| OEHR | 8,626 | 20,108 | 2.4 | 6.4 | 7 | 13 | 1.31 (0.47 - 3.40) | 0.60 | 1.17 (0.43 - 3.17) | 0.75 |
| VA | 1,342 | 12,372 | 0.9 | 7.4 | 0 | <10 | | | | |
| Meta-analysis | 180,439 | 389,250 | 119.7 | 323.7 | 181 | 461 | 1.00 (0.84 - 1.21) | 0.96 | 0.95 (0.79 - 1.15) | 0.63 |
| Meta-analysis (w/o USOC) | 26,583 | 61,305 | 13.7 | 38.3 | 32 | 84 | 0.96 (0.62 - 1.49) | 0.86 | 0.90 (0.58 - 1.40) | 0.66 |
| **PS-matching / on-treatment TAR without escalation** | | | | | | | | | | |
| CCAE | 9,638 | 23,886 | 5.7 | 17.4 | 11 | 34 | 0.56 (0.22 - 1.30) | 0.20 | 0.52 (0.21 - 1.27) | 0.15 |
| ODOD | 7,100 | 17,288 | 3.7 | 10.6 | 7 | 23 | 0.83 (0.22 - 2.51) | 0.76 | 0.85 (0.25 - 2.85) | 0.79 |
| MDCR | 696 | 2,119 | 0.4 | 1.5 | 0 | <5 | | | | |
| Open Claims | 143,625 | 327,916 | 88.0 | 242.8 | 115 | 306 | 0.89 (0.66 - 1.18) | 0.42 | 0.83 (0.62 - 1.10) | 0.20 |
| OEHR | 7,909 | 20,096 | 1.9 | 5.7 | 6 | 11 | 2.28 (0.56 - 9.93) | 0.26 | 2.08 (0.49 - 8.74) | 0.32 |
| VA | 1,172 | 11,059 | 0.6 | 5.7 | 0 | <10 | | | | |
| Meta-analysis | 160,363 | 369,090 | 97.5 | 270.8 | 133 | 363 | 0.85 (0.65 - 1.11) | 0.23 | 0.80 (0.61 - 1.04) | 0.10 |
| Meta-analysis (w/o USOC) | 16,738 | 41,174 | 9.4 | 28.0 | 18 | 57 | 0.64 (0.31 - 1.32) | 0.23 | 0.60 (0.29 - 1.23) | 0.17 |
| **PS-stratification / on-treatment TAR without escalation** | | | | | | | | | | |
| CCAE | 10,266 | 23,890 | 6.0 | 17.4 | 12 | 34 | 0.78 (0.38 - 1.54) | 0.50 | 0.71 (0.35 - 1.45) | 0.35 |
| ODOD | 7,691 | 17,307 | 3.9 | 10.7 | 7 | 23 | 0.91 (0.35 - 2.13) | 0.84 | 0.86 (0.35 - 2.13) | 0.74 |
| MDCR | 727 | 2,119 | 0.4 | 1.5 | 0 | <5 | | | | |
| Open Claims | 153,856 | 327,945 | 92.7 | 242.8 | 123 | 306 | 0.99 (0.79 - 1.24) | 0.96 | 0.94 (0.75 - 1.18) | 0.61 |
| OEHR | 8,626 | 20,108 | 2.1 | 5.7 | 6 | 11 | 1.39 (0.46 - 3.87) | 0.55 | 1.24 (0.43 - 3.62) | 0.69 |
| VA | 1,342 | 12,372 | 0.7 | 6.4 | 0 | <10 | | | | |
| Meta-analysis | 171,813 | 369,142 | 102.6 | 270.9 | 142 | 363 | 0.97 (0.79 - 1.19) | 0.77 | 0.91 (0.74 - 1.12) | 0.39 |
| Meta-analysis (w/o USOC) | 17,957 | 41,197 | 9.9 | 28.0 | 19 | 57 | 0.83 (0.48 - 1.45) | 0.51 | 0.76 (0.44 - 1.34) | 0.35 |

##### 1.1.13 Bone fracture

**Supplementary Table 13:** Exposures, outcomes and risk estimates for Bone fracture in GLP1RA (male) (T) vs SGLT2I (male) (C) new-user cohorts under four different propensity score (PS) adjustment / time-at-risk (TAR) designs across data sources. We report total patient exposure time in 1,000 patient-year (KYr) units, uncalibrated and calibrated hazard ratios (HRs) and their 95% confidence intervals (CIs). Greyed out entries did not pass study diagnostics and are included only for completeness.

| | Patients | | Exposure (KYr) | | Outcomes | | Uncalibrated | | Calibrated | |
|---|---|---|---|---|---|---|---|---|---|---|
| | T | C | T | C | T | C | HR (95% CI) | $p$ | HR (95% CI) | $p$ |
| **PS-matching / on-treatment TAR** | | | | | | | | | | |
| CCAE | 8,868 | 22,294 | 5.9 | 18.2 | 93 | 296 | 0.83 (0.60 - 1.13) | 0.25 | 0.77 (0.56 - 1.06) | 0.12 |
| ODOD | 6,617 | 16,177 | 3.8 | 11.0 | 94 | 211 | 1.15 (0.81 - 1.63) | 0.43 | 1.20 (0.84 - 1.71) | 0.33 |
| MDCR | 634 | 1,967 | 0.4 | 1.6 | 8 | 38 | 1.61 (0.54 - 4.67) | 0.39 | 1.23 (0.42 - 3.61) | 0.71 |
| Open Claims | 126,901 | 295,510 | 88.8 | 256.4 | 943 | 2,775 | 1.02 (0.92 - 1.13) | 0.70 | 0.97 (0.87 - 1.08) | 0.57 |
| OEHR | 7,399 | 18,967 | 2.1 | 6.1 | 21 | 70 | 0.47 (0.21 - 0.95) | 0.05 | 0.44 (0.21 - 0.95) | 0.04 |
| VA | 1,059 | 10,010 | 0.7 | 6.0 | <10 | 50 | 1.62 (0.59 - 4.07) | 0.33 | 1.10 (0.42 - 2.91) | 0.84 |
| Meta-analysis | 150,844 | 362,958 | 101.2 | 297.7 | <1,161 | 3,402 | 1.00 (0.91 - 1.10) | 0.94 | 0.95 (0.86 - 1.06) | 0.39 |
| Meta-analysis (w/o USOC) | 23,943 | 67,448 | 12.5 | 41.4 | <218 | 627 | 0.92 (0.64 - 1.32) | 0.64 | 0.87 (0.60 - 1.26) | 0.48 |
| **PS-stratification / on-treatment TAR** | | | | | | | | | | |
| CCAE | 9,460 | 22,298 | 6.2 | 18.2 | 99 | 296 | 0.94 (0.74 - 1.19) | 0.63 | 0.88 (0.69 - 1.12) | 0.30 |
| ODOD | 7,151 | 16,194 | 4.0 | 11.1 | 103 | 211 | 1.21 (0.94 - 1.56) | 0.13 | 1.17 (0.91 - 1.51) | 0.25 |
| MDCR | 661 | 1,967 | 0.4 | 1.6 | 8 | 38 | 0.75 (0.32 - 1.58) | 0.49 | 0.62 (0.28 - 1.39) | 0.25 |
| Open Claims | 135,417 | 295,535 | 92.9 | 256.4 | 1,015 | 2,775 | 0.99 (0.92 - 1.07) | 0.77 | 0.95 (0.87 - 1.02) | 0.25 |
| OEHR | 8,094 | 18,979 | 2.2 | 6.1 | 25 | 70 | 0.69 (0.41 - 1.11) | 0.14 | 0.62 (0.38 - 1.01) | 0.06 |
| VA | 1,215 | 11,119 | 0.8 | 6.7 | <10 | 52 | 1.71 (0.70 - 3.81) | 0.21 | 1.24 (0.53 - 2.88) | 0.62 |
| Meta-analysis | 160,122 | 353,006 | 105.4 | 291.7 | 1,242 | 3,352 | 0.99 (0.93 - 1.06) | 0.83 | 0.94 (0.86 - 1.03) | 0.20 |
| Meta-analysis (w/o USOC) | 24,705 | 57,471 | 12.5 | 35.4 | 227 | 577 | 0.98 (0.75 - 1.28) | 0.87 | 0.92 (0.70 - 1.21) | 0.55 |
| **PS-matching / on-treatment TAR without escalation** | | | | | | | | | | |
| CCAE | 8,868 | 22,294 | 5.2 | 15.9 | 77 | 254 | 0.86 (0.61 - 1.20) | 0.38 | 0.79 (0.56 - 1.11) | 0.19 |
| ODOD | 6,617 | 16,177 | 3.4 | 9.8 | 82 | 186 | 1.06 (0.71 - 1.54) | 0.79 | 1.08 (0.73 - 1.60) | 0.70 |
| MDCR | 634 | 1,967 | 0.3 | 1.4 | 8 | 32 | 1.95 (0.64 - 5.93) | 0.24 | 1.46 (0.48 - 4.44) | 0.51 |
| Open Claims | 126,901 | 295,510 | 77.4 | 218.3 | 805 | 2,332 | 1.01 (0.90 - 1.13) | 0.92 | 0.94 (0.83 - 1.05) | 0.30 |
| OEHR | 7,399 | 18,967 | 1.8 | 5.3 | 17 | 62 | 0.51 (0.22 - 1.10) | 0.11 | 0.47 (0.21 - 1.06) | 0.07 |
| VA | 1,059 | 10,010 | 0.6 | 5.2 | <10 | 45 | 1.52 (0.46 - 4.27) | 0.46 | 0.98 (0.32 - 2.99) | 0.90 |
| Meta-analysis | 150,844 | 362,958 | 88.3 | 254.6 | <991 | 2,879 | 0.99 (0.89 - 1.09) | 0.82 | 0.93 (0.83 - 1.03) | 0.17 |
| Meta-analysis (w/o USOC) | 23,943 | 67,448 | 10.9 | 36.3 | <186 | 547 | 0.91 (0.70 - 1.19) | 0.49 | 0.85 (0.65 - 1.11) | 0.26 |
| **PS-stratification / on-treatment TAR without escalation** | | | | | | | | | | |
| CCAE | 9,460 | 22,298 | 5.4 | 15.9 | 82 | 254 | 0.90 (0.69 - 1.17) | 0.45 | 0.82 (0.63 - 1.07) | 0.16 |
| ODOD | 7,151 | 16,194 | 3.6 | 9.8 | 91 | 186 | 1.21 (0.92 - 1.58) | 0.17 | 1.14 (0.87 - 1.49) | 0.38 |
| MDCR | 661 | 1,967 | 0.4 | 1.4 | 8 | 32 | 1.00 (0.42 - 2.13) | 1.00 | 0.80 (0.35 - 1.81) | 0.60 |
| Open Claims | 135,417 | 295,535 | 81.2 | 218.3 | 868 | 2,332 | 0.98 (0.90 - 1.07) | 0.66 | 0.93 (0.85 - 1.01) | 0.15 |
| OEHR | 8,094 | 18,979 | 2.0 | 5.3 | 21 | 62 | 0.64 (0.36 - 1.07) | 0.10 | 0.57 (0.33 - 0.98) | 0.04 |
| VA | 1,215 | 11,119 | 0.7 | 5.8 | <10 | 46 | 1.52 (0.53 - 3.76) | 0.40 | 1.10 (0.41 - 2.94) | 0.85 |
| Meta-analysis | 160,122 | 353,006 | 92.2 | 249.4 | 1,062 | 2,834 | 0.98 (0.91 - 1.06) | 0.64 | 0.92 (0.84 - 1.01) | 0.08 |
| Meta-analysis (w/o USOC) | 24,705 | 57,471 | 11.0 | 31.1 | 194 | 502 | 0.94 (0.69 - 1.28) | 0.70 | 0.87 (0.64 - 1.18) | 0.38 |

###### 1.1.14 Diabetic ketoacidosis

**Supplementary Table 14:** Exposures, outcomes and risk estimates for Diabetic ketoacidosis in GLP1RA (male) (T) vs SGLT2I (male) (C) new-user cohorts under four different propensity score (PS) adjustment / time-at-risk (TAR) designs across data sources. We report total patient exposure time in 1,000 patient-year (KYr) units, uncalibrated and calibrated hazard ratios (HRs) and their 95% confidence intervals (CIs). Greyed out entries did not pass study diagnostics and are included only for completeness.

| | Patients | | Exposure (KYr) | | Outcomes | | Uncalibrated | | Calibrated | |
|---|---|---|---|---|---|---|---|---|---|---|
| | T | C | T | C | T | C | HR (95% CI) | $p$ | HR (95% CI) | $p$ |
| **PS-matching / on-treatment TAR** | | | | | | | | | | |
| CCAE | 9,693 | 24,104 | 6.6 | 20.1 | 8 | 40 | 0.61 (0.22 - 1.46) | 0.31 | 0.57 (0.22 - 1.47) | 0.24 |
| ODOD | 7,133 | 17,440 | 4.1 | 12.1 | <5 | 23 | 0.30 (0.05 - 1.11) | 0.14 | 0.31 (0.06 - 1.53) | 0.15 |
| MDCR | 700 | 2,135 | 0.4 | 1.7 | 0 | <5 | | | | |
| Open Claims | 144,918 | 332,168 | 101.8 | 289.1 | 68 | 307 | 0.57 (0.41 - 0.78) | <0.01 | 0.54 (0.39 - 0.75) | <0.01 |
| OEHR | 7,945 | 20,206 | 2.2 | 6.5 | <5 | 14 | 0.86 (0.18 - 3.39) | 0.85 | 0.82 (0.19 - 3.56) | 0.79 |
| VA | 1,185 | 11,176 | 0.8 | 6.7 | <10 | <10 | 0.75 (0.04 - 5.46) | 0.82 | 0.51 (0.04 - 6.24) | 0.60 |
| Meta-analysis | 161,744 | 373,712 | 112.6 | 321.3 | <81 | 370 | 0.56 (0.42 - 0.76) | <0.01 | 0.53 (0.39 - 0.72) | <0.01 |
| Meta-analysis (w/o USOC) | 16,826 | 41,544 | 10.7 | 32.2 | <13 | 63 | 0.51 (0.22 - 1.14) | 0.10 | 0.48 (0.21 - 1.09) | 0.08 |
| **PS-stratification / on-treatment TAR** | | | | | | | | | | |
| CCAE | 10,325 | 24,108 | 6.9 | 20.1 | 8 | 40 | 0.50 (0.21 - 1.05) | 0.09 | 0.46 (0.21 - 1.04) | 0.06 |
| ODOD | 7,721 | 17,459 | 4.4 | 12.1 | <5 | 23 | 0.31 (0.07 - 0.93) | 0.07 | 0.30 (0.08 - 1.07) | 0.06 |
| MDCR | 730 | 2,135 | 0.5 | 1.7 | 0 | <5 | | | | |
| Open Claims | 155,077 | 332,197 | 106.8 | 289.1 | 70 | 307 | 0.53 (0.41 - 0.70) | <0.01 | 0.51 (0.39 - 0.67) | <0.01 |
| OEHR | 8,665 | 20,218 | 2.4 | 6.5 | <5 | 14 | 0.42 (0.09 - 1.36) | 0.21 | 0.38 (0.10 - 1.44) | 0.16 |
| VA | 1,357 | 12,523 | 0.9 | 7.5 | <10 | <10 | 0.77 (0.04 - 6.61) | 0.85 | 0.56 (0.04 - 7.62) | 0.66 |
| Meta-analysis | 173,123 | 373,764 | 118.1 | 321.3 | <83 | 370 | 0.52 (0.40 - 0.67) | <0.01 | 0.49 (0.38 - 0.64) | <0.01 |
| Meta-analysis (w/o USOC) | 18,046 | 41,567 | 11.3 | 32.2 | <13 | 63 | 0.44 (0.22 - 0.86) | 0.02 | 0.41 (0.21 - 0.81) | 0.01 |
| **PS-matching / on-treatment TAR without escalation** | | | | | | | | | | |
| CCAE | 9,693 | 24,104 | 5.8 | 17.5 | 7 | 33 | 0.52 (0.17 - 1.32) | 0.21 | 0.48 (0.17 - 1.34) | 0.16 |
| ODOD | 7,133 | 17,440 | 3.7 | 10.7 | <5 | 20 | 0.14 (0.01 - 0.72) | 0.09 | 0.14 (0.01 - 1.40) | 0.10 |
| MDCR | 700 | 2,135 | 0.4 | 1.5 | 0 | 0 | | | | |
| Open Claims | 144,918 | 332,168 | 88.8 | 245.8 | 57 | 243 | 0.60 (0.42 - 0.83) | <0.01 | 0.55 (0.39 - 0.78) | <0.01 |
| OEHR | 7,945 | 20,206 | 1.9 | 5.7 | <5 | 13 | 0.86 (0.18 - 3.39) | 0.85 | 0.79 (0.18 - 3.44) | 0.75 |
| VA | 1,185 | 11,176 | 0.6 | 5.8 | <10 | <10 | 0.78 (0.04 - 5.90) | 0.85 | 0.50 (0.04 - 6.33) | 0.59 |
| Meta-analysis | 161,744 | 373,712 | 98.2 | 274.1 | <69 | 296 | 0.57 (0.41 - 0.79) | <0.01 | 0.53 (0.39 - 0.74) | <0.01 |
| Meta-analysis (w/o USOC) | 16,826 | 41,544 | 9.5 | 28.3 | <12 | 53 | 0.41 (0.15 - 1.11) | 0.08 | 0.38 (0.14 - 1.04) | 0.06 |
| **PS-stratification / on-treatment TAR without escalation** | | | | | | | | | | |
| CCAE | 10,325 | 24,108 | 6.0 | 17.5 | 7 | 33 | 0.49 (0.19 - 1.09) | 0.11 | 0.45 (0.19 - 1.06) | 0.07 |
| ODOD | 7,721 | 17,459 | 3.9 | 10.8 | <5 | 20 | 0.21 (0.03 - 0.76) | 0.05 | 0.20 (0.04 - 0.95) | 0.04 |
| MDCR | 730 | 2,135 | 0.4 | 1.5 | 0 | 0 | | | | |
| Open Claims | 155,077 | 332,197 | 93.4 | 245.8 | 58 | 243 | 0.54 (0.40 - 0.72) | <0.01 | 0.51 (0.38 - 0.69) | <0.01 |
| OEHR | 8,665 | 20,218 | 2.1 | 5.7 | <5 | 13 | 0.43 (0.10 - 1.40) | 0.22 | 0.38 (0.10 - 1.47) | 0.16 |
| VA | 1,357 | 12,523 | 0.7 | 6.5 | <10 | <10 | 0.81 (0.04 - 7.53) | 0.88 | 0.59 (0.04 - 8.43) | 0.69 |
| Meta-analysis | 173,123 | 373,764 | 103.3 | 274.1 | <70 | 296 | 0.52 (0.39 - 0.68) | <0.01 | 0.49 (0.37 - 0.64) | <0.01 |
| Meta-analysis (w/o USOC) | 18,046 | 41,567 | 10.0 | 28.3 | <12 | 53 | 0.40 (0.19 - 0.86) | 0.02 | 0.37 (0.17 - 0.79) | 0.01 |

##### 1.1.15 Diarrhea

**Supplementary Table 15:** Exposures, outcomes and risk estimates for Diarrhea in GLP1RA (male) (T) vs SGLT2I (male) (C) new-user cohorts under four different propensity score (PS) adjustment / time-at-risk (TAR) designs across data sources. We report total patient exposure time in 1,000 patient-year (KYr) units, uncalibrated and calibrated hazard ratios (HRs) and their 95% confidence intervals (CIs). Greyed out entries did not pass study diagnostics and are included only for completeness.

| | Patients | | Exposure (KYr) | | Outcomes | | Uncalibrated | | Calibrated | |
|---|---|---|---|---|---|---|---|---|---|---|
| | T | C | T | C | T | C | HR (95% CI) | $p$ | HR (95% CI) | $p$ |
| **PS-matching / on-treatment TAR** | | | | | | | | | | |
| CCAE | 8,625 | 21,752 | 5.7 | 17.8 | 223 | 367 | 1.54 (1.22 - 1.94) | <0.01 | 1.43 (1.13 - 1.81) | <0.01 |
| ODOD | 6,363 | 15,584 | 3.6 | 10.6 | 163 | 284 | 1.70 (1.30 - 2.24) | <0.01 | 1.77 (1.35 - 2.33) | <0.01 |
| MDCR | 614 | 1,906 | 0.4 | 1.5 | 10 | 30 | 0.81 (0.28 - 2.09) | 0.68 | 0.62 (0.23 - 1.70) | 0.36 |
| Open Claims | 128,629 | 299,039 | 89.5 | 258.8 | 1,598 | 2,998 | 1.57 (1.44 - 1.71) | <0.01 | 1.48 (1.35 - 1.63) | <0.01 |
| OEHR | 7,231 | 18,494 | 2.0 | 5.8 | 81 | 134 | 1.72 (1.17 - 2.51) | <0.01 | 1.62 (1.11 - 2.38) | 0.02 |
| VA | 1,003 | 9,517 | 0.7 | 5.7 | 16 | 97 | 1.33 (0.65 - 2.57) | 0.42 | 0.90 (0.45 - 1.79) | 0.75 |
| Meta-analysis | 151,851 | 364,386 | 101.5 | 298.8 | 2,081 | 3,880 | 1.58 (1.46 - 1.70) | <0.01 | 1.50 (1.38 - 1.64) | <0.01 |
| Meta-analysis (w/o USOC) | 23,222 | 65,347 | 12.0 | 40.0 | 483 | 882 | 1.61 (1.38 - 1.88) | <0.01 | 1.53 (1.30 - 1.80) | <0.01 |
| **PS-stratification / on-treatment TAR** | | | | | | | | | | |
| CCAE | 9,160 | 21,758 | 6.0 | 17.9 | 243 | 367 | 1.68 (1.42 - 2.00) | <0.01 | 1.57 (1.31 - 1.87) | <0.01 |
| ODOD | 6,875 | 15,599 | 3.8 | 10.6 | 176 | 286 | 1.52 (1.24 - 1.86) | <0.01 | 1.47 (1.20 - 1.80) | <0.01 |
| MDCR | 649 | 1,906 | 0.4 | 1.5 | 10 | 30 | 1.35 (0.60 - 2.78) | 0.44 | 1.11 (0.52 - 2.39) | 0.78 |
| Open Claims | 137,191 | 299,156 | 93.6 | 258.9 | 1,776 | 3,000 | 1.49 (1.40 - 1.59) | <0.01 | 1.43 (1.33 - 1.53) | <0.01 |
| OEHR | 7,828 | 18,506 | 2.1 | 5.8 | 89 | 134 | 1.68 (1.26 - 2.23) | <0.01 | 1.51 (1.13 - 2.01) | <0.01 |
| VA | 1,146 | 10,635 | 0.8 | 6.4 | 21 | 107 | 1.63 (0.91 - 2.81) | 0.09 | 1.17 (0.67 - 2.07) | 0.59 |
| Meta-analysis | 161,054 | 355,019 | 105.6 | 293.2 | 2,284 | 3,787 | 1.53 (1.43 - 1.63) | <0.01 | 1.45 (1.33 - 1.58) | <0.01 |
| Meta-analysis (w/o USOC) | 23,863 | 55,863 | 12.0 | 34.3 | 508 | 787 | 1.63 (1.44 - 1.83) | <0.01 | 1.52 (1.34 - 1.73) | <0.01 |
| **PS-matching / on-treatment TAR without escalation** | | | | | | | | | | |
| CCAE | 8,625 | 21,752 | 5.0 | 15.6 | 200 | 307 | 1.65 (1.28 - 2.11) | <0.01 | 1.52 (1.18 - 1.95) | <0.01 |
| ODOD | 6,363 | 15,584 | 3.2 | 9.5 | 143 | 242 | 1.77 (1.33 - 2.37) | <0.01 | 1.82 (1.36 - 2.43) | <0.01 |
| MDCR | 614 | 1,906 | 0.3 | 1.3 | 9 | 27 | 0.91 (0.31 - 2.40) | 0.86 | 0.68 (0.25 - 1.89) | 0.47 |
| Open Claims | 128,629 | 299,039 | 78.2 | 220.9 | 1,385 | 2,471 | 1.61 (1.46 - 1.76) | <0.01 | 1.49 (1.35 - 1.65) | <0.01 |
| OEHR | 7,231 | 18,494 | 1.7 | 5.2 | 68 | 117 | 1.84 (1.21 - 2.78) | <0.01 | 1.68 (1.11 - 2.54) | 0.02 |
| VA | 1,003 | 9,517 | 0.5 | 4.9 | 13 | 87 | 1.25 (0.56 - 2.61) | 0.57 | 0.81 (0.37 - 1.74) | 0.59 |
| Meta-analysis | 151,851 | 364,386 | 88.7 | 256.1 | 1,809 | 3,224 | 1.63 (1.50 - 1.77) | <0.01 | 1.53 (1.39 - 1.67) | <0.01 |
| Meta-analysis (w/o USOC) | 23,222 | 65,347 | 10.5 | 35.2 | 424 | 753 | 1.70 (1.43 - 2.01) | <0.01 | 1.58 (1.32 - 1.89) | <0.01 |
| **PS-stratification / on-treatment TAR without escalation** | | | | | | | | | | |
| CCAE | 9,160 | 21,758 | 5.3 | 15.7 | 217 | 307 | 1.77 (1.47 - 2.13) | <0.01 | 1.61 (1.33 - 1.94) | <0.01 |
| ODOD | 6,875 | 15,599 | 3.4 | 9.5 | 155 | 244 | 1.64 (1.32 - 2.03) | <0.01 | 1.54 (1.24 - 1.92) | <0.01 |
| MDCR | 649 | 1,906 | 0.4 | 1.3 | 9 | 27 | 1.25 (0.53 - 2.68) | 0.59 | 1.00 (0.45 - 2.25) | 0.89 |
| Open Claims | 137,191 | 299,156 | 82.0 | 221.0 | 1,549 | 2,473 | 1.54 (1.44 - 1.64) | <0.01 | 1.45 (1.35 - 1.56) | <0.01 |
| OEHR | 7,828 | 18,506 | 1.9 | 5.2 | 76 | 117 | 1.62 (1.18 - 2.20) | <0.01 | 1.45 (1.06 - 1.98) | 0.03 |
| VA | 1,146 | 10,635 | 0.6 | 5.5 | 18 | 95 | 1.59 (0.84 - 2.89) | 0.14 | 1.15 (0.62 - 2.13) | 0.67 |
| Meta-analysis | 161,054 | 355,019 | 92.6 | 251.3 | 1,997 | 3,141 | 1.58 (1.47 - 1.71) | <0.01 | 1.49 (1.36 - 1.63) | <0.01 |
| Meta-analysis (w/o USOC) | 23,863 | 55,863 | 10.6 | 30.3 | 448 | 668 | 1.70 (1.49 - 1.93) | <0.01 | 1.56 (1.37 - 1.79) | <0.01 |

##### 1.1.16 Genitourinary infection

**Supplementary Table 16:** Exposures, outcomes and risk estimates for Genitourinary infection in GLP1RA (male) (T) vs SGLT2I (male) (C) new-user cohorts under four different propensity score (PS) adjustment / time-at-risk (TAR) designs across data sources. We report total patient exposure time in 1,000 patient-year (KYr) units, uncalibrated and calibrated hazard ratios (HRs) and their 95% confidence intervals (CIs). Greyed out entries did not pass study diagnostics and are included only for completeness.

| | Patients | | Exposure (KYr) | | Outcomes | | Uncalibrated | | Calibrated | |
|---|---|---|---|---|---|---|---|---|---|---|
| | T | C | T | C | T | C | HR (95% CI) | $p$ | HR (95% CI) | $p$ |
| **PS-matching / on-treatment TAR** | | | | | | | | | | |
| CCAE | 8,832 | 22,098 | 5.9 | 18.2 | 111 | 274 | 1.14 (0.84 - 1.55) | 0.39 | 1.06 (0.78 - 1.45) | 0.70 |
| ODOD | 6,369 | 15,688 | 3.6 | 10.8 | 105 | 287 | 1.17 (0.86 - 1.58) | 0.32 | 1.21 (0.89 - 1.65) | 0.24 |
| MDCR | 584 | 1,870 | 0.3 | 1.5 | 18 | 43 | 1.61 (0.79 - 3.18) | 0.18 | 1.23 (0.61 - 2.47) | 0.57 |
| Open Claims | 129,162 | 296,972 | 90.6 | 259.6 | 1,185 | 2,991 | 1.13 (1.03 - 1.24) | <0.01 | 1.07 (0.97 - 1.18) | 0.23 |
| OEHR | 7,520 | 19,160 | 2.1 | 6.1 | 41 | 109 | 1.17 (0.68 - 1.96) | 0.56 | 1.10 (0.65 - 1.87) | 0.73 |
| VA | 1,061 | 10,108 | 0.7 | 6.1 | 14 | 59 | 1.85 (0.76 - 4.13) | 0.16 | 1.25 (0.54 - 2.92) | 0.61 |
| Meta-analysis | 152,944 | 364,026 | 102.9 | 300.8 | 1,456 | 3,720 | 1.14 (1.05 - 1.24) | <0.01 | 1.08 (0.99 - 1.19) | 0.10 |
| Meta-analysis (w/o USOC) | 23,782 | 67,054 | 12.4 | 41.2 | 271 | 729 | 1.19 (0.98 - 1.44) | 0.09 | 1.13 (0.93 - 1.38) | 0.27 |
| **PS-stratification / on-treatment TAR** | | | | | | | | | | |
| CCAE | 9,362 | 22,104 | 6.1 | 18.3 | 115 | 274 | 1.20 (0.95 - 1.51) | 0.12 | 1.12 (0.89 - 1.41) | 0.36 |
| ODOD | 6,866 | 15,706 | 3.9 | 10.8 | 114 | 287 | 1.14 (0.90 - 1.43) | 0.28 | 1.09 (0.86 - 1.38) | 0.48 |
| MDCR | 621 | 1,870 | 0.4 | 1.5 | 18 | 43 | 1.66 (0.91 - 2.89) | 0.09 | 1.37 (0.77 - 2.44) | 0.29 |
| Open Claims | 137,954 | 297,034 | 94.9 | 259.6 | 1,276 | 2,993 | 1.19 (1.11 - 1.28) | <0.01 | 1.14 (1.06 - 1.23) | <0.01 |
| OEHR | 8,164 | 19,171 | 2.3 | 6.1 | 45 | 109 | 0.99 (0.68 - 1.42) | 0.94 | 0.89 (0.61 - 1.28) | 0.53 |
| VA | 1,210 | 11,310 | 0.8 | 6.8 | 14 | 68 | 2.06 (0.98 - 4.11) | 0.05 | 1.49 (0.73 - 3.06) | 0.28 |
| Meta-analysis | 162,346 | 354,015 | 107.1 | 294.8 | 1,550 | 3,663 | 1.18 (1.11 - 1.26) | <0.01 | 1.12 (1.03 - 1.22) | <0.01 |
| Meta-analysis (w/o USOC) | 24,392 | 56,981 | 12.2 | 35.1 | 274 | 670 | 1.14 (0.98 - 1.32) | 0.09 | 1.07 (0.91 - 1.25) | 0.45 |
| **PS-matching / on-treatment TAR without escalation** | | | | | | | | | | |
| CCAE | 8,832 | 22,098 | 5.2 | 16.0 | 95 | 233 | 1.06 (0.76 - 1.47) | 0.73 | 0.98 (0.70 - 1.36) | 0.85 |
| ODOD | 6,369 | 15,688 | 3.2 | 9.6 | 94 | 248 | 1.14 (0.82 - 1.57) | 0.44 | 1.17 (0.84 - 1.62) | 0.38 |
| MDCR | 584 | 1,870 | 0.3 | 1.3 | 16 | 40 | 1.67 (0.78 - 3.48) | 0.18 | 1.25 (0.59 - 2.65) | 0.57 |
| Open Claims | 129,162 | 296,972 | 79.0 | 220.9 | 1,033 | 2,548 | 1.13 (1.02 - 1.24) | 0.02 | 1.05 (0.94 - 1.16) | 0.43 |
| OEHR | 7,520 | 19,160 | 1.8 | 5.4 | 37 | 95 | 1.43 (0.80 - 2.54) | 0.22 | 1.30 (0.73 - 2.33) | 0.38 |
| VA | 1,061 | 10,108 | 0.6 | 5.3 | 11 | 53 | 1.37 (0.51 - 3.29) | 0.51 | 0.89 (0.35 - 2.25) | 0.80 |
| Meta-analysis | 152,944 | 364,026 | 89.9 | 257.1 | 1,270 | 3,177 | 1.13 (1.03 - 1.24) | <0.01 | 1.06 (0.96 - 1.17) | 0.26 |
| Meta-analysis (w/o USOC) | 23,782 | 67,054 | 10.8 | 36.2 | 237 | 629 | 1.15 (0.93 - 1.42) | 0.19 | 1.07 (0.86 - 1.33) | 0.57 |
| **PS-stratification / on-treatment TAR without escalation** | | | | | | | | | | |
| CCAE | 9,362 | 22,104 | 5.4 | 16.0 | 98 | 233 | 1.21 (0.94 - 1.54) | 0.14 | 1.10 (0.85 - 1.41) | 0.48 |
| ODOD | 6,866 | 15,706 | 3.5 | 9.6 | 103 | 248 | 1.17 (0.91 - 1.50) | 0.21 | 1.10 (0.86 - 1.42) | 0.46 |
| MDCR | 621 | 1,870 | 0.3 | 1.3 | 16 | 40 | 1.70 (0.90 - 3.09) | 0.09 | 1.37 (0.74 - 2.54) | 0.34 |
| Open Claims | 137,954 | 297,034 | 83.0 | 220.9 | 1,114 | 2,550 | 1.19 (1.10 - 1.28) | <0.01 | 1.13 (1.04 - 1.22) | <0.01 |
| OEHR | 8,164 | 19,171 | 2.0 | 5.4 | 41 | 95 | 1.06 (0.71 - 1.56) | 0.76 | 0.95 (0.64 - 1.41) | 0.80 |
| VA | 1,210 | 11,310 | 0.7 | 5.9 | 11 | 59 | 1.99 (0.89 - 4.16) | 0.08 | 1.44 (0.67 - 3.12) | 0.37 |
| Meta-analysis | 162,346 | 354,015 | 93.8 | 251.9 | 1,356 | 3,126 | 1.19 (1.11 - 1.27) | <0.01 | 1.11 (1.03 - 1.21) | 0.01 |
| Meta-analysis (w/o USOC) | 24,392 | 56,981 | 10.8 | 30.9 | 242 | 576 | 1.17 (0.99 - 1.37) | 0.06 | 1.07 (0.91 - 1.27) | 0.43 |

###### 1.1.17 Hyperkalemia

**Supplementary Table 17:** Exposures, outcomes and risk estimates for Hyperkalemia in GLP1RA (male) (T) vs SGLT2I (male) (C) new-user cohorts under four different propensity score (PS) adjustment / time-at-risk (TAR) designs across data sources. We report total patient exposure time in 1,000 patient-year (KYr) units, uncalibrated and calibrated hazard ratios (HRs) and their 95% confidence intervals (CIs). Greyed out entries did not pass study diagnostics and are included only for completeness.

| | Patients | | Exposure (KYr) | | Outcomes | | Uncalibrated | | Calibrated | |
|---|---|---|---|---|---|---|---|---|---|---|
| | T | C | T | C | T | C | HR (95% CI) | $p$ | HR (95% CI) | $p$ |
| **PS-matching / on-treatment TAR** | | | | | | | | | | |
| CCAE | 9,537 | 23,687 | 6.4 | 19.6 | 47 | 134 | 1.18 (0.73 - 1.86) | 0.49 | 1.10 (0.69 - 1.75) | 0.71 |
| ODOD | 6,873 | 16,781 | 3.9 | 11.6 | 62 | 191 | 0.98 (0.63 - 1.48) | 0.91 | 1.02 (0.66 - 1.56) | 0.89 |
| MDCR | 683 | 2,069 | 0.4 | 1.7 | 5 | 20 | 1.37 (0.40 - 4.26) | 0.60 | 1.05 (0.32 - 3.43) | 0.89 |
| Open Claims | 142,419 | 325,969 | 99.9 | 283.4 | 375 | 1,288 | 0.96 (0.82 - 1.11) | 0.57 | 0.91 (0.77 - 1.06) | 0.24 |
| OEHR | 7,810 | 19,852 | 2.2 | 6.3 | 16 | 62 | 0.80 (0.36 - 1.65) | 0.56 | 0.75 (0.35 - 1.62) | 0.47 |
| VA | 1,105 | 10,243 | 0.8 | 6.1 | <10 | 82 | 0.72 (0.23 - 1.80) | 0.52 | 0.49 (0.17 - 1.35) | 0.17 |
| Meta-analysis | 167,744 | 396,532 | 113.2 | 327.0 | <510 | 1,757 | 0.97 (0.84 - 1.10) | 0.61 | 0.92 (0.80 - 1.06) | 0.24 |
| Meta-analysis (w/o USOC) | 25,325 | 70,563 | 13.3 | 43.6 | <135 | 469 | 0.99 (0.75 - 1.31) | 0.96 | 0.94 (0.71 - 1.25) | 0.71 |
| **PS-stratification / on-treatment TAR** | | | | | | | | | | |
| CCAE | 10,165 | 23,691 | 6.7 | 19.6 | 48 | 134 | 1.05 (0.74 - 1.48) | 0.78 | 0.98 (0.69 - 1.39) | 0.89 |
| ODOD | 7,438 | 16,805 | 4.2 | 11.6 | 66 | 192 | 1.00 (0.74 - 1.34) | 0.99 | 0.96 (0.71 - 1.30) | 0.81 |
| MDCR | 714 | 2,069 | 0.5 | 1.7 | 5 | 20 | 1.03 (0.33 - 2.60) | 0.96 | 0.85 (0.30 - 2.36) | 0.75 |
| Open Claims | 152,533 | 325,992 | 104.8 | 283.5 | 393 | 1,288 | 0.93 (0.83 - 1.05) | 0.25 | 0.89 (0.79 - 1.01) | 0.09 |
| OEHR | 8,502 | 19,863 | 2.4 | 6.3 | 18 | 62 | 0.65 (0.37 - 1.11) | 0.13 | 0.59 (0.34 - 1.02) | 0.07 |
| VA | 1,266 | 11,422 | 0.9 | 6.9 | <10 | 104 | 0.83 (0.34 - 1.76) | 0.65 | 0.60 (0.26 - 1.36) | 0.22 |
| Meta-analysis | 178,638 | 386,351 | 118.1 | 320.9 | 525 | 1,676 | 0.94 (0.85 - 1.04) | 0.24 | 0.89 (0.79 - 1.00) | 0.06 |
| Meta-analysis (w/o USOC) | 26,105 | 60,359 | 13.3 | 37.5 | 132 | 388 | 0.96 (0.77 - 1.19) | 0.69 | 0.90 (0.72 - 1.12) | 0.36 |
| **PS-matching / on-treatment TAR without escalation** | | | | | | | | | | |
| CCAE | 9,537 | 23,687 | 5.6 | 17.1 | 36 | 109 | 1.03 (0.61 - 1.69) | 0.90 | 0.95 (0.57 - 1.58) | 0.84 |
| ODOD | 6,873 | 16,781 | 3.5 | 10.3 | 54 | 172 | 1.00 (0.62 - 1.56) | 1.00 | 1.02 (0.64 - 1.63) | 0.89 |
| MDCR | 683 | 2,069 | 0.4 | 1.5 | <5 | 19 | 1.22 (0.31 - 4.23) | 0.76 | 0.92 (0.25 - 3.40) | 0.88 |
| Open Claims | 142,419 | 325,969 | 87.1 | 241.1 | 317 | 1,086 | 0.92 (0.78 - 1.09) | 0.33 | 0.85 (0.72 - 1.01) | 0.09 |
| OEHR | 7,810 | 19,852 | 1.9 | 5.6 | 12 | 47 | 0.72 (0.30 - 1.60) | 0.45 | 0.66 (0.28 - 1.53) | 0.34 |
| VA | 1,105 | 10,243 | 0.6 | 5.3 | <10 | 71 | 0.96 (0.31 - 2.47) | 0.94 | 0.62 (0.22 - 1.76) | 0.37 |
| Meta-analysis | 167,744 | 396,532 | 98.7 | 279.3 | <429 | 1,485 | 0.93 (0.80 - 1.08) | 0.34 | 0.87 (0.75 - 1.01) | 0.08 |
| Meta-analysis (w/o USOC) | 25,325 | 70,563 | 11.7 | 38.3 | <112 | 399 | 0.97 (0.71 - 1.31) | 0.83 | 0.90 (0.66 - 1.23) | 0.52 |
| **PS-stratification / on-treatment TAR without escalation** | | | | | | | | | | |
| CCAE | 10,165 | 23,691 | 5.9 | 17.1 | 37 | 109 | 1.04 (0.70 - 1.53) | 0.83 | 0.95 (0.64 - 1.41) | 0.80 |
| ODOD | 7,438 | 16,805 | 3.8 | 10.3 | 58 | 173 | 1.03 (0.75 - 1.41) | 0.84 | 0.97 (0.71 - 1.33) | 0.85 |
| MDCR | 714 | 2,069 | 0.4 | 1.5 | <5 | 19 | 0.90 (0.26 - 2.48) | 0.86 | 0.73 (0.23 - 2.26) | 0.58 |
| Open Claims | 152,533 | 325,992 | 91.6 | 241.1 | 333 | 1,086 | 0.95 (0.83 - 1.08) | 0.41 | 0.90 (0.79 - 1.02) | 0.14 |
| OEHR | 8,502 | 19,863 | 2.1 | 5.6 | 13 | 47 | 0.67 (0.33 - 1.24) | 0.23 | 0.60 (0.31 - 1.15) | 0.13 |
| VA | 1,266 | 11,422 | 0.7 | 6.0 | <10 | 89 | 0.86 (0.31 - 2.03) | 0.75 | 0.62 (0.24 - 1.59) | 0.32 |
| Meta-analysis | 178,638 | 386,351 | 103.4 | 274.1 | 441 | 1,415 | 0.96 (0.85 - 1.07) | 0.43 | 0.90 (0.79 - 1.01) | 0.09 |
| Meta-analysis (w/o USOC) | 26,105 | 60,359 | 11.7 | 33.0 | 108 | 329 | 0.98 (0.78 - 1.24) | 0.88 | 0.90 (0.72 - 1.14) | 0.41 |

##### 1.1.18 Hypoglycemia

**Supplementary Table 18:** Exposures, outcomes and risk estimates for Hypoglycemia in GLP1RA (male) (T) vs SGLT2I (male) (C) new-user cohorts under four different propensity score (PS) adjustment / time-at-risk (TAR) designs across data sources. We report total patient exposure time in 1,000 patient-year (KYr) units, uncalibrated and calibrated hazard ratios (HRs) and their 95% confidence intervals (CIs). Greyed out entries did not pass study diagnostics and are included only for completeness.

| | Patients | | Exposure (KYr) | | Outcomes | | Uncalibrated | | Calibrated | |
|---|---|---|---|---|---|---|---|---|---|---|
| | T | C | T | C | T | C | HR (95% CI) | $p$ | HR (95% CI) | $p$ |
| **PS-matching / on-treatment TAR** | | | | | | | | | | |
| CCAE | 9,531 | 23,829 | 6.5 | 19.8 | 27 | 49 | 1.26 (0.61 - 2.52) | 0.52 | 1.18 (0.58 - 2.38) | 0.66 |
| ODOD | 7,046 | 17,242 | 4.1 | 12.0 | 21 | 51 | 1.24 (0.64 - 2.38) | 0.52 | 1.29 (0.67 - 2.50) | 0.45 |
| MDCR | 694 | 2,115 | 0.4 | 1.7 | <5 | 5 | 0.49 (0.02 - 4.05) | 0.59 | 0.38 (0.03 - 4.93) | 0.46 |
| Open Claims | 143,099 | 328,434 | 100.5 | 285.7 | 193 | 421 | 1.19 (0.94 - 1.51) | 0.15 | 1.13 (0.89 - 1.44) | 0.34 |
| OEHR | 7,860 | 20,051 | 2.2 | 6.4 | 11 | 18 | 1.40 (0.45 - 4.05) | 0.55 | 1.32 (0.44 - 3.94) | 0.63 |
| VA | 1,170 | 11,030 | 0.8 | 6.6 | 0 | 12 | | | | |
| Meta-analysis | 167,536 | 389,556 | 113.2 | 323.9 | 252 | 539 | 1.21 (0.98 - 1.49) | 0.08 | 1.15 (0.93 - 1.43) | 0.20 |
| Meta-analysis (w/o USOC) | 24,437 | 61,122 | 12.8 | 38.2 | 59 | 118 | 1.27 (0.82 - 1.98) | 0.28 | 1.21 (0.78 - 1.89) | 0.40 |
| **PS-stratification / on-treatment TAR** | | | | | | | | | | |
| CCAE | 10,147 | 23,833 | 6.8 | 19.8 | 27 | 49 | 1.48 (0.89 - 2.41) | 0.13 | 1.37 (0.83 - 2.26) | 0.22 |
| ODOD | 7,628 | 17,261 | 4.4 | 12.0 | 24 | 51 | 0.98 (0.58 - 1.62) | 0.93 | 0.94 (0.56 - 1.58) | 0.82 |
| MDCR | 725 | 2,115 | 0.5 | 1.7 | <5 | 5 | 0.56 (0.03 - 3.63) | 0.64 | 0.46 (0.04 - 5.22) | 0.53 |
| Open Claims | 152,956 | 328,462 | 105.3 | 285.7 | 219 | 421 | 1.31 (1.10 - 1.56) | <0.01 | 1.26 (1.05 - 1.50) | 0.01 |
| OEHR | 8,553 | 20,063 | 2.4 | 6.4 | 15 | 18 | 1.64 (0.79 - 3.38) | 0.18 | 1.47 (0.71 - 3.06) | 0.30 |
| VA | 1,336 | 12,356 | 0.9 | 7.4 | <10 | 14 | 0.18 (0.01 - 0.98) | 0.14 | 0.13 (0.01 - 1.30) | 0.08 |
| Meta-analysis | 179,284 | 389,619 | 118.8 | 323.9 | 285 | 539 | 1.31 (1.12 - 1.52) | <0.01 | 1.24 (1.06 - 1.46) | <0.01 |
| Meta-analysis (w/o USOC) | 26,328 | 61,157 | 13.5 | 38.2 | 66 | 118 | 1.28 (0.93 - 1.77) | 0.13 | 1.20 (0.87 - 1.67) | 0.27 |
| **PS-matching / on-treatment TAR without escalation** | | | | | | | | | | |
| CCAE | 9,531 | 23,829 | 5.7 | 17.3 | 20 | 38 | 1.07 (0.48 - 2.26) | 0.87 | 0.98 (0.45 - 2.14) | 0.94 |
| ODOD | 7,046 | 17,242 | 3.6 | 10.6 | 15 | 36 | 1.21 (0.57 - 2.49) | 0.62 | 1.24 (0.59 - 2.59) | 0.58 |
| MDCR | 694 | 2,115 | 0.4 | 1.5 | 0 | <5 | | | | |
| Open Claims | 143,099 | 328,434 | 87.6 | 243.0 | 149 | 289 | 1.27 (0.97 - 1.66) | 0.08 | 1.18 (0.90 - 1.55) | 0.23 |
| OEHR | 7,860 | 20,051 | 1.9 | 5.6 | 10 | 12 | 1.90 (0.59 - 6.00) | 0.28 | 1.73 (0.54 - 5.53) | 0.36 |
| VA | 1,170 | 11,030 | 0.6 | 5.7 | 0 | <10 | | | | |
| Meta-analysis | 167,536 | 389,556 | 98.8 | 276.6 | 194 | 375 | 1.26 (1.00 - 1.60) | 0.05 | 1.18 (0.93 - 1.50) | 0.16 |
| Meta-analysis (w/o USOC) | 24,437 | 61,122 | 11.2 | 33.6 | 45 | 86 | 1.24 (0.76 - 2.02) | 0.38 | 1.16 (0.71 - 1.89) | 0.57 |
| **PS-stratification / on-treatment TAR without escalation** | | | | | | | | | | |
| CCAE | 10,147 | 23,833 | 5.9 | 17.3 | 20 | 38 | 1.42 (0.78 - 2.49) | 0.24 | 1.29 (0.72 - 2.30) | 0.39 |
| ODOD | 7,628 | 17,261 | 3.9 | 10.6 | 18 | 36 | 1.01 (0.54 - 1.82) | 0.98 | 0.95 (0.52 - 1.74) | 0.87 |
| MDCR | 725 | 2,115 | 0.4 | 1.5 | 0 | <5 | | | | |
| Open Claims | 152,956 | 328,462 | 92.0 | 243.0 | 173 | 289 | 1.42 (1.16 - 1.74) | <0.01 | 1.35 (1.10 - 1.65) | <0.01 |
| OEHR | 8,553 | 20,063 | 2.1 | 5.6 | 12 | 12 | 2.12 (0.91 - 4.95) | 0.08 | 1.90 (0.81 - 4.44) | 0.14 |
| VA | 1,336 | 12,356 | 0.7 | 6.4 | 0 | <10 | | | | |
| Meta-analysis | 179,284 | 389,619 | 103.9 | 276.6 | 223 | 375 | 1.40 (1.18 - 1.68) | <0.01 | 1.32 (1.10 - 1.58) | <0.01 |
| Meta-analysis (w/o USOC) | 26,328 | 61,157 | 11.9 | 33.6 | 50 | 86 | 1.35 (0.92 - 1.96) | 0.12 | 1.24 (0.85 - 1.81) | 0.27 |

##### 1.1.19 Hypotension

**Supplementary Table 19:** Exposures, outcomes and risk estimates for Hypotension in GLP1RA (male) (T) vs SGLT2I (male) (C) new-user cohorts under four different propensity score (PS) adjustment / time-at-risk (TAR) designs across data sources. We report total patient exposure time in 1,000 patient-year (KYr) units, uncalibrated and calibrated hazard ratios (HRs) and their 95% confidence intervals (CIs). Greyed out entries did not pass study diagnostics and are included only for completeness.

| | Patients | | Exposure (KYr) | | Outcomes | | Uncalibrated | | Calibrated | |
|---|---|---|---|---|---|---|---|---|---|---|
| | T | C | T | C | T | C | HR (95% CI) | $p$ | HR (95% CI) | $p$ |
| **PS-matching / on-treatment TAR** | | | | | | | | | | |
| CCAE | 9,511 | 23,650 | 6.4 | 19.6 | 59 | 175 | 0.84 (0.56 - 1.24) | 0.39 | 0.78 (0.53 - 1.16) | 0.23 |
| ODOD | 6,853 | 16,703 | 4.0 | 11.5 | 66 | 229 | 0.83 (0.57 - 1.18) | 0.31 | 0.86 (0.60 - 1.24) | 0.44 |
| MDCR | 665 | 2,017 | 0.4 | 1.6 | 11 | 35 | 1.03 (0.40 - 2.47) | 0.95 | 0.79 (0.32 - 1.96) | 0.61 |
| Open Claims | 140,648 | 321,358 | 98.7 | 279.9 | 585 | 1,986 | 0.91 (0.81 - 1.03) | 0.13 | 0.86 (0.76 - 0.98) | 0.03 |
| OEHR | 7,755 | 19,795 | 2.2 | 6.3 | 30 | 83 | 0.97 (0.55 - 1.66) | 0.91 | 0.91 (0.52 - 1.60) | 0.76 |
| VA | 1,105 | 10,372 | 0.8 | 6.2 | 12 | 129 | 1.20 (0.53 - 2.46) | 0.64 | 0.81 (0.38 - 1.76) | 0.61 |
| Meta-analysis | 165,872 | 391,878 | 112.0 | 323.6 | 752 | 2,602 | 0.90 (0.81 - 1.01) | 0.07 | 0.86 (0.77 - 0.97) | 0.01 |
| Meta-analysis (w/o USOC) | 25,224 | 70,520 | 13.3 | 43.7 | 167 | 616 | 0.88 (0.70 - 1.11) | 0.29 | 0.84 (0.67 - 1.06) | 0.17 |
| **PS-stratification / on-treatment TAR** | | | | | | | | | | |
| CCAE | 10,114 | 23,656 | 6.7 | 19.6 | 59 | 177 | 0.89 (0.65 - 1.20) | 0.44 | 0.82 (0.60 - 1.12) | 0.23 |
| ODOD | 7,418 | 16,736 | 4.2 | 11.6 | 71 | 229 | 0.85 (0.64 - 1.13) | 0.27 | 0.82 (0.62 - 1.09) | 0.19 |
| MDCR | 697 | 2,017 | 0.4 | 1.6 | 11 | 35 | 1.09 (0.52 - 2.13) | 0.81 | 0.90 (0.44 - 1.83) | 0.77 |
| Open Claims | 150,554 | 321,381 | 103.5 | 279.9 | 624 | 1,989 | 0.91 (0.83 - 1.00) | 0.06 | 0.87 (0.79 - 0.96) | 0.02 |
| OEHR | 8,467 | 19,804 | 2.3 | 6.3 | 32 | 83 | 0.93 (0.60 - 1.41) | 0.73 | 0.83 (0.54 - 1.28) | 0.42 |
| VA | 1,263 | 11,420 | 0.9 | 6.9 | 18 | 152 | 1.40 (0.77 - 2.44) | 0.25 | 1.01 (0.57 - 1.80) | 0.87 |
| Meta-analysis | 176,553 | 381,577 | 116.8 | 317.4 | 786 | 2,478 | 0.91 (0.83 - 0.99) | 0.02 | 0.86 (0.78 - 0.95) | <0.01 |
| Meta-analysis (w/o USOC) | 25,999 | 60,196 | 13.3 | 37.5 | 162 | 489 | 0.88 (0.73 - 1.06) | 0.18 | 0.82 (0.68 - 1.00) | 0.06 |
| **PS-matching / on-treatment TAR without escalation** | | | | | | | | | | |
| CCAE | 9,511 | 23,650 | 5.6 | 17.2 | 48 | 144 | 1.00 (0.65 - 1.51) | 1.00 | 0.92 (0.61 - 1.40) | 0.71 |
| ODOD | 6,853 | 16,703 | 3.5 | 10.2 | 57 | 204 | 0.74 (0.49 - 1.09) | 0.14 | 0.76 (0.51 - 1.13) | 0.19 |
| MDCR | 665 | 2,017 | 0.4 | 1.4 | 9 | 29 | 0.82 (0.28 - 2.15) | 0.71 | 0.62 (0.22 - 1.71) | 0.36 |
| Open Claims | 140,648 | 321,358 | 86.1 | 238.2 | 497 | 1,664 | 0.92 (0.80 - 1.04) | 0.19 | 0.85 (0.74 - 0.97) | 0.03 |
| OEHR | 7,755 | 19,795 | 1.9 | 5.5 | 30 | 65 | 1.05 (0.58 - 1.84) | 0.88 | 0.95 (0.53 - 1.70) | 0.83 |
| VA | 1,105 | 10,372 | 0.6 | 5.4 | 10 | 110 | 1.05 (0.41 - 2.32) | 0.91 | 0.68 (0.29 - 1.61) | 0.39 |
| Meta-analysis | 165,872 | 391,878 | 97.7 | 276.5 | 642 | 2,187 | 0.91 (0.81 - 1.02) | 0.12 | 0.85 (0.76 - 0.97) | 0.01 |
| Meta-analysis (w/o USOC) | 25,224 | 70,520 | 11.6 | 38.3 | 145 | 523 | 0.90 (0.70 - 1.15) | 0.41 | 0.84 (0.65 - 1.08) | 0.20 |
| **PS-stratification / on-treatment TAR without escalation** | | | | | | | | | | |
| CCAE | 10,114 | 23,656 | 5.9 | 17.2 | 48 | 146 | 0.89 (0.63 - 1.24) | 0.50 | 0.81 (0.57 - 1.14) | 0.23 |
| ODOD | 7,418 | 16,736 | 3.8 | 10.3 | 62 | 204 | 0.84 (0.62 - 1.13) | 0.26 | 0.79 (0.58 - 1.07) | 0.14 |
| MDCR | 697 | 2,017 | 0.4 | 1.4 | 9 | 29 | 1.03 (0.45 - 2.16) | 0.94 | 0.83 (0.38 - 1.82) | 0.65 |
| Open Claims | 150,554 | 321,381 | 90.5 | 238.2 | 534 | 1,667 | 0.93 (0.83 - 1.03) | 0.14 | 0.88 (0.79 - 0.97) | 0.03 |
| OEHR | 8,467 | 19,804 | 2.0 | 5.5 | 32 | 65 | 1.18 (0.74 - 1.82) | 0.48 | 1.05 (0.67 - 1.65) | 0.81 |
| VA | 1,263 | 11,420 | 0.7 | 5.9 | 15 | 129 | 1.36 (0.70 - 2.50) | 0.35 | 0.98 (0.52 - 1.86) | 0.86 |
| Meta-analysis | 176,553 | 381,577 | 102.2 | 271.2 | 676 | 2,082 | 0.92 (0.84 - 1.01) | 0.09 | 0.87 (0.78 - 0.96) | <0.01 |
| Meta-analysis (w/o USOC) | 25,999 | 60,196 | 11.7 | 33.0 | 142 | 415 | 0.92 (0.75 - 1.12) | 0.40 | 0.84 (0.69 - 1.04) | 0.12 |

##### 1.1.20 Joint pain

**Supplementary Table 20:** Exposures, outcomes and risk estimates for Joint pain in GLP1RA (male) (T) vs SGLT2I (male) (C) new-user cohorts under four different propensity score (PS) adjustment / time-at-risk (TAR) designs across data sources. We report total patient exposure time in 1,000 patient-year (KYr) units, uncalibrated and calibrated hazard ratios (HRs) and their 95% confidence intervals (CIs). Greyed out entries did not pass study diagnostics and are included only for completeness.

| | Patients | | Exposure (KYr) | | Outcomes | | Uncalibrated | | Calibrated | |
|---|---|---|---|---|---|---|---|---|---|---|
| | T | C | T | C | T | C | HR (95% CI) | $p$ | HR (95% CI) | $p$ |
| **PS-matching / on-treatment TAR** | | | | | | | | | | |
| CCAE | 9,238 | 23,071 | 6.2 | 19.1 | 49 | 137 | 0.76 (0.48 - 1.16) | 0.21 | 0.71 (0.46 - 1.09) | 0.12 |
| ODOD | 6,735 | 16,669 | 3.9 | 11.5 | 31 | 103 | 0.70 (0.39 - 1.22) | 0.22 | 0.73 (0.41 - 1.29) | 0.29 |
| MDCR | 674 | 2,062 | 0.4 | 1.6 | <5 | 11 | 1.08 (0.14 - 6.73) | 0.94 | 0.82 (0.12 - 5.77) | 0.84 |
| Open Claims | 137,603 | 317,206 | 96.5 | 276.1 | 444 | 1,151 | 1.01 (0.86 - 1.17) | 0.94 | 0.95 (0.81 - 1.12) | 0.57 |
| OEHR | 7,589 | 19,447 | 2.1 | 6.2 | 14 | 51 | 0.36 (0.10 - 0.98) | 0.07 | 0.34 (0.11 - 1.05) | 0.06 |
| VA | 1,046 | 9,692 | 0.7 | 5.9 | <10 | 39 | 3.27 (1.04 - 9.07) | 0.03 | 2.21 (0.75 - 6.53) | 0.15 |
| Meta-analysis | 162,211 | 386,085 | 109.5 | 318.8 | <548 | 1,481 | 0.89 (0.64 - 1.24) | 0.49 | 0.85 (0.61 - 1.18) | 0.33 |
| Meta-analysis (w/o USOC) | 24,608 | 68,879 | 12.9 | 42.7 | <104 | 330 | 0.85 (0.46 - 1.57) | 0.60 | 0.81 (0.43 - 1.50) | 0.51 |
| **PS-stratification / on-treatment TAR** | | | | | | | | | | |
| CCAE | 9,816 | 23,075 | 6.5 | 19.1 | 52 | 137 | 1.03 (0.73 - 1.43) | 0.87 | 0.96 (0.68 - 1.34) | 0.80 |
| ODOD | 7,306 | 16,688 | 4.2 | 11.6 | 38 | 103 | 0.88 (0.58 - 1.29) | 0.51 | 0.84 (0.57 - 1.25) | 0.40 |
| MDCR | 699 | 2,062 | 0.4 | 1.6 | <5 | 11 | 0.72 (0.15 - 2.47) | 0.64 | 0.59 (0.15 - 2.36) | 0.46 |
| Open Claims | 146,963 | 317,231 | 101.2 | 276.1 | 492 | 1,151 | 1.08 (0.96 - 1.20) | 0.20 | 1.03 (0.92 - 1.15) | 0.66 |
| OEHR | 8,249 | 19,457 | 2.3 | 6.2 | 16 | 51 | 0.80 (0.43 - 1.41) | 0.46 | 0.71 (0.39 - 1.30) | 0.28 |
| VA | 1,201 | 10,977 | 0.8 | 6.6 | 11 | 46 | 1.88 (0.80 - 4.20) | 0.13 | 1.36 (0.59 - 3.11) | 0.48 |
| Meta-analysis | 172,334 | 376,451 | 114.1 | 313.0 | 598 | 1,442 | 1.05 (0.95 - 1.16) | 0.36 | 1.00 (0.89 - 1.12) | 0.90 |
| Meta-analysis (w/o USOC) | 25,371 | 59,220 | 12.9 | 36.9 | 106 | 291 | 0.93 (0.74 - 1.18) | 0.57 | 0.88 (0.69 - 1.11) | 0.30 |
| **PS-matching / on-treatment TAR without escalation** | | | | | | | | | | |
| CCAE | 9,238 | 23,071 | 5.4 | 16.7 | 42 | 115 | 0.89 (0.55 - 1.43) | 0.65 | 0.82 (0.51 - 1.33) | 0.43 |
| ODOD | 6,735 | 16,669 | 3.5 | 10.2 | 27 | 95 | 0.64 (0.34 - 1.15) | 0.15 | 0.65 (0.35 - 1.21) | 0.18 |
| MDCR | 674 | 2,062 | 0.4 | 1.5 | <5 | 10 | 1.56 (0.18 - 13.6) | 0.69 | 1.17 (0.13 - 10.2) | 0.89 |
| Open Claims | 137,603 | 317,206 | 84.2 | 234.9 | 378 | 972 | 1.04 (0.88 - 1.23) | 0.66 | 0.97 (0.81 - 1.15) | 0.70 |
| OEHR | 7,589 | 19,447 | 1.8 | 5.4 | 14 | 45 | 0.38 (0.11 - 1.04) | 0.09 | 0.34 (0.11 - 1.07) | 0.07 |
| VA | 1,046 | 9,692 | 0.6 | 5.0 | <10 | 31 | 4.07 (1.24 - 12.2) | 0.02 | 2.62 (0.84 - 8.23) | 0.10 |
| Meta-analysis | 161,165 | 376,393 | 94.9 | 267.2 | 461 | 1,227 | 0.87 (0.64 - 1.17) | 0.34 | 0.81 (0.60 - 1.10) | 0.17 |
| Meta-analysis (w/o USOC) | 23,562 | 59,187 | 10.8 | 32.4 | 83 | 255 | 0.72 (0.49 - 1.06) | 0.10 | 0.67 (0.46 - 0.99) | 0.05 |
| **PS-stratification / on-treatment TAR without escalation** | | | | | | | | | | |
| CCAE | 9,816 | 23,075 | 5.7 | 16.7 | 44 | 115 | 1.10 (0.76 - 1.58) | 0.60 | 1.00 (0.69 - 1.45) | 0.92 |
| ODOD | 7,306 | 16,688 | 3.7 | 10.3 | 33 | 95 | 0.83 (0.54 - 1.24) | 0.37 | 0.78 (0.51 - 1.18) | 0.25 |
| MDCR | 699 | 2,062 | 0.4 | 1.5 | <5 | 10 | 0.77 (0.16 - 2.81) | 0.72 | 0.62 (0.15 - 2.57) | 0.51 |
| Open Claims | 146,963 | 317,231 | 88.4 | 234.9 | 423 | 972 | 1.06 (0.93 - 1.19) | 0.38 | 1.00 (0.88 - 1.13) | 0.91 |
| OEHR | 8,249 | 19,457 | 2.0 | 5.4 | 16 | 45 | 0.94 (0.49 - 1.70) | 0.84 | 0.84 (0.45 - 1.56) | 0.58 |
| VA | 1,201 | 10,977 | 0.7 | 5.7 | 10 | 36 | 1.85 (0.75 - 4.36) | 0.17 | 1.34 (0.55 - 3.23) | 0.53 |
| Meta-analysis | 172,334 | 376,451 | 99.8 | 267.3 | 516 | 1,227 | 1.04 (0.93 - 1.16) | 0.50 | 0.98 (0.87 - 1.10) | 0.69 |
| Meta-analysis (w/o USOC) | 25,371 | 59,220 | 11.4 | 32.4 | 93 | 255 | 0.97 (0.75 - 1.24) | 0.79 | 0.89 (0.69 - 1.15) | 0.38 |

##### 1.1.21 Lower extremity amputation

**Supplementary Table 21:** Exposures, outcomes and risk estimates for Lower extremity amputation in GLP1RA (male) (T) vs SGLT2I (male) (C) new-user cohorts under four different propensity score (PS) adjustment / time-at-risk (TAR) designs across data sources. We report total patient exposure time in 1,000 patient-year (KYr) units, uncalibrated and calibrated hazard ratios (HRs) and their 95% confidence intervals (CIs). Greyed out entries did not pass study diagnostics and are included only for completeness.

| | Patients | | Exposure (KYr) | | Outcomes | | Uncalibrated | | Calibrated | |
|---|---|---|---|---|---|---|---|---|---|---|
| | T | C | T | C | T | C | HR (95% CI) | $p$ | HR (95% CI) | $p$ |
| **PS-matching / on-treatment TAR** | | | | | | | | | | |
| CCAE | 9,688 | 24,118 | 6.6 | 20.1 | 5 | 19 | 0.43 (0.10 - 1.36) | 0.21 | 0.40 (0.11 - 1.50) | 0.17 |
| ODOD | 7,134 | 17,429 | 4.1 | 12.1 | 6 | 13 | 0.95 (0.24 - 3.38) | 0.95 | 0.99 (0.26 - 3.73) | 0.97 |
| MDCR | 701 | 2,129 | 0.4 | 1.7 | 0 | <5 | | | | |
| Open Claims | 144,948 | 332,213 | 101.8 | 289.1 | 48 | 159 | 0.65 (0.39 - 1.03) | 0.07 | 0.61 (0.38 - 0.99) | 0.05 |
| OEHR | 7,943 | 20,209 | 2.2 | 6.5 | 0 | <5 | | | | |
| VA | 1,179 | 11,159 | 0.8 | 6.7 | 0 | 11 | | | | |
| Meta-analysis | 154,636 | 356,331 | 108.4 | 309.1 | 53 | 178 | 0.62 (0.39 - 0.97) | 0.04 | 0.59 (0.37 - 0.92) | 0.02 |
| Meta-analysis (w/o USOC) | 9,688 | 24,118 | 6.6 | 20.1 | 5 | 19 | 0.43 (0.10 - 1.36) | 0.21 | 0.41 (0.11 - 1.53) | 0.19 |
| **PS-stratification / on-treatment TAR** | | | | | | | | | | |
| CCAE | 10,320 | 24,122 | 6.9 | 20.1 | 6 | 19 | 1.01 (0.36 - 2.48) | 0.99 | 0.94 (0.36 - 2.47) | 0.90 |
| ODOD | 7,717 | 17,448 | 4.4 | 12.1 | 6 | 13 | 0.78 (0.26 - 2.15) | 0.65 | 0.75 (0.26 - 2.17) | 0.60 |
| MDCR | 731 | 2,129 | 0.5 | 1.7 | 0 | <5 | | | | |
| Open Claims | 155,107 | 332,242 | 106.8 | 289.1 | 49 | 159 | 0.84 (0.59 - 1.16) | 0.30 | 0.80 (0.57 - 1.12) | 0.20 |
| OEHR | 8,659 | 20,221 | 2.4 | 6.5 | 0 | <5 | | | | |
| VA | 1,350 | 12,506 | 0.9 | 7.5 | 0 | 11 | | | | |
| Meta-analysis | 165,427 | 356,364 | 113.7 | 309.2 | 55 | 178 | 0.85 (0.62 - 1.17) | 0.33 | 0.81 (0.59 - 1.12) | 0.21 |
| Meta-analysis (w/o USOC) | 10,320 | 24,122 | 6.9 | 20.1 | 6 | 19 | 1.01 (0.36 - 2.48) | 0.99 | 0.95 (0.36 - 2.49) | 0.91 |
| **PS-matching / on-treatment TAR without escalation** | | | | | | | | | | |
| CCAE | 9,688 | 24,118 | 5.7 | 17.5 | 5 | 14 | 0.90 (0.19 - 3.30) | 0.88 | 0.83 (0.20 - 3.44) | 0.79 |
| ODOD | 7,134 | 17,429 | 3.7 | 10.7 | 5 | 10 | 1.35 (0.32 - 5.50) | 0.68 | 1.38 (0.33 - 5.78) | 0.66 |
| MDCR | 701 | 2,129 | 0.4 | 1.5 | 0 | <5 | | | | |
| Open Claims | 144,948 | 332,213 | 88.7 | 245.8 | 38 | 116 | 0.63 (0.36 - 1.06) | 0.10 | 0.59 (0.34 - 1.01) | 0.06 |
| OEHR | 7,943 | 20,209 | 1.9 | 5.7 | 0 | <5 | | | | |
| VA | 1,179 | 11,159 | 0.6 | 5.8 | 0 | 11 | | | | |
| Meta-analysis | 144,948 | 332,213 | 88.7 | 245.8 | 38 | 116 | 0.63 (0.36 - 1.06) | 0.10 | 0.59 (0.34 - 1.02) | 0.06 |
| **PS-stratification / on-treatment TAR without escalation** | | | | | | | | | | |
| CCAE | 10,320 | 24,122 | 6.0 | 17.5 | 5 | 14 | 1.37 (0.43 - 3.73) | 0.57 | 1.25 (0.42 - 3.67) | 0.69 |
| ODOD | 7,717 | 17,448 | 3.9 | 10.8 | 5 | 10 | 0.93 (0.27 - 2.91) | 0.91 | 0.88 (0.27 - 2.89) | 0.83 |
| MDCR | 731 | 2,129 | 0.4 | 1.5 | 0 | <5 | | | | |
| Open Claims | 155,107 | 332,242 | 93.3 | 245.8 | 39 | 116 | 0.89 (0.60 - 1.29) | 0.55 | 0.84 (0.57 - 1.23) | 0.38 |
| OEHR | 8,659 | 20,221 | 2.1 | 5.7 | 0 | <5 | | | | |
| VA | 1,350 | 12,506 | 0.7 | 6.5 | 0 | 11 | | | | |
| Meta-analysis | 155,107 | 332,242 | 93.3 | 245.8 | 39 | 116 | 0.89 (0.60 - 1.29) | 0.55 | 0.84 (0.57 - 1.23) | 0.36 |

##### 1.1.22 Nausea

**Supplementary Table 22:** Exposures, outcomes and risk estimates for Nausea in GLP1RA (male) (T) vs SGLT2I (male) (C) new-user cohorts under four different propensity score (PS) adjustment / time-at-risk (TAR) designs across data sources. We report total patient exposure time in 1,000 patient-year (KYr) units, uncalibrated and calibrated hazard ratios (HRs) and their 95% confidence intervals (CIs). Greyed out entries did not pass study diagnostics and are included only for completeness.

| | Patients | | Exposure (KYr) | | Outcomes | | Uncalibrated | | Calibrated | |
|---|---|---|---|---|---|---|---|---|---|---|
| | T | C | T | C | T | C | HR (95% CI) | $p$ | HR (95% CI) | $p$ |
| **PS-matching / on-treatment TAR** | | | | | | | | | | |
| CCAE | 8,595 | 21,816 | 5.8 | 17.8 | 213 | 432 | 1.40 (1.12 - 1.73) | <0.01 | 1.30 (1.05 - 1.61) | 0.03 |
| ODOD | 6,268 | 15,551 | 3.6 | 10.7 | 165 | 301 | 1.51 (1.17 - 1.95) | <0.01 | 1.57 (1.21 - 2.03) | <0.01 |
| MDCR | 633 | 1,958 | 0.4 | 1.5 | 9 | 38 | 0.60 (0.19 - 1.55) | 0.33 | 0.46 (0.16 - 1.30) | 0.15 |
| Open Claims | 128,477 | 299,122 | 90.1 | 261.1 | 1,849 | 3,189 | 1.62 (1.50 - 1.75) | <0.01 | 1.53 (1.40 - 1.67) | <0.01 |
| OEHR | 7,266 | 18,772 | 2.0 | 5.9 | 113 | 162 | 2.10 (1.51 - 2.92) | <0.01 | 1.98 (1.42 - 2.76) | <0.01 |
| VA | 1,104 | 10,357 | 0.8 | 6.3 | 20 | 45 | 3.29 (1.54 - 7.00) | <0.01 | 2.23 (1.05 - 4.77) | 0.04 |
| Meta-analysis | 151,710 | 365,618 | 102.2 | 301.8 | 2,360 | 4,129 | 1.61 (1.50 - 1.73) | <0.01 | 1.53 (1.42 - 1.66) | <0.01 |
| Meta-analysis (w/o USOC) | 23,233 | 66,496 | 12.1 | 40.7 | 511 | 940 | 1.72 (1.33 - 2.23) | <0.01 | 1.64 (1.26 - 2.13) | <0.01 |
| **PS-stratification / on-treatment TAR** | | | | | | | | | | |
| CCAE | 9,138 | 21,821 | 6.0 | 17.8 | 232 | 432 | 1.38 (1.16 - 1.63) | <0.01 | 1.28 (1.08 - 1.52) | <0.01 |
| ODOD | 6,794 | 15,574 | 3.8 | 10.7 | 182 | 302 | 1.54 (1.26 - 1.87) | <0.01 | 1.48 (1.21 - 1.80) | <0.01 |
| MDCR | 659 | 1,958 | 0.4 | 1.5 | 9 | 38 | 0.80 (0.36 - 1.62) | 0.56 | 0.66 (0.31 - 1.41) | 0.29 |
| Open Claims | 136,903 | 299,241 | 94.3 | 261.1 | 2,002 | 3,193 | 1.58 (1.49 - 1.68) | <0.01 | 1.51 (1.41 - 1.61) | <0.01 |
| OEHR | 7,906 | 18,784 | 2.2 | 5.9 | 122 | 162 | 1.86 (1.44 - 2.39) | <0.01 | 1.67 (1.29 - 2.15) | <0.01 |
| VA | 1,265 | 11,542 | 0.9 | 7.0 | 22 | 53 | 3.30 (1.77 - 6.06) | <0.01 | 2.39 (1.29 - 4.43) | <0.01 |
| Meta-analysis | 160,741 | 355,420 | 106.3 | 295.6 | 2,538 | 4,089 | 1.57 (1.49 - 1.65) | <0.01 | 1.49 (1.38 - 1.61) | <0.01 |
| Meta-analysis (w/o USOC) | 23,838 | 56,179 | 12.0 | 34.5 | 536 | 896 | 1.55 (1.32 - 1.81) | <0.01 | 1.45 (1.23 - 1.71) | <0.01 |
| **PS-matching / on-treatment TAR without escalation** | | | | | | | | | | |
| CCAE | 8,595 | 21,816 | 5.1 | 15.7 | 192 | 359 | 1.45 (1.16 - 1.83) | <0.01 | 1.34 (1.06 - 1.69) | 0.02 |
| ODOD | 6,268 | 15,551 | 3.2 | 9.6 | 151 | 255 | 1.54 (1.17 - 2.02) | <0.01 | 1.58 (1.20 - 2.08) | <0.01 |
| MDCR | 633 | 1,958 | 0.4 | 1.4 | 6 | 35 | 0.46 (0.13 - 1.27) | 0.18 | 0.34 (0.11 - 1.07) | 0.07 |
| Open Claims | 128,477 | 299,122 | 78.8 | 222.8 | 1,622 | 2,538 | 1.73 (1.59 - 1.88) | <0.01 | 1.61 (1.47 - 1.76) | <0.01 |
| OEHR | 7,266 | 18,772 | 1.8 | 5.2 | 98 | 130 | 2.06 (1.44 - 2.96) | <0.01 | 1.88 (1.31 - 2.69) | <0.01 |
| VA | 1,104 | 10,357 | 0.6 | 5.4 | 17 | 35 | 4.42 (1.96 - 10.0) | <0.01 | 2.85 (1.26 - 6.47) | 0.01 |
| Meta-analysis | 151,710 | 365,618 | 89.3 | 258.7 | 2,080 | 3,317 | 1.71 (1.58 - 1.84) | <0.01 | 1.60 (1.47 - 1.74) | <0.01 |
| Meta-analysis (w/o USOC) | 23,233 | 66,496 | 10.6 | 35.9 | 458 | 779 | 1.81 (1.35 - 2.42) | <0.01 | 1.68 (1.25 - 2.26) | <0.01 |
| **PS-stratification / on-treatment TAR without escalation** | | | | | | | | | | |
| CCAE | 9,138 | 21,821 | 5.3 | 15.7 | 207 | 359 | 1.53 (1.27 - 1.83) | <0.01 | 1.39 (1.16 - 1.67) | <0.01 |
| ODOD | 6,794 | 15,574 | 3.4 | 9.6 | 165 | 256 | 1.66 (1.34 - 2.05) | <0.01 | 1.56 (1.26 - 1.93) | <0.01 |
| MDCR | 659 | 1,958 | 0.4 | 1.4 | 6 | 35 | 0.55 (0.21 - 1.26) | 0.20 | 0.45 (0.18 - 1.10) | 0.08 |
| Open Claims | 136,903 | 299,241 | 82.5 | 222.9 | 1,763 | 2,542 | 1.71 (1.60 - 1.83) | <0.01 | 1.62 (1.51 - 1.74) | <0.01 |
| OEHR | 7,906 | 18,784 | 1.9 | 5.2 | 105 | 130 | 1.95 (1.48 - 2.57) | <0.01 | 1.75 (1.32 - 2.31) | <0.01 |
| VA | 1,265 | 11,542 | 0.7 | 6.0 | 19 | 41 | 3.69 (1.84 - 7.25) | <0.01 | 2.67 (1.34 - 5.30) | <0.01 |
| Meta-analysis | 160,741 | 355,420 | 93.1 | 253.3 | 2,240 | 3,287 | 1.70 (1.60 - 1.80) | <0.01 | 1.59 (1.48 - 1.72) | <0.01 |
| Meta-analysis (w/o USOC) | 23,838 | 56,179 | 10.6 | 30.5 | 477 | 745 | 1.65 (1.45 - 1.87) | <0.01 | 1.52 (1.33 - 1.73) | <0.01 |

##### 1.1.23 Peripheral edema

**Supplementary Table 23:** Exposures, outcomes and risk estimates for Peripheral edema in GLP1RA (male) (T) vs SGLT2I (male) (C) new-user cohorts under four different propensity score (PS) adjustment / time-at-risk (TAR) designs across data sources. We report total patient exposure time in 1,000 patient-year (KYr) units, uncalibrated and calibrated hazard ratios (HRs) and their 95% confidence intervals (CIs). Greyed out entries did not pass study diagnostics and are included only for completeness.

| | Patients | | Exposure (KYr) | | Outcomes | | Uncalibrated | | Calibrated | |
|---|---|---|---|---|---|---|---|---|---|---|
| | T | C | T | C | T | C | HR (95% CI) | $p$ | HR (95% CI) | $p$ |
| **PS-matching / on-treatment TAR** | | | | | | | | | | |
| CCAE | 7,982 | 20,325 | 5.3 | 16.4 | 189 | 516 | 1.03 (0.82 - 1.29) | 0.78 | 0.96 (0.76 - 1.21) | 0.73 |
| ODOD | 5,627 | 14,073 | 3.2 | 9.6 | 150 | 405 | 1.33 (1.02 - 1.72) | 0.03 | 1.38 (1.06 - 1.80) | 0.02 |
| MDCR | 526 | 1,632 | 0.3 | 1.3 | 14 | 56 | 0.68 (0.25 - 1.62) | 0.42 | 0.52 (0.20 - 1.34) | 0.18 |
| Open Claims | 114,996 | 269,857 | 80.2 | 235.1 | 1,568 | 4,193 | 1.13 (1.05 - 1.23) | <0.01 | 1.07 (0.98 - 1.17) | 0.16 |
| OEHR | 6,688 | 17,553 | 1.8 | 5.5 | 61 | 130 | 0.93 (0.60 - 1.41) | 0.74 | 0.88 (0.57 - 1.34) | 0.56 |
| VA | 872 | 8,466 | 0.6 | 5.1 | 15 | 161 | 0.82 (0.38 - 1.61) | 0.59 | 0.55 (0.27 - 1.15) | 0.12 |
| Meta-analysis | 136,165 | 330,274 | 91.2 | 271.7 | 1,983 | 5,405 | 1.13 (1.05 - 1.21) | <0.01 | 1.07 (0.99 - 1.16) | 0.11 |
| Meta-analysis (w/o USOC) | 21,169 | 60,417 | 10.9 | 36.7 | 415 | 1,212 | 1.10 (0.92 - 1.31) | 0.31 | 1.04 (0.87 - 1.25) | 0.68 |
| **PS-stratification / on-treatment TAR** | | | | | | | | | | |
| CCAE | 8,446 | 20,331 | 5.5 | 16.5 | 195 | 518 | 1.05 (0.88 - 1.25) | 0.55 | 0.98 (0.82 - 1.17) | 0.81 |
| ODOD | 6,057 | 14,095 | 3.4 | 9.6 | 160 | 406 | 1.11 (0.91 - 1.34) | 0.30 | 1.07 (0.88 - 1.30) | 0.56 |
| MDCR | 557 | 1,633 | 0.3 | 1.3 | 15 | 56 | 0.93 (0.50 - 1.66) | 0.83 | 0.77 (0.42 - 1.41) | 0.41 |
| Open Claims | 122,001 | 269,915 | 83.6 | 235.1 | 1,665 | 4,195 | 1.11 (1.05 - 1.18) | <0.01 | 1.06 (0.99 - 1.13) | 0.15 |
| OEHR | 7,265 | 17,562 | 2.0 | 5.5 | 68 | 130 | 1.30 (0.94 - 1.78) | 0.10 | 1.17 (0.85 - 1.61) | 0.35 |
| VA | 989 | 9,289 | 0.7 | 5.6 | 15 | 177 | 0.66 (0.35 - 1.17) | 0.18 | 0.48 (0.26 - 0.87) | 0.02 |
| Meta-analysis | 143,769 | 321,903 | 94.5 | 266.7 | 2,088 | 5,249 | 1.11 (1.05 - 1.17) | <0.01 | 1.05 (0.98 - 1.14) | 0.19 |
| Meta-analysis (w/o USOC) | 21,768 | 51,988 | 10.9 | 31.6 | 423 | 1,054 | 1.11 (0.98 - 1.25) | 0.09 | 1.04 (0.91 - 1.18) | 0.60 |
| **PS-matching / on-treatment TAR without escalation** | | | | | | | | | | |
| CCAE | 7,982 | 20,325 | 4.7 | 14.5 | 162 | 438 | 1.10 (0.86 - 1.39) | 0.44 | 1.01 (0.79 - 1.29) | 0.84 |
| ODOD | 5,627 | 14,073 | 2.9 | 8.6 | 137 | 336 | 1.44 (1.09 - 1.89) | 0.01 | 1.47 (1.12 - 1.94) | <0.01 |
| MDCR | 526 | 1,632 | 0.3 | 1.1 | 11 | 48 | 0.65 (0.21 - 1.69) | 0.42 | 0.49 (0.17 - 1.38) | 0.18 |
| Open Claims | 114,996 | 269,857 | 70.1 | 200.8 | 1,353 | 3,473 | 1.14 (1.04 - 1.24) | <0.01 | 1.06 (0.97 - 1.16) | 0.29 |
| OEHR | 6,688 | 17,553 | 1.6 | 4.9 | 52 | 105 | 1.27 (0.78 - 2.04) | 0.32 | 1.16 (0.72 - 1.87) | 0.57 |
| VA | 872 | 8,466 | 0.5 | 4.4 | 11 | 136 | 1.04 (0.43 - 2.24) | 0.92 | 0.67 (0.30 - 1.53) | 0.36 |
| Meta-analysis | 136,165 | 330,274 | 79.7 | 233.1 | 1,715 | 4,488 | 1.16 (1.07 - 1.25) | <0.01 | 1.08 (1.00 - 1.18) | 0.07 |
| Meta-analysis (w/o USOC) | 21,169 | 60,417 | 9.6 | 32.3 | 362 | 1,015 | 1.23 (1.04 - 1.45) | 0.01 | 1.14 (0.96 - 1.36) | 0.17 |
| **PS-stratification / on-treatment TAR without escalation** | | | | | | | | | | |
| CCAE | 8,446 | 20,331 | 4.9 | 14.5 | 167 | 440 | 1.04 (0.86 - 1.26) | 0.66 | 0.95 (0.79 - 1.15) | 0.62 |
| ODOD | 6,057 | 14,095 | 3.0 | 8.6 | 147 | 336 | 1.23 (1.00 - 1.51) | 0.05 | 1.16 (0.94 - 1.43) | 0.18 |
| MDCR | 557 | 1,633 | 0.3 | 1.1 | 12 | 48 | 0.84 (0.42 - 1.59) | 0.62 | 0.68 (0.35 - 1.33) | 0.27 |
| Open Claims | 122,001 | 269,915 | 73.2 | 200.8 | 1,445 | 3,475 | 1.14 (1.07 - 1.22) | <0.01 | 1.08 (1.01 - 1.16) | 0.06 |
| OEHR | 7,265 | 17,562 | 1.7 | 4.9 | 56 | 105 | 1.43 (1.00 - 2.02) | 0.05 | 1.28 (0.90 - 1.82) | 0.19 |
| VA | 989 | 9,289 | 0.5 | 4.8 | 11 | 151 | 0.67 (0.32 - 1.27) | 0.25 | 0.48 (0.24 - 0.96) | 0.04 |
| Meta-analysis | 143,769 | 321,903 | 82.9 | 228.8 | 1,815 | 4,356 | 1.15 (1.08 - 1.22) | <0.01 | 1.08 (1.00 - 1.16) | 0.06 |
| Meta-analysis (w/o USOC) | 21,768 | 51,988 | 9.6 | 28.0 | 370 | 881 | 1.18 (1.00 - 1.38) | 0.05 | 1.08 (0.92 - 1.28) | 0.37 |

##### 1.1.24 Photosensitivity

**Supplementary Table 24:** Exposures, outcomes and risk estimates for Photosensitivity in GLP1RA (male) (T) vs SGLT2I (male) (C) new-user cohorts under four different propensity score (PS) adjustment / time-at-risk (TAR) designs across data sources. We report total patient exposure time in 1,000 patient-year (KYr) units, uncalibrated and calibrated hazard ratios (HRs) and their 95% confidence intervals (CIs). Greyed out entries did not pass study diagnostics and are included only for completeness.

| | Patients | | Exposure (KYr) | | Outcomes | | Uncalibrated | | Calibrated | |
|---|---|---|---|---|---|---|---|---|---|---|
| | T | C | T | C | T | C | HR (95% CI) | $p$ | HR (95% CI) | $p$ |
| **PS-matching / on-treatment TAR** | | | | | | | | | | |
| CCAE | 9,624 | 23,941 | 6.5 | 19.9 | 6 | 11 | 0.94 (0.13 - 4.62) | 0.94 | 0.87 (0.15 - 5.20) | 0.88 |
| ODOD | 7,098 | 17,330 | 4.1 | 12.0 | <5 | 8 | 0.69 (0.03 - 7.65) | 0.79 | 0.71 (0.05 - 11.2) | 0.81 |
| MDCR | 689 | 2,105 | 0.4 | 1.7 | <5 | <5 | | | | |
| Open Claims | 143,821 | 329,537 | 100.9 | 286.5 | 26 | 68 | 1.21 (0.64 - 2.21) | 0.55 | 1.15 (0.62 - 2.13) | 0.67 |
| OEHR | 7,905 | 20,095 | 2.2 | 6.4 | <5 | 5 | 1.68 (0.37 - 7.74) | 0.50 | 1.59 (0.35 - 7.24) | 0.55 |
| VA | 1,167 | 11,033 | 0.8 | 6.6 | 0 | <10 | | | | |
| Meta-analysis | 143,821 | 329,537 | 100.9 | 286.5 | 26 | 68 | 1.21 (0.64 - 2.21) | 0.55 | 1.15 (0.62 - 2.14) | 0.66 |
| **PS-stratification / on-treatment TAR** | | | | | | | | | | |
| CCAE | 10,254 | 23,945 | 6.8 | 19.9 | 6 | 11 | 1.50 (0.50 - 4.10) | 0.45 | 1.40 (0.49 - 3.99) | 0.53 |
| ODOD | 7,688 | 17,348 | 4.4 | 12.0 | <5 | 8 | 0.50 (0.07 - 2.14) | 0.42 | 0.48 (0.09 - 2.60) | 0.39 |
| MDCR | 720 | 2,105 | 0.5 | 1.7 | <5 | <5 | 2.34 (0.11 - 25.0) | 0.54 | 1.93 (0.13 - 29.5) | 0.64 |
| Open Claims | 153,926 | 329,565 | 105.8 | 286.5 | 32 | 68 | 1.31 (0.83 - 2.04) | 0.24 | 1.25 (0.80 - 1.97) | 0.33 |
| OEHR | 8,610 | 20,107 | 2.4 | 6.4 | <5 | 5 | 1.59 (0.34 - 7.11) | 0.55 | 1.43 (0.31 - 6.54) | 0.65 |
| VA | 1,337 | 12,352 | 0.9 | 7.4 | 0 | <10 | | | | |
| Meta-analysis | 153,926 | 329,565 | 105.8 | 286.5 | 32 | 68 | 1.31 (0.83 - 2.04) | 0.24 | 1.25 (0.79 - 1.96) | 0.34 |
| **PS-matching / on-treatment TAR without escalation** | | | | | | | | | | |
| CCAE | 9,624 | 23,941 | 5.7 | 17.4 | 5 | 10 | 0.93 (0.13 - 4.57) | 0.94 | 0.86 (0.14 - 5.10) | 0.86 |
| ODOD | 7,098 | 17,330 | 3.7 | 10.7 | <5 | 7 | 0.69 (0.03 - 7.65) | 0.79 | 0.70 (0.04 - 11.1) | 0.80 |
| MDCR | 689 | 2,105 | 0.4 | 1.5 | <5 | <5 | | | | |
| Open Claims | 143,821 | 329,537 | 87.9 | 243.6 | 24 | 55 | 1.22 (0.62 - 2.34) | 0.55 | 1.14 (0.58 - 2.22) | 0.71 |
| OEHR | 7,905 | 20,095 | 1.9 | 5.7 | <5 | <5 | 1.11 (0.21 - 5.53) | 0.90 | 1.01 (0.20 - 5.24) | 0.95 |
| VA | 1,167 | 11,033 | 0.6 | 5.7 | 0 | <10 | | | | |
| Meta-analysis | 143,821 | 329,537 | 87.9 | 243.6 | 24 | 55 | 1.22 (0.62 - 2.34) | 0.55 | 1.15 (0.59 - 2.23) | 0.69 |
| **PS-stratification / on-treatment TAR without escalation** | | | | | | | | | | |
| CCAE | 10,254 | 23,945 | 6.0 | 17.4 | 5 | 10 | 1.23 (0.38 - 3.58) | 0.72 | 1.12 (0.36 - 3.46) | 0.84 |
| ODOD | 7,688 | 17,348 | 3.9 | 10.7 | <5 | 7 | 0.55 (0.08 - 2.50) | 0.50 | 0.52 (0.09 - 2.94) | 0.46 |
| MDCR | 720 | 2,105 | 0.4 | 1.5 | <5 | <5 | 2.41 (0.11 - 25.6) | 0.53 | 1.94 (0.13 - 29.5) | 0.64 |
| Open Claims | 153,926 | 329,565 | 92.5 | 243.6 | 30 | 55 | 1.50 (0.92 - 2.40) | 0.10 | 1.42 (0.87 - 2.29) | 0.16 |
| OEHR | 8,610 | 20,107 | 2.1 | 5.7 | <5 | <5 | 1.11 (0.19 - 6.10) | 0.90 | 1.00 (0.18 - 5.62) | 0.96 |
| VA | 1,337 | 12,352 | 0.7 | 6.4 | 0 | <10 | | | | |
| Meta-analysis | 153,926 | 329,565 | 92.5 | 243.6 | 30 | 55 | 1.50 (0.92 - 2.40) | 0.10 | 1.40 (0.87 - 2.28) | 0.17 |

##### 1.1.25 Venous thromboembolism

**Supplementary Table 25:** Exposures, outcomes and risk estimates for Venous thromboembolism in GLP1RA (male) (T) vs SGLT2I (male) (C) new-user cohorts under four different propensity score (PS) adjustment / time-at-risk (TAR) designs across data sources. We report total patient exposure time in 1,000 patient-year (KYr) units, uncalibrated and calibrated hazard ratios (HRs) and their 95% confidence intervals (CIs). Greyed out entries did not pass study diagnostics and are included only for completeness.

| | Patients | | Exposure (KYr) | | Outcomes | | Uncalibrated | | Calibrated | |
|---|---|---|---|---|---|---|---|---|---|---|
| | T | C | T | C | T | C | HR (95% CI) | $p$ | HR (95% CI) | $p$ |
| **PS-matching / on-treatment TAR** | | | | | | | | | | |
| CCAE | 9,448 | 23,616 | 6.4 | 19.6 | 32 | 77 | 1.53 (0.83 - 2.79) | 0.17 | 1.43 (0.78 - 2.62) | 0.26 |
| ODOD | 6,912 | 16,956 | 4.0 | 11.8 | 21 | 81 | 0.47 (0.24 - 0.87) | 0.02 | 0.49 (0.26 - 0.93) | 0.03 |
| MDCR | 669 | 2,048 | 0.4 | 1.6 | 6 | 15 | 0.78 (0.17 - 2.75) | 0.73 | 0.60 (0.15 - 2.43) | 0.47 |
| Open Claims | 139,783 | 321,841 | 98.2 | 280.3 | 346 | 985 | 1.03 (0.87 - 1.23) | 0.71 | 0.98 (0.82 - 1.16) | 0.81 |
| OEHR | 7,745 | 19,734 | 2.2 | 6.3 | 15 | 42 | 0.99 (0.42 - 2.17) | 0.98 | 0.93 (0.41 - 2.12) | 0.86 |
| VA | 1,133 | 10,718 | 0.8 | 6.5 | <10 | 36 | 0.80 (0.21 - 2.39) | 0.71 | 0.54 (0.16 - 1.81) | 0.32 |
| Meta-analysis | 163,888 | 382,147 | 110.8 | 317.9 | 414 | 1,185 | 0.95 (0.61 - 1.47) | 0.81 | 0.90 (0.58 - 1.40) | 0.64 |
| Meta-analysis (w/o USOC) | 24,105 | 60,306 | 12.6 | 37.6 | 68 | 200 | 0.90 (0.43 - 1.86) | 0.77 | 0.85 (0.41 - 1.78) | 0.67 |
| **PS-stratification / on-treatment TAR** | | | | | | | | | | |
| CCAE | 10,036 | 23,622 | 6.7 | 19.6 | 35 | 77 | 1.38 (0.89 - 2.10) | 0.14 | 1.29 (0.84 - 1.97) | 0.25 |
| ODOD | 7,476 | 16,980 | 4.3 | 11.8 | 24 | 81 | 0.65 (0.39 - 1.04) | 0.08 | 0.63 (0.38 - 1.02) | 0.06 |
| MDCR | 697 | 2,048 | 0.4 | 1.6 | 6 | 15 | 1.25 (0.43 - 3.17) | 0.66 | 1.03 (0.38 - 2.79) | 0.91 |
| Open Claims | 149,433 | 321,867 | 102.9 | 280.3 | 369 | 985 | 0.99 (0.87 - 1.12) | 0.85 | 0.94 (0.83 - 1.07) | 0.43 |
| OEHR | 8,429 | 19,746 | 2.3 | 6.3 | 17 | 42 | 1.06 (0.57 - 1.89) | 0.85 | 0.95 (0.52 - 1.74) | 0.86 |
| VA | 1,292 | 11,951 | 0.9 | 7.2 | <10 | 44 | 1.34 (0.47 - 3.34) | 0.56 | 0.97 (0.36 - 2.60) | 0.91 |
| Meta-analysis | 175,374 | 382,215 | 116.2 | 318.0 | 445 | 1,185 | 0.99 (0.79 - 1.26) | 0.96 | 0.94 (0.74 - 1.20) | 0.65 |
| Meta-analysis (w/o USOC) | 25,941 | 60,348 | 13.3 | 37.7 | 76 | 200 | 0.99 (0.62 - 1.58) | 0.97 | 0.93 (0.58 - 1.48) | 0.76 |
| **PS-matching / on-treatment TAR without escalation** | | | | | | | | | | |
| CCAE | 9,448 | 23,616 | 5.6 | 17.2 | 29 | 64 | 1.92 (1.00 - 3.62) | 0.05 | 1.76 (0.93 - 3.35) | 0.09 |
| ODOD | 6,912 | 16,956 | 3.6 | 10.4 | 20 | 66 | 0.51 (0.25 - 0.96) | 0.05 | 0.52 (0.27 - 1.02) | 0.06 |
| MDCR | 669 | 2,048 | 0.4 | 1.4 | <5 | 15 | 0.57 (0.08 - 2.42) | 0.51 | 0.42 (0.08 - 2.30) | 0.32 |
| Open Claims | 139,783 | 321,841 | 85.6 | 238.4 | 302 | 805 | 1.11 (0.92 - 1.33) | 0.28 | 1.03 (0.85 - 1.24) | 0.77 |
| OEHR | 7,745 | 19,734 | 1.9 | 5.5 | 12 | 31 | 1.19 (0.48 - 2.77) | 0.69 | 1.09 (0.45 - 2.62) | 0.85 |
| VA | 1,133 | 10,718 | 0.6 | 5.6 | <10 | 29 | 1.33 (0.34 - 4.25) | 0.66 | 0.86 (0.24 - 3.03) | 0.81 |
| Meta-analysis | 163,888 | 382,147 | 96.6 | 271.5 | 363 | 966 | 1.07 (0.65 - 1.76) | 0.79 | 1.00 (0.61 - 1.65) | 0.97 |
| Meta-analysis (w/o USOC) | 24,105 | 60,306 | 11.1 | 33.1 | 61 | 161 | 1.05 (0.46 - 2.41) | 0.91 | 0.98 (0.42 - 2.25) | 0.95 |
| **PS-stratification / on-treatment TAR without escalation** | | | | | | | | | | |
| CCAE | 10,036 | 23,622 | 5.9 | 17.2 | 32 | 64 | 1.49 (0.94 - 2.32) | 0.09 | 1.36 (0.86 - 2.14) | 0.20 |
| ODOD | 7,476 | 16,980 | 3.8 | 10.4 | 22 | 66 | 0.68 (0.40 - 1.12) | 0.14 | 0.64 (0.38 - 1.07) | 0.09 |
| MDCR | 697 | 2,048 | 0.4 | 1.4 | <5 | 15 | 0.91 (0.25 - 2.63) | 0.87 | 0.73 (0.22 - 2.36) | 0.60 |
| Open Claims | 149,433 | 321,867 | 89.9 | 238.5 | 323 | 805 | 1.02 (0.89 - 1.16) | 0.81 | 0.96 (0.84 - 1.11) | 0.62 |
| OEHR | 8,429 | 19,746 | 2.0 | 5.5 | 14 | 31 | 1.33 (0.66 - 2.55) | 0.41 | 1.19 (0.61 - 2.34) | 0.61 |
| VA | 1,292 | 11,951 | 0.7 | 6.2 | <10 | 36 | 1.91 (0.63 - 5.00) | 0.22 | 1.38 (0.49 - 3.88) | 0.55 |
| Meta-analysis | 175,374 | 382,215 | 101.7 | 271.6 | 391 | 966 | 1.06 (0.80 - 1.40) | 0.68 | 1.00 (0.75 - 1.32) | 0.96 |
| Meta-analysis (w/o USOC) | 25,941 | 60,348 | 11.7 | 33.1 | 68 | 161 | 1.10 (0.66 - 1.83) | 0.72 | 1.01 (0.60 - 1.69) | 0.95 |

##### 1.1.26 Vomiting

**Supplementary Table 26:** Exposures, outcomes and risk estimates for Vomiting in GLP1RA (male) (T) vs SGLT2I (male) (C) new-user cohorts under four different propensity score (PS) adjustment / time-at-risk (TAR) designs across data sources. We report total patient exposure time in 1,000 patient-year (KYr) units, uncalibrated and calibrated hazard ratios (HRs) and their 95% confidence intervals (CIs). Greyed out entries did not pass study diagnostics and are included only for completeness.

| | Patients | | Exposure (KYr) | | Outcomes | | Uncalibrated | | Calibrated | |
|---|---|---|---|---|---|---|---|---|---|---|
| | T | C | T | C | T | C | HR (95% CI) | $p$ | HR (95% CI) | $p$ |
| **PS-matching / on-treatment TAR** | | | | | | | | | | |
| CCAE | 8,995 | 22,524 | 6.1 | 18.5 | 142 | 308 | 1.48 (1.13 - 1.92) | <0.01 | 1.37 (1.05 - 1.79) | 0.02 |
| ODOD | 6,553 | 16,119 | 3.7 | 11.1 | 102 | 212 | 1.43 (1.03 - 1.97) | 0.03 | 1.48 (1.07 - 2.05) | 0.02 |
| MDCR | 664 | 2,025 | 0.4 | 1.6 | 8 | 29 | 0.78 (0.26 - 2.08) | 0.63 | 0.59 (0.21 - 1.67) | 0.33 |
| Open Claims | 132,807 | 307,109 | 93.4 | 268.3 | 1,364 | 2,347 | 1.71 (1.56 - 1.87) | <0.01 | 1.62 (1.47 - 1.78) | <0.01 |
| OEHR | 7,513 | 19,267 | 2.1 | 6.1 | 82 | 99 | 1.96 (1.29 - 2.97) | <0.01 | 1.85 (1.22 - 2.80) | <0.01 |
| VA | 1,128 | 10,553 | 0.8 | 6.4 | 12 | 30 | 3.08 (1.10 - 8.13) | 0.03 | 2.09 (0.77 - 5.67) | 0.16 |
| Meta-analysis | 155,868 | 365,019 | 105.3 | 304.1 | 1,690 | 2,966 | 1.67 (1.54 - 1.82) | <0.01 | 1.59 (1.45 - 1.75) | <0.01 |
| Meta-analysis (w/o USOC) | 23,061 | 57,910 | 11.9 | 35.8 | 326 | 619 | 1.54 (1.28 - 1.85) | <0.01 | 1.47 (1.22 - 1.77) | <0.01 |
| **PS-stratification / on-treatment TAR** | | | | | | | | | | |
| CCAE | 9,558 | 22,530 | 6.3 | 18.5 | 151 | 308 | 1.23 (1.00 - 1.50) | 0.05 | 1.14 (0.93 - 1.40) | 0.24 |
| ODOD | 7,096 | 16,142 | 4.0 | 11.2 | 111 | 213 | 1.34 (1.04 - 1.71) | 0.02 | 1.29 (1.00 - 1.65) | 0.06 |
| MDCR | 691 | 2,025 | 0.4 | 1.6 | 8 | 29 | 1.02 (0.42 - 2.18) | 0.97 | 0.84 (0.37 - 1.90) | 0.68 |
| Open Claims | 141,776 | 307,189 | 97.8 | 268.3 | 1,490 | 2,351 | 1.61 (1.50 - 1.72) | <0.01 | 1.54 (1.43 - 1.65) | <0.01 |
| OEHR | 8,172 | 19,279 | 2.3 | 6.1 | 88 | 99 | 2.10 (1.54 - 2.85) | <0.01 | 1.88 (1.38 - 2.57) | <0.01 |
| VA | 1,295 | 11,772 | 0.9 | 7.1 | 14 | 40 | 3.16 (1.42 - 6.82) | <0.01 | 2.28 (1.04 - 5.01) | 0.04 |
| Meta-analysis | 166,602 | 365,140 | 110.3 | 304.1 | 1,840 | 2,971 | 1.52 (1.24 - 1.86) | <0.01 | 1.44 (1.17 - 1.78) | <0.01 |
| Meta-analysis (w/o USOC) | 24,826 | 57,951 | 12.6 | 35.8 | 350 | 620 | 1.48 (1.10 - 1.99) | <0.01 | 1.39 (1.03 - 1.88) | 0.04 |
| **PS-matching / on-treatment TAR without escalation** | | | | | | | | | | |
| CCAE | 8,995 | 22,524 | 5.3 | 16.2 | 131 | 255 | 1.56 (1.18 - 2.06) | <0.01 | 1.44 (1.09 - 1.90) | 0.01 |
| ODOD | 6,553 | 16,119 | 3.3 | 9.9 | 95 | 178 | 1.50 (1.07 - 2.11) | 0.02 | 1.54 (1.09 - 2.17) | 0.02 |
| MDCR | 664 | 2,025 | 0.4 | 1.4 | 5 | 27 | 0.58 (0.15 - 1.78) | 0.37 | 0.43 (0.13 - 1.46) | 0.18 |
| Open Claims | 132,807 | 307,109 | 81.6 | 228.8 | 1,178 | 1,863 | 1.82 (1.65 - 2.01) | <0.01 | 1.69 (1.52 - 1.88) | <0.01 |
| OEHR | 7,513 | 19,267 | 1.8 | 5.4 | 67 | 84 | 1.87 (1.19 - 2.95) | <0.01 | 1.70 (1.08 - 2.69) | 0.03 |
| VA | 1,128 | 10,553 | 0.6 | 5.5 | 11 | 22 | 5.68 (1.73 - 17.8) | <0.01 | 3.66 (1.14 - 11.8) | 0.03 |
| Meta-analysis | 155,868 | 365,019 | 92.1 | 260.3 | 1,471 | 2,380 | 1.76 (1.60 - 1.94) | <0.01 | 1.65 (1.49 - 1.83) | <0.01 |
| Meta-analysis (w/o USOC) | 23,061 | 57,910 | 10.5 | 31.5 | 293 | 517 | 1.59 (1.31 - 1.94) | <0.01 | 1.48 (1.21 - 1.82) | <0.01 |
| **PS-stratification / on-treatment TAR without escalation** | | | | | | | | | | |
| CCAE | 9,558 | 22,530 | 5.6 | 16.2 | 139 | 255 | 1.35 (1.08 - 1.68) | <0.01 | 1.23 (0.99 - 1.54) | 0.07 |
| ODOD | 7,096 | 16,142 | 3.6 | 9.9 | 101 | 179 | 1.42 (1.09 - 1.84) | <0.01 | 1.34 (1.02 - 1.74) | 0.04 |
| MDCR | 691 | 2,025 | 0.4 | 1.4 | 5 | 27 | 0.65 (0.22 - 1.60) | 0.40 | 0.52 (0.19 - 1.43) | 0.21 |
| Open Claims | 141,776 | 307,189 | 85.6 | 228.8 | 1,291 | 1,867 | 1.72 (1.60 - 1.86) | <0.01 | 1.63 (1.51 - 1.77) | <0.01 |
| OEHR | 8,172 | 19,279 | 2.0 | 5.4 | 73 | 84 | 1.99 (1.42 - 2.78) | <0.01 | 1.78 (1.27 - 2.50) | <0.01 |
| VA | 1,295 | 11,772 | 0.7 | 6.2 | 13 | 31 | 4.70 (1.92 - 11.2) | <0.01 | 3.39 (1.41 - 8.18) | <0.01 |
| Meta-analysis | 166,602 | 365,140 | 96.7 | 260.4 | 1,604 | 2,385 | 1.60 (1.38 - 1.86) | <0.01 | 1.50 (1.28 - 1.76) | <0.01 |
| Meta-analysis (w/o USOC) | 24,826 | 57,951 | 11.1 | 31.6 | 313 | 518 | 1.52 (1.23 - 1.87) | <0.01 | 1.40 (1.13 - 1.73) | <0.01 |

#### 1.2 SGLT2I (male) vs DPP4I (male)

##### 1.2.1 3-point MACE

**Supplementary Table 27:** Exposures, outcomes and risk estimates for 3-point MACE in SGLT2I (male) (T) vs DPP4I (male) (C) new-user cohorts under four different propensity score (PS) adjustment / time-at-risk (TAR) designs across data sources. We report total patient exposure time in 1,000 patient-year (KYr) units, uncalibrated and calibrated hazard ratios (HRs) and their 95% confidence intervals (CIs). Greyed out entries did not pass study diagnostics and are included only for completeness.

| | Patients | | Exposure (KYr) | | Outcomes | | Uncalibrated | | Calibrated | |
|---|---|---|---|---|---|---|---|---|---|---|
| | T | C | T | C | T | C | HR (95% CI) | $p$ | HR (95% CI) | $p$ |
| **PS-matching / on-treatment TAR** | | | | | | | | | | |
| CCAE | 19,046 | 30,069 | 17.0 | 25.3 | 127 | 233 | 0.59 (0.42 - 0.81) | <0.01 | 0.63 (0.46 - 0.88) | <0.01 |
| DAG | 4,640 | 11,168 | 4.1 | 11.6 | 0 | 0 | | | | |
| ODOD | 11,893 | 20,135 | 9.1 | 15.8 | 133 | 331 | 1.00 (0.76 - 1.32) | 0.99 | 1.03 (0.78 - 1.38) | 0.83 |
| MDCR | 1,609 | 4,821 | 1.4 | 4.5 | 28 | 125 | 0.69 (0.35 - 1.29) | 0.27 | 0.70 (0.36 - 1.34) | 0.29 |
| MDCD | 1,303 | 3,149 | 0.8 | 2.3 | 17 | 48 | 1.14 (0.54 - 2.30) | 0.72 | 1.03 (0.50 - 2.11) | 0.88 |
| Open Claims | 230,169 | 399,585 | 223.7 | 382.0 | 1,716 | 4,171 | 0.87 (0.80 - 0.94) | <0.01 | 0.95 (0.83 - 1.08) | 0.45 |
| OEHR | 16,401 | 31,503 | 5.3 | 16.2 | 43 | 172 | 0.85 (0.53 - 1.32) | 0.47 | 0.85 (0.54 - 1.34) | 0.50 |
| VA | 7,323 | 23,032 | 5.0 | 20.8 | 23 | 112 | 1.06 (0.56 - 1.93) | 0.86 | 1.06 (0.57 - 1.97) | 0.85 |
| SIDIAP | 4,064 | 19,143 | 5.9 | 43.3 | 26 | 237 | 0.78 (0.48 - 1.22) | 0.30 | 0.85 (0.53 - 1.36) | 0.53 |
| Meta-analysis | 290,505 | 528,288 | 267.4 | 508.0 | 2,096 | 5,381 | 0.84 (0.73 - 0.96) | 0.01 | 0.89 (0.75 - 1.05) | 0.17 |
| Meta-analysis (w/o USOC) | 60,336 | 128,703 | 43.7 | 126.0 | 380 | 1,210 | 0.81 (0.66 - 0.99) | 0.04 | 0.83 (0.68 - 1.02) | 0.09 |
| **PS-stratification / on-treatment TAR** | | | | | | | | | | |
| CCAE | 23,133 | 30,091 | 19.3 | 25.3 | 140 | 233 | 0.79 (0.63 - 0.98) | 0.04 | 0.82 (0.66 - 1.03) | 0.09 |
| DAG | 5,763 | 11,200 | 4.7 | 11.7 | 0 | 0 | | | | |
| ODOD | 16,422 | 20,238 | 11.4 | 15.9 | 170 | 339 | 0.89 (0.72 - 1.09) | 0.25 | 0.89 (0.73 - 1.10) | 0.32 |
| MDCR | 1,977 | 4,866 | 1.6 | 4.6 | 36 | 127 | 0.91 (0.59 - 1.36) | 0.65 | 0.94 (0.62 - 1.42) | 0.77 |
| MDCD | 1,365 | 3,169 | 0.8 | 2.3 | 17 | 50 | 1.17 (0.64 - 2.04) | 0.61 | 1.05 (0.59 - 1.89) | 0.85 |
| Open Claims | 308,612 | 399,821 | 270.1 | 382.2 | 2,093 | 4,185 | 0.89 (0.84 - 0.94) | <0.01 | 0.96 (0.84 - 1.11) | 0.61 |
| OEHR | 19,539 | 31,569 | 6.2 | 16.2 | 48 | 175 | 0.89 (0.63 - 1.25) | 0.52 | 0.89 (0.63 - 1.27) | 0.55 |
| VA | 11,375 | 24,034 | 6.9 | 22.3 | 50 | 122 | 1.07 (0.68 - 1.68) | 0.75 | 1.12 (0.72 - 1.77) | 0.61 |
| SIDIAP | 4,643 | 19,567 | 6.4 | 44.9 | 29 | 248 | 0.77 (0.50 - 1.15) | 0.22 | 0.79 (0.52 - 1.21) | 0.31 |
| Meta-analysis | 374,326 | 506,152 | 315.0 | 489.1 | 2,516 | 5,307 | 0.88 (0.84 - 0.93) | <0.01 | 0.93 (0.82 - 1.06) | 0.28 |
| Meta-analysis (w/o USOC) | 65,714 | 106,331 | 44.8 | 106.9 | 423 | 1,122 | 0.85 (0.75 - 0.96) | <0.01 | 0.86 (0.76 - 0.98) | 0.04 |
| **PS-matching / on-treatment TAR without escalation** | | | | | | | | | | |
| CCAE | 19,046 | 30,069 | 14.7 | 20.5 | 104 | 187 | 0.58 (0.40 - 0.83) | <0.01 | 0.62 (0.43 - 0.89) | 0.01 |
| DAG | 4,640 | 11,168 | 3.7 | 10.5 | 0 | 0 | | | | |
| ODOD | 11,893 | 20,135 | 8.0 | 12.9 | 116 | 279 | 0.90 (0.66 - 1.23) | 0.53 | 0.93 (0.66 - 1.32) | 0.74 |
| MDCR | 1,609 | 4,821 | 1.3 | 3.9 | 24 | 108 | 0.69 (0.33 - 1.34) | 0.30 | 0.70 (0.34 - 1.41) | 0.33 |
| MDCD | 1,303 | 3,149 | 0.7 | 1.7 | 15 | 38 | 1.05 (0.47 - 2.27) | 0.90 | 1.00 (0.46 - 2.21) | 0.88 |
| Open Claims | 230,169 | 399,585 | 187.6 | 296.5 | 1,438 | 3,248 | 0.87 (0.79 - 0.95) | <0.01 | 0.93 (0.81 - 1.07) | 0.35 |
| OEHR | 16,401 | 31,503 | 4.7 | 13.1 | 33 | 123 | 0.82 (0.47 - 1.38) | 0.47 | 0.82 (0.48 - 1.41) | 0.50 |
| VA | 7,323 | 23,032 | 4.2 | 17.0 | 20 | 78 | 1.31 (0.63 - 2.60) | 0.46 | 1.30 (0.64 - 2.64) | 0.47 |
| SIDIAP | 4,064 | 19,143 | 5.2 | 34.5 | 24 | 180 | 0.80 (0.46 - 1.33) | 0.40 | 0.94 (0.55 - 1.60) | 0.80 |
| Meta-analysis | 290,505 | 528,288 | 225.7 | 398.4 | 1,759 | 4,203 | 0.84 (0.76 - 0.93) | <0.01 | 0.89 (0.76 - 1.05) | 0.16 |
| Meta-analysis (w/o USOC) | 60,336 | 128,703 | 38.1 | 101.9 | 321 | 955 | 0.79 (0.65 - 0.97) | 0.02 | 0.81 (0.66 - 1.00) | 0.05 |
| **PS-stratification / on-treatment TAR without escalation** | | | | | | | | | | |
| CCAE | 23,133 | 30,091 | 16.8 | 20.5 | 116 | 187 | 0.77 (0.61 - 0.98) | 0.04 | 0.80 (0.63 - 1.03) | 0.09 |
| DAG | 5,763 | 11,200 | 4.3 | 10.6 | 0 | 0 | | | | |
| ODOD | 16,422 | 20,238 | 10.1 | 13.0 | 150 | 285 | 0.88 (0.71 - 1.10) | 0.27 | 0.88 (0.70 - 1.09) | 0.27 |
| MDCR | 1,977 | 4,866 | 1.4 | 3.9 | 32 | 109 | 0.87 (0.55 - 1.36) | 0.56 | 0.91 (0.58 - 1.43) | 0.69 |
| MDCD | 1,365 | 3,169 | 0.7 | 1.7 | 15 | 39 | 1.13 (0.59 - 2.07) | 0.70 | 1.04 (0.55 - 1.95) | 0.87 |
| Open Claims | 308,612 | 399,821 | 229.5 | 296.7 | 1,786 | 3,260 | 0.90 (0.84 - 0.96) | <0.01 | 0.97 (0.87 - 1.09) | 0.64 |
| OEHR | 19,539 | 31,569 | 5.5 | 13.1 | 38 | 124 | 0.85 (0.57 - 1.26) | 0.43 | 0.85 (0.57 - 1.26) | 0.42 |
| VA | 11,375 | 24,034 | 6.0 | 18.1 | 42 | 85 | 1.31 (0.78 - 2.20) | 0.30 | 1.36 (0.81 - 2.30) | 0.25 |
| SIDIAP | 4,643 | 19,567 | 5.7 | 35.7 | 27 | 189 | 0.85 (0.54 - 1.30) | 0.47 | 0.93 (0.60 - 1.45) | 0.77 |
| Meta-analysis | 374,326 | 506,152 | 269.0 | 382.9 | 2,149 | 4,154 | 0.89 (0.84 - 0.94) | <0.01 | 0.94 (0.85 - 1.03) | 0.18 |
| Meta-analysis (w/o USOC) | 65,714 | 106,331 | 39.5 | 86.2 | 363 | 894 | 0.84 (0.73 - 0.96) | 0.01 | 0.86 (0.74 - 0.99) | 0.05 |

##### 1.2.2 4-point MACE

**Supplementary Table 28:** Exposures, outcomes and risk estimates for 4-point MACE in SGLT2I (male) (T) vs DPP4I (male) (C) new-user cohorts under four different propensity score (PS) adjustment / time-at-risk (TAR) designs across data sources. We report total patient exposure time in 1,000 patient-year (KYr) units, uncalibrated and calibrated hazard ratios (HRs) and their 95% confidence intervals (CIs). Greyed out entries did not pass study diagnostics and are included only for completeness.

| | Patients | | Exposure (KYr) | | Outcomes | | Uncalibrated | | Calibrated | |
|---|---|---|---|---|---|---|---|---|---|---|
| | T | C | T | C | T | C | HR (95% CI) | $p$ | HR (95% CI) | $p$ |
| **PS-matching / on-treatment TAR** | | | | | | | | | | |
| CCAE | 18,788 | 29,650 | 16.8 | 24.9 | 183 | 325 | 0.74 (0.56 - 0.96) | 0.02 | 0.79 (0.61 - 1.04) | 0.10 |
| DAG | 4,640 | 11,168 | 4.1 | 11.6 | 0 | 0 | | | | |
| ODOD | 11,493 | 19,247 | 8.8 | 15.0 | 191 | 491 | 0.95 (0.75 - 1.19) | 0.65 | 0.98 (0.77 - 1.25) | 0.85 |
| MDCR | 1,531 | 4,540 | 1.4 | 4.3 | 43 | 196 | 0.69 (0.41 - 1.11) | 0.14 | 0.69 (0.42 - 1.13) | 0.16 |
| MDCD | 1,212 | 2,924 | 0.7 | 2.1 | 20 | 60 | 1.14 (0.56 - 2.23) | 0.70 | 1.03 (0.52 - 2.06) | 0.87 |
| Open Claims | 224,326 | 386,854 | 218.7 | 369.1 | 2,394 | 6,506 | 0.79 (0.73 - 0.84) | <0.01 | 0.86 (0.76 - 0.97) | 0.02 |
| OEHR | 16,132 | 30,890 | 5.3 | 15.7 | 77 | 308 | 1.05 (0.75 - 1.45) | 0.79 | 1.05 (0.76 - 1.47) | 0.76 |
| VA | 7,057 | 22,438 | 4.8 | 20.2 | 43 | 244 | 0.74 (0.48 - 1.11) | 0.15 | 0.74 (0.49 - 1.12) | 0.17 |
| SIDIAP | 4,038 | 19,055 | 5.8 | 43.1 | 31 | 296 | 0.67 (0.41 - 1.04) | 0.08 | 0.73 (0.46 - 1.16) | 0.22 |
| Meta-analysis | 283,365 | 512,674 | 261.5 | 492.3 | 2,962 | 8,366 | 0.80 (0.75 - 0.85) | <0.01 | 0.84 (0.75 - 0.95) | <0.01 |
| Meta-analysis (w/o USOC) | 59,039 | 125,820 | 42.8 | 123.2 | 568 | 1,860 | 0.83 (0.72 - 0.96) | 0.01 | 0.86 (0.74 - 0.99) | 0.05 |
| **PS-stratification / on-treatment TAR** | | | | | | | | | | |
| CCAE | 22,780 | 29,670 | 19.0 | 24.9 | 211 | 325 | 0.86 (0.71 - 1.03) | 0.10 | 0.89 (0.74 - 1.07) | 0.24 |
| DAG | 5,763 | 11,200 | 4.7 | 11.7 | 0 | 0 | | | | |
| ODOD | 15,713 | 19,318 | 10.9 | 15.1 | 257 | 500 | 0.87 (0.73 - 1.02) | 0.09 | 0.87 (0.74 - 1.04) | 0.15 |
| MDCR | 1,862 | 4,574 | 1.5 | 4.3 | 52 | 198 | 0.86 (0.60 - 1.20) | 0.38 | 0.88 (0.63 - 1.25) | 0.51 |
| MDCD | 1,263 | 2,941 | 0.7 | 2.1 | 20 | 63 | 1.17 (0.67 - 1.93) | 0.57 | 1.05 (0.62 - 1.79) | 0.84 |
| Open Claims | 297,226 | 387,040 | 262.0 | 369.2 | 2,981 | 6,531 | 0.82 (0.78 - 0.86) | <0.01 | 0.88 (0.77 - 1.01) | 0.08 |
| OEHR | 19,146 | 30,946 | 6.1 | 15.7 | 93 | 312 | 0.96 (0.74 - 1.23) | 0.76 | 0.96 (0.75 - 1.25) | 0.79 |
| VA | 10,518 | 23,414 | 6.4 | 21.6 | 86 | 260 | 0.71 (0.52 - 0.97) | 0.03 | 0.74 (0.54 - 1.02) | 0.07 |
| SIDIAP | 4,579 | 19,474 | 6.3 | 44.6 | 33 | 314 | 0.71 (0.47 - 1.03) | 0.09 | 0.73 (0.49 - 1.08) | 0.14 |
| Meta-analysis | 361,306 | 491,022 | 305.8 | 473.9 | 3,627 | 8,180 | 0.82 (0.79 - 0.86) | <0.01 | 0.87 (0.77 - 0.99) | 0.03 |
| Meta-analysis (w/o USOC) | 64,080 | 103,982 | 43.8 | 104.7 | 646 | 1,649 | 0.87 (0.78 - 0.96) | <0.01 | 0.88 (0.79 - 0.98) | 0.04 |
| **PS-matching / on-treatment TAR without escalation** | | | | | | | | | | |
| CCAE | 18,788 | 29,650 | 14.5 | 20.2 | 152 | 261 | 0.74 (0.54 - 0.99) | 0.04 | 0.78 (0.58 - 1.06) | 0.12 |
| DAG | 4,640 | 11,168 | 3.7 | 10.5 | 0 | 0 | | | | |
| ODOD | 11,493 | 19,247 | 7.7 | 12.3 | 162 | 412 | 0.83 (0.63 - 1.07) | 0.15 | 0.85 (0.63 - 1.16) | 0.40 |
| MDCR | 1,531 | 4,540 | 1.2 | 3.6 | 37 | 174 | 0.66 (0.37 - 1.11) | 0.13 | 0.66 (0.38 - 1.15) | 0.16 |
| MDCD | 1,212 | 2,924 | 0.6 | 1.6 | 17 | 46 | 1.11 (0.50 - 2.36) | 0.79 | 1.07 (0.49 - 2.32) | 0.85 |
| Open Claims | 224,326 | 386,854 | 183.6 | 286.7 | 1,970 | 5,063 | 0.77 (0.72 - 0.83) | <0.01 | 0.83 (0.73 - 0.95) | 0.01 |
| OEHR | 16,132 | 30,890 | 4.6 | 12.8 | 60 | 222 | 1.03 (0.70 - 1.48) | 0.90 | 1.03 (0.71 - 1.51) | 0.83 |
| VA | 7,057 | 22,438 | 4.1 | 16.5 | 36 | 179 | 0.83 (0.52 - 1.32) | 0.45 | 0.83 (0.52 - 1.33) | 0.44 |
| SIDIAP | 4,038 | 19,055 | 5.2 | 34.4 | 27 | 224 | 0.71 (0.42 - 1.16) | 0.19 | 0.84 (0.50 - 1.40) | 0.53 |
| Meta-analysis | 283,365 | 512,674 | 220.9 | 386.4 | 2,444 | 6,535 | 0.78 (0.73 - 0.84) | <0.01 | 0.83 (0.72 - 0.95) | <0.01 |
| Meta-analysis (w/o USOC) | 59,039 | 125,820 | 37.4 | 99.7 | 474 | 1,472 | 0.81 (0.69 - 0.94) | <0.01 | 0.83 (0.71 - 0.97) | 0.02 |
| **PS-stratification / on-treatment TAR without escalation** | | | | | | | | | | |
| CCAE | 22,780 | 29,670 | 16.6 | 20.2 | 176 | 261 | 0.84 (0.68 - 1.02) | 0.08 | 0.87 (0.71 - 1.07) | 0.19 |
| DAG | 5,763 | 11,200 | 4.3 | 10.6 | 0 | 0 | | | | |
| ODOD | 15,713 | 19,318 | 9.7 | 12.4 | 226 | 419 | 0.84 (0.70 - 1.01) | 0.07 | 0.84 (0.70 - 1.00) | 0.08 |
| MDCR | 1,862 | 4,574 | 1.3 | 3.7 | 46 | 175 | 0.79 (0.54 - 1.13) | 0.21 | 0.82 (0.57 - 1.19) | 0.32 |
| MDCD | 1,263 | 2,941 | 0.6 | 1.6 | 17 | 48 | 1.13 (0.62 - 1.98) | 0.67 | 1.04 (0.58 - 1.86) | 0.86 |
| Open Claims | 297,226 | 387,040 | 222.7 | 286.8 | 2,508 | 5,085 | 0.81 (0.77 - 0.85) | <0.01 | 0.88 (0.79 - 0.97) | 0.02 |
| OEHR | 19,146 | 30,946 | 5.4 | 12.8 | 75 | 223 | 0.96 (0.72 - 1.28) | 0.80 | 0.96 (0.71 - 1.28) | 0.77 |
| VA | 10,518 | 23,414 | 5.5 | 17.6 | 71 | 192 | 0.76 (0.53 - 1.07) | 0.12 | 0.79 (0.55 - 1.12) | 0.20 |
| SIDIAP | 4,579 | 19,474 | 5.6 | 35.5 | 29 | 239 | 0.74 (0.48 - 1.11) | 0.17 | 0.81 (0.53 - 1.24) | 0.38 |
| Meta-analysis | 361,306 | 491,022 | 261.3 | 371.4 | 3,060 | 6,402 | 0.82 (0.78 - 0.86) | <0.01 | 0.86 (0.79 - 0.94) | <0.01 |
| Meta-analysis (w/o USOC) | 64,080 | 103,982 | 38.6 | 84.5 | 552 | 1,317 | 0.85 (0.76 - 0.95) | <0.01 | 0.86 (0.77 - 0.97) | 0.03 |

##### 1.2.3 Acute myocardial infarction

**Supplementary Table 29:** Exposures, outcomes and risk estimates for Acute myocardial infarction in SGLT2I (male) (T) vs DPP4I (male) (C) new-user cohorts under four different propensity score (PS) adjustment / time-at-risk (TAR) designs across data sources. We report total patient exposure time in 1,000 patient-year (KYr) units, uncalibrated and calibrated hazard ratios (HRs) and their 95% confidence intervals (CIs). Greyed out entries did not pass study diagnostics and are included only for completeness.

| | Patients | | Exposure (KYr) | | Outcomes | | Uncalibrated | | Calibrated | |
|---|---|---|---|---|---|---|---|---|---|---|
| | T | C | T | C | T | C | HR (95% CI) | $p$ | HR (95% CI) | $p$ |
| **PS-matching / on-treatment TAR** | | | | | | | | | | |
| CCAE | 19,282 | 30,461 | 17.2 | 25.7 | 75 | 137 | 0.49 (0.30 - 0.75) | <0.01 | 0.52 (0.33 - 0.82) | <0.01 |
| DAG | 4,640 | 11,168 | 4.1 | 11.6 | 0 | 0 | | | | |
| ODOD | 12,151 | 20,752 | 9.3 | 16.3 | 73 | 171 | 1.02 (0.70 - 1.48) | 0.92 | 1.05 (0.72 - 1.54) | 0.81 |
| MDCR | 1,652 | 5,026 | 1.5 | 4.8 | 9 | 64 | 0.55 (0.19 - 1.33) | 0.22 | 0.55 (0.21 - 1.44) | 0.23 |
| MDCD | 1,349 | 3,285 | 0.8 | 2.4 | 12 | 28 | 1.47 (0.59 - 3.71) | 0.41 | 1.33 (0.53 - 3.35) | 0.55 |
| Open Claims | 235,510 | 410,866 | 228.5 | 393.0 | 958 | 2,165 | 0.86 (0.77 - 0.96) | <0.01 | 0.94 (0.80 - 1.09) | 0.43 |
| OEHR | 16,549 | 31,819 | 5.4 | 16.4 | 22 | 99 | 0.63 (0.34 - 1.14) | 0.15 | 0.64 (0.35 - 1.18) | 0.16 |
| VA | 7,456 | 23,381 | 5.1 | 21.1 | 14 | 61 | 0.93 (0.39 - 2.06) | 0.86 | 0.93 (0.40 - 2.15) | 0.87 |
| SIDIAP | 4,136 | 19,402 | 6.0 | 44.0 | 14 | 124 | 0.81 (0.40 - 1.50) | 0.52 | 0.88 (0.45 - 1.70) | 0.72 |
| Meta-analysis | 296,736 | 541,707 | 272.9 | 521.3 | 1,165 | 2,821 | 0.77 (0.62 - 0.96) | 0.02 | 0.82 (0.64 - 1.04) | 0.11 |
| Meta-analysis (w/o USOC) | 61,226 | 130,841 | 44.4 | 128.3 | 207 | 656 | 0.72 (0.54 - 0.96) | 0.03 | 0.75 (0.56 - 1.00) | 0.05 |
| **PS-stratification / on-treatment TAR** | | | | | | | | | | |
| CCAE | 23,425 | 30,485 | 19.5 | 25.7 | 82 | 137 | 0.79 (0.59 - 1.05) | 0.10 | 0.82 (0.61 - 1.09) | 0.18 |
| DAG | 5,763 | 11,200 | 4.7 | 11.7 | 0 | 0 | | | | |
| ODOD | 16,779 | 20,863 | 11.7 | 16.4 | 101 | 174 | 0.97 (0.73 - 1.28) | 0.83 | 0.98 (0.74 - 1.29) | 0.88 |
| MDCR | 2,032 | 5,078 | 1.6 | 4.8 | 15 | 66 | 0.61 (0.31 - 1.13) | 0.13 | 0.63 (0.33 - 1.20) | 0.17 |
| MDCD | 1,420 | 3,306 | 0.8 | 2.4 | 12 | 29 | 1.42 (0.67 - 2.83) | 0.34 | 1.28 (0.63 - 2.63) | 0.50 |
| Open Claims | 316,207 | 411,152 | 276.1 | 393.2 | 1,187 | 2,173 | 0.94 (0.87 - 1.02) | 0.13 | 1.02 (0.88 - 1.18) | 0.83 |
| OEHR | 19,723 | 31,887 | 6.3 | 16.4 | 26 | 99 | 0.73 (0.45 - 1.15) | 0.19 | 0.73 (0.46 - 1.17) | 0.20 |
| VA | 11,651 | 24,407 | 7.0 | 22.7 | 35 | 63 | 1.30 (0.72 - 2.33) | 0.38 | 1.36 (0.75 - 2.45) | 0.31 |
| SIDIAP | 4,738 | 19,837 | 6.5 | 45.6 | 16 | 127 | 0.74 (0.41 - 1.27) | 0.31 | 0.76 (0.43 - 1.35) | 0.37 |
| Meta-analysis | 382,904 | 519,302 | 321.7 | 502.2 | 1,427 | 2,776 | 0.90 (0.81 - 1.00) | 0.04 | 0.94 (0.80 - 1.11) | 0.48 |
| Meta-analysis (w/o USOC) | 66,697 | 108,150 | 45.6 | 109.0 | 240 | 603 | 0.82 (0.70 - 0.97) | 0.02 | 0.84 (0.71 - 1.00) | 0.06 |
| **PS-matching / on-treatment TAR without escalation** | | | | | | | | | | |
| CCAE | 19,282 | 30,461 | 14.9 | 20.8 | 63 | 110 | 0.47 (0.28 - 0.77) | <0.01 | 0.50 (0.30 - 0.83) | <0.01 |
| DAG | 4,640 | 11,168 | 3.7 | 10.5 | 0 | 0 | | | | |
| ODOD | 12,151 | 20,752 | 8.1 | 13.4 | 65 | 138 | 0.89 (0.58 - 1.36) | 0.61 | 0.92 (0.59 - 1.45) | 0.75 |
| MDCR | 1,652 | 5,026 | 1.3 | 4.1 | 8 | 59 | 0.53 (0.17 - 1.38) | 0.24 | 0.54 (0.19 - 1.53) | 0.25 |
| MDCD | 1,349 | 3,285 | 0.7 | 1.8 | 12 | 26 | 1.20 (0.44 - 3.23) | 0.72 | 1.15 (0.42 - 3.12) | 0.79 |
| Open Claims | 235,510 | 410,866 | 191.6 | 305.0 | 791 | 1,667 | 0.85 (0.75 - 0.96) | <0.01 | 0.91 (0.77 - 1.07) | 0.28 |
| OEHR | 16,549 | 31,819 | 4.7 | 13.3 | 21 | 67 | 0.75 (0.37 - 1.43) | 0.40 | 0.75 (0.38 - 1.48) | 0.42 |
| VA | 7,456 | 23,381 | 4.3 | 17.2 | 13 | 46 | 1.10 (0.42 - 2.64) | 0.84 | 1.09 (0.44 - 2.73) | 0.85 |
| SIDIAP | 4,136 | 19,402 | 5.3 | 35.0 | 13 | 100 | 0.86 (0.39 - 1.71) | 0.68 | 1.01 (0.48 - 2.11) | 0.89 |
| Meta-analysis | 296,736 | 541,707 | 230.3 | 408.7 | 974 | 2,187 | 0.80 (0.67 - 0.94) | <0.01 | 0.84 (0.68 - 1.04) | 0.11 |
| Meta-analysis (w/o USOC) | 61,226 | 130,841 | 38.7 | 103.7 | 183 | 520 | 0.73 (0.56 - 0.94) | 0.02 | 0.75 (0.58 - 0.97) | 0.03 |
| **PS-stratification / on-treatment TAR without escalation** | | | | | | | | | | |
| CCAE | 23,425 | 30,485 | 17.0 | 20.8 | 70 | 110 | 0.79 (0.58 - 1.08) | 0.15 | 0.82 (0.60 - 1.13) | 0.24 |
| DAG | 5,763 | 11,200 | 4.3 | 10.6 | 0 | 0 | | | | |
| ODOD | 16,779 | 20,863 | 10.3 | 13.5 | 90 | 139 | 1.02 (0.75 - 1.37) | 0.91 | 1.01 (0.75 - 1.36) | 0.91 |
| MDCR | 2,032 | 5,078 | 1.5 | 4.1 | 14 | 60 | 0.61 (0.31 - 1.14) | 0.14 | 0.63 (0.33 - 1.22) | 0.18 |
| MDCD | 1,420 | 3,306 | 0.7 | 1.8 | 12 | 26 | 1.35 (0.64 - 2.71) | 0.42 | 1.23 (0.60 - 2.55) | 0.58 |
| Open Claims | 316,207 | 411,152 | 234.6 | 305.1 | 1,000 | 1,674 | 0.96 (0.88 - 1.04) | 0.31 | 1.03 (0.91 - 1.17) | 0.62 |
| OEHR | 19,723 | 31,887 | 5.5 | 13.3 | 25 | 67 | 0.86 (0.51 - 1.41) | 0.57 | 0.86 (0.52 - 1.42) | 0.56 |
| VA | 11,651 | 24,407 | 6.1 | 18.4 | 31 | 48 | 1.72 (0.89 - 3.31) | 0.11 | 1.79 (0.92 - 3.46) | 0.09 |
| SIDIAP | 4,738 | 19,837 | 5.8 | 36.2 | 15 | 101 | 0.81 (0.43 - 1.41) | 0.48 | 0.88 (0.49 - 1.60) | 0.70 |
| Meta-analysis | 382,904 | 519,302 | 274.8 | 393.0 | 1,214 | 2,151 | 0.94 (0.87 - 1.01) | 0.11 | 0.99 (0.89 - 1.10) | 0.86 |
| Meta-analysis (w/o USOC) | 66,697 | 108,150 | 40.2 | 87.9 | 214 | 477 | 0.86 (0.72 - 1.03) | 0.11 | 0.88 (0.73 - 1.06) | 0.20 |

##### 1.2.4 Acute renal failure

**Supplementary Table 30:** Exposures, outcomes and risk estimates for Acute renal failure in SGLT2I (male) (T) vs DPP4I (male) (C) new-user cohorts under four different propensity score (PS) adjustment / time-at-risk (TAR) designs across data sources. We report total patient exposure time in 1,000 patient-year (KYr) units, uncalibrated and calibrated hazard ratios (HRs) and their 95% confidence intervals (CIs). Greyed out entries did not pass study diagnostics and are included only for completeness.

| | Patients | | Exposure (KYr) | | Outcomes | | Uncalibrated | | Calibrated | |
|---|---|---|---|---|---|---|---|---|---|---|
| | T | C | T | C | T | C | HR (95% CI) | $p$ | HR (95% CI) | $p$ |
| **PS-matching / on-treatment TAR** | | | | | | | | | | |
| CCAE | 19,381 | 30,494 | 17.3 | 25.7 | 111 | 260 | 0.61 (0.44 - 0.84) | <0.01 | 0.66 (0.47 - 0.91) | 0.01 |
| DAG | 4,640 | 11,168 | 4.1 | 11.6 | 0 | 0 | | | | |
| ODOD | 11,987 | 20,055 | 9.2 | 15.7 | 152 | 513 | 0.67 (0.52 - 0.86) | <0.01 | 0.69 (0.54 - 0.90) | 0.02 |
| MDCR | 1,627 | 4,845 | 1.5 | 4.5 | 29 | 172 | 0.80 (0.46 - 1.34) | 0.41 | 0.80 (0.47 - 1.37) | 0.44 |
| MDCD | 1,337 | 3,140 | 0.8 | 2.3 | 17 | 81 | 0.77 (0.39 - 1.43) | 0.42 | 0.69 (0.36 - 1.33) | 0.29 |
| Open Claims | 236,488 | 408,696 | 230.3 | 390.0 | 1,787 | 5,910 | 0.69 (0.64 - 0.74) | <0.01 | 0.75 (0.66 - 0.85) | <0.01 |
| OEHR | 16,609 | 31,733 | 5.4 | 16.2 | 58 | 292 | 0.92 (0.63 - 1.31) | 0.64 | 0.92 (0.64 - 1.33) | 0.68 |
| VA | 7,454 | 23,043 | 5.1 | 20.8 | 53 | 265 | 0.74 (0.50 - 1.08) | 0.13 | 0.74 (0.50 - 1.09) | 0.15 |
| SIDIAP | 4,334 | 19,811 | 6.3 | 45.1 | <5 | 19 | 1.35 (0.19 - 5.77) | 0.73 | 1.47 (0.27 - 8.00) | 0.66 |
| Meta-analysis | 293,546 | 518,866 | 268.8 | 472.9 | 2,190 | 7,412 | 0.69 (0.65 - 0.74) | <0.01 | 0.73 (0.65 - 0.83) | <0.01 |
| Meta-analysis (w/o USOC) | 57,058 | 110,170 | 38.5 | 82.9 | 403 | 1,502 | 0.71 (0.61 - 0.83) | <0.01 | 0.74 (0.63 - 0.86) | <0.01 |
| **PS-stratification / on-treatment TAR** | | | | | | | | | | |
| CCAE | 23,699 | 30,511 | 19.7 | 25.7 | 127 | 261 | 0.62 (0.49 - 0.77) | <0.01 | 0.64 (0.51 - 0.80) | <0.01 |
| DAG | 5,763 | 11,200 | 4.7 | 11.7 | 0 | 0 | | | | |
| ODOD | 16,747 | 20,115 | 11.7 | 15.7 | 199 | 521 | 0.62 (0.51 - 0.73) | <0.01 | 0.62 (0.52 - 0.74) | <0.01 |
| MDCR | 2,014 | 4,884 | 1.6 | 4.6 | 36 | 177 | 0.70 (0.47 - 1.03) | 0.08 | 0.72 (0.49 - 1.08) | 0.13 |
| MDCD | 1,409 | 3,155 | 0.9 | 2.3 | 17 | 83 | 0.62 (0.35 - 1.04) | 0.09 | 0.56 (0.32 - 0.97) | 0.04 |
| Open Claims | 320,733 | 408,870 | 280.5 | 390.1 | 2,329 | 5,925 | 0.66 (0.62 - 0.69) | <0.01 | 0.71 (0.62 - 0.82) | <0.01 |
| OEHR | 19,913 | 31,795 | 6.4 | 16.3 | 66 | 300 | 0.72 (0.54 - 0.96) | 0.03 | 0.73 (0.54 - 0.97) | 0.04 |
| VA | 11,731 | 24,015 | 7.1 | 22.2 | 93 | 284 | 0.72 (0.53 - 0.98) | 0.04 | 0.76 (0.56 - 1.02) | 0.08 |
| SIDIAP | 5,159 | 20,247 | 7.1 | 46.7 | <5 | 21 | 1.57 (0.35 - 5.04) | 0.50 | 1.62 (0.43 - 6.12) | 0.48 |
| Meta-analysis | 388,265 | 516,422 | 326.9 | 499.1 | <2,762 | 7,205 | 0.66 (0.63 - 0.69) | <0.01 | 0.69 (0.61 - 0.79) | <0.01 |
| Meta-analysis (w/o USOC) | 67,532 | 107,552 | 46.4 | 108.9 | <433 | 1,280 | 0.65 (0.57 - 0.73) | <0.01 | 0.66 (0.58 - 0.74) | <0.01 |
| **PS-matching / on-treatment TAR without escalation** | | | | | | | | | | |
| CCAE | 19,381 | 30,494 | 15.0 | 20.8 | 89 | 214 | 0.62 (0.43 - 0.88) | <0.01 | 0.66 (0.46 - 0.94) | 0.02 |
| DAG | 4,640 | 11,168 | 3.7 | 10.5 | 0 | 0 | | | | |
| ODOD | 11,987 | 20,055 | 8.1 | 12.8 | 124 | 419 | 0.61 (0.46 - 0.80) | <0.01 | 0.63 (0.46 - 0.87) | 0.02 |
| MDCR | 1,627 | 4,845 | 1.3 | 3.8 | 25 | 146 | 0.79 (0.43 - 1.38) | 0.43 | 0.80 (0.45 - 1.43) | 0.48 |
| MDCD | 1,337 | 3,140 | 0.7 | 1.7 | 16 | 64 | 0.79 (0.37 - 1.62) | 0.54 | 0.76 (0.36 - 1.60) | 0.48 |
| Open Claims | 236,488 | 408,696 | 193.3 | 302.9 | 1,420 | 4,596 | 0.66 (0.60 - 0.71) | <0.01 | 0.71 (0.62 - 0.81) | <0.01 |
| OEHR | 16,609 | 31,733 | 4.8 | 13.2 | 40 | 206 | 0.71 (0.46 - 1.08) | 0.12 | 0.72 (0.47 - 1.10) | 0.14 |
| VA | 7,454 | 23,043 | 4.3 | 16.9 | 43 | 187 | 0.82 (0.52 - 1.26) | 0.39 | 0.82 (0.53 - 1.27) | 0.38 |
| SIDIAP | 4,334 | 19,811 | 5.6 | 35.8 | <5 | 13 | 1.39 (0.20 - 6.17) | 0.70 | 1.64 (0.29 - 9.12) | 0.58 |
| Meta-analysis | 293,546 | 518,866 | 226.8 | 370.6 | 1,741 | 5,768 | 0.66 (0.61 - 0.71) | <0.01 | 0.70 (0.61 - 0.80) | <0.01 |
| Meta-analysis (w/o USOC) | 57,058 | 110,170 | 33.6 | 67.7 | 321 | 1,172 | 0.67 (0.57 - 0.80) | <0.01 | 0.69 (0.58 - 0.82) | <0.01 |
| **PS-stratification / on-treatment TAR without escalation** | | | | | | | | | | |
| CCAE | 23,699 | 30,511 | 17.3 | 20.8 | 102 | 215 | 0.56 (0.44 - 0.72) | <0.01 | 0.59 (0.46 - 0.75) | <0.01 |
| DAG | 5,763 | 11,200 | 4.3 | 10.6 | 0 | 0 | | | | |
| ODOD | 16,747 | 20,115 | 10.4 | 12.9 | 167 | 427 | 0.60 (0.49 - 0.73) | <0.01 | 0.59 (0.49 - 0.72) | <0.01 |
| MDCR | 2,014 | 4,884 | 1.5 | 3.9 | 32 | 150 | 0.73 (0.47 - 1.11) | 0.15 | 0.76 (0.50 - 1.17) | 0.23 |
| MDCD | 1,409 | 3,155 | 0.7 | 1.7 | 16 | 65 | 0.66 (0.36 - 1.13) | 0.15 | 0.60 (0.34 - 1.07) | 0.09 |
| Open Claims | 320,733 | 408,870 | 238.7 | 303.0 | 1,920 | 4,609 | 0.63 (0.60 - 0.67) | <0.01 | 0.69 (0.62 - 0.76) | <0.01 |
| OEHR | 19,913 | 31,795 | 5.6 | 13.2 | 46 | 211 | 0.61 (0.43 - 0.85) | <0.01 | 0.61 (0.43 - 0.86) | <0.01 |
| VA | 11,731 | 24,015 | 6.2 | 18.0 | 80 | 201 | 0.84 (0.60 - 1.19) | 0.33 | 0.88 (0.62 - 1.24) | 0.47 |
| SIDIAP | 5,159 | 20,247 | 6.3 | 37.0 | <5 | 13 | 1.64 (0.24 - 6.56) | 0.56 | 1.79 (0.34 - 9.32) | 0.49 |
| Meta-analysis | 383,106 | 496,175 | 273.3 | 353.9 | 2,267 | 5,612 | 0.63 (0.60 - 0.66) | <0.01 | 0.66 (0.61 - 0.73) | <0.01 |
| Meta-analysis (w/o USOC) | 62,373 | 87,305 | 34.7 | 50.8 | 347 | 1,003 | 0.60 (0.53 - 0.69) | <0.01 | 0.61 (0.53 - 0.70) | <0.01 |

##### 1.2.5 Glycemic control

**Supplementary Table 31:** Exposures, outcomes and risk estimates for Glycemic control in SGLT2I (male) (T) vs DPP4I (male) (C) new-user cohorts under four different propensity score (PS) adjustment / time-at-risk (TAR) designs across data sources. We report total patient exposure time in 1,000 patient-year (KYr) units, uncalibrated and calibrated hazard ratios (HRs) and their 95% confidence intervals (CIs). Greyed out entries did not pass study diagnostics and are included only for completeness.

| | Patients | | Exposure (KYr) | | Outcomes | | Uncalibrated | | Calibrated | |
|---|---|---|---|---|---|---|---|---|---|---|
| | T | C | T | C | T | C | HR (95% CI) | $p$ | HR (95% CI) | $p$ |
| **PS-matching / on-treatment TAR** | | | | | | | | | | |
| CCAE | 19,610 | 30,810 | 17.5 | 26.1 | 26 | 40 | 1.96 (0.96 - 4.04) | 0.07 | 2.11 (1.03 - 4.35) | 0.04 |
| DAG | 1,654 | 4,152 | 1.2 | 3.7 | 180 | 560 | 0.91 (0.73 - 1.12) | 0.38 | 0.89 (0.71 - 1.10) | 0.36 |
| ODOD | 8,133 | 13,765 | 5.6 | 9.6 | 611 | 1,079 | 1.03 (0.89 - 1.19) | 0.69 | 1.06 (0.91 - 1.25) | 0.57 |
| MDCR | 1,714 | 5,229 | 1.5 | 5.0 | 0 | 5 | | | | |
| MDCD | 1,426 | 3,470 | 0.9 | 2.6 | 0 | 0 | | | | |
| Open Claims | 244,206 | 426,386 | 237.2 | 409.0 | 0 | 0 | | | | |
| OEHR | 16,869 | 32,344 | 5.5 | 16.7 | 0 | 0 | | | | |
| VA | 7,441 | 23,209 | 5.1 | 21.0 | <10 | 21 | 0.93 (0.20 - 3.36) | 0.92 | 0.93 (0.23 - 3.86) | 0.92 |
| SIDIAP | 496 | 2,248 | 0.5 | 3.2 | 162 | 832 | 1.21 (0.98 - 1.48) | 0.08 | 1.31 (1.05 - 1.64) | 0.04 |
| Meta-analysis | 37,334 | 74,184 | 29.9 | 63.6 | <989 | 2,532 | 1.07 (0.92 - 1.23) | 0.40 | 1.13 (0.95 - 1.35) | 0.18 |
| Meta-analysis (w/o USOC) | 37,334 | 74,184 | 29.9 | 63.6 | <989 | 2,532 | 1.07 (0.91 - 1.25) | 0.40 | 1.10 (0.94 - 1.29) | 0.25 |
| **PS-stratification / on-treatment TAR** | | | | | | | | | | |
| CCAE | 23,976 | 30,829 | 20.0 | 26.1 | 26 | 40 | 1.48 (0.87 - 2.49) | 0.15 | 1.54 (0.91 - 2.60) | 0.11 |
| DAG | 1,994 | 4,184 | 1.4 | 3.7 | 210 | 560 | 0.90 (0.75 - 1.07) | 0.22 | 0.86 (0.72 - 1.03) | 0.16 |
| ODOD | 11,471 | 13,903 | 7.3 | 9.7 | 761 | 1,088 | 1.05 (0.95 - 1.16) | 0.38 | 1.06 (0.95 - 1.17) | 0.40 |
| MDCR | 2,128 | 5,285 | 1.7 | 5.0 | 0 | 5 | | | | |
| MDCD | 1,507 | 3,484 | 0.9 | 2.6 | 0 | 0 | | | | |
| Open Claims | 332,801 | 426,715 | 289.7 | 409.2 | 0 | 0 | | | | |
| OEHR | 20,231 | 32,415 | 6.5 | 16.8 | 0 | 0 | | | | |
| VA | 11,946 | 24,227 | 7.2 | 22.5 | 12 | 21 | 1.47 (0.61 - 3.44) | 0.39 | 1.54 (0.65 - 3.66) | 0.33 |
| SIDIAP | 541 | 2,295 | 0.5 | 3.3 | 180 | 857 | 1.27 (1.06 - 1.51) | <0.01 | 1.31 (1.09 - 1.56) | <0.01 |
| Meta-analysis | 37,982 | 51,211 | 29.2 | 42.8 | 1,177 | 2,545 | 1.09 (0.91 - 1.30) | 0.34 | 1.15 (0.93 - 1.42) | 0.20 |
| Meta-analysis (w/o USOC) | 37,982 | 51,211 | 29.2 | 42.8 | 1,177 | 2,545 | 1.09 (0.92 - 1.28) | 0.32 | 1.11 (0.94 - 1.31) | 0.25 |
| **PS-matching / on-treatment TAR without escalation** | | | | | | | | | | |
| CCAE | 19,610 | 30,810 | 15.2 | 21.1 | 25 | 33 | 2.11 (0.99 - 4.52) | 0.05 | 2.24 (1.05 - 4.77) | 0.04 |
| DAG | 1,654 | 4,152 | 1.1 | 3.3 | 157 | 507 | 0.91 (0.72 - 1.14) | 0.41 | 0.88 (0.70 - 1.11) | 0.36 |
| ODOD | 8,133 | 13,765 | 5.0 | 7.9 | 532 | 871 | 1.02 (0.87 - 1.20) | 0.78 | 1.06 (0.84 - 1.32) | 0.72 |
| MDCR | 1,714 | 5,229 | 1.4 | 4.2 | 0 | 5 | | | | |
| MDCD | 1,426 | 3,470 | 0.7 | 1.9 | 0 | 0 | | | | |
| Open Claims | 244,206 | 426,386 | 198.9 | 317.1 | 0 | 0 | | | | |
| OEHR | 16,869 | 32,344 | 4.8 | 13.5 | 0 | 0 | | | | |
| VA | 7,441 | 23,209 | 4.3 | 17.2 | <10 | 18 | 1.63 (0.32 - 6.91) | 0.53 | 1.62 (0.35 - 7.51) | 0.54 |
| SIDIAP | 496 | 2,248 | 0.4 | 2.5 | 150 | 656 | 1.26 (1.00 - 1.57) | 0.05 | 1.47 (1.17 - 1.86) | <0.01 |
| Meta-analysis | 37,334 | 74,184 | 26.0 | 51.9 | <874 | 2,085 | 1.10 (0.91 - 1.32) | 0.32 | 1.16 (0.93 - 1.45) | 0.19 |
| Meta-analysis (w/o USOC) | 37,334 | 74,184 | 26.0 | 51.9 | <874 | 2,085 | 1.10 (0.91 - 1.32) | 0.32 | 1.13 (0.94 - 1.36) | 0.21 |
| **PS-stratification / on-treatment TAR without escalation** | | | | | | | | | | |
| CCAE | 23,976 | 30,829 | 17.4 | 21.1 | 25 | 33 | 1.59 (0.91 - 2.75) | 0.10 | 1.65 (0.95 - 2.88) | 0.08 |
| DAG | 1,994 | 4,184 | 1.3 | 3.3 | 185 | 507 | 0.87 (0.72 - 1.05) | 0.15 | 0.84 (0.70 - 1.02) | 0.13 |
| ODOD | 11,471 | 13,903 | 6.5 | 8.0 | 669 | 878 | 1.08 (0.96 - 1.20) | 0.20 | 1.07 (0.95 - 1.19) | 0.36 |
| MDCR | 2,128 | 5,285 | 1.5 | 4.3 | 0 | 5 | | | | |
| MDCD | 1,507 | 3,484 | 0.8 | 1.9 | 0 | 0 | | | | |
| Open Claims | 332,801 | 426,715 | 246.2 | 317.3 | 0 | 0 | | | | |
| OEHR | 20,231 | 32,415 | 5.7 | 13.6 | 0 | 0 | | | | |
| VA | 11,946 | 24,227 | 6.2 | 18.3 | <10 | 18 | 1.33 (0.49 - 3.41) | 0.57 | 1.38 (0.52 - 3.63) | 0.52 |
| SIDIAP | 541 | 2,295 | 0.4 | 2.5 | 165 | 677 | 1.37 (1.14 - 1.65) | <0.01 | 1.51 (1.24 - 1.83) | <0.01 |
| Meta-analysis | 37,982 | 51,211 | 25.7 | 34.9 | 1,044 | 2,095 | 1.14 (0.90 - 1.44) | 0.28 | 1.20 (0.94 - 1.53) | 0.14 |
| Meta-analysis (w/o USOC) | 37,982 | 51,211 | 25.7 | 34.9 | 1,044 | 2,095 | 1.13 (0.92 - 1.40) | 0.25 | 1.15 (0.93 - 1.43) | 0.20 |

##### 1.2.6 Hospitalization with heart failure

**Supplementary Table 32:** Exposures, outcomes and risk estimates for Hospitalization with heart failure in SGLT2I (male) (T) vs DPP4I (male) (C) new-user cohorts under four different propensity score (PS) adjustment / time-at-risk (TAR) designs across data sources. We report total patient exposure time in 1,000 patient-year (KYr) units, uncalibrated and calibrated hazard ratios (HRs) and their 95% confidence intervals (CIs). Greyed out entries did not pass study diagnostics and are included only for completeness.

| | Patients | | Exposure (KYr) | | Outcomes | | Uncalibrated | | Calibrated | |
|---|---|---|---|---|---|---|---|---|---|---|
| | T | C | T | C | T | C | HR (95% CI) | $p$ | HR (95% CI) | $p$ |
| **PS-matching / on-treatment TAR** | | | | | | | | | | |
| CCAE | 19,363 | 30,576 | 17.3 | 25.8 | 96 | 151 | 0.89 (0.60 - 1.30) | 0.54 | 0.95 (0.65 - 1.41) | 0.82 |
| DAG | 4,640 | 11,168 | 4.1 | 11.6 | 0 | 0 | | | | |
| ODOD | 11,932 | 20,118 | 9.2 | 15.8 | 117 | 344 | 0.79 (0.58 - 1.05) | 0.11 | 0.81 (0.60 - 1.10) | 0.23 |
| MDCR | 1,605 | 4,825 | 1.4 | 4.6 | 28 | 137 | 0.74 (0.42 - 1.28) | 0.30 | 0.75 (0.43 - 1.32) | 0.33 |
| MDCD | 1,287 | 3,127 | 0.8 | 2.3 | 11 | 41 | 1.18 (0.51 - 2.61) | 0.68 | 1.07 (0.47 - 2.42) | 0.86 |
| Open Claims | 235,537 | 407,798 | 229.8 | 390.2 | 1,198 | 4,114 | 0.66 (0.60 - 0.72) | <0.01 | 0.71 (0.62 - 0.82) | <0.01 |
| OEHR | 16,513 | 31,579 | 5.4 | 16.1 | 50 | 202 | 1.14 (0.74 - 1.73) | 0.54 | 1.15 (0.75 - 1.75) | 0.53 |
| VA | 7,403 | 23,147 | 5.0 | 20.9 | 31 | 188 | 0.60 (0.36 - 0.96) | 0.04 | 0.60 (0.37 - 0.98) | 0.05 |
| SIDIAP | 4,309 | 19,733 | 6.3 | 44.9 | 5 | 70 | 0.32 (0.05 - 1.15) | 0.15 | 0.35 (0.07 - 1.67) | 0.19 |
| Meta-analysis | 296,662 | 537,776 | 274.4 | 518.3 | 1,525 | 5,206 | 0.75 (0.63 - 0.89) | <0.01 | 0.80 (0.65 - 0.97) | 0.03 |
| Meta-analysis (w/o USOC) | 61,125 | 129,978 | 44.6 | 128.1 | 327 | 1,092 | 0.82 (0.67 - 0.99) | 0.04 | 0.84 (0.69 - 1.02) | 0.09 |
| **PS-stratification / on-treatment TAR** | | | | | | | | | | |
| CCAE | 23,598 | 30,597 | 19.7 | 25.8 | 113 | 151 | 0.95 (0.74 - 1.24) | 0.73 | 0.99 (0.76 - 1.29) | 0.92 |
| DAG | 5,763 | 11,200 | 4.7 | 11.7 | 0 | 0 | | | | |
| ODOD | 16,421 | 20,195 | 11.5 | 15.9 | 163 | 349 | 0.78 (0.63 - 0.96) | 0.02 | 0.79 (0.64 - 0.98) | 0.04 |
| MDCR | 1,962 | 4,867 | 1.6 | 4.6 | 33 | 140 | 0.82 (0.53 - 1.24) | 0.37 | 0.85 (0.55 - 1.30) | 0.47 |
| MDCD | 1,350 | 3,145 | 0.8 | 2.3 | 11 | 42 | 0.94 (0.45 - 1.81) | 0.86 | 0.85 (0.42 - 1.70) | 0.65 |
| Open Claims | 315,211 | 408,030 | 277.3 | 390.3 | 1,631 | 4,137 | 0.70 (0.66 - 0.75) | <0.01 | 0.75 (0.65 - 0.87) | <0.01 |
| OEHR | 19,700 | 31,638 | 6.3 | 16.2 | 64 | 205 | 1.04 (0.75 - 1.40) | 0.83 | 1.04 (0.76 - 1.42) | 0.81 |
| VA | 11,226 | 24,158 | 6.8 | 22.3 | 73 | 194 | 0.63 (0.44 - 0.88) | <0.01 | 0.65 (0.46 - 0.92) | 0.02 |
| SIDIAP | 5,095 | 20,166 | 7.0 | 46.5 | 5 | 77 | 0.38 (0.12 - 0.92) | 0.06 | 0.39 (0.14 - 1.05) | 0.07 |
| Meta-analysis | 381,987 | 515,493 | 323.4 | 499.3 | 2,009 | 5,059 | 0.80 (0.69 - 0.94) | <0.01 | 0.85 (0.70 - 1.03) | 0.10 |
| Meta-analysis (w/o USOC) | 66,776 | 107,463 | 46.0 | 109.0 | 378 | 922 | 0.87 (0.73 - 1.03) | 0.09 | 0.88 (0.74 - 1.05) | 0.17 |
| **PS-matching / on-treatment TAR without escalation** | | | | | | | | | | |
| CCAE | 19,363 | 30,576 | 15.0 | 20.9 | 79 | 125 | 0.93 (0.60 - 1.43) | 0.75 | 0.99 (0.64 - 1.53) | 0.93 |
| DAG | 4,640 | 11,168 | 3.7 | 10.5 | 0 | 0 | | | | |
| ODOD | 11,932 | 20,118 | 8.1 | 12.9 | 99 | 291 | 0.76 (0.55 - 1.05) | 0.10 | 0.79 (0.55 - 1.13) | 0.26 |
| MDCR | 1,605 | 4,825 | 1.3 | 3.9 | 24 | 123 | 0.70 (0.36 - 1.28) | 0.27 | 0.71 (0.38 - 1.34) | 0.31 |
| MDCD | 1,287 | 3,127 | 0.7 | 1.7 | 10 | 33 | 1.12 (0.41 - 2.79) | 0.82 | 1.07 (0.41 - 2.79) | 0.88 |
| Open Claims | 235,537 | 407,798 | 192.8 | 302.8 | 969 | 3,212 | 0.64 (0.57 - 0.71) | <0.01 | 0.68 (0.59 - 0.79) | <0.01 |
| OEHR | 16,513 | 31,579 | 4.7 | 13.1 | 37 | 147 | 1.04 (0.63 - 1.67) | 0.88 | 1.05 (0.64 - 1.70) | 0.84 |
| VA | 7,403 | 23,147 | 4.3 | 17.0 | 27 | 147 | 0.66 (0.38 - 1.11) | 0.13 | 0.66 (0.38 - 1.12) | 0.13 |
| SIDIAP | 4,309 | 19,733 | 5.6 | 35.7 | <5 | 52 | 0.34 (0.05 - 1.28) | 0.19 | 0.40 (0.08 - 1.99) | 0.27 |
| Meta-analysis | 296,662 | 537,776 | 231.8 | 406.3 | <1,240 | 4,097 | 0.73 (0.61 - 0.86) | <0.01 | 0.77 (0.62 - 0.95) | 0.01 |
| Meta-analysis (w/o USOC) | 61,125 | 129,978 | 39.0 | 103.5 | <271 | 885 | 0.80 (0.66 - 0.98) | 0.03 | 0.83 (0.68 - 1.01) | 0.07 |
| **PS-stratification / on-treatment TAR without escalation** | | | | | | | | | | |
| CCAE | 23,598 | 30,597 | 17.2 | 20.9 | 93 | 125 | 0.87 (0.66 - 1.16) | 0.35 | 0.91 (0.68 - 1.21) | 0.52 |
| DAG | 5,763 | 11,200 | 4.3 | 10.6 | 0 | 0 | | | | |
| ODOD | 16,421 | 20,195 | 10.2 | 13.0 | 144 | 295 | 0.75 (0.60 - 0.94) | 0.01 | 0.75 (0.59 - 0.94) | 0.02 |
| MDCR | 1,962 | 4,867 | 1.4 | 3.9 | 28 | 126 | 0.71 (0.44 - 1.12) | 0.15 | 0.74 (0.46 - 1.18) | 0.22 |
| MDCD | 1,350 | 3,145 | 0.7 | 1.7 | 10 | 34 | 0.92 (0.42 - 1.85) | 0.82 | 0.84 (0.40 - 1.77) | 0.66 |
| Open Claims | 315,211 | 408,030 | 235.8 | 302.9 | 1,369 | 3,233 | 0.69 (0.64 - 0.74) | <0.01 | 0.74 (0.66 - 0.84) | <0.01 |
| OEHR | 19,700 | 31,638 | 5.5 | 13.1 | 50 | 148 | 0.98 (0.68 - 1.39) | 0.90 | 0.97 (0.68 - 1.39) | 0.86 |
| VA | 11,226 | 24,158 | 5.9 | 18.2 | 66 | 153 | 0.65 (0.45 - 0.94) | 0.02 | 0.67 (0.47 - 0.98) | 0.04 |
| SIDIAP | 5,095 | 20,166 | 6.2 | 36.8 | <5 | 58 | 0.28 (0.06 - 0.83) | 0.05 | 0.30 (0.08 - 1.10) | 0.07 |
| Meta-analysis | 381,987 | 515,493 | 276.4 | 390.7 | <1,689 | 3,985 | 0.75 (0.66 - 0.85) | <0.01 | 0.79 (0.68 - 0.91) | <0.01 |
| Meta-analysis (w/o USOC) | 66,776 | 107,463 | 40.6 | 87.8 | <320 | 752 | 0.81 (0.68 - 0.95) | 0.01 | 0.82 (0.69 - 0.98) | 0.04 |

##### 1.2.7 Measured renal dysfunction

**Supplementary Table 33:** Exposures, outcomes and risk estimates for Measured renal dysfunction in SGLT2I (male) (T) vs DPP4I (male) (C) new-user cohorts under four different propensity score (PS) adjustment / time-at-risk (TAR) designs across data sources. We report total patient exposure time in 1,000 patient-year (KYr) units, uncalibrated and calibrated hazard ratios (HRs) and their 95% confidence intervals (CIs). Greyed out entries did not pass study diagnostics and are included only for completeness.

| | Patients | | Exposure (KYr) | | Outcomes | | Uncalibrated | | Calibrated | |
|---|---|---|---|---|---|---|---|---|---|---|
| | T | C | T | C | T | C | HR (95% CI) | $p$ | HR (95% CI) | $p$ |
| **PS-matching / on-treatment TAR** | | | | | | | | | | |
| CCAE | 19,702 | 31,115 | 17.6 | 26.3 | 5 | 11 | 0.49 (0.11 - 1.72) | 0.32 | 0.53 (0.13 - 2.12) | 0.37 |
| DAG | 4,640 | 11,168 | 4.1 | 11.6 | 0 | 0 | | | | |
| ODOD | 12,499 | 21,418 | 9.5 | 16.9 | 6 | 42 | 0.68 (0.22 - 1.79) | 0.48 | 0.71 (0.25 - 2.02) | 0.52 |
| MDCR | 1,721 | 5,267 | 1.5 | 5.0 | 0 | <5 | | | | |
| MDCD | 1,426 | 3,470 | 0.9 | 2.6 | 0 | 0 | | | | |
| Open Claims | 244,206 | 426,386 | 237.2 | 409.0 | 0 | 0 | | | | |
| OEHR | 16,869 | 32,344 | 5.5 | 16.7 | 0 | 0 | | | | |
| VA | 7,710 | 23,670 | 5.3 | 21.4 | 10 | 89 | 0.44 (0.20 - 0.86) | 0.02 | 0.44 (0.22 - 0.90) | 0.03 |
| SIDIAP | 4,322 | 19,658 | 6.3 | 44.5 | 13 | 276 | 0.42 (0.20 - 0.78) | 0.01 | 0.45 (0.23 - 0.90) | 0.03 |
| Meta-analysis | 24,531 | 64,746 | 21.1 | 82.8 | 29 | 407 | 0.46 (0.30 - 0.73) | <0.01 | 0.49 (0.31 - 0.78) | <0.01 |
| Meta-analysis (w/o USOC) | 24,531 | 64,746 | 21.1 | 82.8 | 29 | 407 | 0.46 (0.30 - 0.73) | <0.01 | 0.48 (0.31 - 0.75) | <0.01 |
| **PS-stratification / on-treatment TAR** | | | | | | | | | | |
| CCAE | 24,082 | 31,138 | 20.0 | 26.3 | 5 | 11 | 1.00 (0.31 - 2.91) | 0.99 | 1.04 (0.34 - 3.22) | 0.94 |
| DAG | 5,763 | 11,200 | 4.7 | 11.7 | 0 | 0 | | | | |
| ODOD | 17,471 | 21,538 | 12.1 | 17.0 | 7 | 44 | 0.40 (0.16 - 0.89) | 0.04 | 0.41 (0.17 - 0.96) | 0.04 |
| MDCR | 2,136 | 5,326 | 1.7 | 5.0 | 0 | <5 | | | | |
| MDCD | 1,507 | 3,484 | 0.9 | 2.6 | 0 | 0 | | | | |
| Open Claims | 332,801 | 426,715 | 289.7 | 409.2 | 0 | 0 | | | | |
| OEHR | 20,231 | 32,415 | 6.5 | 16.8 | 0 | 0 | | | | |
| VA | 12,350 | 24,682 | 7.4 | 22.9 | 23 | 95 | 0.42 (0.23 - 0.73) | <0.01 | 0.44 (0.25 - 0.77) | <0.01 |
| SIDIAP | 5,147 | 20,075 | 7.0 | 46.0 | 16 | 299 | 0.40 (0.22 - 0.68) | <0.01 | 0.41 (0.23 - 0.72) | <0.01 |
| Meta-analysis | 22,618 | 41,613 | 19.2 | 62.9 | 23 | 343 | 0.40 (0.25 - 0.64) | <0.01 | 0.42 (0.26 - 0.68) | <0.01 |
| Meta-analysis (w/o USOC) | 22,618 | 41,613 | 19.2 | 62.9 | 23 | 343 | 0.40 (0.25 - 0.64) | <0.01 | 0.41 (0.25 - 0.65) | <0.01 |
| **PS-matching / on-treatment TAR without escalation** | | | | | | | | | | |
| CCAE | 19,702 | 31,115 | 15.2 | 21.3 | 5 | 10 | 0.49 (0.11 - 1.72) | 0.32 | 0.52 (0.13 - 2.09) | 0.36 |
| DAG | 4,640 | 11,168 | 3.7 | 10.5 | 0 | 0 | | | | |
| ODOD | 12,499 | 21,418 | 8.4 | 13.8 | 6 | 37 | 0.68 (0.22 - 1.82) | 0.48 | 0.71 (0.24 - 2.07) | 0.53 |
| MDCR | 1,721 | 5,267 | 1.4 | 4.2 | 0 | 0 | | | | |
| MDCD | 1,426 | 3,470 | 0.7 | 1.9 | 0 | 0 | | | | |
| Open Claims | 244,206 | 426,386 | 198.9 | 317.1 | 0 | 0 | | | | |
| OEHR | 16,869 | 32,344 | 4.8 | 13.5 | 0 | 0 | | | | |
| VA | 7,710 | 23,670 | 4.5 | 17.4 | <10 | 69 | 0.38 (0.16 - 0.79) | 0.02 | 0.38 (0.17 - 0.83) | 0.02 |
| SIDIAP | 4,322 | 19,658 | 5.6 | 35.3 | 11 | 230 | 0.32 (0.13 - 0.66) | <0.01 | 0.37 (0.17 - 0.84) | 0.02 |
| Meta-analysis | 24,531 | 64,746 | 18.5 | 66.5 | <27 | 336 | 0.40 (0.24 - 0.66) | <0.01 | 0.43 (0.25 - 0.71) | <0.01 |
| Meta-analysis (w/o USOC) | 24,531 | 64,746 | 18.5 | 66.5 | <27 | 336 | 0.40 (0.24 - 0.66) | <0.01 | 0.42 (0.25 - 0.68) | <0.01 |
| **PS-stratification / on-treatment TAR without escalation** | | | | | | | | | | |
| CCAE | 24,082 | 31,138 | 17.5 | 21.3 | 5 | 10 | 1.04 (0.31 - 3.08) | 0.95 | 1.08 (0.34 - 3.39) | 0.90 |
| DAG | 5,763 | 11,200 | 4.3 | 10.6 | 0 | 0 | | | | |
| ODOD | 17,471 | 21,538 | 10.8 | 13.9 | 7 | 39 | 0.44 (0.17 - 0.99) | 0.07 | 0.44 (0.18 - 1.05) | 0.07 |
| MDCR | 2,136 | 5,326 | 1.5 | 4.3 | 0 | 0 | | | | |
| MDCD | 1,507 | 3,484 | 0.8 | 1.9 | 0 | 0 | | | | |
| Open Claims | 332,801 | 426,715 | 246.2 | 317.3 | 0 | 0 | | | | |
| OEHR | 20,231 | 32,415 | 5.7 | 13.6 | 0 | 0 | | | | |
| VA | 12,350 | 24,682 | 6.4 | 18.6 | 18 | 75 | 0.41 (0.21 - 0.77) | <0.01 | 0.43 (0.23 - 0.81) | <0.01 |
| SIDIAP | 5,147 | 20,075 | 6.3 | 36.4 | 14 | 246 | 0.36 (0.19 - 0.64) | <0.01 | 0.40 (0.22 - 0.73) | <0.01 |
| Meta-analysis | 22,618 | 41,613 | 17.0 | 50.3 | 21 | 285 | 0.39 (0.23 - 0.64) | <0.01 | 0.41 (0.25 - 0.67) | <0.01 |
| Meta-analysis (w/o USOC) | 22,618 | 41,613 | 17.0 | 50.3 | 21 | 285 | 0.39 (0.23 - 0.64) | <0.01 | 0.39 (0.24 - 0.65) | <0.01 |

##### 1.2.8 Stroke

**Supplementary Table 34:** Exposures, outcomes and risk estimates for Stroke in SGLT2I (male) (T) vs DPP4I (male) (C) new-user cohorts under four different propensity score (PS) adjustment / time-at-risk (TAR) designs across data sources. We report total patient exposure time in 1,000 patient-year (KYr) units, uncalibrated and calibrated hazard ratios (HRs) and their 95% confidence intervals (CIs). Greyed out entries did not pass study diagnostics and are included only for completeness.

| | Patients | | Exposure (KYr) | | Outcomes | | Uncalibrated | | Calibrated | |
|---|---|---|---|---|---|---|---|---|---|---|
| | T | C | T | C | T | C | HR (95% CI) | $p$ | HR (95% CI) | $p$ |
| **PS-matching / on-treatment TAR** | | | | | | | | | | |
| CCAE | 19,517 | 30,782 | 17.4 | 26.0 | 51 | 93 | 0.70 (0.41 - 1.15) | 0.16 | 0.75 (0.45 - 1.25) | 0.27 |
| DAG | 4,640 | 11,168 | 4.1 | 11.6 | 0 | 0 | | | | |
| ODOD | 12,233 | 20,802 | 9.3 | 16.4 | 67 | 161 | 0.93 (0.61 - 1.38) | 0.71 | 0.96 (0.64 - 1.44) | 0.84 |
| MDCR | 1,670 | 5,029 | 1.5 | 4.7 | 16 | 60 | 0.61 (0.22 - 1.47) | 0.31 | 0.61 (0.24 - 1.59) | 0.32 |
| MDCD | 1,375 | 3,307 | 0.9 | 2.4 | 6 | 26 | 0.66 (0.14 - 2.29) | 0.56 | 0.60 (0.15 - 2.39) | 0.47 |
| Open Claims | 238,515 | 414,277 | 232.2 | 397.2 | 781 | 2,049 | 0.91 (0.81 - 1.02) | 0.12 | 0.99 (0.85 - 1.16) | 0.90 |
| OEHR | 16,716 | 32,022 | 5.5 | 16.5 | 18 | 72 | 1.40 (0.63 - 3.00) | 0.39 | 1.41 (0.65 - 3.07) | 0.39 |
| VA | 7,643 | 23,577 | 5.2 | 21.4 | 11 | 55 | 1.72 (0.71 - 3.97) | 0.22 | 1.73 (0.73 - 4.10) | 0.22 |
| SIDIAP | 4,251 | 19,534 | 6.2 | 44.4 | 13 | 131 | 0.77 (0.39 - 1.41) | 0.43 | 0.84 (0.44 - 1.60) | 0.61 |
| Meta-analysis | 300,545 | 546,023 | 277.3 | 526.6 | 957 | 2,621 | 0.91 (0.82 - 1.01) | 0.08 | 0.96 (0.83 - 1.11) | 0.63 |
| Meta-analysis (w/o USOC) | 62,030 | 131,746 | 45.1 | 129.4 | 176 | 572 | 0.90 (0.70 - 1.16) | 0.42 | 0.93 (0.72 - 1.20) | 0.58 |
| **PS-stratification / on-treatment TAR** | | | | | | | | | | |
| CCAE | 23,843 | 30,803 | 19.9 | 26.0 | 58 | 93 | 0.87 (0.62 - 1.22) | 0.43 | 0.90 (0.64 - 1.28) | 0.57 |
| DAG | 5,763 | 11,200 | 4.7 | 11.7 | 0 | 0 | | | | |
| ODOD | 17,093 | 20,912 | 11.8 | 16.5 | 81 | 167 | 0.92 (0.68 - 1.24) | 0.60 | 0.93 (0.69 - 1.26) | 0.66 |
| MDCR | 2,074 | 5,079 | 1.7 | 4.8 | 18 | 60 | 1.12 (0.61 - 1.98) | 0.70 | 1.16 (0.64 - 2.10) | 0.63 |
| MDCD | 1,446 | 3,329 | 0.9 | 2.4 | 6 | 27 | 0.78 (0.28 - 1.86) | 0.61 | 0.71 (0.28 - 1.81) | 0.48 |
| Open Claims | 324,481 | 414,551 | 283.2 | 397.4 | 934 | 2,055 | 0.86 (0.79 - 0.93) | <0.01 | 0.92 (0.79 - 1.08) | 0.34 |
| OEHR | 20,037 | 32,091 | 6.4 | 16.6 | 19 | 73 | 1.06 (0.60 - 1.81) | 0.83 | 1.06 (0.61 - 1.85) | 0.83 |
| VA | 12,216 | 24,595 | 7.4 | 22.9 | 19 | 63 | 1.00 (0.49 - 1.98) | 1.00 | 1.05 (0.52 - 2.09) | 0.90 |
| SIDIAP | 5,058 | 19,960 | 6.9 | 45.9 | 14 | 141 | 0.69 (0.36 - 1.22) | 0.23 | 0.71 (0.39 - 1.30) | 0.29 |
| Meta-analysis | 392,586 | 523,396 | 329.9 | 507.2 | 1,124 | 2,589 | 0.87 (0.80 - 0.94) | <0.01 | 0.91 (0.79 - 1.05) | 0.20 |
| Meta-analysis (w/o USOC) | 68,105 | 108,845 | 46.7 | 109.8 | 190 | 534 | 0.92 (0.76 - 1.10) | 0.35 | 0.93 (0.77 - 1.13) | 0.49 |
| **PS-matching / on-treatment TAR without escalation** | | | | | | | | | | |
| CCAE | 19,517 | 30,782 | 15.1 | 21.1 | 41 | 73 | 0.77 (0.44 - 1.30) | 0.33 | 0.81 (0.47 - 1.40) | 0.46 |
| DAG | 4,640 | 11,168 | 3.7 | 10.5 | 0 | 0 | | | | |
| ODOD | 12,233 | 20,802 | 8.2 | 13.4 | 59 | 138 | 0.84 (0.53 - 1.28) | 0.42 | 0.86 (0.54 - 1.37) | 0.57 |
| MDCR | 1,670 | 5,029 | 1.3 | 4.0 | 13 | 50 | 0.55 (0.18 - 1.42) | 0.26 | 0.56 (0.20 - 1.58) | 0.28 |
| MDCD | 1,375 | 3,307 | 0.7 | 1.8 | <5 | 19 | 0.68 (0.10 - 2.99) | 0.65 | 0.65 (0.12 - 3.59) | 0.62 |
| Open Claims | 238,515 | 414,277 | 194.7 | 308.1 | 664 | 1,629 | 0.90 (0.79 - 1.02) | 0.09 | 0.96 (0.81 - 1.13) | 0.66 |
| OEHR | 16,716 | 32,022 | 4.8 | 13.4 | 11 | 53 | 0.80 (0.30 - 1.93) | 0.65 | 0.81 (0.32 - 2.05) | 0.66 |
| VA | 7,643 | 23,577 | 4.5 | 17.4 | <10 | 36 | 2.19 (0.78 - 5.82) | 0.13 | 2.18 (0.80 - 5.95) | 0.13 |
| SIDIAP | 4,251 | 19,534 | 5.5 | 35.2 | 12 | 96 | 0.73 (0.35 - 1.41) | 0.38 | 0.86 (0.43 - 1.73) | 0.69 |
| Meta-analysis | 300,545 | 546,023 | 234.1 | 412.6 | <810 | 2,075 | 0.88 (0.79 - 0.99) | 0.04 | 0.93 (0.79 - 1.10) | 0.43 |
| Meta-analysis (w/o USOC) | 62,030 | 131,746 | 39.4 | 104.5 | <146 | 446 | 0.83 (0.64 - 1.09) | 0.18 | 0.86 (0.65 - 1.13) | 0.28 |
| **PS-stratification / on-treatment TAR without escalation** | | | | | | | | | | |
| CCAE | 23,843 | 30,803 | 17.3 | 21.1 | 46 | 73 | 0.84 (0.57 - 1.23) | 0.37 | 0.87 (0.59 - 1.28) | 0.49 |
| DAG | 5,763 | 11,200 | 4.3 | 10.6 | 0 | 0 | | | | |
| ODOD | 17,093 | 20,912 | 10.5 | 13.5 | 72 | 144 | 0.92 (0.67 - 1.26) | 0.62 | 0.91 (0.66 - 1.26) | 0.59 |
| MDCR | 2,074 | 5,079 | 1.5 | 4.1 | 15 | 50 | 1.06 (0.54 - 1.99) | 0.86 | 1.10 (0.57 - 2.12) | 0.77 |
| MDCD | 1,446 | 3,329 | 0.7 | 1.8 | <5 | 19 | 0.65 (0.18 - 1.83) | 0.47 | 0.60 (0.19 - 1.89) | 0.38 |
| Open Claims | 324,481 | 414,551 | 240.8 | 308.3 | 806 | 1,634 | 0.85 (0.78 - 0.94) | <0.01 | 0.92 (0.81 - 1.05) | 0.25 |
| OEHR | 20,037 | 32,091 | 5.6 | 13.4 | 12 | 53 | 0.79 (0.39 - 1.52) | 0.51 | 0.79 (0.40 - 1.56) | 0.50 |
| VA | 12,216 | 24,595 | 6.4 | 18.6 | 14 | 41 | 1.07 (0.46 - 2.42) | 0.86 | 1.12 (0.49 - 2.55) | 0.80 |
| SIDIAP | 5,058 | 19,960 | 6.2 | 36.4 | 13 | 104 | 0.74 (0.37 - 1.35) | 0.35 | 0.81 (0.43 - 1.54) | 0.54 |
| Meta-analysis | 392,586 | 523,396 | 281.9 | 396.7 | 964 | 2,058 | 0.86 (0.79 - 0.93) | <0.01 | 0.91 (0.81 - 1.01) | 0.08 |
| Meta-analysis (w/o USOC) | 68,105 | 108,845 | 41.2 | 88.4 | 158 | 424 | 0.88 (0.71 - 1.08) | 0.21 | 0.90 (0.73 - 1.10) | 0.32 |

##### 1.2.9 Sudden cardiac death

**Supplementary Table 35:** Exposures, outcomes and risk estimates for Sudden cardiac death in SGLT2I (male) (T) vs DPP4I (male) (C) new-user cohorts under four different propensity score (PS) adjustment / time-at-risk (TAR) designs across data sources. We report total patient exposure time in 1,000 patient-year (KYr) units, uncalibrated and calibrated hazard ratios (HRs) and their 95% confidence intervals (CIs). Greyed out entries did not pass study diagnostics and are included only for completeness.

| | Patients | | Exposure (KYr) | | Outcomes | | Uncalibrated | | Calibrated | |
|---|---|---|---|---|---|---|---|---|---|---|
| | T | C | T | C | T | C | HR (95% CI) | $p$ | HR (95% CI) | $p$ |
| **PS-matching / on-treatment TAR** | | | | | | | | | | |
| CCAE | 19,730 | 31,143 | 17.6 | 26.4 | 15 | 27 | 0.65 (0.24 - 1.59) | 0.37 | 0.70 (0.27 - 1.79) | 0.46 |
| DAG | 4,640 | 11,168 | 4.1 | 11.6 | 0 | 0 | | | | |
| ODOD | 12,497 | 21,421 | 9.6 | 16.9 | 15 | 70 | 0.34 (0.13 - 0.80) | 0.02 | 0.35 (0.14 - 0.90) | 0.03 |
| MDCR | 1,715 | 5,246 | 1.5 | 5.0 | 9 | 21 | 2.19 (0.60 - 7.46) | 0.22 | 2.20 (0.62 - 7.79) | 0.22 |
| MDCD | 1,419 | 3,454 | 0.9 | 2.5 | <5 | 8 | 0.77 (0.04 - 6.26) | 0.84 | 0.69 (0.05 - 9.10) | 0.78 |
| Open Claims | 243,645 | 425,371 | 236.7 | 407.8 | 188 | 643 | 0.66 (0.51 - 0.84) | <0.01 | 0.71 (0.55 - 0.93) | 0.02 |
| OEHR | 16,851 | 32,316 | 5.5 | 16.7 | <5 | 22 | 0.50 (0.14 - 1.47) | 0.25 | 0.50 (0.15 - 1.65) | 0.26 |
| VA | 7,788 | 23,941 | 5.3 | 21.7 | 0 | <10 | | | | |
| SIDIAP | 4,338 | 19,828 | 6.3 | 45.1 | 0 | <5 | | | | |
| Meta-analysis | 294,438 | 515,497 | 270.9 | 472.7 | <232 | 783 | 0.65 (0.52 - 0.81) | <0.01 | 0.69 (0.54 - 0.88) | <0.01 |
| Meta-analysis (w/o USOC) | 50,793 | 90,126 | 34.2 | 64.9 | <44 | 140 | 0.65 (0.32 - 1.35) | 0.25 | 0.67 (0.33 - 1.39) | 0.29 |
| **PS-stratification / on-treatment TAR** | | | | | | | | | | |
| CCAE | 24,111 | 31,166 | 20.1 | 26.4 | 15 | 27 | 0.69 (0.35 - 1.33) | 0.28 | 0.72 (0.37 - 1.40) | 0.34 |
| DAG | 5,763 | 11,200 | 4.7 | 11.7 | 0 | 0 | | | | |
| ODOD | 17,442 | 21,543 | 12.1 | 17.0 | 19 | 70 | 0.42 (0.23 - 0.71) | <0.01 | 0.42 (0.24 - 0.73) | <0.01 |
| MDCR | 2,130 | 5,305 | 1.7 | 5.0 | 11 | 21 | 2.07 (0.87 - 4.67) | 0.09 | 2.13 (0.92 - 4.94) | 0.08 |
| MDCD | 1,499 | 3,468 | 0.9 | 2.6 | <5 | 8 | 1.29 (0.18 - 5.85) | 0.78 | 1.16 (0.20 - 6.69) | 0.87 |
| Open Claims | 331,685 | 425,694 | 288.8 | 408.0 | 259 | 648 | 0.67 (0.57 - 0.78) | <0.01 | 0.72 (0.59 - 0.88) | <0.01 |
| OEHR | 20,205 | 32,387 | 6.5 | 16.8 | 5 | 24 | 0.70 (0.23 - 1.80) | 0.50 | 0.70 (0.25 - 1.98) | 0.51 |
| VA | 12,496 | 24,984 | 7.5 | 23.2 | <10 | <10 | 1.02 (0.23 - 4.49) | 0.98 | 1.07 (0.24 - 4.73) | 0.93 |
| SIDIAP | 5,160 | 20,267 | 7.1 | 46.7 | 0 | <5 | | | | |
| Meta-analysis | 395,573 | 516,095 | 329.2 | 473.2 | 309 | 790 | 0.73 (0.46 - 1.15) | 0.17 | 0.76 (0.48 - 1.23) | 0.27 |
| Meta-analysis (w/o USOC) | 63,888 | 90,401 | 40.4 | 65.1 | 50 | 142 | 0.77 (0.39 - 1.52) | 0.46 | 0.79 (0.40 - 1.55) | 0.49 |
| **PS-matching / on-treatment TAR without escalation** | | | | | | | | | | |
| CCAE | 19,730 | 31,143 | 15.3 | 21.3 | 11 | 22 | 0.52 (0.16 - 1.40) | 0.23 | 0.55 (0.19 - 1.61) | 0.28 |
| DAG | 4,640 | 11,168 | 3.7 | 10.5 | 0 | 0 | | | | |
| ODOD | 12,497 | 21,421 | 8.4 | 13.8 | 12 | 64 | 0.28 (0.08 - 0.77) | 0.03 | 0.29 (0.09 - 0.91) | 0.04 |
| MDCR | 1,715 | 5,246 | 1.4 | 4.2 | 8 | 13 | 2.04 (0.55 - 7.02) | 0.27 | 2.07 (0.58 - 7.38) | 0.27 |
| MDCD | 1,419 | 3,454 | 0.7 | 1.9 | <5 | 7 | 0.63 (0.03 - 5.99) | 0.74 | 0.61 (0.04 - 8.67) | 0.71 |
| Open Claims | 243,645 | 425,371 | 198.4 | 316.3 | 160 | 495 | 0.67 (0.51 - 0.88) | <0.01 | 0.72 (0.54 - 0.97) | 0.03 |
| OEHR | 16,851 | 32,316 | 4.8 | 13.5 | <5 | 16 | 0.35 (0.05 - 1.44) | 0.21 | 0.35 (0.07 - 1.85) | 0.22 |
| VA | 7,788 | 23,941 | 4.5 | 17.7 | 0 | <10 | | | | |
| SIDIAP | 4,338 | 19,828 | 5.6 | 35.8 | 0 | <5 | | | | |
| Meta-analysis | 292,723 | 510,251 | 226.9 | 364.9 | <188 | 597 | 0.56 (0.37 - 0.86) | <0.01 | 0.59 (0.38 - 0.92) | 0.02 |
| Meta-analysis (w/o USOC) | 49,078 | 84,880 | 28.5 | 48.7 | <28 | 102 | 0.38 (0.19 - 0.77) | <0.01 | 0.39 (0.19 - 0.79) | <0.01 |
| **PS-stratification / on-treatment TAR without escalation** | | | | | | | | | | |
| CCAE | 24,111 | 31,166 | 17.5 | 21.3 | 11 | 22 | 0.59 (0.26 - 1.24) | 0.18 | 0.61 (0.28 - 1.33) | 0.22 |
| DAG | 5,763 | 11,200 | 4.3 | 10.6 | 0 | 0 | | | | |
| ODOD | 17,442 | 21,543 | 10.7 | 13.9 | 16 | 64 | 0.35 (0.19 - 0.62) | <0.01 | 0.35 (0.19 - 0.63) | <0.01 |
| MDCR | 2,130 | 5,305 | 1.5 | 4.3 | 10 | 13 | 2.71 (0.99 - 7.09) | 0.05 | 2.81 (1.05 - 7.51) | 0.04 |
| MDCD | 1,499 | 3,468 | 0.8 | 1.9 | <5 | 7 | 1.14 (0.16 - 5.36) | 0.88 | 1.05 (0.18 - 6.15) | 0.95 |
| Open Claims | 331,685 | 425,694 | 245.5 | 316.4 | 228 | 500 | 0.69 (0.58 - 0.82) | <0.01 | 0.75 (0.61 - 0.91) | <0.01 |
| OEHR | 20,205 | 32,387 | 5.7 | 13.5 | <5 | 17 | 0.51 (0.11 - 1.63) | 0.32 | 0.51 (0.13 - 1.92) | 0.32 |
| VA | 12,496 | 24,984 | 6.5 | 18.9 | <10 | <10 | 1.09 (0.19 - 6.49) | 0.93 | 1.13 (0.19 - 6.59) | 0.89 |
| SIDIAP | 5,160 | 20,267 | 6.3 | 37.0 | 0 | <5 | | | | |
| Meta-analysis | 395,573 | 516,095 | 280.9 | 369.5 | <270 | 616 | 0.69 (0.37 - 1.29) | 0.25 | 0.73 (0.39 - 1.37) | 0.33 |
| Meta-analysis (w/o USOC) | 63,888 | 90,401 | 35.5 | 53.1 | <42 | 116 | 0.71 (0.29 - 1.71) | 0.44 | 0.72 (0.30 - 1.75) | 0.47 |

##### 1.2.10 Abnormal weight gain

**Supplementary Table 36:** Exposures, outcomes and risk estimates for Abnormal weight gain in SGLT2I (male) (T) vs DPP4I (male) (C) new-user cohorts under four different propensity score (PS) adjustment / time-at-risk (TAR) designs across data sources. We report total patient exposure time in 1,000 patient-year (KYr) units, uncalibrated and calibrated hazard ratios (HRs) and their 95% confidence intervals (CIs). Greyed out entries did not pass study diagnostics and are included only for completeness.

| | Patients | | Exposure (KYr) | | Outcomes | | Uncalibrated | | Calibrated | |
|---|---|---|---|---|---|---|---|---|---|---|
| | T | C | T | C | T | C | HR (95% CI) | $p$ | HR (95% CI) | $p$ |
| **PS-matching / on-treatment TAR** | | | | | | | | | | |
| CCAE | 19,056 | 30,297 | 17.0 | 25.6 | 59 | 85 | 1.38 (0.87 - 2.20) | 0.17 | 1.49 (0.93 - 2.38) | 0.10 |
| DAG | 4,609 | 11,102 | 4.0 | 11.6 | 0 | 13 | | | | |
| ODOD | 12,108 | 20,883 | 9.2 | 16.4 | 28 | 92 | 0.48 (0.26 - 0.86) | 0.02 | 0.50 (0.27 - 0.92) | 0.03 |
| MDCR | 1,687 | 5,199 | 1.5 | 4.9 | <5 | 10 | 1.04 (0.05 - 12.4) | 0.98 | 1.05 (0.06 - 17.4) | 0.97 |
| MDCD | 1,334 | 3,301 | 0.8 | 2.4 | <5 | 16 | 0.61 (0.16 - 1.86) | 0.42 | 0.55 (0.16 - 1.85) | 0.34 |
| Open Claims | 236,160 | 414,950 | 228.8 | 397.2 | 269 | 700 | 0.73 (0.59 - 0.89) | <0.01 | 0.79 (0.63 - 1.00) | 0.05 |
| OEHR | 16,309 | 31,521 | 5.3 | 16.2 | 26 | 67 | 1.38 (0.70 - 2.71) | 0.36 | 1.38 (0.70 - 2.73) | 0.35 |
| VA | 7,634 | 23,500 | 5.2 | 21.3 | <10 | 11 | 2.22 (0.45 - 12.1) | 0.34 | 2.23 (0.43 - 11.5) | 0.34 |
| SIDIAP | 4,284 | 19,581 | 6.2 | 44.6 | 8 | 53 | 1.58 (0.61 - 3.56) | 0.31 | 1.72 (0.71 - 4.16) | 0.24 |
| Meta-analysis | 287,917 | 517,232 | 266.6 | 500.0 | 390 | 997 | 0.96 (0.63 - 1.48) | 0.86 | 1.02 (0.66 - 1.58) | 0.93 |
| Meta-analysis (w/o USOC) | 51,757 | 102,282 | 37.8 | 102.8 | 121 | 297 | 1.08 (0.62 - 1.87) | 0.80 | 1.11 (0.64 - 1.93) | 0.72 |
| **PS-stratification / on-treatment TAR** | | | | | | | | | | |
| CCAE | 23,300 | 30,318 | 19.3 | 25.6 | 68 | 86 | 1.04 (0.74 - 1.45) | 0.82 | 1.08 (0.77 - 1.51) | 0.65 |
| DAG | 5,722 | 11,134 | 4.7 | 11.6 | 0 | 13 | | | | |
| ODOD | 16,867 | 21,000 | 11.7 | 16.5 | 32 | 92 | 0.51 (0.33 - 0.78) | <0.01 | 0.52 (0.33 - 0.80) | <0.01 |
| MDCR | 2,099 | 5,258 | 1.7 | 5.0 | <5 | 10 | 0.64 (0.09 - 2.93) | 0.62 | 0.66 (0.11 - 3.85) | 0.65 |
| MDCD | 1,408 | 3,314 | 0.8 | 2.4 | <5 | 16 | 0.62 (0.17 - 1.81) | 0.43 | 0.56 (0.17 - 1.83) | 0.34 |
| Open Claims | 321,381 | 415,274 | 279.0 | 397.5 | 317 | 701 | 0.73 (0.63 - 0.84) | <0.01 | 0.79 (0.65 - 0.95) | 0.02 |
| OEHR | 19,556 | 31,592 | 6.2 | 16.3 | 30 | 67 | 1.21 (0.74 - 1.93) | 0.44 | 1.21 (0.75 - 1.96) | 0.44 |
| VA | 12,226 | 24,501 | 7.4 | 22.8 | <10 | 11 | 1.69 (0.56 - 5.19) | 0.35 | 1.77 (0.58 - 5.37) | 0.31 |
| SIDIAP | 5,107 | 20,006 | 7.0 | 46.1 | 8 | 60 | 1.66 (0.71 - 3.46) | 0.21 | 1.71 (0.77 - 3.78) | 0.19 |
| Meta-analysis | 386,211 | 518,190 | 323.2 | 501.9 | 455 | 1,006 | 0.89 (0.63 - 1.26) | 0.51 | 0.94 (0.65 - 1.35) | 0.73 |
| Meta-analysis (w/o USOC) | 64,830 | 102,916 | 44.2 | 104.5 | 138 | 305 | 0.97 (0.61 - 1.52) | 0.88 | 0.99 (0.62 - 1.55) | 0.95 |
| **PS-matching / on-treatment TAR without escalation** | | | | | | | | | | |
| CCAE | 19,056 | 30,297 | 14.7 | 20.7 | 51 | 69 | 1.71 (1.02 - 2.90) | 0.04 | 1.82 (1.08 - 3.07) | 0.03 |
| DAG | 4,609 | 11,102 | 3.7 | 10.4 | 0 | 11 | | | | |
| ODOD | 12,108 | 20,883 | 8.1 | 13.5 | 23 | 70 | 0.55 (0.27 - 1.06) | 0.09 | 0.57 (0.28 - 1.15) | 0.13 |
| MDCR | 1,687 | 5,199 | 1.3 | 4.2 | <5 | 7 | 1.04 (0.05 - 12.4) | 0.98 | 1.05 (0.06 - 17.5) | 0.97 |
| MDCD | 1,334 | 3,301 | 0.7 | 1.8 | <5 | 10 | 0.82 (0.21 - 2.75) | 0.77 | 0.79 (0.22 - 2.84) | 0.72 |
| Open Claims | 236,160 | 414,950 | 192.0 | 308.7 | 213 | 526 | 0.70 (0.55 - 0.88) | <0.01 | 0.75 (0.58 - 0.97) | 0.04 |
| OEHR | 16,309 | 31,521 | 4.7 | 13.1 | 21 | 52 | 1.42 (0.65 - 3.08) | 0.38 | 1.43 (0.66 - 3.09) | 0.37 |
| VA | 7,634 | 23,500 | 4.4 | 17.4 | <10 | <10 | 2.19 (0.45 - 11.7) | 0.35 | 2.17 (0.43 - 11.1) | 0.35 |
| SIDIAP | 4,284 | 19,581 | 5.6 | 35.4 | 8 | 45 | 1.76 (0.67 - 4.09) | 0.22 | 2.06 (0.83 - 5.12) | 0.13 |
| Meta-analysis | 287,917 | 517,232 | 225.0 | 391.4 | 316 | 762 | 1.05 (0.65 - 1.69) | 0.84 | 1.11 (0.68 - 1.82) | 0.67 |
| Meta-analysis (w/o USOC) | 51,757 | 102,282 | 33.0 | 82.7 | 103 | 236 | 1.23 (0.70 - 2.15) | 0.47 | 1.27 (0.72 - 2.22) | 0.41 |
| **PS-stratification / on-treatment TAR without escalation** | | | | | | | | | | |
| CCAE | 23,300 | 30,318 | 16.9 | 20.7 | 60 | 70 | 1.07 (0.74 - 1.55) | 0.71 | 1.12 (0.77 - 1.61) | 0.56 |
| DAG | 5,722 | 11,134 | 4.3 | 10.5 | 0 | 11 | | | | |
| ODOD | 16,867 | 21,000 | 10.4 | 13.5 | 25 | 70 | 0.48 (0.29 - 0.78) | <0.01 | 0.48 (0.29 - 0.79) | <0.01 |
| MDCR | 2,099 | 5,258 | 1.5 | 4.2 | <5 | 7 | 0.87 (0.11 - 4.38) | 0.89 | 0.91 (0.14 - 5.71) | 0.92 |
| MDCD | 1,408 | 3,314 | 0.7 | 1.8 | <5 | 10 | 0.86 (0.22 - 2.80) | 0.82 | 0.79 (0.22 - 2.79) | 0.71 |
| Open Claims | 321,381 | 415,274 | 237.4 | 308.9 | 254 | 527 | 0.69 (0.58 - 0.81) | <0.01 | 0.74 (0.62 - 0.90) | <0.01 |
| OEHR | 19,556 | 31,592 | 5.5 | 13.2 | 24 | 52 | 1.21 (0.70 - 2.04) | 0.49 | 1.20 (0.70 - 2.05) | 0.51 |
| VA | 12,226 | 24,501 | 6.4 | 18.5 | <10 | <10 | 1.67 (0.53 - 5.43) | 0.39 | 1.73 (0.54 - 5.54) | 0.36 |
| SIDIAP | 5,107 | 20,006 | 6.2 | 36.5 | 8 | 50 | 1.80 (0.76 - 3.81) | 0.15 | 1.97 (0.88 - 4.43) | 0.10 |
| Meta-analysis | 386,211 | 518,190 | 276.4 | 392.8 | 371 | 769 | 0.89 (0.60 - 1.32) | 0.57 | 0.94 (0.63 - 1.41) | 0.77 |
| Meta-analysis (w/o USOC) | 64,830 | 102,916 | 39.0 | 84.0 | 117 | 242 | 0.98 (0.60 - 1.62) | 0.95 | 1.01 (0.61 - 1.65) | 0.96 |

##### 1.2.11 Abnormal weight loss

**Supplementary Table 37:** Exposures, outcomes and risk estimates for Abnormal weight loss in SGLT2I (male) (T) vs DPP4I (male) (C) new-user cohorts under four different propensity score (PS) adjustment / time-at-risk (TAR) designs across data sources. We report total patient exposure time in 1,000 patient-year (KYr) units, uncalibrated and calibrated hazard ratios (HRs) and their 95% confidence intervals (CIs). Greyed out entries did not pass study diagnostics and are included only for completeness.

| | Patients | | Exposure (KYr) | | Outcomes | | Uncalibrated | | Calibrated | |
|---|---|---|---|---|---|---|---|---|---|---|
| | T | C | T | C | T | C | HR (95% CI) | $p$ | HR (95% CI) | $p$ |
| **PS-matching / on-treatment TAR** | | | | | | | | | | |
| CCAE | 19,278 | 30,384 | 17.2 | 25.6 | 123 | 205 | 1.01 (0.74 - 1.37) | 0.95 | 1.09 (0.80 - 1.48) | 0.60 |
| DAG | 4,550 | 10,926 | 4.0 | 11.4 | 33 | 79 | 1.10 (0.62 - 1.89) | 0.75 | 1.07 (0.61 - 1.87) | 0.77 |
| ODOD | 12,051 | 20,464 | 9.1 | 16.0 | 158 | 265 | 1.41 (1.05 - 1.90) | 0.02 | 1.46 (1.08 - 1.97) | 0.03 |
| MDCR | 1,655 | 5,015 | 1.5 | 4.7 | 33 | 72 | 1.91 (1.09 - 3.32) | 0.02 | 1.92 (1.10 - 3.36) | 0.02 |
| MDCD | 1,361 | 3,296 | 0.8 | 2.4 | 14 | 38 | 1.29 (0.57 - 2.86) | 0.53 | 1.17 (0.52 - 2.62) | 0.72 |
| Open Claims | 236,402 | 410,998 | 229.1 | 393.1 | 1,313 | 2,502 | 1.15 (1.04 - 1.26) | <0.01 | 1.25 (1.09 - 1.44) | <0.01 |
| OEHR | 16,368 | 31,327 | 5.3 | 16.1 | 74 | 193 | 1.26 (0.85 - 1.87) | 0.25 | 1.27 (0.86 - 1.89) | 0.25 |
| VA | 7,516 | 23,081 | 5.1 | 20.9 | 69 | 188 | 1.42 (0.94 - 2.11) | 0.09 | 1.42 (0.95 - 2.14) | 0.10 |
| SIDIAP | 4,288 | 19,572 | 6.3 | 44.5 | 29 | 186 | 0.98 (0.56 - 1.64) | 0.93 | 1.06 (0.62 - 1.83) | 0.81 |
| Meta-analysis | 304,043 | 559,339 | 279.5 | 541.8 | 1,839 | 3,720 | 1.18 (1.08 - 1.27) | <0.01 | 1.25 (1.10 - 1.42) | <0.01 |
| Meta-analysis (w/o USOC) | 67,641 | 148,341 | 50.4 | 148.8 | 526 | 1,218 | 1.24 (1.07 - 1.44) | <0.01 | 1.28 (1.10 - 1.49) | <0.01 |
| **PS-stratification / on-treatment TAR** | | | | | | | | | | |
| CCAE | 23,568 | 30,406 | 19.6 | 25.6 | 145 | 206 | 0.89 (0.71 - 1.11) | 0.30 | 0.92 (0.74 - 1.15) | 0.49 |
| DAG | 5,651 | 10,957 | 4.6 | 11.4 | 39 | 79 | 1.14 (0.74 - 1.75) | 0.55 | 1.10 (0.71 - 1.69) | 0.68 |
| ODOD | 16,888 | 20,567 | 11.6 | 16.1 | 184 | 267 | 1.21 (0.98 - 1.49) | 0.07 | 1.22 (0.99 - 1.51) | 0.08 |
| MDCR | 2,062 | 5,063 | 1.6 | 4.8 | 35 | 73 | 1.54 (0.98 - 2.38) | 0.06 | 1.59 (1.02 - 2.48) | 0.05 |
| MDCD | 1,443 | 3,308 | 0.9 | 2.4 | 14 | 38 | 1.10 (0.56 - 2.04) | 0.78 | 0.99 (0.52 - 1.90) | 0.90 |
| Open Claims | 322,505 | 411,312 | 280.1 | 393.3 | 1,595 | 2,511 | 1.08 (1.01 - 1.16) | 0.02 | 1.17 (1.01 - 1.35) | 0.04 |
| OEHR | 19,644 | 31,392 | 6.3 | 16.1 | 79 | 195 | 1.19 (0.89 - 1.57) | 0.24 | 1.19 (0.89 - 1.59) | 0.25 |
| VA | 12,060 | 24,052 | 7.2 | 22.3 | 100 | 206 | 1.69 (1.24 - 2.30) | <0.01 | 1.77 (1.30 - 2.42) | <0.01 |
| SIDIAP | 5,093 | 19,998 | 7.0 | 46.0 | 36 | 192 | 1.04 (0.68 - 1.56) | 0.85 | 1.07 (0.70 - 1.62) | 0.75 |
| Meta-analysis | 395,411 | 529,695 | 330.8 | 513.2 | 2,113 | 3,523 | 1.09 (1.03 - 1.16) | <0.01 | 1.15 (1.00 - 1.31) | 0.04 |
| Meta-analysis (w/o USOC) | 72,906 | 118,383 | 50.7 | 120.0 | 518 | 1,012 | 1.12 (0.97 - 1.29) | 0.14 | 1.14 (0.98 - 1.32) | 0.11 |
| **PS-matching / on-treatment TAR without escalation** | | | | | | | | | | |
| CCAE | 19,278 | 30,384 | 14.9 | 20.7 | 106 | 159 | 1.04 (0.75 - 1.43) | 0.83 | 1.10 (0.79 - 1.53) | 0.57 |
| DAG | 4,550 | 10,926 | 3.6 | 10.3 | 32 | 68 | 1.31 (0.72 - 2.32) | 0.37 | 1.26 (0.70 - 2.27) | 0.44 |
| ODOD | 12,051 | 20,464 | 8.0 | 13.1 | 135 | 216 | 1.53 (1.10 - 2.13) | 0.01 | 1.58 (1.09 - 2.28) | 0.03 |
| MDCR | 1,655 | 5,015 | 1.3 | 4.0 | 30 | 65 | 1.67 (0.94 - 2.93) | 0.08 | 1.69 (0.96 - 2.99) | 0.08 |
| MDCD | 1,361 | 3,296 | 0.7 | 1.8 | 13 | 24 | 1.62 (0.66 - 3.91) | 0.29 | 1.55 (0.64 - 3.77) | 0.35 |
| Open Claims | 236,402 | 410,998 | 192.3 | 304.9 | 1,128 | 1,972 | 1.20 (1.08 - 1.33) | <0.01 | 1.29 (1.11 - 1.50) | <0.01 |
| OEHR | 16,368 | 31,327 | 4.7 | 13.0 | 66 | 144 | 1.28 (0.84 - 1.96) | 0.25 | 1.29 (0.84 - 1.98) | 0.25 |
| VA | 7,516 | 23,081 | 4.4 | 17.0 | 58 | 156 | 1.32 (0.82 - 2.08) | 0.25 | 1.31 (0.82 - 2.08) | 0.26 |
| SIDIAP | 4,288 | 19,572 | 5.6 | 35.3 | 25 | 152 | 0.87 (0.46 - 1.53) | 0.64 | 1.02 (0.56 - 1.86) | 0.86 |
| Meta-analysis | 304,043 | 559,339 | 236.4 | 426.4 | 1,587 | 2,962 | 1.22 (1.11 - 1.33) | <0.01 | 1.29 (1.11 - 1.50) | <0.01 |
| Meta-analysis (w/o USOC) | 67,641 | 148,341 | 44.2 | 121.5 | 459 | 990 | 1.26 (1.07 - 1.48) | <0.01 | 1.30 (1.10 - 1.53) | <0.01 |
| **PS-stratification / on-treatment TAR without escalation** | | | | | | | | | | |
| CCAE | 23,568 | 30,406 | 17.1 | 20.7 | 126 | 159 | 0.95 (0.74 - 1.21) | 0.67 | 0.99 (0.77 - 1.26) | 0.89 |
| DAG | 5,651 | 10,957 | 4.3 | 10.3 | 36 | 68 | 1.22 (0.77 - 1.91) | 0.39 | 1.18 (0.75 - 1.86) | 0.49 |
| ODOD | 16,888 | 20,567 | 10.3 | 13.2 | 160 | 218 | 1.21 (0.96 - 1.51) | 0.10 | 1.20 (0.95 - 1.50) | 0.15 |
| MDCR | 2,062 | 5,063 | 1.5 | 4.1 | 31 | 66 | 1.46 (0.91 - 2.32) | 0.11 | 1.52 (0.95 - 2.44) | 0.09 |
| MDCD | 1,443 | 3,308 | 0.7 | 1.8 | 13 | 24 | 1.63 (0.77 - 3.28) | 0.19 | 1.49 (0.72 - 3.07) | 0.29 |
| Open Claims | 322,505 | 411,312 | 238.3 | 305.1 | 1,386 | 1,979 | 1.13 (1.04 - 1.22) | <0.01 | 1.22 (1.08 - 1.37) | <0.01 |
| OEHR | 19,644 | 31,392 | 5.5 | 13.1 | 70 | 145 | 1.32 (0.96 - 1.81) | 0.08 | 1.31 (0.96 - 1.80) | 0.10 |
| VA | 12,060 | 24,052 | 6.3 | 18.1 | 87 | 170 | 1.62 (1.16 - 2.27) | <0.01 | 1.69 (1.20 - 2.36) | <0.01 |
| SIDIAP | 5,093 | 19,998 | 6.2 | 36.4 | 32 | 157 | 1.02 (0.64 - 1.57) | 0.94 | 1.11 (0.71 - 1.75) | 0.65 |
| Meta-analysis | 395,411 | 529,695 | 283.2 | 402.9 | 1,841 | 2,792 | 1.13 (1.06 - 1.21) | <0.01 | 1.20 (1.08 - 1.32) | <0.01 |
| Meta-analysis (w/o USOC) | 72,906 | 118,383 | 44.9 | 97.8 | 455 | 813 | 1.15 (1.01 - 1.31) | 0.03 | 1.17 (1.03 - 1.34) | 0.03 |

##### 1.2.12 Acute pancreatitis

**Supplementary Table 38:** Exposures, outcomes and risk estimates for Acute pancreatitis in SGLT2I (male) (T) vs DPP4I (male) (C) new-user cohorts under four different propensity score (PS) adjustment / time-at-risk (TAR) designs across data sources. We report total patient exposure time in 1,000 patient-year (KYr) units, uncalibrated and calibrated hazard ratios (HRs) and their 95% confidence intervals (CIs). Greyed out entries did not pass study diagnostics and are included only for completeness.

| | Patients | | Exposure (KYr) | | Outcomes | | Uncalibrated | | Calibrated | |
|---|---|---|---|---|---|---|---|---|---|---|
| | T | C | T | C | T | C | HR (95% CI) | $p$ | HR (95% CI) | $p$ |
| **PS-matching / on-treatment TAR** | | | | | | | | | | |
| CCAE | 19,555 | 30,931 | 17.5 | 26.2 | 36 | 61 | 0.76 (0.43 - 1.33) | 0.35 | 0.82 (0.47 - 1.45) | 0.50 |
| DAG | 4,640 | 11,168 | 4.1 | 11.6 | 0 | 0 | | | | |
| ODOD | 12,395 | 21,291 | 9.5 | 16.8 | 25 | 47 | 1.12 (0.58 - 2.12) | 0.73 | 1.16 (0.61 - 2.22) | 0.67 |
| MDCR | 1,708 | 5,225 | 1.5 | 4.9 | <5 | 17 | 0.56 (0.08 - 2.59) | 0.52 | 0.57 (0.10 - 3.24) | 0.53 |
| MDCD | 1,404 | 3,403 | 0.9 | 2.5 | 6 | 15 | 1.62 (0.39 - 5.86) | 0.49 | 1.46 (0.38 - 5.64) | 0.59 |
| Open Claims | 240,896 | 421,406 | 233.9 | 404.3 | 305 | 803 | 0.71 (0.58 - 0.86) | <0.01 | 0.77 (0.62 - 0.96) | 0.02 |
| OEHR | 16,778 | 32,198 | 5.5 | 16.6 | 12 | 38 | 1.21 (0.51 - 2.78) | 0.66 | 1.22 (0.52 - 2.85) | 0.65 |
| VA | 7,703 | 23,745 | 5.3 | 21.5 | <10 | 38 | 0.23 (0.01 - 1.25) | 0.22 | 0.24 (0.02 - 2.34) | 0.22 |
| SIDIAP | 4,320 | 19,781 | 6.3 | 45.0 | <5 | 18 | 1.74 (0.32 - 7.91) | 0.50 | 1.89 (0.38 - 9.46) | 0.44 |
| Meta-analysis | 299,035 | 534,796 | 273.1 | 490.4 | <393 | 1,004 | 0.75 (0.62 - 0.90) | <0.01 | 0.80 (0.65 - 0.98) | 0.03 |
| Meta-analysis (w/o USOC) | 58,139 | 113,390 | 39.2 | 86.1 | <88 | 201 | 0.90 (0.62 - 1.30) | 0.58 | 0.93 (0.64 - 1.34) | 0.70 |
| **PS-stratification / on-treatment TAR** | | | | | | | | | | |
| CCAE | 23,890 | 30,952 | 19.9 | 26.2 | 42 | 61 | 0.81 (0.54 - 1.23) | 0.33 | 0.85 (0.56 - 1.28) | 0.44 |
| DAG | 5,763 | 11,200 | 4.7 | 11.7 | 0 | 0 | | | | |
| ODOD | 17,307 | 21,412 | 12.0 | 16.9 | 29 | 47 | 1.12 (0.67 - 1.84) | 0.66 | 1.13 (0.68 - 1.87) | 0.64 |
| MDCR | 2,119 | 5,282 | 1.7 | 5.0 | <5 | 17 | 0.51 (0.08 - 1.89) | 0.40 | 0.52 (0.11 - 2.58) | 0.43 |
| MDCD | 1,481 | 3,416 | 0.9 | 2.5 | 6 | 16 | 1.48 (0.50 - 3.87) | 0.45 | 1.34 (0.48 - 3.72) | 0.58 |
| Open Claims | 327,945 | 421,728 | 285.4 | 404.5 | 377 | 804 | 0.78 (0.68 - 0.89) | <0.01 | 0.84 (0.70 - 1.01) | 0.08 |
| OEHR | 20,108 | 32,269 | 6.4 | 16.7 | 13 | 38 | 1.15 (0.57 - 2.20) | 0.69 | 1.15 (0.58 - 2.27) | 0.69 |
| VA | 12,372 | 24,770 | 7.4 | 23.0 | <10 | 40 | 0.46 (0.12 - 1.38) | 0.21 | 0.48 (0.14 - 1.63) | 0.24 |
| SIDIAP | 5,142 | 20,218 | 7.0 | 46.6 | <5 | 20 | 1.73 (0.39 - 5.61) | 0.42 | 1.78 (0.47 - 6.79) | 0.40 |
| Meta-analysis | 391,369 | 511,643 | 325.4 | 469.2 | <466 | 967 | 0.81 (0.71 - 0.93) | <0.01 | 0.85 (0.72 - 1.02) | 0.08 |
| Meta-analysis (w/o USOC) | 63,424 | 89,915 | 40.0 | 64.8 | <89 | 163 | 0.94 (0.71 - 1.26) | 0.69 | 0.96 (0.72 - 1.28) | 0.79 |
| **PS-matching / on-treatment TAR without escalation** | | | | | | | | | | |
| CCAE | 19,555 | 30,931 | 15.1 | 21.2 | 29 | 45 | 0.70 (0.36 - 1.31) | 0.28 | 0.75 (0.39 - 1.42) | 0.38 |
| DAG | 4,640 | 11,168 | 3.7 | 10.5 | 0 | 0 | | | | |
| ODOD | 12,395 | 21,291 | 8.3 | 13.7 | 19 | 33 | 1.34 (0.63 - 2.83) | 0.44 | 1.38 (0.64 - 2.98) | 0.43 |
| MDCR | 1,708 | 5,225 | 1.3 | 4.2 | <5 | 16 | 0.56 (0.08 - 2.55) | 0.51 | 0.57 (0.10 - 3.21) | 0.52 |
| MDCD | 1,404 | 3,403 | 0.7 | 1.9 | 5 | 10 | 3.25 (0.70 - 15.0) | 0.13 | 3.10 (0.67 - 14.4) | 0.15 |
| Open Claims | 240,896 | 421,406 | 196.3 | 313.7 | 239 | 587 | 0.70 (0.56 - 0.87) | <0.01 | 0.75 (0.59 - 0.95) | 0.02 |
| OEHR | 16,778 | 32,198 | 4.8 | 13.5 | 10 | 26 | 1.17 (0.44 - 2.94) | 0.75 | 1.18 (0.46 - 3.04) | 0.74 |
| VA | 7,703 | 23,745 | 4.5 | 17.5 | <10 | 27 | 0.26 (0.01 - 1.41) | 0.25 | 0.26 (0.03 - 2.60) | 0.25 |
| SIDIAP | 4,320 | 19,781 | 5.6 | 35.8 | <5 | 16 | 1.01 (0.14 - 5.07) | 0.99 | 1.19 (0.20 - 7.26) | 0.85 |
| Meta-analysis | 297,327 | 529,571 | 229.0 | 379.6 | <307 | 718 | 0.77 (0.59 - 0.99) | 0.04 | 0.81 (0.61 - 1.07) | 0.14 |
| Meta-analysis (w/o USOC) | 56,431 | 108,165 | 32.7 | 65.9 | <68 | 131 | 0.93 (0.60 - 1.43) | 0.73 | 0.96 (0.62 - 1.47) | 0.84 |
| **PS-stratification / on-treatment TAR without escalation** | | | | | | | | | | |
| CCAE | 23,890 | 30,952 | 17.4 | 21.2 | 34 | 45 | 0.79 (0.49 - 1.26) | 0.33 | 0.82 (0.51 - 1.32) | 0.42 |
| DAG | 5,763 | 11,200 | 4.3 | 10.6 | 0 | 0 | | | | |
| ODOD | 17,307 | 21,412 | 10.7 | 13.8 | 23 | 33 | 1.16 (0.65 - 2.06) | 0.61 | 1.15 (0.65 - 2.06) | 0.63 |
| MDCR | 2,119 | 5,282 | 1.5 | 4.2 | <5 | 16 | 0.51 (0.08 - 1.93) | 0.41 | 0.53 (0.11 - 2.64) | 0.44 |
| MDCD | 1,481 | 3,416 | 0.7 | 1.9 | 5 | 11 | 1.87 (0.54 - 5.68) | 0.30 | 1.72 (0.53 - 5.57) | 0.37 |
| Open Claims | 327,945 | 421,728 | 242.8 | 313.8 | 306 | 588 | 0.79 (0.68 - 0.92) | <0.01 | 0.85 (0.72 - 1.02) | 0.09 |
| OEHR | 20,108 | 32,269 | 5.7 | 13.5 | 11 | 26 | 1.17 (0.53 - 2.44) | 0.69 | 1.16 (0.54 - 2.49) | 0.71 |
| VA | 12,372 | 24,770 | 6.4 | 18.7 | <10 | 29 | 0.43 (0.09 - 1.49) | 0.24 | 0.45 (0.11 - 1.82) | 0.26 |
| SIDIAP | 5,142 | 20,218 | 6.3 | 37.0 | <5 | 18 | 1.59 (0.35 - 5.24) | 0.50 | 1.75 (0.45 - 6.75) | 0.42 |
| Meta-analysis | 391,369 | 511,643 | 278.0 | 366.6 | <379 | 708 | 0.81 (0.71 - 0.93) | <0.01 | 0.86 (0.74 - 1.01) | 0.06 |
| Meta-analysis (w/o USOC) | 63,424 | 89,915 | 35.2 | 52.8 | <73 | 120 | 0.94 (0.68 - 1.30) | 0.70 | 0.96 (0.69 - 1.33) | 0.80 |

##### 1.2.13 Bone fracture

**Supplementary Table 39:** Exposures, outcomes and risk estimates for Bone fracture in SGLT2I (male) (T) vs DPP4I (male) (C) new-user cohorts under four different propensity score (PS) adjustment / time-at-risk (TAR) designs across data sources. We report total patient exposure time in 1,000 patient-year (KYr) units, uncalibrated and calibrated hazard ratios (HRs) and their 95% confidence intervals (CIs). Greyed out entries did not pass study diagnostics and are included only for completeness.

| | Patients | | Exposure (KYr) | | Outcomes | | Uncalibrated | | Calibrated | |
|---|---|---|---|---|---|---|---|---|---|---|
| | T | C | T | C | T | C | HR (95% CI) | $p$ | HR (95% CI) | $p$ |
| **PS-matching / on-treatment TAR** | | | | | | | | | | |
| CCAE | 18,252 | 28,862 | 16.0 | 24.1 | 258 | 342 | 1.10 (0.87 - 1.39) | 0.41 | 1.19 (0.94 - 1.50) | 0.16 |
| DAG | 4,145 | 9,946 | 3.6 | 10.2 | 51 | 174 | 0.93 (0.61 - 1.40) | 0.74 | 0.91 (0.60 - 1.39) | 0.66 |
| ODOD | 11,627 | 19,777 | 8.7 | 15.3 | 173 | 313 | 1.06 (0.80 - 1.39) | 0.68 | 1.09 (0.83 - 1.45) | 0.58 |
| MDCR | 1,574 | 4,723 | 1.4 | 4.4 | 37 | 111 | 1.13 (0.68 - 1.83) | 0.63 | 1.13 (0.69 - 1.86) | 0.64 |
| MDCD | 1,250 | 3,021 | 0.8 | 2.2 | 35 | 61 | 1.43 (0.80 - 2.51) | 0.22 | 1.29 (0.73 - 2.28) | 0.40 |
| Open Claims | 218,226 | 382,506 | 210.7 | 363.0 | 2,328 | 4,935 | 0.95 (0.88 - 1.02) | 0.17 | 1.03 (0.91 - 1.17) | 0.63 |
| OEHR | 15,884 | 30,462 | 5.2 | 15.6 | 62 | 240 | 0.83 (0.56 - 1.21) | 0.33 | 0.83 (0.57 - 1.23) | 0.37 |
| VA | 6,972 | 21,479 | 4.8 | 19.5 | 34 | 183 | 0.61 (0.35 - 1.01) | 0.06 | 0.61 (0.36 - 1.04) | 0.07 |
| SIDIAP | 3,903 | 18,011 | 5.6 | 40.8 | 51 | 394 | 1.32 (0.91 - 1.90) | 0.14 | 1.44 (0.99 - 2.10) | 0.07 |
| Meta-analysis | 282,383 | 522,911 | 257.8 | 501.9 | 3,004 | 6,747 | 0.97 (0.90 - 1.04) | 0.34 | 1.03 (0.91 - 1.16) | 0.67 |
| Meta-analysis (w/o USOC) | 64,157 | 140,405 | 47.1 | 138.9 | 676 | 1,812 | 0.99 (0.84 - 1.17) | 0.93 | 1.02 (0.86 - 1.21) | 0.80 |
| **PS-stratification / on-treatment TAR** | | | | | | | | | | |
| CCAE | 22,298 | 28,882 | 18.2 | 24.1 | 296 | 342 | 1.17 (1.00 - 1.38) | 0.05 | 1.22 (1.03 - 1.44) | 0.02 |
| DAG | 5,164 | 9,967 | 4.2 | 10.2 | 56 | 175 | 0.97 (0.69 - 1.35) | 0.87 | 0.93 (0.67 - 1.30) | 0.68 |
| ODOD | 16,194 | 19,885 | 11.1 | 15.4 | 211 | 317 | 1.01 (0.83 - 1.22) | 0.96 | 1.01 (0.84 - 1.23) | 0.86 |
| MDCR | 1,967 | 4,786 | 1.6 | 4.5 | 38 | 113 | 1.08 (0.71 - 1.60) | 0.72 | 1.11 (0.74 - 1.68) | 0.62 |
| MDCD | 1,321 | 3,032 | 0.8 | 2.2 | 35 | 62 | 1.83 (1.17 - 2.81) | <0.01 | 1.65 (1.06 - 2.56) | 0.03 |
| Open Claims | 295,535 | 382,793 | 256.4 | 363.2 | 2,775 | 4,949 | 0.94 (0.89 - 0.99) | 0.01 | 1.01 (0.88 - 1.16) | 0.87 |
| OEHR | 18,979 | 30,527 | 6.1 | 15.6 | 70 | 242 | 0.86 (0.64 - 1.13) | 0.29 | 0.86 (0.64 - 1.15) | 0.32 |
| VA | 11,119 | 22,469 | 6.7 | 20.9 | 52 | 209 | 0.75 (0.51 - 1.09) | 0.14 | 0.79 (0.54 - 1.15) | 0.23 |
| SIDIAP | 4,634 | 18,465 | 6.3 | 42.4 | 55 | 417 | 1.10 (0.79 - 1.50) | 0.55 | 1.13 (0.82 - 1.56) | 0.47 |
| Meta-analysis | 364,771 | 495,305 | 303.8 | 475.4 | 3,501 | 6,555 | 1.00 (0.91 - 1.10) | 0.99 | 1.05 (0.91 - 1.23) | 0.50 |
| Meta-analysis (w/o USOC) | 69,236 | 112,512 | 47.4 | 112.2 | 726 | 1,606 | 1.05 (0.95 - 1.16) | 0.30 | 1.07 (0.97 - 1.19) | 0.22 |
| **PS-matching / on-treatment TAR without escalation** | | | | | | | | | | |
| CCAE | 18,252 | 28,862 | 13.9 | 19.6 | 218 | 279 | 1.17 (0.91 - 1.50) | 0.22 | 1.24 (0.97 - 1.60) | 0.10 |
| DAG | 4,145 | 9,946 | 3.3 | 9.2 | 47 | 161 | 0.91 (0.58 - 1.41) | 0.68 | 0.88 (0.56 - 1.38) | 0.60 |
| ODOD | 11,627 | 19,777 | 7.7 | 12.6 | 150 | 251 | 1.03 (0.76 - 1.39) | 0.83 | 1.07 (0.76 - 1.50) | 0.76 |
| MDCR | 1,574 | 4,723 | 1.2 | 3.8 | 31 | 95 | 1.21 (0.71 - 2.01) | 0.48 | 1.22 (0.72 - 2.06) | 0.48 |
| MDCD | 1,250 | 3,021 | 0.6 | 1.7 | 31 | 44 | 1.49 (0.79 - 2.79) | 0.21 | 1.43 (0.76 - 2.69) | 0.29 |
| Open Claims | 218,226 | 382,506 | 177.0 | 282.3 | 1,930 | 3,805 | 0.95 (0.87 - 1.03) | 0.21 | 1.02 (0.89 - 1.16) | 0.81 |
| OEHR | 15,884 | 30,462 | 4.5 | 12.6 | 55 | 181 | 0.86 (0.55 - 1.31) | 0.49 | 0.86 (0.56 - 1.34) | 0.53 |
| VA | 6,972 | 21,479 | 4.1 | 15.9 | 30 | 140 | 0.70 (0.39 - 1.20) | 0.21 | 0.69 (0.39 - 1.22) | 0.21 |
| SIDIAP | 3,903 | 18,011 | 5.0 | 32.5 | 46 | 311 | 1.45 (0.97 - 2.13) | 0.07 | 1.70 (1.14 - 2.54) | 0.02 |
| Meta-analysis | 282,383 | 522,911 | 218.3 | 396.0 | 2,516 | 5,266 | 1.00 (0.90 - 1.12) | 0.96 | 1.06 (0.90 - 1.25) | 0.48 |
| Meta-analysis (w/o USOC) | 64,157 | 140,405 | 41.3 | 113.7 | 586 | 1,461 | 1.05 (0.90 - 1.23) | 0.55 | 1.08 (0.92 - 1.27) | 0.36 |
| **PS-stratification / on-treatment TAR without escalation** | | | | | | | | | | |
| CCAE | 22,298 | 28,882 | 15.9 | 19.6 | 254 | 279 | 1.16 (0.97 - 1.39) | 0.10 | 1.21 (1.01 - 1.45) | 0.05 |
| DAG | 5,164 | 9,967 | 3.8 | 9.3 | 52 | 162 | 0.97 (0.68 - 1.37) | 0.88 | 0.94 (0.66 - 1.34) | 0.71 |
| ODOD | 16,194 | 19,885 | 9.8 | 12.7 | 186 | 254 | 1.06 (0.86 - 1.30) | 0.60 | 1.05 (0.85 - 1.29) | 0.68 |
| MDCR | 1,967 | 4,786 | 1.4 | 3.8 | 32 | 97 | 1.10 (0.70 - 1.69) | 0.67 | 1.14 (0.73 - 1.78) | 0.57 |
| MDCD | 1,321 | 3,032 | 0.7 | 1.7 | 31 | 45 | 1.95 (1.20 - 3.15) | <0.01 | 1.79 (1.10 - 2.91) | 0.02 |
| Open Claims | 295,535 | 382,793 | 218.3 | 282.5 | 2,332 | 3,817 | 0.94 (0.89 - 0.99) | 0.03 | 1.02 (0.91 - 1.13) | 0.79 |
| OEHR | 18,979 | 30,527 | 5.3 | 12.7 | 62 | 181 | 0.92 (0.67 - 1.25) | 0.58 | 0.91 (0.66 - 1.25) | 0.57 |
| VA | 11,119 | 22,469 | 5.8 | 17.0 | 46 | 161 | 0.82 (0.53 - 1.23) | 0.35 | 0.85 (0.56 - 1.29) | 0.46 |
| SIDIAP | 4,634 | 18,465 | 5.6 | 33.7 | 50 | 326 | 1.13 (0.80 - 1.57) | 0.49 | 1.24 (0.88 - 1.74) | 0.25 |
| Meta-analysis | 364,771 | 495,305 | 260.3 | 374.1 | 2,968 | 5,116 | 1.01 (0.92 - 1.11) | 0.80 | 1.07 (0.95 - 1.21) | 0.28 |
| Meta-analysis (w/o USOC) | 69,236 | 112,512 | 41.9 | 91.7 | 636 | 1,299 | 1.08 (0.97 - 1.20) | 0.18 | 1.10 (0.98 - 1.23) | 0.14 |

##### 1.2.14 Diabetic ketoacidosis

**Supplementary Table 40:** Exposures, outcomes and risk estimates for Diabetic ketoacidosis in SGLT2I (male) (T) vs DPP4I (male) (C) new-user cohorts under four different propensity score (PS) adjustment / time-at-risk (TAR) designs across data sources. We report total patient exposure time in 1,000 patient-year (KYr) units, uncalibrated and calibrated hazard ratios (HRs) and their 95% confidence intervals (CIs). Greyed out entries did not pass study diagnostics and are included only for completeness.

| | Patients | | Exposure (KYr) | | Outcomes | | Uncalibrated | | Calibrated | |
|---|---|---|---|---|---|---|---|---|---|---|
| | T | C | T | C | T | C | HR (95% CI) | $p$ | HR (95% CI) | $p$ |
| **PS-matching / on-treatment TAR** | | | | | | | | | | |
| CCAE | 19,732 | 31,147 | 17.6 | 26.4 | 36 | 24 | 2.29 (1.16 - 4.71) | 0.02 | 2.47 (1.22 - 4.96) | 0.01 |
| DAG | 4,640 | 11,168 | 4.1 | 11.6 | 0 | 0 | | | | |
| ODOD | 12,497 | 21,443 | 9.6 | 16.9 | 16 | 28 | 1.41 (0.60 - 3.29) | 0.43 | 1.46 (0.62 - 3.43) | 0.40 |
| MDCR | 1,717 | 5,257 | 1.5 | 5.0 | <5 | 5 | | | | |
| MDCD | 1,418 | 3,456 | 0.9 | 2.5 | <5 | 5 | | 0.09 | | 0.10 |
| Open Claims | 243,735 | 425,574 | 236.8 | 408.1 | 233 | 302 | 1.39 (1.09 - 1.76) | <0.01 | 1.51 (1.17 - 1.96) | <0.01 |
| OEHR | 16,856 | 32,318 | 5.5 | 16.7 | 13 | 23 | 1.91 (0.71 - 5.44) | 0.21 | 1.92 (0.69 - 5.31) | 0.21 |
| VA | 7,793 | 23,947 | 5.3 | 21.7 | <10 | 10 | 2.46 (0.66 - 8.85) | 0.17 | 2.48 (0.68 - 9.06) | 0.17 |
| SIDIAP | 4,340 | 19,836 | 6.3 | 45.1 | 0 | 0 | | | | |
| Meta-analysis | 292,820 | 510,482 | 269.5 | 468.1 | 298 | 377 | 1.48 (1.19 - 1.85) | <0.01 | 1.57 (1.23 - 2.01) | <0.01 |
| Meta-analysis (w/o USOC) | 49,085 | 84,908 | 32.7 | 60.0 | 65 | 75 | 1.89 (1.17 - 3.05) | <0.01 | 1.95 (1.21 - 3.14) | <0.01 |
| **PS-stratification / on-treatment TAR** | | | | | | | | | | |
| CCAE | 24,108 | 31,170 | 20.1 | 26.4 | 40 | 24 | 1.97 (1.17 - 3.41) | 0.01 | 2.05 (1.20 - 3.50) | <0.01 |
| DAG | 5,763 | 11,200 | 4.7 | 11.7 | 0 | 0 | | | | |
| ODOD | 17,459 | 21,565 | 12.1 | 17.0 | 23 | 28 | 1.16 (0.62 - 2.17) | 0.65 | 1.17 (0.62 - 2.19) | 0.63 |
| MDCR | 2,135 | 5,316 | 1.7 | 5.0 | <5 | 5 | 1.25 (0.06 - 9.69) | 0.86 | 1.29 (0.10 - 16.4) | 0.84 |
| MDCD | 1,499 | 3,470 | 0.9 | 2.6 | <5 | 5 | 2.23 (0.52 - 9.02) | 0.27 | 2.01 (0.48 - 8.36) | 0.34 |
| Open Claims | 332,197 | 425,900 | 289.1 | 408.4 | 307 | 302 | 1.44 (1.21 - 1.71) | <0.01 | 1.55 (1.25 - 1.93) | <0.01 |
| OEHR | 20,218 | 32,389 | 6.5 | 16.8 | 14 | 23 | 1.62 (0.75 - 3.42) | 0.22 | 1.62 (0.76 - 3.48) | 0.21 |
| VA | 12,523 | 24,988 | 7.5 | 23.2 | <10 | 11 | 2.77 (0.88 - 8.65) | 0.08 | 2.90 (0.93 - 9.08) | 0.07 |
| SIDIAP | 5,168 | 20,274 | 7.1 | 46.8 | 0 | 0 | | | | |
| Meta-analysis | 393,982 | 511,024 | 327.8 | 468.5 | 384 | 377 | 1.47 (1.25 - 1.72) | <0.01 | 1.54 (1.27 - 1.88) | <0.01 |
| Meta-analysis (w/o USOC) | 61,785 | 85,124 | 38.7 | 60.2 | 77 | 75 | 1.58 (1.11 - 2.27) | 0.01 | 1.62 (1.13 - 2.32) | <0.01 |
| **PS-matching / on-treatment TAR without escalation** | | | | | | | | | | |
| CCAE | 19,732 | 31,147 | 15.3 | 21.3 | 29 | 19 | 2.39 (1.16 - 5.18) | 0.02 | 2.53 (1.20 - 5.36) | 0.02 |
| DAG | 4,640 | 11,168 | 3.7 | 10.5 | 0 | 0 | | | | |
| ODOD | 12,497 | 21,443 | 8.4 | 13.8 | 13 | 21 | 1.70 (0.67 - 4.35) | 0.27 | 1.76 (0.68 - 4.55) | 0.26 |
| MDCR | 1,717 | 5,257 | 1.4 | 4.2 | 0 | <5 | | | | |
| MDCD | 1,418 | 3,456 | 0.7 | 1.9 | <5 | 5 | | 0.12 | | 0.13 |
| Open Claims | 243,735 | 425,574 | 198.5 | 316.6 | 176 | 217 | 1.37 (1.05 - 1.79) | 0.02 | 1.47 (1.10 - 1.96) | 0.01 |
| OEHR | 16,856 | 32,318 | 4.8 | 13.5 | 12 | 16 | 2.23 (0.72 - 7.78) | 0.19 | 2.25 (0.68 - 7.42) | 0.18 |
| VA | 7,793 | 23,947 | 4.5 | 17.7 | <10 | <10 | 4.27 (1.05 - 18.5) | 0.05 | 4.25 (1.01 - 17.9) | 0.05 |
| SIDIAP | 4,340 | 19,836 | 5.6 | 35.8 | 0 | 0 | | | | |
| Meta-analysis | 292,820 | 510,482 | 227.0 | 365.3 | 230 | 273 | 1.59 (1.15 - 2.21) | <0.01 | 1.68 (1.19 - 2.39) | <0.01 |
| Meta-analysis (w/o USOC) | 49,085 | 84,908 | 28.5 | 48.7 | 54 | 56 | 2.12 (1.25 - 3.58) | <0.01 | 2.18 (1.29 - 3.70) | <0.01 |
| **PS-stratification / on-treatment TAR without escalation** | | | | | | | | | | |
| CCAE | 24,108 | 31,170 | 17.5 | 21.4 | 33 | 19 | 1.85 (1.03 - 3.40) | 0.04 | 1.92 (1.06 - 3.48) | 0.03 |
| DAG | 5,763 | 11,200 | 4.3 | 10.6 | 0 | 0 | | | | |
| ODOD | 17,459 | 21,565 | 10.8 | 13.9 | 20 | 21 | 1.22 (0.60 - 2.48) | 0.58 | 1.21 (0.59 - 2.45) | 0.60 |
| MDCR | 2,135 | 5,316 | 1.5 | 4.3 | 0 | <5 | | | | |
| MDCD | 1,499 | 3,470 | 0.8 | 1.9 | <5 | 5 | 1.57 (0.31 - 6.90) | 0.57 | 1.44 (0.30 - 6.85) | 0.65 |
| Open Claims | 332,197 | 425,900 | 245.8 | 316.8 | 243 | 217 | 1.50 (1.23 - 1.84) | <0.01 | 1.62 (1.30 - 2.03) | <0.01 |
| OEHR | 20,218 | 32,389 | 5.7 | 13.5 | 13 | 16 | 1.73 (0.75 - 3.95) | 0.20 | 1.72 (0.75 - 3.94) | 0.20 |
| VA | 12,523 | 24,988 | 6.5 | 18.9 | <10 | <10 | 6.23 (1.62 - 26.5) | 0.01 | 6.47 (1.60 - 26.2) | <0.01 |
| SIDIAP | 5,168 | 20,274 | 6.3 | 37.1 | 0 | 0 | | | | |
| Meta-analysis | 393,982 | 511,024 | 279.8 | 365.6 | 309 | 273 | 1.52 (1.27 - 1.82) | <0.01 | 1.60 (1.32 - 1.95) | <0.01 |
| Meta-analysis (w/o USOC) | 61,785 | 85,124 | 34.0 | 48.8 | 66 | 56 | 1.59 (1.07 - 2.38) | 0.02 | 1.63 (1.09 - 2.43) | 0.02 |

##### 1.2.15 Diarrhea

**Supplementary Table 41:** Exposures, outcomes and risk estimates for Diarrhea in SGLT2I (male) (T) vs DPP4I (male) (C) new-user cohorts under four different propensity score (PS) adjustment / time-at-risk (TAR) designs across data sources. We report total patient exposure time in 1,000 patient-year (KYr) units, uncalibrated and calibrated hazard ratios (HRs) and their 95% confidence intervals (CIs). Greyed out entries did not pass study diagnostics and are included only for completeness.

| | Patients | | Exposure (KYr) | | Outcomes | | Uncalibrated | | Calibrated | |
|---|---|---|---|---|---|---|---|---|---|---|
| | T | C | T | C | T | C | HR (95% CI) | $p$ | HR (95% CI) | $p$ |
| **PS-matching / on-treatment TAR** | | | | | | | | | | |
| CCAE | 17,875 | 28,266 | 15.7 | 23.4 | 315 | 540 | 0.78 (0.64 - 0.96) | 0.02 | 0.84 (0.69 - 1.03) | 0.12 |
| DAG | 4,563 | 11,024 | 4.0 | 11.5 | 6 | 20 | 1.58 (0.45 - 5.09) | 0.46 | 1.55 (0.46 - 5.21) | 0.48 |
| ODOD | 11,129 | 18,887 | 8.4 | 14.5 | 222 | 452 | 0.92 (0.73 - 1.16) | 0.50 | 0.95 (0.75 - 1.21) | 0.74 |
| MDCR | 1,531 | 4,626 | 1.3 | 4.3 | 27 | 149 | 0.73 (0.41 - 1.24) | 0.27 | 0.73 (0.42 - 1.28) | 0.29 |
| MDCD | 1,215 | 2,921 | 0.7 | 2.1 | 34 | 94 | 1.30 (0.75 - 2.24) | 0.34 | 1.17 (0.68 - 2.04) | 0.59 |
| Open Claims | 219,670 | 383,846 | 211.8 | 363.1 | 2,498 | 5,465 | 0.85 (0.79 - 0.91) | <0.01 | 0.93 (0.82 - 1.05) | 0.28 |
| OEHR | 15,443 | 29,633 | 5.0 | 14.9 | 116 | 400 | 0.98 (0.73 - 1.29) | 0.87 | 0.98 (0.74 - 1.31) | 0.83 |
| VA | 6,699 | 20,641 | 4.5 | 18.5 | 69 | 369 | 0.61 (0.42 - 0.87) | <0.01 | 0.62 (0.43 - 0.88) | 0.01 |
| SIDIAP | 4,182 | 18,936 | 6.1 | 42.8 | 36 | 412 | 0.49 (0.31 - 0.73) | <0.01 | 0.53 (0.35 - 0.82) | <0.01 |
| Meta-analysis | 282,764 | 522,125 | 258.6 | 500.7 | 3,311 | 7,952 | 0.80 (0.71 - 0.90) | <0.01 | 0.85 (0.73 - 0.99) | 0.04 |
| Meta-analysis (w/o USOC) | 63,094 | 138,279 | 46.7 | 137.7 | 813 | 2,487 | 0.77 (0.65 - 0.91) | <0.01 | 0.79 (0.67 - 0.95) | 0.01 |
| **PS-stratification / on-treatment TAR** | | | | | | | | | | |
| CCAE | 21,758 | 28,286 | 17.9 | 23.4 | 367 | 540 | 0.87 (0.75 - 1.00) | 0.04 | 0.90 (0.78 - 1.04) | 0.17 |
| DAG | 5,660 | 11,056 | 4.6 | 11.5 | 6 | 20 | 0.92 (0.30 - 2.50) | 0.87 | 0.88 (0.30 - 2.55) | 0.81 |
| ODOD | 15,599 | 19,076 | 10.6 | 14.6 | 286 | 462 | 0.89 (0.76 - 1.05) | 0.18 | 0.90 (0.77 - 1.07) | 0.27 |
| MDCR | 1,906 | 4,679 | 1.5 | 4.3 | 30 | 149 | 0.64 (0.41 - 0.96) | 0.04 | 0.66 (0.43 - 1.00) | 0.06 |
| MDCD | 1,283 | 2,930 | 0.8 | 2.1 | 34 | 95 | 1.11 (0.72 - 1.67) | 0.64 | 1.00 (0.65 - 1.53) | 0.86 |
| Open Claims | 299,156 | 384,186 | 258.9 | 363.3 | 3,000 | 5,481 | 0.88 (0.84 - 0.92) | <0.01 | 0.95 (0.83 - 1.09) | 0.47 |
| OEHR | 18,506 | 29,693 | 5.8 | 15.0 | 134 | 402 | 0.94 (0.76 - 1.16) | 0.56 | 0.94 (0.76 - 1.17) | 0.62 |
| VA | 10,635 | 21,651 | 6.4 | 20.0 | 107 | 398 | 0.72 (0.55 - 0.94) | 0.02 | 0.76 (0.58 - 0.99) | 0.05 |
| SIDIAP | 4,981 | 19,435 | 6.8 | 44.5 | 46 | 436 | 0.57 (0.40 - 0.79) | <0.01 | 0.58 (0.42 - 0.82) | <0.01 |
| Meta-analysis | 367,566 | 496,411 | 306.2 | 476.7 | 3,869 | 7,490 | 0.87 (0.84 - 0.91) | <0.01 | 0.92 (0.81 - 1.04) | 0.18 |
| Meta-analysis (w/o USOC) | 68,410 | 112,225 | 47.3 | 113.4 | 869 | 2,009 | 0.83 (0.72 - 0.94) | <0.01 | 0.84 (0.73 - 0.97) | 0.02 |
| **PS-matching / on-treatment TAR without escalation** | | | | | | | | | | |
| CCAE | 17,875 | 28,266 | 13.7 | 19.1 | 263 | 435 | 0.76 (0.61 - 0.94) | 0.01 | 0.81 (0.65 - 1.00) | 0.06 |
| DAG | 4,563 | 11,024 | 3.7 | 10.4 | 6 | 18 | 1.47 (0.42 - 4.75) | 0.53 | 1.42 (0.42 - 4.81) | 0.57 |
| ODOD | 11,129 | 18,887 | 7.4 | 11.9 | 188 | 373 | 0.93 (0.72 - 1.20) | 0.57 | 0.96 (0.71 - 1.29) | 0.81 |
| MDCR | 1,531 | 4,626 | 1.2 | 3.7 | 24 | 129 | 0.74 (0.40 - 1.29) | 0.30 | 0.75 (0.41 - 1.34) | 0.35 |
| MDCD | 1,215 | 2,921 | 0.6 | 1.6 | 29 | 67 | 1.57 (0.85 - 2.85) | 0.14 | 1.50 (0.82 - 2.75) | 0.21 |
| Open Claims | 219,670 | 383,846 | 178.3 | 283.1 | 2,022 | 4,233 | 0.87 (0.80 - 0.94) | <0.01 | 0.93 (0.82 - 1.06) | 0.34 |
| OEHR | 15,443 | 29,633 | 4.4 | 12.2 | 102 | 290 | 1.08 (0.79 - 1.47) | 0.63 | 1.09 (0.79 - 1.49) | 0.62 |
| VA | 6,699 | 20,641 | 3.9 | 15.2 | 60 | 303 | 0.63 (0.42 - 0.92) | 0.02 | 0.63 (0.43 - 0.92) | 0.02 |
| SIDIAP | 4,182 | 18,936 | 5.4 | 34.0 | 31 | 336 | 0.46 (0.28 - 0.71) | <0.01 | 0.54 (0.34 - 0.85) | 0.01 |
| Meta-analysis | 282,764 | 522,125 | 219.5 | 396.1 | 2,717 | 6,237 | 0.80 (0.69 - 0.93) | <0.01 | 0.85 (0.70 - 1.03) | 0.10 |
| Meta-analysis (w/o USOC) | 63,094 | 138,279 | 41.1 | 113.0 | 695 | 2,004 | 0.78 (0.64 - 0.94) | 0.01 | 0.80 (0.66 - 0.97) | 0.03 |
| **PS-stratification / on-treatment TAR without escalation** | | | | | | | | | | |
| CCAE | 21,758 | 28,286 | 15.7 | 19.1 | 307 | 435 | 0.85 (0.73 - 0.99) | 0.03 | 0.88 (0.75 - 1.03) | 0.13 |
| DAG | 5,660 | 11,056 | 4.3 | 10.4 | 6 | 18 | 0.95 (0.30 - 2.63) | 0.92 | 0.92 (0.31 - 2.70) | 0.87 |
| ODOD | 15,599 | 19,076 | 9.5 | 12.1 | 244 | 382 | 0.87 (0.73 - 1.04) | 0.12 | 0.86 (0.72 - 1.03) | 0.13 |
| MDCR | 1,906 | 4,679 | 1.3 | 3.7 | 27 | 129 | 0.60 (0.38 - 0.93) | 0.03 | 0.63 (0.40 - 0.98) | 0.05 |
| MDCD | 1,283 | 2,930 | 0.6 | 1.6 | 29 | 68 | 1.23 (0.76 - 1.96) | 0.39 | 1.13 (0.70 - 1.82) | 0.63 |
| Open Claims | 299,156 | 384,186 | 221.0 | 283.3 | 2,473 | 4,246 | 0.87 (0.83 - 0.92) | <0.01 | 0.94 (0.85 - 1.05) | 0.32 |
| OEHR | 18,506 | 29,693 | 5.2 | 12.2 | 117 | 290 | 1.09 (0.86 - 1.38) | 0.46 | 1.09 (0.86 - 1.38) | 0.52 |
| VA | 10,635 | 21,651 | 5.5 | 16.3 | 95 | 321 | 0.72 (0.54 - 0.95) | 0.02 | 0.75 (0.56 - 1.00) | 0.06 |
| SIDIAP | 4,981 | 19,435 | 6.1 | 35.3 | 41 | 353 | 0.52 (0.36 - 0.74) | <0.01 | 0.57 (0.40 - 0.82) | <0.01 |
| Meta-analysis | 367,566 | 496,411 | 263.0 | 376.2 | 3,215 | 5,853 | 0.82 (0.70 - 0.97) | 0.02 | 0.87 (0.73 - 1.04) | 0.13 |
| Meta-analysis (w/o USOC) | 68,410 | 112,225 | 42.0 | 92.8 | 742 | 1,607 | 0.81 (0.67 - 0.98) | 0.03 | 0.83 (0.68 - 1.00) | 0.06 |

##### 1.2.16 Genitourinary infection

**Supplementary Table 42:** Exposures, outcomes and risk estimates for Genitourinary infection in SGLT2I (male) (T) vs DPP4I (male) (C) new-user cohorts under four different propensity score (PS) adjustment / time-at-risk (TAR) designs across data sources. We report total patient exposure time in 1,000 patient-year (KYr) units, uncalibrated and calibrated hazard ratios (HRs) and their 95% confidence intervals (CIs). Greyed out entries did not pass study diagnostics and are included only for completeness.

| | Patients | | Exposure (KYr) | | Outcomes | | Uncalibrated | | Calibrated | |
|---|---|---|---|---|---|---|---|---|---|---|
| | T | C | T | C | T | C | HR (95% CI) | $p$ | HR (95% CI) | $p$ |
| **PS-matching / on-treatment TAR** | | | | | | | | | | |
| CCAE | 18,063 | 28,312 | 16.0 | 23.6 | 243 | 468 | 0.88 (0.71 - 1.09) | 0.26 | 0.95 (0.77 - 1.18) | 0.67 |
| DAG | 4,031 | 9,692 | 3.5 | 9.8 | 79 | 233 | 1.05 (0.75 - 1.47) | 0.76 | 1.03 (0.73 - 1.45) | 0.69 |
| ODOD | 11,059 | 18,283 | 8.4 | 14.0 | 243 | 556 | 0.92 (0.74 - 1.14) | 0.44 | 0.95 (0.76 - 1.18) | 0.70 |
| MDCR | 1,499 | 4,364 | 1.3 | 4.1 | 41 | 166 | 1.07 (0.67 - 1.65) | 0.78 | 1.07 (0.68 - 1.68) | 0.77 |
| MDCD | 1,306 | 3,061 | 0.8 | 2.2 | 15 | 62 | 0.93 (0.44 - 1.85) | 0.84 | 0.84 (0.41 - 1.72) | 0.64 |
| Open Claims | 217,569 | 374,695 | 212.3 | 354.2 | 2,496 | 6,880 | 0.83 (0.78 - 0.89) | <0.01 | 0.91 (0.81 - 1.03) | 0.16 |
| OEHR | 15,971 | 30,272 | 5.2 | 15.4 | 95 | 329 | 1.30 (0.94 - 1.78) | 0.11 | 1.31 (0.95 - 1.80) | 0.11 |
| VA | 7,042 | 21,383 | 4.8 | 19.3 | 39 | 268 | 0.67 (0.42 - 1.05) | 0.09 | 0.68 (0.43 - 1.07) | 0.11 |
| SIDIAP | 3,828 | 17,257 | 5.7 | 38.5 | 119 | 976 | 0.97 (0.76 - 1.22) | 0.78 | 1.05 (0.82 - 1.35) | 0.69 |
| Meta-analysis | 280,892 | 511,290 | 259.0 | 487.8 | 3,365 | 9,956 | 0.92 (0.83 - 1.02) | 0.10 | 0.97 (0.84 - 1.12) | 0.71 |
| Meta-analysis (w/o USOC) | 63,323 | 136,595 | 46.7 | 133.6 | 869 | 3,076 | 0.96 (0.86 - 1.07) | 0.43 | 0.99 (0.89 - 1.10) | 0.80 |
| **PS-stratification / on-treatment TAR** | | | | | | | | | | |
| CCAE | 22,104 | 28,325 | 18.3 | 23.6 | 274 | 470 | 0.78 (0.66 - 0.91) | <0.01 | 0.81 (0.69 - 0.95) | 0.01 |
| DAG | 4,973 | 9,717 | 4.0 | 9.8 | 96 | 236 | 1.12 (0.86 - 1.46) | 0.40 | 1.08 (0.82 - 1.41) | 0.59 |
| ODOD | 15,706 | 18,423 | 10.8 | 14.1 | 287 | 568 | 0.88 (0.75 - 1.03) | 0.12 | 0.89 (0.76 - 1.04) | 0.20 |
| MDCR | 1,870 | 4,406 | 1.5 | 4.1 | 43 | 168 | 0.82 (0.56 - 1.17) | 0.28 | 0.84 (0.58 - 1.22) | 0.39 |
| MDCD | 1,377 | 3,076 | 0.8 | 2.2 | 15 | 64 | 0.78 (0.42 - 1.36) | 0.40 | 0.70 (0.39 - 1.27) | 0.25 |
| Open Claims | 297,034 | 374,885 | 259.6 | 354.3 | 2,993 | 6,901 | 0.78 (0.74 - 0.81) | <0.01 | 0.84 (0.73 - 0.96) | 0.01 |
| OEHR | 19,171 | 30,328 | 6.1 | 15.4 | 109 | 336 | 1.13 (0.89 - 1.43) | 0.32 | 1.13 (0.89 - 1.44) | 0.33 |
| VA | 11,310 | 22,289 | 6.8 | 20.6 | 68 | 289 | 0.62 (0.45 - 0.86) | <0.01 | 0.65 (0.47 - 0.91) | 0.01 |
| SIDIAP | 4,522 | 17,643 | 6.3 | 39.8 | 131 | 1,029 | 0.91 (0.74 - 1.11) | 0.37 | 0.94 (0.76 - 1.15) | 0.60 |
| Meta-analysis | 365,380 | 483,727 | 306.5 | 461.3 | 3,933 | 9,708 | 0.88 (0.79 - 0.99) | 0.04 | 0.93 (0.79 - 1.10) | 0.40 |
| Meta-analysis (w/o USOC) | 68,346 | 108,842 | 46.9 | 106.9 | 940 | 2,807 | 0.92 (0.81 - 1.04) | 0.18 | 0.94 (0.82 - 1.06) | 0.35 |
| **PS-matching / on-treatment TAR without escalation** | | | | | | | | | | |
| CCAE | 18,063 | 28,312 | 13.9 | 19.2 | 206 | 381 | 0.99 (0.78 - 1.25) | 0.93 | 1.05 (0.83 - 1.33) | 0.68 |
| DAG | 4,031 | 9,692 | 3.2 | 8.9 | 71 | 207 | 1.08 (0.74 - 1.55) | 0.68 | 1.05 (0.72 - 1.51) | 0.68 |
| ODOD | 11,059 | 18,283 | 7.4 | 11.5 | 208 | 465 | 0.92 (0.73 - 1.15) | 0.46 | 0.95 (0.72 - 1.25) | 0.75 |
| MDCR | 1,499 | 4,364 | 1.2 | 3.5 | 38 | 144 | 1.08 (0.67 - 1.69) | 0.75 | 1.09 (0.69 - 1.73) | 0.73 |
| MDCD | 1,306 | 3,061 | 0.7 | 1.6 | 12 | 49 | 1.18 (0.50 - 2.61) | 0.70 | 1.12 (0.49 - 2.57) | 0.78 |
| Open Claims | 217,569 | 374,695 | 178.2 | 274.9 | 2,090 | 5,470 | 0.85 (0.79 - 0.91) | <0.01 | 0.91 (0.80 - 1.03) | 0.19 |
| OEHR | 15,971 | 30,272 | 4.5 | 12.5 | 82 | 254 | 1.18 (0.83 - 1.67) | 0.36 | 1.19 (0.83 - 1.69) | 0.36 |
| VA | 7,042 | 21,383 | 4.1 | 15.7 | 31 | 220 | 0.70 (0.42 - 1.15) | 0.17 | 0.70 (0.42 - 1.16) | 0.18 |
| SIDIAP | 3,828 | 17,257 | 5.0 | 30.7 | 103 | 771 | 0.99 (0.76 - 1.29) | 0.96 | 1.17 (0.89 - 1.53) | 0.35 |
| Meta-analysis | 280,892 | 511,290 | 219.2 | 384.2 | 2,839 | 7,978 | 0.93 (0.84 - 1.02) | 0.14 | 0.98 (0.84 - 1.15) | 0.82 |
| Meta-analysis (w/o USOC) | 63,323 | 136,595 | 41.0 | 109.3 | 749 | 2,508 | 0.99 (0.88 - 1.10) | 0.80 | 1.02 (0.90 - 1.14) | 0.79 |
| **PS-stratification / on-treatment TAR without escalation** | | | | | | | | | | |
| CCAE | 22,104 | 28,325 | 16.0 | 19.2 | 233 | 383 | 0.77 (0.65 - 0.91) | <0.01 | 0.80 (0.67 - 0.95) | 0.02 |
| DAG | 4,973 | 9,717 | 3.7 | 8.9 | 88 | 210 | 1.15 (0.86 - 1.53) | 0.33 | 1.12 (0.84 - 1.49) | 0.49 |
| ODOD | 15,706 | 18,423 | 9.6 | 11.6 | 248 | 475 | 0.86 (0.72 - 1.02) | 0.08 | 0.85 (0.72 - 1.01) | 0.09 |
| MDCR | 1,870 | 4,406 | 1.3 | 3.5 | 40 | 145 | 0.81 (0.55 - 1.18) | 0.29 | 0.84 (0.57 - 1.24) | 0.41 |
| MDCD | 1,377 | 3,076 | 0.7 | 1.7 | 12 | 50 | 0.73 (0.36 - 1.37) | 0.36 | 0.67 (0.34 - 1.31) | 0.25 |
| Open Claims | 297,034 | 374,885 | 220.9 | 275.0 | 2,550 | 5,488 | 0.77 (0.73 - 0.81) | <0.01 | 0.84 (0.75 - 0.93) | <0.01 |
| OEHR | 19,171 | 30,328 | 5.4 | 12.5 | 95 | 261 | 1.15 (0.88 - 1.48) | 0.30 | 1.14 (0.88 - 1.48) | 0.35 |
| VA | 11,310 | 22,289 | 5.9 | 16.7 | 59 | 238 | 0.58 (0.41 - 0.83) | <0.01 | 0.61 (0.42 - 0.87) | <0.01 |
| SIDIAP | 4,522 | 17,643 | 5.6 | 31.7 | 115 | 809 | 0.91 (0.73 - 1.13) | 0.40 | 1.00 (0.80 - 1.25) | 0.76 |
| Meta-analysis | 365,380 | 483,727 | 262.5 | 362.5 | 3,369 | 7,771 | 0.88 (0.78 - 1.00) | 0.04 | 0.93 (0.81 - 1.07) | 0.33 |
| Meta-analysis (w/o USOC) | 68,346 | 108,842 | 41.6 | 87.4 | 819 | 2,283 | 0.92 (0.80 - 1.05) | 0.20 | 0.94 (0.81 - 1.08) | 0.39 |

##### 1.2.17 Hyperkalemia

**Supplementary Table 43:** Exposures, outcomes and risk estimates for Hyperkalemia in SGLT2I (male) (T) vs DPP4I (male) (C) new-user cohorts under four different propensity score (PS) adjustment / time-at-risk (TAR) designs across data sources. We report total patient exposure time in 1,000 patient-year (KYr) units, uncalibrated and calibrated hazard ratios (HRs) and their 95% confidence intervals (CIs). Greyed out entries did not pass study diagnostics and are included only for completeness.

| | Patients | | Exposure (KYr) | | Outcomes | | Uncalibrated | | Calibrated | |
|---|---|---|---|---|---|---|---|---|---|---|
| | T | C | T | C | T | C | HR (95% CI) | $p$ | HR (95% CI) | $p$ |
| **PS-matching / on-treatment TAR** | | | | | | | | | | |
| CCAE | 19,405 | 30,586 | 17.2 | 25.6 | 118 | 218 | 0.79 (0.57 - 1.08) | 0.15 | 0.85 (0.62 - 1.17) | 0.33 |
| DAG | 4,358 | 10,400 | 3.8 | 10.7 | 44 | 162 | 0.83 (0.52 - 1.30) | 0.43 | 0.81 (0.51 - 1.29) | 0.40 |
| ODOD | 12,017 | 20,373 | 9.1 | 15.8 | 152 | 394 | 0.96 (0.73 - 1.25) | 0.75 | 0.99 (0.75 - 1.30) | 0.88 |
| MDCR | 1,664 | 5,089 | 1.5 | 4.7 | 18 | 103 | 1.20 (0.59 - 2.31) | 0.60 | 1.20 (0.61 - 2.38) | 0.60 |
| MDCD | 1,384 | 3,314 | 0.8 | 2.4 | 14 | 42 | 1.11 (0.48 - 2.46) | 0.80 | 1.00 (0.44 - 2.28) | 0.90 |
| Open Claims | 239,626 | 416,521 | 232.3 | 396.4 | 1,007 | 3,251 | 0.71 (0.65 - 0.79) | <0.01 | 0.78 (0.67 - 0.90) | <0.01 |
| OEHR | 16,549 | 31,585 | 5.4 | 16.1 | 51 | 263 | 0.64 (0.41 - 0.95) | 0.03 | 0.64 (0.42 - 0.97) | 0.04 |
| VA | 7,242 | 22,546 | 4.9 | 20.1 | 64 | 287 | 1.10 (0.75 - 1.58) | 0.63 | 1.10 (0.76 - 1.60) | 0.62 |
| SIDIAP | 4,139 | 18,612 | 6.0 | 41.8 | 60 | 528 | 0.83 (0.58 - 1.17) | 0.31 | 0.91 (0.63 - 1.30) | 0.65 |
| Meta-analysis | 305,000 | 555,712 | 280.1 | 531.3 | 1,514 | 5,206 | 0.82 (0.72 - 0.94) | <0.01 | 0.87 (0.73 - 1.03) | 0.10 |
| Meta-analysis (w/o USOC) | 65,374 | 139,191 | 47.9 | 134.9 | 507 | 1,955 | 0.88 (0.76 - 1.01) | 0.06 | 0.90 (0.78 - 1.04) | 0.18 |
| **PS-stratification / on-treatment TAR** | | | | | | | | | | |
| CCAE | 23,691 | 30,605 | 19.6 | 25.6 | 134 | 218 | 0.88 (0.70 - 1.11) | 0.28 | 0.92 (0.73 - 1.15) | 0.47 |
| DAG | 5,392 | 10,429 | 4.4 | 10.7 | 52 | 164 | 0.78 (0.54 - 1.10) | 0.16 | 0.75 (0.52 - 1.06) | 0.13 |
| ODOD | 16,805 | 20,471 | 11.6 | 15.8 | 192 | 401 | 0.86 (0.71 - 1.04) | 0.12 | 0.87 (0.72 - 1.05) | 0.18 |
| MDCR | 2,069 | 5,141 | 1.7 | 4.8 | 20 | 103 | 0.73 (0.42 - 1.21) | 0.24 | 0.75 (0.44 - 1.28) | 0.31 |
| MDCD | 1,462 | 3,327 | 0.9 | 2.4 | 15 | 42 | 1.07 (0.56 - 1.95) | 0.83 | 0.97 (0.52 - 1.80) | 0.89 |
| Open Claims | 325,992 | 416,779 | 283.5 | 396.6 | 1,288 | 3,267 | 0.68 (0.64 - 0.73) | <0.01 | 0.74 (0.64 - 0.86) | <0.01 |
| OEHR | 19,863 | 31,647 | 6.3 | 16.1 | 62 | 266 | 0.71 (0.52 - 0.95) | 0.03 | 0.71 (0.53 - 0.96) | 0.03 |
| VA | 11,422 | 23,467 | 6.9 | 21.4 | 104 | 307 | 0.84 (0.63 - 1.11) | 0.23 | 0.88 (0.66 - 1.17) | 0.39 |
| SIDIAP | 4,910 | 18,967 | 6.7 | 43.1 | 78 | 556 | 0.88 (0.67 - 1.15) | 0.37 | 0.91 (0.69 - 1.19) | 0.55 |
| Meta-analysis | 398,722 | 534,039 | 333.7 | 512.8 | 1,826 | 4,975 | 0.78 (0.69 - 0.87) | <0.01 | 0.82 (0.69 - 0.96) | 0.02 |
| Meta-analysis (w/o USOC) | 72,730 | 117,260 | 50.2 | 116.3 | 538 | 1,708 | 0.83 (0.75 - 0.93) | <0.01 | 0.85 (0.76 - 0.95) | <0.01 |
| **PS-matching / on-treatment TAR without escalation** | | | | | | | | | | |
| CCAE | 19,405 | 30,586 | 14.9 | 20.8 | 97 | 178 | 0.77 (0.54 - 1.09) | 0.15 | 0.82 (0.58 - 1.17) | 0.29 |
| DAG | 4,358 | 10,400 | 3.5 | 9.7 | 38 | 143 | 0.83 (0.50 - 1.34) | 0.45 | 0.80 (0.49 - 1.32) | 0.41 |
| ODOD | 12,017 | 20,373 | 8.0 | 12.9 | 136 | 319 | 1.01 (0.76 - 1.34) | 0.96 | 1.04 (0.75 - 1.44) | 0.83 |
| MDCR | 1,664 | 5,089 | 1.3 | 4.0 | 17 | 84 | 1.31 (0.63 - 2.59) | 0.46 | 1.32 (0.65 - 2.69) | 0.45 |
| MDCD | 1,384 | 3,314 | 0.7 | 1.8 | 13 | 30 | 1.10 (0.45 - 2.55) | 0.82 | 1.06 (0.44 - 2.51) | 0.87 |
| Open Claims | 239,626 | 416,521 | 194.9 | 307.8 | 829 | 2,555 | 0.69 (0.62 - 0.78) | <0.01 | 0.74 (0.64 - 0.87) | <0.01 |
| OEHR | 16,549 | 31,585 | 4.7 | 13.1 | 39 | 203 | 0.75 (0.47 - 1.18) | 0.23 | 0.76 (0.48 - 1.20) | 0.26 |
| VA | 7,242 | 22,546 | 4.2 | 16.4 | 56 | 220 | 1.05 (0.69 - 1.57) | 0.82 | 1.04 (0.69 - 1.57) | 0.83 |
| SIDIAP | 4,139 | 18,612 | 5.4 | 33.2 | 46 | 412 | 0.69 (0.46 - 1.01) | 0.06 | 0.81 (0.54 - 1.20) | 0.35 |
| Meta-analysis | 305,000 | 555,712 | 236.8 | 417.9 | 1,258 | 4,114 | 0.82 (0.70 - 0.95) | <0.01 | 0.86 (0.71 - 1.05) | 0.14 |
| Meta-analysis (w/o USOC) | 65,374 | 139,191 | 42.0 | 110.1 | 429 | 1,559 | 0.88 (0.75 - 1.02) | 0.09 | 0.90 (0.78 - 1.06) | 0.22 |
| **PS-stratification / on-treatment TAR without escalation** | | | | | | | | | | |
| CCAE | 23,691 | 30,605 | 17.1 | 20.8 | 109 | 178 | 0.84 (0.65 - 1.08) | 0.18 | 0.87 (0.68 - 1.13) | 0.31 |
| DAG | 5,392 | 10,429 | 4.1 | 9.7 | 46 | 145 | 0.74 (0.51 - 1.08) | 0.12 | 0.72 (0.49 - 1.05) | 0.11 |
| ODOD | 16,805 | 20,471 | 10.3 | 13.0 | 173 | 326 | 0.88 (0.72 - 1.08) | 0.23 | 0.87 (0.71 - 1.07) | 0.23 |
| MDCR | 2,069 | 5,141 | 1.5 | 4.1 | 19 | 84 | 0.89 (0.50 - 1.50) | 0.66 | 0.92 (0.53 - 1.59) | 0.77 |
| MDCD | 1,462 | 3,327 | 0.7 | 1.8 | 14 | 30 | 1.23 (0.62 - 2.37) | 0.54 | 1.13 (0.58 - 2.22) | 0.73 |
| Open Claims | 325,992 | 416,779 | 241.1 | 308.0 | 1,086 | 2,569 | 0.68 (0.62 - 0.73) | <0.01 | 0.73 (0.65 - 0.82) | <0.01 |
| OEHR | 19,863 | 31,647 | 5.6 | 13.1 | 47 | 205 | 0.62 (0.44 - 0.87) | <0.01 | 0.62 (0.44 - 0.87) | <0.01 |
| VA | 11,422 | 23,467 | 6.0 | 17.4 | 89 | 238 | 0.87 (0.63 - 1.19) | 0.39 | 0.91 (0.66 - 1.24) | 0.56 |
| SIDIAP | 4,910 | 18,967 | 6.0 | 34.2 | 62 | 436 | 0.81 (0.59 - 1.09) | 0.18 | 0.89 (0.65 - 1.21) | 0.52 |
| Meta-analysis | 398,722 | 534,039 | 285.6 | 402.9 | 1,542 | 3,943 | 0.76 (0.67 - 0.85) | <0.01 | 0.80 (0.69 - 0.92) | <0.01 |
| Meta-analysis (w/o USOC) | 72,730 | 117,260 | 44.5 | 94.9 | 456 | 1,374 | 0.81 (0.72 - 0.91) | <0.01 | 0.83 (0.73 - 0.94) | <0.01 |

##### 1.2.18 Hypoglycemia

**Supplementary Table 44:** Exposures, outcomes and risk estimates for Hypoglycemia in SGLT2I (male) (T) vs DPP4I (male) (C) new-user cohorts under four different propensity score (PS) adjustment / time-at-risk (TAR) designs across data sources. We report total patient exposure time in 1,000 patient-year (KYr) units, uncalibrated and calibrated hazard ratios (HRs) and their 95% confidence intervals (CIs). Greyed out entries did not pass study diagnostics and are included only for completeness.

| | Patients | | Exposure (KYr) | | Outcomes | | Uncalibrated | | Calibrated | |
|---|---|---|---|---|---|---|---|---|---|---|
| | T | C | T | C | T | C | HR (95% CI) | $p$ | HR (95% CI) | $p$ |
| **PS-matching / on-treatment TAR** | | | | | | | | | | |
| CCAE | 19,492 | 30,753 | 17.4 | 26.0 | 42 | 80 | 0.81 (0.46 - 1.39) | 0.45 | 0.87 (0.50 - 1.51) | 0.63 |
| DAG | 4,612 | 11,097 | 4.0 | 11.6 | <5 | 17 | 0.70 (0.10 - 3.50) | 0.69 | 0.68 (0.11 - 4.12) | 0.68 |
| ODOD | 12,339 | 21,176 | 9.4 | 16.7 | 35 | 85 | 0.72 (0.39 - 1.30) | 0.29 | 0.75 (0.41 - 1.37) | 0.36 |
| MDCR | 1,702 | 5,209 | 1.5 | 4.9 | 5 | 18 | 1.06 (0.21 - 4.38) | 0.94 | 1.07 (0.24 - 4.87) | 0.93 |
| MDCD | 1,388 | 3,405 | 0.9 | 2.5 | 6 | 20 | 1.46 (0.42 - 4.73) | 0.54 | 1.31 (0.39 - 4.43) | 0.66 |
| Open Claims | 241,000 | 420,640 | 234.0 | 403.0 | 353 | 960 | 0.78 (0.65 - 0.93) | <0.01 | 0.85 (0.69 - 1.05) | 0.14 |
| OEHR | 16,739 | 32,099 | 5.5 | 16.5 | 15 | 93 | 1.21 (0.56 - 2.55) | 0.62 | 1.22 (0.57 - 2.60) | 0.61 |
| VA | 7,705 | 23,718 | 5.3 | 21.5 | 10 | 71 | 0.38 (0.13 - 0.94) | 0.06 | 0.38 (0.14 - 1.04) | 0.06 |
| SIDIAP | 4,333 | 19,814 | 6.3 | 45.1 | <5 | 22 | 0.43 (0.02 - 2.67) | 0.49 | 0.47 (0.04 - 5.18) | 0.54 |
| Meta-analysis | 303,589 | 544,692 | 277.1 | 500.1 | <465 | 1,324 | 0.78 (0.67 - 0.92) | <0.01 | 0.83 (0.69 - 1.00) | 0.05 |
| Meta-analysis (w/o USOC) | 62,589 | 124,052 | 43.1 | 97.1 | <112 | 364 | 0.79 (0.57 - 1.09) | 0.14 | 0.81 (0.59 - 1.12) | 0.21 |
| **PS-stratification / on-treatment TAR** | | | | | | | | | | |
| CCAE | 23,833 | 30,776 | 19.8 | 26.0 | 49 | 80 | 0.71 (0.49 - 1.02) | 0.07 | 0.73 (0.51 - 1.06) | 0.11 |
| DAG | 5,735 | 11,129 | 4.7 | 11.6 | 5 | 17 | 1.02 (0.31 - 2.82) | 0.97 | 0.98 (0.32 - 2.95) | 0.92 |
| ODOD | 17,261 | 21,296 | 12.0 | 16.8 | 51 | 85 | 0.88 (0.59 - 1.31) | 0.53 | 0.89 (0.60 - 1.32) | 0.57 |
| MDCR | 2,115 | 5,268 | 1.7 | 5.0 | 5 | 18 | 0.97 (0.30 - 2.66) | 0.96 | 1.01 (0.34 - 2.98) | 0.95 |
| MDCD | 1,468 | 3,418 | 0.9 | 2.5 | 6 | 20 | 0.87 (0.31 - 2.14) | 0.78 | 0.79 (0.30 - 2.07) | 0.64 |
| Open Claims | 328,462 | 420,953 | 285.7 | 403.2 | 421 | 962 | 0.71 (0.63 - 0.81) | <0.01 | 0.77 (0.64 - 0.92) | <0.01 |
| OEHR | 20,063 | 32,169 | 6.4 | 16.6 | 18 | 93 | 0.63 (0.36 - 1.06) | 0.09 | 0.63 (0.37 - 1.09) | 0.10 |
| VA | 12,356 | 24,768 | 7.4 | 23.0 | 14 | 77 | 0.61 (0.30 - 1.17) | 0.16 | 0.64 (0.32 - 1.27) | 0.20 |
| SIDIAP | 5,159 | 20,250 | 7.1 | 46.7 | 5 | 23 | 1.27 (0.35 - 3.92) | 0.70 | 1.31 (0.39 - 4.37) | 0.67 |
| Meta-analysis | 402,628 | 541,841 | 337.4 | 525.8 | 554 | 1,278 | 0.73 (0.65 - 0.81) | <0.01 | 0.77 (0.65 - 0.90) | <0.01 |
| Meta-analysis (w/o USOC) | 74,166 | 120,888 | 51.7 | 122.6 | 133 | 316 | 0.78 (0.62 - 0.98) | 0.03 | 0.80 (0.63 - 1.00) | 0.06 |
| **PS-matching / on-treatment TAR without escalation** | | | | | | | | | | |
| CCAE | 19,492 | 30,753 | 15.1 | 21.1 | 32 | 53 | 0.78 (0.40 - 1.49) | 0.46 | 0.83 (0.43 - 1.60) | 0.58 |
| DAG | 4,612 | 11,097 | 3.7 | 10.4 | <5 | 14 | 0.69 (0.10 - 3.47) | 0.69 | 0.67 (0.11 - 4.05) | 0.67 |
| ODOD | 12,339 | 21,176 | 8.3 | 13.7 | 23 | 48 | 0.95 (0.47 - 1.88) | 0.88 | 0.98 (0.48 - 2.00) | 0.93 |
| MDCR | 1,702 | 5,209 | 1.3 | 4.2 | <5 | 12 | 0.93 (0.13 - 4.75) | 0.94 | 0.95 (0.15 - 5.82) | 0.95 |
| MDCD | 1,388 | 3,405 | 0.7 | 1.9 | <5 | 9 | 1.70 (0.31 - 9.44) | 0.54 | 1.63 (0.29 - 9.03) | 0.58 |
| Open Claims | 241,000 | 420,640 | 196.3 | 312.9 | 240 | 532 | 0.86 (0.69 - 1.06) | 0.16 | 0.92 (0.72 - 1.17) | 0.51 |
| OEHR | 16,739 | 32,099 | 4.8 | 13.4 | 9 | 47 | 1.06 (0.40 - 2.69) | 0.90 | 1.07 (0.41 - 2.76) | 0.89 |
| VA | 7,705 | 23,718 | 4.5 | 17.5 | <10 | 40 | 0.50 (0.14 - 1.45) | 0.25 | 0.50 (0.15 - 1.62) | 0.25 |
| SIDIAP | 4,333 | 19,814 | 5.6 | 35.8 | <5 | 12 | 0.53 (0.03 - 3.82) | 0.62 | 0.62 (0.05 - 7.57) | 0.71 |
| Meta-analysis | 297,275 | 528,386 | 228.9 | 378.5 | <314 | 720 | 0.85 (0.70 - 1.03) | 0.10 | 0.90 (0.72 - 1.13) | 0.37 |
| Meta-analysis (w/o USOC) | 56,275 | 107,746 | 32.6 | 65.6 | <74 | 188 | 0.84 (0.56 - 1.25) | 0.38 | 0.86 (0.58 - 1.29) | 0.47 |
| **PS-stratification / on-treatment TAR without escalation** | | | | | | | | | | |
| CCAE | 23,833 | 30,776 | 17.3 | 21.1 | 38 | 53 | 0.74 (0.48 - 1.14) | 0.18 | 0.77 (0.50 - 1.19) | 0.25 |
| DAG | 5,735 | 11,129 | 4.3 | 10.5 | 5 | 14 | 1.20 (0.35 - 3.47) | 0.75 | 1.16 (0.37 - 3.63) | 0.80 |
| ODOD | 17,261 | 21,296 | 10.6 | 13.8 | 36 | 48 | 1.08 (0.65 - 1.77) | 0.77 | 1.07 (0.65 - 1.76) | 0.80 |
| MDCR | 2,115 | 5,268 | 1.5 | 4.2 | <5 | 12 | 0.93 (0.24 - 2.98) | 0.91 | 0.97 (0.28 - 3.39) | 0.95 |
| MDCD | 1,468 | 3,418 | 0.7 | 1.9 | <5 | 9 | 1.16 (0.30 - 3.76) | 0.81 | 1.07 (0.30 - 3.74) | 0.92 |
| Open Claims | 328,462 | 420,953 | 243.0 | 313.0 | 289 | 533 | 0.76 (0.65 - 0.89) | <0.01 | 0.82 (0.69 - 0.99) | 0.04 |
| OEHR | 20,063 | 32,169 | 5.6 | 13.4 | 12 | 47 | 0.69 (0.33 - 1.32) | 0.29 | 0.68 (0.34 - 1.36) | 0.28 |
| VA | 12,356 | 24,768 | 6.4 | 18.7 | <10 | 43 | 0.47 (0.17 - 1.16) | 0.12 | 0.49 (0.19 - 1.27) | 0.15 |
| SIDIAP | 5,159 | 20,250 | 6.3 | 37.0 | <5 | 12 | 0.98 (0.18 - 4.17) | 0.98 | 1.07 (0.22 - 5.17) | 0.92 |
| Meta-analysis | 395,354 | 516,323 | 281.0 | 371.8 | 380 | 695 | 0.78 (0.68 - 0.90) | <0.01 | 0.83 (0.71 - 0.97) | 0.02 |
| Meta-analysis (w/o USOC) | 66,892 | 95,370 | 37.9 | 58.7 | 91 | 162 | 0.85 (0.64 - 1.14) | 0.28 | 0.87 (0.65 - 1.16) | 0.36 |

##### 1.2.19 Hypotension

**Supplementary Table 45:** Exposures, outcomes and risk estimates for Hypotension in SGLT2I (male) (T) vs DPP4I (male) (C) new-user cohorts under four different propensity score (PS) adjustment / time-at-risk (TAR) designs across data sources. We report total patient exposure time in 1,000 patient-year (KYr) units, uncalibrated and calibrated hazard ratios (HRs) and their 95% confidence intervals (CIs). Greyed out entries did not pass study diagnostics and are included only for completeness.

| | Patients | | Exposure (KYr) | | Outcomes | | Uncalibrated | | Calibrated | |
|---|---|---|---|---|---|---|---|---|---|---|
| | T | C | T | C | T | C | HR (95% CI) | $p$ | HR (95% CI) | $p$ |
| **PS-matching / on-treatment TAR** | | | | | | | | | | |
| CCAE | 19,373 | 30,593 | 17.2 | 25.7 | 150 | 234 | 1.06 (0.79 - 1.42) | 0.68 | 1.14 (0.85 - 1.54) | 0.38 |
| DAG | 4,506 | 10,904 | 3.9 | 11.3 | 17 | 59 | 0.86 (0.39 - 1.78) | 0.69 | 0.84 (0.39 - 1.79) | 0.66 |
| ODOD | 11,998 | 20,309 | 9.1 | 15.8 | 187 | 360 | 1.13 (0.88 - 1.46) | 0.33 | 1.17 (0.90 - 1.52) | 0.30 |
| MDCR | 1,613 | 4,915 | 1.4 | 4.6 | 29 | 133 | 1.21 (0.69 - 2.05) | 0.49 | 1.22 (0.71 - 2.09) | 0.50 |
| MDCD | 1,343 | 3,198 | 0.8 | 2.3 | 20 | 75 | 0.98 (0.49 - 1.88) | 0.96 | 0.89 (0.45 - 1.74) | 0.74 |
| Open Claims | 236,828 | 411,272 | 229.8 | 392.3 | 1,604 | 3,732 | 1.02 (0.93 - 1.11) | 0.70 | 1.11 (0.97 - 1.26) | 0.17 |
| OEHR | 16,524 | 31,578 | 5.4 | 16.1 | 74 | 287 | 0.86 (0.60 - 1.22) | 0.41 | 0.87 (0.61 - 1.23) | 0.45 |
| VA | 7,264 | 22,448 | 4.9 | 20.1 | 91 | 259 | 1.18 (0.83 - 1.67) | 0.34 | 1.19 (0.84 - 1.69) | 0.35 |
| SIDIAP | 4,311 | 19,685 | 6.3 | 44.7 | 19 | 93 | 1.51 (0.76 - 2.83) | 0.22 | 1.64 (0.85 - 3.17) | 0.15 |
| Meta-analysis | 302,417 | 551,704 | 278.1 | 530.6 | 2,171 | 5,157 | 1.03 (0.96 - 1.11) | 0.36 | 1.10 (0.97 - 1.24) | 0.14 |
| Meta-analysis (w/o USOC) | 65,589 | 140,432 | 48.3 | 138.4 | 567 | 1,425 | 1.09 (0.94 - 1.25) | 0.25 | 1.12 (0.97 - 1.29) | 0.14 |
| **PS-stratification / on-treatment TAR** | | | | | | | | | | |
| CCAE | 23,656 | 30,615 | 19.6 | 25.7 | 177 | 234 | 1.02 (0.83 - 1.25) | 0.87 | 1.06 (0.86 - 1.30) | 0.61 |
| DAG | 5,584 | 10,935 | 4.6 | 11.4 | 25 | 60 | 0.93 (0.53 - 1.58) | 0.78 | 0.89 (0.52 - 1.53) | 0.68 |
| ODOD | 16,736 | 20,394 | 11.6 | 15.9 | 229 | 369 | 1.07 (0.89 - 1.28) | 0.49 | 1.08 (0.89 - 1.30) | 0.47 |
| MDCR | 2,017 | 4,966 | 1.6 | 4.6 | 35 | 138 | 1.04 (0.67 - 1.56) | 0.87 | 1.07 (0.70 - 1.63) | 0.76 |
| MDCD | 1,415 | 3,209 | 0.8 | 2.3 | 21 | 77 | 0.98 (0.57 - 1.60) | 0.93 | 0.88 (0.53 - 1.47) | 0.65 |
| Open Claims | 321,381 | 411,524 | 279.9 | 392.4 | 1,989 | 3,746 | 0.95 (0.89 - 1.01) | 0.09 | 1.02 (0.89 - 1.18) | 0.76 |
| OEHR | 19,804 | 31,642 | 6.3 | 16.2 | 83 | 295 | 0.91 (0.69 - 1.18) | 0.47 | 0.91 (0.70 - 1.19) | 0.51 |
| VA | 11,420 | 23,485 | 6.9 | 21.6 | 152 | 277 | 1.22 (0.94 - 1.58) | 0.13 | 1.28 (0.98 - 1.65) | 0.08 |
| SIDIAP | 5,126 | 20,114 | 7.0 | 46.3 | 23 | 96 | 1.44 (0.81 - 2.47) | 0.20 | 1.48 (0.85 - 2.59) | 0.18 |
| Meta-analysis | 394,304 | 530,190 | 330.6 | 512.5 | 2,561 | 4,938 | 0.97 (0.91 - 1.02) | 0.23 | 1.02 (0.89 - 1.16) | 0.79 |
| Meta-analysis (w/o USOC) | 72,923 | 118,666 | 50.7 | 120.0 | 572 | 1,192 | 1.03 (0.92 - 1.15) | 0.65 | 1.05 (0.93 - 1.18) | 0.48 |
| **PS-matching / on-treatment TAR without escalation** | | | | | | | | | | |
| CCAE | 19,373 | 30,593 | 14.9 | 20.8 | 121 | 191 | 1.05 (0.76 - 1.44) | 0.78 | 1.11 (0.81 - 1.54) | 0.53 |
| DAG | 4,506 | 10,904 | 3.6 | 10.2 | 14 | 53 | 0.98 (0.42 - 2.14) | 0.96 | 0.95 (0.42 - 2.13) | 0.85 |
| ODOD | 11,998 | 20,309 | 8.0 | 13.0 | 167 | 274 | 1.21 (0.92 - 1.59) | 0.17 | 1.25 (0.91 - 1.72) | 0.25 |
| MDCR | 1,613 | 4,915 | 1.3 | 3.9 | 24 | 115 | 1.14 (0.63 - 2.01) | 0.65 | 1.16 (0.65 - 2.07) | 0.64 |
| MDCD | 1,343 | 3,198 | 0.7 | 1.7 | 16 | 58 | 0.65 (0.27 - 1.43) | 0.32 | 0.63 (0.27 - 1.44) | 0.29 |
| Open Claims | 236,828 | 411,272 | 192.9 | 304.4 | 1,321 | 2,896 | 1.01 (0.92 - 1.11) | 0.87 | 1.08 (0.94 - 1.25) | 0.32 |
| OEHR | 16,524 | 31,578 | 4.7 | 13.1 | 58 | 204 | 0.81 (0.54 - 1.20) | 0.30 | 0.82 (0.55 - 1.21) | 0.34 |
| VA | 7,264 | 22,448 | 4.2 | 16.4 | 73 | 189 | 1.26 (0.85 - 1.85) | 0.25 | 1.25 (0.85 - 1.85) | 0.27 |
| SIDIAP | 4,311 | 19,685 | 5.6 | 35.5 | 17 | 79 | 1.23 (0.58 - 2.44) | 0.57 | 1.45 (0.71 - 2.96) | 0.33 |
| Meta-analysis | 302,417 | 551,704 | 235.3 | 417.3 | 1,795 | 4,001 | 1.03 (0.95 - 1.12) | 0.45 | 1.09 (0.94 - 1.26) | 0.24 |
| Meta-analysis (w/o USOC) | 65,589 | 140,432 | 42.3 | 112.9 | 474 | 1,105 | 1.10 (0.94 - 1.28) | 0.24 | 1.13 (0.97 - 1.32) | 0.14 |
| **PS-stratification / on-treatment TAR without escalation** | | | | | | | | | | |
| CCAE | 23,656 | 30,615 | 17.2 | 20.8 | 146 | 191 | 0.97 (0.77 - 1.22) | 0.78 | 1.01 (0.80 - 1.27) | 0.90 |
| DAG | 5,584 | 10,935 | 4.2 | 10.3 | 22 | 54 | 0.93 (0.51 - 1.64) | 0.80 | 0.90 (0.50 - 1.61) | 0.73 |
| ODOD | 16,736 | 20,394 | 10.3 | 13.0 | 204 | 282 | 1.19 (0.97 - 1.46) | 0.09 | 1.18 (0.96 - 1.45) | 0.13 |
| MDCR | 2,017 | 4,966 | 1.4 | 3.9 | 29 | 120 | 0.96 (0.59 - 1.50) | 0.86 | 1.00 (0.63 - 1.59) | 0.88 |
| MDCD | 1,415 | 3,209 | 0.7 | 1.8 | 17 | 59 | 0.86 (0.47 - 1.50) | 0.61 | 0.79 (0.44 - 1.41) | 0.43 |
| Open Claims | 321,381 | 411,524 | 238.2 | 304.6 | 1,667 | 2,908 | 0.95 (0.89 - 1.02) | 0.17 | 1.03 (0.92 - 1.15) | 0.62 |
| OEHR | 19,804 | 31,642 | 5.5 | 13.1 | 65 | 208 | 0.93 (0.68 - 1.26) | 0.65 | 0.93 (0.68 - 1.26) | 0.64 |
| VA | 11,420 | 23,485 | 5.9 | 17.6 | 129 | 203 | 1.27 (0.95 - 1.69) | 0.11 | 1.32 (0.99 - 1.76) | 0.08 |
| SIDIAP | 5,126 | 20,114 | 6.3 | 36.7 | 20 | 82 | 1.23 (0.66 - 2.18) | 0.50 | 1.35 (0.74 - 2.45) | 0.34 |
| Meta-analysis | 394,304 | 530,190 | 283.1 | 402.4 | 2,153 | 3,845 | 0.99 (0.91 - 1.09) | 0.90 | 1.05 (0.93 - 1.18) | 0.42 |
| Meta-analysis (w/o USOC) | 72,923 | 118,666 | 44.9 | 97.9 | 486 | 937 | 1.05 (0.93 - 1.19) | 0.45 | 1.07 (0.94 - 1.22) | 0.34 |

##### 1.2.20 Joint pain

**Supplementary Table 46:** Exposures, outcomes and risk estimates for Joint pain in SGLT2I (male) (T) vs DPP4I (male) (C) new-user cohorts under four different propensity score (PS) adjustment / time-at-risk (TAR) designs across data sources. We report total patient exposure time in 1,000 patient-year (KYr) units, uncalibrated and calibrated hazard ratios (HRs) and their 95% confidence intervals (CIs). Greyed out entries did not pass study diagnostics and are included only for completeness.

| | Patients | | Exposure (KYr) | | Outcomes | | Uncalibrated | | Calibrated | |
|---|---|---|---|---|---|---|---|---|---|---|
| | T | C | T | C | T | C | HR (95% CI) | $p$ | HR (95% CI) | $p$ |
| **PS-matching / on-treatment TAR** | | | | | | | | | | |
| CCAE | 18,888 | 29,889 | 16.9 | 25.1 | 118 | 229 | 0.72 (0.52 - 0.98) | 0.04 | 0.77 (0.56 - 1.06) | 0.12 |
| DAG | 3,957 | 9,569 | 3.4 | 9.8 | 58 | 213 | 0.57 (0.37 - 0.85) | <0.01 | 0.56 (0.37 - 0.84) | <0.01 |
| ODOD | 11,934 | 20,464 | 9.1 | 16.0 | 83 | 177 | 0.86 (0.60 - 1.24) | 0.44 | 0.89 (0.62 - 1.29) | 0.58 |
| MDCR | 1,669 | 5,087 | 1.5 | 4.8 | 10 | 49 | 0.98 (0.40 - 2.23) | 0.96 | 0.98 (0.42 - 2.31) | 0.92 |
| MDCD | 1,311 | 3,225 | 0.8 | 2.3 | 8 | 43 | 0.58 (0.21 - 1.42) | 0.27 | 0.52 (0.20 - 1.37) | 0.20 |
| Open Claims | 232,773 | 407,388 | 226.1 | 389.3 | 977 | 2,059 | 0.86 (0.76 - 0.96) | <0.01 | 0.93 (0.80 - 1.08) | 0.39 |
| OEHR | 16,259 | 31,055 | 5.3 | 15.9 | 47 | 120 | 1.37 (0.79 - 2.34) | 0.26 | 1.38 (0.80 - 2.37) | 0.25 |
| VA | 6,862 | 21,136 | 4.7 | 19.3 | 27 | 92 | 0.81 (0.43 - 1.48) | 0.51 | 0.81 (0.44 - 1.52) | 0.52 |
| SIDIAP | 3,476 | 16,242 | 5.1 | 36.3 | 81 | 640 | 0.97 (0.73 - 1.27) | 0.83 | 1.06 (0.79 - 1.40) | 0.71 |
| Meta-analysis | 297,656 | 548,054 | 273.8 | 525.7 | 1,411 | 3,619 | 0.85 (0.78 - 0.93) | <0.01 | 0.90 (0.79 - 1.03) | 0.13 |
| Meta-analysis (w/o USOC) | 64,883 | 140,666 | 47.7 | 136.5 | 434 | 1,560 | 0.84 (0.71 - 1.00) | 0.05 | 0.87 (0.73 - 1.04) | 0.13 |
| **PS-stratification / on-treatment TAR** | | | | | | | | | | |
| CCAE | 23,075 | 29,911 | 19.1 | 25.1 | 137 | 229 | 0.79 (0.63 - 0.98) | 0.04 | 0.82 (0.65 - 1.02) | 0.09 |
| DAG | 4,896 | 9,596 | 4.0 | 9.8 | 60 | 213 | 0.79 (0.58 - 1.07) | 0.14 | 0.76 (0.56 - 1.04) | 0.11 |
| ODOD | 16,688 | 20,572 | 11.6 | 16.1 | 103 | 177 | 0.88 (0.67 - 1.14) | 0.33 | 0.88 (0.68 - 1.16) | 0.39 |
| MDCR | 2,062 | 5,143 | 1.6 | 4.8 | 11 | 49 | 0.69 (0.33 - 1.33) | 0.30 | 0.71 (0.35 - 1.43) | 0.35 |
| MDCD | 1,397 | 3,238 | 0.8 | 2.3 | 9 | 43 | 0.54 (0.24 - 1.08) | 0.11 | 0.49 (0.23 - 1.03) | 0.06 |
| Open Claims | 317,231 | 407,695 | 276.1 | 389.5 | 1,151 | 2,059 | 0.89 (0.83 - 0.97) | <0.01 | 0.96 (0.83 - 1.12) | 0.65 |
| OEHR | 19,457 | 31,120 | 6.2 | 15.9 | 51 | 120 | 1.26 (0.87 - 1.79) | 0.21 | 1.26 (0.88 - 1.81) | 0.22 |
| VA | 10,977 | 21,929 | 6.6 | 20.5 | 46 | 99 | 1.00 (0.64 - 1.55) | 0.99 | 1.05 (0.67 - 1.63) | 0.84 |
| SIDIAP | 4,147 | 16,590 | 5.7 | 37.6 | 87 | 668 | 1.05 (0.82 - 1.34) | 0.69 | 1.08 (0.84 - 1.39) | 0.57 |
| Meta-analysis | 387,556 | 520,627 | 324.3 | 498.9 | 1,600 | 3,515 | 0.90 (0.84 - 0.96) | <0.01 | 0.94 (0.82 - 1.08) | 0.40 |
| Meta-analysis (w/o USOC) | 70,325 | 112,932 | 48.2 | 109.4 | 449 | 1,456 | 0.91 (0.78 - 1.06) | 0.21 | 0.92 (0.79 - 1.08) | 0.35 |
| **PS-matching / on-treatment TAR without escalation** | | | | | | | | | | |
| CCAE | 18,888 | 29,889 | 14.6 | 20.4 | 100 | 183 | 0.69 (0.48 - 0.98) | 0.04 | 0.73 (0.51 - 1.05) | 0.09 |
| DAG | 3,957 | 9,569 | 3.1 | 8.8 | 52 | 201 | 0.54 (0.34 - 0.82) | <0.01 | 0.52 (0.34 - 0.81) | <0.01 |
| ODOD | 11,934 | 20,464 | 8.0 | 13.1 | 77 | 152 | 0.90 (0.61 - 1.32) | 0.60 | 0.93 (0.61 - 1.41) | 0.76 |
| MDCR | 1,669 | 5,087 | 1.3 | 4.1 | 9 | 43 | 1.27 (0.48 - 3.22) | 0.62 | 1.29 (0.49 - 3.34) | 0.61 |
| MDCD | 1,311 | 3,225 | 0.7 | 1.8 | 7 | 29 | 0.86 (0.30 - 2.27) | 0.78 | 0.83 (0.30 - 2.29) | 0.72 |
| Open Claims | 232,773 | 407,388 | 189.7 | 302.3 | 813 | 1,564 | 0.86 (0.76 - 0.98) | 0.02 | 0.93 (0.79 - 1.09) | 0.41 |
| OEHR | 16,259 | 31,055 | 4.6 | 12.9 | 41 | 91 | 1.55 (0.85 - 2.83) | 0.15 | 1.56 (0.85 - 2.85) | 0.16 |
| VA | 6,862 | 21,136 | 4.0 | 15.8 | 20 | 71 | 0.77 (0.36 - 1.55) | 0.48 | 0.76 (0.37 - 1.58) | 0.47 |
| SIDIAP | 3,476 | 16,242 | 4.6 | 28.9 | 71 | 516 | 0.94 (0.69 - 1.27) | 0.70 | 1.11 (0.81 - 1.51) | 0.57 |
| Meta-analysis | 297,656 | 548,054 | 231.5 | 413.9 | 1,193 | 2,851 | 0.86 (0.78 - 0.95) | <0.01 | 0.91 (0.77 - 1.06) | 0.22 |
| Meta-analysis (w/o USOC) | 64,883 | 140,666 | 41.8 | 111.6 | 380 | 1,287 | 0.86 (0.69 - 1.07) | 0.17 | 0.88 (0.71 - 1.10) | 0.29 |
| **PS-stratification / on-treatment TAR without escalation** | | | | | | | | | | |
| CCAE | 23,075 | 29,911 | 16.7 | 20.4 | 115 | 183 | 0.78 (0.61 - 0.99) | 0.05 | 0.81 (0.63 - 1.04) | 0.10 |
| DAG | 4,896 | 9,596 | 3.6 | 8.9 | 54 | 201 | 0.73 (0.53 - 1.01) | 0.06 | 0.71 (0.51 - 0.98) | 0.05 |
| ODOD | 16,688 | 20,572 | 10.3 | 13.2 | 95 | 152 | 0.92 (0.70 - 1.22) | 0.58 | 0.92 (0.69 - 1.21) | 0.55 |
| MDCR | 2,062 | 5,143 | 1.5 | 4.1 | 10 | 43 | 0.68 (0.31 - 1.37) | 0.31 | 0.71 (0.34 - 1.49) | 0.37 |
| MDCD | 1,397 | 3,238 | 0.7 | 1.8 | 8 | 29 | 0.66 (0.27 - 1.42) | 0.32 | 0.60 (0.26 - 1.37) | 0.23 |
| Open Claims | 317,231 | 407,695 | 234.9 | 302.4 | 972 | 1,564 | 0.91 (0.83 - 0.99) | 0.04 | 0.98 (0.87 - 1.12) | 0.82 |
| OEHR | 19,457 | 31,120 | 5.4 | 12.9 | 45 | 91 | 1.43 (0.96 - 2.10) | 0.08 | 1.42 (0.96 - 2.10) | 0.09 |
| VA | 10,977 | 21,929 | 5.7 | 16.7 | 36 | 78 | 1.01 (0.61 - 1.65) | 0.98 | 1.05 (0.64 - 1.72) | 0.86 |
| SIDIAP | 4,147 | 16,590 | 5.1 | 29.8 | 76 | 536 | 1.03 (0.78 - 1.33) | 0.83 | 1.13 (0.86 - 1.48) | 0.42 |
| Meta-analysis | 387,556 | 520,627 | 277.5 | 391.8 | 1,367 | 2,770 | 0.91 (0.85 - 0.98) | <0.01 | 0.96 (0.87 - 1.07) | 0.44 |
| Meta-analysis (w/o USOC) | 70,325 | 112,932 | 42.6 | 89.3 | 395 | 1,206 | 0.91 (0.76 - 1.10) | 0.34 | 0.93 (0.77 - 1.13) | 0.49 |

##### 1.2.21 Lower extremity amputation

**Supplementary Table 47:** Exposures, outcomes and risk estimates for Lower extremity amputation in SGLT2I (male) (T) vs DPP4I (male) (C) new-user cohorts under four different propensity score (PS) adjustment / time-at-risk (TAR) designs across data sources. We report total patient exposure time in 1,000 patient-year (KYr) units, uncalibrated and calibrated hazard ratios (HRs) and their 95% confidence intervals (CIs). Greyed out entries did not pass study diagnostics and are included only for completeness.

| | Patients | | Exposure (KYr) | | Outcomes | | Uncalibrated | | Calibrated | |
|---|---|---|---|---|---|---|---|---|---|---|
| | T | C | T | C | T | C | HR (95% CI) | $p$ | HR (95% CI) | $p$ |
| **PS-matching / on-treatment TAR** | | | | | | | | | | |
| CCAE | 19,732 | 31,141 | 17.6 | 26.4 | 19 | 20 | 1.71 (0.73 - 4.10) | 0.22 | 1.84 (0.78 - 4.38) | 0.17 |
| DAG | 4,640 | 11,168 | 4.1 | 11.6 | 0 | 0 | | | | |
| ODOD | 12,486 | 21,414 | 9.6 | 16.9 | 10 | 22 | 0.35 (0.05 - 1.42) | 0.22 | 0.37 (0.07 - 1.89) | 0.23 |
| MDCR | 1,717 | 5,253 | 1.5 | 5.0 | <5 | <5 | | 0.38 | | 0.38 |
| MDCD | 1,423 | 3,450 | 0.9 | 2.5 | <5 | 9 | 0.62 (0.03 - 6.63) | 0.73 | 0.56 (0.04 - 8.53) | 0.68 |
| Open Claims | 243,789 | 425,547 | 236.7 | 408.0 | 135 | 242 | 1.18 (0.87 - 1.59) | 0.30 | 1.28 (0.93 - 1.77) | 0.14 |
| OEHR | 16,859 | 32,311 | 5.5 | 16.7 | <5 | 13 | 0.91 (0.12 - 4.56) | 0.92 | 0.92 (0.15 - 5.56) | 0.92 |
| VA | 7,780 | 23,914 | 5.3 | 21.7 | <10 | 13 | 3.13 (1.05 - 9.49) | 0.04 | 3.15 (1.05 - 9.45) | 0.04 |
| SIDIAP | 4,337 | 19,807 | 6.3 | 45.0 | 0 | 14 | | | | |
| Meta-analysis | 283,787 | 502,016 | 269.2 | 472.9 | <174 | 297 | 1.34 (0.86 - 2.08) | 0.20 | 1.42 (0.90 - 2.23) | 0.13 |
| Meta-analysis (w/o USOC) | 39,998 | 76,469 | 32.5 | 64.9 | <39 | 55 | 1.46 (0.53 - 4.08) | 0.47 | 1.51 (0.54 - 4.20) | 0.43 |
| **PS-stratification / on-treatment TAR** | | | | | | | | | | |
| CCAE | 24,122 | 31,164 | 20.1 | 26.4 | 19 | 20 | 1.57 (0.81 - 3.03) | 0.18 | 1.63 (0.84 - 3.15) | 0.15 |
| DAG | 5,763 | 11,200 | 4.7 | 11.7 | 0 | 0 | | | | |
| ODOD | 17,448 | 21,537 | 12.1 | 17.0 | 13 | 22 | 0.88 (0.41 - 1.86) | 0.75 | 0.89 (0.42 - 1.90) | 0.77 |
| MDCR | 2,129 | 5,312 | 1.7 | 5.0 | <5 | <5 | 3.26 (0.34 - 24.5) | 0.28 | 3.37 (0.40 - 28.7) | 0.27 |
| MDCD | 1,503 | 3,464 | 0.9 | 2.5 | <5 | 9 | 1.01 (0.22 - 3.51) | 0.99 | 0.91 (0.23 - 3.65) | 0.90 |
| Open Claims | 332,242 | 425,871 | 289.1 | 408.2 | 159 | 243 | 1.16 (0.93 - 1.45) | 0.18 | 1.26 (0.97 - 1.62) | 0.08 |
| OEHR | 20,221 | 32,382 | 6.5 | 16.8 | <5 | 13 | 0.60 (0.13 - 1.96) | 0.45 | 0.60 (0.15 - 2.31) | 0.46 |
| VA | 12,506 | 24,952 | 7.5 | 23.2 | 11 | 13 | 2.85 (1.11 - 7.24) | 0.03 | 2.98 (1.17 - 7.59) | 0.02 |
| SIDIAP | 5,166 | 20,241 | 7.1 | 46.6 | 0 | 16 | | | | |
| Meta-analysis | 394,033 | 510,954 | 327.7 | 468.4 | <196 | 298 | 1.16 (0.95 - 1.41) | 0.15 | 1.22 (0.97 - 1.54) | 0.10 |
| Meta-analysis (w/o USOC) | 61,791 | 85,083 | 38.7 | 60.1 | <37 | 55 | 1.11 (0.67 - 1.83) | 0.69 | 1.13 (0.69 - 1.86) | 0.63 |
| **PS-matching / on-treatment TAR without escalation** | | | | | | | | | | |
| CCAE | 19,732 | 31,141 | 15.3 | 21.3 | 14 | 17 | 1.07 (0.38 - 2.91) | 0.89 | 1.14 (0.41 - 3.17) | 0.80 |
| DAG | 4,640 | 11,168 | 3.7 | 10.5 | 0 | 0 | | | | |
| ODOD | 12,486 | 21,414 | 8.4 | 13.8 | 8 | 16 | 0.42 (0.06 - 1.73) | 0.31 | 0.43 (0.08 - 2.30) | 0.33 |
| MDCR | 1,717 | 5,253 | 1.4 | 4.2 | <5 | <5 | 3.46 (0.13 - 93.8) | 0.46 | 3.51 (0.13 - 95.0) | 0.46 |
| MDCD | 1,423 | 3,450 | 0.7 | 1.9 | <5 | 7 | 1.41 (0.05 - 36.7) | 0.83 | 1.35 (0.05 - 35.1) | 0.86 |
| Open Claims | 243,789 | 425,547 | 198.5 | 316.5 | 97 | 183 | 1.10 (0.77 - 1.56) | 0.60 | 1.18 (0.82 - 1.70) | 0.39 |
| OEHR | 16,859 | 32,311 | 4.8 | 13.5 | <5 | 10 | 0.91 (0.12 - 4.56) | 0.92 | 0.92 (0.15 - 5.56) | 0.92 |
| VA | 7,780 | 23,914 | 4.5 | 17.6 | <10 | 11 | 3.48 (1.11 - 12.1) | 0.04 | 3.46 (1.05 - 11.4) | 0.04 |
| SIDIAP | 4,337 | 19,807 | 5.6 | 35.8 | 0 | 11 | | | | |
| Meta-analysis | 276,007 | 478,102 | 222.2 | 351.6 | 119 | 216 | 1.06 (0.76 - 1.46) | 0.75 | 1.12 (0.79 - 1.58) | 0.54 |
| Meta-analysis (w/o USOC) | 32,218 | 52,555 | 23.7 | 35.2 | 22 | 33 | 0.83 (0.35 - 1.98) | 0.68 | 0.86 (0.36 - 2.04) | 0.73 |
| **PS-stratification / on-treatment TAR without escalation** | | | | | | | | | | |
| CCAE | 24,122 | 31,164 | 17.5 | 21.3 | 14 | 17 | 1.30 (0.61 - 2.70) | 0.49 | 1.35 (0.64 - 2.83) | 0.43 |
| DAG | 5,763 | 11,200 | 4.3 | 10.6 | 0 | 0 | | | | |
| ODOD | 17,448 | 21,537 | 10.8 | 13.9 | 10 | 16 | 0.97 (0.39 - 2.32) | 0.95 | 0.96 (0.40 - 2.34) | 0.93 |
| MDCR | 2,129 | 5,312 | 1.5 | 4.3 | <5 | <5 | 3.16 (0.33 - 23.4) | 0.29 | 3.29 (0.39 - 27.6) | 0.27 |
| MDCD | 1,503 | 3,464 | 0.8 | 1.9 | <5 | 7 | 1.15 (0.24 - 4.34) | 0.85 | 1.05 (0.25 - 4.48) | 0.93 |
| Open Claims | 332,242 | 425,871 | 245.8 | 316.7 | 116 | 184 | 1.07 (0.82 - 1.38) | 0.63 | 1.15 (0.88 - 1.51) | 0.32 |
| OEHR | 20,221 | 32,382 | 5.7 | 13.5 | <5 | 10 | 0.47 (0.07 - 1.91) | 0.37 | 0.47 (0.09 - 2.45) | 0.37 |
| VA | 12,506 | 24,952 | 6.5 | 18.8 | 11 | 11 | 3.12 (1.18 - 8.37) | 0.02 | 3.25 (1.22 - 8.63) | 0.02 |
| SIDIAP | 5,166 | 20,241 | 6.3 | 37.0 | 0 | 13 | | | | |
| Meta-analysis | 373,812 | 478,572 | 274.1 | 351.9 | 140 | 217 | 1.08 (0.85 - 1.37) | 0.52 | 1.14 (0.89 - 1.46) | 0.30 |
| Meta-analysis (w/o USOC) | 41,570 | 52,701 | 28.3 | 35.3 | 24 | 33 | 1.15 (0.65 - 2.03) | 0.63 | 1.18 (0.66 - 2.08) | 0.58 |

###### 1.2.22 Nausea

**Supplementary Table 48:** Exposures, outcomes and risk estimates for Nausea in SGLT2I (male) (T) vs DPP4I (male) (C) new-user cohorts under four different propensity score (PS) adjustment / time-at-risk (TAR) designs across data sources. We report total patient exposure time in 1,000 patient-year (KYr) units, uncalibrated and calibrated hazard ratios (HRs) and their 95% confidence intervals (CIs). Greyed out entries did not pass study diagnostics and are included only for completeness.

| | Patients | | Exposure (KYr) | | Outcomes | | Uncalibrated | | Calibrated | |
|---|---|---|---|---|---|---|---|---|---|---|
| | T | C | T | C | T | C | HR (95% CI) | $p$ | HR (95% CI) | $p$ |
| **PS-matching / on-treatment TAR** | | | | | | | | | | |
| CCAE | 17,863 | 28,402 | 15.7 | 23.6 | 384 | 640 | 0.93 (0.78 - 1.11) | 0.45 | 1.01 (0.84 - 1.20) | 0.84 |
| DAG | 4,388 | 10,521 | 3.8 | 10.9 | 28 | 94 | 1.00 (0.58 - 1.69) | 0.99 | 0.98 (0.57 - 1.68) | 0.81 |
| ODOD | 11,162 | 19,099 | 8.5 | 14.8 | 238 | 494 | 0.94 (0.75 - 1.17) | 0.58 | 0.97 (0.77 - 1.22) | 0.82 |
| MDCR | 1,574 | 4,783 | 1.4 | 4.5 | 35 | 117 | 1.33 (0.80 - 2.16) | 0.27 | 1.33 (0.81 - 2.20) | 0.28 |
| MDCD | 1,144 | 2,754 | 0.7 | 2.0 | 43 | 122 | 1.24 (0.79 - 1.93) | 0.34 | 1.12 (0.71 - 1.76) | 0.64 |
| Open Claims | 220,752 | 387,306 | 214.5 | 369.1 | 2,628 | 5,718 | 0.88 (0.82 - 0.94) | <0.01 | 0.95 (0.85 - 1.08) | 0.50 |
| OEHR | 15,676 | 30,166 | 5.1 | 15.3 | 142 | 398 | 1.19 (0.91 - 1.55) | 0.21 | 1.20 (0.92 - 1.56) | 0.21 |
| VA | 7,229 | 22,387 | 4.9 | 20.3 | 31 | 168 | 0.83 (0.49 - 1.36) | 0.48 | 0.84 (0.50 - 1.39) | 0.50 |
| SIDIAP | 4,206 | 19,292 | 6.2 | 43.8 | 16 | 224 | 0.55 (0.27 - 1.01) | 0.08 | 0.60 (0.31 - 1.16) | 0.15 |
| Meta-analysis | 282,850 | 521,956 | 260.1 | 502.3 | 3,502 | 7,853 | 0.93 (0.85 - 1.02) | 0.12 | 0.99 (0.86 - 1.13) | 0.83 |
| Meta-analysis (w/o USOC) | 62,098 | 134,650 | 45.6 | 133.2 | 874 | 2,135 | 0.98 (0.86 - 1.12) | 0.74 | 1.01 (0.88 - 1.15) | 0.86 |
| **PS-stratification / on-treatment TAR** | | | | | | | | | | |
| CCAE | 21,821 | 28,422 | 17.8 | 23.6 | 432 | 640 | 0.88 (0.78 - 1.00) | 0.06 | 0.92 (0.80 - 1.05) | 0.22 |
| DAG | 5,453 | 10,548 | 4.5 | 11.0 | 34 | 94 | 1.07 (0.69 - 1.64) | 0.76 | 1.03 (0.67 - 1.59) | 0.80 |
| ODOD | 15,574 | 19,195 | 10.7 | 14.8 | 302 | 502 | 0.92 (0.78 - 1.07) | 0.28 | 0.92 (0.79 - 1.09) | 0.38 |
| MDCR | 1,958 | 4,835 | 1.5 | 4.5 | 38 | 120 | 1.16 (0.76 - 1.73) | 0.48 | 1.19 (0.79 - 1.80) | 0.41 |
| MDCD | 1,195 | 2,764 | 0.7 | 2.0 | 43 | 122 | 1.09 (0.75 - 1.55) | 0.65 | 0.98 (0.68 - 1.42) | 0.83 |
| Open Claims | 299,241 | 387,604 | 261.1 | 369.3 | 3,193 | 5,727 | 0.87 (0.83 - 0.91) | <0.01 | 0.94 (0.82 - 1.08) | 0.41 |
| OEHR | 18,784 | 30,230 | 5.9 | 15.3 | 162 | 402 | 1.13 (0.92 - 1.38) | 0.23 | 1.13 (0.93 - 1.39) | 0.26 |
| VA | 11,542 | 23,328 | 7.0 | 21.7 | 53 | 181 | 0.69 (0.47 - 1.02) | 0.07 | 0.73 (0.49 - 1.07) | 0.12 |
| SIDIAP | 5,004 | 19,705 | 6.9 | 45.2 | 20 | 236 | 0.57 (0.33 - 0.92) | 0.03 | 0.58 (0.35 - 0.97) | 0.05 |
| Meta-analysis | 367,835 | 500,539 | 308.5 | 483.8 | 4,181 | 7,721 | 0.92 (0.84 - 1.00) | 0.05 | 0.97 (0.83 - 1.12) | 0.65 |
| Meta-analysis (w/o USOC) | 68,594 | 112,935 | 47.4 | 114.5 | 988 | 1,994 | 0.95 (0.83 - 1.09) | 0.47 | 0.97 (0.84 - 1.11) | 0.69 |
| **PS-matching / on-treatment TAR without escalation** | | | | | | | | | | |
| CCAE | 17,863 | 28,402 | 13.7 | 19.2 | 318 | 526 | 0.93 (0.76 - 1.12) | 0.43 | 0.98 (0.81 - 1.19) | 0.83 |
| DAG | 4,388 | 10,521 | 3.5 | 9.9 | 26 | 84 | 1.04 (0.59 - 1.79) | 0.88 | 1.01 (0.58 - 1.76) | 0.80 |
| ODOD | 11,162 | 19,099 | 7.5 | 12.2 | 201 | 399 | 0.88 (0.69 - 1.12) | 0.29 | 0.90 (0.68 - 1.21) | 0.58 |
| MDCR | 1,574 | 4,783 | 1.2 | 3.8 | 32 | 105 | 1.39 (0.81 - 2.32) | 0.22 | 1.41 (0.83 - 2.37) | 0.23 |
| MDCD | 1,144 | 2,754 | 0.6 | 1.5 | 37 | 97 | 1.13 (0.69 - 1.85) | 0.62 | 1.08 (0.66 - 1.78) | 0.75 |
| Open Claims | 220,752 | 387,306 | 180.6 | 287.5 | 2,065 | 4,359 | 0.84 (0.78 - 0.91) | <0.01 | 0.91 (0.80 - 1.03) | 0.18 |
| OEHR | 15,676 | 30,166 | 4.5 | 12.5 | 114 | 291 | 1.15 (0.84 - 1.56) | 0.37 | 1.16 (0.85 - 1.58) | 0.38 |
| VA | 7,229 | 22,387 | 4.2 | 16.6 | 22 | 137 | 0.74 (0.40 - 1.28) | 0.30 | 0.73 (0.41 - 1.31) | 0.30 |
| SIDIAP | 4,206 | 19,292 | 5.5 | 34.8 | 14 | 186 | 0.52 (0.24 - 1.02) | 0.08 | 0.61 (0.29 - 1.27) | 0.20 |
| Meta-analysis | 282,850 | 521,956 | 220.7 | 396.5 | 2,792 | 6,087 | 0.90 (0.81 - 0.99) | 0.03 | 0.95 (0.81 - 1.11) | 0.50 |
| Meta-analysis (w/o USOC) | 62,098 | 134,650 | 40.1 | 108.9 | 727 | 1,728 | 0.95 (0.82 - 1.10) | 0.50 | 0.98 (0.84 - 1.14) | 0.79 |
| **PS-stratification / on-treatment TAR without escalation** | | | | | | | | | | |
| CCAE | 21,821 | 28,422 | 15.7 | 19.2 | 359 | 526 | 0.84 (0.73 - 0.97) | 0.02 | 0.88 (0.76 - 1.01) | 0.09 |
| DAG | 5,453 | 10,548 | 4.1 | 9.9 | 31 | 84 | 1.06 (0.67 - 1.66) | 0.79 | 1.03 (0.65 - 1.63) | 0.81 |
| ODOD | 15,574 | 19,195 | 9.6 | 12.2 | 256 | 407 | 0.89 (0.75 - 1.06) | 0.20 | 0.88 (0.74 - 1.05) | 0.21 |
| MDCR | 1,958 | 4,835 | 1.4 | 3.9 | 35 | 108 | 1.17 (0.76 - 1.78) | 0.46 | 1.22 (0.79 - 1.88) | 0.38 |
| MDCD | 1,195 | 2,764 | 0.6 | 1.5 | 37 | 97 | 1.14 (0.76 - 1.67) | 0.53 | 1.04 (0.70 - 1.55) | 0.80 |
| Open Claims | 299,241 | 387,604 | 222.9 | 287.7 | 2,542 | 4,368 | 0.85 (0.81 - 0.90) | <0.01 | 0.92 (0.83 - 1.03) | 0.17 |
| OEHR | 18,784 | 30,230 | 5.2 | 12.5 | 130 | 294 | 1.09 (0.87 - 1.37) | 0.43 | 1.09 (0.86 - 1.37) | 0.50 |
| VA | 11,542 | 23,328 | 6.0 | 17.7 | 41 | 148 | 0.61 (0.39 - 0.94) | 0.03 | 0.63 (0.41 - 0.98) | 0.05 |
| SIDIAP | 5,004 | 19,705 | 6.1 | 35.9 | 18 | 196 | 0.53 (0.30 - 0.89) | 0.02 | 0.58 (0.34 - 1.00) | 0.06 |
| Meta-analysis | 367,835 | 500,539 | 265.0 | 381.4 | 3,371 | 5,983 | 0.88 (0.82 - 0.94) | <0.01 | 0.93 (0.84 - 1.02) | 0.13 |
| Meta-analysis (w/o USOC) | 68,594 | 112,935 | 42.1 | 93.6 | 829 | 1,615 | 0.92 (0.80 - 1.07) | 0.28 | 0.94 (0.81 - 1.10) | 0.47 |

##### 1.2.23 Peripheral edema

**Supplementary Table 49:** Exposures, outcomes and risk estimates for Peripheral edema in SGLT2I (male) (T) vs DPP4I (male) (C) new-user cohorts under four different propensity score (PS) adjustment / time-at-risk (TAR) designs across data sources. We report total patient exposure time in 1,000 patient-year (KYr) units, uncalibrated and calibrated hazard ratios (HRs) and their 95% confidence intervals (CIs). Greyed out entries did not pass study diagnostics and are included only for completeness.

| | Patients | | Exposure (KYr) | | Outcomes | | Uncalibrated | | Calibrated | |
|---|---|---|---|---|---|---|---|---|---|---|
| | T | C | T | C | T | C | HR (95% CI) | $p$ | HR (95% CI) | $p$ |
| **PS-matching / on-treatment TAR** | | | | | | | | | | |
| CCAE | 16,678 | 26,547 | 14.5 | 21.8 | 457 | 817 | 0.85 (0.72 - 1.00) | 0.04 | 0.91 (0.77 - 1.08) | 0.31 |
| DAG | 4,104 | 9,849 | 3.5 | 10.1 | 48 | 235 | 0.70 (0.46 - 1.04) | 0.09 | 0.69 (0.46 - 1.03) | 0.09 |
| ODOD | 10,187 | 16,998 | 7.7 | 12.9 | 338 | 813 | 0.73 (0.60 - 0.88) | <0.01 | 0.75 (0.62 - 0.92) | 0.02 |
| MDCR | 1,301 | 3,926 | 1.1 | 3.6 | 50 | 285 | 0.60 (0.39 - 0.91) | 0.02 | 0.60 (0.39 - 0.93) | 0.03 |
| MDCD | 1,057 | 2,501 | 0.6 | 1.7 | 37 | 135 | 0.68 (0.42 - 1.08) | 0.12 | 0.62 (0.38 - 0.99) | 0.06 |
| Open Claims | 200,483 | 347,984 | 193.9 | 325.3 | 3,470 | 9,842 | 0.71 (0.68 - 0.76) | <0.01 | 0.78 (0.69 - 0.87) | <0.01 |
| OEHR | 14,800 | 28,212 | 4.8 | 14.0 | 107 | 540 | 0.61 (0.45 - 0.81) | <0.01 | 0.61 (0.46 - 0.82) | <0.01 |
| VA | 5,997 | 18,905 | 4.0 | 16.8 | 108 | 570 | 0.91 (0.70 - 1.19) | 0.51 | 0.92 (0.70 - 1.20) | 0.57 |
| SIDIAP | 3,910 | 18,105 | 5.7 | 40.6 | 59 | 694 | 0.68 (0.49 - 0.94) | 0.02 | 0.74 (0.53 - 1.04) | 0.12 |
| Meta-analysis | 259,139 | 476,856 | 236.9 | 452.9 | 4,652 | 13,933 | 0.74 (0.68 - 0.80) | <0.01 | 0.78 (0.69 - 0.89) | <0.01 |
| Meta-analysis (w/o USOC) | 58,656 | 128,872 | 43.0 | 127.6 | 1,182 | 4,091 | 0.76 (0.68 - 0.84) | <0.01 | 0.78 (0.70 - 0.87) | <0.01 |
| **PS-stratification / on-treatment TAR** | | | | | | | | | | |
| CCAE | 20,331 | 26,564 | 16.5 | 21.8 | 518 | 820 | 0.87 (0.78 - 0.98) | 0.02 | 0.91 (0.80 - 1.02) | 0.13 |
| DAG | 5,059 | 9,872 | 4.1 | 10.1 | 57 | 236 | 0.65 (0.47 - 0.88) | <0.01 | 0.62 (0.45 - 0.85) | <0.01 |
| ODOD | 14,095 | 17,061 | 9.6 | 12.9 | 406 | 820 | 0.77 (0.68 - 0.88) | <0.01 | 0.78 (0.68 - 0.89) | <0.01 |
| MDCR | 1,633 | 3,973 | 1.3 | 3.6 | 56 | 294 | 0.62 (0.45 - 0.84) | <0.01 | 0.64 (0.46 - 0.87) | <0.01 |
| MDCD | 1,095 | 2,510 | 0.6 | 1.7 | 37 | 138 | 0.83 (0.56 - 1.21) | 0.35 | 0.75 (0.51 - 1.10) | 0.17 |
| Open Claims | 269,915 | 348,152 | 235.1 | 325.4 | 4,195 | 9,863 | 0.73 (0.70 - 0.76) | <0.01 | 0.79 (0.69 - 0.90) | <0.01 |
| OEHR | 17,562 | 28,266 | 5.5 | 14.0 | 130 | 544 | 0.75 (0.61 - 0.92) | <0.01 | 0.75 (0.61 - 0.93) | 0.01 |
| VA | 9,289 | 19,618 | 5.6 | 17.8 | 177 | 603 | 0.82 (0.66 - 1.01) | 0.06 | 0.86 (0.69 - 1.06) | 0.18 |
| SIDIAP | 4,631 | 18,482 | 6.4 | 41.9 | 66 | 729 | 0.63 (0.47 - 0.83) | <0.01 | 0.65 (0.49 - 0.86) | <0.01 |
| Meta-analysis | 333,226 | 452,370 | 278.5 | 429.8 | 5,428 | 13,306 | 0.75 (0.69 - 0.81) | <0.01 | 0.79 (0.68 - 0.91) | <0.01 |
| Meta-analysis (w/o USOC) | 63,311 | 104,218 | 43.4 | 104.3 | 1,233 | 3,443 | 0.75 (0.67 - 0.84) | <0.01 | 0.76 (0.68 - 0.86) | <0.01 |
| **PS-matching / on-treatment TAR without escalation** | | | | | | | | | | |
| CCAE | 16,678 | 26,547 | 12.6 | 17.7 | 381 | 664 | 0.89 (0.74 - 1.06) | 0.18 | 0.94 (0.79 - 1.13) | 0.54 |
| DAG | 4,104 | 9,849 | 3.2 | 9.1 | 45 | 222 | 0.70 (0.46 - 1.04) | 0.09 | 0.67 (0.45 - 1.02) | 0.08 |
| ODOD | 10,187 | 16,998 | 6.8 | 10.6 | 271 | 674 | 0.71 (0.58 - 0.87) | <0.01 | 0.73 (0.56 - 0.96) | 0.07 |
| MDCR | 1,301 | 3,926 | 1.0 | 3.1 | 42 | 244 | 0.54 (0.33 - 0.85) | 0.01 | 0.55 (0.34 - 0.88) | 0.02 |
| MDCD | 1,057 | 2,501 | 0.5 | 1.3 | 30 | 117 | 0.58 (0.33 - 0.97) | 0.05 | 0.56 (0.33 - 0.95) | 0.04 |
| Open Claims | 200,483 | 347,984 | 163.5 | 253.6 | 2,832 | 7,676 | 0.72 (0.68 - 0.76) | <0.01 | 0.77 (0.68 - 0.87) | <0.01 |
| OEHR | 14,800 | 28,212 | 4.2 | 11.5 | 88 | 399 | 0.62 (0.44 - 0.85) | <0.01 | 0.62 (0.45 - 0.87) | <0.01 |
| VA | 5,997 | 18,905 | 3.5 | 13.7 | 89 | 450 | 0.86 (0.63 - 1.15) | 0.31 | 0.85 (0.63 - 1.15) | 0.32 |
| SIDIAP | 3,910 | 18,105 | 5.1 | 32.3 | 53 | 551 | 0.65 (0.45 - 0.92) | 0.02 | 0.77 (0.53 - 1.10) | 0.21 |
| Meta-analysis | 259,139 | 476,856 | 201.4 | 358.2 | 3,816 | 10,997 | 0.74 (0.68 - 0.80) | <0.01 | 0.78 (0.67 - 0.90) | <0.01 |
| Meta-analysis (w/o USOC) | 58,656 | 128,872 | 37.9 | 104.6 | 984 | 3,321 | 0.75 (0.66 - 0.84) | <0.01 | 0.77 (0.68 - 0.87) | <0.01 |
| **PS-stratification / on-treatment TAR without escalation** | | | | | | | | | | |
| CCAE | 20,331 | 26,564 | 14.5 | 17.8 | 440 | 667 | 0.85 (0.74 - 0.96) | <0.01 | 0.88 (0.77 - 1.00) | 0.07 |
| DAG | 5,059 | 9,872 | 3.8 | 9.1 | 53 | 223 | 0.63 (0.45 - 0.87) | <0.01 | 0.61 (0.44 - 0.85) | <0.01 |
| ODOD | 14,095 | 17,061 | 8.6 | 10.7 | 336 | 680 | 0.72 (0.62 - 0.83) | <0.01 | 0.71 (0.62 - 0.83) | <0.01 |
| MDCR | 1,633 | 3,973 | 1.1 | 3.1 | 48 | 252 | 0.60 (0.43 - 0.84) | <0.01 | 0.63 (0.45 - 0.88) | 0.01 |
| MDCD | 1,095 | 2,510 | 0.5 | 1.3 | 30 | 120 | 0.69 (0.45 - 1.04) | 0.09 | 0.64 (0.42 - 0.97) | 0.04 |
| Open Claims | 269,915 | 348,152 | 200.8 | 253.7 | 3,475 | 7,695 | 0.72 (0.69 - 0.75) | <0.01 | 0.78 (0.70 - 0.86) | <0.01 |
| OEHR | 17,562 | 28,266 | 4.9 | 11.5 | 105 | 399 | 0.75 (0.60 - 0.95) | 0.02 | 0.75 (0.59 - 0.95) | 0.02 |
| VA | 9,289 | 19,618 | 4.8 | 14.5 | 151 | 478 | 0.84 (0.66 - 1.06) | 0.14 | 0.87 (0.69 - 1.10) | 0.29 |
| SIDIAP | 4,631 | 18,482 | 5.7 | 33.3 | 60 | 578 | 0.63 (0.47 - 0.85) | <0.01 | 0.70 (0.52 - 0.94) | 0.03 |
| Meta-analysis | 333,226 | 452,370 | 239.4 | 339.2 | 4,517 | 10,494 | 0.73 (0.68 - 0.79) | <0.01 | 0.77 (0.69 - 0.86) | <0.01 |
| Meta-analysis (w/o USOC) | 63,311 | 104,218 | 38.6 | 85.5 | 1,042 | 2,799 | 0.73 (0.66 - 0.81) | <0.01 | 0.75 (0.67 - 0.84) | <0.01 |

##### 1.2.24 Photosensitivity

**Supplementary Table 50:** Exposures, outcomes and risk estimates for Photosensitivity in SGLT2I (male) (T) vs DPP4I (male) (C) new-user cohorts under four different propensity score (PS) adjustment / time-at-risk (TAR) designs across data sources. We report total patient exposure time in 1,000 patient-year (KYr) units, uncalibrated and calibrated hazard ratios (HRs) and their 95% confidence intervals (CIs). Greyed out entries did not pass study diagnostics and are included only for completeness.

| | Patients | | Exposure (KYr) | | Outcomes | | Uncalibrated | | Calibrated | |
|---|---|---|---|---|---|---|---|---|---|---|
| | T | C | T | C | T | C | HR (95% CI) | $p$ | HR (95% CI) | $p$ |
| **PS-matching / on-treatment TAR** | | | | | | | | | | |
| CCAE | 19,583 | 30,894 | 17.5 | 26.1 | 10 | 23 | 1.00 (0.36 - 2.52) | 0.99 | 1.07 (0.41 - 2.82) | 0.89 |
| DAG | 4,625 | 11,133 | 4.0 | 11.6 | 0 | <5 | | | | |
| ODOD | 12,399 | 21,234 | 9.5 | 16.7 | 6 | 23 | 0.22 (0.01 - 1.21) | 0.20 | 0.23 (0.02 - 2.32) | 0.21 |
| MDCR | 1,690 | 5,139 | 1.5 | 4.8 | <5 | 14 | 1.12 (0.06 - 6.96) | 0.92 | 1.13 (0.10 - 12.4) | 0.92 |
| MDCD | 1,413 | 3,447 | 0.9 | 2.5 | 0 | 0 | | | | |
| Open Claims | 241,800 | 421,909 | 234.6 | 403.9 | 65 | 213 | 0.91 (0.61 - 1.33) | 0.64 | 0.99 (0.67 - 1.48) | 0.96 |
| OEHR | 16,774 | 32,082 | 5.5 | 16.6 | 5 | 21 | 1.16 (0.30 - 3.76) | 0.82 | 1.17 (0.33 - 4.13) | 0.81 |
| VA | 7,694 | 23,539 | 5.2 | 21.2 | <10 | <10 | 0.82 (0.04 - 9.91) | 0.89 | 0.82 (0.05 - 13.8) | 0.89 |
| SIDIAP | 4,334 | 19,796 | 6.3 | 45.0 | <5 | <5 | | 0.18 | | 0.16 |
| Meta-analysis | 290,556 | 506,119 | 267.0 | 463.3 | 86 | 280 | 0.91 (0.65 - 1.28) | 0.59 | 0.97 (0.68 - 1.38) | 0.85 |
| Meta-analysis (w/o USOC) | 48,756 | 84,210 | 32.4 | 59.4 | 21 | 67 | 0.90 (0.44 - 1.87) | 0.78 | 0.93 (0.45 - 1.93) | 0.85 |
| **PS-stratification / on-treatment TAR** | | | | | | | | | | |
| CCAE | 23,945 | 30,916 | 19.9 | 26.1 | 11 | 23 | 0.89 (0.41 - 1.85) | 0.77 | 0.93 (0.43 - 1.98) | 0.84 |
| DAG | 5,747 | 11,165 | 4.7 | 11.7 | 0 | <5 | | | | |
| ODOD | 17,348 | 21,351 | 12.0 | 16.8 | 8 | 23 | 0.43 (0.17 - 1.03) | 0.07 | 0.43 (0.17 - 1.08) | 0.07 |
| MDCR | 2,105 | 5,194 | 1.7 | 4.9 | <5 | 15 | 0.86 (0.13 - 3.31) | 0.86 | 0.89 (0.18 - 4.48) | 0.89 |
| MDCD | 1,495 | 3,460 | 0.9 | 2.5 | 0 | 0 | | | | |
| Open Claims | 329,565 | 422,234 | 286.5 | 404.2 | 68 | 214 | 0.85 (0.63 - 1.14) | 0.30 | 0.92 (0.67 - 1.27) | 0.62 |
| OEHR | 20,107 | 32,216 | 6.4 | 16.6 | 5 | 21 | 1.11 (0.35 - 2.98) | 0.84 | 1.12 (0.38 - 3.26) | 0.84 |
| VA | 12,352 | 24,648 | 7.4 | 22.9 | <10 | <10 | 0.76 (0.03 - 7.17) | 0.84 | 0.79 (0.05 - 11.5) | 0.86 |
| SIDIAP | 5,159 | 20,233 | 7.1 | 46.6 | <5 | <5 | 4.24 (0.50 - 30.9) | 0.17 | 4.36 (0.55 - 34.4) | 0.16 |
| Meta-analysis | 393,070 | 511,911 | 326.6 | 468.6 | <97 | 296 | 0.83 (0.64 - 1.06) | 0.14 | 0.87 (0.66 - 1.15) | 0.33 |
| Meta-analysis (w/o USOC) | 63,505 | 89,677 | 40.0 | 64.4 | <29 | 82 | 0.76 (0.46 - 1.23) | 0.26 | 0.77 (0.47 - 1.26) | 0.30 |
| **PS-matching / on-treatment TAR without escalation** | | | | | | | | | | |
| CCAE | 19,583 | 30,894 | 15.1 | 21.1 | 9 | 19 | 0.97 (0.33 - 2.65) | 0.96 | 1.03 (0.36 - 2.94) | 0.95 |
| DAG | 4,625 | 11,133 | 3.7 | 10.5 | 0 | <5 | | | | |
| ODOD | 12,399 | 21,234 | 8.3 | 13.7 | 5 | 18 | 0.27 (0.01 - 1.57) | 0.28 | 0.28 (0.03 - 2.95) | 0.29 |
| MDCR | 1,690 | 5,139 | 1.3 | 4.1 | <5 | 13 | 1.42 (0.07 - 9.38) | 0.78 | 1.44 (0.13 - 16.4) | 0.77 |
| MDCD | 1,413 | 3,447 | 0.7 | 1.9 | 0 | 0 | | | | |
| Open Claims | 241,800 | 421,909 | 196.7 | 313.3 | 52 | 180 | 0.99 (0.65 - 1.50) | 0.98 | 1.07 (0.69 - 1.64) | 0.77 |
| OEHR | 16,774 | 32,082 | 4.8 | 13.4 | <5 | 19 | 1.75 (0.44 - 6.07) | 0.40 | 1.76 (0.47 - 6.56) | 0.40 |
| VA | 7,694 | 23,539 | 4.4 | 17.3 | <10 | <10 | 0.82 (0.04 - 9.90) | 0.89 | 0.81 (0.05 - 13.7) | 0.89 |
| SIDIAP | 4,334 | 19,796 | 5.6 | 35.8 | <5 | <5 | | 0.19 | | 0.15 |
| Meta-analysis | 290,556 | 506,119 | 224.9 | 361.4 | <71 | 236 | 1.00 (0.69 - 1.45) | 0.98 | 1.06 (0.72 - 1.56) | 0.76 |
| Meta-analysis (w/o USOC) | 48,756 | 84,210 | 28.2 | 48.2 | <19 | 56 | 1.04 (0.48 - 2.25) | 0.92 | 1.07 (0.49 - 2.32) | 0.86 |
| **PS-stratification / on-treatment TAR without escalation** | | | | | | | | | | |
| CCAE | 23,945 | 30,916 | 17.4 | 21.1 | 10 | 19 | 0.93 (0.40 - 2.03) | 0.85 | 0.96 (0.43 - 2.17) | 0.93 |
| DAG | 5,747 | 11,165 | 4.3 | 10.5 | 0 | <5 | | | | |
| ODOD | 17,348 | 21,351 | 10.7 | 13.8 | 7 | 18 | 0.44 (0.15 - 1.18) | 0.12 | 0.43 (0.15 - 1.22) | 0.12 |
| MDCR | 2,105 | 5,194 | 1.5 | 4.2 | <5 | 14 | 0.92 (0.14 - 3.63) | 0.92 | 0.95 (0.19 - 4.90) | 0.95 |
| MDCD | 1,495 | 3,460 | 0.8 | 1.9 | 0 | 0 | | | | |
| Open Claims | 329,565 | 422,234 | 243.6 | 313.5 | 55 | 181 | 0.79 (0.56 - 1.09) | 0.15 | 0.85 (0.61 - 1.20) | 0.36 |
| OEHR | 20,107 | 32,216 | 5.7 | 13.4 | <5 | 19 | 0.93 (0.26 - 2.67) | 0.90 | 0.92 (0.28 - 2.98) | 0.89 |
| VA | 12,352 | 24,648 | 6.4 | 18.6 | <10 | <10 | 0.75 (0.03 - 7.18) | 0.83 | 0.78 (0.05 - 11.5) | 0.86 |
| SIDIAP | 5,159 | 20,233 | 6.3 | 37.0 | <5 | <5 | 4.09 (0.48 - 29.3) | 0.18 | 4.48 (0.57 - 35.0) | 0.15 |
| Meta-analysis | 390,965 | 506,717 | 277.3 | 361.8 | <77 | 237 | 0.78 (0.58 - 1.03) | 0.08 | 0.82 (0.61 - 1.10) | 0.18 |
| Meta-analysis (w/o USOC) | 61,400 | 84,483 | 33.7 | 48.3 | <22 | 56 | 0.74 (0.42 - 1.30) | 0.30 | 0.76 (0.43 - 1.33) | 0.34 |

##### 1.2.25 Venous thromboembolism

**Supplementary Table 51:** Exposures, outcomes and risk estimates for Venous thromboembolism in SGLT2I (male) (T) vs DPP4I (male) (C) new-user cohorts under four different propensity score (PS) adjustment / time-at-risk (TAR) designs across data sources. We report total patient exposure time in 1,000 patient-year (KYr) units, uncalibrated and calibrated hazard ratios (HRs) and their 95% confidence intervals (CIs). Greyed out entries did not pass study diagnostics and are included only for completeness.

| | Patients | | Exposure (KYr) | | Outcomes | | Uncalibrated | | Calibrated | |
|---|---|---|---|---|---|---|---|---|---|---|
| | T | C | T | C | T | C | HR (95% CI) | $p$ | HR (95% CI) | $p$ |
| **PS-matching / on-treatment TAR** | | | | | | | | | | |
| CCAE | 19,330 | 30,468 | 17.2 | 25.7 | 66 | 124 | 0.94 (0.61 - 1.43) | 0.76 | 1.01 (0.66 - 1.55) | 0.93 |
| DAG | 4,400 | 10,540 | 3.8 | 10.9 | 24 | 71 | 1.04 (0.56 - 1.86) | 0.90 | 1.02 (0.56 - 1.85) | 0.82 |
| ODOD | 12,183 | 20,779 | 9.3 | 16.3 | 59 | 147 | 0.88 (0.59 - 1.31) | 0.54 | 0.91 (0.61 - 1.37) | 0.68 |
| MDCR | 1,645 | 5,019 | 1.4 | 4.7 | 12 | 57 | 1.05 (0.46 - 2.31) | 0.90 | 1.06 (0.47 - 2.39) | 0.88 |
| MDCD | 1,375 | 3,292 | 0.8 | 2.4 | 10 | 33 | 1.08 (0.42 - 2.66) | 0.87 | 0.98 (0.38 - 2.47) | 0.91 |
| Open Claims | 236,728 | 412,478 | 229.8 | 394.5 | 783 | 1,936 | 0.84 (0.74 - 0.94) | <0.01 | 0.91 (0.78 - 1.07) | 0.28 |
| OEHR | 16,473 | 31,512 | 5.3 | 16.2 | 36 | 102 | 1.40 (0.82 - 2.37) | 0.21 | 1.41 (0.83 - 2.41) | 0.21 |
| VA | 7,470 | 23,118 | 5.1 | 20.9 | 25 | 97 | 1.18 (0.63 - 2.11) | 0.60 | 1.18 (0.65 - 2.16) | 0.59 |
| SIDIAP | 4,220 | 19,368 | 6.1 | 44.0 | 22 | 170 | 0.90 (0.52 - 1.50) | 0.70 | 0.98 (0.57 - 1.68) | 0.87 |
| Meta-analysis | 304,347 | 560,749 | 280.1 | 542.7 | 1,035 | 2,736 | 0.93 (0.80 - 1.07) | 0.29 | 0.98 (0.83 - 1.17) | 0.84 |
| Meta-analysis (w/o USOC) | 67,619 | 148,271 | 50.2 | 148.3 | 252 | 800 | 1.03 (0.84 - 1.25) | 0.80 | 1.06 (0.87 - 1.29) | 0.59 |
| **PS-stratification / on-treatment TAR** | | | | | | | | | | |
| CCAE | 23,622 | 30,488 | 19.6 | 25.7 | 77 | 126 | 0.79 (0.58 - 1.06) | 0.12 | 0.82 (0.61 - 1.10) | 0.19 |
| DAG | 5,471 | 10,567 | 4.5 | 10.9 | 27 | 72 | 1.05 (0.63 - 1.69) | 0.85 | 1.01 (0.62 - 1.64) | 0.84 |
| ODOD | 16,980 | 20,888 | 11.8 | 16.4 | 81 | 148 | 0.89 (0.65 - 1.20) | 0.45 | 0.90 (0.66 - 1.22) | 0.51 |
| MDCR | 2,048 | 5,074 | 1.6 | 4.8 | 15 | 57 | 1.11 (0.56 - 2.08) | 0.76 | 1.14 (0.59 - 2.20) | 0.69 |
| MDCD | 1,452 | 3,305 | 0.9 | 2.4 | 10 | 33 | 0.85 (0.38 - 1.73) | 0.67 | 0.76 (0.36 - 1.63) | 0.50 |
| Open Claims | 321,867 | 412,766 | 280.3 | 394.7 | 985 | 1,944 | 0.88 (0.81 - 0.96) | <0.01 | 0.95 (0.81 - 1.11) | 0.53 |
| OEHR | 19,746 | 31,577 | 6.3 | 16.2 | 42 | 102 | 1.29 (0.87 - 1.89) | 0.20 | 1.29 (0.87 - 1.92) | 0.21 |
| VA | 11,951 | 24,130 | 7.2 | 22.4 | 44 | 99 | 1.05 (0.67 - 1.65) | 0.82 | 1.10 (0.70 - 1.74) | 0.68 |
| SIDIAP | 5,024 | 19,780 | 6.9 | 45.5 | 26 | 179 | 1.00 (0.61 - 1.58) | 1.00 | 1.03 (0.64 - 1.65) | 0.85 |
| Meta-analysis | 394,758 | 531,140 | 330.9 | 514.3 | 1,253 | 2,628 | 0.90 (0.83 - 0.97) | <0.01 | 0.94 (0.82 - 1.09) | 0.41 |
| Meta-analysis (w/o USOC) | 72,891 | 118,374 | 50.6 | 119.6 | 268 | 684 | 0.95 (0.81 - 1.12) | 0.56 | 0.97 (0.83 - 1.14) | 0.75 |
| **PS-matching / on-treatment TAR without escalation** | | | | | | | | | | |
| CCAE | 19,330 | 30,468 | 14.9 | 20.8 | 53 | 106 | 0.90 (0.55 - 1.44) | 0.66 | 0.95 (0.59 - 1.54) | 0.85 |
| DAG | 4,400 | 10,540 | 3.5 | 9.8 | 23 | 66 | 1.03 (0.54 - 1.89) | 0.93 | 1.00 (0.53 - 1.87) | 0.82 |
| ODOD | 12,183 | 20,779 | 8.2 | 13.4 | 44 | 125 | 0.78 (0.50 - 1.19) | 0.26 | 0.80 (0.50 - 1.28) | 0.39 |
| MDCR | 1,645 | 5,019 | 1.3 | 4.0 | 12 | 45 | 1.35 (0.55 - 3.16) | 0.50 | 1.37 (0.57 - 3.29) | 0.49 |
| MDCD | 1,375 | 3,292 | 0.7 | 1.8 | 9 | 28 | 1.06 (0.38 - 2.72) | 0.91 | 1.01 (0.38 - 2.70) | 0.90 |
| Open Claims | 236,728 | 412,478 | 192.9 | 306.3 | 630 | 1,507 | 0.84 (0.73 - 0.96) | <0.01 | 0.90 (0.76 - 1.06) | 0.25 |
| OEHR | 16,473 | 31,512 | 4.7 | 13.1 | 26 | 78 | 1.52 (0.80 - 2.84) | 0.19 | 1.53 (0.81 - 2.89) | 0.19 |
| VA | 7,470 | 23,118 | 4.3 | 17.0 | 21 | 72 | 1.08 (0.55 - 2.04) | 0.83 | 1.07 (0.55 - 2.07) | 0.84 |
| SIDIAP | 4,220 | 19,368 | 5.5 | 34.9 | 19 | 120 | 0.94 (0.51 - 1.65) | 0.83 | 1.10 (0.61 - 2.00) | 0.76 |
| Meta-analysis | 304,347 | 560,749 | 237.0 | 427.3 | 835 | 2,147 | 0.89 (0.78 - 1.00) | 0.05 | 0.94 (0.79 - 1.11) | 0.46 |
| Meta-analysis (w/o USOC) | 67,619 | 148,271 | 44.1 | 121.0 | 205 | 640 | 0.99 (0.80 - 1.23) | 0.94 | 1.02 (0.82 - 1.27) | 0.84 |
| **PS-stratification / on-treatment TAR without escalation** | | | | | | | | | | |
| CCAE | 23,622 | 30,488 | 17.2 | 20.8 | 64 | 108 | 0.71 (0.51 - 0.98) | 0.04 | 0.74 (0.53 - 1.02) | 0.07 |
| DAG | 5,471 | 10,567 | 4.1 | 9.9 | 26 | 67 | 1.11 (0.66 - 1.81) | 0.69 | 1.07 (0.65 - 1.78) | 0.77 |
| ODOD | 16,980 | 20,888 | 10.4 | 13.5 | 66 | 126 | 0.80 (0.57 - 1.11) | 0.19 | 0.79 (0.56 - 1.11) | 0.18 |
| MDCR | 2,048 | 5,074 | 1.4 | 4.1 | 15 | 45 | 1.41 (0.70 - 2.70) | 0.31 | 1.47 (0.75 - 2.88) | 0.27 |
| MDCD | 1,452 | 3,305 | 0.7 | 1.8 | 9 | 28 | 0.95 (0.41 - 2.04) | 0.90 | 0.87 (0.39 - 1.95) | 0.74 |
| Open Claims | 321,867 | 412,766 | 238.5 | 306.5 | 805 | 1,514 | 0.86 (0.78 - 0.95) | <0.01 | 0.93 (0.82 - 1.06) | 0.33 |
| OEHR | 19,746 | 31,577 | 5.5 | 13.1 | 31 | 78 | 1.09 (0.68 - 1.70) | 0.72 | 1.08 (0.68 - 1.71) | 0.74 |
| VA | 11,951 | 24,130 | 6.2 | 18.2 | 36 | 72 | 0.99 (0.60 - 1.63) | 0.97 | 1.03 (0.62 - 1.70) | 0.90 |
| SIDIAP | 5,024 | 19,780 | 6.1 | 36.0 | 23 | 127 | 0.97 (0.57 - 1.59) | 0.91 | 1.06 (0.63 - 1.79) | 0.80 |
| Meta-analysis | 394,758 | 531,140 | 283.3 | 403.8 | 1,030 | 2,065 | 0.87 (0.80 - 0.94) | <0.01 | 0.92 (0.82 - 1.03) | 0.13 |
| Meta-analysis (w/o USOC) | 72,891 | 118,374 | 44.8 | 97.4 | 225 | 551 | 0.90 (0.75 - 1.09) | 0.29 | 0.92 (0.76 - 1.12) | 0.43 |

##### 1.2.26 Vomiting

**Supplementary Table 52:** Exposures, outcomes and risk estimates for Vomiting in SGLT2I (male) (T) vs DPP4I (male) (C) new-user cohorts under four different propensity score (PS) adjustment / time-at-risk (TAR) designs across data sources. We report total patient exposure time in 1,000 patient-year (KYr) units, uncalibrated and calibrated hazard ratios (HRs) and their 95% confidence intervals (CIs). Greyed out entries did not pass study diagnostics and are included only for completeness.

| | Patients | | Exposure (KYr) | | Outcomes | | Uncalibrated | | Calibrated | |
|---|---|---|---|---|---|---|---|---|---|---|
| | T | C | T | C | T | C | HR (95% CI) | $p$ | HR (95% CI) | $p$ |
| **PS-matching / on-treatment TAR** | | | | | | | | | | |
| CCAE | 18,420 | 29,193 | 16.3 | 24.5 | 269 | 451 | 0.89 (0.72 - 1.10) | 0.28 | 0.96 (0.78 - 1.18) | 0.71 |
| DAG | 4,388 | 10,512 | 3.8 | 10.9 | 28 | 94 | 0.98 (0.56 - 1.66) | 0.95 | 0.96 (0.56 - 1.65) | 0.80 |
| ODOD | 11,564 | 19,749 | 8.8 | 15.3 | 178 | 392 | 0.96 (0.74 - 1.23) | 0.72 | 0.99 (0.76 - 1.28) | 0.87 |
| MDCR | 1,614 | 4,910 | 1.4 | 4.6 | 26 | 101 | 1.03 (0.55 - 1.85) | 0.93 | 1.03 (0.56 - 1.90) | 0.88 |
| MDCD | 1,218 | 2,886 | 0.7 | 2.1 | 29 | 94 | 1.14 (0.66 - 1.92) | 0.63 | 1.03 (0.60 - 1.76) | 0.84 |
| Open Claims | 225,990 | 395,523 | 220.1 | 377.7 | 1,941 | 4,573 | 0.83 (0.77 - 0.90) | <0.01 | 0.91 (0.80 - 1.03) | 0.17 |
| OEHR | 16,094 | 30,889 | 5.2 | 15.8 | 85 | 254 | 1.30 (0.93 - 1.81) | 0.12 | 1.31 (0.94 - 1.83) | 0.13 |
| VA | 7,360 | 22,787 | 5.0 | 20.7 | 22 | 136 | 0.61 (0.33 - 1.09) | 0.11 | 0.62 (0.34 - 1.12) | 0.12 |
| SIDIAP | 4,195 | 19,235 | 6.2 | 43.6 | 18 | 238 | 0.53 (0.28 - 0.94) | 0.04 | 0.58 (0.32 - 1.07) | 0.10 |
| Meta-analysis | 289,625 | 532,798 | 266.8 | 513.1 | 2,567 | 6,239 | 0.89 (0.79 - 1.00) | 0.06 | 0.94 (0.81 - 1.10) | 0.48 |
| Meta-analysis (w/o USOC) | 63,635 | 137,275 | 46.8 | 135.4 | 626 | 1,666 | 0.92 (0.77 - 1.11) | 0.39 | 0.95 (0.79 - 1.14) | 0.61 |
| **PS-stratification / on-treatment TAR** | | | | | | | | | | |
| CCAE | 22,530 | 29,213 | 18.5 | 24.5 | 308 | 451 | 0.92 (0.79 - 1.07) | 0.29 | 0.96 (0.82 - 1.12) | 0.59 |
| DAG | 5,453 | 10,539 | 4.5 | 11.0 | 34 | 94 | 1.05 (0.68 - 1.61) | 0.81 | 1.01 (0.65 - 1.56) | 0.81 |
| ODOD | 16,142 | 19,845 | 11.2 | 15.4 | 213 | 397 | 0.87 (0.72 - 1.04) | 0.13 | 0.87 (0.73 - 1.05) | 0.19 |
| MDCR | 2,025 | 4,964 | 1.6 | 4.7 | 29 | 103 | 1.01 (0.62 - 1.59) | 0.97 | 1.04 (0.65 - 1.67) | 0.85 |
| MDCD | 1,271 | 2,896 | 0.8 | 2.1 | 29 | 94 | 0.96 (0.61 - 1.47) | 0.87 | 0.87 (0.56 - 1.35) | 0.55 |
| Open Claims | 307,189 | 395,776 | 268.3 | 377.9 | 2,351 | 4,583 | 0.84 (0.79 - 0.89) | <0.01 | 0.91 (0.79 - 1.04) | 0.18 |
| OEHR | 19,279 | 30,954 | 6.1 | 15.8 | 99 | 256 | 1.19 (0.92 - 1.52) | 0.19 | 1.19 (0.92 - 1.54) | 0.20 |
| VA | 11,772 | 23,743 | 7.1 | 22.1 | 40 | 149 | 0.65 (0.42 - 1.01) | 0.06 | 0.68 (0.44 - 1.06) | 0.10 |
| SIDIAP | 4,990 | 19,644 | 6.9 | 45.1 | 22 | 252 | 0.57 (0.34 - 0.90) | 0.02 | 0.58 (0.36 - 0.95) | 0.04 |
| Meta-analysis | 377,608 | 510,935 | 317.1 | 494.3 | 3,056 | 6,136 | 0.90 (0.81 - 1.00) | 0.05 | 0.95 (0.81 - 1.11) | 0.50 |
| Meta-analysis (w/o USOC) | 70,419 | 115,159 | 48.7 | 116.4 | 705 | 1,553 | 0.93 (0.81 - 1.08) | 0.37 | 0.95 (0.82 - 1.11) | 0.55 |
| **PS-matching / on-treatment TAR without escalation** | | | | | | | | | | |
| CCAE | 18,420 | 29,193 | 14.1 | 19.8 | 219 | 372 | 0.82 (0.65 - 1.03) | 0.10 | 0.87 (0.69 - 1.10) | 0.27 |
| DAG | 4,388 | 10,512 | 3.5 | 9.9 | 26 | 85 | 0.99 (0.56 - 1.70) | 0.98 | 0.96 (0.55 - 1.68) | 0.79 |
| ODOD | 11,564 | 19,749 | 7.8 | 12.6 | 149 | 323 | 0.90 (0.68 - 1.18) | 0.46 | 0.93 (0.67 - 1.28) | 0.70 |
| MDCR | 1,614 | 4,910 | 1.3 | 3.9 | 24 | 90 | 0.99 (0.51 - 1.83) | 0.96 | 1.00 (0.52 - 1.90) | 0.89 |
| MDCD | 1,218 | 2,886 | 0.6 | 1.6 | 27 | 78 | 1.08 (0.60 - 1.90) | 0.79 | 1.03 (0.58 - 1.84) | 0.83 |
| Open Claims | 225,990 | 395,523 | 185.1 | 294.0 | 1,518 | 3,461 | 0.83 (0.76 - 0.90) | <0.01 | 0.89 (0.77 - 1.02) | 0.12 |
| OEHR | 16,094 | 30,889 | 4.6 | 12.8 | 72 | 171 | 1.52 (1.04 - 2.20) | 0.03 | 1.53 (1.05 - 2.22) | 0.03 |
| VA | 7,360 | 22,787 | 4.3 | 16.9 | 15 | 111 | 0.48 (0.22 - 0.95) | 0.05 | 0.48 (0.23 - 0.99) | 0.05 |
| SIDIAP | 4,195 | 19,235 | 5.5 | 34.7 | 16 | 200 | 0.46 (0.22 - 0.87) | 0.03 | 0.54 (0.27 - 1.08) | 0.09 |
| Meta-analysis | 289,625 | 532,798 | 226.2 | 404.6 | 2,039 | 4,813 | 0.87 (0.72 - 1.06) | 0.18 | 0.92 (0.73 - 1.16) | 0.50 |
| Meta-analysis (w/o USOC) | 63,635 | 137,275 | 41.1 | 110.7 | 521 | 1,352 | 0.88 (0.68 - 1.14) | 0.32 | 0.90 (0.70 - 1.17) | 0.46 |
| **PS-stratification / on-treatment TAR without escalation** | | | | | | | | | | |
| CCAE | 22,530 | 29,213 | 16.2 | 19.9 | 255 | 372 | 0.86 (0.73 - 1.02) | 0.09 | 0.90 (0.76 - 1.06) | 0.23 |
| DAG | 5,453 | 10,539 | 4.1 | 9.9 | 31 | 85 | 1.04 (0.65 - 1.61) | 0.88 | 1.00 (0.64 - 1.58) | 0.82 |
| ODOD | 16,142 | 19,845 | 9.9 | 12.7 | 179 | 327 | 0.83 (0.68 - 1.02) | 0.08 | 0.83 (0.67 - 1.01) | 0.08 |
| MDCR | 2,025 | 4,964 | 1.4 | 4.0 | 27 | 92 | 1.06 (0.64 - 1.71) | 0.81 | 1.11 (0.68 - 1.81) | 0.70 |
| MDCD | 1,271 | 2,896 | 0.6 | 1.6 | 27 | 78 | 1.02 (0.64 - 1.60) | 0.92 | 0.94 (0.59 - 1.49) | 0.78 |
| Open Claims | 307,189 | 395,776 | 228.8 | 294.1 | 1,867 | 3,469 | 0.82 (0.77 - 0.87) | <0.01 | 0.89 (0.79 - 0.99) | 0.05 |
| OEHR | 19,279 | 30,954 | 5.4 | 12.9 | 84 | 172 | 1.38 (1.03 - 1.84) | 0.03 | 1.37 (1.02 - 1.83) | 0.04 |
| VA | 11,772 | 23,743 | 6.2 | 18.0 | 31 | 122 | 0.58 (0.35 - 0.95) | 0.03 | 0.61 (0.37 - 1.00) | 0.05 |
| SIDIAP | 4,990 | 19,644 | 6.1 | 35.8 | 20 | 212 | 0.53 (0.31 - 0.86) | 0.02 | 0.58 (0.35 - 0.97) | 0.05 |
| Meta-analysis | 377,608 | 510,935 | 272.0 | 389.2 | 2,463 | 4,729 | 0.90 (0.76 - 1.07) | 0.25 | 0.95 (0.79 - 1.15) | 0.63 |
| Meta-analysis (w/o USOC) | 70,419 | 115,159 | 43.3 | 95.1 | 596 | 1,260 | 0.93 (0.75 - 1.15) | 0.49 | 0.95 (0.77 - 1.17) | 0.64 |

#### 1.3 SGLT2I (male) vs SU (male)

##### 1.3.1 3-point MACE

**Supplementary Table 53:** Exposures, outcomes and risk estimates for 3-point MACE in SGLT2I (male) (T) vs SU (male) (C) new-user cohorts under four different propensity score (PS) adjustment / time-at-risk (TAR) designs across data sources. We report total patient exposure time in 1,000 patient-year (KYr) units, uncalibrated and calibrated hazard ratios (HRs) and their 95% confidence intervals (CIs). Greyed out entries did not pass study diagnostics and are included only for completeness.

| | Patients | | Exposure (KYr) | | Outcomes | | Uncalibrated | | Calibrated | |
|---|---|---|---|---|---|---|---|---|---|---|
| | T | C | T | C | T | C | HR (95% CI) | $p$ | HR (95% CI) | $p$ |
| **PS-matching / on-treatment TAR** | | | | | | | | | | |
| CCAE | 19,864 | 45,123 | 17.0 | 37.5 | 123 | 429 | 0.70 (0.52 - 0.91) | 0.01 | 0.71 (0.53 - 0.93) | 0.02 |
| DAG | 5,471 | 973 | 4.6 | 0.9 | 0 | 0 | | | | |
| ODOD | 14,875 | 45,890 | 10.6 | 41.3 | 155 | 1,052 | 0.77 (0.61 - 0.96) | 0.02 | 0.76 (0.56 - 1.01) | 0.09 |
| MDCR | 1,661 | 7,509 | 1.4 | 7.0 | 33 | 214 | 0.69 (0.38 - 1.19) | 0.21 | 0.69 (0.39 - 1.23) | 0.23 |
| MDCD | 1,331 | 6,621 | 0.8 | 4.6 | 17 | 132 | 0.89 (0.46 - 1.61) | 0.72 | 0.79 (0.42 - 1.48) | 0.48 |
| Open Claims | 285,069 | 830,545 | 254.9 | 893.3 | 1,943 | 12,369 | 0.72 (0.67 - 0.77) | <0.01 | 0.77 (0.63 - 0.93) | <0.01 |
| OEHR | 17,735 | 71,834 | 5.6 | 39.8 | 44 | 557 | 0.81 (0.54 - 1.17) | 0.28 | 0.82 (0.56 - 1.21) | 0.34 |
| VA | 0 | 0 | 0.0 | 0.0 | 0 | 0 | | | | |
| SIDIAP | 4,099 | 1,187 | 6.0 | 2.2 | 27 | 12 | 1.10 (0.43 - 3.02) | 0.86 | 1.03 (0.39 - 2.74) | 0.85 |
| Meta-analysis | 344,634 | 1,008,709 | 296.2 | 1025.6 | 2,342 | 14,765 | 0.73 (0.68 - 0.77) | <0.01 | 0.75 (0.62 - 0.90) | <0.01 |
| Meta-analysis (w/o USOC) | 59,565 | 178,164 | 41.3 | 132.4 | 399 | 2,396 | 0.76 (0.66 - 0.88) | <0.01 | 0.76 (0.64 - 0.89) | <0.01 |
| **PS-stratification / on-treatment TAR** | | | | | | | | | | |
| CCAE | 23,133 | 45,722 | 19.3 | 37.8 | 140 | 434 | 0.71 (0.58 - 0.87) | <0.01 | 0.74 (0.60 - 0.91) | <0.01 |
| DAG | 5,702 | 1,403 | 4.7 | 1.4 | 0 | 0 | | | | |
| ODOD | 16,422 | 46,390 | 11.4 | 41.8 | 170 | 1,084 | 0.78 (0.65 - 0.93) | <0.01 | 0.77 (0.60 - 0.97) | 0.05 |
| MDCR | 1,977 | 7,751 | 1.6 | 7.2 | 36 | 230 | 0.94 (0.62 - 1.40) | 0.78 | 0.91 (0.61 - 1.38) | 0.70 |
| MDCD | 1,365 | 6,824 | 0.8 | 4.8 | 17 | 140 | 0.91 (0.51 - 1.51) | 0.72 | 0.86 (0.50 - 1.47) | 0.59 |
| Open Claims | 308,612 | 835,362 | 270.1 | 898.3 | 2,093 | 12,566 | 0.74 (0.70 - 0.77) | <0.01 | 0.79 (0.64 - 0.96) | 0.02 |
| OEHR | 19,539 | 74,077 | 6.2 | 41.3 | 48 | 604 | 0.70 (0.50 - 0.95) | 0.03 | 0.75 (0.54 - 1.03) | 0.09 |
| VA | 11,375 | 105,972 | 6.9 | 96.9 | 50 | 842 | 0.82 (0.61 - 1.08) | 0.17 | 0.93 (0.59 - 1.47) | 0.78 |
| SIDIAP | 4,643 | 2,634 | 6.4 | 6.1 | 29 | 39 | 0.75 (0.38 - 1.45) | 0.39 | 0.69 (0.35 - 1.34) | 0.28 |
| Meta-analysis | 367,706 | 1,001,551 | 307.0 | 1019.2 | 2,451 | 14,688 | 0.74 (0.70 - 0.77) | <0.01 | 0.77 (0.64 - 0.92) | <0.01 |
| Meta-analysis (w/o USOC) | 59,094 | 166,189 | 36.9 | 120.9 | 358 | 2,122 | 0.74 (0.66 - 0.84) | <0.01 | 0.76 (0.65 - 0.88) | <0.01 |
| **PS-matching / on-treatment TAR without escalation** | | | | | | | | | | |
| CCAE | 19,864 | 45,123 | 14.8 | 32.6 | 100 | 373 | 0.69 (0.51 - 0.92) | 0.01 | 0.71 (0.53 - 0.96) | 0.03 |
| DAG | 5,471 | 973 | 4.2 | 0.8 | 0 | 0 | | | | |
| ODOD | 14,875 | 45,890 | 9.4 | 37.0 | 137 | 942 | 0.76 (0.59 - 0.96) | 0.02 | 0.74 (0.57 - 0.98) | 0.06 |
| MDCR | 1,661 | 7,509 | 1.3 | 6.4 | 30 | 193 | 0.64 (0.34 - 1.14) | 0.15 | 0.64 (0.35 - 1.16) | 0.16 |
| MDCD | 1,331 | 6,621 | 0.7 | 3.8 | 15 | 111 | 0.88 (0.43 - 1.66) | 0.71 | 0.78 (0.40 - 1.52) | 0.47 |
| Open Claims | 285,069 | 830,545 | 215.9 | 758.2 | 1,649 | 10,619 | 0.73 (0.68 - 0.78) | <0.01 | 0.78 (0.65 - 0.94) | 0.01 |
| OEHR | 17,735 | 71,834 | 4.9 | 34.7 | 34 | 405 | 0.77 (0.49 - 1.18) | 0.25 | 0.81 (0.52 - 1.26) | 0.36 |
| VA | 0 | 0 | 0.0 | 0.0 | 0 | 0 | | | | |
| SIDIAP | 4,099 | 1,187 | 5.3 | 1.9 | 25 | 11 | 1.22 (0.45 - 3.64) | 0.71 | 1.20 (0.42 - 3.42) | 0.74 |
| Meta-analysis | 344,634 | 1,008,709 | 252.3 | 874.6 | 1,990 | 12,654 | 0.73 (0.68 - 0.78) | <0.01 | 0.76 (0.64 - 0.90) | <0.01 |
| Meta-analysis (w/o USOC) | 59,565 | 178,164 | 36.3 | 116.4 | 341 | 2,035 | 0.75 (0.64 - 0.87) | <0.01 | 0.75 (0.63 - 0.89) | <0.01 |
| **PS-stratification / on-treatment TAR without escalation** | | | | | | | | | | |
| CCAE | 23,133 | 45,722 | 16.8 | 32.9 | 116 | 378 | 0.67 (0.53 - 0.83) | <0.01 | 0.70 (0.56 - 0.88) | <0.01 |
| DAG | 5,702 | 1,403 | 4.3 | 1.3 | 0 | 0 | | | | |
| ODOD | 16,422 | 46,390 | 10.1 | 37.4 | 150 | 971 | 0.77 (0.63 - 0.93) | <0.01 | 0.75 (0.60 - 0.94) | 0.03 |
| MDCR | 1,977 | 7,751 | 1.4 | 6.6 | 32 | 209 | 0.95 (0.61 - 1.44) | 0.81 | 0.91 (0.59 - 1.41) | 0.69 |
| MDCD | 1,365 | 6,824 | 0.7 | 4.0 | 15 | 116 | 0.89 (0.48 - 1.53) | 0.68 | 0.84 (0.47 - 1.49) | 0.55 |
| Open Claims | 308,612 | 835,362 | 229.5 | 762.6 | 1,786 | 10,794 | 0.74 (0.70 - 0.78) | <0.01 | 0.79 (0.64 - 0.98) | 0.03 |
| OEHR | 19,539 | 74,077 | 5.5 | 35.9 | 38 | 440 | 0.71 (0.49 - 1.01) | 0.07 | 0.76 (0.53 - 1.10) | 0.16 |
| VA | 11,375 | 105,972 | 6.0 | 85.9 | 42 | 727 | 0.82 (0.59 - 1.10) | 0.21 | 0.92 (0.66 - 1.29) | 0.70 |
| SIDIAP | 4,643 | 2,634 | 5.7 | 5.2 | 27 | 33 | 0.81 (0.40 - 1.68) | 0.58 | 0.79 (0.38 - 1.64) | 0.55 |
| Meta-analysis | 367,706 | 1,001,551 | 261.9 | 868.8 | 2,090 | 12,583 | 0.73 (0.70 - 0.77) | <0.01 | 0.77 (0.64 - 0.92) | <0.01 |
| Meta-analysis (w/o USOC) | 59,094 | 166,189 | 32.4 | 106.2 | 304 | 1,789 | 0.72 (0.63 - 0.83) | <0.01 | 0.74 (0.63 - 0.86) | <0.01 |

##### 1.3.2 4-point MACE

**Supplementary Table 54:** Exposures, outcomes and risk estimates for 4-point MACE in SGLT2I (male) (T) vs SU (male) (C) new-user cohorts under four different propensity score (PS) adjustment / time-at-risk (TAR) designs across data sources. We report total patient exposure time in 1,000 patient-year (KYr) units, uncalibrated and calibrated hazard ratios (HRs) and their 95% confidence intervals (CIs). Greyed out entries did not pass study diagnostics and are included only for completeness.

| | Patients | | Exposure (KYr) | | Outcomes | | Uncalibrated | | Calibrated | |
|---|---|---|---|---|---|---|---|---|---|---|
| | T | C | T | C | T | C | HR (95% CI) | $p$ | HR (95% CI) | $p$ |
| **PS-matching / on-treatment TAR** | | | | | | | | | | |
| CCAE | 19,596 | 44,420 | 16.7 | 36.8 | 181 | 597 | 0.71 (0.56 - 0.89) | <0.01 | 0.72 (0.57 - 0.90) | <0.01 |
| DAG | 5,471 | 973 | 4.6 | 0.9 | 0 | 0 | | | | |
| ODOD | 14,341 | 43,929 | 10.2 | 39.1 | 236 | 1,675 | 0.71 (0.59 - 0.85) | <0.01 | 0.70 (0.54 - 0.91) | 0.02 |
| MDCR | 1,571 | 6,993 | 1.4 | 6.5 | 46 | 326 | 0.81 (0.51 - 1.25) | 0.35 | 0.81 (0.52 - 1.28) | 0.40 |
| MDCD | 1,234 | 6,134 | 0.7 | 4.2 | 20 | 194 | 0.72 (0.40 - 1.21) | 0.24 | 0.64 (0.37 - 1.11) | 0.13 |
| Open Claims | 276,531 | 802,166 | 248.4 | 858.7 | 2,753 | 19,306 | 0.67 (0.63 - 0.70) | <0.01 | 0.71 (0.59 - 0.86) | <0.01 |
| OEHR | 17,413 | 70,491 | 5.5 | 38.6 | 84 | 988 | 0.82 (0.62 - 1.08) | 0.17 | 0.84 (0.63 - 1.11) | 0.25 |
| VA | 0 | 0 | 0.0 | 0.0 | 0 | 0 | | | | |
| SIDIAP | 4,065 | 1,186 | 5.9 | 2.2 | 32 | 12 | 1.20 (0.48 - 3.29) | 0.71 | 1.13 (0.43 - 2.97) | 0.79 |
| Meta-analysis | 334,751 | 975,319 | 288.8 | 986.0 | 3,352 | 23,098 | 0.69 (0.64 - 0.75) | <0.01 | 0.71 (0.59 - 0.86) | <0.01 |
| Meta-analysis (w/o USOC) | 58,220 | 173,153 | 40.4 | 127.3 | 599 | 3,792 | 0.74 (0.66 - 0.84) | <0.01 | 0.74 (0.64 - 0.85) | <0.01 |
| **PS-stratification / on-treatment TAR** | | | | | | | | | | |
| CCAE | 22,780 | 44,992 | 19.0 | 37.1 | 211 | 603 | 0.76 (0.63 - 0.90) | <0.01 | 0.78 (0.66 - 0.93) | <0.01 |
| DAG | 5,702 | 1,403 | 4.7 | 1.4 | 0 | 0 | | | | |
| ODOD | 15,713 | 44,282 | 10.9 | 39.4 | 257 | 1,716 | 0.70 (0.60 - 0.80) | <0.01 | 0.68 (0.55 - 0.85) | <0.01 |
| MDCR | 1,862 | 7,168 | 1.5 | 6.6 | 52 | 344 | 0.88 (0.62 - 1.22) | 0.46 | 0.85 (0.61 - 1.20) | 0.41 |
| MDCD | 1,263 | 6,287 | 0.7 | 4.4 | 20 | 204 | 0.72 (0.43 - 1.14) | 0.18 | 0.68 (0.42 - 1.11) | 0.13 |
| Open Claims | 297,226 | 806,034 | 262.0 | 862.9 | 2,981 | 19,609 | 0.67 (0.65 - 0.70) | <0.01 | 0.72 (0.59 - 0.88) | <0.01 |
| OEHR | 19,146 | 72,526 | 6.1 | 39.9 | 93 | 1,074 | 0.75 (0.60 - 0.95) | 0.02 | 0.81 (0.64 - 1.02) | 0.09 |
| VA | 10,518 | 102,634 | 6.4 | 93.0 | 86 | 1,665 | 0.73 (0.58 - 0.90) | <0.01 | 0.83 (0.55 - 1.27) | 0.42 |
| SIDIAP | 4,579 | 2,630 | 6.3 | 6.1 | 33 | 44 | 0.78 (0.41 - 1.47) | 0.44 | 0.72 (0.38 - 1.35) | 0.31 |
| Meta-analysis | 354,865 | 967,834 | 298.0 | 979.3 | 3,542 | 23,002 | 0.69 (0.65 - 0.72) | <0.01 | 0.71 (0.60 - 0.86) | <0.01 |
| Meta-analysis (w/o USOC) | 57,639 | 161,800 | 36.0 | 116.4 | 561 | 3,393 | 0.73 (0.66 - 0.80) | <0.01 | 0.74 (0.65 - 0.84) | <0.01 |
| **PS-matching / on-treatment TAR without escalation** | | | | | | | | | | |
| CCAE | 19,596 | 44,420 | 14.6 | 32.0 | 149 | 517 | 0.70 (0.54 - 0.89) | <0.01 | 0.72 (0.56 - 0.92) | 0.01 |
| DAG | 5,471 | 973 | 4.2 | 0.8 | 0 | 0 | | | | |
| ODOD | 14,341 | 43,929 | 9.1 | 35.0 | 207 | 1,494 | 0.70 (0.57 - 0.84) | <0.01 | 0.68 (0.54 - 0.86) | <0.01 |
| MDCR | 1,571 | 6,993 | 1.2 | 6.0 | 41 | 303 | 0.80 (0.49 - 1.25) | 0.34 | 0.79 (0.49 - 1.27) | 0.36 |
| MDCD | 1,234 | 6,134 | 0.6 | 3.5 | 17 | 161 | 0.69 (0.36 - 1.23) | 0.24 | 0.61 (0.33 - 1.13) | 0.13 |
| Open Claims | 276,531 | 802,166 | 210.6 | 729.7 | 2,307 | 16,514 | 0.66 (0.62 - 0.70) | <0.01 | 0.71 (0.59 - 0.86) | <0.01 |
| OEHR | 17,413 | 70,491 | 4.8 | 33.8 | 67 | 767 | 0.80 (0.58 - 1.09) | 0.17 | 0.84 (0.61 - 1.15) | 0.30 |
| VA | 0 | 0 | 0.0 | 0.0 | 0 | 0 | | | | |
| SIDIAP | 4,065 | 1,186 | 5.2 | 1.9 | 28 | 11 | 1.26 (0.47 - 3.73) | 0.65 | 1.24 (0.44 - 3.49) | 0.68 |
| Meta-analysis | 334,751 | 975,319 | 246.1 | 841.8 | 2,816 | 19,767 | 0.67 (0.64 - 0.71) | <0.01 | 0.70 (0.59 - 0.83) | <0.01 |
| Meta-analysis (w/o USOC) | 58,220 | 173,153 | 35.6 | 112.2 | 509 | 3,253 | 0.73 (0.64 - 0.83) | <0.01 | 0.73 (0.63 - 0.84) | <0.01 |
| **PS-stratification / on-treatment TAR without escalation** | | | | | | | | | | |
| CCAE | 22,780 | 44,992 | 16.6 | 32.3 | 176 | 523 | 0.73 (0.60 - 0.87) | <0.01 | 0.76 (0.63 - 0.92) | <0.01 |
| DAG | 5,702 | 1,403 | 4.3 | 1.3 | 0 | 0 | | | | |
| ODOD | 15,713 | 44,282 | 9.7 | 35.3 | 226 | 1,530 | 0.69 (0.59 - 0.80) | <0.01 | 0.67 (0.55 - 0.81) | <0.01 |
| MDCR | 1,862 | 7,168 | 1.3 | 6.1 | 46 | 321 | 0.85 (0.59 - 1.21) | 0.38 | 0.82 (0.57 - 1.17) | 0.30 |
| MDCD | 1,263 | 6,287 | 0.6 | 3.6 | 17 | 168 | 0.67 (0.39 - 1.11) | 0.14 | 0.64 (0.37 - 1.08) | 0.10 |
| Open Claims | 297,226 | 806,034 | 222.7 | 733.3 | 2,508 | 16,789 | 0.67 (0.64 - 0.70) | <0.01 | 0.72 (0.58 - 0.88) | <0.01 |
| OEHR | 19,146 | 72,526 | 5.4 | 34.9 | 75 | 836 | 0.73 (0.56 - 0.95) | 0.02 | 0.79 (0.61 - 1.02) | 0.09 |
| VA | 10,518 | 102,634 | 5.5 | 82.6 | 71 | 1,428 | 0.72 (0.57 - 0.91) | <0.01 | 0.82 (0.62 - 1.07) | 0.30 |
| SIDIAP | 4,579 | 2,630 | 5.6 | 5.2 | 29 | 38 | 0.84 (0.42 - 1.68) | 0.63 | 0.82 (0.41 - 1.65) | 0.60 |
| Meta-analysis | 354,865 | 967,834 | 254.4 | 835.8 | 2,985 | 19,678 | 0.67 (0.64 - 0.70) | <0.01 | 0.70 (0.59 - 0.84) | <0.01 |
| Meta-analysis (w/o USOC) | 57,639 | 161,800 | 31.7 | 102.5 | 477 | 2,889 | 0.71 (0.63 - 0.79) | <0.01 | 0.72 (0.63 - 0.83) | <0.01 |

##### 1.3.3 Acute myocardial infarction

**Supplementary Table 55:** Exposures, outcomes and risk estimates for Acute myocardial infarction in SGLT2I (male) (T) vs SU (male) (C) new-user cohorts under four different propensity score (PS) adjustment / time-at-risk (TAR) designs across data sources. We report total patient exposure time in 1,000 patient-year (KYr) units, uncalibrated and calibrated hazard ratios (HRs) and their 95% confidence intervals (CIs). Greyed out entries did not pass study diagnostics and are included only for completeness.

| | Patients | | Exposure (KYr) | | Outcomes | | Uncalibrated | | Calibrated | |
|---|---|---|---|---|---|---|---|---|---|---|
| | T | C | T | C | T | C | HR (95% CI) | $p$ | HR (95% CI) | $p$ |
| **PS-matching / on-treatment TAR** | | | | | | | | | | |
| CCAE | 20,117 | 45,808 | 17.1 | 38.1 | 72 | 255 | 0.67 (0.47 - 0.94) | 0.03 | 0.68 (0.47 - 0.97) | 0.03 |
| DAG | 5,471 | 973 | 4.6 | 0.9 | 0 | 0 | | | | |
| ODOD | 15,190 | 47,220 | 10.8 | 42.8 | 90 | 580 | 0.79 (0.58 - 1.06) | 0.13 | 0.78 (0.55 - 1.11) | 0.20 |
| MDCR | 1,705 | 7,915 | 1.5 | 7.4 | 12 | 112 | 0.66 (0.28 - 1.40) | 0.32 | 0.67 (0.30 - 1.49) | 0.34 |
| MDCD | 1,383 | 6,895 | 0.8 | 4.8 | 12 | 67 | 1.08 (0.46 - 2.31) | 0.86 | 0.96 (0.43 - 2.15) | 0.89 |
| Open Claims | 291,920 | 854,971 | 260.5 | 921.5 | 1,091 | 6,456 | 0.72 (0.66 - 0.79) | <0.01 | 0.77 (0.63 - 0.94) | 0.01 |
| OEHR | 17,905 | 72,582 | 5.7 | 40.4 | 21 | 318 | 0.64 (0.36 - 1.08) | 0.11 | 0.65 (0.38 - 1.13) | 0.14 |
| VA | 0 | 0 | 0.0 | 0.0 | 0 | 0 | | | | |
| SIDIAP | 4,168 | 1,203 | 6.1 | 2.2 | 15 | 8 | 1.16 (0.35 - 4.50) | 0.82 | 1.09 (0.30 - 3.92) | 0.86 |
| Meta-analysis | 348,220 | 1,035,391 | 296.4 | 1055.1 | 1,298 | 7,788 | 0.73 (0.67 - 0.79) | <0.01 | 0.75 (0.62 - 0.91) | <0.01 |
| Meta-analysis (w/o USOC) | 56,300 | 180,420 | 36.0 | 133.5 | 207 | 1,332 | 0.74 (0.61 - 0.90) | <0.01 | 0.73 (0.60 - 0.91) | <0.01 |
| **PS-stratification / on-treatment TAR** | | | | | | | | | | |
| CCAE | 23,425 | 46,429 | 19.5 | 38.5 | 82 | 256 | 0.68 (0.51 - 0.88) | <0.01 | 0.70 (0.53 - 0.92) | 0.01 |
| DAG | 5,702 | 1,403 | 4.7 | 1.4 | 0 | 0 | | | | |
| ODOD | 16,779 | 47,825 | 11.7 | 43.4 | 101 | 601 | 0.79 (0.63 - 1.00) | 0.05 | 0.78 (0.59 - 1.03) | 0.11 |
| MDCR | 2,032 | 8,121 | 1.6 | 7.6 | 15 | 121 | 0.81 (0.42 - 1.45) | 0.50 | 0.78 (0.42 - 1.45) | 0.45 |
| MDCD | 1,420 | 7,116 | 0.8 | 5.0 | 12 | 73 | 1.33 (0.65 - 2.51) | 0.40 | 1.26 (0.64 - 2.47) | 0.51 |
| Open Claims | 316,207 | 860,148 | 276.1 | 926.9 | 1,187 | 6,563 | 0.76 (0.71 - 0.82) | <0.01 | 0.82 (0.66 - 1.01) | 0.06 |
| OEHR | 19,723 | 74,941 | 6.3 | 42.0 | 26 | 347 | 0.58 (0.37 - 0.87) | 0.01 | 0.62 (0.40 - 0.96) | 0.03 |
| VA | 11,651 | 107,705 | 7.0 | 98.7 | 35 | 520 | 0.94 (0.66 - 1.31) | 0.75 | 1.08 (0.65 - 1.77) | 0.79 |
| SIDIAP | 4,738 | 2,664 | 6.5 | 6.2 | 16 | 23 | 0.51 (0.21 - 1.24) | 0.14 | 0.47 (0.19 - 1.15) | 0.10 |
| Meta-analysis | 376,134 | 1,029,343 | 313.6 | 1050.8 | 1,396 | 7,767 | 0.76 (0.71 - 0.80) | <0.01 | 0.79 (0.65 - 0.95) | 0.01 |
| Meta-analysis (w/o USOC) | 59,927 | 169,195 | 37.5 | 123.8 | 209 | 1,204 | 0.72 (0.61 - 0.84) | <0.01 | 0.73 (0.61 - 0.88) | <0.01 |
| **PS-matching / on-treatment TAR without escalation** | | | | | | | | | | |
| CCAE | 20,117 | 45,808 | 14.9 | 33.1 | 60 | 216 | 0.69 (0.47 - 1.00) | 0.05 | 0.71 (0.48 - 1.04) | 0.09 |
| DAG | 5,471 | 973 | 4.2 | 0.8 | 0 | 0 | | | | |
| ODOD | 15,190 | 47,220 | 9.6 | 38.3 | 80 | 524 | 0.73 (0.52 - 0.99) | 0.05 | 0.71 (0.50 - 1.01) | 0.08 |
| MDCR | 1,705 | 7,915 | 1.3 | 6.8 | 12 | 98 | 0.71 (0.29 - 1.51) | 0.41 | 0.70 (0.31 - 1.59) | 0.41 |
| MDCD | 1,383 | 6,895 | 0.7 | 4.0 | 12 | 57 | 1.24 (0.52 - 2.71) | 0.61 | 1.10 (0.48 - 2.50) | 0.83 |
| Open Claims | 291,920 | 854,971 | 220.6 | 782.1 | 913 | 5,507 | 0.74 (0.67 - 0.81) | <0.01 | 0.79 (0.65 - 0.97) | 0.03 |
| OEHR | 17,905 | 72,582 | 5.0 | 35.2 | 20 | 219 | 0.84 (0.45 - 1.46) | 0.55 | 0.87 (0.49 - 1.57) | 0.66 |
| VA | 0 | 0 | 0.0 | 0.0 | 0 | 0 | | | | |
| SIDIAP | 4,168 | 1,203 | 5.4 | 1.9 | 14 | 7 | 0.96 (0.28 - 3.80) | 0.95 | 0.94 (0.26 - 3.50) | 0.87 |
| Meta-analysis | 348,220 | 1,035,391 | 252.1 | 899.5 | 1,097 | 6,621 | 0.74 (0.68 - 0.81) | <0.01 | 0.77 (0.64 - 0.92) | <0.01 |
| Meta-analysis (w/o USOC) | 56,300 | 180,420 | 31.5 | 117.4 | 184 | 1,114 | 0.75 (0.61 - 0.93) | <0.01 | 0.76 (0.61 - 0.94) | 0.02 |
| **PS-stratification / on-treatment TAR without escalation** | | | | | | | | | | |
| CCAE | 23,425 | 46,429 | 17.0 | 33.4 | 70 | 217 | 0.67 (0.50 - 0.89) | <0.01 | 0.70 (0.52 - 0.94) | 0.02 |
| DAG | 5,702 | 1,403 | 4.3 | 1.3 | 0 | 0 | | | | |
| ODOD | 16,779 | 47,825 | 10.3 | 38.8 | 90 | 541 | 0.78 (0.61 - 1.00) | 0.05 | 0.76 (0.58 - 1.00) | 0.08 |
| MDCR | 2,032 | 8,121 | 1.5 | 7.0 | 14 | 107 | 0.87 (0.44 - 1.60) | 0.66 | 0.83 (0.44 - 1.58) | 0.59 |
| MDCD | 1,420 | 7,116 | 0.7 | 4.1 | 12 | 61 | 1.52 (0.74 - 2.91) | 0.23 | 1.43 (0.72 - 2.84) | 0.32 |
| Open Claims | 316,207 | 860,148 | 234.6 | 786.8 | 1,000 | 5,603 | 0.76 (0.71 - 0.82) | <0.01 | 0.82 (0.66 - 1.01) | 0.07 |
| OEHR | 19,723 | 74,941 | 5.5 | 36.5 | 25 | 240 | 0.77 (0.48 - 1.19) | 0.25 | 0.82 (0.52 - 1.30) | 0.40 |
| VA | 11,651 | 107,705 | 6.1 | 87.5 | 31 | 443 | 1.00 (0.68 - 1.42) | 0.98 | 1.13 (0.77 - 1.67) | 0.61 |
| SIDIAP | 4,738 | 2,664 | 5.8 | 5.3 | 15 | 18 | 0.56 (0.21 - 1.48) | 0.25 | 0.55 (0.21 - 1.45) | 0.24 |
| Meta-analysis | 376,134 | 1,029,343 | 267.5 | 895.5 | 1,185 | 6,601 | 0.76 (0.71 - 0.81) | <0.01 | 0.79 (0.66 - 0.96) | 0.01 |
| Meta-analysis (w/o USOC) | 59,927 | 169,195 | 32.9 | 108.7 | 185 | 998 | 0.74 (0.62 - 0.88) | <0.01 | 0.75 (0.62 - 0.91) | <0.01 |

##### 1.3.4 Acute renal failure

**Supplementary Table 56:** Exposures, outcomes and risk estimates for Acute renal failure in SGLT2I (male) (T) vs SU (male) (C) new-user cohorts under four different propensity score (PS) adjustment / time-at-risk (TAR) designs across data sources. We report total patient exposure time in 1,000 patient-year (KYr) units, uncalibrated and calibrated hazard ratios (HRs) and their 95% confidence intervals (CIs). Greyed out entries did not pass study diagnostics and are included only for completeness.

| | Patients | | Exposure (KYr) | | Outcomes | | Uncalibrated | | Calibrated | |
|---|---|---|---|---|---|---|---|---|---|---|
| | T | C | T | C | T | C | HR (95% CI) | $p$ | HR (95% CI) | $p$ |
| **PS-matching / on-treatment TAR** | | | | | | | | | | |
| CCAE | 20,248 | 45,772 | 17.3 | 38.2 | 112 | 452 | 0.62 (0.46 - 0.82) | <0.01 | 0.63 (0.47 - 0.84) | <0.01 |
| DAG | 5,471 | 973 | 4.6 | 0.9 | 0 | 0 | | | | |
| ODOD | 15,107 | 45,548 | 10.8 | 40.7 | 180 | 1,562 | 0.65 (0.53 - 0.79) | <0.01 | 0.64 (0.49 - 0.84) | <0.01 |
| MDCR | 1,684 | 7,647 | 1.4 | 7.1 | 30 | 293 | 0.67 (0.40 - 1.08) | 0.12 | 0.68 (0.41 - 1.11) | 0.14 |
| MDCD | 1,366 | 6,610 | 0.8 | 4.6 | 16 | 193 | 0.64 (0.33 - 1.16) | 0.17 | 0.57 (0.31 - 1.07) | 0.09 |
| Open Claims | 294,583 | 849,930 | 263.5 | 912.8 | 2,103 | 16,419 | 0.62 (0.58 - 0.66) | <0.01 | 0.66 (0.54 - 0.80) | <0.01 |
| OEHR | 17,982 | 72,411 | 5.7 | 40.0 | 60 | 853 | 0.84 (0.60 - 1.14) | 0.27 | 0.85 (0.62 - 1.17) | 0.36 |
| VA | 0 | 0 | 0.0 | 0.0 | 0 | 0 | | | | |
| SIDIAP | 4,546 | 1,216 | 6.6 | 2.2 | <5 | <5 | 0.00 (0.00 - 5.83) | 0.21 | 0.00 (0.00 - 37681) | 0.20 |
| Meta-analysis | 350,970 | 1,027,918 | 299.6 | 1043.4 | 2,501 | 19,772 | 0.63 (0.60 - 0.66) | <0.01 | 0.65 (0.54 - 0.78) | <0.01 |
| Meta-analysis (w/o USOC) | 56,387 | 177,988 | 36.1 | 130.6 | 398 | 3,353 | 0.68 (0.59 - 0.77) | <0.01 | 0.67 (0.58 - 0.78) | <0.01 |
| **PS-stratification / on-treatment TAR** | | | | | | | | | | |
| CCAE | 23,699 | 46,349 | 19.7 | 38.5 | 127 | 459 | 0.63 (0.51 - 0.78) | <0.01 | 0.65 (0.52 - 0.81) | <0.01 |
| DAG | 5,702 | 1,403 | 4.7 | 1.4 | 0 | 0 | | | | |
| ODOD | 16,747 | 46,175 | 11.7 | 41.3 | 199 | 1,629 | 0.60 (0.51 - 0.71) | <0.01 | 0.59 (0.47 - 0.74) | <0.01 |
| MDCR | 2,014 | 7,786 | 1.6 | 7.2 | 36 | 301 | 0.70 (0.46 - 1.03) | 0.08 | 0.68 (0.45 - 1.01) | 0.07 |
| MDCD | 1,409 | 6,762 | 0.9 | 4.7 | 17 | 204 | 0.61 (0.35 - 1.01) | 0.07 | 0.58 (0.34 - 0.98) | 0.05 |
| Open Claims | 320,733 | 854,446 | 280.5 | 917.6 | 2,329 | 16,716 | 0.60 (0.57 - 0.63) | <0.01 | 0.64 (0.52 - 0.79) | <0.01 |
| OEHR | 19,913 | 74,597 | 6.4 | 41.4 | 66 | 934 | 0.65 (0.50 - 0.85) | <0.01 | 0.70 (0.54 - 0.92) | 0.01 |
| VA | 11,731 | 106,207 | 7.1 | 96.6 | 93 | 1,559 | 0.79 (0.64 - 0.97) | 0.03 | 0.90 (0.59 - 1.36) | 0.64 |
| SIDIAP | 5,159 | 2,688 | 7.1 | 6.3 | <5 | <5 | 1.28 (0.14 - 28.2) | 0.85 | 1.18 (0.08 - 16.5) | 0.90 |
| Meta-analysis | 381,092 | 1,021,567 | 318.2 | 1038.9 | 2,721 | 19,738 | 0.60 (0.58 - 0.63) | <0.01 | 0.63 (0.52 - 0.75) | <0.01 |
| Meta-analysis (w/o USOC) | 60,359 | 167,121 | 37.8 | 121.3 | 392 | 3,022 | 0.62 (0.55 - 0.70) | <0.01 | 0.63 (0.55 - 0.73) | <0.01 |
| **PS-matching / on-treatment TAR without escalation** | | | | | | | | | | |
| CCAE | 20,248 | 45,772 | 15.1 | 33.1 | 87 | 380 | 0.64 (0.47 - 0.87) | <0.01 | 0.66 (0.48 - 0.90) | 0.01 |
| DAG | 5,471 | 973 | 4.2 | 0.8 | 0 | 0 | | | | |
| ODOD | 15,107 | 45,548 | 9.6 | 36.5 | 151 | 1,403 | 0.65 (0.52 - 0.80) | <0.01 | 0.64 (0.50 - 0.81) | <0.01 |
| MDCR | 1,684 | 7,647 | 1.3 | 6.5 | 27 | 266 | 0.78 (0.46 - 1.27) | 0.34 | 0.77 (0.46 - 1.29) | 0.35 |
| MDCD | 1,366 | 6,610 | 0.7 | 3.8 | 15 | 157 | 0.67 (0.33 - 1.27) | 0.25 | 0.59 (0.30 - 1.17) | 0.14 |
| Open Claims | 294,583 | 849,930 | 223.5 | 775.7 | 1,709 | 13,780 | 0.60 (0.56 - 0.64) | <0.01 | 0.65 (0.54 - 0.78) | <0.01 |
| OEHR | 17,982 | 72,411 | 5.0 | 35.0 | 40 | 630 | 0.70 (0.47 - 1.01) | 0.07 | 0.73 (0.50 - 1.07) | 0.12 |
| VA | 0 | 0 | 0.0 | 0.0 | 0 | 0 | | | | |
| SIDIAP | 4,546 | 1,216 | 5.9 | 2.0 | <5 | 0 | | | | |
| Meta-analysis | 350,970 | 1,027,918 | 255.2 | 890.7 | 2,029 | 16,616 | 0.61 (0.58 - 0.65) | <0.01 | 0.64 (0.54 - 0.75) | <0.01 |
| Meta-analysis (w/o USOC) | 56,387 | 177,988 | 31.7 | 115.0 | 320 | 2,836 | 0.66 (0.57 - 0.77) | <0.01 | 0.67 (0.57 - 0.78) | <0.01 |
| **PS-stratification / on-treatment TAR without escalation** | | | | | | | | | | |
| CCAE | 23,699 | 46,349 | 17.3 | 33.4 | 102 | 386 | 0.59 (0.46 - 0.74) | <0.01 | 0.62 (0.49 - 0.79) | <0.01 |
| DAG | 5,702 | 1,403 | 4.3 | 1.3 | 0 | 0 | | | | |
| ODOD | 16,747 | 46,175 | 10.4 | 37.1 | 167 | 1,464 | 0.57 (0.48 - 0.68) | <0.01 | 0.56 (0.45 - 0.69) | <0.01 |
| MDCR | 2,014 | 7,786 | 1.5 | 6.6 | 32 | 272 | 0.76 (0.49 - 1.13) | 0.19 | 0.72 (0.48 - 1.10) | 0.15 |
| MDCD | 1,409 | 6,762 | 0.7 | 3.9 | 16 | 165 | 0.68 (0.38 - 1.14) | 0.17 | 0.64 (0.37 - 1.11) | 0.12 |
| Open Claims | 320,733 | 854,446 | 238.7 | 779.9 | 1,920 | 14,041 | 0.59 (0.56 - 0.62) | <0.01 | 0.63 (0.51 - 0.78) | <0.01 |
| OEHR | 19,913 | 74,597 | 5.6 | 36.2 | 46 | 694 | 0.57 (0.41 - 0.78) | <0.01 | 0.61 (0.44 - 0.85) | <0.01 |
| VA | 11,731 | 106,207 | 6.2 | 85.7 | 80 | 1,275 | 0.86 (0.68 - 1.07) | 0.18 | 0.97 (0.74 - 1.26) | 0.83 |
| SIDIAP | 5,159 | 2,688 | 6.3 | 5.4 | <5 | 0 | | | | |
| Meta-analysis | 381,092 | 1,021,567 | 271.9 | 886.6 | 2,235 | 16,585 | 0.59 (0.56 - 0.61) | <0.01 | 0.61 (0.51 - 0.73) | <0.01 |
| Meta-analysis (w/o USOC) | 60,359 | 167,121 | 33.2 | 106.7 | 315 | 2,544 | 0.58 (0.51 - 0.66) | <0.01 | 0.59 (0.51 - 0.69) | <0.01 |

##### 1.3.5 Glycemic control

**Supplementary Table 57:** Exposures, outcomes and risk estimates for Glycemic control in SGLT2I (male) (T) vs SU (male) (C) new-user cohorts under four different propensity score (PS) adjustment / time-at-risk (TAR) designs across data sources. We report total patient exposure time in 1,000 patient-year (KYr) units, uncalibrated and calibrated hazard ratios (HRs) and their 95% confidence intervals (CIs). Greyed out entries did not pass study diagnostics and are included only for completeness.

| | Patients | | Exposure (KYr) | | Outcomes | | Uncalibrated | | Calibrated | |
|---|---|---|---|---|---|---|---|---|---|---|
| | T | C | T | C | T | C | HR (95% CI) | $p$ | HR (95% CI) | $p$ |
| **PS-matching / on-treatment TAR** | | | | | | | | | | |
| CCAE | 20,489 | 46,544 | 17.5 | 38.9 | 22 | 60 | 1.15 (0.57 - 2.24) | 0.69 | 1.17 (0.59 - 2.32) | 0.66 |
| DAG | 1,909 | 406 | 1.4 | 0.3 | 199 | 54 | 0.90 (0.61 - 1.35) | 0.61 | 1.69 (1.13 - 2.52) | 0.03 |
| ODOD | 10,546 | 33,733 | 6.9 | 26.6 | 716 | 2,844 | 0.98 (0.88 - 1.10) | 0.78 | 0.97 (0.78 - 1.21) | 0.79 |
| MDCR | 1,772 | 8,248 | 1.5 | 7.8 | 0 | 11 | | | | |
| MDCD | 1,464 | 7,245 | 0.9 | 5.1 | 0 | 0 | | | | |
| Open Claims | 304,948 | 889,205 | 271.7 | 962.4 | 0 | 0 | | | | |
| OEHR | 18,285 | 73,829 | 5.8 | 41.4 | 0 | 0 | | | | |
| VA | 0 | 0 | 0.0 | 0.0 | 0 | 0 | | | | |
| SIDIAP | 487 | 169 | 0.5 | 0.2 | 161 | 46 | 1.36 (0.89 - 2.13) | 0.17 | 1.28 (0.82 - 1.98) | 0.31 |
| Meta-analysis | 33,431 | 80,852 | 26.2 | 66.1 | 1,098 | 3,004 | 1.00 (0.90 - 1.11) | 1.00 | 1.03 (0.84 - 1.26) | 0.78 |
| Meta-analysis (w/o USOC) | 33,431 | 80,852 | 26.2 | 66.1 | 1,098 | 3,004 | 1.00 (0.90 - 1.11) | 1.00 | 1.00 (0.88 - 1.13) | 0.86 |
| **PS-stratification / on-treatment TAR** | | | | | | | | | | |
| CCAE | 23,976 | 47,040 | 20.0 | 39.3 | 26 | 73 | 1.00 (0.60 - 1.64) | 0.99 | 1.04 (0.63 - 1.72) | 0.88 |
| DAG | 1,981 | 611 | 1.4 | 0.5 | 210 | 78 | 0.83 (0.60 - 1.16) | 0.27 | 1.09 (0.78 - 1.52) | 0.53 |
| ODOD | 11,471 | 34,473 | 7.3 | 27.2 | 761 | 2,931 | 1.01 (0.93 - 1.11) | 0.75 | 0.99 (0.83 - 1.19) | 0.86 |
| MDCR | 2,128 | 8,484 | 1.7 | 8.0 | 0 | 12 | | | | |
| MDCD | 1,507 | 7,493 | 0.9 | 5.3 | 0 | 0 | | | | |
| Open Claims | 332,801 | 894,802 | 289.7 | 968.3 | 0 | 0 | | | | |
| OEHR | 20,231 | 76,288 | 6.5 | 43.1 | 0 | 0 | | | | |
| VA | 11,946 | 105,730 | 7.2 | 96.4 | 12 | 146 | 0.98 (0.51 - 1.69) | 0.94 | 1.11 (0.56 - 2.23) | 0.78 |
| SIDIAP | 541 | 437 | 0.5 | 0.6 | 180 | 150 | 1.39 (1.02 - 1.90) | 0.04 | 1.28 (0.93 - 1.75) | 0.14 |
| Meta-analysis | 35,447 | 81,513 | 27.2 | 66.5 | 787 | 3,004 | 1.01 (0.93 - 1.11) | 0.75 | 1.05 (0.87 - 1.28) | 0.60 |
| Meta-analysis (w/o USOC) | 35,447 | 81,513 | 27.2 | 66.5 | 787 | 3,004 | 1.01 (0.93 - 1.11) | 0.75 | 1.03 (0.92 - 1.17) | 0.64 |
| **PS-matching / on-treatment TAR without escalation** | | | | | | | | | | |
| CCAE | 20,489 | 46,544 | 15.2 | 33.7 | 21 | 56 | 1.15 (0.55 - 2.28) | 0.70 | 1.19 (0.58 - 2.41) | 0.64 |
| DAG | 1,909 | 406 | 1.3 | 0.3 | 174 | 48 | 0.95 (0.63 - 1.45) | 0.80 | 1.86 (1.23 - 2.82) | <0.01 |
| ODOD | 10,546 | 33,733 | 6.1 | 23.9 | 628 | 2,598 | 0.95 (0.84 - 1.08) | 0.45 | 0.94 (0.78 - 1.12) | 0.57 |
| MDCR | 1,772 | 8,248 | 1.4 | 7.2 | 0 | 10 | | | | |
| MDCD | 1,464 | 7,245 | 0.8 | 4.2 | 0 | 0 | | | | |
| Open Claims | 304,948 | 889,205 | 230.2 | 816.1 | 0 | 0 | | | | |
| OEHR | 18,285 | 73,829 | 5.1 | 35.9 | 0 | 0 | | | | |
| VA | 0 | 0 | 0.0 | 0.0 | 0 | 0 | | | | |
| SIDIAP | 487 | 169 | 0.4 | 0.2 | 150 | 42 | 1.48 (0.95 - 2.39) | 0.09 | 1.46 (0.92 - 2.32) | 0.14 |
| Meta-analysis | 33,431 | 80,852 | 23.0 | 58.0 | 973 | 2,744 | 1.02 (0.85 - 1.23) | 0.81 | 1.06 (0.83 - 1.35) | 0.64 |
| Meta-analysis (w/o USOC) | 33,431 | 80,852 | 23.0 | 58.0 | 973 | 2,744 | 1.01 (0.86 - 1.19) | 0.89 | 1.01 (0.85 - 1.21) | 0.85 |
| **PS-stratification / on-treatment TAR without escalation** | | | | | | | | | | |
| CCAE | 23,976 | 47,040 | 17.4 | 34.0 | 25 | 68 | 1.04 (0.61 - 1.72) | 0.89 | 1.09 (0.65 - 1.83) | 0.75 |
| DAG | 1,981 | 611 | 1.3 | 0.5 | 185 | 71 | 0.83 (0.58 - 1.18) | 0.29 | 1.15 (0.81 - 1.65) | 0.46 |
| ODOD | 11,471 | 34,473 | 6.5 | 24.4 | 669 | 2,677 | 0.99 (0.90 - 1.09) | 0.88 | 0.97 (0.83 - 1.13) | 0.73 |
| MDCR | 2,128 | 8,484 | 1.5 | 7.3 | 0 | 11 | | | | |
| MDCD | 1,507 | 7,493 | 0.8 | 4.4 | 0 | 0 | | | | |
| Open Claims | 332,801 | 894,802 | 246.2 | 821.2 | 0 | 0 | | | | |
| OEHR | 20,231 | 76,288 | 5.7 | 37.3 | 0 | 0 | | | | |
| VA | 11,946 | 105,730 | 6.2 | 85.3 | <10 | 133 | 0.83 (0.39 - 1.54) | 0.60 | 0.94 (0.47 - 1.88) | 0.86 |
| SIDIAP | 541 | 437 | 0.4 | 0.5 | 165 | 133 | 1.44 (1.04 - 2.00) | 0.03 | 1.41 (1.01 - 1.96) | 0.08 |
| Meta-analysis | 35,447 | 81,513 | 23.9 | 58.4 | 694 | 2,745 | 0.99 (0.91 - 1.09) | 0.91 | 1.04 (0.86 - 1.27) | 0.68 |
| Meta-analysis (w/o USOC) | 35,447 | 81,513 | 23.9 | 58.4 | 694 | 2,745 | 0.99 (0.91 - 1.09) | 0.91 | 1.01 (0.90 - 1.15) | 0.83 |

##### 1.3.6 Hospitalization with heart failure

**Supplementary Table 58:** Exposures, outcomes and risk estimates for Hospitalization with heart failure in SGLT2I (male) (T) vs SU (male) (C) new-user cohorts under four different propensity score (PS) adjustment / time-at-risk (TAR) designs across data sources. We report total patient exposure time in 1,000 patient-year (KYr) units, uncalibrated and calibrated hazard ratios (HRs) and their 95% confidence intervals (CIs). Greyed out entries did not pass study diagnostics and are included only for completeness.

| | Patients | | Exposure (KYr) | | Outcomes | | Uncalibrated | | Calibrated | |
|---|---|---|---|---|---|---|---|---|---|---|
| | T | C | T | C | T | C | HR (95% CI) | $p$ | HR (95% CI) | $p$ |
| **PS-matching / on-treatment TAR** | | | | | | | | | | |
| CCAE | 20,240 | 45,906 | 17.3 | 38.3 | 93 | 286 | 0.75 (0.53 - 1.04) | 0.09 | 0.76 (0.54 - 1.07) | 0.12 |
| DAG | 5,471 | 973 | 4.6 | 0.9 | 0 | 0 | | | | |
| ODOD | 14,941 | 45,955 | 10.7 | 41.2 | 145 | 1,161 | 0.62 (0.49 - 0.77) | <0.01 | 0.61 (0.45 - 0.81) | <0.01 |
| MDCR | 1,653 | 7,560 | 1.4 | 7.1 | 31 | 227 | 0.78 (0.43 - 1.34) | 0.38 | 0.78 (0.44 - 1.38) | 0.42 |
| MDCD | 1,314 | 6,533 | 0.8 | 4.5 | 11 | 122 | 0.58 (0.27 - 1.12) | 0.13 | 0.51 (0.25 - 1.04) | 0.07 |
| Open Claims | 292,046 | 847,603 | 262.1 | 912.3 | 1,442 | 12,311 | 0.58 (0.54 - 0.62) | <0.01 | 0.62 (0.51 - 0.75) | <0.01 |
| OEHR | 17,851 | 72,117 | 5.7 | 39.8 | 55 | 662 | 0.75 (0.53 - 1.04) | 0.10 | 0.76 (0.54 - 1.07) | 0.14 |
| VA | 0 | 0 | 0.0 | 0.0 | 0 | 0 | | | | |
| SIDIAP | 4,513 | 1,218 | 6.6 | 2.2 | 5 | 0 | | | | |
| Meta-analysis | 348,045 | 1,025,674 | 298.0 | 1043.2 | 1,777 | 14,769 | 0.62 (0.56 - 0.70) | <0.01 | 0.64 (0.52 - 0.80) | <0.01 |
| Meta-analysis (w/o USOC) | 55,999 | 178,071 | 35.9 | 131.0 | 335 | 2,458 | 0.68 (0.58 - 0.79) | <0.01 | 0.67 (0.57 - 0.80) | <0.01 |
| **PS-stratification / on-treatment TAR** | | | | | | | | | | |
| CCAE | 23,598 | 46,509 | 19.7 | 38.6 | 113 | 292 | 0.76 (0.60 - 0.97) | 0.03 | 0.79 (0.62 - 1.01) | 0.06 |
| DAG | 5,702 | 1,403 | 4.7 | 1.4 | 0 | 0 | | | | |
| ODOD | 16,421 | 46,384 | 11.5 | 41.6 | 163 | 1,194 | 0.63 (0.53 - 0.76) | <0.01 | 0.62 (0.49 - 0.79) | <0.01 |
| MDCR | 1,962 | 7,706 | 1.6 | 7.2 | 33 | 237 | 0.87 (0.56 - 1.31) | 0.53 | 0.84 (0.55 - 1.29) | 0.47 |
| MDCD | 1,350 | 6,710 | 0.8 | 4.7 | 11 | 129 | 0.60 (0.30 - 1.09) | 0.12 | 0.56 (0.29 - 1.08) | 0.09 |
| Open Claims | 315,211 | 852,161 | 277.3 | 917.1 | 1,631 | 12,575 | 0.59 (0.55 - 0.62) | <0.01 | 0.63 (0.51 - 0.77) | <0.01 |
| OEHR | 19,700 | 74,317 | 6.3 | 41.3 | 64 | 730 | 0.78 (0.59 - 1.02) | 0.08 | 0.84 (0.63 - 1.11) | 0.24 |
| VA | 11,226 | 106,553 | 6.8 | 97.0 | 73 | 1,224 | 0.83 (0.65 - 1.05) | 0.13 | 0.95 (0.62 - 1.46) | 0.82 |
| SIDIAP | 5,095 | 2,687 | 7.0 | 6.3 | 5 | 5 | 1.08 (0.12 - 7.64) | 0.94 | 0.99 (0.13 - 7.82) | 0.95 |
| Meta-analysis | 374,930 | 1,019,371 | 314.8 | 1038.6 | 1,971 | 14,791 | 0.66 (0.57 - 0.76) | <0.01 | 0.68 (0.55 - 0.86) | <0.01 |
| Meta-analysis (w/o USOC) | 59,719 | 167,210 | 37.5 | 121.5 | 340 | 2,216 | 0.70 (0.61 - 0.81) | <0.01 | 0.71 (0.61 - 0.84) | <0.01 |
| **PS-matching / on-treatment TAR without escalation** | | | | | | | | | | |
| CCAE | 20,240 | 45,906 | 15.1 | 33.2 | 75 | 242 | 0.76 (0.52 - 1.10) | 0.15 | 0.79 (0.54 - 1.14) | 0.22 |
| DAG | 5,471 | 973 | 4.2 | 0.8 | 0 | 0 | | | | |
| ODOD | 14,941 | 45,955 | 9.5 | 36.9 | 127 | 1,029 | 0.60 (0.46 - 0.76) | <0.01 | 0.59 (0.44 - 0.77) | <0.01 |
| MDCR | 1,653 | 7,560 | 1.3 | 6.5 | 26 | 212 | 0.78 (0.42 - 1.36) | 0.41 | 0.77 (0.43 - 1.39) | 0.42 |
| MDCD | 1,314 | 6,533 | 0.7 | 3.8 | 10 | 101 | 0.61 (0.26 - 1.26) | 0.22 | 0.54 (0.24 - 1.19) | 0.13 |
| Open Claims | 292,046 | 847,603 | 222.3 | 774.6 | 1,197 | 10,491 | 0.58 (0.54 - 0.63) | <0.01 | 0.62 (0.51 - 0.76) | <0.01 |
| OEHR | 17,851 | 72,117 | 5.0 | 34.8 | 42 | 511 | 0.73 (0.49 - 1.05) | 0.10 | 0.76 (0.52 - 1.11) | 0.17 |
| VA | 0 | 0 | 0.0 | 0.0 | 0 | 0 | | | | |
| SIDIAP | 4,513 | 1,218 | 5.8 | 1.9 | <5 | 0 | | | | |
| Meta-analysis | 348,045 | 1,025,674 | 253.7 | 889.9 | 1,477 | 12,586 | 0.61 (0.55 - 0.68) | <0.01 | 0.63 (0.52 - 0.77) | <0.01 |
| Meta-analysis (w/o USOC) | 55,999 | 178,071 | 31.5 | 115.3 | 280 | 2,095 | 0.67 (0.56 - 0.79) | <0.01 | 0.67 (0.56 - 0.80) | <0.01 |
| **PS-stratification / on-treatment TAR without escalation** | | | | | | | | | | |
| CCAE | 23,598 | 46,509 | 17.2 | 33.5 | 93 | 248 | 0.77 (0.59 - 0.99) | 0.05 | 0.80 (0.62 - 1.05) | 0.11 |
| DAG | 5,702 | 1,403 | 4.3 | 1.3 | 0 | 0 | | | | |
| ODOD | 16,421 | 46,384 | 10.2 | 37.3 | 144 | 1,056 | 0.63 (0.52 - 0.76) | <0.01 | 0.61 (0.48 - 0.77) | <0.01 |
| MDCR | 1,962 | 7,706 | 1.4 | 6.6 | 28 | 221 | 0.84 (0.52 - 1.31) | 0.46 | 0.81 (0.51 - 1.28) | 0.38 |
| MDCD | 1,350 | 6,710 | 0.7 | 3.9 | 10 | 106 | 0.61 (0.29 - 1.15) | 0.16 | 0.57 (0.29 - 1.15) | 0.12 |
| Open Claims | 315,211 | 852,161 | 235.8 | 778.8 | 1,369 | 10,730 | 0.58 (0.54 - 0.61) | <0.01 | 0.62 (0.50 - 0.77) | <0.01 |
| OEHR | 19,700 | 74,317 | 5.5 | 36.0 | 50 | 561 | 0.74 (0.53 - 1.00) | 0.06 | 0.79 (0.58 - 1.09) | 0.16 |
| VA | 11,226 | 106,553 | 5.9 | 86.0 | 66 | 1,051 | 0.90 (0.69 - 1.14) | 0.41 | 1.01 (0.76 - 1.35) | 0.89 |
| SIDIAP | 5,095 | 2,687 | 6.2 | 5.4 | <5 | 5 | 1.05 (0.11 - 7.66) | 0.96 | 1.03 (0.12 - 8.59) | 0.95 |
| Meta-analysis | 374,930 | 1,019,371 | 268.7 | 885.7 | 1,656 | 12,595 | 0.64 (0.56 - 0.74) | <0.01 | 0.67 (0.54 - 0.84) | <0.01 |
| Meta-analysis (w/o USOC) | 59,719 | 167,210 | 32.9 | 106.8 | 287 | 1,865 | 0.68 (0.60 - 0.79) | <0.01 | 0.70 (0.59 - 0.82) | <0.01 |

##### 1.3.7 Measured renal dysfunction

**Supplementary Table 59:** Exposures, outcomes and risk estimates for Measured renal dysfunction in SGLT2I (male) (T) vs SU (male) (C) new-user cohorts under four different propensity score (PS) adjustment / time-at-risk (TAR) designs across data sources. We report total patient exposure time in 1,000 patient-year (KYr) units, uncalibrated and calibrated hazard ratios (HRs) and their 95% confidence intervals (CIs). Greyed out entries did not pass study diagnostics and are included only for completeness.

| | Patients | | Exposure (KYr) | | Outcomes | | Uncalibrated | | Calibrated | |
|---|---|---|---|---|---|---|---|---|---|---|
| | T | C | T | C | T | C | HR (95% CI) | $p$ | HR (95% CI) | $p$ |
| **PS-matching / on-treatment TAR** | | | | | | | | | | |
| CCAE | 20,587 | 46,816 | 17.6 | 39.1 | <5 | 23 | 0.42 (0.09 - 1.43) | 0.21 | 0.42 (0.11 - 1.67) | 0.22 |
| DAG | 5,471 | 973 | 4.6 | 0.9 | 0 | 0 | | | | |
| ODOD | 15,718 | 48,808 | 11.2 | 44.4 | 7 | 116 | 0.87 (0.32 - 1.99) | 0.76 | 0.85 (0.33 - 2.17) | 0.74 |
| MDCR | 1,777 | 8,314 | 1.5 | 7.9 | 0 | <5 | | | | |
| MDCD | 1,464 | 7,245 | 0.9 | 5.1 | 0 | 0 | | | | |
| Open Claims | 304,948 | 889,205 | 271.7 | 962.4 | 0 | 0 | | | | |
| OEHR | 18,285 | 73,829 | 5.8 | 41.4 | 0 | 0 | | | | |
| VA | 0 | 0 | 0.0 | 0.0 | 0 | 0 | | | | |
| SIDIAP | 4,535 | 1,216 | 6.6 | 2.2 | 13 | <5 | 1.40 (0.29 - 10.1) | 0.71 | 1.31 (0.22 - 7.69) | 0.76 |
| Meta-analysis | 36,305 | 95,624 | 28.8 | 83.5 | <12 | 139 | 0.69 (0.32 - 1.48) | 0.34 | 0.71 (0.32 - 1.56) | 0.39 |
| Meta-analysis (w/o USOC) | 36,305 | 95,624 | 28.8 | 83.5 | <12 | 139 | 0.69 (0.32 - 1.48) | 0.34 | 0.69 (0.32 - 1.48) | 0.34 |
| **PS-stratification / on-treatment TAR** | | | | | | | | | | |
| CCAE | 24,082 | 47,464 | 20.0 | 39.5 | 5 | 23 | 0.35 (0.11 - 0.92) | 0.05 | 0.36 (0.12 - 1.04) | 0.06 |
| DAG | 5,702 | 1,403 | 4.7 | 1.4 | 0 | 0 | | | | |
| ODOD | 17,471 | 49,460 | 12.1 | 45.0 | 7 | 117 | 0.43 (0.18 - 0.89) | 0.04 | 0.42 (0.18 - 0.96) | 0.04 |
| MDCR | 2,136 | 8,601 | 1.7 | 8.1 | 0 | <5 | | | | |
| MDCD | 1,507 | 7,493 | 0.9 | 5.3 | 0 | 0 | | | | |
| Open Claims | 332,801 | 894,802 | 289.7 | 968.3 | 0 | 0 | | | | |
| OEHR | 20,231 | 76,288 | 6.5 | 43.1 | 0 | 0 | | | | |
| VA | 12,350 | 109,470 | 7.4 | 100.4 | 23 | 492 | 0.63 (0.40 - 0.94) | 0.03 | 0.72 (0.41 - 1.25) | 0.27 |
| SIDIAP | 5,147 | 2,685 | 7.0 | 6.3 | 16 | 7 | 1.85 (0.53 - 6.99) | 0.35 | 1.70 (0.47 - 6.19) | 0.42 |
| Meta-analysis | 41,553 | 96,924 | 32.2 | 84.5 | 12 | 140 | 0.40 (0.21 - 0.76) | <0.01 | 0.41 (0.21 - 0.80) | <0.01 |
| Meta-analysis (w/o USOC) | 41,553 | 96,924 | 32.2 | 84.5 | 12 | 140 | 0.40 (0.21 - 0.76) | <0.01 | 0.40 (0.21 - 0.77) | <0.01 |
| **PS-matching / on-treatment TAR without escalation** | | | | | | | | | | |
| CCAE | 20,587 | 46,816 | 15.3 | 33.9 | <5 | 21 | 0.43 (0.06 - 1.95) | 0.34 | 0.45 (0.08 - 2.50) | 0.36 |
| DAG | 5,471 | 973 | 4.2 | 0.8 | 0 | 0 | | | | |
| ODOD | 15,718 | 48,808 | 9.9 | 39.7 | 7 | 102 | 1.03 (0.37 - 2.40) | 0.95 | 1.01 (0.40 - 2.59) | 0.96 |
| MDCR | 1,777 | 8,314 | 1.4 | 7.2 | 0 | <5 | | | | |
| MDCD | 1,464 | 7,245 | 0.8 | 4.2 | 0 | 0 | | | | |
| Open Claims | 304,948 | 889,205 | 230.2 | 816.1 | 0 | 0 | | | | |
| OEHR | 18,285 | 73,829 | 5.1 | 35.9 | 0 | 0 | | | | |
| VA | 0 | 0 | 0.0 | 0.0 | 0 | 0 | | | | |
| SIDIAP | 4,535 | 1,216 | 5.9 | 1.9 | 11 | <5 | 1.24 (0.24 - 9.19) | 0.82 | 1.22 (0.20 - 7.52) | 0.83 |
| Meta-analysis | 36,305 | 95,624 | 25.2 | 73.6 | <12 | 123 | 0.85 (0.37 - 1.92) | 0.69 | 0.88 (0.38 - 2.02) | 0.76 |
| Meta-analysis (w/o USOC) | 36,305 | 95,624 | 25.2 | 73.6 | <12 | 123 | 0.85 (0.37 - 1.92) | 0.69 | 0.85 (0.37 - 1.93) | 0.70 |
| **PS-stratification / on-treatment TAR without escalation** | | | | | | | | | | |
| CCAE | 24,082 | 47,464 | 17.5 | 34.2 | 5 | 21 | 0.39 (0.12 - 1.07) | 0.09 | 0.41 (0.14 - 1.22) | 0.11 |
| DAG | 5,702 | 1,403 | 4.3 | 1.3 | 0 | 0 | | | | |
| ODOD | 17,471 | 49,460 | 10.8 | 40.3 | 7 | 103 | 0.54 (0.22 - 1.13) | 0.14 | 0.52 (0.23 - 1.20) | 0.13 |
| MDCR | 2,136 | 8,601 | 1.5 | 7.4 | 0 | <5 | | | | |
| MDCD | 1,507 | 7,493 | 0.8 | 4.4 | 0 | 0 | | | | |
| Open Claims | 332,801 | 894,802 | 246.2 | 821.2 | 0 | 0 | | | | |
| OEHR | 20,231 | 76,288 | 5.7 | 37.3 | 0 | 0 | | | | |
| VA | 12,350 | 109,470 | 6.4 | 88.9 | 18 | 407 | 0.61 (0.37 - 0.95) | 0.04 | 0.69 (0.42 - 1.13) | 0.20 |
| SIDIAP | 5,147 | 2,685 | 6.3 | 5.3 | 14 | <5 | 3.10 (0.71 - 16.4) | 0.16 | 3.02 (0.63 - 14.5) | 0.17 |
| Meta-analysis | 41,553 | 96,924 | 28.3 | 74.5 | 12 | 124 | 0.48 (0.25 - 0.92) | 0.03 | 0.50 (0.25 - 0.99) | 0.05 |
| Meta-analysis (w/o USOC) | 41,553 | 96,924 | 28.3 | 74.5 | 12 | 124 | 0.48 (0.25 - 0.92) | 0.03 | 0.49 (0.25 - 0.95) | 0.03 |

##### 1.3.8 Stroke

**Supplementary Table 60:** Exposures, outcomes and risk estimates for Stroke in SGLT2I (male) (T) vs SU (male) (C) new-user cohorts under four different propensity score (PS) adjustment / time-at-risk (TAR) designs across data sources. We report total patient exposure time in 1,000 patient-year (KYr) units, uncalibrated and calibrated hazard ratios (HRs) and their 95% confidence intervals (CIs). Greyed out entries did not pass study diagnostics and are included only for completeness.

| | Patients | | Exposure (KYr) | | Outcomes | | Uncalibrated | | Calibrated | |
|---|---|---|---|---|---|---|---|---|---|---|
| | T | C | T | C | T | C | HR (95% CI) | $p$ | HR (95% CI) | $p$ |
| **PS-matching / on-treatment TAR** | | | | | | | | | | |
| CCAE | 20,375 | 46,202 | 17.4 | 38.7 | 50 | 157 | 0.86 (0.55 - 1.32) | 0.49 | 0.87 (0.56 - 1.35) | 0.54 |
| DAG | 5,471 | 973 | 4.6 | 0.9 | 0 | 0 | | | | |
| ODOD | 15,392 | 47,482 | 10.9 | 43.0 | 73 | 506 | 0.82 (0.58 - 1.14) | 0.25 | 0.81 (0.55 - 1.19) | 0.31 |
| MDCR | 1,731 | 7,871 | 1.5 | 7.4 | 18 | 115 | 0.84 (0.38 - 1.72) | 0.64 | 0.84 (0.39 - 1.80) | 0.67 |
| MDCD | 1,404 | 6,941 | 0.9 | 4.8 | 6 | 67 | 0.82 (0.27 - 2.09) | 0.71 | 0.73 (0.26 - 2.05) | 0.56 |
| Open Claims | 297,554 | 862,949 | 265.9 | 932.4 | 874 | 6,002 | 0.75 (0.68 - 0.82) | <0.01 | 0.80 (0.65 - 0.98) | 0.03 |
| OEHR | 18,104 | 73,055 | 5.8 | 40.8 | 19 | 220 | 0.94 (0.51 - 1.63) | 0.83 | 0.95 (0.53 - 1.71) | 0.87 |
| VA | 0 | 0 | 0.0 | 0.0 | 0 | 0 | | | | |
| SIDIAP | 4,471 | 1,203 | 6.5 | 2.2 | 13 | 5 | 0.92 (0.24 - 3.80) | 0.90 | 0.86 (0.22 - 3.43) | 0.83 |
| Meta-analysis | 354,560 | 1,044,500 | 302.3 | 1067.1 | 1,040 | 7,067 | 0.76 (0.70 - 0.83) | <0.01 | 0.78 (0.65 - 0.95) | 0.01 |
| Meta-analysis (w/o USOC) | 57,006 | 181,551 | 36.4 | 134.7 | 166 | 1,065 | 0.85 (0.68 - 1.06) | 0.15 | 0.85 (0.67 - 1.07) | 0.20 |
| **PS-stratification / on-treatment TAR** | | | | | | | | | | |
| CCAE | 23,843 | 46,828 | 19.9 | 39.0 | 58 | 161 | 0.85 (0.60 - 1.17) | 0.32 | 0.87 (0.63 - 1.22) | 0.43 |
| DAG | 5,702 | 1,403 | 4.7 | 1.4 | 0 | 0 | | | | |
| ODOD | 17,093 | 48,024 | 11.8 | 43.5 | 81 | 519 | 0.82 (0.63 - 1.05) | 0.13 | 0.80 (0.59 - 1.09) | 0.19 |
| MDCR | 2,074 | 8,165 | 1.7 | 7.7 | 18 | 127 | 1.03 (0.57 - 1.78) | 0.92 | 1.00 (0.56 - 1.77) | 0.91 |
| MDCD | 1,446 | 7,166 | 0.9 | 5.0 | 6 | 70 | 0.79 (0.30 - 1.75) | 0.60 | 0.75 (0.31 - 1.82) | 0.53 |
| Open Claims | 324,481 | 868,102 | 283.2 | 937.7 | 934 | 6,104 | 0.72 (0.66 - 0.77) | <0.01 | 0.77 (0.62 - 0.95) | 0.02 |
| OEHR | 20,037 | 75,392 | 6.4 | 42.4 | 19 | 239 | 0.83 (0.48 - 1.33) | 0.46 | 0.89 (0.53 - 1.47) | 0.65 |
| VA | 12,216 | 109,267 | 7.4 | 100.4 | 19 | 364 | 0.67 (0.41 - 1.04) | 0.10 | 0.77 (0.43 - 1.38) | 0.39 |
| SIDIAP | 5,058 | 2,662 | 6.9 | 6.2 | 14 | 17 | 1.21 (0.43 - 3.40) | 0.71 | 1.12 (0.40 - 3.14) | 0.83 |
| Meta-analysis | 385,454 | 1,038,346 | 321.3 | 1062.6 | 1,092 | 7,023 | 0.74 (0.67 - 0.82) | <0.01 | 0.77 (0.63 - 0.94) | 0.01 |
| Meta-analysis (w/o USOC) | 60,973 | 170,244 | 38.1 | 124.9 | 158 | 919 | 0.83 (0.68 - 1.00) | 0.05 | 0.84 (0.68 - 1.04) | 0.13 |
| **PS-matching / on-treatment TAR without escalation** | | | | | | | | | | |
| CCAE | 20,375 | 46,202 | 15.2 | 33.5 | 38 | 140 | 0.80 (0.49 - 1.26) | 0.35 | 0.83 (0.52 - 1.32) | 0.43 |
| DAG | 5,471 | 973 | 4.2 | 0.8 | 0 | 0 | | | | |
| ODOD | 15,392 | 47,482 | 9.7 | 38.5 | 65 | 452 | 0.90 (0.63 - 1.27) | 0.55 | 0.88 (0.61 - 1.28) | 0.54 |
| MDCR | 1,731 | 7,871 | 1.3 | 6.8 | 15 | 108 | 0.68 (0.28 - 1.49) | 0.37 | 0.68 (0.29 - 1.56) | 0.37 |
| MDCD | 1,404 | 6,941 | 0.7 | 4.0 | <5 | 52 | 0.64 (0.15 - 1.95) | 0.50 | 0.57 (0.16 - 2.06) | 0.39 |
| Open Claims | 297,554 | 862,949 | 225.3 | 790.7 | 750 | 5,201 | 0.75 (0.68 - 0.83) | <0.01 | 0.81 (0.66 - 0.99) | 0.04 |
| OEHR | 18,104 | 73,055 | 5.0 | 35.4 | 12 | 169 | 0.75 (0.35 - 1.47) | 0.44 | 0.79 (0.39 - 1.61) | 0.52 |
| VA | 0 | 0 | 0.0 | 0.0 | 0 | 0 | | | | |
| SIDIAP | 4,471 | 1,203 | 5.8 | 1.9 | 12 | 5 | 1.30 (0.31 - 6.48) | 0.73 | 1.28 (0.28 - 5.82) | 0.75 |
| Meta-analysis | 354,560 | 1,044,500 | 257.2 | 908.9 | <885 | 6,122 | 0.76 (0.69 - 0.84) | <0.01 | 0.79 (0.66 - 0.95) | 0.01 |
| Meta-analysis (w/o USOC) | 57,006 | 181,551 | 31.9 | 118.2 | <135 | 921 | 0.82 (0.65 - 1.05) | 0.12 | 0.83 (0.64 - 1.06) | 0.16 |
| **PS-stratification / on-treatment TAR without escalation** | | | | | | | | | | |
| CCAE | 23,843 | 46,828 | 17.3 | 33.8 | 46 | 144 | 0.74 (0.51 - 1.05) | 0.10 | 0.77 (0.54 - 1.11) | 0.17 |
| DAG | 5,702 | 1,403 | 4.3 | 1.3 | 0 | 0 | | | | |
| ODOD | 17,093 | 48,024 | 10.5 | 38.9 | 72 | 463 | 0.82 (0.62 - 1.07) | 0.15 | 0.79 (0.59 - 1.08) | 0.17 |
| MDCR | 2,074 | 8,165 | 1.5 | 7.0 | 15 | 120 | 0.93 (0.49 - 1.68) | 0.82 | 0.89 (0.48 - 1.66) | 0.73 |
| MDCD | 1,446 | 7,166 | 0.7 | 4.2 | <5 | 53 | 0.60 (0.18 - 1.52) | 0.35 | 0.56 (0.19 - 1.66) | 0.30 |
| Open Claims | 324,481 | 868,102 | 240.8 | 795.4 | 806 | 5,285 | 0.72 (0.66 - 0.78) | <0.01 | 0.77 (0.62 - 0.96) | 0.02 |
| OEHR | 20,037 | 75,392 | 5.6 | 36.7 | 12 | 183 | 0.66 (0.34 - 1.19) | 0.20 | 0.71 (0.38 - 1.34) | 0.29 |
| VA | 12,216 | 109,267 | 6.4 | 88.9 | 14 | 317 | 0.59 (0.33 - 0.97) | 0.06 | 0.66 (0.38 - 1.16) | 0.20 |
| SIDIAP | 5,058 | 2,662 | 6.2 | 5.3 | 13 | 17 | 1.12 (0.38 - 3.25) | 0.83 | 1.10 (0.37 - 3.20) | 0.86 |
| Meta-analysis | 385,454 | 1,038,346 | 274.3 | 904.9 | 936 | 6,075 | 0.72 (0.67 - 0.78) | <0.01 | 0.76 (0.63 - 0.92) | <0.01 |
| Meta-analysis (w/o USOC) | 60,973 | 170,244 | 33.5 | 109.5 | 130 | 790 | 0.77 (0.63 - 0.95) | 0.01 | 0.79 (0.63 - 0.99) | 0.05 |

##### 1.3.9 Sudden cardiac death

**Supplementary Table 61:** Exposures, outcomes and risk estimates for Sudden cardiac death in SGLT2I (male) (T) vs SU (male) (C) new-user cohorts under four different propensity score (PS) adjustment / time-at-risk (TAR) designs across data sources. We report total patient exposure time in 1,000 patient-year (KYr) units, uncalibrated and calibrated hazard ratios (HRs) and their 95% confidence intervals (CIs). Greyed out entries did not pass study diagnostics and are included only for completeness.

| | Patients | | Exposure (KYr) | | Outcomes | | Uncalibrated | | Calibrated | |
|---|---|---|---|---|---|---|---|---|---|---|
| | T | C | T | C | T | C | HR (95% CI) | $p$ | HR (95% CI) | $p$ |
| **PS-matching / on-treatment TAR** | | | | | | | | | | |
| CCAE | 20,619 | 46,876 | 17.6 | 39.2 | 13 | 72 | 0.52 (0.23 - 1.07) | 0.10 | 0.53 (0.24 - 1.14) | 0.10 |
| DAG | 5,471 | 973 | 4.6 | 0.9 | 0 | 0 | | | | |
| ODOD | 15,698 | 48,810 | 11.2 | 44.5 | 18 | 167 | 0.47 (0.22 - 0.91) | 0.04 | 0.46 (0.22 - 0.96) | 0.04 |
| MDCR | 1,774 | 8,290 | 1.5 | 7.8 | 10 | 25 | 2.32 (0.63 - 7.81) | 0.19 | 2.33 (0.66 - 8.23) | 0.19 |
| MDCD | 1,457 | 7,215 | 0.9 | 5.1 | <5 | 22 | 0.70 (0.03 - 4.80) | 0.78 | 0.62 (0.05 - 7.35) | 0.71 |
| Open Claims | 304,126 | 887,080 | 271.0 | 959.7 | 231 | 1,802 | 0.58 (0.48 - 0.69) | <0.01 | 0.61 (0.47 - 0.80) | <0.01 |
| OEHR | 18,264 | 73,759 | 5.8 | 41.4 | 5 | 79 | 0.66 (0.21 - 1.73) | 0.44 | 0.67 (0.24 - 1.93) | 0.47 |
| VA | 0 | 0 | 0.0 | 0.0 | 0 | 0 | | | | |
| SIDIAP | 4,551 | 1,220 | 6.6 | 2.2 | 0 | 0 | | | | |
| Meta-analysis | 360,481 | 1,064,815 | 307.2 | 1092.6 | 277 | 2,145 | 0.58 (0.49 - 0.69) | <0.01 | 0.60 (0.47 - 0.77) | <0.01 |
| Meta-analysis (w/o USOC) | 56,355 | 177,735 | 36.2 | 132.9 | 46 | 343 | 0.67 (0.37 - 1.21) | 0.19 | 0.67 (0.37 - 1.21) | 0.19 |
| **PS-stratification / on-treatment TAR** | | | | | | | | | | |
| CCAE | 24,111 | 47,518 | 20.1 | 39.6 | 15 | 72 | 0.47 (0.25 - 0.85) | 0.02 | 0.49 (0.27 - 0.90) | 0.02 |
| DAG | 5,702 | 1,403 | 4.7 | 1.4 | 0 | 0 | | | | |
| ODOD | 17,442 | 49,468 | 12.1 | 45.1 | 19 | 176 | 0.54 (0.31 - 0.89) | 0.02 | 0.53 (0.31 - 0.92) | 0.03 |
| MDCR | 2,130 | 8,575 | 1.7 | 8.0 | 11 | 26 | 2.70 (1.08 - 6.43) | 0.03 | 2.62 (1.07 - 6.38) | 0.04 |
| MDCD | 1,499 | 7,461 | 0.9 | 5.3 | <5 | 24 | 0.47 (0.07 - 1.76) | 0.36 | 0.45 (0.09 - 2.20) | 0.32 |
| Open Claims | 331,685 | 892,613 | 288.8 | 965.5 | 259 | 1,851 | 0.62 (0.54 - 0.71) | <0.01 | 0.66 (0.52 - 0.84) | <0.01 |
| OEHR | 20,205 | 76,222 | 6.5 | 43.0 | 5 | 85 | 0.55 (0.18 - 1.37) | 0.24 | 0.59 (0.21 - 1.63) | 0.31 |
| VA | 12,496 | 111,025 | 7.5 | 102.2 | <10 | 47 | 1.55 (0.53 - 3.57) | 0.37 | 1.76 (0.64 - 4.86) | 0.28 |
| SIDIAP | 5,160 | 2,691 | 7.1 | 6.3 | 0 | <5 | | | | |
| Meta-analysis | 393,443 | 1,065,821 | 327.5 | 1093.2 | 298 | 2,184 | 0.60 (0.53 - 0.69) | <0.01 | 0.63 (0.50 - 0.78) | <0.01 |
| Meta-analysis (w/o USOC) | 61,758 | 173,208 | 38.7 | 127.7 | 39 | 333 | 0.52 (0.36 - 0.75) | <0.01 | 0.53 (0.36 - 0.77) | <0.01 |
| **PS-matching / on-treatment TAR without escalation** | | | | | | | | | | |
| CCAE | 20,619 | 46,876 | 15.3 | 34.0 | 9 | 64 | 0.45 (0.18 - 1.00) | 0.07 | 0.46 (0.19 - 1.10) | 0.08 |
| DAG | 5,471 | 973 | 4.2 | 0.8 | 0 | 0 | | | | |
| ODOD | 15,698 | 48,810 | 9.9 | 39.8 | 15 | 149 | 0.49 (0.22 - 1.00) | 0.07 | 0.48 (0.22 - 1.05) | 0.07 |
| MDCR | 1,774 | 8,290 | 1.4 | 7.2 | 9 | 19 | 5.28 (1.30 - 19.7) | 0.02 | 5.21 (1.34 - 20.3) | 0.02 |
| MDCD | 1,457 | 7,215 | 0.8 | 4.2 | <5 | 20 | 0.86 (0.04 - 6.42) | 0.90 | 0.75 (0.06 - 9.50) | 0.83 |
| Open Claims | 304,126 | 887,080 | 229.6 | 813.9 | 202 | 1,532 | 0.60 (0.49 - 0.73) | <0.01 | 0.64 (0.49 - 0.84) | <0.01 |
| OEHR | 18,264 | 73,759 | 5.1 | 35.9 | <5 | 55 | 0.51 (0.11 - 1.68) | 0.33 | 0.53 (0.14 - 2.06) | 0.36 |
| VA | 0 | 0 | 0.0 | 0.0 | 0 | 0 | | | | |
| SIDIAP | 4,551 | 1,220 | 5.9 | 2.0 | 0 | 0 | | | | |
| Meta-analysis | 358,707 | 1,056,525 | 260.0 | 923.5 | <231 | 1,800 | 0.58 (0.48 - 0.70) | <0.01 | 0.60 (0.47 - 0.77) | <0.01 |
| Meta-analysis (w/o USOC) | 54,581 | 169,445 | 30.4 | 109.6 | <29 | 268 | 0.48 (0.28 - 0.81) | <0.01 | 0.48 (0.28 - 0.82) | <0.01 |
| **PS-stratification / on-treatment TAR without escalation** | | | | | | | | | | |
| CCAE | 24,111 | 47,518 | 17.5 | 34.3 | 11 | 64 | 0.36 (0.17 - 0.70) | <0.01 | 0.38 (0.19 - 0.77) | <0.01 |
| DAG | 5,702 | 1,403 | 4.3 | 1.3 | 0 | 0 | | | | |
| ODOD | 17,442 | 49,468 | 10.7 | 40.3 | 16 | 156 | 0.52 (0.29 - 0.88) | 0.02 | 0.50 (0.28 - 0.89) | 0.02 |
| MDCR | 2,130 | 8,575 | 1.5 | 7.4 | 10 | 20 | 3.91 (1.43 - 10.2) | <0.01 | 3.75 (1.41 - 10.00) | <0.01 |
| MDCD | 1,499 | 7,461 | 0.8 | 4.4 | <5 | 20 | 0.49 (0.08 - 1.87) | 0.39 | 0.47 (0.09 - 2.32) | 0.35 |
| Open Claims | 331,685 | 892,613 | 245.5 | 818.9 | 228 | 1,575 | 0.64 (0.55 - 0.75) | <0.01 | 0.69 (0.54 - 0.89) | <0.01 |
| OEHR | 20,205 | 76,222 | 5.7 | 37.3 | <5 | 60 | 0.38 (0.09 - 1.18) | 0.15 | 0.40 (0.11 - 1.50) | 0.18 |
| VA | 12,496 | 111,025 | 6.5 | 90.4 | <10 | 38 | 1.55 (0.46 - 3.91) | 0.42 | 1.75 (0.60 - 5.13) | 0.32 |
| SIDIAP | 5,160 | 2,691 | 6.3 | 5.4 | 0 | <5 | | | | |
| Meta-analysis | 393,443 | 1,065,821 | 279.4 | 930.8 | <260 | 1,855 | 0.56 (0.42 - 0.74) | <0.01 | 0.58 (0.42 - 0.81) | <0.01 |
| Meta-analysis (w/o USOC) | 61,758 | 173,208 | 34.0 | 111.9 | <32 | 280 | 0.44 (0.29 - 0.67) | <0.01 | 0.45 (0.30 - 0.69) | <0.01 |

##### 1.3.10 Abnormal weight gain

**Supplementary Table 62:** Exposures, outcomes and risk estimates for Abnormal weight gain in SGLT2I (male) (T) vs SU (male) (C) new-user cohorts under four different propensity score (PS) adjustment / time-at-risk (TAR) designs across data sources. We report total patient exposure time in 1,000 patient-year (KYr) units, uncalibrated and calibrated hazard ratios (HRs) and their 95% confidence intervals (CIs). Greyed out entries did not pass study diagnostics and are included only for completeness.

| | Patients | | Exposure (KYr) | | Outcomes | | Uncalibrated | | Calibrated | |
|---|---|---|---|---|---|---|---|---|---|---|
| | T | C | T | C | T | C | HR (95% CI) | $p$ | HR (95% CI) | $p$ |
| **PS-matching / on-treatment TAR** | | | | | | | | | | |
| CCAE | 19,985 | 45,765 | 17.0 | 38.2 | 53 | 175 | 0.69 (0.44 - 1.04) | 0.09 | 0.70 (0.45 - 1.07) | 0.10 |
| DAG | 5,432 | 968 | 4.5 | 0.9 | 0 | <5 | | | | |
| ODOD | 15,208 | 47,704 | 10.8 | 43.3 | 32 | 244 | 0.40 (0.23 - 0.63) | <0.01 | 0.39 (0.23 - 0.66) | <0.01 |
| MDCR | 1,747 | 8,227 | 1.5 | 7.7 | <5 | 31 | 0.16 (0.01 - 0.83) | 0.11 | 0.16 (0.02 - 1.57) | 0.12 |
| MDCD | 1,367 | 6,949 | 0.8 | 4.8 | <5 | 49 | 0.50 (0.11 - 1.53) | 0.29 | 0.44 (0.12 - 1.62) | 0.22 |
| Open Claims | 294,899 | 868,913 | 262.1 | 939.0 | 290 | 1,795 | 0.55 (0.46 - 0.65) | <0.01 | 0.58 (0.45 - 0.75) | <0.01 |
| OEHR | 17,680 | 72,048 | 5.6 | 40.2 | 26 | 216 | 0.89 (0.51 - 1.47) | 0.65 | 0.90 (0.53 - 1.53) | 0.71 |
| VA | 0 | 0 | 0.0 | 0.0 | 0 | 0 | | | | |
| SIDIAP | 4,500 | 1,199 | 6.5 | 2.2 | 8 | <5 | 1.85 (0.38 - 13.3) | 0.50 | 1.74 (0.30 - 10.2) | 0.54 |
| Meta-analysis | 350,886 | 1,049,606 | 297.8 | 1073.2 | <411 | 2,510 | 0.57 (0.46 - 0.72) | <0.01 | 0.59 (0.45 - 0.78) | <0.01 |
| Meta-analysis (w/o USOC) | 55,987 | 180,693 | 35.7 | 134.2 | <121 | 715 | 0.59 (0.40 - 0.87) | <0.01 | 0.59 (0.40 - 0.87) | <0.01 |
| **PS-stratification / on-treatment TAR** | | | | | | | | | | |
| CCAE | 23,300 | 46,404 | 19.3 | 38.5 | 68 | 176 | 0.72 (0.52 - 0.97) | 0.03 | 0.74 (0.54 - 1.01) | 0.06 |
| DAG | 5,661 | 1,392 | 4.7 | 1.4 | 0 | <5 | | | | |
| ODOD | 16,867 | 48,341 | 11.7 | 43.9 | 32 | 250 | 0.47 (0.31 - 0.68) | <0.01 | 0.46 (0.30 - 0.70) | <0.01 |
| MDCR | 2,099 | 8,500 | 1.7 | 7.9 | <5 | 34 | 0.26 (0.04 - 1.00) | 0.10 | 0.25 (0.05 - 1.27) | 0.10 |
| MDCD | 1,408 | 7,185 | 0.8 | 5.0 | <5 | 51 | 0.43 (0.13 - 1.10) | 0.13 | 0.40 (0.14 - 1.20) | 0.11 |
| Open Claims | 321,381 | 874,572 | 279.0 | 944.9 | 317 | 1,820 | 0.62 (0.54 - 0.70) | <0.01 | 0.66 (0.52 - 0.84) | <0.01 |
| OEHR | 19,556 | 74,612 | 6.2 | 41.9 | 30 | 229 | 0.88 (0.57 - 1.31) | 0.53 | 0.94 (0.62 - 1.43) | 0.77 |
| VA | 12,226 | 109,285 | 7.4 | 100.4 | <10 | 172 | 0.68 (0.32 - 1.26) | 0.27 | 0.78 (0.36 - 1.67) | 0.53 |
| SIDIAP | 5,107 | 2,666 | 7.0 | 6.2 | 8 | 8 | 2.52 (0.74 - 8.29) | 0.13 | 2.31 (0.69 - 7.75) | 0.18 |
| Meta-analysis | 381,104 | 1,043,929 | 316.2 | 1069.2 | 447 | 2,475 | 0.64 (0.55 - 0.75) | <0.01 | 0.66 (0.53 - 0.84) | <0.01 |
| Meta-analysis (w/o USOC) | 59,723 | 169,357 | 37.2 | 124.4 | 130 | 655 | 0.66 (0.47 - 0.93) | 0.02 | 0.68 (0.48 - 0.96) | 0.03 |
| **PS-matching / on-treatment TAR without escalation** | | | | | | | | | | |
| CCAE | 19,985 | 45,765 | 14.8 | 33.1 | 47 | 147 | 0.74 (0.47 - 1.15) | 0.19 | 0.76 (0.49 - 1.20) | 0.25 |
| DAG | 5,432 | 968 | 4.2 | 0.8 | 0 | <5 | | | | |
| ODOD | 15,208 | 47,704 | 9.6 | 38.8 | 25 | 211 | 0.39 (0.21 - 0.65) | <0.01 | 0.38 (0.21 - 0.67) | <0.01 |
| MDCR | 1,747 | 8,227 | 1.3 | 7.1 | <5 | 28 | 0.17 (0.01 - 0.88) | 0.13 | 0.16 (0.02 - 1.63) | 0.12 |
| MDCD | 1,367 | 6,949 | 0.7 | 4.0 | <5 | 35 | 0.69 (0.15 - 2.27) | 0.59 | 0.61 (0.16 - 2.35) | 0.48 |
| Open Claims | 294,899 | 868,913 | 222.2 | 797.2 | 229 | 1,485 | 0.52 (0.43 - 0.63) | <0.01 | 0.56 (0.44 - 0.73) | <0.01 |
| OEHR | 17,680 | 72,048 | 4.9 | 34.9 | 21 | 178 | 0.86 (0.47 - 1.51) | 0.62 | 0.90 (0.50 - 1.62) | 0.74 |
| VA | 0 | 0 | 0.0 | 0.0 | 0 | 0 | | | | |
| SIDIAP | 4,500 | 1,199 | 5.8 | 1.9 | 8 | <5 | 3.61 (0.55 - 70.5) | 0.30 | 3.55 (0.31 - 40.0) | 0.31 |
| Meta-analysis | 350,886 | 1,049,606 | 253.5 | 915.1 | <332 | 2,084 | 0.57 (0.45 - 0.74) | <0.01 | 0.60 (0.44 - 0.80) | <0.01 |
| Meta-analysis (w/o USOC) | 55,987 | 180,693 | 31.3 | 118.0 | <103 | 599 | 0.61 (0.41 - 0.91) | 0.01 | 0.61 (0.41 - 0.91) | 0.02 |
| **PS-stratification / on-treatment TAR without escalation** | | | | | | | | | | |
| CCAE | 23,300 | 46,404 | 16.9 | 33.4 | 60 | 148 | 0.75 (0.54 - 1.04) | 0.08 | 0.79 (0.56 - 1.09) | 0.16 |
| DAG | 5,661 | 1,392 | 4.3 | 1.3 | 0 | <5 | | | | |
| ODOD | 16,867 | 48,341 | 10.4 | 39.3 | 25 | 217 | 0.43 (0.27 - 0.65) | <0.01 | 0.41 (0.26 - 0.66) | <0.01 |
| MDCR | 2,099 | 8,500 | 1.5 | 7.3 | <5 | 31 | 0.27 (0.04 - 1.03) | 0.11 | 0.26 (0.05 - 1.30) | 0.10 |
| MDCD | 1,408 | 7,185 | 0.7 | 4.2 | <5 | 36 | 0.64 (0.18 - 1.72) | 0.43 | 0.60 (0.20 - 1.84) | 0.38 |
| Open Claims | 321,381 | 874,572 | 237.4 | 802.3 | 254 | 1,507 | 0.59 (0.51 - 0.68) | <0.01 | 0.63 (0.49 - 0.81) | <0.01 |
| OEHR | 19,556 | 74,612 | 5.5 | 36.4 | 24 | 188 | 0.79 (0.49 - 1.24) | 0.33 | 0.85 (0.53 - 1.36) | 0.50 |
| VA | 12,226 | 109,285 | 6.4 | 88.9 | <10 | 131 | 0.80 (0.36 - 1.54) | 0.55 | 0.91 (0.43 - 1.89) | 0.80 |
| SIDIAP | 5,107 | 2,666 | 6.2 | 5.3 | 8 | 6 | 3.28 (0.92 - 11.7) | 0.07 | 3.19 (0.89 - 11.4) | 0.08 |
| Meta-analysis | 381,104 | 1,043,929 | 270.1 | 911.5 | 363 | 2,060 | 0.62 (0.50 - 0.75) | <0.01 | 0.65 (0.50 - 0.84) | <0.01 |
| Meta-analysis (w/o USOC) | 59,723 | 169,357 | 32.7 | 109.2 | 109 | 553 | 0.64 (0.44 - 0.93) | 0.02 | 0.65 (0.45 - 0.95) | 0.03 |

##### 1.3.11 Abnormal weight loss

**Supplementary Table 63:** Exposures, outcomes and risk estimates for Abnormal weight loss in SGLT2I (male) (T) vs SU (male) (C) new-user cohorts under four different propensity score (PS) adjustment / time-at-risk (TAR) designs across data sources. We report total patient exposure time in 1,000 patient-year (KYr) units, uncalibrated and calibrated hazard ratios (HRs) and their 95% confidence intervals (CIs). Greyed out entries did not pass study diagnostics and are included only for completeness.

| | Patients | | Exposure (KYr) | | Outcomes | | Uncalibrated | | Calibrated | |
|---|---|---|---|---|---|---|---|---|---|---|
| | T | C | T | C | T | C | HR (95% CI) | $p$ | HR (95% CI) | $p$ |
| **PS-matching / on-treatment TAR** | | | | | | | | | | |
| CCAE | 20,137 | 45,662 | 17.1 | 38.2 | 124 | 211 | 1.51 (1.11 - 2.04) | <0.01 | 1.53 (1.12 - 2.08) | <0.01 |
| DAG | 5,363 | 957 | 4.5 | 0.9 | 39 | <5 | 1.76 (0.45 - 11.8) | 0.50 | 3.30 (0.65 - 16.8) | 0.16 |
| ODOD | 15,188 | 46,639 | 10.8 | 42.3 | 173 | 569 | 1.42 (1.12 - 1.80) | <0.01 | 1.40 (1.03 - 1.89) | 0.05 |
| MDCR | 1,712 | 7,847 | 1.5 | 7.4 | 32 | 113 | 1.42 (0.81 - 2.44) | 0.21 | 1.43 (0.82 - 2.48) | 0.22 |
| MDCD | 1,400 | 6,848 | 0.8 | 4.8 | 14 | 50 | 2.22 (0.92 - 5.17) | 0.07 | 1.98 (0.83 - 4.70) | 0.13 |
| Open Claims | 295,525 | 858,080 | 262.7 | 927.2 | 1,500 | 4,775 | 1.44 (1.33 - 1.56) | <0.01 | 1.54 (1.26 - 1.88) | <0.01 |
| OEHR | 17,739 | 71,325 | 5.6 | 39.8 | 72 | 410 | 1.79 (1.26 - 2.53) | <0.01 | 1.82 (1.28 - 2.59) | <0.01 |
| VA | 0 | 0 | 0.0 | 0.0 | 0 | 0 | | | | |
| SIDIAP | 4,493 | 1,206 | 6.5 | 2.2 | 29 | 8 | 1.49 (0.45 - 6.76) | 0.57 | 1.40 (0.36 - 5.41) | 0.63 |
| Meta-analysis | 361,557 | 1,038,564 | 309.5 | 1062.8 | 1,983 | <6,141 | 1.46 (1.36 - 1.57) | <0.01 | 1.51 (1.25 - 1.82) | <0.01 |
| Meta-analysis (w/o USOC) | 66,032 | 180,484 | 46.8 | 135.6 | 483 | <1,366 | 1.53 (1.32 - 1.79) | <0.01 | 1.53 (1.29 - 1.81) | <0.01 |
| **PS-stratification / on-treatment TAR** | | | | | | | | | | |
| CCAE | 23,568 | 46,288 | 19.6 | 38.5 | 145 | 215 | 1.41 (1.11 - 1.79) | <0.01 | 1.46 (1.15 - 1.85) | <0.01 |
| DAG | 5,592 | 1,377 | 4.6 | 1.4 | 39 | 7 | 1.03 (0.40 - 3.10) | 0.95 | 1.35 (0.48 - 3.79) | 0.57 |
| ODOD | 16,888 | 47,195 | 11.6 | 42.9 | 184 | 582 | 1.42 (1.17 - 1.71) | <0.01 | 1.39 (1.09 - 1.78) | 0.02 |
| MDCR | 2,062 | 8,142 | 1.6 | 7.7 | 35 | 119 | 1.54 (0.97 - 2.38) | 0.06 | 1.49 (0.95 - 2.33) | 0.09 |
| MDCD | 1,443 | 7,139 | 0.9 | 5.0 | 14 | 55 | 1.92 (0.96 - 3.63) | 0.05 | 1.82 (0.94 - 3.53) | 0.08 |
| Open Claims | 322,505 | 863,615 | 280.1 | 933.0 | 1,595 | 4,867 | 1.36 (1.27 - 1.44) | <0.01 | 1.45 (1.18 - 1.79) | <0.01 |
| OEHR | 19,644 | 73,670 | 6.3 | 41.3 | 79 | 446 | 1.51 (1.14 - 1.97) | <0.01 | 1.62 (1.23 - 2.12) | <0.01 |
| VA | 12,060 | 106,937 | 7.2 | 98.2 | 100 | 668 | 1.98 (1.59 - 2.43) | <0.01 | 2.25 (1.48 - 3.41) | <0.01 |
| SIDIAP | 5,093 | 2,666 | 7.0 | 6.2 | 36 | 18 | 0.79 (0.34 - 1.90) | 0.59 | 0.73 (0.31 - 1.72) | 0.47 |
| Meta-analysis | 382,605 | 1,030,768 | 317.5 | 1055.8 | 2,003 | 6,110 | 1.37 (1.30 - 1.45) | <0.01 | 1.42 (1.19 - 1.71) | <0.01 |
| Meta-analysis (w/o USOC) | 60,100 | 167,153 | 37.5 | 122.7 | 408 | 1,243 | 1.44 (1.26 - 1.64) | <0.01 | 1.46 (1.25 - 1.71) | <0.01 |
| **PS-matching / on-treatment TAR without escalation** | | | | | | | | | | |
| CCAE | 20,137 | 45,662 | 14.9 | 33.1 | 109 | 184 | 1.52 (1.10 - 2.11) | 0.01 | 1.57 (1.14 - 2.18) | <0.01 |
| DAG | 5,363 | 957 | 4.1 | 0.8 | 36 | <5 | 1.88 (0.47 - 12.7) | 0.45 | 3.69 (0.71 - 19.1) | 0.13 |
| ODOD | 15,188 | 46,639 | 9.5 | 37.9 | 150 | 497 | 1.45 (1.12 - 1.87) | <0.01 | 1.42 (1.07 - 1.89) | 0.03 |
| MDCR | 1,712 | 7,847 | 1.3 | 6.8 | 29 | 100 | 1.60 (0.90 - 2.78) | 0.10 | 1.58 (0.90 - 2.79) | 0.13 |
| MDCD | 1,400 | 6,848 | 0.7 | 4.0 | 13 | 36 | 2.22 (0.87 - 5.43) | 0.09 | 1.96 (0.79 - 4.90) | 0.16 |
| Open Claims | 295,525 | 858,080 | 222.8 | 786.7 | 1,301 | 4,071 | 1.48 (1.35 - 1.61) | <0.01 | 1.58 (1.30 - 1.93) | <0.01 |
| OEHR | 17,739 | 71,325 | 4.9 | 34.5 | 65 | 336 | 1.74 (1.19 - 2.52) | <0.01 | 1.82 (1.25 - 2.65) | <0.01 |
| VA | 0 | 0 | 0.0 | 0.0 | 0 | 0 | | | | |
| SIDIAP | 4,493 | 1,206 | 5.8 | 1.9 | 25 | 6 | 2.17 (0.55 - 14.5) | 0.35 | 2.13 (0.42 - 10.9) | 0.37 |
| Meta-analysis | 361,557 | 1,038,564 | 264.1 | 905.8 | 1,728 | <5,235 | 1.49 (1.38 - 1.61) | <0.01 | 1.55 (1.30 - 1.85) | <0.01 |
| Meta-analysis (w/o USOC) | 66,032 | 180,484 | 41.3 | 119.1 | 427 | <1,164 | 1.57 (1.33 - 1.85) | <0.01 | 1.57 (1.32 - 1.87) | <0.01 |
| **PS-stratification / on-treatment TAR without escalation** | | | | | | | | | | |
| CCAE | 23,568 | 46,288 | 17.1 | 33.4 | 126 | 188 | 1.42 (1.10 - 1.84) | <0.01 | 1.49 (1.15 - 1.93) | <0.01 |
| DAG | 5,592 | 1,377 | 4.2 | 1.2 | 36 | 5 | 1.35 (0.45 - 4.94) | 0.62 | 1.88 (0.57 - 6.23) | 0.31 |
| ODOD | 16,888 | 47,195 | 10.3 | 38.4 | 160 | 510 | 1.47 (1.20 - 1.80) | <0.01 | 1.43 (1.13 - 1.82) | <0.01 |
| MDCR | 2,062 | 8,142 | 1.5 | 7.1 | 31 | 105 | 1.58 (0.97 - 2.50) | 0.06 | 1.51 (0.94 - 2.43) | 0.09 |
| MDCD | 1,443 | 7,139 | 0.7 | 4.2 | 13 | 41 | 2.27 (1.09 - 4.48) | 0.02 | 2.13 (1.05 - 4.34) | 0.04 |
| Open Claims | 322,505 | 863,615 | 238.3 | 791.7 | 1,386 | 4,156 | 1.41 (1.32 - 1.51) | <0.01 | 1.52 (1.23 - 1.88) | <0.01 |
| OEHR | 19,644 | 73,670 | 5.5 | 35.8 | 70 | 368 | 1.60 (1.19 - 2.12) | <0.01 | 1.71 (1.28 - 2.29) | <0.01 |
| VA | 12,060 | 106,937 | 6.3 | 86.9 | 87 | 571 | 2.05 (1.62 - 2.56) | <0.01 | 2.31 (1.78 - 3.01) | <0.01 |
| SIDIAP | 5,093 | 2,666 | 6.2 | 5.3 | 32 | 15 | 0.95 (0.38 - 2.46) | 0.92 | 0.93 (0.36 - 2.37) | 0.85 |
| Meta-analysis | 382,605 | 1,030,768 | 271.2 | 899.4 | 1,742 | 5,222 | 1.42 (1.34 - 1.51) | <0.01 | 1.49 (1.24 - 1.79) | <0.01 |
| Meta-analysis (w/o USOC) | 60,100 | 167,153 | 32.9 | 107.6 | 356 | 1,066 | 1.48 (1.29 - 1.71) | <0.01 | 1.51 (1.29 - 1.78) | <0.01 |

##### 1.3.12 Acute pancreatitis

**Supplementary Table 64:** Exposures, outcomes and risk estimates for Acute pancreatitis in SGLT2I (male) (T) vs SU (male) (C) new-user cohorts under four different propensity score (PS) adjustment / time-at-risk (TAR) designs across data sources. We report total patient exposure time in 1,000 patient-year (KYr) units, uncalibrated and calibrated hazard ratios (HRs) and their 95% confidence intervals (CIs). Greyed out entries did not pass study diagnostics and are included only for completeness.

| | Patients | | Exposure (KYr) | | Outcomes | | Uncalibrated | | Calibrated | |
|---|---|---|---|---|---|---|---|---|---|---|
| | T | C | T | C | T | C | HR (95% CI) | $p$ | HR (95% CI) | $p$ |
| **PS-matching / on-treatment TAR** | | | | | | | | | | |
| CCAE | 20,418 | 46,370 | 17.4 | 38.8 | 36 | 108 | 0.55 (0.32 - 0.91) | 0.02 | 0.55 (0.33 - 0.94) | 0.03 |
| DAG | 5,471 | 973 | 4.6 | 0.9 | 0 | 0 | | | | |
| ODOD | 15,572 | 48,378 | 11.1 | 44.0 | 27 | 164 | 1.35 (0.75 - 2.32) | 0.30 | 1.32 (0.73 - 2.40) | 0.37 |
| MDCR | 1,762 | 8,208 | 1.5 | 7.7 | <5 | 27 | 0.55 (0.08 - 2.27) | 0.48 | 0.55 (0.10 - 2.94) | 0.49 |
| MDCD | 1,436 | 7,035 | 0.9 | 4.9 | 6 | 38 | 0.98 (0.31 - 2.68) | 0.98 | 0.87 (0.30 - 2.59) | 0.81 |
| Open Claims | 300,492 | 875,565 | 267.7 | 947.3 | 357 | 2,128 | 0.68 (0.58 - 0.79) | <0.01 | 0.72 (0.57 - 0.92) | <0.01 |
| OEHR | 18,169 | 73,312 | 5.8 | 41.1 | 11 | 126 | 0.81 (0.36 - 1.66) | 0.59 | 0.82 (0.38 - 1.77) | 0.62 |
| VA | 0 | 0 | 0.0 | 0.0 | 0 | 0 | | | | |
| SIDIAP | 4,533 | 1,218 | 6.6 | 2.2 | <5 | 0 | | | | |
| Meta-analysis | 357,849 | 1,058,868 | 304.4 | 1083.8 | <442 | 2,591 | 0.76 (0.57 - 1.03) | 0.07 | 0.79 (0.56 - 1.11) | 0.17 |
| Meta-analysis (w/o USOC) | 57,357 | 183,303 | 36.7 | 136.5 | <85 | 463 | 0.83 (0.55 - 1.25) | 0.37 | 0.83 (0.55 - 1.25) | 0.38 |
| **PS-stratification / on-treatment TAR** | | | | | | | | | | |
| CCAE | 23,890 | 47,008 | 19.9 | 39.1 | 42 | 109 | 0.67 (0.45 - 0.98) | 0.04 | 0.69 (0.47 - 1.02) | 0.07 |
| DAG | 5,702 | 1,403 | 4.7 | 1.4 | 0 | 0 | | | | |
| ODOD | 17,307 | 49,005 | 12.0 | 44.6 | 29 | 167 | 0.84 (0.54 - 1.28) | 0.45 | 0.83 (0.52 - 1.31) | 0.44 |
| MDCR | 2,119 | 8,479 | 1.7 | 8.0 | <5 | 27 | 0.55 (0.08 - 2.02) | 0.46 | 0.53 (0.11 - 2.61) | 0.44 |
| MDCD | 1,481 | 7,259 | 0.9 | 5.1 | 6 | 41 | 1.20 (0.43 - 2.85) | 0.70 | 1.14 (0.44 - 2.92) | 0.79 |
| Open Claims | 327,945 | 880,953 | 285.4 | 952.9 | 377 | 2,155 | 0.70 (0.62 - 0.79) | <0.01 | 0.75 (0.59 - 0.94) | 0.02 |
| OEHR | 20,108 | 75,742 | 6.4 | 42.7 | 13 | 133 | 0.87 (0.45 - 1.55) | 0.65 | 0.93 (0.50 - 1.73) | 0.82 |
| VA | 12,372 | 109,694 | 7.4 | 100.9 | <10 | 214 | 0.25 (0.08 - 0.58) | <0.01 | 0.28 (0.10 - 0.82) | 0.02 |
| SIDIAP | 5,142 | 2,685 | 7.0 | 6.3 | <5 | <5 | 2.46 (0.15 - 36.1) | 0.52 | 2.26 (0.15 - 34.9) | 0.56 |
| Meta-analysis | 389,250 | 1,052,708 | 323.7 | 1079.3 | 461 | 2,564 | 0.71 (0.64 - 0.79) | <0.01 | 0.74 (0.60 - 0.91) | <0.01 |
| Meta-analysis (w/o USOC) | 61,305 | 171,755 | 38.3 | 126.4 | 84 | 409 | 0.76 (0.59 - 0.99) | 0.04 | 0.78 (0.59 - 1.03) | 0.09 |
| **PS-matching / on-treatment TAR without escalation** | | | | | | | | | | |
| CCAE | 20,418 | 46,370 | 15.2 | 33.6 | 28 | 91 | 0.56 (0.31 - 0.97) | 0.05 | 0.58 (0.33 - 1.03) | 0.07 |
| DAG | 5,471 | 973 | 4.2 | 0.8 | 0 | 0 | | | | |
| ODOD | 15,572 | 48,378 | 9.8 | 39.4 | 21 | 146 | 1.52 (0.83 - 2.69) | 0.16 | 1.49 (0.82 - 2.73) | 0.20 |
| MDCR | 1,762 | 8,208 | 1.3 | 7.1 | <5 | 24 | 0.71 (0.10 - 3.26) | 0.70 | 0.70 (0.12 - 4.03) | 0.69 |
| MDCD | 1,436 | 7,035 | 0.7 | 4.1 | 5 | 33 | 0.75 (0.21 - 2.19) | 0.64 | 0.67 (0.20 - 2.17) | 0.50 |
| Open Claims | 300,492 | 875,565 | 227.1 | 804.0 | 288 | 1,765 | 0.68 (0.57 - 0.80) | <0.01 | 0.73 (0.57 - 0.93) | 0.01 |
| OEHR | 18,169 | 73,312 | 5.1 | 35.7 | 9 | 90 | 0.88 (0.34 - 2.00) | 0.78 | 0.92 (0.38 - 2.23) | 0.86 |
| VA | 0 | 0 | 0.0 | 0.0 | 0 | 0 | | | | |
| SIDIAP | 4,533 | 1,218 | 5.8 | 2.0 | <5 | 0 | | | | |
| Meta-analysis | 357,849 | 1,058,868 | 259.2 | 923.8 | <356 | 2,149 | 0.79 (0.56 - 1.13) | 0.20 | 0.82 (0.56 - 1.21) | 0.33 |
| Meta-analysis (w/o USOC) | 57,357 | 183,303 | 32.1 | 119.8 | <68 | 384 | 0.88 (0.56 - 1.38) | 0.57 | 0.88 (0.56 - 1.39) | 0.59 |
| **PS-stratification / on-treatment TAR without escalation** | | | | | | | | | | |
| CCAE | 23,890 | 47,008 | 17.4 | 34.0 | 34 | 92 | 0.66 (0.42 - 1.00) | 0.06 | 0.69 (0.44 - 1.06) | 0.09 |
| DAG | 5,702 | 1,403 | 4.3 | 1.3 | 0 | 0 | | | | |
| ODOD | 17,307 | 49,005 | 10.7 | 39.9 | 23 | 149 | 0.76 (0.46 - 1.21) | 0.27 | 0.74 (0.45 - 1.22) | 0.25 |
| MDCR | 2,119 | 8,479 | 1.5 | 7.3 | <5 | 24 | 0.59 (0.09 - 2.23) | 0.52 | 0.57 (0.11 - 2.83) | 0.49 |
| MDCD | 1,481 | 7,259 | 0.7 | 4.2 | 5 | 36 | 1.09 (0.35 - 2.77) | 0.87 | 1.03 (0.37 - 2.88) | 0.93 |
| Open Claims | 327,945 | 880,953 | 242.8 | 808.8 | 306 | 1,785 | 0.68 (0.60 - 0.78) | <0.01 | 0.73 (0.58 - 0.93) | 0.01 |
| OEHR | 20,108 | 75,742 | 5.7 | 37.0 | 11 | 96 | 1.03 (0.50 - 1.94) | 0.94 | 1.10 (0.56 - 2.17) | 0.79 |
| VA | 12,372 | 109,694 | 6.4 | 89.3 | <10 | 175 | 0.23 (0.06 - 0.61) | 0.01 | 0.26 (0.08 - 0.85) | 0.03 |
| SIDIAP | 5,142 | 2,685 | 6.3 | 5.4 | <5 | <5 | | 0.29 | | 0.30 |
| Meta-analysis | 389,250 | 1,052,708 | 276.5 | 919.7 | 374 | 2,122 | 0.69 (0.61 - 0.78) | <0.01 | 0.73 (0.59 - 0.90) | <0.01 |
| Meta-analysis (w/o USOC) | 61,305 | 171,755 | 33.7 | 110.9 | 68 | 337 | 0.75 (0.56 - 1.01) | 0.05 | 0.77 (0.57 - 1.04) | 0.10 |

##### 1.3.13 Bone fracture

**Supplementary Table 65:** Exposures, outcomes and risk estimates for Bone fracture in SGLT2I (male) (T) vs SU (male) (C) new-user cohorts under four different propensity score (PS) adjustment / time-at-risk (TAR) designs across data sources. We report total patient exposure time in 1,000 patient-year (KYr) units, uncalibrated and calibrated hazard ratios (HRs) and their 95% confidence intervals (CIs). Greyed out entries did not pass study diagnostics and are included only for completeness.

| | Patients | | Exposure (KYr) | | Outcomes | | Uncalibrated | | Calibrated | |
|---|---|---|---|---|---|---|---|---|---|---|
| | T | C | T | C | T | C | HR (95% CI) | $p$ | HR (95% CI) | $p$ |
| **PS-matching / on-treatment TAR** | | | | | | | | | | |
| CCAE | 19,098 | 43,539 | 16.0 | 35.9 | 249 | 567 | 1.11 (0.89 - 1.38) | 0.34 | 1.13 (0.91 - 1.40) | 0.30 |
| DAG | 4,918 | 876 | 4.0 | 0.8 | 56 | 16 | 0.80 (0.38 - 1.76) | 0.56 | 1.49 (0.69 - 3.21) | 0.35 |
| ODOD | 14,591 | 45,502 | 10.3 | 40.7 | 197 | 930 | 1.01 (0.81 - 1.25) | 0.96 | 0.99 (0.74 - 1.32) | 0.89 |
| MDCR | 1,640 | 7,527 | 1.4 | 7.0 | 33 | 185 | 0.98 (0.56 - 1.65) | 0.94 | 0.99 (0.58 - 1.69) | 0.88 |
| MDCD | 1,290 | 6,310 | 0.8 | 4.3 | 35 | 139 | 1.48 (0.87 - 2.45) | 0.14 | 1.32 (0.78 - 2.21) | 0.33 |
| Open Claims | 271,263 | 798,298 | 240.8 | 855.6 | 2,620 | 12,263 | 0.91 (0.85 - 0.96) | <0.01 | 0.96 (0.80 - 1.17) | 0.72 |
| OEHR | 17,159 | 68,847 | 5.5 | 38.0 | 64 | 636 | 1.17 (0.84 - 1.63) | 0.35 | 1.19 (0.85 - 1.67) | 0.33 |
| VA | 0 | 0 | 0.0 | 0.0 | 0 | 0 | | | | |
| SIDIAP | 4,092 | 1,110 | 5.9 | 2.0 | 51 | 23 | 0.63 (0.31 - 1.26) | 0.19 | 0.59 (0.29 - 1.18) | 0.15 |
| Meta-analysis | 334,051 | 972,009 | 284.6 | 984.3 | 3,305 | 14,759 | 0.99 (0.89 - 1.12) | 0.92 | 1.02 (0.83 - 1.26) | 0.82 |
| Meta-analysis (w/o USOC) | 62,788 | 173,711 | 43.8 | 128.7 | 685 | 2,496 | 1.06 (0.94 - 1.21) | 0.33 | 1.06 (0.92 - 1.22) | 0.50 |
| **PS-stratification / on-treatment TAR** | | | | | | | | | | |
| CCAE | 22,298 | 44,128 | 18.2 | 36.2 | 296 | 574 | 1.10 (0.94 - 1.29) | 0.21 | 1.14 (0.97 - 1.34) | 0.12 |
| DAG | 5,113 | 1,271 | 4.2 | 1.3 | 56 | 26 | 0.91 (0.50 - 1.68) | 0.76 | 1.19 (0.65 - 2.18) | 0.58 |
| ODOD | 16,194 | 45,989 | 11.1 | 41.2 | 211 | 946 | 0.96 (0.81 - 1.13) | 0.60 | 0.94 (0.75 - 1.18) | 0.63 |
| MDCR | 1,967 | 7,745 | 1.6 | 7.1 | 38 | 194 | 1.01 (0.67 - 1.50) | 0.95 | 0.98 (0.66 - 1.47) | 0.87 |
| MDCD | 1,321 | 6,508 | 0.8 | 4.5 | 35 | 149 | 1.42 (0.94 - 2.11) | 0.09 | 1.34 (0.89 - 2.02) | 0.17 |
| Open Claims | 295,535 | 803,838 | 256.4 | 861.3 | 2,775 | 12,461 | 0.89 (0.85 - 0.93) | <0.01 | 0.95 (0.77 - 1.16) | 0.63 |
| OEHR | 18,979 | 71,259 | 6.1 | 39.6 | 70 | 681 | 0.90 (0.68 - 1.17) | 0.44 | 0.97 (0.74 - 1.26) | 0.81 |
| VA | 11,119 | 99,994 | 6.7 | 91.5 | 52 | 926 | 0.77 (0.57 - 1.01) | 0.07 | 0.88 (0.56 - 1.38) | 0.59 |
| SIDIAP | 4,634 | 2,478 | 6.3 | 5.7 | 55 | 62 | 0.67 (0.40 - 1.13) | 0.13 | 0.62 (0.37 - 1.04) | 0.08 |
| Meta-analysis | 353,006 | 965,214 | 291.7 | 978.3 | 3,352 | 14,662 | 0.95 (0.85 - 1.06) | 0.37 | 0.99 (0.81 - 1.21) | 0.90 |
| Meta-analysis (w/o USOC) | 57,471 | 161,376 | 35.4 | 117.0 | 577 | 2,201 | 1.01 (0.90 - 1.13) | 0.89 | 1.03 (0.89 - 1.19) | 0.74 |
| **PS-matching / on-treatment TAR without escalation** | | | | | | | | | | |
| CCAE | 19,098 | 43,539 | 13.9 | 31.2 | 212 | 485 | 1.12 (0.89 - 1.40) | 0.35 | 1.15 (0.91 - 1.45) | 0.26 |
| DAG | 4,918 | 876 | 3.7 | 0.8 | 52 | 16 | 0.79 (0.38 - 1.76) | 0.56 | 1.56 (0.72 - 3.37) | 0.29 |
| ODOD | 14,591 | 45,502 | 9.1 | 36.5 | 174 | 838 | 1.00 (0.79 - 1.26) | 0.99 | 0.98 (0.75 - 1.28) | 0.87 |
| MDCR | 1,640 | 7,527 | 1.2 | 6.4 | 28 | 175 | 1.00 (0.57 - 1.71) | 0.99 | 0.99 (0.57 - 1.73) | 0.87 |
| MDCD | 1,290 | 6,310 | 0.7 | 3.6 | 31 | 115 | 1.76 (1.01 - 2.99) | 0.04 | 1.55 (0.90 - 2.68) | 0.13 |
| Open Claims | 271,263 | 798,298 | 204.4 | 727.4 | 2,196 | 10,360 | 0.88 (0.82 - 0.94) | <0.01 | 0.94 (0.78 - 1.14) | 0.56 |
| OEHR | 17,159 | 68,847 | 4.8 | 33.1 | 57 | 539 | 1.18 (0.82 - 1.67) | 0.35 | 1.23 (0.87 - 1.76) | 0.26 |
| VA | 0 | 0 | 0.0 | 0.0 | 0 | 0 | | | | |
| SIDIAP | 4,092 | 1,110 | 5.2 | 1.8 | 46 | 19 | 0.73 (0.34 - 1.61) | 0.43 | 0.72 (0.33 - 1.57) | 0.43 |
| Meta-analysis | 334,051 | 972,009 | 243.1 | 840.7 | 2,796 | 12,547 | 1.01 (0.88 - 1.16) | 0.88 | 1.05 (0.85 - 1.30) | 0.67 |
| Meta-analysis (w/o USOC) | 62,788 | 173,711 | 38.7 | 113.3 | 600 | 2,187 | 1.08 (0.95 - 1.24) | 0.24 | 1.09 (0.94 - 1.26) | 0.32 |
| **PS-stratification / on-treatment TAR without escalation** | | | | | | | | | | |
| CCAE | 22,298 | 44,128 | 15.9 | 31.5 | 254 | 492 | 1.09 (0.92 - 1.29) | 0.31 | 1.14 (0.97 - 1.36) | 0.14 |
| DAG | 5,113 | 1,271 | 3.8 | 1.1 | 52 | 26 | 0.87 (0.48 - 1.62) | 0.65 | 1.21 (0.66 - 2.24) | 0.56 |
| ODOD | 16,194 | 45,989 | 9.8 | 36.9 | 186 | 853 | 0.96 (0.80 - 1.14) | 0.65 | 0.93 (0.75 - 1.16) | 0.59 |
| MDCR | 1,967 | 7,745 | 1.4 | 6.5 | 32 | 183 | 0.91 (0.58 - 1.37) | 0.65 | 0.87 (0.57 - 1.33) | 0.54 |
| MDCD | 1,321 | 6,508 | 0.7 | 3.8 | 31 | 123 | 1.62 (1.03 - 2.47) | 0.03 | 1.52 (0.98 - 2.36) | 0.07 |
| Open Claims | 295,535 | 803,838 | 218.3 | 732.3 | 2,332 | 10,533 | 0.89 (0.84 - 0.93) | <0.01 | 0.95 (0.78 - 1.17) | 0.66 |
| OEHR | 18,979 | 71,259 | 5.3 | 34.4 | 62 | 577 | 0.93 (0.69 - 1.23) | 0.63 | 1.00 (0.75 - 1.33) | 0.88 |
| VA | 11,119 | 99,994 | 5.8 | 81.2 | 46 | 816 | 0.79 (0.58 - 1.05) | 0.13 | 0.89 (0.64 - 1.24) | 0.59 |
| SIDIAP | 4,634 | 2,478 | 5.6 | 4.9 | 50 | 55 | 0.67 (0.39 - 1.15) | 0.15 | 0.65 (0.38 - 1.13) | 0.15 |
| Meta-analysis | 353,006 | 965,214 | 249.4 | 835.1 | 2,834 | 12,455 | 0.95 (0.85 - 1.05) | 0.33 | 0.99 (0.81 - 1.22) | 0.91 |
| Meta-analysis (w/o USOC) | 57,471 | 161,376 | 31.1 | 102.8 | 502 | 1,922 | 1.01 (0.90 - 1.13) | 0.84 | 1.03 (0.90 - 1.19) | 0.70 |

##### 1.3.14 Diabetic ketoacidosis

**Supplementary Table 66:** Exposures, outcomes and risk estimates for Diabetic ketoacidosis in SGLT2I (male) (T) vs SU (male) (C) new-user cohorts under four different propensity score (PS) adjustment / time-at-risk (TAR) designs across data sources. We report total patient exposure time in 1,000 patient-year (KYr) units, uncalibrated and calibrated hazard ratios (HRs) and their 95% confidence intervals (CIs). Greyed out entries did not pass study diagnostics and are included only for completeness.

| | Patients | | Exposure (KYr) | | Outcomes | | Uncalibrated | | Calibrated | |
|---|---|---|---|---|---|---|---|---|---|---|
| | T | C | T | C | T | C | HR (95% CI) | $p$ | HR (95% CI) | $p$ |
| **PS-matching / on-treatment TAR** | | | | | | | | | | |
| CCAE | 20,611 | 46,816 | 17.6 | 39.2 | 38 | 28 | 3.20 (1.56 - 6.91) | <0.01 | 3.25 (1.54 - 6.84) | <0.01 |
| DAG | 5,471 | 973 | 4.6 | 0.9 | 0 | 0 | | | | |
| ODOD | 15,705 | 48,826 | 11.2 | 44.5 | 22 | 65 | 1.79 (0.91 - 3.44) | 0.09 | 1.76 (0.88 - 3.51) | 0.12 |
| MDCR | 1,776 | 8,308 | 1.5 | 7.8 | 0 | 11 | | | | |
| MDCD | 1,456 | 7,196 | 0.9 | 5.0 | <5 | 30 | 1.27 (0.34 - 3.83) | 0.70 | 1.13 (0.33 - 3.79) | 0.85 |
| Open Claims | 304,375 | 886,843 | 271.2 | 960.3 | 287 | 771 | 1.78 (1.48 - 2.15) | <0.01 | 1.90 (1.46 - 2.47) | <0.01 |
| OEHR | 18,273 | 73,770 | 5.8 | 41.4 | 12 | 50 | 3.22 (1.31 - 7.76) | <0.01 | 3.27 (1.34 - 7.98) | <0.01 |
| VA | 0 | 0 | 0.0 | 0.0 | 0 | 0 | | | | |
| SIDIAP | 4,555 | 1,220 | 6.6 | 2.2 | 0 | <5 | | | | |
| Meta-analysis | 360,420 | 1,063,451 | 306.7 | 1090.5 | <364 | 944 | 1.92 (1.54 - 2.38) | <0.01 | 1.97 (1.49 - 2.61) | <0.01 |
| Meta-analysis (w/o USOC) | 56,045 | 176,608 | 35.5 | 130.1 | <77 | 173 | 2.32 (1.54 - 3.49) | <0.01 | 2.31 (1.53 - 3.49) | <0.01 |
| **PS-stratification / on-treatment TAR** | | | | | | | | | | |
| CCAE | 24,108 | 47,460 | 20.1 | 39.6 | 40 | 29 | 2.79 (1.66 - 4.75) | <0.01 | 2.88 (1.71 - 4.87) | <0.01 |
| DAG | 5,702 | 1,403 | 4.7 | 1.4 | 0 | 0 | | | | |
| ODOD | 17,459 | 49,485 | 12.1 | 45.1 | 23 | 66 | 1.43 (0.81 - 2.43) | 0.20 | 1.40 (0.79 - 2.47) | 0.26 |
| MDCR | 2,135 | 8,594 | 1.7 | 8.1 | <5 | 11 | | | | |
| MDCD | 1,499 | 7,439 | 0.9 | 5.2 | <5 | 32 | 0.90 (0.25 - 2.55) | 0.86 | 0.85 (0.27 - 2.74) | 0.79 |
| Open Claims | 332,197 | 892,383 | 289.1 | 966.1 | 307 | 783 | 1.59 (1.37 - 1.84) | <0.01 | 1.70 (1.33 - 2.18) | <0.01 |
| OEHR | 20,218 | 76,232 | 6.5 | 43.1 | 14 | 52 | 2.10 (1.02 - 4.12) | 0.04 | 2.26 (1.13 - 4.53) | 0.02 |
| VA | 12,523 | 111,064 | 7.5 | 102.3 | <10 | 58 | 1.57 (0.69 - 3.12) | 0.24 | 1.79 (0.78 - 4.12) | 0.18 |
| SIDIAP | 5,168 | 2,692 | 7.1 | 6.3 | 0 | <5 | | | | |
| Meta-analysis | 393,982 | 1,065,560 | 327.8 | 1093.9 | 384 | 930 | 1.79 (1.37 - 2.35) | <0.01 | 1.86 (1.35 - 2.57) | <0.01 |
| Meta-analysis (w/o USOC) | 61,785 | 173,177 | 38.7 | 127.8 | 77 | 147 | 2.04 (1.35 - 3.07) | <0.01 | 2.07 (1.36 - 3.16) | <0.01 |
| **PS-matching / on-treatment TAR without escalation** | | | | | | | | | | |
| CCAE | 20,611 | 46,816 | 15.3 | 34.0 | 31 | 21 | 3.39 (1.62 - 7.55) | <0.01 | 3.50 (1.62 - 7.57) | <0.01 |
| DAG | 5,471 | 973 | 4.2 | 0.8 | 0 | 0 | | | | |
| ODOD | 15,705 | 48,826 | 9.9 | 39.8 | 19 | 50 | 2.09 (1.01 - 4.22) | 0.04 | 2.05 (0.99 - 4.24) | 0.06 |
| MDCR | 1,776 | 8,308 | 1.4 | 7.2 | 0 | 9 | | | | |
| MDCD | 1,456 | 7,196 | 0.7 | 4.2 | <5 | 23 | 1.06 (0.23 - 3.56) | 0.93 | 0.94 (0.24 - 3.67) | 0.92 |
| Open Claims | 304,375 | 886,843 | 229.9 | 814.7 | 227 | 594 | 1.72 (1.40 - 2.12) | <0.01 | 1.85 (1.41 - 2.43) | <0.01 |
| OEHR | 18,273 | 73,770 | 5.1 | 35.9 | 11 | 29 | 4.29 (1.59 - 11.6) | <0.01 | 4.48 (1.66 - 12.1) | <0.01 |
| VA | 0 | 0 | 0.0 | 0.0 | 0 | 0 | | | | |
| SIDIAP | 4,555 | 1,220 | 5.9 | 2.0 | 0 | <5 | | | | |
| Meta-analysis | 358,964 | 1,056,255 | 260.2 | 924.3 | 288 | 694 | 2.30 (1.52 - 3.50) | <0.01 | 2.39 (1.53 - 3.73) | <0.01 |
| Meta-analysis (w/o USOC) | 54,589 | 169,412 | 30.4 | 109.7 | 61 | 100 | 2.91 (1.83 - 4.63) | <0.01 | 2.92 (1.83 - 4.66) | <0.01 |
| **PS-stratification / on-treatment TAR without escalation** | | | | | | | | | | |
| CCAE | 24,108 | 47,460 | 17.5 | 34.3 | 33 | 22 | 3.12 (1.74 - 5.74) | <0.01 | 3.28 (1.80 - 5.96) | <0.01 |
| DAG | 5,702 | 1,403 | 4.3 | 1.3 | 0 | 0 | | | | |
| ODOD | 17,459 | 49,485 | 10.8 | 40.4 | 20 | 51 | 1.57 (0.85 - 2.82) | 0.14 | 1.52 (0.82 - 2.82) | 0.19 |
| MDCR | 2,135 | 8,594 | 1.5 | 7.4 | 0 | 9 | | | | |
| MDCD | 1,499 | 7,439 | 0.8 | 4.3 | <5 | 25 | 0.87 (0.19 - 2.78) | 0.83 | 0.81 (0.21 - 3.10) | 0.77 |
| Open Claims | 332,197 | 892,383 | 245.8 | 819.7 | 243 | 603 | 1.59 (1.34 - 1.88) | <0.01 | 1.71 (1.32 - 2.23) | <0.01 |
| OEHR | 20,218 | 76,232 | 5.7 | 37.3 | 13 | 30 | 3.20 (1.42 - 6.98) | <0.01 | 3.43 (1.54 - 7.60) | <0.01 |
| VA | 12,523 | 111,064 | 6.5 | 90.5 | <10 | 38 | 2.02 (0.82 - 4.28) | 0.09 | 2.28 (0.99 - 5.25) | 0.06 |
| SIDIAP | 5,168 | 2,692 | 6.3 | 5.4 | 0 | <5 | | | | |
| Meta-analysis | 393,982 | 1,065,560 | 279.8 | 931.7 | 309 | 706 | 2.04 (1.39 - 3.00) | <0.01 | 2.14 (1.41 - 3.26) | <0.01 |
| Meta-analysis (w/o USOC) | 61,785 | 173,177 | 34.0 | 112.0 | 66 | 103 | 2.44 (1.51 - 3.93) | <0.01 | 2.48 (1.53 - 4.04) | <0.01 |

##### 1.3.15 Diarrhea

**Supplementary Table 67:** Exposures, outcomes and risk estimates for Diarrhea in SGLT2I (male) (T) vs SU (male) (C) new-user cohorts under four different propensity score (PS) adjustment / time-at-risk (TAR) designs across data sources. We report total patient exposure time in 1,000 patient-year (KYr) units, uncalibrated and calibrated hazard ratios (HRs) and their 95% confidence intervals (CIs). Greyed out entries did not pass study diagnostics and are included only for completeness.

| | Patients | | Exposure (KYr) | | Outcomes | | Uncalibrated | | Calibrated | |
|---|---|---|---|---|---|---|---|---|---|---|
| | T | C | T | C | T | C | HR (95% CI) | $p$ | HR (95% CI) | $p$ |
| **PS-matching / on-treatment TAR** | | | | | | | | | | |
| CCAE | 18,695 | 42,551 | 15.8 | 35.0 | 321 | 823 | 0.79 (0.66 - 0.95) | 0.01 | 0.80 (0.67 - 0.97) | 0.03 |
| DAG | 5,370 | 960 | 4.5 | 0.9 | 6 | <5 | 1.27 (0.22 - 9.73) | 0.80 | 2.38 (0.36 - 15.9) | 0.38 |
| ODOD | 14,057 | 43,197 | 9.9 | 38.3 | 265 | 1,228 | 0.90 (0.75 - 1.08) | 0.27 | 0.89 (0.68 - 1.15) | 0.43 |
| MDCR | 1,592 | 7,280 | 1.3 | 6.7 | 27 | 198 | 0.66 (0.35 - 1.19) | 0.19 | 0.67 (0.36 - 1.24) | 0.21 |
| MDCD | 1,250 | 6,122 | 0.7 | 4.2 | 34 | 190 | 0.93 (0.58 - 1.44) | 0.74 | 0.82 (0.52 - 1.30) | 0.43 |
| Open Claims | 274,570 | 806,193 | 243.1 | 861.5 | 2,783 | 12,671 | 0.83 (0.78 - 0.88) | <0.01 | 0.88 (0.73 - 1.07) | 0.21 |
| OEHR | 16,714 | 67,327 | 5.2 | 36.4 | 122 | 1,112 | 0.92 (0.72 - 1.16) | 0.49 | 0.93 (0.73 - 1.19) | 0.62 |
| VA | 0 | 0 | 0.0 | 0.0 | 0 | 0 | | | | |
| SIDIAP | 4,396 | 1,173 | 6.4 | 2.1 | 39 | 20 | 0.34 (0.15 - 0.74) | <0.01 | 0.32 (0.14 - 0.70) | <0.01 |
| Meta-analysis | 331,274 | 973,843 | 282.4 | 984.2 | 3,591 | 16,242 | 0.83 (0.79 - 0.87) | <0.01 | 0.86 (0.71 - 1.03) | 0.09 |
| Meta-analysis (w/o USOC) | 56,704 | 167,650 | 39.4 | 122.7 | 808 | 3,571 | 0.83 (0.72 - 0.96) | 0.01 | 0.83 (0.70 - 0.98) | 0.05 |
| **PS-stratification / on-treatment TAR** | | | | | | | | | | |
| CCAE | 21,758 | 43,117 | 17.9 | 35.4 | 367 | 829 | 0.84 (0.73 - 0.96) | <0.01 | 0.87 (0.76 - 0.99) | 0.05 |
| DAG | 5,600 | 1,385 | 4.6 | 1.4 | 6 | <5 | 1.22 (0.21 - 7.62) | 0.83 | 1.60 (0.27 - 9.58) | 0.61 |
| ODOD | 15,599 | 44,007 | 10.6 | 39.1 | 286 | 1,274 | 0.88 (0.76 - 1.02) | 0.08 | 0.86 (0.70 - 1.07) | 0.25 |
| MDCR | 1,906 | 7,505 | 1.5 | 6.9 | 30 | 207 | 0.72 (0.46 - 1.10) | 0.14 | 0.70 (0.45 - 1.08) | 0.13 |
| MDCD | 1,283 | 6,366 | 0.8 | 4.3 | 34 | 198 | 0.99 (0.65 - 1.44) | 0.94 | 0.93 (0.63 - 1.39) | 0.74 |
| Open Claims | 299,156 | 811,534 | 258.9 | 866.9 | 3,000 | 12,838 | 0.86 (0.82 - 0.89) | <0.01 | 0.91 (0.75 - 1.12) | 0.40 |
| OEHR | 18,506 | 69,602 | 5.8 | 37.8 | 134 | 1,196 | 0.84 (0.69 - 1.02) | 0.09 | 0.91 (0.74 - 1.10) | 0.37 |
| VA | 10,635 | 96,741 | 6.4 | 87.3 | 107 | 1,717 | 0.82 (0.67 - 1.00) | 0.05 | 0.94 (0.62 - 1.41) | 0.77 |
| SIDIAP | 4,981 | 2,590 | 6.8 | 6.0 | 46 | 47 | 0.46 (0.25 - 0.84) | 0.01 | 0.42 (0.23 - 0.77) | <0.01 |
| Meta-analysis | 355,019 | 968,260 | 293.2 | 979.1 | 3,787 | 16,137 | 0.86 (0.82 - 0.89) | <0.01 | 0.89 (0.74 - 1.06) | 0.20 |
| Meta-analysis (w/o USOC) | 55,863 | 156,726 | 34.3 | 112.2 | 787 | 3,299 | 0.86 (0.78 - 0.93) | <0.01 | 0.87 (0.77 - 0.98) | 0.06 |
| **PS-matching / on-treatment TAR without escalation** | | | | | | | | | | |
| CCAE | 18,695 | 42,551 | 13.8 | 30.5 | 266 | 709 | 0.79 (0.65 - 0.96) | 0.02 | 0.82 (0.67 - 1.00) | 0.07 |
| DAG | 5,370 | 960 | 4.1 | 0.8 | 6 | <5 | 1.28 (0.22 - 9.80) | 0.80 | 2.52 (0.38 - 16.7) | 0.34 |
| ODOD | 14,057 | 43,197 | 8.8 | 34.4 | 225 | 1,088 | 0.93 (0.77 - 1.13) | 0.50 | 0.92 (0.73 - 1.16) | 0.54 |
| MDCR | 1,592 | 7,280 | 1.2 | 6.2 | 24 | 185 | 0.73 (0.38 - 1.31) | 0.31 | 0.72 (0.38 - 1.34) | 0.32 |
| MDCD | 1,250 | 6,122 | 0.6 | 3.5 | 29 | 158 | 0.99 (0.60 - 1.57) | 0.96 | 0.87 (0.54 - 1.41) | 0.59 |
| Open Claims | 274,570 | 806,193 | 206.8 | 734.1 | 2,284 | 10,660 | 0.80 (0.75 - 0.85) | <0.01 | 0.86 (0.71 - 1.03) | 0.12 |
| OEHR | 16,714 | 67,327 | 4.6 | 31.9 | 106 | 881 | 0.95 (0.73 - 1.22) | 0.69 | 0.99 (0.76 - 1.29) | 0.83 |
| VA | 0 | 0 | 0.0 | 0.0 | 0 | 0 | | | | |
| SIDIAP | 4,396 | 1,173 | 5.7 | 1.9 | 34 | 17 | 0.39 (0.16 - 0.93) | 0.03 | 0.39 (0.16 - 0.92) | 0.04 |
| Meta-analysis | 331,274 | 973,843 | 241.6 | 842.4 | 2,968 | 13,698 | 0.82 (0.76 - 0.89) | <0.01 | 0.85 (0.72 - 1.02) | 0.08 |
| Meta-analysis (w/o USOC) | 56,704 | 167,650 | 34.7 | 108.4 | 684 | 3,038 | 0.87 (0.76 - 0.98) | 0.03 | 0.87 (0.75 - 1.00) | 0.09 |
| **PS-stratification / on-treatment TAR without escalation** | | | | | | | | | | |
| CCAE | 21,758 | 43,117 | 15.7 | 30.8 | 307 | 715 | 0.80 (0.69 - 0.92) | <0.01 | 0.84 (0.72 - 0.97) | 0.02 |
| DAG | 5,600 | 1,385 | 4.3 | 1.2 | 6 | <5 | 1.23 (0.21 - 7.74) | 0.82 | 1.71 (0.28 - 10.4) | 0.56 |
| ODOD | 15,599 | 44,007 | 9.5 | 35.1 | 244 | 1,130 | 0.86 (0.73 - 1.00) | 0.05 | 0.83 (0.68 - 1.01) | 0.12 |
| MDCR | 1,906 | 7,505 | 1.3 | 6.3 | 27 | 194 | 0.74 (0.46 - 1.16) | 0.21 | 0.71 (0.45 - 1.13) | 0.17 |
| MDCD | 1,283 | 6,366 | 0.6 | 3.7 | 29 | 166 | 1.01 (0.64 - 1.52) | 0.98 | 0.95 (0.61 - 1.46) | 0.80 |
| Open Claims | 299,156 | 811,534 | 221.0 | 738.8 | 2,473 | 10,811 | 0.84 (0.80 - 0.88) | <0.01 | 0.91 (0.74 - 1.12) | 0.37 |
| OEHR | 18,506 | 69,602 | 5.2 | 33.1 | 117 | 945 | 0.90 (0.72 - 1.10) | 0.32 | 0.96 (0.78 - 1.19) | 0.73 |
| VA | 10,635 | 96,741 | 5.5 | 77.8 | 95 | 1,496 | 0.87 (0.70 - 1.06) | 0.18 | 0.98 (0.76 - 1.25) | 0.85 |
| SIDIAP | 4,981 | 2,590 | 6.1 | 5.1 | 41 | 41 | 0.40 (0.20 - 0.77) | <0.01 | 0.39 (0.20 - 0.76) | <0.01 |
| Meta-analysis | 355,019 | 968,260 | 251.3 | 837.8 | 3,141 | 13,601 | 0.84 (0.81 - 0.88) | <0.01 | 0.88 (0.74 - 1.05) | 0.17 |
| Meta-analysis (w/o USOC) | 55,863 | 156,726 | 30.3 | 99.0 | 668 | 2,790 | 0.84 (0.76 - 0.92) | <0.01 | 0.86 (0.75 - 0.97) | 0.04 |

##### 1.3.16 Genitourinary infection

**Supplementary Table 68:** Exposures, outcomes and risk estimates for Genitourinary infection in SGLT2I (male) (T) vs SU (male) (C) new-user cohorts under four different propensity score (PS) adjustment / time-at-risk (TAR) designs across data sources. We report total patient exposure time in 1,000 patient-year (KYr) units, uncalibrated and calibrated hazard ratios (HRs) and their 95% confidence intervals (CIs). Greyed out entries did not pass study diagnostics and are included only for completeness.

| | Patients | | Exposure (KYr) | | Outcomes | | Uncalibrated | | Calibrated | |
|---|---|---|---|---|---|---|---|---|---|---|
| | T | C | T | C | T | C | HR (95% CI) | $p$ | HR (95% CI) | $p$ |
| **PS-matching / on-treatment TAR** | | | | | | | | | | |
| CCAE | 18,933 | 42,878 | 16.0 | 35.3 | 242 | 712 | 0.82 (0.67 - 1.01) | 0.06 | 0.83 (0.68 - 1.03) | 0.10 |
| DAG | 4,678 | 844 | 3.9 | 0.8 | 93 | 16 | 1.27 (0.66 - 2.63) | 0.49 | 2.39 (1.19 - 4.78) | 0.02 |
| ODOD | 14,139 | 42,339 | 10.0 | 37.3 | 266 | 1,516 | 0.96 (0.80 - 1.14) | 0.64 | 0.94 (0.73 - 1.22) | 0.69 |
| MDCR | 1,575 | 7,087 | 1.3 | 6.5 | 37 | 287 | 0.76 (0.47 - 1.19) | 0.26 | 0.77 (0.48 - 1.22) | 0.30 |
| MDCD | 1,333 | 6,465 | 0.8 | 4.4 | 15 | 126 | 0.69 (0.33 - 1.34) | 0.31 | 0.62 (0.31 - 1.24) | 0.19 |
| Open Claims | 272,767 | 789,830 | 244.0 | 841.1 | 2,747 | 16,133 | 0.79 (0.75 - 0.84) | <0.01 | 0.84 (0.70 - 1.02) | 0.09 |
| OEHR | 17,285 | 69,322 | 5.5 | 38.0 | 97 | 895 | 1.07 (0.81 - 1.40) | 0.61 | 1.09 (0.83 - 1.43) | 0.57 |
| VA | 0 | 0 | 0.0 | 0.0 | 0 | 0 | | | | |
| SIDIAP | 3,966 | 1,104 | 5.8 | 2.0 | 123 | 39 | 1.04 (0.63 - 1.76) | 0.89 | 0.98 (0.58 - 1.64) | 0.72 |
| Meta-analysis | 334,676 | 959,869 | 287.3 | 965.3 | 3,620 | 19,724 | 0.88 (0.78 - 0.98) | 0.02 | 0.90 (0.73 - 1.11) | 0.33 |
| Meta-analysis (w/o USOC) | 61,909 | 170,039 | 43.3 | 124.2 | 873 | 3,591 | 0.93 (0.83 - 1.03) | 0.17 | 0.92 (0.81 - 1.05) | 0.31 |
| **PS-stratification / on-treatment TAR** | | | | | | | | | | |
| CCAE | 22,104 | 43,443 | 18.3 | 35.6 | 274 | 724 | 0.74 (0.64 - 0.86) | <0.01 | 0.77 (0.66 - 0.90) | <0.01 |
| DAG | 4,928 | 1,221 | 4.0 | 1.2 | 96 | 20 | 1.45 (0.82 - 2.69) | 0.22 | 1.90 (1.05 - 3.45) | 0.04 |
| ODOD | 15,706 | 42,920 | 10.8 | 37.8 | 287 | 1,564 | 0.87 (0.75 - 1.00) | 0.05 | 0.85 (0.69 - 1.05) | 0.20 |
| MDCR | 1,870 | 7,208 | 1.5 | 6.6 | 43 | 293 | 0.82 (0.56 - 1.16) | 0.27 | 0.79 (0.55 - 1.14) | 0.24 |
| MDCD | 1,377 | 6,646 | 0.8 | 4.6 | 15 | 131 | 0.66 (0.36 - 1.13) | 0.16 | 0.63 (0.35 - 1.11) | 0.12 |
| Open Claims | 297,034 | 794,829 | 259.6 | 846.1 | 2,993 | 16,427 | 0.76 (0.73 - 0.79) | <0.01 | 0.81 (0.66 - 0.99) | 0.04 |
| OEHR | 19,171 | 71,370 | 6.1 | 39.3 | 109 | 961 | 1.01 (0.81 - 1.25) | 0.92 | 1.08 (0.87 - 1.35) | 0.50 |
| VA | 11,310 | 99,225 | 6.8 | 89.8 | 68 | 1,456 | 0.62 (0.48 - 0.78) | <0.01 | 0.70 (0.46 - 1.08) | 0.14 |
| SIDIAP | 4,522 | 2,420 | 6.3 | 5.5 | 131 | 111 | 1.03 (0.71 - 1.49) | 0.88 | 0.95 (0.65 - 1.37) | 0.70 |
| Meta-analysis | 354,015 | 952,562 | 294.8 | 958.8 | 3,663 | 19,676 | 0.82 (0.73 - 0.92) | <0.01 | 0.85 (0.69 - 1.05) | 0.13 |
| Meta-analysis (w/o USOC) | 56,981 | 157,733 | 35.1 | 112.7 | 670 | 3,249 | 0.85 (0.73 - 1.00) | 0.06 | 0.87 (0.73 - 1.04) | 0.17 |
| **PS-matching / on-treatment TAR without escalation** | | | | | | | | | | |
| CCAE | 18,933 | 42,878 | 14.0 | 30.6 | 205 | 627 | 0.84 (0.67 - 1.04) | 0.11 | 0.86 (0.69 - 1.08) | 0.22 |
| DAG | 4,678 | 844 | 3.6 | 0.7 | 86 | 15 | 1.41 (0.72 - 3.00) | 0.35 | 2.77 (1.35 - 5.68) | <0.01 |
| ODOD | 14,139 | 42,339 | 8.9 | 33.4 | 228 | 1,374 | 0.93 (0.77 - 1.11) | 0.42 | 0.91 (0.72 - 1.14) | 0.48 |
| MDCR | 1,575 | 7,087 | 1.2 | 6.0 | 34 | 263 | 0.86 (0.53 - 1.34) | 0.52 | 0.85 (0.53 - 1.36) | 0.53 |
| MDCD | 1,333 | 6,465 | 0.7 | 3.7 | 12 | 104 | 0.83 (0.36 - 1.75) | 0.65 | 0.74 (0.33 - 1.62) | 0.45 |
| Open Claims | 272,767 | 789,830 | 206.9 | 714.5 | 2,327 | 13,847 | 0.79 (0.74 - 0.84) | <0.01 | 0.85 (0.70 - 1.02) | 0.09 |
| OEHR | 17,285 | 69,322 | 4.8 | 33.1 | 84 | 712 | 1.17 (0.86 - 1.56) | 0.30 | 1.22 (0.91 - 1.65) | 0.21 |
| VA | 0 | 0 | 0.0 | 0.0 | 0 | 0 | | | | |
| SIDIAP | 3,966 | 1,104 | 5.2 | 1.7 | 107 | 35 | 0.98 (0.58 - 1.70) | 0.94 | 0.96 (0.56 - 1.66) | 0.70 |
| Meta-analysis | 334,676 | 959,869 | 245.2 | 823.8 | 3,083 | 16,977 | 0.89 (0.79 - 1.01) | 0.08 | 0.93 (0.75 - 1.13) | 0.45 |
| Meta-analysis (w/o USOC) | 61,909 | 170,039 | 38.3 | 109.2 | 756 | 3,130 | 0.94 (0.83 - 1.06) | 0.30 | 0.94 (0.83 - 1.08) | 0.46 |
| **PS-stratification / on-treatment TAR without escalation** | | | | | | | | | | |
| CCAE | 22,104 | 43,443 | 16.0 | 30.9 | 233 | 639 | 0.70 (0.59 - 0.83) | <0.01 | 0.74 (0.62 - 0.87) | <0.01 |
| DAG | 4,928 | 1,221 | 3.7 | 1.1 | 88 | 19 | 1.40 (0.78 - 2.64) | 0.28 | 1.95 (1.06 - 3.60) | 0.04 |
| ODOD | 15,706 | 42,920 | 9.6 | 33.9 | 248 | 1,417 | 0.84 (0.73 - 0.98) | 0.03 | 0.82 (0.68 - 1.00) | 0.09 |
| MDCR | 1,870 | 7,208 | 1.3 | 6.1 | 40 | 269 | 0.85 (0.58 - 1.23) | 0.41 | 0.82 (0.56 - 1.20) | 0.33 |
| MDCD | 1,377 | 6,646 | 0.7 | 3.8 | 12 | 108 | 0.62 (0.31 - 1.13) | 0.14 | 0.58 (0.31 - 1.11) | 0.11 |
| Open Claims | 297,034 | 794,829 | 220.9 | 718.9 | 2,550 | 14,105 | 0.76 (0.73 - 0.80) | <0.01 | 0.82 (0.67 - 1.01) | 0.07 |
| OEHR | 19,171 | 71,370 | 5.4 | 34.2 | 95 | 758 | 1.10 (0.86 - 1.39) | 0.42 | 1.18 (0.93 - 1.50) | 0.20 |
| VA | 11,310 | 99,225 | 5.9 | 79.6 | 59 | 1,250 | 0.64 (0.49 - 0.83) | <0.01 | 0.73 (0.54 - 0.97) | 0.12 |
| SIDIAP | 4,522 | 2,420 | 5.6 | 4.7 | 115 | 97 | 0.97 (0.65 - 1.45) | 0.88 | 0.95 (0.63 - 1.42) | 0.73 |
| Meta-analysis | 354,015 | 952,562 | 251.9 | 818.0 | 3,126 | 16,919 | 0.82 (0.70 - 0.97) | 0.02 | 0.86 (0.68 - 1.09) | 0.22 |
| Meta-analysis (w/o USOC) | 56,981 | 157,733 | 30.9 | 99.0 | 576 | 2,814 | 0.86 (0.68 - 1.07) | 0.18 | 0.87 (0.69 - 1.11) | 0.29 |

##### 1.3.17 Hyperkalemia

**Supplementary Table 69:** Exposures, outcomes and risk estimates for Hyperkalemia in SGLT2I (male) (T) vs SU (male) (C) new-user cohorts under four different propensity score (PS) adjustment / time-at-risk (TAR) designs across data sources. We report total patient exposure time in 1,000 patient-year (KYr) units, uncalibrated and calibrated hazard ratios (HRs) and their 95% confidence intervals (CIs). Greyed out entries did not pass study diagnostics and are included only for completeness.

| | Patients | | Exposure (KYr) | | Outcomes | | Uncalibrated | | Calibrated | |
|---|---|---|---|---|---|---|---|---|---|---|
| | T | C | T | C | T | C | HR (95% CI) | $p$ | HR (95% CI) | $p$ |
| **PS-matching / on-treatment TAR** | | | | | | | | | | |
| CCAE | 20,266 | 46,044 | 17.2 | 38.2 | 115 | 367 | 0.76 (0.57 - 1.02) | 0.07 | 0.77 (0.58 - 1.04) | 0.10 |
| DAG | 5,131 | 910 | 4.3 | 0.9 | 50 | 12 | 0.72 (0.28 - 2.02) | 0.51 | 1.34 (0.50 - 3.61) | 0.57 |
| ODOD | 15,150 | 46,307 | 10.7 | 41.1 | 178 | 1,101 | 0.81 (0.65 - 1.00) | 0.06 | 0.80 (0.60 - 1.06) | 0.17 |
| MDCR | 1,729 | 7,998 | 1.5 | 7.5 | 17 | 172 | 0.81 (0.39 - 1.57) | 0.56 | 0.82 (0.40 - 1.65) | 0.59 |
| MDCD | 1,418 | 6,953 | 0.9 | 4.8 | 15 | 112 | 1.19 (0.58 - 2.28) | 0.62 | 1.06 (0.53 - 2.10) | 0.86 |
| Open Claims | 299,090 | 868,896 | 266.1 | 931.6 | 1,170 | 7,952 | 0.72 (0.66 - 0.78) | <0.01 | 0.77 (0.63 - 0.94) | 0.01 |
| OEHR | 17,953 | 72,312 | 5.7 | 39.9 | 57 | 598 | 1.02 (0.72 - 1.43) | 0.91 | 1.04 (0.73 - 1.46) | 0.83 |
| VA | 0 | 0 | 0.0 | 0.0 | 0 | 0 | | | | |
| SIDIAP | 4,355 | 1,181 | 6.3 | 2.2 | 68 | 18 | 1.54 (0.68 - 3.81) | 0.32 | 1.45 (0.61 - 3.42) | 0.41 |
| Meta-analysis | 365,092 | 1,050,601 | 312.6 | 1066.1 | 1,670 | 10,332 | 0.81 (0.71 - 0.92) | <0.01 | 0.83 (0.67 - 1.04) | 0.10 |
| Meta-analysis (w/o USOC) | 66,002 | 181,705 | 46.5 | 134.5 | 500 | 2,380 | 0.86 (0.74 - 0.99) | 0.04 | 0.86 (0.73 - 1.01) | 0.10 |
| **PS-stratification / on-treatment TAR** | | | | | | | | | | |
| CCAE | 23,691 | 46,675 | 19.6 | 38.6 | 134 | 371 | 0.75 (0.60 - 0.92) | <0.01 | 0.77 (0.62 - 0.96) | 0.02 |
| DAG | 5,339 | 1,317 | 4.4 | 1.3 | 52 | 14 | 0.84 (0.41 - 1.83) | 0.65 | 1.10 (0.52 - 2.34) | 0.75 |
| ODOD | 16,805 | 46,980 | 11.6 | 41.7 | 192 | 1,138 | 0.80 (0.68 - 0.95) | 0.01 | 0.79 (0.62 - 0.99) | 0.08 |
| MDCR | 2,069 | 8,235 | 1.7 | 7.6 | 20 | 181 | 0.73 (0.42 - 1.19) | 0.23 | 0.70 (0.42 - 1.19) | 0.21 |
| MDCD | 1,462 | 7,170 | 0.9 | 5.0 | 15 | 117 | 1.16 (0.63 - 2.00) | 0.61 | 1.10 (0.62 - 1.96) | 0.76 |
| Open Claims | 325,992 | 874,103 | 283.5 | 937.0 | 1,288 | 8,117 | 0.70 (0.65 - 0.74) | <0.01 | 0.75 (0.60 - 0.92) | <0.01 |
| OEHR | 19,863 | 74,522 | 6.3 | 41.4 | 62 | 645 | 0.82 (0.62 - 1.09) | 0.18 | 0.88 (0.66 - 1.18) | 0.42 |
| VA | 11,422 | 104,088 | 6.9 | 94.0 | 104 | 1,465 | 0.97 (0.79 - 1.18) | 0.77 | 1.10 (0.73 - 1.66) | 0.67 |
| SIDIAP | 4,910 | 2,594 | 6.7 | 6.0 | 78 | 59 | 0.95 (0.58 - 1.55) | 0.82 | 0.87 (0.53 - 1.43) | 0.59 |
| Meta-analysis | 386,351 | 1,042,280 | 320.9 | 1058.7 | 1,676 | 10,271 | 0.74 (0.67 - 0.81) | <0.01 | 0.77 (0.63 - 0.93) | <0.01 |
| Meta-analysis (w/o USOC) | 60,359 | 168,177 | 37.5 | 121.7 | 388 | 2,154 | 0.79 (0.70 - 0.89) | <0.01 | 0.80 (0.69 - 0.93) | <0.01 |
| **PS-matching / on-treatment TAR without escalation** | | | | | | | | | | |
| CCAE | 20,266 | 46,044 | 15.0 | 33.2 | 93 | 316 | 0.69 (0.49 - 0.95) | 0.03 | 0.71 (0.51 - 0.99) | 0.05 |
| DAG | 5,131 | 910 | 3.9 | 0.8 | 45 | 12 | 0.72 (0.28 - 2.06) | 0.52 | 1.42 (0.52 - 3.87) | 0.51 |
| ODOD | 15,150 | 46,307 | 9.5 | 36.9 | 161 | 977 | 0.82 (0.65 - 1.02) | 0.09 | 0.80 (0.62 - 1.04) | 0.15 |
| MDCR | 1,729 | 7,998 | 1.3 | 6.8 | 17 | 159 | 0.81 (0.37 - 1.61) | 0.58 | 0.80 (0.39 - 1.67) | 0.57 |
| MDCD | 1,418 | 6,953 | 0.7 | 4.0 | 14 | 89 | 1.26 (0.60 - 2.46) | 0.52 | 1.11 (0.55 - 2.25) | 0.77 |
| Open Claims | 299,090 | 868,896 | 225.6 | 791.1 | 975 | 6,657 | 0.71 (0.65 - 0.77) | <0.01 | 0.76 (0.62 - 0.93) | <0.01 |
| OEHR | 17,953 | 72,312 | 5.0 | 34.7 | 43 | 449 | 1.01 (0.68 - 1.48) | 0.94 | 1.06 (0.72 - 1.56) | 0.77 |
| VA | 0 | 0 | 0.0 | 0.0 | 0 | 0 | | | | |
| SIDIAP | 4,355 | 1,181 | 5.6 | 1.9 | 53 | 14 | 1.21 (0.46 - 3.54) | 0.72 | 1.19 (0.43 - 3.29) | 0.74 |
| Meta-analysis | 365,092 | 1,050,601 | 266.7 | 909.4 | 1,401 | 8,673 | 0.78 (0.68 - 0.89) | <0.01 | 0.81 (0.66 - 1.00) | 0.05 |
| Meta-analysis (w/o USOC) | 66,002 | 181,705 | 41.1 | 118.3 | 426 | 2,016 | 0.84 (0.72 - 0.98) | 0.03 | 0.84 (0.71 - 0.99) | 0.07 |
| **PS-stratification / on-treatment TAR without escalation** | | | | | | | | | | |
| CCAE | 23,691 | 46,675 | 17.1 | 33.5 | 109 | 320 | 0.69 (0.54 - 0.87) | <0.01 | 0.72 (0.57 - 0.92) | 0.01 |
| DAG | 5,339 | 1,317 | 4.0 | 1.2 | 46 | 14 | 0.75 (0.36 - 1.67) | 0.47 | 1.05 (0.49 - 2.28) | 0.77 |
| ODOD | 16,805 | 46,980 | 10.3 | 37.4 | 173 | 1,008 | 0.84 (0.70 - 1.00) | 0.05 | 0.82 (0.66 - 1.01) | 0.11 |
| MDCR | 2,069 | 8,235 | 1.5 | 7.0 | 19 | 168 | 0.83 (0.48 - 1.38) | 0.50 | 0.80 (0.47 - 1.36) | 0.42 |
| MDCD | 1,462 | 7,170 | 0.7 | 4.2 | 14 | 92 | 1.24 (0.65 - 2.19) | 0.49 | 1.16 (0.63 - 2.14) | 0.63 |
| Open Claims | 325,992 | 874,103 | 241.1 | 795.8 | 1,086 | 6,803 | 0.70 (0.65 - 0.75) | <0.01 | 0.75 (0.61 - 0.93) | 0.01 |
| OEHR | 19,863 | 74,522 | 5.6 | 36.0 | 47 | 484 | 0.77 (0.55 - 1.06) | 0.12 | 0.83 (0.60 - 1.15) | 0.28 |
| VA | 11,422 | 104,088 | 6.0 | 83.4 | 89 | 1,239 | 1.00 (0.80 - 1.24) | 0.97 | 1.13 (0.88 - 1.46) | 0.49 |
| SIDIAP | 4,910 | 2,594 | 6.0 | 5.1 | 62 | 48 | 0.86 (0.48 - 1.51) | 0.59 | 0.83 (0.47 - 1.48) | 0.56 |
| Meta-analysis | 386,351 | 1,042,280 | 274.1 | 902.7 | 1,415 | 8,615 | 0.73 (0.66 - 0.81) | <0.01 | 0.77 (0.63 - 0.94) | <0.01 |
| Meta-analysis (w/o USOC) | 60,359 | 168,177 | 33.0 | 106.9 | 329 | 1,812 | 0.78 (0.68 - 0.89) | <0.01 | 0.80 (0.68 - 0.93) | <0.01 |

##### 1.3.18 Hypoglycemia

**Supplementary Table 70:** Exposures, outcomes and risk estimates for Hypoglycemia in SGLT2I (male) (T) vs SU (male) (C) new-user cohorts under four different propensity score (PS) adjustment / time-at-risk (TAR) designs across data sources. We report total patient exposure time in 1,000 patient-year (KYr) units, uncalibrated and calibrated hazard ratios (HRs) and their 95% confidence intervals (CIs). Greyed out entries did not pass study diagnostics and are included only for completeness.

| | Patients | | Exposure (KYr) | | Outcomes | | Uncalibrated | | Calibrated | |
|---|---|---|---|---|---|---|---|---|---|---|
| | T | C | T | C | T | C | HR (95% CI) | $p$ | HR (95% CI) | $p$ |
| **PS-matching / on-treatment TAR** | | | | | | | | | | |
| CCAE | 20,386 | 46,405 | 17.4 | 38.7 | 37 | 361 | 0.22 (0.14 - 0.33) | <0.01 | 0.22 (0.15 - 0.34) | <0.01 |
| DAG | 5,443 | 969 | 4.5 | 0.9 | 5 | <5 | 0.15 (0.00 - 5.29) | 0.30 | 0.29 (0.01 - 9.92) | 0.49 |
| ODOD | 15,530 | 48,361 | 11.1 | 43.7 | 46 | 888 | 0.24 (0.16 - 0.35) | <0.01 | 0.24 (0.16 - 0.36) | <0.01 |
| MDCR | 1,756 | 8,229 | 1.5 | 7.7 | 5 | 149 | 0.32 (0.11 - 0.74) | 0.02 | 0.32 (0.12 - 0.83) | 0.02 |
| MDCD | 1,427 | 7,084 | 0.9 | 4.9 | 6 | 128 | 0.45 (0.17 - 1.01) | 0.08 | 0.40 (0.17 - 0.98) | 0.05 |
| Open Claims | 301,109 | 878,668 | 268.1 | 947.5 | 395 | 7,185 | 0.24 (0.21 - 0.27) | <0.01 | 0.25 (0.20 - 0.32) | <0.01 |
| OEHR | 18,137 | 73,263 | 5.8 | 40.8 | 15 | 616 | 0.20 (0.11 - 0.33) | <0.01 | 0.20 (0.11 - 0.35) | <0.01 |
| VA | 0 | 0 | 0.0 | 0.0 | 0 | 0 | | | | |
| SIDIAP | 4,547 | 1,220 | 6.6 | 2.2 | <5 | 10 | 0.08 (0.01 - 0.32) | <0.01 | 0.08 (0.01 - 0.39) | <0.01 |
| Meta-analysis | 358,345 | 1,062,010 | 304.7 | 1083.2 | 504 | 9,327 | 0.24 (0.21 - 0.27) | <0.01 | 0.25 (0.20 - 0.30) | <0.01 |
| Meta-analysis (w/o USOC) | 57,236 | 183,342 | 36.6 | 135.7 | 109 | 2,142 | 0.24 (0.19 - 0.31) | <0.01 | 0.24 (0.19 - 0.31) | <0.01 |
| **PS-stratification / on-treatment TAR** | | | | | | | | | | |
| CCAE | 23,833 | 47,046 | 19.8 | 39.0 | 49 | 366 | 0.24 (0.17 - 0.32) | <0.01 | 0.25 (0.18 - 0.34) | <0.01 |
| DAG | 5,675 | 1,395 | 4.7 | 1.4 | 5 | 5 | 0.45 (0.08 - 2.40) | 0.35 | 0.59 (0.11 - 3.15) | 0.54 |
| ODOD | 17,261 | 48,998 | 12.0 | 44.3 | 51 | 915 | 0.26 (0.19 - 0.35) | <0.01 | 0.26 (0.19 - 0.36) | <0.01 |
| MDCR | 2,115 | 8,517 | 1.7 | 7.9 | 5 | 157 | 0.22 (0.07 - 0.50) | <0.01 | 0.21 (0.08 - 0.55) | <0.01 |
| MDCD | 1,468 | 7,319 | 0.9 | 5.1 | 6 | 136 | 0.31 (0.12 - 0.65) | <0.01 | 0.29 (0.12 - 0.68) | <0.01 |
| Open Claims | 328,462 | 883,945 | 285.7 | 953.0 | 421 | 7,333 | 0.23 (0.21 - 0.26) | <0.01 | 0.25 (0.20 - 0.31) | <0.01 |
| OEHR | 20,063 | 75,672 | 6.4 | 42.4 | 18 | 652 | 0.20 (0.12 - 0.32) | <0.01 | 0.22 (0.13 - 0.35) | <0.01 |
| VA | 12,356 | 110,156 | 7.4 | 100.9 | 14 | 1,038 | 0.17 (0.10 - 0.28) | <0.01 | 0.19 (0.10 - 0.37) | <0.01 |
| SIDIAP | 5,159 | 2,687 | 7.1 | 6.3 | 5 | 20 | 0.10 (0.02 - 0.33) | <0.01 | 0.09 (0.02 - 0.34) | <0.01 |
| Meta-analysis | 389,619 | 1,055,661 | 323.9 | 1078.7 | 539 | 9,266 | 0.24 (0.22 - 0.26) | <0.01 | 0.25 (0.20 - 0.30) | <0.01 |
| Meta-analysis (w/o USOC) | 61,157 | 171,716 | 38.2 | 125.7 | 118 | 1,933 | 0.24 (0.20 - 0.30) | <0.01 | 0.25 (0.20 - 0.31) | <0.01 |
| **PS-matching / on-treatment TAR without escalation** | | | | | | | | | | |
| CCAE | 20,386 | 46,405 | 15.2 | 33.5 | 27 | 303 | 0.18 (0.11 - 0.29) | <0.01 | 0.19 (0.11 - 0.31) | <0.01 |
| DAG | 5,443 | 969 | 4.2 | 0.8 | 5 | <5 | 0.15 (0.00 - 5.29) | 0.30 | 0.30 (0.01 - 10.4) | 0.51 |
| ODOD | 15,530 | 48,361 | 9.8 | 39.1 | 32 | 765 | 0.22 (0.14 - 0.33) | <0.01 | 0.22 (0.14 - 0.34) | <0.01 |
| MDCR | 1,756 | 8,229 | 1.3 | 7.1 | <5 | 141 | 0.27 (0.08 - 0.68) | 0.02 | 0.27 (0.09 - 0.77) | 0.02 |
| MDCD | 1,427 | 7,084 | 0.7 | 4.1 | <5 | 100 | 0.27 (0.07 - 0.78) | 0.04 | 0.24 (0.07 - 0.84) | 0.03 |
| Open Claims | 301,109 | 878,668 | 227.3 | 804.4 | 269 | 5,922 | 0.19 (0.17 - 0.22) | <0.01 | 0.21 (0.17 - 0.26) | <0.01 |
| OEHR | 18,137 | 73,263 | 5.1 | 35.5 | 10 | 463 | 0.16 (0.08 - 0.30) | <0.01 | 0.17 (0.08 - 0.34) | <0.01 |
| VA | 0 | 0 | 0.0 | 0.0 | 0 | 0 | | | | |
| SIDIAP | 4,547 | 1,220 | 5.9 | 1.9 | <5 | 9 | 0.00 (0.00 - 0.10) | 0.16 | 0.00 (0.00 - 3613) | 0.16 |
| Meta-analysis | 358,345 | 1,062,010 | 259.4 | 923.7 | <348 | 7,694 | 0.20 (0.17 - 0.22) | <0.01 | 0.20 (0.17 - 0.25) | <0.01 |
| Meta-analysis (w/o USOC) | 57,236 | 183,342 | 32.1 | 119.3 | <79 | 1,772 | 0.20 (0.15 - 0.27) | <0.01 | 0.20 (0.15 - 0.27) | <0.01 |
| **PS-stratification / on-treatment TAR without escalation** | | | | | | | | | | |
| CCAE | 23,833 | 47,046 | 17.3 | 33.9 | 38 | 308 | 0.21 (0.14 - 0.29) | <0.01 | 0.22 (0.15 - 0.31) | <0.01 |
| DAG | 5,675 | 1,395 | 4.3 | 1.3 | 5 | 5 | 0.45 (0.08 - 2.40) | 0.35 | 0.62 (0.12 - 3.36) | 0.59 |
| ODOD | 17,261 | 48,998 | 10.6 | 39.7 | 36 | 789 | 0.22 (0.16 - 0.31) | <0.01 | 0.22 (0.15 - 0.32) | <0.01 |
| MDCR | 2,115 | 8,517 | 1.5 | 7.3 | <5 | 149 | 0.18 (0.05 - 0.45) | <0.01 | 0.17 (0.06 - 0.50) | <0.01 |
| MDCD | 1,468 | 7,319 | 0.7 | 4.3 | <5 | 106 | 0.25 (0.08 - 0.62) | 0.01 | 0.24 (0.08 - 0.68) | <0.01 |
| Open Claims | 328,462 | 883,945 | 243.0 | 809.2 | 289 | 6,052 | 0.19 (0.17 - 0.22) | <0.01 | 0.21 (0.16 - 0.26) | <0.01 |
| OEHR | 20,063 | 75,672 | 5.6 | 36.8 | 12 | 487 | 0.16 (0.09 - 0.28) | <0.01 | 0.17 (0.10 - 0.32) | <0.01 |
| VA | 12,356 | 110,156 | 6.4 | 89.4 | <10 | 842 | 0.10 (0.04 - 0.20) | <0.01 | 0.12 (0.05 - 0.25) | <0.01 |
| SIDIAP | 5,159 | 2,687 | 6.3 | 5.4 | <5 | 16 | 0.07 (0.01 - 0.29) | <0.01 | 0.07 (0.01 - 0.36) | <0.01 |
| Meta-analysis | 389,619 | 1,055,661 | 276.6 | 919.5 | 375 | 7,636 | 0.20 (0.18 - 0.22) | <0.01 | 0.21 (0.17 - 0.25) | <0.01 |
| Meta-analysis (w/o USOC) | 61,157 | 171,716 | 33.6 | 110.4 | 86 | 1,584 | 0.21 (0.17 - 0.26) | <0.01 | 0.21 (0.17 - 0.27) | <0.01 |

##### 1.3.19 Hypotension

**Supplementary Table 71:** Exposures, outcomes and risk estimates for Hypotension in SGLT2I (male) (T) vs SU (male) (C) new-user cohorts under four different propensity score (PS) adjustment / time-at-risk (TAR) designs across data sources. We report total patient exposure time in 1,000 patient-year (KYr) units, uncalibrated and calibrated hazard ratios (HRs) and their 95% confidence intervals (CIs). Greyed out entries did not pass study diagnostics and are included only for completeness.

| | Patients | | Exposure (KYr) | | Outcomes | | Uncalibrated | | Calibrated | |
|---|---|---|---|---|---|---|---|---|---|---|
| | T | C | T | C | T | C | HR (95% CI) | $p$ | HR (95% CI) | $p$ |
| **PS-matching / on-treatment TAR** | | | | | | | | | | |
| CCAE | 20,246 | 46,020 | 17.2 | 38.4 | 151 | 314 | 1.27 (0.96 - 1.67) | 0.10 | 1.28 (0.97 - 1.70) | 0.09 |
| DAG | 5,260 | 947 | 4.4 | 0.9 | 25 | 5 | 0.81 (0.22 - 3.95) | 0.77 | 1.52 (0.36 - 6.42) | 0.58 |
| ODOD | 15,100 | 46,353 | 10.7 | 41.6 | 208 | 1,056 | 1.16 (0.94 - 1.41) | 0.16 | 1.14 (0.86 - 1.50) | 0.41 |
| MDCR | 1,678 | 7,772 | 1.4 | 7.3 | 30 | 195 | 1.04 (0.60 - 1.76) | 0.88 | 1.05 (0.61 - 1.80) | 0.85 |
| MDCD | 1,375 | 6,741 | 0.8 | 4.6 | 21 | 123 | 1.16 (0.58 - 2.17) | 0.66 | 1.03 (0.53 - 1.98) | 0.88 |
| Open Claims | 295,340 | 857,867 | 263.1 | 923.3 | 1,790 | 8,573 | 1.09 (1.01 - 1.17) | 0.02 | 1.16 (0.95 - 1.41) | 0.15 |
| OEHR | 17,909 | 71,996 | 5.7 | 39.8 | 74 | 680 | 1.03 (0.75 - 1.40) | 0.84 | 1.05 (0.77 - 1.43) | 0.77 |
| VA | 0 | 0 | 0.0 | 0.0 | 0 | 0 | | | | |
| SIDIAP | 4,522 | 1,213 | 6.6 | 2.2 | 23 | <5 | 3.48 (0.59 - 66.3) | 0.30 | 3.27 (0.31 - 34.7) | 0.33 |
| Meta-analysis | 356,170 | 1,037,962 | 305.6 | 1057.3 | 2,297 | <10,946 | 1.10 (1.03 - 1.17) | <0.01 | 1.13 (0.94 - 1.37) | 0.18 |
| Meta-analysis (w/o USOC) | 60,830 | 180,095 | 42.5 | 134.0 | 507 | <2,373 | 1.15 (1.01 - 1.32) | 0.04 | 1.15 (0.98 - 1.34) | 0.12 |
| **PS-stratification / on-treatment TAR** | | | | | | | | | | |
| CCAE | 23,656 | 46,632 | 19.6 | 38.7 | 177 | 320 | 1.21 (0.98 - 1.49) | 0.08 | 1.25 (1.01 - 1.54) | 0.05 |
| DAG | 5,526 | 1,358 | 4.5 | 1.3 | 25 | 7 | 0.71 (0.24 - 2.37) | 0.55 | 0.93 (0.30 - 2.90) | 0.84 |
| ODOD | 16,736 | 46,829 | 11.6 | 42.0 | 229 | 1,089 | 1.01 (0.86 - 1.18) | 0.89 | 0.99 (0.79 - 1.24) | 0.88 |
| MDCR | 2,017 | 7,968 | 1.6 | 7.4 | 35 | 205 | 1.09 (0.70 - 1.65) | 0.69 | 1.06 (0.69 - 1.62) | 0.81 |
| MDCD | 1,415 | 6,935 | 0.8 | 4.8 | 21 | 131 | 1.20 (0.70 - 1.95) | 0.49 | 1.13 (0.68 - 1.88) | 0.65 |
| Open Claims | 321,381 | 863,294 | 279.9 | 929.0 | 1,989 | 8,819 | 1.01 (0.96 - 1.07) | 0.72 | 1.08 (0.88 - 1.33) | 0.48 |
| OEHR | 19,804 | 74,383 | 6.3 | 41.3 | 83 | 748 | 1.04 (0.80 - 1.33) | 0.78 | 1.11 (0.86 - 1.43) | 0.44 |
| VA | 11,420 | 104,433 | 6.9 | 94.8 | 152 | 1,252 | 1.68 (1.41 - 1.98) | <0.01 | 1.91 (1.29 - 2.84) | <0.01 |
| SIDIAP | 5,126 | 2,678 | 7.0 | 6.2 | 23 | 7 | 2.33 (0.71 - 8.84) | 0.19 | 2.14 (0.61 - 7.57) | 0.24 |
| Meta-analysis | 381,577 | 1,031,138 | 317.4 | 1051.1 | 2,478 | 10,976 | 1.02 (0.97 - 1.07) | 0.41 | 1.06 (0.88 - 1.27) | 0.52 |
| Meta-analysis (w/o USOC) | 60,196 | 167,844 | 37.5 | 122.1 | 489 | 2,157 | 1.07 (0.96 - 1.20) | 0.24 | 1.09 (0.95 - 1.26) | 0.28 |
| **PS-matching / on-treatment TAR without escalation** | | | | | | | | | | |
| CCAE | 20,246 | 46,020 | 15.0 | 33.3 | 123 | 255 | 1.37 (1.02 - 1.85) | 0.04 | 1.42 (1.05 - 1.92) | 0.03 |
| DAG | 5,260 | 947 | 4.0 | 0.8 | 22 | <5 | 1.08 (0.24 - 7.80) | 0.93 | 2.11 (0.37 - 12.1) | 0.41 |
| ODOD | 15,100 | 46,353 | 9.5 | 37.2 | 185 | 956 | 1.23 (0.99 - 1.52) | 0.06 | 1.20 (0.94 - 1.55) | 0.21 |
| MDCR | 1,678 | 7,772 | 1.3 | 6.7 | 24 | 173 | 1.03 (0.57 - 1.80) | 0.92 | 1.02 (0.57 - 1.82) | 0.88 |
| MDCD | 1,375 | 6,741 | 0.7 | 3.9 | 17 | 102 | 0.85 (0.37 - 1.78) | 0.69 | 0.75 (0.34 - 1.65) | 0.48 |
| Open Claims | 295,340 | 857,867 | 223.2 | 783.7 | 1,486 | 7,180 | 1.09 (1.01 - 1.17) | 0.03 | 1.17 (0.96 - 1.42) | 0.13 |
| OEHR | 17,909 | 71,996 | 5.0 | 34.7 | 56 | 474 | 1.06 (0.74 - 1.49) | 0.76 | 1.10 (0.78 - 1.57) | 0.59 |
| VA | 0 | 0 | 0.0 | 0.0 | 0 | 0 | | | | |
| SIDIAP | 4,522 | 1,213 | 5.8 | 1.9 | 20 | <5 | 2.08 (0.32 - 40.9) | 0.55 | 2.04 (0.18 - 23.1) | 0.57 |
| Meta-analysis | 356,170 | 1,037,962 | 260.6 | 901.4 | 1,911 | <9,145 | 1.12 (1.03 - 1.21) | <0.01 | 1.16 (0.97 - 1.38) | 0.11 |
| Meta-analysis (w/o USOC) | 60,830 | 180,095 | 37.3 | 117.7 | 425 | <1,965 | 1.20 (1.04 - 1.39) | 0.02 | 1.20 (1.03 - 1.41) | 0.04 |
| **PS-stratification / on-treatment TAR without escalation** | | | | | | | | | | |
| CCAE | 23,656 | 46,632 | 17.2 | 33.6 | 146 | 259 | 1.25 (0.99 - 1.57) | 0.06 | 1.31 (1.04 - 1.65) | 0.03 |
| DAG | 5,526 | 1,358 | 4.2 | 1.2 | 22 | 6 | 0.88 (0.27 - 3.30) | 0.84 | 1.23 (0.35 - 4.29) | 0.75 |
| ODOD | 16,736 | 46,829 | 10.3 | 37.6 | 204 | 986 | 1.02 (0.86 - 1.21) | 0.78 | 1.00 (0.81 - 1.23) | 0.88 |
| MDCR | 2,017 | 7,968 | 1.4 | 6.8 | 29 | 181 | 1.09 (0.67 - 1.72) | 0.71 | 1.05 (0.65 - 1.68) | 0.83 |
| MDCD | 1,415 | 6,935 | 0.7 | 4.0 | 17 | 108 | 1.11 (0.61 - 1.90) | 0.72 | 1.04 (0.59 - 1.84) | 0.86 |
| Open Claims | 321,381 | 863,294 | 238.2 | 788.6 | 1,667 | 7,409 | 1.02 (0.96 - 1.08) | 0.53 | 1.10 (0.89 - 1.35) | 0.40 |
| OEHR | 19,804 | 74,383 | 5.5 | 36.0 | 65 | 520 | 1.08 (0.80 - 1.43) | 0.62 | 1.15 (0.86 - 1.54) | 0.36 |
| VA | 11,420 | 104,433 | 5.9 | 84.1 | 129 | 1,000 | 1.80 (1.49 - 2.16) | <0.01 | 2.03 (1.62 - 2.55) | <0.01 |
| SIDIAP | 5,126 | 2,678 | 6.3 | 5.3 | 20 | 6 | 2.13 (0.61 - 8.86) | 0.27 | 2.08 (0.55 - 7.92) | 0.29 |
| Meta-analysis | 381,577 | 1,031,138 | 271.2 | 895.9 | 2,082 | 9,174 | 1.03 (0.98 - 1.09) | 0.24 | 1.08 (0.90 - 1.30) | 0.39 |
| Meta-analysis (w/o USOC) | 60,196 | 167,844 | 33.0 | 107.2 | 415 | 1,765 | 1.09 (0.97 - 1.24) | 0.15 | 1.12 (0.96 - 1.30) | 0.20 |

##### 1.3.20 Joint pain

**Supplementary Table 72:** Exposures, outcomes and risk estimates for Joint pain in SGLT2I (male) (T) vs SU (male) (C) new-user cohorts under four different propensity score (PS) adjustment / time-at-risk (TAR) designs across data sources. We report total patient exposure time in 1,000 patient-year (KYr) units, uncalibrated and calibrated hazard ratios (HRs) and their 95% confidence intervals (CIs). Greyed out entries did not pass study diagnostics and are included only for completeness.

| | Patients | | Exposure (KYr) | | Outcomes | | Uncalibrated | | Calibrated | |
|---|---|---|---|---|---|---|---|---|---|---|
| | T | C | T | C | T | C | HR (95% CI) | $p$ | HR (95% CI) | $p$ |
| **PS-matching / on-treatment TAR** | | | | | | | | | | |
| CCAE | 19,768 | 44,945 | 16.8 | 37.2 | 116 | 324 | 0.79 (0.58 - 1.07) | 0.13 | 0.80 (0.59 - 1.09) | 0.17 |
| DAG | 4,662 | 846 | 3.8 | 0.8 | 59 | 13 | 1.42 (0.55 - 4.40) | 0.51 | 2.66 (0.94 - 7.54) | 0.08 |
| ODOD | 15,016 | 46,626 | 10.7 | 42.2 | 98 | 430 | 0.88 (0.64 - 1.20) | 0.43 | 0.87 (0.60 - 1.25) | 0.48 |
| MDCR | 1,723 | 7,929 | 1.5 | 7.4 | 11 | 66 | 0.80 (0.33 - 1.76) | 0.61 | 0.81 (0.35 - 1.87) | 0.63 |
| MDCD | 1,350 | 6,763 | 0.8 | 4.7 | 9 | 62 | 0.92 (0.34 - 2.20) | 0.85 | 0.81 (0.32 - 2.06) | 0.67 |
| Open Claims | 290,971 | 853,349 | 259.2 | 920.9 | 1,062 | 4,402 | 0.88 (0.80 - 0.97) | <0.01 | 0.94 (0.76 - 1.15) | 0.54 |
| OEHR | 17,579 | 71,027 | 5.5 | 39.5 | 45 | 319 | 1.39 (0.87 - 2.17) | 0.16 | 1.41 (0.89 - 2.23) | 0.15 |
| VA | 0 | 0 | 0.0 | 0.0 | 0 | 0 | | | | |
| SIDIAP | 3,648 | 1,010 | 5.3 | 1.8 | 82 | 46 | 0.69 (0.42 - 1.14) | 0.15 | 0.65 (0.40 - 1.08) | 0.11 |
| Meta-analysis | 354,717 | 1,032,495 | 303.7 | 1054.5 | 1,482 | 5,662 | 0.88 (0.81 - 0.96) | <0.01 | 0.91 (0.75 - 1.10) | 0.33 |
| Meta-analysis (w/o USOC) | 63,746 | 179,146 | 44.5 | 133.5 | 420 | 1,260 | 0.89 (0.75 - 1.06) | 0.20 | 0.89 (0.74 - 1.07) | 0.27 |
| **PS-stratification / on-treatment TAR** | | | | | | | | | | |
| CCAE | 23,075 | 45,566 | 19.1 | 37.6 | 137 | 326 | 0.80 (0.64 - 0.99) | 0.05 | 0.83 (0.67 - 1.03) | 0.10 |
| DAG | 4,851 | 1,219 | 3.9 | 1.2 | 60 | 19 | 1.01 (0.53 - 2.01) | 0.98 | 1.32 (0.68 - 2.58) | 0.44 |
| ODOD | 16,688 | 47,302 | 11.6 | 42.9 | 103 | 435 | 0.88 (0.69 - 1.11) | 0.29 | 0.86 (0.65 - 1.15) | 0.35 |
| MDCR | 2,062 | 8,283 | 1.6 | 7.7 | 11 | 68 | 0.71 (0.34 - 1.40) | 0.35 | 0.69 (0.34 - 1.41) | 0.32 |
| MDCD | 1,397 | 6,982 | 0.8 | 4.9 | 9 | 64 | 0.87 (0.38 - 1.78) | 0.72 | 0.82 (0.38 - 1.77) | 0.63 |
| Open Claims | 317,231 | 859,008 | 276.1 | 926.8 | 1,151 | 4,442 | 0.90 (0.84 - 0.97) | <0.01 | 0.96 (0.78 - 1.19) | 0.72 |
| OEHR | 19,457 | 73,365 | 6.2 | 41.0 | 51 | 338 | 1.14 (0.82 - 1.57) | 0.42 | 1.23 (0.88 - 1.70) | 0.24 |
| VA | 10,977 | 97,215 | 6.6 | 88.9 | 46 | 581 | 1.03 (0.75 - 1.38) | 0.84 | 1.17 (0.73 - 1.87) | 0.53 |
| SIDIAP | 4,147 | 2,263 | 5.7 | 5.0 | 87 | 118 | 0.80 (0.55 - 1.17) | 0.25 | 0.74 (0.51 - 1.08) | 0.13 |
| Meta-analysis | 376,451 | 1,025,241 | 313.0 | 1048.3 | 1,442 | 5,541 | 0.90 (0.84 - 0.96) | <0.01 | 0.93 (0.77 - 1.12) | 0.46 |
| Meta-analysis (w/o USOC) | 59,220 | 166,233 | 36.9 | 121.5 | 291 | 1,099 | 0.90 (0.75 - 1.09) | 0.28 | 0.92 (0.75 - 1.13) | 0.45 |
| **PS-matching / on-treatment TAR without escalation** | | | | | | | | | | |
| CCAE | 19,768 | 44,945 | 14.6 | 32.3 | 96 | 278 | 0.71 (0.51 - 0.98) | 0.04 | 0.73 (0.53 - 1.02) | 0.07 |
| DAG | 4,662 | 846 | 3.5 | 0.7 | 53 | 13 | 1.80 (0.64 - 6.46) | 0.32 | 3.54 (1.11 - 11.3) | 0.04 |
| ODOD | 15,016 | 46,626 | 9.5 | 37.8 | 90 | 383 | 0.92 (0.66 - 1.28) | 0.64 | 0.91 (0.64 - 1.29) | 0.62 |
| MDCR | 1,723 | 7,929 | 1.3 | 6.8 | 10 | 62 | 0.76 (0.29 - 1.75) | 0.55 | 0.75 (0.31 - 1.85) | 0.54 |
| MDCD | 1,350 | 6,763 | 0.7 | 3.9 | 8 | 51 | 1.29 (0.43 - 3.45) | 0.63 | 1.14 (0.40 - 3.22) | 0.81 |
| Open Claims | 290,971 | 853,349 | 219.8 | 781.7 | 892 | 3,728 | 0.89 (0.80 - 0.98) | 0.02 | 0.95 (0.78 - 1.17) | 0.65 |
| OEHR | 17,579 | 71,027 | 4.8 | 34.3 | 39 | 267 | 1.39 (0.85 - 2.22) | 0.18 | 1.45 (0.90 - 2.35) | 0.13 |
| VA | 0 | 0 | 0.0 | 0.0 | 0 | 0 | | | | |
| SIDIAP | 3,648 | 1,010 | 4.7 | 1.5 | 72 | 38 | 0.71 (0.40 - 1.25) | 0.23 | 0.69 (0.39 - 1.23) | 0.24 |
| Meta-analysis | 354,717 | 1,032,495 | 259.0 | 899.1 | 1,260 | 4,820 | 0.89 (0.81 - 0.97) | 0.01 | 0.92 (0.77 - 1.11) | 0.38 |
| Meta-analysis (w/o USOC) | 63,746 | 179,146 | 39.2 | 117.4 | 368 | 1,092 | 0.92 (0.73 - 1.15) | 0.46 | 0.92 (0.72 - 1.17) | 0.52 |
| **PS-stratification / on-treatment TAR without escalation** | | | | | | | | | | |
| CCAE | 23,075 | 45,566 | 16.7 | 32.6 | 115 | 280 | 0.77 (0.61 - 0.97) | 0.03 | 0.81 (0.64 - 1.03) | 0.09 |
| DAG | 4,851 | 1,219 | 3.6 | 1.1 | 54 | 19 | 0.94 (0.49 - 1.89) | 0.86 | 1.31 (0.66 - 2.59) | 0.47 |
| ODOD | 16,688 | 47,302 | 10.3 | 38.4 | 95 | 387 | 0.93 (0.72 - 1.19) | 0.57 | 0.90 (0.68 - 1.20) | 0.52 |
| MDCR | 2,062 | 8,283 | 1.5 | 7.1 | 10 | 64 | 0.72 (0.32 - 1.46) | 0.39 | 0.69 (0.33 - 1.46) | 0.34 |
| MDCD | 1,397 | 6,982 | 0.7 | 4.0 | 8 | 51 | 0.94 (0.39 - 2.03) | 0.89 | 0.89 (0.39 - 2.03) | 0.78 |
| Open Claims | 317,231 | 859,008 | 234.9 | 786.8 | 972 | 3,761 | 0.89 (0.82 - 0.96) | <0.01 | 0.96 (0.77 - 1.19) | 0.70 |
| OEHR | 19,457 | 73,365 | 5.4 | 35.6 | 45 | 283 | 1.19 (0.83 - 1.66) | 0.34 | 1.27 (0.90 - 1.80) | 0.19 |
| VA | 10,977 | 97,215 | 5.7 | 78.8 | 36 | 524 | 0.93 (0.65 - 1.29) | 0.69 | 1.05 (0.73 - 1.52) | 0.83 |
| SIDIAP | 4,147 | 2,263 | 5.1 | 4.3 | 76 | 104 | 0.84 (0.56 - 1.26) | 0.41 | 0.82 (0.55 - 1.23) | 0.40 |
| Meta-analysis | 376,451 | 1,025,241 | 267.3 | 893.4 | 1,227 | 4,711 | 0.89 (0.83 - 0.96) | <0.01 | 0.93 (0.78 - 1.13) | 0.48 |
| Meta-analysis (w/o USOC) | 59,220 | 166,233 | 32.4 | 106.6 | 255 | 950 | 0.92 (0.73 - 1.16) | 0.48 | 0.94 (0.74 - 1.20) | 0.63 |

##### 1.3.21 Lower extremity amputation

**Supplementary Table 73:** Exposures, outcomes and risk estimates for Lower extremity amputation in SGLT2I (male) (T) vs SU (male) (C) new-user cohorts under four different propensity score (PS) adjustment / time-at-risk (TAR) designs across data sources. We report total patient exposure time in 1,000 patient-year (KYr) units, uncalibrated and calibrated hazard ratios (HRs) and their 95% confidence intervals (CIs). Greyed out entries did not pass study diagnostics and are included only for completeness.

| | Patients | | Exposure (KYr) | | Outcomes | | Uncalibrated | | Calibrated | |
|---|---|---|---|---|---|---|---|---|---|---|
| | T | C | T | C | T | C | HR (95% CI) | $p$ | HR (95% CI) | $p$ |
| **PS-matching / on-treatment TAR** | | | | | | | | | | |
| CCAE | 20,621 | 46,859 | 17.6 | 39.2 | 19 | 43 | 1.27 (0.54 - 2.90) | 0.57 | 1.29 (0.56 - 3.01) | 0.55 |
| DAG | 5,471 | 973 | 4.6 | 0.9 | 0 | 0 | | | | |
| ODOD | 15,700 | 48,812 | 11.2 | 44.5 | 13 | 78 | 0.70 (0.30 - 1.48) | 0.38 | 0.69 (0.31 - 1.56) | 0.38 |
| MDCR | 1,774 | 8,300 | 1.5 | 7.8 | <5 | 13 | 0.41 (0.02 - 2.85) | 0.48 | 0.41 (0.03 - 4.84) | 0.48 |
| MDCD | 1,460 | 7,201 | 0.9 | 5.1 | <5 | 11 | 1.65 (0.07 - 17.2) | 0.72 | 1.47 (0.10 - 22.3) | 0.78 |
| Open Claims | 304,425 | 886,839 | 271.2 | 959.5 | 154 | 777 | 0.93 (0.73 - 1.18) | 0.58 | 0.99 (0.73 - 1.35) | 0.94 |
| OEHR | 18,275 | 73,767 | 5.8 | 41.4 | <5 | 53 | 0.44 (0.07 - 1.59) | 0.30 | 0.44 (0.09 - 2.16) | 0.31 |
| VA | 0 | 0 | 0.0 | 0.0 | 0 | 0 | | | | |
| SIDIAP | 4,552 | 1,219 | 6.6 | 2.2 | 0 | 0 | | | | |
| Meta-analysis | 359,021 | 1,056,277 | 305.8 | 1084.5 | <191 | 951 | 0.92 (0.74 - 1.15) | 0.45 | 0.95 (0.71 - 1.25) | 0.70 |
| Meta-analysis (w/o USOC) | 54,596 | 169,438 | 34.6 | 125.0 | <37 | 174 | 0.85 (0.49 - 1.46) | 0.55 | 0.84 (0.49 - 1.46) | 0.55 |
| **PS-stratification / on-treatment TAR** | | | | | | | | | | |
| CCAE | 24,122 | 47,502 | 20.1 | 39.6 | 19 | 45 | 1.11 (0.61 - 1.97) | 0.73 | 1.15 (0.64 - 2.07) | 0.65 |
| DAG | 5,702 | 1,403 | 4.7 | 1.4 | 0 | 0 | | | | |
| ODOD | 17,448 | 49,467 | 12.1 | 45.1 | 13 | 80 | 0.74 (0.38 - 1.36) | 0.36 | 0.73 (0.38 - 1.41) | 0.35 |
| MDCR | 2,129 | 8,585 | 1.7 | 8.1 | <5 | 13 | 0.59 (0.08 - 2.52) | 0.54 | 0.57 (0.10 - 3.16) | 0.52 |
| MDCD | 1,503 | 7,445 | 0.9 | 5.2 | <5 | 11 | 2.63 (0.56 - 9.35) | 0.18 | 2.49 (0.61 - 10.2) | 0.21 |
| Open Claims | 332,242 | 892,529 | 289.1 | 965.4 | 159 | 807 | 0.98 (0.81 - 1.18) | 0.85 | 1.05 (0.80 - 1.38) | 0.73 |
| OEHR | 20,221 | 76,212 | 6.5 | 43.0 | <5 | 58 | 0.56 (0.13 - 1.64) | 0.37 | 0.60 (0.17 - 2.13) | 0.43 |
| VA | 12,506 | 110,856 | 7.5 | 102.0 | 11 | 89 | 1.73 (0.87 - 3.12) | 0.09 | 1.97 (0.95 - 4.10) | 0.08 |
| SIDIAP | 5,166 | 2,688 | 7.1 | 6.3 | 0 | <5 | | | | |
| Meta-analysis | 394,033 | 1,065,710 | 327.7 | 1093.1 | <196 | 990 | 0.96 (0.81 - 1.14) | 0.66 | 1.00 (0.78 - 1.28) | 0.93 |
| Meta-analysis (w/o USOC) | 61,791 | 173,181 | 38.7 | 127.7 | <37 | 183 | 0.88 (0.58 - 1.32) | 0.53 | 0.89 (0.59 - 1.36) | 0.60 |
| **PS-matching / on-treatment TAR without escalation** | | | | | | | | | | |
| CCAE | 20,621 | 46,859 | 15.3 | 33.9 | 14 | 33 | 1.24 (0.44 - 3.24) | 0.67 | 1.28 (0.48 - 3.47) | 0.62 |
| DAG | 5,471 | 973 | 4.2 | 0.8 | 0 | 0 | | | | |
| ODOD | 15,700 | 48,812 | 9.9 | 39.8 | 10 | 66 | 0.53 (0.19 - 1.26) | 0.19 | 0.52 (0.20 - 1.35) | 0.19 |
| MDCR | 1,774 | 8,300 | 1.4 | 7.2 | <5 | 12 | 0.56 (0.03 - 4.93) | 0.66 | 0.55 (0.04 - 7.55) | 0.66 |
| MDCD | 1,460 | 7,201 | 0.7 | 4.2 | <5 | 8 | | 0.21 | | 0.24 |
| Open Claims | 304,425 | 886,839 | 229.8 | 813.7 | 111 | 626 | 0.91 (0.69 - 1.19) | 0.51 | 0.98 (0.71 - 1.36) | 0.90 |
| OEHR | 18,275 | 73,767 | 5.1 | 35.9 | <5 | 29 | 0.99 (0.14 - 4.05) | 0.99 | 1.03 (0.19 - 5.49) | 0.96 |
| VA | 0 | 0 | 0.0 | 0.0 | 0 | 0 | | | | |
| SIDIAP | 4,552 | 1,219 | 5.9 | 2.0 | 0 | 0 | | | | |
| Meta-analysis | 359,021 | 1,056,277 | 260.2 | 923.3 | <140 | 754 | 0.90 (0.70 - 1.15) | 0.40 | 0.93 (0.69 - 1.25) | 0.63 |
| Meta-analysis (w/o USOC) | 54,596 | 169,438 | 30.3 | 109.6 | <29 | 128 | 0.82 (0.44 - 1.55) | 0.54 | 0.82 (0.44 - 1.56) | 0.55 |
| **PS-stratification / on-treatment TAR without escalation** | | | | | | | | | | |
| CCAE | 24,122 | 47,502 | 17.5 | 34.3 | 14 | 35 | 1.13 (0.56 - 2.17) | 0.73 | 1.18 (0.60 - 2.33) | 0.63 |
| DAG | 5,702 | 1,403 | 4.3 | 1.3 | 0 | 0 | | | | |
| ODOD | 17,448 | 49,467 | 10.8 | 40.3 | 10 | 67 | 0.77 (0.35 - 1.51) | 0.47 | 0.74 (0.36 - 1.55) | 0.44 |
| MDCR | 2,129 | 8,585 | 1.5 | 7.4 | <5 | 12 | 0.68 (0.09 - 3.07) | 0.67 | 0.66 (0.11 - 3.78) | 0.64 |
| MDCD | 1,503 | 7,445 | 0.8 | 4.3 | <5 | 8 | 2.99 (0.61 - 11.8) | 0.15 | 2.81 (0.64 - 12.4) | 0.17 |
| Open Claims | 332,242 | 892,529 | 245.8 | 818.9 | 116 | 654 | 0.88 (0.71 - 1.09) | 0.24 | 0.95 (0.70 - 1.27) | 0.72 |
| OEHR | 20,221 | 76,212 | 5.7 | 37.3 | <5 | 31 | 0.69 (0.10 - 2.55) | 0.64 | 0.74 (0.15 - 3.63) | 0.71 |
| VA | 12,506 | 110,856 | 6.5 | 90.3 | 11 | 71 | 2.27 (1.13 - 4.14) | 0.01 | 2.56 (1.32 - 4.97) | <0.01 |
| SIDIAP | 5,166 | 2,688 | 6.3 | 5.4 | 0 | 0 | | | | |
| Meta-analysis | 394,033 | 1,065,710 | 279.8 | 930.7 | <145 | 787 | 0.88 (0.73 - 1.08) | 0.22 | 0.93 (0.71 - 1.20) | 0.57 |
| Meta-analysis (w/o USOC) | 61,791 | 173,181 | 34.0 | 111.9 | <29 | 133 | 0.91 (0.57 - 1.47) | 0.71 | 0.93 (0.58 - 1.51) | 0.78 |

##### 1.3.22 Nausea

**Supplementary Table 74:** Exposures, outcomes and risk estimates for Nausea in SGLT2I (male) (T) vs SU (male) (C) new-user cohorts under four different propensity score (PS) adjustment / time-at-risk (TAR) designs across data sources. We report total patient exposure time in 1,000 patient-year (KYr) units, uncalibrated and calibrated hazard ratios (HRs) and their 95% confidence intervals (CIs). Greyed out entries did not pass study diagnostics and are included only for completeness.

| | Patients | | Exposure (KYr) | | Outcomes | | Uncalibrated | | Calibrated | |
|---|---|---|---|---|---|---|---|---|---|---|
| | T | C | T | C | T | C | HR (95% CI) | $p$ | HR (95% CI) | $p$ |
| **PS-matching / on-treatment TAR** | | | | | | | | | | |
| CCAE | 18,644 | 42,437 | 15.6 | 35.2 | 381 | 816 | 1.13 (0.95 - 1.35) | 0.16 | 1.15 (0.96 - 1.37) | 0.14 |
| DAG | 5,175 | 921 | 4.3 | 0.9 | 33 | 8 | 1.35 (0.50 - 4.32) | 0.59 | 2.53 (0.85 - 7.48) | 0.11 |
| ODOD | 14,037 | 43,462 | 9.9 | 39.0 | 281 | 1,284 | 0.99 (0.83 - 1.19) | 0.95 | 0.98 (0.75 - 1.27) | 0.85 |
| MDCR | 1,627 | 7,529 | 1.4 | 7.1 | 36 | 226 | 0.96 (0.59 - 1.52) | 0.87 | 0.97 (0.60 - 1.56) | 0.85 |
| MDCD | 1,157 | 5,722 | 0.7 | 4.0 | 43 | 252 | 1.05 (0.68 - 1.58) | 0.82 | 0.93 (0.61 - 1.43) | 0.75 |
| Open Claims | 274,852 | 807,270 | 245.4 | 871.4 | 2,980 | 13,407 | 0.90 (0.85 - 0.95) | <0.01 | 0.96 (0.79 - 1.16) | 0.67 |
| OEHR | 16,955 | 68,391 | 5.3 | 37.5 | 147 | 1,001 | 1.32 (1.05 - 1.67) | 0.02 | 1.35 (1.06 - 1.70) | 0.02 |
| VA | 0 | 0 | 0.0 | 0.0 | 0 | 0 | | | | |
| SIDIAP | 4,418 | 1,186 | 6.5 | 2.2 | 19 | 5 | 0.78 (0.24 - 2.73) | 0.68 | 0.73 (0.22 - 2.47) | 0.62 |
| Meta-analysis | 332,447 | 975,732 | 282.7 | 995.0 | 3,901 | 16,994 | 1.04 (0.91 - 1.19) | 0.52 | 1.08 (0.86 - 1.34) | 0.52 |
| Meta-analysis (w/o USOC) | 57,595 | 168,462 | 37.3 | 123.5 | 921 | 3,587 | 1.11 (1.00 - 1.23) | 0.05 | 1.10 (0.97 - 1.25) | 0.21 |
| **PS-stratification / on-treatment TAR** | | | | | | | | | | |
| CCAE | 21,821 | 43,009 | 17.8 | 35.5 | 432 | 829 | 1.08 (0.95 - 1.22) | 0.25 | 1.11 (0.98 - 1.27) | 0.13 |
| DAG | 5,395 | 1,334 | 4.5 | 1.3 | 34 | 12 | 1.06 (0.45 - 2.65) | 0.90 | 1.39 (0.57 - 3.39) | 0.48 |
| ODOD | 15,574 | 44,119 | 10.7 | 39.6 | 302 | 1,322 | 0.87 (0.76 - 1.00) | 0.06 | 0.86 (0.69 - 1.06) | 0.21 |
| MDCR | 1,958 | 7,697 | 1.5 | 7.2 | 38 | 233 | 0.90 (0.60 - 1.32) | 0.61 | 0.88 (0.59 - 1.30) | 0.54 |
| MDCD | 1,195 | 5,861 | 0.7 | 4.1 | 43 | 262 | 1.00 (0.70 - 1.41) | 0.98 | 0.95 (0.67 - 1.35) | 0.77 |
| Open Claims | 299,241 | 812,448 | 261.1 | 876.8 | 3,193 | 13,602 | 0.88 (0.85 - 0.92) | <0.01 | 0.94 (0.77 - 1.16) | 0.59 |
| OEHR | 18,784 | 70,594 | 5.9 | 38.9 | 162 | 1,064 | 1.20 (0.99 - 1.44) | 0.05 | 1.29 (1.07 - 1.55) | 0.01 |
| VA | 11,542 | 103,745 | 7.0 | 95.1 | 53 | 948 | 0.75 (0.56 - 0.97) | 0.04 | 0.85 (0.54 - 1.34) | 0.50 |
| SIDIAP | 5,004 | 2,630 | 6.9 | 6.1 | 20 | 25 | 0.77 (0.30 - 1.87) | 0.56 | 0.70 (0.28 - 1.75) | 0.46 |
| Meta-analysis | 355,420 | 970,170 | 295.6 | 990.8 | 4,089 | 16,817 | 0.99 (0.85 - 1.14) | 0.85 | 1.02 (0.82 - 1.29) | 0.83 |
| Meta-analysis (w/o USOC) | 56,179 | 157,722 | 34.5 | 114.0 | 896 | 3,215 | 1.03 (0.87 - 1.23) | 0.71 | 1.05 (0.87 - 1.28) | 0.62 |
| **PS-matching / on-treatment TAR without escalation** | | | | | | | | | | |
| CCAE | 18,644 | 42,437 | 13.7 | 30.6 | 316 | 707 | 1.10 (0.91 - 1.33) | 0.31 | 1.14 (0.94 - 1.37) | 0.22 |
| DAG | 5,175 | 921 | 4.0 | 0.8 | 30 | 8 | 1.32 (0.47 - 4.26) | 0.62 | 2.59 (0.86 - 7.77) | 0.10 |
| ODOD | 14,037 | 43,462 | 8.9 | 35.0 | 239 | 1,138 | 0.97 (0.80 - 1.17) | 0.79 | 0.96 (0.76 - 1.20) | 0.74 |
| MDCR | 1,627 | 7,529 | 1.2 | 6.5 | 33 | 200 | 1.03 (0.62 - 1.65) | 0.90 | 1.02 (0.62 - 1.67) | 0.86 |
| MDCD | 1,157 | 5,722 | 0.6 | 3.4 | 37 | 216 | 0.97 (0.61 - 1.52) | 0.91 | 0.86 (0.54 - 1.36) | 0.54 |
| Open Claims | 274,852 | 807,270 | 208.8 | 742.1 | 2,361 | 11,117 | 0.87 (0.82 - 0.93) | <0.01 | 0.94 (0.78 - 1.13) | 0.50 |
| OEHR | 16,955 | 68,391 | 4.7 | 32.8 | 117 | 772 | 1.32 (1.01 - 1.71) | 0.04 | 1.38 (1.06 - 1.80) | 0.02 |
| VA | 0 | 0 | 0.0 | 0.0 | 0 | 0 | | | | |
| SIDIAP | 4,418 | 1,186 | 5.7 | 1.9 | 17 | <5 | 0.72 (0.15 - 3.86) | 0.69 | 0.71 (0.14 - 3.55) | 0.68 |
| Meta-analysis | 332,447 | 975,732 | 241.8 | 851.2 | 3,133 | 14,158 | 1.02 (0.89 - 1.17) | 0.80 | 1.06 (0.85 - 1.30) | 0.61 |
| Meta-analysis (w/o USOC) | 57,595 | 168,462 | 33.0 | 109.0 | 772 | 3,041 | 1.08 (0.97 - 1.21) | 0.17 | 1.08 (0.95 - 1.23) | 0.29 |
| **PS-stratification / on-treatment TAR without escalation** | | | | | | | | | | |
| CCAE | 21,821 | 43,009 | 15.7 | 30.9 | 359 | 720 | 1.03 (0.89 - 1.18) | 0.72 | 1.08 (0.93 - 1.24) | 0.34 |
| DAG | 5,395 | 1,334 | 4.1 | 1.2 | 31 | 12 | 1.07 (0.44 - 2.72) | 0.89 | 1.49 (0.60 - 3.69) | 0.41 |
| ODOD | 15,574 | 44,119 | 9.6 | 35.5 | 256 | 1,170 | 0.86 (0.74 - 1.00) | 0.05 | 0.84 (0.69 - 1.02) | 0.13 |
| MDCR | 1,958 | 7,697 | 1.4 | 6.6 | 35 | 206 | 1.05 (0.69 - 1.57) | 0.81 | 1.01 (0.67 - 1.53) | 0.87 |
| MDCD | 1,195 | 5,861 | 0.6 | 3.4 | 37 | 226 | 0.94 (0.63 - 1.35) | 0.73 | 0.88 (0.60 - 1.29) | 0.54 |
| Open Claims | 299,241 | 812,448 | 222.9 | 746.8 | 2,542 | 11,285 | 0.85 (0.81 - 0.89) | <0.01 | 0.92 (0.75 - 1.13) | 0.43 |
| OEHR | 18,784 | 70,594 | 5.2 | 34.0 | 130 | 821 | 1.17 (0.95 - 1.44) | 0.13 | 1.26 (1.02 - 1.55) | 0.04 |
| VA | 11,542 | 103,745 | 6.0 | 84.3 | 41 | 765 | 0.71 (0.51 - 0.96) | 0.04 | 0.80 (0.57 - 1.13) | 0.33 |
| SIDIAP | 5,004 | 2,630 | 6.1 | 5.2 | 18 | 22 | 0.56 (0.21 - 1.47) | 0.24 | 0.55 (0.21 - 1.44) | 0.24 |
| Meta-analysis | 355,420 | 970,170 | 253.3 | 847.2 | 3,287 | 13,996 | 0.95 (0.83 - 1.09) | 0.48 | 1.00 (0.80 - 1.24) | 0.92 |
| Meta-analysis (w/o USOC) | 56,179 | 157,722 | 30.5 | 100.5 | 745 | 2,711 | 1.00 (0.85 - 1.18) | 0.99 | 1.02 (0.85 - 1.23) | 0.83 |

##### 1.3.23 Peripheral edema

**Supplementary Table 75:** Exposures, outcomes and risk estimates for Peripheral edema in SGLT2I (male) (T) vs SU (male) (C) new-user cohorts under four different propensity score (PS) adjustment / time-at-risk (TAR) designs across data sources. We report total patient exposure time in 1,000 patient-year (KYr) units, uncalibrated and calibrated hazard ratios (HRs) and their 95% confidence intervals (CIs). Greyed out entries did not pass study diagnostics and are included only for completeness.

| | Patients | | Exposure (KYr) | | Outcomes | | Uncalibrated | | Calibrated | |
|---|---|---|---|---|---|---|---|---|---|---|
| | T | C | T | C | T | C | HR (95% CI) | $p$ | HR (95% CI) | $p$ |
| **PS-matching / on-treatment TAR** | | | | | | | | | | |
| CCAE | 17,523 | 39,972 | 14.5 | 32.3 | 451 | 1,346 | 0.82 (0.70 - 0.95) | <0.01 | 0.83 (0.71 - 0.97) | 0.03 |
| DAG | 4,810 | 854 | 4.0 | 0.8 | 57 | 21 | 0.49 (0.24 - 1.03) | 0.06 | 0.92 (0.44 - 1.91) | 0.68 |
| ODOD | 12,839 | 38,577 | 9.0 | 32.8 | 377 | 2,478 | 0.59 (0.51 - 0.68) | <0.01 | 0.58 (0.46 - 0.74) | <0.01 |
| MDCR | 1,383 | 6,216 | 1.1 | 5.4 | 52 | 475 | 0.64 (0.43 - 0.94) | 0.03 | 0.65 (0.44 - 0.96) | 0.04 |
| MDCD | 1,069 | 5,088 | 0.6 | 3.3 | 37 | 278 | 0.95 (0.59 - 1.49) | 0.84 | 0.85 (0.53 - 1.35) | 0.51 |
| Open Claims | 249,361 | 729,685 | 221.8 | 764.2 | 3,909 | 25,663 | 0.66 (0.63 - 0.69) | <0.01 | 0.70 (0.58 - 0.85) | <0.01 |
| OEHR | 15,892 | 63,880 | 5.0 | 33.8 | 116 | 1,739 | 0.57 (0.45 - 0.72) | <0.01 | 0.58 (0.46 - 0.74) | <0.01 |
| VA | 0 | 0 | 0.0 | 0.0 | 0 | 0 | | | | |
| SIDIAP | 4,070 | 1,122 | 6.0 | 2.0 | 60 | 29 | 0.69 (0.38 - 1.27) | 0.23 | 0.65 (0.35 - 1.19) | 0.18 |
| Meta-analysis | 306,947 | 885,394 | 262.0 | 874.7 | 5,059 | 32,029 | 0.67 (0.59 - 0.75) | <0.01 | 0.69 (0.56 - 0.85) | <0.01 |
| Meta-analysis (w/o USOC) | 57,586 | 155,709 | 40.2 | 110.5 | 1,150 | 6,366 | 0.67 (0.57 - 0.79) | <0.01 | 0.67 (0.56 - 0.80) | <0.01 |
| **PS-stratification / on-treatment TAR** | | | | | | | | | | |
| CCAE | 20,331 | 40,503 | 16.5 | 32.5 | 518 | 1,366 | 0.79 (0.71 - 0.89) | <0.01 | 0.82 (0.73 - 0.92) | <0.01 |
| DAG | 5,011 | 1,246 | 4.1 | 1.2 | 57 | 39 | 0.52 (0.30 - 0.92) | 0.02 | 0.68 (0.39 - 1.20) | 0.21 |
| ODOD | 14,095 | 39,119 | 9.6 | 33.3 | 406 | 2,552 | 0.66 (0.59 - 0.74) | <0.01 | 0.64 (0.53 - 0.78) | <0.01 |
| MDCR | 1,633 | 6,377 | 1.3 | 5.5 | 56 | 501 | 0.62 (0.45 - 0.84) | <0.01 | 0.60 (0.44 - 0.82) | <0.01 |
| MDCD | 1,095 | 5,322 | 0.6 | 3.5 | 37 | 296 | 0.85 (0.58 - 1.21) | 0.40 | 0.81 (0.56 - 1.17) | 0.28 |
| Open Claims | 269,915 | 733,872 | 235.1 | 768.3 | 4,195 | 26,074 | 0.67 (0.64 - 0.69) | <0.01 | 0.71 (0.58 - 0.87) | <0.01 |
| OEHR | 17,562 | 65,731 | 5.5 | 34.9 | 130 | 1,856 | 0.57 (0.47 - 0.69) | <0.01 | 0.61 (0.50 - 0.74) | <0.01 |
| VA | 9,289 | 87,849 | 5.6 | 77.1 | 177 | 3,099 | 0.78 (0.67 - 0.91) | <0.01 | 0.89 (0.60 - 1.31) | 0.58 |
| SIDIAP | 4,631 | 2,504 | 6.4 | 5.7 | 66 | 79 | 0.63 (0.39 - 1.00) | 0.05 | 0.58 (0.36 - 0.93) | 0.03 |
| Meta-analysis | 321,903 | 879,225 | 266.7 | 869.0 | 5,249 | 31,848 | 0.68 (0.60 - 0.76) | <0.01 | 0.70 (0.57 - 0.87) | <0.01 |
| Meta-analysis (w/o USOC) | 51,988 | 145,353 | 31.6 | 100.8 | 1,054 | 5,774 | 0.68 (0.57 - 0.81) | <0.01 | 0.69 (0.57 - 0.84) | <0.01 |
| **PS-matching / on-treatment TAR without escalation** | | | | | | | | | | |
| CCAE | 17,523 | 39,972 | 12.7 | 28.2 | 378 | 1,176 | 0.76 (0.65 - 0.90) | <0.01 | 0.79 (0.67 - 0.93) | <0.01 |
| DAG | 4,810 | 854 | 3.7 | 0.7 | 53 | 21 | 0.49 (0.24 - 1.05) | 0.06 | 0.97 (0.46 - 2.02) | 0.68 |
| ODOD | 12,839 | 38,577 | 8.0 | 29.6 | 307 | 2,209 | 0.55 (0.46 - 0.64) | <0.01 | 0.53 (0.43 - 0.66) | <0.01 |
| MDCR | 1,383 | 6,216 | 1.0 | 5.0 | 44 | 436 | 0.57 (0.37 - 0.86) | 0.01 | 0.56 (0.37 - 0.87) | 0.02 |
| MDCD | 1,069 | 5,088 | 0.5 | 2.8 | 30 | 229 | 0.76 (0.44 - 1.28) | 0.33 | 0.67 (0.39 - 1.16) | 0.17 |
| Open Claims | 249,361 | 729,685 | 188.8 | 652.7 | 3,222 | 21,880 | 0.65 (0.62 - 0.68) | <0.01 | 0.70 (0.58 - 0.84) | <0.01 |
| OEHR | 15,892 | 63,880 | 4.4 | 29.7 | 95 | 1,393 | 0.57 (0.44 - 0.74) | <0.01 | 0.60 (0.46 - 0.78) | <0.01 |
| VA | 0 | 0 | 0.0 | 0.0 | 0 | 0 | | | | |
| SIDIAP | 4,070 | 1,122 | 5.3 | 1.8 | 54 | 26 | 0.56 (0.30 - 1.07) | 0.08 | 0.55 (0.29 - 1.05) | 0.08 |
| Meta-analysis | 306,947 | 885,394 | 224.5 | 750.5 | 4,183 | 27,370 | 0.63 (0.56 - 0.71) | <0.01 | 0.65 (0.54 - 0.80) | <0.01 |
| Meta-analysis (w/o USOC) | 57,586 | 155,709 | 35.7 | 97.9 | 961 | 5,490 | 0.62 (0.53 - 0.72) | <0.01 | 0.62 (0.53 - 0.73) | <0.01 |
| **PS-stratification / on-treatment TAR without escalation** | | | | | | | | | | |
| CCAE | 20,331 | 40,503 | 14.5 | 28.4 | 440 | 1,195 | 0.76 (0.67 - 0.86) | <0.01 | 0.80 (0.70 - 0.90) | <0.01 |
| DAG | 5,011 | 1,246 | 3.8 | 1.1 | 53 | 39 | 0.48 (0.27 - 0.84) | 0.01 | 0.66 (0.38 - 1.17) | 0.19 |
| ODOD | 14,095 | 39,119 | 8.6 | 30.1 | 336 | 2,277 | 0.61 (0.53 - 0.69) | <0.01 | 0.59 (0.49 - 0.70) | <0.01 |
| MDCR | 1,633 | 6,377 | 1.1 | 5.1 | 48 | 460 | 0.60 (0.43 - 0.83) | <0.01 | 0.58 (0.41 - 0.80) | <0.01 |
| MDCD | 1,095 | 5,322 | 0.5 | 3.0 | 30 | 246 | 0.77 (0.50 - 1.13) | 0.20 | 0.72 (0.48 - 1.09) | 0.14 |
| Open Claims | 269,915 | 733,872 | 200.8 | 656.3 | 3,475 | 22,252 | 0.65 (0.63 - 0.68) | <0.01 | 0.70 (0.57 - 0.86) | <0.01 |
| OEHR | 17,562 | 65,731 | 4.9 | 30.7 | 105 | 1,472 | 0.57 (0.46 - 0.71) | <0.01 | 0.61 (0.50 - 0.76) | <0.01 |
| VA | 9,289 | 87,849 | 4.8 | 68.7 | 151 | 2,720 | 0.78 (0.66 - 0.92) | <0.01 | 0.88 (0.71 - 1.09) | 0.47 |
| SIDIAP | 4,631 | 2,504 | 5.7 | 4.9 | 60 | 67 | 0.64 (0.38 - 1.06) | 0.08 | 0.62 (0.37 - 1.03) | 0.09 |
| Meta-analysis | 321,903 | 879,225 | 228.8 | 745.4 | 4,356 | 27,196 | 0.65 (0.59 - 0.73) | <0.01 | 0.68 (0.56 - 0.84) | <0.01 |
| Meta-analysis (w/o USOC) | 51,988 | 145,353 | 28.0 | 89.2 | 881 | 4,944 | 0.65 (0.54 - 0.77) | <0.01 | 0.66 (0.54 - 0.80) | <0.01 |

##### 1.3.24 Photosensitivity

**Supplementary Table 76:** Exposures, outcomes and risk estimates for Photosensitivity in SGLT2I (male) (T) vs SU (male) (C) new-user cohorts under four different propensity score (PS) adjustment / time-at-risk (TAR) designs across data sources. We report total patient exposure time in 1,000 patient-year (KYr) units, uncalibrated and calibrated hazard ratios (HRs) and their 95% confidence intervals (CIs). Greyed out entries did not pass study diagnostics and are included only for completeness.

| | Patients | | Exposure (KYr) | | Outcomes | | Uncalibrated | | Calibrated | |
|---|---|---|---|---|---|---|---|---|---|---|
| | T | C | T | C | T | C | HR (95% CI) | $p$ | HR (95% CI) | $p$ |
| **PS-matching / on-treatment TAR** | | | | | | | | | | |
| CCAE | 20,476 | 46,543 | 17.4 | 38.9 | 10 | 42 | 0.78 (0.24 - 2.20) | 0.66 | 0.79 (0.26 - 2.40) | 0.68 |
| DAG | 5,457 | 970 | 4.6 | 0.9 | 0 | 0 | | | | |
| ODOD | 15,608 | 48,467 | 11.1 | 44.0 | 5 | 42 | 0.64 (0.14 - 2.07) | 0.52 | 0.63 (0.16 - 2.45) | 0.51 |
| MDCR | 1,749 | 8,143 | 1.5 | 7.6 | <5 | 25 | 0.59 (0.03 - 3.59) | 0.66 | 0.59 (0.05 - 6.46) | 0.67 |
| MDCD | 1,453 | 7,219 | 0.9 | 5.1 | 0 | <5 | | | | |
| Open Claims | 302,021 | 881,219 | 268.8 | 951.5 | 67 | 446 | 0.89 (0.63 - 1.25) | 0.52 | 0.95 (0.65 - 1.40) | 0.81 |
| OEHR | 18,182 | 73,261 | 5.8 | 41.1 | 5 | 68 | 1.68 (0.53 - 4.40) | 0.33 | 1.71 (0.60 - 4.91) | 0.32 |
| VA | 0 | 0 | 0.0 | 0.0 | 0 | 0 | | | | |
| SIDIAP | 4,549 | 1,219 | 6.6 | 2.2 | <5 | <5 | 0.83 (0.07 - 19.5) | 0.90 | 0.78 (0.05 - 13.1) | 0.86 |
| Meta-analysis | 358,036 | 1,057,633 | 304.7 | 1083.2 | <92 | 623 | 0.91 (0.67 - 1.23) | 0.54 | 0.94 (0.66 - 1.33) | 0.72 |
| Meta-analysis (w/o USOC) | 56,015 | 176,414 | 35.9 | 131.6 | <25 | 177 | 0.97 (0.51 - 1.84) | 0.93 | 0.97 (0.51 - 1.84) | 0.92 |
| **PS-stratification / on-treatment TAR** | | | | | | | | | | |
| CCAE | 23,945 | 47,187 | 19.9 | 39.3 | 11 | 42 | 0.77 (0.35 - 1.56) | 0.49 | 0.79 (0.38 - 1.68) | 0.55 |
| DAG | 5,686 | 1,398 | 4.7 | 1.4 | 0 | <5 | | | | |
| ODOD | 17,348 | 49,106 | 12.0 | 44.7 | 8 | 43 | 0.75 (0.30 - 1.70) | 0.53 | 0.74 (0.31 - 1.79) | 0.51 |
| MDCR | 2,105 | 8,426 | 1.7 | 7.9 | <5 | 25 | 0.76 (0.11 - 2.94) | 0.74 | 0.74 (0.14 - 3.76) | 0.71 |
| MDCD | 1,495 | 7,464 | 0.9 | 5.3 | 0 | <5 | | | | |
| Open Claims | 329,565 | 886,842 | 286.5 | 957.4 | 68 | 465 | 0.89 (0.67 - 1.16) | 0.39 | 0.95 (0.67 - 1.33) | 0.76 |
| OEHR | 20,107 | 75,737 | 6.4 | 42.7 | 5 | 79 | 1.14 (0.38 - 2.73) | 0.79 | 1.22 (0.46 - 3.27) | 0.69 |
| VA | 12,352 | 109,597 | 7.4 | 100.6 | <10 | 100 | 0.11 (0.01 - 0.51) | 0.05 | 0.13 (0.01 - 1.18) | 0.07 |
| SIDIAP | 5,159 | 2,688 | 7.1 | 6.3 | <5 | 6 | 0.39 (0.04 - 2.80) | 0.39 | 0.36 (0.04 - 3.05) | 0.35 |
| Meta-analysis | 390,965 | 1,058,872 | 324.9 | 1084.0 | 92 | 629 | 0.88 (0.69 - 1.11) | 0.28 | 0.91 (0.68 - 1.22) | 0.54 |
| Meta-analysis (w/o USOC) | 61,400 | 172,030 | 38.4 | 126.6 | 24 | 164 | 0.84 (0.52 - 1.38) | 0.49 | 0.86 (0.52 - 1.41) | 0.55 |
| **PS-matching / on-treatment TAR without escalation** | | | | | | | | | | |
| CCAE | 20,476 | 46,543 | 15.2 | 33.7 | 9 | 37 | 0.70 (0.19 - 2.15) | 0.57 | 0.72 (0.21 - 2.45) | 0.60 |
| DAG | 5,457 | 970 | 4.2 | 0.8 | 0 | 0 | | | | |
| ODOD | 15,608 | 48,467 | 9.9 | 39.4 | <5 | 40 | 0.75 (0.16 - 2.49) | 0.68 | 0.73 (0.19 - 2.89) | 0.66 |
| MDCR | 1,749 | 8,143 | 1.3 | 7.0 | <5 | 24 | 0.57 (0.03 - 3.49) | 0.65 | 0.57 (0.05 - 6.19) | 0.64 |
| MDCD | 1,453 | 7,219 | 0.7 | 4.2 | 0 | <5 | | | | |
| Open Claims | 302,021 | 881,219 | 227.8 | 806.9 | 54 | 407 | 0.93 (0.64 - 1.31) | 0.67 | 0.99 (0.67 - 1.48) | 0.95 |
| OEHR | 18,182 | 73,261 | 5.1 | 35.6 | <5 | 62 | 1.86 (0.52 - 5.29) | 0.30 | 1.94 (0.61 - 6.21) | 0.27 |
| VA | 0 | 0 | 0.0 | 0.0 | 0 | 0 | | | | |
| SIDIAP | 4,549 | 1,219 | 5.9 | 1.9 | <5 | <5 | 0.83 (0.07 - 19.5) | 0.90 | 0.82 (0.05 - 13.7) | 0.89 |
| Meta-analysis | 358,036 | 1,057,633 | 259.2 | 922.5 | <78 | 570 | 0.94 (0.68 - 1.29) | 0.69 | 0.97 (0.68 - 1.39) | 0.88 |
| Meta-analysis (w/o USOC) | 56,015 | 176,414 | 31.4 | 115.7 | <24 | 163 | 0.98 (0.50 - 1.95) | 0.96 | 0.99 (0.50 - 1.97) | 0.96 |
| **PS-stratification / on-treatment TAR without escalation** | | | | | | | | | | |
| CCAE | 23,945 | 47,187 | 17.4 | 34.0 | 10 | 37 | 0.88 (0.39 - 1.85) | 0.75 | 0.92 (0.42 - 2.02) | 0.84 |
| DAG | 5,686 | 1,398 | 4.3 | 1.3 | 0 | <5 | | | | |
| ODOD | 17,348 | 49,106 | 10.7 | 39.9 | 7 | 41 | 0.73 (0.27 - 1.75) | 0.51 | 0.71 (0.28 - 1.83) | 0.49 |
| MDCR | 2,105 | 8,426 | 1.5 | 7.2 | <5 | 24 | 0.80 (0.12 - 3.13) | 0.79 | 0.77 (0.15 - 3.95) | 0.76 |
| MDCD | 1,495 | 7,464 | 0.8 | 4.4 | 0 | <5 | | | | |
| Open Claims | 329,565 | 886,842 | 243.6 | 811.9 | 55 | 426 | 0.82 (0.60 - 1.10) | 0.19 | 0.88 (0.61 - 1.27) | 0.49 |
| OEHR | 20,107 | 75,737 | 5.7 | 37.0 | <5 | 73 | 0.94 (0.27 - 2.45) | 0.92 | 1.01 (0.34 - 3.02) | 0.96 |
| VA | 12,352 | 109,597 | 6.4 | 89.1 | <10 | 92 | 0.13 (0.01 - 0.57) | 0.06 | 0.14 (0.02 - 1.27) | 0.08 |
| SIDIAP | 5,159 | 2,688 | 6.3 | 5.3 | <5 | 5 | 0.39 (0.04 - 2.88) | 0.40 | 0.38 (0.04 - 3.30) | 0.39 |
| Meta-analysis | 390,965 | 1,058,872 | 277.3 | 922.9 | <77 | 577 | 0.82 (0.63 - 1.07) | 0.15 | 0.86 (0.63 - 1.18) | 0.36 |
| Meta-analysis (w/o USOC) | 61,400 | 172,030 | 33.7 | 110.9 | <22 | 151 | 0.84 (0.50 - 1.43) | 0.53 | 0.86 (0.50 - 1.47) | 0.59 |

##### 1.3.25 Venous thromboembolism

**Supplementary Table 77:** Exposures, outcomes and risk estimates for Venous thromboembolism in SGLT2I (male) (T) vs SU (male) (C) new-user cohorts under four different propensity score (PS) adjustment / time-at-risk (TAR) designs across data sources. We report total patient exposure time in 1,000 patient-year (KYr) units, uncalibrated and calibrated hazard ratios (HRs) and their 95% confidence intervals (CIs). Greyed out entries did not pass study diagnostics and are included only for completeness.

| | Patients | | Exposure (KYr) | | Outcomes | | Uncalibrated | | Calibrated | |
|---|---|---|---|---|---|---|---|---|---|---|
| | T | C | T | C | T | C | HR (95% CI) | $p$ | HR (95% CI) | $p$ |
| **PS-matching / on-treatment TAR** | | | | | | | | | | |
| CCAE | 20,188 | 45,819 | 17.2 | 38.2 | 69 | 224 | 0.72 (0.49 - 1.05) | 0.10 | 0.74 (0.50 - 1.08) | 0.12 |
| DAG | 5,202 | 928 | 4.3 | 0.9 | 27 | 6 | 1.14 (0.25 - 8.15) | 0.88 | 2.14 (0.38 - 12.1) | 0.40 |
| ODOD | 15,300 | 47,283 | 10.9 | 42.7 | 72 | 511 | 0.66 (0.47 - 0.91) | 0.01 | 0.65 (0.44 - 0.95) | 0.04 |
| MDCR | 1,698 | 7,910 | 1.4 | 7.4 | 12 | 113 | 1.16 (0.51 - 2.42) | 0.71 | 1.16 (0.53 - 2.53) | 0.71 |
| MDCD | 1,409 | 6,928 | 0.9 | 4.8 | 10 | 50 | 1.89 (0.67 - 4.92) | 0.21 | 1.68 (0.62 - 4.56) | 0.31 |
| Open Claims | 295,267 | 859,629 | 263.2 | 926.0 | 921 | 5,559 | 0.72 (0.65 - 0.79) | <0.01 | 0.76 (0.62 - 0.94) | 0.01 |
| OEHR | 17,845 | 71,845 | 5.6 | 39.9 | 39 | 358 | 0.91 (0.60 - 1.37) | 0.67 | 0.93 (0.61 - 1.41) | 0.74 |
| VA | 0 | 0 | 0.0 | 0.0 | 0 | 0 | | | | |
| SIDIAP | 4,441 | 1,186 | 6.4 | 2.2 | 24 | 11 | 0.41 (0.14 - 1.19) | 0.10 | 0.38 (0.13 - 1.11) | 0.08 |
| Meta-analysis | 361,350 | 1,041,528 | 309.9 | 1062.1 | 1,174 | 6,832 | 0.73 (0.67 - 0.79) | <0.01 | 0.75 (0.62 - 0.91) | <0.01 |
| Meta-analysis (w/o USOC) | 66,083 | 181,899 | 46.8 | 136.0 | 253 | 1,273 | 0.80 (0.63 - 1.01) | 0.06 | 0.79 (0.62 - 1.02) | 0.09 |
| **PS-stratification / on-treatment TAR** | | | | | | | | | | |
| CCAE | 23,622 | 46,448 | 19.6 | 38.5 | 77 | 231 | 0.69 (0.52 - 0.91) | 0.01 | 0.71 (0.54 - 0.95) | 0.02 |
| DAG | 5,411 | 1,343 | 4.5 | 1.3 | 27 | 14 | 1.28 (0.52 - 3.15) | 0.59 | 1.67 (0.68 - 4.11) | 0.27 |
| ODOD | 16,980 | 47,874 | 11.8 | 43.2 | 81 | 533 | 0.70 (0.54 - 0.90) | <0.01 | 0.68 (0.50 - 0.92) | 0.02 |
| MDCR | 2,048 | 8,160 | 1.6 | 7.6 | 15 | 120 | 0.72 (0.38 - 1.29) | 0.29 | 0.70 (0.38 - 1.29) | 0.27 |
| MDCD | 1,452 | 7,144 | 0.9 | 5.0 | 10 | 51 | 1.63 (0.74 - 3.33) | 0.20 | 1.55 (0.73 - 3.29) | 0.26 |
| Open Claims | 321,867 | 865,397 | 280.3 | 932.0 | 985 | 5,672 | 0.74 (0.69 - 0.79) | <0.01 | 0.79 (0.64 - 0.97) | 0.03 |
| OEHR | 19,746 | 74,195 | 6.3 | 41.4 | 42 | 402 | 0.86 (0.60 - 1.21) | 0.41 | 0.93 (0.65 - 1.32) | 0.68 |
| VA | 11,951 | 107,388 | 7.2 | 98.3 | 44 | 537 | 1.08 (0.78 - 1.45) | 0.63 | 1.23 (0.77 - 1.97) | 0.42 |
| SIDIAP | 5,024 | 2,636 | 6.9 | 6.1 | 26 | 22 | 0.77 (0.35 - 1.67) | 0.50 | 0.71 (0.32 - 1.53) | 0.39 |
| Meta-analysis | 382,215 | 1,033,914 | 318.0 | 1055.3 | 1,185 | 6,838 | 0.74 (0.69 - 0.79) | <0.01 | 0.77 (0.63 - 0.92) | <0.01 |
| Meta-analysis (w/o USOC) | 60,348 | 168,517 | 37.7 | 123.2 | 200 | 1,166 | 0.73 (0.62 - 0.86) | <0.01 | 0.74 (0.62 - 0.90) | <0.01 |
| **PS-matching / on-treatment TAR without escalation** | | | | | | | | | | |
| CCAE | 20,188 | 45,819 | 15.0 | 33.1 | 57 | 181 | 0.67 (0.44 - 1.01) | 0.06 | 0.69 (0.46 - 1.05) | 0.09 |
| DAG | 5,202 | 928 | 4.0 | 0.8 | 26 | 6 | 1.13 (0.25 - 8.08) | 0.89 | 2.21 (0.39 - 12.6) | 0.38 |
| ODOD | 15,300 | 47,283 | 9.7 | 38.2 | 58 | 453 | 0.62 (0.42 - 0.89) | 0.01 | 0.61 (0.41 - 0.90) | 0.02 |
| MDCR | 1,698 | 7,910 | 1.3 | 6.8 | 12 | 104 | 1.25 (0.53 - 2.72) | 0.59 | 1.24 (0.54 - 2.82) | 0.62 |
| MDCD | 1,409 | 6,928 | 0.7 | 4.0 | 9 | 39 | 2.04 (0.64 - 6.04) | 0.21 | 1.80 (0.58 - 5.56) | 0.31 |
| Open Claims | 295,267 | 859,629 | 223.2 | 785.9 | 752 | 4,759 | 0.71 (0.64 - 0.78) | <0.01 | 0.76 (0.62 - 0.93) | 0.01 |
| OEHR | 17,845 | 71,845 | 4.9 | 34.7 | 28 | 270 | 0.80 (0.47 - 1.30) | 0.38 | 0.83 (0.50 - 1.38) | 0.50 |
| VA | 0 | 0 | 0.0 | 0.0 | 0 | 0 | | | | |
| SIDIAP | 4,441 | 1,186 | 5.7 | 1.9 | 21 | 9 | 0.36 (0.12 - 1.07) | 0.07 | 0.35 (0.12 - 1.06) | 0.07 |
| Meta-analysis | 361,350 | 1,041,528 | 264.5 | 905.5 | 963 | 5,821 | 0.71 (0.65 - 0.78) | <0.01 | 0.74 (0.61 - 0.89) | <0.01 |
| Meta-analysis (w/o USOC) | 66,083 | 181,899 | 41.3 | 119.5 | 211 | 1,062 | 0.75 (0.57 - 0.99) | 0.04 | 0.75 (0.57 - 1.00) | 0.06 |
| **PS-stratification / on-treatment TAR without escalation** | | | | | | | | | | |
| CCAE | 23,622 | 46,448 | 17.2 | 33.5 | 64 | 188 | 0.73 (0.53 - 0.99) | 0.05 | 0.76 (0.55 - 1.04) | 0.10 |
| DAG | 5,411 | 1,343 | 4.1 | 1.2 | 26 | 14 | 1.25 (0.50 - 3.10) | 0.64 | 1.74 (0.70 - 4.32) | 0.25 |
| ODOD | 16,980 | 47,874 | 10.4 | 38.7 | 66 | 474 | 0.66 (0.50 - 0.87) | <0.01 | 0.64 (0.47 - 0.88) | <0.01 |
| MDCR | 2,048 | 8,160 | 1.4 | 7.0 | 15 | 111 | 0.82 (0.43 - 1.49) | 0.54 | 0.79 (0.42 - 1.47) | 0.47 |
| MDCD | 1,452 | 7,144 | 0.7 | 4.2 | 9 | 40 | 1.74 (0.74 - 3.74) | 0.18 | 1.64 (0.73 - 3.69) | 0.24 |
| Open Claims | 321,867 | 865,397 | 238.5 | 791.1 | 805 | 4,864 | 0.71 (0.65 - 0.77) | <0.01 | 0.76 (0.62 - 0.95) | 0.02 |
| OEHR | 19,746 | 74,195 | 5.5 | 36.0 | 31 | 297 | 0.78 (0.51 - 1.16) | 0.23 | 0.84 (0.55 - 1.26) | 0.40 |
| VA | 11,951 | 107,388 | 6.2 | 87.1 | 36 | 447 | 1.08 (0.75 - 1.50) | 0.66 | 1.22 (0.84 - 1.76) | 0.38 |
| SIDIAP | 5,024 | 2,636 | 6.1 | 5.2 | 23 | 19 | 0.65 (0.28 - 1.51) | 0.31 | 0.63 (0.27 - 1.47) | 0.31 |
| Meta-analysis | 382,215 | 1,033,914 | 271.6 | 899.3 | 966 | 5,823 | 0.71 (0.66 - 0.76) | <0.01 | 0.74 (0.62 - 0.90) | <0.01 |
| Meta-analysis (w/o USOC) | 60,348 | 168,517 | 33.1 | 108.1 | 161 | 959 | 0.71 (0.59 - 0.85) | <0.01 | 0.72 (0.59 - 0.89) | <0.01 |

##### 1.3.26 Vomiting

**Supplementary Table 78:** Exposures, outcomes and risk estimates for Vomiting in SGLT2I (male) (T) vs SU (male) (C) new-user cohorts under four different propensity score (PS) adjustment / time-at-risk (TAR) designs across data sources. We report total patient exposure time in 1,000 patient-year (KYr) units, uncalibrated and calibrated hazard ratios (HRs) and their 95% confidence intervals (CIs). Greyed out entries did not pass study diagnostics and are included only for completeness.

| | Patients | | Exposure (KYr) | | Outcomes | | Uncalibrated | | Calibrated | |
|---|---|---|---|---|---|---|---|---|---|---|
| | T | C | T | C | T | C | HR (95% CI) | $p$ | HR (95% CI) | $p$ |
| **PS-matching / on-treatment TAR** | | | | | | | | | | |
| CCAE | 19,268 | 43,723 | 16.2 | 36.3 | 277 | 588 | 1.14 (0.93 - 1.40) | 0.20 | 1.16 (0.94 - 1.42) | 0.18 |
| DAG | 5,175 | 921 | 4.3 | 0.9 | 33 | 8 | 1.35 (0.50 - 4.32) | 0.59 | 2.53 (0.85 - 7.48) | 0.11 |
| ODOD | 14,548 | 44,916 | 10.3 | 40.5 | 202 | 924 | 0.91 (0.73 - 1.13) | 0.40 | 0.90 (0.67 - 1.19) | 0.50 |
| MDCR | 1,684 | 7,707 | 1.4 | 7.2 | 28 | 177 | 1.19 (0.66 - 2.06) | 0.54 | 1.20 (0.68 - 2.12) | 0.54 |
| MDCD | 1,233 | 6,000 | 0.7 | 4.2 | 29 | 206 | 0.80 (0.49 - 1.27) | 0.36 | 0.71 (0.44 - 1.15) | 0.19 |
| Open Claims | 281,769 | 822,887 | 252.0 | 891.1 | 2,192 | 10,560 | 0.87 (0.81 - 0.92) | <0.01 | 0.92 (0.76 - 1.12) | 0.43 |
| OEHR | 17,371 | 70,182 | 5.5 | 38.8 | 91 | 713 | 1.41 (1.05 - 1.86) | 0.02 | 1.43 (1.07 - 1.90) | 0.02 |
| VA | 0 | 0 | 0.0 | 0.0 | 0 | 0 | | | | |
| SIDIAP | 4,404 | 1,181 | 6.4 | 2.2 | 21 | 8 | 0.35 (0.11 - 1.08) | 0.08 | 0.33 (0.10 - 1.05) | 0.06 |
| Meta-analysis | 345,452 | 997,517 | 296.9 | 1021.2 | 2,873 | 13,184 | 1.00 (0.85 - 1.19) | 0.98 | 1.03 (0.81 - 1.32) | 0.80 |
| Meta-analysis (w/o USOC) | 63,683 | 174,630 | 45.0 | 130.1 | 681 | 2,624 | 1.06 (0.86 - 1.29) | 0.60 | 1.05 (0.85 - 1.30) | 0.67 |
| **PS-stratification / on-treatment TAR** | | | | | | | | | | |
| CCAE | 22,530 | 44,307 | 18.5 | 36.7 | 308 | 599 | 1.10 (0.94 - 1.28) | 0.23 | 1.13 (0.97 - 1.32) | 0.13 |
| DAG | 5,395 | 1,334 | 4.5 | 1.3 | 34 | 12 | 1.06 (0.45 - 2.65) | 0.90 | 1.39 (0.57 - 3.39) | 0.48 |
| ODOD | 16,142 | 45,579 | 11.2 | 41.2 | 213 | 954 | 0.90 (0.76 - 1.06) | 0.21 | 0.88 (0.70 - 1.11) | 0.34 |
| MDCR | 2,025 | 7,919 | 1.6 | 7.4 | 29 | 191 | 1.00 (0.63 - 1.54) | 1.00 | 0.97 (0.62 - 1.52) | 0.87 |
| MDCD | 1,271 | 6,147 | 0.8 | 4.3 | 29 | 215 | 0.78 (0.51 - 1.17) | 0.25 | 0.74 (0.49 - 1.12) | 0.17 |
| Open Claims | 307,189 | 827,933 | 268.3 | 896.3 | 2,351 | 10,719 | 0.87 (0.83 - 0.91) | <0.01 | 0.93 (0.76 - 1.14) | 0.50 |
| OEHR | 19,279 | 72,408 | 6.1 | 40.2 | 99 | 760 | 1.14 (0.90 - 1.43) | 0.26 | 1.22 (0.97 - 1.54) | 0.10 |
| VA | 11,772 | 105,490 | 7.1 | 96.9 | 40 | 713 | 0.75 (0.53 - 1.01) | 0.07 | 0.85 (0.53 - 1.37) | 0.53 |
| SIDIAP | 4,990 | 2,623 | 6.9 | 6.1 | 22 | 28 | 0.59 (0.25 - 1.38) | 0.23 | 0.54 (0.23 - 1.29) | 0.17 |
| Meta-analysis | 365,140 | 990,227 | 304.1 | 1014.4 | 2,971 | 13,032 | 0.97 (0.85 - 1.11) | 0.70 | 1.01 (0.81 - 1.26) | 0.90 |
| Meta-analysis (w/o USOC) | 57,951 | 162,294 | 35.8 | 118.1 | 620 | 2,313 | 1.03 (0.89 - 1.19) | 0.68 | 1.05 (0.89 - 1.25) | 0.59 |
| **PS-matching / on-treatment TAR without escalation** | | | | | | | | | | |
| CCAE | 19,268 | 43,723 | 14.2 | 31.6 | 228 | 517 | 1.12 (0.90 - 1.39) | 0.30 | 1.16 (0.93 - 1.44) | 0.22 |
| DAG | 5,175 | 921 | 4.0 | 0.8 | 30 | 8 | 1.32 (0.47 - 4.26) | 0.62 | 2.59 (0.86 - 7.77) | 0.10 |
| ODOD | 14,548 | 44,916 | 9.2 | 36.3 | 171 | 816 | 0.93 (0.73 - 1.16) | 0.52 | 0.91 (0.70 - 1.19) | 0.54 |
| MDCR | 1,684 | 7,707 | 1.3 | 6.6 | 26 | 150 | 1.37 (0.74 - 2.41) | 0.30 | 1.35 (0.75 - 2.45) | 0.34 |
| MDCD | 1,233 | 6,000 | 0.6 | 3.5 | 27 | 173 | 0.89 (0.53 - 1.43) | 0.63 | 0.78 (0.47 - 1.29) | 0.36 |
| Open Claims | 281,769 | 822,887 | 214.1 | 758.0 | 1,724 | 8,742 | 0.85 (0.79 - 0.91) | <0.01 | 0.91 (0.76 - 1.10) | 0.36 |
| OEHR | 17,371 | 70,182 | 4.8 | 33.8 | 79 | 543 | 1.61 (1.18 - 2.18) | <0.01 | 1.68 (1.23 - 2.29) | <0.01 |
| VA | 0 | 0 | 0.0 | 0.0 | 0 | 0 | | | | |
| SIDIAP | 4,404 | 1,181 | 5.7 | 1.9 | 19 | 6 | 0.28 (0.06 - 1.19) | 0.09 | 0.28 (0.06 - 1.20) | 0.09 |
| Meta-analysis | 345,452 | 997,517 | 253.9 | 872.6 | 2,304 | 10,955 | 1.04 (0.85 - 1.28) | 0.68 | 1.08 (0.84 - 1.40) | 0.55 |
| Meta-analysis (w/o USOC) | 63,683 | 174,630 | 39.8 | 114.6 | 580 | 2,213 | 1.11 (0.89 - 1.40) | 0.35 | 1.12 (0.88 - 1.42) | 0.38 |
| **PS-stratification / on-treatment TAR without escalation** | | | | | | | | | | |
| CCAE | 22,530 | 44,307 | 16.2 | 31.9 | 255 | 528 | 1.02 (0.86 - 1.20) | 0.81 | 1.07 (0.91 - 1.27) | 0.44 |
| DAG | 5,395 | 1,334 | 4.1 | 1.2 | 31 | 12 | 1.07 (0.44 - 2.72) | 0.89 | 1.49 (0.60 - 3.69) | 0.41 |
| ODOD | 16,142 | 45,579 | 9.9 | 36.9 | 179 | 841 | 0.89 (0.74 - 1.06) | 0.18 | 0.86 (0.69 - 1.07) | 0.24 |
| MDCR | 2,025 | 7,919 | 1.4 | 6.8 | 27 | 163 | 1.21 (0.75 - 1.92) | 0.42 | 1.17 (0.73 - 1.87) | 0.54 |
| MDCD | 1,271 | 6,147 | 0.6 | 3.6 | 27 | 181 | 0.85 (0.54 - 1.30) | 0.48 | 0.80 (0.52 - 1.25) | 0.35 |
| Open Claims | 307,189 | 827,933 | 228.8 | 762.5 | 1,867 | 8,881 | 0.85 (0.80 - 0.89) | <0.01 | 0.91 (0.74 - 1.12) | 0.40 |
| OEHR | 19,279 | 72,408 | 5.4 | 35.1 | 84 | 576 | 1.27 (0.98 - 1.63) | 0.07 | 1.36 (1.05 - 1.75) | 0.02 |
| VA | 11,772 | 105,490 | 6.2 | 85.9 | 31 | 587 | 0.71 (0.48 - 1.00) | 0.06 | 0.80 (0.54 - 1.17) | 0.34 |
| SIDIAP | 4,990 | 2,623 | 6.1 | 5.2 | 20 | 24 | 0.45 (0.18 - 1.14) | 0.09 | 0.44 (0.18 - 1.11) | 0.09 |
| Meta-analysis | 365,140 | 990,227 | 260.4 | 866.4 | 2,385 | 10,826 | 0.97 (0.82 - 1.14) | 0.68 | 1.01 (0.80 - 1.28) | 0.90 |
| Meta-analysis (w/o USOC) | 57,951 | 162,294 | 31.6 | 103.9 | 518 | 1,945 | 1.03 (0.86 - 1.23) | 0.77 | 1.05 (0.86 - 1.28) | 0.66 |

#### 1.4 GLP1RA (male) vs DPP4I (male)

##### 1.4.1 3-point MACE

**Supplementary Table 79:** Exposures, outcomes and risk estimates for 3-point MACE in GLP1RA (male) (T) vs DPP4I (male) (C) new-user cohorts under four different propensity score (PS) adjustment / time-at-risk (TAR) designs across data sources. We report total patient exposure time in 1,000 patient-year (KYr) units, uncalibrated and calibrated hazard ratios (HRs) and their 95% confidence intervals (CIs). Greyed out entries did not pass study diagnostics and are included only for completeness.

| | Patients | | Exposure (KYr) | | Outcomes | | Uncalibrated | | Calibrated | |
|---|---|---|---|---|---|---|---|---|---|---|
| | T | C | T | C | T | C | HR (95% CI) | $p$ | HR (95% CI) | $p$ |
| **PS-matching / on-treatment TAR** | | | | | | | | | | |
| CCAE | 10,198 | 50,982 | 7.0 | 44.6 | 56 | 383 | 1.17 (0.79 - 1.69) | 0.42 | 1.13 (0.77 - 1.66) | 0.54 |
| ODOD | 6,134 | 28,677 | 3.8 | 24.1 | 37 | 435 | 0.52 (0.31 - 0.83) | <0.01 | 0.51 (0.31 - 0.84) | <0.01 |
| MDCR | 800 | 8,308 | 0.5 | 8.7 | 11 | 233 | 0.72 (0.32 - 1.42) | 0.38 | 0.69 (0.33 - 1.45) | 0.34 |
| Open Claims | 109,355 | 401,048 | 84.8 | 384.3 | 561 | 4,126 | 0.84 (0.74 - 0.94) | <0.01 | 0.85 (0.75 - 0.96) | 0.02 |
| OEHR | 7,631 | 37,284 | 2.3 | 18.5 | 23 | 204 | 1.09 (0.62 - 1.85) | 0.75 | 0.95 (0.55 - 1.65) | 0.86 |
| VA | 1,153 | 17,084 | 0.9 | 14.5 | <10 | 67 | 0.20 (0.01 - 1.02) | 0.16 | 0.16 (0.02 - 1.57) | 0.12 |
| Meta-analysis | 135,271 | 543,383 | 99.3 | 494.7 | <698 | 5,448 | 0.84 (0.65 - 1.08) | 0.18 | 0.83 (0.62 - 1.10) | 0.20 |
| Meta-analysis (w/o USOC) | 25,916 | 142,335 | 14.5 | 110.4 | <137 | 1,322 | 0.81 (0.53 - 1.23) | 0.32 | 0.78 (0.51 - 1.18) | 0.25 |
| **PS-stratification / on-treatment TAR** | | | | | | | | | | |
| CCAE | 12,059 | 51,105 | 7.9 | 44.7 | 60 | 383 | 0.95 (0.69 - 1.27) | 0.72 | 0.93 (0.69 - 1.27) | 0.67 |
| ODOD | 8,175 | 29,196 | 4.7 | 24.5 | 42 | 458 | 0.56 (0.39 - 0.79) | <0.01 | 0.54 (0.38 - 0.77) | <0.01 |
| MDCR | 893 | 8,780 | 0.6 | 9.2 | 11 | 259 | 0.72 (0.36 - 1.33) | 0.33 | 0.67 (0.35 - 1.30) | 0.24 |
| Open Claims | 147,591 | 402,784 | 101.8 | 385.9 | 647 | 4,214 | 0.82 (0.75 - 0.91) | <0.01 | 0.85 (0.77 - 0.94) | 0.02 |
| OEHR | 8,896 | 37,369 | 2.5 | 18.6 | 26 | 209 | 1.03 (0.64 - 1.60) | 0.90 | 0.90 (0.57 - 1.42) | 0.67 |
| VA | 1,334 | 24,642 | 0.9 | 23.0 | <10 | 124 | 0.87 (0.29 - 2.17) | 0.79 | 0.77 (0.28 - 2.11) | 0.61 |
| Meta-analysis | 176,721 | 520,454 | 116.9 | 473.7 | 775 | 5,264 | 0.82 (0.66 - 1.00) | 0.05 | 0.82 (0.65 - 1.05) | 0.12 |
| Meta-analysis (w/o USOC) | 29,130 | 117,670 | 15.1 | 87.8 | 128 | 1,050 | 0.81 (0.56 - 1.18) | 0.27 | 0.78 (0.53 - 1.14) | 0.21 |
| **PS-matching / on-treatment TAR without escalation** | | | | | | | | | | |
| CCAE | 10,198 | 50,982 | 6.1 | 36.1 | 49 | 306 | 1.26 (0.83 - 1.88) | 0.26 | 1.24 (0.82 - 1.86) | 0.32 |
| ODOD | 6,134 | 28,677 | 3.4 | 19.7 | 30 | 369 | 0.43 (0.24 - 0.74) | <0.01 | 0.41 (0.23 - 0.73) | <0.01 |
| MDCR | 800 | 8,308 | 0.5 | 7.4 | 7 | 191 | 0.51 (0.17 - 1.20) | 0.17 | 0.44 (0.17 - 1.16) | 0.10 |
| Open Claims | 109,355 | 401,048 | 73.2 | 298.0 | 464 | 3,205 | 0.81 (0.71 - 0.92) | <0.01 | 0.80 (0.70 - 0.92) | <0.01 |
| OEHR | 7,631 | 37,284 | 1.9 | 15.1 | 19 | 144 | 0.97 (0.50 - 1.77) | 0.92 | 0.84 (0.44 - 1.58) | 0.59 |
| VA | 1,153 | 17,084 | 0.7 | 11.8 | <10 | 46 | 0.37 (0.02 - 1.95) | 0.39 | 0.28 (0.03 - 2.77) | 0.28 |
| Meta-analysis | 134,118 | 526,299 | 85.1 | 376.3 | 569 | 4,215 | 0.79 (0.54 - 1.14) | 0.20 | 0.77 (0.53 - 1.12) | 0.17 |
| Meta-analysis (w/o USOC) | 24,763 | 125,251 | 11.9 | 78.3 | 105 | 1,010 | 0.76 (0.43 - 1.33) | 0.33 | 0.71 (0.40 - 1.25) | 0.24 |
| **PS-stratification / on-treatment TAR without escalation** | | | | | | | | | | |
| CCAE | 12,059 | 51,105 | 6.9 | 36.2 | 53 | 306 | 1.02 (0.73 - 1.41) | 0.89 | 0.99 (0.72 - 1.38) | 0.91 |
| ODOD | 8,175 | 29,196 | 4.2 | 20.0 | 35 | 390 | 0.50 (0.34 - 0.74) | <0.01 | 0.47 (0.32 - 0.70) | <0.01 |
| MDCR | 893 | 8,780 | 0.5 | 7.8 | 7 | 215 | 0.52 (0.21 - 1.08) | 0.11 | 0.47 (0.21 - 1.08) | 0.08 |
| Open Claims | 147,591 | 402,784 | 89.0 | 299.3 | 538 | 3,277 | 0.79 (0.71 - 0.88) | <0.01 | 0.81 (0.72 - 0.90) | <0.01 |
| OEHR | 8,896 | 37,369 | 2.2 | 15.1 | 22 | 147 | 1.08 (0.63 - 1.76) | 0.77 | 0.91 (0.55 - 1.52) | 0.73 |
| VA | 1,334 | 24,642 | 0.7 | 18.7 | <10 | 86 | 1.35 (0.43 - 3.53) | 0.58 | 1.13 (0.39 - 3.23) | 0.82 |
| Meta-analysis | 176,721 | 520,454 | 102.2 | 370.6 | 648 | 4,120 | 0.81 (0.60 - 1.09) | 0.16 | 0.80 (0.59 - 1.10) | 0.18 |
| Meta-analysis (w/o USOC) | 29,130 | 117,670 | 13.2 | 71.3 | 110 | 843 | 0.82 (0.50 - 1.33) | 0.42 | 0.77 (0.47 - 1.26) | 0.31 |

##### 1.4.2 4-point MACE

**Supplementary Table 80:** Exposures, outcomes and risk estimates for 4-point MACE in GLP1RA (male) (T) vs DPP4I (male) (C) new-user cohorts under four different propensity score (PS) adjustment / time-at-risk (TAR) designs across data sources. We report total patient exposure time in 1,000 patient-year (KYr) units, uncalibrated and calibrated hazard ratios (HRs) and their 95% confidence intervals (CIs). Greyed out entries did not pass study diagnostics and are included only for completeness.

| | Patients | | Exposure (KYr) | | Outcomes | | Uncalibrated | | Calibrated | |
|---|---|---|---|---|---|---|---|---|---|---|
| | T | C | T | C | T | C | HR (95% CI) | $p$ | HR (95% CI) | $p$ |
| **PS-matching / on-treatment TAR** | | | | | | | | | | |
| CCAE | 10,031 | 50,301 | 6.9 | 43.9 | 87 | 553 | 1.00 (0.73 - 1.35) | 0.99 | 0.97 (0.71 - 1.32) | 0.83 |
| ODOD | 5,924 | 27,592 | 3.7 | 23.1 | 66 | 650 | 0.72 (0.50 - 1.02) | 0.07 | 0.71 (0.49 - 1.01) | 0.07 |
| MDCR | 758 | 7,793 | 0.5 | 8.1 | 21 | 365 | 1.11 (0.64 - 1.84) | 0.69 | 1.07 (0.63 - 1.83) | 0.77 |
| Open Claims | 106,379 | 388,374 | 82.6 | 371.4 | 852 | 6,449 | 0.82 (0.75 - 0.91) | <0.01 | 0.83 (0.75 - 0.93) | <0.01 |
| OEHR | 7,520 | 36,606 | 2.2 | 18.0 | 28 | 361 | 0.64 (0.38 - 1.01) | 0.07 | 0.55 (0.34 - 0.91) | 0.02 |
| VA | 1,114 | 16,661 | 0.8 | 14.1 | <10 | 158 | 0.35 (0.08 - 1.03) | 0.11 | 0.29 (0.08 - 1.02) | 0.06 |
| Meta-analysis | 131,726 | 527,327 | 96.8 | 478.5 | <1,064 | 8,536 | 0.83 (0.76 - 0.90) | <0.01 | 0.82 (0.70 - 0.96) | 0.01 |
| Meta-analysis (w/o USOC) | 25,347 | 138,953 | 14.2 | 107.1 | <212 | 2,087 | 0.82 (0.63 - 1.06) | 0.13 | 0.79 (0.61 - 1.02) | 0.07 |
| **PS-stratification / on-treatment TAR** | | | | | | | | | | |
| CCAE | 11,840 | 50,414 | 7.7 | 44.0 | 96 | 553 | 0.96 (0.75 - 1.22) | 0.73 | 0.95 (0.74 - 1.21) | 0.67 |
| ODOD | 7,889 | 28,074 | 4.5 | 23.5 | 77 | 680 | 0.70 (0.53 - 0.91) | <0.01 | 0.68 (0.52 - 0.88) | <0.01 |
| MDCR | 847 | 8,212 | 0.5 | 8.5 | 22 | 407 | 0.95 (0.58 - 1.48) | 0.83 | 0.88 (0.55 - 1.41) | 0.61 |
| Open Claims | 143,665 | 389,938 | 99.2 | 372.9 | 977 | 6,586 | 0.80 (0.74 - 0.86) | <0.01 | 0.83 (0.76 - 0.90) | <0.01 |
| OEHR | 8,765 | 36,683 | 2.5 | 18.0 | 33 | 371 | 0.73 (0.48 - 1.06) | 0.11 | 0.64 (0.43 - 0.94) | 0.03 |
| VA | 1,281 | 24,006 | 0.9 | 22.3 | <10 | 268 | 0.43 (0.15 - 0.98) | 0.08 | 0.38 (0.15 - 0.97) | 0.05 |
| Meta-analysis | 172,159 | 505,109 | 113.9 | 458.4 | 1,183 | 8,190 | 0.80 (0.75 - 0.86) | <0.01 | 0.81 (0.70 - 0.94) | <0.01 |
| Meta-analysis (w/o USOC) | 28,494 | 115,171 | 14.7 | 85.5 | 206 | 1,604 | 0.80 (0.65 - 0.99) | 0.04 | 0.77 (0.62 - 0.96) | 0.03 |
| **PS-matching / on-treatment TAR without escalation** | | | | | | | | | | |
| CCAE | 10,031 | 50,301 | 6.0 | 35.5 | 77 | 441 | 1.08 (0.77 - 1.50) | 0.65 | 1.06 (0.76 - 1.49) | 0.74 |
| ODOD | 5,924 | 27,592 | 3.3 | 18.9 | 57 | 545 | 0.70 (0.46 - 1.03) | 0.08 | 0.67 (0.45 - 1.01) | 0.06 |
| MDCR | 758 | 7,793 | 0.5 | 6.9 | 17 | 310 | 0.98 (0.52 - 1.74) | 0.95 | 0.85 (0.46 - 1.55) | 0.60 |
| Open Claims | 106,379 | 388,374 | 71.3 | 288.2 | 718 | 5,009 | 0.82 (0.73 - 0.91) | <0.01 | 0.81 (0.72 - 0.91) | <0.01 |
| OEHR | 7,520 | 36,606 | 1.9 | 14.7 | 23 | 261 | 0.67 (0.38 - 1.11) | 0.14 | 0.58 (0.34 - 0.99) | 0.05 |
| VA | 1,114 | 16,661 | 0.6 | 11.5 | <10 | 118 | 0.53 (0.12 - 1.62) | 0.33 | 0.40 (0.11 - 1.48) | 0.17 |
| Meta-analysis | 131,726 | 527,327 | 83.6 | 375.7 | <902 | 6,684 | 0.82 (0.75 - 0.91) | <0.01 | 0.80 (0.71 - 0.91) | <0.01 |
| Meta-analysis (w/o USOC) | 25,347 | 138,953 | 12.3 | 87.5 | <184 | 1,675 | 0.85 (0.68 - 1.07) | 0.17 | 0.80 (0.64 - 1.01) | 0.07 |
| **PS-stratification / on-treatment TAR without escalation** | | | | | | | | | | |
| CCAE | 11,840 | 50,414 | 6.7 | 35.6 | 85 | 441 | 1.05 (0.80 - 1.36) | 0.71 | 1.02 (0.78 - 1.33) | 0.86 |
| ODOD | 7,889 | 28,074 | 4.0 | 19.2 | 67 | 573 | 0.68 (0.51 - 0.91) | <0.01 | 0.64 (0.48 - 0.86) | <0.01 |
| MDCR | 847 | 8,212 | 0.5 | 7.3 | 18 | 349 | 0.88 (0.51 - 1.45) | 0.65 | 0.81 (0.48 - 1.37) | 0.45 |
| Open Claims | 143,665 | 389,938 | 86.8 | 289.4 | 830 | 5,120 | 0.79 (0.73 - 0.86) | <0.01 | 0.81 (0.74 - 0.89) | <0.01 |
| OEHR | 8,765 | 36,683 | 2.2 | 14.8 | 28 | 267 | 0.79 (0.50 - 1.20) | 0.29 | 0.67 (0.43 - 1.03) | 0.08 |
| VA | 1,281 | 24,006 | 0.7 | 18.2 | <10 | 197 | 0.60 (0.20 - 1.41) | 0.30 | 0.50 (0.19 - 1.32) | 0.16 |
| Meta-analysis | 172,159 | 505,109 | 99.7 | 359.0 | 1,010 | 6,401 | 0.82 (0.70 - 0.96) | 0.02 | 0.82 (0.68 - 0.98) | 0.03 |
| Meta-analysis (w/o USOC) | 28,494 | 115,171 | 12.9 | 69.6 | 180 | 1,281 | 0.84 (0.63 - 1.11) | 0.22 | 0.79 (0.59 - 1.06) | 0.12 |

##### 1.4.3 Acute myocardial infarction

**Supplementary Table 81:** Exposures, outcomes and risk estimates for Acute myocardial infarction in GLP1RA (male) (T) vs DPP4I (male) (C) new-user cohorts under four different propensity score (PS) adjustment / time-at-risk (TAR) designs across data sources. We report total patient exposure time in 1,000 patient-year (KYr) units, uncalibrated and calibrated hazard ratios (HRs) and their 95% confidence intervals (CIs). Greyed out entries did not pass study diagnostics and are included only for completeness.

| | Patients | | Exposure (KYr) | | Outcomes | | Uncalibrated | | Calibrated | |
|---|---|---|---|---|---|---|---|---|---|---|
| | T | C | T | C | T | C | HR (95% CI) | $p$ | HR (95% CI) | $p$ |
| **PS-matching / on-treatment TAR** | | | | | | | | | | |
| CCAE | 10,327 | 51,541 | 7.2 | 45.1 | 35 | 230 | 0.93 (0.56 - 1.51) | 0.79 | 0.90 (0.55 - 1.49) | 0.70 |
| ODOD | 6,255 | 29,323 | 3.9 | 24.7 | 17 | 220 | 0.39 (0.18 - 0.77) | 0.01 | 0.38 (0.18 - 0.80) | 0.01 |
| MDCR | 826 | 8,574 | 0.6 | 9.0 | 7 | 107 | 0.80 (0.26 - 2.03) | 0.67 | 0.77 (0.28 - 2.17) | 0.63 |
| Open Claims | 111,826 | 412,257 | 86.5 | 395.3 | 301 | 2,145 | 0.82 (0.70 - 0.97) | 0.02 | 0.83 (0.70 - 0.98) | 0.05 |
| OEHR | 7,690 | 37,619 | 2.3 | 18.8 | 7 | 113 | 0.64 (0.24 - 1.47) | 0.34 | 0.56 (0.22 - 1.40) | 0.22 |
| VA | 1,172 | 17,282 | 0.9 | 14.7 | <10 | 35 | 0.84 (0.12 - 3.37) | 0.83 | 0.68 (0.13 - 3.54) | 0.65 |
| Meta-analysis | 136,924 | 539,314 | 100.5 | 492.9 | 367 | 2,815 | 0.80 (0.69 - 0.93) | <0.01 | 0.79 (0.65 - 0.96) | 0.02 |
| Meta-analysis (w/o USOC) | 25,098 | 127,057 | 13.9 | 97.6 | 66 | 670 | 0.69 (0.45 - 1.04) | 0.08 | 0.66 (0.43 - 1.00) | 0.05 |
| **PS-stratification / on-treatment TAR** | | | | | | | | | | |
| CCAE | 12,209 | 51,682 | 8.0 | 45.2 | 38 | 231 | 1.01 (0.68 - 1.47) | 0.96 | 1.00 (0.68 - 1.47) | 0.93 |
| ODOD | 8,318 | 29,940 | 4.8 | 25.2 | 19 | 235 | 0.48 (0.27 - 0.78) | <0.01 | 0.46 (0.27 - 0.78) | <0.01 |
| MDCR | 923 | 9,130 | 0.6 | 9.6 | 7 | 120 | 0.97 (0.38 - 2.13) | 0.95 | 0.90 (0.38 - 2.14) | 0.82 |
| Open Claims | 150,626 | 414,162 | 103.8 | 397.0 | 346 | 2,192 | 0.84 (0.74 - 0.96) | <0.01 | 0.87 (0.76 - 1.00) | 0.09 |
| OEHR | 8,966 | 37,708 | 2.6 | 18.8 | 10 | 116 | 0.73 (0.34 - 1.39) | 0.37 | 0.63 (0.31 - 1.29) | 0.21 |
| VA | 1,360 | 25,018 | 0.9 | 23.4 | <10 | 65 | 1.94 (0.59 - 5.35) | 0.24 | 1.70 (0.57 - 5.13) | 0.34 |
| Meta-analysis | 180,119 | 533,492 | 119.1 | 486.3 | 413 | 2,774 | 0.79 (0.60 - 1.03) | 0.08 | 0.79 (0.59 - 1.07) | 0.13 |
| Meta-analysis (w/o USOC) | 29,493 | 119,330 | 15.3 | 89.3 | 67 | 582 | 0.72 (0.44 - 1.17) | 0.19 | 0.69 (0.42 - 1.13) | 0.15 |
| **PS-matching / on-treatment TAR without escalation** | | | | | | | | | | |
| CCAE | 10,327 | 51,541 | 6.2 | 36.5 | 32 | 181 | 1.19 (0.68 - 2.01) | 0.53 | 1.17 (0.68 - 2.01) | 0.58 |
| ODOD | 6,255 | 29,323 | 3.5 | 20.2 | 14 | 178 | 0.37 (0.15 - 0.78) | 0.02 | 0.35 (0.15 - 0.81) | 0.01 |
| MDCR | 826 | 8,574 | 0.5 | 7.7 | <5 | 88 | 0.68 (0.15 - 2.14) | 0.57 | 0.59 (0.16 - 2.20) | 0.44 |
| Open Claims | 111,826 | 412,257 | 74.7 | 306.4 | 251 | 1,648 | 0.82 (0.68 - 0.98) | 0.04 | 0.82 (0.68 - 0.98) | 0.04 |
| OEHR | 7,690 | 37,619 | 2.0 | 15.3 | 6 | 76 | 0.69 (0.23 - 1.71) | 0.47 | 0.59 (0.21 - 1.63) | 0.32 |
| VA | 1,172 | 17,282 | 0.7 | 12.0 | <10 | 26 | 1.15 (0.17 - 4.94) | 0.87 | 0.87 (0.16 - 4.78) | 0.88 |
| Meta-analysis | 136,924 | 539,314 | 86.8 | 386.1 | <308 | 2,171 | 0.79 (0.58 - 1.09) | 0.15 | 0.77 (0.56 - 1.07) | 0.12 |
| Meta-analysis (w/o USOC) | 25,098 | 127,057 | 12.1 | 79.7 | <57 | 523 | 0.71 (0.39 - 1.29) | 0.27 | 0.67 (0.37 - 1.22) | 0.19 |
| **PS-stratification / on-treatment TAR without escalation** | | | | | | | | | | |
| CCAE | 12,209 | 51,682 | 7.0 | 36.6 | 35 | 182 | 1.17 (0.76 - 1.74) | 0.46 | 1.14 (0.75 - 1.72) | 0.55 |
| ODOD | 8,318 | 29,940 | 4.2 | 20.6 | 16 | 190 | 0.43 (0.23 - 0.74) | <0.01 | 0.40 (0.23 - 0.71) | <0.01 |
| MDCR | 923 | 9,130 | 0.5 | 8.2 | <5 | 100 | 0.59 (0.17 - 1.60) | 0.36 | 0.54 (0.17 - 1.67) | 0.29 |
| Open Claims | 150,626 | 414,162 | 90.8 | 307.8 | 289 | 1,685 | 0.83 (0.72 - 0.96) | 0.01 | 0.85 (0.73 - 0.98) | 0.04 |
| OEHR | 8,966 | 37,708 | 2.2 | 15.3 | 9 | 78 | 0.82 (0.36 - 1.67) | 0.61 | 0.69 (0.32 - 1.50) | 0.36 |
| VA | 1,360 | 25,018 | 0.7 | 19.0 | <10 | 50 | 2.71 (0.79 - 8.09) | 0.09 | 2.27 (0.71 - 7.27) | 0.17 |
| Meta-analysis | 180,119 | 533,492 | 104.2 | 380.3 | 349 | 2,135 | 0.79 (0.54 - 1.16) | 0.23 | 0.79 (0.53 - 1.17) | 0.24 |
| Meta-analysis (w/o USOC) | 29,493 | 119,330 | 13.4 | 72.5 | 60 | 450 | 0.76 (0.39 - 1.44) | 0.40 | 0.72 (0.37 - 1.37) | 0.32 |

##### 1.4.4 Acute renal failure

**Supplementary Table 82:** Exposures, outcomes and risk estimates for Acute renal failure in GLP1RA (male) (T) vs DPP4I (male) (C) new-user cohorts under four different propensity score (PS) adjustment / time-at-risk (TAR) designs across data sources. We report total patient exposure time in 1,000 patient-year (KYr) units, uncalibrated and calibrated hazard ratios (HRs) and their 95% confidence intervals (CIs). Greyed out entries did not pass study diagnostics and are included only for completeness.

| | Patients | | Exposure (KYr) | | Outcomes | | Uncalibrated | | Calibrated | |
|---|---|---|---|---|---|---|---|---|---|---|
| | T | C | T | C | T | C | HR (95% CI) | $p$ | HR (95% CI) | $p$ |
| **PS-matching / on-treatment TAR** | | | | | | | | | | |
| CCAE | 10,390 | 51,877 | 7.2 | 45.5 | 54 | 385 | 0.64 (0.43 - 0.93) | 0.02 | 0.62 (0.42 - 0.91) | 0.02 |
| ODOD | 6,146 | 28,625 | 3.8 | 24.0 | 76 | 624 | 0.65 (0.46 - 0.91) | 0.01 | 0.64 (0.46 - 0.91) | 0.01 |
| MDCR | 816 | 8,402 | 0.6 | 8.7 | 23 | 301 | 1.01 (0.55 - 1.73) | 0.98 | 0.97 (0.55 - 1.73) | 0.85 |
| Open Claims | 111,443 | 410,208 | 86.2 | 392.3 | 773 | 5,862 | 0.78 (0.70 - 0.86) | <0.01 | 0.79 (0.71 - 0.88) | <0.01 |
| OEHR | 7,691 | 37,538 | 2.3 | 18.6 | 30 | 334 | 0.89 (0.55 - 1.39) | 0.63 | 0.78 (0.49 - 1.24) | 0.31 |
| VA | 1,157 | 17,032 | 0.9 | 14.4 | <10 | 181 | 0.80 (0.32 - 1.72) | 0.60 | 0.65 (0.28 - 1.52) | 0.34 |
| Meta-analysis | 137,643 | 553,682 | 100.9 | 503.5 | <966 | 7,687 | 0.77 (0.70 - 0.84) | <0.01 | 0.76 (0.65 - 0.89) | <0.01 |
| Meta-analysis (w/o USOC) | 26,200 | 143,474 | 14.7 | 111.2 | <193 | 1,825 | 0.73 (0.60 - 0.90) | <0.01 | 0.71 (0.58 - 0.87) | <0.01 |
| **PS-stratification / on-treatment TAR** | | | | | | | | | | |
| CCAE | 12,261 | 51,990 | 8.0 | 45.6 | 61 | 388 | 0.67 (0.49 - 0.89) | <0.01 | 0.66 (0.48 - 0.89) | <0.01 |
| ODOD | 8,198 | 29,178 | 4.7 | 24.5 | 91 | 659 | 0.74 (0.57 - 0.95) | 0.02 | 0.72 (0.55 - 0.93) | 0.01 |
| MDCR | 910 | 8,907 | 0.6 | 9.2 | 23 | 339 | 1.02 (0.62 - 1.61) | 0.92 | 0.95 (0.59 - 1.53) | 0.81 |
| Open Claims | 150,166 | 411,910 | 103.4 | 394.0 | 898 | 5,966 | 0.74 (0.69 - 0.81) | <0.01 | 0.77 (0.71 - 0.84) | <0.01 |
| OEHR | 8,969 | 37,635 | 2.6 | 18.6 | 34 | 341 | 0.81 (0.54 - 1.18) | 0.30 | 0.71 (0.48 - 1.05) | 0.10 |
| VA | 1,342 | 24,628 | 0.9 | 22.9 | <10 | 295 | 0.73 (0.32 - 1.46) | 0.42 | 0.64 (0.30 - 1.38) | 0.27 |
| Meta-analysis | 179,594 | 530,713 | 118.7 | 482.7 | 1,084 | 7,354 | 0.74 (0.69 - 0.80) | <0.01 | 0.75 (0.64 - 0.87) | <0.01 |
| Meta-analysis (w/o USOC) | 29,428 | 118,803 | 15.2 | 88.7 | 186 | 1,388 | 0.73 (0.61 - 0.87) | <0.01 | 0.70 (0.58 - 0.84) | <0.01 |
| **PS-matching / on-treatment TAR without escalation** | | | | | | | | | | |
| CCAE | 10,390 | 51,877 | 6.2 | 36.8 | 46 | 304 | 0.65 (0.41 - 0.98) | 0.05 | 0.63 (0.41 - 0.97) | 0.04 |
| ODOD | 6,146 | 28,625 | 3.4 | 19.6 | 68 | 499 | 0.71 (0.49 - 1.00) | 0.06 | 0.68 (0.47 - 0.97) | 0.04 |
| MDCR | 816 | 8,402 | 0.5 | 7.5 | 19 | 249 | 0.83 (0.41 - 1.53) | 0.57 | 0.72 (0.37 - 1.38) | 0.33 |
| Open Claims | 111,443 | 410,208 | 74.4 | 304.4 | 639 | 4,547 | 0.75 (0.66 - 0.84) | <0.01 | 0.74 (0.66 - 0.83) | <0.01 |
| OEHR | 7,691 | 37,538 | 2.0 | 15.2 | 25 | 229 | 0.91 (0.53 - 1.49) | 0.71 | 0.78 (0.47 - 1.31) | 0.37 |
| VA | 1,157 | 17,032 | 0.7 | 11.8 | <10 | 132 | 0.97 (0.35 - 2.32) | 0.95 | 0.74 (0.29 - 1.92) | 0.54 |
| Meta-analysis | 137,643 | 553,682 | 87.1 | 395.2 | <807 | 5,960 | 0.75 (0.67 - 0.83) | <0.01 | 0.73 (0.64 - 0.83) | <0.01 |
| Meta-analysis (w/o USOC) | 26,200 | 143,474 | 12.7 | 90.9 | <168 | 1,413 | 0.75 (0.60 - 0.93) | 0.01 | 0.70 (0.56 - 0.88) | <0.01 |
| **PS-stratification / on-treatment TAR without escalation** | | | | | | | | | | |
| CCAE | 12,261 | 51,990 | 7.0 | 36.9 | 53 | 307 | 0.69 (0.49 - 0.95) | 0.03 | 0.67 (0.48 - 0.93) | 0.02 |
| ODOD | 8,198 | 29,178 | 4.2 | 20.0 | 80 | 531 | 0.75 (0.56 - 0.98) | 0.04 | 0.70 (0.53 - 0.92) | 0.02 |
| MDCR | 910 | 8,907 | 0.5 | 7.9 | 19 | 283 | 0.97 (0.56 - 1.60) | 0.91 | 0.89 (0.53 - 1.51) | 0.67 |
| Open Claims | 150,166 | 411,910 | 90.4 | 305.7 | 753 | 4,632 | 0.72 (0.66 - 0.79) | <0.01 | 0.74 (0.67 - 0.81) | <0.01 |
| OEHR | 8,969 | 37,635 | 2.2 | 15.2 | 29 | 234 | 0.87 (0.55 - 1.31) | 0.51 | 0.73 (0.47 - 1.13) | 0.17 |
| VA | 1,342 | 24,628 | 0.7 | 18.6 | <10 | 212 | 0.96 (0.39 - 2.04) | 0.92 | 0.80 (0.35 - 1.84) | 0.61 |
| Meta-analysis | 179,594 | 530,713 | 103.8 | 377.8 | 915 | 5,704 | 0.73 (0.67 - 0.79) | <0.01 | 0.73 (0.64 - 0.82) | <0.01 |
| Meta-analysis (w/o USOC) | 29,428 | 118,803 | 13.4 | 72.1 | 162 | 1,072 | 0.75 (0.62 - 0.90) | <0.01 | 0.71 (0.58 - 0.86) | <0.01 |

##### 1.4.5 Glycemic control

**Supplementary Table 83:** Exposures, outcomes and risk estimates for Glycemic control in GLP1RA (male) (T) vs DPP4I (male) (C) new-user cohorts under four different propensity score (PS) adjustment / time-at-risk (TAR) designs across data sources. We report total patient exposure time in 1,000 patient-year (KYr) units, uncalibrated and calibrated hazard ratios (HRs) and their 95% confidence intervals (CIs). Greyed out entries did not pass study diagnostics and are included only for completeness.

| | Patients | | Exposure (KYr) | | Outcomes | | Uncalibrated | | Calibrated | |
|---|---|---|---|---|---|---|---|---|---|---|
| | T | C | T | C | T | C | HR (95% CI) | $p$ | HR (95% CI) | $p$ |
| **PS-matching / on-treatment TAR** | | | | | | | | | | |
| CCAE | 10,544 | 52,228 | 7.3 | 45.9 | 6 | 130 | 0.68 (0.23 - 1.67) | 0.45 | 0.66 (0.24 - 1.80) | 0.42 |
| ODOD | 4,318 | 20,609 | 2.4 | 15.5 | 277 | 1,651 | 1.23 (1.02 - 1.47) | 0.02 | 1.21 (1.01 - 1.45) | 0.07 |
| MDCR | 855 | 8,869 | 0.6 | 9.4 | <5 | 11 | 3.72 (0.16 - 28.9) | 0.32 | 3.60 (0.27 - 47.7) | 0.33 |
| Open Claims | 115,660 | 427,485 | 89.4 | 411.0 | 0 | 0 | | | | |
| OEHR | 7,806 | 38,195 | 2.3 | 19.2 | 0 | 0 | | | | |
| VA | 1,158 | 16,768 | 0.9 | 14.2 | 0 | 12 | | | | |
| Meta-analysis | 14,862 | 72,837 | 9.7 | 61.4 | 283 | 1,781 | 1.14 (0.77 - 1.68) | 0.53 | 1.12 (0.74 - 1.69) | 0.59 |
| Meta-analysis (w/o USOC) | 14,862 | 72,837 | 9.7 | 61.4 | 283 | 1,781 | 1.14 (0.77 - 1.68) | 0.53 | 1.09 (0.74 - 1.61) | 0.67 |
| **PS-stratification / on-treatment TAR** | | | | | | | | | | |
| CCAE | 12,456 | 52,351 | 8.1 | 46.0 | 6 | 131 | 0.52 (0.20 - 1.12) | 0.14 | 0.51 (0.21 - 1.22) | 0.13 |
| ODOD | 5,649 | 20,906 | 3.0 | 15.7 | 329 | 1,677 | 1.22 (1.06 - 1.40) | <0.01 | 1.18 (1.02 - 1.36) | 0.04 |
| MDCR | 952 | 9,515 | 0.6 | 10.0 | <5 | 12 | 2.22 (0.12 - 12.6) | 0.50 | 2.06 (0.20 - 21.3) | 0.54 |
| Open Claims | 155,612 | 429,807 | 107.2 | 413.1 | 0 | 0 | | | | |
| OEHR | 9,105 | 38,288 | 2.6 | 19.2 | 0 | 0 | | | | |
| VA | 1,341 | 24,844 | 0.9 | 23.3 | 0 | 21 | | | | |
| Meta-analysis | 18,105 | 73,257 | 11.1 | 61.7 | 335 | 1,808 | 0.89 (0.39 - 2.00) | 0.78 | 0.90 (0.39 - 2.05) | 0.80 |
| Meta-analysis (w/o USOC) | 18,105 | 73,257 | 11.1 | 61.7 | 335 | 1,808 | 0.89 (0.39 - 2.00) | 0.78 | 0.85 (0.38 - 1.93) | 0.71 |
| **PS-matching / on-treatment TAR without escalation** | | | | | | | | | | |
| CCAE | 10,544 | 52,228 | 6.3 | 37.0 | 5 | 107 | 0.55 (0.13 - 1.61) | 0.35 | 0.54 (0.15 - 1.90) | 0.34 |
| ODOD | 4,318 | 20,609 | 2.1 | 12.7 | 249 | 1,340 | 1.33 (1.10 - 1.61) | <0.01 | 1.27 (1.05 - 1.55) | 0.03 |
| MDCR | 855 | 8,869 | 0.5 | 8.0 | <5 | 10 | 3.51 (0.14 - 32.6) | 0.37 | 3.03 (0.20 - 46.5) | 0.43 |
| Open Claims | 115,660 | 427,485 | 77.1 | 318.3 | 0 | 0 | | | | |
| OEHR | 7,806 | 38,195 | 2.0 | 15.5 | 0 | 0 | | | | |
| VA | 1,158 | 16,768 | 0.7 | 11.6 | 0 | 11 | | | | |
| Meta-analysis | 14,862 | 72,837 | 8.5 | 49.7 | 254 | 1,447 | 1.07 (0.51 - 2.26) | 0.85 | 1.05 (0.49 - 2.22) | 0.90 |
| Meta-analysis (w/o USOC) | 14,862 | 72,837 | 8.5 | 49.7 | 254 | 1,447 | 1.07 (0.51 - 2.26) | 0.85 | 1.01 (0.48 - 2.13) | 0.96 |
| **PS-stratification / on-treatment TAR without escalation** | | | | | | | | | | |
| CCAE | 12,456 | 52,351 | 7.1 | 37.1 | 5 | 108 | 0.51 (0.17 - 1.20) | 0.17 | 0.50 (0.19 - 1.31) | 0.16 |
| ODOD | 5,649 | 20,906 | 2.6 | 12.9 | 297 | 1,360 | 1.30 (1.12 - 1.51) | <0.01 | 1.22 (1.05 - 1.42) | 0.02 |
| MDCR | 952 | 9,515 | 0.6 | 8.5 | <5 | 11 | 2.84 (0.15 - 16.7) | 0.39 | 2.60 (0.25 - 27.6) | 0.43 |
| Open Claims | 155,612 | 429,807 | 93.6 | 320.0 | 0 | 0 | | | | |
| OEHR | 9,105 | 38,288 | 2.3 | 15.6 | 0 | 0 | | | | |
| VA | 1,341 | 24,844 | 0.7 | 18.9 | 0 | 18 | | | | |
| Meta-analysis | 18,105 | 73,257 | 9.7 | 50.0 | 302 | 1,468 | 0.93 (0.38 - 2.25) | 0.86 | 0.92 (0.38 - 2.25) | 0.86 |
| Meta-analysis (w/o USOC) | 18,105 | 73,257 | 9.7 | 50.0 | 302 | 1,468 | 0.93 (0.38 - 2.25) | 0.86 | 0.88 (0.36 - 2.13) | 0.77 |

##### 1.4.6 Hospitalization with heart failure

**Supplementary Table 84:** Exposures, outcomes and risk estimates for Hospitalization with heart failure in GLP1RA (male) (T) vs DPP4I (male) (C) new-user cohorts under four different propensity score (PS) adjustment / time-at-risk (TAR) designs across data sources. We report total patient exposure time in 1,000 patient-year (KYr) units, uncalibrated and calibrated hazard ratios (HRs) and their 95% confidence intervals (CIs). Greyed out entries did not pass study diagnostics and are included only for completeness.

| | Patients | | Exposure (KYr) | | Outcomes | | Uncalibrated | | Calibrated | |
|---|---|---|---|---|---|---|---|---|---|---|
| | T | C | T | C | T | C | HR (95% CI) | $p$ | HR (95% CI) | $p$ |
| **PS-matching / on-treatment TAR** | | | | | | | | | | |
| CCAE | 10,377 | 51,798 | 7.2 | 45.4 | 44 | 254 | 0.88 (0.55 - 1.37) | 0.58 | 0.85 (0.54 - 1.34) | 0.50 |
| ODOD | 6,125 | 28,604 | 3.8 | 24.1 | 46 | 421 | 0.84 (0.55 - 1.26) | 0.41 | 0.83 (0.55 - 1.25) | 0.39 |
| MDCR | 798 | 8,221 | 0.6 | 8.6 | 16 | 247 | 1.37 (0.70 - 2.51) | 0.33 | 1.33 (0.70 - 2.52) | 0.40 |
| Open Claims | 111,489 | 409,329 | 86.4 | 392.5 | 472 | 4,073 | 0.76 (0.67 - 0.86) | <0.01 | 0.77 (0.67 - 0.88) | <0.01 |
| OEHR | 7,657 | 37,350 | 2.3 | 18.5 | 14 | 232 | 0.50 (0.25 - 0.93) | 0.04 | 0.44 (0.23 - 0.85) | 0.02 |
| VA | 1,160 | 17,097 | 0.9 | 14.5 | <10 | 117 | 0.37 (0.06 - 1.30) | 0.21 | 0.30 (0.06 - 1.43) | 0.14 |
| Meta-analysis | 137,606 | 552,399 | 101.0 | 503.6 | <602 | 5,344 | 0.78 (0.69 - 0.87) | <0.01 | 0.77 (0.64 - 0.91) | <0.01 |
| Meta-analysis (w/o USOC) | 26,117 | 143,070 | 14.7 | 111.1 | <130 | 1,271 | 0.82 (0.60 - 1.13) | 0.23 | 0.79 (0.58 - 1.09) | 0.16 |
| **PS-stratification / on-treatment TAR** | | | | | | | | | | |
| CCAE | 12,239 | 51,915 | 8.0 | 45.6 | 52 | 256 | 0.96 (0.68 - 1.33) | 0.80 | 0.94 (0.67 - 1.33) | 0.75 |
| ODOD | 8,142 | 29,193 | 4.7 | 24.6 | 55 | 448 | 0.76 (0.54 - 1.04) | 0.09 | 0.73 (0.53 - 1.01) | 0.07 |
| MDCR | 889 | 8,741 | 0.6 | 9.1 | 17 | 284 | 1.14 (0.64 - 1.89) | 0.63 | 1.06 (0.62 - 1.82) | 0.82 |
| Open Claims | 150,190 | 411,035 | 103.6 | 394.2 | 538 | 4,180 | 0.73 (0.66 - 0.81) | <0.01 | 0.76 (0.68 - 0.84) | <0.01 |
| OEHR | 8,933 | 37,434 | 2.5 | 18.5 | 17 | 239 | 0.62 (0.35 - 1.02) | 0.08 | 0.54 (0.32 - 0.92) | 0.03 |
| VA | 1,339 | 24,760 | 0.9 | 23.1 | <10 | 201 | 0.34 (0.08 - 0.94) | 0.08 | 0.30 (0.09 - 1.01) | 0.05 |
| Meta-analysis | 179,504 | 529,577 | 118.8 | 482.8 | 662 | 5,123 | 0.75 (0.68 - 0.82) | <0.01 | 0.75 (0.64 - 0.89) | <0.01 |
| Meta-analysis (w/o USOC) | 29,314 | 118,542 | 15.2 | 88.7 | 124 | 943 | 0.80 (0.64 - 1.00) | 0.05 | 0.77 (0.61 - 0.97) | 0.04 |
| **PS-matching / on-treatment TAR without escalation** | | | | | | | | | | |
| CCAE | 10,377 | 51,798 | 6.2 | 36.7 | 40 | 207 | 0.92 (0.56 - 1.48) | 0.75 | 0.90 (0.55 - 1.47) | 0.69 |
| ODOD | 6,125 | 28,604 | 3.4 | 19.7 | 43 | 349 | 0.96 (0.61 - 1.48) | 0.86 | 0.92 (0.59 - 1.44) | 0.73 |
| MDCR | 798 | 8,221 | 0.5 | 7.4 | 13 | 211 | 1.27 (0.58 - 2.51) | 0.52 | 1.09 (0.53 - 2.28) | 0.80 |
| Open Claims | 111,489 | 409,329 | 74.5 | 304.3 | 403 | 3,168 | 0.79 (0.68 - 0.91) | <0.01 | 0.78 (0.68 - 0.91) | <0.01 |
| OEHR | 7,657 | 37,350 | 2.0 | 15.1 | 13 | 172 | 0.64 (0.31 - 1.21) | 0.20 | 0.55 (0.28 - 1.10) | 0.10 |
| VA | 1,160 | 17,097 | 0.7 | 11.8 | <10 | 94 | 0.64 (0.10 - 2.41) | 0.58 | 0.49 (0.10 - 2.44) | 0.38 |
| Meta-analysis | 137,606 | 552,399 | 87.2 | 395.0 | <522 | 4,201 | 0.81 (0.72 - 0.92) | <0.01 | 0.79 (0.69 - 0.92) | <0.01 |
| Meta-analysis (w/o USOC) | 26,117 | 143,070 | 12.7 | 90.7 | <119 | 1,033 | 0.91 (0.70 - 1.20) | 0.52 | 0.86 (0.65 - 1.13) | 0.29 |
| **PS-stratification / on-treatment TAR without escalation** | | | | | | | | | | |
| CCAE | 12,239 | 51,915 | 7.0 | 36.8 | 47 | 209 | 1.08 (0.75 - 1.54) | 0.68 | 1.05 (0.73 - 1.51) | 0.79 |
| ODOD | 8,142 | 29,193 | 4.1 | 20.1 | 51 | 374 | 0.81 (0.57 - 1.13) | 0.23 | 0.76 (0.54 - 1.07) | 0.13 |
| MDCR | 889 | 8,741 | 0.5 | 7.8 | 14 | 246 | 1.04 (0.55 - 1.83) | 0.89 | 0.95 (0.52 - 1.74) | 0.85 |
| Open Claims | 150,190 | 411,035 | 90.5 | 305.6 | 463 | 3,260 | 0.75 (0.67 - 0.84) | <0.01 | 0.77 (0.68 - 0.86) | <0.01 |
| OEHR | 8,933 | 37,434 | 2.2 | 15.1 | 16 | 176 | 0.75 (0.41 - 1.28) | 0.32 | 0.63 (0.36 - 1.12) | 0.12 |
| VA | 1,339 | 24,760 | 0.7 | 18.8 | <10 | 158 | 0.42 (0.10 - 1.22) | 0.18 | 0.36 (0.10 - 1.24) | 0.11 |
| Meta-analysis | 179,504 | 529,577 | 103.9 | 377.6 | 577 | 4,019 | 0.81 (0.68 - 0.95) | <0.01 | 0.80 (0.67 - 0.97) | 0.02 |
| Meta-analysis (w/o USOC) | 29,314 | 118,542 | 13.3 | 72.0 | 114 | 759 | 0.90 (0.71 - 1.13) | 0.35 | 0.85 (0.68 - 1.07) | 0.18 |

##### 1.4.7 Measured renal dysfunction

**Supplementary Table 85:** Exposures, outcomes and risk estimates for Measured renal dysfunction in GLP1RA (male) (T) vs DPP4I (male) (C) new-user cohorts under four different propensity score (PS) adjustment / time-at-risk (TAR) designs across data sources. We report total patient exposure time in 1,000 patient-year (KYr) units, uncalibrated and calibrated hazard ratios (HRs) and their 95% confidence intervals (CIs). Greyed out entries did not pass study diagnostics and are included only for completeness.

| | Patients | | Exposure (KYr) | | Outcomes | | Uncalibrated | | Calibrated | |
|---|---|---|---|---|---|---|---|---|---|---|
| | T | C | T | C | T | C | HR (95% CI) | $p$ | HR (95% CI) | $p$ |
| **PS-matching / on-treatment TAR** | | | | | | | | | | |
| CCAE | 10,598 | 52,681 | 7.3 | 46.3 | 0 | 16 | | | | |
| ODOD | 6,403 | 30,170 | 4.0 | 25.5 | <5 | 46 | 0.70 (0.15 - 2.37) | 0.61 | 0.68 (0.17 - 2.72) | 0.59 |
| MDCR | 856 | 8,910 | 0.6 | 9.4 | 0 | <5 | | | | |
| Open Claims | 115,660 | 427,485 | 89.4 | 411.0 | 0 | 0 | | | | |
| OEHR | 7,806 | 38,195 | 2.3 | 19.2 | 0 | 0 | | | | |
| VA | 1,194 | 17,479 | 0.9 | 14.8 | <10 | 69 | 1.55 (0.42 - 4.49) | 0.47 | 1.27 (0.39 - 4.14) | 0.70 |
| Meta-analysis | 7,597 | 47,649 | 4.9 | 40.3 | <15 | 115 | 1.11 (0.45 - 2.71) | 0.83 | 1.09 (0.44 - 2.71) | 0.85 |
| Meta-analysis (w/o USOC) | 7,597 | 47,649 | 4.9 | 40.3 | <15 | 115 | 1.11 (0.45 - 2.71) | 0.83 | 1.06 (0.43 - 2.61) | 0.90 |
| **PS-stratification / on-treatment TAR** | | | | | | | | | | |
| CCAE | 12,522 | 52,816 | 8.2 | 46.4 | 0 | 17 | | | | |
| ODOD | 8,508 | 30,791 | 4.9 | 26.0 | 6 | 54 | 0.67 (0.24 - 1.62) | 0.41 | 0.65 (0.25 - 1.69) | 0.38 |
| MDCR | 953 | 9,568 | 0.6 | 10.0 | 0 | <5 | | | | |
| Open Claims | 155,612 | 429,807 | 107.2 | 413.1 | 0 | 0 | | | | |
| OEHR | 9,105 | 38,288 | 2.6 | 19.2 | 0 | 0 | | | | |
| VA | 1,388 | 25,292 | 1.0 | 23.6 | <10 | 98 | 1.25 (0.41 - 3.06) | 0.67 | 1.10 (0.40 - 3.00) | 0.85 |
| Meta-analysis | 8,508 | 30,791 | 4.9 | 26.0 | 6 | 54 | 0.67 (0.24 - 1.62) | 0.41 | 0.68 (0.26 - 1.79) | 0.43 |
| Meta-analysis (w/o USOC) | 8,508 | 30,791 | 4.9 | 26.0 | 6 | 54 | 0.67 (0.24 - 1.62) | 0.41 | 0.64 (0.25 - 1.69) | 0.37 |
| **PS-matching / on-treatment TAR without escalation** | | | | | | | | | | |
| CCAE | 10,598 | 52,681 | 6.3 | 37.4 | 0 | 12 | | | | |
| ODOD | 6,403 | 30,170 | 3.5 | 20.8 | <5 | 40 | 0.50 (0.07 - 1.98) | 0.41 | 0.48 (0.09 - 2.47) | 0.38 |
| MDCR | 856 | 8,910 | 0.5 | 8.0 | 0 | 0 | | | | |
| Open Claims | 115,660 | 427,485 | 77.1 | 318.3 | 0 | 0 | | | | |
| OEHR | 7,806 | 38,195 | 2.0 | 15.5 | 0 | 0 | | | | |
| VA | 1,194 | 17,479 | 0.7 | 12.1 | <10 | 54 | 2.19 (0.57 - 6.76) | 0.21 | 1.67 (0.48 - 5.77) | 0.42 |
| Meta-analysis | 6,403 | 30,170 | 3.5 | 20.8 | <5 | 40 | 0.50 (0.07 - 1.98) | 0.41 | 0.49 (0.09 - 2.52) | 0.39 |
| Meta-analysis (w/o USOC) | 6,403 | 30,170 | 3.5 | 20.8 | <5 | 40 | 0.50 (0.07 - 1.98) | 0.41 | 0.47 (0.09 - 2.42) | 0.37 |
| **PS-stratification / on-treatment TAR without escalation** | | | | | | | | | | |
| CCAE | 12,522 | 52,816 | 7.1 | 37.5 | 0 | 13 | | | | |
| ODOD | 8,508 | 30,791 | 4.3 | 21.2 | <5 | 48 | 0.52 (0.15 - 1.44) | 0.27 | 0.49 (0.16 - 1.53) | 0.22 |
| MDCR | 953 | 9,568 | 0.6 | 8.6 | 0 | 0 | | | | |
| Open Claims | 155,612 | 429,807 | 93.6 | 320.0 | 0 | 0 | | | | |
| OEHR | 9,105 | 38,288 | 2.3 | 15.6 | 0 | 0 | | | | |
| VA | 1,388 | 25,292 | 0.8 | 19.2 | <10 | 77 | 1.52 (0.43 - 4.14) | 0.47 | 1.27 (0.41 - 3.97) | 0.68 |
| Meta-analysis | 8,508 | 30,791 | 4.3 | 21.2 | <5 | 48 | 0.52 (0.15 - 1.44) | 0.27 | 0.52 (0.17 - 1.63) | 0.26 |
| Meta-analysis (w/o USOC) | 8,508 | 30,791 | 4.3 | 21.2 | <5 | 48 | 0.52 (0.15 - 1.44) | 0.27 | 0.50 (0.16 - 1.55) | 0.23 |

##### 1.4.8 Stroke

**Supplementary Table 86:** Exposures, outcomes and risk estimates for Stroke in GLP1RA (male) (T) vs DPP4I (male) (C) new-user cohorts under four different propensity score (PS) adjustment / time-at-risk (TAR) designs across data sources. We report total patient exposure time in 1,000 patient-year (KYr) units, uncalibrated and calibrated hazard ratios (HRs) and their 95% confidence intervals (CIs). Greyed out entries did not pass study diagnostics and are included only for completeness.

| | Patients | | Exposure (KYr) | | Outcomes | | Uncalibrated | | Calibrated | |
|---|---|---|---|---|---|---|---|---|---|---|
| | T | C | T | C | T | C | HR (95% CI) | $p$ | HR (95% CI) | $p$ |
| **PS-matching / on-treatment TAR** | | | | | | | | | | |
| CCAE | 10,484 | 52,183 | 7.2 | 45.8 | 23 | 152 | 1.18 (0.66 - 2.02) | 0.57 | 1.14 (0.65 - 2.00) | 0.65 |
| ODOD | 6,282 | 29,513 | 3.9 | 24.9 | 20 | 213 | 0.80 (0.42 - 1.43) | 0.47 | 0.79 (0.42 - 1.46) | 0.45 |
| MDCR | 828 | 8,611 | 0.6 | 9.0 | 5 | 128 | 0.69 (0.20 - 1.84) | 0.52 | 0.67 (0.22 - 2.04) | 0.48 |
| Open Claims | 113,043 | 415,501 | 87.5 | 399.3 | 255 | 2,015 | 0.79 (0.66 - 0.94) | <0.01 | 0.80 (0.66 - 0.96) | 0.02 |
| OEHR | 7,745 | 37,856 | 2.3 | 18.9 | 11 | 85 | 1.49 (0.65 - 3.16) | 0.33 | 1.29 (0.59 - 2.86) | 0.53 |
| VA | 1,192 | 17,454 | 0.9 | 14.8 | <10 | 32 | | | | |
| Meta-analysis | 138,382 | 543,664 | 101.5 | 498.1 | 314 | 2,593 | 0.87 (0.70 - 1.09) | 0.23 | 0.86 (0.66 - 1.12) | 0.26 |
| Meta-analysis (w/o USOC) | 25,339 | 128,163 | 14.0 | 98.7 | 59 | 578 | 1.03 (0.73 - 1.47) | 0.86 | 0.99 (0.70 - 1.41) | 0.93 |
| **PS-stratification / on-treatment TAR** | | | | | | | | | | |
| CCAE | 12,388 | 52,301 | 8.1 | 46.0 | 24 | 152 | 1.05 (0.64 - 1.65) | 0.85 | 1.03 (0.64 - 1.66) | 0.89 |
| ODOD | 8,371 | 30,043 | 4.8 | 25.4 | 24 | 223 | 0.69 (0.42 - 1.10) | 0.14 | 0.67 (0.41 - 1.09) | 0.11 |
| MDCR | 922 | 9,165 | 0.6 | 9.6 | 5 | 141 | 0.69 (0.23 - 1.64) | 0.46 | 0.64 (0.24 - 1.71) | 0.38 |
| Open Claims | 152,434 | 417,598 | 105.1 | 401.3 | 292 | 2,067 | 0.78 (0.68 - 0.90) | <0.01 | 0.81 (0.70 - 0.93) | 0.01 |
| OEHR | 9,031 | 37,945 | 2.6 | 19.0 | 12 | 87 | 1.33 (0.64 - 2.54) | 0.42 | 1.16 (0.58 - 2.31) | 0.67 |
| VA | 1,382 | 25,218 | 1.0 | 23.6 | <10 | 64 | 0.28 (0.02 - 1.47) | 0.28 | 0.25 (0.03 - 2.43) | 0.23 |
| Meta-analysis | 182,224 | 537,887 | 120.5 | 491.6 | 352 | 2,529 | 0.82 (0.70 - 0.95) | <0.01 | 0.82 (0.67 - 1.01) | 0.06 |
| Meta-analysis (w/o USOC) | 29,790 | 120,289 | 15.4 | 90.3 | 60 | 462 | 0.94 (0.66 - 1.35) | 0.75 | 0.91 (0.63 - 1.30) | 0.60 |
| **PS-matching / on-treatment TAR without escalation** | | | | | | | | | | |
| CCAE | 10,484 | 52,183 | 6.3 | 37.0 | 20 | 121 | 1.09 (0.58 - 1.95) | 0.77 | 1.07 (0.58 - 1.96) | 0.82 |
| ODOD | 6,282 | 29,513 | 3.5 | 20.3 | 17 | 184 | 0.62 (0.29 - 1.21) | 0.19 | 0.59 (0.29 - 1.21) | 0.16 |
| MDCR | 828 | 8,611 | 0.5 | 7.7 | <5 | 107 | 0.40 (0.06 - 1.39) | 0.24 | 0.34 (0.07 - 1.61) | 0.18 |
| Open Claims | 113,043 | 415,501 | 75.5 | 309.4 | 212 | 1,599 | 0.79 (0.65 - 0.96) | 0.02 | 0.78 (0.64 - 0.95) | 0.02 |
| OEHR | 7,745 | 37,856 | 2.0 | 15.4 | 9 | 63 | 1.33 (0.49 - 3.19) | 0.55 | 1.15 (0.45 - 2.92) | 0.77 |
| VA | 1,192 | 17,454 | 0.7 | 12.1 | <10 | 22 | | | | |
| Meta-analysis | 138,382 | 543,664 | 87.7 | 389.9 | <263 | 2,074 | 0.81 (0.68 - 0.96) | 0.02 | 0.79 (0.65 - 0.95) | 0.01 |
| Meta-analysis (w/o USOC) | 25,339 | 128,163 | 12.2 | 80.5 | <51 | 475 | 0.88 (0.58 - 1.34) | 0.56 | 0.83 (0.54 - 1.26) | 0.39 |
| **PS-stratification / on-treatment TAR without escalation** | | | | | | | | | | |
| CCAE | 12,388 | 52,301 | 7.0 | 37.1 | 21 | 121 | 1.08 (0.63 - 1.77) | 0.77 | 1.05 (0.63 - 1.76) | 0.85 |
| ODOD | 8,371 | 30,043 | 4.3 | 20.7 | 21 | 194 | 0.67 (0.39 - 1.11) | 0.14 | 0.63 (0.37 - 1.06) | 0.09 |
| MDCR | 922 | 9,165 | 0.5 | 8.2 | <5 | 118 | 0.50 (0.12 - 1.47) | 0.29 | 0.46 (0.13 - 1.63) | 0.23 |
| Open Claims | 152,434 | 417,598 | 91.8 | 311.0 | 245 | 1,640 | 0.74 (0.63 - 0.86) | <0.01 | 0.76 (0.65 - 0.89) | <0.01 |
| OEHR | 9,031 | 37,945 | 2.2 | 15.4 | 10 | 64 | 1.36 (0.60 - 2.83) | 0.43 | 1.15 (0.53 - 2.50) | 0.72 |
| VA | 1,382 | 25,218 | 0.8 | 19.2 | <10 | 41 | 0.46 (0.02 - 2.57) | 0.52 | 0.39 (0.04 - 3.95) | 0.43 |
| Meta-analysis | 182,224 | 537,887 | 105.3 | 384.2 | 297 | 2,019 | 0.81 (0.65 - 1.02) | 0.07 | 0.81 (0.63 - 1.03) | 0.09 |
| Meta-analysis (w/o USOC) | 29,790 | 120,289 | 13.5 | 73.2 | 52 | 379 | 0.94 (0.63 - 1.41) | 0.78 | 0.89 (0.60 - 1.33) | 0.59 |

##### 1.4.9 Sudden cardiac death

**Supplementary Table 87:** Exposures, outcomes and risk estimates for Sudden cardiac death in GLP1RA (male) (T) vs DPP4I (male) (C) new-user cohorts under four different propensity score (PS) adjustment / time-at-risk (TAR) designs across data sources. We report total patient exposure time in 1,000 patient-year (KYr) units, uncalibrated and calibrated hazard ratios (HRs) and their 95% confidence intervals (CIs). Greyed out entries did not pass study diagnostics and are included only for completeness.

| | Patients | | Exposure (KYr) | | Outcomes | | Uncalibrated | | Calibrated | |
|---|---|---|---|---|---|---|---|---|---|---|
| | T | C | T | C | T | C | HR (95% CI) | $p$ | HR (95% CI) | $p$ |
| **PS-matching / on-treatment TAR** | | | | | | | | | | |
| CCAE | 10,601 | 52,701 | 7.3 | 46.4 | <5 | 36 | 0.48 (0.11 - 1.54) | 0.28 | 0.46 (0.12 - 1.76) | 0.26 |
| ODOD | 6,400 | 30,099 | 4.0 | 25.4 | 9 | 81 | 0.76 (0.27 - 1.85) | 0.57 | 0.75 (0.28 - 1.96) | 0.55 |
| MDCR | 851 | 8,878 | 0.6 | 9.4 | <5 | 35 | | | | |
| Open Claims | 115,407 | 426,516 | 89.2 | 409.8 | 76 | 621 | 0.77 (0.55 - 1.05) | 0.11 | 0.77 (0.56 - 1.07) | 0.14 |
| OEHR | 7,802 | 38,165 | 2.3 | 19.1 | 5 | 29 | 0.92 (0.19 - 3.50) | 0.91 | 0.80 (0.18 - 3.44) | 0.76 |
| VA | 1,212 | 17,668 | 0.9 | 15.0 | 0 | <10 | | | | |
| Meta-analysis | 140,210 | 547,481 | 102.9 | 500.8 | <95 | 767 | 0.75 (0.56 - 1.01) | 0.06 | 0.74 (0.54 - 1.03) | 0.07 |
| Meta-analysis (w/o USOC) | 24,803 | 120,965 | 13.6 | 90.9 | <19 | 146 | 0.70 (0.35 - 1.39) | 0.31 | 0.67 (0.34 - 1.34) | 0.26 |
| **PS-stratification / on-treatment TAR** | | | | | | | | | | |
| CCAE | 12,525 | 52,834 | 8.2 | 46.5 | <5 | 36 | 0.44 (0.12 - 1.21) | 0.16 | 0.43 (0.14 - 1.36) | 0.15 |
| ODOD | 8,502 | 30,796 | 4.9 | 26.0 | 10 | 87 | 0.52 (0.24 - 1.04) | 0.08 | 0.50 (0.24 - 1.05) | 0.07 |
| MDCR | 949 | 9,537 | 0.6 | 10.0 | <5 | 40 | 0.32 (0.02 - 1.68) | 0.33 | 0.30 (0.03 - 2.95) | 0.30 |
| Open Claims | 155,278 | 428,778 | 106.9 | 412.0 | 87 | 650 | 0.80 (0.62 - 1.03) | 0.09 | 0.83 (0.64 - 1.07) | 0.17 |
| OEHR | 9,101 | 38,258 | 2.6 | 19.2 | 5 | 30 | 0.82 (0.25 - 2.25) | 0.72 | 0.71 (0.24 - 2.14) | 0.55 |
| VA | 1,405 | 25,610 | 1.0 | 24.0 | 0 | <10 | | | | |
| Meta-analysis | 185,406 | 550,666 | 122.6 | 503.6 | <107 | 803 | 0.74 (0.58 - 0.95) | 0.02 | 0.75 (0.57 - 0.99) | 0.05 |
| Meta-analysis (w/o USOC) | 30,128 | 121,888 | 15.7 | 91.7 | <20 | 153 | 0.56 (0.32 - 0.96) | 0.03 | 0.54 (0.31 - 0.93) | 0.03 |
| **PS-matching / on-treatment TAR without escalation** | | | | | | | | | | |
| CCAE | 10,601 | 52,701 | 6.3 | 37.5 | <5 | 30 | 0.26 (0.01 - 1.40) | 0.26 | 0.26 (0.03 - 2.57) | 0.25 |
| ODOD | 6,400 | 30,099 | 3.5 | 20.8 | 8 | 73 | 0.81 (0.26 - 2.17) | 0.70 | 0.78 (0.27 - 2.26) | 0.65 |
| MDCR | 851 | 8,878 | 0.5 | 8.0 | <5 | 23 | | | | |
| Open Claims | 115,407 | 426,516 | 76.9 | 317.5 | 64 | 475 | 0.74 (0.51 - 1.05) | 0.10 | 0.74 (0.51 - 1.06) | 0.10 |
| OEHR | 7,802 | 38,165 | 2.0 | 15.5 | <5 | 19 | 1.25 (0.24 - 5.81) | 0.78 | 1.08 (0.22 - 5.35) | 0.92 |
| VA | 1,212 | 17,668 | 0.7 | 12.3 | 0 | <10 | | | | |
| Meta-analysis | 132,408 | 509,316 | 86.8 | 375.7 | <77 | 578 | 0.73 (0.52 - 1.03) | 0.07 | 0.71 (0.50 - 1.01) | 0.06 |
| Meta-analysis (w/o USOC) | 17,001 | 82,800 | 9.9 | 58.2 | <13 | 103 | 0.66 (0.25 - 1.75) | 0.41 | 0.62 (0.24 - 1.64) | 0.34 |
| **PS-stratification / on-treatment TAR without escalation** | | | | | | | | | | |
| CCAE | 12,525 | 52,834 | 7.1 | 37.6 | <5 | 30 | 0.27 (0.04 - 1.00) | 0.11 | 0.27 (0.05 - 1.30) | 0.10 |
| ODOD | 8,502 | 30,796 | 4.3 | 21.2 | 9 | 78 | 0.47 (0.20 - 0.98) | 0.06 | 0.44 (0.20 - 0.97) | 0.04 |
| MDCR | 949 | 9,537 | 0.6 | 8.5 | <5 | 27 | 0.36 (0.02 - 1.96) | 0.39 | 0.33 (0.03 - 3.35) | 0.35 |
| Open Claims | 155,278 | 428,778 | 93.4 | 319.2 | 72 | 501 | 0.80 (0.60 - 1.06) | 0.12 | 0.82 (0.62 - 1.09) | 0.18 |
| OEHR | 9,101 | 38,258 | 2.3 | 15.6 | <5 | 20 | 0.74 (0.19 - 2.33) | 0.63 | 0.62 (0.18 - 2.17) | 0.46 |
| VA | 1,405 | 25,610 | 0.8 | 19.5 | 0 | <10 | | | | |
| Meta-analysis | 176,305 | 512,408 | 104.9 | 378.0 | <86 | 609 | 0.62 (0.37 - 1.05) | 0.08 | 0.62 (0.36 - 1.06) | 0.08 |
| Meta-analysis (w/o USOC) | 21,027 | 83,630 | 11.5 | 58.8 | <14 | 108 | 0.42 (0.21 - 0.85) | 0.02 | 0.40 (0.20 - 0.81) | 0.01 |

##### 1.4.10 Abnormal weight gain

**Supplementary Table 88:** Exposures, outcomes and risk estimates for Abnormal weight gain in GLP1RA (male) (T) vs DPP4I (male) (C) new-user cohorts under four different propensity score (PS) adjustment / time-at-risk (TAR) designs across data sources. We report total patient exposure time in 1,000 patient-year (KYr) units, uncalibrated and calibrated hazard ratios (HRs) and their 95% confidence intervals (CIs). Greyed out entries did not pass study diagnostics and are included only for completeness.

| | Patients | | Exposure (KYr) | | Outcomes | | Uncalibrated | | Calibrated | |
|---|---|---|---|---|---|---|---|---|---|---|
| | T | C | T | C | T | C | HR (95% CI) | $p$ | HR (95% CI) | $p$ |
| **PS-matching / on-treatment TAR** | | | | | | | | | | |
| CCAE | 10,111 | 51,366 | 7.0 | 45.1 | 37 | 152 | 1.61 (0.91 - 2.76) | 0.09 | 1.56 (0.89 - 2.71) | 0.12 |
| ODOD | 6,116 | 29,345 | 3.8 | 24.6 | 28 | 156 | 0.84 (0.45 - 1.51) | 0.58 | 0.83 (0.45 - 1.52) | 0.55 |
| MDCR | 835 | 8,821 | 0.6 | 9.3 | 5 | 20 | 8.83 (2.00 - 38.3) | <0.01 | 8.53 (1.95 - 37.3) | <0.01 |
| Open Claims | 110,691 | 415,969 | 85.3 | 399.1 | 175 | 708 | 1.03 (0.81 - 1.30) | 0.79 | 1.04 (0.82 - 1.33) | 0.75 |
| OEHR | 7,454 | 37,259 | 2.2 | 18.6 | 20 | 84 | 1.90 (0.90 - 3.84) | 0.08 | 1.65 (0.80 - 3.41) | 0.18 |
| VA | 1,177 | 17,367 | 0.9 | 14.8 | <10 | <10 | 1.76 (0.08 - 19.3) | 0.69 | 1.43 (0.09 - 22.4) | 0.80 |
| Meta-analysis | 134,372 | 533,939 | 98.3 | 487.5 | 260 | 1,100 | 1.19 (0.88 - 1.61) | 0.27 | 1.17 (0.84 - 1.63) | 0.35 |
| Meta-analysis (w/o USOC) | 23,681 | 117,970 | 13.0 | 88.4 | 85 | 392 | 1.35 (0.84 - 2.18) | 0.22 | 1.30 (0.80 - 2.09) | 0.29 |
| **PS-stratification / on-treatment TAR** | | | | | | | | | | |
| CCAE | 11,829 | 51,500 | 7.8 | 45.3 | 41 | 152 | 1.31 (0.87 - 1.92) | 0.18 | 1.29 (0.87 - 1.91) | 0.21 |
| ODOD | 8,011 | 29,955 | 4.6 | 25.1 | 40 | 157 | 1.08 (0.70 - 1.62) | 0.72 | 1.04 (0.69 - 1.58) | 0.85 |
| MDCR | 928 | 9,468 | 0.6 | 10.0 | 5 | 22 | 4.40 (1.27 - 12.8) | 0.01 | 4.09 (1.29 - 13.0) | 0.02 |
| Open Claims | 146,922 | 418,287 | 101.3 | 401.3 | 235 | 710 | 1.11 (0.93 - 1.32) | 0.25 | 1.15 (0.96 - 1.37) | 0.17 |
| OEHR | 8,589 | 37,352 | 2.4 | 18.6 | 23 | 84 | 2.15 (1.27 - 3.53) | <0.01 | 1.88 (1.13 - 3.15) | 0.02 |
| VA | 1,353 | 25,119 | 0.9 | 23.5 | <10 | 11 | 2.72 (0.50 - 13.7) | 0.24 | 2.40 (0.46 - 12.6) | 0.30 |
| Meta-analysis | 175,351 | 537,094 | 116.1 | 490.3 | 339 | 1,103 | 1.27 (0.99 - 1.63) | 0.06 | 1.29 (0.97 - 1.71) | 0.08 |
| Meta-analysis (w/o USOC) | 28,429 | 118,807 | 14.8 | 89.0 | 104 | 393 | 1.41 (0.97 - 2.04) | 0.07 | 1.35 (0.93 - 1.97) | 0.12 |
| **PS-matching / on-treatment TAR without escalation** | | | | | | | | | | |
| CCAE | 10,111 | 51,366 | 6.1 | 36.5 | 30 | 121 | 1.93 (1.05 - 3.46) | 0.03 | 1.89 (1.04 - 3.44) | 0.04 |
| ODOD | 6,116 | 29,345 | 3.4 | 20.2 | 21 | 121 | 0.75 (0.35 - 1.48) | 0.43 | 0.72 (0.35 - 1.47) | 0.38 |
| MDCR | 835 | 8,821 | 0.5 | 7.9 | <5 | 14 | 7.74 (1.41 - 42.7) | 0.02 | 6.68 (1.21 - 36.8) | 0.03 |
| Open Claims | 110,691 | 415,969 | 73.6 | 309.8 | 149 | 532 | 0.95 (0.72 - 1.24) | 0.69 | 0.94 (0.72 - 1.24) | 0.67 |
| OEHR | 7,454 | 37,259 | 1.9 | 15.1 | 18 | 66 | 2.07 (0.92 - 4.44) | 0.07 | 1.79 (0.81 - 3.92) | 0.15 |
| VA | 1,177 | 17,367 | 0.7 | 12.1 | <10 | <10 | | | | |
| Meta-analysis | 134,372 | 533,939 | 84.9 | 381.6 | 218 | 840 | 1.25 (0.78 - 1.98) | 0.36 | 1.22 (0.76 - 1.95) | 0.42 |
| Meta-analysis (w/o USOC) | 23,681 | 117,970 | 11.3 | 71.8 | 69 | 308 | 1.45 (0.77 - 2.72) | 0.25 | 1.36 (0.72 - 2.56) | 0.34 |
| **PS-stratification / on-treatment TAR without escalation** | | | | | | | | | | |
| CCAE | 11,829 | 51,500 | 6.8 | 36.6 | 34 | 121 | 1.25 (0.80 - 1.91) | 0.32 | 1.21 (0.78 - 1.88) | 0.40 |
| ODOD | 8,011 | 29,955 | 4.1 | 20.6 | 33 | 122 | 0.99 (0.61 - 1.58) | 0.98 | 0.93 (0.58 - 1.49) | 0.77 |
| MDCR | 928 | 9,468 | 0.5 | 8.5 | <5 | 16 | 3.83 (0.92 - 13.0) | 0.05 | 3.51 (0.93 - 13.2) | 0.06 |
| Open Claims | 146,922 | 418,287 | 88.4 | 311.5 | 206 | 534 | 1.12 (0.92 - 1.36) | 0.25 | 1.15 (0.94 - 1.40) | 0.19 |
| OEHR | 8,589 | 37,352 | 2.1 | 15.2 | 19 | 66 | 1.96 (1.09 - 3.38) | 0.02 | 1.65 (0.94 - 2.92) | 0.09 |
| VA | 1,353 | 25,119 | 0.7 | 19.1 | <10 | <10 | 2.03 (0.25 - 13.5) | 0.49 | 1.69 (0.23 - 12.4) | 0.60 |
| Meta-analysis | 175,351 | 537,094 | 101.4 | 383.9 | 292 | 843 | 1.17 (1.00 - 1.38) | 0.05 | 1.17 (0.97 - 1.41) | 0.10 |
| Meta-analysis (w/o USOC) | 28,429 | 118,807 | 13.0 | 72.4 | 86 | 309 | 1.30 (0.91 - 1.87) | 0.15 | 1.24 (0.86 - 1.77) | 0.26 |

##### 1.4.11 Abnormal weight loss

**Supplementary Table 89:** Exposures, outcomes and risk estimates for Abnormal weight loss in GLP1RA (male) (T) vs DPP4I (male) (C) new-user cohorts under four different propensity score (PS) adjustment / time-at-risk (TAR) designs across data sources. We report total patient exposure time in 1,000 patient-year (KYr) units, uncalibrated and calibrated hazard ratios (HRs) and their 95% confidence intervals (CIs). Greyed out entries did not pass study diagnostics and are included only for completeness.

| | Patients | | Exposure (KYr) | | Outcomes | | Uncalibrated | | Calibrated | |
|---|---|---|---|---|---|---|---|---|---|---|
| | T | C | T | C | T | C | HR (95% CI) | $p$ | HR (95% CI) | $p$ |
| **PS-matching / on-treatment TAR** | | | | | | | | | | |
| CCAE | 10,401 | 51,534 | 7.2 | 45.2 | 56 | 297 | 1.23 (0.84 - 1.78) | 0.28 | 1.19 (0.82 - 1.74) | 0.37 |
| ODOD | 6,222 | 29,041 | 3.9 | 24.3 | 50 | 368 | 1.02 (0.68 - 1.52) | 0.91 | 1.01 (0.67 - 1.51) | 0.90 |
| MDCR | 835 | 8,595 | 0.6 | 9.1 | 5 | 122 | 0.74 (0.24 - 1.86) | 0.56 | 0.72 (0.26 - 1.99) | 0.53 |
| Open Claims | 112,193 | 412,516 | 86.7 | 395.4 | 390 | 2,494 | 0.89 (0.77 - 1.03) | 0.13 | 0.90 (0.78 - 1.05) | 0.23 |
| OEHR | 7,600 | 37,056 | 2.2 | 18.4 | 24 | 221 | 0.95 (0.54 - 1.58) | 0.85 | 0.83 (0.48 - 1.41) | 0.50 |
| VA | 1,178 | 17,162 | 0.9 | 14.6 | 10 | 109 | 0.73 (0.23 - 1.83) | 0.54 | 0.59 (0.21 - 1.66) | 0.34 |
| Meta-analysis | 138,429 | 555,904 | 101.5 | 507.0 | 535 | 3,611 | 0.94 (0.82 - 1.08) | 0.37 | 0.93 (0.77 - 1.12) | 0.44 |
| Meta-analysis (w/o USOC) | 26,236 | 143,388 | 14.7 | 111.6 | 145 | 1,117 | 1.05 (0.83 - 1.32) | 0.70 | 1.01 (0.80 - 1.27) | 0.90 |
| **PS-stratification / on-treatment TAR** | | | | | | | | | | |
| CCAE | 12,276 | 51,674 | 8.0 | 45.3 | 65 | 298 | 1.19 (0.87 - 1.61) | 0.27 | 1.17 (0.86 - 1.60) | 0.32 |
| ODOD | 8,275 | 29,539 | 4.7 | 24.7 | 61 | 379 | 1.06 (0.76 - 1.45) | 0.73 | 1.02 (0.74 - 1.42) | 0.87 |
| MDCR | 935 | 9,144 | 0.6 | 9.6 | 5 | 137 | 0.46 (0.16 - 1.10) | 0.12 | 0.43 (0.16 - 1.14) | 0.09 |
| Open Claims | 151,256 | 414,312 | 104.1 | 397.1 | 462 | 2,535 | 0.90 (0.80 - 1.01) | 0.07 | 0.93 (0.82 - 1.05) | 0.32 |
| OEHR | 8,866 | 37,138 | 2.5 | 18.5 | 25 | 221 | 1.02 (0.64 - 1.56) | 0.94 | 0.89 (0.57 - 1.39) | 0.62 |
| VA | 1,367 | 24,656 | 1.0 | 23.0 | 11 | 212 | 1.28 (0.61 - 2.46) | 0.49 | 1.13 (0.56 - 2.26) | 0.74 |
| Meta-analysis | 180,673 | 532,663 | 119.3 | 485.6 | 613 | 3,433 | 0.99 (0.85 - 1.15) | 0.88 | 1.00 (0.81 - 1.22) | 0.92 |
| Meta-analysis (w/o USOC) | 29,417 | 118,351 | 15.2 | 88.5 | 151 | 898 | 1.10 (0.90 - 1.35) | 0.33 | 1.06 (0.86 - 1.31) | 0.61 |
| **PS-matching / on-treatment TAR without escalation** | | | | | | | | | | |
| CCAE | 10,401 | 51,534 | 6.2 | 36.5 | 51 | 231 | 1.20 (0.79 - 1.80) | 0.37 | 1.18 (0.78 - 1.78) | 0.44 |
| ODOD | 6,222 | 29,041 | 3.4 | 19.9 | 44 | 310 | 1.01 (0.63 - 1.57) | 0.97 | 0.97 (0.61 - 1.52) | 0.87 |
| MDCR | 835 | 8,595 | 0.5 | 7.7 | 5 | 109 | 0.76 (0.25 - 1.94) | 0.61 | 0.66 (0.24 - 1.85) | 0.43 |
| Open Claims | 112,193 | 412,516 | 74.8 | 306.4 | 325 | 1,964 | 0.83 (0.70 - 0.98) | 0.03 | 0.83 (0.70 - 0.98) | 0.04 |
| OEHR | 7,600 | 37,056 | 1.9 | 15.0 | 19 | 164 | 0.93 (0.49 - 1.68) | 0.83 | 0.81 (0.43 - 1.50) | 0.51 |
| VA | 1,178 | 17,162 | 0.7 | 11.9 | <10 | 92 | 0.55 (0.12 - 1.74) | 0.37 | 0.42 (0.11 - 1.57) | 0.20 |
| Meta-analysis | 138,429 | 555,904 | 87.5 | 397.5 | <454 | 2,870 | 0.90 (0.76 - 1.08) | 0.26 | 0.88 (0.73 - 1.07) | 0.20 |
| Meta-analysis (w/o USOC) | 26,236 | 143,388 | 12.7 | 91.1 | <129 | 906 | 1.03 (0.79 - 1.33) | 0.85 | 0.96 (0.74 - 1.25) | 0.79 |
| **PS-stratification / on-treatment TAR without escalation** | | | | | | | | | | |
| CCAE | 12,276 | 51,674 | 7.0 | 36.6 | 59 | 232 | 1.26 (0.90 - 1.74) | 0.17 | 1.22 (0.88 - 1.71) | 0.24 |
| ODOD | 8,275 | 29,539 | 4.2 | 20.2 | 53 | 318 | 1.04 (0.73 - 1.47) | 0.81 | 0.98 (0.69 - 1.39) | 0.88 |
| MDCR | 935 | 9,144 | 0.5 | 8.2 | 5 | 121 | 0.53 (0.18 - 1.29) | 0.21 | 0.49 (0.18 - 1.32) | 0.16 |
| Open Claims | 151,256 | 414,312 | 91.0 | 307.7 | 394 | 1,998 | 0.89 (0.78 - 1.01) | 0.06 | 0.91 (0.79 - 1.03) | 0.17 |
| OEHR | 8,866 | 37,138 | 2.2 | 15.0 | 20 | 164 | 1.07 (0.63 - 1.74) | 0.80 | 0.90 (0.54 - 1.50) | 0.70 |
| VA | 1,367 | 24,656 | 0.8 | 18.7 | <10 | 174 | 0.93 (0.37 - 2.06) | 0.87 | 0.78 (0.33 - 1.84) | 0.57 |
| Meta-analysis | 180,673 | 532,663 | 104.3 | 379.6 | 526 | 2,712 | 1.01 (0.83 - 1.21) | 0.95 | 1.00 (0.81 - 1.24) | 0.92 |
| Meta-analysis (w/o USOC) | 29,417 | 118,351 | 13.3 | 71.9 | 132 | 714 | 1.14 (0.92 - 1.41) | 0.25 | 1.08 (0.87 - 1.34) | 0.52 |

##### 1.4.12 Acute pancreatitis

**Supplementary Table 90:** Exposures, outcomes and risk estimates for Acute pancreatitis in GLP1RA (male) (T) vs DPP4I (male) (C) new-user cohorts under four different propensity score (PS) adjustment / time-at-risk (TAR) designs across data sources. We report total patient exposure time in 1,000 patient-year (KYr) units, uncalibrated and calibrated hazard ratios (HRs) and their 95% confidence intervals (CIs). Greyed out entries did not pass study diagnostics and are included only for completeness.

| | Patients | | Exposure (KYr) | | Outcomes | | Uncalibrated | | Calibrated | |
|---|---|---|---|---|---|---|---|---|---|---|
| | T | C | T | C | T | C | HR (95% CI) | $p$ | HR (95% CI) | $p$ |
| **PS-matching / on-treatment TAR** | | | | | | | | | | |
| CCAE | 10,542 | 52,351 | 7.3 | 46.0 | 16 | 112 | 0.79 (0.37 - 1.57) | 0.52 | 0.77 (0.37 - 1.57) | 0.47 |
| ODOD | 6,368 | 29,991 | 4.0 | 25.4 | 8 | 67 | 0.76 (0.27 - 1.80) | 0.56 | 0.75 (0.29 - 1.92) | 0.55 |
| MDCR | 851 | 8,848 | 0.6 | 9.3 | 0 | 35 | | | | |
| Open Claims | 114,402 | 422,380 | 88.5 | 406.1 | 124 | 802 | 0.79 (0.61 - 1.01) | 0.07 | 0.79 (0.61 - 1.03) | 0.10 |
| OEHR | 7,767 | 38,036 | 2.3 | 19.1 | 6 | 44 | 1.39 (0.36 - 4.28) | 0.60 | 1.21 (0.35 - 4.18) | 0.76 |
| VA | 1,203 | 17,550 | 0.9 | 14.9 | 0 | 21 | | | | |
| Meta-analysis | 139,079 | 542,758 | 102.1 | 496.6 | 154 | 1,025 | 0.80 (0.64 - 1.01) | 0.06 | 0.79 (0.61 - 1.03) | 0.08 |
| Meta-analysis (w/o USOC) | 24,677 | 120,378 | 13.6 | 90.5 | 30 | 223 | 0.86 (0.51 - 1.45) | 0.58 | 0.83 (0.49 - 1.39) | 0.48 |
| **PS-stratification / on-treatment TAR** | | | | | | | | | | |
| CCAE | 12,453 | 52,487 | 8.1 | 46.2 | 18 | 112 | 0.73 (0.41 - 1.25) | 0.27 | 0.72 (0.41 - 1.26) | 0.26 |
| ODOD | 8,472 | 30,602 | 4.9 | 25.9 | 9 | 67 | 0.95 (0.41 - 1.97) | 0.90 | 0.92 (0.42 - 2.01) | 0.83 |
| MDCR | 948 | 9,484 | 0.6 | 9.9 | 0 | 38 | | | | |
| Open Claims | 154,048 | 424,790 | 106.2 | 408.4 | 149 | 812 | 0.82 (0.67 - 0.99) | 0.05 | 0.84 (0.69 - 1.03) | 0.12 |
| OEHR | 9,062 | 38,128 | 2.6 | 19.1 | 7 | 44 | 1.15 (0.44 - 2.62) | 0.76 | 1.01 (0.41 - 2.47) | 0.95 |
| VA | 1,395 | 25,396 | 1.0 | 23.8 | 0 | 42 | | | | |
| Meta-analysis | 184,035 | 546,007 | 121.8 | 499.5 | 183 | 1,035 | 0.82 (0.69 - 0.99) | 0.03 | 0.83 (0.67 - 1.04) | 0.11 |
| Meta-analysis (w/o USOC) | 29,987 | 121,217 | 15.6 | 91.1 | 34 | 223 | 0.86 (0.57 - 1.29) | 0.47 | 0.83 (0.55 - 1.25) | 0.38 |
| **PS-matching / on-treatment TAR without escalation** | | | | | | | | | | |
| CCAE | 10,542 | 52,351 | 6.3 | 37.2 | 12 | 81 | 0.63 (0.26 - 1.35) | 0.27 | 0.61 (0.27 - 1.40) | 0.25 |
| ODOD | 6,368 | 29,991 | 3.5 | 20.7 | 7 | 52 | 0.86 (0.27 - 2.25) | 0.78 | 0.82 (0.29 - 2.36) | 0.72 |
| MDCR | 851 | 8,848 | 0.5 | 7.9 | 0 | 33 | | | | |
| Open Claims | 114,402 | 422,380 | 76.3 | 314.8 | 99 | 588 | 0.79 (0.59 - 1.05) | 0.11 | 0.79 (0.59 - 1.05) | 0.11 |
| OEHR | 7,767 | 38,036 | 2.0 | 15.5 | 5 | 32 | 1.60 (0.40 - 5.15) | 0.47 | 1.38 (0.38 - 4.95) | 0.62 |
| VA | 1,203 | 17,550 | 0.7 | 12.2 | 0 | 15 | | | | |
| Meta-analysis | 139,079 | 542,758 | 88.1 | 388.2 | 123 | 753 | 0.80 (0.62 - 1.04) | 0.09 | 0.78 (0.60 - 1.02) | 0.07 |
| Meta-analysis (w/o USOC) | 24,677 | 120,378 | 11.8 | 73.4 | 24 | 165 | 0.84 (0.47 - 1.49) | 0.54 | 0.78 (0.44 - 1.40) | 0.42 |
| **PS-stratification / on-treatment TAR without escalation** | | | | | | | | | | |
| CCAE | 12,453 | 52,487 | 7.1 | 37.3 | 13 | 81 | 0.64 (0.32 - 1.19) | 0.18 | 0.62 (0.32 - 1.20) | 0.16 |
| ODOD | 8,472 | 30,602 | 4.3 | 21.1 | 8 | 52 | 1.12 (0.45 - 2.47) | 0.79 | 1.05 (0.45 - 2.46) | 0.91 |
| MDCR | 948 | 9,484 | 0.6 | 8.5 | 0 | 36 | | | | |
| Open Claims | 154,048 | 424,790 | 92.8 | 316.6 | 123 | 595 | 0.82 (0.65 - 1.03) | 0.09 | 0.84 (0.67 - 1.06) | 0.15 |
| OEHR | 9,062 | 38,128 | 2.3 | 15.5 | 6 | 32 | 1.32 (0.45 - 3.31) | 0.59 | 1.11 (0.41 - 3.03) | 0.84 |
| VA | 1,395 | 25,396 | 0.8 | 19.3 | 0 | 31 | | | | |
| Meta-analysis | 184,035 | 546,007 | 106.4 | 390.5 | 150 | 760 | 0.83 (0.68 - 1.02) | 0.08 | 0.83 (0.67 - 1.04) | 0.11 |
| Meta-analysis (w/o USOC) | 29,987 | 121,217 | 13.7 | 74.0 | 27 | 165 | 0.88 (0.55 - 1.40) | 0.59 | 0.83 (0.52 - 1.32) | 0.45 |

##### 1.4.13 Bone fracture

**Supplementary Table 91:** Exposures, outcomes and risk estimates for Bone fracture in GLP1RA (male) (T) vs DPP4I (male) (C) new-user cohorts under four different propensity score (PS) adjustment / time-at-risk (TAR) designs across data sources. We report total patient exposure time in 1,000 patient-year (KYr) units, uncalibrated and calibrated hazard ratios (HRs) and their 95% confidence intervals (CIs). Greyed out entries did not pass study diagnostics and are included only for completeness.

| | Patients | | Exposure (KYr) | | Outcomes | | Uncalibrated | | Calibrated | |
|---|---|---|---|---|---|---|---|---|---|---|
| | T | C | T | C | T | C | HR (95% CI) | $p$ | HR (95% CI) | $p$ |
| **PS-matching / on-treatment TAR** | | | | | | | | | | |
| CCAE | 9,745 | 49,178 | 6.6 | 42.5 | 111 | 632 | 1.08 (0.81 - 1.43) | 0.58 | 1.05 (0.79 - 1.39) | 0.76 |
| ODOD | 5,938 | 28,138 | 3.7 | 23.4 | 85 | 449 | 1.36 (0.96 - 1.92) | 0.08 | 1.34 (0.95 - 1.90) | 0.12 |
| MDCR | 769 | 8,059 | 0.5 | 8.3 | 11 | 227 | 0.81 (0.36 - 1.63) | 0.59 | 0.79 (0.37 - 1.67) | 0.54 |
| Open Claims | 101,595 | 383,240 | 78.0 | 364.6 | 852 | 4,916 | 0.99 (0.89 - 1.09) | 0.82 | 1.00 (0.89 - 1.11) | 0.88 |
| OEHR | 7,309 | 36,130 | 2.2 | 17.9 | 27 | 281 | 0.79 (0.45 - 1.31) | 0.39 | 0.69 (0.40 - 1.17) | 0.19 |
| VA | 1,086 | 15,990 | 0.8 | 13.7 | <10 | 123 | 0.85 (0.30 - 2.04) | 0.74 | 0.69 (0.27 - 1.80) | 0.47 |
| Meta-analysis | 126,442 | 520,735 | 91.8 | 470.3 | <1,096 | 6,628 | 1.01 (0.92 - 1.10) | 0.86 | 1.00 (0.85 - 1.17) | 0.88 |
| Meta-analysis (w/o USOC) | 24,847 | 137,495 | 13.8 | 105.7 | <244 | 1,712 | 1.08 (0.89 - 1.31) | 0.42 | 1.04 (0.86 - 1.26) | 0.71 |
| **PS-stratification / on-treatment TAR** | | | | | | | | | | |
| CCAE | 11,492 | 49,297 | 7.4 | 42.6 | 122 | 633 | 1.06 (0.85 - 1.32) | 0.59 | 1.05 (0.84 - 1.31) | 0.69 |
| ODOD | 7,882 | 28,638 | 4.5 | 23.8 | 111 | 462 | 1.34 (1.04 - 1.72) | 0.02 | 1.30 (1.01 - 1.67) | 0.05 |
| MDCR | 864 | 8,670 | 0.5 | 8.8 | 11 | 242 | 0.77 (0.38 - 1.41) | 0.44 | 0.72 (0.37 - 1.38) | 0.33 |
| Open Claims | 135,593 | 385,620 | 93.0 | 366.8 | 1,016 | 4,993 | 0.93 (0.86 - 1.00) | 0.06 | 0.96 (0.88 - 1.05) | 0.48 |
| OEHR | 8,521 | 36,214 | 2.4 | 17.9 | 30 | 284 | 0.84 (0.55 - 1.24) | 0.41 | 0.74 (0.49 - 1.11) | 0.15 |
| VA | 1,264 | 23,057 | 0.9 | 21.6 | <10 | 216 | 0.99 (0.45 - 1.97) | 0.99 | 0.87 (0.42 - 1.84) | 0.73 |
| Meta-analysis | 163,488 | 499,769 | 107.3 | 451.1 | 1,279 | 6,372 | 1.03 (0.86 - 1.24) | 0.73 | 1.04 (0.83 - 1.30) | 0.72 |
| Meta-analysis (w/o USOC) | 27,895 | 114,149 | 14.3 | 84.3 | 263 | 1,379 | 1.10 (0.87 - 1.39) | 0.41 | 1.06 (0.83 - 1.34) | 0.66 |
| **PS-matching / on-treatment TAR without escalation** | | | | | | | | | | |
| CCAE | 9,745 | 49,178 | 5.7 | 34.5 | 89 | 510 | 1.04 (0.75 - 1.43) | 0.79 | 1.02 (0.74 - 1.41) | 0.86 |
| ODOD | 5,938 | 28,138 | 3.2 | 19.2 | 74 | 360 | 1.35 (0.91 - 1.97) | 0.13 | 1.29 (0.88 - 1.90) | 0.21 |
| MDCR | 769 | 8,059 | 0.5 | 7.1 | 10 | 197 | 0.95 (0.40 - 2.01) | 0.91 | 0.82 (0.37 - 1.85) | 0.64 |
| Open Claims | 101,595 | 383,240 | 67.3 | 283.1 | 724 | 3,775 | 0.98 (0.88 - 1.10) | 0.77 | 0.98 (0.87 - 1.10) | 0.73 |
| OEHR | 7,309 | 36,130 | 1.9 | 14.6 | 21 | 207 | 0.88 (0.47 - 1.57) | 0.69 | 0.76 (0.42 - 1.39) | 0.39 |
| VA | 1,086 | 15,990 | 0.6 | 11.2 | <10 | 95 | 0.72 (0.23 - 1.86) | 0.53 | 0.55 (0.19 - 1.55) | 0.27 |
| Meta-analysis | 126,442 | 520,735 | 79.2 | 369.7 | <928 | 5,144 | 1.00 (0.91 - 1.11) | 0.94 | 0.98 (0.87 - 1.11) | 0.76 |
| Meta-analysis (w/o USOC) | 24,847 | 137,495 | 11.9 | 86.5 | <204 | 1,369 | 1.08 (0.87 - 1.34) | 0.47 | 1.02 (0.82 - 1.26) | 0.85 |
| **PS-stratification / on-treatment TAR without escalation** | | | | | | | | | | |
| CCAE | 11,492 | 49,297 | 6.4 | 34.6 | 99 | 511 | 1.02 (0.80 - 1.30) | 0.87 | 0.99 (0.78 - 1.27) | 0.89 |
| ODOD | 7,882 | 28,638 | 4.0 | 19.5 | 99 | 372 | 1.38 (1.05 - 1.81) | 0.02 | 1.29 (0.98 - 1.70) | 0.08 |
| MDCR | 864 | 8,670 | 0.5 | 7.6 | 10 | 211 | 0.83 (0.39 - 1.57) | 0.60 | 0.76 (0.38 - 1.52) | 0.45 |
| Open Claims | 135,593 | 385,620 | 81.3 | 284.9 | 869 | 3,843 | 0.92 (0.85 - 1.01) | 0.08 | 0.94 (0.86 - 1.04) | 0.31 |
| OEHR | 8,521 | 36,214 | 2.1 | 14.6 | 24 | 209 | 0.83 (0.51 - 1.29) | 0.42 | 0.70 (0.44 - 1.11) | 0.14 |
| VA | 1,264 | 23,057 | 0.7 | 17.6 | <10 | 166 | 0.95 (0.35 - 2.17) | 0.92 | 0.80 (0.32 - 1.98) | 0.63 |
| Meta-analysis | 163,488 | 499,769 | 93.8 | 353.7 | 1,091 | 4,935 | 1.03 (0.84 - 1.25) | 0.80 | 1.02 (0.82 - 1.27) | 0.84 |
| Meta-analysis (w/o USOC) | 27,895 | 114,149 | 12.5 | 68.8 | 222 | 1,092 | 1.09 (0.84 - 1.43) | 0.52 | 1.03 (0.79 - 1.36) | 0.81 |

##### 1.4.14 Diabetic ketoacidosis

**Supplementary Table 92:** Exposures, outcomes and risk estimates for Diabetic ketoacidosis in GLP1RA (male) (T) vs DPP4I (male) (C) new-user cohorts under four different propensity score (PS) adjustment / time-at-risk (TAR) designs across data sources. We report total patient exposure time in 1,000 patient-year (KYr) units, uncalibrated and calibrated hazard ratios (HRs) and their 95% confidence intervals (CIs). Greyed out entries did not pass study diagnostics and are included only for completeness.

| | Patients | | Exposure (KYr) | | Outcomes | | Uncalibrated | | Calibrated | |
|---|---|---|---|---|---|---|---|---|---|---|
| | T | C | T | C | T | C | HR (95% CI) | $p$ | HR (95% CI) | $p$ |
| **PS-matching / on-treatment TAR** | | | | | | | | | | |
| CCAE | 10,603 | 52,701 | 7.3 | 46.4 | 7 | 36 | 0.52 (0.14 - 1.48) | 0.27 | 0.50 (0.16 - 1.61) | 0.25 |
| ODOD | 6,402 | 30,186 | 4.0 | 25.5 | <5 | 37 | 0.68 (0.10 - 3.02) | 0.66 | 0.67 (0.12 - 3.74) | 0.65 |
| MDCR | 854 | 8,895 | 0.6 | 9.4 | 0 | <5 | | | | |
| Open Claims | 115,399 | 426,651 | 89.2 | 410.1 | 60 | 303 | 0.78 (0.55 - 1.10) | 0.16 | 0.79 (0.55 - 1.12) | 0.20 |
| OEHR | 7,800 | 38,169 | 2.3 | 19.1 | <5 | 24 | 0.66 (0.14 - 2.28) | 0.56 | 0.58 (0.15 - 2.30) | 0.44 |
| VA | 1,214 | 17,669 | 0.9 | 15.0 | <10 | <10 | 4.05 (0.11 - 68.6) | 0.40 | 3.29 (0.13 - 83.7) | 0.47 |
| Meta-analysis | 140,204 | 547,707 | 102.9 | 501.2 | <77 | 400 | 0.75 (0.54 - 1.03) | 0.07 | 0.74 (0.52 - 1.04) | 0.08 |
| Meta-analysis (w/o USOC) | 24,805 | 121,056 | 13.7 | 91.1 | <17 | 97 | 0.59 (0.27 - 1.31) | 0.20 | 0.57 (0.26 - 1.26) | 0.17 |
| **PS-stratification / on-treatment TAR** | | | | | | | | | | |
| CCAE | 12,523 | 52,836 | 8.2 | 46.5 | 8 | 36 | 0.90 (0.36 - 2.04) | 0.81 | 0.89 (0.37 - 2.12) | 0.79 |
| ODOD | 8,503 | 30,811 | 4.9 | 26.0 | <5 | 38 | 0.40 (0.09 - 1.27) | 0.17 | 0.38 (0.10 - 1.46) | 0.16 |
| MDCR | 952 | 9,553 | 0.6 | 10.0 | 0 | 7 | | | | |
| Open Claims | 155,271 | 428,987 | 107.0 | 412.3 | 70 | 304 | 0.80 (0.59 - 1.06) | 0.13 | 0.82 (0.61 - 1.11) | 0.22 |
| OEHR | 9,102 | 38,262 | 2.6 | 19.2 | <5 | 24 | 0.61 (0.13 - 2.01) | 0.48 | 0.54 (0.14 - 2.08) | 0.37 |
| VA | 1,410 | 25,614 | 1.0 | 24.0 | <10 | 11 | 5.01 (0.18 - 51.0) | 0.26 | 4.41 (0.26 - 74.3) | 0.30 |
| Meta-analysis | 185,399 | 550,896 | 122.6 | 504.0 | <88 | 402 | 0.78 (0.59 - 1.01) | 0.06 | 0.78 (0.58 - 1.06) | 0.11 |
| Meta-analysis (w/o USOC) | 30,128 | 121,909 | 15.7 | 91.7 | <18 | 98 | 0.68 (0.36 - 1.30) | 0.24 | 0.66 (0.34 - 1.25) | 0.20 |
| **PS-matching / on-treatment TAR without escalation** | | | | | | | | | | |
| CCAE | 10,603 | 52,701 | 6.4 | 37.5 | 6 | 28 | 0.44 (0.10 - 1.41) | 0.22 | 0.43 (0.11 - 1.63) | 0.21 |
| ODOD | 6,402 | 30,186 | 3.5 | 20.8 | <5 | 25 | 0.28 (0.01 - 1.68) | 0.30 | 0.27 (0.03 - 2.89) | 0.28 |
| MDCR | 854 | 8,895 | 0.5 | 8.0 | 0 | <5 | | | | |
| Open Claims | 115,399 | 426,651 | 76.9 | 317.8 | 49 | 216 | 0.82 (0.55 - 1.21) | 0.34 | 0.82 (0.55 - 1.22) | 0.33 |
| OEHR | 7,800 | 38,169 | 2.0 | 15.5 | <5 | 17 | 1.28 (0.25 - 5.36) | 0.75 | 1.10 (0.24 - 5.09) | 0.90 |
| VA | 1,214 | 17,669 | 0.7 | 12.3 | <10 | <10 | | 0.31 | | 0.37 |
| Meta-analysis | 132,404 | 509,538 | 86.8 | 376.1 | <60 | 269 | 0.75 (0.49 - 1.14) | 0.18 | 0.73 (0.48 - 1.12) | 0.15 |
| Meta-analysis (w/o USOC) | 17,005 | 82,887 | 9.9 | 58.3 | <11 | 53 | 0.39 (0.12 - 1.26) | 0.12 | 0.37 (0.12 - 1.18) | 0.09 |
| **PS-stratification / on-treatment TAR without escalation** | | | | | | | | | | |
| CCAE | 12,523 | 52,836 | 7.1 | 37.6 | 7 | 28 | 0.89 (0.32 - 2.16) | 0.80 | 0.86 (0.33 - 2.22) | 0.76 |
| ODOD | 8,503 | 30,811 | 4.3 | 21.3 | <5 | 26 | 0.26 (0.04 - 1.05) | 0.11 | 0.25 (0.05 - 1.28) | 0.10 |
| MDCR | 952 | 9,553 | 0.6 | 8.6 | 0 | 6 | | | | |
| Open Claims | 155,271 | 428,987 | 93.5 | 319.5 | 58 | 217 | 0.89 (0.63 - 1.23) | 0.49 | 0.91 (0.65 - 1.27) | 0.58 |
| OEHR | 9,102 | 38,262 | 2.3 | 15.6 | <5 | 17 | 0.72 (0.15 - 2.50) | 0.64 | 0.60 (0.15 - 2.46) | 0.48 |
| VA | 1,410 | 25,614 | 0.8 | 19.5 | <10 | <10 | 6.72 (0.21 - 96.9) | 0.22 | 5.63 (0.26 - 121) | 0.27 |
| Meta-analysis | 176,297 | 512,634 | 104.9 | 378.4 | <70 | 271 | 0.85 (0.63 - 1.16) | 0.31 | 0.85 (0.62 - 1.17) | 0.32 |
| Meta-analysis (w/o USOC) | 21,026 | 83,647 | 11.5 | 58.9 | <12 | 54 | 0.59 (0.19 - 1.80) | 0.35 | 0.56 (0.18 - 1.71) | 0.31 |

##### 1.4.15 Diarrhea

**Supplementary Table 93:** Exposures, outcomes and risk estimates for Diarrhea in GLP1RA (male) (T) vs DPP4I (male) (C) new-user cohorts under four different propensity score (PS) adjustment / time-at-risk (TAR) designs across data sources. We report total patient exposure time in 1,000 patient-year (KYr) units, uncalibrated and calibrated hazard ratios (HRs) and their 95% confidence intervals (CIs). Greyed out entries did not pass study diagnostics and are included only for completeness.

| | Patients | | Exposure (KYr) | | Outcomes | | Uncalibrated | | Calibrated | |
|---|---|---|---|---|---|---|---|---|---|---|
| | T | C | T | C | T | C | HR (95% CI) | $p$ | HR (95% CI) | $p$ |
| **PS-matching / on-treatment TAR** | | | | | | | | | | |
| CCAE | 9,532 | 48,344 | 6.4 | 41.7 | 237 | 869 | 1.40 (1.14 - 1.71) | <0.01 | 1.35 (1.10 - 1.66) | <0.01 |
| ODOD | 5,742 | 26,979 | 3.5 | 22.1 | 151 | 661 | 1.33 (1.02 - 1.71) | 0.03 | 1.31 (1.01 - 1.69) | 0.06 |
| MDCR | 764 | 7,959 | 0.5 | 8.3 | 17 | 237 | 1.09 (0.58 - 1.92) | 0.79 | 1.05 (0.57 - 1.92) | 0.83 |
| Open Claims | 102,627 | 385,298 | 78.6 | 365.3 | 1,387 | 5,462 | 1.30 (1.20 - 1.42) | <0.01 | 1.32 (1.20 - 1.45) | <0.01 |
| OEHR | 7,127 | 35,171 | 2.1 | 17.2 | 82 | 466 | 1.75 (1.29 - 2.36) | <0.01 | 1.52 (1.12 - 2.07) | 0.01 |
| VA | 1,041 | 15,382 | 0.8 | 13.0 | 21 | 258 | 1.23 (0.67 - 2.15) | 0.48 | 1.00 (0.56 - 1.80) | 0.86 |
| Meta-analysis | 126,833 | 519,133 | 91.9 | 467.6 | 1,895 | 7,953 | 1.33 (1.24 - 1.43) | <0.01 | 1.32 (1.13 - 1.53) | <0.01 |
| Meta-analysis (w/o USOC) | 24,206 | 133,835 | 13.3 | 102.3 | 508 | 2,491 | 1.41 (1.23 - 1.61) | <0.01 | 1.36 (1.18 - 1.55) | <0.01 |
| **PS-stratification / on-treatment TAR** | | | | | | | | | | |
| CCAE | 11,200 | 48,527 | 7.2 | 41.9 | 273 | 874 | 1.43 (1.22 - 1.67) | <0.01 | 1.41 (1.20 - 1.66) | <0.01 |
| ODOD | 7,599 | 27,566 | 4.3 | 22.6 | 191 | 676 | 1.31 (1.08 - 1.59) | <0.01 | 1.27 (1.04 - 1.55) | 0.03 |
| MDCR | 850 | 8,555 | 0.5 | 8.8 | 17 | 264 | 1.01 (0.57 - 1.67) | 0.98 | 0.94 (0.55 - 1.60) | 0.80 |
| Open Claims | 137,360 | 387,058 | 93.7 | 366.9 | 1,776 | 5,531 | 1.27 (1.19 - 1.35) | <0.01 | 1.31 (1.22 - 1.41) | <0.01 |
| OEHR | 8,239 | 35,251 | 2.3 | 17.2 | 99 | 470 | 1.57 (1.23 - 1.99) | <0.01 | 1.37 (1.08 - 1.75) | 0.01 |
| VA | 1,194 | 22,232 | 0.8 | 20.7 | 23 | 408 | 1.27 (0.77 - 1.99) | 0.33 | 1.11 (0.69 - 1.79) | 0.66 |
| Meta-analysis | 164,398 | 498,402 | 107.5 | 448.6 | 2,339 | 7,551 | 1.34 (1.23 - 1.46) | <0.01 | 1.35 (1.15 - 1.59) | <0.01 |
| Meta-analysis (w/o USOC) | 27,038 | 111,344 | 13.7 | 81.7 | 563 | 2,020 | 1.42 (1.27 - 1.58) | <0.01 | 1.36 (1.20 - 1.54) | <0.01 |
| **PS-matching / on-treatment TAR without escalation** | | | | | | | | | | |
| CCAE | 9,532 | 48,344 | 5.6 | 33.9 | 214 | 707 | 1.54 (1.24 - 1.92) | <0.01 | 1.51 (1.21 - 1.88) | <0.01 |
| ODOD | 5,742 | 26,979 | 3.1 | 18.2 | 132 | 546 | 1.36 (1.03 - 1.78) | 0.03 | 1.30 (0.99 - 1.71) | 0.09 |
| MDCR | 764 | 7,959 | 0.5 | 7.1 | 15 | 202 | 1.23 (0.65 - 2.22) | 0.50 | 1.06 (0.57 - 1.97) | 0.81 |
| Open Claims | 102,627 | 385,298 | 68.0 | 284.6 | 1,192 | 4,223 | 1.37 (1.25 - 1.50) | <0.01 | 1.36 (1.23 - 1.50) | <0.01 |
| OEHR | 7,127 | 35,171 | 1.8 | 14.1 | 70 | 330 | 1.91 (1.36 - 2.67) | <0.01 | 1.65 (1.18 - 2.32) | <0.01 |
| VA | 1,041 | 15,382 | 0.6 | 10.7 | 18 | 213 | 1.28 (0.63 - 2.45) | 0.47 | 0.98 (0.50 - 1.93) | 0.86 |
| Meta-analysis | 126,833 | 519,133 | 79.5 | 368.5 | 1,641 | 6,221 | 1.42 (1.30 - 1.56) | <0.01 | 1.39 (1.23 - 1.56) | <0.01 |
| Meta-analysis (w/o USOC) | 24,206 | 133,835 | 11.5 | 84.0 | 449 | 1,998 | 1.52 (1.31 - 1.75) | <0.01 | 1.42 (1.23 - 1.65) | <0.01 |
| **PS-stratification / on-treatment TAR without escalation** | | | | | | | | | | |
| CCAE | 11,200 | 48,527 | 6.3 | 34.1 | 246 | 711 | 1.50 (1.26 - 1.77) | <0.01 | 1.45 (1.22 - 1.73) | <0.01 |
| ODOD | 7,599 | 27,566 | 3.8 | 18.6 | 165 | 561 | 1.30 (1.05 - 1.60) | 0.02 | 1.21 (0.98 - 1.50) | 0.10 |
| MDCR | 850 | 8,555 | 0.5 | 7.6 | 15 | 227 | 0.97 (0.52 - 1.67) | 0.91 | 0.89 (0.49 - 1.58) | 0.69 |
| Open Claims | 137,360 | 387,058 | 82.1 | 285.9 | 1,549 | 4,278 | 1.32 (1.23 - 1.41) | <0.01 | 1.35 (1.24 - 1.46) | <0.01 |
| OEHR | 8,239 | 35,251 | 2.0 | 14.1 | 85 | 332 | 1.84 (1.40 - 2.41) | <0.01 | 1.56 (1.18 - 2.05) | <0.01 |
| VA | 1,194 | 22,232 | 0.6 | 16.9 | 20 | 329 | 1.28 (0.75 - 2.09) | 0.34 | 1.07 (0.64 - 1.79) | 0.78 |
| Meta-analysis | 164,398 | 498,402 | 94.2 | 352.6 | 2,045 | 5,882 | 1.42 (1.26 - 1.61) | <0.01 | 1.42 (1.21 - 1.66) | <0.01 |
| Meta-analysis (w/o USOC) | 27,038 | 111,344 | 12.1 | 66.8 | 496 | 1,604 | 1.50 (1.26 - 1.79) | <0.01 | 1.43 (1.19 - 1.70) | <0.01 |

##### 1.4.16 Genitourinary infection

**Supplementary Table 94:** Exposures, outcomes and risk estimates for Genitourinary infection in GLP1RA (male) (T) vs DPP4I (male) (C) new-user cohorts under four different propensity score (PS) adjustment / time-at-risk (TAR) designs across data sources. We report total patient exposure time in 1,000 patient-year (KYr) units, uncalibrated and calibrated hazard ratios (HRs) and their 95% confidence intervals (CIs). Greyed out entries did not pass study diagnostics and are included only for completeness.

| | Patients | | Exposure (KYr) | | Outcomes | | Uncalibrated | | Calibrated | |
|---|---|---|---|---|---|---|---|---|---|---|
| | T | C | T | C | T | C | HR (95% CI) | $p$ | HR (95% CI) | $p$ |
| **PS-matching / on-treatment TAR** | | | | | | | | | | |
| CCAE | 9,683 | 47,980 | 6.6 | 41.3 | 137 | 863 | 1.01 (0.79 - 1.29) | 0.91 | 0.98 (0.77 - 1.26) | 0.85 |
| ODOD | 5,664 | 26,092 | 3.5 | 21.4 | 95 | 762 | 0.74 (0.54 - 0.98) | 0.04 | 0.72 (0.54 - 0.97) | 0.04 |
| MDCR | 736 | 7,646 | 0.5 | 7.9 | 27 | 334 | 1.47 (0.89 - 2.36) | 0.12 | 1.42 (0.87 - 2.32) | 0.17 |
| Open Claims | 102,361 | 374,892 | 79.1 | 355.3 | 1,075 | 6,758 | 0.94 (0.86 - 1.03) | 0.19 | 0.95 (0.86 - 1.05) | 0.43 |
| OEHR | 7,365 | 35,861 | 2.2 | 17.6 | 44 | 395 | 1.29 (0.86 - 1.89) | 0.21 | 1.12 (0.75 - 1.67) | 0.59 |
| VA | 1,084 | 15,730 | 0.8 | 13.3 | 14 | 186 | 1.10 (0.51 - 2.14) | 0.79 | 0.90 (0.44 - 1.84) | 0.79 |
| Meta-analysis | 126,893 | 508,201 | 92.6 | 456.8 | 1,392 | 9,298 | 1.00 (0.85 - 1.17) | 0.96 | 0.98 (0.80 - 1.21) | 0.87 |
| Meta-analysis (w/o USOC) | 24,532 | 133,309 | 13.5 | 101.6 | 317 | 2,540 | 1.04 (0.82 - 1.33) | 0.73 | 1.00 (0.78 - 1.28) | 0.91 |
| **PS-stratification / on-treatment TAR** | | | | | | | | | | |
| CCAE | 11,380 | 48,125 | 7.3 | 41.4 | 145 | 868 | 1.01 (0.82 - 1.22) | 0.95 | 0.99 (0.81 - 1.21) | 0.87 |
| ODOD | 7,578 | 26,713 | 4.3 | 21.9 | 121 | 803 | 0.87 (0.69 - 1.08) | 0.22 | 0.84 (0.67 - 1.05) | 0.15 |
| MDCR | 819 | 8,073 | 0.5 | 8.3 | 27 | 365 | 1.32 (0.84 - 1.99) | 0.21 | 1.22 (0.79 - 1.89) | 0.37 |
| Open Claims | 138,123 | 377,644 | 95.0 | 357.7 | 1,277 | 6,968 | 0.93 (0.87 - 1.00) | 0.04 | 0.96 (0.89 - 1.04) | 0.49 |
| OEHR | 8,578 | 35,940 | 2.4 | 17.7 | 50 | 402 | 1.18 (0.85 - 1.62) | 0.31 | 1.03 (0.75 - 1.43) | 0.81 |
| VA | 1,260 | 22,854 | 0.9 | 21.3 | 15 | 298 | 1.10 (0.60 - 1.91) | 0.74 | 0.97 (0.54 - 1.74) | 0.88 |
| Meta-analysis | 165,659 | 488,422 | 109.0 | 438.8 | 1,593 | 9,041 | 0.94 (0.88 - 1.00) | 0.05 | 0.95 (0.82 - 1.10) | 0.50 |
| Meta-analysis (w/o USOC) | 27,536 | 110,778 | 14.0 | 81.1 | 316 | 2,073 | 0.98 (0.85 - 1.15) | 0.84 | 0.95 (0.80 - 1.11) | 0.56 |
| **PS-matching / on-treatment TAR without escalation** | | | | | | | | | | |
| CCAE | 9,683 | 47,980 | 5.7 | 33.6 | 118 | 696 | 1.08 (0.82 - 1.41) | 0.57 | 1.06 (0.81 - 1.39) | 0.69 |
| ODOD | 5,664 | 26,092 | 3.1 | 17.6 | 86 | 632 | 0.77 (0.56 - 1.05) | 0.10 | 0.74 (0.54 - 1.01) | 0.07 |
| MDCR | 736 | 7,646 | 0.4 | 6.7 | 21 | 283 | 1.17 (0.64 - 2.01) | 0.59 | 1.01 (0.57 - 1.78) | 0.84 |
| Open Claims | 102,361 | 374,892 | 68.3 | 275.4 | 925 | 5,348 | 0.94 (0.85 - 1.03) | 0.18 | 0.93 (0.84 - 1.03) | 0.21 |
| OEHR | 7,365 | 35,861 | 1.9 | 14.4 | 40 | 305 | 1.42 (0.91 - 2.16) | 0.11 | 1.23 (0.80 - 1.89) | 0.36 |
| VA | 1,084 | 15,730 | 0.6 | 10.9 | 11 | 152 | 1.25 (0.52 - 2.63) | 0.60 | 0.95 (0.42 - 2.14) | 0.87 |
| Meta-analysis | 126,893 | 508,201 | 80.0 | 358.6 | 1,201 | 7,416 | 0.99 (0.86 - 1.15) | 0.94 | 0.97 (0.82 - 1.14) | 0.72 |
| Meta-analysis (w/o USOC) | 24,532 | 133,309 | 11.7 | 83.2 | 276 | 2,068 | 1.06 (0.84 - 1.32) | 0.64 | 0.99 (0.79 - 1.25) | 0.88 |
| **PS-stratification / on-treatment TAR without escalation** | | | | | | | | | | |
| CCAE | 11,380 | 48,125 | 6.4 | 33.7 | 125 | 701 | 1.03 (0.83 - 1.27) | 0.80 | 1.00 (0.81 - 1.24) | 0.88 |
| ODOD | 7,578 | 26,713 | 3.8 | 18.0 | 110 | 670 | 0.89 (0.69 - 1.12) | 0.33 | 0.83 (0.65 - 1.06) | 0.16 |
| MDCR | 819 | 8,073 | 0.4 | 7.1 | 21 | 309 | 1.07 (0.64 - 1.70) | 0.79 | 0.98 (0.60 - 1.60) | 0.85 |
| Open Claims | 138,123 | 377,644 | 83.1 | 277.4 | 1,115 | 5,531 | 0.92 (0.86 - 1.00) | 0.04 | 0.94 (0.87 - 1.03) | 0.26 |
| OEHR | 8,578 | 35,940 | 2.1 | 14.4 | 45 | 311 | 1.28 (0.89 - 1.80) | 0.16 | 1.08 (0.76 - 1.54) | 0.67 |
| VA | 1,260 | 22,854 | 0.7 | 17.3 | 12 | 243 | 1.17 (0.59 - 2.16) | 0.63 | 0.98 (0.51 - 1.89) | 0.88 |
| Meta-analysis | 165,659 | 488,422 | 95.4 | 343.5 | 1,395 | 7,213 | 0.95 (0.88 - 1.02) | 0.17 | 0.94 (0.84 - 1.07) | 0.36 |
| Meta-analysis (w/o USOC) | 27,536 | 110,778 | 12.3 | 66.1 | 280 | 1,682 | 1.02 (0.85 - 1.22) | 0.82 | 0.97 (0.81 - 1.16) | 0.74 |

##### 1.4.17 Hyperkalemia

**Supplementary Table 95:** Exposures, outcomes and risk estimates for Hyperkalemia in GLP1RA (male) (T) vs DPP4I (male) (C) new-user cohorts under four different propensity score (PS) adjustment / time-at-risk (TAR) designs across data sources. We report total patient exposure time in 1,000 patient-year (KYr) units, uncalibrated and calibrated hazard ratios (HRs) and their 95% confidence intervals (CIs). Greyed out entries did not pass study diagnostics and are included only for completeness.

| | Patients | | Exposure (KYr) | | Outcomes | | Uncalibrated | | Calibrated | |
|---|---|---|---|---|---|---|---|---|---|---|
| | T | C | T | C | T | C | HR (95% CI) | $p$ | HR (95% CI) | $p$ |
| **PS-matching / on-treatment TAR** | | | | | | | | | | |
| CCAE | 10,450 | 51,940 | 7.2 | 45.3 | 47 | 321 | 0.81 (0.52 - 1.22) | 0.32 | 0.78 (0.51 - 1.19) | 0.26 |
| ODOD | 6,159 | 29,035 | 3.8 | 24.1 | 58 | 498 | 0.86 (0.58 - 1.24) | 0.44 | 0.85 (0.58 - 1.24) | 0.41 |
| MDCR | 834 | 8,711 | 0.6 | 9.1 | 7 | 134 | 0.63 (0.20 - 1.57) | 0.37 | 0.60 (0.22 - 1.68) | 0.34 |
| Open Claims | 113,360 | 417,653 | 87.4 | 398.4 | 351 | 3,208 | 0.71 (0.61 - 0.83) | <0.01 | 0.72 (0.62 - 0.84) | <0.01 |
| OEHR | 7,661 | 37,332 | 2.3 | 18.4 | 19 | 293 | 0.64 (0.36 - 1.09) | 0.12 | 0.56 (0.32 - 0.98) | 0.05 |
| VA | 1,134 | 16,675 | 0.8 | 14.0 | <10 | 189 | 0.83 (0.27 - 2.03) | 0.71 | 0.67 (0.24 - 1.85) | 0.46 |
| Meta-analysis | 139,598 | 561,346 | 102.1 | 509.3 | <492 | 4,643 | 0.73 (0.65 - 0.83) | <0.01 | 0.72 (0.60 - 0.87) | <0.01 |
| Meta-analysis (w/o USOC) | 26,238 | 143,693 | 14.7 | 110.9 | <141 | 1,435 | 0.78 (0.62 - 1.00) | 0.05 | 0.75 (0.59 - 0.96) | 0.02 |
| **PS-stratification / on-treatment TAR** | | | | | | | | | | |
| CCAE | 12,345 | 52,064 | 8.0 | 45.4 | 52 | 323 | 0.95 (0.68 - 1.30) | 0.74 | 0.93 (0.67 - 1.30) | 0.70 |
| ODOD | 8,212 | 29,533 | 4.7 | 24.6 | 70 | 515 | 0.87 (0.65 - 1.16) | 0.36 | 0.84 (0.63 - 1.13) | 0.27 |
| MDCR | 932 | 9,303 | 0.6 | 9.7 | 7 | 160 | 0.64 (0.26 - 1.38) | 0.30 | 0.60 (0.26 - 1.38) | 0.23 |
| Open Claims | 152,722 | 419,830 | 104.9 | 400.4 | 394 | 3,291 | 0.69 (0.62 - 0.78) | <0.01 | 0.72 (0.63 - 0.81) | <0.01 |
| OEHR | 8,937 | 37,431 | 2.5 | 18.5 | 20 | 294 | 0.55 (0.33 - 0.87) | 0.01 | 0.48 (0.30 - 0.78) | <0.01 |
| VA | 1,317 | 24,038 | 0.9 | 22.1 | <10 | 315 | 0.68 (0.30 - 1.37) | 0.33 | 0.60 (0.28 - 1.29) | 0.20 |
| Meta-analysis | 182,216 | 538,858 | 120.2 | 488.9 | 536 | 4,423 | 0.76 (0.63 - 0.91) | <0.01 | 0.77 (0.61 - 0.96) | 0.02 |
| Meta-analysis (w/o USOC) | 29,494 | 119,028 | 15.2 | 88.4 | 142 | 1,132 | 0.81 (0.62 - 1.06) | 0.13 | 0.78 (0.59 - 1.03) | 0.09 |
| **PS-matching / on-treatment TAR without escalation** | | | | | | | | | | |
| CCAE | 10,450 | 51,940 | 6.2 | 36.7 | 37 | 256 | 0.81 (0.50 - 1.29) | 0.40 | 0.80 (0.49 - 1.29) | 0.36 |
| ODOD | 6,159 | 29,035 | 3.4 | 19.8 | 51 | 406 | 0.88 (0.58 - 1.32) | 0.56 | 0.85 (0.56 - 1.28) | 0.45 |
| MDCR | 834 | 8,711 | 0.5 | 7.8 | 6 | 110 | 0.60 (0.17 - 1.65) | 0.38 | 0.52 (0.17 - 1.62) | 0.26 |
| Open Claims | 113,360 | 417,653 | 75.4 | 309.0 | 295 | 2,522 | 0.71 (0.60 - 0.83) | <0.01 | 0.70 (0.59 - 0.83) | <0.01 |
| OEHR | 7,661 | 37,332 | 2.0 | 15.0 | 14 | 222 | 0.60 (0.29 - 1.12) | 0.13 | 0.52 (0.27 - 1.01) | 0.06 |
| VA | 1,134 | 16,675 | 0.7 | 11.4 | <10 | 142 | 1.02 (0.29 - 2.75) | 0.97 | 0.78 (0.25 - 2.40) | 0.67 |
| Meta-analysis | 139,598 | 561,346 | 88.1 | 399.6 | <413 | 3,658 | 0.73 (0.64 - 0.84) | <0.01 | 0.71 (0.61 - 0.84) | <0.01 |
| Meta-analysis (w/o USOC) | 26,238 | 143,693 | 12.7 | 90.6 | <118 | 1,136 | 0.80 (0.61 - 1.04) | 0.10 | 0.75 (0.57 - 0.98) | 0.04 |
| **PS-stratification / on-treatment TAR without escalation** | | | | | | | | | | |
| CCAE | 12,345 | 52,064 | 7.0 | 36.7 | 40 | 258 | 0.93 (0.64 - 1.34) | 0.71 | 0.91 (0.62 - 1.32) | 0.62 |
| ODOD | 8,212 | 29,533 | 4.2 | 20.1 | 61 | 420 | 0.85 (0.61 - 1.16) | 0.31 | 0.80 (0.58 - 1.09) | 0.18 |
| MDCR | 932 | 9,303 | 0.5 | 8.3 | 6 | 134 | 0.68 (0.25 - 1.56) | 0.41 | 0.63 (0.25 - 1.56) | 0.32 |
| Open Claims | 152,722 | 419,830 | 91.7 | 310.6 | 334 | 2,584 | 0.68 (0.60 - 0.78) | <0.01 | 0.70 (0.61 - 0.80) | <0.01 |
| OEHR | 8,937 | 37,431 | 2.2 | 15.1 | 15 | 223 | 0.49 (0.27 - 0.82) | 0.01 | 0.41 (0.23 - 0.72) | <0.01 |
| VA | 1,317 | 24,038 | 0.7 | 18.0 | <10 | 243 | 0.64 (0.24 - 1.43) | 0.33 | 0.54 (0.22 - 1.31) | 0.18 |
| Meta-analysis | 182,216 | 538,858 | 105.1 | 382.6 | 450 | 3,485 | 0.74 (0.61 - 0.88) | <0.01 | 0.73 (0.60 - 0.90) | <0.01 |
| Meta-analysis (w/o USOC) | 29,494 | 119,028 | 13.4 | 71.9 | 116 | 901 | 0.78 (0.56 - 1.07) | 0.12 | 0.74 (0.53 - 1.01) | 0.07 |

##### 1.4.18 Hypoglycemia

**Supplementary Table 96:** Exposures, outcomes and risk estimates for Hypoglycemia in GLP1RA (male) (T) vs DPP4I (male) (C) new-user cohorts under four different propensity score (PS) adjustment / time-at-risk (TAR) designs across data sources. We report total patient exposure time in 1,000 patient-year (KYr) units, uncalibrated and calibrated hazard ratios (HRs) and their 95% confidence intervals (CIs). Greyed out entries did not pass study diagnostics and are included only for completeness.

| | Patients | | Exposure (KYr) | | Outcomes | | Uncalibrated | | Calibrated | |
|---|---|---|---|---|---|---|---|---|---|---|
| | T | C | T | C | T | C | HR (95% CI) | $p$ | HR (95% CI) | $p$ |
| **PS-matching / on-treatment TAR** | | | | | | | | | | |
| CCAE | 10,442 | 52,040 | 7.2 | 45.7 | 34 | 125 | 1.48 (0.86 - 2.50) | 0.15 | 1.43 (0.84 - 2.44) | 0.19 |
| ODOD | 6,324 | 29,836 | 3.9 | 25.1 | 22 | 124 | 1.44 (0.74 - 2.74) | 0.27 | 1.42 (0.74 - 2.74) | 0.30 |
| MDCR | 847 | 8,828 | 0.6 | 9.3 | <5 | 28 | 0.78 (0.04 - 5.18) | 0.84 | 0.75 (0.06 - 8.78) | 0.82 |
| Open Claims | 113,979 | 421,712 | 88.0 | 405.0 | 177 | 959 | 0.99 (0.79 - 1.24) | 0.95 | 1.00 (0.80 - 1.26) | 0.93 |
| OEHR | 7,722 | 37,931 | 2.3 | 18.9 | 13 | 103 | 0.73 (0.35 - 1.44) | 0.39 | 0.64 (0.31 - 1.30) | 0.23 |
| VA | 1,200 | 17,540 | 0.9 | 14.9 | 0 | 50 | | | | |
| Meta-analysis | 138,467 | 541,519 | 101.5 | 494.7 | 246 | 1,311 | 1.08 (0.85 - 1.36) | 0.53 | 1.06 (0.81 - 1.39) | 0.66 |
| Meta-analysis (w/o USOC) | 24,488 | 119,807 | 13.4 | 89.7 | 69 | 352 | 1.21 (0.80 - 1.85) | 0.37 | 1.16 (0.76 - 1.78) | 0.48 |
| **PS-stratification / on-treatment TAR** | | | | | | | | | | |
| CCAE | 12,307 | 52,171 | 8.0 | 45.8 | 36 | 125 | 1.35 (0.87 - 2.05) | 0.17 | 1.33 (0.87 - 2.05) | 0.20 |
| ODOD | 8,400 | 30,447 | 4.8 | 25.6 | 25 | 127 | 0.93 (0.55 - 1.54) | 0.79 | 0.90 (0.54 - 1.51) | 0.70 |
| MDCR | 944 | 9,475 | 0.6 | 9.9 | <5 | 34 | 0.54 (0.03 - 3.13) | 0.60 | 0.50 (0.05 - 5.28) | 0.56 |
| Open Claims | 153,146 | 424,002 | 105.4 | 407.1 | 219 | 972 | 0.91 (0.76 - 1.07) | 0.27 | 0.94 (0.79 - 1.12) | 0.52 |
| OEHR | 8,985 | 38,024 | 2.6 | 19.0 | 15 | 104 | 1.08 (0.58 - 1.89) | 0.80 | 0.95 (0.52 - 1.71) | 0.85 |
| VA | 1,388 | 25,385 | 1.0 | 23.8 | <10 | 78 | 0.21 (0.01 - 1.08) | 0.18 | 0.19 (0.02 - 1.81) | 0.15 |
| Meta-analysis | 182,838 | 544,644 | 120.8 | 497.5 | 295 | 1,328 | 0.99 (0.82 - 1.21) | 0.95 | 1.00 (0.79 - 1.27) | 0.93 |
| Meta-analysis (w/o USOC) | 29,692 | 120,642 | 15.4 | 90.4 | 76 | 356 | 1.14 (0.85 - 1.52) | 0.37 | 1.10 (0.82 - 1.47) | 0.56 |
| **PS-matching / on-treatment TAR without escalation** | | | | | | | | | | |
| CCAE | 10,442 | 52,040 | 6.2 | 37.0 | 25 | 83 | 1.40 (0.76 - 2.51) | 0.27 | 1.37 (0.76 - 2.49) | 0.30 |
| ODOD | 6,324 | 29,836 | 3.5 | 20.6 | 16 | 65 | 1.63 (0.69 - 3.61) | 0.25 | 1.56 (0.68 - 3.56) | 0.30 |
| MDCR | 847 | 8,828 | 0.5 | 7.9 | 0 | 16 | | | | |
| Open Claims | 113,979 | 421,712 | 76.0 | 314.0 | 133 | 534 | 1.16 (0.89 - 1.51) | 0.26 | 1.16 (0.89 - 1.51) | 0.30 |
| OEHR | 7,722 | 37,931 | 2.0 | 15.4 | 11 | 50 | 0.84 (0.29 - 2.12) | 0.73 | 0.73 (0.27 - 1.96) | 0.53 |
| VA | 1,200 | 17,540 | 0.7 | 12.2 | 0 | 30 | | | | |
| Meta-analysis | 138,467 | 541,519 | 87.7 | 387.0 | 185 | 732 | 1.20 (0.96 - 1.51) | 0.11 | 1.18 (0.93 - 1.49) | 0.18 |
| Meta-analysis (w/o USOC) | 24,488 | 119,807 | 11.7 | 72.9 | 52 | 198 | 1.32 (0.86 - 2.04) | 0.21 | 1.24 (0.80 - 1.92) | 0.33 |
| **PS-stratification / on-treatment TAR without escalation** | | | | | | | | | | |
| CCAE | 12,307 | 52,171 | 7.0 | 37.1 | 27 | 83 | 1.36 (0.81 - 2.22) | 0.23 | 1.32 (0.80 - 2.19) | 0.29 |
| ODOD | 8,400 | 30,447 | 4.3 | 21.0 | 19 | 67 | 1.21 (0.64 - 2.22) | 0.54 | 1.14 (0.61 - 2.12) | 0.69 |
| MDCR | 944 | 9,475 | 0.5 | 8.5 | 0 | 20 | | | | |
| Open Claims | 153,146 | 424,002 | 92.1 | 315.7 | 173 | 537 | 1.03 (0.84 - 1.26) | 0.79 | 1.05 (0.85 - 1.29) | 0.66 |
| OEHR | 8,985 | 38,024 | 2.2 | 15.4 | 12 | 51 | 1.66 (0.80 - 3.24) | 0.16 | 1.40 (0.70 - 2.83) | 0.35 |
| VA | 1,388 | 25,385 | 0.8 | 19.3 | 0 | 44 | | | | |
| Meta-analysis | 182,838 | 544,644 | 105.6 | 389.2 | 231 | 738 | 1.14 (0.92 - 1.42) | 0.22 | 1.14 (0.90 - 1.44) | 0.28 |
| Meta-analysis (w/o USOC) | 29,692 | 120,642 | 13.5 | 73.5 | 58 | 201 | 1.38 (0.98 - 1.94) | 0.07 | 1.31 (0.93 - 1.84) | 0.13 |

##### 1.4.19 Hypotension

**Supplementary Table 97:** Exposures, outcomes and risk estimates for Hypotension in GLP1RA (male) (T) vs DPP4I (male) (C) new-user cohorts under four different propensity score (PS) adjustment / time-at-risk (TAR) designs across data sources. We report total patient exposure time in 1,000 patient-year (KYr) units, uncalibrated and calibrated hazard ratios (HRs) and their 95% confidence intervals (CIs). Greyed out entries did not pass study diagnostics and are included only for completeness.

| | Patients | | Exposure (KYr) | | Outcomes | | Uncalibrated | | Calibrated | |
|---|---|---|---|---|---|---|---|---|---|---|
| | T | C | T | C | T | C | HR (95% CI) | $p$ | HR (95% CI) | $p$ |
| **PS-matching / on-treatment TAR** | | | | | | | | | | |
| CCAE | 10,416 | 51,890 | 7.2 | 45.3 | 70 | 346 | 1.18 (0.81 - 1.69) | 0.38 | 1.14 (0.79 - 1.65) | 0.49 |
| ODOD | 6,168 | 28,752 | 3.8 | 24.0 | 66 | 487 | 0.99 (0.68 - 1.40) | 0.95 | 0.97 (0.68 - 1.39) | 0.86 |
| MDCR | 818 | 8,345 | 0.6 | 8.7 | 15 | 225 | 0.73 (0.34 - 1.43) | 0.40 | 0.71 (0.35 - 1.45) | 0.35 |
| Open Claims | 111,950 | 412,509 | 86.4 | 394.4 | 541 | 3,678 | 0.91 (0.81 - 1.03) | 0.15 | 0.92 (0.81 - 1.05) | 0.29 |
| OEHR | 7,635 | 37,341 | 2.3 | 18.4 | 37 | 338 | 1.06 (0.70 - 1.58) | 0.77 | 0.92 (0.61 - 1.40) | 0.73 |
| VA | 1,130 | 16,781 | 0.8 | 14.1 | 15 | 178 | 1.22 (0.57 - 2.41) | 0.58 | 1.00 (0.48 - 2.06) | 0.88 |
| Meta-analysis | 138,117 | 555,618 | 101.1 | 505.0 | 744 | 5,252 | 0.95 (0.86 - 1.05) | 0.33 | 0.94 (0.79 - 1.11) | 0.45 |
| Meta-analysis (w/o USOC) | 26,167 | 143,109 | 14.7 | 110.6 | 203 | 1,574 | 1.05 (0.86 - 1.29) | 0.61 | 1.01 (0.83 - 1.24) | 0.87 |
| **PS-stratification / on-treatment TAR** | | | | | | | | | | |
| CCAE | 12,285 | 52,015 | 8.0 | 45.5 | 75 | 348 | 1.20 (0.90 - 1.58) | 0.21 | 1.18 (0.89 - 1.57) | 0.27 |
| ODOD | 8,191 | 29,398 | 4.7 | 24.6 | 76 | 520 | 0.99 (0.74 - 1.30) | 0.92 | 0.95 (0.72 - 1.27) | 0.75 |
| MDCR | 914 | 8,981 | 0.6 | 9.3 | 16 | 263 | 1.18 (0.66 - 1.98) | 0.56 | 1.10 (0.63 - 1.91) | 0.75 |
| Open Claims | 150,746 | 414,545 | 103.6 | 396.2 | 624 | 3,770 | 0.87 (0.79 - 0.96) | <0.01 | 0.90 (0.81 - 1.00) | 0.12 |
| OEHR | 8,903 | 37,424 | 2.5 | 18.5 | 38 | 345 | 1.00 (0.68 - 1.41) | 0.98 | 0.87 (0.61 - 1.25) | 0.47 |
| VA | 1,316 | 24,071 | 0.9 | 22.3 | 18 | 285 | 1.50 (0.85 - 2.53) | 0.14 | 1.32 (0.77 - 2.29) | 0.33 |
| Meta-analysis | 180,125 | 533,382 | 118.8 | 484.7 | 813 | 4,983 | 0.97 (0.83 - 1.13) | 0.67 | 0.98 (0.80 - 1.20) | 0.82 |
| Meta-analysis (w/o USOC) | 29,379 | 118,837 | 15.2 | 88.5 | 189 | 1,213 | 1.06 (0.89 - 1.27) | 0.49 | 1.02 (0.85 - 1.23) | 0.82 |
| **PS-matching / on-treatment TAR without escalation** | | | | | | | | | | |
| CCAE | 10,416 | 51,890 | 6.2 | 36.7 | 57 | 276 | 1.15 (0.77 - 1.68) | 0.48 | 1.13 (0.76 - 1.66) | 0.55 |
| ODOD | 6,168 | 28,752 | 3.4 | 19.7 | 56 | 378 | 1.00 (0.66 - 1.48) | 0.99 | 0.96 (0.64 - 1.43) | 0.83 |
| MDCR | 818 | 8,345 | 0.5 | 7.4 | 12 | 189 | 0.56 (0.21 - 1.27) | 0.21 | 0.48 (0.19 - 1.20) | 0.12 |
| Open Claims | 111,950 | 412,509 | 74.5 | 305.7 | 458 | 2,844 | 0.89 (0.77 - 1.02) | 0.09 | 0.88 (0.77 - 1.01) | 0.10 |
| OEHR | 7,635 | 37,341 | 1.9 | 15.1 | 36 | 235 | 1.32 (0.85 - 2.00) | 0.21 | 1.14 (0.74 - 1.75) | 0.57 |
| VA | 1,130 | 16,781 | 0.7 | 11.6 | 13 | 134 | 1.25 (0.55 - 2.61) | 0.57 | 0.95 (0.44 - 2.08) | 0.87 |
| Meta-analysis | 138,117 | 555,618 | 87.2 | 396.1 | 632 | 4,056 | 0.99 (0.83 - 1.17) | 0.89 | 0.96 (0.80 - 1.16) | 0.70 |
| Meta-analysis (w/o USOC) | 26,167 | 143,109 | 12.7 | 90.4 | 174 | 1,212 | 1.10 (0.89 - 1.37) | 0.38 | 1.04 (0.83 - 1.29) | 0.76 |
| **PS-stratification / on-treatment TAR without escalation** | | | | | | | | | | |
| CCAE | 12,285 | 52,015 | 6.9 | 36.8 | 61 | 278 | 1.15 (0.83 - 1.57) | 0.39 | 1.12 (0.81 - 1.54) | 0.50 |
| ODOD | 8,191 | 29,398 | 4.1 | 20.1 | 65 | 407 | 0.99 (0.72 - 1.35) | 0.96 | 0.93 (0.68 - 1.28) | 0.67 |
| MDCR | 914 | 8,981 | 0.5 | 7.9 | 13 | 226 | 1.01 (0.52 - 1.81) | 0.97 | 0.93 (0.50 - 1.73) | 0.81 |
| Open Claims | 150,746 | 414,545 | 90.6 | 307.2 | 534 | 2,921 | 0.89 (0.80 - 0.99) | 0.04 | 0.91 (0.81 - 1.02) | 0.14 |
| OEHR | 8,903 | 37,424 | 2.2 | 15.1 | 37 | 240 | 1.34 (0.90 - 1.95) | 0.14 | 1.13 (0.77 - 1.67) | 0.54 |
| VA | 1,316 | 24,071 | 0.7 | 18.1 | 15 | 210 | 1.60 (0.84 - 2.88) | 0.13 | 1.34 (0.72 - 2.48) | 0.36 |
| Meta-analysis | 180,125 | 533,382 | 103.9 | 379.2 | 697 | 3,846 | 1.02 (0.85 - 1.23) | 0.82 | 1.02 (0.83 - 1.25) | 0.86 |
| Meta-analysis (w/o USOC) | 29,379 | 118,837 | 13.3 | 72.0 | 163 | 925 | 1.13 (0.93 - 1.37) | 0.21 | 1.07 (0.88 - 1.30) | 0.51 |

##### 1.4.20 Joint pain

**Supplementary Table 98:** Exposures, outcomes and risk estimates for Joint pain in GLP1RA (male) (T) vs DPP4I (male) (C) new-user cohorts under four different propensity score (PS) adjustment / time-at-risk (TAR) designs across data sources. We report total patient exposure time in 1,000 patient-year (KYr) units, uncalibrated and calibrated hazard ratios (HRs) and their 95% confidence intervals (CIs). Greyed out entries did not pass study diagnostics and are included only for completeness.

| | Patients | | Exposure (KYr) | | Outcomes | | Uncalibrated | | Calibrated | |
|---|---|---|---|---|---|---|---|---|---|---|
| | T | C | T | C | T | C | HR (95% CI) | $p$ | HR (95% CI) | $p$ |
| **PS-matching / on-treatment TAR** | | | | | | | | | | |
| CCAE | 10,133 | 50,771 | 6.9 | 44.3 | 55 | 376 | 0.78 (0.53 - 1.13) | 0.20 | 0.76 (0.52 - 1.11) | 0.16 |
| ODOD | 6,076 | 28,925 | 3.8 | 24.2 | 31 | 261 | 0.95 (0.57 - 1.52) | 0.84 | 0.94 (0.57 - 1.53) | 0.80 |
| MDCR | 823 | 8,563 | 0.6 | 9.0 | <5 | 83 | 0.18 (0.01 - 0.92) | 0.14 | 0.18 (0.02 - 1.69) | 0.13 |
| Open Claims | 109,630 | 408,535 | 84.6 | 391.2 | 397 | 2,067 | 0.86 (0.74 - 1.01) | 0.06 | 0.87 (0.74 - 1.02) | 0.13 |
| OEHR | 7,460 | 36,748 | 2.2 | 18.2 | 13 | 151 | 0.58 (0.26 - 1.18) | 0.16 | 0.50 (0.24 - 1.08) | 0.09 |
| VA | 1,078 | 15,142 | 0.8 | 12.9 | <10 | 71 | 2.19 (0.82 - 5.32) | 0.10 | 1.78 (0.70 - 4.53) | 0.24 |
| Meta-analysis | 135,200 | 548,684 | 98.9 | 499.9 | <511 | 3,009 | 0.86 (0.75 - 0.98) | 0.03 | 0.85 (0.70 - 1.03) | 0.09 |
| Meta-analysis (w/o USOC) | 25,570 | 140,149 | 14.3 | 108.7 | <114 | 942 | 0.86 (0.57 - 1.30) | 0.49 | 0.83 (0.55 - 1.25) | 0.38 |
| **PS-stratification / on-treatment TAR** | | | | | | | | | | |
| CCAE | 11,936 | 50,908 | 7.7 | 44.5 | 62 | 376 | 0.81 (0.60 - 1.09) | 0.17 | 0.80 (0.59 - 1.08) | 0.17 |
| ODOD | 8,069 | 29,493 | 4.6 | 24.7 | 44 | 265 | 0.83 (0.56 - 1.20) | 0.33 | 0.80 (0.55 - 1.17) | 0.26 |
| MDCR | 913 | 9,268 | 0.6 | 9.7 | <5 | 90 | 0.49 (0.14 - 1.27) | 0.20 | 0.46 (0.15 - 1.36) | 0.16 |
| Open Claims | 147,146 | 410,683 | 101.3 | 393.3 | 492 | 2,082 | 0.98 (0.87 - 1.09) | 0.69 | 1.01 (0.90 - 1.14) | 0.87 |
| OEHR | 8,667 | 36,838 | 2.5 | 18.3 | 16 | 152 | 0.80 (0.44 - 1.35) | 0.42 | 0.70 (0.40 - 1.22) | 0.21 |
| VA | 1,247 | 22,503 | 0.9 | 21.2 | 12 | 108 | 2.30 (1.09 - 4.54) | 0.02 | 2.02 (0.99 - 4.13) | 0.06 |
| Meta-analysis | 175,818 | 527,922 | 116.1 | 480.7 | 614 | 2,875 | 0.92 (0.81 - 1.05) | 0.21 | 0.93 (0.77 - 1.12) | 0.45 |
| Meta-analysis (w/o USOC) | 28,672 | 117,239 | 14.8 | 87.4 | 122 | 793 | 0.82 (0.66 - 1.01) | 0.06 | 0.78 (0.63 - 0.98) | 0.05 |
| **PS-matching / on-treatment TAR without escalation** | | | | | | | | | | |
| CCAE | 10,133 | 50,771 | 6.0 | 35.9 | 46 | 310 | 0.83 (0.53 - 1.25) | 0.39 | 0.81 (0.53 - 1.25) | 0.35 |
| ODOD | 6,076 | 28,925 | 3.3 | 19.9 | 27 | 222 | 0.86 (0.50 - 1.41) | 0.57 | 0.82 (0.49 - 1.39) | 0.48 |
| MDCR | 823 | 8,563 | 0.5 | 7.7 | <5 | 70 | 0.19 (0.01 - 0.98) | 0.15 | 0.16 (0.02 - 1.60) | 0.12 |
| Open Claims | 109,630 | 408,535 | 73.0 | 303.5 | 333 | 1,569 | 0.89 (0.75 - 1.06) | 0.19 | 0.88 (0.74 - 1.06) | 0.20 |
| OEHR | 7,460 | 36,748 | 1.9 | 14.8 | 13 | 119 | 0.57 (0.24 - 1.21) | 0.17 | 0.49 (0.22 - 1.11) | 0.09 |
| VA | 1,078 | 15,142 | 0.6 | 10.5 | <10 | 56 | 3.21 (1.13 - 8.49) | 0.02 | 2.45 (0.89 - 6.70) | 0.09 |
| Meta-analysis | 135,200 | 548,684 | 85.4 | 392.3 | <434 | 2,346 | 0.89 (0.76 - 1.03) | 0.11 | 0.86 (0.73 - 1.02) | 0.09 |
| Meta-analysis (w/o USOC) | 25,570 | 140,149 | 12.4 | 88.8 | <101 | 777 | 0.90 (0.54 - 1.51) | 0.69 | 0.85 (0.50 - 1.42) | 0.53 |
| **PS-stratification / on-treatment TAR without escalation** | | | | | | | | | | |
| CCAE | 11,936 | 50,908 | 6.8 | 36.0 | 52 | 310 | 0.78 (0.56 - 1.08) | 0.14 | 0.76 (0.55 - 1.06) | 0.12 |
| ODOD | 8,069 | 29,493 | 4.1 | 20.2 | 39 | 226 | 0.84 (0.56 - 1.24) | 0.40 | 0.79 (0.53 - 1.18) | 0.26 |
| MDCR | 913 | 9,268 | 0.5 | 8.3 | <5 | 76 | 0.52 (0.15 - 1.37) | 0.25 | 0.48 (0.16 - 1.44) | 0.19 |
| Open Claims | 147,146 | 410,683 | 88.5 | 305.0 | 423 | 1,578 | 1.00 (0.88 - 1.13) | 0.98 | 1.02 (0.89 - 1.16) | 0.78 |
| OEHR | 8,667 | 36,838 | 2.1 | 14.9 | 16 | 120 | 0.98 (0.53 - 1.68) | 0.94 | 0.83 (0.47 - 1.47) | 0.53 |
| VA | 1,247 | 22,503 | 0.7 | 17.2 | 11 | 85 | 2.66 (1.20 - 5.52) | 0.01 | 2.22 (1.03 - 4.78) | 0.04 |
| Meta-analysis | 175,818 | 527,922 | 101.5 | 376.2 | 530 | 2,234 | 0.94 (0.82 - 1.08) | 0.39 | 0.94 (0.79 - 1.11) | 0.45 |
| Meta-analysis (w/o USOC) | 28,672 | 117,239 | 13.0 | 71.1 | 107 | 656 | 0.83 (0.66 - 1.05) | 0.12 | 0.79 (0.62 - 1.00) | 0.05 |

##### 1.4.21 Lower extremity amputation

**Supplementary Table 99:** Exposures, outcomes and risk estimates for Lower extremity amputation in GLP1RA (male) (T) vs DPP4I (male) (C) new-user cohorts under four different propensity score (PS) adjustment / time-at-risk (TAR) designs across data sources. We report total patient exposure time in 1,000 patient-year (KYr) units, uncalibrated and calibrated hazard ratios (HRs) and their 95% confidence intervals (CIs). Greyed out entries did not pass study diagnostics and are included only for completeness.

| | Patients | | Exposure (KYr) | | Outcomes | | Uncalibrated | | Calibrated | |
|---|---|---|---|---|---|---|---|---|---|---|
| | T | C | T | C | T | C | HR (95% CI) | $p$ | HR (95% CI) | $p$ |
| **PS-matching / on-treatment TAR** | | | | | | | | | | |
| CCAE | 10,595 | 52,689 | 7.3 | 46.3 | 6 | 33 | 0.97 (0.31 - 2.55) | 0.95 | 0.94 (0.32 - 2.70) | 0.90 |
| ODOD | 6,392 | 30,161 | 4.0 | 25.5 | <5 | 26 | 1.09 (0.15 - 5.24) | 0.93 | 1.07 (0.18 - 6.34) | 0.94 |
| MDCR | 854 | 8,887 | 0.6 | 9.4 | 0 | 8 | | | | |
| Open Claims | 115,432 | 426,667 | 89.2 | 410.0 | 46 | 239 | 0.80 (0.49 - 1.27) | 0.37 | 0.81 (0.50 - 1.31) | 0.40 |
| OEHR | 7,797 | 38,157 | 2.3 | 19.1 | 0 | 16 | | | | |
| VA | 1,210 | 17,651 | 0.9 | 15.0 | 0 | <10 | | | | |
| Meta-analysis | 132,419 | 509,517 | 100.5 | 481.9 | <57 | 298 | 0.84 (0.55 - 1.28) | 0.42 | 0.83 (0.53 - 1.29) | 0.41 |
| Meta-analysis (w/o USOC) | 16,987 | 82,850 | 11.3 | 71.8 | <11 | 59 | 1.00 (0.40 - 2.48) | 1.00 | 0.96 (0.39 - 2.38) | 0.93 |
| **PS-stratification / on-treatment TAR** | | | | | | | | | | |
| CCAE | 12,517 | 52,823 | 8.2 | 46.5 | 7 | 33 | 1.53 (0.58 - 3.56) | 0.36 | 1.51 (0.61 - 3.75) | 0.38 |
| ODOD | 8,500 | 30,785 | 4.9 | 26.0 | 6 | 28 | 0.84 (0.28 - 2.24) | 0.74 | 0.81 (0.28 - 2.31) | 0.70 |
| MDCR | 952 | 9,542 | 0.6 | 10.0 | 0 | 10 | | | | |
| Open Claims | 155,300 | 428,959 | 107.0 | 412.2 | 49 | 244 | 0.93 (0.65 - 1.30) | 0.66 | 0.96 (0.67 - 1.36) | 0.81 |
| OEHR | 9,096 | 38,250 | 2.6 | 19.2 | 0 | 16 | | | | |
| VA | 1,403 | 25,577 | 1.0 | 24.0 | 0 | 13 | | | | |
| Meta-analysis | 176,317 | 512,567 | 120.0 | 484.7 | 62 | 305 | 0.97 (0.71 - 1.33) | 0.86 | 0.98 (0.70 - 1.38) | 0.92 |
| Meta-analysis (w/o USOC) | 21,017 | 83,608 | 13.0 | 72.5 | 13 | 61 | 1.18 (0.60 - 2.35) | 0.63 | 1.14 (0.57 - 2.26) | 0.72 |
| **PS-matching / on-treatment TAR without escalation** | | | | | | | | | | |
| CCAE | 10,595 | 52,689 | 6.3 | 37.5 | 6 | 25 | 1.16 (0.36 - 3.24) | 0.79 | 1.14 (0.38 - 3.43) | 0.82 |
| ODOD | 6,392 | 30,161 | 3.5 | 20.8 | <5 | 19 | 1.09 (0.14 - 5.88) | 0.93 | 1.05 (0.16 - 6.72) | 0.96 |
| MDCR | 854 | 8,887 | 0.5 | 8.0 | 0 | 8 | | | | |
| Open Claims | 115,432 | 426,667 | 76.9 | 317.7 | 37 | 182 | 0.86 (0.49 - 1.43) | 0.57 | 0.85 (0.50 - 1.45) | 0.56 |
| OEHR | 7,797 | 38,157 | 2.0 | 15.5 | 0 | 11 | | | | |
| VA | 1,210 | 17,651 | 0.7 | 12.2 | 0 | <10 | | | | |
| Meta-analysis | 126,027 | 479,356 | 83.3 | 355.2 | 43 | 207 | 0.91 (0.56 - 1.47) | 0.69 | 0.89 (0.54 - 1.44) | 0.63 |
| Meta-analysis (w/o USOC) | 10,595 | 52,689 | 6.3 | 37.5 | 6 | 25 | 1.16 (0.36 - 3.24) | 0.79 | 1.09 (0.36 - 3.29) | 0.87 |
| **PS-stratification / on-treatment TAR without escalation** | | | | | | | | | | |
| CCAE | 12,517 | 52,823 | 7.1 | 37.6 | 6 | 25 | 1.85 (0.65 - 4.60) | 0.22 | 1.80 (0.67 - 4.80) | 0.24 |
| ODOD | 8,500 | 30,785 | 4.3 | 21.3 | 5 | 21 | 0.91 (0.25 - 2.79) | 0.87 | 0.85 (0.26 - 2.82) | 0.79 |
| MDCR | 952 | 9,542 | 0.6 | 8.5 | 0 | 10 | | | | |
| Open Claims | 155,300 | 428,959 | 93.4 | 319.4 | 39 | 185 | 0.94 (0.63 - 1.38) | 0.76 | 0.96 (0.65 - 1.43) | 0.85 |
| OEHR | 9,096 | 38,250 | 2.3 | 15.6 | 0 | 11 | | | | |
| VA | 1,403 | 25,577 | 0.8 | 19.5 | 0 | 11 | | | | |
| Meta-analysis | 176,317 | 512,567 | 104.9 | 378.2 | 50 | 231 | 1.02 (0.72 - 1.45) | 0.91 | 1.02 (0.71 - 1.46) | 0.92 |
| Meta-analysis (w/o USOC) | 21,017 | 83,608 | 11.4 | 58.8 | 11 | 46 | 1.39 (0.65 - 2.97) | 0.40 | 1.32 (0.62 - 2.81) | 0.48 |

##### 1.4.22 Nausea

**Supplementary Table 100:** Exposures, outcomes and risk estimates for Nausea in GLP1RA (male) (T) vs DPP4I (male) (C) new-user cohorts under four different propensity score (PS) adjustment / time-at-risk (TAR) designs across data sources. We report total patient exposure time in 1,000 patient-year (KYr) units, uncalibrated and calibrated hazard ratios (HRs) and their 95% confidence intervals (CIs). Greyed out entries did not pass study diagnostics and are included only for completeness.

| | Patients | | Exposure (KYr) | | Outcomes | | Uncalibrated | | Calibrated | |
|---|---|---|---|---|---|---|---|---|---|---|
| | T | C | T | C | T | C | HR (95% CI) | $p$ | HR (95% CI) | $p$ |
| **PS-matching / on-treatment TAR** | | | | | | | | | | |
| CCAE | 9,578 | 48,902 | 6.5 | 42.4 | 221 | 971 | 1.43 (1.17 - 1.75) | <0.01 | 1.39 (1.13 - 1.70) | <0.01 |
| ODOD | 5,655 | 27,190 | 3.5 | 22.5 | 151 | 701 | 1.32 (1.03 - 1.69) | 0.03 | 1.30 (1.01 - 1.67) | 0.06 |
| MDCR | 783 | 8,162 | 0.5 | 8.5 | 12 | 209 | 1.16 (0.55 - 2.25) | 0.67 | 1.12 (0.56 - 2.27) | 0.74 |
| Open Claims | 102,841 | 388,233 | 79.3 | 370.9 | 1,572 | 5,699 | 1.45 (1.34 - 1.57) | <0.01 | 1.46 (1.34 - 1.60) | <0.01 |
| OEHR | 7,171 | 35,809 | 2.1 | 17.5 | 106 | 457 | 1.77 (1.33 - 2.33) | <0.01 | 1.54 (1.16 - 2.04) | <0.01 |
| VA | 1,133 | 16,631 | 0.8 | 14.2 | 22 | 121 | 1.98 (0.96 - 3.86) | 0.06 | 1.61 (0.80 - 3.24) | 0.20 |
| Meta-analysis | 127,161 | 524,927 | 92.8 | 476.1 | 2,084 | 8,158 | 1.46 (1.36 - 1.56) | <0.01 | 1.44 (1.24 - 1.67) | <0.01 |
| Meta-analysis (w/o USOC) | 24,320 | 136,694 | 13.5 | 105.2 | 512 | 2,459 | 1.47 (1.29 - 1.68) | <0.01 | 1.41 (1.24 - 1.62) | <0.01 |
| **PS-stratification / on-treatment TAR** | | | | | | | | | | |
| CCAE | 11,233 | 49,023 | 7.3 | 42.5 | 258 | 977 | 1.27 (1.08 - 1.48) | <0.01 | 1.25 (1.07 - 1.47) | 0.01 |
| ODOD | 7,529 | 27,806 | 4.2 | 23.0 | 197 | 724 | 1.34 (1.10 - 1.62) | <0.01 | 1.29 (1.06 - 1.57) | 0.02 |
| MDCR | 870 | 8,802 | 0.6 | 9.1 | 12 | 238 | 0.99 (0.51 - 1.78) | 0.98 | 0.92 (0.49 - 1.73) | 0.80 |
| Open Claims | 137,078 | 390,524 | 94.4 | 373.0 | 2,003 | 5,784 | 1.35 (1.27 - 1.44) | <0.01 | 1.40 (1.30 - 1.50) | <0.01 |
| OEHR | 8,322 | 35,895 | 2.3 | 17.6 | 125 | 462 | 1.90 (1.51 - 2.37) | <0.01 | 1.66 (1.33 - 2.08) | <0.01 |
| VA | 1,316 | 23,930 | 0.9 | 22.4 | 22 | 192 | 2.38 (1.37 - 3.99) | <0.01 | 2.09 (1.23 - 3.58) | <0.01 |
| Meta-analysis | 164,162 | 503,248 | 108.2 | 456.1 | 2,583 | 7,947 | 1.42 (1.22 - 1.66) | <0.01 | 1.43 (1.17 - 1.76) | <0.01 |
| Meta-analysis (w/o USOC) | 27,084 | 112,724 | 13.8 | 83.1 | 580 | 2,163 | 1.46 (1.16 - 1.84) | <0.01 | 1.40 (1.11 - 1.78) | <0.01 |
| **PS-matching / on-treatment TAR without escalation** | | | | | | | | | | |
| CCAE | 9,578 | 48,902 | 5.7 | 34.4 | 197 | 800 | 1.44 (1.16 - 1.78) | <0.01 | 1.41 (1.14 - 1.75) | <0.01 |
| ODOD | 5,655 | 27,190 | 3.1 | 18.5 | 140 | 576 | 1.47 (1.12 - 1.90) | <0.01 | 1.40 (1.07 - 1.83) | 0.02 |
| MDCR | 783 | 8,162 | 0.5 | 7.2 | 9 | 180 | 1.04 (0.44 - 2.18) | 0.93 | 0.89 (0.40 - 2.00) | 0.79 |
| Open Claims | 102,841 | 388,233 | 68.6 | 288.6 | 1,358 | 4,337 | 1.54 (1.41 - 1.68) | <0.01 | 1.53 (1.40 - 1.67) | <0.01 |
| OEHR | 7,171 | 35,809 | 1.8 | 14.4 | 88 | 336 | 1.75 (1.27 - 2.38) | <0.01 | 1.51 (1.10 - 2.06) | 0.01 |
| VA | 1,133 | 16,631 | 0.6 | 11.6 | 19 | 97 | 1.77 (0.76 - 3.80) | 0.16 | 1.35 (0.60 - 3.01) | 0.48 |
| Meta-analysis | 127,161 | 524,927 | 80.3 | 374.8 | 1,811 | 6,326 | 1.53 (1.42 - 1.65) | <0.01 | 1.49 (1.35 - 1.66) | <0.01 |
| Meta-analysis (w/o USOC) | 24,320 | 136,694 | 11.7 | 86.2 | 453 | 1,989 | 1.50 (1.30 - 1.73) | <0.01 | 1.41 (1.22 - 1.63) | <0.01 |
| **PS-stratification / on-treatment TAR without escalation** | | | | | | | | | | |
| CCAE | 11,233 | 49,023 | 6.3 | 34.5 | 230 | 801 | 1.32 (1.11 - 1.56) | <0.01 | 1.29 (1.08 - 1.53) | <0.01 |
| ODOD | 7,529 | 27,806 | 3.8 | 18.9 | 179 | 596 | 1.41 (1.15 - 1.74) | <0.01 | 1.33 (1.08 - 1.63) | 0.01 |
| MDCR | 870 | 8,802 | 0.5 | 7.8 | 9 | 207 | 0.90 (0.41 - 1.75) | 0.78 | 0.83 (0.40 - 1.71) | 0.62 |
| Open Claims | 137,078 | 390,524 | 82.6 | 290.3 | 1,764 | 4,408 | 1.45 (1.35 - 1.55) | <0.01 | 1.48 (1.37 - 1.60) | <0.01 |
| OEHR | 8,322 | 35,895 | 2.0 | 14.4 | 107 | 340 | 1.96 (1.52 - 2.51) | <0.01 | 1.65 (1.29 - 2.13) | <0.01 |
| VA | 1,316 | 23,930 | 0.7 | 18.3 | 19 | 158 | 2.42 (1.33 - 4.28) | <0.01 | 2.03 (1.13 - 3.64) | 0.02 |
| Meta-analysis | 164,162 | 503,248 | 94.8 | 358.1 | 2,280 | 6,145 | 1.48 (1.31 - 1.68) | <0.01 | 1.48 (1.26 - 1.72) | <0.01 |
| Meta-analysis (w/o USOC) | 27,084 | 112,724 | 12.2 | 67.9 | 516 | 1,737 | 1.52 (1.22 - 1.89) | <0.01 | 1.44 (1.16 - 1.79) | <0.01 |

##### 1.4.23 Peripheral edema

**Supplementary Table 101:** Exposures, outcomes and risk estimates for Peripheral edema in GLP1RA (male) (T) vs DPP4I (male) (C) new-user cohorts under four different propensity score (PS) adjustment / time-at-risk (TAR) designs across data sources. We report total patient exposure time in 1,000 patient-year (KYr) units, uncalibrated and calibrated hazard ratios (HRs) and their 95% confidence intervals (CIs). Greyed out entries did not pass study diagnostics and are included only for completeness.

| | Patients | | Exposure (KYr) | | Outcomes | | Uncalibrated | | Calibrated | |
|---|---|---|---|---|---|---|---|---|---|---|
| | T | C | T | C | T | C | HR (95% CI) | $p$ | HR (95% CI) | $p$ |
| **PS-matching / on-treatment TAR** | | | | | | | | | | |
| CCAE | 8,834 | 45,734 | 6.0 | 38.9 | 206 | 1,437 | 0.79 (0.64 - 0.96) | 0.02 | 0.76 (0.62 - 0.93) | 0.01 |
| ODOD | 5,129 | 24,462 | 3.1 | 19.8 | 155 | 1,155 | 0.94 (0.75 - 1.17) | 0.59 | 0.93 (0.74 - 1.16) | 0.55 |
| MDCR | 646 | 6,936 | 0.4 | 6.9 | 26 | 526 | 0.79 (0.46 - 1.28) | 0.36 | 0.76 (0.46 - 1.27) | 0.31 |
| Open Claims | 92,057 | 348,757 | 70.6 | 326.9 | 1,404 | 9,744 | 0.86 (0.80 - 0.93) | <0.01 | 0.87 (0.80 - 0.95) | 0.01 |
| OEHR | 6,629 | 33,499 | 1.9 | 16.1 | 73 | 624 | 0.95 (0.68 - 1.29) | 0.73 | 0.82 (0.60 - 1.14) | 0.29 |
| VA | 921 | 13,982 | 0.7 | 11.6 | 16 | 377 | 0.54 (0.26 - 1.00) | 0.07 | 0.44 (0.22 - 0.87) | 0.03 |
| Meta-analysis | 114,216 | 473,370 | 82.7 | 420.2 | 1,880 | 13,863 | 0.86 (0.80 - 0.91) | <0.01 | 0.85 (0.73 - 0.98) | 0.02 |
| Meta-analysis (w/o USOC) | 22,159 | 124,613 | 12.1 | 93.3 | 476 | 4,119 | 0.85 (0.75 - 0.97) | 0.01 | 0.82 (0.72 - 0.93) | <0.01 |
| **PS-stratification / on-treatment TAR** | | | | | | | | | | |
| CCAE | 10,344 | 45,839 | 6.6 | 39.0 | 229 | 1,443 | 0.86 (0.73 - 1.01) | 0.07 | 0.85 (0.73 - 1.00) | 0.07 |
| ODOD | 6,702 | 24,995 | 3.8 | 20.2 | 185 | 1,204 | 0.85 (0.71 - 1.02) | 0.08 | 0.82 (0.68 - 0.99) | 0.05 |
| MDCR | 730 | 7,379 | 0.5 | 7.3 | 28 | 597 | 0.87 (0.56 - 1.28) | 0.50 | 0.81 (0.53 - 1.22) | 0.32 |
| Open Claims | 122,153 | 350,794 | 83.8 | 328.6 | 1,666 | 9,964 | 0.82 (0.77 - 0.87) | <0.01 | 0.84 (0.79 - 0.91) | <0.01 |
| OEHR | 7,667 | 33,645 | 2.1 | 16.2 | 79 | 637 | 1.08 (0.84 - 1.39) | 0.53 | 0.95 (0.74 - 1.22) | 0.70 |
| VA | 1,029 | 20,139 | 0.7 | 18.4 | 16 | 620 | 0.57 (0.33 - 0.94) | 0.04 | 0.51 (0.30 - 0.86) | 0.01 |
| Meta-analysis | 146,866 | 455,273 | 96.3 | 404.0 | 2,159 | 13,248 | 0.85 (0.79 - 0.92) | <0.01 | 0.86 (0.74 - 1.00) | 0.06 |
| Meta-analysis (w/o USOC) | 24,713 | 104,479 | 12.5 | 75.4 | 493 | 3,284 | 0.90 (0.79 - 1.02) | 0.11 | 0.87 (0.75 - 1.00) | 0.08 |
| **PS-matching / on-treatment TAR without escalation** | | | | | | | | | | |
| CCAE | 8,834 | 45,734 | 5.2 | 31.7 | 172 | 1,155 | 0.83 (0.66 - 1.03) | 0.09 | 0.81 (0.65 - 1.01) | 0.08 |
| ODOD | 5,129 | 24,462 | 2.8 | 16.3 | 139 | 954 | 0.96 (0.76 - 1.22) | 0.75 | 0.92 (0.73 - 1.17) | 0.56 |
| MDCR | 646 | 6,936 | 0.4 | 6.0 | 22 | 440 | 0.73 (0.41 - 1.24) | 0.27 | 0.63 (0.36 - 1.10) | 0.12 |
| Open Claims | 92,057 | 348,757 | 61.0 | 254.5 | 1,203 | 7,568 | 0.87 (0.80 - 0.94) | <0.01 | 0.86 (0.79 - 0.94) | <0.01 |
| OEHR | 6,629 | 33,499 | 1.6 | 13.3 | 60 | 461 | 1.09 (0.76 - 1.54) | 0.63 | 0.94 (0.66 - 1.35) | 0.74 |
| VA | 921 | 13,982 | 0.5 | 9.5 | 12 | 303 | 0.75 (0.35 - 1.45) | 0.44 | 0.57 (0.28 - 1.18) | 0.14 |
| Meta-analysis | 114,216 | 473,370 | 71.6 | 331.3 | 1,608 | 10,881 | 0.88 (0.82 - 0.94) | <0.01 | 0.86 (0.77 - 0.95) | <0.01 |
| Meta-analysis (w/o USOC) | 22,159 | 124,613 | 10.5 | 76.8 | 405 | 3,313 | 0.90 (0.78 - 1.03) | 0.14 | 0.85 (0.73 - 0.98) | 0.03 |
| **PS-stratification / on-treatment TAR without escalation** | | | | | | | | | | |
| CCAE | 10,344 | 45,839 | 5.8 | 31.8 | 194 | 1,159 | 0.85 (0.71 - 1.00) | 0.06 | 0.82 (0.69 - 0.98) | 0.05 |
| ODOD | 6,702 | 24,995 | 3.4 | 16.7 | 168 | 994 | 0.88 (0.72 - 1.06) | 0.18 | 0.82 (0.68 - 1.00) | 0.07 |
| MDCR | 730 | 7,379 | 0.4 | 6.3 | 24 | 502 | 0.87 (0.54 - 1.34) | 0.56 | 0.80 (0.51 - 1.26) | 0.36 |
| Open Claims | 122,153 | 350,794 | 73.3 | 256.0 | 1,445 | 7,757 | 0.84 (0.78 - 0.89) | <0.01 | 0.86 (0.79 - 0.92) | <0.01 |
| OEHR | 7,667 | 33,645 | 1.9 | 13.3 | 65 | 467 | 1.10 (0.82 - 1.45) | 0.50 | 0.93 (0.70 - 1.24) | 0.65 |
| VA | 1,029 | 20,139 | 0.6 | 15.0 | 12 | 491 | 0.57 (0.30 - 1.00) | 0.07 | 0.48 (0.26 - 0.88) | 0.02 |
| Meta-analysis | 146,866 | 455,273 | 84.3 | 317.8 | 1,872 | 10,377 | 0.85 (0.80 - 0.90) | <0.01 | 0.85 (0.76 - 0.95) | <0.01 |
| Meta-analysis (w/o USOC) | 24,713 | 104,479 | 11.0 | 61.9 | 427 | 2,620 | 0.90 (0.79 - 1.03) | 0.13 | 0.86 (0.75 - 0.98) | 0.04 |

##### 1.4.24 Photosensitivity

**Supplementary Table 102:** Exposures, outcomes and risk estimates for Photosensitivity in GLP1RA (male) (T) vs DPP4I (male) (C) new-user cohorts under four different propensity score (PS) adjustment / time-at-risk (TAR) designs across data sources. We report total patient exposure time in 1,000 patient-year (KYr) units, uncalibrated and calibrated hazard ratios (HRs) and their 95% confidence intervals (CIs). Greyed out entries did not pass study diagnostics and are included only for completeness.

| | Patients | | Exposure (KYr) | | Outcomes | | Uncalibrated | | Calibrated | |
|---|---|---|---|---|---|---|---|---|---|---|
| | T | C | T | C | T | C | HR (95% CI) | $p$ | HR (95% CI) | $p$ |
| **PS-matching / on-treatment TAR** | | | | | | | | | | |
| CCAE | 10,525 | 52,283 | 7.3 | 45.9 | 9 | 64 | 1.39 (0.52 - 3.30) | 0.49 | 1.34 (0.53 - 3.39) | 0.54 |
| ODOD | 6,375 | 29,904 | 4.0 | 25.2 | <5 | 47 | 0.52 (0.08 - 1.93) | 0.42 | 0.51 (0.10 - 2.52) | 0.41 |
| MDCR | 836 | 8,723 | 0.6 | 9.1 | <5 | 43 | 0.83 (0.05 - 4.09) | 0.87 | 0.80 (0.08 - 7.56) | 0.85 |
| Open Claims | 114,514 | 423,044 | 88.4 | 406.0 | 27 | 220 | 1.39 (0.80 - 2.33) | 0.23 | 1.40 (0.82 - 2.41) | 0.22 |
| OEHR | 7,758 | 37,977 | 2.3 | 19.0 | 5 | 23 | 3.48 (0.78 - 12.9) | 0.08 | 3.03 (0.74 - 12.4) | 0.12 |
| VA | 1,195 | 17,491 | 0.9 | 14.9 | 0 | <10 | | | | |
| Meta-analysis | 131,414 | 505,231 | 99.6 | 477.1 | <41 | 331 | 1.29 (0.82 - 2.01) | 0.27 | 1.27 (0.80 - 2.02) | 0.31 |
| Meta-analysis (w/o USOC) | 16,900 | 82,187 | 11.3 | 71.1 | <14 | 111 | 1.06 (0.45 - 2.49) | 0.89 | 1.02 (0.44 - 2.39) | 0.96 |
| **PS-stratification / on-treatment TAR** | | | | | | | | | | |
| CCAE | 12,437 | 52,413 | 8.1 | 46.0 | 9 | 65 | 1.37 (0.60 - 2.79) | 0.41 | 1.36 (0.63 - 2.92) | 0.44 |
| ODOD | 8,466 | 30,499 | 4.8 | 25.7 | <5 | 50 | 0.37 (0.10 - 1.10) | 0.11 | 0.36 (0.11 - 1.19) | 0.10 |
| MDCR | 936 | 9,340 | 0.6 | 9.7 | <5 | 47 | 0.54 (0.03 - 2.60) | 0.59 | 0.50 (0.05 - 4.68) | 0.55 |
| Open Claims | 154,117 | 425,286 | 106.0 | 408.1 | 32 | 221 | 1.19 (0.75 - 1.81) | 0.44 | 1.23 (0.79 - 1.90) | 0.37 |
| OEHR | 9,044 | 38,070 | 2.6 | 19.1 | 5 | 23 | 2.64 (0.77 - 7.53) | 0.10 | 2.31 (0.74 - 7.24) | 0.15 |
| VA | 1,389 | 25,264 | 1.0 | 23.7 | 0 | <10 | | | | |
| Meta-analysis | 184,064 | 546,268 | 121.5 | 498.8 | <51 | 359 | 1.20 (0.85 - 1.69) | 0.31 | 1.21 (0.83 - 1.75) | 0.32 |
| Meta-analysis (w/o USOC) | 29,947 | 120,982 | 15.5 | 90.8 | <19 | 138 | 1.14 (0.42 - 3.09) | 0.79 | 1.10 (0.41 - 2.97) | 0.85 |
| **PS-matching / on-treatment TAR without escalation** | | | | | | | | | | |
| CCAE | 10,525 | 52,283 | 6.3 | 37.1 | 8 | 54 | 1.34 (0.47 - 3.31) | 0.56 | 1.31 (0.49 - 3.49) | 0.58 |
| ODOD | 6,375 | 29,904 | 3.5 | 20.6 | <5 | 40 | 0.80 (0.12 - 3.04) | 0.78 | 0.76 (0.15 - 3.82) | 0.74 |
| MDCR | 836 | 8,723 | 0.5 | 7.8 | <5 | 36 | 0.97 (0.05 - 5.04) | 0.98 | 0.84 (0.09 - 8.20) | 0.88 |
| Open Claims | 114,514 | 423,044 | 76.2 | 314.5 | 25 | 186 | 1.31 (0.71 - 2.28) | 0.37 | 1.30 (0.73 - 2.32) | 0.38 |
| OEHR | 7,758 | 37,977 | 2.0 | 15.4 | <5 | 20 | 1.44 (0.18 - 7.23) | 0.70 | 1.24 (0.20 - 7.82) | 0.81 |
| VA | 1,195 | 17,491 | 0.7 | 12.1 | 0 | <10 | | | | |
| Meta-analysis | 131,414 | 505,231 | 86.0 | 372.2 | <38 | 280 | 1.26 (0.78 - 2.03) | 0.34 | 1.23 (0.76 - 1.99) | 0.40 |
| Meta-analysis (w/o USOC) | 16,900 | 82,187 | 9.8 | 57.7 | <13 | 94 | 1.17 (0.51 - 2.69) | 0.72 | 1.10 (0.47 - 2.52) | 0.83 |
| **PS-stratification / on-treatment TAR without escalation** | | | | | | | | | | |
| CCAE | 12,437 | 52,413 | 7.1 | 37.2 | 8 | 54 | 1.40 (0.58 - 3.00) | 0.42 | 1.36 (0.60 - 3.11) | 0.46 |
| ODOD | 8,466 | 30,499 | 4.3 | 21.0 | <5 | 42 | 0.44 (0.11 - 1.38) | 0.20 | 0.42 (0.12 - 1.45) | 0.17 |
| MDCR | 936 | 9,340 | 0.5 | 8.3 | <5 | 40 | 0.74 (0.04 - 3.64) | 0.79 | 0.68 (0.07 - 6.40) | 0.74 |
| Open Claims | 154,117 | 425,286 | 92.6 | 316.1 | 30 | 187 | 1.17 (0.72 - 1.84) | 0.50 | 1.20 (0.75 - 1.91) | 0.45 |
| OEHR | 9,044 | 38,070 | 2.2 | 15.5 | <5 | 20 | 1.69 (0.33 - 6.14) | 0.48 | 1.43 (0.33 - 6.15) | 0.63 |
| VA | 1,389 | 25,264 | 0.8 | 19.2 | 0 | <10 | | | | |
| Meta-analysis | 175,020 | 508,198 | 103.9 | 374.3 | <43 | 283 | 1.11 (0.76 - 1.63) | 0.59 | 1.11 (0.75 - 1.65) | 0.61 |
| Meta-analysis (w/o USOC) | 20,903 | 82,912 | 11.4 | 58.2 | <13 | 96 | 0.87 (0.29 - 2.64) | 0.81 | 0.83 (0.27 - 2.50) | 0.74 |

##### 1.4.25 Venous thromboembolism

**Supplementary Table 103:** Exposures, outcomes and risk estimates for Venous thromboembolism in GLP1RA (male) (T) vs DPP4I (male) (C) new-user cohorts under four different propensity score (PS) adjustment / time-at-risk (TAR) designs across data sources. We report total patient exposure time in 1,000 patient-year (KYr) units, uncalibrated and calibrated hazard ratios (HRs) and their 95% confidence intervals (CIs). Greyed out entries did not pass study diagnostics and are included only for completeness.

| | Patients | | Exposure (KYr) | | Outcomes | | Uncalibrated | | Calibrated | |
|---|---|---|---|---|---|---|---|---|---|---|
| | T | C | T | C | T | C | HR (95% CI) | $p$ | HR (95% CI) | $p$ |
| **PS-matching / on-treatment TAR** | | | | | | | | | | |
| CCAE | 10,331 | 51,708 | 7.1 | 45.3 | 34 | 203 | 0.85 (0.50 - 1.39) | 0.54 | 0.82 (0.49 - 1.38) | 0.46 |
| ODOD | 6,230 | 29,301 | 3.9 | 24.7 | 24 | 214 | 0.65 (0.37 - 1.10) | 0.12 | 0.64 (0.37 - 1.11) | 0.12 |
| MDCR | 814 | 8,491 | 0.6 | 8.9 | 8 | 110 | 1.13 (0.40 - 2.71) | 0.81 | 1.09 (0.42 - 2.83) | 0.85 |
| Open Claims | 111,380 | 413,513 | 86.1 | 396.4 | 317 | 1,916 | 0.92 (0.78 - 1.07) | 0.28 | 0.92 (0.78 - 1.09) | 0.40 |
| OEHR | 7,598 | 37,256 | 2.3 | 18.5 | 19 | 122 | 1.16 (0.58 - 2.18) | 0.67 | 1.01 (0.52 - 1.96) | 0.91 |
| VA | 1,162 | 17,157 | 0.9 | 14.5 | <10 | 64 | 0.94 (0.29 - 2.48) | 0.91 | 0.76 (0.26 - 2.22) | 0.64 |
| Meta-analysis | 137,515 | 557,426 | 100.7 | 508.4 | <412 | 2,629 | 0.90 (0.78 - 1.04) | 0.16 | 0.89 (0.74 - 1.08) | 0.25 |
| Meta-analysis (w/o USOC) | 26,135 | 143,913 | 14.7 | 112.0 | <95 | 713 | 0.87 (0.64 - 1.16) | 0.34 | 0.83 (0.62 - 1.12) | 0.23 |
| **PS-stratification / on-treatment TAR** | | | | | | | | | | |
| CCAE | 12,188 | 51,836 | 7.9 | 45.4 | 42 | 204 | 1.04 (0.70 - 1.50) | 0.84 | 1.03 (0.70 - 1.50) | 0.89 |
| ODOD | 8,239 | 29,963 | 4.7 | 25.2 | 27 | 223 | 0.63 (0.38 - 0.98) | 0.05 | 0.60 (0.38 - 0.97) | 0.04 |
| MDCR | 910 | 9,176 | 0.6 | 9.6 | 9 | 128 | 1.48 (0.66 - 2.93) | 0.30 | 1.38 (0.65 - 2.89) | 0.40 |
| Open Claims | 149,623 | 415,788 | 103.1 | 398.5 | 370 | 1,959 | 0.88 (0.77 - 1.00) | 0.05 | 0.91 (0.80 - 1.04) | 0.23 |
| OEHR | 8,858 | 37,344 | 2.5 | 18.6 | 19 | 124 | 1.16 (0.66 - 1.92) | 0.58 | 1.01 (0.60 - 1.73) | 0.91 |
| VA | 1,344 | 24,743 | 0.9 | 23.1 | <10 | 103 | 1.11 (0.44 - 2.43) | 0.82 | 0.97 (0.41 - 2.30) | 0.92 |
| Meta-analysis | 178,908 | 534,931 | 118.2 | 487.8 | 458 | 2,510 | 0.89 (0.79 - 0.99) | 0.04 | 0.90 (0.75 - 1.07) | 0.22 |
| Meta-analysis (w/o USOC) | 29,285 | 119,143 | 15.2 | 89.3 | 88 | 551 | 0.91 (0.64 - 1.30) | 0.61 | 0.87 (0.61 - 1.26) | 0.48 |
| **PS-matching / on-treatment TAR without escalation** | | | | | | | | | | |
| CCAE | 10,331 | 51,708 | 6.2 | 36.6 | 31 | 167 | 0.98 (0.56 - 1.63) | 0.93 | 0.96 (0.56 - 1.63) | 0.87 |
| ODOD | 6,230 | 29,301 | 3.4 | 20.1 | 21 | 174 | 0.78 (0.42 - 1.36) | 0.40 | 0.74 (0.41 - 1.34) | 0.34 |
| MDCR | 814 | 8,491 | 0.5 | 7.6 | <5 | 90 | 0.55 (0.12 - 1.74) | 0.38 | 0.48 (0.13 - 1.79) | 0.27 |
| Open Claims | 111,380 | 413,513 | 74.2 | 307.4 | 275 | 1,487 | 1.00 (0.83 - 1.19) | 0.99 | 0.99 (0.83 - 1.19) | 0.90 |
| OEHR | 7,598 | 37,256 | 1.9 | 15.1 | 15 | 94 | 1.27 (0.58 - 2.58) | 0.53 | 1.10 (0.52 - 2.31) | 0.81 |
| VA | 1,162 | 17,157 | 0.7 | 11.9 | <10 | 50 | 1.05 (0.28 - 3.06) | 0.94 | 0.80 (0.24 - 2.63) | 0.71 |
| Meta-analysis | 136,353 | 540,269 | 86.3 | 386.8 | <347 | 2,012 | 0.98 (0.84 - 1.15) | 0.81 | 0.96 (0.80 - 1.14) | 0.63 |
| Meta-analysis (w/o USOC) | 24,973 | 126,756 | 12.0 | 79.4 | <72 | 525 | 0.92 (0.66 - 1.29) | 0.63 | 0.86 (0.62 - 1.21) | 0.41 |
| **PS-stratification / on-treatment TAR without escalation** | | | | | | | | | | |
| CCAE | 12,188 | 51,836 | 6.9 | 36.7 | 39 | 168 | 1.05 (0.69 - 1.55) | 0.82 | 1.02 (0.68 - 1.52) | 0.91 |
| ODOD | 8,239 | 29,963 | 4.2 | 20.6 | 24 | 183 | 0.62 (0.37 - 1.01) | 0.07 | 0.58 (0.35 - 0.97) | 0.04 |
| MDCR | 910 | 9,176 | 0.5 | 8.2 | 5 | 105 | 0.99 (0.33 - 2.38) | 0.98 | 0.90 (0.33 - 2.43) | 0.84 |
| Open Claims | 149,623 | 415,788 | 90.0 | 309.1 | 324 | 1,522 | 0.92 (0.80 - 1.06) | 0.25 | 0.94 (0.81 - 1.09) | 0.44 |
| OEHR | 8,858 | 37,344 | 2.2 | 15.1 | 15 | 96 | 1.10 (0.58 - 1.94) | 0.77 | 0.93 (0.51 - 1.69) | 0.81 |
| VA | 1,344 | 24,743 | 0.7 | 18.8 | <10 | 74 | 1.28 (0.50 - 2.92) | 0.58 | 1.07 (0.44 - 2.61) | 0.87 |
| Meta-analysis | 178,908 | 534,931 | 103.4 | 381.6 | 402 | 1,969 | 0.92 (0.81 - 1.04) | 0.18 | 0.91 (0.78 - 1.07) | 0.26 |
| Meta-analysis (w/o USOC) | 29,285 | 119,143 | 13.3 | 72.5 | 78 | 447 | 0.90 (0.63 - 1.27) | 0.54 | 0.85 (0.60 - 1.20) | 0.37 |

##### 1.4.26 Vomiting

**Supplementary Table 104:** Exposures, outcomes and risk estimates for Vomiting in GLP1RA (male) (T) vs DPP4I (male) (C) new-user cohorts under four different propensity score (PS) adjustment / time-at-risk (TAR) designs across data sources. We report total patient exposure time in 1,000 patient-year (KYr) units, uncalibrated and calibrated hazard ratios (HRs) and their 95% confidence intervals (CIs). Greyed out entries did not pass study diagnostics and are included only for completeness.

| | Patients | | Exposure (KYr) | | Outcomes | | Uncalibrated | | Calibrated | |
|---|---|---|---|---|---|---|---|---|---|---|
| | T | C | T | C | T | C | HR (95% CI) | $p$ | HR (95% CI) | $p$ |
| **PS-matching / on-treatment TAR** | | | | | | | | | | |
| CCAE | 9,911 | 49,940 | 6.8 | 43.5 | 153 | 713 | 1.51 (1.19 - 1.90) | <0.01 | 1.46 (1.15 - 1.85) | <0.01 |
| ODOD | 5,885 | 28,044 | 3.6 | 23.4 | 97 | 533 | 1.24 (0.92 - 1.66) | 0.15 | 1.22 (0.91 - 1.65) | 0.22 |
| MDCR | 808 | 8,326 | 0.6 | 8.7 | 11 | 175 | 1.05 (0.48 - 2.10) | 0.89 | 1.02 (0.49 - 2.13) | 0.88 |
| Open Claims | 105,894 | 396,335 | 81.9 | 379.5 | 1,196 | 4,527 | 1.47 (1.35 - 1.61) | <0.01 | 1.49 (1.35 - 1.64) | <0.01 |
| OEHR | 7,405 | 36,590 | 2.2 | 18.1 | 79 | 290 | 2.15 (1.51 - 3.05) | <0.01 | 1.87 (1.31 - 2.67) | <0.01 |
| VA | 1,157 | 16,781 | 0.8 | 14.3 | 14 | 94 | 1.76 (0.74 - 3.90) | 0.18 | 1.43 (0.62 - 3.30) | 0.41 |
| Meta-analysis | 131,060 | 536,016 | 95.9 | 487.4 | 1,550 | 6,332 | 1.48 (1.37 - 1.60) | <0.01 | 1.46 (1.26 - 1.71) | <0.01 |
| Meta-analysis (w/o USOC) | 25,166 | 139,681 | 14.0 | 108.0 | 354 | 1,805 | 1.52 (1.22 - 1.91) | <0.01 | 1.46 (1.17 - 1.83) | <0.01 |
| **PS-stratification / on-treatment TAR** | | | | | | | | | | |
| CCAE | 11,678 | 50,059 | 7.6 | 43.6 | 176 | 717 | 1.25 (1.03 - 1.51) | 0.02 | 1.23 (1.02 - 1.49) | 0.04 |
| ODOD | 7,839 | 28,613 | 4.4 | 23.8 | 125 | 555 | 1.30 (1.02 - 1.64) | 0.03 | 1.25 (0.99 - 1.59) | 0.07 |
| MDCR | 905 | 8,966 | 0.6 | 9.3 | 11 | 202 | 0.99 (0.48 - 1.84) | 0.98 | 0.92 (0.47 - 1.80) | 0.81 |
| Open Claims | 141,956 | 398,725 | 97.9 | 381.6 | 1,492 | 4,622 | 1.37 (1.27 - 1.47) | <0.01 | 1.41 (1.31 - 1.53) | <0.01 |
| OEHR | 8,595 | 36,696 | 2.4 | 18.1 | 90 | 295 | 2.28 (1.74 - 2.97) | <0.01 | 1.99 (1.52 - 2.61) | <0.01 |
| VA | 1,348 | 24,357 | 0.9 | 22.9 | 14 | 156 | 1.71 (0.87 - 3.19) | 0.10 | 1.51 (0.79 - 2.89) | 0.22 |
| Meta-analysis | 170,068 | 514,093 | 112.3 | 467.2 | 1,883 | 6,189 | 1.48 (1.15 - 1.91) | <0.01 | 1.49 (1.12 - 1.99) | <0.01 |
| Meta-analysis (w/o USOC) | 28,112 | 115,368 | 14.4 | 85.6 | 391 | 1,567 | 1.53 (1.08 - 2.18) | 0.02 | 1.47 (1.03 - 2.10) | 0.04 |
| **PS-matching / on-treatment TAR without escalation** | | | | | | | | | | |
| CCAE | 9,911 | 49,940 | 5.9 | 35.3 | 138 | 593 | 1.52 (1.18 - 1.95) | <0.01 | 1.49 (1.16 - 1.92) | <0.01 |
| ODOD | 5,885 | 28,044 | 3.2 | 19.2 | 91 | 447 | 1.31 (0.95 - 1.79) | 0.10 | 1.26 (0.91 - 1.73) | 0.19 |
| MDCR | 808 | 8,326 | 0.5 | 7.4 | 8 | 147 | 0.89 (0.35 - 1.98) | 0.80 | 0.77 (0.32 - 1.84) | 0.56 |
| Open Claims | 105,894 | 396,335 | 70.8 | 295.0 | 1,016 | 3,420 | 1.52 (1.38 - 1.67) | <0.01 | 1.51 (1.36 - 1.67) | <0.01 |
| OEHR | 7,405 | 36,590 | 1.9 | 14.8 | 63 | 195 | 2.02 (1.35 - 2.99) | <0.01 | 1.75 (1.17 - 2.60) | <0.01 |
| VA | 1,157 | 16,781 | 0.7 | 11.7 | 13 | 74 | 2.25 (0.86 - 5.50) | 0.09 | 1.71 (0.68 - 4.34) | 0.26 |
| Meta-analysis | 131,060 | 536,016 | 82.9 | 383.3 | 1,329 | 4,876 | 1.52 (1.40 - 1.65) | <0.01 | 1.48 (1.33 - 1.66) | <0.01 |
| Meta-analysis (w/o USOC) | 25,166 | 139,681 | 12.1 | 88.3 | 313 | 1,456 | 1.53 (1.25 - 1.88) | <0.01 | 1.44 (1.17 - 1.77) | <0.01 |
| **PS-stratification / on-treatment TAR without escalation** | | | | | | | | | | |
| CCAE | 11,678 | 50,059 | 6.6 | 35.4 | 159 | 593 | 1.27 (1.04 - 1.55) | 0.02 | 1.24 (1.01 - 1.52) | 0.06 |
| ODOD | 7,839 | 28,613 | 3.9 | 19.6 | 114 | 466 | 1.35 (1.05 - 1.72) | 0.02 | 1.26 (0.98 - 1.62) | 0.09 |
| MDCR | 905 | 8,966 | 0.5 | 7.9 | 8 | 172 | 0.91 (0.39 - 1.85) | 0.81 | 0.83 (0.38 - 1.82) | 0.65 |
| Open Claims | 141,956 | 398,725 | 85.7 | 296.7 | 1,293 | 3,494 | 1.44 (1.34 - 1.56) | <0.01 | 1.47 (1.35 - 1.61) | <0.01 |
| OEHR | 8,595 | 36,696 | 2.1 | 14.8 | 74 | 198 | 2.52 (1.84 - 3.42) | <0.01 | 2.13 (1.56 - 2.90) | <0.01 |
| VA | 1,348 | 24,357 | 0.7 | 18.6 | 13 | 128 | 2.20 (1.06 - 4.33) | 0.03 | 1.84 (0.91 - 3.73) | 0.09 |
| Meta-analysis | 170,068 | 514,093 | 98.3 | 366.4 | 1,640 | 4,751 | 1.55 (1.17 - 2.05) | <0.01 | 1.55 (1.15 - 2.08) | <0.01 |
| Meta-analysis (w/o USOC) | 28,112 | 115,368 | 12.7 | 69.7 | 347 | 1,257 | 1.60 (1.09 - 2.36) | 0.02 | 1.52 (1.03 - 2.24) | 0.04 |

#### 1.5 GLP1RA (male) vs SU (male)

##### 1.5.1 3-point MACE

**Supplementary Table 105:** Exposures, outcomes and risk estimates for 3-point MACE in GLP1RA (male) (T) vs SU (male) (C) new-user cohorts under four different propensity score (PS) adjustment / time-at-risk (TAR) designs across data sources. We report total patient exposure time in 1,000 patient-year (KYr) units, uncalibrated and calibrated hazard ratios (HRs) and their 95% confidence intervals (CIs). Greyed out entries did not pass study diagnostics and are included only for completeness.

| | Patients | | Exposure (KYr) | | Outcomes | | Uncalibrated | | Calibrated | |
|---|---|---|---|---|---|---|---|---|---|---|
| | T | C | T | C | T | C | HR (95% CI) | $p$ | HR (95% CI) | $p$ |
| **PS-matching / on-treatment TAR** | | | | | | | | | | |
| CCAE | 11,015 | 82,939 | 7.4 | 71.1 | 55 | 722 | 0.79 (0.55 - 1.11) | 0.19 | 0.77 (0.54 - 1.09) | 0.15 |
| ODOD | 7,287 | 59,676 | 4.3 | 53.9 | 39 | 1,228 | 0.52 (0.35 - 0.76) | <0.01 | 0.50 (0.34 - 0.74) | <0.01 |
| MDCR | 871 | 14,448 | 0.6 | 15.5 | 11 | 458 | 0.57 (0.23 - 1.19) | 0.18 | 0.56 (0.25 - 1.26) | 0.16 |
| Open Claims | 132,622 | 817,979 | 94.6 | 881.7 | 604 | 11,673 | 0.66 (0.60 - 0.73) | <0.01 | 0.67 (0.57 - 0.80) | <0.01 |
| OEHR | 8,180 | 84,320 | 2.4 | 46.0 | 25 | 600 | 0.95 (0.58 - 1.50) | 0.85 | 0.89 (0.56 - 1.43) | 0.64 |
| VA | 1,240 | 31,321 | 0.9 | 23.6 | <10 | 152 | 0.55 (0.13 - 1.65) | 0.36 | 0.43 (0.12 - 1.55) | 0.20 |
| Meta-analysis | 159,975 | 1,059,362 | 109.3 | 1068.1 | 734 | 14,681 | 0.67 (0.61 - 0.73) | <0.01 | 0.66 (0.55 - 0.80) | <0.01 |
| Meta-analysis (w/o USOC) | 27,353 | 241,383 | 14.6 | 186.4 | 130 | 3,008 | 0.71 (0.53 - 0.93) | 0.01 | 0.68 (0.51 - 0.90) | <0.01 |
| **PS-stratification / on-treatment TAR** | | | | | | | | | | |
| CCAE | 12,495 | 87,190 | 8.1 | 74.5 | 62 | 764 | 0.76 (0.57 - 1.01) | 0.06 | 0.74 (0.55 - 0.99) | 0.05 |
| ODOD | 8,175 | 66,607 | 4.7 | 61.8 | 42 | 1,568 | 0.49 (0.35 - 0.67) | <0.01 | 0.47 (0.34 - 0.65) | <0.01 |
| MDCR | 945 | 18,402 | 0.6 | 19.5 | 11 | 654 | 0.59 (0.30 - 1.05) | 0.10 | 0.55 (0.29 - 1.03) | 0.06 |
| Open Claims | 147,591 | 843,396 | 101.8 | 911.1 | 647 | 12,750 | 0.69 (0.63 - 0.75) | <0.01 | 0.70 (0.57 - 0.85) | <0.01 |
| OEHR | 8,896 | 93,416 | 2.5 | 51.3 | 26 | 750 | 0.93 (0.60 - 1.39) | 0.74 | 0.86 (0.57 - 1.31) | 0.51 |
| VA | 1,335 | 230,206 | 0.9 | 244.4 | <10 | 2,056 | 0.80 (0.28 - 1.76) | 0.64 | 0.68 (0.27 - 1.69) | 0.41 |
| Meta-analysis | 177,157 | 1,090,609 | 117.1 | 1098.7 | 777 | 15,832 | 0.69 (0.56 - 0.84) | <0.01 | 0.68 (0.52 - 0.90) | <0.01 |
| Meta-analysis (w/o USOC) | 29,566 | 247,213 | 15.3 | 187.6 | 130 | 3,082 | 0.70 (0.49 - 0.99) | 0.05 | 0.66 (0.46 - 0.96) | 0.04 |
| **PS-matching / on-treatment TAR without escalation** | | | | | | | | | | |
| CCAE | 11,015 | 82,939 | 6.4 | 61.7 | 49 | 629 | 0.76 (0.52 - 1.09) | 0.14 | 0.74 (0.51 - 1.07) | 0.12 |
| ODOD | 7,287 | 59,676 | 3.8 | 47.9 | 32 | 1,095 | 0.45 (0.28 - 0.68) | <0.01 | 0.42 (0.27 - 0.66) | <0.01 |
| MDCR | 871 | 14,448 | 0.5 | 14.1 | 7 | 406 | 0.73 (0.27 - 1.61) | 0.48 | 0.72 (0.29 - 1.75) | 0.46 |
| Open Claims | 132,622 | 817,979 | 82.3 | 747.0 | 497 | 9,969 | 0.62 (0.56 - 0.70) | <0.01 | 0.62 (0.52 - 0.75) | <0.01 |
| OEHR | 8,180 | 84,320 | 2.0 | 40.1 | 21 | 442 | 1.05 (0.61 - 1.71) | 0.87 | 0.98 (0.58 - 1.65) | 0.88 |
| VA | 1,240 | 31,321 | 0.7 | 20.7 | <10 | 136 | 0.42 (0.06 - 1.48) | 0.27 | 0.31 (0.07 - 1.49) | 0.15 |
| Meta-analysis | 159,975 | 1,059,362 | 95.1 | 910.8 | 606 | 12,541 | 0.67 (0.53 - 0.84) | <0.01 | 0.65 (0.49 - 0.87) | <0.01 |
| Meta-analysis (w/o USOC) | 27,353 | 241,383 | 12.8 | 163.8 | 109 | 2,572 | 0.70 (0.48 - 1.02) | 0.06 | 0.67 (0.46 - 0.98) | 0.05 |
| **PS-stratification / on-treatment TAR without escalation** | | | | | | | | | | |
| CCAE | 12,495 | 87,190 | 7.1 | 64.8 | 55 | 670 | 0.78 (0.57 - 1.04) | 0.10 | 0.76 (0.56 - 1.03) | 0.08 |
| ODOD | 8,175 | 66,607 | 4.2 | 55.2 | 35 | 1,402 | 0.45 (0.31 - 0.63) | <0.01 | 0.42 (0.29 - 0.60) | <0.01 |
| MDCR | 945 | 18,402 | 0.6 | 17.8 | 7 | 584 | 0.44 (0.18 - 0.89) | 0.04 | 0.40 (0.18 - 0.88) | 0.02 |
| Open Claims | 147,591 | 843,396 | 89.0 | 772.2 | 538 | 10,929 | 0.66 (0.60 - 0.72) | <0.01 | 0.66 (0.54 - 0.81) | <0.01 |
| OEHR | 8,896 | 93,416 | 2.2 | 44.7 | 22 | 548 | 1.05 (0.65 - 1.63) | 0.83 | 0.95 (0.60 - 1.51) | 0.84 |
| VA | 1,335 | 230,206 | 0.7 | 217.9 | <10 | 1,754 | 1.03 (0.36 - 2.28) | 0.95 | 0.82 (0.33 - 2.06) | 0.68 |
| Meta-analysis | 177,157 | 1,090,609 | 102.5 | 936.9 | 650 | 13,549 | 0.69 (0.51 - 0.92) | 0.01 | 0.67 (0.47 - 0.96) | 0.03 |
| Meta-analysis (w/o USOC) | 29,566 | 247,213 | 13.5 | 164.7 | 112 | 2,620 | 0.71 (0.45 - 1.12) | 0.14 | 0.67 (0.41 - 1.07) | 0.10 |

##### 1.5.2 4-point MACE

**Supplementary Table 106:** Exposures, outcomes and risk estimates for 4-point MACE in GLP1RA (male) (T) vs SU (male) (C) new-user cohorts under four different propensity score (PS) adjustment / time-at-risk (TAR) designs across data sources. We report total patient exposure time in 1,000 patient-year (KYr) units, uncalibrated and calibrated hazard ratios (HRs) and their 95% confidence intervals (CIs). Greyed out entries did not pass study diagnostics and are included only for completeness.

| | Patients | | Exposure (KYr) | | Outcomes | | Uncalibrated | | Calibrated | |
|---|---|---|---|---|---|---|---|---|---|---|
| | T | C | T | C | T | C | HR (95% CI) | $p$ | HR (95% CI) | $p$ |
| **PS-matching / on-treatment TAR** | | | | | | | | | | |
| CCAE | 10,829 | 81,745 | 7.3 | 69.8 | 85 | 1,071 | 0.78 (0.58 - 1.04) | 0.10 | 0.76 (0.57 - 1.02) | 0.07 |
| ODOD | 7,033 | 57,505 | 4.1 | 51.3 | 71 | 1,953 | 0.55 (0.40 - 0.73) | <0.01 | 0.52 (0.39 - 0.70) | <0.01 |
| MDCR | 823 | 13,580 | 0.6 | 14.4 | 21 | 742 | 0.82 (0.46 - 1.37) | 0.49 | 0.81 (0.47 - 1.39) | 0.45 |
| Open Claims | 128,962 | 790,564 | 92.1 | 847.8 | 921 | 18,264 | 0.67 (0.61 - 0.72) | <0.01 | 0.68 (0.58 - 0.80) | <0.01 |
| OEHR | 8,057 | 82,802 | 2.3 | 44.5 | 31 | 1,099 | 0.64 (0.42 - 0.95) | 0.03 | 0.60 (0.40 - 0.90) | 0.02 |
| VA | 1,184 | 30,622 | 0.9 | 22.9 | <10 | 364 | 0.37 (0.11 - 0.91) | 0.06 | 0.29 (0.10 - 0.82) | 0.02 |
| Meta-analysis | 155,704 | 1,026,196 | 106.4 | 1027.9 | 1,129 | 23,129 | 0.67 (0.62 - 0.72) | <0.01 | 0.66 (0.55 - 0.79) | <0.01 |
| Meta-analysis (w/o USOC) | 26,742 | 235,632 | 14.3 | 180.1 | 208 | 4,865 | 0.67 (0.55 - 0.82) | <0.01 | 0.64 (0.53 - 0.78) | <0.01 |
| **PS-stratification / on-treatment TAR** | | | | | | | | | | |
| CCAE | 12,270 | 85,777 | 8.0 | 73.1 | 98 | 1,132 | 0.77 (0.61 - 0.96) | 0.02 | 0.74 (0.59 - 0.94) | 0.02 |
| ODOD | 7,889 | 63,755 | 4.5 | 58.4 | 77 | 2,460 | 0.52 (0.41 - 0.66) | <0.01 | 0.50 (0.39 - 0.64) | <0.01 |
| MDCR | 895 | 17,003 | 0.6 | 17.9 | 23 | 1,035 | 0.79 (0.49 - 1.19) | 0.29 | 0.73 (0.47 - 1.14) | 0.17 |
| Open Claims | 143,665 | 813,841 | 99.2 | 875.2 | 977 | 19,937 | 0.65 (0.61 - 0.70) | <0.01 | 0.66 (0.55 - 0.80) | <0.01 |
| OEHR | 8,765 | 91,617 | 2.5 | 49.6 | 33 | 1,350 | 0.65 (0.45 - 0.92) | 0.02 | 0.60 (0.42 - 0.87) | <0.01 |
| VA | 1,282 | 223,743 | 0.9 | 235.5 | <10 | 4,077 | 0.35 (0.12 - 0.75) | 0.02 | 0.29 (0.12 - 0.73) | <0.01 |
| Meta-analysis | 172,589 | 1,054,990 | 114.2 | 1056.3 | 1,185 | 24,879 | 0.65 (0.57 - 0.74) | <0.01 | 0.64 (0.51 - 0.81) | <0.01 |
| Meta-analysis (w/o USOC) | 28,924 | 241,149 | 15.0 | 181.1 | 208 | 4,942 | 0.64 (0.50 - 0.82) | <0.01 | 0.61 (0.47 - 0.80) | <0.01 |
| **PS-matching / on-treatment TAR without escalation** | | | | | | | | | | |
| CCAE | 10,829 | 81,745 | 6.3 | 60.6 | 77 | 925 | 0.82 (0.60 - 1.10) | 0.20 | 0.80 (0.58 - 1.08) | 0.16 |
| ODOD | 7,033 | 57,505 | 3.7 | 45.8 | 61 | 1,725 | 0.56 (0.40 - 0.76) | <0.01 | 0.53 (0.38 - 0.73) | <0.01 |
| MDCR | 823 | 13,580 | 0.5 | 13.1 | 16 | 663 | 0.80 (0.42 - 1.40) | 0.47 | 0.79 (0.43 - 1.44) | 0.45 |
| Open Claims | 128,962 | 790,564 | 80.2 | 719.1 | 778 | 15,559 | 0.66 (0.61 - 0.73) | <0.01 | 0.67 (0.56 - 0.78) | <0.01 |
| OEHR | 8,057 | 82,802 | 2.0 | 39.1 | 26 | 848 | 0.70 (0.43 - 1.07) | 0.12 | 0.65 (0.41 - 1.03) | 0.07 |
| VA | 1,184 | 30,622 | 0.7 | 20.1 | <10 | 298 | 0.43 (0.13 - 1.09) | 0.12 | 0.32 (0.11 - 0.94) | 0.04 |
| Meta-analysis | 155,704 | 1,026,196 | 92.7 | 877.6 | 958 | 19,720 | 0.67 (0.62 - 0.73) | <0.01 | 0.66 (0.55 - 0.79) | <0.01 |
| Meta-analysis (w/o USOC) | 26,742 | 235,632 | 12.5 | 158.6 | 180 | 4,161 | 0.70 (0.57 - 0.85) | <0.01 | 0.67 (0.55 - 0.81) | <0.01 |
| **PS-stratification / on-treatment TAR without escalation** | | | | | | | | | | |
| CCAE | 12,270 | 85,777 | 7.0 | 63.6 | 87 | 979 | 0.80 (0.62 - 1.01) | 0.07 | 0.78 (0.61 - 0.99) | 0.06 |
| ODOD | 7,889 | 63,755 | 4.0 | 52.2 | 67 | 2,184 | 0.51 (0.39 - 0.66) | <0.01 | 0.48 (0.37 - 0.62) | <0.01 |
| MDCR | 895 | 17,003 | 0.5 | 16.3 | 18 | 937 | 0.69 (0.41 - 1.10) | 0.15 | 0.63 (0.38 - 1.04) | 0.08 |
| Open Claims | 143,665 | 813,841 | 86.8 | 742.6 | 830 | 17,034 | 0.65 (0.60 - 0.70) | <0.01 | 0.65 (0.53 - 0.79) | <0.01 |
| OEHR | 8,765 | 91,617 | 2.2 | 43.5 | 28 | 1,045 | 0.69 (0.45 - 1.00) | 0.06 | 0.62 (0.42 - 0.93) | 0.03 |
| VA | 1,282 | 223,743 | 0.7 | 210.3 | <10 | 3,467 | 0.47 (0.17 - 1.02) | 0.10 | 0.37 (0.15 - 0.93) | 0.04 |
| Meta-analysis | 172,589 | 1,054,990 | 99.9 | 902.0 | 1,012 | 21,242 | 0.65 (0.55 - 0.76) | <0.01 | 0.64 (0.49 - 0.82) | <0.01 |
| Meta-analysis (w/o USOC) | 28,924 | 241,149 | 13.2 | 159.4 | 182 | 4,208 | 0.65 (0.49 - 0.87) | <0.01 | 0.62 (0.45 - 0.85) | <0.01 |

##### 1.5.3 Acute myocardial infarction

**Supplementary Table 107:** Exposures, outcomes and risk estimates for Acute myocardial infarction in GLP1RA (male) (T) vs SU (male) (C) new-user cohorts under four different propensity score (PS) adjustment / time-at-risk (TAR) designs across data sources. We report total patient exposure time in 1,000 patient-year (KYr) units, uncalibrated and calibrated hazard ratios (HRs) and their 95% confidence intervals (CIs). Greyed out entries did not pass study diagnostics and are included only for completeness.

| | Patients | | Exposure (KYr) | | Outcomes | | Uncalibrated | | Calibrated | |
|---|---|---|---|---|---|---|---|---|---|---|
| | T | C | T | C | T | C | HR (95% CI) | $p$ | HR (95% CI) | $p$ |
| **PS-matching / on-treatment TAR** | | | | | | | | | | |
| CCAE | 11,160 | 83,945 | 7.5 | 72.1 | 34 | 434 | 0.83 (0.53 - 1.27) | 0.41 | 0.81 (0.52 - 1.26) | 0.35 |
| ODOD | 7,417 | 61,088 | 4.4 | 55.5 | 17 | 658 | 0.42 (0.22 - 0.73) | <0.01 | 0.40 (0.22 - 0.72) | <0.01 |
| MDCR | 898 | 14,949 | 0.6 | 16.3 | 7 | 222 | 0.78 (0.23 - 2.01) | 0.66 | 0.77 (0.26 - 2.27) | 0.63 |
| Open Claims | 135,489 | 841,364 | 96.5 | 908.9 | 319 | 6,108 | 0.66 (0.58 - 0.76) | <0.01 | 0.67 (0.55 - 0.82) | <0.01 |
| OEHR | 8,240 | 85,150 | 2.4 | 46.6 | 9 | 344 | 0.55 (0.25 - 1.08) | 0.11 | 0.51 (0.25 - 1.07) | 0.08 |
| VA | 1,264 | 31,658 | 0.9 | 23.9 | <10 | 99 | 1.36 (0.37 - 3.84) | 0.60 | 1.06 (0.33 - 3.41) | 0.91 |
| Meta-analysis | 163,204 | 1,086,496 | 111.4 | 1099.4 | 386 | 7,766 | 0.66 (0.58 - 0.75) | <0.01 | 0.65 (0.53 - 0.80) | <0.01 |
| Meta-analysis (w/o USOC) | 27,715 | 245,132 | 14.9 | 190.5 | 67 | 1,658 | 0.63 (0.44 - 0.90) | 0.01 | 0.60 (0.42 - 0.86) | <0.01 |
| **PS-stratification / on-treatment TAR** | | | | | | | | | | |
| CCAE | 12,650 | 88,414 | 8.3 | 75.7 | 40 | 456 | 0.85 (0.59 - 1.20) | 0.38 | 0.83 (0.58 - 1.18) | 0.31 |
| ODOD | 8,318 | 68,419 | 4.8 | 63.9 | 19 | 825 | 0.40 (0.24 - 0.63) | <0.01 | 0.39 (0.24 - 0.63) | <0.01 |
| MDCR | 975 | 19,284 | 0.6 | 20.6 | 7 | 323 | 0.88 (0.36 - 1.81) | 0.75 | 0.81 (0.36 - 1.82) | 0.62 |
| Open Claims | 150,626 | 868,394 | 103.8 | 940.1 | 346 | 6,663 | 0.68 (0.61 - 0.76) | <0.01 | 0.69 (0.56 - 0.86) | <0.01 |
| OEHR | 8,966 | 94,391 | 2.6 | 52.1 | 10 | 434 | 0.54 (0.26 - 0.99) | 0.07 | 0.49 (0.25 - 0.96) | 0.04 |
| VA | 1,361 | 233,198 | 0.9 | 248.0 | <10 | 1,249 | 1.36 (0.48 - 3.04) | 0.51 | 1.15 (0.46 - 2.91) | 0.77 |
| Meta-analysis | 180,560 | 1,119,618 | 119.4 | 1131.9 | 415 | 8,378 | 0.63 (0.47 - 0.84) | <0.01 | 0.62 (0.44 - 0.89) | <0.01 |
| Meta-analysis (w/o USOC) | 29,934 | 251,224 | 15.6 | 191.8 | 69 | 1,715 | 0.58 (0.36 - 0.96) | 0.04 | 0.56 (0.33 - 0.92) | 0.03 |
| **PS-matching / on-treatment TAR without escalation** | | | | | | | | | | |
| CCAE | 11,160 | 83,945 | 6.5 | 62.5 | 32 | 372 | 0.79 (0.49 - 1.23) | 0.32 | 0.77 (0.48 - 1.22) | 0.27 |
| ODOD | 7,417 | 61,088 | 3.9 | 49.4 | 14 | 591 | 0.35 (0.16 - 0.66) | <0.01 | 0.33 (0.16 - 0.66) | <0.01 |
| MDCR | 898 | 14,949 | 0.5 | 14.7 | <5 | 196 | 0.75 (0.17 - 2.23) | 0.66 | 0.74 (0.21 - 2.66) | 0.64 |
| Open Claims | 135,489 | 841,364 | 84.0 | 769.9 | 263 | 5,189 | 0.62 (0.53 - 0.72) | <0.01 | 0.62 (0.51 - 0.77) | <0.01 |
| OEHR | 8,240 | 85,150 | 2.1 | 40.7 | 8 | 246 | 0.55 (0.22 - 1.18) | 0.16 | 0.52 (0.22 - 1.19) | 0.12 |
| VA | 1,264 | 31,658 | 0.7 | 20.9 | <10 | 88 | 1.37 (0.29 - 4.49) | 0.65 | 1.02 (0.26 - 4.03) | 0.93 |
| Meta-analysis | 163,204 | 1,086,496 | 97.0 | 937.2 | <322 | 6,594 | 0.62 (0.54 - 0.71) | <0.01 | 0.61 (0.49 - 0.76) | <0.01 |
| Meta-analysis (w/o USOC) | 27,715 | 245,132 | 13.0 | 167.3 | <59 | 1,405 | 0.59 (0.39 - 0.89) | 0.01 | 0.57 (0.38 - 0.86) | <0.01 |
| **PS-stratification / on-treatment TAR without escalation** | | | | | | | | | | |
| CCAE | 12,650 | 88,414 | 7.2 | 65.8 | 37 | 393 | 0.90 (0.61 - 1.30) | 0.58 | 0.88 (0.60 - 1.28) | 0.51 |
| ODOD | 8,318 | 68,419 | 4.2 | 57.1 | 16 | 737 | 0.37 (0.21 - 0.61) | <0.01 | 0.35 (0.21 - 0.59) | <0.01 |
| MDCR | 975 | 19,284 | 0.6 | 18.7 | <5 | 283 | 0.59 (0.17 - 1.46) | 0.32 | 0.54 (0.19 - 1.55) | 0.25 |
| Open Claims | 150,626 | 868,394 | 90.8 | 796.7 | 289 | 5,680 | 0.66 (0.58 - 0.75) | <0.01 | 0.66 (0.53 - 0.83) | <0.01 |
| OEHR | 8,966 | 94,391 | 2.2 | 45.4 | 9 | 303 | 0.66 (0.30 - 1.28) | 0.26 | 0.60 (0.29 - 1.23) | 0.17 |
| VA | 1,361 | 233,198 | 0.7 | 221.0 | <10 | 1,055 | 1.72 (0.60 - 3.88) | 0.25 | 1.37 (0.54 - 3.49) | 0.51 |
| Meta-analysis | 180,560 | 1,119,618 | 104.4 | 964.9 | 351 | 7,113 | 0.64 (0.46 - 0.89) | <0.01 | 0.63 (0.43 - 0.92) | 0.02 |
| Meta-analysis (w/o USOC) | 29,934 | 251,224 | 13.7 | 168.2 | 62 | 1,433 | 0.62 (0.35 - 1.09) | 0.10 | 0.58 (0.32 - 1.04) | 0.07 |

##### 1.5.4 Acute renal failure

**Supplementary Table 108:** Exposures, outcomes and risk estimates for Acute renal failure in GLP1RA (male) (T) vs SU (male) (C) new-user cohorts under four different propensity score (PS) adjustment / time-at-risk (TAR) designs across data sources. We report total patient exposure time in 1,000 patient-year (KYr) units, uncalibrated and calibrated hazard ratios (HRs) and their 95% confidence intervals (CIs). Greyed out entries did not pass study diagnostics and are included only for completeness.

| | Patients | | Exposure (KYr) | | Outcomes | | Uncalibrated | | Calibrated | |
|---|---|---|---|---|---|---|---|---|---|---|
| | T | C | T | C | T | C | HR (95% CI) | $p$ | HR (95% CI) | $p$ |
| **PS-matching / on-treatment TAR** | | | | | | | | | | |
| CCAE | 11,229 | 84,870 | 7.5 | 73.0 | 53 | 715 | 0.60 (0.41 - 0.86) | <0.01 | 0.59 (0.40 - 0.85) | <0.01 |
| ODOD | 7,315 | 60,157 | 4.3 | 54.2 | 83 | 1,773 | 0.64 (0.48 - 0.84) | <0.01 | 0.61 (0.46 - 0.81) | <0.01 |
| MDCR | 892 | 14,768 | 0.6 | 15.9 | 23 | 533 | 0.91 (0.52 - 1.53) | 0.74 | 0.89 (0.52 - 1.54) | 0.69 |
| Open Claims | 135,101 | 837,375 | 96.2 | 901.1 | 860 | 15,464 | 0.70 (0.64 - 0.76) | <0.01 | 0.71 (0.60 - 0.83) | <0.01 |
| OEHR | 8,242 | 85,033 | 2.4 | 46.2 | 34 | 891 | 0.90 (0.59 - 1.31) | 0.59 | 0.84 (0.56 - 1.25) | 0.40 |
| VA | 1,249 | 31,250 | 0.9 | 23.4 | <10 | 361 | 0.70 (0.28 - 1.46) | 0.39 | 0.54 (0.24 - 1.23) | 0.15 |
| Meta-analysis | 162,779 | 1,082,203 | 111.0 | 1090.4 | 1,053 | 19,376 | 0.70 (0.65 - 0.75) | <0.01 | 0.69 (0.58 - 0.83) | <0.01 |
| Meta-analysis (w/o USOC) | 27,678 | 244,828 | 14.8 | 189.3 | 193 | 3,912 | 0.71 (0.58 - 0.87) | <0.01 | 0.68 (0.56 - 0.83) | <0.01 |
| **PS-stratification / on-treatment TAR** | | | | | | | | | | |
| CCAE | 12,718 | 88,812 | 8.3 | 76.2 | 61 | 777 | 0.62 (0.46 - 0.82) | <0.01 | 0.60 (0.45 - 0.80) | <0.01 |
| ODOD | 8,198 | 66,571 | 4.7 | 61.5 | 91 | 2,227 | 0.66 (0.52 - 0.83) | <0.01 | 0.63 (0.50 - 0.80) | <0.01 |
| MDCR | 963 | 18,886 | 0.6 | 20.0 | 23 | 791 | 1.10 (0.68 - 1.69) | 0.69 | 1.02 (0.65 - 1.61) | 0.85 |
| Open Claims | 150,166 | 862,791 | 103.4 | 930.8 | 898 | 16,968 | 0.67 (0.62 - 0.72) | <0.01 | 0.68 (0.56 - 0.82) | <0.01 |
| OEHR | 8,969 | 94,051 | 2.6 | 51.5 | 34 | 1,133 | 0.79 (0.54 - 1.12) | 0.21 | 0.73 (0.51 - 1.05) | 0.11 |
| VA | 1,343 | 232,074 | 0.9 | 244.9 | <10 | 3,743 | 0.61 (0.28 - 1.15) | 0.17 | 0.52 (0.25 - 1.05) | 0.07 |
| Meta-analysis | 180,051 | 1,112,225 | 118.9 | 1119.9 | 1,084 | 21,105 | 0.67 (0.63 - 0.71) | <0.01 | 0.66 (0.54 - 0.81) | <0.01 |
| Meta-analysis (w/o USOC) | 29,885 | 249,434 | 15.5 | 189.2 | 186 | 4,137 | 0.67 (0.57 - 0.79) | <0.01 | 0.64 (0.53 - 0.77) | <0.01 |
| **PS-matching / on-treatment TAR without escalation** | | | | | | | | | | |
| CCAE | 11,229 | 84,870 | 6.5 | 63.3 | 47 | 609 | 0.66 (0.45 - 0.96) | 0.04 | 0.64 (0.44 - 0.95) | 0.03 |
| ODOD | 7,315 | 60,157 | 3.8 | 48.3 | 74 | 1,587 | 0.70 (0.52 - 0.93) | 0.02 | 0.66 (0.49 - 0.89) | <0.01 |
| MDCR | 892 | 14,768 | 0.5 | 14.4 | 19 | 467 | 0.90 (0.48 - 1.57) | 0.72 | 0.88 (0.49 - 1.60) | 0.68 |
| Open Claims | 135,101 | 837,375 | 83.7 | 764.2 | 717 | 12,926 | 0.70 (0.63 - 0.76) | <0.01 | 0.70 (0.59 - 0.82) | <0.01 |
| OEHR | 8,242 | 85,033 | 2.1 | 40.4 | 29 | 655 | 1.02 (0.65 - 1.54) | 0.94 | 0.95 (0.61 - 1.47) | 0.81 |
| VA | 1,249 | 31,250 | 0.7 | 20.5 | <10 | 293 | 0.69 (0.26 - 1.55) | 0.43 | 0.52 (0.21 - 1.28) | 0.16 |
| Meta-analysis | 162,779 | 1,082,203 | 96.6 | 930.6 | 886 | 16,244 | 0.71 (0.65 - 0.77) | <0.01 | 0.70 (0.58 - 0.84) | <0.01 |
| Meta-analysis (w/o USOC) | 27,678 | 244,828 | 13.0 | 166.4 | 169 | 3,318 | 0.77 (0.63 - 0.93) | <0.01 | 0.73 (0.60 - 0.89) | <0.01 |
| **PS-stratification / on-treatment TAR without escalation** | | | | | | | | | | |
| CCAE | 12,718 | 88,812 | 7.2 | 66.2 | 53 | 663 | 0.64 (0.47 - 0.87) | <0.01 | 0.63 (0.46 - 0.86) | <0.01 |
| ODOD | 8,198 | 66,571 | 4.2 | 55.0 | 80 | 1,986 | 0.66 (0.52 - 0.84) | <0.01 | 0.62 (0.48 - 0.79) | <0.01 |
| MDCR | 963 | 18,886 | 0.6 | 18.3 | 19 | 704 | 1.02 (0.60 - 1.63) | 0.94 | 0.93 (0.57 - 1.54) | 0.78 |
| Open Claims | 150,166 | 862,791 | 90.4 | 789.9 | 753 | 14,219 | 0.66 (0.61 - 0.71) | <0.01 | 0.66 (0.54 - 0.80) | <0.01 |
| OEHR | 8,969 | 94,051 | 2.2 | 45.0 | 29 | 834 | 0.87 (0.57 - 1.26) | 0.48 | 0.79 (0.53 - 1.17) | 0.26 |
| VA | 1,343 | 232,074 | 0.7 | 218.6 | <10 | 3,030 | 0.72 (0.31 - 1.42) | 0.40 | 0.57 (0.27 - 1.24) | 0.16 |
| Meta-analysis | 180,051 | 1,112,225 | 104.1 | 956.0 | 915 | 17,702 | 0.66 (0.62 - 0.71) | <0.01 | 0.65 (0.52 - 0.80) | <0.01 |
| Meta-analysis (w/o USOC) | 29,885 | 249,434 | 13.6 | 166.2 | 162 | 3,483 | 0.69 (0.58 - 0.82) | <0.01 | 0.65 (0.52 - 0.81) | <0.01 |

##### 1.5.5 Glycemic control

**Supplementary Table 109:** Exposures, outcomes and risk estimates for Glycemic control in GLP1RA (male) (T) vs SU (male) (C) new-user cohorts under four different propensity score (PS) adjustment / time-at-risk (TAR) designs across data sources. We report total patient exposure time in 1,000 patient-year (KYr) units, uncalibrated and calibrated hazard ratios (HRs) and their 95% confidence intervals (CIs). Greyed out entries did not pass study diagnostics and are included only for completeness.

| | Patients | | Exposure (KYr) | | Outcomes | | Uncalibrated | | Calibrated | |
|---|---|---|---|---|---|---|---|---|---|---|
| | T | C | T | C | T | C | HR (95% CI) | $p$ | HR (95% CI) | $p$ |
| **PS-matching / on-treatment TAR** | | | | | | | | | | |
| CCAE | 11,397 | 85,482 | 7.7 | 73.6 | 6 | 301 | 0.32 (0.08 - 0.89) | 0.07 | 0.31 (0.09 - 1.06) | 0.06 |
| ODOD | 5,201 | 45,690 | 2.8 | 36.9 | 311 | 3,538 | 1.21 (1.04 - 1.41) | 0.01 | 1.16 (0.99 - 1.35) | 0.09 |
| MDCR | 931 | 15,433 | 0.6 | 16.9 | <5 | 77 | 0.72 (0.04 - 3.75) | 0.77 | 0.70 (0.07 - 6.90) | 0.76 |
| Open Claims | 140,133 | 873,951 | 99.8 | 948.1 | 0 | 0 | | | | |
| OEHR | 8,369 | 86,443 | 2.4 | 47.7 | 0 | 0 | | | | |
| VA | 1,245 | 30,995 | 0.9 | 23.1 | 0 | 45 | | | | |
| Meta-analysis | 17,529 | 146,605 | 11.1 | 127.3 | <322 | 3,916 | 0.76 (0.29 - 1.98) | 0.58 | 0.76 (0.29 - 1.99) | 0.57 |
| Meta-analysis (w/o USOC) | 17,529 | 146,605 | 11.1 | 127.3 | <322 | 3,916 | 0.76 (0.30 - 1.97) | 0.58 | 0.73 (0.28 - 1.89) | 0.52 |
| **PS-stratification / on-treatment TAR** | | | | | | | | | | |
| CCAE | 12,911 | 89,467 | 8.4 | 76.6 | 6 | 512 | 0.42 (0.16 - 0.91) | 0.05 | 0.41 (0.17 - 0.98) | 0.05 |
| ODOD | 5,649 | 52,045 | 3.0 | 43.2 | 329 | 4,236 | 1.17 (1.03 - 1.33) | 0.01 | 1.12 (0.99 - 1.28) | 0.14 |
| MDCR | 1,007 | 20,047 | 0.7 | 21.4 | <5 | 178 | 1.12 (0.06 - 5.16) | 0.92 | 1.04 (0.12 - 9.43) | 0.96 |
| Open Claims | 155,612 | 903,312 | 107.2 | 981.9 | 0 | 0 | | | | |
| OEHR | 9,105 | 95,902 | 2.6 | 53.4 | 0 | 0 | | | | |
| VA | 1,342 | 227,080 | 0.9 | 239.3 | 0 | 536 | | | | |
| Meta-analysis | 18,560 | 141,512 | 11.4 | 119.8 | 335 | 4,748 | 0.78 (0.29 - 2.07) | 0.61 | 0.77 (0.28 - 2.09) | 0.60 |
| Meta-analysis (w/o USOC) | 18,560 | 141,512 | 11.4 | 119.8 | 335 | 4,748 | 0.78 (0.29 - 2.07) | 0.61 | 0.74 (0.27 - 1.98) | 0.55 |
| **PS-matching / on-treatment TAR without escalation** | | | | | | | | | | |
| CCAE | 11,397 | 85,482 | 6.6 | 63.6 | 5 | 270 | 0.39 (0.09 - 1.07) | 0.13 | 0.37 (0.11 - 1.27) | 0.12 |
| ODOD | 5,201 | 45,690 | 2.5 | 32.9 | 280 | 3,207 | 1.19 (1.01 - 1.39) | 0.03 | 1.13 (0.96 - 1.32) | 0.20 |
| MDCR | 931 | 15,433 | 0.6 | 15.2 | <5 | 70 | 1.58 (0.09 - 7.73) | 0.69 | 1.56 (0.17 - 14.7) | 0.70 |
| Open Claims | 140,133 | 873,951 | 86.7 | 802.3 | 0 | 0 | | | | |
| OEHR | 8,369 | 86,443 | 2.1 | 41.5 | 0 | 0 | | | | |
| VA | 1,245 | 30,995 | 0.7 | 20.3 | 0 | 43 | | | | |
| Meta-analysis | 16,598 | 131,172 | 9.1 | 96.5 | 285 | 3,477 | 0.80 (0.28 - 2.30) | 0.68 | 0.79 (0.27 - 2.29) | 0.66 |
| Meta-analysis (w/o USOC) | 16,598 | 131,172 | 9.1 | 96.5 | 285 | 3,477 | 0.80 (0.28 - 2.30) | 0.68 | 0.77 (0.27 - 2.21) | 0.63 |
| **PS-stratification / on-treatment TAR without escalation** | | | | | | | | | | |
| CCAE | 12,911 | 89,467 | 7.3 | 66.4 | 5 | 467 | 0.41 (0.14 - 0.95) | 0.07 | 0.40 (0.16 - 1.04) | 0.06 |
| ODOD | 5,649 | 52,045 | 2.6 | 38.6 | 297 | 3,853 | 1.20 (1.05 - 1.36) | <0.01 | 1.12 (0.97 - 1.28) | 0.17 |
| MDCR | 1,007 | 20,047 | 0.6 | 19.4 | <5 | 168 | 1.45 (0.08 - 6.66) | 0.74 | 1.32 (0.15 - 12.0) | 0.80 |
| Open Claims | 155,612 | 903,312 | 93.6 | 831.4 | 0 | 0 | | | | |
| OEHR | 9,105 | 95,902 | 2.3 | 46.3 | 0 | 0 | | | | |
| VA | 1,342 | 227,080 | 0.7 | 213.0 | 0 | 500 | | | | |
| Meta-analysis | 18,560 | 141,512 | 10.0 | 105.0 | 302 | 4,320 | 0.78 (0.28 - 2.17) | 0.64 | 0.77 (0.27 - 2.17) | 0.62 |
| Meta-analysis (w/o USOC) | 18,560 | 141,512 | 10.0 | 105.0 | 302 | 4,320 | 0.78 (0.28 - 2.17) | 0.64 | 0.74 (0.26 - 2.07) | 0.57 |

##### 1.5.6 Hospitalization with heart failure

**Supplementary Table 110:** Exposures, outcomes and risk estimates for Hospitalization with heart failure in GLP1RA (male) (T) vs SU (male) (C) new-user cohorts under four different propensity score (PS) adjustment / time-at-risk (TAR) designs across data sources. We report total patient exposure time in 1,000 patient-year (KYr) units, uncalibrated and calibrated hazard ratios (HRs) and their 95% confidence intervals (CIs). Greyed out entries did not pass study diagnostics and are included only for completeness.

| | Patients | | Exposure (KYr) | | Outcomes | | Uncalibrated | | Calibrated | |
|---|---|---|---|---|---|---|---|---|---|---|
| | T | C | T | C | T | C | HR (95% CI) | $p$ | HR (95% CI) | $p$ |
| **PS-matching / on-treatment TAR** | | | | | | | | | | |
| CCAE | 11,207 | 84,502 | 7.5 | 72.7 | 45 | 537 | 0.76 (0.51 - 1.12) | 0.18 | 0.74 (0.50 - 1.10) | 0.15 |
| ODOD | 7,267 | 59,738 | 4.3 | 53.9 | 51 | 1,333 | 0.57 (0.40 - 0.79) | <0.01 | 0.54 (0.38 - 0.77) | <0.01 |
| MDCR | 867 | 14,380 | 0.6 | 15.5 | 17 | 494 | 1.10 (0.58 - 1.92) | 0.75 | 1.08 (0.59 - 1.96) | 0.81 |
| Open Claims | 135,009 | 834,445 | 96.3 | 899.5 | 514 | 11,503 | 0.64 (0.57 - 0.71) | <0.01 | 0.65 (0.54 - 0.77) | <0.01 |
| OEHR | 8,208 | 84,581 | 2.4 | 45.9 | 16 | 734 | 0.52 (0.29 - 0.88) | 0.02 | 0.49 (0.28 - 0.85) | 0.01 |
| VA | 1,240 | 31,306 | 0.9 | 23.3 | <10 | 289 | 0.41 (0.10 - 1.14) | 0.15 | 0.32 (0.09 - 1.08) | 0.07 |
| Meta-analysis | 162,558 | 1,077,646 | 111.1 | 1087.4 | 643 | 14,601 | 0.64 (0.58 - 0.71) | <0.01 | 0.64 (0.53 - 0.77) | <0.01 |
| Meta-analysis (w/o USOC) | 27,549 | 243,201 | 14.8 | 187.9 | 129 | 3,098 | 0.68 (0.51 - 0.91) | <0.01 | 0.65 (0.49 - 0.87) | <0.01 |
| **PS-stratification / on-treatment TAR** | | | | | | | | | | |
| CCAE | 12,687 | 88,628 | 8.3 | 76.0 | 53 | 573 | 0.72 (0.52 - 0.98) | 0.04 | 0.70 (0.51 - 0.96) | 0.03 |
| ODOD | 8,142 | 66,524 | 4.7 | 61.6 | 55 | 1,697 | 0.53 (0.40 - 0.71) | <0.01 | 0.51 (0.38 - 0.68) | <0.01 |
| MDCR | 939 | 18,277 | 0.6 | 19.5 | 18 | 738 | 1.02 (0.60 - 1.65) | 0.93 | 0.95 (0.57 - 1.58) | 0.82 |
| Open Claims | 150,190 | 860,350 | 103.6 | 930.0 | 538 | 12,800 | 0.60 (0.54 - 0.66) | <0.01 | 0.61 (0.49 - 0.74) | <0.01 |
| OEHR | 8,933 | 93,635 | 2.5 | 51.2 | 17 | 920 | 0.51 (0.30 - 0.82) | <0.01 | 0.48 (0.29 - 0.78) | <0.01 |
| VA | 1,340 | 231,003 | 0.9 | 244.2 | <10 | 2,973 | 0.26 (0.07 - 0.69) | 0.03 | 0.22 (0.07 - 0.73) | 0.01 |
| Meta-analysis | 179,952 | 1,109,137 | 119.1 | 1118.8 | 663 | 15,990 | 0.60 (0.55 - 0.65) | <0.01 | 0.59 (0.48 - 0.73) | <0.01 |
| Meta-analysis (w/o USOC) | 29,762 | 248,787 | 15.5 | 188.8 | 125 | 3,190 | 0.60 (0.48 - 0.74) | <0.01 | 0.57 (0.45 - 0.72) | <0.01 |
| **PS-matching / on-treatment TAR without escalation** | | | | | | | | | | |
| CCAE | 11,207 | 84,502 | 6.5 | 63.0 | 42 | 457 | 0.83 (0.53 - 1.25) | 0.39 | 0.81 (0.53 - 1.24) | 0.33 |
| ODOD | 7,267 | 59,738 | 3.8 | 47.9 | 47 | 1,173 | 0.63 (0.44 - 0.89) | 0.01 | 0.60 (0.42 - 0.86) | <0.01 |
| MDCR | 867 | 14,380 | 0.5 | 14.1 | 13 | 443 | 0.90 (0.43 - 1.70) | 0.77 | 0.89 (0.44 - 1.78) | 0.74 |
| Open Claims | 135,009 | 834,445 | 83.7 | 762.3 | 441 | 9,760 | 0.66 (0.58 - 0.74) | <0.01 | 0.66 (0.55 - 0.79) | <0.01 |
| OEHR | 8,208 | 84,581 | 2.1 | 40.2 | 15 | 565 | 0.59 (0.32 - 1.02) | 0.08 | 0.56 (0.31 - 0.99) | 0.05 |
| VA | 1,240 | 31,306 | 0.7 | 20.5 | <10 | 238 | 0.49 (0.12 - 1.38) | 0.26 | 0.37 (0.11 - 1.26) | 0.11 |
| Meta-analysis | 162,558 | 1,077,646 | 96.6 | 927.6 | 558 | 12,398 | 0.67 (0.60 - 0.74) | <0.01 | 0.66 (0.54 - 0.80) | <0.01 |
| Meta-analysis (w/o USOC) | 27,549 | 243,201 | 12.9 | 165.2 | 117 | 2,638 | 0.71 (0.56 - 0.89) | <0.01 | 0.68 (0.53 - 0.86) | <0.01 |
| **PS-stratification / on-treatment TAR without escalation** | | | | | | | | | | |
| CCAE | 12,687 | 88,628 | 7.2 | 66.0 | 48 | 487 | 0.81 (0.58 - 1.12) | 0.22 | 0.79 (0.57 - 1.11) | 0.18 |
| ODOD | 8,142 | 66,524 | 4.1 | 55.0 | 51 | 1,497 | 0.57 (0.41 - 0.76) | <0.01 | 0.53 (0.39 - 0.72) | <0.01 |
| MDCR | 939 | 18,277 | 0.6 | 17.7 | 14 | 673 | 0.90 (0.49 - 1.53) | 0.71 | 0.82 (0.46 - 1.46) | 0.50 |
| Open Claims | 150,190 | 860,350 | 90.5 | 788.6 | 463 | 10,898 | 0.60 (0.55 - 0.67) | <0.01 | 0.60 (0.49 - 0.74) | <0.01 |
| OEHR | 8,933 | 93,635 | 2.2 | 44.8 | 16 | 704 | 0.60 (0.34 - 0.97) | 0.05 | 0.54 (0.32 - 0.91) | 0.02 |
| VA | 1,340 | 231,003 | 0.7 | 217.9 | <10 | 2,550 | 0.36 (0.09 - 0.95) | 0.09 | 0.29 (0.09 - 0.94) | 0.04 |
| Meta-analysis | 179,952 | 1,109,137 | 104.1 | 954.4 | 578 | 13,586 | 0.61 (0.56 - 0.67) | <0.01 | 0.60 (0.48 - 0.74) | <0.01 |
| Meta-analysis (w/o USOC) | 29,762 | 248,787 | 13.6 | 165.8 | 115 | 2,688 | 0.66 (0.52 - 0.83) | <0.01 | 0.62 (0.47 - 0.81) | <0.01 |

##### 1.5.7 Measured renal dysfunction

**Supplementary Table 111:** Exposures, outcomes and risk estimates for Measured renal dysfunction in GLP1RA (male) (T) vs SU (male) (C) new-user cohorts under four different propensity score (PS) adjustment / time-at-risk (TAR) designs across data sources. We report total patient exposure time in 1,000 patient-year (KYr) units, uncalibrated and calibrated hazard ratios (HRs) and their 95% confidence intervals (CIs). Greyed out entries did not pass study diagnostics and are included only for completeness.

| | Patients | | Exposure (KYr) | | Outcomes | | Uncalibrated | | Calibrated | |
|---|---|---|---|---|---|---|---|---|---|---|
| | T | C | T | C | T | C | HR (95% CI) | $p$ | HR (95% CI) | $p$ |
| **PS-matching / on-treatment TAR** | | | | | | | | | | |
| CCAE | 11,450 | 86,141 | 7.7 | 74.3 | 0 | 30 | | | | |
| ODOD | 7,591 | 62,908 | 4.5 | 57.5 | 6 | 109 | 0.68 (0.15 - 2.12) | 0.57 | 0.65 (0.18 - 2.43) | 0.53 |
| MDCR | 932 | 15,503 | 0.6 | 17.0 | 0 | 5 | | | | |
| Open Claims | 140,133 | 873,951 | 99.8 | 948.1 | 0 | 0 | | | | |
| OEHR | 8,369 | 86,443 | 2.4 | 47.7 | 0 | 0 | | | | |
| VA | 1,293 | 31,902 | 0.9 | 23.9 | <10 | 143 | 1.15 (0.37 - 2.92) | 0.79 | 0.89 (0.32 - 2.51) | 0.83 |
| Meta-analysis | 7,591 | 62,908 | 4.5 | 57.5 | 6 | 109 | 0.68 (0.15 - 2.12) | 0.57 | 0.68 (0.18 - 2.55) | 0.57 |
| Meta-analysis (w/o USOC) | 7,591 | 62,908 | 4.5 | 57.5 | 6 | 109 | 0.68 (0.15 - 2.12) | 0.57 | 0.66 (0.18 - 2.44) | 0.53 |
| **PS-stratification / on-treatment TAR** | | | | | | | | | | |
| CCAE | 12,979 | 90,487 | 8.5 | 77.8 | 0 | 31 | | | | |
| ODOD | 8,508 | 70,635 | 4.9 | 66.4 | 6 | 146 | 0.80 (0.29 - 1.81) | 0.63 | 0.76 (0.31 - 1.90) | 0.56 |
| MDCR | 1,008 | 20,367 | 0.7 | 21.9 | 0 | 8 | | | | |
| Open Claims | 155,612 | 903,312 | 107.2 | 981.9 | 0 | 0 | | | | |
| OEHR | 9,105 | 95,902 | 2.6 | 53.4 | 0 | 0 | | | | |
| VA | 1,389 | 235,040 | 1.0 | 249.7 | <10 | 1,309 | 0.85 (0.30 - 1.88) | 0.73 | 0.72 (0.29 - 1.80) | 0.49 |
| Meta-analysis | 8,508 | 70,635 | 4.9 | 66.4 | 6 | 146 | 0.80 (0.29 - 1.81) | 0.63 | 0.79 (0.31 - 2.00) | 0.62 |
| Meta-analysis (w/o USOC) | 8,508 | 70,635 | 4.9 | 66.4 | 6 | 146 | 0.80 (0.29 - 1.81) | 0.63 | 0.76 (0.31 - 1.90) | 0.56 |
| **PS-matching / on-treatment TAR without escalation** | | | | | | | | | | |
| CCAE | 11,450 | 86,141 | 6.7 | 64.3 | 0 | 26 | | | | |
| ODOD | 7,591 | 62,908 | 4.0 | 51.1 | <5 | 94 | 0.65 (0.10 - 2.35) | 0.59 | 0.62 (0.13 - 2.97) | 0.55 |
| MDCR | 932 | 15,503 | 0.6 | 15.3 | 0 | <5 | | | | |
| Open Claims | 140,133 | 873,951 | 86.7 | 802.3 | 0 | 0 | | | | |
| OEHR | 8,369 | 86,443 | 2.1 | 41.5 | 0 | 0 | | | | |
| VA | 1,293 | 31,902 | 0.7 | 21.0 | <10 | 119 | 1.31 (0.37 - 3.58) | 0.64 | 0.98 (0.31 - 3.07) | 0.92 |
| Meta-analysis | 7,591 | 62,908 | 4.0 | 51.1 | <5 | 94 | 0.65 (0.10 - 2.35) | 0.59 | 0.64 (0.13 - 3.12) | 0.58 |
| Meta-analysis (w/o USOC) | 7,591 | 62,908 | 4.0 | 51.1 | <5 | 94 | 0.65 (0.10 - 2.35) | 0.59 | 0.62 (0.13 - 3.02) | 0.56 |
| **PS-stratification / on-treatment TAR without escalation** | | | | | | | | | | |
| CCAE | 12,979 | 90,487 | 7.4 | 67.5 | 0 | 27 | | | | |
| ODOD | 8,508 | 70,635 | 4.3 | 59.1 | <5 | 126 | 0.73 (0.21 - 1.88) | 0.57 | 0.68 (0.23 - 2.02) | 0.49 |
| MDCR | 1,008 | 20,367 | 0.6 | 19.9 | 0 | 7 | | | | |
| Open Claims | 155,612 | 903,312 | 93.6 | 831.4 | 0 | 0 | | | | |
| OEHR | 9,105 | 95,902 | 2.3 | 46.3 | 0 | 0 | | | | |
| VA | 1,389 | 235,040 | 0.8 | 222.5 | <10 | 1,073 | 0.91 (0.28 - 2.17) | 0.86 | 0.72 (0.26 - 2.03) | 0.54 |
| Meta-analysis | 8,508 | 70,635 | 4.3 | 59.1 | <5 | 126 | 0.73 (0.21 - 1.88) | 0.57 | 0.71 (0.23 - 2.16) | 0.55 |
| Meta-analysis (w/o USOC) | 8,508 | 70,635 | 4.3 | 59.1 | <5 | 126 | 0.73 (0.21 - 1.88) | 0.57 | 0.69 (0.23 - 2.06) | 0.50 |

##### 1.5.8 Stroke

**Supplementary Table 112:** Exposures, outcomes and risk estimates for Stroke in GLP1RA (male) (T) vs SU (male) (C) new-user cohorts under four different propensity score (PS) adjustment / time-at-risk (TAR) designs across data sources. We report total patient exposure time in 1,000 patient-year (KYr) units, uncalibrated and calibrated hazard ratios (HRs) and their 95% confidence intervals (CIs). Greyed out entries did not pass study diagnostics and are included only for completeness.

| | Patients | | Exposure (KYr) | | Outcomes | | Uncalibrated | | Calibrated | |
|---|---|---|---|---|---|---|---|---|---|---|
| | T | C | T | C | T | C | HR (95% CI) | $p$ | HR (95% CI) | $p$ |
| **PS-matching / on-treatment TAR** | | | | | | | | | | |
| CCAE | 11,318 | 85,192 | 7.6 | 73.5 | 23 | 259 | 1.00 (0.58 - 1.64) | 0.99 | 0.97 (0.57 - 1.63) | 0.90 |
| ODOD | 7,468 | 61,465 | 4.4 | 55.9 | 23 | 595 | 0.85 (0.49 - 1.39) | 0.55 | 0.81 (0.48 - 1.37) | 0.44 |
| MDCR | 905 | 14,960 | 0.6 | 16.2 | 5 | 251 | 0.28 (0.04 - 0.94) | 0.10 | 0.27 (0.06 - 1.25) | 0.09 |
| Open Claims | 137,113 | 848,723 | 97.7 | 919.0 | 279 | 5,620 | 0.72 (0.62 - 0.83) | <0.01 | 0.73 (0.59 - 0.89) | <0.01 |
| OEHR | 8,308 | 85,605 | 2.4 | 47.0 | 12 | 230 | 1.62 (0.77 - 3.11) | 0.17 | 1.51 (0.75 - 3.04) | 0.25 |
| VA | 1,283 | 31,903 | 0.9 | 24.0 | <10 | 61 | 0.30 (0.02 - 1.67) | 0.31 | 0.23 (0.02 - 2.38) | 0.22 |
| Meta-analysis | 165,112 | 1,095,945 | 112.7 | 1111.6 | 342 | 6,955 | 0.85 (0.64 - 1.13) | 0.26 | 0.84 (0.60 - 1.17) | 0.31 |
| Meta-analysis (w/o USOC) | 27,999 | 247,222 | 15.0 | 192.6 | 63 | 1,335 | 0.97 (0.63 - 1.50) | 0.89 | 0.93 (0.60 - 1.44) | 0.75 |
| **PS-stratification / on-treatment TAR** | | | | | | | | | | |
| CCAE | 12,840 | 89,313 | 8.3 | 76.8 | 24 | 279 | 0.77 (0.48 - 1.21) | 0.28 | 0.75 (0.47 - 1.19) | 0.23 |
| ODOD | 8,371 | 68,804 | 4.8 | 64.3 | 24 | 770 | 0.62 (0.39 - 0.94) | 0.03 | 0.59 (0.38 - 0.92) | 0.02 |
| MDCR | 977 | 19,375 | 0.6 | 20.7 | 5 | 362 | 0.48 (0.17 - 1.08) | 0.12 | 0.44 (0.17 - 1.13) | 0.09 |
| Open Claims | 152,434 | 876,391 | 105.1 | 951.0 | 292 | 6,193 | 0.69 (0.60 - 0.78) | <0.01 | 0.70 (0.56 - 0.87) | <0.01 |
| OEHR | 9,031 | 94,894 | 2.6 | 52.5 | 12 | 296 | 1.36 (0.70 - 2.42) | 0.32 | 1.26 (0.68 - 2.34) | 0.47 |
| VA | 1,383 | 236,388 | 1.0 | 252.0 | <10 | 880 | 0.33 (0.02 - 1.51) | 0.33 | 0.28 (0.03 - 2.52) | 0.26 |
| Meta-analysis | 182,676 | 1,129,402 | 120.8 | 1144.6 | 352 | 7,538 | 0.71 (0.62 - 0.81) | <0.01 | 0.70 (0.55 - 0.89) | <0.01 |
| Meta-analysis (w/o USOC) | 30,242 | 253,011 | 15.7 | 193.6 | 60 | 1,345 | 0.83 (0.54 - 1.26) | 0.38 | 0.79 (0.51 - 1.21) | 0.30 |
| **PS-matching / on-treatment TAR without escalation** | | | | | | | | | | |
| CCAE | 11,318 | 85,192 | 6.6 | 63.6 | 20 | 234 | 0.95 (0.53 - 1.61) | 0.86 | 0.92 (0.53 - 1.60) | 0.78 |
| ODOD | 7,468 | 61,465 | 3.9 | 49.7 | 20 | 528 | 0.75 (0.41 - 1.28) | 0.32 | 0.71 (0.40 - 1.25) | 0.24 |
| MDCR | 905 | 14,960 | 0.5 | 14.7 | <5 | 227 | 0.35 (0.06 - 1.21) | 0.18 | 0.34 (0.07 - 1.61) | 0.18 |
| Open Claims | 137,113 | 848,723 | 84.9 | 777.9 | 232 | 4,844 | 0.68 (0.57 - 0.80) | <0.01 | 0.68 (0.55 - 0.84) | <0.01 |
| OEHR | 8,308 | 85,605 | 2.1 | 40.9 | 10 | 174 | 2.03 (0.90 - 4.12) | 0.07 | 1.90 (0.88 - 4.07) | 0.10 |
| VA | 1,283 | 31,903 | 0.7 | 21.0 | <10 | 51 | 0.00 (0.00 - 0.76) | 0.19 | 0.00 (0.00 - 15043) | 0.18 |
| Meta-analysis | 165,112 | 1,095,945 | 98.0 | 946.8 | <287 | 6,007 | 0.85 (0.58 - 1.24) | 0.40 | 0.84 (0.55 - 1.26) | 0.40 |
| Meta-analysis (w/o USOC) | 27,999 | 247,222 | 13.1 | 168.9 | <55 | 1,163 | 0.97 (0.57 - 1.64) | 0.90 | 0.92 (0.54 - 1.57) | 0.78 |
| **PS-stratification / on-treatment TAR without escalation** | | | | | | | | | | |
| CCAE | 12,840 | 89,313 | 7.3 | 66.6 | 21 | 253 | 0.76 (0.45 - 1.22) | 0.28 | 0.74 (0.45 - 1.22) | 0.25 |
| ODOD | 8,371 | 68,804 | 4.3 | 57.3 | 21 | 687 | 0.59 (0.36 - 0.93) | 0.03 | 0.55 (0.34 - 0.89) | 0.02 |
| MDCR | 977 | 19,375 | 0.6 | 18.8 | <5 | 331 | 0.34 (0.08 - 0.93) | 0.08 | 0.31 (0.09 - 1.04) | 0.06 |
| Open Claims | 152,434 | 876,391 | 91.8 | 805.3 | 245 | 5,349 | 0.66 (0.57 - 0.76) | <0.01 | 0.66 (0.53 - 0.83) | <0.01 |
| OEHR | 9,031 | 94,894 | 2.2 | 45.6 | 10 | 225 | 1.44 (0.68 - 2.72) | 0.30 | 1.31 (0.66 - 2.61) | 0.45 |
| VA | 1,383 | 236,388 | 0.8 | 224.4 | <10 | 757 | 0.44 (0.02 - 2.02) | 0.47 | 0.35 (0.04 - 3.17) | 0.35 |
| Meta-analysis | 182,676 | 1,129,402 | 105.6 | 974.9 | 297 | 6,514 | 0.69 (0.59 - 0.82) | <0.01 | 0.68 (0.52 - 0.87) | <0.01 |
| Meta-analysis (w/o USOC) | 30,242 | 253,011 | 13.8 | 169.6 | 52 | 1,165 | 0.82 (0.51 - 1.30) | 0.40 | 0.77 (0.48 - 1.25) | 0.30 |

##### 1.5.9 Sudden cardiac death

**Supplementary Table 113:** Exposures, outcomes and risk estimates for Sudden cardiac death in GLP1RA (male) (T) vs SU (male) (C) new-user cohorts under four different propensity score (PS) adjustment / time-at-risk (TAR) designs across data sources. We report total patient exposure time in 1,000 patient-year (KYr) units, uncalibrated and calibrated hazard ratios (HRs) and their 95% confidence intervals (CIs). Greyed out entries did not pass study diagnostics and are included only for completeness.

| | Patients | | Exposure (KYr) | | Outcomes | | Uncalibrated | | Calibrated | |
|---|---|---|---|---|---|---|---|---|---|---|
| | T | C | T | C | T | C | HR (95% CI) | $p$ | HR (95% CI) | $p$ |
| **PS-matching / on-treatment TAR** | | | | | | | | | | |
| CCAE | 11,460 | 86,179 | 7.7 | 74.4 | <5 | 109 | 0.09 (0.00 - 0.42) | 0.03 | 0.08 (0.01 - 0.79) | 0.03 |
| ODOD | 7,587 | 62,755 | 4.5 | 57.4 | 10 | 177 | 0.68 (0.27 - 1.48) | 0.37 | 0.65 (0.28 - 1.52) | 0.32 |
| MDCR | 927 | 15,472 | 0.6 | 17.0 | <5 | 48 | 0.42 (0.02 - 2.58) | 0.48 | 0.41 (0.04 - 4.53) | 0.47 |
| Open Claims | 139,827 | 871,935 | 99.5 | 945.5 | 80 | 1,647 | 0.64 (0.48 - 0.84) | <0.01 | 0.65 (0.48 - 0.89) | <0.01 |
| OEHR | 8,363 | 86,388 | 2.4 | 47.6 | 5 | 84 | 1.14 (0.31 - 3.27) | 0.83 | 1.06 (0.33 - 3.45) | 0.92 |
| VA | 1,309 | 32,260 | 0.9 | 24.3 | 0 | <10 | | | | |
| Meta-analysis | 167,237 | 1,107,257 | 114.1 | 1124.9 | <100 | 2,017 | 0.65 (0.50 - 0.84) | <0.01 | 0.64 (0.47 - 0.87) | <0.01 |
| Meta-analysis (w/o USOC) | 27,410 | 235,322 | 14.6 | 179.4 | <20 | 370 | 0.58 (0.20 - 1.66) | 0.31 | 0.56 (0.20 - 1.59) | 0.28 |
| **PS-stratification / on-treatment TAR** | | | | | | | | | | |
| CCAE | 12,982 | 90,509 | 8.5 | 77.8 | <5 | 115 | 0.33 (0.10 - 0.83) | 0.04 | 0.32 (0.11 - 0.93) | 0.04 |
| ODOD | 8,502 | 70,624 | 4.9 | 66.4 | 10 | 253 | 0.79 (0.37 - 1.50) | 0.50 | 0.75 (0.37 - 1.51) | 0.43 |
| MDCR | 1,004 | 20,293 | 0.7 | 21.8 | <5 | 77 | 0.39 (0.02 - 1.99) | 0.42 | 0.36 (0.04 - 3.51) | 0.38 |
| Open Claims | 155,278 | 901,112 | 106.9 | 979.1 | 87 | 1,869 | 0.65 (0.51 - 0.81) | <0.01 | 0.66 (0.49 - 0.88) | <0.01 |
| OEHR | 9,101 | 95,825 | 2.6 | 53.3 | 5 | 104 | 1.32 (0.43 - 3.20) | 0.59 | 1.21 (0.45 - 3.30) | 0.70 |
| VA | 1,406 | 239,362 | 1.0 | 255.6 | 0 | 134 | | | | |
| Meta-analysis | 185,863 | 1,158,070 | 122.9 | 1176.7 | <107 | 2,341 | 0.66 (0.54 - 0.82) | <0.01 | 0.66 (0.49 - 0.88) | <0.01 |
| Meta-analysis (w/o USOC) | 30,585 | 256,958 | 15.9 | 197.6 | <20 | 472 | 0.72 (0.36 - 1.45) | 0.36 | 0.69 (0.34 - 1.39) | 0.31 |
| **PS-matching / on-treatment TAR without escalation** | | | | | | | | | | |
| CCAE | 11,460 | 86,179 | 6.7 | 64.4 | <5 | 94 | 0.11 (0.01 - 0.54) | 0.05 | 0.11 (0.01 - 1.00) | 0.05 |
| ODOD | 7,587 | 62,755 | 4.0 | 51.0 | 9 | 157 | 0.64 (0.24 - 1.47) | 0.34 | 0.61 (0.24 - 1.52) | 0.29 |
| MDCR | 927 | 15,472 | 0.6 | 15.3 | <5 | 39 | 0.77 (0.04 - 4.98) | 0.83 | 0.76 (0.07 - 8.71) | 0.82 |
| Open Claims | 139,827 | 871,935 | 86.5 | 800.2 | 66 | 1,383 | 0.65 (0.47 - 0.87) | <0.01 | 0.65 (0.47 - 0.91) | 0.01 |
| OEHR | 8,363 | 86,388 | 2.1 | 41.4 | <5 | 59 | 1.37 (0.36 - 4.21) | 0.62 | 1.28 (0.37 - 4.38) | 0.70 |
| VA | 1,309 | 32,260 | 0.7 | 21.3 | 0 | <10 | | | | |
| Meta-analysis | 167,237 | 1,107,257 | 99.2 | 957.0 | <85 | 1,693 | 0.66 (0.50 - 0.87) | <0.01 | 0.64 (0.47 - 0.89) | <0.01 |
| Meta-analysis (w/o USOC) | 27,410 | 235,322 | 12.7 | 156.8 | <19 | 310 | 0.63 (0.22 - 1.81) | 0.39 | 0.60 (0.21 - 1.73) | 0.35 |
| **PS-stratification / on-treatment TAR without escalation** | | | | | | | | | | |
| CCAE | 12,982 | 90,509 | 7.4 | 67.5 | <5 | 100 | 0.19 (0.03 - 0.66) | 0.04 | 0.19 (0.04 - 0.87) | 0.03 |
| ODOD | 8,502 | 70,624 | 4.3 | 59.2 | 9 | 223 | 0.80 (0.36 - 1.59) | 0.56 | 0.75 (0.36 - 1.57) | 0.45 |
| MDCR | 1,004 | 20,293 | 0.6 | 19.8 | <5 | 66 | 0.47 (0.03 - 2.48) | 0.52 | 0.43 (0.04 - 4.24) | 0.47 |
| Open Claims | 155,278 | 901,112 | 93.4 | 829.1 | 72 | 1,585 | 0.64 (0.49 - 0.82) | <0.01 | 0.64 (0.47 - 0.88) | <0.01 |
| OEHR | 9,101 | 95,825 | 2.3 | 46.3 | <5 | 73 | 1.31 (0.37 - 3.59) | 0.65 | 1.18 (0.38 - 3.71) | 0.77 |
| VA | 1,406 | 239,362 | 0.8 | 227.5 | 0 | 102 | | | | |
| Meta-analysis | 185,863 | 1,158,070 | 107.4 | 1002.1 | <91 | 1,981 | 0.65 (0.52 - 0.83) | <0.01 | 0.64 (0.47 - 0.87) | <0.01 |
| Meta-analysis (w/o USOC) | 30,585 | 256,958 | 14.0 | 173.0 | <19 | 396 | 0.68 (0.28 - 1.66) | 0.40 | 0.64 (0.26 - 1.58) | 0.34 |

##### 1.5.10 Abnormal weight gain

**Supplementary Table 114:** Exposures, outcomes and risk estimates for Abnormal weight gain in GLP1RA (male) (T) vs SU (male) (C) new-user cohorts under four different propensity score (PS) adjustment / time-at-risk (TAR) designs across data sources. We report total patient exposure time in 1,000 patient-year (KYr) units, uncalibrated and calibrated hazard ratios (HRs) and their 95% confidence intervals (CIs). Greyed out entries did not pass study diagnostics and are included only for completeness.

| | Patients | | Exposure (KYr) | | Outcomes | | Uncalibrated | | Calibrated | |
|---|---|---|---|---|---|---|---|---|---|---|
| | T | C | T | C | T | C | HR (95% CI) | $p$ | HR (95% CI) | $p$ |
| **PS-matching / on-treatment TAR** | | | | | | | | | | |
| CCAE | 10,954 | 84,463 | 7.4 | 72.7 | 35 | 288 | 0.96 (0.58 - 1.54) | 0.87 | 0.93 (0.57 - 1.52) | 0.78 |
| ODOD | 7,230 | 61,302 | 4.2 | 55.8 | 36 | 305 | 0.80 (0.45 - 1.36) | 0.43 | 0.76 (0.44 - 1.33) | 0.34 |
| MDCR | 913 | 15,392 | 0.6 | 16.8 | 5 | 48 | 2.53 (0.75 - 7.18) | 0.11 | 2.47 (0.80 - 7.65) | 0.12 |
| Open Claims | 133,544 | 853,326 | 94.9 | 924.2 | 203 | 1,746 | 0.96 (0.79 - 1.17) | 0.70 | 0.98 (0.77 - 1.24) | 0.86 |
| OEHR | 7,961 | 84,542 | 2.3 | 46.3 | 18 | 259 | 1.54 (0.79 - 2.83) | 0.19 | 1.44 (0.76 - 2.74) | 0.27 |
| VA | 1,269 | 31,600 | 0.9 | 23.7 | <10 | 57 | 0.94 (0.14 - 3.75) | 0.95 | 0.73 (0.14 - 3.81) | 0.71 |
| Meta-analysis | 159,689 | 1,083,633 | 108.8 | 1099.0 | 292 | 2,598 | 0.98 (0.83 - 1.15) | 0.78 | 0.97 (0.77 - 1.22) | 0.78 |
| Meta-analysis (w/o USOC) | 26,145 | 230,307 | 13.9 | 174.8 | 89 | 852 | 1.02 (0.72 - 1.45) | 0.91 | 0.98 (0.69 - 1.39) | 0.90 |
| **PS-stratification / on-treatment TAR** | | | | | | | | | | |
| CCAE | 12,272 | 88,789 | 8.0 | 76.2 | 42 | 301 | 1.09 (0.75 - 1.56) | 0.64 | 1.06 (0.73 - 1.53) | 0.77 |
| ODOD | 8,011 | 69,036 | 4.6 | 64.6 | 40 | 374 | 1.12 (0.76 - 1.62) | 0.56 | 1.07 (0.73 - 1.56) | 0.74 |
| MDCR | 983 | 20,211 | 0.7 | 21.7 | 5 | 73 | 1.90 (0.61 - 4.80) | 0.22 | 1.76 (0.63 - 4.94) | 0.28 |
| Open Claims | 146,922 | 882,934 | 101.3 | 958.2 | 235 | 1,841 | 1.05 (0.90 - 1.22) | 0.54 | 1.06 (0.84 - 1.35) | 0.61 |
| OEHR | 8,589 | 93,991 | 2.4 | 52.0 | 23 | 286 | 1.60 (0.98 - 2.49) | 0.05 | 1.47 (0.92 - 2.35) | 0.11 |
| VA | 1,354 | 235,922 | 0.9 | 251.3 | <10 | 415 | 1.43 (0.35 - 3.92) | 0.57 | 1.21 (0.36 - 4.07) | 0.76 |
| Meta-analysis | 175,794 | 1,134,750 | 116.3 | 1151.0 | 340 | 2,802 | 1.10 (0.97 - 1.25) | 0.16 | 1.09 (0.86 - 1.37) | 0.49 |
| Meta-analysis (w/o USOC) | 28,872 | 251,816 | 15.1 | 192.8 | 105 | 961 | 1.21 (0.96 - 1.52) | 0.10 | 1.15 (0.90 - 1.48) | 0.32 |
| **PS-matching / on-treatment TAR without escalation** | | | | | | | | | | |
| CCAE | 10,954 | 84,463 | 6.4 | 63.0 | 28 | 243 | 0.95 (0.54 - 1.59) | 0.85 | 0.92 (0.54 - 1.58) | 0.77 |
| ODOD | 7,230 | 61,302 | 3.8 | 49.7 | 29 | 265 | 0.87 (0.47 - 1.53) | 0.65 | 0.82 (0.46 - 1.49) | 0.52 |
| MDCR | 913 | 15,392 | 0.6 | 15.2 | <5 | 44 | 1.85 (0.48 - 5.70) | 0.33 | 1.83 (0.53 - 6.28) | 0.34 |
| Open Claims | 133,544 | 853,326 | 82.5 | 783.1 | 175 | 1,441 | 0.97 (0.78 - 1.20) | 0.78 | 0.97 (0.76 - 1.25) | 0.83 |
| OEHR | 7,961 | 84,542 | 2.0 | 40.4 | 15 | 217 | 1.85 (0.88 - 3.59) | 0.09 | 1.73 (0.86 - 3.48) | 0.13 |
| VA | 1,269 | 31,600 | 0.7 | 20.8 | <10 | 42 | 1.19 (0.06 - 7.08) | 0.89 | 0.89 (0.08 - 9.53) | 0.92 |
| Meta-analysis | 159,689 | 1,083,633 | 94.6 | 936.2 | 247 | 2,166 | 1.00 (0.83 - 1.20) | 1.00 | 0.98 (0.77 - 1.25) | 0.88 |
| Meta-analysis (w/o USOC) | 26,145 | 230,307 | 12.1 | 153.1 | 72 | 725 | 1.10 (0.72 - 1.69) | 0.65 | 1.06 (0.69 - 1.62) | 0.80 |
| **PS-stratification / on-treatment TAR without escalation** | | | | | | | | | | |
| CCAE | 12,272 | 88,789 | 7.0 | 66.1 | 34 | 256 | 1.06 (0.69 - 1.58) | 0.78 | 1.04 (0.69 - 1.56) | 0.87 |
| ODOD | 8,011 | 69,036 | 4.1 | 57.7 | 33 | 326 | 1.12 (0.73 - 1.67) | 0.60 | 1.04 (0.69 - 1.58) | 0.85 |
| MDCR | 983 | 20,211 | 0.6 | 19.7 | <5 | 67 | 1.62 (0.45 - 4.49) | 0.41 | 1.48 (0.47 - 4.67) | 0.50 |
| Open Claims | 146,922 | 882,934 | 88.4 | 812.4 | 206 | 1,524 | 1.07 (0.90 - 1.26) | 0.45 | 1.07 (0.83 - 1.37) | 0.61 |
| OEHR | 8,589 | 93,991 | 2.1 | 45.2 | 19 | 237 | 1.51 (0.88 - 2.46) | 0.11 | 1.37 (0.82 - 2.29) | 0.24 |
| VA | 1,354 | 235,922 | 0.7 | 223.8 | <10 | 339 | 1.41 (0.23 - 4.65) | 0.66 | 1.12 (0.25 - 5.07) | 0.88 |
| Meta-analysis | 175,794 | 1,134,750 | 101.7 | 981.4 | 292 | 2,343 | 1.10 (0.96 - 1.26) | 0.18 | 1.07 (0.84 - 1.37) | 0.56 |
| Meta-analysis (w/o USOC) | 28,872 | 251,816 | 13.2 | 169.1 | 86 | 819 | 1.18 (0.92 - 1.52) | 0.20 | 1.11 (0.83 - 1.48) | 0.49 |

##### 1.5.11 Abnormal weight loss

**Supplementary Table 115:** Exposures, outcomes and risk estimates for Abnormal weight loss in GLP1RA (male) (T) vs SU (male) (C) new-user cohorts under four different propensity score (PS) adjustment / time-at-risk (TAR) designs across data sources. We report total patient exposure time in 1,000 patient-year (KYr) units, uncalibrated and calibrated hazard ratios (HRs) and their 95% confidence intervals (CIs). Greyed out entries did not pass study diagnostics and are included only for completeness.

| | Patients | | Exposure (KYr) | | Outcomes | | Uncalibrated | | Calibrated | |
|---|---|---|---|---|---|---|---|---|---|---|
| | T | C | T | C | T | C | HR (95% CI) | $p$ | HR (95% CI) | $p$ |
| **PS-matching / on-treatment TAR** | | | | | | | | | | |
| CCAE | 11,234 | 84,259 | 7.5 | 72.7 | 55 | 336 | 1.36 (0.93 - 1.95) | 0.11 | 1.32 (0.90 - 1.91) | 0.16 |
| ODOD | 7,392 | 60,690 | 4.3 | 55.3 | 55 | 667 | 1.19 (0.83 - 1.68) | 0.34 | 1.14 (0.79 - 1.62) | 0.50 |
| MDCR | 914 | 14,983 | 0.6 | 16.4 | 5 | 183 | 0.44 (0.13 - 1.15) | 0.14 | 0.43 (0.14 - 1.29) | 0.13 |
| Open Claims | 136,139 | 845,324 | 96.9 | 915.3 | 419 | 4,538 | 1.18 (1.03 - 1.34) | 0.01 | 1.20 (0.99 - 1.45) | 0.07 |
| OEHR | 8,138 | 84,145 | 2.4 | 46.1 | 23 | 464 | 1.03 (0.59 - 1.69) | 0.91 | 0.96 (0.57 - 1.63) | 0.87 |
| VA | 1,273 | 31,466 | 0.9 | 23.7 | <10 | 162 | 1.32 (0.52 - 2.87) | 0.53 | 1.02 (0.43 - 2.40) | 0.91 |
| Meta-analysis | 163,817 | 1,089,401 | 111.7 | 1105.9 | 557 | 6,188 | 1.18 (1.05 - 1.32) | <0.01 | 1.17 (0.96 - 1.42) | 0.13 |
| Meta-analysis (w/o USOC) | 27,678 | 244,077 | 14.8 | 190.5 | 138 | 1,650 | 1.14 (0.87 - 1.50) | 0.34 | 1.09 (0.83 - 1.44) | 0.55 |
| **PS-stratification / on-treatment TAR** | | | | | | | | | | |
| CCAE | 12,729 | 88,509 | 8.3 | 76.1 | 65 | 361 | 1.46 (1.07 - 1.98) | 0.02 | 1.42 (1.04 - 1.94) | 0.03 |
| ODOD | 8,275 | 67,640 | 4.7 | 63.4 | 61 | 834 | 1.15 (0.85 - 1.53) | 0.36 | 1.10 (0.81 - 1.48) | 0.57 |
| MDCR | 989 | 19,461 | 0.7 | 20.9 | 5 | 256 | 0.46 (0.16 - 1.05) | 0.11 | 0.43 (0.17 - 1.10) | 0.08 |
| Open Claims | 151,256 | 871,889 | 104.1 | 946.2 | 462 | 4,959 | 1.16 (1.04 - 1.29) | <0.01 | 1.17 (0.95 - 1.45) | 0.14 |
| OEHR | 8,866 | 92,813 | 2.5 | 51.3 | 25 | 552 | 1.14 (0.73 - 1.72) | 0.55 | 1.05 (0.68 - 1.62) | 0.81 |
| VA | 1,368 | 231,885 | 1.0 | 247.0 | 11 | 1,510 | 1.54 (0.79 - 2.72) | 0.17 | 1.30 (0.70 - 2.43) | 0.41 |
| Meta-analysis | 181,126 | 1,120,851 | 119.6 | 1136.9 | 613 | 6,706 | 1.18 (1.08 - 1.30) | <0.01 | 1.17 (0.94 - 1.45) | 0.16 |
| Meta-analysis (w/o USOC) | 29,870 | 248,962 | 15.5 | 190.7 | 151 | 1,747 | 1.26 (1.04 - 1.52) | 0.02 | 1.20 (0.96 - 1.48) | 0.15 |
| **PS-matching / on-treatment TAR without escalation** | | | | | | | | | | |
| CCAE | 11,234 | 84,259 | 6.5 | 63.0 | 49 | 287 | 1.37 (0.91 - 2.02) | 0.13 | 1.33 (0.89 - 1.98) | 0.18 |
| ODOD | 7,392 | 60,690 | 3.8 | 49.2 | 48 | 581 | 1.08 (0.72 - 1.57) | 0.71 | 1.02 (0.69 - 1.50) | 0.89 |
| MDCR | 914 | 14,983 | 0.6 | 14.8 | 5 | 161 | 0.58 (0.17 - 1.53) | 0.33 | 0.57 (0.19 - 1.73) | 0.32 |
| Open Claims | 136,139 | 845,324 | 84.2 | 775.1 | 354 | 3,853 | 1.12 (0.97 - 1.29) | 0.12 | 1.13 (0.92 - 1.37) | 0.26 |
| OEHR | 8,138 | 84,145 | 2.0 | 40.2 | 19 | 374 | 1.06 (0.58 - 1.81) | 0.85 | 0.99 (0.56 - 1.74) | 0.90 |
| VA | 1,273 | 31,466 | 0.7 | 20.8 | <10 | 139 | 1.44 (0.47 - 3.53) | 0.48 | 1.08 (0.39 - 2.95) | 0.88 |
| Meta-analysis | 163,817 | 1,089,401 | 97.2 | 942.3 | 475 | 5,256 | 1.13 (1.00 - 1.27) | 0.06 | 1.11 (0.90 - 1.36) | 0.33 |
| Meta-analysis (w/o USOC) | 27,678 | 244,077 | 13.0 | 167.2 | 121 | 1,403 | 1.14 (0.89 - 1.45) | 0.30 | 1.09 (0.85 - 1.40) | 0.52 |
| **PS-stratification / on-treatment TAR without escalation** | | | | | | | | | | |
| CCAE | 12,729 | 88,509 | 7.2 | 66.1 | 59 | 311 | 1.52 (1.08 - 2.10) | 0.01 | 1.48 (1.06 - 2.06) | 0.02 |
| ODOD | 8,275 | 67,640 | 4.2 | 56.5 | 53 | 733 | 1.14 (0.82 - 1.56) | 0.43 | 1.06 (0.77 - 1.47) | 0.73 |
| MDCR | 989 | 19,461 | 0.6 | 19.1 | 5 | 228 | 0.48 (0.16 - 1.11) | 0.13 | 0.44 (0.17 - 1.14) | 0.09 |
| Open Claims | 151,256 | 871,889 | 91.0 | 801.6 | 394 | 4,225 | 1.15 (1.02 - 1.29) | 0.02 | 1.15 (0.92 - 1.43) | 0.23 |
| OEHR | 8,866 | 92,813 | 2.2 | 44.6 | 20 | 448 | 1.10 (0.66 - 1.74) | 0.69 | 1.00 (0.62 - 1.63) | 0.90 |
| VA | 1,368 | 231,885 | 0.8 | 220.0 | <10 | 1,312 | 1.30 (0.55 - 2.60) | 0.50 | 1.04 (0.48 - 2.26) | 0.89 |
| Meta-analysis | 181,126 | 1,120,851 | 104.6 | 968.7 | 526 | 5,717 | 1.17 (1.06 - 1.30) | <0.01 | 1.15 (0.92 - 1.43) | 0.22 |
| Meta-analysis (w/o USOC) | 29,870 | 248,962 | 13.6 | 167.1 | 132 | 1,492 | 1.27 (1.03 - 1.56) | 0.03 | 1.20 (0.93 - 1.53) | 0.18 |

##### 1.5.12 Acute pancreatitis

**Supplementary Table 116:** Exposures, outcomes and risk estimates for Acute pancreatitis in GLP1RA (male) (T) vs SU (male) (C) new-user cohorts under four different propensity score (PS) adjustment / time-at-risk (TAR) designs across data sources. We report total patient exposure time in 1,000 patient-year (KYr) units, uncalibrated and calibrated hazard ratios (HRs) and their 95% confidence intervals (CIs). Greyed out entries did not pass study diagnostics and are included only for completeness.

| | Patients | | Exposure (KYr) | | Outcomes | | Uncalibrated | | Calibrated | |
|---|---|---|---|---|---|---|---|---|---|---|
| | T | C | T | C | T | C | HR (95% CI) | $p$ | HR (95% CI) | $p$ |
| **PS-matching / on-treatment TAR** | | | | | | | | | | |
| CCAE | 11,384 | 85,387 | 7.7 | 73.6 | 14 | 200 | 0.38 (0.16 - 0.77) | 0.01 | 0.37 (0.17 - 0.80) | 0.01 |
| ODOD | 7,559 | 62,479 | 4.5 | 57.1 | 9 | 193 | 0.81 (0.33 - 1.74) | 0.63 | 0.78 (0.34 - 1.79) | 0.55 |
| MDCR | 927 | 15,310 | 0.6 | 16.7 | 0 | 52 | | | | |
| Open Claims | 138,619 | 861,514 | 98.7 | 934.2 | 139 | 2,056 | 0.74 (0.60 - 0.92) | <0.01 | 0.75 (0.58 - 0.97) | 0.03 |
| OEHR | 8,327 | 85,885 | 2.4 | 47.3 | 7 | 135 | 0.89 (0.30 - 2.16) | 0.82 | 0.83 (0.31 - 2.24) | 0.72 |
| VA | 1,297 | 32,059 | 0.9 | 24.1 | 0 | 44 | | | | |
| Meta-analysis | 165,889 | 1,095,265 | 113.3 | 1112.2 | 169 | 2,584 | 0.72 (0.59 - 0.88) | <0.01 | 0.71 (0.55 - 0.92) | <0.01 |
| Meta-analysis (w/o USOC) | 27,270 | 233,751 | 14.5 | 178.0 | 30 | 528 | 0.62 (0.35 - 1.08) | 0.09 | 0.59 (0.34 - 1.03) | 0.07 |
| **PS-stratification / on-treatment TAR** | | | | | | | | | | |
| CCAE | 12,906 | 89,619 | 8.4 | 77.0 | 18 | 213 | 0.63 (0.36 - 1.05) | 0.09 | 0.61 (0.35 - 1.04) | 0.07 |
| ODOD | 8,472 | 69,985 | 4.9 | 65.7 | 9 | 238 | 0.70 (0.32 - 1.36) | 0.33 | 0.67 (0.32 - 1.38) | 0.28 |
| MDCR | 1,003 | 20,114 | 0.7 | 21.6 | 0 | 77 | | | | |
| Open Claims | 154,048 | 889,363 | 106.2 | 966.4 | 149 | 2,182 | 0.75 (0.62 - 0.89) | <0.01 | 0.76 (0.59 - 0.98) | 0.04 |
| OEHR | 9,062 | 95,290 | 2.6 | 52.9 | 7 | 164 | 0.96 (0.39 - 1.99) | 0.91 | 0.88 (0.39 - 2.00) | 0.77 |
| VA | 1,396 | 237,077 | 1.0 | 252.9 | 0 | 521 | | | | |
| Meta-analysis | 184,488 | 1,144,257 | 122.0 | 1162.1 | 183 | 2,797 | 0.74 (0.63 - 0.87) | <0.01 | 0.73 (0.57 - 0.95) | 0.02 |
| Meta-analysis (w/o USOC) | 30,440 | 254,894 | 15.9 | 195.7 | 34 | 615 | 0.71 (0.48 - 1.04) | 0.08 | 0.67 (0.45 - 1.00) | 0.06 |
| **PS-matching / on-treatment TAR without escalation** | | | | | | | | | | |
| CCAE | 11,384 | 85,387 | 6.6 | 63.8 | 9 | 168 | 0.33 (0.12 - 0.74) | 0.02 | 0.32 (0.13 - 0.79) | 0.01 |
| ODOD | 7,559 | 62,479 | 4.0 | 50.7 | 8 | 171 | 0.74 (0.27 - 1.66) | 0.51 | 0.70 (0.28 - 1.72) | 0.44 |
| MDCR | 927 | 15,310 | 0.6 | 15.1 | 0 | 45 | | | | |
| Open Claims | 138,619 | 861,514 | 85.8 | 791.2 | 113 | 1,699 | 0.75 (0.59 - 0.95) | 0.02 | 0.75 (0.57 - 0.99) | 0.05 |
| OEHR | 8,327 | 85,885 | 2.1 | 41.2 | 6 | 99 | 1.38 (0.45 - 3.44) | 0.53 | 1.29 (0.47 - 3.57) | 0.62 |
| VA | 1,297 | 32,059 | 0.7 | 21.1 | 0 | 37 | | | | |
| Meta-analysis | 165,889 | 1,095,265 | 98.5 | 946.9 | 136 | 2,137 | 0.73 (0.59 - 0.91) | <0.01 | 0.72 (0.55 - 0.95) | 0.02 |
| Meta-analysis (w/o USOC) | 27,270 | 233,751 | 12.7 | 155.7 | 23 | 438 | 0.68 (0.31 - 1.51) | 0.34 | 0.65 (0.29 - 1.45) | 0.29 |
| **PS-stratification / on-treatment TAR without escalation** | | | | | | | | | | |
| CCAE | 12,906 | 89,619 | 7.4 | 66.9 | 13 | 181 | 0.54 (0.28 - 0.97) | 0.05 | 0.53 (0.28 - 0.99) | 0.05 |
| ODOD | 8,472 | 69,985 | 4.3 | 58.6 | 8 | 208 | 0.67 (0.29 - 1.36) | 0.31 | 0.62 (0.29 - 1.36) | 0.24 |
| MDCR | 1,003 | 20,114 | 0.6 | 19.6 | 0 | 67 | | | | |
| Open Claims | 154,048 | 889,363 | 92.8 | 819.0 | 123 | 1,807 | 0.73 (0.60 - 0.89) | <0.01 | 0.73 (0.56 - 0.96) | 0.03 |
| OEHR | 9,062 | 95,290 | 2.3 | 46.0 | 6 | 120 | 1.19 (0.45 - 2.65) | 0.70 | 1.08 (0.44 - 2.64) | 0.86 |
| VA | 1,396 | 237,077 | 0.8 | 225.3 | 0 | 431 | | | | |
| Meta-analysis | 184,488 | 1,144,257 | 106.7 | 990.4 | 150 | 2,316 | 0.73 (0.61 - 0.87) | <0.01 | 0.71 (0.54 - 0.93) | 0.01 |
| Meta-analysis (w/o USOC) | 30,440 | 254,894 | 13.9 | 171.5 | 27 | 509 | 0.69 (0.45 - 1.07) | 0.10 | 0.65 (0.41 - 1.03) | 0.07 |

##### 1.5.13 Bone fracture

**Supplementary Table 117:** Exposures, outcomes and risk estimates for Bone fracture in GLP1RA (male) (T) vs SU (male) (C) new-user cohorts under four different propensity score (PS) adjustment / time-at-risk (TAR) designs across data sources. We report total patient exposure time in 1,000 patient-year (KYr) units, uncalibrated and calibrated hazard ratios (HRs) and their 95% confidence intervals (CIs). Greyed out entries did not pass study diagnostics and are included only for completeness.

| | Patients | | Exposure (KYr) | | Outcomes | | Uncalibrated | | Calibrated | |
|---|---|---|---|---|---|---|---|---|---|---|
| | T | C | T | C | T | C | HR (95% CI) | $p$ | HR (95% CI) | $p$ |
| **PS-matching / on-treatment TAR** | | | | | | | | | | |
| CCAE | 10,570 | 80,684 | 7.0 | 68.6 | 114 | 1,068 | 0.93 (0.71 - 1.21) | 0.60 | 0.90 (0.69 - 1.18) | 0.48 |
| ODOD | 7,048 | 58,762 | 4.1 | 52.8 | 101 | 1,109 | 1.20 (0.90 - 1.57) | 0.20 | 1.14 (0.86 - 1.51) | 0.37 |
| MDCR | 850 | 14,248 | 0.6 | 15.3 | 11 | 380 | 0.80 (0.35 - 1.61) | 0.57 | 0.79 (0.37 - 1.69) | 0.54 |
| Open Claims | 122,443 | 783,604 | 86.8 | 841.6 | 932 | 11,844 | 0.89 (0.82 - 0.97) | 0.01 | 0.91 (0.77 - 1.07) | 0.27 |
| OEHR | 7,836 | 81,327 | 2.3 | 44.0 | 29 | 729 | 0.90 (0.56 - 1.37) | 0.63 | 0.84 (0.54 - 1.31) | 0.45 |
| VA | 1,173 | 29,601 | 0.8 | 22.2 | <10 | 248 | 1.42 (0.60 - 2.94) | 0.39 | 1.10 (0.49 - 2.44) | 0.82 |
| Meta-analysis | 148,747 | 1,018,625 | 100.7 | 1022.4 | 1,187 | 15,130 | 0.94 (0.83 - 1.06) | 0.31 | 0.93 (0.76 - 1.14) | 0.49 |
| Meta-analysis (w/o USOC) | 26,304 | 235,021 | 13.9 | 180.8 | 255 | 3,286 | 1.01 (0.85 - 1.20) | 0.89 | 0.97 (0.81 - 1.16) | 0.76 |
| **PS-stratification / on-treatment TAR** | | | | | | | | | | |
| CCAE | 11,928 | 84,627 | 7.7 | 71.8 | 126 | 1,110 | 1.01 (0.82 - 1.23) | 0.96 | 0.98 (0.79 - 1.20) | 0.82 |
| ODOD | 7,882 | 66,029 | 4.5 | 61.0 | 111 | 1,355 | 1.14 (0.91 - 1.42) | 0.23 | 1.09 (0.87 - 1.37) | 0.47 |
| MDCR | 916 | 18,429 | 0.6 | 19.4 | 11 | 515 | 0.68 (0.34 - 1.23) | 0.24 | 0.64 (0.34 - 1.20) | 0.17 |
| Open Claims | 135,593 | 811,681 | 93.0 | 873.6 | 1,016 | 12,691 | 0.89 (0.83 - 0.95) | <0.01 | 0.90 (0.74 - 1.09) | 0.30 |
| OEHR | 8,521 | 90,116 | 2.4 | 49.2 | 30 | 894 | 0.85 (0.57 - 1.23) | 0.42 | 0.79 (0.53 - 1.16) | 0.25 |
| VA | 1,265 | 219,434 | 0.9 | 232.5 | <10 | 2,502 | 1.05 (0.50 - 1.94) | 0.88 | 0.89 (0.45 - 1.75) | 0.74 |
| Meta-analysis | 163,924 | 1,052,453 | 107.6 | 1055.5 | 1,283 | 16,050 | 0.96 (0.84 - 1.09) | 0.54 | 0.95 (0.75 - 1.20) | 0.67 |
| Meta-analysis (w/o USOC) | 28,331 | 240,772 | 14.5 | 181.9 | 267 | 3,359 | 1.04 (0.90 - 1.19) | 0.61 | 0.99 (0.83 - 1.17) | 0.87 |
| **PS-matching / on-treatment TAR without escalation** | | | | | | | | | | |
| CCAE | 10,570 | 80,684 | 6.1 | 59.6 | 91 | 927 | 0.96 (0.71 - 1.26) | 0.76 | 0.93 (0.69 - 1.24) | 0.62 |
| ODOD | 7,048 | 58,762 | 3.6 | 47.1 | 90 | 991 | 1.10 (0.81 - 1.47) | 0.54 | 1.04 (0.77 - 1.40) | 0.81 |
| MDCR | 850 | 14,248 | 0.5 | 13.8 | 10 | 348 | 0.88 (0.36 - 1.83) | 0.76 | 0.87 (0.38 - 1.96) | 0.73 |
| Open Claims | 122,443 | 783,604 | 75.4 | 713.9 | 792 | 9,958 | 0.88 (0.80 - 0.97) | <0.01 | 0.88 (0.75 - 1.04) | 0.17 |
| OEHR | 7,836 | 81,327 | 2.0 | 38.5 | 23 | 611 | 0.88 (0.52 - 1.40) | 0.60 | 0.82 (0.50 - 1.35) | 0.44 |
| VA | 1,173 | 29,601 | 0.7 | 19.5 | <10 | 220 | 1.56 (0.57 - 3.55) | 0.34 | 1.17 (0.47 - 2.93) | 0.74 |
| Meta-analysis | 148,747 | 1,018,625 | 87.6 | 872.9 | 1,006 | 12,835 | 0.90 (0.83 - 0.98) | 0.02 | 0.89 (0.74 - 1.07) | 0.21 |
| Meta-analysis (w/o USOC) | 26,304 | 235,021 | 12.2 | 159.0 | 214 | 2,877 | 0.99 (0.82 - 1.19) | 0.94 | 0.95 (0.79 - 1.15) | 0.64 |
| **PS-stratification / on-treatment TAR without escalation** | | | | | | | | | | |
| CCAE | 11,928 | 84,627 | 6.7 | 62.4 | 102 | 965 | 0.92 (0.73 - 1.15) | 0.48 | 0.90 (0.71 - 1.13) | 0.39 |
| ODOD | 7,882 | 66,029 | 4.0 | 54.5 | 99 | 1,222 | 1.14 (0.90 - 1.44) | 0.27 | 1.06 (0.84 - 1.35) | 0.64 |
| MDCR | 916 | 18,429 | 0.5 | 17.7 | 10 | 471 | 0.68 (0.33 - 1.26) | 0.26 | 0.62 (0.32 - 1.22) | 0.17 |
| Open Claims | 135,593 | 811,681 | 81.3 | 741.6 | 869 | 10,693 | 0.88 (0.82 - 0.95) | <0.01 | 0.88 (0.72 - 1.08) | 0.23 |
| OEHR | 8,521 | 90,116 | 2.1 | 42.9 | 24 | 745 | 0.80 (0.51 - 1.21) | 0.32 | 0.73 (0.47 - 1.13) | 0.17 |
| VA | 1,265 | 219,434 | 0.7 | 207.6 | <10 | 2,203 | 0.90 (0.35 - 1.87) | 0.81 | 0.72 (0.31 - 1.65) | 0.44 |
| Meta-analysis | 163,924 | 1,052,453 | 94.0 | 901.4 | 1,094 | 13,625 | 0.93 (0.82 - 1.05) | 0.23 | 0.91 (0.72 - 1.14) | 0.41 |
| Meta-analysis (w/o USOC) | 28,331 | 240,772 | 12.8 | 159.8 | 225 | 2,932 | 0.98 (0.82 - 1.18) | 0.87 | 0.93 (0.74 - 1.17) | 0.56 |

##### 1.5.14 Diabetic ketoacidosis

**Supplementary Table 118:** Exposures, outcomes and risk estimates for Diabetic ketoacidosis in GLP1RA (male) (T) vs SU (male) (C) new-user cohorts under four different propensity score (PS) adjustment / time-at-risk (TAR) designs across data sources. We report total patient exposure time in 1,000 patient-year (KYr) units, uncalibrated and calibrated hazard ratios (HRs) and their 95% confidence intervals (CIs). Greyed out entries did not pass study diagnostics and are included only for completeness.

| | Patients | | Exposure (KYr) | | Outcomes | | Uncalibrated | | Calibrated | |
|---|---|---|---|---|---|---|---|---|---|---|
| | T | C | T | C | T | C | HR (95% CI) | $p$ | HR (95% CI) | $p$ |
| **PS-matching / on-treatment TAR** | | | | | | | | | | |
| CCAE | 11,456 | 86,109 | 7.7 | 74.3 | 8 | 52 | 1.04 (0.39 - 2.48) | 0.94 | 1.01 (0.40 - 2.56) | 0.96 |
| ODOD | 7,588 | 62,916 | 4.5 | 57.6 | <5 | 71 | 0.47 (0.07 - 1.71) | 0.35 | 0.45 (0.09 - 2.18) | 0.32 |
| MDCR | 932 | 15,496 | 0.6 | 17.0 | 0 | 16 | | | | |
| Open Claims | 139,827 | 871,590 | 99.6 | 946.0 | 67 | 727 | 0.96 (0.70 - 1.29) | 0.79 | 0.97 (0.70 - 1.36) | 0.88 |
| OEHR | 8,366 | 86,386 | 2.4 | 47.6 | <5 | 56 | 1.59 (0.34 - 5.22) | 0.50 | 1.48 (0.38 - 5.79) | 0.57 |
| VA | 1,312 | 32,262 | 0.9 | 24.3 | <10 | 19 | 1.73 (0.08 - 12.1) | 0.67 | 1.34 (0.11 - 16.2) | 0.82 |
| Meta-analysis | 167,237 | 1,107,001 | 114.2 | 1125.5 | <85 | 906 | 0.96 (0.73 - 1.28) | 0.80 | 0.96 (0.69 - 1.32) | 0.79 |
| Meta-analysis (w/o USOC) | 27,410 | 235,411 | 14.6 | 179.6 | <18 | 179 | 1.00 (0.50 - 1.99) | 0.99 | 0.95 (0.48 - 1.90) | 0.89 |
| **PS-stratification / on-treatment TAR** | | | | | | | | | | |
| CCAE | 12,980 | 90,423 | 8.5 | 77.8 | 8 | 55 | 0.96 (0.38 - 2.14) | 0.92 | 0.93 (0.39 - 2.19) | 0.86 |
| ODOD | 8,503 | 70,638 | 4.9 | 66.5 | <5 | 87 | 0.48 (0.11 - 1.44) | 0.26 | 0.46 (0.13 - 1.65) | 0.24 |
| MDCR | 1,007 | 20,353 | 0.7 | 21.9 | 0 | 23 | | | | |
| Open Claims | 155,271 | 900,860 | 107.0 | 979.7 | 70 | 788 | 0.89 (0.67 - 1.16) | 0.40 | 0.90 (0.65 - 1.25) | 0.54 |
| OEHR | 9,102 | 95,832 | 2.6 | 53.3 | <5 | 62 | 0.98 (0.23 - 2.84) | 0.98 | 0.90 (0.26 - 3.18) | 0.88 |
| VA | 1,411 | 239,435 | 1.0 | 255.7 | <10 | 127 | 1.15 (0.06 - 5.58) | 0.91 | 0.97 (0.10 - 9.08) | 0.97 |
| Meta-analysis | 185,856 | 1,157,753 | 122.9 | 1177.4 | <88 | 992 | 0.88 (0.68 - 1.13) | 0.30 | 0.87 (0.63 - 1.19) | 0.38 |
| Meta-analysis (w/o USOC) | 30,585 | 256,893 | 15.9 | 197.6 | <18 | 204 | 0.82 (0.44 - 1.52) | 0.53 | 0.78 (0.42 - 1.46) | 0.45 |
| **PS-matching / on-treatment TAR without escalation** | | | | | | | | | | |
| CCAE | 11,456 | 86,109 | 6.7 | 64.4 | 7 | 40 | 1.17 (0.37 - 3.14) | 0.77 | 1.14 (0.39 - 3.33) | 0.82 |
| ODOD | 7,588 | 62,916 | 4.0 | 51.2 | <5 | 52 | 0.35 (0.02 - 1.79) | 0.36 | 0.33 (0.03 - 3.19) | 0.34 |
| MDCR | 932 | 15,496 | 0.6 | 15.3 | 0 | 12 | | | | |
| Open Claims | 139,827 | 871,590 | 86.5 | 800.8 | 56 | 557 | 0.93 (0.66 - 1.28) | 0.66 | 0.93 (0.65 - 1.34) | 0.70 |
| OEHR | 8,366 | 86,386 | 2.1 | 41.4 | <5 | 30 | 1.80 (0.38 - 6.18) | 0.41 | 1.68 (0.42 - 6.78) | 0.47 |
| VA | 1,312 | 32,262 | 0.7 | 21.3 | <10 | 12 | 1.68 (0.08 - 13.0) | 0.69 | 1.26 (0.10 - 16.1) | 0.86 |
| Meta-analysis | 158,871 | 1,020,615 | 97.2 | 916.4 | <68 | 649 | 0.93 (0.68 - 1.27) | 0.64 | 0.91 (0.64 - 1.30) | 0.62 |
| Meta-analysis (w/o USOC) | 19,044 | 149,025 | 10.6 | 115.6 | <12 | 92 | 0.94 (0.35 - 2.48) | 0.90 | 0.90 (0.34 - 2.38) | 0.83 |
| **PS-stratification / on-treatment TAR without escalation** | | | | | | | | | | |
| CCAE | 12,980 | 90,423 | 7.4 | 67.5 | 7 | 43 | 1.02 (0.38 - 2.47) | 0.96 | 1.00 (0.39 - 2.55) | 0.97 |
| ODOD | 8,503 | 70,638 | 4.3 | 59.2 | <5 | 65 | 0.43 (0.07 - 1.58) | 0.30 | 0.40 (0.08 - 1.96) | 0.26 |
| MDCR | 1,007 | 20,353 | 0.6 | 19.9 | 0 | 19 | | | | |
| Open Claims | 155,271 | 900,860 | 93.5 | 829.9 | 58 | 606 | 0.89 (0.65 - 1.19) | 0.44 | 0.89 (0.62 - 1.26) | 0.52 |
| OEHR | 9,102 | 95,832 | 2.3 | 46.3 | <5 | 35 | 1.74 (0.39 - 5.42) | 0.41 | 1.58 (0.42 - 5.85) | 0.50 |
| VA | 1,411 | 239,435 | 0.8 | 227.6 | <10 | 91 | 1.80 (0.10 - 9.23) | 0.61 | 1.44 (0.15 - 13.9) | 0.75 |
| Meta-analysis | 176,754 | 1,061,921 | 105.2 | 956.7 | <70 | 714 | 0.88 (0.66 - 1.16) | 0.37 | 0.86 (0.61 - 1.21) | 0.39 |
| Meta-analysis (w/o USOC) | 21,483 | 161,061 | 11.7 | 126.8 | <12 | 108 | 0.82 (0.36 - 1.83) | 0.62 | 0.77 (0.34 - 1.75) | 0.54 |

##### 1.5.15 Diarrhea

**Supplementary Table 119:** Exposures, outcomes and risk estimates for Diarrhea in GLP1RA (male) (T) vs SU (male) (C) new-user cohorts under four different propensity score (PS) adjustment / time-at-risk (TAR) designs across data sources. We report total patient exposure time in 1,000 patient-year (KYr) units, uncalibrated and calibrated hazard ratios (HRs) and their 95% confidence intervals (CIs). Greyed out entries did not pass study diagnostics and are included only for completeness.

| | Patients | | Exposure (KYr) | | Outcomes | | Uncalibrated | | Calibrated | |
|---|---|---|---|---|---|---|---|---|---|---|
| | T | C | T | C | T | C | HR (95% CI) | $p$ | HR (95% CI) | $p$ |
| **PS-matching / on-treatment TAR** | | | | | | | | | | |
| CCAE | 10,329 | 79,209 | 6.8 | 67.3 | 243 | 1,390 | 1.32 (1.10 - 1.59) | <0.01 | 1.29 (1.06 - 1.56) | 0.02 |
| ODOD | 6,819 | 56,936 | 3.9 | 50.7 | 166 | 1,603 | 1.33 (1.08 - 1.64) | <0.01 | 1.27 (1.03 - 1.58) | 0.04 |
| MDCR | 836 | 13,964 | 0.6 | 14.9 | 18 | 393 | 1.23 (0.65 - 2.18) | 0.50 | 1.21 (0.66 - 2.21) | 0.56 |
| Open Claims | 124,238 | 792,429 | 87.6 | 848.8 | 1,593 | 12,255 | 1.34 (1.25 - 1.43) | <0.01 | 1.36 (1.16 - 1.59) | <0.01 |
| OEHR | 7,621 | 79,157 | 2.2 | 42.0 | 91 | 1,318 | 1.43 (1.09 - 1.86) | <0.01 | 1.34 (1.03 - 1.75) | 0.04 |
| VA | 1,109 | 27,701 | 0.8 | 20.7 | 22 | 389 | 1.30 (0.75 - 2.15) | 0.33 | 1.01 (0.59 - 1.71) | 0.86 |
| Meta-analysis | 149,843 | 1,021,695 | 101.0 | 1023.7 | 2,111 | 16,959 | 1.34 (1.26 - 1.42) | <0.01 | 1.33 (1.12 - 1.58) | <0.01 |
| Meta-analysis (w/o USOC) | 25,605 | 229,266 | 13.4 | 174.9 | 518 | 4,704 | 1.35 (1.19 - 1.52) | <0.01 | 1.29 (1.13 - 1.47) | <0.01 |
| **PS-stratification / on-treatment TAR** | | | | | | | | | | |
| CCAE | 11,634 | 83,498 | 7.4 | 70.7 | 278 | 1,463 | 1.34 (1.16 - 1.56) | <0.01 | 1.30 (1.12 - 1.52) | <0.01 |
| ODOD | 7,599 | 63,534 | 4.3 | 58.0 | 191 | 1,910 | 1.30 (1.09 - 1.54) | <0.01 | 1.24 (1.04 - 1.48) | 0.03 |
| MDCR | 901 | 18,340 | 0.6 | 19.2 | 18 | 543 | 1.00 (0.59 - 1.61) | 0.99 | 0.93 (0.56 - 1.54) | 0.78 |
| Open Claims | 137,360 | 819,429 | 93.7 | 879.3 | 1,776 | 13,027 | 1.27 (1.20 - 1.34) | <0.01 | 1.28 (1.06 - 1.55) | 0.01 |
| OEHR | 8,239 | 88,182 | 2.3 | 47.1 | 99 | 1,557 | 1.34 (1.07 - 1.66) | 0.01 | 1.23 (0.99 - 1.54) | 0.09 |
| VA | 1,195 | 214,202 | 0.8 | 224.2 | 23 | 4,038 | 1.50 (0.95 - 2.23) | 0.06 | 1.26 (0.83 - 1.94) | 0.30 |
| Meta-analysis | 164,832 | 1,054,643 | 107.7 | 1055.2 | 2,344 | 17,957 | 1.28 (1.22 - 1.35) | <0.01 | 1.27 (1.04 - 1.55) | 0.02 |
| Meta-analysis (w/o USOC) | 27,472 | 235,214 | 14.0 | 175.8 | 568 | 4,930 | 1.33 (1.20 - 1.47) | <0.01 | 1.26 (1.10 - 1.45) | 0.01 |
| **PS-matching / on-treatment TAR without escalation** | | | | | | | | | | |
| CCAE | 10,329 | 79,209 | 5.9 | 58.5 | 218 | 1,208 | 1.38 (1.13 - 1.68) | <0.01 | 1.34 (1.09 - 1.63) | <0.01 |
| ODOD | 6,819 | 56,936 | 3.5 | 45.3 | 145 | 1,424 | 1.28 (1.02 - 1.61) | 0.03 | 1.21 (0.96 - 1.53) | 0.12 |
| MDCR | 836 | 13,964 | 0.5 | 13.5 | 16 | 356 | 1.38 (0.71 - 2.47) | 0.32 | 1.36 (0.73 - 2.54) | 0.35 |
| Open Claims | 124,238 | 792,429 | 76.3 | 721.7 | 1,375 | 10,270 | 1.33 (1.24 - 1.44) | <0.01 | 1.34 (1.14 - 1.57) | <0.01 |
| OEHR | 7,621 | 79,157 | 1.9 | 36.9 | 78 | 1,057 | 1.53 (1.14 - 2.03) | <0.01 | 1.43 (1.07 - 1.91) | 0.02 |
| VA | 1,109 | 27,701 | 0.6 | 18.2 | 19 | 337 | 1.26 (0.68 - 2.21) | 0.44 | 0.95 (0.52 - 1.71) | 0.82 |
| Meta-analysis | 149,843 | 1,021,695 | 88.1 | 876.1 | 1,832 | 14,315 | 1.35 (1.26 - 1.44) | <0.01 | 1.32 (1.11 - 1.58) | <0.01 |
| Meta-analysis (w/o USOC) | 25,605 | 229,266 | 11.8 | 154.3 | 457 | 4,045 | 1.38 (1.21 - 1.57) | <0.01 | 1.32 (1.15 - 1.51) | <0.01 |
| **PS-stratification / on-treatment TAR without escalation** | | | | | | | | | | |
| CCAE | 11,634 | 83,498 | 6.5 | 61.7 | 250 | 1,273 | 1.38 (1.18 - 1.61) | <0.01 | 1.35 (1.15 - 1.58) | <0.01 |
| ODOD | 7,599 | 63,534 | 3.8 | 52.0 | 165 | 1,712 | 1.29 (1.07 - 1.55) | <0.01 | 1.20 (0.99 - 1.45) | 0.08 |
| MDCR | 901 | 18,340 | 0.5 | 17.5 | 16 | 498 | 1.04 (0.59 - 1.72) | 0.89 | 0.95 (0.55 - 1.63) | 0.83 |
| Open Claims | 137,360 | 819,429 | 82.1 | 748.1 | 1,549 | 10,955 | 1.30 (1.23 - 1.39) | <0.01 | 1.30 (1.07 - 1.58) | <0.01 |
| OEHR | 8,239 | 88,182 | 2.0 | 41.4 | 85 | 1,243 | 1.40 (1.09 - 1.77) | <0.01 | 1.26 (0.99 - 1.61) | 0.08 |
| VA | 1,195 | 214,202 | 0.6 | 200.8 | 20 | 3,535 | 1.66 (1.02 - 2.54) | 0.03 | 1.32 (0.84 - 2.09) | 0.25 |
| Meta-analysis | 164,832 | 1,054,643 | 94.4 | 903.1 | 2,049 | 15,183 | 1.32 (1.25 - 1.39) | <0.01 | 1.29 (1.05 - 1.58) | 0.02 |
| Meta-analysis (w/o USOC) | 27,472 | 235,214 | 12.3 | 155.0 | 500 | 4,228 | 1.35 (1.21 - 1.50) | <0.01 | 1.27 (1.07 - 1.51) | 0.01 |

##### 1.5.16 Genitourinary infection

**Supplementary Table 120:** Exposures, outcomes and risk estimates for Genitourinary infection in GLP1RA (male) (T) vs SU (male) (C) new-user cohorts under four different propensity score (PS) adjustment / time-at-risk (TAR) designs across data sources. We report total patient exposure time in 1,000 patient-year (KYr) units, uncalibrated and calibrated hazard ratios (HRs) and their 95% confidence intervals (CIs). Greyed out entries did not pass study diagnostics and are included only for completeness.

| | Patients | | Exposure (KYr) | | Outcomes | | Uncalibrated | | Calibrated | |
|---|---|---|---|---|---|---|---|---|---|---|
| | T | C | T | C | T | C | HR (95% CI) | $p$ | HR (95% CI) | $p$ |
| **PS-matching / on-treatment TAR** | | | | | | | | | | |
| CCAE | 10,455 | 78,994 | 6.9 | 67.0 | 144 | 1,346 | 0.96 (0.76 - 1.20) | 0.74 | 0.93 (0.74 - 1.18) | 0.59 |
| ODOD | 6,788 | 54,840 | 3.9 | 48.3 | 108 | 1,836 | 0.91 (0.71 - 1.16) | 0.45 | 0.87 (0.68 - 1.11) | 0.28 |
| MDCR | 814 | 13,541 | 0.5 | 14.4 | 28 | 555 | 1.36 (0.85 - 2.10) | 0.18 | 1.33 (0.85 - 2.10) | 0.24 |
| Open Claims | 124,593 | 776,506 | 88.5 | 828.9 | 1,171 | 15,299 | 0.91 (0.84 - 0.98) | 0.02 | 0.93 (0.79 - 1.08) | 0.36 |
| OEHR | 7,896 | 81,313 | 2.3 | 43.7 | 47 | 1,007 | 1.09 (0.74 - 1.55) | 0.66 | 1.02 (0.70 - 1.47) | 0.86 |
| VA | 1,170 | 28,977 | 0.9 | 21.6 | 15 | 325 | 1.22 (0.58 - 2.31) | 0.57 | 0.95 (0.47 - 1.90) | 0.86 |
| Meta-analysis | 150,546 | 1,005,194 | 102.1 | 1002.3 | 1,498 | 20,043 | 0.93 (0.87 - 0.99) | 0.03 | 0.92 (0.77 - 1.10) | 0.36 |
| Meta-analysis (w/o USOC) | 25,953 | 228,688 | 13.6 | 173.4 | 327 | 4,744 | 1.00 (0.86 - 1.15) | 0.96 | 0.95 (0.82 - 1.11) | 0.60 |
| **PS-stratification / on-treatment TAR** | | | | | | | | | | |
| CCAE | 11,792 | 83,057 | 7.5 | 70.2 | 153 | 1,414 | 0.97 (0.81 - 1.17) | 0.78 | 0.94 (0.78 - 1.14) | 0.59 |
| ODOD | 7,578 | 61,676 | 4.3 | 55.7 | 121 | 2,374 | 0.85 (0.69 - 1.04) | 0.12 | 0.81 (0.66 - 1.00) | 0.07 |
| MDCR | 868 | 17,360 | 0.5 | 18.2 | 28 | 795 | 1.32 (0.86 - 1.95) | 0.19 | 1.23 (0.81 - 1.85) | 0.35 |
| Open Claims | 138,123 | 802,465 | 95.0 | 857.9 | 1,277 | 16,698 | 0.94 (0.88 - 1.00) | 0.04 | 0.95 (0.78 - 1.15) | 0.61 |
| OEHR | 8,578 | 90,094 | 2.4 | 48.8 | 50 | 1,242 | 1.12 (0.82 - 1.50) | 0.45 | 1.04 (0.76 - 1.41) | 0.80 |
| VA | 1,261 | 216,571 | 0.9 | 227.0 | 15 | 3,830 | 1.12 (0.63 - 1.81) | 0.68 | 0.94 (0.56 - 1.59) | 0.83 |
| Meta-analysis | 166,071 | 1,037,292 | 109.3 | 1032.6 | 1,601 | 21,728 | 0.94 (0.89 - 0.99) | 0.03 | 0.93 (0.76 - 1.14) | 0.48 |
| Meta-analysis (w/o USOC) | 27,948 | 234,827 | 14.3 | 174.7 | 324 | 5,030 | 0.95 (0.83 - 1.09) | 0.47 | 0.90 (0.77 - 1.07) | 0.34 |
| **PS-matching / on-treatment TAR without escalation** | | | | | | | | | | |
| CCAE | 10,455 | 78,994 | 6.0 | 58.2 | 125 | 1,176 | 0.99 (0.77 - 1.26) | 0.95 | 0.96 (0.75 - 1.23) | 0.77 |
| ODOD | 6,788 | 54,840 | 3.5 | 43.0 | 97 | 1,658 | 0.91 (0.70 - 1.18) | 0.49 | 0.86 (0.66 - 1.12) | 0.29 |
| MDCR | 814 | 13,541 | 0.5 | 13.1 | 22 | 508 | 1.27 (0.73 - 2.08) | 0.37 | 1.25 (0.74 - 2.11) | 0.41 |
| Open Claims | 124,593 | 776,506 | 77.0 | 702.7 | 1,013 | 13,064 | 0.90 (0.83 - 0.98) | 0.02 | 0.90 (0.77 - 1.06) | 0.25 |
| OEHR | 7,896 | 81,313 | 2.0 | 38.2 | 42 | 798 | 1.33 (0.88 - 1.93) | 0.16 | 1.24 (0.84 - 1.83) | 0.30 |
| VA | 1,170 | 28,977 | 0.7 | 18.9 | 12 | 281 | 1.28 (0.55 - 2.60) | 0.53 | 0.96 (0.44 - 2.09) | 0.88 |
| Meta-analysis | 150,546 | 1,005,194 | 89.0 | 855.2 | 1,299 | 17,204 | 0.96 (0.86 - 1.07) | 0.46 | 0.94 (0.77 - 1.15) | 0.56 |
| Meta-analysis (w/o USOC) | 25,953 | 228,688 | 11.9 | 152.5 | 286 | 4,140 | 1.03 (0.88 - 1.21) | 0.69 | 0.99 (0.84 - 1.17) | 0.86 |
| **PS-stratification / on-treatment TAR without escalation** | | | | | | | | | | |
| CCAE | 11,792 | 83,057 | 6.6 | 61.1 | 133 | 1,238 | 0.96 (0.78 - 1.16) | 0.66 | 0.93 (0.76 - 1.14) | 0.54 |
| ODOD | 7,578 | 61,676 | 3.8 | 49.7 | 110 | 2,156 | 0.89 (0.72 - 1.10) | 0.31 | 0.83 (0.67 - 1.04) | 0.13 |
| MDCR | 868 | 17,360 | 0.5 | 16.6 | 22 | 718 | 1.13 (0.70 - 1.75) | 0.59 | 1.04 (0.66 - 1.65) | 0.83 |
| Open Claims | 138,123 | 802,465 | 83.1 | 727.8 | 1,115 | 14,311 | 0.96 (0.89 - 1.02) | 0.22 | 0.96 (0.79 - 1.17) | 0.68 |
| OEHR | 8,578 | 90,094 | 2.1 | 42.6 | 45 | 981 | 1.31 (0.93 - 1.79) | 0.11 | 1.19 (0.85 - 1.64) | 0.33 |
| VA | 1,261 | 216,571 | 0.7 | 202.7 | 12 | 3,300 | 1.13 (0.60 - 1.94) | 0.67 | 0.90 (0.50 - 1.63) | 0.74 |
| Meta-analysis | 166,071 | 1,037,292 | 95.6 | 881.3 | 1,403 | 18,686 | 0.96 (0.91 - 1.02) | 0.22 | 0.94 (0.77 - 1.16) | 0.56 |
| Meta-analysis (w/o USOC) | 27,948 | 234,827 | 12.5 | 153.5 | 288 | 4,375 | 1.00 (0.83 - 1.21) | 0.98 | 0.94 (0.75 - 1.19) | 0.66 |

##### 1.5.17 Hyperkalemia

**Supplementary Table 121:** Exposures, outcomes and risk estimates for Hyperkalemia in GLP1RA (male) (T) vs SU (male) (C) new-user cohorts under four different propensity score (PS) adjustment / time-at-risk (TAR) designs across data sources. We report total patient exposure time in 1,000 patient-year (KYr) units, uncalibrated and calibrated hazard ratios (HRs) and their 95% confidence intervals (CIs). Greyed out entries did not pass study diagnostics and are included only for completeness.

| | Patients | | Exposure (KYr) | | Outcomes | | Uncalibrated | | Calibrated | |
|---|---|---|---|---|---|---|---|---|---|---|
| | T | C | T | C | T | C | HR (95% CI) | $p$ | HR (95% CI) | $p$ |
| **PS-matching / on-treatment TAR** | | | | | | | | | | |
| CCAE | 11,300 | 84,787 | 7.5 | 72.7 | 49 | 564 | 0.71 (0.47 - 1.03) | 0.08 | 0.69 (0.47 - 1.01) | 0.06 |
| ODOD | 7,326 | 60,481 | 4.3 | 54.2 | 70 | 1,220 | 0.74 (0.53 - 1.00) | 0.06 | 0.70 (0.51 - 0.97) | 0.04 |
| MDCR | 911 | 15,191 | 0.6 | 16.5 | 7 | 249 | 0.67 (0.27 - 1.44) | 0.35 | 0.65 (0.28 - 1.52) | 0.33 |
| Open Claims | 137,448 | 854,895 | 97.6 | 919.0 | 378 | 7,452 | 0.69 (0.61 - 0.78) | <0.01 | 0.70 (0.58 - 0.85) | <0.01 |
| OEHR | 8,215 | 84,844 | 2.4 | 46.0 | 20 | 677 | 0.80 (0.46 - 1.30) | 0.39 | 0.75 (0.45 - 1.25) | 0.28 |
| VA | 1,219 | 30,847 | 0.9 | 23.1 | <10 | 347 | 0.54 (0.20 - 1.17) | 0.16 | 0.42 (0.17 - 1.00) | 0.05 |
| Meta-analysis | 165,200 | 1,100,198 | 112.4 | 1108.4 | 524 | 10,162 | 0.70 (0.63 - 0.78) | <0.01 | 0.69 (0.57 - 0.85) | <0.01 |
| Meta-analysis (w/o USOC) | 27,752 | 245,303 | 14.8 | 189.4 | 146 | 2,710 | 0.73 (0.59 - 0.91) | <0.01 | 0.70 (0.56 - 0.87) | <0.01 |
| **PS-stratification / on-treatment TAR** | | | | | | | | | | |
| CCAE | 12,798 | 89,234 | 8.3 | 76.3 | 55 | 596 | 0.78 (0.57 - 1.05) | 0.11 | 0.75 (0.55 - 1.02) | 0.08 |
| ODOD | 8,212 | 67,531 | 4.7 | 62.0 | 70 | 1,534 | 0.79 (0.60 - 1.02) | 0.08 | 0.75 (0.58 - 0.98) | 0.05 |
| MDCR | 986 | 19,737 | 0.7 | 21.0 | 7 | 369 | 0.60 (0.25 - 1.23) | 0.21 | 0.56 (0.25 - 1.24) | 0.16 |
| Open Claims | 152,722 | 882,468 | 104.9 | 950.2 | 394 | 8,247 | 0.70 (0.62 - 0.78) | <0.01 | 0.71 (0.57 - 0.87) | <0.01 |
| OEHR | 8,937 | 93,845 | 2.5 | 51.2 | 20 | 841 | 0.65 (0.39 - 1.00) | 0.07 | 0.60 (0.37 - 0.95) | 0.04 |
| VA | 1,318 | 225,516 | 0.9 | 235.1 | <10 | 4,092 | 0.59 (0.27 - 1.11) | 0.15 | 0.50 (0.24 - 1.02) | 0.06 |
| Meta-analysis | 182,669 | 1,133,078 | 120.4 | 1139.7 | 539 | 11,218 | 0.71 (0.65 - 0.78) | <0.01 | 0.71 (0.57 - 0.88) | <0.01 |
| Meta-analysis (w/o USOC) | 29,947 | 250,610 | 15.5 | 189.5 | 145 | 2,971 | 0.76 (0.63 - 0.91) | <0.01 | 0.72 (0.59 - 0.89) | <0.01 |
| **PS-matching / on-treatment TAR without escalation** | | | | | | | | | | |
| CCAE | 11,300 | 84,787 | 6.6 | 63.0 | 38 | 490 | 0.72 (0.47 - 1.08) | 0.13 | 0.70 (0.46 - 1.07) | 0.10 |
| ODOD | 7,326 | 60,481 | 3.8 | 48.2 | 61 | 1,081 | 0.76 (0.53 - 1.05) | 0.11 | 0.71 (0.51 - 1.00) | 0.06 |
| MDCR | 911 | 15,191 | 0.6 | 14.9 | 6 | 223 | 0.65 (0.24 - 1.49) | 0.36 | 0.64 (0.26 - 1.60) | 0.35 |
| Open Claims | 137,448 | 854,895 | 84.9 | 778.8 | 320 | 6,209 | 0.69 (0.60 - 0.79) | <0.01 | 0.69 (0.57 - 0.84) | <0.01 |
| OEHR | 8,215 | 84,844 | 2.1 | 40.2 | 15 | 517 | 0.68 (0.35 - 1.22) | 0.23 | 0.64 (0.34 - 1.19) | 0.17 |
| VA | 1,219 | 30,847 | 0.7 | 20.2 | <10 | 291 | 0.68 (0.23 - 1.59) | 0.43 | 0.51 (0.19 - 1.34) | 0.17 |
| Meta-analysis | 165,200 | 1,100,198 | 97.8 | 945.1 | 440 | 8,520 | 0.70 (0.62 - 0.79) | <0.01 | 0.69 (0.56 - 0.84) | <0.01 |
| Meta-analysis (w/o USOC) | 27,752 | 245,303 | 13.0 | 166.3 | 120 | 2,311 | 0.73 (0.57 - 0.92) | <0.01 | 0.70 (0.55 - 0.88) | <0.01 |
| **PS-stratification / on-treatment TAR without escalation** | | | | | | | | | | |
| CCAE | 12,798 | 89,234 | 7.2 | 66.2 | 43 | 517 | 0.71 (0.50 - 0.99) | 0.05 | 0.69 (0.49 - 0.98) | 0.04 |
| ODOD | 8,212 | 67,531 | 4.2 | 55.4 | 61 | 1,356 | 0.80 (0.60 - 1.06) | 0.13 | 0.75 (0.56 - 0.99) | 0.06 |
| MDCR | 986 | 19,737 | 0.6 | 19.1 | 6 | 336 | 0.60 (0.23 - 1.29) | 0.24 | 0.55 (0.23 - 1.30) | 0.17 |
| Open Claims | 152,722 | 882,468 | 91.7 | 805.8 | 334 | 6,891 | 0.69 (0.61 - 0.78) | <0.01 | 0.69 (0.56 - 0.86) | <0.01 |
| OEHR | 8,937 | 93,845 | 2.2 | 44.7 | 15 | 626 | 0.59 (0.33 - 0.98) | 0.06 | 0.54 (0.31 - 0.93) | 0.03 |
| VA | 1,318 | 225,516 | 0.7 | 210.0 | <10 | 3,455 | 0.59 (0.23 - 1.22) | 0.22 | 0.47 (0.21 - 1.08) | 0.08 |
| Meta-analysis | 182,669 | 1,133,078 | 105.3 | 972.1 | 453 | 9,390 | 0.70 (0.63 - 0.78) | <0.01 | 0.69 (0.55 - 0.86) | <0.01 |
| Meta-analysis (w/o USOC) | 29,947 | 250,610 | 13.6 | 166.3 | 119 | 2,499 | 0.74 (0.60 - 0.90) | <0.01 | 0.69 (0.54 - 0.89) | <0.01 |

##### 1.5.18 Hypoglycemia

**Supplementary Table 122:** Exposures, outcomes and risk estimates for Hypoglycemia in GLP1RA (male) (T) vs SU (male) (C) new-user cohorts under four different propensity score (PS) adjustment / time-at-risk (TAR) designs across data sources. We report total patient exposure time in 1,000 patient-year (KYr) units, uncalibrated and calibrated hazard ratios (HRs) and their 95% confidence intervals (CIs). Greyed out entries did not pass study diagnostics and are included only for completeness.

| | Patients | | Exposure (KYr) | | Outcomes | | Uncalibrated | | Calibrated | |
|---|---|---|---|---|---|---|---|---|---|---|
| | T | C | T | C | T | C | HR (95% CI) | $p$ | HR (95% CI) | $p$ |
| **PS-matching / on-treatment TAR** | | | | | | | | | | |
| CCAE | 11,282 | 85,231 | 7.6 | 73.3 | 34 | 537 | 0.44 (0.28 - 0.68) | <0.01 | 0.43 (0.27 - 0.68) | <0.01 |
| ODOD | 7,516 | 62,401 | 4.4 | 56.7 | 20 | 957 | 0.27 (0.15 - 0.43) | <0.01 | 0.25 (0.15 - 0.43) | <0.01 |
| MDCR | 924 | 15,389 | 0.6 | 16.8 | <5 | 233 | 0.13 (0.01 - 0.62) | 0.07 | 0.13 (0.01 - 1.17) | 0.07 |
| Open Claims | 138,137 | 863,757 | 98.3 | 933.6 | 196 | 6,745 | 0.33 (0.28 - 0.39) | <0.01 | 0.34 (0.27 - 0.42) | <0.01 |
| OEHR | 8,267 | 85,803 | 2.4 | 46.9 | 12 | 648 | 0.36 (0.18 - 0.64) | <0.01 | 0.33 (0.18 - 0.63) | <0.01 |
| VA | 1,293 | 31,887 | 0.9 | 23.9 | 0 | 253 | | | | |
| Meta-analysis | 166,126 | 1,112,581 | 113.3 | 1127.2 | <267 | 9,120 | 0.34 (0.29 - 0.39) | <0.01 | 0.33 (0.27 - 0.41) | <0.01 |
| Meta-analysis (w/o USOC) | 27,989 | 248,824 | 15.0 | 193.7 | <71 | 2,375 | 0.35 (0.26 - 0.47) | <0.01 | 0.33 (0.25 - 0.45) | <0.01 |
| **PS-stratification / on-treatment TAR** | | | | | | | | | | |
| CCAE | 12,757 | 89,567 | 8.3 | 76.7 | 37 | 570 | 0.41 (0.28 - 0.58) | <0.01 | 0.40 (0.28 - 0.57) | <0.01 |
| ODOD | 8,400 | 69,983 | 4.8 | 65.3 | 25 | 1,202 | 0.27 (0.18 - 0.41) | <0.01 | 0.26 (0.17 - 0.40) | <0.01 |
| MDCR | 998 | 20,199 | 0.7 | 21.6 | <5 | 356 | 0.11 (0.01 - 0.48) | 0.05 | 0.10 (0.01 - 0.89) | 0.04 |
| Open Claims | 153,146 | 892,316 | 105.4 | 966.3 | 219 | 7,417 | 0.33 (0.29 - 0.38) | <0.01 | 0.34 (0.27 - 0.42) | <0.01 |
| OEHR | 8,985 | 95,219 | 2.6 | 52.5 | 15 | 763 | 0.43 (0.24 - 0.70) | <0.01 | 0.39 (0.23 - 0.67) | <0.01 |
| VA | 1,389 | 237,705 | 1.0 | 252.5 | <10 | 2,179 | 0.10 (0.01 - 0.45) | 0.04 | 0.08 (0.01 - 0.75) | 0.03 |
| Meta-analysis | 183,288 | 1,147,085 | 121.1 | 1160.9 | 296 | 9,952 | 0.34 (0.30 - 0.38) | <0.01 | 0.33 (0.27 - 0.42) | <0.01 |
| Meta-analysis (w/o USOC) | 30,142 | 254,769 | 15.7 | 194.6 | 77 | 2,535 | 0.36 (0.27 - 0.48) | <0.01 | 0.34 (0.26 - 0.46) | <0.01 |
| **PS-matching / on-treatment TAR without escalation** | | | | | | | | | | |
| CCAE | 11,282 | 85,231 | 6.6 | 63.5 | 25 | 445 | 0.43 (0.25 - 0.69) | <0.01 | 0.41 (0.25 - 0.69) | <0.01 |
| ODOD | 7,516 | 62,401 | 3.9 | 50.4 | 14 | 825 | 0.24 (0.12 - 0.41) | <0.01 | 0.23 (0.12 - 0.41) | <0.01 |
| MDCR | 924 | 15,389 | 0.6 | 15.2 | 0 | 207 | | | | |
| Open Claims | 138,137 | 863,757 | 85.4 | 791.0 | 151 | 5,522 | 0.33 (0.27 - 0.39) | <0.01 | 0.33 (0.26 - 0.41) | <0.01 |
| OEHR | 8,267 | 85,803 | 2.1 | 41.0 | 10 | 486 | 0.40 (0.18 - 0.75) | <0.01 | 0.37 (0.18 - 0.75) | <0.01 |
| VA | 1,293 | 31,887 | 0.7 | 20.9 | 0 | 207 | | | | |
| Meta-analysis | 165,202 | 1,097,192 | 98.0 | 945.9 | 200 | 7,278 | 0.33 (0.28 - 0.39) | <0.01 | 0.33 (0.26 - 0.41) | <0.01 |
| Meta-analysis (w/o USOC) | 27,065 | 233,435 | 12.6 | 154.9 | 49 | 1,756 | 0.35 (0.24 - 0.50) | <0.01 | 0.33 (0.23 - 0.48) | <0.01 |
| **PS-stratification / on-treatment TAR without escalation** | | | | | | | | | | |
| CCAE | 12,757 | 89,567 | 7.3 | 66.6 | 28 | 475 | 0.36 (0.23 - 0.53) | <0.01 | 0.35 (0.23 - 0.52) | <0.01 |
| ODOD | 8,400 | 69,983 | 4.3 | 58.3 | 19 | 1,045 | 0.25 (0.15 - 0.39) | <0.01 | 0.23 (0.14 - 0.37) | <0.01 |
| MDCR | 998 | 20,199 | 0.6 | 19.6 | 0 | 322 | | | | |
| Open Claims | 153,146 | 892,316 | 92.1 | 819.2 | 173 | 6,105 | 0.31 (0.26 - 0.36) | <0.01 | 0.31 (0.24 - 0.40) | <0.01 |
| OEHR | 8,985 | 95,219 | 2.2 | 45.7 | 12 | 569 | 0.43 (0.22 - 0.74) | <0.01 | 0.39 (0.21 - 0.70) | <0.01 |
| VA | 1,389 | 237,705 | 0.8 | 225.1 | 0 | 1,785 | | | | |
| Meta-analysis | 183,288 | 1,147,085 | 105.9 | 989.9 | 232 | 8,194 | 0.31 (0.27 - 0.36) | <0.01 | 0.31 (0.24 - 0.39) | <0.01 |
| Meta-analysis (w/o USOC) | 30,142 | 254,769 | 13.8 | 170.7 | 59 | 2,089 | 0.33 (0.25 - 0.44) | <0.01 | 0.31 (0.23 - 0.43) | <0.01 |

##### 1.5.19 Hypotension

**Supplementary Table 123:** Exposures, outcomes and risk estimates for Hypotension in GLP1RA (male) (T) vs SU (male) (C) new-user cohorts under four different propensity score (PS) adjustment / time-at-risk (TAR) designs across data sources. We report total patient exposure time in 1,000 patient-year (KYr) units, uncalibrated and calibrated hazard ratios (HRs) and their 95% confidence intervals (CIs). Greyed out entries did not pass study diagnostics and are included only for completeness.

| | Patients | | Exposure (KYr) | | Outcomes | | Uncalibrated | | Calibrated | |
|---|---|---|---|---|---|---|---|---|---|---|
| | T | C | T | C | T | C | HR (95% CI) | $p$ | HR (95% CI) | $p$ |
| **PS-matching / on-treatment TAR** | | | | | | | | | | |
| CCAE | 11,253 | 84,856 | 7.5 | 73.0 | 66 | 510 | 1.18 (0.83 - 1.66) | 0.33 | 1.15 (0.81 - 1.62) | 0.44 |
| ODOD | 7,303 | 60,032 | 4.3 | 54.1 | 71 | 1,178 | 0.76 (0.54 - 1.04) | 0.10 | 0.72 (0.52 - 1.00) | 0.06 |
| MDCR | 894 | 14,745 | 0.6 | 16.0 | 16 | 333 | 1.27 (0.62 - 2.39) | 0.48 | 1.25 (0.64 - 2.44) | 0.53 |
| Open Claims | 135,689 | 844,420 | 96.5 | 910.9 | 575 | 7,991 | 0.94 (0.84 - 1.04) | 0.24 | 0.95 (0.80 - 1.14) | 0.60 |
| OEHR | 8,178 | 84,514 | 2.4 | 45.9 | 37 | 752 | 1.16 (0.78 - 1.67) | 0.46 | 1.08 (0.73 - 1.59) | 0.71 |
| VA | 1,221 | 30,932 | 0.9 | 23.1 | 17 | 305 | 1.08 (0.59 - 1.88) | 0.78 | 0.84 (0.47 - 1.50) | 0.57 |
| Meta-analysis | 163,317 | 1,088,567 | 111.2 | 1099.9 | 765 | 10,764 | 0.97 (0.85 - 1.10) | 0.60 | 0.96 (0.78 - 1.18) | 0.69 |
| Meta-analysis (w/o USOC) | 27,628 | 244,147 | 14.8 | 189.1 | 190 | 2,773 | 1.03 (0.80 - 1.32) | 0.82 | 0.99 (0.76 - 1.27) | 0.90 |
| **PS-stratification / on-treatment TAR** | | | | | | | | | | |
| CCAE | 12,737 | 89,064 | 8.3 | 76.4 | 75 | 555 | 1.21 (0.91 - 1.59) | 0.19 | 1.17 (0.88 - 1.55) | 0.30 |
| ODOD | 8,191 | 67,236 | 4.7 | 62.2 | 76 | 1,548 | 0.84 (0.64 - 1.08) | 0.18 | 0.80 (0.62 - 1.04) | 0.11 |
| MDCR | 967 | 19,153 | 0.6 | 20.4 | 17 | 515 | 1.35 (0.77 - 2.23) | 0.27 | 1.25 (0.73 - 2.14) | 0.42 |
| Open Claims | 150,746 | 871,592 | 103.6 | 942.1 | 624 | 8,952 | 0.92 (0.84 - 1.01) | 0.07 | 0.93 (0.76 - 1.14) | 0.52 |
| OEHR | 8,903 | 93,699 | 2.5 | 51.2 | 38 | 957 | 1.15 (0.80 - 1.61) | 0.43 | 1.06 (0.75 - 1.51) | 0.74 |
| VA | 1,317 | 227,052 | 0.9 | 238.7 | 18 | 3,318 | 1.38 (0.82 - 2.16) | 0.19 | 1.17 (0.72 - 1.89) | 0.54 |
| Meta-analysis | 180,577 | 1,121,591 | 119.1 | 1132.0 | 813 | 12,012 | 0.98 (0.85 - 1.13) | 0.78 | 0.97 (0.76 - 1.24) | 0.80 |
| Meta-analysis (w/o USOC) | 29,831 | 249,999 | 15.5 | 189.8 | 189 | 3,060 | 1.04 (0.82 - 1.32) | 0.76 | 0.99 (0.76 - 1.28) | 0.91 |
| **PS-matching / on-treatment TAR without escalation** | | | | | | | | | | |
| CCAE | 11,253 | 84,856 | 6.5 | 63.3 | 55 | 419 | 1.25 (0.85 - 1.79) | 0.24 | 1.21 (0.83 - 1.75) | 0.33 |
| ODOD | 7,303 | 60,032 | 3.8 | 48.1 | 60 | 1,057 | 0.69 (0.47 - 0.98) | 0.04 | 0.65 (0.45 - 0.94) | 0.02 |
| MDCR | 894 | 14,745 | 0.5 | 14.5 | 13 | 291 | 1.01 (0.44 - 2.05) | 0.99 | 0.99 (0.46 - 2.14) | 0.89 |
| Open Claims | 135,689 | 844,420 | 83.9 | 771.6 | 488 | 6,645 | 0.95 (0.84 - 1.07) | 0.43 | 0.96 (0.80 - 1.15) | 0.64 |
| OEHR | 8,178 | 84,514 | 2.0 | 40.1 | 36 | 519 | 1.43 (0.93 - 2.12) | 0.09 | 1.33 (0.88 - 2.01) | 0.19 |
| VA | 1,221 | 30,932 | 0.7 | 20.3 | 15 | 249 | 1.38 (0.70 - 2.54) | 0.33 | 1.03 (0.54 - 1.97) | 0.86 |
| Meta-analysis | 163,317 | 1,088,567 | 96.8 | 937.6 | 652 | 8,931 | 1.02 (0.79 - 1.30) | 0.90 | 1.00 (0.74 - 1.34) | 0.93 |
| Meta-analysis (w/o USOC) | 27,628 | 244,147 | 12.9 | 166.0 | 164 | 2,286 | 1.05 (0.73 - 1.52) | 0.78 | 1.01 (0.70 - 1.46) | 0.93 |
| **PS-stratification / on-treatment TAR without escalation** | | | | | | | | | | |
| CCAE | 12,737 | 89,064 | 7.2 | 66.4 | 61 | 461 | 1.26 (0.91 - 1.71) | 0.15 | 1.23 (0.90 - 1.68) | 0.21 |
| ODOD | 8,191 | 67,236 | 4.1 | 55.5 | 65 | 1,397 | 0.81 (0.61 - 1.06) | 0.14 | 0.76 (0.57 - 1.00) | 0.06 |
| MDCR | 967 | 19,153 | 0.6 | 18.6 | 14 | 452 | 1.19 (0.64 - 2.07) | 0.57 | 1.09 (0.60 - 1.96) | 0.78 |
| Open Claims | 150,746 | 871,592 | 90.6 | 798.5 | 534 | 7,508 | 0.93 (0.84 - 1.02) | 0.14 | 0.93 (0.75 - 1.14) | 0.50 |
| OEHR | 8,903 | 93,699 | 2.2 | 44.7 | 37 | 667 | 1.58 (1.08 - 2.24) | 0.01 | 1.43 (0.99 - 2.06) | 0.06 |
| VA | 1,317 | 227,052 | 0.7 | 213.1 | 15 | 2,734 | 1.62 (0.92 - 2.64) | 0.07 | 1.29 (0.76 - 2.20) | 0.36 |
| Meta-analysis | 180,577 | 1,121,591 | 104.1 | 965.1 | 697 | 10,033 | 1.08 (0.82 - 1.42) | 0.61 | 1.05 (0.75 - 1.48) | 0.77 |
| Meta-analysis (w/o USOC) | 29,831 | 249,999 | 13.6 | 166.6 | 163 | 2,525 | 1.16 (0.78 - 1.71) | 0.47 | 1.09 (0.72 - 1.65) | 0.69 |

##### 1.5.20 Joint pain

**Supplementary Table 124:** Exposures, outcomes and risk estimates for Joint pain in GLP1RA (male) (T) vs SU (male) (C) new-user cohorts under four different propensity score (PS) adjustment / time-at-risk (TAR) designs across data sources. We report total patient exposure time in 1,000 patient-year (KYr) units, uncalibrated and calibrated hazard ratios (HRs) and their 95% confidence intervals (CIs). Greyed out entries did not pass study diagnostics and are included only for completeness.

| | Patients | | Exposure (KYr) | | Outcomes | | Uncalibrated | | Calibrated | |
|---|---|---|---|---|---|---|---|---|---|---|
| | T | C | T | C | T | C | HR (95% CI) | $p$ | HR (95% CI) | $p$ |
| **PS-matching / on-treatment TAR** | | | | | | | | | | |
| CCAE | 10,943 | 83,101 | 7.3 | 71.2 | 61 | 578 | 0.92 (0.63 - 1.32) | 0.66 | 0.89 (0.61 - 1.30) | 0.56 |
| ODOD | 7,211 | 60,514 | 4.2 | 54.9 | 37 | 571 | 0.87 (0.56 - 1.33) | 0.54 | 0.83 (0.54 - 1.29) | 0.42 |
| MDCR | 896 | 14,874 | 0.6 | 16.2 | <5 | 140 | 0.36 (0.06 - 1.27) | 0.20 | 0.35 (0.07 - 1.68) | 0.19 |
| Open Claims | 132,805 | 838,401 | 94.4 | 906.9 | 441 | 4,276 | 0.92 (0.80 - 1.05) | 0.20 | 0.93 (0.77 - 1.13) | 0.49 |
| OEHR | 7,966 | 83,548 | 2.3 | 45.6 | 14 | 382 | 0.66 (0.32 - 1.22) | 0.21 | 0.61 (0.31 - 1.19) | 0.16 |
| VA | 1,157 | 28,186 | 0.8 | 20.9 | 11 | 137 | 1.98 (0.80 - 4.33) | 0.11 | 1.54 (0.66 - 3.58) | 0.33 |
| Meta-analysis | 159,821 | 1,080,438 | 108.9 | 1094.8 | <558 | 5,947 | 0.90 (0.80 - 1.01) | 0.08 | 0.89 (0.73 - 1.09) | 0.26 |
| Meta-analysis (w/o USOC) | 27,016 | 242,037 | 14.4 | 187.9 | <117 | 1,671 | 0.84 (0.65 - 1.08) | 0.18 | 0.80 (0.62 - 1.04) | 0.12 |
| **PS-stratification / on-treatment TAR** | | | | | | | | | | |
| CCAE | 12,378 | 87,314 | 8.0 | 74.5 | 64 | 600 | 0.92 (0.69 - 1.21) | 0.56 | 0.89 (0.67 - 1.19) | 0.45 |
| ODOD | 8,069 | 67,870 | 4.6 | 63.3 | 44 | 646 | 0.80 (0.56 - 1.10) | 0.19 | 0.76 (0.54 - 1.07) | 0.13 |
| MDCR | 967 | 19,773 | 0.6 | 21.2 | <5 | 176 | 0.61 (0.18 - 1.52) | 0.36 | 0.57 (0.20 - 1.64) | 0.29 |
| Open Claims | 147,146 | 867,272 | 101.3 | 940.0 | 492 | 4,493 | 0.98 (0.88 - 1.08) | 0.67 | 0.99 (0.80 - 1.22) | 0.90 |
| OEHR | 8,667 | 92,518 | 2.5 | 50.9 | 16 | 431 | 0.83 (0.47 - 1.35) | 0.48 | 0.76 (0.45 - 1.29) | 0.33 |
| VA | 1,248 | 212,284 | 0.9 | 224.9 | 12 | 1,767 | 2.05 (1.07 - 3.56) | 0.02 | 1.74 (0.95 - 3.17) | 0.08 |
| Meta-analysis | 176,260 | 1,114,974 | 116.4 | 1128.7 | 616 | 6,170 | 0.95 (0.87 - 1.04) | 0.29 | 0.94 (0.76 - 1.17) | 0.58 |
| Meta-analysis (w/o USOC) | 29,114 | 247,702 | 15.1 | 188.7 | 124 | 1,677 | 0.86 (0.70 - 1.05) | 0.14 | 0.82 (0.65 - 1.02) | 0.12 |
| **PS-matching / on-treatment TAR without escalation** | | | | | | | | | | |
| CCAE | 10,943 | 83,101 | 6.3 | 61.7 | 51 | 504 | 0.86 (0.56 - 1.26) | 0.45 | 0.83 (0.55 - 1.24) | 0.38 |
| ODOD | 7,211 | 60,514 | 3.8 | 48.9 | 33 | 506 | 0.80 (0.49 - 1.27) | 0.37 | 0.76 (0.47 - 1.22) | 0.26 |
| MDCR | 896 | 14,874 | 0.5 | 14.7 | <5 | 127 | 0.38 (0.06 - 1.35) | 0.23 | 0.37 (0.08 - 1.80) | 0.22 |
| Open Claims | 132,805 | 838,401 | 82.1 | 768.3 | 375 | 3,616 | 0.86 (0.74 - 0.99) | 0.04 | 0.86 (0.71 - 1.05) | 0.16 |
| OEHR | 7,966 | 83,548 | 2.0 | 39.7 | 14 | 318 | 0.67 (0.32 - 1.30) | 0.27 | 0.63 (0.31 - 1.28) | 0.21 |
| VA | 1,157 | 28,186 | 0.7 | 18.4 | 11 | 123 | 2.26 (0.89 - 5.09) | 0.07 | 1.69 (0.71 - 4.05) | 0.24 |
| Meta-analysis | 159,821 | 1,080,438 | 94.7 | 933.3 | <478 | 5,071 | 0.84 (0.74 - 0.96) | <0.01 | 0.83 (0.67 - 1.02) | 0.08 |
| Meta-analysis (w/o USOC) | 27,016 | 242,037 | 12.6 | 165.0 | <103 | 1,455 | 0.79 (0.60 - 1.04) | 0.09 | 0.75 (0.57 - 1.00) | 0.06 |
| **PS-stratification / on-treatment TAR without escalation** | | | | | | | | | | |
| CCAE | 12,378 | 87,314 | 7.0 | 64.7 | 54 | 525 | 0.85 (0.61 - 1.15) | 0.29 | 0.83 (0.60 - 1.13) | 0.24 |
| ODOD | 8,069 | 67,870 | 4.1 | 56.5 | 39 | 576 | 0.81 (0.56 - 1.15) | 0.25 | 0.76 (0.53 - 1.09) | 0.14 |
| MDCR | 967 | 19,773 | 0.6 | 19.3 | <5 | 161 | 0.65 (0.19 - 1.63) | 0.43 | 0.59 (0.20 - 1.73) | 0.34 |
| Open Claims | 147,146 | 867,272 | 88.5 | 796.8 | 423 | 3,799 | 0.95 (0.85 - 1.07) | 0.42 | 0.95 (0.77 - 1.18) | 0.68 |
| OEHR | 8,667 | 92,518 | 2.1 | 44.3 | 16 | 354 | 1.01 (0.57 - 1.67) | 0.97 | 0.92 (0.53 - 1.57) | 0.76 |
| VA | 1,248 | 212,284 | 0.7 | 200.9 | 11 | 1,590 | 2.30 (1.17 - 4.09) | <0.01 | 1.83 (0.98 - 3.44) | 0.06 |
| Meta-analysis | 176,260 | 1,114,974 | 101.7 | 962.3 | 532 | 5,254 | 0.93 (0.84 - 1.03) | 0.17 | 0.91 (0.73 - 1.14) | 0.41 |
| Meta-analysis (w/o USOC) | 29,114 | 247,702 | 13.2 | 165.5 | 109 | 1,455 | 0.86 (0.69 - 1.06) | 0.16 | 0.81 (0.63 - 1.04) | 0.12 |

##### 1.5.21 Lower extremity amputation

**Supplementary Table 125:** Exposures, outcomes and risk estimates for Lower extremity amputation in GLP1RA (male) (T) vs SU (male) (C) new-user cohorts under four different propensity score (PS) adjustment / time-at-risk (TAR) designs across data sources. We report total patient exposure time in 1,000 patient-year (KYr) units, uncalibrated and calibrated hazard ratios (HRs) and their 95% confidence intervals (CIs). Greyed out entries did not pass study diagnostics and are included only for completeness.

| | Patients | | Exposure (KYr) | | Outcomes | | Uncalibrated | | Calibrated | |
|---|---|---|---|---|---|---|---|---|---|---|
| | T | C | T | C | T | C | HR (95% CI) | $p$ | HR (95% CI) | $p$ |
| **PS-matching / on-treatment TAR** | | | | | | | | | | |
| CCAE | 11,450 | 86,132 | 7.7 | 74.3 | 7 | 72 | 1.03 (0.32 - 2.75) | 0.96 | 1.00 (0.34 - 2.92) | 0.97 |
| ODOD | 7,586 | 62,895 | 4.5 | 57.5 | 6 | 88 | 0.59 (0.13 - 1.79) | 0.42 | 0.56 (0.15 - 2.05) | 0.38 |
| MDCR | 931 | 15,477 | 0.6 | 17.0 | 0 | 23 | | | | |
| Open Claims | 139,861 | 871,656 | 99.6 | 945.1 | 49 | 762 | 0.76 (0.51 - 1.09) | 0.15 | 0.77 (0.51 - 1.15) | 0.21 |
| OEHR | 8,358 | 86,380 | 2.4 | 47.6 | 0 | 68 | | | | |
| VA | 1,306 | 32,239 | 0.9 | 24.2 | 0 | 23 | | | | |
| Meta-analysis | 158,897 | 1,020,683 | 111.7 | 1076.9 | 62 | 922 | 0.77 (0.54 - 1.09) | 0.14 | 0.76 (0.52 - 1.12) | 0.16 |
| Meta-analysis (w/o USOC) | 19,036 | 149,027 | 12.1 | 131.8 | 13 | 160 | 0.82 (0.36 - 1.87) | 0.64 | 0.79 (0.34 - 1.79) | 0.57 |
| **PS-stratification / on-treatment TAR** | | | | | | | | | | |
| CCAE | 12,974 | 90,464 | 8.5 | 77.8 | 7 | 80 | 1.01 (0.39 - 2.24) | 0.99 | 0.98 (0.41 - 2.34) | 0.95 |
| ODOD | 8,500 | 70,613 | 4.9 | 66.4 | 6 | 102 | 0.93 (0.34 - 2.09) | 0.87 | 0.88 (0.36 - 2.19) | 0.79 |
| MDCR | 1,007 | 20,312 | 0.7 | 21.8 | 0 | 31 | | | | |
| Open Claims | 155,300 | 901,023 | 107.0 | 979.0 | 49 | 832 | 0.75 (0.54 - 1.02) | 0.07 | 0.76 (0.53 - 1.09) | 0.14 |
| OEHR | 9,096 | 95,815 | 2.6 | 53.3 | 0 | 83 | | | | |
| VA | 1,404 | 239,038 | 1.0 | 255.1 | 0 | 204 | | | | |
| Meta-analysis | 176,774 | 1,062,100 | 120.3 | 1123.2 | 62 | 1,014 | 0.79 (0.60 - 1.04) | 0.10 | 0.78 (0.56 - 1.10) | 0.16 |
| Meta-analysis (w/o USOC) | 21,474 | 161,077 | 13.3 | 144.2 | 13 | 182 | 0.97 (0.52 - 1.81) | 0.92 | 0.92 (0.49 - 1.74) | 0.80 |
| **PS-matching / on-treatment TAR without escalation** | | | | | | | | | | |
| CCAE | 11,450 | 86,132 | 6.7 | 64.3 | 6 | 57 | 1.19 (0.37 - 3.21) | 0.75 | 1.15 (0.39 - 3.40) | 0.80 |
| ODOD | 7,586 | 62,895 | 4.0 | 51.1 | 5 | 73 | 0.68 (0.15 - 2.12) | 0.57 | 0.65 (0.18 - 2.39) | 0.51 |
| MDCR | 931 | 15,477 | 0.6 | 15.3 | 0 | 19 | | | | |
| Open Claims | 139,861 | 871,656 | 86.5 | 799.9 | 39 | 612 | 0.76 (0.49 - 1.13) | 0.20 | 0.76 (0.49 - 1.18) | 0.23 |
| OEHR | 8,358 | 86,380 | 2.1 | 41.4 | 0 | 35 | | | | |
| VA | 1,306 | 32,239 | 0.7 | 21.3 | 0 | 16 | | | | |
| Meta-analysis | 158,897 | 1,020,683 | 97.1 | 915.4 | 50 | 742 | 0.79 (0.55 - 1.15) | 0.23 | 0.78 (0.52 - 1.17) | 0.24 |
| Meta-analysis (w/o USOC) | 19,036 | 149,027 | 10.6 | 115.5 | 11 | 130 | 0.95 (0.41 - 2.19) | 0.90 | 0.91 (0.40 - 2.09) | 0.83 |
| **PS-stratification / on-treatment TAR without escalation** | | | | | | | | | | |
| CCAE | 12,974 | 90,464 | 7.4 | 67.5 | 6 | 64 | 1.22 (0.43 - 2.94) | 0.68 | 1.19 (0.46 - 3.12) | 0.72 |
| ODOD | 8,500 | 70,613 | 4.3 | 59.2 | 5 | 86 | 1.15 (0.38 - 2.81) | 0.78 | 1.08 (0.40 - 2.93) | 0.89 |
| MDCR | 1,007 | 20,312 | 0.6 | 19.8 | 0 | 26 | | | | |
| Open Claims | 155,300 | 901,023 | 93.4 | 829.1 | 39 | 670 | 0.73 (0.50 - 1.02) | 0.07 | 0.73 (0.49 - 1.08) | 0.12 |
| OEHR | 9,096 | 95,815 | 2.3 | 46.3 | 0 | 44 | | | | |
| VA | 1,404 | 239,038 | 0.8 | 227.1 | 0 | 153 | | | | |
| Meta-analysis | 176,774 | 1,062,100 | 105.1 | 955.7 | 50 | 820 | 0.82 (0.58 - 1.16) | 0.26 | 0.80 (0.54 - 1.19) | 0.27 |
| Meta-analysis (w/o USOC) | 21,474 | 161,077 | 11.7 | 126.7 | 11 | 150 | 1.19 (0.60 - 2.38) | 0.62 | 1.12 (0.55 - 2.27) | 0.75 |

##### 1.5.22 Nausea

**Supplementary Table 126:** Exposures, outcomes and risk estimates for Nausea in GLP1RA (male) (T) vs SU (male) (C) new-user cohorts under four different propensity score (PS) adjustment / time-at-risk (TAR) designs across data sources. We report total patient exposure time in 1,000 patient-year (KYr) units, uncalibrated and calibrated hazard ratios (HRs) and their 95% confidence intervals (CIs). Greyed out entries did not pass study diagnostics and are included only for completeness.

| | Patients | | Exposure (KYr) | | Outcomes | | Uncalibrated | | Calibrated | |
|---|---|---|---|---|---|---|---|---|---|---|
| | T | C | T | C | T | C | HR (95% CI) | $p$ | HR (95% CI) | $p$ |
| **PS-matching / on-treatment TAR** | | | | | | | | | | |
| CCAE | 10,329 | 79,722 | 6.9 | 68.4 | 223 | 1,359 | 1.47 (1.21 - 1.78) | <0.01 | 1.43 (1.17 - 1.74) | <0.01 |
| ODOD | 6,728 | 56,420 | 3.9 | 50.6 | 178 | 1,601 | 1.30 (1.05 - 1.59) | 0.01 | 1.24 (1.01 - 1.52) | 0.06 |
| MDCR | 859 | 14,193 | 0.6 | 15.4 | 14 | 411 | 0.92 (0.47 - 1.65) | 0.79 | 0.90 (0.48 - 1.68) | 0.74 |
| Open Claims | 123,951 | 793,055 | 88.2 | 857.8 | 1,820 | 12,893 | 1.46 (1.37 - 1.56) | <0.01 | 1.49 (1.28 - 1.74) | <0.01 |
| OEHR | 7,668 | 80,799 | 2.2 | 43.4 | 118 | 1,161 | 2.13 (1.66 - 2.72) | <0.01 | 1.99 (1.55 - 2.55) | <0.01 |
| VA | 1,224 | 30,316 | 0.9 | 22.8 | 22 | 215 | 3.04 (1.67 - 5.27) | <0.01 | 2.36 (1.33 - 4.20) | <0.01 |
| Meta-analysis | 149,535 | 1,024,189 | 101.7 | 1035.6 | 2,353 | 17,425 | 1.49 (1.23 - 1.79) | <0.01 | 1.47 (1.15 - 1.89) | <0.01 |
| Meta-analysis (w/o USOC) | 25,584 | 231,134 | 13.5 | 177.8 | 533 | 4,532 | 1.48 (1.14 - 1.93) | <0.01 | 1.42 (1.08 - 1.86) | 0.01 |
| **PS-stratification / on-treatment TAR** | | | | | | | | | | |
| CCAE | 11,671 | 83,812 | 7.5 | 71.6 | 263 | 1,451 | 1.43 (1.22 - 1.66) | <0.01 | 1.38 (1.18 - 1.62) | <0.01 |
| ODOD | 7,529 | 63,655 | 4.2 | 58.8 | 197 | 1,934 | 1.38 (1.17 - 1.63) | <0.01 | 1.32 (1.11 - 1.57) | <0.01 |
| MDCR | 924 | 18,733 | 0.6 | 20.0 | 14 | 565 | 0.74 (0.41 - 1.26) | 0.31 | 0.69 (0.39 - 1.22) | 0.21 |
| Open Claims | 137,078 | 820,444 | 94.4 | 889.4 | 2,003 | 13,802 | 1.40 (1.33 - 1.48) | <0.01 | 1.42 (1.18 - 1.72) | <0.01 |
| OEHR | 8,322 | 89,549 | 2.3 | 48.5 | 125 | 1,328 | 1.94 (1.58 - 2.37) | <0.01 | 1.79 (1.46 - 2.20) | <0.01 |
| VA | 1,317 | 226,602 | 0.9 | 240.4 | 22 | 2,291 | 2.59 (1.61 - 3.93) | <0.01 | 2.19 (1.40 - 3.42) | <0.01 |
| Meta-analysis | 164,600 | 1,057,460 | 108.5 | 1068.4 | 2,588 | 18,515 | 1.50 (1.31 - 1.71) | <0.01 | 1.48 (1.17 - 1.88) | <0.01 |
| Meta-analysis (w/o USOC) | 27,522 | 237,016 | 14.1 | 178.9 | 585 | 4,713 | 1.55 (1.28 - 1.88) | <0.01 | 1.47 (1.19 - 1.83) | <0.01 |
| **PS-matching / on-treatment TAR without escalation** | | | | | | | | | | |
| CCAE | 10,329 | 79,722 | 6.0 | 59.4 | 199 | 1,170 | 1.53 (1.24 - 1.88) | <0.01 | 1.48 (1.20 - 1.83) | <0.01 |
| ODOD | 6,728 | 56,420 | 3.4 | 45.2 | 161 | 1,406 | 1.31 (1.05 - 1.63) | 0.02 | 1.24 (0.99 - 1.55) | 0.07 |
| MDCR | 859 | 14,193 | 0.5 | 14.0 | 10 | 364 | 0.70 (0.32 - 1.39) | 0.35 | 0.69 (0.33 - 1.46) | 0.34 |
| Open Claims | 123,951 | 793,055 | 76.8 | 729.0 | 1,588 | 10,636 | 1.49 (1.39 - 1.60) | <0.01 | 1.50 (1.28 - 1.75) | <0.01 |
| OEHR | 7,668 | 80,799 | 1.9 | 38.1 | 100 | 893 | 2.17 (1.65 - 2.83) | <0.01 | 2.03 (1.55 - 2.66) | <0.01 |
| VA | 1,224 | 30,316 | 0.7 | 20.0 | 19 | 169 | 3.29 (1.58 - 6.41) | <0.01 | 2.46 (1.22 - 4.97) | 0.01 |
| Meta-analysis | 149,535 | 1,024,189 | 88.7 | 885.6 | 2,058 | 14,469 | 1.50 (1.22 - 1.84) | <0.01 | 1.48 (1.14 - 1.92) | <0.01 |
| Meta-analysis (w/o USOC) | 25,584 | 231,134 | 11.9 | 156.7 | 470 | 3,833 | 1.48 (1.10 - 1.99) | <0.01 | 1.42 (1.05 - 1.91) | 0.03 |
| **PS-stratification / on-treatment TAR without escalation** | | | | | | | | | | |
| CCAE | 11,671 | 83,812 | 6.6 | 62.4 | 235 | 1,251 | 1.48 (1.26 - 1.74) | <0.01 | 1.45 (1.23 - 1.71) | <0.01 |
| ODOD | 7,529 | 63,655 | 3.8 | 52.6 | 179 | 1,706 | 1.46 (1.22 - 1.74) | <0.01 | 1.36 (1.13 - 1.63) | <0.01 |
| MDCR | 924 | 18,733 | 0.5 | 18.2 | 10 | 505 | 0.61 (0.30 - 1.12) | 0.14 | 0.56 (0.29 - 1.09) | 0.09 |
| Open Claims | 137,078 | 820,444 | 82.6 | 756.3 | 1,764 | 11,431 | 1.47 (1.38 - 1.55) | <0.01 | 1.47 (1.21 - 1.78) | <0.01 |
| OEHR | 8,322 | 89,549 | 2.0 | 42.5 | 107 | 1,021 | 2.04 (1.62 - 2.54) | <0.01 | 1.85 (1.48 - 2.32) | <0.01 |
| VA | 1,317 | 226,602 | 0.7 | 214.6 | 19 | 1,925 | 3.08 (1.84 - 4.84) | <0.01 | 2.45 (1.51 - 3.98) | <0.01 |
| Meta-analysis | 164,600 | 1,057,460 | 95.0 | 913.8 | 2,285 | 15,409 | 1.56 (1.37 - 1.77) | <0.01 | 1.52 (1.20 - 1.93) | <0.01 |
| Meta-analysis (w/o USOC) | 27,522 | 237,016 | 12.4 | 157.5 | 521 | 3,978 | 1.62 (1.34 - 1.96) | <0.01 | 1.53 (1.21 - 1.93) | <0.01 |

##### 1.5.23 Peripheral edema

**Supplementary Table 127:** Exposures, outcomes and risk estimates for Peripheral edema in GLP1RA (male) (T) vs SU (male) (C) new-user cohorts under four different propensity score (PS) adjustment / time-at-risk (TAR) designs across data sources. We report total patient exposure time in 1,000 patient-year (KYr) units, uncalibrated and calibrated hazard ratios (HRs) and their 95% confidence intervals (CIs). Greyed out entries did not pass study diagnostics and are included only for completeness.

| | Patients | | Exposure (KYr) | | Outcomes | | Uncalibrated | | Calibrated | |
|---|---|---|---|---|---|---|---|---|---|---|
| | T | C | T | C | T | C | HR (95% CI) | $p$ | HR (95% CI) | $p$ |
| **PS-matching / on-treatment TAR** | | | | | | | | | | |
| CCAE | 9,596 | 75,118 | 6.3 | 62.5 | 215 | 2,537 | 0.80 (0.66 - 0.96) | 0.02 | 0.77 (0.64 - 0.93) | 0.01 |
| ODOD | 6,069 | 50,874 | 3.5 | 43.4 | 168 | 3,085 | 0.80 (0.66 - 0.96) | 0.02 | 0.76 (0.63 - 0.93) | <0.01 |
| MDCR | 718 | 12,217 | 0.5 | 12.5 | 30 | 958 | 0.95 (0.59 - 1.45) | 0.81 | 0.93 (0.59 - 1.45) | 0.73 |
| Open Claims | 110,824 | 717,174 | 78.4 | 752.6 | 1,562 | 24,428 | 0.78 (0.74 - 0.84) | <0.01 | 0.80 (0.68 - 0.93) | <0.01 |
| OEHR | 7,080 | 75,134 | 2.0 | 39.0 | 76 | 2,006 | 0.80 (0.60 - 1.04) | 0.11 | 0.75 (0.57 - 0.98) | 0.05 |
| VA | 969 | 25,382 | 0.7 | 18.3 | 16 | 797 | 0.53 (0.28 - 0.91) | 0.03 | 0.41 (0.23 - 0.74) | <0.01 |
| Meta-analysis | 134,287 | 930,517 | 90.7 | 910.1 | 2,051 | 33,014 | 0.79 (0.75 - 0.84) | <0.01 | 0.78 (0.66 - 0.93) | <0.01 |
| Meta-analysis (w/o USOC) | 23,463 | 213,343 | 12.2 | 157.5 | 489 | 8,586 | 0.81 (0.72 - 0.91) | <0.01 | 0.77 (0.68 - 0.88) | <0.01 |
| **PS-stratification / on-treatment TAR** | | | | | | | | | | |
| CCAE | 10,734 | 78,888 | 6.8 | 65.5 | 244 | 2,707 | 0.81 (0.70 - 0.93) | <0.01 | 0.78 (0.68 - 0.91) | <0.01 |
| ODOD | 6,702 | 57,013 | 3.8 | 49.9 | 185 | 3,814 | 0.74 (0.63 - 0.87) | <0.01 | 0.70 (0.60 - 0.83) | <0.01 |
| MDCR | 772 | 15,931 | 0.5 | 16.0 | 32 | 1,343 | 0.91 (0.61 - 1.29) | 0.60 | 0.84 (0.58 - 1.23) | 0.38 |
| Open Claims | 122,153 | 741,055 | 83.8 | 779.2 | 1,666 | 26,510 | 0.75 (0.71 - 0.79) | <0.01 | 0.76 (0.63 - 0.92) | <0.01 |
| OEHR | 7,667 | 83,664 | 2.1 | 43.6 | 79 | 2,420 | 0.82 (0.64 - 1.03) | 0.09 | 0.75 (0.59 - 0.96) | 0.03 |
| VA | 1,030 | 195,946 | 0.7 | 198.7 | 16 | 7,866 | 0.49 (0.29 - 0.78) | <0.01 | 0.42 (0.25 - 0.69) | <0.01 |
| Meta-analysis | 147,256 | 960,620 | 96.5 | 938.1 | 2,174 | 35,451 | 0.76 (0.72 - 0.80) | <0.01 | 0.75 (0.61 - 0.92) | <0.01 |
| Meta-analysis (w/o USOC) | 25,103 | 219,565 | 12.7 | 158.9 | 508 | 8,941 | 0.78 (0.71 - 0.86) | <0.01 | 0.74 (0.65 - 0.85) | <0.01 |
| **PS-matching / on-treatment TAR without escalation** | | | | | | | | | | |
| CCAE | 9,596 | 75,118 | 5.5 | 54.5 | 179 | 2,212 | 0.80 (0.66 - 0.98) | 0.03 | 0.78 (0.64 - 0.95) | 0.02 |
| ODOD | 6,069 | 50,874 | 3.1 | 39.0 | 153 | 2,721 | 0.85 (0.69 - 1.03) | 0.11 | 0.80 (0.65 - 0.98) | 0.04 |
| MDCR | 718 | 12,217 | 0.4 | 11.4 | 26 | 864 | 1.00 (0.60 - 1.58) | 0.99 | 0.99 (0.61 - 1.60) | 0.82 |
| Open Claims | 110,824 | 717,174 | 68.3 | 641.4 | 1,346 | 20,754 | 0.77 (0.72 - 0.83) | <0.01 | 0.78 (0.66 - 0.90) | <0.01 |
| OEHR | 7,080 | 75,134 | 1.7 | 34.4 | 62 | 1,624 | 0.79 (0.57 - 1.06) | 0.13 | 0.74 (0.54 - 1.00) | 0.06 |
| VA | 969 | 25,382 | 0.5 | 16.2 | 12 | 697 | 0.60 (0.29 - 1.11) | 0.14 | 0.45 (0.23 - 0.88) | 0.02 |
| Meta-analysis | 134,287 | 930,517 | 79.1 | 780.8 | 1,766 | 28,175 | 0.79 (0.74 - 0.84) | <0.01 | 0.77 (0.65 - 0.92) | <0.01 |
| Meta-analysis (w/o USOC) | 23,463 | 213,343 | 10.8 | 139.4 | 420 | 7,421 | 0.83 (0.73 - 0.94) | <0.01 | 0.79 (0.70 - 0.90) | <0.01 |
| **PS-stratification / on-treatment TAR without escalation** | | | | | | | | | | |
| CCAE | 10,734 | 78,888 | 6.0 | 57.2 | 207 | 2,366 | 0.79 (0.67 - 0.93) | <0.01 | 0.77 (0.66 - 0.91) | <0.01 |
| ODOD | 6,702 | 57,013 | 3.4 | 44.9 | 168 | 3,392 | 0.75 (0.63 - 0.89) | <0.01 | 0.70 (0.59 - 0.83) | <0.01 |
| MDCR | 772 | 15,931 | 0.4 | 14.7 | 28 | 1,224 | 0.92 (0.60 - 1.35) | 0.69 | 0.84 (0.56 - 1.26) | 0.42 |
| Open Claims | 122,153 | 741,055 | 73.3 | 664.6 | 1,445 | 22,577 | 0.76 (0.72 - 0.81) | <0.01 | 0.76 (0.63 - 0.92) | <0.01 |
| OEHR | 7,667 | 83,664 | 1.9 | 38.4 | 65 | 1,940 | 0.82 (0.63 - 1.06) | 0.15 | 0.75 (0.57 - 0.97) | 0.05 |
| VA | 1,030 | 195,946 | 0.6 | 178.7 | 12 | 6,851 | 0.48 (0.26 - 0.81) | 0.01 | 0.38 (0.22 - 0.68) | <0.01 |
| Meta-analysis | 147,256 | 960,620 | 84.6 | 805.1 | 1,885 | 30,275 | 0.76 (0.73 - 0.80) | <0.01 | 0.75 (0.61 - 0.92) | <0.01 |
| Meta-analysis (w/o USOC) | 25,103 | 219,565 | 11.2 | 140.6 | 440 | 7,698 | 0.78 (0.70 - 0.87) | <0.01 | 0.74 (0.62 - 0.87) | <0.01 |

##### 1.5.24 Photosensitivity

**Supplementary Table 128:** Exposures, outcomes and risk estimates for Photosensitivity in GLP1RA (male) (T) vs SU (male) (C) new-user cohorts under four different propensity score (PS) adjustment / time-at-risk (TAR) designs across data sources. We report total patient exposure time in 1,000 patient-year (KYr) units, uncalibrated and calibrated hazard ratios (HRs) and their 95% confidence intervals (CIs). Greyed out entries did not pass study diagnostics and are included only for completeness.

| | Patients | | Exposure (KYr) | | Outcomes | | Uncalibrated | | Calibrated | |
|---|---|---|---|---|---|---|---|---|---|---|
| | T | C | T | C | T | C | HR (95% CI) | $p$ | HR (95% CI) | $p$ |
| **PS-matching / on-treatment TAR** | | | | | | | | | | |
| CCAE | 11,381 | 85,596 | 7.6 | 73.8 | 11 | 101 | 2.14 (0.89 - 4.77) | 0.08 | 2.08 (0.90 - 4.82) | 0.09 |
| ODOD | 7,555 | 62,423 | 4.4 | 56.9 | <5 | 96 | 0.51 (0.08 - 1.87) | 0.41 | 0.49 (0.10 - 2.38) | 0.38 |
| MDCR | 915 | 15,256 | 0.6 | 16.7 | <5 | 57 | 0.97 (0.15 - 3.71) | 0.97 | 0.95 (0.19 - 4.77) | 0.94 |
| Open Claims | 138,800 | 866,440 | 98.7 | 937.8 | 29 | 434 | 1.16 (0.70 - 1.84) | 0.56 | 1.18 (0.71 - 1.95) | 0.53 |
| OEHR | 8,315 | 85,941 | 2.4 | 47.3 | 5 | 86 | 2.18 (0.69 - 5.69) | 0.15 | 2.04 (0.71 - 5.84) | 0.19 |
| VA | 1,290 | 31,833 | 0.9 | 23.9 | 0 | 13 | | | | |
| Meta-analysis | 166,051 | 1,100,400 | 113.1 | 1115.8 | <50 | 717 | 1.38 (0.90 - 2.12) | 0.14 | 1.37 (0.87 - 2.17) | 0.17 |
| Meta-analysis (w/o USOC) | 27,251 | 233,960 | 14.5 | 178.0 | <21 | 283 | 1.66 (0.80 - 3.45) | 0.17 | 1.59 (0.76 - 3.31) | 0.22 |
| **PS-stratification / on-treatment TAR** | | | | | | | | | | |
| CCAE | 12,891 | 89,977 | 8.4 | 77.3 | 11 | 104 | 1.57 (0.75 - 2.99) | 0.20 | 1.52 (0.76 - 3.04) | 0.24 |
| ODOD | 8,466 | 70,086 | 4.8 | 65.7 | <5 | 120 | 0.86 (0.24 - 2.30) | 0.79 | 0.82 (0.27 - 2.52) | 0.73 |
| MDCR | 991 | 19,987 | 0.7 | 21.4 | <5 | 83 | 0.85 (0.13 - 3.04) | 0.84 | 0.79 (0.17 - 3.79) | 0.77 |
| Open Claims | 154,117 | 895,273 | 106.0 | 970.9 | 32 | 484 | 1.25 (0.82 - 1.83) | 0.28 | 1.26 (0.82 - 1.96) | 0.29 |
| OEHR | 9,044 | 95,253 | 2.6 | 52.9 | 5 | 110 | 1.81 (0.60 - 4.35) | 0.24 | 1.67 (0.62 - 4.50) | 0.31 |
| VA | 1,390 | 236,875 | 1.0 | 252.2 | 0 | 376 | | | | |
| Meta-analysis | 184,518 | 1,150,589 | 121.8 | 1166.7 | <53 | 818 | 1.32 (0.96 - 1.80) | 0.08 | 1.30 (0.90 - 1.89) | 0.16 |
| Meta-analysis (w/o USOC) | 30,401 | 255,316 | 15.8 | 195.8 | <21 | 334 | 1.44 (0.87 - 2.39) | 0.16 | 1.37 (0.82 - 2.30) | 0.24 |
| **PS-matching / on-treatment TAR without escalation** | | | | | | | | | | |
| CCAE | 11,381 | 85,596 | 6.6 | 63.9 | 10 | 92 | 2.68 (1.09 - 6.18) | 0.03 | 2.60 (1.09 - 6.21) | 0.03 |
| ODOD | 7,555 | 62,423 | 3.9 | 50.6 | <5 | 90 | 0.50 (0.08 - 1.87) | 0.40 | 0.47 (0.10 - 2.34) | 0.36 |
| MDCR | 915 | 15,256 | 0.6 | 15.1 | <5 | 51 | 1.06 (0.16 - 4.17) | 0.95 | 1.04 (0.20 - 5.36) | 0.94 |
| Open Claims | 138,800 | 866,440 | 85.7 | 793.6 | 27 | 393 | 1.19 (0.70 - 1.94) | 0.50 | 1.20 (0.71 - 2.03) | 0.51 |
| OEHR | 8,315 | 85,941 | 2.1 | 41.2 | <5 | 76 | 1.31 (0.29 - 4.07) | 0.69 | 1.22 (0.33 - 4.54) | 0.77 |
| VA | 1,290 | 31,833 | 0.7 | 21.0 | 0 | <10 | | | | |
| Meta-analysis | 166,051 | 1,100,400 | 98.4 | 949.2 | <47 | 651 | 1.38 (0.82 - 2.31) | 0.23 | 1.35 (0.79 - 2.33) | 0.28 |
| Meta-analysis (w/o USOC) | 27,251 | 233,960 | 12.6 | 155.6 | <20 | 258 | 1.45 (0.58 - 3.68) | 0.43 | 1.39 (0.55 - 3.52) | 0.49 |
| **PS-stratification / on-treatment TAR without escalation** | | | | | | | | | | |
| CCAE | 12,891 | 89,977 | 7.3 | 67.0 | 10 | 95 | 1.74 (0.79 - 3.44) | 0.14 | 1.69 (0.81 - 3.53) | 0.16 |
| ODOD | 8,466 | 70,086 | 4.3 | 58.5 | <5 | 112 | 0.88 (0.25 - 2.38) | 0.83 | 0.82 (0.27 - 2.54) | 0.73 |
| MDCR | 991 | 19,987 | 0.6 | 19.5 | <5 | 77 | 0.93 (0.14 - 3.42) | 0.93 | 0.85 (0.17 - 4.17) | 0.85 |
| Open Claims | 154,117 | 895,273 | 92.6 | 822.1 | 30 | 443 | 1.35 (0.87 - 2.01) | 0.16 | 1.35 (0.86 - 2.12) | 0.20 |
| OEHR | 9,044 | 95,253 | 2.2 | 45.9 | <5 | 98 | 1.18 (0.27 - 3.49) | 0.80 | 1.07 (0.30 - 3.85) | 0.92 |
| VA | 1,390 | 236,875 | 0.8 | 224.6 | 0 | 344 | | | | |
| Meta-analysis | 184,518 | 1,150,589 | 106.4 | 993.5 | <50 | 748 | 1.36 (0.97 - 1.89) | 0.07 | 1.33 (0.90 - 1.95) | 0.15 |
| Meta-analysis (w/o USOC) | 30,401 | 255,316 | 13.9 | 171.5 | <20 | 305 | 1.37 (0.79 - 2.39) | 0.26 | 1.29 (0.73 - 2.29) | 0.38 |

##### 1.5.25 Venous thromboembolism

**Supplementary Table 129:** Exposures, outcomes and risk estimates for Venous thromboembolism in GLP1RA (male) (T) vs SU (male) (C) new-user cohorts under four different propensity score (PS) adjustment / time-at-risk (TAR) designs across data sources. We report total patient exposure time in 1,000 patient-year (KYr) units, uncalibrated and calibrated hazard ratios (HRs) and their 95% confidence intervals (CIs). Greyed out entries did not pass study diagnostics and are included only for completeness.

| | Patients | | Exposure (KYr) | | Outcomes | | Uncalibrated | | Calibrated | |
|---|---|---|---|---|---|---|---|---|---|---|
| | T | C | T | C | T | C | HR (95% CI) | $p$ | HR (95% CI) | $p$ |
| **PS-matching / on-treatment TAR** | | | | | | | | | | |
| CCAE | 11,172 | 84,495 | 7.5 | 72.6 | 38 | 408 | 0.99 (0.62 - 1.52) | 0.96 | 0.96 (0.61 - 1.50) | 0.86 |
| ODOD | 7,358 | 61,099 | 4.3 | 55.4 | 26 | 611 | 0.51 (0.30 - 0.82) | <0.01 | 0.49 (0.29 - 0.81) | <0.01 |
| MDCR | 892 | 14,897 | 0.6 | 16.2 | 9 | 226 | 0.99 (0.41 - 2.07) | 0.97 | 0.96 (0.43 - 2.16) | 0.89 |
| Open Claims | 134,797 | 845,418 | 95.9 | 912.8 | 348 | 5,254 | 0.75 (0.65 - 0.85) | <0.01 | 0.76 (0.62 - 0.92) | <0.01 |
| OEHR | 8,157 | 84,240 | 2.4 | 45.9 | 19 | 421 | 0.87 (0.48 - 1.47) | 0.63 | 0.81 (0.47 - 1.42) | 0.48 |
| VA | 1,255 | 31,500 | 0.9 | 23.6 | <10 | 142 | 1.55 (0.59 - 3.52) | 0.34 | 1.20 (0.49 - 2.94) | 0.69 |
| Meta-analysis | 162,376 | 1,090,149 | 110.7 | 1103.0 | 440 | 6,920 | 0.76 (0.67 - 0.85) | <0.01 | 0.75 (0.61 - 0.92) | <0.01 |
| Meta-analysis (w/o USOC) | 27,579 | 244,731 | 14.8 | 190.2 | 92 | 1,666 | 0.80 (0.57 - 1.10) | 0.17 | 0.76 (0.55 - 1.06) | 0.12 |
| **PS-stratification / on-treatment TAR** | | | | | | | | | | |
| CCAE | 12,632 | 88,756 | 8.2 | 76.1 | 43 | 443 | 0.84 (0.59 - 1.18) | 0.33 | 0.81 (0.57 - 1.16) | 0.27 |
| ODOD | 8,239 | 68,583 | 4.7 | 63.9 | 27 | 787 | 0.55 (0.35 - 0.81) | <0.01 | 0.52 (0.34 - 0.79) | <0.01 |
| MDCR | 962 | 19,507 | 0.6 | 20.8 | 9 | 312 | 1.08 (0.49 - 2.09) | 0.83 | 1.00 (0.49 - 2.07) | 0.90 |
| Open Claims | 149,623 | 873,663 | 103.1 | 945.2 | 370 | 5,765 | 0.73 (0.65 - 0.82) | <0.01 | 0.74 (0.60 - 0.92) | <0.01 |
| OEHR | 8,858 | 93,438 | 2.5 | 51.4 | 19 | 501 | 0.90 (0.54 - 1.43) | 0.68 | 0.83 (0.51 - 1.36) | 0.48 |
| VA | 1,345 | 233,342 | 0.9 | 247.7 | <10 | 1,323 | 1.23 (0.52 - 2.44) | 0.60 | 1.04 (0.48 - 2.26) | 0.90 |
| Meta-analysis | 179,352 | 1,124,440 | 118.5 | 1136.5 | 459 | 7,496 | 0.74 (0.66 - 0.82) | <0.01 | 0.73 (0.58 - 0.91) | <0.01 |
| Meta-analysis (w/o USOC) | 29,729 | 250,777 | 15.4 | 191.3 | 89 | 1,731 | 0.74 (0.55 - 1.00) | 0.05 | 0.71 (0.51 - 0.97) | 0.05 |
| **PS-matching / on-treatment TAR without escalation** | | | | | | | | | | |
| CCAE | 11,172 | 84,495 | 6.5 | 63.0 | 35 | 340 | 0.98 (0.60 - 1.55) | 0.93 | 0.95 (0.59 - 1.53) | 0.84 |
| ODOD | 7,358 | 61,099 | 3.8 | 49.3 | 23 | 529 | 0.60 (0.35 - 0.98) | 0.06 | 0.57 (0.34 - 0.96) | 0.04 |
| MDCR | 892 | 14,897 | 0.5 | 14.7 | 5 | 200 | 0.55 (0.16 - 1.41) | 0.28 | 0.54 (0.18 - 1.61) | 0.27 |
| Open Claims | 134,797 | 845,418 | 83.4 | 773.2 | 305 | 4,468 | 0.78 (0.67 - 0.90) | <0.01 | 0.78 (0.64 - 0.96) | 0.02 |
| OEHR | 8,157 | 84,240 | 2.1 | 40.1 | 15 | 316 | 0.93 (0.48 - 1.67) | 0.82 | 0.87 (0.47 - 1.62) | 0.66 |
| VA | 1,255 | 31,500 | 0.7 | 20.7 | <10 | 114 | 1.90 (0.69 - 4.57) | 0.18 | 1.42 (0.55 - 3.66) | 0.47 |
| Meta-analysis | 162,376 | 1,090,149 | 96.3 | 940.2 | 383 | 5,853 | 0.78 (0.68 - 0.89) | <0.01 | 0.77 (0.62 - 0.95) | 0.01 |
| Meta-analysis (w/o USOC) | 27,579 | 244,731 | 12.9 | 167.0 | 78 | 1,385 | 0.79 (0.59 - 1.06) | 0.12 | 0.76 (0.57 - 1.02) | 0.09 |
| **PS-stratification / on-treatment TAR without escalation** | | | | | | | | | | |
| CCAE | 12,632 | 88,756 | 7.2 | 66.1 | 40 | 370 | 0.98 (0.67 - 1.40) | 0.92 | 0.96 (0.66 - 1.38) | 0.82 |
| ODOD | 8,239 | 68,583 | 4.2 | 57.0 | 24 | 691 | 0.56 (0.35 - 0.85) | <0.01 | 0.52 (0.33 - 0.81) | <0.01 |
| MDCR | 962 | 19,507 | 0.6 | 18.9 | 5 | 282 | 0.62 (0.21 - 1.42) | 0.32 | 0.56 (0.22 - 1.46) | 0.24 |
| Open Claims | 149,623 | 873,663 | 90.0 | 801.0 | 324 | 4,927 | 0.75 (0.66 - 0.85) | <0.01 | 0.75 (0.60 - 0.94) | 0.01 |
| OEHR | 8,858 | 93,438 | 2.2 | 44.7 | 15 | 370 | 0.92 (0.51 - 1.55) | 0.78 | 0.84 (0.48 - 1.46) | 0.54 |
| VA | 1,345 | 233,342 | 0.7 | 220.8 | <10 | 1,087 | 1.74 (0.73 - 3.50) | 0.16 | 1.39 (0.64 - 3.04) | 0.42 |
| Meta-analysis | 179,352 | 1,124,440 | 103.6 | 968.8 | 403 | 6,358 | 0.76 (0.68 - 0.86) | <0.01 | 0.75 (0.59 - 0.94) | 0.01 |
| Meta-analysis (w/o USOC) | 29,729 | 250,777 | 13.6 | 167.8 | 79 | 1,431 | 0.80 (0.55 - 1.15) | 0.23 | 0.75 (0.51 - 1.11) | 0.17 |

##### 1.5.26 Vomiting

**Supplementary Table 130:** Exposures, outcomes and risk estimates for Vomiting in GLP1RA (male) (T) vs SU (male) (C) new-user cohorts under four different propensity score (PS) adjustment / time-at-risk (TAR) designs across data sources. We report total patient exposure time in 1,000 patient-year (KYr) units, uncalibrated and calibrated hazard ratios (HRs) and their 95% confidence intervals (CIs). Greyed out entries did not pass study diagnostics and are included only for completeness.

| | Patients | | Exposure (KYr) | | Outcomes | | Uncalibrated | | Calibrated | |
|---|---|---|---|---|---|---|---|---|---|---|
| | T | C | T | C | T | C | HR (95% CI) | $p$ | HR (95% CI) | $p$ |
| **PS-matching / on-treatment TAR** | | | | | | | | | | |
| CCAE | 10,715 | 81,419 | 7.2 | 69.9 | 161 | 1,000 | 1.55 (1.24 - 1.94) | <0.01 | 1.51 (1.20 - 1.89) | <0.01 |
| ODOD | 7,003 | 58,306 | 4.0 | 52.6 | 116 | 1,150 | 1.34 (1.04 - 1.71) | 0.02 | 1.28 (0.99 - 1.64) | 0.07 |
| MDCR | 889 | 14,632 | 0.6 | 15.9 | 12 | 356 | 0.79 (0.36 - 1.54) | 0.52 | 0.77 (0.37 - 1.60) | 0.49 |
| Open Claims | 128,040 | 808,320 | 91.3 | 877.2 | 1,378 | 10,087 | 1.48 (1.37 - 1.60) | <0.01 | 1.50 (1.28 - 1.76) | <0.01 |
| OEHR | 7,894 | 82,571 | 2.3 | 44.7 | 85 | 833 | 2.03 (1.51 - 2.70) | <0.01 | 1.89 (1.41 - 2.54) | <0.01 |
| VA | 1,250 | 30,824 | 0.9 | 23.1 | 14 | 167 | 2.00 (0.93 - 3.95) | 0.06 | 1.55 (0.75 - 3.20) | 0.24 |
| Meta-analysis | 154,541 | 1,045,248 | 105.3 | 1060.3 | 1,752 | 13,426 | 1.49 (1.40 - 1.60) | <0.01 | 1.48 (1.24 - 1.76) | <0.01 |
| Meta-analysis (w/o USOC) | 26,501 | 236,928 | 14.1 | 183.1 | 374 | 3,339 | 1.50 (1.16 - 1.93) | <0.01 | 1.43 (1.11 - 1.85) | <0.01 |
| **PS-stratification / on-treatment TAR** | | | | | | | | | | |
| CCAE | 12,118 | 85,512 | 7.8 | 73.2 | 180 | 1,068 | 1.38 (1.15 - 1.65) | <0.01 | 1.34 (1.11 - 1.61) | <0.01 |
| ODOD | 7,839 | 65,485 | 4.4 | 60.9 | 125 | 1,409 | 1.30 (1.05 - 1.60) | 0.02 | 1.24 (1.00 - 1.53) | 0.07 |
| MDCR | 960 | 19,051 | 0.6 | 20.3 | 12 | 494 | 0.84 (0.43 - 1.47) | 0.56 | 0.78 (0.42 - 1.43) | 0.42 |
| Open Claims | 141,956 | 835,989 | 97.9 | 909.2 | 1,492 | 10,876 | 1.42 (1.33 - 1.51) | <0.01 | 1.44 (1.19 - 1.74) | <0.01 |
| OEHR | 8,595 | 91,557 | 2.4 | 50.0 | 90 | 954 | 2.08 (1.63 - 2.62) | <0.01 | 1.92 (1.50 - 2.44) | <0.01 |
| VA | 1,349 | 229,643 | 0.9 | 244.3 | 14 | 1,751 | 2.10 (1.16 - 3.52) | <0.01 | 1.78 (1.02 - 3.10) | 0.05 |
| Meta-analysis | 170,508 | 1,078,543 | 112.6 | 1093.4 | 1,887 | 14,307 | 1.50 (1.25 - 1.79) | <0.01 | 1.48 (1.13 - 1.93) | <0.01 |
| Meta-analysis (w/o USOC) | 28,552 | 242,554 | 14.7 | 184.2 | 395 | 3,431 | 1.54 (1.18 - 2.01) | <0.01 | 1.46 (1.10 - 1.94) | 0.02 |
| **PS-matching / on-treatment TAR without escalation** | | | | | | | | | | |
| CCAE | 10,715 | 81,419 | 6.2 | 60.7 | 145 | 877 | 1.65 (1.30 - 2.08) | <0.01 | 1.60 (1.26 - 2.03) | <0.01 |
| ODOD | 7,003 | 58,306 | 3.6 | 46.9 | 106 | 1,002 | 1.42 (1.09 - 1.85) | <0.01 | 1.35 (1.03 - 1.76) | 0.04 |
| MDCR | 889 | 14,632 | 0.5 | 14.4 | 8 | 309 | 0.49 (0.17 - 1.15) | 0.15 | 0.49 (0.19 - 1.28) | 0.15 |
| Open Claims | 128,040 | 808,320 | 79.5 | 744.5 | 1,183 | 8,312 | 1.50 (1.38 - 1.62) | <0.01 | 1.50 (1.28 - 1.76) | <0.01 |
| OEHR | 7,894 | 82,571 | 2.0 | 39.1 | 69 | 632 | 1.86 (1.33 - 2.56) | <0.01 | 1.74 (1.25 - 2.41) | <0.01 |
| VA | 1,250 | 30,824 | 0.7 | 20.2 | 13 | 133 | 2.29 (0.92 - 5.10) | 0.06 | 1.71 (0.72 - 4.04) | 0.23 |
| Meta-analysis | 154,541 | 1,045,248 | 91.8 | 905.7 | 1,511 | 11,132 | 1.51 (1.40 - 1.62) | <0.01 | 1.49 (1.24 - 1.78) | <0.01 |
| Meta-analysis (w/o USOC) | 26,501 | 236,928 | 12.3 | 161.1 | 328 | 2,820 | 1.50 (1.15 - 1.96) | <0.01 | 1.44 (1.10 - 1.88) | 0.01 |
| **PS-stratification / on-treatment TAR without escalation** | | | | | | | | | | |
| CCAE | 12,118 | 85,512 | 6.9 | 63.7 | 163 | 938 | 1.41 (1.16 - 1.71) | <0.01 | 1.38 (1.14 - 1.68) | <0.01 |
| ODOD | 7,839 | 65,485 | 3.9 | 54.4 | 114 | 1,234 | 1.37 (1.09 - 1.70) | <0.01 | 1.28 (1.02 - 1.60) | 0.05 |
| MDCR | 960 | 19,051 | 0.6 | 18.5 | 8 | 433 | 0.63 (0.28 - 1.22) | 0.22 | 0.57 (0.27 - 1.21) | 0.15 |
| Open Claims | 141,956 | 835,989 | 85.7 | 772.2 | 1,293 | 8,987 | 1.46 (1.37 - 1.57) | <0.01 | 1.46 (1.20 - 1.78) | <0.01 |
| OEHR | 8,595 | 91,557 | 2.1 | 43.7 | 74 | 718 | 2.22 (1.68 - 2.88) | <0.01 | 2.01 (1.53 - 2.63) | <0.01 |
| VA | 1,349 | 229,643 | 0.7 | 217.9 | 13 | 1,469 | 2.63 (1.41 - 4.51) | <0.01 | 2.10 (1.17 - 3.76) | 0.01 |
| Meta-analysis | 170,508 | 1,078,543 | 98.6 | 934.1 | 1,644 | 11,877 | 1.55 (1.29 - 1.87) | <0.01 | 1.52 (1.16 - 1.99) | <0.01 |
| Meta-analysis (w/o USOC) | 28,552 | 242,554 | 12.9 | 161.9 | 351 | 2,890 | 1.61 (1.22 - 2.12) | <0.01 | 1.51 (1.11 - 2.06) | 0.01 |

#### 1.6 DPP4I (male) vs SU (male)

##### 1.6.1 3-point MACE

**Supplementary Table 131:** Exposures, outcomes and risk estimates for 3-point MACE in DPP4I (male) (T) vs SU (male) (C) new-user cohorts under four different propensity score (PS) adjustment / time-at-risk (TAR) designs across data sources. We report total patient exposure time in 1,000 patient-year (KYr) units, uncalibrated and calibrated hazard ratios (HRs) and their 95% confidence intervals (CIs). Greyed out entries did not pass study diagnostics and are included only for completeness.

| | Patients | | Exposure (KYr) | | Outcomes | | Uncalibrated | | Calibrated | |
|---|---|---|---|---|---|---|---|---|---|---|
| | T | C | T | C | T | C | HR (95% CI) | $p$ | HR (95% CI) | $p$ |
| **PS-matching / on-treatment TAR** | | | | | | | | | | |
| CCAE | 45,796 | 81,969 | 39.9 | 69.6 | 354 | 722 | 0.87 (0.73 - 1.04) | 0.13 | 0.88 (0.74 - 1.06) | 0.21 |
| DAG | 13,419 | 2,361 | 14.3 | 2.4 | 0 | 0 | | | | |
| ODOD | 27,622 | 66,613 | 23.1 | 61.8 | 442 | 1,567 | 0.85 (0.73 - 0.98) | 0.02 | 0.88 (0.76 - 1.02) | 0.12 |
| MDCR | 8,273 | 16,421 | 8.6 | 17.1 | 247 | 571 | 0.88 (0.71 - 1.09) | 0.25 | 0.87 (0.70 - 1.08) | 0.21 |
| MDCD | 3,727 | 8,994 | 2.8 | 6.4 | 65 | 195 | 0.84 (0.56 - 1.23) | 0.37 | 0.87 (0.59 - 1.29) | 0.51 |
| Open Claims | 395,491 | 843,428 | 376.8 | 911.1 | 4,098 | 12,752 | 0.81 (0.77 - 0.85) | <0.01 | 0.81 (0.71 - 0.91) | <0.01 |
| OEHR | 35,578 | 93,394 | 17.5 | 51.3 | 194 | 749 | 1.08 (0.87 - 1.33) | 0.48 | 1.15 (0.90 - 1.47) | 0.32 |
| VA | 0 | 0 | 0.0 | 0.0 | 0 | 0 | | | | |
| SIDIAP | 25,996 | 7,090 | 75.8 | 20.0 | 375 | 72 | 1.10 (0.80 - 1.54) | 0.57 | 1.05 (0.75 - 1.46) | 0.80 |
| Meta-analysis | 542,483 | 1,117,909 | 544.6 | 1137.4 | 5,775 | 16,628 | 0.88 (0.80 - 0.96) | <0.01 | 0.88 (0.77 - 1.01) | 0.07 |
| Meta-analysis (w/o USOC) | 146,992 | 274,481 | 167.7 | 226.2 | 1,677 | 3,876 | 0.91 (0.83 - 0.99) | 0.03 | 0.93 (0.83 - 1.04) | 0.28 |
| **PS-stratification / on-treatment TAR** | | | | | | | | | | |
| CCAE | 51,105 | 83,416 | 44.7 | 70.7 | 383 | 728 | 0.87 (0.77 - 0.99) | 0.04 | 0.88 (0.77 - 1.01) | 0.08 |
| DAG | 13,447 | 2,736 | 14.3 | 2.8 | 0 | 0 | | | | |
| ODOD | 29,200 | 66,618 | 24.5 | 61.8 | 458 | 1,568 | 0.85 (0.76 - 0.94) | <0.01 | 0.86 (0.76 - 0.96) | 0.01 |
| MDCR | 8,780 | 16,871 | 9.2 | 17.6 | 259 | 589 | 0.92 (0.79 - 1.07) | 0.28 | 0.93 (0.80 - 1.09) | 0.39 |
| MDCD | 3,809 | 9,012 | 2.9 | 6.4 | 67 | 195 | 0.81 (0.61 - 1.08) | 0.17 | 0.84 (0.62 - 1.12) | 0.25 |
| Open Claims | 402,801 | 843,441 | 386.0 | 911.2 | 4,214 | 12,752 | 0.82 (0.79 - 0.85) | <0.01 | 0.82 (0.70 - 0.95) | <0.01 |
| OEHR | 37,370 | 93,423 | 18.6 | 51.3 | 209 | 750 | 0.92 (0.78 - 1.08) | 0.33 | 1.01 (0.86 - 1.19) | 0.85 |
| VA | 24,642 | 224,563 | 23.0 | 237.8 | 124 | 2,007 | 0.63 (0.53 - 0.76) | <0.01 | 0.65 (0.22 - 1.98) | 0.46 |
| SIDIAP | 26,076 | 9,118 | 75.9 | 27.1 | 376 | 92 | 1.12 (0.87 - 1.46) | 0.39 | 1.10 (0.85 - 1.43) | 0.49 |
| Meta-analysis | 559,141 | 1,121,899 | 561.7 | 1146.1 | 5,966 | 16,674 | 0.86 (0.82 - 0.91) | <0.01 | 0.87 (0.79 - 0.97) | <0.01 |
| Meta-analysis (w/o USOC) | 156,340 | 278,458 | 175.8 | 235.0 | 1,752 | 3,922 | 0.89 (0.84 - 0.95) | <0.01 | 0.91 (0.82 - 1.00) | 0.07 |
| **PS-matching / on-treatment TAR without escalation** | | | | | | | | | | |
| CCAE | 45,796 | 81,969 | 32.2 | 60.5 | 281 | 632 | 0.83 (0.68 - 1.01) | 0.06 | 0.85 (0.70 - 1.03) | 0.12 |
| DAG | 13,419 | 2,361 | 12.9 | 2.2 | 0 | 0 | | | | |
| ODOD | 27,622 | 66,613 | 18.9 | 55.2 | 377 | 1,401 | 0.88 (0.75 - 1.03) | 0.10 | 0.93 (0.79 - 1.08) | 0.38 |
| MDCR | 8,273 | 16,421 | 7.3 | 15.6 | 206 | 509 | 0.88 (0.70 - 1.10) | 0.27 | 0.88 (0.70 - 1.11) | 0.30 |
| MDCD | 3,727 | 8,994 | 2.2 | 5.4 | 47 | 165 | 0.71 (0.45 - 1.09) | 0.13 | 0.75 (0.48 - 1.16) | 0.21 |
| Open Claims | 395,491 | 843,428 | 292.4 | 772.3 | 3,195 | 10,930 | 0.81 (0.77 - 0.86) | <0.01 | 0.82 (0.73 - 0.92) | <0.01 |
| OEHR | 35,578 | 93,394 | 14.2 | 44.7 | 135 | 547 | 1.05 (0.82 - 1.33) | 0.70 | 1.14 (0.86 - 1.51) | 0.42 |
| VA | 0 | 0 | 0.0 | 0.0 | 0 | 0 | | | | |
| SIDIAP | 25,996 | 7,090 | 55.7 | 16.8 | 260 | 60 | 1.18 (0.81 - 1.73) | 0.40 | 1.09 (0.75 - 1.60) | 0.67 |
| Meta-analysis | 542,483 | 1,117,909 | 422.9 | 970.4 | 4,501 | 14,244 | 0.86 (0.80 - 0.93) | <0.01 | 0.87 (0.76 - 0.99) | 0.04 |
| Meta-analysis (w/o USOC) | 146,992 | 274,481 | 130.5 | 198.2 | 1,306 | 3,314 | 0.90 (0.82 - 0.99) | 0.03 | 0.93 (0.81 - 1.06) | 0.31 |
| **PS-stratification / on-treatment TAR without escalation** | | | | | | | | | | |
| CCAE | 51,105 | 83,416 | 36.2 | 61.6 | 306 | 638 | 0.83 (0.72 - 0.96) | 0.01 | 0.84 (0.73 - 0.98) | 0.03 |
| DAG | 13,447 | 2,736 | 12.9 | 2.6 | 0 | 0 | | | | |
| ODOD | 29,200 | 66,618 | 20.0 | 55.2 | 390 | 1,402 | 0.87 (0.77 - 0.98) | 0.02 | 0.88 (0.78 - 1.00) | 0.08 |
| MDCR | 8,780 | 16,871 | 7.8 | 16.1 | 215 | 524 | 0.91 (0.77 - 1.08) | 0.29 | 0.92 (0.78 - 1.09) | 0.35 |
| MDCD | 3,809 | 9,012 | 2.2 | 5.4 | 49 | 165 | 0.75 (0.53 - 1.03) | 0.09 | 0.76 (0.55 - 1.06) | 0.13 |
| Open Claims | 402,801 | 843,441 | 299.3 | 772.3 | 3,277 | 10,930 | 0.81 (0.78 - 0.84) | <0.01 | 0.81 (0.69 - 0.95) | 0.01 |
| OEHR | 37,370 | 93,423 | 15.1 | 44.7 | 147 | 548 | 0.97 (0.80 - 1.17) | 0.74 | 1.08 (0.88 - 1.31) | 0.49 |
| VA | 24,642 | 224,563 | 18.7 | 212.0 | 86 | 1,719 | 0.56 (0.45 - 0.69) | <0.01 | 0.56 (0.18 - 1.73) | 0.32 |
| SIDIAP | 26,076 | 9,118 | 55.7 | 22.4 | 261 | 76 | 1.01 (0.75 - 1.37) | 0.95 | 0.97 (0.72 - 1.31) | 0.84 |
| Meta-analysis | 559,141 | 1,121,899 | 436.4 | 977.7 | 4,645 | 14,283 | 0.85 (0.80 - 0.90) | <0.01 | 0.86 (0.76 - 0.97) | 0.02 |
| Meta-analysis (w/o USOC) | 156,340 | 278,458 | 137.1 | 205.4 | 1,368 | 3,353 | 0.88 (0.82 - 0.95) | <0.01 | 0.90 (0.81 - 1.01) | 0.09 |

##### 1.6.2 4-point MACE

**Supplementary Table 132:** Exposures, outcomes and risk estimates for 4-point MACE in DPP4I (male) (T) vs SU (male) (C) new-user cohorts under four different propensity score (PS) adjustment / time-at-risk (TAR) designs across data sources. We report total patient exposure time in 1,000 patient-year (KYr) units, uncalibrated and calibrated hazard ratios (HRs) and their 95% confidence intervals (CIs). Greyed out entries did not pass study diagnostics and are included only for completeness.

| | Patients | | Exposure (KYr) | | Outcomes | | Uncalibrated | | Calibrated | |
|---|---|---|---|---|---|---|---|---|---|---|
| | T | C | T | C | T | C | HR (95% CI) | $p$ | HR (95% CI) | $p$ |
| **PS-matching / on-treatment TAR** | | | | | | | | | | |
| CCAE | 45,149 | 80,662 | 39.2 | 68.3 | 514 | 1,061 | 0.88 (0.75 - 1.02) | 0.08 | 0.89 (0.77 - 1.03) | 0.16 |
| DAG | 13,419 | 2,361 | 14.3 | 2.4 | 0 | 0 | | | | |
| ODOD | 26,531 | 63,762 | 22.1 | 58.5 | 649 | 2,459 | 0.77 (0.69 - 0.87) | <0.01 | 0.80 (0.71 - 0.91) | <0.01 |
| MDCR | 7,728 | 15,158 | 7.9 | 15.7 | 386 | 912 | 0.88 (0.74 - 1.04) | 0.14 | 0.87 (0.73 - 1.03) | 0.11 |
| MDCD | 3,462 | 8,289 | 2.6 | 5.8 | 91 | 286 | 0.59 (0.41 - 0.82) | <0.01 | 0.61 (0.43 - 0.87) | <0.01 |
| Open Claims | 382,594 | 813,874 | 363.8 | 875.2 | 6,389 | 19,940 | 0.82 (0.79 - 0.85) | <0.01 | 0.82 (0.72 - 0.93) | <0.01 |
| OEHR | 34,918 | 91,595 | 17.0 | 49.6 | 344 | 1,349 | 0.97 (0.83 - 1.14) | 0.75 | 1.04 (0.85 - 1.27) | 0.75 |
| VA | 0 | 0 | 0.0 | 0.0 | 0 | 0 | | | | |
| SIDIAP | 25,887 | 7,076 | 75.5 | 20.0 | 471 | 95 | 1.05 (0.79 - 1.42) | 0.73 | 1.00 (0.74 - 1.34) | 0.87 |
| Meta-analysis | 526,269 | 1,080,416 | 528.2 | 1093.1 | 8,844 | 26,102 | 0.85 (0.79 - 0.91) | <0.01 | 0.85 (0.75 - 0.97) | 0.01 |
| Meta-analysis (w/o USOC) | 143,675 | 266,542 | 164.4 | 217.8 | 2,455 | 6,162 | 0.86 (0.76 - 0.96) | <0.01 | 0.88 (0.76 - 1.01) | 0.10 |
| **PS-stratification / on-treatment TAR** | | | | | | | | | | |
| CCAE | 50,414 | 82,081 | 44.0 | 69.5 | 553 | 1,070 | 0.87 (0.78 - 0.96) | <0.01 | 0.87 (0.78 - 0.97) | 0.03 |
| DAG | 13,447 | 2,736 | 14.3 | 2.8 | 0 | 0 | | | | |
| ODOD | 28,078 | 63,766 | 23.5 | 58.5 | 680 | 2,460 | 0.81 (0.74 - 0.88) | <0.01 | 0.81 (0.74 - 0.89) | <0.01 |
| MDCR | 8,212 | 15,576 | 8.5 | 16.1 | 407 | 935 | 0.92 (0.81 - 1.04) | 0.18 | 0.93 (0.82 - 1.05) | 0.30 |
| MDCD | 3,536 | 8,304 | 2.7 | 5.9 | 92 | 286 | 0.76 (0.59 - 0.97) | 0.03 | 0.78 (0.61 - 1.00) | 0.06 |
| Open Claims | 389,953 | 813,885 | 372.9 | 875.2 | 6,586 | 19,940 | 0.83 (0.80 - 0.85) | <0.01 | 0.82 (0.71 - 0.95) | 0.01 |
| OEHR | 36,683 | 91,624 | 18.0 | 49.6 | 371 | 1,350 | 0.90 (0.79 - 1.01) | 0.07 | 0.98 (0.87 - 1.11) | 0.80 |
| VA | 24,006 | 218,190 | 22.3 | 229.1 | 268 | 3,973 | 0.69 (0.61 - 0.78) | <0.01 | 0.71 (0.24 - 2.13) | 0.55 |
| SIDIAP | 25,965 | 9,103 | 75.6 | 27.0 | 472 | 123 | 1.05 (0.84 - 1.32) | 0.66 | 1.03 (0.82 - 1.30) | 0.79 |
| Meta-analysis | 542,841 | 1,084,339 | 545.2 | 1101.8 | 9,161 | 26,164 | 0.85 (0.81 - 0.89) | <0.01 | 0.86 (0.78 - 0.94) | <0.01 |
| Meta-analysis (w/o USOC) | 152,888 | 270,454 | 172.3 | 226.5 | 2,575 | 6,224 | 0.87 (0.81 - 0.93) | <0.01 | 0.89 (0.80 - 0.98) | 0.03 |
| **PS-matching / on-treatment TAR without escalation** | | | | | | | | | | |
| CCAE | 45,149 | 80,662 | 31.7 | 59.5 | 409 | 919 | 0.86 (0.73 - 1.01) | 0.07 | 0.88 (0.75 - 1.04) | 0.16 |
| DAG | 13,419 | 2,361 | 12.9 | 2.2 | 0 | 0 | | | | |
| ODOD | 26,531 | 63,762 | 18.1 | 52.2 | 546 | 2,183 | 0.80 (0.70 - 0.91) | <0.01 | 0.84 (0.74 - 0.96) | 0.02 |
| MDCR | 7,728 | 15,158 | 6.8 | 14.3 | 332 | 826 | 0.90 (0.75 - 1.07) | 0.25 | 0.90 (0.75 - 1.08) | 0.29 |
| MDCD | 3,462 | 8,289 | 2.0 | 4.9 | 68 | 240 | 0.59 (0.40 - 0.86) | <0.01 | 0.62 (0.42 - 0.91) | 0.02 |
| Open Claims | 382,594 | 813,874 | 282.5 | 742.6 | 4,974 | 17,036 | 0.82 (0.78 - 0.85) | <0.01 | 0.82 (0.73 - 0.92) | <0.01 |
| OEHR | 34,918 | 91,595 | 13.9 | 43.5 | 247 | 1,044 | 0.94 (0.78 - 1.11) | 0.46 | 1.01 (0.81 - 1.27) | 0.86 |
| VA | 0 | 0 | 0.0 | 0.0 | 0 | 0 | | | | |
| SIDIAP | 25,887 | 7,076 | 55.4 | 16.7 | 330 | 80 | 1.10 (0.79 - 1.55) | 0.59 | 1.02 (0.73 - 1.43) | 0.86 |
| Meta-analysis | 526,269 | 1,080,416 | 410.4 | 933.8 | 6,906 | 22,328 | 0.83 (0.80 - 0.86) | <0.01 | 0.84 (0.75 - 0.94) | <0.01 |
| Meta-analysis (w/o USOC) | 143,675 | 266,542 | 127.9 | 191.2 | 1,932 | 5,292 | 0.86 (0.78 - 0.95) | <0.01 | 0.89 (0.77 - 1.02) | 0.12 |
| **PS-stratification / on-treatment TAR without escalation** | | | | | | | | | | |
| CCAE | 50,414 | 82,081 | 35.6 | 60.6 | 441 | 928 | 0.84 (0.74 - 0.94) | <0.01 | 0.85 (0.75 - 0.96) | 0.02 |
| DAG | 13,447 | 2,736 | 12.9 | 2.6 | 0 | 0 | | | | |
| ODOD | 28,078 | 63,766 | 19.2 | 52.2 | 573 | 2,184 | 0.83 (0.75 - 0.91) | <0.01 | 0.84 (0.76 - 0.93) | <0.01 |
| MDCR | 8,212 | 15,576 | 7.3 | 14.8 | 349 | 847 | 0.92 (0.81 - 1.05) | 0.21 | 0.92 (0.81 - 1.05) | 0.28 |
| MDCD | 3,536 | 8,304 | 2.0 | 4.9 | 69 | 240 | 0.73 (0.55 - 0.95) | 0.02 | 0.74 (0.56 - 0.98) | 0.04 |
| Open Claims | 389,953 | 813,885 | 289.4 | 742.6 | 5,120 | 17,036 | 0.82 (0.79 - 0.85) | <0.01 | 0.81 (0.69 - 0.96) | 0.01 |
| OEHR | 36,683 | 91,624 | 14.8 | 43.5 | 267 | 1,045 | 0.88 (0.76 - 1.01) | 0.08 | 0.98 (0.85 - 1.14) | 0.81 |
| VA | 24,006 | 218,190 | 18.2 | 204.6 | 197 | 3,386 | 0.65 (0.56 - 0.74) | <0.01 | 0.65 (0.22 - 1.98) | 0.46 |
| SIDIAP | 25,965 | 9,103 | 55.5 | 22.3 | 331 | 101 | 0.97 (0.75 - 1.26) | 0.81 | 0.93 (0.72 - 1.21) | 0.62 |
| Meta-analysis | 542,841 | 1,084,339 | 423.8 | 941.0 | 7,150 | 22,381 | 0.83 (0.80 - 0.86) | <0.01 | 0.84 (0.75 - 0.94) | <0.01 |
| Meta-analysis (w/o USOC) | 152,888 | 270,454 | 134.4 | 198.4 | 2,030 | 5,345 | 0.86 (0.81 - 0.91) | <0.01 | 0.88 (0.79 - 0.97) | 0.02 |

##### 1.6.3 Acute myocardial infarction

**Supplementary Table 133:** Exposures, outcomes and risk estimates for Acute myocardial infarction in DPP4I (male) (T) vs SU (male) (C) new-user cohorts under four different propensity score (PS) adjustment / time-at-risk (TAR) designs across data sources. We report total patient exposure time in 1,000 patient-year (KYr) units, uncalibrated and calibrated hazard ratios (HRs) and their 95% confidence intervals (CIs). Greyed out entries did not pass study diagnostics and are included only for completeness.

| | Patients | | Exposure (KYr) | | Outcomes | | Uncalibrated | | Calibrated | |
|---|---|---|---|---|---|---|---|---|---|---|
| | T | C | T | C | T | C | HR (95% CI) | $p$ | HR (95% CI) | $p$ |
| **PS-matching / on-treatment TAR** | | | | | | | | | | |
| CCAE | 46,342 | 83,132 | 40.4 | 70.6 | 215 | 429 | 0.88 (0.70 - 1.11) | 0.28 | 0.89 (0.71 - 1.13) | 0.37 |
| DAG | 13,419 | 2,361 | 14.3 | 2.4 | 0 | 0 | | | | |
| ODOD | 28,348 | 68,425 | 23.8 | 64.0 | 227 | 825 | 0.83 (0.68 - 1.02) | 0.08 | 0.87 (0.71 - 1.06) | 0.19 |
| MDCR | 8,611 | 17,208 | 9.0 | 18.0 | 115 | 280 | 0.85 (0.62 - 1.16) | 0.31 | 0.84 (0.61 - 1.14) | 0.27 |
| MDCD | 3,879 | 9,389 | 3.0 | 6.7 | 34 | 99 | 0.58 (0.32 - 0.99) | 0.06 | 0.60 (0.34 - 1.06) | 0.08 |
| Open Claims | 406,847 | 868,427 | 387.9 | 940.2 | 2,133 | 6,664 | 0.80 (0.75 - 0.86) | <0.01 | 0.80 (0.70 - 0.92) | <0.01 |
| OEHR | 35,898 | 94,369 | 17.7 | 52.1 | 106 | 433 | 0.98 (0.73 - 1.29) | 0.88 | 1.04 (0.77 - 1.42) | 0.80 |
| VA | 0 | 0 | 0.0 | 0.0 | 0 | 0 | | | | |
| SIDIAP | 26,300 | 7,143 | 76.9 | 20.2 | 180 | 42 | 0.83 (0.54 - 1.30) | 0.41 | 0.79 (0.51 - 1.22) | 0.30 |
| Meta-analysis | 556,225 | 1,148,093 | 558.6 | 1171.9 | 3,010 | 8,772 | 0.82 (0.77 - 0.87) | <0.01 | 0.82 (0.73 - 0.93) | <0.01 |
| Meta-analysis (w/o USOC) | 149,378 | 279,666 | 170.7 | 231.7 | 877 | 2,108 | 0.86 (0.76 - 0.96) | <0.01 | 0.88 (0.76 - 1.01) | 0.10 |
| **PS-stratification / on-treatment TAR** | | | | | | | | | | |
| CCAE | 51,682 | 84,598 | 45.2 | 71.9 | 231 | 432 | 0.87 (0.74 - 1.03) | 0.11 | 0.88 (0.74 - 1.04) | 0.15 |
| DAG | 13,447 | 2,736 | 14.3 | 2.8 | 0 | 0 | | | | |
| ODOD | 29,945 | 68,430 | 25.2 | 64.0 | 235 | 825 | 0.82 (0.70 - 0.95) | <0.01 | 0.83 (0.71 - 0.96) | 0.02 |
| MDCR | 9,130 | 17,670 | 9.6 | 18.6 | 120 | 288 | 0.88 (0.70 - 1.10) | 0.26 | 0.89 (0.71 - 1.11) | 0.32 |
| MDCD | 3,971 | 9,409 | 3.0 | 6.7 | 37 | 99 | 0.99 (0.66 - 1.46) | 0.96 | 1.02 (0.68 - 1.52) | 0.91 |
| Open Claims | 414,179 | 868,440 | 397.1 | 940.2 | 2,192 | 6,664 | 0.82 (0.78 - 0.86) | <0.01 | 0.81 (0.70 - 0.95) | <0.01 |
| OEHR | 37,709 | 94,398 | 18.8 | 52.1 | 116 | 434 | 0.87 (0.70 - 1.07) | 0.20 | 0.95 (0.77 - 1.19) | 0.69 |
| VA | 25,018 | 227,520 | 23.4 | 241.4 | 65 | 1,220 | 0.55 (0.42 - 0.70) | <0.01 | 0.57 (0.19 - 1.74) | 0.33 |
| SIDIAP | 26,383 | 9,182 | 76.9 | 27.3 | 181 | 52 | 0.96 (0.68 - 1.38) | 0.82 | 0.94 (0.66 - 1.34) | 0.74 |
| Meta-analysis | 572,999 | 1,152,127 | 575.9 | 1180.8 | 3,112 | 8,794 | 0.83 (0.79 - 0.87) | <0.01 | 0.84 (0.76 - 0.92) | <0.01 |
| Meta-analysis (w/o USOC) | 158,820 | 283,687 | 178.8 | 240.6 | 920 | 2,130 | 0.86 (0.79 - 0.94) | <0.01 | 0.88 (0.79 - 0.99) | 0.04 |
| **PS-matching / on-treatment TAR without escalation** | | | | | | | | | | |
| CCAE | 46,342 | 83,132 | 32.7 | 61.5 | 169 | 370 | 0.85 (0.66 - 1.09) | 0.21 | 0.87 (0.68 - 1.12) | 0.29 |
| DAG | 13,419 | 2,361 | 12.9 | 2.2 | 0 | 0 | | | | |
| ODOD | 28,348 | 68,425 | 19.5 | 57.1 | 184 | 737 | 0.85 (0.69 - 1.06) | 0.15 | 0.90 (0.72 - 1.12) | 0.36 |
| MDCR | 8,611 | 17,208 | 7.7 | 16.4 | 96 | 246 | 0.85 (0.60 - 1.19) | 0.34 | 0.85 (0.60 - 1.20) | 0.36 |
| MDCD | 3,879 | 9,389 | 2.3 | 5.6 | 28 | 82 | 0.63 (0.34 - 1.11) | 0.12 | 0.66 (0.36 - 1.19) | 0.17 |
| Open Claims | 406,847 | 868,427 | 300.8 | 796.8 | 1,643 | 5,681 | 0.82 (0.76 - 0.88) | <0.01 | 0.82 (0.72 - 0.93) | <0.01 |
| OEHR | 35,898 | 94,369 | 14.4 | 45.4 | 70 | 302 | 0.95 (0.68 - 1.31) | 0.78 | 1.03 (0.72 - 1.47) | 0.86 |
| VA | 0 | 0 | 0.0 | 0.0 | 0 | 0 | | | | |
| SIDIAP | 26,300 | 7,143 | 56.4 | 16.9 | 132 | 32 | 0.82 (0.50 - 1.39) | 0.45 | 0.76 (0.46 - 1.27) | 0.31 |
| Meta-analysis | 556,225 | 1,148,093 | 433.7 | 999.7 | 2,322 | 7,450 | 0.82 (0.77 - 0.88) | <0.01 | 0.83 (0.73 - 0.95) | <0.01 |
| Meta-analysis (w/o USOC) | 149,378 | 279,666 | 132.9 | 202.9 | 679 | 1,769 | 0.85 (0.75 - 0.97) | 0.01 | 0.88 (0.75 - 1.03) | 0.13 |
| **PS-stratification / on-treatment TAR without escalation** | | | | | | | | | | |
| CCAE | 51,682 | 84,598 | 36.6 | 62.6 | 182 | 373 | 0.83 (0.69 - 1.00) | 0.05 | 0.85 (0.70 - 1.02) | 0.10 |
| DAG | 13,447 | 2,736 | 12.9 | 2.6 | 0 | 0 | | | | |
| ODOD | 29,945 | 68,430 | 20.6 | 57.1 | 190 | 737 | 0.79 (0.67 - 0.93) | <0.01 | 0.80 (0.68 - 0.95) | 0.02 |
| MDCR | 9,130 | 17,670 | 8.2 | 17.0 | 100 | 252 | 0.89 (0.69 - 1.13) | 0.33 | 0.89 (0.70 - 1.14) | 0.37 |
| MDCD | 3,971 | 9,409 | 2.3 | 5.7 | 31 | 82 | 1.03 (0.66 - 1.57) | 0.89 | 1.05 (0.68 - 1.62) | 0.85 |
| Open Claims | 414,179 | 868,440 | 307.8 | 796.8 | 1,685 | 5,681 | 0.80 (0.76 - 0.85) | <0.01 | 0.80 (0.67 - 0.94) | <0.01 |
| OEHR | 37,709 | 94,398 | 15.3 | 45.4 | 78 | 303 | 0.89 (0.68 - 1.16) | 0.40 | 0.99 (0.76 - 1.30) | 0.91 |
| VA | 25,018 | 227,520 | 19.0 | 215.1 | 50 | 1,037 | 0.54 (0.40 - 0.71) | <0.01 | 0.55 (0.18 - 1.71) | 0.31 |
| SIDIAP | 26,383 | 9,182 | 56.5 | 22.6 | 133 | 40 | 0.97 (0.65 - 1.48) | 0.89 | 0.93 (0.62 - 1.41) | 0.75 |
| Meta-analysis | 572,999 | 1,152,127 | 447.3 | 1007.0 | 2,399 | 7,468 | 0.81 (0.78 - 0.85) | <0.01 | 0.82 (0.73 - 0.93) | <0.01 |
| Meta-analysis (w/o USOC) | 158,820 | 283,687 | 139.5 | 210.2 | 714 | 1,787 | 0.85 (0.77 - 0.94) | <0.01 | 0.87 (0.76 - 0.99) | 0.04 |

##### 1.6.4 Acute renal failure

**Supplementary Table 134:** Exposures, outcomes and risk estimates for Acute renal failure in DPP4I (male) (T) vs SU (male) (C) new-user cohorts under four different propensity score (PS) adjustment / time-at-risk (TAR) designs across data sources. We report total patient exposure time in 1,000 patient-year (KYr) units, uncalibrated and calibrated hazard ratios (HRs) and their 95% confidence intervals (CIs). Greyed out entries did not pass study diagnostics and are included only for completeness.

| | Patients | | Exposure (KYr) | | Outcomes | | Uncalibrated | | Calibrated | |
|---|---|---|---|---|---|---|---|---|---|---|
| | T | C | T | C | T | C | HR (95% CI) | $p$ | HR (95% CI) | $p$ |
| **PS-matching / on-treatment TAR** | | | | | | | | | | |
| CCAE | 46,613 | 83,347 | 40.7 | 71.0 | 345 | 734 | 0.91 (0.76 - 1.09) | 0.30 | 0.92 (0.77 - 1.11) | 0.42 |
| DAG | 13,419 | 2,361 | 14.3 | 2.4 | 0 | 0 | | | | |
| ODOD | 27,580 | 66,579 | 23.0 | 61.5 | 630 | 2,226 | 0.86 (0.76 - 0.98) | 0.02 | 0.90 (0.79 - 1.02) | 0.14 |
| MDCR | 8,428 | 16,789 | 8.7 | 17.5 | 323 | 727 | 0.93 (0.77 - 1.12) | 0.46 | 0.92 (0.76 - 1.11) | 0.39 |
| MDCD | 3,730 | 8,975 | 2.8 | 6.4 | 101 | 277 | 0.81 (0.59 - 1.11) | 0.20 | 0.85 (0.62 - 1.17) | 0.33 |
| Open Claims | 404,418 | 862,829 | 384.6 | 930.8 | 5,819 | 16,972 | 0.89 (0.85 - 0.93) | <0.01 | 0.89 (0.78 - 1.01) | 0.07 |
| OEHR | 35,818 | 94,028 | 17.6 | 51.4 | 329 | 1,132 | 0.91 (0.77 - 1.09) | 0.31 | 0.98 (0.79 - 1.20) | 0.83 |
| VA | 0 | 0 | 0.0 | 0.0 | 0 | 0 | | | | |
| SIDIAP | 26,766 | 7,212 | 78.4 | 20.5 | 34 | <5 | 2.66 (0.82 - 11.9) | 0.15 | 2.52 (0.66 - 9.58) | 0.18 |
| Meta-analysis | 553,353 | 1,139,759 | 555.7 | 1159.2 | 7,581 | <22,073 | 0.89 (0.86 - 0.92) | <0.01 | 0.89 (0.80 - 1.00) | 0.05 |
| Meta-analysis (w/o USOC) | 148,935 | 276,930 | 171.1 | 228.3 | 1,762 | <5,101 | 0.89 (0.83 - 0.97) | <0.01 | 0.91 (0.82 - 1.02) | 0.17 |
| **PS-stratification / on-treatment TAR** | | | | | | | | | | |
| CCAE | 51,990 | 84,940 | 45.6 | 72.3 | 388 | 744 | 0.89 (0.78 - 1.01) | 0.08 | 0.90 (0.78 - 1.02) | 0.13 |
| DAG | 13,447 | 2,736 | 14.3 | 2.8 | 0 | 0 | | | | |
| ODOD | 29,183 | 66,582 | 24.5 | 61.5 | 659 | 2,227 | 0.84 (0.77 - 0.92) | <0.01 | 0.85 (0.78 - 0.94) | <0.01 |
| MDCR | 8,907 | 17,252 | 9.2 | 18.0 | 339 | 741 | 0.97 (0.84 - 1.11) | 0.63 | 0.98 (0.85 - 1.12) | 0.77 |
| MDCD | 3,816 | 8,991 | 2.9 | 6.4 | 103 | 277 | 0.89 (0.70 - 1.12) | 0.33 | 0.91 (0.72 - 1.16) | 0.49 |
| Open Claims | 411,928 | 862,839 | 394.0 | 930.8 | 5,966 | 16,973 | 0.89 (0.86 - 0.92) | <0.01 | 0.88 (0.76 - 1.03) | 0.11 |
| OEHR | 37,636 | 94,058 | 18.6 | 51.5 | 341 | 1,133 | 0.99 (0.87 - 1.13) | 0.92 | 1.09 (0.96 - 1.24) | 0.22 |
| VA | 24,628 | 226,342 | 22.9 | 238.2 | 295 | 3,666 | 0.84 (0.75 - 0.95) | <0.01 | 0.87 (0.29 - 2.61) | 0.81 |
| SIDIAP | 26,859 | 9,265 | 78.5 | 27.6 | 34 | 8 | 1.61 (0.70 - 4.10) | 0.29 | 1.58 (0.65 - 3.82) | 0.32 |
| Meta-analysis | 570,319 | 1,143,927 | 573.3 | 1168.2 | 7,830 | 22,103 | 0.89 (0.87 - 0.92) | <0.01 | 0.90 (0.82 - 0.99) | 0.03 |
| Meta-analysis (w/o USOC) | 158,391 | 281,088 | 179.3 | 237.4 | 1,864 | 5,130 | 0.91 (0.85 - 0.98) | 0.01 | 0.93 (0.84 - 1.03) | 0.21 |
| **PS-matching / on-treatment TAR without escalation** | | | | | | | | | | |
| CCAE | 46,613 | 83,347 | 32.9 | 61.7 | 274 | 625 | 0.94 (0.77 - 1.14) | 0.53 | 0.96 (0.79 - 1.17) | 0.70 |
| DAG | 13,419 | 2,361 | 12.9 | 2.2 | 0 | 0 | | | | |
| ODOD | 27,580 | 66,579 | 18.9 | 55.0 | 506 | 1,985 | 0.89 (0.78 - 1.01) | 0.08 | 0.94 (0.82 - 1.07) | 0.38 |
| MDCR | 8,428 | 16,789 | 7.4 | 16.0 | 269 | 648 | 0.92 (0.75 - 1.13) | 0.42 | 0.92 (0.75 - 1.14) | 0.47 |
| MDCD | 3,730 | 8,975 | 2.1 | 5.4 | 78 | 226 | 0.74 (0.51 - 1.05) | 0.10 | 0.77 (0.54 - 1.11) | 0.18 |
| Open Claims | 404,418 | 862,829 | 298.6 | 789.9 | 4,531 | 14,223 | 0.89 (0.85 - 0.94) | <0.01 | 0.90 (0.80 - 1.00) | 0.07 |
| OEHR | 35,818 | 94,028 | 14.3 | 45.0 | 224 | 833 | 0.92 (0.76 - 1.12) | 0.42 | 1.00 (0.79 - 1.27) | 0.87 |
| VA | 0 | 0 | 0.0 | 0.0 | 0 | 0 | | | | |
| SIDIAP | 26,766 | 7,212 | 57.4 | 17.1 | 22 | <5 | 1.88 (0.42 - 13.2) | 0.47 | 1.75 (0.31 - 9.75) | 0.52 |
| Meta-analysis | 553,353 | 1,139,759 | 431.7 | 990.1 | 5,904 | <18,545 | 0.89 (0.86 - 0.93) | <0.01 | 0.90 (0.81 - 1.02) | 0.09 |
| Meta-analysis (w/o USOC) | 148,935 | 276,930 | 133.1 | 200.2 | 1,373 | <4,322 | 0.90 (0.83 - 0.98) | 0.02 | 0.93 (0.82 - 1.06) | 0.31 |
| **PS-stratification / on-treatment TAR without escalation** | | | | | | | | | | |
| CCAE | 51,990 | 84,940 | 36.9 | 63.0 | 307 | 635 | 0.89 (0.77 - 1.02) | 0.10 | 0.90 (0.78 - 1.04) | 0.20 |
| DAG | 13,447 | 2,736 | 12.9 | 2.6 | 0 | 0 | | | | |
| ODOD | 29,183 | 66,582 | 20.0 | 55.0 | 531 | 1,986 | 0.83 (0.75 - 0.92) | <0.01 | 0.85 (0.76 - 0.94) | <0.01 |
| MDCR | 8,907 | 17,252 | 7.9 | 16.5 | 283 | 661 | 0.97 (0.84 - 1.12) | 0.67 | 0.97 (0.84 - 1.13) | 0.73 |
| MDCD | 3,816 | 8,991 | 2.2 | 5.4 | 78 | 226 | 0.89 (0.68 - 1.16) | 0.40 | 0.90 (0.69 - 1.19) | 0.50 |
| Open Claims | 411,928 | 862,839 | 305.7 | 789.9 | 4,632 | 14,223 | 0.90 (0.87 - 0.93) | <0.01 | 0.90 (0.76 - 1.05) | 0.20 |
| OEHR | 37,636 | 94,058 | 15.2 | 45.0 | 234 | 834 | 0.99 (0.84 - 1.15) | 0.85 | 1.10 (0.94 - 1.29) | 0.31 |
| VA | 24,628 | 226,342 | 18.6 | 212.7 | 212 | 2,979 | 0.81 (0.70 - 0.93) | <0.01 | 0.82 (0.27 - 2.49) | 0.73 |
| SIDIAP | 26,859 | 9,265 | 57.5 | 22.8 | 22 | 7 | 1.51 (0.57 - 4.38) | 0.43 | 1.45 (0.52 - 4.02) | 0.48 |
| Meta-analysis | 570,319 | 1,143,927 | 445.4 | 997.6 | 6,087 | 18,572 | 0.90 (0.87 - 0.93) | <0.01 | 0.91 (0.81 - 1.02) | 0.11 |
| Meta-analysis (w/o USOC) | 158,391 | 281,088 | 139.7 | 207.7 | 1,455 | 4,349 | 0.90 (0.84 - 0.97) | <0.01 | 0.92 (0.83 - 1.03) | 0.18 |

##### 1.6.5 Glycemic control

**Supplementary Table 135:** Exposures, outcomes and risk estimates for Glycemic control in DPP4I (male) (T) vs SU (male) (C) new-user cohorts under four different propensity score (PS) adjustment / time-at-risk (TAR) designs across data sources. We report total patient exposure time in 1,000 patient-year (KYr) units, uncalibrated and calibrated hazard ratios (HRs) and their 95% confidence intervals (CIs). Greyed out entries did not pass study diagnostics and are included only for completeness.

| | Patients | | Exposure (KYr) | | Outcomes | | Uncalibrated | | Calibrated | |
|---|---|---|---|---|---|---|---|---|---|---|
| | T | C | T | C | T | C | HR (95% CI) | $p$ | HR (95% CI) | $p$ |
| **PS-matching / on-treatment TAR** | | | | | | | | | | |
| CCAE | 47,014 | 84,418 | 41.1 | 71.9 | 113 | 318 | 0.98 (0.70 - 1.35) | 0.89 | 0.99 (0.72 - 1.37) | 0.93 |
| DAG | 5,039 | 1,014 | 4.5 | 0.9 | 675 | 131 | 0.98 (0.79 - 1.24) | 0.89 | 1.06 (0.84 - 1.35) | 0.56 |
| ODOD | 20,061 | 52,049 | 15.1 | 43.2 | 1,578 | 4,237 | 0.93 (0.86 - 1.00) | 0.07 | 0.96 (0.89 - 1.05) | 0.50 |
| MDCR | 9,004 | 18,079 | 9.4 | 19.0 | 11 | 80 | 0.43 (0.14 - 1.06) | 0.10 | 0.42 (0.16 - 1.15) | 0.09 |
| MDCD | 4,087 | 9,851 | 3.1 | 7.1 | 0 | 0 | | | | |
| Open Claims | 422,431 | 903,346 | 403.8 | 982.0 | 0 | 0 | | | | |
| OEHR | 36,445 | 95,879 | 18.1 | 53.4 | 0 | 0 | | | | |
| VA | 0 | 0 | 0.0 | 0.0 | 0 | 0 | | | | |
| SIDIAP | 4,064 | 1,774 | 6.9 | 2.9 | 1,719 | 644 | 1.12 (0.99 - 1.28) | 0.07 | 1.07 (0.94 - 1.21) | 0.41 |
| Meta-analysis | 85,182 | 157,334 | 77.0 | 137.7 | 4,096 | 5,410 | 0.99 (0.88 - 1.11) | 0.87 | 0.99 (0.85 - 1.16) | 0.92 |
| Meta-analysis (w/o USOC) | 85,182 | 157,334 | 77.0 | 137.7 | 4,096 | 5,410 | 0.99 (0.87 - 1.12) | 0.87 | 1.01 (0.87 - 1.17) | 0.87 |
| **PS-stratification / on-treatment TAR** | | | | | | | | | | |
| CCAE | 52,351 | 85,573 | 46.0 | 72.7 | 131 | 490 | 0.71 (0.58 - 0.88) | <0.01 | 0.72 (0.58 - 0.89) | <0.01 |
| DAG | 5,066 | 1,195 | 4.5 | 1.0 | 683 | 161 | 0.93 (0.77 - 1.14) | 0.49 | 1.00 (0.82 - 1.22) | 0.62 |
| ODOD | 20,910 | 52,056 | 15.7 | 43.2 | 1,678 | 4,237 | 0.93 (0.88 - 0.99) | 0.02 | 0.94 (0.88 - 1.01) | 0.17 |
| MDCR | 9,515 | 18,352 | 10.0 | 19.3 | 12 | 174 | 0.34 (0.18 - 0.59) | <0.01 | 0.34 (0.19 - 0.63) | <0.01 |
| MDCD | 4,183 | 9,871 | 3.2 | 7.1 | 0 | 0 | | | | |
| Open Claims | 429,825 | 903,360 | 413.2 | 982.0 | 0 | 0 | | | | |
| OEHR | 38,289 | 95,909 | 19.2 | 53.4 | 0 | 0 | | | | |
| VA | 24,844 | 221,635 | 23.3 | 233.0 | 21 | 499 | 0.39 (0.24 - 0.58) | <0.01 | 0.40 (0.12 - 1.30) | 0.13 |
| SIDIAP | 4,096 | 2,569 | 6.9 | 4.3 | 1,729 | 944 | 1.13 (1.03 - 1.24) | <0.01 | 1.11 (1.01 - 1.23) | 0.06 |
| Meta-analysis | 91,938 | 159,745 | 83.1 | 140.5 | 4,233 | 6,006 | 0.81 (0.60 - 1.11) | 0.19 | 0.82 (0.60 - 1.13) | 0.23 |
| Meta-analysis (w/o USOC) | 91,938 | 159,745 | 83.1 | 140.5 | 4,233 | 6,006 | 0.87 (0.73 - 1.04) | 0.13 | 0.89 (0.73 - 1.08) | 0.24 |
| **PS-matching / on-treatment TAR without escalation** | | | | | | | | | | |
| CCAE | 47,014 | 84,418 | 33.2 | 62.4 | 90 | 290 | 0.95 (0.67 - 1.34) | 0.76 | 0.97 (0.68 - 1.37) | 0.86 |
| DAG | 5,039 | 1,014 | 4.1 | 0.8 | 618 | 123 | 1.01 (0.80 - 1.29) | 0.92 | 1.10 (0.86 - 1.41) | 0.50 |
| ODOD | 20,061 | 52,049 | 12.4 | 38.6 | 1,280 | 3,854 | 0.91 (0.83 - 0.99) | 0.03 | 0.96 (0.88 - 1.05) | 0.45 |
| MDCR | 9,004 | 18,079 | 8.0 | 17.3 | 10 | 74 | 0.52 (0.17 - 1.32) | 0.21 | 0.52 (0.19 - 1.46) | 0.21 |
| MDCD | 4,087 | 9,851 | 2.4 | 5.9 | 0 | 0 | | | | |
| Open Claims | 422,431 | 903,346 | 313.0 | 831.5 | 0 | 0 | | | | |
| OEHR | 36,445 | 95,879 | 14.7 | 46.3 | 0 | 0 | | | | |
| VA | 0 | 0 | 0.0 | 0.0 | 0 | 0 | | | | |
| SIDIAP | 4,064 | 1,774 | 4.9 | 2.4 | 1,302 | 569 | 1.10 (0.96 - 1.26) | 0.19 | 1.02 (0.88 - 1.17) | 0.73 |
| Meta-analysis | 85,182 | 157,334 | 62.6 | 121.5 | 3,300 | 4,910 | 0.98 (0.87 - 1.10) | 0.70 | 0.99 (0.85 - 1.16) | 0.89 |
| Meta-analysis (w/o USOC) | 85,182 | 157,334 | 62.6 | 121.5 | 3,300 | 4,910 | 0.98 (0.87 - 1.10) | 0.71 | 1.01 (0.87 - 1.17) | 0.87 |
| **PS-stratification / on-treatment TAR without escalation** | | | | | | | | | | |
| CCAE | 52,351 | 85,573 | 37.1 | 63.2 | 108 | 451 | 0.67 (0.53 - 0.85) | <0.01 | 0.69 (0.54 - 0.86) | <0.01 |
| DAG | 5,066 | 1,195 | 4.1 | 0.9 | 625 | 152 | 0.94 (0.77 - 1.15) | 0.55 | 1.02 (0.83 - 1.26) | 0.62 |
| ODOD | 20,910 | 52,056 | 12.9 | 38.6 | 1,361 | 3,854 | 0.90 (0.84 - 0.95) | <0.01 | 0.91 (0.84 - 0.98) | 0.05 |
| MDCR | 9,515 | 18,352 | 8.5 | 17.6 | 11 | 164 | 0.36 (0.18 - 0.65) | <0.01 | 0.36 (0.19 - 0.69) | <0.01 |
| MDCD | 4,183 | 9,871 | 2.4 | 6.0 | 0 | 0 | | | | |
| Open Claims | 429,825 | 903,360 | 320.1 | 831.5 | 0 | 0 | | | | |
| OEHR | 38,289 | 95,909 | 15.6 | 46.3 | 0 | 0 | | | | |
| VA | 24,844 | 221,635 | 18.9 | 207.4 | 18 | 467 | 0.38 (0.23 - 0.59) | <0.01 | 0.39 (0.12 - 1.28) | 0.13 |
| SIDIAP | 4,096 | 2,569 | 4.9 | 3.6 | 1,312 | 828 | 1.09 (0.99 - 1.21) | 0.09 | 1.05 (0.94 - 1.17) | 0.47 |
| Meta-analysis | 91,938 | 159,745 | 67.5 | 123.8 | 3,417 | 5,449 | 0.81 (0.62 - 1.07) | 0.13 | 0.82 (0.61 - 1.10) | 0.19 |
| Meta-analysis (w/o USOC) | 91,938 | 159,745 | 67.5 | 123.8 | 3,417 | 5,449 | 0.85 (0.71 - 1.02) | 0.08 | 0.87 (0.72 - 1.07) | 0.19 |

##### 1.6.6 Hospitalization with heart failure

**Supplementary Table 136:** Exposures, outcomes and risk estimates for Hospitalization with heart failure in DPP4I (male) (T) vs SU (male) (C) new-user cohorts under four different propensity score (PS) adjustment / time-at-risk (TAR) designs across data sources. We report total patient exposure time in 1,000 patient-year (KYr) units, uncalibrated and calibrated hazard ratios (HRs) and their 95% confidence intervals (CIs). Greyed out entries did not pass study diagnostics and are included only for completeness.

| | Patients | | Exposure (KYr) | | Outcomes | | Uncalibrated | | Calibrated | |
|---|---|---|---|---|---|---|---|---|---|---|
| | T | C | T | C | T | C | HR (95% CI) | $p$ | HR (95% CI) | $p$ |
| **PS-matching / on-treatment TAR** | | | | | | | | | | |
| CCAE | 46,539 | 83,340 | 40.7 | 71.0 | 238 | 531 | 0.85 (0.68 - 1.05) | 0.13 | 0.86 (0.69 - 1.07) | 0.20 |
| DAG | 13,419 | 2,361 | 14.3 | 2.4 | 0 | 0 | | | | |
| ODOD | 27,628 | 66,531 | 23.1 | 61.6 | 428 | 1,697 | 0.81 (0.70 - 0.94) | <0.01 | 0.84 (0.73 - 0.98) | 0.04 |
| MDCR | 8,253 | 16,385 | 8.5 | 17.1 | 272 | 657 | 0.95 (0.77 - 1.15) | 0.58 | 0.93 (0.76 - 1.14) | 0.49 |
| MDCD | 3,699 | 8,827 | 2.8 | 6.3 | 62 | 187 | 0.57 (0.37 - 0.85) | <0.01 | 0.59 (0.39 - 0.90) | 0.02 |
| Open Claims | 403,516 | 860,385 | 384.8 | 930.1 | 4,057 | 12,803 | 0.84 (0.80 - 0.88) | <0.01 | 0.84 (0.74 - 0.95) | <0.01 |
| OEHR | 35,630 | 93,613 | 17.4 | 51.1 | 225 | 920 | 0.95 (0.78 - 1.15) | 0.60 | 1.01 (0.81 - 1.27) | 0.87 |
| VA | 0 | 0 | 0.0 | 0.0 | 0 | 0 | | | | |
| SIDIAP | 26,675 | 7,201 | 78.1 | 20.5 | 108 | 25 | 0.93 (0.51 - 1.71) | 0.80 | 0.88 (0.48 - 1.60) | 0.67 |
| Meta-analysis | 551,940 | 1,136,282 | 555.5 | 1157.7 | 5,390 | 16,820 | 0.84 (0.81 - 0.88) | <0.01 | 0.85 (0.75 - 0.95) | <0.01 |
| Meta-analysis (w/o USOC) | 148,424 | 275,897 | 170.8 | 227.6 | 1,333 | 4,017 | 0.86 (0.77 - 0.95) | <0.01 | 0.88 (0.77 - 1.00) | 0.08 |
| **PS-stratification / on-treatment TAR** | | | | | | | | | | |
| CCAE | 51,915 | 84,822 | 45.6 | 72.2 | 256 | 536 | 0.81 (0.69 - 0.94) | <0.01 | 0.81 (0.69 - 0.95) | 0.02 |
| DAG | 13,447 | 2,736 | 14.3 | 2.8 | 0 | 0 | | | | |
| ODOD | 29,198 | 66,535 | 24.6 | 61.6 | 448 | 1,697 | 0.79 (0.71 - 0.88) | <0.01 | 0.80 (0.72 - 0.90) | <0.01 |
| MDCR | 8,741 | 16,748 | 9.1 | 17.5 | 284 | 671 | 0.96 (0.83 - 1.11) | 0.61 | 0.97 (0.84 - 1.13) | 0.74 |
| MDCD | 3,781 | 8,844 | 2.9 | 6.3 | 62 | 187 | 0.80 (0.59 - 1.07) | 0.15 | 0.82 (0.61 - 1.11) | 0.23 |
| Open Claims | 411,051 | 860,397 | 394.2 | 930.1 | 4,180 | 12,803 | 0.84 (0.81 - 0.87) | <0.01 | 0.83 (0.71 - 0.96) | 0.02 |
| OEHR | 37,434 | 93,642 | 18.5 | 51.2 | 239 | 920 | 0.87 (0.74 - 1.00) | 0.06 | 0.95 (0.82 - 1.11) | 0.56 |
| VA | 24,760 | 225,345 | 23.1 | 237.6 | 201 | 2,899 | 0.72 (0.62 - 0.82) | <0.01 | 0.74 (0.25 - 2.23) | 0.60 |
| SIDIAP | 26,764 | 9,253 | 78.2 | 27.6 | 108 | 33 | 0.84 (0.55 - 1.33) | 0.45 | 0.83 (0.53 - 1.29) | 0.40 |
| Meta-analysis | 568,884 | 1,140,241 | 573.1 | 1166.5 | 5,577 | 16,847 | 0.84 (0.81 - 0.87) | <0.01 | 0.85 (0.77 - 0.93) | <0.01 |
| Meta-analysis (w/o USOC) | 157,833 | 279,844 | 178.9 | 236.5 | 1,397 | 4,044 | 0.84 (0.79 - 0.90) | <0.01 | 0.86 (0.78 - 0.95) | <0.01 |
| **PS-matching / on-treatment TAR without escalation** | | | | | | | | | | |
| CCAE | 46,539 | 83,340 | 32.9 | 61.8 | 194 | 454 | 0.89 (0.70 - 1.12) | 0.33 | 0.91 (0.72 - 1.15) | 0.45 |
| DAG | 13,419 | 2,361 | 12.9 | 2.2 | 0 | 0 | | | | |
| ODOD | 27,628 | 66,531 | 19.0 | 55.0 | 356 | 1,497 | 0.85 (0.73 - 0.99) | 0.03 | 0.89 (0.76 - 1.04) | 0.18 |
| MDCR | 8,253 | 16,385 | 7.3 | 15.6 | 235 | 601 | 1.03 (0.83 - 1.26) | 0.81 | 1.03 (0.83 - 1.27) | 0.78 |
| MDCD | 3,699 | 8,827 | 2.1 | 5.3 | 51 | 156 | 0.60 (0.38 - 0.93) | 0.03 | 0.63 (0.40 - 0.99) | 0.05 |
| Open Claims | 403,516 | 860,385 | 298.5 | 788.6 | 3,166 | 10,900 | 0.83 (0.79 - 0.88) | <0.01 | 0.83 (0.74 - 0.94) | <0.01 |
| OEHR | 35,630 | 93,613 | 14.2 | 44.8 | 164 | 704 | 0.92 (0.74 - 1.13) | 0.42 | 0.99 (0.77 - 1.28) | 0.88 |
| VA | 0 | 0 | 0.0 | 0.0 | 0 | 0 | | | | |
| SIDIAP | 26,675 | 7,201 | 57.2 | 17.1 | 79 | 22 | 1.11 (0.56 - 2.30) | 0.77 | 1.03 (0.51 - 2.09) | 0.92 |
| Meta-analysis | 551,940 | 1,136,282 | 431.2 | 988.1 | 4,245 | 14,334 | 0.86 (0.80 - 0.91) | <0.01 | 0.86 (0.76 - 0.98) | 0.02 |
| Meta-analysis (w/o USOC) | 148,424 | 275,897 | 132.7 | 199.5 | 1,079 | 3,434 | 0.89 (0.81 - 0.99) | 0.03 | 0.92 (0.80 - 1.06) | 0.28 |
| **PS-stratification / on-treatment TAR without escalation** | | | | | | | | | | |
| CCAE | 51,915 | 84,822 | 36.8 | 62.9 | 209 | 459 | 0.83 (0.69 - 0.98) | 0.03 | 0.84 (0.70 - 1.00) | 0.06 |
| DAG | 13,447 | 2,736 | 12.9 | 2.6 | 0 | 0 | | | | |
| ODOD | 29,198 | 66,535 | 20.1 | 55.0 | 374 | 1,497 | 0.82 (0.73 - 0.92) | <0.01 | 0.83 (0.74 - 0.94) | <0.01 |
| MDCR | 8,741 | 16,748 | 7.8 | 16.1 | 246 | 614 | 0.96 (0.82 - 1.12) | 0.63 | 0.97 (0.83 - 1.13) | 0.69 |
| MDCD | 3,781 | 8,844 | 2.2 | 5.3 | 51 | 156 | 0.85 (0.60 - 1.17) | 0.32 | 0.86 (0.62 - 1.20) | 0.40 |
| Open Claims | 411,051 | 860,397 | 305.6 | 788.6 | 3,260 | 10,900 | 0.83 (0.80 - 0.87) | <0.01 | 0.83 (0.70 - 0.98) | 0.03 |
| OEHR | 37,434 | 93,642 | 15.1 | 44.8 | 176 | 704 | 0.88 (0.73 - 1.04) | 0.14 | 0.97 (0.81 - 1.17) | 0.80 |
| VA | 24,760 | 225,345 | 18.8 | 212.0 | 158 | 2,488 | 0.71 (0.60 - 0.84) | <0.01 | 0.72 (0.24 - 2.20) | 0.57 |
| SIDIAP | 26,764 | 9,253 | 57.3 | 22.8 | 79 | 27 | 0.85 (0.52 - 1.42) | 0.52 | 0.81 (0.49 - 1.35) | 0.43 |
| Meta-analysis | 568,884 | 1,140,241 | 444.9 | 995.5 | 4,395 | 14,357 | 0.84 (0.81 - 0.87) | <0.01 | 0.85 (0.75 - 0.95) | <0.01 |
| Meta-analysis (w/o USOC) | 157,833 | 279,844 | 139.3 | 206.8 | 1,135 | 3,457 | 0.86 (0.80 - 0.93) | <0.01 | 0.88 (0.79 - 0.99) | 0.04 |

##### 1.6.7 Measured renal dysfunction

**Supplementary Table 137:** Exposures, outcomes and risk estimates for Measured renal dysfunction in DPP4I (male) (T) vs SU (male) (C) new-user cohorts under four different propensity score (PS) adjustment / time-at-risk (TAR) designs across data sources. We report total patient exposure time in 1,000 patient-year (KYr) units, uncalibrated and calibrated hazard ratios (HRs) and their 95% confidence intervals (CIs). Greyed out entries did not pass study diagnostics and are included only for completeness.

| | Patients | | Exposure (KYr) | | Outcomes | | Uncalibrated | | Calibrated | |
|---|---|---|---|---|---|---|---|---|---|---|
| | T | C | T | C | T | C | HR (95% CI) | $p$ | HR (95% CI) | $p$ |
| **PS-matching / on-treatment TAR** | | | | | | | | | | |
| CCAE | 47,371 | 85,067 | 41.5 | 72.6 | 14 | 30 | 1.10 (0.42 - 2.79) | 0.84 | 1.12 (0.43 - 2.89) | 0.82 |
| DAG | 13,419 | 2,361 | 14.3 | 2.4 | 0 | 0 | | | | |
| ODOD | 29,183 | 70,640 | 24.5 | 66.4 | 51 | 146 | 1.13 (0.72 - 1.75) | 0.60 | 1.17 (0.75 - 1.84) | 0.49 |
| MDCR | 9,055 | 18,278 | 9.4 | 19.3 | <5 | 8 | 0.33 (0.02 - 2.60) | 0.39 | 0.33 (0.03 - 4.12) | 0.39 |
| MDCD | 4,087 | 9,851 | 3.1 | 7.1 | 0 | 0 | | | | |
| Open Claims | 422,431 | 903,346 | 403.8 | 982.0 | 0 | 0 | | | | |
| OEHR | 36,445 | 95,879 | 18.1 | 53.4 | 0 | 0 | | | | |
| VA | 0 | 0 | 0.0 | 0.0 | 0 | 0 | | | | |
| SIDIAP | 26,580 | 7,201 | 77.4 | 20.4 | 421 | 28 | 2.46 (1.53 - 4.18) | <0.01 | 2.33 (1.41 - 3.86) | <0.01 |
| Meta-analysis | 103,134 | 162,908 | 143.4 | 159.4 | 486 | 204 | 1.51 (0.86 - 2.65) | 0.15 | 1.52 (0.86 - 2.68) | 0.15 |
| Meta-analysis (w/o USOC) | 103,134 | 162,908 | 143.4 | 159.4 | 486 | 204 | 1.51 (0.85 - 2.68) | 0.16 | 1.54 (0.86 - 2.76) | 0.15 |
| **PS-stratification / on-treatment TAR** | | | | | | | | | | |
| CCAE | 52,816 | 86,589 | 46.4 | 73.9 | 17 | 30 | 0.78 (0.41 - 1.45) | 0.44 | 0.78 (0.42 - 1.48) | 0.45 |
| DAG | 13,447 | 2,736 | 14.3 | 2.8 | 0 | 0 | | | | |
| ODOD | 30,796 | 70,646 | 26.0 | 66.4 | 54 | 146 | 0.94 (0.67 - 1.29) | 0.70 | 0.95 (0.68 - 1.32) | 0.75 |
| MDCR | 9,568 | 18,672 | 10.0 | 19.7 | <5 | 8 | 0.25 (0.01 - 1.48) | 0.25 | 0.26 (0.02 - 2.71) | 0.26 |
| MDCD | 4,183 | 9,871 | 3.2 | 7.1 | 0 | 0 | | | | |
| Open Claims | 429,825 | 903,360 | 413.2 | 982.0 | 0 | 0 | | | | |
| OEHR | 38,289 | 95,909 | 19.2 | 53.4 | 0 | 0 | | | | |
| VA | 25,292 | 229,407 | 23.6 | 243.1 | 98 | 1,265 | 0.80 (0.65 - 0.98) | 0.03 | 0.83 (0.27 - 2.52) | 0.74 |
| SIDIAP | 26,665 | 9,253 | 77.5 | 27.6 | 424 | 40 | 2.22 (1.57 - 3.21) | <0.01 | 2.18 (1.52 - 3.11) | <0.01 |
| Meta-analysis | 110,277 | 166,488 | 149.9 | 167.8 | 495 | 216 | 1.21 (0.64 - 2.28) | 0.57 | 1.22 (0.64 - 2.32) | 0.55 |
| Meta-analysis (w/o USOC) | 110,277 | 166,488 | 149.9 | 167.8 | 495 | 216 | 1.20 (0.62 - 2.33) | 0.58 | 1.23 (0.63 - 2.38) | 0.54 |
| **PS-matching / on-treatment TAR without escalation** | | | | | | | | | | |
| CCAE | 47,371 | 85,067 | 33.5 | 63.1 | 11 | 26 | 1.38 (0.50 - 3.68) | 0.53 | 1.41 (0.52 - 3.80) | 0.50 |
| DAG | 13,419 | 2,361 | 12.9 | 2.2 | 0 | 0 | | | | |
| ODOD | 29,183 | 70,640 | 20.0 | 59.1 | 45 | 126 | 1.21 (0.75 - 1.94) | 0.42 | 1.28 (0.79 - 2.06) | 0.32 |
| MDCR | 9,055 | 18,278 | 8.1 | 17.6 | 0 | 7 | | | | |
| MDCD | 4,087 | 9,851 | 2.4 | 5.9 | 0 | 0 | | | | |
| Open Claims | 422,431 | 903,346 | 313.0 | 831.5 | 0 | 0 | | | | |
| OEHR | 36,445 | 95,879 | 14.7 | 46.3 | 0 | 0 | | | | |
| VA | 0 | 0 | 0.0 | 0.0 | 0 | 0 | | | | |
| SIDIAP | 26,580 | 7,201 | 56.7 | 17.1 | 320 | 19 | 3.25 (1.78 - 6.53) | <0.01 | 3.01 (1.57 - 5.78) | <0.01 |
| Meta-analysis | 103,134 | 162,908 | 110.2 | 139.3 | 376 | 171 | 1.76 (0.92 - 3.39) | 0.09 | 1.78 (0.92 - 3.46) | 0.09 |
| Meta-analysis (w/o USOC) | 103,134 | 162,908 | 110.2 | 139.3 | 376 | 171 | 1.76 (0.90 - 3.45) | 0.10 | 1.82 (0.92 - 3.59) | 0.09 |
| **PS-stratification / on-treatment TAR without escalation** | | | | | | | | | | |
| CCAE | 52,816 | 86,589 | 37.5 | 64.3 | 13 | 26 | 0.75 (0.36 - 1.50) | 0.43 | 0.76 (0.37 - 1.56) | 0.46 |
| DAG | 13,447 | 2,736 | 12.9 | 2.6 | 0 | 0 | | | | |
| ODOD | 30,796 | 70,646 | 21.2 | 59.1 | 48 | 126 | 1.05 (0.74 - 1.49) | 0.77 | 1.07 (0.75 - 1.53) | 0.71 |
| MDCR | 9,568 | 18,672 | 8.6 | 18.0 | 0 | 7 | | | | |
| MDCD | 4,183 | 9,871 | 2.4 | 6.0 | 0 | 0 | | | | |
| Open Claims | 429,825 | 903,360 | 320.1 | 831.5 | 0 | 0 | | | | |
| OEHR | 38,289 | 95,909 | 15.6 | 46.3 | 0 | 0 | | | | |
| VA | 25,292 | 229,407 | 19.2 | 216.6 | 77 | 1,037 | 0.83 (0.66 - 1.04) | 0.13 | 0.84 (0.27 - 2.60) | 0.77 |
| SIDIAP | 26,665 | 9,253 | 56.8 | 22.8 | 323 | 26 | 2.76 (1.81 - 4.38) | <0.01 | 2.66 (1.71 - 4.13) | <0.01 |
| Meta-analysis | 110,277 | 166,488 | 115.5 | 146.2 | 384 | 178 | 1.33 (0.63 - 2.84) | 0.46 | 1.34 (0.63 - 2.89) | 0.45 |
| Meta-analysis (w/o USOC) | 110,277 | 166,488 | 115.5 | 146.2 | 384 | 178 | 1.33 (0.63 - 2.81) | 0.45 | 1.37 (0.64 - 2.90) | 0.42 |

##### 1.6.8 Stroke

**Supplementary Table 138:** Exposures, outcomes and risk estimates for Stroke in DPP4I (male) (T) vs SU (male) (C) new-user cohorts under four different propensity score (PS) adjustment / time-at-risk (TAR) designs across data sources. We report total patient exposure time in 1,000 patient-year (KYr) units, uncalibrated and calibrated hazard ratios (HRs) and their 95% confidence intervals (CIs). Greyed out entries did not pass study diagnostics and are included only for completeness.

| | Patients | | Exposure (KYr) | | Outcomes | | Uncalibrated | | Calibrated | |
|---|---|---|---|---|---|---|---|---|---|---|
| | T | C | T | C | T | C | HR (95% CI) | $p$ | HR (95% CI) | $p$ |
| **PS-matching / on-treatment TAR** | | | | | | | | | | |
| CCAE | 46,880 | 83,957 | 41.1 | 71.6 | 138 | 265 | 1.00 (0.74 - 1.33) | 0.98 | 1.01 (0.75 - 1.36) | 0.92 |
| DAG | 13,419 | 2,361 | 14.3 | 2.4 | 0 | 0 | | | | |
| ODOD | 28,456 | 68,809 | 23.9 | 64.3 | 212 | 769 | 0.87 (0.71 - 1.07) | 0.19 | 0.90 (0.73 - 1.11) | 0.37 |
| MDCR | 8,667 | 17,383 | 9.0 | 18.2 | 133 | 318 | 0.98 (0.74 - 1.31) | 0.92 | 0.97 (0.73 - 1.29) | 0.82 |
| MDCD | 3,910 | 9,416 | 3.0 | 6.7 | 33 | 100 | 0.97 (0.54 - 1.70) | 0.92 | 1.01 (0.57 - 1.80) | 0.92 |
| Open Claims | 410,222 | 876,424 | 392.0 | 951.0 | 2,009 | 6,194 | 0.84 (0.79 - 0.90) | <0.01 | 0.84 (0.73 - 0.96) | 0.02 |
| OEHR | 36,122 | 94,871 | 17.9 | 52.5 | 82 | 296 | 1.26 (0.90 - 1.75) | 0.17 | 1.35 (0.94 - 1.92) | 0.13 |
| VA | 0 | 0 | 0.0 | 0.0 | 0 | 0 | | | | |
| SIDIAP | 26,445 | 7,160 | 77.2 | 20.3 | 222 | 33 | 1.52 (0.96 - 2.52) | 0.09 | 1.44 (0.89 - 2.35) | 0.15 |
| Meta-analysis | 560,702 | 1,158,020 | 564.0 | 1184.8 | 2,829 | 7,975 | 0.97 (0.84 - 1.12) | 0.68 | 0.98 (0.82 - 1.16) | 0.78 |
| Meta-analysis (w/o USOC) | 150,480 | 281,596 | 172.1 | 233.7 | 820 | 1,781 | 1.02 (0.88 - 1.19) | 0.76 | 1.05 (0.89 - 1.24) | 0.62 |
| **PS-stratification / on-treatment TAR** | | | | | | | | | | |
| CCAE | 52,301 | 85,457 | 46.0 | 72.9 | 152 | 268 | 0.97 (0.79 - 1.20) | 0.79 | 0.98 (0.79 - 1.21) | 0.85 |
| DAG | 13,447 | 2,736 | 14.3 | 2.8 | 0 | 0 | | | | |
| ODOD | 30,047 | 68,815 | 25.4 | 64.3 | 223 | 770 | 0.85 (0.73 - 0.99) | 0.04 | 0.86 (0.73 - 1.01) | 0.08 |
| MDCR | 9,165 | 17,767 | 9.6 | 18.7 | 141 | 324 | 0.92 (0.75 - 1.13) | 0.44 | 0.93 (0.76 - 1.15) | 0.52 |
| MDCD | 3,997 | 9,434 | 3.1 | 6.8 | 33 | 100 | 0.76 (0.50 - 1.13) | 0.18 | 0.78 (0.52 - 1.17) | 0.24 |
| Open Claims | 417,616 | 876,438 | 401.3 | 951.0 | 2,067 | 6,194 | 0.83 (0.79 - 0.88) | <0.01 | 0.83 (0.71 - 0.96) | 0.02 |
| OEHR | 37,946 | 94,901 | 19.0 | 52.5 | 87 | 296 | 0.98 (0.76 - 1.25) | 0.87 | 1.08 (0.84 - 1.39) | 0.58 |
| VA | 25,218 | 230,647 | 23.6 | 245.2 | 64 | 859 | 0.76 (0.59 - 0.98) | 0.04 | 0.79 (0.26 - 2.42) | 0.68 |
| SIDIAP | 26,532 | 9,197 | 77.3 | 27.4 | 222 | 47 | 1.34 (0.95 - 1.93) | 0.11 | 1.31 (0.92 - 1.88) | 0.14 |
| Meta-analysis | 577,604 | 1,162,009 | 581.6 | 1193.6 | 2,925 | 7,999 | 0.89 (0.82 - 0.98) | 0.01 | 0.90 (0.80 - 1.02) | 0.10 |
| Meta-analysis (w/o USOC) | 159,988 | 285,571 | 180.3 | 242.6 | 858 | 1,805 | 0.94 (0.84 - 1.04) | 0.24 | 0.96 (0.84 - 1.09) | 0.52 |
| **PS-matching / on-treatment TAR without escalation** | | | | | | | | | | |
| CCAE | 46,880 | 83,957 | 33.1 | 62.3 | 108 | 239 | 0.97 (0.70 - 1.32) | 0.83 | 0.99 (0.72 - 1.35) | 0.92 |
| DAG | 13,419 | 2,361 | 12.9 | 2.2 | 0 | 0 | | | | |
| ODOD | 28,456 | 68,809 | 19.5 | 57.3 | 186 | 686 | 0.93 (0.74 - 1.15) | 0.51 | 0.98 (0.78 - 1.22) | 0.85 |
| MDCR | 8,667 | 17,383 | 7.7 | 16.6 | 113 | 290 | 0.94 (0.69 - 1.27) | 0.68 | 0.94 (0.69 - 1.28) | 0.70 |
| MDCD | 3,910 | 9,416 | 2.3 | 5.6 | 23 | 81 | 0.78 (0.38 - 1.51) | 0.48 | 0.82 (0.41 - 1.63) | 0.57 |
| Open Claims | 410,222 | 876,424 | 303.9 | 805.4 | 1,601 | 5,349 | 0.84 (0.78 - 0.91) | <0.01 | 0.84 (0.74 - 0.96) | 0.01 |
| OEHR | 36,122 | 94,871 | 14.5 | 45.6 | 60 | 225 | 1.26 (0.86 - 1.82) | 0.23 | 1.36 (0.91 - 2.04) | 0.15 |
| VA | 0 | 0 | 0.0 | 0.0 | 0 | 0 | | | | |
| SIDIAP | 26,445 | 7,160 | 56.6 | 17.0 | 148 | 31 | 1.59 (0.93 - 2.82) | 0.10 | 1.47 (0.85 - 2.56) | 0.18 |
| Meta-analysis | 560,702 | 1,158,020 | 437.7 | 1009.8 | 2,239 | 6,901 | 0.94 (0.83 - 1.08) | 0.39 | 0.96 (0.81 - 1.13) | 0.60 |
| Meta-analysis (w/o USOC) | 150,480 | 281,596 | 133.7 | 204.4 | 638 | 1,552 | 1.00 (0.87 - 1.16) | 0.95 | 1.04 (0.87 - 1.23) | 0.70 |
| **PS-stratification / on-treatment TAR without escalation** | | | | | | | | | | |
| CCAE | 52,301 | 85,457 | 37.1 | 63.4 | 121 | 242 | 0.89 (0.71 - 1.12) | 0.34 | 0.91 (0.72 - 1.15) | 0.44 |
| DAG | 13,447 | 2,736 | 12.9 | 2.6 | 0 | 0 | | | | |
| ODOD | 30,047 | 68,815 | 20.7 | 57.3 | 194 | 687 | 0.90 (0.76 - 1.06) | 0.23 | 0.92 (0.77 - 1.09) | 0.35 |
| MDCR | 9,165 | 17,767 | 8.2 | 17.0 | 118 | 296 | 0.91 (0.72 - 1.13) | 0.38 | 0.91 (0.73 - 1.14) | 0.43 |
| MDCD | 3,997 | 9,434 | 2.3 | 5.7 | 23 | 81 | 0.68 (0.42 - 1.08) | 0.12 | 0.69 (0.43 - 1.12) | 0.15 |
| Open Claims | 417,616 | 876,438 | 311.0 | 805.4 | 1,640 | 5,349 | 0.83 (0.79 - 0.88) | <0.01 | 0.83 (0.70 - 0.98) | 0.03 |
| OEHR | 37,946 | 94,901 | 15.4 | 45.6 | 64 | 225 | 1.06 (0.79 - 1.42) | 0.68 | 1.18 (0.88 - 1.59) | 0.29 |
| VA | 25,218 | 230,647 | 19.2 | 218.4 | 41 | 740 | 0.62 (0.44 - 0.83) | <0.01 | 0.62 (0.20 - 1.96) | 0.42 |
| SIDIAP | 26,532 | 9,197 | 56.7 | 22.7 | 148 | 40 | 1.09 (0.73 - 1.65) | 0.69 | 1.04 (0.69 - 1.58) | 0.84 |
| Meta-analysis | 577,604 | 1,162,009 | 451.4 | 1017.1 | 2,308 | 6,920 | 0.87 (0.81 - 0.93) | <0.01 | 0.88 (0.77 - 1.00) | 0.05 |
| Meta-analysis (w/o USOC) | 159,988 | 285,571 | 140.4 | 211.7 | 668 | 1,571 | 0.92 (0.83 - 1.02) | 0.10 | 0.94 (0.82 - 1.08) | 0.39 |

##### 1.6.9 Sudden cardiac death

**Supplementary Table 139:** Exposures, outcomes and risk estimates for Sudden cardiac death in DPP4I (male) (T) vs SU (male) (C) new-user cohorts under four different propensity score (PS) adjustment / time-at-risk (TAR) designs across data sources. We report total patient exposure time in 1,000 patient-year (KYr) units, uncalibrated and calibrated hazard ratios (HRs) and their 95% confidence intervals (CIs). Greyed out entries did not pass study diagnostics and are included only for completeness.

| | Patients | | Exposure (KYr) | | Outcomes | | Uncalibrated | | Calibrated | |
|---|---|---|---|---|---|---|---|---|---|---|
| | T | C | T | C | T | C | HR (95% CI) | $p$ | HR (95% CI) | $p$ |
| **PS-matching / on-treatment TAR** | | | | | | | | | | |
| CCAE | 47,385 | 85,092 | 41.5 | 72.7 | 35 | 111 | 0.57 (0.32 - 0.95) | 0.04 | 0.57 (0.33 - 0.99) | 0.05 |
| DAG | 13,419 | 2,361 | 14.3 | 2.4 | 0 | 0 | | | | |
| ODOD | 29,198 | 70,628 | 24.5 | 66.4 | 83 | 253 | 0.98 (0.69 - 1.38) | 0.92 | 1.02 (0.72 - 1.44) | 0.90 |
| MDCR | 9,027 | 18,209 | 9.4 | 19.2 | 40 | 69 | 1.02 (0.52 - 1.93) | 0.96 | 1.00 (0.52 - 1.94) | 0.95 |
| MDCD | 4,070 | 9,811 | 3.1 | 7.1 | 12 | 32 | 0.75 (0.26 - 1.94) | 0.58 | 0.78 (0.28 - 2.14) | 0.63 |
| Open Claims | 421,389 | 901,146 | 402.7 | 979.2 | 635 | 1,870 | 0.87 (0.77 - 0.99) | 0.04 | 0.87 (0.73 - 1.04) | 0.13 |
| OEHR | 36,417 | 95,802 | 18.1 | 53.3 | 29 | 104 | 0.91 (0.50 - 1.60) | 0.75 | 0.97 (0.54 - 1.75) | 0.92 |
| VA | 0 | 0 | 0.0 | 0.0 | 0 | 0 | | | | |
| SIDIAP | 26,792 | 7,215 | 78.5 | 20.5 | 5 | <5 | 0.87 (0.07 - 20.2) | 0.92 | 0.82 (0.05 - 13.6) | 0.89 |
| Meta-analysis | 547,486 | 1,180,688 | 499.3 | 1197.8 | 834 | 2,439 | 0.87 (0.78 - 0.98) | 0.02 | 0.88 (0.75 - 1.02) | 0.09 |
| Meta-analysis (w/o USOC) | 126,097 | 279,542 | 96.7 | 218.6 | 199 | 569 | 0.87 (0.68 - 1.09) | 0.23 | 0.89 (0.69 - 1.13) | 0.35 |
| **PS-stratification / on-treatment TAR** | | | | | | | | | | |
| CCAE | 52,834 | 86,613 | 46.5 | 73.9 | 36 | 112 | 0.52 (0.35 - 0.76) | <0.01 | 0.52 (0.35 - 0.77) | <0.01 |
| DAG | 13,447 | 2,736 | 14.3 | 2.8 | 0 | 0 | | | | |
| ODOD | 30,801 | 70,635 | 26.0 | 66.4 | 87 | 253 | 1.03 (0.79 - 1.32) | 0.84 | 1.04 (0.80 - 1.34) | 0.79 |
| MDCR | 9,537 | 18,603 | 10.0 | 19.7 | 40 | 71 | 1.24 (0.82 - 1.84) | 0.31 | 1.25 (0.83 - 1.88) | 0.29 |
| MDCD | 4,163 | 9,831 | 3.2 | 7.1 | 12 | 32 | 0.89 (0.43 - 1.73) | 0.74 | 0.92 (0.46 - 1.83) | 0.80 |
| Open Claims | 428,796 | 901,160 | 412.0 | 979.2 | 650 | 1,870 | 0.89 (0.82 - 0.98) | 0.02 | 0.89 (0.75 - 1.05) | 0.18 |
| OEHR | 38,259 | 95,832 | 19.2 | 53.3 | 30 | 104 | 0.99 (0.64 - 1.51) | 0.98 | 1.09 (0.71 - 1.68) | 0.69 |
| VA | 25,610 | 233,587 | 24.0 | 248.8 | <10 | 132 | 0.56 (0.23 - 1.10) | 0.14 | 0.57 (0.15 - 2.19) | 0.42 |
| SIDIAP | 26,884 | 9,267 | 78.6 | 27.7 | 5 | <5 | 0.63 (0.10 - 5.73) | 0.66 | 0.62 (0.08 - 4.62) | 0.64 |
| Meta-analysis | 564,390 | 1,182,674 | 516.9 | 1199.6 | 855 | 2,442 | 0.90 (0.72 - 1.12) | 0.35 | 0.91 (0.72 - 1.15) | 0.43 |
| Meta-analysis (w/o USOC) | 135,594 | 281,514 | 104.9 | 220.4 | 205 | 572 | 0.90 (0.67 - 1.22) | 0.50 | 0.92 (0.67 - 1.26) | 0.61 |
| **PS-matching / on-treatment TAR without escalation** | | | | | | | | | | |
| CCAE | 47,385 | 85,092 | 33.5 | 63.1 | 29 | 96 | 0.55 (0.31 - 0.94) | 0.04 | 0.56 (0.32 - 0.98) | 0.04 |
| DAG | 13,419 | 2,361 | 12.9 | 2.2 | 0 | 0 | | | | |
| ODOD | 29,198 | 70,628 | 20.1 | 59.2 | 74 | 223 | 1.10 (0.76 - 1.58) | 0.61 | 1.16 (0.80 - 1.68) | 0.44 |
| MDCR | 9,027 | 18,209 | 8.0 | 17.5 | 27 | 58 | 1.05 (0.52 - 2.05) | 0.90 | 1.05 (0.53 - 2.09) | 0.89 |
| MDCD | 4,070 | 9,811 | 2.4 | 5.9 | 9 | 27 | 0.54 (0.15 - 1.59) | 0.31 | 0.56 (0.17 - 1.85) | 0.34 |
| Open Claims | 421,389 | 901,146 | 312.2 | 829.2 | 490 | 1,586 | 0.86 (0.74 - 0.99) | 0.04 | 0.86 (0.72 - 1.03) | 0.11 |
| OEHR | 36,417 | 95,802 | 14.6 | 46.2 | 19 | 73 | 1.01 (0.49 - 1.96) | 0.98 | 1.09 (0.54 - 2.21) | 0.81 |
| VA | 0 | 0 | 0.0 | 0.0 | 0 | 0 | | | | |
| SIDIAP | 26,792 | 7,215 | 57.5 | 17.1 | <5 | <5 | 1.31 (0.11 - 29.8) | 0.85 | 1.21 (0.07 - 20.1) | 0.89 |
| Meta-analysis | 547,486 | 1,180,688 | 390.8 | 1021.1 | 648 | 2,063 | 0.87 (0.77 - 0.99) | 0.03 | 0.88 (0.75 - 1.04) | 0.13 |
| Meta-analysis (w/o USOC) | 126,097 | 279,542 | 78.6 | 192.0 | 158 | 477 | 0.88 (0.65 - 1.20) | 0.41 | 0.91 (0.66 - 1.25) | 0.56 |
| **PS-stratification / on-treatment TAR without escalation** | | | | | | | | | | |
| CCAE | 52,834 | 86,613 | 37.6 | 64.3 | 30 | 97 | 0.53 (0.34 - 0.81) | <0.01 | 0.54 (0.35 - 0.83) | <0.01 |
| DAG | 13,447 | 2,736 | 12.9 | 2.6 | 0 | 0 | | | | |
| ODOD | 30,801 | 70,635 | 21.3 | 59.2 | 78 | 223 | 1.14 (0.86 - 1.49) | 0.35 | 1.16 (0.88 - 1.53) | 0.31 |
| MDCR | 9,537 | 18,603 | 8.5 | 18.0 | 27 | 60 | 1.06 (0.65 - 1.69) | 0.81 | 1.06 (0.66 - 1.71) | 0.80 |
| MDCD | 4,163 | 9,831 | 2.4 | 5.9 | 9 | 27 | 0.82 (0.36 - 1.71) | 0.62 | 0.83 (0.38 - 1.82) | 0.65 |
| Open Claims | 428,796 | 901,160 | 319.2 | 829.2 | 501 | 1,586 | 0.89 (0.80 - 0.98) | 0.03 | 0.88 (0.73 - 1.07) | 0.20 |
| OEHR | 38,259 | 95,832 | 15.6 | 46.3 | 20 | 73 | 1.04 (0.60 - 1.72) | 0.90 | 1.15 (0.68 - 1.96) | 0.61 |
| VA | 25,610 | 233,587 | 19.5 | 221.5 | <10 | 100 | 0.45 (0.14 - 1.08) | 0.13 | 0.46 (0.10 - 2.06) | 0.31 |
| SIDIAP | 26,884 | 9,267 | 57.6 | 22.8 | <5 | <5 | 2.21 (0.21 - 55.2) | 0.58 | 2.13 (0.13 - 34.2) | 0.59 |
| Meta-analysis | 564,390 | 1,182,674 | 404.5 | 1022.8 | 665 | 2,066 | 0.90 (0.73 - 1.12) | 0.35 | 0.91 (0.71 - 1.16) | 0.45 |
| Meta-analysis (w/o USOC) | 135,594 | 281,514 | 85.3 | 193.6 | 164 | 480 | 0.90 (0.66 - 1.23) | 0.51 | 0.92 (0.67 - 1.27) | 0.63 |

##### 1.6.10 Abnormal weight gain

**Supplementary Table 140:** Exposures, outcomes and risk estimates for Abnormal weight gain in DPP4I (male) (T) vs SU (male) (C) new-user cohorts under four different propensity score (PS) adjustment / time-at-risk (TAR) designs across data sources. We report total patient exposure time in 1,000 patient-year (KYr) units, uncalibrated and calibrated hazard ratios (HRs) and their 95% confidence intervals (CIs). Greyed out entries did not pass study diagnostics and are included only for completeness.

| | Patients | | Exposure (KYr) | | Outcomes | | Uncalibrated | | Calibrated | |
|---|---|---|---|---|---|---|---|---|---|---|
| | T | C | T | C | T | C | HR (95% CI) | $p$ | HR (95% CI) | $p$ |
| **PS-matching / on-treatment TAR** | | | | | | | | | | |
| CCAE | 46,233 | 83,342 | 40.5 | 71.0 | 132 | 292 | 0.70 (0.53 - 0.92) | 0.01 | 0.71 (0.53 - 0.94) | 0.02 |
| DAG | 13,346 | 2,345 | 14.2 | 2.4 | 16 | <5 | 0.55 (0.15 - 2.72) | 0.43 | 0.59 (0.14 - 2.58) | 0.49 |
| ODOD | 28,438 | 69,041 | 23.8 | 64.6 | 143 | 374 | 0.90 (0.68 - 1.18) | 0.45 | 0.94 (0.71 - 1.23) | 0.64 |
| MDCR | 8,956 | 18,124 | 9.4 | 19.0 | 19 | 68 | 0.42 (0.20 - 0.84) | 0.02 | 0.42 (0.20 - 0.87) | 0.02 |
| MDCD | 3,898 | 9,467 | 3.0 | 6.7 | 20 | 67 | 0.59 (0.29 - 1.13) | 0.13 | 0.61 (0.31 - 1.21) | 0.16 |
| Open Claims | 411,272 | 882,968 | 392.5 | 958.3 | 687 | 1,841 | 0.82 (0.73 - 0.93) | <0.01 | 0.82 (0.69 - 0.97) | 0.03 |
| OEHR | 35,591 | 93,969 | 17.6 | 52.0 | 78 | 286 | 0.63 (0.44 - 0.90) | 0.01 | 0.68 (0.46 - 0.99) | 0.06 |
| VA | 0 | 0 | 0.0 | 0.0 | 0 | 0 | | | | |
| SIDIAP | 26,515 | 7,179 | 77.6 | 20.3 | 105 | 37 | 1.03 (0.62 - 1.76) | 0.90 | 0.98 (0.58 - 1.65) | 0.91 |
| Meta-analysis | 560,903 | 1,164,090 | 564.2 | 1191.9 | 1,184 | 2,965 | 0.79 (0.71 - 0.88) | <0.01 | 0.79 (0.68 - 0.92) | <0.01 |
| Meta-analysis (w/o USOC) | 149,631 | 281,122 | 171.8 | 233.7 | 497 | 1,124 | 0.73 (0.60 - 0.89) | <0.01 | 0.75 (0.61 - 0.93) | 0.01 |
| **PS-stratification / on-treatment TAR** | | | | | | | | | | |
| CCAE | 51,500 | 84,920 | 45.3 | 72.3 | 152 | 292 | 0.78 (0.63 - 0.96) | 0.02 | 0.79 (0.64 - 0.97) | 0.03 |
| DAG | 13,373 | 2,715 | 14.2 | 2.8 | 16 | <5 | 0.59 (0.19 - 2.40) | 0.43 | 0.63 (0.18 - 2.28) | 0.49 |
| ODOD | 29,960 | 69,047 | 25.1 | 64.6 | 157 | 374 | 1.01 (0.83 - 1.23) | 0.89 | 1.02 (0.84 - 1.25) | 0.82 |
| MDCR | 9,468 | 18,518 | 10.0 | 19.5 | 22 | 69 | 0.52 (0.31 - 0.85) | 0.01 | 0.53 (0.32 - 0.88) | 0.01 |
| MDCD | 3,988 | 9,487 | 3.0 | 6.8 | 22 | 67 | 0.72 (0.43 - 1.16) | 0.19 | 0.74 (0.45 - 1.22) | 0.24 |
| Open Claims | 418,304 | 882,982 | 401.3 | 958.3 | 710 | 1,841 | 0.86 (0.78 - 0.94) | <0.01 | 0.85 (0.72 - 1.01) | 0.07 |
| OEHR | 37,353 | 93,998 | 18.6 | 52.0 | 84 | 286 | 0.78 (0.60 - 1.00) | 0.05 | 0.85 (0.66 - 1.10) | 0.24 |
| VA | 25,119 | 230,220 | 23.5 | 244.6 | 11 | 407 | 0.27 (0.14 - 0.47) | <0.01 | 0.28 (0.08 - 0.97) | 0.05 |
| SIDIAP | 26,606 | 9,227 | 77.7 | 27.4 | 105 | 45 | 0.94 (0.63 - 1.41) | 0.75 | 0.92 (0.61 - 1.37) | 0.68 |
| Meta-analysis | 577,179 | 1,168,179 | 581.1 | 1200.9 | 1,252 | 2,974 | 0.85 (0.78 - 0.92) | <0.01 | 0.86 (0.76 - 0.96) | <0.01 |
| Meta-analysis (w/o USOC) | 158,875 | 285,197 | 179.8 | 242.6 | 542 | 1,133 | 0.82 (0.70 - 0.96) | 0.01 | 0.84 (0.70 - 1.00) | 0.05 |
| **PS-matching / on-treatment TAR without escalation** | | | | | | | | | | |
| CCAE | 46,233 | 83,342 | 32.7 | 61.7 | 105 | 251 | 0.71 (0.52 - 0.96) | 0.03 | 0.73 (0.53 - 0.99) | 0.05 |
| DAG | 13,346 | 2,345 | 12.8 | 2.2 | 14 | <5 | 0.51 (0.13 - 2.59) | 0.38 | 0.56 (0.12 - 2.50) | 0.45 |
| ODOD | 28,438 | 69,041 | 19.5 | 57.7 | 110 | 326 | 0.84 (0.63 - 1.13) | 0.27 | 0.89 (0.66 - 1.20) | 0.45 |
| MDCR | 8,956 | 18,124 | 8.0 | 17.3 | 13 | 62 | 0.41 (0.18 - 0.83) | 0.02 | 0.41 (0.19 - 0.87) | 0.02 |
| MDCD | 3,898 | 9,467 | 2.3 | 5.7 | 13 | 49 | 0.49 (0.21 - 1.06) | 0.09 | 0.52 (0.23 - 1.17) | 0.12 |
| Open Claims | 411,272 | 882,968 | 304.8 | 812.4 | 517 | 1,524 | 0.80 (0.70 - 0.92) | <0.01 | 0.80 (0.68 - 0.95) | 0.02 |
| OEHR | 35,591 | 93,969 | 14.3 | 45.2 | 60 | 237 | 0.65 (0.44 - 0.95) | 0.03 | 0.71 (0.47 - 1.07) | 0.12 |
| VA | 0 | 0 | 0.0 | 0.0 | 0 | 0 | | | | |
| SIDIAP | 26,515 | 7,179 | 56.8 | 17.0 | 81 | 31 | 1.06 (0.60 - 1.92) | 0.85 | 0.98 (0.55 - 1.76) | 0.92 |
| Meta-analysis | 560,903 | 1,164,090 | 438.4 | 1017.0 | 899 | 2,480 | 0.77 (0.70 - 0.86) | <0.01 | 0.78 (0.67 - 0.91) | <0.01 |
| Meta-analysis (w/o USOC) | 149,631 | 281,122 | 133.6 | 204.6 | 382 | 956 | 0.73 (0.60 - 0.88) | <0.01 | 0.75 (0.60 - 0.93) | 0.01 |
| **PS-stratification / on-treatment TAR without escalation** | | | | | | | | | | |
| CCAE | 51,500 | 84,920 | 36.6 | 62.9 | 121 | 251 | 0.74 (0.59 - 0.93) | <0.01 | 0.75 (0.60 - 0.95) | 0.02 |
| DAG | 13,373 | 2,715 | 12.8 | 2.6 | 14 | <5 | 0.55 (0.17 - 2.25) | 0.36 | 0.59 (0.16 - 2.19) | 0.44 |
| ODOD | 29,960 | 69,047 | 20.6 | 57.7 | 122 | 326 | 0.97 (0.77 - 1.20) | 0.76 | 0.98 (0.78 - 1.23) | 0.87 |
| MDCR | 9,468 | 18,518 | 8.5 | 17.8 | 16 | 63 | 0.44 (0.24 - 0.75) | <0.01 | 0.44 (0.25 - 0.78) | <0.01 |
| MDCD | 3,988 | 9,487 | 2.3 | 5.7 | 14 | 49 | 0.70 (0.37 - 1.26) | 0.25 | 0.71 (0.38 - 1.32) | 0.29 |
| Open Claims | 418,304 | 882,982 | 311.5 | 812.4 | 534 | 1,524 | 0.83 (0.75 - 0.92) | <0.01 | 0.83 (0.69 - 1.00) | 0.05 |
| OEHR | 37,353 | 93,998 | 15.2 | 45.2 | 66 | 237 | 0.73 (0.55 - 0.97) | 0.03 | 0.82 (0.61 - 1.09) | 0.19 |
| VA | 25,119 | 230,220 | 19.1 | 217.9 | <10 | 332 | 0.29 (0.14 - 0.54) | <0.01 | 0.30 (0.08 - 1.08) | 0.07 |
| SIDIAP | 26,606 | 9,227 | 56.9 | 22.7 | 81 | 39 | 1.04 (0.67 - 1.63) | 0.86 | 1.00 (0.64 - 1.56) | 0.93 |
| Meta-analysis | 577,179 | 1,168,179 | 451.6 | 1024.4 | 954 | 2,489 | 0.82 (0.76 - 0.89) | <0.01 | 0.83 (0.72 - 0.95) | <0.01 |
| Meta-analysis (w/o USOC) | 158,875 | 285,197 | 140.1 | 212.0 | 420 | 965 | 0.78 (0.65 - 0.95) | 0.01 | 0.80 (0.65 - 0.99) | 0.04 |

##### 1.6.11 Abnormal weight loss

**Supplementary Table 141:** Exposures, outcomes and risk estimates for Abnormal weight loss in DPP4I (male) (T) vs SU (male) (C) new-user cohorts under four different propensity score (PS) adjustment / time-at-risk (TAR) designs across data sources. We report total patient exposure time in 1,000 patient-year (KYr) units, uncalibrated and calibrated hazard ratios (HRs) and their 95% confidence intervals (CIs). Greyed out entries did not pass study diagnostics and are included only for completeness.

| | Patients | | Exposure (KYr) | | Outcomes | | Uncalibrated | | Calibrated | |
|---|---|---|---|---|---|---|---|---|---|---|
| | T | C | T | C | T | C | HR (95% CI) | $p$ | HR (95% CI) | $p$ |
| **PS-matching / on-treatment TAR** | | | | | | | | | | |
| CCAE | 46,342 | 83,167 | 40.5 | 71.0 | 261 | 346 | 1.19 (0.95 - 1.48) | 0.13 | 1.21 (0.96 - 1.51) | 0.12 |
| DAG | 13,155 | 2,317 | 14.0 | 2.4 | 89 | 13 | 2.21 (0.99 - 5.91) | 0.08 | 2.38 (0.97 - 5.85) | 0.07 |
| ODOD | 27,995 | 67,645 | 23.3 | 63.4 | 353 | 834 | 1.13 (0.95 - 1.35) | 0.17 | 1.18 (0.98 - 1.41) | 0.10 |
| MDCR | 8,637 | 17,435 | 9.0 | 18.4 | 128 | 243 | 1.04 (0.77 - 1.41) | 0.78 | 1.03 (0.76 - 1.39) | 0.86 |
| MDCD | 3,890 | 9,392 | 3.0 | 6.7 | 45 | 81 | 1.19 (0.68 - 2.08) | 0.54 | 1.24 (0.71 - 2.18) | 0.46 |
| Open Claims | 407,051 | 871,924 | 388.0 | 946.2 | 2,452 | 4,960 | 1.16 (1.09 - 1.24) | <0.01 | 1.16 (1.01 - 1.33) | 0.04 |
| OEHR | 35,345 | 92,794 | 17.4 | 51.2 | 203 | 551 | 1.21 (0.97 - 1.50) | 0.08 | 1.29 (1.01 - 1.66) | 0.07 |
| VA | 0 | 0 | 0.0 | 0.0 | 0 | 0 | | | | |
| SIDIAP | 26,520 | 7,185 | 77.5 | 20.4 | 259 | 39 | 0.99 (0.66 - 1.54) | 0.98 | 0.94 (0.62 - 1.44) | 0.79 |
| Meta-analysis | 568,935 | 1,151,859 | 572.7 | 1179.7 | 3,790 | 7,067 | 1.16 (1.10 - 1.23) | <0.01 | 1.17 (1.03 - 1.31) | 0.01 |
| Meta-analysis (w/o USOC) | 161,884 | 279,935 | 184.7 | 233.5 | 1,338 | 2,107 | 1.15 (1.04 - 1.28) | <0.01 | 1.18 (1.04 - 1.34) | 0.03 |
| **PS-stratification / on-treatment TAR** | | | | | | | | | | |
| CCAE | 51,674 | 84,669 | 45.3 | 72.2 | 298 | 348 | 1.36 (1.15 - 1.60) | <0.01 | 1.37 (1.16 - 1.61) | <0.01 |
| DAG | 13,183 | 2,676 | 14.0 | 2.8 | 89 | 15 | 1.26 (0.71 - 2.39) | 0.45 | 1.35 (0.73 - 2.47) | 0.35 |
| ODOD | 29,544 | 67,651 | 24.8 | 63.4 | 379 | 834 | 1.23 (1.08 - 1.40) | <0.01 | 1.24 (1.09 - 1.42) | <0.01 |
| MDCR | 9,144 | 17,821 | 9.6 | 18.9 | 137 | 244 | 1.08 (0.86 - 1.34) | 0.50 | 1.09 (0.87 - 1.36) | 0.46 |
| MDCD | 3,979 | 9,412 | 3.0 | 6.7 | 46 | 81 | 1.23 (0.83 - 1.79) | 0.29 | 1.26 (0.86 - 1.85) | 0.26 |
| Open Claims | 414,330 | 871,935 | 397.1 | 946.3 | 2,536 | 4,960 | 1.21 (1.16 - 1.28) | <0.01 | 1.20 (1.03 - 1.40) | 0.02 |
| OEHR | 37,139 | 92,820 | 18.5 | 51.3 | 221 | 552 | 1.18 (1.00 - 1.40) | 0.05 | 1.30 (1.10 - 1.54) | <0.01 |
| VA | 24,656 | 226,229 | 23.0 | 240.4 | 212 | 1,476 | 1.50 (1.30 - 1.73) | <0.01 | 1.55 (0.52 - 4.68) | 0.44 |
| SIDIAP | 26,609 | 9,240 | 77.6 | 27.5 | 259 | 43 | 1.15 (0.81 - 1.66) | 0.45 | 1.13 (0.79 - 1.61) | 0.53 |
| Meta-analysis | 585,602 | 1,156,224 | 589.9 | 1189.0 | 3,965 | 7,077 | 1.22 (1.17 - 1.27) | <0.01 | 1.23 (1.12 - 1.35) | <0.01 |
| Meta-analysis (w/o USOC) | 171,272 | 284,289 | 192.8 | 242.8 | 1,429 | 2,117 | 1.22 (1.13 - 1.32) | <0.01 | 1.25 (1.12 - 1.39) | <0.01 |
| **PS-matching / on-treatment TAR without escalation** | | | | | | | | | | |
| CCAE | 46,342 | 83,167 | 32.7 | 61.7 | 202 | 298 | 1.24 (0.97 - 1.57) | 0.08 | 1.27 (1.00 - 1.61) | 0.06 |
| DAG | 13,155 | 2,317 | 12.6 | 2.2 | 78 | 11 | 2.19 (0.91 - 6.54) | 0.12 | 2.38 (0.88 - 6.39) | 0.10 |
| ODOD | 27,995 | 67,645 | 19.1 | 56.5 | 298 | 733 | 1.11 (0.91 - 1.34) | 0.31 | 1.16 (0.96 - 1.42) | 0.15 |
| MDCR | 8,637 | 17,435 | 7.7 | 16.8 | 112 | 215 | 1.05 (0.76 - 1.45) | 0.77 | 1.05 (0.76 - 1.46) | 0.76 |
| MDCD | 3,890 | 9,392 | 2.3 | 5.6 | 29 | 62 | 1.16 (0.60 - 2.20) | 0.66 | 1.21 (0.63 - 2.33) | 0.57 |
| Open Claims | 407,051 | 871,924 | 300.9 | 801.7 | 1,928 | 4,225 | 1.15 (1.06 - 1.23) | <0.01 | 1.15 (1.01 - 1.31) | 0.04 |
| OEHR | 35,345 | 92,794 | 14.2 | 44.6 | 148 | 447 | 1.20 (0.93 - 1.52) | 0.15 | 1.30 (0.98 - 1.72) | 0.11 |
| VA | 0 | 0 | 0.0 | 0.0 | 0 | 0 | | | | |
| SIDIAP | 26,520 | 7,185 | 56.8 | 17.0 | 194 | 29 | 1.11 (0.68 - 1.87) | 0.69 | 1.03 (0.62 - 1.70) | 0.89 |
| Meta-analysis | 568,935 | 1,151,859 | 446.2 | 1006.0 | 2,989 | 6,020 | 1.15 (1.08 - 1.22) | <0.01 | 1.16 (1.03 - 1.32) | 0.02 |
| Meta-analysis (w/o USOC) | 161,884 | 279,935 | 145.3 | 204.4 | 1,061 | 1,795 | 1.16 (1.03 - 1.30) | 0.01 | 1.20 (1.03 - 1.39) | 0.03 |
| **PS-stratification / on-treatment TAR without escalation** | | | | | | | | | | |
| CCAE | 51,674 | 84,669 | 36.6 | 62.9 | 232 | 300 | 1.33 (1.11 - 1.60) | <0.01 | 1.35 (1.13 - 1.63) | <0.01 |
| DAG | 13,183 | 2,676 | 12.6 | 2.5 | 78 | 12 | 1.44 (0.76 - 2.97) | 0.29 | 1.57 (0.80 - 3.11) | 0.21 |
| ODOD | 29,544 | 67,651 | 20.3 | 56.5 | 318 | 733 | 1.28 (1.11 - 1.47) | <0.01 | 1.30 (1.12 - 1.50) | <0.01 |
| MDCR | 9,144 | 17,821 | 8.2 | 17.2 | 121 | 216 | 1.14 (0.90 - 1.43) | 0.29 | 1.14 (0.90 - 1.44) | 0.30 |
| MDCD | 3,979 | 9,412 | 2.3 | 5.6 | 30 | 62 | 1.21 (0.76 - 1.90) | 0.41 | 1.23 (0.78 - 1.95) | 0.39 |
| Open Claims | 414,330 | 871,935 | 307.7 | 801.7 | 1,999 | 4,225 | 1.22 (1.15 - 1.28) | <0.01 | 1.21 (1.02 - 1.43) | 0.03 |
| OEHR | 37,139 | 92,820 | 15.0 | 44.6 | 164 | 448 | 1.16 (0.96 - 1.41) | 0.12 | 1.29 (1.06 - 1.57) | 0.02 |
| VA | 24,656 | 226,229 | 18.7 | 214.1 | 174 | 1,280 | 1.54 (1.31 - 1.80) | <0.01 | 1.56 (0.51 - 4.75) | 0.44 |
| SIDIAP | 26,609 | 9,240 | 56.9 | 22.8 | 194 | 32 | 1.14 (0.76 - 1.76) | 0.55 | 1.09 (0.72 - 1.67) | 0.69 |
| Meta-analysis | 585,602 | 1,156,224 | 459.7 | 1013.8 | 3,136 | 6,028 | 1.22 (1.17 - 1.28) | <0.01 | 1.23 (1.09 - 1.39) | <0.01 |
| Meta-analysis (w/o USOC) | 171,272 | 284,289 | 151.9 | 212.1 | 1,137 | 1,803 | 1.24 (1.14 - 1.35) | <0.01 | 1.27 (1.12 - 1.43) | <0.01 |

##### 1.6.12 Acute pancreatitis

**Supplementary Table 142:** Exposures, outcomes and risk estimates for Acute pancreatitis in DPP4I (male) (T) vs SU (male) (C) new-user cohorts under four different propensity score (PS) adjustment / time-at-risk (TAR) designs across data sources. We report total patient exposure time in 1,000 patient-year (KYr) units, uncalibrated and calibrated hazard ratios (HRs) and their 95% confidence intervals (CIs). Greyed out entries did not pass study diagnostics and are included only for completeness.

| | Patients | | Exposure (KYr) | | Outcomes | | Uncalibrated | | Calibrated | |
|---|---|---|---|---|---|---|---|---|---|---|
| | T | C | T | C | T | C | HR (95% CI) | $p$ | HR (95% CI) | $p$ |
| **PS-matching / on-treatment TAR** | | | | | | | | | | |
| CCAE | 47,045 | 84,234 | 41.2 | 71.9 | 104 | 199 | 0.91 (0.65 - 1.26) | 0.56 | 0.92 (0.66 - 1.28) | 0.63 |
| DAG | 13,419 | 2,361 | 14.3 | 2.4 | 0 | 0 | | | | |
| ODOD | 28,998 | 69,988 | 24.4 | 65.7 | 62 | 238 | 0.88 (0.60 - 1.27) | 0.50 | 0.91 (0.63 - 1.33) | 0.65 |
| MDCR | 8,967 | 18,042 | 9.3 | 19.0 | 38 | 70 | 1.25 (0.71 - 2.15) | 0.44 | 1.23 (0.70 - 2.14) | 0.48 |
| MDCD | 4,011 | 9,564 | 3.1 | 6.9 | 16 | 56 | 0.61 (0.26 - 1.30) | 0.22 | 0.64 (0.29 - 1.41) | 0.27 |
| Open Claims | 417,329 | 889,404 | 398.9 | 966.5 | 793 | 2,182 | 0.94 (0.84 - 1.04) | 0.24 | 0.93 (0.80 - 1.10) | 0.42 |
| OEHR | 36,266 | 95,267 | 18.0 | 52.9 | 43 | 164 | 1.09 (0.68 - 1.69) | 0.72 | 1.16 (0.72 - 1.86) | 0.56 |
| VA | 0 | 0 | 0.0 | 0.0 | 0 | 0 | | | | |
| SIDIAP | 26,706 | 7,204 | 78.2 | 20.5 | 30 | 8 | 0.93 (0.31 - 3.10) | 0.90 | 0.88 (0.28 - 2.78) | 0.83 |
| Meta-analysis | 569,322 | 1,173,703 | 573.1 | 1203.3 | 1,086 | 2,917 | 0.94 (0.85 - 1.03) | 0.19 | 0.94 (0.82 - 1.09) | 0.41 |
| Meta-analysis (w/o USOC) | 151,993 | 284,299 | 174.2 | 236.9 | 293 | 735 | 0.94 (0.78 - 1.15) | 0.56 | 0.97 (0.78 - 1.19) | 0.75 |
| **PS-stratification / on-treatment TAR** | | | | | | | | | | |
| CCAE | 52,487 | 85,745 | 46.2 | 73.2 | 112 | 201 | 0.92 (0.72 - 1.17) | 0.48 | 0.92 (0.72 - 1.18) | 0.53 |
| DAG | 13,447 | 2,736 | 14.3 | 2.8 | 0 | 0 | | | | |
| ODOD | 30,607 | 69,996 | 25.9 | 65.7 | 67 | 238 | 0.75 (0.56 - 0.99) | 0.05 | 0.76 (0.57 - 1.01) | 0.06 |
| MDCR | 9,484 | 18,431 | 9.9 | 19.5 | 38 | 72 | 1.10 (0.72 - 1.66) | 0.65 | 1.12 (0.73 - 1.70) | 0.61 |
| MDCD | 4,102 | 9,580 | 3.1 | 6.9 | 18 | 56 | 0.73 (0.41 - 1.26) | 0.28 | 0.75 (0.43 - 1.32) | 0.33 |
| Open Claims | 424,808 | 889,411 | 408.4 | 966.5 | 813 | 2,182 | 0.89 (0.82 - 0.96) | <0.01 | 0.88 (0.74 - 1.04) | 0.14 |
| OEHR | 38,129 | 95,297 | 19.1 | 52.9 | 44 | 164 | 0.95 (0.66 - 1.34) | 0.79 | 1.05 (0.74 - 1.49) | 0.80 |
| VA | 25,396 | 231,330 | 23.8 | 246.1 | 42 | 510 | 0.84 (0.61 - 1.14) | 0.29 | 0.87 (0.28 - 2.72) | 0.82 |
| SIDIAP | 26,821 | 9,254 | 78.3 | 27.6 | 30 | 12 | 0.84 (0.40 - 1.86) | 0.65 | 0.82 (0.38 - 1.77) | 0.62 |
| Meta-analysis | 586,438 | 1,177,714 | 591.0 | 1212.2 | 1,122 | 2,925 | 0.89 (0.82 - 0.95) | <0.01 | 0.89 (0.80 - 1.00) | 0.05 |
| Meta-analysis (w/o USOC) | 161,630 | 288,303 | 182.6 | 245.8 | 309 | 743 | 0.88 (0.76 - 1.02) | 0.08 | 0.90 (0.76 - 1.06) | 0.20 |
| **PS-matching / on-treatment TAR without escalation** | | | | | | | | | | |
| CCAE | 47,045 | 84,234 | 33.3 | 62.5 | 75 | 168 | 0.90 (0.62 - 1.28) | 0.55 | 0.92 (0.64 - 1.32) | 0.64 |
| DAG | 13,419 | 2,361 | 12.9 | 2.2 | 0 | 0 | | | | |
| ODOD | 28,998 | 69,988 | 20.0 | 58.6 | 48 | 208 | 0.77 (0.50 - 1.15) | 0.22 | 0.81 (0.54 - 1.23) | 0.33 |
| MDCR | 8,967 | 18,042 | 8.0 | 17.3 | 36 | 61 | 1.38 (0.77 - 2.45) | 0.27 | 1.39 (0.78 - 2.47) | 0.27 |
| MDCD | 4,011 | 9,564 | 2.3 | 5.8 | 12 | 50 | 0.62 (0.24 - 1.45) | 0.30 | 0.65 (0.26 - 1.60) | 0.35 |
| Open Claims | 417,329 | 889,404 | 309.4 | 819.0 | 582 | 1,807 | 0.91 (0.80 - 1.03) | 0.12 | 0.91 (0.77 - 1.07) | 0.27 |
| OEHR | 36,266 | 95,267 | 14.6 | 46.0 | 31 | 120 | 1.22 (0.72 - 2.02) | 0.45 | 1.32 (0.77 - 2.26) | 0.32 |
| VA | 0 | 0 | 0.0 | 0.0 | 0 | 0 | | | | |
| SIDIAP | 26,706 | 7,204 | 57.3 | 17.1 | 26 | 7 | 0.93 (0.30 - 3.17) | 0.90 | 0.86 (0.26 - 2.81) | 0.80 |
| Meta-analysis | 569,322 | 1,173,703 | 444.9 | 1026.3 | 810 | 2,421 | 0.92 (0.82 - 1.02) | 0.11 | 0.93 (0.80 - 1.08) | 0.33 |
| Meta-analysis (w/o USOC) | 151,993 | 284,299 | 135.5 | 207.3 | 228 | 614 | 0.94 (0.76 - 1.17) | 0.59 | 0.97 (0.77 - 1.23) | 0.83 |
| **PS-stratification / on-treatment TAR without escalation** | | | | | | | | | | |
| CCAE | 52,487 | 85,745 | 37.3 | 63.7 | 81 | 170 | 0.85 (0.64 - 1.11) | 0.24 | 0.86 (0.65 - 1.14) | 0.30 |
| DAG | 13,447 | 2,736 | 12.9 | 2.6 | 0 | 0 | | | | |
| ODOD | 30,607 | 69,996 | 21.1 | 58.6 | 52 | 208 | 0.71 (0.51 - 0.97) | 0.04 | 0.72 (0.52 - 0.99) | 0.05 |
| MDCR | 9,484 | 18,431 | 8.5 | 17.8 | 36 | 63 | 1.30 (0.84 - 2.00) | 0.24 | 1.31 (0.84 - 2.02) | 0.24 |
| MDCD | 4,102 | 9,580 | 2.4 | 5.8 | 13 | 50 | 0.67 (0.34 - 1.24) | 0.23 | 0.68 (0.36 - 1.30) | 0.26 |
| Open Claims | 424,808 | 889,411 | 316.6 | 819.0 | 596 | 1,807 | 0.85 (0.77 - 0.94) | <0.01 | 0.85 (0.70 - 1.02) | 0.08 |
| OEHR | 38,129 | 95,297 | 15.5 | 46.0 | 32 | 120 | 1.03 (0.67 - 1.53) | 0.89 | 1.15 (0.76 - 1.73) | 0.52 |
| VA | 25,396 | 231,330 | 19.3 | 219.3 | 31 | 424 | 0.81 (0.55 - 1.14) | 0.26 | 0.82 (0.26 - 2.61) | 0.74 |
| SIDIAP | 26,821 | 9,254 | 57.4 | 22.8 | 26 | 11 | 0.88 (0.40 - 2.09) | 0.77 | 0.85 (0.37 - 1.95) | 0.70 |
| Meta-analysis | 586,438 | 1,177,714 | 458.9 | 1033.7 | 836 | 2,429 | 0.86 (0.79 - 0.93) | <0.01 | 0.86 (0.75 - 0.99) | 0.04 |
| Meta-analysis (w/o USOC) | 161,630 | 288,303 | 142.3 | 214.6 | 240 | 622 | 0.88 (0.73 - 1.06) | 0.18 | 0.90 (0.73 - 1.11) | 0.34 |

##### 1.6.13 Bone fracture

**Supplementary Table 143:** Exposures, outcomes and risk estimates for Bone fracture in DPP4I (male) (T) vs SU (male) (C) new-user cohorts under four different propensity score (PS) adjustment / time-at-risk (TAR) designs across data sources. We report total patient exposure time in 1,000 patient-year (KYr) units, uncalibrated and calibrated hazard ratios (HRs) and their 95% confidence intervals (CIs). Greyed out entries did not pass study diagnostics and are included only for completeness.

| | Patients | | Exposure (KYr) | | Outcomes | | Uncalibrated | | Calibrated | |
|---|---|---|---|---|---|---|---|---|---|---|
| | T | C | T | C | T | C | HR (95% CI) | $p$ | HR (95% CI) | $p$ |
| **PS-matching / on-treatment TAR** | | | | | | | | | | |
| CCAE | 44,226 | 79,432 | 38.1 | 66.8 | 555 | 1,030 | 0.86 (0.74 - 1.00) | 0.05 | 0.87 (0.75 - 1.02) | 0.12 |
| DAG | 11,999 | 2,144 | 12.6 | 2.2 | 208 | 41 | 0.92 (0.61 - 1.44) | 0.71 | 1.00 (0.64 - 1.55) | 0.74 |
| ODOD | 27,120 | 66,034 | 22.4 | 61.0 | 438 | 1,355 | 0.88 (0.75 - 1.03) | 0.11 | 0.92 (0.78 - 1.07) | 0.32 |
| MDCR | 8,218 | 16,511 | 8.3 | 17.0 | 228 | 465 | 0.98 (0.78 - 1.23) | 0.85 | 0.96 (0.76 - 1.21) | 0.75 |
| MDCD | 3,584 | 8,620 | 2.7 | 6.1 | 84 | 204 | 0.98 (0.68 - 1.40) | 0.92 | 1.02 (0.71 - 1.47) | 0.86 |
| Open Claims | 378,952 | 811,712 | 358.5 | 873.7 | 4,866 | 12,692 | 0.95 (0.91 - 1.00) | 0.04 | 0.95 (0.84 - 1.08) | 0.44 |
| OEHR | 34,463 | 90,094 | 16.9 | 49.2 | 271 | 893 | 1.02 (0.84 - 1.25) | 0.81 | 1.09 (0.87 - 1.38) | 0.51 |
| VA | 0 | 0 | 0.0 | 0.0 | 0 | 0 | | | | |
| SIDIAP | 24,737 | 6,841 | 72.1 | 19.2 | 682 | 202 | 0.78 (0.63 - 0.97) | 0.02 | 0.74 (0.60 - 0.92) | 0.01 |
| Meta-analysis | 533,299 | 1,081,388 | 531.6 | 1095.1 | 7,332 | 16,882 | 0.93 (0.89 - 0.98) | <0.01 | 0.94 (0.83 - 1.05) | 0.26 |
| Meta-analysis (w/o USOC) | 154,347 | 269,676 | 173.1 | 221.4 | 2,466 | 4,190 | 0.90 (0.83 - 0.97) | <0.01 | 0.92 (0.83 - 1.03) | 0.20 |
| **PS-stratification / on-treatment TAR** | | | | | | | | | | |
| CCAE | 49,297 | 80,914 | 42.6 | 68.1 | 633 | 1,044 | 0.95 (0.86 - 1.05) | 0.34 | 0.96 (0.86 - 1.07) | 0.47 |
| DAG | 12,024 | 2,488 | 12.6 | 2.6 | 209 | 44 | 0.87 (0.61 - 1.25) | 0.43 | 0.92 (0.64 - 1.33) | 0.67 |
| ODOD | 28,643 | 66,040 | 23.8 | 61.0 | 462 | 1,355 | 0.92 (0.82 - 1.02) | 0.12 | 0.92 (0.82 - 1.04) | 0.23 |
| MDCR | 8,670 | 16,876 | 8.8 | 17.5 | 242 | 473 | 0.98 (0.83 - 1.15) | 0.79 | 0.99 (0.84 - 1.17) | 0.86 |
| MDCD | 3,661 | 8,636 | 2.8 | 6.1 | 85 | 204 | 0.91 (0.70 - 1.19) | 0.51 | 0.94 (0.72 - 1.22) | 0.66 |
| Open Claims | 385,636 | 811,724 | 366.8 | 873.7 | 4,994 | 12,692 | 0.95 (0.92 - 0.98) | <0.01 | 0.94 (0.81 - 1.09) | 0.44 |
| OEHR | 36,215 | 90,122 | 17.9 | 49.2 | 284 | 894 | 1.04 (0.90 - 1.20) | 0.56 | 1.15 (0.99 - 1.32) | 0.08 |
| VA | 23,057 | 213,933 | 21.6 | 226.0 | 216 | 2,435 | 0.93 (0.81 - 1.07) | 0.31 | 0.96 (0.32 - 2.89) | 0.92 |
| SIDIAP | 24,839 | 8,816 | 72.2 | 25.8 | 682 | 266 | 0.88 (0.75 - 1.04) | 0.13 | 0.86 (0.73 - 1.02) | 0.10 |
| Meta-analysis | 548,985 | 1,085,616 | 547.6 | 1103.9 | 7,591 | 16,972 | 0.95 (0.92 - 0.98) | <0.01 | 0.96 (0.87 - 1.05) | 0.34 |
| Meta-analysis (w/o USOC) | 163,349 | 273,892 | 180.7 | 230.2 | 2,597 | 4,280 | 0.95 (0.89 - 1.00) | 0.05 | 0.97 (0.88 - 1.06) | 0.49 |
| **PS-matching / on-treatment TAR without escalation** | | | | | | | | | | |
| CCAE | 44,226 | 79,432 | 30.9 | 58.3 | 449 | 895 | 0.85 (0.72 - 1.00) | 0.05 | 0.87 (0.74 - 1.02) | 0.11 |
| DAG | 11,999 | 2,144 | 11.4 | 2.0 | 194 | 41 | 0.83 (0.54 - 1.31) | 0.41 | 0.90 (0.57 - 1.41) | 0.65 |
| ODOD | 27,120 | 66,034 | 18.4 | 54.5 | 352 | 1,222 | 0.84 (0.71 - 0.99) | 0.04 | 0.88 (0.74 - 1.05) | 0.18 |
| MDCR | 8,218 | 16,511 | 7.1 | 15.5 | 199 | 430 | 0.95 (0.75 - 1.21) | 0.69 | 0.96 (0.75 - 1.22) | 0.72 |
| MDCD | 3,584 | 8,620 | 2.1 | 5.2 | 62 | 171 | 1.05 (0.70 - 1.57) | 0.80 | 1.10 (0.74 - 1.65) | 0.65 |
| Open Claims | 378,952 | 811,712 | 278.7 | 741.7 | 3,739 | 10,693 | 0.94 (0.89 - 0.99) | 0.02 | 0.94 (0.84 - 1.06) | 0.33 |
| OEHR | 34,463 | 90,094 | 13.7 | 42.9 | 198 | 745 | 0.99 (0.79 - 1.23) | 0.89 | 1.07 (0.82 - 1.39) | 0.67 |
| VA | 0 | 0 | 0.0 | 0.0 | 0 | 0 | | | | |
| SIDIAP | 24,737 | 6,841 | 53.0 | 16.0 | 502 | 171 | 0.76 (0.60 - 0.96) | 0.02 | 0.70 (0.55 - 0.90) | <0.01 |
| Meta-analysis | 533,299 | 1,081,388 | 415.3 | 936.0 | 5,695 | 14,368 | 0.91 (0.86 - 0.96) | <0.01 | 0.92 (0.81 - 1.04) | 0.18 |
| Meta-analysis (w/o USOC) | 154,347 | 269,676 | 136.7 | 194.3 | 1,956 | 3,675 | 0.87 (0.80 - 0.95) | <0.01 | 0.90 (0.79 - 1.02) | 0.14 |
| **PS-stratification / on-treatment TAR without escalation** | | | | | | | | | | |
| CCAE | 49,297 | 80,914 | 34.6 | 59.4 | 511 | 909 | 0.94 (0.84 - 1.06) | 0.32 | 0.96 (0.85 - 1.08) | 0.53 |
| DAG | 12,024 | 2,488 | 11.4 | 2.3 | 195 | 43 | 0.82 (0.57 - 1.19) | 0.28 | 0.89 (0.62 - 1.29) | 0.57 |
| ODOD | 28,643 | 66,040 | 19.6 | 54.5 | 372 | 1,222 | 0.88 (0.78 - 1.00) | 0.05 | 0.90 (0.79 - 1.02) | 0.13 |
| MDCR | 8,670 | 16,876 | 7.6 | 16.0 | 211 | 437 | 0.99 (0.83 - 1.17) | 0.87 | 0.99 (0.83 - 1.18) | 0.86 |
| MDCD | 3,661 | 8,636 | 2.1 | 5.2 | 63 | 171 | 0.90 (0.66 - 1.21) | 0.49 | 0.91 (0.68 - 1.24) | 0.59 |
| Open Claims | 385,636 | 811,724 | 285.0 | 741.7 | 3,843 | 10,693 | 0.94 (0.91 - 0.98) | <0.01 | 0.94 (0.80 - 1.10) | 0.44 |
| OEHR | 36,215 | 90,122 | 14.6 | 42.9 | 209 | 745 | 0.97 (0.82 - 1.13) | 0.68 | 1.08 (0.91 - 1.27) | 0.45 |
| VA | 23,057 | 213,933 | 17.6 | 201.8 | 166 | 2,146 | 0.88 (0.75 - 1.03) | 0.13 | 0.89 (0.29 - 2.72) | 0.85 |
| SIDIAP | 24,839 | 8,816 | 53.1 | 21.4 | 502 | 225 | 0.86 (0.71 - 1.03) | 0.11 | 0.83 (0.68 - 1.00) | 0.07 |
| Meta-analysis | 548,985 | 1,085,616 | 427.9 | 943.3 | 5,906 | 14,445 | 0.94 (0.91 - 0.97) | <0.01 | 0.94 (0.84 - 1.06) | 0.34 |
| Meta-analysis (w/o USOC) | 163,349 | 273,892 | 142.9 | 201.7 | 2,063 | 3,752 | 0.92 (0.87 - 0.98) | <0.01 | 0.94 (0.85 - 1.05) | 0.31 |

##### 1.6.14 Diabetic ketoacidosis

**Supplementary Table 144:** Exposures, outcomes and risk estimates for Diabetic ketoacidosis in DPP4I (male) (T) vs SU (male) (C) new-user cohorts under four different propensity score (PS) adjustment / time-at-risk (TAR) designs across data sources. We report total patient exposure time in 1,000 patient-year (KYr) units, uncalibrated and calibrated hazard ratios (HRs) and their 95% confidence intervals (CIs). Greyed out entries did not pass study diagnostics and are included only for completeness.

| | Patients | | Exposure (KYr) | | Outcomes | | Uncalibrated | | Calibrated | |
|---|---|---|---|---|---|---|---|---|---|---|
| | T | C | T | C | T | C | HR (95% CI) | $p$ | HR (95% CI) | $p$ |
| **PS-matching / on-treatment TAR** | | | | | | | | | | |
| CCAE | 47,391 | 85,013 | 41.6 | 72.6 | 34 | 50 | 1.99 (1.09 - 3.60) | 0.02 | 2.02 (1.11 - 3.66) | 0.02 |
| DAG | 13,419 | 2,361 | 14.3 | 2.4 | 0 | 0 | | | | |
| ODOD | 29,199 | 70,642 | 24.5 | 66.5 | 37 | 87 | 1.67 (0.98 - 2.80) | 0.06 | 1.73 (1.02 - 2.93) | 0.04 |
| MDCR | 9,041 | 18,266 | 9.4 | 19.2 | 7 | 23 | 1.02 (0.30 - 3.12) | 0.98 | 1.00 (0.31 - 3.23) | 0.97 |
| MDCD | 4,072 | 9,788 | 3.1 | 7.0 | 5 | 39 | 0.51 (0.17 - 1.28) | 0.19 | 0.53 (0.19 - 1.47) | 0.22 |
| Open Claims | 421,605 | 900,893 | 403.0 | 979.8 | 299 | 788 | 1.06 (0.89 - 1.27) | 0.52 | 1.06 (0.86 - 1.31) | 0.61 |
| OEHR | 36,419 | 95,809 | 18.1 | 53.3 | 24 | 62 | 1.26 (0.64 - 2.37) | 0.49 | 1.34 (0.69 - 2.61) | 0.40 |
| VA | 0 | 0 | 0.0 | 0.0 | 0 | 0 | | | | |
| SIDIAP | 26,798 | 7,217 | 78.5 | 20.5 | 0 | <5 | | | | |
| Meta-analysis | 547,727 | 1,180,411 | 499.7 | 1198.5 | 406 | 1,049 | 1.23 (0.93 - 1.64) | 0.15 | 1.24 (0.91 - 1.68) | 0.17 |
| Meta-analysis (w/o USOC) | 126,122 | 279,518 | 96.7 | 218.7 | 107 | 261 | 1.35 (0.91 - 2.00) | 0.13 | 1.38 (0.93 - 2.06) | 0.12 |
| **PS-stratification / on-treatment TAR** | | | | | | | | | | |
| CCAE | 52,836 | 86,530 | 46.5 | 73.9 | 36 | 52 | 1.36 (0.86 - 2.13) | 0.18 | 1.37 (0.87 - 2.16) | 0.18 |
| DAG | 13,447 | 2,736 | 14.3 | 2.8 | 0 | 0 | | | | |
| ODOD | 30,816 | 70,649 | 26.1 | 66.5 | 38 | 87 | 1.28 (0.85 - 1.89) | 0.23 | 1.29 (0.86 - 1.93) | 0.22 |
| MDCR | 9,553 | 18,658 | 10.0 | 19.7 | 7 | 23 | 0.70 (0.27 - 1.60) | 0.43 | 0.70 (0.29 - 1.72) | 0.44 |
| MDCD | 4,168 | 9,808 | 3.2 | 7.1 | 5 | 39 | 0.31 (0.11 - 0.75) | 0.02 | 0.32 (0.12 - 0.86) | 0.02 |
| Open Claims | 429,005 | 900,907 | 412.3 | 979.8 | 304 | 788 | 0.99 (0.86 - 1.13) | 0.85 | 0.98 (0.80 - 1.19) | 0.84 |
| OEHR | 38,263 | 95,839 | 19.2 | 53.3 | 24 | 62 | 1.30 (0.78 - 2.11) | 0.30 | 1.43 (0.87 - 2.36) | 0.16 |
| VA | 25,614 | 233,668 | 24.0 | 248.9 | 11 | 126 | 0.87 (0.44 - 1.54) | 0.67 | 0.90 (0.26 - 3.17) | 0.88 |
| SIDIAP | 26,891 | 9,270 | 78.6 | 27.7 | 0 | <5 | | | | |
| Meta-analysis | 564,641 | 1,182,391 | 517.3 | 1200.3 | 414 | 1,051 | 1.05 (0.88 - 1.26) | 0.58 | 1.06 (0.87 - 1.29) | 0.56 |
| Meta-analysis (w/o USOC) | 135,636 | 281,484 | 105.0 | 220.5 | 110 | 263 | 1.03 (0.70 - 1.52) | 0.88 | 1.05 (0.71 - 1.56) | 0.80 |
| **PS-matching / on-treatment TAR without escalation** | | | | | | | | | | |
| CCAE | 47,391 | 85,013 | 33.5 | 63.1 | 27 | 38 | 1.88 (0.99 - 3.55) | 0.05 | 1.92 (1.01 - 3.64) | 0.05 |
| DAG | 13,419 | 2,361 | 12.9 | 2.2 | 0 | 0 | | | | |
| ODOD | 29,199 | 70,642 | 20.1 | 59.2 | 26 | 65 | 1.61 (0.88 - 2.91) | 0.12 | 1.70 (0.93 - 3.10) | 0.09 |
| MDCR | 9,041 | 18,266 | 8.0 | 17.5 | 6 | 19 | 0.65 (0.14 - 2.33) | 0.55 | 0.66 (0.16 - 2.68) | 0.56 |
| MDCD | 4,072 | 9,788 | 2.4 | 5.9 | 5 | 31 | 0.59 (0.19 - 1.53) | 0.32 | 0.62 (0.22 - 1.76) | 0.37 |
| Open Claims | 421,605 | 900,893 | 312.5 | 830.0 | 212 | 606 | 1.03 (0.84 - 1.26) | 0.74 | 1.04 (0.83 - 1.30) | 0.75 |
| OEHR | 36,419 | 95,809 | 14.7 | 46.3 | 17 | 35 | 1.65 (0.77 - 3.42) | 0.19 | 1.79 (0.84 - 3.83) | 0.14 |
| VA | 0 | 0 | 0.0 | 0.0 | 0 | 0 | | | | |
| SIDIAP | 26,798 | 7,217 | 57.5 | 17.1 | 0 | <5 | | | | |
| Meta-analysis | 547,727 | 1,180,411 | 391.2 | 1022.0 | 293 | 794 | 1.22 (0.91 - 1.64) | 0.18 | 1.24 (0.90 - 1.69) | 0.19 |
| Meta-analysis (w/o USOC) | 126,122 | 279,518 | 78.7 | 192.1 | 81 | 188 | 1.40 (0.94 - 2.07) | 0.10 | 1.44 (0.96 - 2.16) | 0.08 |
| **PS-stratification / on-treatment TAR without escalation** | | | | | | | | | | |
| CCAE | 52,836 | 86,530 | 37.6 | 64.3 | 28 | 40 | 1.52 (0.91 - 2.53) | 0.11 | 1.55 (0.93 - 2.58) | 0.10 |
| DAG | 13,447 | 2,736 | 12.9 | 2.6 | 0 | 0 | | | | |
| ODOD | 30,816 | 70,649 | 21.3 | 59.3 | 26 | 65 | 1.26 (0.77 - 2.00) | 0.34 | 1.28 (0.80 - 2.07) | 0.31 |
| MDCR | 9,553 | 18,658 | 8.6 | 18.0 | 6 | 19 | 0.75 (0.26 - 1.85) | 0.56 | 0.75 (0.28 - 1.99) | 0.57 |
| MDCD | 4,168 | 9,808 | 2.4 | 5.9 | 5 | 31 | 0.44 (0.15 - 1.08) | 0.11 | 0.45 (0.17 - 1.22) | 0.12 |
| Open Claims | 429,005 | 900,907 | 319.5 | 830.0 | 217 | 606 | 0.98 (0.84 - 1.15) | 0.84 | 0.98 (0.78 - 1.22) | 0.84 |
| OEHR | 38,263 | 95,839 | 15.6 | 46.3 | 17 | 35 | 1.76 (0.94 - 3.20) | 0.07 | 1.96 (1.06 - 3.63) | 0.03 |
| VA | 25,614 | 233,668 | 19.5 | 221.6 | <10 | 91 | 0.69 (0.27 - 1.46) | 0.39 | 0.70 (0.18 - 2.81) | 0.62 |
| SIDIAP | 26,891 | 9,270 | 57.6 | 22.9 | 0 | <5 | | | | |
| Meta-analysis | 564,641 | 1,182,391 | 404.9 | 1023.7 | 299 | 796 | 1.12 (0.87 - 1.44) | 0.39 | 1.13 (0.85 - 1.49) | 0.40 |
| Meta-analysis (w/o USOC) | 135,636 | 281,484 | 85.4 | 193.8 | 82 | 190 | 1.19 (0.80 - 1.75) | 0.39 | 1.21 (0.81 - 1.81) | 0.34 |

##### 1.6.15 Diarrhea

**Supplementary Table 145:** Exposures, outcomes and risk estimates for Diarrhea in DPP4I (male) (T) vs SU (male) (C) new-user cohorts under four different propensity score (PS) adjustment / time-at-risk (TAR) designs across data sources. We report total patient exposure time in 1,000 patient-year (KYr) units, uncalibrated and calibrated hazard ratios (HRs) and their 95% confidence intervals (CIs). Greyed out entries did not pass study diagnostics and are included only for completeness.

| | Patients | | Exposure (KYr) | | Outcomes | | Uncalibrated | | Calibrated | |
|---|---|---|---|---|---|---|---|---|---|---|
| | T | C | T | C | T | C | HR (95% CI) | $p$ | HR (95% CI) | $p$ |
| **PS-matching / on-treatment TAR** | | | | | | | | | | |
| CCAE | 43,507 | 78,291 | 37.5 | 65.8 | 767 | 1,393 | 0.98 (0.87 - 1.11) | 0.79 | 1.00 (0.88 - 1.13) | 0.86 |
| DAG | 13,237 | 2,325 | 14.1 | 2.4 | 28 | 8 | 0.92 (0.32 - 3.05) | 0.89 | 1.00 (0.32 - 3.07) | 0.89 |
| ODOD | 26,135 | 63,538 | 21.3 | 58.0 | 630 | 1,910 | 0.96 (0.85 - 1.09) | 0.51 | 1.00 (0.88 - 1.13) | 0.86 |
| MDCR | 8,110 | 16,353 | 8.3 | 16.7 | 251 | 494 | 0.93 (0.75 - 1.15) | 0.51 | 0.92 (0.74 - 1.14) | 0.44 |
| MDCD | 3,468 | 8,433 | 2.6 | 5.9 | 111 | 269 | 0.87 (0.63 - 1.19) | 0.40 | 0.91 (0.66 - 1.25) | 0.58 |
| Open Claims | 380,477 | 819,460 | 358.9 | 879.4 | 5,372 | 13,028 | 0.97 (0.93 - 1.01) | 0.17 | 0.97 (0.85 - 1.10) | 0.63 |
| OEHR | 33,533 | 88,165 | 16.2 | 47.1 | 447 | 1,556 | 0.90 (0.78 - 1.05) | 0.18 | 0.96 (0.80 - 1.17) | 0.76 |
| VA | 0 | 0 | 0.0 | 0.0 | 0 | 0 | | | | |
| SIDIAP | 25,832 | 7,060 | 75.5 | 20.0 | 573 | 91 | 1.11 (0.84 - 1.49) | 0.47 | 1.05 (0.79 - 1.41) | 0.73 |
| Meta-analysis | 534,299 | 1,083,625 | 534.3 | 1095.2 | 8,179 | 18,749 | 0.96 (0.93 - 1.00) | 0.06 | 0.97 (0.87 - 1.08) | 0.58 |
| Meta-analysis (w/o USOC) | 153,822 | 264,165 | 175.4 | 215.9 | 2,807 | 5,721 | 0.95 (0.89 - 1.02) | 0.18 | 0.98 (0.88 - 1.08) | 0.70 |
| **PS-stratification / on-treatment TAR** | | | | | | | | | | |
| CCAE | 48,527 | 79,796 | 41.9 | 67.0 | 874 | 1,409 | 0.98 (0.89 - 1.07) | 0.58 | 0.98 (0.89 - 1.08) | 0.73 |
| DAG | 13,265 | 2,696 | 14.1 | 2.8 | 28 | 8 | 0.65 (0.29 - 1.62) | 0.32 | 0.69 (0.29 - 1.65) | 0.41 |
| ODOD | 27,571 | 63,544 | 22.6 | 58.0 | 676 | 1,910 | 0.91 (0.83 - 1.00) | 0.05 | 0.92 (0.84 - 1.02) | 0.15 |
| MDCR | 8,555 | 16,728 | 8.8 | 17.2 | 264 | 507 | 0.98 (0.83 - 1.14) | 0.75 | 0.99 (0.84 - 1.16) | 0.84 |
| MDCD | 3,545 | 8,450 | 2.7 | 5.9 | 111 | 269 | 0.92 (0.73 - 1.16) | 0.49 | 0.95 (0.75 - 1.20) | 0.67 |
| Open Claims | 387,075 | 819,471 | 367.0 | 879.4 | 5,531 | 13,028 | 0.98 (0.95 - 1.01) | 0.18 | 0.97 (0.84 - 1.13) | 0.70 |
| OEHR | 35,251 | 88,189 | 17.2 | 47.1 | 470 | 1,557 | 0.90 (0.81 - 1.00) | 0.05 | 0.99 (0.88 - 1.10) | 0.81 |
| VA | 22,232 | 208,766 | 20.7 | 218.0 | 408 | 3,934 | 1.07 (0.97 - 1.19) | 0.17 | 1.11 (0.37 - 3.33) | 0.85 |
| SIDIAP | 25,996 | 9,089 | 75.6 | 26.9 | 575 | 112 | 1.10 (0.88 - 1.39) | 0.41 | 1.08 (0.86 - 1.36) | 0.54 |
| Meta-analysis | 549,785 | 1,087,963 | 549.8 | 1104.3 | 8,529 | 18,800 | 0.96 (0.93 - 1.00) | 0.02 | 0.97 (0.89 - 1.07) | 0.56 |
| Meta-analysis (w/o USOC) | 162,710 | 268,492 | 182.9 | 225.0 | 2,998 | 5,772 | 0.94 (0.90 - 0.99) | 0.02 | 0.96 (0.88 - 1.05) | 0.43 |
| **PS-matching / on-treatment TAR without escalation** | | | | | | | | | | |
| CCAE | 43,507 | 78,291 | 30.4 | 57.5 | 628 | 1,216 | 1.00 (0.87 - 1.14) | 0.96 | 1.02 (0.89 - 1.17) | 0.79 |
| DAG | 13,237 | 2,325 | 12.7 | 2.2 | 25 | 7 | 0.83 (0.28 - 2.78) | 0.75 | 0.90 (0.29 - 2.83) | 0.84 |
| ODOD | 26,135 | 63,538 | 17.6 | 52.0 | 525 | 1,712 | 0.97 (0.84 - 1.10) | 0.62 | 1.02 (0.89 - 1.17) | 0.81 |
| MDCR | 8,110 | 16,353 | 7.1 | 15.3 | 216 | 457 | 0.95 (0.75 - 1.19) | 0.63 | 0.95 (0.75 - 1.20) | 0.67 |
| MDCD | 3,468 | 8,433 | 2.0 | 5.0 | 81 | 230 | 0.91 (0.64 - 1.29) | 0.62 | 0.96 (0.67 - 1.36) | 0.79 |
| Open Claims | 380,477 | 819,460 | 279.7 | 748.2 | 4,159 | 10,955 | 0.95 (0.91 - 1.00) | 0.06 | 0.96 (0.85 - 1.08) | 0.48 |
| OEHR | 33,533 | 88,165 | 13.3 | 41.4 | 313 | 1,242 | 0.82 (0.69 - 0.96) | 0.02 | 0.89 (0.71 - 1.10) | 0.36 |
| VA | 0 | 0 | 0.0 | 0.0 | 0 | 0 | | | | |
| SIDIAP | 25,832 | 7,060 | 55.4 | 16.7 | 436 | 75 | 1.05 (0.76 - 1.48) | 0.76 | 0.98 (0.70 - 1.36) | 0.85 |
| Meta-analysis | 521,062 | 1,081,300 | 405.5 | 936.0 | 6,358 | 15,887 | 0.95 (0.91 - 0.99) | 0.02 | 0.96 (0.86 - 1.08) | 0.51 |
| Meta-analysis (w/o USOC) | 140,585 | 261,840 | 125.8 | 187.8 | 2,199 | 4,932 | 0.94 (0.88 - 1.02) | 0.12 | 0.97 (0.86 - 1.10) | 0.69 |
| **PS-stratification / on-treatment TAR without escalation** | | | | | | | | | | |
| CCAE | 48,527 | 79,796 | 34.1 | 58.6 | 711 | 1,229 | 0.96 (0.87 - 1.06) | 0.42 | 0.98 (0.88 - 1.08) | 0.68 |
| DAG | 13,265 | 2,696 | 12.7 | 2.6 | 25 | 7 | 0.68 (0.29 - 1.79) | 0.40 | 0.74 (0.30 - 1.84) | 0.52 |
| ODOD | 27,571 | 63,544 | 18.6 | 52.0 | 561 | 1,712 | 0.92 (0.83 - 1.01) | 0.08 | 0.93 (0.84 - 1.03) | 0.25 |
| MDCR | 8,555 | 16,728 | 7.6 | 15.8 | 227 | 470 | 0.97 (0.82 - 1.14) | 0.72 | 0.97 (0.82 - 1.15) | 0.77 |
| MDCD | 3,545 | 8,450 | 2.1 | 5.0 | 81 | 230 | 0.88 (0.67 - 1.14) | 0.34 | 0.89 (0.68 - 1.16) | 0.43 |
| Open Claims | 387,075 | 819,471 | 285.9 | 748.2 | 4,278 | 10,955 | 0.97 (0.94 - 1.01) | 0.12 | 0.96 (0.82 - 1.13) | 0.67 |
| OEHR | 35,251 | 88,189 | 14.1 | 41.4 | 332 | 1,243 | 0.83 (0.73 - 0.95) | <0.01 | 0.93 (0.81 - 1.06) | 0.37 |
| VA | 22,232 | 208,766 | 16.9 | 195.3 | 329 | 3,449 | 1.08 (0.96 - 1.21) | 0.19 | 1.09 (0.36 - 3.30) | 0.88 |
| SIDIAP | 25,996 | 9,089 | 55.5 | 22.3 | 438 | 91 | 1.05 (0.81 - 1.37) | 0.71 | 1.01 (0.78 - 1.31) | 0.88 |
| Meta-analysis | 549,785 | 1,087,963 | 430.5 | 945.7 | 6,653 | 15,937 | 0.94 (0.90 - 0.99) | 0.01 | 0.95 (0.85 - 1.07) | 0.43 |
| Meta-analysis (w/o USOC) | 162,710 | 268,492 | 144.6 | 197.6 | 2,375 | 4,982 | 0.92 (0.87 - 0.97) | <0.01 | 0.94 (0.85 - 1.05) | 0.31 |

##### 1.6.16 Genitourinary infection

**Supplementary Table 146:** Exposures, outcomes and risk estimates for Genitourinary infection in DPP4I (male) (T) vs SU (male) (C) new-user cohorts under four different propensity score (PS) adjustment / time-at-risk (TAR) designs across data sources. We report total patient exposure time in 1,000 patient-year (KYr) units, uncalibrated and calibrated hazard ratios (HRs) and their 95% confidence intervals (CIs). Greyed out entries did not pass study diagnostics and are included only for completeness.

| | Patients | | Exposure (KYr) | | Outcomes | | Uncalibrated | | Calibrated | |
|---|---|---|---|---|---|---|---|---|---|---|
| | T | C | T | C | T | C | HR (95% CI) | $p$ | HR (95% CI) | $p$ |
| **PS-matching / on-treatment TAR** | | | | | | | | | | |
| CCAE | 43,222 | 78,021 | 37.1 | 65.5 | 762 | 1,325 | 1.00 (0.88 - 1.14) | 1.00 | 1.01 (0.89 - 1.16) | 0.83 |
| DAG | 11,640 | 2,076 | 12.0 | 2.1 | 282 | 45 | 1.09 (0.74 - 1.65) | 0.66 | 1.18 (0.79 - 1.77) | 0.49 |
| ODOD | 25,325 | 61,682 | 20.7 | 55.7 | 756 | 2,375 | 0.91 (0.81 - 1.01) | 0.08 | 0.94 (0.84 - 1.06) | 0.37 |
| MDCR | 7,685 | 15,540 | 7.8 | 15.9 | 346 | 718 | 0.97 (0.81 - 1.16) | 0.74 | 0.95 (0.80 - 1.14) | 0.62 |
| MDCD | 3,601 | 8,687 | 2.7 | 6.1 | 82 | 192 | 0.99 (0.68 - 1.43) | 0.94 | 1.03 (0.71 - 1.49) | 0.85 |
| Open Claims | 371,458 | 802,493 | 350.2 | 858.0 | 6,759 | 16,701 | 0.99 (0.95 - 1.03) | 0.69 | 0.99 (0.88 - 1.12) | 0.87 |
| OEHR | 34,221 | 90,073 | 16.7 | 48.8 | 379 | 1,241 | 0.98 (0.84 - 1.15) | 0.85 | 1.05 (0.86 - 1.28) | 0.68 |
| VA | 0 | 0 | 0.0 | 0.0 | 0 | 0 | | | | |
| SIDIAP | 23,919 | 6,791 | 68.7 | 18.6 | 1,582 | 340 | 1.07 (0.92 - 1.26) | 0.38 | 1.02 (0.87 - 1.20) | 0.77 |
| Meta-analysis | 521,071 | 1,065,363 | 515.9 | 1070.5 | 10,948 | 22,937 | 0.99 (0.95 - 1.02) | 0.46 | 0.99 (0.89 - 1.11) | 0.86 |
| Meta-analysis (w/o USOC) | 149,613 | 262,870 | 165.6 | 212.6 | 4,189 | 6,236 | 0.98 (0.92 - 1.04) | 0.45 | 1.00 (0.91 - 1.10) | 0.88 |
| **PS-stratification / on-treatment TAR** | | | | | | | | | | |
| CCAE | 48,125 | 79,426 | 41.4 | 66.6 | 868 | 1,336 | 1.00 (0.91 - 1.09) | 0.95 | 1.00 (0.91 - 1.10) | 0.87 |
| DAG | 11,719 | 2,418 | 12.1 | 2.4 | 285 | 51 | 1.20 (0.87 - 1.68) | 0.29 | 1.28 (0.91 - 1.78) | 0.19 |
| ODOD | 26,717 | 61,687 | 21.9 | 55.7 | 803 | 2,375 | 0.89 (0.82 - 0.97) | <0.01 | 0.90 (0.82 - 0.99) | 0.05 |
| MDCR | 8,073 | 15,881 | 8.3 | 16.4 | 365 | 729 | 1.02 (0.90 - 1.17) | 0.74 | 1.03 (0.90 - 1.18) | 0.65 |
| MDCD | 3,670 | 8,703 | 2.7 | 6.1 | 83 | 192 | 0.94 (0.71 - 1.22) | 0.64 | 0.96 (0.73 - 1.26) | 0.79 |
| Open Claims | 377,661 | 802,507 | 357.8 | 858.0 | 6,968 | 16,701 | 0.98 (0.95 - 1.00) | 0.10 | 0.97 (0.83 - 1.12) | 0.67 |
| OEHR | 35,941 | 90,101 | 17.7 | 48.8 | 402 | 1,242 | 0.97 (0.86 - 1.09) | 0.65 | 1.07 (0.95 - 1.21) | 0.33 |
| VA | 22,854 | 211,170 | 21.3 | 220.7 | 298 | 3,737 | 0.83 (0.74 - 0.93) | <0.01 | 0.86 (0.29 - 2.57) | 0.79 |
| SIDIAP | 24,002 | 8,800 | 68.7 | 25.3 | 1,585 | 443 | 1.03 (0.92 - 1.17) | 0.59 | 1.01 (0.90 - 1.15) | 0.77 |
| Meta-analysis | 535,908 | 1,069,523 | 530.7 | 1079.2 | 11,359 | 23,069 | 0.97 (0.95 - 1.00) | 0.04 | 0.98 (0.90 - 1.08) | 0.71 |
| Meta-analysis (w/o USOC) | 158,247 | 267,016 | 172.9 | 221.2 | 4,391 | 6,368 | 0.98 (0.93 - 1.03) | 0.36 | 1.00 (0.91 - 1.09) | 0.88 |
| **PS-matching / on-treatment TAR without escalation** | | | | | | | | | | |
| CCAE | 43,222 | 78,021 | 30.1 | 57.1 | 621 | 1,159 | 0.95 (0.83 - 1.10) | 0.51 | 0.98 (0.85 - 1.12) | 0.75 |
| DAG | 11,640 | 2,076 | 10.9 | 1.9 | 248 | 43 | 1.01 (0.68 - 1.55) | 0.95 | 1.10 (0.72 - 1.67) | 0.65 |
| ODOD | 25,325 | 61,682 | 17.0 | 49.7 | 634 | 2,157 | 0.89 (0.79 - 1.01) | 0.07 | 0.94 (0.83 - 1.07) | 0.40 |
| MDCR | 7,685 | 15,540 | 6.7 | 14.6 | 293 | 650 | 0.97 (0.80 - 1.17) | 0.73 | 0.97 (0.80 - 1.18) | 0.76 |
| MDCD | 3,601 | 8,687 | 2.0 | 5.1 | 64 | 162 | 0.91 (0.59 - 1.36) | 0.64 | 0.95 (0.62 - 1.44) | 0.80 |
| Open Claims | 371,458 | 802,493 | 271.8 | 727.9 | 5,351 | 14,313 | 0.97 (0.93 - 1.02) | 0.22 | 0.98 (0.87 - 1.09) | 0.69 |
| OEHR | 34,221 | 90,073 | 13.6 | 42.6 | 292 | 980 | 1.01 (0.85 - 1.20) | 0.88 | 1.10 (0.88 - 1.37) | 0.48 |
| VA | 0 | 0 | 0.0 | 0.0 | 0 | 0 | | | | |
| SIDIAP | 23,919 | 6,791 | 50.5 | 15.6 | 1,151 | 278 | 1.04 (0.86 - 1.26) | 0.69 | 0.96 (0.80 - 1.17) | 0.72 |
| Meta-analysis | 521,071 | 1,065,363 | 402.6 | 914.4 | 8,654 | 19,742 | 0.97 (0.93 - 1.00) | 0.08 | 0.98 (0.87 - 1.10) | 0.71 |
| Meta-analysis (w/o USOC) | 149,613 | 262,870 | 130.9 | 186.6 | 3,303 | 5,429 | 0.96 (0.89 - 1.02) | 0.19 | 0.99 (0.88 - 1.11) | 0.82 |
| **PS-stratification / on-treatment TAR without escalation** | | | | | | | | | | |
| CCAE | 48,125 | 79,426 | 33.7 | 58.2 | 701 | 1,169 | 0.98 (0.89 - 1.08) | 0.72 | 1.00 (0.90 - 1.11) | 0.87 |
| DAG | 11,719 | 2,418 | 11.0 | 2.2 | 251 | 49 | 1.08 (0.77 - 1.53) | 0.66 | 1.18 (0.83 - 1.66) | 0.40 |
| ODOD | 26,717 | 61,687 | 18.0 | 49.7 | 670 | 2,157 | 0.88 (0.81 - 0.97) | <0.01 | 0.90 (0.81 - 0.99) | 0.06 |
| MDCR | 8,073 | 15,881 | 7.1 | 15.0 | 309 | 660 | 0.99 (0.86 - 1.14) | 0.88 | 0.99 (0.86 - 1.15) | 0.84 |
| MDCD | 3,670 | 8,703 | 2.1 | 5.1 | 65 | 162 | 0.98 (0.72 - 1.31) | 0.88 | 0.99 (0.73 - 1.34) | 0.87 |
| Open Claims | 377,661 | 802,507 | 277.4 | 727.9 | 5,531 | 14,313 | 0.98 (0.95 - 1.01) | 0.23 | 0.97 (0.83 - 1.14) | 0.75 |
| OEHR | 35,941 | 90,101 | 14.4 | 42.6 | 311 | 981 | 1.01 (0.88 - 1.15) | 0.89 | 1.12 (0.98 - 1.29) | 0.16 |
| VA | 22,854 | 211,170 | 17.3 | 197.1 | 243 | 3,218 | 0.86 (0.76 - 0.98) | 0.03 | 0.87 (0.29 - 2.65) | 0.81 |
| SIDIAP | 24,002 | 8,800 | 50.6 | 21.0 | 1,154 | 355 | 1.04 (0.90 - 1.19) | 0.61 | 1.00 (0.86 - 1.15) | 0.80 |
| Meta-analysis | 535,908 | 1,069,523 | 414.2 | 921.7 | 8,992 | 19,846 | 0.97 (0.95 - 1.00) | 0.10 | 0.98 (0.88 - 1.10) | 0.77 |
| Meta-analysis (w/o USOC) | 158,247 | 267,016 | 136.8 | 193.8 | 3,461 | 5,533 | 0.96 (0.92 - 1.01) | 0.16 | 0.99 (0.89 - 1.09) | 0.82 |

##### 1.6.17 Hyperkalemia

**Supplementary Table 147:** Exposures, outcomes and risk estimates for Hyperkalemia in DPP4I (male) (T) vs SU (male) (C) new-user cohorts under four different propensity score (PS) adjustment / time-at-risk (TAR) designs across data sources. We report total patient exposure time in 1,000 patient-year (KYr) units, uncalibrated and calibrated hazard ratios (HRs) and their 95% confidence intervals (CIs). Greyed out entries did not pass study diagnostics and are included only for completeness.

| | Patients | | Exposure (KYr) | | Outcomes | | Uncalibrated | | Calibrated | |
|---|---|---|---|---|---|---|---|---|---|---|
| | T | C | T | C | T | C | HR (95% CI) | $p$ | HR (95% CI) | $p$ |
| **PS-matching / on-treatment TAR** | | | | | | | | | | |
| CCAE | 46,699 | 83,866 | 40.6 | 71.2 | 296 | 570 | 0.94 (0.76 - 1.16) | 0.56 | 0.95 (0.77 - 1.18) | 0.67 |
| DAG | 12,541 | 2,235 | 13.1 | 2.3 | 207 | 34 | 0.98 (0.63 - 1.60) | 0.94 | 1.06 (0.66 - 1.70) | 0.73 |
| ODOD | 27,972 | 67,536 | 23.1 | 62.0 | 485 | 1,534 | 1.00 (0.87 - 1.15) | 0.97 | 1.04 (0.90 - 1.20) | 0.65 |
| MDCR | 8,781 | 17,691 | 9.1 | 18.4 | 150 | 353 | 0.76 (0.58 - 1.00) | 0.06 | 0.75 (0.57 - 0.99) | 0.05 |
| MDCD | 3,914 | 9,426 | 2.9 | 6.7 | 51 | 158 | 0.79 (0.49 - 1.23) | 0.31 | 0.82 (0.52 - 1.30) | 0.41 |
| Open Claims | 412,547 | 882,505 | 391.3 | 950.3 | 3,221 | 8,249 | 1.01 (0.96 - 1.07) | 0.66 | 1.01 (0.89 - 1.15) | 0.85 |
| OEHR | 35,645 | 93,824 | 17.4 | 51.2 | 279 | 839 | 1.15 (0.95 - 1.38) | 0.16 | 1.22 (0.98 - 1.52) | 0.13 |
| VA | 0 | 0 | 0.0 | 0.0 | 0 | 0 | | | | |
| SIDIAP | 25,263 | 6,967 | 72.5 | 19.5 | 934 | 207 | 1.04 (0.85 - 1.28) | 0.71 | 0.99 (0.80 - 1.21) | 0.81 |
| Meta-analysis | 573,362 | 1,164,050 | 570.1 | 1181.6 | 5,623 | 11,944 | 1.01 (0.96 - 1.05) | 0.82 | 1.01 (0.90 - 1.13) | 0.86 |
| Meta-analysis (w/o USOC) | 160,815 | 281,545 | 178.8 | 231.3 | 2,402 | 3,695 | 0.98 (0.90 - 1.08) | 0.75 | 1.01 (0.89 - 1.14) | 0.88 |
| **PS-stratification / on-treatment TAR** | | | | | | | | | | |
| CCAE | 52,064 | 85,370 | 45.4 | 72.4 | 323 | 573 | 0.89 (0.77 - 1.03) | 0.12 | 0.90 (0.78 - 1.04) | 0.17 |
| DAG | 12,561 | 2,595 | 13.1 | 2.7 | 208 | 38 | 1.12 (0.77 - 1.68) | 0.57 | 1.20 (0.81 - 1.77) | 0.41 |
| ODOD | 29,538 | 67,542 | 24.6 | 62.0 | 515 | 1,534 | 0.93 (0.83 - 1.03) | 0.15 | 0.94 (0.84 - 1.04) | 0.29 |
| MDCR | 9,303 | 18,074 | 9.7 | 18.9 | 160 | 357 | 0.88 (0.72 - 1.06) | 0.19 | 0.89 (0.73 - 1.08) | 0.25 |
| MDCD | 4,006 | 9,470 | 3.0 | 6.7 | 52 | 158 | 0.83 (0.59 - 1.14) | 0.26 | 0.85 (0.61 - 1.18) | 0.35 |
| Open Claims | 419,848 | 882,516 | 400.5 | 950.3 | 3,291 | 8,250 | 1.00 (0.96 - 1.04) | 0.91 | 0.99 (0.85 - 1.15) | 0.89 |
| OEHR | 37,432 | 93,852 | 18.5 | 51.2 | 294 | 841 | 1.11 (0.97 - 1.28) | 0.14 | 1.22 (1.06 - 1.41) | <0.01 |
| VA | 24,038 | 219,997 | 22.1 | 228.7 | 315 | 3,958 | 0.82 (0.73 - 0.92) | <0.01 | 0.85 (0.28 - 2.55) | 0.78 |
| SIDIAP | 25,338 | 8,957 | 72.6 | 26.3 | 935 | 288 | 1.09 (0.94 - 1.28) | 0.24 | 1.07 (0.92 - 1.25) | 0.41 |
| Meta-analysis | 590,090 | 1,168,376 | 587.3 | 1190.5 | 5,778 | 12,039 | 0.98 (0.92 - 1.04) | 0.55 | 0.99 (0.89 - 1.10) | 0.85 |
| Meta-analysis (w/o USOC) | 170,242 | 285,860 | 186.8 | 240.2 | 2,487 | 3,789 | 0.97 (0.89 - 1.06) | 0.55 | 0.99 (0.89 - 1.11) | 0.90 |
| **PS-matching / on-treatment TAR without escalation** | | | | | | | | | | |
| CCAE | 46,699 | 83,866 | 32.8 | 61.9 | 236 | 496 | 0.93 (0.74 - 1.17) | 0.54 | 0.95 (0.76 - 1.20) | 0.69 |
| DAG | 12,541 | 2,235 | 11.8 | 2.1 | 182 | 32 | 0.92 (0.58 - 1.50) | 0.74 | 1.00 (0.62 - 1.61) | 0.76 |
| ODOD | 27,972 | 67,536 | 19.0 | 55.4 | 395 | 1,356 | 0.98 (0.84 - 1.14) | 0.83 | 1.04 (0.89 - 1.21) | 0.69 |
| MDCR | 8,781 | 17,691 | 7.8 | 16.8 | 126 | 321 | 0.81 (0.60 - 1.09) | 0.16 | 0.81 (0.60 - 1.10) | 0.18 |
| MDCD | 3,914 | 9,426 | 2.2 | 5.6 | 37 | 129 | 0.78 (0.46 - 1.27) | 0.33 | 0.81 (0.49 - 1.35) | 0.44 |
| Open Claims | 412,547 | 882,505 | 303.8 | 805.8 | 2,531 | 6,892 | 1.03 (0.97 - 1.10) | 0.35 | 1.03 (0.91 - 1.17) | 0.61 |
| OEHR | 35,645 | 93,824 | 14.2 | 44.7 | 212 | 625 | 1.22 (0.98 - 1.50) | 0.07 | 1.32 (1.02 - 1.70) | 0.06 |
| VA | 0 | 0 | 0.0 | 0.0 | 0 | 0 | | | | |
| SIDIAP | 25,263 | 6,967 | 53.3 | 16.3 | 674 | 167 | 1.13 (0.89 - 1.44) | 0.33 | 1.05 (0.82 - 1.34) | 0.73 |
| Meta-analysis | 573,362 | 1,164,050 | 445.0 | 1008.6 | 4,393 | 10,018 | 1.02 (0.97 - 1.08) | 0.41 | 1.03 (0.92 - 1.17) | 0.59 |
| Meta-analysis (w/o USOC) | 160,815 | 281,545 | 141.2 | 202.8 | 1,862 | 3,126 | 1.00 (0.90 - 1.11) | 0.99 | 1.03 (0.89 - 1.19) | 0.68 |
| **PS-stratification / on-treatment TAR without escalation** | | | | | | | | | | |
| CCAE | 52,064 | 85,370 | 36.7 | 63.1 | 258 | 499 | 0.89 (0.76 - 1.04) | 0.15 | 0.91 (0.77 - 1.06) | 0.25 |
| DAG | 12,561 | 2,595 | 11.9 | 2.4 | 183 | 36 | 1.03 (0.69 - 1.56) | 0.90 | 1.12 (0.74 - 1.68) | 0.62 |
| ODOD | 29,538 | 67,542 | 20.1 | 55.4 | 420 | 1,356 | 0.93 (0.83 - 1.05) | 0.24 | 0.95 (0.84 - 1.07) | 0.45 |
| MDCR | 9,303 | 18,074 | 8.3 | 17.2 | 134 | 325 | 0.88 (0.71 - 1.09) | 0.25 | 0.89 (0.72 - 1.10) | 0.29 |
| MDCD | 4,006 | 9,470 | 2.3 | 5.7 | 37 | 129 | 0.80 (0.54 - 1.15) | 0.24 | 0.81 (0.55 - 1.18) | 0.29 |
| Open Claims | 419,848 | 882,516 | 310.7 | 805.8 | 2,584 | 6,893 | 1.02 (0.97 - 1.07) | 0.48 | 1.01 (0.86 - 1.19) | 0.89 |
| OEHR | 37,432 | 93,852 | 15.1 | 44.7 | 223 | 626 | 1.21 (1.03 - 1.43) | 0.02 | 1.35 (1.14 - 1.60) | <0.01 |
| VA | 24,038 | 219,997 | 18.0 | 204.4 | 243 | 3,343 | 0.82 (0.72 - 0.94) | <0.01 | 0.83 (0.28 - 2.53) | 0.75 |
| SIDIAP | 25,338 | 8,957 | 53.4 | 21.8 | 675 | 233 | 1.09 (0.92 - 1.30) | 0.33 | 1.05 (0.88 - 1.26) | 0.64 |
| Meta-analysis | 590,090 | 1,168,376 | 458.4 | 1016.1 | 4,514 | 10,097 | 0.99 (0.92 - 1.08) | 0.85 | 1.00 (0.87 - 1.15) | 0.92 |
| Meta-analysis (w/o USOC) | 170,242 | 285,860 | 147.7 | 210.3 | 1,930 | 3,204 | 0.98 (0.88 - 1.09) | 0.75 | 1.01 (0.88 - 1.15) | 0.90 |

##### 1.6.18 Hypoglycemia

**Supplementary Table 148:** Exposures, outcomes and risk estimates for Hypoglycemia in DPP4I (male) (T) vs SU (male) (C) new-user cohorts under four different propensity score (PS) adjustment / time-at-risk (TAR) designs across data sources. We report total patient exposure time in 1,000 patient-year (KYr) units, uncalibrated and calibrated hazard ratios (HRs) and their 95% confidence intervals (CIs). Greyed out entries did not pass study diagnostics and are included only for completeness.

| | Patients | | Exposure (KYr) | | Outcomes | | Uncalibrated | | Calibrated | |
|---|---|---|---|---|---|---|---|---|---|---|
| | T | C | T | C | T | C | HR (95% CI) | $p$ | HR (95% CI) | $p$ |
| **PS-matching / on-treatment TAR** | | | | | | | | | | |
| CCAE | 46,846 | 84,213 | 40.9 | 71.6 | 112 | 544 | 0.28 (0.21 - 0.37) | <0.01 | 0.28 (0.21 - 0.38) | <0.01 |
| DAG | 13,335 | 2,343 | 14.2 | 2.4 | 23 | 12 | 0.35 (0.14 - 0.89) | 0.03 | 0.38 (0.15 - 0.95) | 0.05 |
| ODOD | 28,883 | 69,988 | 24.2 | 65.3 | 121 | 1,203 | 0.24 (0.19 - 0.31) | <0.01 | 0.25 (0.20 - 0.32) | <0.01 |
| MDCR | 8,973 | 18,118 | 9.4 | 19.0 | 33 | 322 | 0.15 (0.08 - 0.25) | <0.01 | 0.15 (0.09 - 0.25) | <0.01 |
| MDCD | 4,004 | 9,629 | 3.0 | 6.9 | 25 | 176 | 0.30 (0.16 - 0.52) | <0.01 | 0.31 (0.17 - 0.56) | <0.01 |
| Open Claims | 416,792 | 892,347 | 397.9 | 966.4 | 949 | 7,417 | 0.27 (0.25 - 0.30) | <0.01 | 0.27 (0.24 - 0.32) | <0.01 |
| OEHR | 36,212 | 95,196 | 17.9 | 52.5 | 100 | 761 | 0.32 (0.24 - 0.43) | <0.01 | 0.34 (0.25 - 0.47) | <0.01 |
| VA | 0 | 0 | 0.0 | 0.0 | 0 | 0 | | | | |
| SIDIAP | 26,767 | 7,208 | 78.3 | 20.4 | 51 | 64 | 0.15 (0.08 - 0.27) | <0.01 | 0.15 (0.08 - 0.26) | <0.01 |
| Meta-analysis | 581,812 | 1,179,042 | 585.9 | 1204.5 | 1,414 | 10,499 | 0.27 (0.25 - 0.29) | <0.01 | 0.27 (0.24 - 0.31) | <0.01 |
| Meta-analysis (w/o USOC) | 165,020 | 286,695 | 187.9 | 238.1 | 465 | 3,082 | 0.25 (0.20 - 0.31) | <0.01 | 0.26 (0.21 - 0.32) | <0.01 |
| **PS-stratification / on-treatment TAR** | | | | | | | | | | |
| CCAE | 52,171 | 85,721 | 45.8 | 72.9 | 125 | 549 | 0.34 (0.28 - 0.42) | <0.01 | 0.35 (0.28 - 0.42) | <0.01 |
| DAG | 13,363 | 2,719 | 14.2 | 2.8 | 23 | 18 | 0.33 (0.16 - 0.71) | <0.01 | 0.36 (0.17 - 0.75) | <0.01 |
| ODOD | 30,452 | 69,994 | 25.6 | 65.3 | 127 | 1,203 | 0.28 (0.23 - 0.33) | <0.01 | 0.28 (0.23 - 0.34) | <0.01 |
| MDCR | 9,475 | 18,512 | 9.9 | 19.4 | 34 | 326 | 0.21 (0.15 - 0.30) | <0.01 | 0.22 (0.15 - 0.31) | <0.01 |
| MDCD | 4,097 | 9,647 | 3.1 | 6.9 | 25 | 176 | 0.32 (0.21 - 0.49) | <0.01 | 0.33 (0.22 - 0.51) | <0.01 |
| Open Claims | 424,020 | 892,361 | 407.1 | 966.4 | 972 | 7,418 | 0.31 (0.29 - 0.33) | <0.01 | 0.31 (0.26 - 0.36) | <0.01 |
| OEHR | 38,025 | 95,226 | 19.0 | 52.5 | 104 | 763 | 0.39 (0.32 - 0.48) | <0.01 | 0.43 (0.35 - 0.54) | <0.01 |
| VA | 25,385 | 231,973 | 23.8 | 245.8 | 78 | 2,126 | 0.37 (0.29 - 0.46) | <0.01 | 0.39 (0.13 - 1.18) | 0.10 |
| SIDIAP | 26,860 | 9,258 | 78.4 | 27.5 | 51 | 77 | 0.23 (0.15 - 0.34) | <0.01 | 0.22 (0.15 - 0.34) | <0.01 |
| Meta-analysis | 598,463 | 1,183,438 | 603.3 | 1213.8 | 1,461 | 10,530 | 0.31 (0.27 - 0.35) | <0.01 | 0.31 (0.26 - 0.36) | <0.01 |
| Meta-analysis (w/o USOC) | 174,443 | 291,077 | 196.1 | 247.4 | 489 | 3,112 | 0.30 (0.25 - 0.36) | <0.01 | 0.31 (0.26 - 0.37) | <0.01 |
| **PS-matching / on-treatment TAR without escalation** | | | | | | | | | | |
| CCAE | 46,846 | 84,213 | 33.1 | 62.3 | 75 | 454 | 0.26 (0.18 - 0.35) | <0.01 | 0.26 (0.19 - 0.37) | <0.01 |
| DAG | 13,335 | 2,343 | 12.8 | 2.2 | 16 | 11 | 0.30 (0.12 - 0.77) | 0.01 | 0.32 (0.13 - 0.83) | 0.02 |
| ODOD | 28,883 | 69,988 | 19.9 | 58.3 | 63 | 1,046 | 0.18 (0.13 - 0.24) | <0.01 | 0.19 (0.14 - 0.25) | <0.01 |
| MDCR | 8,973 | 18,118 | 8.0 | 17.3 | 19 | 294 | 0.11 (0.05 - 0.20) | <0.01 | 0.11 (0.06 - 0.21) | <0.01 |
| MDCD | 4,004 | 9,629 | 2.3 | 5.8 | 10 | 142 | 0.16 (0.06 - 0.34) | <0.01 | 0.17 (0.07 - 0.39) | <0.01 |
| Open Claims | 416,792 | 892,347 | 308.8 | 819.3 | 524 | 6,105 | 0.20 (0.18 - 0.23) | <0.01 | 0.20 (0.18 - 0.24) | <0.01 |
| OEHR | 36,212 | 95,196 | 14.5 | 45.7 | 49 | 567 | 0.19 (0.12 - 0.28) | <0.01 | 0.20 (0.13 - 0.31) | <0.01 |
| VA | 0 | 0 | 0.0 | 0.0 | 0 | 0 | | | | |
| SIDIAP | 26,767 | 7,208 | 57.4 | 17.0 | 16 | 50 | 0.08 (0.03 - 0.16) | <0.01 | 0.07 (0.03 - 0.16) | <0.01 |
| Meta-analysis | 568,477 | 1,176,699 | 444.0 | 1025.7 | 756 | 8,658 | 0.18 (0.15 - 0.22) | <0.01 | 0.18 (0.15 - 0.23) | <0.01 |
| Meta-analysis (w/o USOC) | 151,685 | 284,352 | 135.2 | 206.4 | 232 | 2,553 | 0.17 (0.12 - 0.23) | <0.01 | 0.17 (0.13 - 0.24) | <0.01 |
| **PS-stratification / on-treatment TAR without escalation** | | | | | | | | | | |
| CCAE | 52,171 | 85,721 | 37.1 | 63.5 | 83 | 459 | 0.29 (0.22 - 0.36) | <0.01 | 0.29 (0.23 - 0.37) | <0.01 |
| DAG | 13,363 | 2,719 | 12.8 | 2.6 | 16 | 17 | 0.27 (0.12 - 0.61) | <0.01 | 0.29 (0.13 - 0.67) | <0.01 |
| ODOD | 30,452 | 69,994 | 21.0 | 58.3 | 67 | 1,046 | 0.18 (0.14 - 0.23) | <0.01 | 0.18 (0.14 - 0.23) | <0.01 |
| MDCR | 9,475 | 18,512 | 8.5 | 17.8 | 20 | 297 | 0.14 (0.09 - 0.22) | <0.01 | 0.14 (0.09 - 0.23) | <0.01 |
| MDCD | 4,097 | 9,647 | 2.4 | 5.8 | 10 | 142 | 0.19 (0.09 - 0.34) | <0.01 | 0.19 (0.10 - 0.37) | <0.01 |
| Open Claims | 424,020 | 892,361 | 315.7 | 819.3 | 537 | 6,106 | 0.22 (0.20 - 0.24) | <0.01 | 0.22 (0.18 - 0.26) | <0.01 |
| OEHR | 38,025 | 95,226 | 15.4 | 45.7 | 51 | 569 | 0.27 (0.20 - 0.36) | <0.01 | 0.30 (0.22 - 0.40) | <0.01 |
| VA | 25,385 | 231,973 | 19.3 | 219.1 | 44 | 1,738 | 0.27 (0.20 - 0.36) | <0.01 | 0.27 (0.09 - 0.86) | 0.03 |
| SIDIAP | 26,860 | 9,258 | 57.5 | 22.8 | 16 | 60 | 0.08 (0.04 - 0.16) | <0.01 | 0.08 (0.04 - 0.15) | <0.01 |
| Meta-analysis | 598,463 | 1,183,438 | 470.5 | 1035.6 | 800 | 8,696 | 0.20 (0.16 - 0.26) | <0.01 | 0.20 (0.16 - 0.26) | <0.01 |
| Meta-analysis (w/o USOC) | 174,443 | 291,077 | 154.7 | 216.4 | 263 | 2,590 | 0.20 (0.15 - 0.26) | <0.01 | 0.20 (0.15 - 0.27) | <0.01 |

##### 1.6.19 Hypotension

**Supplementary Table 149:** Exposures, outcomes and risk estimates for Hypotension in DPP4I (male) (T) vs SU (male) (C) new-user cohorts under four different propensity score (PS) adjustment / time-at-risk (TAR) designs across data sources. We report total patient exposure time in 1,000 patient-year (KYr) units, uncalibrated and calibrated hazard ratios (HRs) and their 95% confidence intervals (CIs). Greyed out entries did not pass study diagnostics and are included only for completeness.

| | Patients | | Exposure (KYr) | | Outcomes | | Uncalibrated | | Calibrated | |
|---|---|---|---|---|---|---|---|---|---|---|
| | T | C | T | C | T | C | HR (95% CI) | $p$ | HR (95% CI) | $p$ |
| **PS-matching / on-treatment TAR** | | | | | | | | | | |
| CCAE | 46,654 | 83,703 | 40.6 | 71.3 | 317 | 525 | 1.21 (0.99 - 1.48) | 0.06 | 1.23 (1.01 - 1.51) | 0.05 |
| DAG | 13,110 | 2,308 | 13.9 | 2.4 | 69 | 14 | 0.81 (0.40 - 1.77) | 0.58 | 0.87 (0.41 - 1.85) | 0.73 |
| ODOD | 27,836 | 67,242 | 23.1 | 62.2 | 491 | 1,548 | 0.94 (0.81 - 1.08) | 0.39 | 0.98 (0.84 - 1.13) | 0.77 |
| MDCR | 8,487 | 17,145 | 8.7 | 17.9 | 242 | 474 | 1.05 (0.84 - 1.31) | 0.69 | 1.03 (0.82 - 1.29) | 0.80 |
| MDCD | 3,796 | 9,127 | 2.9 | 6.4 | 94 | 183 | 1.12 (0.78 - 1.60) | 0.52 | 1.17 (0.82 - 1.67) | 0.41 |
| Open Claims | 407,233 | 871,628 | 387.0 | 942.2 | 3,663 | 8,953 | 1.02 (0.97 - 1.08) | 0.40 | 1.02 (0.90 - 1.16) | 0.74 |
| OEHR | 35,607 | 93,678 | 17.4 | 51.2 | 325 | 954 | 1.05 (0.88 - 1.25) | 0.57 | 1.12 (0.91 - 1.38) | 0.37 |
| VA | 0 | 0 | 0.0 | 0.0 | 0 | 0 | | | | |
| SIDIAP | 26,630 | 7,194 | 77.8 | 20.4 | 147 | 20 | 1.19 (0.68 - 2.18) | 0.55 | 1.13 (0.63 - 2.02) | 0.68 |
| Meta-analysis | 569,353 | 1,152,025 | 571.5 | 1174.1 | 5,348 | 12,671 | 1.03 (0.98 - 1.08) | 0.24 | 1.03 (0.92 - 1.16) | 0.59 |
| Meta-analysis (w/o USOC) | 162,120 | 280,397 | 184.5 | 231.9 | 1,685 | 3,718 | 1.04 (0.95 - 1.13) | 0.38 | 1.06 (0.95 - 1.19) | 0.37 |
| **PS-stratification / on-treatment TAR** | | | | | | | | | | |
| CCAE | 52,015 | 85,196 | 45.5 | 72.5 | 348 | 530 | 1.14 (0.98 - 1.31) | 0.08 | 1.14 (0.99 - 1.32) | 0.09 |
| DAG | 13,137 | 2,665 | 14.0 | 2.8 | 69 | 16 | 0.86 (0.48 - 1.65) | 0.64 | 0.92 (0.50 - 1.71) | 0.79 |
| ODOD | 29,403 | 67,247 | 24.6 | 62.2 | 520 | 1,548 | 0.94 (0.85 - 1.04) | 0.26 | 0.95 (0.85 - 1.06) | 0.42 |
| MDCR | 8,981 | 17,519 | 9.3 | 18.3 | 263 | 486 | 1.13 (0.96 - 1.32) | 0.13 | 1.14 (0.97 - 1.34) | 0.12 |
| MDCD | 3,870 | 9,169 | 2.9 | 6.5 | 95 | 186 | 1.22 (0.94 - 1.58) | 0.14 | 1.25 (0.96 - 1.63) | 0.12 |
| Open Claims | 414,563 | 871,639 | 396.3 | 942.2 | 3,770 | 8,954 | 1.03 (0.99 - 1.08) | 0.09 | 1.03 (0.88 - 1.19) | 0.73 |
| OEHR | 37,425 | 93,706 | 18.5 | 51.2 | 345 | 957 | 1.17 (1.03 - 1.34) | 0.02 | 1.29 (1.13 - 1.47) | <0.01 |
| VA | 24,071 | 221,458 | 22.3 | 232.2 | 285 | 3,230 | 0.92 (0.81 - 1.04) | 0.18 | 0.95 (0.32 - 2.86) | 0.91 |
| SIDIAP | 26,719 | 9,245 | 77.9 | 27.6 | 147 | 24 | 1.52 (0.96 - 2.50) | 0.08 | 1.49 (0.92 - 2.41) | 0.10 |
| Meta-analysis | 586,113 | 1,156,386 | 588.9 | 1183.3 | 5,557 | 12,701 | 1.08 (1.00 - 1.17) | 0.04 | 1.09 (0.97 - 1.22) | 0.14 |
| Meta-analysis (w/o USOC) | 171,550 | 284,747 | 192.6 | 241.1 | 1,787 | 3,747 | 1.10 (1.00 - 1.22) | 0.05 | 1.13 (0.99 - 1.28) | 0.07 |
| **PS-matching / on-treatment TAR without escalation** | | | | | | | | | | |
| CCAE | 46,654 | 83,703 | 32.8 | 62.0 | 256 | 432 | 1.24 (0.99 - 1.54) | 0.06 | 1.26 (1.01 - 1.58) | 0.05 |
| DAG | 13,110 | 2,308 | 12.6 | 2.2 | 61 | 13 | 0.71 (0.33 - 1.65) | 0.41 | 0.77 (0.35 - 1.73) | 0.55 |
| ODOD | 27,836 | 67,242 | 19.0 | 55.5 | 383 | 1,397 | 0.89 (0.76 - 1.03) | 0.13 | 0.93 (0.80 - 1.09) | 0.43 |
| MDCR | 8,487 | 17,145 | 7.4 | 16.3 | 208 | 415 | 1.10 (0.87 - 1.40) | 0.42 | 1.11 (0.87 - 1.41) | 0.42 |
| MDCD | 3,796 | 9,127 | 2.2 | 5.4 | 74 | 153 | 1.13 (0.76 - 1.66) | 0.55 | 1.18 (0.80 - 1.75) | 0.43 |
| Open Claims | 407,233 | 871,628 | 300.2 | 798.5 | 2,840 | 7,508 | 1.04 (0.98 - 1.11) | 0.16 | 1.05 (0.93 - 1.18) | 0.48 |
| OEHR | 35,607 | 93,678 | 14.2 | 44.7 | 222 | 666 | 1.06 (0.86 - 1.30) | 0.61 | 1.14 (0.89 - 1.47) | 0.36 |
| VA | 0 | 0 | 0.0 | 0.0 | 0 | 0 | | | | |
| SIDIAP | 26,630 | 7,194 | 57.0 | 17.1 | 116 | 18 | 1.26 (0.66 - 2.58) | 0.51 | 1.17 (0.59 - 2.31) | 0.67 |
| Meta-analysis | 569,353 | 1,152,025 | 445.4 | 1001.8 | 4,160 | 10,602 | 1.04 (0.96 - 1.13) | 0.33 | 1.05 (0.92 - 1.21) | 0.46 |
| Meta-analysis (w/o USOC) | 162,120 | 280,397 | 145.2 | 203.2 | 1,320 | 3,094 | 1.04 (0.93 - 1.17) | 0.47 | 1.08 (0.93 - 1.25) | 0.36 |
| **PS-stratification / on-treatment TAR without escalation** | | | | | | | | | | |
| CCAE | 52,015 | 85,196 | 36.8 | 63.2 | 278 | 437 | 1.21 (1.03 - 1.42) | 0.02 | 1.23 (1.04 - 1.44) | 0.02 |
| DAG | 13,137 | 2,665 | 12.6 | 2.5 | 61 | 15 | 0.81 (0.43 - 1.59) | 0.52 | 0.88 (0.46 - 1.69) | 0.71 |
| ODOD | 29,403 | 67,247 | 20.1 | 55.5 | 407 | 1,397 | 0.88 (0.78 - 0.98) | 0.03 | 0.89 (0.79 - 1.01) | 0.10 |
| MDCR | 8,981 | 17,519 | 7.9 | 16.8 | 226 | 426 | 1.18 (1.00 - 1.40) | 0.05 | 1.19 (1.00 - 1.41) | 0.07 |
| MDCD | 3,870 | 9,169 | 2.2 | 5.5 | 75 | 156 | 1.27 (0.95 - 1.69) | 0.11 | 1.29 (0.96 - 1.73) | 0.11 |
| Open Claims | 414,563 | 871,639 | 307.2 | 798.5 | 2,921 | 7,509 | 1.04 (0.99 - 1.08) | 0.12 | 1.03 (0.87 - 1.21) | 0.74 |
| OEHR | 37,425 | 93,706 | 15.1 | 44.7 | 240 | 667 | 1.22 (1.04 - 1.42) | 0.02 | 1.35 (1.15 - 1.59) | <0.01 |
| VA | 24,071 | 221,458 | 18.1 | 207.3 | 210 | 2,664 | 0.90 (0.78 - 1.03) | 0.13 | 0.91 (0.30 - 2.75) | 0.86 |
| SIDIAP | 26,719 | 9,245 | 57.1 | 22.8 | 116 | 21 | 1.41 (0.86 - 2.41) | 0.20 | 1.35 (0.80 - 2.27) | 0.26 |
| Meta-analysis | 586,113 | 1,156,386 | 459.0 | 1009.5 | 4,324 | 10,628 | 1.10 (0.99 - 1.23) | 0.09 | 1.11 (0.95 - 1.30) | 0.19 |
| Meta-analysis (w/o USOC) | 171,550 | 284,747 | 151.8 | 210.9 | 1,403 | 3,119 | 1.13 (0.97 - 1.31) | 0.12 | 1.15 (0.97 - 1.37) | 0.12 |

##### 1.6.20 Joint pain

**Supplementary Table 150:** Exposures, outcomes and risk estimates for Joint pain in DPP4I (male) (T) vs SU (male) (C) new-user cohorts under four different propensity score (PS) adjustment / time-at-risk (TAR) designs across data sources. We report total patient exposure time in 1,000 patient-year (KYr) units, uncalibrated and calibrated hazard ratios (HRs) and their 95% confidence intervals (CIs). Greyed out entries did not pass study diagnostics and are included only for completeness.

| | Patients | | Exposure (KYr) | | Outcomes | | Uncalibrated | | Calibrated | |
|---|---|---|---|---|---|---|---|---|---|---|
| | T | C | T | C | T | C | HR (95% CI) | $p$ | HR (95% CI) | $p$ |
| **PS-matching / on-treatment TAR** | | | | | | | | | | |
| CCAE | 45,710 | 82,050 | 39.8 | 69.5 | 329 | 581 | 1.10 (0.90 - 1.35) | 0.33 | 1.12 (0.92 - 1.37) | 0.29 |
| DAG | 11,582 | 2,086 | 12.0 | 2.1 | 264 | 33 | 1.68 (1.03 - 2.88) | 0.05 | 1.81 (1.08 - 3.05) | 0.04 |
| ODOD | 27,948 | 67,874 | 23.3 | 63.3 | 250 | 646 | 1.09 (0.88 - 1.35) | 0.41 | 1.14 (0.92 - 1.40) | 0.26 |
| MDCR | 8,776 | 17,714 | 9.1 | 18.6 | 82 | 162 | 1.01 (0.70 - 1.45) | 0.96 | 0.99 (0.69 - 1.44) | 0.91 |
| MDCD | 3,800 | 9,203 | 2.9 | 6.5 | 49 | 85 | 1.42 (0.87 - 2.32) | 0.16 | 1.48 (0.91 - 2.42) | 0.13 |
| Open Claims | 403,632 | 867,294 | 384.4 | 940.1 | 2,011 | 4,493 | 1.02 (0.95 - 1.10) | 0.60 | 1.02 (0.89 - 1.17) | 0.79 |
| OEHR | 35,054 | 92,497 | 17.2 | 50.9 | 139 | 431 | 1.07 (0.82 - 1.38) | 0.63 | 1.14 (0.85 - 1.52) | 0.43 |
| VA | 0 | 0 | 0.0 | 0.0 | 0 | 0 | | | | |
| SIDIAP | 22,584 | 6,504 | 65.1 | 17.6 | 1,213 | 378 | 0.90 (0.78 - 1.06) | 0.20 | 0.86 (0.73 - 1.00) | 0.09 |
| Meta-analysis | 559,086 | 1,145,222 | 553.8 | 1168.5 | 4,337 | 6,809 | 1.03 (0.97 - 1.09) | 0.35 | 1.03 (0.92 - 1.16) | 0.60 |
| Meta-analysis (w/o USOC) | 155,454 | 277,928 | 169.4 | 228.5 | 2,326 | 2,316 | 1.07 (0.95 - 1.20) | 0.29 | 1.09 (0.95 - 1.26) | 0.28 |
| **PS-stratification / on-treatment TAR** | | | | | | | | | | |
| CCAE | 50,908 | 83,525 | 44.5 | 70.7 | 376 | 582 | 0.98 (0.86 - 1.13) | 0.82 | 0.99 (0.86 - 1.14) | 0.88 |
| DAG | 11,607 | 2,420 | 12.1 | 2.5 | 264 | 40 | 1.43 (1.00 - 2.10) | 0.06 | 1.53 (1.05 - 2.22) | 0.04 |
| ODOD | 29,498 | 67,881 | 24.7 | 63.3 | 265 | 646 | 1.02 (0.87 - 1.18) | 0.83 | 1.03 (0.88 - 1.20) | 0.75 |
| MDCR | 9,268 | 18,102 | 9.7 | 19.1 | 90 | 162 | 1.03 (0.78 - 1.34) | 0.85 | 1.04 (0.79 - 1.36) | 0.79 |
| MDCD | 3,898 | 9,221 | 2.9 | 6.5 | 51 | 87 | 1.17 (0.81 - 1.67) | 0.40 | 1.20 (0.83 - 1.72) | 0.35 |
| Open Claims | 410,700 | 867,316 | 393.3 | 940.1 | 2,082 | 4,493 | 1.03 (0.98 - 1.09) | 0.22 | 1.03 (0.88 - 1.20) | 0.74 |
| OEHR | 36,839 | 92,525 | 18.3 | 50.9 | 152 | 431 | 1.02 (0.84 - 1.24) | 0.85 | 1.12 (0.92 - 1.37) | 0.28 |
| VA | 22,503 | 206,997 | 21.2 | 218.8 | 108 | 1,714 | 0.64 (0.52 - 0.77) | <0.01 | 0.66 (0.22 - 2.00) | 0.47 |
| SIDIAP | 22,781 | 8,424 | 65.3 | 23.9 | 1,214 | 511 | 0.92 (0.82 - 1.04) | 0.18 | 0.90 (0.80 - 1.02) | 0.15 |
| Meta-analysis | 575,499 | 1,149,414 | 570.7 | 1176.9 | 4,494 | 6,952 | 1.02 (0.98 - 1.06) | 0.39 | 1.03 (0.93 - 1.13) | 0.58 |
| Meta-analysis (w/o USOC) | 164,799 | 282,098 | 177.4 | 236.8 | 2,412 | 2,459 | 1.00 (0.93 - 1.07) | 0.92 | 1.02 (0.92 - 1.13) | 0.75 |
| **PS-matching / on-treatment TAR without escalation** | | | | | | | | | | |
| CCAE | 45,710 | 82,050 | 32.2 | 60.4 | 267 | 507 | 1.07 (0.86 - 1.32) | 0.54 | 1.09 (0.88 - 1.36) | 0.43 |
| DAG | 11,582 | 2,086 | 10.9 | 1.9 | 249 | 32 | 1.64 (1.00 - 2.81) | 0.06 | 1.77 (1.05 - 2.99) | 0.05 |
| ODOD | 27,948 | 67,874 | 19.1 | 56.5 | 213 | 576 | 1.15 (0.92 - 1.44) | 0.22 | 1.21 (0.97 - 1.52) | 0.11 |
| MDCR | 8,776 | 17,714 | 7.8 | 16.9 | 68 | 147 | 0.99 (0.67 - 1.46) | 0.98 | 1.00 (0.67 - 1.48) | 0.91 |
| MDCD | 3,800 | 9,203 | 2.2 | 5.5 | 35 | 70 | 1.42 (0.80 - 2.52) | 0.23 | 1.49 (0.84 - 2.65) | 0.19 |
| Open Claims | 403,632 | 867,294 | 298.3 | 796.8 | 1,525 | 3,799 | 0.99 (0.91 - 1.07) | 0.76 | 0.99 (0.86 - 1.13) | 0.87 |
| OEHR | 35,054 | 92,497 | 14.0 | 44.3 | 109 | 354 | 1.10 (0.82 - 1.47) | 0.50 | 1.20 (0.87 - 1.65) | 0.32 |
| VA | 0 | 0 | 0.0 | 0.0 | 0 | 0 | | | | |
| SIDIAP | 22,584 | 6,504 | 47.9 | 14.8 | 875 | 320 | 0.91 (0.76 - 1.09) | 0.31 | 0.85 (0.71 - 1.01) | 0.11 |
| Meta-analysis | 559,086 | 1,145,222 | 432.4 | 997.2 | 3,341 | 5,805 | 1.02 (0.95 - 1.09) | 0.59 | 1.03 (0.91 - 1.17) | 0.65 |
| Meta-analysis (w/o USOC) | 155,454 | 277,928 | 134.1 | 200.4 | 1,816 | 2,006 | 1.07 (0.95 - 1.20) | 0.26 | 1.10 (0.95 - 1.28) | 0.23 |
| **PS-stratification / on-treatment TAR without escalation** | | | | | | | | | | |
| CCAE | 50,908 | 83,525 | 36.0 | 61.6 | 310 | 508 | 0.99 (0.85 - 1.15) | 0.89 | 1.01 (0.86 - 1.17) | 0.90 |
| DAG | 11,607 | 2,420 | 10.9 | 2.2 | 249 | 39 | 1.42 (0.98 - 2.09) | 0.07 | 1.54 (1.05 - 2.26) | 0.04 |
| ODOD | 29,498 | 67,881 | 20.3 | 56.5 | 226 | 576 | 1.05 (0.89 - 1.23) | 0.58 | 1.06 (0.90 - 1.26) | 0.50 |
| MDCR | 9,268 | 18,102 | 8.3 | 17.4 | 76 | 147 | 1.00 (0.75 - 1.33) | 1.00 | 1.00 (0.75 - 1.34) | 0.92 |
| MDCD | 3,898 | 9,221 | 2.3 | 5.5 | 36 | 72 | 1.13 (0.74 - 1.71) | 0.55 | 1.15 (0.76 - 1.75) | 0.53 |
| Open Claims | 410,700 | 867,316 | 305.1 | 796.8 | 1,578 | 3,799 | 0.99 (0.94 - 1.06) | 0.87 | 0.99 (0.83 - 1.17) | 0.88 |
| OEHR | 36,839 | 92,525 | 14.9 | 44.3 | 120 | 354 | 1.03 (0.82 - 1.28) | 0.80 | 1.14 (0.92 - 1.43) | 0.26 |
| VA | 22,503 | 206,997 | 17.2 | 195.4 | 85 | 1,548 | 0.61 (0.49 - 0.76) | <0.01 | 0.62 (0.20 - 1.90) | 0.41 |
| SIDIAP | 22,781 | 8,424 | 48.1 | 19.9 | 875 | 424 | 0.89 (0.78 - 1.02) | 0.08 | 0.85 (0.74 - 0.98) | 0.05 |
| Meta-analysis | 575,499 | 1,149,414 | 445.7 | 1004.3 | 3,470 | 5,919 | 0.99 (0.95 - 1.04) | 0.79 | 1.00 (0.89 - 1.13) | 0.91 |
| Meta-analysis (w/o USOC) | 164,799 | 282,098 | 140.7 | 207.4 | 1,892 | 2,120 | 1.00 (0.92 - 1.08) | 0.97 | 1.02 (0.91 - 1.15) | 0.73 |

##### 1.6.21 Lower extremity amputation

**Supplementary Table 151:** Exposures, outcomes and risk estimates for Lower extremity amputation in DPP4I (male) (T) vs SU (male) (C) new-user cohorts under four different propensity score (PS) adjustment / time-at-risk (TAR) designs across data sources. We report total patient exposure time in 1,000 patient-year (KYr) units, uncalibrated and calibrated hazard ratios (HRs) and their 95% confidence intervals (CIs). Greyed out entries did not pass study diagnostics and are included only for completeness.

| | Patients | | Exposure (KYr) | | Outcomes | | Uncalibrated | | Calibrated | |
|---|---|---|---|---|---|---|---|---|---|---|
| | T | C | T | C | T | C | HR (95% CI) | $p$ | HR (95% CI) | $p$ |
| **PS-matching / on-treatment TAR** | | | | | | | | | | |
| CCAE | 47,380 | 85,055 | 41.5 | 72.6 | 33 | 75 | 1.17 (0.64 - 2.09) | 0.61 | 1.19 (0.66 - 2.15) | 0.58 |
| DAG | 13,419 | 2,361 | 14.3 | 2.4 | 0 | 0 | | | | |
| ODOD | 29,180 | 70,617 | 24.5 | 66.4 | 27 | 102 | 0.82 (0.45 - 1.45) | 0.51 | 0.85 (0.47 - 1.54) | 0.60 |
| MDCR | 9,031 | 18,228 | 9.4 | 19.2 | 10 | 28 | 0.56 (0.19 - 1.41) | 0.25 | 0.55 (0.20 - 1.47) | 0.23 |
| MDCD | 4,059 | 9,787 | 3.1 | 7.1 | 9 | 13 | 1.89 (0.54 - 7.05) | 0.33 | 1.97 (0.54 - 7.13) | 0.30 |
| Open Claims | 421,580 | 901,057 | 402.8 | 979.0 | 240 | 832 | 0.82 (0.67 - 0.99) | 0.04 | 0.82 (0.65 - 1.03) | 0.09 |
| OEHR | 36,410 | 95,792 | 18.1 | 53.3 | 15 | 83 | 0.69 (0.34 - 1.29) | 0.27 | 0.73 (0.37 - 1.44) | 0.38 |
| VA | 0 | 0 | 0.0 | 0.0 | 0 | 0 | | | | |
| SIDIAP | 26,751 | 7,203 | 78.3 | 20.4 | 32 | 11 | 1.48 (0.62 - 3.90) | 0.40 | 1.40 (0.56 - 3.52) | 0.47 |
| Meta-analysis | 574,391 | 1,187,739 | 577.7 | 1218.0 | 366 | 1,144 | 0.85 (0.72 - 1.00) | 0.05 | 0.85 (0.70 - 1.04) | 0.12 |
| Meta-analysis (w/o USOC) | 152,811 | 286,682 | 174.9 | 239.0 | 126 | 312 | 0.94 (0.69 - 1.27) | 0.67 | 0.96 (0.70 - 1.31) | 0.79 |
| **PS-stratification / on-treatment TAR** | | | | | | | | | | |
| CCAE | 52,823 | 86,574 | 46.5 | 73.9 | 33 | 76 | 0.83 (0.53 - 1.26) | 0.39 | 0.83 (0.54 - 1.29) | 0.41 |
| DAG | 13,447 | 2,736 | 14.3 | 2.8 | 0 | 0 | | | | |
| ODOD | 30,790 | 70,624 | 26.0 | 66.4 | 28 | 102 | 0.86 (0.55 - 1.31) | 0.50 | 0.87 (0.56 - 1.35) | 0.53 |
| MDCR | 9,542 | 18,619 | 10.0 | 19.7 | 10 | 28 | 0.76 (0.34 - 1.57) | 0.48 | 0.77 (0.36 - 1.65) | 0.50 |
| MDCD | 4,154 | 9,807 | 3.2 | 7.1 | 9 | 13 | 1.70 (0.66 - 4.27) | 0.27 | 1.75 (0.69 - 4.45) | 0.25 |
| Open Claims | 428,977 | 901,071 | 412.2 | 979.1 | 244 | 832 | 0.85 (0.73 - 0.98) | 0.03 | 0.84 (0.68 - 1.03) | 0.11 |
| OEHR | 38,251 | 95,822 | 19.2 | 53.3 | 16 | 83 | 0.67 (0.37 - 1.14) | 0.16 | 0.73 (0.42 - 1.29) | 0.28 |
| VA | 25,577 | 233,270 | 24.0 | 248.3 | 13 | 201 | 0.68 (0.37 - 1.15) | 0.19 | 0.71 (0.21 - 2.42) | 0.59 |
| SIDIAP | 26,844 | 9,248 | 78.4 | 27.6 | 32 | 20 | 1.06 (0.57 - 2.01) | 0.85 | 1.04 (0.56 - 1.95) | 0.90 |
| Meta-analysis | 591,381 | 1,191,765 | 595.4 | 1226.9 | 372 | 1,154 | 0.85 (0.75 - 0.96) | 0.01 | 0.86 (0.74 - 1.00) | 0.05 |
| Meta-analysis (w/o USOC) | 162,404 | 290,694 | 183.2 | 247.9 | 128 | 322 | 0.86 (0.69 - 1.09) | 0.21 | 0.88 (0.69 - 1.12) | 0.31 |
| **PS-matching / on-treatment TAR without escalation** | | | | | | | | | | |
| CCAE | 47,380 | 85,055 | 33.5 | 63.1 | 25 | 59 | 1.21 (0.66 - 2.20) | 0.53 | 1.24 (0.68 - 2.26) | 0.49 |
| DAG | 13,419 | 2,361 | 12.9 | 2.2 | 0 | 0 | | | | |
| ODOD | 29,180 | 70,617 | 20.1 | 59.2 | 20 | 86 | 0.71 (0.35 - 1.35) | 0.32 | 0.74 (0.38 - 1.47) | 0.40 |
| MDCR | 9,031 | 18,228 | 8.0 | 17.5 | 10 | 25 | 0.61 (0.21 - 1.57) | 0.33 | 0.61 (0.22 - 1.67) | 0.34 |
| MDCD | 4,059 | 9,787 | 2.4 | 5.9 | 7 | 10 | 1.04 (0.20 - 4.63) | 0.96 | 1.09 (0.23 - 5.21) | 0.92 |
| Open Claims | 421,580 | 901,057 | 312.3 | 829.1 | 181 | 670 | 0.82 (0.65 - 1.01) | 0.07 | 0.82 (0.64 - 1.04) | 0.11 |
| OEHR | 36,410 | 95,792 | 14.6 | 46.2 | 10 | 44 | 0.94 (0.38 - 2.04) | 0.87 | 1.01 (0.43 - 2.36) | 0.95 |
| VA | 0 | 0 | 0.0 | 0.0 | 0 | 0 | | | | |
| SIDIAP | 26,751 | 7,203 | 57.3 | 17.1 | 21 | 7 | 2.11 (0.73 - 7.57) | 0.21 | 1.95 (0.61 - 6.28) | 0.26 |
| Meta-analysis | 570,332 | 1,177,952 | 446.0 | 1032.2 | 267 | 891 | 0.86 (0.71 - 1.03) | 0.10 | 0.87 (0.70 - 1.07) | 0.19 |
| Meta-analysis (w/o USOC) | 148,752 | 276,895 | 133.6 | 203.1 | 86 | 221 | 0.97 (0.68 - 1.38) | 0.86 | 1.00 (0.69 - 1.44) | 0.95 |
| **PS-stratification / on-treatment TAR without escalation** | | | | | | | | | | |
| CCAE | 52,823 | 86,574 | 37.6 | 64.3 | 25 | 60 | 0.84 (0.50 - 1.36) | 0.48 | 0.85 (0.51 - 1.40) | 0.52 |
| DAG | 13,447 | 2,736 | 12.9 | 2.6 | 0 | 0 | | | | |
| ODOD | 30,790 | 70,624 | 21.3 | 59.2 | 21 | 86 | 0.83 (0.49 - 1.33) | 0.46 | 0.84 (0.51 - 1.39) | 0.50 |
| MDCR | 9,542 | 18,619 | 8.5 | 17.9 | 10 | 25 | 0.89 (0.39 - 1.86) | 0.76 | 0.89 (0.41 - 1.94) | 0.77 |
| MDCD | 4,154 | 9,807 | 2.4 | 5.9 | 7 | 10 | 1.73 (0.58 - 4.96) | 0.32 | 1.75 (0.60 - 5.12) | 0.31 |
| Open Claims | 428,977 | 901,071 | 319.4 | 829.1 | 185 | 670 | 0.86 (0.73 - 1.02) | 0.08 | 0.86 (0.68 - 1.08) | 0.19 |
| OEHR | 38,251 | 95,822 | 15.6 | 46.3 | 11 | 44 | 0.91 (0.43 - 1.77) | 0.78 | 1.01 (0.50 - 2.05) | 0.96 |
| VA | 25,577 | 233,270 | 19.5 | 221.1 | 11 | 150 | 0.84 (0.43 - 1.48) | 0.59 | 0.85 (0.24 - 3.02) | 0.81 |
| SIDIAP | 26,844 | 9,248 | 57.4 | 22.8 | 21 | 12 | 1.28 (0.57 - 2.96) | 0.56 | 1.23 (0.54 - 2.80) | 0.63 |
| Meta-analysis | 587,227 | 1,181,958 | 459.8 | 1039.6 | 273 | 897 | 0.87 (0.75 - 1.00) | 0.06 | 0.88 (0.73 - 1.05) | 0.16 |
| Meta-analysis (w/o USOC) | 158,250 | 280,887 | 140.4 | 210.5 | 88 | 227 | 0.89 (0.68 - 1.17) | 0.41 | 0.91 (0.68 - 1.22) | 0.54 |

##### 1.6.22 Nausea

**Supplementary Table 152:** Exposures, outcomes and risk estimates for Nausea in DPP4I (male) (T) vs SU (male) (C) new-user cohorts under four different propensity score (PS) adjustment / time-at-risk (TAR) designs across data sources. We report total patient exposure time in 1,000 patient-year (KYr) units, uncalibrated and calibrated hazard ratios (HRs) and their 95% confidence intervals (CIs). Greyed out entries did not pass study diagnostics and are included only for completeness.

| | Patients | | Exposure (KYr) | | Outcomes | | Uncalibrated | | Calibrated | |
|---|---|---|---|---|---|---|---|---|---|---|
| | T | C | T | C | T | C | HR (95% CI) | $p$ | HR (95% CI) | $p$ |
| **PS-matching / on-treatment TAR** | | | | | | | | | | |
| CCAE | 43,894 | 78,606 | 38.0 | 66.6 | 863 | 1,393 | 1.10 (0.98 - 1.24) | 0.11 | 1.12 (0.99 - 1.26) | 0.10 |
| DAG | 12,654 | 2,255 | 13.4 | 2.3 | 115 | 25 | 0.77 (0.45 - 1.36) | 0.34 | 0.83 (0.47 - 1.44) | 0.54 |
| ODOD | 26,343 | 63,660 | 21.7 | 58.8 | 677 | 1,934 | 0.97 (0.86 - 1.09) | 0.62 | 1.01 (0.89 - 1.14) | 0.85 |
| MDCR | 8,327 | 16,640 | 8.6 | 17.4 | 229 | 533 | 0.79 (0.64 - 0.99) | 0.04 | 0.78 (0.63 - 0.97) | 0.03 |
| MDCD | 3,262 | 7,837 | 2.4 | 5.5 | 134 | 341 | 0.87 (0.65 - 1.14) | 0.32 | 0.90 (0.68 - 1.20) | 0.51 |
| Open Claims | 383,482 | 820,475 | 364.1 | 889.5 | 5,625 | 13,804 | 1.01 (0.97 - 1.05) | 0.65 | 1.01 (0.89 - 1.14) | 0.87 |
| OEHR | 34,141 | 89,528 | 16.6 | 48.5 | 443 | 1,327 | 1.14 (0.99 - 1.32) | 0.07 | 1.22 (1.01 - 1.47) | 0.09 |
| VA | 0 | 0 | 0.0 | 0.0 | 0 | 0 | | | | |
| SIDIAP | 26,189 | 7,117 | 76.4 | 20.1 | 345 | 73 | 1.10 (0.79 - 1.56) | 0.58 | 1.05 (0.74 - 1.47) | 0.81 |
| Meta-analysis | 538,292 | 1,086,118 | 541.3 | 1108.7 | 8,431 | 19,430 | 1.01 (0.93 - 1.08) | 0.88 | 1.01 (0.89 - 1.15) | 0.87 |
| Meta-analysis (w/o USOC) | 154,810 | 265,643 | 177.1 | 219.2 | 2,806 | 5,626 | 1.00 (0.89 - 1.11) | 0.93 | 1.02 (0.89 - 1.16) | 0.81 |
| **PS-stratification / on-treatment TAR** | | | | | | | | | | |
| CCAE | 49,023 | 80,057 | 42.5 | 67.8 | 977 | 1,411 | 1.11 (1.02 - 1.21) | 0.02 | 1.11 (1.02 - 1.22) | 0.03 |
| DAG | 12,680 | 2,609 | 13.4 | 2.7 | 115 | 31 | 0.86 (0.56 - 1.37) | 0.51 | 0.92 (0.58 - 1.44) | 0.71 |
| ODOD | 27,811 | 63,666 | 23.0 | 58.8 | 724 | 1,934 | 0.98 (0.90 - 1.07) | 0.67 | 0.99 (0.90 - 1.09) | 0.83 |
| MDCR | 8,802 | 17,104 | 9.1 | 17.9 | 238 | 541 | 0.84 (0.72 - 0.98) | 0.03 | 0.85 (0.72 - 1.00) | 0.06 |
| MDCD | 3,340 | 7,870 | 2.5 | 5.6 | 138 | 343 | 0.94 (0.76 - 1.15) | 0.54 | 0.96 (0.78 - 1.19) | 0.74 |
| Open Claims | 390,539 | 820,488 | 373.0 | 889.5 | 5,784 | 13,805 | 1.00 (0.97 - 1.03) | 0.88 | 0.99 (0.85 - 1.15) | 0.88 |
| OEHR | 35,896 | 89,556 | 17.6 | 48.5 | 462 | 1,328 | 1.05 (0.94 - 1.17) | 0.41 | 1.15 (1.03 - 1.29) | 0.03 |
| VA | 23,930 | 221,000 | 22.4 | 233.9 | 192 | 2,238 | 0.88 (0.76 - 1.02) | 0.09 | 0.91 (0.30 - 2.74) | 0.87 |
| SIDIAP | 26,278 | 9,165 | 76.5 | 27.1 | 345 | 101 | 0.96 (0.75 - 1.25) | 0.77 | 0.94 (0.73 - 1.22) | 0.67 |
| Meta-analysis | 554,369 | 1,090,515 | 557.7 | 1118.0 | 8,783 | 19,494 | 1.00 (0.94 - 1.06) | 0.93 | 1.01 (0.91 - 1.12) | 0.89 |
| Meta-analysis (w/o USOC) | 163,830 | 270,027 | 184.7 | 228.5 | 2,999 | 5,689 | 0.99 (0.92 - 1.07) | 0.84 | 1.01 (0.91 - 1.13) | 0.82 |
| **PS-matching / on-treatment TAR without escalation** | | | | | | | | | | |
| CCAE | 43,894 | 78,606 | 30.8 | 58.2 | 711 | 1,201 | 1.10 (0.96 - 1.25) | 0.16 | 1.12 (0.98 - 1.28) | 0.11 |
| DAG | 12,654 | 2,255 | 12.1 | 2.1 | 103 | 24 | 0.80 (0.45 - 1.46) | 0.44 | 0.86 (0.48 - 1.55) | 0.64 |
| ODOD | 26,343 | 63,660 | 17.9 | 52.6 | 558 | 1,706 | 0.96 (0.84 - 1.09) | 0.55 | 1.01 (0.89 - 1.16) | 0.83 |
| MDCR | 8,327 | 16,640 | 7.3 | 15.9 | 200 | 476 | 0.80 (0.64 - 1.01) | 0.06 | 0.81 (0.64 - 1.02) | 0.08 |
| MDCD | 3,262 | 7,837 | 1.9 | 4.7 | 108 | 296 | 0.83 (0.61 - 1.12) | 0.22 | 0.86 (0.64 - 1.17) | 0.38 |
| Open Claims | 383,482 | 820,475 | 283.6 | 756.3 | 4,294 | 11,432 | 1.01 (0.96 - 1.06) | 0.75 | 1.01 (0.90 - 1.13) | 0.86 |
| OEHR | 34,141 | 89,528 | 13.6 | 42.5 | 327 | 1,021 | 1.08 (0.92 - 1.27) | 0.35 | 1.17 (0.95 - 1.45) | 0.23 |
| VA | 0 | 0 | 0.0 | 0.0 | 0 | 0 | | | | |
| SIDIAP | 26,189 | 7,117 | 56.0 | 16.8 | 260 | 60 | 1.38 (0.93 - 2.09) | 0.12 | 1.28 (0.85 - 1.92) | 0.26 |
| Meta-analysis | 538,292 | 1,086,118 | 423.2 | 949.1 | 6,561 | 16,216 | 1.01 (0.97 - 1.05) | 0.71 | 1.02 (0.91 - 1.14) | 0.75 |
| Meta-analysis (w/o USOC) | 154,810 | 265,643 | 139.6 | 192.8 | 2,267 | 4,784 | 0.99 (0.89 - 1.11) | 0.89 | 1.02 (0.88 - 1.19) | 0.76 |
| **PS-stratification / on-treatment TAR without escalation** | | | | | | | | | | |
| CCAE | 49,023 | 80,057 | 34.5 | 59.3 | 801 | 1,218 | 1.13 (1.03 - 1.24) | 0.01 | 1.15 (1.04 - 1.27) | 0.02 |
| DAG | 12,680 | 2,609 | 12.1 | 2.4 | 103 | 30 | 0.79 (0.51 - 1.28) | 0.33 | 0.87 (0.54 - 1.38) | 0.57 |
| ODOD | 27,811 | 63,666 | 18.9 | 52.6 | 596 | 1,706 | 0.98 (0.89 - 1.08) | 0.65 | 0.99 (0.89 - 1.10) | 0.85 |
| MDCR | 8,802 | 17,104 | 7.8 | 16.4 | 207 | 484 | 0.88 (0.74 - 1.04) | 0.12 | 0.88 (0.74 - 1.04) | 0.16 |
| MDCD | 3,340 | 7,870 | 1.9 | 4.7 | 111 | 298 | 0.94 (0.75 - 1.18) | 0.63 | 0.96 (0.76 - 1.21) | 0.75 |
| Open Claims | 390,539 | 820,488 | 290.3 | 756.3 | 4,408 | 11,433 | 0.99 (0.95 - 1.03) | 0.55 | 0.98 (0.84 - 1.15) | 0.83 |
| OEHR | 35,896 | 89,556 | 14.4 | 42.5 | 340 | 1,021 | 1.04 (0.91 - 1.18) | 0.60 | 1.15 (1.01 - 1.32) | 0.07 |
| VA | 23,930 | 221,000 | 18.3 | 208.8 | 158 | 1,880 | 0.94 (0.79 - 1.10) | 0.43 | 0.95 (0.31 - 2.88) | 0.91 |
| SIDIAP | 26,278 | 9,165 | 56.1 | 22.5 | 260 | 78 | 1.03 (0.77 - 1.39) | 0.86 | 0.99 (0.73 - 1.33) | 0.88 |
| Meta-analysis | 554,369 | 1,090,515 | 436.1 | 956.8 | 6,826 | 16,268 | 1.00 (0.95 - 1.06) | 0.95 | 1.01 (0.89 - 1.15) | 0.86 |
| Meta-analysis (w/o USOC) | 163,830 | 270,027 | 145.8 | 200.5 | 2,418 | 4,835 | 1.00 (0.93 - 1.08) | 0.92 | 1.03 (0.91 - 1.15) | 0.66 |

##### 1.6.23 Peripheral edema

**Supplementary Table 153:** Exposures, outcomes and risk estimates for Peripheral edema in DPP4I (male) (T) vs SU (male) (C) new-user cohorts under four different propensity score (PS) adjustment / time-at-risk (TAR) designs across data sources. We report total patient exposure time in 1,000 patient-year (KYr) units, uncalibrated and calibrated hazard ratios (HRs) and their 95% confidence intervals (CIs). Greyed out entries did not pass study diagnostics and are included only for completeness.

| | Patients | | Exposure (KYr) | | Outcomes | | Uncalibrated | | Calibrated | |
|---|---|---|---|---|---|---|---|---|---|---|
| | T | C | T | C | T | C | HR (95% CI) | $p$ | HR (95% CI) | $p$ |
| **PS-matching / on-treatment TAR** | | | | | | | | | | |
| CCAE | 41,136 | 73,891 | 34.9 | 60.8 | 1,297 | 2,567 | 0.90 (0.82 - 0.99) | 0.03 | 0.91 (0.83 - 1.01) | 0.12 |
| DAG | 11,877 | 2,119 | 12.4 | 2.1 | 281 | 58 | 0.95 (0.65 - 1.41) | 0.78 | 1.02 (0.69 - 1.51) | 0.72 |
| ODOD | 23,587 | 57,021 | 19.0 | 49.9 | 1,128 | 3,813 | 0.81 (0.74 - 0.89) | <0.01 | 0.84 (0.76 - 0.93) | <0.01 |
| MDCR | 6,975 | 14,141 | 6.9 | 13.9 | 563 | 1,213 | 1.01 (0.88 - 1.16) | 0.84 | 1.00 (0.87 - 1.15) | 0.80 |
| MDCD | 2,950 | 7,106 | 2.1 | 4.7 | 165 | 426 | 0.95 (0.74 - 1.23) | 0.72 | 0.99 (0.77 - 1.29) | 0.83 |
| Open Claims | 344,503 | 741,086 | 321.0 | 779.2 | 9,680 | 26,509 | 0.92 (0.89 - 0.95) | <0.01 | 0.92 (0.81 - 1.03) | 0.17 |
| OEHR | 31,993 | 83,626 | 15.2 | 43.5 | 600 | 2,420 | 0.77 (0.68 - 0.86) | <0.01 | 0.82 (0.69 - 0.97) | 0.06 |
| VA | 0 | 0 | 0.0 | 0.0 | 0 | 0 | | | | |
| SIDIAP | 24,872 | 6,927 | 71.8 | 19.3 | 1,087 | 241 | 0.95 (0.79 - 1.15) | 0.61 | 0.90 (0.75 - 1.09) | 0.34 |
| Meta-analysis | 487,893 | 985,917 | 483.3 | 973.5 | 14,801 | 37,247 | 0.89 (0.83 - 0.95) | <0.01 | 0.89 (0.79 - 1.01) | 0.08 |
| Meta-analysis (w/o USOC) | 143,390 | 244,831 | 162.3 | 194.3 | 5,121 | 10,738 | 0.88 (0.81 - 0.96) | <0.01 | 0.90 (0.81 - 1.01) | 0.13 |
| **PS-stratification / on-treatment TAR** | | | | | | | | | | |
| CCAE | 45,839 | 75,314 | 39.0 | 62.0 | 1,443 | 2,594 | 0.89 (0.83 - 0.95) | <0.01 | 0.90 (0.83 - 0.97) | 0.01 |
| DAG | 11,913 | 2,451 | 12.5 | 2.4 | 281 | 72 | 0.81 (0.61 - 1.09) | 0.15 | 0.86 (0.64 - 1.16) | 0.39 |
| ODOD | 24,999 | 57,024 | 20.2 | 49.9 | 1,204 | 3,814 | 0.83 (0.78 - 0.89) | <0.01 | 0.84 (0.78 - 0.90) | <0.01 |
| MDCR | 7,379 | 14,485 | 7.3 | 14.3 | 597 | 1,238 | 0.96 (0.87 - 1.06) | 0.45 | 0.97 (0.87 - 1.08) | 0.64 |
| MDCD | 3,021 | 7,122 | 2.1 | 4.7 | 171 | 426 | 0.92 (0.76 - 1.11) | 0.39 | 0.95 (0.78 - 1.14) | 0.61 |
| Open Claims | 350,809 | 741,094 | 328.7 | 779.2 | 9,964 | 26,510 | 0.91 (0.88 - 0.93) | <0.01 | 0.90 (0.78 - 1.04) | 0.16 |
| OEHR | 33,646 | 83,671 | 16.2 | 43.6 | 637 | 2,420 | 0.80 (0.73 - 0.88) | <0.01 | 0.88 (0.80 - 0.97) | 0.02 |
| VA | 20,139 | 190,948 | 18.4 | 193.3 | 620 | 7,649 | 0.85 (0.78 - 0.92) | <0.01 | 0.88 (0.29 - 2.63) | 0.83 |
| SIDIAP | 24,956 | 8,914 | 71.9 | 26.1 | 1,089 | 305 | 0.98 (0.85 - 1.14) | 0.83 | 0.97 (0.83 - 1.12) | 0.66 |
| Meta-analysis | 502,562 | 990,075 | 497.9 | 982.3 | 15,386 | 37,379 | 0.89 (0.84 - 0.93) | <0.01 | 0.89 (0.81 - 0.99) | 0.03 |
| Meta-analysis (w/o USOC) | 151,753 | 248,981 | 169.2 | 203.0 | 5,422 | 10,869 | 0.88 (0.83 - 0.93) | <0.01 | 0.90 (0.82 - 0.99) | 0.04 |
| **PS-matching / on-treatment TAR without escalation** | | | | | | | | | | |
| CCAE | 41,136 | 73,891 | 28.4 | 53.3 | 1,044 | 2,254 | 0.90 (0.82 - 1.00) | 0.06 | 0.92 (0.83 - 1.03) | 0.21 |
| DAG | 11,877 | 2,119 | 11.2 | 1.9 | 265 | 57 | 0.91 (0.62 - 1.37) | 0.64 | 0.99 (0.66 - 1.48) | 0.73 |
| ODOD | 23,587 | 57,021 | 15.7 | 44.9 | 931 | 3,391 | 0.79 (0.72 - 0.88) | <0.01 | 0.84 (0.76 - 0.93) | <0.01 |
| MDCR | 6,975 | 14,141 | 5.9 | 12.7 | 473 | 1,109 | 1.02 (0.88 - 1.18) | 0.84 | 1.02 (0.88 - 1.19) | 0.76 |
| MDCD | 2,950 | 7,106 | 1.6 | 4.0 | 138 | 361 | 0.93 (0.70 - 1.22) | 0.59 | 0.97 (0.73 - 1.28) | 0.78 |
| Open Claims | 344,503 | 741,086 | 250.2 | 664.6 | 7,527 | 22,576 | 0.90 (0.86 - 0.93) | <0.01 | 0.90 (0.80 - 1.00) | 0.07 |
| OEHR | 31,993 | 83,626 | 12.5 | 38.4 | 442 | 1,940 | 0.73 (0.64 - 0.83) | <0.01 | 0.79 (0.65 - 0.96) | 0.05 |
| VA | 0 | 0 | 0.0 | 0.0 | 0 | 0 | | | | |
| SIDIAP | 24,872 | 6,927 | 52.8 | 16.2 | 792 | 192 | 0.94 (0.76 - 1.18) | 0.61 | 0.88 (0.70 - 1.09) | 0.29 |
| Meta-analysis | 487,893 | 985,917 | 378.3 | 836.1 | 11,612 | 31,880 | 0.87 (0.81 - 0.95) | <0.01 | 0.88 (0.77 - 1.01) | 0.07 |
| Meta-analysis (w/o USOC) | 143,390 | 244,831 | 128.2 | 171.5 | 4,085 | 9,304 | 0.87 (0.79 - 0.96) | <0.01 | 0.90 (0.78 - 1.03) | 0.15 |
| **PS-stratification / on-treatment TAR without escalation** | | | | | | | | | | |
| CCAE | 45,839 | 75,314 | 31.8 | 54.4 | 1,159 | 2,279 | 0.87 (0.81 - 0.94) | <0.01 | 0.88 (0.81 - 0.96) | 0.01 |
| DAG | 11,913 | 2,451 | 11.3 | 2.3 | 265 | 70 | 0.79 (0.59 - 1.07) | 0.13 | 0.87 (0.64 - 1.17) | 0.41 |
| ODOD | 24,999 | 57,024 | 16.7 | 44.9 | 994 | 3,392 | 0.83 (0.77 - 0.89) | <0.01 | 0.84 (0.77 - 0.91) | <0.01 |
| MDCR | 7,379 | 14,485 | 6.3 | 13.1 | 502 | 1,132 | 0.94 (0.84 - 1.05) | 0.26 | 0.94 (0.84 - 1.05) | 0.36 |
| MDCD | 3,021 | 7,122 | 1.6 | 4.0 | 143 | 361 | 0.98 (0.80 - 1.20) | 0.87 | 1.00 (0.81 - 1.23) | 0.83 |
| Open Claims | 350,809 | 741,094 | 256.0 | 664.6 | 7,757 | 22,577 | 0.89 (0.87 - 0.92) | <0.01 | 0.89 (0.76 - 1.04) | 0.15 |
| OEHR | 33,646 | 83,671 | 13.3 | 38.4 | 467 | 1,940 | 0.77 (0.69 - 0.85) | <0.01 | 0.85 (0.76 - 0.96) | 0.03 |
| VA | 20,139 | 190,948 | 15.0 | 173.7 | 491 | 6,668 | 0.85 (0.77 - 0.93) | <0.01 | 0.86 (0.28 - 2.59) | 0.79 |
| SIDIAP | 24,956 | 8,914 | 52.8 | 21.6 | 794 | 246 | 0.97 (0.82 - 1.14) | 0.68 | 0.93 (0.78 - 1.10) | 0.44 |
| Meta-analysis | 502,562 | 990,075 | 389.9 | 843.4 | 12,081 | 31,997 | 0.87 (0.83 - 0.92) | <0.01 | 0.88 (0.78 - 0.99) | 0.04 |
| Meta-analysis (w/o USOC) | 151,753 | 248,981 | 133.9 | 178.8 | 4,324 | 9,420 | 0.87 (0.81 - 0.92) | <0.01 | 0.89 (0.80 - 0.99) | 0.04 |

##### 1.6.24 Photosensitivity

**Supplementary Table 154:** Exposures, outcomes and risk estimates for Photosensitivity in DPP4I (male) (T) vs SU (male) (C) new-user cohorts under four different propensity score (PS) adjustment / time-at-risk (TAR) designs across data sources. We report total patient exposure time in 1,000 patient-year (KYr) units, uncalibrated and calibrated hazard ratios (HRs) and their 95% confidence intervals (CIs). Greyed out entries did not pass study diagnostics and are included only for completeness.

| | Patients | | Exposure (KYr) | | Outcomes | | Uncalibrated | | Calibrated | |
|---|---|---|---|---|---|---|---|---|---|---|
| | T | C | T | C | T | C | HR (95% CI) | $p$ | HR (95% CI) | $p$ |
| **PS-matching / on-treatment TAR** | | | | | | | | | | |
| CCAE | 47,032 | 84,575 | 41.1 | 72.1 | 53 | 95 | 0.91 (0.55 - 1.50) | 0.72 | 0.93 (0.56 - 1.53) | 0.77 |
| DAG | 13,381 | 2,353 | 14.3 | 2.4 | 5 | <5 | 0.50 (0.02 - 14.1) | 0.68 | 0.54 (0.02 - 15.2) | 0.72 |
| ODOD | 28,904 | 70,090 | 24.2 | 65.7 | 44 | 120 | 0.85 (0.52 - 1.38) | 0.53 | 0.89 (0.54 - 1.45) | 0.64 |
| MDCR | 8,842 | 17,915 | 9.2 | 18.8 | 43 | 75 | 1.06 (0.64 - 1.72) | 0.83 | 1.04 (0.63 - 1.70) | 0.88 |
| MDCD | 4,062 | 9,815 | 3.1 | 7.1 | <5 | <5 | 1.39 (0.16 - 12.5) | 0.77 | 1.44 (0.16 - 12.9) | 0.74 |
| Open Claims | 418,047 | 895,307 | 399.0 | 970.9 | 213 | 484 | 1.07 (0.86 - 1.32) | 0.54 | 1.07 (0.84 - 1.36) | 0.60 |
| OEHR | 36,240 | 95,230 | 18.0 | 52.9 | 23 | 110 | 0.77 (0.41 - 1.38) | 0.40 | 0.82 (0.44 - 1.53) | 0.55 |
| VA | 0 | 0 | 0.0 | 0.0 | 0 | 0 | | | | |
| SIDIAP | 26,751 | 7,209 | 78.4 | 20.5 | 7 | 9 | 0.07 (0.01 - 0.32) | <0.01 | 0.07 (0.01 - 0.38) | <0.01 |
| Meta-analysis | 539,065 | 1,163,117 | 491.4 | 1180.4 | 376 | 884 | 1.00 (0.85 - 1.18) | 1.00 | 1.00 (0.83 - 1.22) | 0.94 |
| Meta-analysis (w/o USOC) | 121,018 | 267,810 | 92.5 | 209.5 | 163 | 400 | 0.91 (0.70 - 1.17) | 0.45 | 0.93 (0.71 - 1.21) | 0.59 |
| **PS-stratification / on-treatment TAR** | | | | | | | | | | |
| CCAE | 52,413 | 86,087 | 46.0 | 73.4 | 65 | 96 | 0.94 (0.67 - 1.31) | 0.72 | 0.95 (0.68 - 1.32) | 0.75 |
| DAG | 13,408 | 2,730 | 14.3 | 2.8 | 5 | <5 | 1.20 (0.20 - 9.69) | 0.86 | 1.28 (0.18 - 9.00) | 0.81 |
| ODOD | 30,504 | 70,097 | 25.7 | 65.7 | 51 | 120 | 1.04 (0.73 - 1.46) | 0.82 | 1.05 (0.74 - 1.49) | 0.78 |
| MDCR | 9,340 | 18,307 | 9.7 | 19.3 | 47 | 77 | 1.17 (0.80 - 1.71) | 0.41 | 1.19 (0.81 - 1.74) | 0.39 |
| MDCD | 4,156 | 9,835 | 3.2 | 7.1 | <5 | <5 | 1.36 (0.25 - 6.67) | 0.72 | 1.39 (0.27 - 7.15) | 0.69 |
| Open Claims | 425,304 | 895,320 | 408.1 | 970.9 | 221 | 484 | 1.02 (0.87 - 1.20) | 0.77 | 1.02 (0.82 - 1.26) | 0.88 |
| OEHR | 38,071 | 95,260 | 19.1 | 52.9 | 23 | 110 | 0.77 (0.47 - 1.21) | 0.28 | 0.85 (0.53 - 1.36) | 0.50 |
| VA | 25,264 | 231,142 | 23.7 | 245.5 | <10 | 367 | 0.22 (0.10 - 0.42) | <0.01 | 0.23 (0.06 - 0.84) | 0.03 |
| SIDIAP | 26,844 | 9,260 | 78.5 | 27.6 | 7 | 11 | 0.18 (0.06 - 0.50) | <0.01 | 0.17 (0.06 - 0.51) | <0.01 |
| Meta-analysis | 555,632 | 1,165,071 | 508.6 | 1182.2 | 407 | 887 | 1.01 (0.89 - 1.14) | 0.88 | 1.02 (0.88 - 1.18) | 0.81 |
| Meta-analysis (w/o USOC) | 130,328 | 269,751 | 100.5 | 211.2 | 186 | 403 | 0.99 (0.82 - 1.19) | 0.92 | 1.01 (0.83 - 1.24) | 0.91 |
| **PS-matching / on-treatment TAR without escalation** | | | | | | | | | | |
| CCAE | 47,032 | 84,575 | 33.2 | 62.7 | 43 | 88 | 0.78 (0.44 - 1.36) | 0.40 | 0.80 (0.46 - 1.41) | 0.44 |
| DAG | 13,381 | 2,353 | 12.9 | 2.2 | 5 | <5 | 0.50 (0.02 - 14.1) | 0.68 | 0.54 (0.02 - 15.3) | 0.72 |
| ODOD | 28,904 | 70,090 | 19.8 | 58.5 | 37 | 112 | 0.94 (0.55 - 1.58) | 0.83 | 0.99 (0.59 - 1.68) | 0.96 |
| MDCR | 8,842 | 17,915 | 7.8 | 17.1 | 36 | 70 | 1.04 (0.61 - 1.73) | 0.90 | 1.04 (0.62 - 1.75) | 0.89 |
| MDCD | 4,062 | 9,815 | 2.4 | 5.9 | <5 | <5 | 0.73 (0.03 - 8.48) | 0.83 | 0.77 (0.05 - 12.4) | 0.85 |
| Open Claims | 418,047 | 895,307 | 309.3 | 822.1 | 180 | 443 | 0.98 (0.78 - 1.23) | 0.89 | 0.99 (0.77 - 1.27) | 0.91 |
| OEHR | 36,240 | 95,230 | 14.6 | 45.9 | 20 | 98 | 0.87 (0.45 - 1.59) | 0.66 | 0.94 (0.49 - 1.80) | 0.86 |
| VA | 0 | 0 | 0.0 | 0.0 | 0 | 0 | | | | |
| SIDIAP | 26,751 | 7,209 | 57.4 | 17.1 | 6 | 7 | 0.10 (0.01 - 0.53) | 0.01 | 0.10 (0.02 - 0.59) | 0.01 |
| Meta-analysis | 539,065 | 1,163,117 | 384.7 | 1006.3 | 316 | 811 | 0.95 (0.80 - 1.14) | 0.60 | 0.96 (0.78 - 1.19) | 0.73 |
| Meta-analysis (w/o USOC) | 121,018 | 267,810 | 75.4 | 184.2 | 136 | 368 | 0.91 (0.69 - 1.20) | 0.51 | 0.94 (0.70 - 1.26) | 0.69 |
| **PS-stratification / on-treatment TAR without escalation** | | | | | | | | | | |
| CCAE | 52,413 | 86,087 | 37.2 | 63.8 | 54 | 89 | 0.90 (0.62 - 1.28) | 0.55 | 0.91 (0.63 - 1.31) | 0.62 |
| DAG | 13,408 | 2,730 | 12.9 | 2.6 | 5 | <5 | 1.22 (0.20 - 9.91) | 0.84 | 1.33 (0.19 - 9.40) | 0.78 |
| ODOD | 30,504 | 70,097 | 21.0 | 58.5 | 43 | 112 | 0.98 (0.67 - 1.42) | 0.94 | 1.00 (0.69 - 1.45) | 0.96 |
| MDCR | 9,340 | 18,307 | 8.3 | 17.6 | 40 | 72 | 1.07 (0.71 - 1.60) | 0.73 | 1.08 (0.72 - 1.61) | 0.72 |
| MDCD | 4,156 | 9,835 | 2.4 | 5.9 | <5 | <5 | 0.98 (0.13 - 5.38) | 0.99 | 1.00 (0.16 - 6.40) | 0.98 |
| Open Claims | 425,304 | 895,320 | 316.2 | 822.1 | 187 | 443 | 1.01 (0.84 - 1.20) | 0.95 | 1.00 (0.79 - 1.26) | 0.94 |
| OEHR | 38,071 | 95,260 | 15.5 | 45.9 | 20 | 98 | 0.79 (0.46 - 1.28) | 0.36 | 0.88 (0.53 - 1.46) | 0.63 |
| VA | 25,264 | 231,142 | 19.2 | 218.6 | <10 | 336 | 0.27 (0.12 - 0.50) | <0.01 | 0.27 (0.07 - 1.01) | 0.05 |
| SIDIAP | 26,844 | 9,260 | 57.5 | 22.8 | 6 | 8 | 0.19 (0.06 - 0.61) | <0.01 | 0.18 (0.06 - 0.61) | <0.01 |
| Meta-analysis | 555,632 | 1,165,071 | 398.1 | 1008.0 | 344 | 814 | 0.98 (0.86 - 1.12) | 0.74 | 0.99 (0.83 - 1.17) | 0.88 |
| Meta-analysis (w/o USOC) | 130,328 | 269,751 | 82.0 | 185.9 | 157 | 371 | 0.94 (0.77 - 1.15) | 0.57 | 0.97 (0.78 - 1.20) | 0.77 |

##### 1.6.25 Venous thromboembolism

**Supplementary Table 155:** Exposures, outcomes and risk estimates for Venous thromboembolism in DPP4I (male) (T) vs SU (male) (C) new-user cohorts under four different propensity score (PS) adjustment / time-at-risk (TAR) designs across data sources. We report total patient exposure time in 1,000 patient-year (KYr) units, uncalibrated and calibrated hazard ratios (HRs) and their 95% confidence intervals (CIs). Greyed out entries did not pass study diagnostics and are included only for completeness.

| | Patients | | Exposure (KYr) | | Outcomes | | Uncalibrated | | Calibrated | |
|---|---|---|---|---|---|---|---|---|---|---|
| | T | C | T | C | T | C | HR (95% CI) | $p$ | HR (95% CI) | $p$ |
| **PS-matching / on-treatment TAR** | | | | | | | | | | |
| CCAE | 46,500 | 83,393 | 40.6 | 70.9 | 184 | 427 | 0.86 (0.67 - 1.09) | 0.21 | 0.87 (0.68 - 1.11) | 0.28 |
| DAG | 12,703 | 2,266 | 13.4 | 2.3 | 90 | 24 | 0.96 (0.53 - 1.83) | 0.89 | 1.03 (0.55 - 1.94) | 0.81 |
| ODOD | 28,387 | 68,587 | 23.8 | 63.9 | 214 | 787 | 0.86 (0.69 - 1.05) | 0.14 | 0.89 (0.72 - 1.10) | 0.30 |
| MDCR | 8,678 | 17,475 | 9.0 | 18.3 | 125 | 293 | 0.75 (0.55 - 1.00) | 0.06 | 0.74 (0.55 - 0.99) | 0.05 |
| MDCD | 3,893 | 9,404 | 3.0 | 6.8 | 44 | 71 | 1.72 (0.99 - 2.97) | 0.05 | 1.79 (1.04 - 3.10) | 0.04 |
| Open Claims | 408,514 | 873,694 | 389.4 | 945.3 | 1,917 | 5,766 | 0.83 (0.77 - 0.89) | <0.01 | 0.83 (0.72 - 0.95) | <0.01 |
| OEHR | 35,569 | 93,391 | 17.5 | 51.3 | 120 | 499 | 0.80 (0.62 - 1.04) | 0.10 | 0.86 (0.64 - 1.14) | 0.35 |
| VA | 0 | 0 | 0.0 | 0.0 | 0 | 0 | | | | |
| SIDIAP | 26,225 | 7,125 | 76.6 | 20.2 | 275 | 61 | 0.84 (0.59 - 1.22) | 0.36 | 0.80 (0.56 - 1.15) | 0.24 |
| Meta-analysis | 570,469 | 1,155,335 | 573.2 | 1178.9 | 2,969 | 7,928 | 0.84 (0.79 - 0.89) | <0.01 | 0.84 (0.74 - 0.95) | <0.01 |
| Meta-analysis (w/o USOC) | 161,955 | 281,641 | 183.9 | 233.7 | 1,052 | 2,162 | 0.86 (0.76 - 0.98) | 0.02 | 0.88 (0.76 - 1.02) | 0.14 |
| **PS-stratification / on-treatment TAR** | | | | | | | | | | |
| CCAE | 51,836 | 84,899 | 45.4 | 72.2 | 204 | 430 | 0.77 (0.65 - 0.92) | <0.01 | 0.78 (0.65 - 0.93) | <0.01 |
| DAG | 12,728 | 2,625 | 13.5 | 2.7 | 91 | 28 | 0.71 (0.45 - 1.16) | 0.16 | 0.76 (0.47 - 1.22) | 0.28 |
| ODOD | 29,968 | 68,594 | 25.2 | 63.9 | 223 | 787 | 0.80 (0.69 - 0.94) | <0.01 | 0.81 (0.69 - 0.95) | 0.02 |
| MDCR | 9,176 | 17,858 | 9.6 | 18.7 | 128 | 298 | 0.86 (0.69 - 1.07) | 0.18 | 0.87 (0.70 - 1.09) | 0.23 |
| MDCD | 3,976 | 9,424 | 3.0 | 6.8 | 45 | 71 | 1.70 (1.13 - 2.52) | <0.01 | 1.74 (1.17 - 2.60) | <0.01 |
| Open Claims | 415,805 | 873,708 | 398.6 | 945.3 | 1,959 | 5,766 | 0.85 (0.80 - 0.89) | <0.01 | 0.84 (0.72 - 0.98) | 0.03 |
| OEHR | 37,345 | 93,445 | 18.6 | 51.4 | 124 | 501 | 0.76 (0.62 - 0.93) | 0.01 | 0.84 (0.68 - 1.03) | 0.11 |
| VA | 24,743 | 227,637 | 23.1 | 241.0 | 103 | 1,296 | 0.82 (0.67 - 1.00) | 0.06 | 0.85 (0.28 - 2.59) | 0.78 |
| SIDIAP | 26,327 | 9,159 | 76.7 | 27.2 | 276 | 77 | 1.09 (0.82 - 1.46) | 0.57 | 1.07 (0.80 - 1.42) | 0.68 |
| Meta-analysis | 587,161 | 1,159,712 | 590.6 | 1188.1 | 3,050 | 7,958 | 0.84 (0.81 - 0.88) | <0.01 | 0.85 (0.77 - 0.94) | <0.01 |
| Meta-analysis (w/o USOC) | 171,356 | 286,004 | 192.1 | 242.9 | 1,091 | 2,192 | 0.88 (0.76 - 1.03) | 0.12 | 0.90 (0.76 - 1.07) | 0.25 |
| **PS-matching / on-treatment TAR without escalation** | | | | | | | | | | |
| CCAE | 46,500 | 83,393 | 32.8 | 61.7 | 154 | 358 | 0.90 (0.69 - 1.17) | 0.42 | 0.92 (0.70 - 1.20) | 0.54 |
| DAG | 12,703 | 2,266 | 12.1 | 2.1 | 85 | 23 | 1.02 (0.55 - 1.98) | 0.96 | 1.10 (0.58 - 2.10) | 0.76 |
| ODOD | 28,387 | 68,587 | 19.4 | 57.0 | 177 | 691 | 0.86 (0.68 - 1.07) | 0.19 | 0.91 (0.72 - 1.14) | 0.41 |
| MDCR | 8,678 | 17,475 | 7.7 | 16.7 | 103 | 264 | 0.73 (0.53 - 1.00) | 0.05 | 0.73 (0.53 - 1.01) | 0.06 |
| MDCD | 3,893 | 9,404 | 2.3 | 5.7 | 36 | 57 | 1.78 (0.96 - 3.28) | 0.07 | 1.86 (1.00 - 3.45) | 0.05 |
| Open Claims | 408,514 | 873,694 | 302.2 | 801.1 | 1,485 | 4,927 | 0.81 (0.75 - 0.87) | <0.01 | 0.81 (0.71 - 0.92) | <0.01 |
| OEHR | 35,569 | 93,391 | 14.2 | 44.7 | 92 | 369 | 0.81 (0.60 - 1.09) | 0.18 | 0.88 (0.64 - 1.22) | 0.50 |
| VA | 0 | 0 | 0.0 | 0.0 | 0 | 0 | | | | |
| SIDIAP | 26,225 | 7,125 | 56.2 | 16.9 | 180 | 49 | 0.82 (0.53 - 1.27) | 0.36 | 0.76 (0.49 - 1.17) | 0.23 |
| Meta-analysis | 570,469 | 1,155,335 | 446.8 | 1005.8 | 2,312 | 6,738 | 0.82 (0.77 - 0.88) | <0.01 | 0.83 (0.73 - 0.94) | <0.01 |
| Meta-analysis (w/o USOC) | 161,955 | 281,641 | 144.7 | 204.7 | 827 | 1,811 | 0.87 (0.76 - 1.00) | 0.04 | 0.90 (0.76 - 1.06) | 0.22 |
| **PS-stratification / on-treatment TAR without escalation** | | | | | | | | | | |
| CCAE | 51,836 | 84,899 | 36.7 | 62.9 | 168 | 361 | 0.84 (0.69 - 1.01) | 0.07 | 0.85 (0.70 - 1.03) | 0.12 |
| DAG | 12,728 | 2,625 | 12.1 | 2.5 | 86 | 27 | 0.69 (0.43 - 1.14) | 0.14 | 0.75 (0.46 - 1.23) | 0.29 |
| ODOD | 29,968 | 68,594 | 20.6 | 57.0 | 183 | 691 | 0.82 (0.69 - 0.97) | 0.02 | 0.83 (0.70 - 0.99) | 0.05 |
| MDCR | 9,176 | 17,858 | 8.2 | 17.1 | 105 | 269 | 0.84 (0.66 - 1.05) | 0.13 | 0.84 (0.66 - 1.06) | 0.16 |
| MDCD | 3,976 | 9,424 | 2.3 | 5.7 | 37 | 57 | 1.92 (1.23 - 2.97) | <0.01 | 1.95 (1.26 - 3.03) | <0.01 |
| Open Claims | 415,805 | 873,708 | 309.1 | 801.1 | 1,522 | 4,927 | 0.83 (0.78 - 0.88) | <0.01 | 0.82 (0.70 - 0.98) | 0.03 |
| OEHR | 37,345 | 93,445 | 15.1 | 44.7 | 96 | 370 | 0.83 (0.65 - 1.05) | 0.13 | 0.93 (0.73 - 1.18) | 0.57 |
| VA | 24,743 | 227,637 | 18.8 | 214.8 | 74 | 1,065 | 0.78 (0.61 - 0.99) | 0.04 | 0.79 (0.26 - 2.45) | 0.69 |
| SIDIAP | 26,327 | 9,159 | 56.3 | 22.5 | 180 | 59 | 0.94 (0.67 - 1.33) | 0.71 | 0.90 (0.64 - 1.28) | 0.57 |
| Meta-analysis | 587,161 | 1,159,712 | 460.5 | 1013.5 | 2,377 | 6,761 | 0.84 (0.80 - 0.88) | <0.01 | 0.85 (0.75 - 0.96) | <0.01 |
| Meta-analysis (w/o USOC) | 171,356 | 286,004 | 151.4 | 212.4 | 855 | 1,834 | 0.89 (0.76 - 1.05) | 0.16 | 0.91 (0.76 - 1.09) | 0.34 |

##### 1.6.26 Vomiting

**Supplementary Table 156:** Exposures, outcomes and risk estimates for Vomiting in DPP4I (male) (T) vs SU (male) (C) new-user cohorts under four different propensity score (PS) adjustment / time-at-risk (TAR) designs across data sources. We report total patient exposure time in 1,000 patient-year (KYr) units, uncalibrated and calibrated hazard ratios (HRs) and their 95% confidence intervals (CIs). Greyed out entries did not pass study diagnostics and are included only for completeness.

| | Patients | | Exposure (KYr) | | Outcomes | | Uncalibrated | | Calibrated | |
|---|---|---|---|---|---|---|---|---|---|---|
| | T | C | T | C | T | C | HR (95% CI) | $p$ | HR (95% CI) | $p$ |
| **PS-matching / on-treatment TAR** | | | | | | | | | | |
| CCAE | 44,822 | 80,290 | 39.0 | 68.2 | 634 | 1,015 | 1.12 (0.98 - 1.29) | 0.10 | 1.14 (0.99 - 1.31) | 0.09 |
| DAG | 12,643 | 2,255 | 13.4 | 2.3 | 116 | 25 | 0.77 (0.45 - 1.36) | 0.35 | 0.83 (0.48 - 1.45) | 0.55 |
| ODOD | 27,096 | 65,489 | 22.4 | 60.9 | 519 | 1,409 | 1.09 (0.95 - 1.25) | 0.24 | 1.13 (0.98 - 1.30) | 0.12 |
| MDCR | 8,485 | 16,955 | 8.7 | 17.7 | 190 | 460 | 0.78 (0.61 - 0.99) | 0.04 | 0.77 (0.60 - 0.98) | 0.04 |
| MDCD | 3,406 | 8,130 | 2.6 | 5.8 | 111 | 279 | 0.95 (0.69 - 1.29) | 0.74 | 0.99 (0.73 - 1.35) | 0.85 |
| Open Claims | 391,579 | 836,022 | 372.6 | 909.3 | 4,488 | 10,879 | 1.00 (0.96 - 1.05) | 0.87 | 1.00 (0.88 - 1.14) | 0.90 |
| OEHR | 34,905 | 91,536 | 17.1 | 50.0 | 282 | 954 | 0.98 (0.82 - 1.17) | 0.80 | 1.04 (0.84 - 1.29) | 0.74 |
| VA | 0 | 0 | 0.0 | 0.0 | 0 | 0 | | | | |
| SIDIAP | 26,125 | 7,109 | 76.3 | 20.1 | 368 | 83 | 1.06 (0.78 - 1.47) | 0.71 | 1.01 (0.73 - 1.39) | 0.88 |
| Meta-analysis | 549,061 | 1,107,786 | 552.2 | 1134.3 | 6,708 | 15,104 | 1.01 (0.97 - 1.05) | 0.61 | 1.02 (0.91 - 1.14) | 0.80 |
| Meta-analysis (w/o USOC) | 157,482 | 271,764 | 179.5 | 225.0 | 2,220 | 4,225 | 1.01 (0.91 - 1.11) | 0.89 | 1.03 (0.91 - 1.17) | 0.69 |
| **PS-stratification / on-treatment TAR** | | | | | | | | | | |
| CCAE | 50,059 | 81,748 | 43.6 | 69.5 | 717 | 1,031 | 1.14 (1.03 - 1.26) | 0.01 | 1.15 (1.03 - 1.27) | 0.02 |
| DAG | 12,669 | 2,608 | 13.4 | 2.7 | 116 | 31 | 0.87 (0.56 - 1.38) | 0.55 | 0.93 (0.59 - 1.46) | 0.73 |
| ODOD | 28,618 | 65,496 | 23.8 | 60.9 | 555 | 1,409 | 1.05 (0.95 - 1.16) | 0.36 | 1.06 (0.95 - 1.18) | 0.34 |
| MDCR | 8,966 | 17,423 | 9.3 | 18.3 | 202 | 467 | 0.85 (0.72 - 1.01) | 0.07 | 0.86 (0.73 - 1.03) | 0.11 |
| MDCD | 3,484 | 8,165 | 2.6 | 5.8 | 117 | 280 | 0.94 (0.75 - 1.18) | 0.61 | 0.97 (0.77 - 1.22) | 0.78 |
| Open Claims | 398,740 | 836,034 | 381.7 | 909.3 | 4,622 | 10,880 | 1.02 (0.98 - 1.05) | 0.39 | 1.01 (0.87 - 1.17) | 0.90 |
| OEHR | 36,697 | 91,564 | 18.1 | 50.0 | 295 | 954 | 0.95 (0.83 - 1.09) | 0.47 | 1.04 (0.91 - 1.20) | 0.58 |
| VA | 24,357 | 224,001 | 22.9 | 237.7 | 156 | 1,701 | 0.94 (0.80 - 1.11) | 0.47 | 0.97 (0.32 - 2.94) | 0.92 |
| SIDIAP | 26,215 | 9,149 | 76.4 | 27.1 | 368 | 114 | 0.92 (0.72 - 1.17) | 0.49 | 0.90 (0.71 - 1.15) | 0.41 |
| Meta-analysis | 565,448 | 1,112,187 | 569.0 | 1143.5 | 6,992 | 15,166 | 1.00 (0.94 - 1.07) | 0.97 | 1.01 (0.91 - 1.13) | 0.85 |
| Meta-analysis (w/o USOC) | 166,708 | 276,153 | 187.3 | 234.2 | 2,370 | 4,286 | 0.99 (0.91 - 1.08) | 0.82 | 1.01 (0.90 - 1.13) | 0.85 |
| **PS-matching / on-treatment TAR without escalation** | | | | | | | | | | |
| CCAE | 44,822 | 80,290 | 31.6 | 59.5 | 527 | 891 | 1.10 (0.95 - 1.28) | 0.19 | 1.13 (0.97 - 1.31) | 0.13 |
| DAG | 12,643 | 2,255 | 12.1 | 2.1 | 105 | 24 | 0.80 (0.46 - 1.47) | 0.46 | 0.87 (0.49 - 1.56) | 0.66 |
| ODOD | 27,096 | 65,489 | 18.5 | 54.4 | 435 | 1,234 | 1.09 (0.94 - 1.27) | 0.27 | 1.15 (0.98 - 1.34) | 0.10 |
| MDCR | 8,485 | 16,955 | 7.5 | 16.2 | 164 | 404 | 0.78 (0.60 - 1.01) | 0.06 | 0.79 (0.61 - 1.02) | 0.07 |
| MDCD | 3,406 | 8,130 | 2.0 | 4.9 | 94 | 239 | 0.96 (0.69 - 1.33) | 0.81 | 1.00 (0.72 - 1.40) | 0.85 |
| Open Claims | 391,579 | 836,022 | 289.9 | 772.3 | 3,390 | 8,989 | 0.99 (0.94 - 1.04) | 0.66 | 0.99 (0.88 - 1.12) | 0.86 |
| OEHR | 34,905 | 91,536 | 13.9 | 43.7 | 189 | 718 | 0.92 (0.75 - 1.13) | 0.43 | 1.00 (0.78 - 1.28) | 0.88 |
| VA | 0 | 0 | 0.0 | 0.0 | 0 | 0 | | | | |
| SIDIAP | 26,125 | 7,109 | 55.9 | 16.8 | 280 | 67 | 1.38 (0.95 - 2.04) | 0.10 | 1.28 (0.87 - 1.88) | 0.23 |
| Meta-analysis | 549,061 | 1,107,786 | 431.3 | 969.8 | 5,184 | 12,566 | 1.00 (0.94 - 1.07) | 0.88 | 1.02 (0.89 - 1.15) | 0.80 |
| Meta-analysis (w/o USOC) | 157,482 | 271,764 | 141.4 | 197.5 | 1,794 | 3,577 | 1.01 (0.90 - 1.13) | 0.89 | 1.04 (0.89 - 1.21) | 0.63 |
| **PS-stratification / on-treatment TAR without escalation** | | | | | | | | | | |
| CCAE | 50,059 | 81,748 | 35.4 | 60.6 | 593 | 907 | 1.15 (1.03 - 1.28) | 0.01 | 1.17 (1.04 - 1.31) | 0.02 |
| DAG | 12,669 | 2,608 | 12.1 | 2.4 | 105 | 30 | 0.81 (0.52 - 1.30) | 0.37 | 0.88 (0.55 - 1.40) | 0.62 |
| ODOD | 28,618 | 65,496 | 19.6 | 54.4 | 466 | 1,234 | 1.09 (0.97 - 1.22) | 0.14 | 1.11 (0.98 - 1.24) | 0.14 |
| MDCR | 8,966 | 17,423 | 7.9 | 16.7 | 172 | 411 | 0.89 (0.73 - 1.06) | 0.20 | 0.89 (0.74 - 1.07) | 0.24 |
| MDCD | 3,484 | 8,165 | 2.0 | 4.9 | 98 | 240 | 1.02 (0.79 - 1.30) | 0.89 | 1.03 (0.81 - 1.33) | 0.80 |
| Open Claims | 398,740 | 836,034 | 296.7 | 772.3 | 3,494 | 8,990 | 1.00 (0.96 - 1.04) | 0.84 | 0.99 (0.84 - 1.16) | 0.88 |
| OEHR | 36,697 | 91,564 | 14.8 | 43.7 | 198 | 718 | 0.90 (0.76 - 1.06) | 0.19 | 1.00 (0.84 - 1.18) | 0.87 |
| VA | 24,357 | 224,001 | 18.6 | 212.1 | 128 | 1,427 | 1.00 (0.83 - 1.19) | 1.00 | 1.01 (0.33 - 3.09) | 0.93 |
| SIDIAP | 26,215 | 9,149 | 56.0 | 22.4 | 280 | 87 | 1.00 (0.76 - 1.33) | 1.00 | 0.96 (0.72 - 1.28) | 0.79 |
| Meta-analysis | 565,448 | 1,112,187 | 444.6 | 977.6 | 5,406 | 12,617 | 1.01 (0.94 - 1.09) | 0.79 | 1.02 (0.89 - 1.16) | 0.79 |
| Meta-analysis (w/o USOC) | 166,708 | 276,153 | 147.8 | 205.3 | 1,912 | 3,627 | 1.01 (0.92 - 1.11) | 0.83 | 1.03 (0.91 - 1.18) | 0.62 |

### 2 Baseline patient characteristics across data sources

#### 2.1 GLP1RA (male) vs SGLT2I (male)

##### 2.1.1 CCAE

**Supplementary Table 157:** Baseline patient characteristics for GLP1RA (male) (T) and SGLT2I (male) (C) new-users in the CCAE data source. We report proportion of initiators satisfying selected base-line characteristics and the standardized difference of population proportions (StdDiff) before and after propensity score adjustment. Less extreme StdDiffs through matching and stratification suggest improved balance between patient cohorts through adjustment.

| | Before adjustment | | | After matching | | | After stratification | | |
|---|---|---|---|---|---|---|---|---|---|
| Characteristic | T (%) | C (%) | StdDiff | T (%) | C (%) | StdDiff | T (%) | C (%) | StdDiff |
| **Age group** | | | | | | | | | |
| 20 - 24 | 0.4 | 0.3 | 0.02 | 0.4 | 0.4 | 0.00 | 0.4 | 0.3 | 0.01 |
| 25 - 29 | 0.8 | 0.5 | 0.04 | 0.8 | 0.6 | 0.03 | 0.7 | 0.5 | 0.03 |
| 30 - 34 | 2.6 | 1.9 | 0.05 | 2.5 | 2.3 | 0.01 | 2.3 | 2.0 | 0.02 |
| 35 - 39 | 5.7 | 4.6 | 0.05 | 5.6 | 5.4 | 0.01 | 5.2 | 4.9 | 0.01 |
| 40 - 44 | 10.8 | 9.1 | 0.06 | 10.4 | 10.3 | 0.00 | 9.5 | 9.5 | 0.00 |
| 45 - 49 | 16.1 | 15.4 | 0.02 | 15.9 | 15.6 | 0.01 | 15.9 | 15.6 | 0.01 |
| 50 - 54 | 20.8 | 20.3 | 0.01 | 20.7 | 20.4 | 0.01 | 20.7 | 20.3 | 0.01 |
| 55 - 59 | 21.8 | 23.8 | -0.05 | 22.1 | 23.3 | -0.03 | 22.3 | 23.6 | -0.03 |
| 60 - 64 | 19.3 | 22.2 | -0.07 | 19.8 | 19.5 | 0.01 | 21.1 | 21.3 | 0.00 |
| 65 - 69 | 1.7 | 1.9 | -0.01 | 1.6 | 1.8 | -0.01 | 1.7 | 1.8 | -0.01 |
| **Medical history: General** | | | | | | | | | |
| Acute respiratory disease | 24.7 | 23.1 | 0.04 | 24.6 | 24.7 | 0.00 | 24.6 | 24.7 | 0.00 |
| Attention deficit hyperactivity disorder | 1.6 | 1.2 | 0.03 | 1.6 | 1.7 | -0.01 | 1.4 | 1.4 | 0.00 |
| Chronic liver disease | 1.5 | 1.6 | -0.01 | 1.1 | 1.3 | -0.02 | 1.2 | 1.5 | -0.03 |
| Chronic obstructive lung disease | 2.6 | 2.3 | 0.02 | 2.4 | 2.6 | -0.01 | 2.2 | 2.4 | -0.01 |
| Crohn’s disease | 0.2 | 0.1 | 0.02 | 0.2 | 0.1 | 0.02 | 0.2 | 0.1 | 0.02 |
| Dementia | 0.1 | 0.1 | 0.00 | 0.1 | 0.1 | 0.00 | 0.1 | 0.1 | 0.00 |
| Depressive disorder | 9.0 | 7.0 | 0.07 | 8.9 | 9.3 | -0.01 | 7.8 | 7.9 | 0.00 |
| Gastroesophageal reflux disease | 11.2 | 11.3 | 0.00 | 12.1 | 12.3 | -0.01 | 11.5 | 11.7 | -0.01 |
| Gastrointestinal hemorrhage | 1.9 | 1.4 | 0.04 | 1.8 | 1.6 | 0.02 | 1.6 | 1.5 | 0.01 |
| Human immunodeficiency virus infection | 0.4 | 0.5 | -0.02 | 0.4 | 0.6 | -0.02 | 0.3 | 0.5 | -0.03 |
| Hyperlipidemia | 71.3 | 75.7 | -0.10 | 74.3 | 74.2 | 0.00 | 75.3 | 75.2 | 0.00 |
| Hypertensive disorder | 70.8 | 72.2 | -0.03 | 73.2 | 73.1 | 0.00 | 73.5 | 72.6 | 0.02 |
| Lesion of liver | 0.6 | 0.6 | 0.00 | 0.6 | 0.8 | -0.02 | 0.5 | 0.7 | -0.02 |
| Obesity | 33.0 | 25.3 | 0.17 | 35.2 | 37.1 | -0.04 | 28.8 | 29.5 | -0.01 |
| Osteoarthritis | 18.5 | 15.8 | 0.07 | 18.1 | 18.2 | 0.00 | 16.8 | 16.7 | 0.00 |
| Pneumonia | 2.1 | 1.9 | 0.01 | 2.0 | 2.2 | -0.01 | 1.8 | 2.0 | -0.02 |
| Psoriasis | 2.0 | 1.7 | 0.02 | 2.0 | 1.9 | 0.01 | 1.9 | 1.7 | 0.01 |
| Renal impairment | 4.1 | 3.6 | 0.03 | 4.5 | 4.4 | 0.00 | 4.0 | 3.9 | 0.01 |
| Rheumatoid arthritis | 0.5 | 0.6 | 0.00 | 0.5 | 0.6 | -0.01 | 0.5 | 0.6 | -0.01 |
| Schizophrenia | 0.1 | 0.1 | 0.00 | 0.1 | 0.1 | 0.00 | 0.1 | 0.1 | 0.00 |
| Ulcerative colitis | 0.2 | 0.3 | -0.01 | 0.2 | 0.3 | -0.01 | 0.2 | 0.3 | -0.02 |
| Urinary tract infectious disease | 2.8 | 2.1 | 0.04 | 2.5 | 2.3 | 0.02 | 2.5 | 2.2 | 0.02 |
| Viral hepatitis C | 0.2 | 0.3 | -0.01 | 0.2 | 0.3 | -0.02 | 0.2 | 0.3 | -0.02 |
| Visual system disorder | 22.0 | 22.7 | -0.02 | 22.4 | 22.6 | 0.00 | 22.4 | 22.7 | -0.01 |
| **Medical history: Cardiovascular disease** | | | | | | | | | |
| Atrial fibrillation | 2.8 | 2.7 | 0.01 | 2.9 | 2.8 | 0.01 | 2.8 | 2.8 | 0.00 |
| Cerebrovascular disease | 2.2 | 2.5 | -0.01 | 2.1 | 2.3 | -0.01 | 2.1 | 2.4 | -0.02 |
| Coronary arteriosclerosis | 10.2 | 11.3 | -0.04 | 9.6 | 9.4 | 0.01 | 11.2 | 10.7 | 0.01 |
| Heart disease | 20.4 | 20.3 | 0.00 | 19.4 | 19.0 | 0.01 | 20.6 | 20.0 | 0.01 |
| Heart failure | 2.5 | 2.5 | 0.00 | 2.5 | 2.7 | -0.01 | 2.4 | 2.6 | -0.01 |
| Ischemic heart disease | 5.0 | 5.4 | -0.02 | 4.7 | 4.7 | 0.00 | 5.1 | 5.2 | -0.01 |
| Peripheral vascular disease | 4.1 | 4.2 | 0.00 | 4.3 | 4.5 | -0.01 | 4.2 | 4.3 | 0.00 |
| Pulmonary embolism | 0.7 | 0.4 | 0.03 | 0.7 | 0.5 | 0.02 | 0.6 | 0.5 | 0.02 |
| Venous thrombosis | 1.2 | 0.9 | 0.02 | 1.1 | 1.0 | 0.01 | 1.0 | 1.0 | 0.01 |
| **Medical history: Neoplasms** | | | | | | | | | |
| Hematologic neoplasm | 0.7 | 0.7 | 0.00 | 0.8 | 0.8 | 0.01 | 0.8 | 0.7 | 0.01 |
| Malignant lymphoma | 0.3 | 0.4 | 0.00 | 0.4 | 0.4 | 0.00 | 0.4 | 0.4 | 0.01 |
| Malignant neoplasm of anorectum | 0.1 | 0.1 | 0.00 | 0.1 | 0.1 | -0.01 | 0.1 | 0.1 | -0.01 |
| Malignant neoplastic disease | 4.6 | 4.7 | 0.00 | 4.5 | 5.0 | -0.02 | 4.7 | 4.8 | 0.00 |
| Malignant tumor of colon | 0.2 | 0.3 | -0.02 | 0.2 | 0.3 | -0.01 | 0.3 | 0.3 | 0.00 |

| (Continued) | Before ajustment | | | After matching | | | After stratification | | |
|---|---|---|---|---|---|---|---|---|---|
| Characteristic | T (%) | C (%) | StdDiff | T (%) | C (%) | StdDiff | T (%) | C (%) | StdDiff |
| Malignant tumor of lung | 0.1 | 0.1 | -0.01 | 0.1 | 0.1 | 0.00 | 0.1 | 0.1 | 0.00 |
| Malignant tumor of urinary bladder | 0.2 | 0.2 | 0.01 | 0.2 | 0.2 | 0.00 | 0.2 | 0.2 | 0.00 |
| Primary malignant neoplasm of prostate | 1.0 | 1.0 | 0.00 | 1.0 | 1.0 | 0.00 | 1.1 | 1.0 | 0.01 |
| **Medication use** | | | | | | | | | |
| Agents acting on the renin-angiotensin system | 71.3 | 71.2 | 0.00 | 71.1 | 71.3 | 0.00 | 71.9 | 71.3 | 0.01 |
| Antibacterials for systemic use | 53.6 | 50.9 | 0.05 | 52.0 | 52.3 | -0.01 | 51.6 | 51.5 | 0.00 |
| Antidepressants | 24.9 | 18.8 | 0.15 | 23.3 | 23.9 | -0.01 | 20.4 | 20.7 | -0.01 |
| Antiepileptics | 11.8 | 9.9 | 0.06 | 11.6 | 12.0 | -0.01 | 10.5 | 10.7 | -0.01 |
| Antiinflammatory and antirheumatic products | 31.7 | 30.2 | 0.03 | 31.9 | 32.2 | -0.01 | 30.5 | 30.9 | -0.01 |
| Antineoplastic agents | 3.9 | 3.1 | 0.04 | 3.4 | 3.4 | 0.00 | 3.2 | 3.3 | -0.01 |
| Antipsoriatics | 0.8 | 0.5 | 0.03 | 0.7 | 0.5 | 0.03 | 0.8 | 0.5 | 0.03 |
| Antithrombotic agents | 11.5 | 12.1 | -0.02 | 11.4 | 11.5 | 0.00 | 12.0 | 12.0 | 0.00 |
| Calcium channel blockers | 23.5 | 22.8 | 0.02 | 23.7 | 23.4 | 0.01 | 23.7 | 23.4 | 0.01 |
| Diuretics | 36.7 | 32.5 | 0.09 | 34.4 | 34.6 | 0.00 | 33.7 | 33.3 | 0.01 |
| Drugs for acid related disorders | 22.7 | 20.6 | 0.05 | 22.0 | 21.8 | 0.01 | 21.1 | 21.1 | 0.00 |
| Drugs for obstructive airway diseases | 35.3 | 31.8 | 0.07 | 34.6 | 34.9 | -0.01 | 32.7 | 33.0 | -0.01 |
| Immunosuppressants | 2.0 | 1.7 | 0.02 | 2.1 | 2.1 | 0.00 | 1.9 | 1.9 | 0.01 |
| Lipid modifying agents | 71.1 | 72.9 | -0.04 | 71.5 | 71.4 | 0.00 | 72.7 | 72.4 | 0.01 |
| Opioids | 22.9 | 21.4 | 0.04 | 22.4 | 22.1 | 0.01 | 22.0 | 21.8 | 0.01 |
| Psycholeptics | 19.9 | 17.6 | 0.06 | 19.5 | 19.6 | 0.00 | 18.2 | 18.4 | 0.00 |
| Psychostimulants | 4.0 | 2.7 | 0.07 | 3.8 | 3.7 | 0.01 | 3.3 | 3.0 | 0.01 |

##### 2.1.2 ODOD

**Supplementary Table 158:** Baseline patient characteristics for GLP1RA (male) (T) and SGLT2I (male) (C) new-users in the ODOD data source. We report proportion of initiators satisfying selected base-line characteristics and the standardized difference of population proportions (StdDiff) before and after propensity score adjustment. Less extreme StdDiffs through matching and stratification suggest improved balance between patient cohorts through adjustment.

| | Before adjustment | | | After matching | | | After stratification | | |
|---|---|---|---|---|---|---|---|---|---|
| Characteristic | T (%) | C (%) | StdDiff | T (%) | C (%) | StdDiff | T (%) | C (%) | StdDiff |
| **Age group** | | | | | | | | | |
| 20 - 24 | 0.3 | 0.1 | 0.05 | 0.3 | 0.2 | 0.03 | 0.2 | 0.1 | 0.03 |
| 25 - 29 | 0.4 | 0.4 | 0.00 | 0.3 | 0.5 | -0.03 | 0.3 | 0.4 | -0.03 |
| 30 - 34 | 2.1 | 1.3 | 0.06 | 1.8 | 1.6 | 0.02 | 1.7 | 1.4 | 0.03 |
| 35 - 39 | 4.2 | 3.2 | 0.05 | 4.2 | 3.6 | 0.03 | 3.6 | 3.4 | 0.01 |
| 40 - 44 | 7.9 | 6.1 | 0.07 | 7.4 | 7.2 | 0.01 | 6.7 | 6.5 | 0.01 |
| 45 - 49 | 12.1 | 9.5 | 0.08 | 11.3 | 11.3 | 0.00 | 10.3 | 10.2 | 0.00 |
| 50 - 54 | 14.4 | 13.3 | 0.03 | 13.7 | 14.3 | -0.02 | 13.2 | 13.7 | -0.01 |
| 55 - 59 | 16.4 | 15.0 | 0.04 | 16.2 | 15.4 | 0.02 | 16.1 | 15.1 | 0.03 |
| 60 - 64 | 14.3 | 14.6 | -0.01 | 14.4 | 14.5 | 0.00 | 14.6 | 14.6 | 0.00 |
| 65 - 69 | 12.8 | 14.7 | -0.06 | 13.6 | 13.9 | -0.01 | 14.1 | 14.3 | -0.01 |
| 70 - 74 | 9.1 | 12.1 | -0.10 | 10.0 | 10.0 | 0.00 | 10.0 | 10.0 | 0.00 |
| 75 - 79 | 4.3 | 6.4 | -0.09 | 4.8 | 4.9 | 0.00 | 5.7 | 5.9 | -0.01 |
| 80 - 84 | 1.4 | 2.3 | -0.07 | 1.5 | 1.8 | -0.02 | 1.7 | 2.1 | -0.03 |
| 85 - 89 | 0.3 | 0.9 | -0.07 | 0.4 | 0.7 | -0.05 | 0.4 | 0.8 | -0.05 |
| 90 - 94 | <0.1 | 0.1 | -0.03 | <0.1 | 0.1 | -0.03 | <0.1 | 0.1 | -0.03 |
| **Medical history: General** | | | | | | | | | |
| Acute respiratory disease | 22.6 | 21.6 | 0.03 | 21.7 | 22.1 | -0.01 | 21.5 | 21.8 | -0.01 |
| Attention deficit hyperactivity disorder | 1.8 | 0.9 | 0.07 | 1.5 | 1.4 | 0.01 | 1.3 | 1.1 | 0.01 |
| Chronic liver disease | 1.7 | 1.7 | 0.00 | 1.5 | 1.4 | 0.00 | 1.5 | 1.6 | -0.01 |
| Chronic obstructive lung disease | 7.3 | 7.4 | 0.00 | 7.3 | 7.5 | -0.01 | 7.3 | 7.5 | -0.01 |
| Crohn’s disease | 0.2 | 0.3 | -0.01 | 0.2 | 0.3 | -0.02 | 0.2 | 0.3 | -0.01 |
| Dementia | 0.6 | 0.8 | -0.02 | 0.7 | 0.7 | -0.01 | 0.7 | 0.8 | 0.00 |
| Depressive disorder | 12.7 | 9.8 | 0.09 | 12.1 | 12.8 | -0.02 | 10.8 | 11.0 | 0.00 |
| Gastroesophageal reflux disease | 16.4 | 16.0 | 0.01 | 16.4 | 17.2 | -0.02 | 16.0 | 16.5 | -0.01 |
| Gastrointestinal hemorrhage | 2.1 | 2.0 | 0.01 | 2.0 | 2.0 | 0.00 | 2.0 | 2.0 | 0.01 |
| Human immunodeficiency virus infection | 0.6 | 0.5 | 0.01 | 0.6 | 0.7 | -0.01 | 0.5 | 0.6 | 0.00 |
| Hyperlipidemia | 79.8 | 81.8 | -0.05 | 79.4 | 79.4 | 0.00 | 81.1 | 81.0 | 0.00 |
| Hypertensive disorder | 79.1 | 78.8 | 0.01 | 78.8 | 79.1 | -0.01 | 79.3 | 79.0 | 0.01 |
| Lesion of liver | 1.0 | 1.2 | -0.02 | 1.0 | 1.3 | -0.02 | 0.9 | 1.2 | -0.03 |
| Obesity | 41.6 | 28.9 | 0.27 | 38.9 | 41.5 | -0.05 | 31.9 | 33.7 | -0.04 |
| Osteoarthritis | 25.3 | 22.2 | 0.07 | 24.3 | 24.9 | -0.01 | 23.1 | 23.2 | 0.00 |
| Pneumonia | 3.2 | 3.0 | 0.01 | 3.1 | 3.2 | 0.00 | 3.1 | 3.1 | 0.00 |
| Psoriasis | 2.2 | 2.0 | 0.01 | 2.1 | 2.0 | 0.01 | 2.1 | 2.0 | 0.01 |
| Renal impairment | 9.9 | 10.3 | -0.01 | 10.1 | 10.1 | 0.00 | 10.8 | 10.3 | 0.02 |
| Rheumatoid arthritis | 0.9 | 0.9 | 0.00 | 0.8 | 0.9 | -0.01 | 0.8 | 0.9 | -0.01 |
| Schizophrenia | 0.4 | 0.3 | 0.03 | 0.4 | 0.4 | 0.01 | 0.4 | 0.3 | 0.02 |
| Ulcerative colitis | 0.3 | 0.4 | -0.02 | 0.3 | 0.4 | -0.03 | 0.3 | 0.4 | -0.01 |
| Urinary tract infectious disease | 4.3 | 3.4 | 0.05 | 3.9 | 3.7 | 0.01 | 3.9 | 3.5 | 0.02 |
| Viral hepatitis C | 0.6 | 0.5 | 0.01 | 0.6 | 0.5 | 0.01 | 0.7 | 0.5 | 0.02 |
| Visual system disorder | 31.6 | 33.7 | -0.04 | 32.1 | 32.0 | 0.00 | 33.5 | 33.1 | 0.01 |
| **Medical history: Cardiovascular disease** | | | | | | | | | |
| Atrial fibrillation | 6.1 | 7.4 | -0.05 | 6.4 | 6.0 | 0.01 | 7.1 | 7.0 | 0.00 |
| Cerebrovascular disease | 4.0 | 5.4 | -0.07 | 4.1 | 4.3 | -0.01 | 5.0 | 5.1 | 0.00 |
| Coronary arteriosclerosis | 15.5 | 21.5 | -0.16 | 16.0 | 15.6 | 0.01 | 20.4 | 19.6 | 0.02 |
| Heart disease | 28.3 | 33.4 | -0.11 | 28.6 | 28.3 | 0.01 | 32.7 | 31.8 | 0.02 |
| Heart failure | 5.9 | 8.2 | -0.09 | 6.0 | 6.1 | -0.01 | 7.6 | 7.5 | 0.00 |
| Ischemic heart disease | 7.8 | 11.2 | -0.12 | 8.2 | 8.1 | 0.00 | 8.2 | 8.1 | 0.00 |
| Peripheral vascular disease | 9.9 | 10.8 | -0.03 | 10.4 | 10.1 | 0.01 | 10.3 | 10.6 | -0.01 |
| Pulmonary embolism | 0.8 | 0.7 | 0.01 | 0.8 | 0.7 | 0.00 | 0.7 | 0.7 | -0.01 |
| Venous thrombosis | 1.4 | 1.5 | -0.01 | 1.2 | 1.4 | -0.01 | 1.3 | 1.5 | -0.02 |
| **Medical history: Neoplasms** | | | | | | | | | |
| Hematologic neoplasm | 0.8 | 0.9 | -0.02 | 0.6 | 0.9 | -0.03 | 0.7 | 0.9 | -0.02 |
| Malignant lymphoma | 0.4 | 0.5 | -0.01 | 0.4 | 0.5 | -0.02 | 0.4 | 0.5 | -0.02 |
| Malignant neoplasm of anorectum | 0.2 | 0.2 | 0.00 | 0.1 | 0.2 | -0.01 | 0.1 | 0.2 | -0.02 |
| Malignant neoplastic disease | 7.9 | 8.7 | -0.03 | 8.1 | 8.3 | -0.01 | 8.1 | 8.3 | -0.01 |
| Malignant tumor of colon | 0.5 | 0.5 | 0.00 | 0.4 | 0.4 | 0.00 | 0.4 | 0.5 | 0.00 |
| Malignant tumor of lung | 0.2 | 0.2 | 0.01 | 0.3 | 0.1 | 0.03 | 0.4 | 0.2 | 0.04 |

| (Continued) | Before ajustment | | | After matching | | | After stratification | | |
|---|---|---|---|---|---|---|---|---|---|
| Characteristic | T (%) | C (%) | StdDiff | T (%) | C (%) | StdDiff | T (%) | C (%) | StdDiff |
| Malignant tumor of urinary bladder | 0.4 | 0.4 | -0.01 | 0.4 | 0.4 | 0.01 | 0.4 | 0.4 | 0.01 |
| Primary malignant neoplasm of prostate | 2.6 | 3.0 | -0.02 | 2.8 | 2.8 | 0.00 | 3.0 | 2.9 | 0.01 |
| **Medication use** | | | | | | | | | |
| Agents acting on the renin-angiotensin system | 72.9 | 73.1 | 0.00 | 72.8 | 73.0 | 0.00 | 73.3 | 73.1 | 0.00 |
| Antibacterials for systemic use | 53.5 | 49.7 | 0.08 | 51.9 | 52.3 | -0.01 | 50.8 | 50.8 | 0.00 |
| Antidepressants | 25.9 | 19.9 | 0.14 | 24.4 | 25.4 | -0.02 | 21.6 | 22.0 | -0.01 |
| Antiepileptics | 16.6 | 13.8 | 0.08 | 16.2 | 16.9 | -0.02 | 15.0 | 15.0 | 0.00 |
| Antiinflammatory and antirheumatic products | 30.9 | 27.6 | 0.07 | 30.0 | 30.8 | -0.02 | 28.7 | 28.9 | 0.00 |
| Antineoplastic agents | 4.7 | 4.2 | 0.02 | 4.5 | 4.5 | 0.00 | 4.5 | 4.4 | 0.01 |
| Antipsoriatics | 0.6 | 0.4 | 0.02 | 0.5 | 0.5 | 0.01 | 0.5 | 0.5 | 0.01 |
| Antithrombotic agents | 15.7 | 19.5 | -0.10 | 16.1 | 16.1 | 0.00 | 18.6 | 18.4 | 0.00 |
| Beta blocking agents | 31.3 | 35.3 | -0.08 | 31.6 | 31.1 | 0.01 | 34.1 | 33.9 | 0.00 |
| Calcium channel blockers | 25.6 | 25.8 | 0.00 | 25.8 | 26.5 | -0.01 | 25.3 | 26.0 | -0.01 |
| Diuretics | 40.5 | 36.9 | 0.07 | 39.1 | 39.5 | -0.01 | 37.3 | 37.9 | -0.01 |
| Drugs for acid related disorders | 22.2 | 22.2 | 0.00 | 22.3 | 22.7 | -0.01 | 22.3 | 22.7 | -0.01 |
| Drugs for obstructive airway diseases | 36.5 | 33.2 | 0.07 | 35.3 | 35.9 | -0.01 | 34.7 | 34.3 | 0.01 |
| Immunosuppressants | 2.0 | 1.8 | 0.02 | 2.0 | 2.0 | 0.00 | 1.9 | 1.9 | 0.00 |
| Lipid modifying agents | 74.7 | 78.4 | -0.09 | 75.5 | 75.2 | 0.01 | 77.8 | 77.3 | 0.01 |
| Opioids | 24.9 | 23.9 | 0.02 | 24.0 | 25.2 | -0.03 | 24.0 | 24.5 | -0.01 |
| Psycholeptics | 22.0 | 19.8 | 0.05 | 21.0 | 21.3 | 0.00 | 20.2 | 20.4 | -0.01 |
| Psychostimulants | 3.1 | 1.9 | 0.08 | 2.4 | 2.2 | 0.01 | 2.4 | 2.2 | 0.01 |

##### 2.1.3 MDCR

**Supplementary Table 159:** Baseline patient characteristics for GLP1RA (male) (T) and SGLT2I (male) (C) new-users in the MDCR data source. We report proportion of initiators satisfying selected base-line characteristics and the standardized difference of population proportions (StdDiff) before and after propensity score adjustment. Less extreme StdDiffs through matching and stratification suggest improved balance between patient cohorts through adjustment.

| | Before adjustment | | | After matching | | | After stratification | | |
|---|---|---|---|---|---|---|---|---|---|
| Characteristic | T (%) | C (%) | StdDiff | T (%) | C (%) | StdDiff | T (%) | C (%) | StdDiff |
| **Age group** | | | | | | | | | |
| 45 - 49 | <0.5 | <0.2 | 0.03 | <0.6 | <0.2 | -0.01 | <0.6 | <0.2 | -0.01 |
| 50 - 54 | 0.6 | 0.3 | 0.04 | 0.7 | 0.4 | 0.04 | <0.6 | 0.4 | 0.03 |
| 55 - 59 | 1.5 | 1.0 | 0.05 | 1.4 | 1.3 | 0.01 | 1.1 | 1.1 | 0.00 |
| 60 - 64 | 2.8 | 2.4 | 0.02 | 2.1 | 3.0 | -0.05 | 2.2 | 2.6 | -0.03 |
| 65 - 69 | 40.4 | 36.1 | 0.09 | 37.9 | 37.8 | 0.00 | 37.1 | 36.6 | 0.01 |
| 70 - 74 | 35.1 | 35.8 | -0.01 | 36.4 | 35.7 | 0.01 | 35.5 | 35.8 | -0.01 |
| 75 - 79 | 12.9 | 16.8 | -0.11 | 13.1 | 15.2 | -0.06 | 14.1 | 16.3 | -0.06 |
| 80 - 84 | 4.9 | 5.6 | -0.03 | 6.0 | 4.8 | 0.05 | 6.4 | 5.3 | 0.05 |
| 85 - 89 | 1.3 | 1.6 | -0.03 | 1.7 | 1.4 | 0.03 | 2.3 | 1.6 | 0.05 |
| 90 - 94 | <0.5 | 0.3 | 0.02 | <0.7 | <0.2 | 0.06 | 0.7 | 0.3 | 0.06 |
| **Medical history: General** | | | | | | | | | |
| Acute respiratory disease | 21.6 | 21.2 | 0.01 | 21.3 | 22.8 | -0.04 | 19.7 | 21.6 | -0.05 |
| Attention deficit hyperactivity disorder | <0.5 | 0.2 | -0.01 | <0.7 | 0.2 | -0.02 | <0.6 | <0.2 | -0.04 |
| Chronic liver disease | 1.7 | 1.8 | -0.01 | 1.6 | 2.1 | -0.04 | 1.7 | 1.8 | -0.01 |
| Chronic obstructive lung disease | 13.0 | 10.2 | 0.09 | 13.0 | 11.5 | 0.05 | 12.7 | 10.6 | 0.07 |
| Crohn's disease | <0.5 | 0.4 | -0.04 | <0.7 | 0.4 | -0.05 | <0.6 | 0.4 | -0.02 |
| Dementia | 1.4 | 1.1 | 0.02 | 1.7 | 1.3 | 0.03 | 1.7 | 1.2 | 0.04 |
| Depressive disorder | 8.8 | 6.5 | 0.09 | 10.0 | 9.2 | 0.03 | 8.2 | 7.2 | 0.04 |
| Gastroesophageal reflux disease | 11.9 | 15.2 | -0.10 | 14.1 | 17.9 | -0.10 | 13.1 | 16.0 | -0.08 |
| Gastrointestinal hemorrhage | 2.0 | 2.6 | -0.04 | 1.6 | 2.6 | -0.07 | 1.4 | 2.6 | -0.09 |
| Human immunodeficiency virus infection | <0.5 | <0.2 | -0.02 | <0.7 | <0.2 | -0.01 | <0.6 | <0.2 | -0.01 |
| Hyperlipidemia | 70.7 | 83.0 | -0.29 | 83.3 | 84.4 | -0.03 | 82.6 | 83.3 | -0.02 |
| Hypertensive disorder | 80.2 | 84.2 | -0.11 | 85.6 | 85.1 | 0.01 | 84.8 | 84.5 | 0.01 |
| Lesion of liver | 0.9 | 1.3 | -0.04 | 0.9 | 1.2 | -0.04 | 1.1 | 1.3 | -0.02 |
| Obesity | 27.7 | 22.9 | 0.11 | 33.2 | 36.8 | -0.07 | 24.1 | 27.0 | -0.07 |
| Osteoarthritis | 33.3 | 29.4 | 0.08 | 35.2 | 34.2 | 0.02 | 31.4 | 30.8 | 0.01 |
| Pneumonia | 4.1 | 4.0 | 0.00 | 3.9 | 4.5 | -0.03 | 3.8 | 4.1 | -0.02 |
| Psoriasis | 3.3 | 2.1 | 0.07 | 3.6 | 1.8 | 0.11 | 3.6 | 2.0 | 0.10 |
| Renal impairment | 11.1 | 13.1 | -0.06 | 13.1 | 14.4 | -0.04 | 12.9 | 13.5 | -0.02 |
| Rheumatoid arthritis | 1.2 | 1.2 | 0.00 | 1.1 | 1.6 | -0.04 | 0.8 | 1.3 | -0.04 |
| Schizophrenia | <0.5 | <0.2 | 0.03 | <0.7 | <0.2 | 0.02 | <0.6 | <0.2 | 0.02 |
| Ulcerative colitis | <0.5 | 0.3 | -0.03 | <0.7 | 0.4 | -0.01 | <0.6 | 0.3 | 0.01 |
| Urinary tract infectious disease | 4.9 | 4.1 | 0.04 | 5.8 | 4.8 | 0.05 | 5.5 | 4.3 | 0.06 |
| Viral hepatitis C | <0.5 | 0.7 | -0.06 | <0.7 | 0.8 | -0.05 | <0.6 | 0.7 | -0.05 |
| Visual system disorder | 49.9 | 52.9 | -0.06 | 52.4 | 53.7 | -0.03 | 52.8 | 53.1 | -0.01 |
| **Medical history: Cardiovascular disease** | | | | | | | | | |
| Atrial fibrillation | 11.4 | 15.0 | -0.11 | 11.7 | 14.4 | -0.08 | 12.5 | 14.9 | -0.07 |
| Cerebrovascular disease | 10.1 | 11.2 | -0.04 | 9.7 | 10.5 | -0.03 | 10.3 | 11.0 | -0.02 |
| Coronary arteriosclerosis | 30.3 | 36.4 | -0.13 | 30.8 | 31.1 | -0.01 | 34.6 | 35.0 | -0.01 |
| Heart disease | 47.7 | 54.2 | -0.13 | 47.5 | 49.3 | -0.04 | 52.2 | 52.9 | -0.01 |
| Heart failure | 8.1 | 10.6 | -0.09 | 8.7 | 10.3 | -0.06 | 9.4 | 10.5 | -0.04 |
| Ischemic heart disease | 11.4 | 13.8 | -0.07 | 11.1 | 12.6 | -0.04 | 11.3 | 13.5 | -0.07 |
| Peripheral vascular disease | 12.5 | 15.0 | -0.07 | 12.3 | 15.5 | -0.09 | 12.8 | 15.2 | -0.07 |
| Pulmonary embolism | 1.3 | 0.9 | 0.03 | 1.1 | 1.3 | -0.01 | 0.8 | 1.0 | -0.02 |
| Venous thrombosis | 1.7 | 1.9 | -0.01 | 1.6 | 2.0 | -0.03 | 1.2 | 1.9 | -0.06 |
| **Medical history: Neoplasms** | | | | | | | | | |
| Hematologic neoplasm | 2.0 | 2.0 | 0.00 | 2.4 | 2.0 | 0.03 | 1.7 | 2.1 | -0.03 |
| Malignant lymphoma | 1.2 | 1.4 | -0.02 | 1.6 | 1.4 | 0.01 | 1.1 | 1.5 | -0.04 |
| Malignant neoplastic disease | 21.4 | 18.4 | 0.07 | 22.0 | 19.3 | 0.06 | 21.7 | 18.8 | 0.07 |
| Malignant tumor of breast | <0.5 | <0.2 | 0.04 | <0.7 | <0.2 | 0.03 | <0.6 | <0.2 | 0.04 |
| Malignant tumor of colon | <0.5 | 0.5 | -0.02 | <0.7 | 0.5 | 0.01 | <0.6 | 0.5 | -0.01 |
| Malignant tumor of lung | 0.8 | 0.3 | 0.06 | 1.0 | 0.5 | 0.06 | 1.1 | 0.4 | 0.08 |
| Malignant tumor of urinary bladder | 1.9 | 1.4 | 0.04 | 1.9 | 1.6 | 0.02 | 2.0 | 1.5 | 0.04 |
| Primary malignant neoplasm of prostate | 8.4 | 7.1 | 0.05 | 8.4 | 7.6 | 0.03 | 8.9 | 7.3 | 0.06 |
| **Medication use** | | | | | | | | | |
| Agents acting on the renin-angiotensin system | 77.4 | 77.6 | 0.00 | 76.2 | 77.6 | -0.03 | 76.1 | 77.5 | -0.03 |
| Antibacterials for systemic use | 59.4 | 59.8 | -0.01 | 59.8 | 62.5 | -0.06 | 57.6 | 60.6 | -0.06 |

| (Continued) | Before ajustment | | | After matching | | | After stratification | | |
|---|---|---|---|---|---|---|---|---|---|
| Characteristic | T (%) | C (%) | StdDiff | T (%) | C (%) | StdDiff | T (%) | C (%) | StdDiff |
| Antidepressants | 27.0 | 18.4 | 0.21 | 25.4 | 26.8 | -0.03 | 20.0 | 20.8 | -0.02 |
| Antiepileptics | 17.1 | 14.3 | 0.08 | 17.7 | 18.0 | -0.01 | 15.3 | 15.4 | 0.00 |
| Antiinflammatory and antirheumatic products | 28.3 | 26.0 | 0.05 | 27.2 | 28.6 | -0.03 | 24.3 | 26.8 | -0.06 |
| Antineoplastic agents | 8.7 | 7.7 | 0.04 | 8.3 | 8.1 | 0.01 | 8.5 | 7.8 | 0.03 |
| Antipsoriatics | 0.9 | 0.5 | 0.04 | <0.7 | 0.4 | 0.03 | 0.6 | 0.5 | 0.02 |
| Antithrombotic agents | 27.8 | 30.2 | -0.05 | 26.7 | 28.1 | -0.03 | 29.0 | 29.7 | -0.01 |
| Beta blocking agents | 48.6 | 51.1 | -0.05 | 49.2 | 48.5 | 0.01 | 51.4 | 50.4 | 0.02 |
| Calcium channel blockers | 32.7 | 32.9 | 0.00 | 31.4 | 33.5 | -0.04 | 31.4 | 33.0 | -0.04 |
| Diuretics | 50.7 | 45.1 | 0.11 | 47.6 | 48.0 | -0.01 | 45.8 | 46.0 | 0.00 |
| Drugs for acid related disorders | 31.2 | 30.5 | 0.01 | 31.5 | 32.8 | -0.03 | 29.5 | 31.2 | -0.04 |
| Drugs for obstructive airway diseases | 45.1 | 41.1 | 0.08 | 45.4 | 43.5 | 0.04 | 43.2 | 41.8 | 0.03 |
| Immunosuppressants | 3.5 | 2.7 | 0.05 | 3.9 | 3.5 | 0.02 | 3.9 | 2.9 | 0.06 |
| Lipid modifying agents | 85.0 | 85.8 | -0.02 | 85.4 | 84.2 | 0.03 | 85.1 | 85.3 | -0.01 |
| Opioids | 27.3 | 24.1 | 0.07 | 26.4 | 27.5 | -0.03 | 24.8 | 25.2 | -0.01 |
| Psycholeptics | 23.0 | 20.5 | 0.06 | 22.3 | 23.3 | -0.02 | 19.6 | 21.3 | -0.04 |
| Psychostimulants | 1.5 | 1.3 | 0.01 | 1.7 | 2.0 | -0.02 | 1.2 | 1.5 | -0.02 |

##### 2.1.4 Open Claims

**Supplementary Table 160:** Baseline patient characteristics for GLP1RA (male) (T) and SGLT2I (male) (C) new-users in the Open Claims data source. We report proportion of initiators satisfying selected base-line characteristics and the standardized difference of population proportions (StdDiff) before and after propensity score adjustment. Less extreme StdDiffs through matching and stratification suggest improved balance between patient cohorts through adjustment.

| Characteristic | Before adjustment | | | After matching | | | After stratification | | |
|---|---|---|---|---|---|---|---|---|---|
| | T (%) | C (%) | StdDiff | T (%) | C (%) | StdDiff | T (%) | C (%) | StdDiff |
| **Age group** | | | | | | | | | |
| 15 - 19 | 0.2 | 0.1 | 0.03 | 0.2 | 0.1 | 0.02 | 0.1 | 0.1 | 0.02 |
| 20 - 24 | 0.5 | 0.3 | 0.04 | 0.4 | 0.4 | 0.00 | 0.4 | 0.3 | 0.01 |
| 25 - 29 | 1.0 | 0.6 | 0.05 | 0.9 | 0.9 | 0.00 | 0.7 | 0.7 | 0.01 |
| 30 - 34 | 2.3 | 1.5 | 0.06 | 2.1 | 2.1 | 0.00 | 1.7 | 1.7 | 0.00 |
| 35 - 39 | 4.7 | 3.4 | 0.07 | 4.4 | 4.4 | 0.00 | 3.7 | 3.8 | 0.00 |
| 45 - 49 | 11.8 | 10.0 | 0.06 | 11.6 | 11.5 | 0.00 | 10.5 | 10.6 | 0.00 |
| 50 - 54 | 15.1 | 13.9 | 0.03 | 15.0 | 14.9 | 0.00 | 14.2 | 14.2 | 0.00 |
| 55 - 59 | 16.8 | 16.5 | 0.01 | 16.9 | 16.9 | 0.00 | 16.5 | 16.7 | 0.00 |
| 60 - 64 | 15.5 | 17.1 | -0.04 | 16.0 | 15.8 | 0.00 | 16.7 | 16.6 | 0.00 |
| 65 - 69 | 11.6 | 13.3 | -0.05 | 12.0 | 12.0 | 0.00 | 12.8 | 12.7 | 0.00 |
| 70 - 74 | 7.1 | 8.9 | -0.07 | 7.4 | 7.6 | -0.01 | 8.3 | 8.4 | 0.00 |
| 75 - 79 | 3.6 | 5.4 | -0.09 | 4.8 | 4.9 | 0.00 | 4.8 | 4.9 | 0.00 |
| 80 - 84 | 1.6 | 2.9 | -0.09 | 1.7 | 1.7 | 0.00 | 2.5 | 2.4 | 0.00 |
| 85 - 89 | 0.1 | 0.2 | -0.03 | 0.1 | 0.2 | -0.01 | 0.1 | 0.2 | -0.02 |
| **Medical history: General** | | | | | | | | | |
| Acute respiratory disease | 11.3 | 11.1 | 0.01 | 10.9 | 11.1 | -0.01 | 11.1 | 11.2 | 0.00 |
| Attention deficit hyperactivity disorder | 1.0 | 0.6 | 0.05 | 0.8 | 0.9 | -0.01 | 0.7 | 0.7 | 0.00 |
| Chronic liver disease | 0.8 | 0.9 | -0.02 | 0.8 | 0.8 | 0.00 | 0.9 | 0.9 | 0.00 |
| Chronic obstructive lung disease | 3.9 | 4.2 | -0.02 | 3.8 | 3.9 | 0.00 | 4.2 | 4.2 | 0.00 |
| Crohn’s disease | 0.1 | 0.1 | 0.00 | 0.1 | 0.1 | -0.01 | 0.1 | 0.1 | 0.00 |
| Dementia | 0.3 | 0.4 | -0.01 | 0.3 | 0.3 | 0.00 | 0.3 | 0.3 | 0.00 |
| Depressive disorder | 6.6 | 4.7 | 0.08 | 5.8 | 6.1 | -0.01 | 5.3 | 5.3 | 0.00 |
| Gastroesophageal reflux disease | 7.5 | 6.9 | 0.02 | 7.1 | 7.4 | -0.01 | 7.1 | 7.1 | 0.00 |
| Gastrointestinal hemorrhage | 1.1 | 1.1 | 0.00 | 1.0 | 1.1 | -0.01 | 1.1 | 1.2 | 0.00 |
| Human immunodeficiency virus infection | 0.5 | 0.3 | 0.02 | 0.5 | 0.5 | 0.00 | 0.4 | 0.4 | 0.00 |
| Hyperlipidemia | 42.8 | 46.0 | -0.06 | 42.6 | 43.0 | -0.01 | 44.8 | 45.1 | 0.00 |
| Hypertensive disorder | 49.1 | 49.8 | -0.01 | 48.3 | 48.7 | -0.01 | 49.7 | 49.7 | 0.00 |
| Lesion of liver | 0.6 | 0.6 | 0.00 | 0.6 | 0.6 | 0.00 | 0.6 | 0.6 | -0.01 |
| Obesity | 18.0 | 11.4 | 0.19 | 15.5 | 15.7 | -0.01 | 13.8 | 13.2 | 0.02 |
| Osteoarthritis | 13.3 | 11.8 | 0.05 | 12.7 | 12.7 | 0.00 | 12.5 | 12.2 | 0.01 |
| Pneumonia | 2.0 | 2.0 | 0.00 | 1.9 | 2.0 | 0.00 | 2.0 | 2.0 | 0.00 |
| Psoriasis | 1.3 | 1.0 | 0.03 | 1.2 | 1.1 | 0.00 | 1.2 | 1.1 | 0.01 |
| Renal impairment | 4.7 | 5.3 | -0.03 | 4.7 | 4.8 | -0.01 | 5.3 | 5.2 | 0.00 |
| Rheumatoid arthritis | 0.6 | 0.5 | 0.00 | 0.5 | 0.6 | 0.00 | 0.5 | 0.6 | 0.00 |
| Schizophrenia | 0.3 | 0.3 | 0.01 | 0.3 | 0.3 | 0.00 | 0.3 | 0.3 | 0.00 |
| Ulcerative colitis | 0.2 | 0.2 | 0.00 | 0.2 | 0.2 | 0.00 | 0.2 | 0.2 | 0.00 |
| Urinary tract infectious disease | 1.9 | 1.8 | 0.01 | 1.9 | 1.8 | 0.00 | 1.9 | 1.8 | 0.00 |
| Viral hepatitis C | 0.3 | 0.3 | -0.01 | 0.3 | 0.3 | 0.00 | 0.3 | 0.3 | 0.01 |
| Visual system disorder | 14.5 | 15.5 | -0.03 | 14.5 | 14.6 | 0.00 | 15.2 | 15.2 | 0.00 |
| **Medical history: Cardiovascular disease** | | | | | | | | | |
| Atrial fibrillation | 3.6 | 4.6 | -0.05 | 3.6 | 3.6 | 0.00 | 4.2 | 4.3 | 0.00 |
| Cerebrovascular disease | 1.9 | 2.7 | -0.05 | 2.0 | 2.1 | -0.01 | 2.4 | 2.5 | -0.01 |
| Coronary arteriosclerosis | 8.2 | 11.9 | -0.13 | 8.4 | 8.6 | -0.01 | 10.6 | 10.9 | -0.01 |
| Heart disease | 16.3 | 21.1 | -0.12 | 16.5 | 16.7 | -0.01 | 19.4 | 19.8 | -0.01 |
| Heart failure | 2.9 | 5.2 | -0.12 | 3.0 | 3.2 | -0.01 | 4.1 | 4.6 | -0.03 |
| Ischemic heart disease | 3.5 | 5.5 | -0.10 | 3.6 | 3.8 | -0.01 | 4.7 | 5.0 | -0.01 |
| Peripheral vascular disease | 4.2 | 4.7 | -0.02 | 4.1 | 4.2 | -0.01 | 4.5 | 4.6 | 0.00 |
| Pulmonary embolism | 0.5 | 0.4 | 0.01 | 0.5 | 0.4 | 0.00 | 0.5 | 0.4 | 0.00 |
| Venous thrombosis | 0.7 | 0.8 | -0.01 | 0.7 | 0.7 | 0.00 | 0.7 | 0.8 | 0.00 |
| **Medical history: Neoplasms** | | | | | | | | | |
| Hematologic neoplasm | 0.5 | 0.6 | 0.00 | 0.5 | 0.5 | 0.00 | 0.6 | 0.6 | 0.00 |
| Malignant lymphoma | 0.3 | 0.4 | -0.01 | 0.3 | 0.3 | 0.00 | 0.3 | 0.4 | 0.00 |
| Malignant neoplasm of anorectum | 0.1 | 0.1 | -0.01 | 0.1 | 0.1 | -0.01 | 0.1 | 0.1 | -0.01 |
| Malignant neoplastic disease | 4.4 | 4.9 | -0.03 | 4.4 | 4.5 | -0.01 | 4.8 | 4.7 | 0.00 |
| Malignant tumor of colon | 0.2 | 0.3 | -0.01 | 0.2 | 0.2 | 0.00 | 0.2 | 0.2 | 0.00 |
| Malignant tumor of lung | 0.1 | 0.1 | -0.01 | 0.1 | 0.1 | 0.00 | 0.1 | 0.1 | 0.00 |
| Malignant tumor of urinary bladder | 0.3 | 0.3 | 0.00 | 0.3 | 0.2 | 0.00 | 0.3 | 0.3 | 0.01 |

| (Continued) | Before ajustment | | | After matching | | | After stratification | | |
|---|---|---|---|---|---|---|---|---|---|
| Characteristic | T (%) | C (%) | StdDiff | T (%) | C (%) | StdDiff | T (%) | C (%) | StdDiff |
| Primary malignant neoplasm of prostate | 1.5 | 1.7 | -0.01 | 1.5 | 1.6 | 0.00 | 1.7 | 1.6 | 0.00 |
| **Medication use** | | | | | | | | | |
| Agents acting on the renin-angiotensin system | 69.0 | 71.6 | -0.06 | 69.3 | 69.6 | -0.01 | 70.8 | 70.9 | 0.00 |
| Antibacterials for systemic use | 50.5 | 48.9 | 0.03 | 49.8 | 49.7 | 0.00 | 49.7 | 49.5 | 0.00 |
| Antidepressants | 27.4 | 21.1 | 0.15 | 25.7 | 26.2 | -0.01 | 23.3 | 23.2 | 0.00 |
| Antiepileptics | 17.7 | 14.7 | 0.08 | 16.8 | 17.0 | 0.00 | 16.0 | 15.6 | 0.01 |
| Antiinflammatory and antirheumatic products | 33.2 | 32.1 | 0.03 | 32.7 | 32.5 | 0.00 | 32.6 | 32.3 | 0.00 |
| Antineoplastic agents | 4.5 | 4.0 | 0.02 | 4.3 | 4.3 | 0.00 | 4.2 | 4.1 | 0.00 |
| Antipsoriatics | 0.6 | 0.5 | 0.02 | 0.6 | 0.5 | 0.01 | 0.6 | 0.5 | 0.00 |
| Antithrombotic agents | 16.8 | 21.2 | -0.11 | 17.1 | 17.3 | 0.00 | 19.7 | 20.0 | -0.01 |
| Beta blocking agents | 30.3 | 35.0 | -0.10 | 30.6 | 30.8 | -0.01 | 33.6 | 33.7 | 0.00 |
| Calcium channel blockers | 25.6 | 25.8 | 0.00 | 25.6 | 25.7 | 0.00 | 25.8 | 25.9 | 0.00 |
| Diuretics | 37.3 | 36.2 | 0.02 | 36.7 | 37.2 | -0.01 | 36.5 | 36.7 | 0.00 |
| Drugs for acid related disorders | 28.3 | 27.4 | 0.02 | 27.8 | 28.2 | -0.01 | 27.8 | 28.2 | -0.01 |
| Drugs for obstructive airway diseases | 34.4 | 31.6 | 0.06 | 33.5 | 33.6 | 0.00 | 32.7 | 32.4 | 0.01 |
| Immunosuppressants | 2.2 | 1.9 | 0.03 | 2.2 | 2.1 | 0.00 | 2.1 | 2.0 | 0.00 |
| Lipid modifying agents | 71.1 | 75.3 | -0.10 | 72.0 | 72.2 | 0.00 | 74.1 | 74.1 | 0.00 |
| Opioids | 21.7 | 20.7 | 0.03 | 21.1 | 21.3 | 0.00 | 21.3 | 21.1 | 0.00 |
| Psycholeptics | 20.0 | 17.4 | 0.06 | 19.0 | 19.4 | -0.01 | 18.3 | 18.3 | 0.00 |
| Psychostimulants | 3.7 | 2.3 | 0.09 | 3.2 | 3.3 | -0.01 | 3.2 | 3.3 | -0.01 |

##### 2.1.5 OEHR

**Supplementary Table 161:** Baseline patient characteristics for GLP1RA (male) (T) and SGLT2I (male) (C) new-users in the OEHR data source. We report proportion of initiators satisfying selected base-line characteristics and the standardized difference of population proportions (StdDiff) before and after propensity score adjustment. Less extreme StdDiffs through matching and stratification suggest improved balance between patient cohorts through adjustment.

| | Before adjustment | | | After matching | | | After stratification | | |
|---|---|---|---|---|---|---|---|---|---|
| Characteristic | T (%) | C (%) | StdDiff | T (%) | C (%) | StdDiff | T (%) | C (%) | StdDiff |
| **Age group** | | | | | | | | | |
| 15 - 19 | 0.1 | 0.1 | 0.02 | 0.1 | 0.1 | 0.01 | 0.1 | 0.1 | 0.01 |
| 20 - 24 | 0.5 | 0.2 | 0.04 | 0.5 | 0.3 | 0.03 | 0.4 | 0.2 | 0.03 |
| 25 - 29 | 1.2 | 0.7 | 0.05 | 1.0 | 0.9 | 0.02 | 1.0 | 0.7 | 0.03 |
| 30 - 34 | 2.8 | 1.9 | 0.06 | 2.5 | 2.5 | 0.00 | 2.3 | 2.1 | 0.01 |
| 35 - 39 | 5.7 | 4.3 | 0.07 | 5.4 | 5.3 | 0.00 | 4.9 | 4.7 | 0.01 |
| 40 - 44 | 9.1 | 8.0 | 0.04 | 9.0 | 8.8 | 0.00 | 8.7 | 8.3 | 0.01 |
| 45 - 49 | 13.3 | 11.9 | 0.04 | 13.2 | 13.2 | 0.00 | 12.5 | 12.4 | 0.00 |
| 50 - 54 | 16.7 | 15.7 | 0.03 | 16.3 | 15.8 | 0.01 | 15.5 | 15.7 | -0.01 |
| 55 - 59 | 17.5 | 18.1 | -0.02 | 17.4 | 17.8 | -0.01 | 17.8 | 18.0 | 0.00 |
| 60 - 64 | 15.7 | 17.3 | -0.04 | 16.5 | 16.3 | 0.00 | 16.8 | 16.9 | 0.00 |
| 65 - 69 | 9.4 | 10.5 | -0.03 | 9.8 | 10.0 | -0.01 | 9.8 | 10.0 | -0.01 |
| 70 - 74 | 5.0 | 6.5 | -0.06 | 5.1 | 5.2 | 0.00 | 5.8 | 6.0 | -0.01 |
| 75 - 79 | 2.2 | 3.2 | -0.06 | 2.4 | 2.5 | -0.01 | 2.9 | 3.0 | 0.00 |
| 80 - 84 | 0.6 | 1.4 | -0.08 | 0.7 | 0.9 | -0.02 | 1.0 | 1.2 | -0.03 |
| 85 - 89 | 0.2 | 0.4 | -0.04 | 0.2 | 0.4 | -0.03 | 0.2 | 0.3 | -0.03 |
| **Race** | | | | | | | | | |
| Asian | 1.6 | 2.3 | -0.05 | 1.7 | 1.9 | -0.01 | 1.8 | 2.2 | -0.03 |
| Black or African American | 7.4 | 8.2 | -0.03 | 7.7 | 7.8 | -0.01 | 7.8 | 8.1 | -0.01 |
| White | 83.4 | 81.8 | 0.04 | 83.0 | 83.1 | 0.00 | 82.5 | 82.2 | 0.01 |
| **Ethnicity** | | | | | | | | | |
| Hispanic or Latino | 5.0 | 5.2 | -0.01 | 5.0 | 5.1 | 0.00 | 5.3 | 5.2 | 0.00 |
| Not Hispanic or Latino | 85.8 | 85.6 | 0.01 | 85.7 | 85.8 | 0.00 | 86.0 | 85.7 | 0.01 |
| **Medical history: General** | | | | | | | | | |
| Acute respiratory disease | 15.6 | 15.0 | 0.02 | 15.3 | 15.1 | 0.00 | 15.0 | 15.2 | 0.00 |
| Attention deficit hyperactivity disorder | 1.4 | 1.0 | 0.04 | 1.3 | 1.4 | -0.01 | 1.1 | 1.2 | 0.00 |
| Chronic liver disease | 2.3 | 1.9 | 0.03 | 2.2 | 2.0 | 0.01 | 2.2 | 2.0 | 0.01 |
| Chronic obstructive lung disease | 4.3 | 4.3 | 0.00 | 4.1 | 4.5 | -0.02 | 4.3 | 4.4 | -0.01 |
| Crohn's disease | 0.2 | 0.2 | 0.00 | 0.2 | 0.2 | 0.00 | 0.2 | 0.2 | -0.01 |
| Dementia | 0.3 | 0.3 | -0.02 | 0.2 | 0.3 | -0.01 | 0.3 | 0.3 | -0.01 |
| Depressive disorder | 12.6 | 9.3 | 0.10 | 11.5 | 12.1 | -0.02 | 10.1 | 10.4 | -0.01 |
| Gastroesophageal reflux disease | 15.5 | 14.9 | 0.02 | 15.0 | 15.6 | -0.02 | 14.8 | 15.2 | -0.01 |
| Gastrointestinal hemorrhage | 1.4 | 1.2 | 0.03 | 1.4 | 1.2 | 0.01 | 1.4 | 1.2 | 0.01 |
| Human immunodeficiency virus infection | 0.4 | 0.3 | 0.02 | 0.3 | 0.4 | 0.00 | 0.3 | 0.3 | 0.00 |
| Hyperlipidemia | 67.8 | 69.9 | -0.04 | 67.4 | 66.6 | 0.02 | 69.2 | 68.9 | 0.01 |
| Hypertensive disorder | 68.4 | 68.7 | 0.00 | 68.9 | 68.6 | 0.00 | 68.9 | 68.6 | 0.00 |
| Lesion of liver | 0.7 | 0.7 | 0.01 | 0.8 | 0.8 | -0.01 | 0.7 | 0.7 | 0.00 |
| Obesity | 41.8 | 31.3 | 0.22 | 39.0 | 41.4 | -0.05 | 33.6 | 35.1 | -0.03 |
| Osteoarthritis | 15.5 | 13.4 | 0.06 | 14.7 | 14.8 | 0.00 | 14.7 | 14.8 | 0.00 |
| Pneumonia | 1.8 | 1.5 | 0.03 | 1.6 | 1.6 | 0.00 | 1.6 | 1.5 | 0.00 |
| Psoriasis | 1.7 | 1.4 | 0.03 | 1.7 | 1.7 | 0.00 | 1.7 | 1.5 | 0.02 |
| Renal impairment | 5.4 | 4.8 | 0.03 | 5.3 | 5.6 | -0.01 | 5.3 | 5.6 | -0.01 |
| Rheumatoid arthritis | 0.6 | 0.6 | 0.00 | 0.5 | 0.6 | -0.01 | 0.5 | 0.6 | -0.01 |
| Schizophrenia | 0.3 | 0.2 | 0.00 | 0.2 | 0.3 | -0.02 | 0.2 | 0.3 | -0.01 |
| Ulcerative colitis | 0.3 | 0.3 | 0.00 | 0.3 | 0.2 | 0.01 | 0.3 | 0.3 | 0.00 |
| Urinary tract infectious disease | 1.4 | 1.1 | 0.02 | 1.2 | 1.1 | 0.01 | 1.2 | 1.1 | 0.01 |
| Viral hepatitis C | 0.6 | 0.4 | 0.03 | 0.6 | 0.5 | 0.02 | 0.6 | 0.5 | 0.03 |
| Visual system disorder | 11.1 | 10.1 | 0.03 | 10.9 | 10.7 | 0.01 | 10.4 | 10.3 | 0.00 |
| **Medical history: Cardiovascular disease** | | | | | | | | | |
| Atrial fibrillation | 4.2 | 4.6 | -0.02 | 4.3 | 4.0 | 0.01 | 4.8 | 4.4 | 0.02 |
| Cerebrovascular disease | 2.1 | 2.6 | -0.04 | 2.1 | 2.1 | 0.00 | 2.3 | 2.5 | -0.01 |
| Coronary arteriosclerosis | 11.0 | 15.7 | -0.14 | 11.6 | 11.1 | 0.02 | 15.1 | 14.2 | 0.03 |
| Heart disease | 20.8 | 24.7 | -0.09 | 21.1 | 20.2 | 0.02 | 24.7 | 23.3 | 0.03 |
| Heart failure | 2.9 | 4.0 | -0.06 | 3.0 | 3.1 | 0.00 | 3.7 | 3.7 | 0.00 |
| Ischemic heart disease | 5.1 | 7.0 | -0.08 | 5.3 | 5.2 | 0.01 | 6.8 | 6.5 | 0.01 |
| Peripheral vascular disease | 5.1 | 5.8 | -0.03 | 5.2 | 5.3 | -0.01 | 5.4 | 5.7 | -0.01 |
| Pulmonary embolism | 0.5 | 0.4 | 0.02 | 0.5 | 0.5 | 0.00 | 0.4 | 0.4 | 0.00 |
| Venous thrombosis | 1.0 | 1.0 | 0.00 | 1.0 | 1.1 | -0.01 | 0.9 | 1.0 | -0.01 |

| (Continued) | Before ajustment | | | After matching | | | After stratification | | |
|---|---|---|---|---|---|---|---|---|---|
| Characteristic | T (%) | C (%) | StdDiff | T (%) | C (%) | StdDiff | T (%) | C (%) | StdDiff |
| **Medical history: Neoplasms** | | | | | | | | | |
| Hematologic neoplasm | 0.6 | 0.6 | 0.00 | 0.5 | 0.7 | -0.02 | 0.6 | 0.7 | -0.01 |
| Malignant lymphoma | 0.4 | 0.3 | 0.01 | 0.4 | 0.4 | 0.01 | 0.4 | 0.3 | 0.01 |
| Malignant neoplasm of anorectum | 0.1 | 0.1 | 0.00 | 0.1 | 0.1 | -0.01 | 0.1 | 0.1 | -0.01 |
| Malignant neoplastic disease | 4.8 | 4.7 | 0.00 | 4.8 | 4.7 | 0.01 | 4.9 | 4.7 | 0.01 |
| Malignant tumor of colon | 0.2 | 0.3 | -0.01 | 0.2 | 0.3 | -0.02 | 0.2 | 0.3 | -0.02 |
| Malignant tumor of lung | 0.1 | 0.1 | -0.01 | 0.1 | 0.1 | -0.01 | 0.1 | 0.1 | -0.01 |
| Malignant tumor of urinary bladder | 0.3 | 0.3 | 0.01 | 0.3 | 0.2 | 0.02 | 0.4 | 0.3 | 0.02 |
| Primary malignant neoplasm of prostate | 1.3 | 1.6 | -0.02 | 1.3 | 1.4 | -0.01 | 1.5 | 1.5 | 0.00 |
| **Medication use** | | | | | | | | | |
| Agents acting on the renin-angiotensin system | 69.3 | 69.5 | 0.00 | 69.2 | 69.4 | 0.00 | 70.1 | 69.5 | 0.01 |
| Antibacterials for systemic use | 38.4 | 36.4 | 0.04 | 37.6 | 38.0 | -0.01 | 36.8 | 37.1 | -0.01 |
| Antidepressants | 27.3 | 21.4 | 0.14 | 25.5 | 26.6 | -0.03 | 22.5 | 23.3 | -0.02 |
| Antiepileptics | 15.7 | 13.2 | 0.07 | 15.1 | 15.5 | -0.01 | 14.2 | 14.0 | 0.00 |
| Antiinflammatory and antirheumatic products | 54.5 | 54.6 | 0.00 | 53.9 | 54.1 | 0.00 | 54.4 | 54.6 | 0.00 |
| Antineoplastic agents | 3.9 | 3.0 | 0.05 | 3.5 | 3.5 | 0.00 | 3.5 | 3.2 | 0.01 |
| Antipsoriatics | 0.5 | 0.3 | 0.02 | 0.5 | 0.4 | 0.00 | 0.4 | 0.4 | 0.01 |
| Antithrombotic agents | 38.4 | 41.3 | -0.06 | 38.5 | 37.8 | 0.01 | 40.6 | 40.2 | 0.01 |
| Beta blocking agents | 30.0 | 32.6 | -0.06 | 30.4 | 29.9 | 0.01 | 32.2 | 31.8 | 0.01 |
| Calcium channel blockers | 23.0 | 23.2 | 0.00 | 23.1 | 23.4 | -0.01 | 23.4 | 23.3 | 0.00 |
| Diuretics | 37.1 | 34.3 | 0.06 | 36.3 | 36.6 | 0.00 | 35.4 | 35.2 | 0.00 |
| Drugs for acid related disorders | 31.3 | 29.7 | 0.04 | 30.3 | 30.9 | -0.01 | 29.7 | 30.2 | -0.01 |
| Drugs for obstructive airway diseases | 39.4 | 35.0 | 0.09 | 37.8 | 39.0 | -0.03 | 35.8 | 36.5 | -0.01 |
| Immunosuppressants | 2.5 | 1.8 | 0.05 | 2.4 | 2.2 | 0.01 | 2.2 | 1.9 | 0.02 |
| Lipid modifying agents | 72.1 | 75.2 | -0.07 | 72.3 | 71.6 | 0.01 | 74.6 | 74.0 | 0.01 |
| Opioids | 23.4 | 21.4 | 0.05 | 22.5 | 22.5 | 0.00 | 22.3 | 22.0 | 0.01 |
| Psycholeptics | 24.6 | 21.6 | 0.07 | 23.3 | 24.0 | -0.02 | 22.4 | 22.6 | 0.00 |
| Psychostimulants | 4.1 | 2.8 | 0.07 | 3.8 | 3.7 | 0.00 | 3.3 | 3.2 | 0.00 |

### 2.1.6 VA

**Supplementary Table 162:** Baseline patient characteristics for GLP1RA (male) (T) and SGLT2I (male) (C) new-users in the VA data source. We report proportion of initiators satisfying selected base-line characteristics and the standardized difference of population proportions (StdDiff) before and after propensity score adjustment. Less extreme StdDiffs through matching and stratification suggest improved balance between patient cohorts through adjustment.

| | Before adjustment | | | After matching | | | After stratification | | |
|---|---|---|---|---|---|---|---|---|---|
| Characteristic | T (%) | C (%) | StdDiff | T (%) | C (%) | StdDiff | T (%) | C (%) | StdDiff |
| **Age group** | | | | | | | | | |
| 25 - 29 | <0.7 | <0.1 | 0.03 | <0.8 | 0.1 | 0.04 | <0.3 | <0.1 | 0.01 |
| 30 - 34 | 0.8 | 0.4 | 0.05 | <0.8 | 0.9 | -0.01 | <0.3 | 0.4 | -0.04 |
| 35 - 39 | 1.8 | 1.5 | 0.02 | 1.5 | 2.0 | -0.04 | 1.3 | 1.6 | -0.02 |
| 40 - 44 | 4.1 | 2.5 | 0.09 | 4.0 | 4.0 | 0.00 | 2.6 | 2.7 | -0.01 |
| 45 - 49 | 7.1 | 4.5 | 0.11 | 6.3 | 6.1 | 0.01 | 3.2 | 4.6 | -0.07 |
| 50 - 54 | 10.6 | 8.3 | 0.08 | 10.8 | 10.7 | 0.00 | 7.4 | 8.5 | -0.04 |
| 55 - 59 | 12.0 | 10.8 | 0.04 | 12.0 | 12.3 | -0.01 | 14.4 | 11.0 | 0.10 |
| 60 - 64 | 12.4 | 13.3 | -0.03 | 11.3 | 12.2 | -0.03 | 11.1 | 13.2 | -0.06 |
| 65 - 69 | 15.2 | 15.5 | -0.01 | 15.4 | 14.5 | 0.02 | 17.6 | 15.4 | 0.06 |
| 70 - 74 | 22.6 | 26.7 | -0.10 | 23.4 | 23.4 | 0.00 | 23.4 | 26.4 | -0.07 |
| 75 - 79 | 8.8 | 11.6 | -0.09 | 9.5 | 9.3 | 0.00 | 13.3 | 11.4 | 0.06 |
| 80 - 84 | 3.0 | 3.1 | 0.00 | 3.4 | 3.0 | 0.02 | 3.9 | 3.1 | 0.04 |
| 85 - 89 | 1.1 | 1.3 | -0.02 | 1.2 | 1.1 | 0.01 | 1.2 | 1.3 | 0.00 |
| 90 - 94 | <0.7 | 0.3 | -0.01 | <0.8 | 0.3 | 0.01 | <0.3 | 0.3 | -0.01 |
| 95 - 99 | <0.7 | <0.1 | 0.02 | <0.3 | <0.1 | 0.02 | <0.3 | <0.1 | 0.02 |
| **Race** | | | | | | | | | |
| Asian | 1.4 | 1.3 | 0.01 | 1.5 | 1.5 | 0.00 | 1.7 | 1.3 | 0.03 |
| Black or African American | 12.2 | 15.5 | -0.10 | 12.0 | 12.2 | -0.01 | 15.5 | 15.2 | 0.01 |
| White | 77.3 | 75.3 | 0.05 | 77.0 | 77.8 | -0.02 | 76.9 | 75.5 | 0.03 |
| Unknown | 7.2 | 6.0 | 0.05 | 7.3 | 6.4 | 0.04 | 4.8 | 6.0 | -0.05 |
| Native Hawaiian or Other Pacific Islander | 1.4 | 1.0 | 0.03 | 1.6 | 1.4 | 0.02 | 0.8 | 1.1 | -0.03 |
| American Indian or Alaska Native | <0.7 | 0.9 | -0.04 | <0.8 | 0.7 | -0.02 | <0.3 | 0.9 | -0.09 |
| **Ethnicity** | | | | | | | | | |
| Hispanic or Latino | 6.2 | 6.6 | -0.02 | 6.7 | 6.6 | 0.00 | 9.7 | 6.6 | 0.11 |
| Not Hispanic or Latino | 90.5 | 90.8 | -0.01 | 90.1 | 90.7 | -0.02 | 88.2 | 90.8 | -0.08 |
| **Medical history: General** | | | | | | | | | |
| Acute respiratory disease | 6.9 | 8.1 | -0.04 | 6.6 | 6.6 | 0.00 | 6.8 | 8.0 | -0.04 |
| Attention deficit hyperactivity disorder | 1.3 | 0.7 | 0.06 | 1.1 | 1.1 | 0.00 | <0.3 | 0.7 | -0.05 |
| Chronic liver disease | 2.4 | 2.2 | 0.01 | 2.1 | 1.9 | 0.01 | 1.5 | 2.2 | -0.05 |
| Chronic obstructive lung disease | 11.2 | 13.2 | -0.06 | 11.1 | 10.9 | 0.01 | 10.3 | 13.0 | -0.09 |
| Crohn’s disease | <0.7 | 0.2 | 0.03 | <0.8 | 0.3 | 0.01 | <0.3 | 0.2 | 0.01 |
| Dementia | 1.8 | 1.2 | 0.05 | 1.9 | 1.1 | 0.06 | 1.4 | 1.2 | 0.01 |
| Depressive disorder | 26.3 | 21.7 | 0.11 | 24.1 | 26.8 | -0.06 | 16.6 | 22.2 | -0.14 |
| Gastroesophageal reflux disease | 21.3 | 20.8 | 0.01 | 20.4 | 21.0 | -0.01 | 20.4 | 20.9 | -0.01 |
| Gastrointestinal hemorrhage | 1.1 | 1.4 | -0.03 | 1.1 | 1.5 | -0.03 | 1.5 | 1.4 | 0.01 |
| Human immunodeficiency virus infection | 0.7 | 0.6 | 0.02 | <0.8 | 0.6 | 0.01 | 0.4 | 0.6 | -0.02 |
| Hyperlipidemia | 71.1 | 74.2 | -0.07 | 70.9 | 69.5 | 0.03 | 80.1 | 73.8 | 0.15 |
| Hypertensive disorder | 75.0 | 77.7 | -0.06 | 73.8 | 72.8 | 0.03 | 83.6 | 77.2 | 0.16 |
| Lesion of liver | 2.2 | 2.0 | 0.01 | 2.2 | 2.0 | 0.01 | 0.9 | 2.0 | -0.09 |
| Obesity | 46.9 | 32.4 | 0.30 | 41.0 | 45.8 | -0.10 | 27.3 | 33.8 | -0.14 |
| Osteoarthritis | 24.9 | 20.7 | 0.10 | 22.4 | 23.6 | -0.03 | 23.7 | 20.9 | 0.06 |
| Pneumonia | 1.0 | 2.0 | -0.08 | 0.8 | 1.3 | -0.04 | 0.6 | 1.9 | -0.12 |
| Psoriasis | 2.0 | 1.9 | 0.01 | 1.9 | 1.7 | 0.02 | 2.4 | 1.8 | 0.04 |
| Renal impairment | 10.5 | 10.1 | 0.01 | 10.5 | 10.2 | 0.01 | 9.4 | 10.0 | -0.02 |
| Rheumatoid arthritis | 0.7 | 0.8 | -0.01 | <0.8 | 0.9 | -0.03 | 0.4 | 0.8 | -0.06 |
| Schizophrenia | <0.7 | 0.6 | -0.02 | <0.8 | 0.6 | -0.04 | <0.3 | 0.6 | -0.07 |
| Ulcerative colitis | <0.7 | 0.4 | -0.05 | <0.8 | 0.5 | -0.05 | <0.3 | 0.4 | -0.07 |
| Urinary tract infectious disease | 2.3 | 1.3 | 0.07 | 2.2 | 1.4 | 0.06 | 1.5 | 1.3 | 0.01 |
| Viral hepatitis C | 0.9 | 1.0 | -0.02 | 0.8 | 0.7 | 0.01 | 1.1 | 1.0 | 0.01 |
| Visual system disorder | 40.8 | 42.7 | -0.04 | 39.7 | 37.0 | 0.06 | 41.6 | 42.2 | -0.01 |
| **Medical history: Cardiovascular disease** | | | | | | | | | |
| Atrial fibrillation | 9.9 | 12.1 | -0.07 | 10.5 | 9.1 | 0.05 | 12.4 | 11.7 | 0.02 |
| Cerebrovascular disease | 3.1 | 4.6 | -0.08 | 3.2 | 3.8 | -0.03 | 4.1 | 4.5 | -0.02 |
| Coronary arteriosclerosis | 23.2 | 32.7 | -0.21 | 25.1 | 22.1 | 0.07 | 43.3 | 31.7 | 0.24 |
| Heart disease | 35.9 | 48.3 | -0.25 | 37.7 | 35.0 | 0.06 | 56.2 | 47.0 | 0.18 |
| Heart failure | 6.5 | 13.6 | -0.24 | 6.9 | 6.4 | 0.02 | 16.9 | 12.8 | 0.11 |
| Ischemic heart disease | 8.2 | 14.3 | -0.20 | 8.2 | 7.2 | 0.04 | 13.8 | 13.6 | 0.01 |

| (Continued) | Before ajustment | | | After matching | | | After stratification | | |
|---|---|---|---|---|---|---|---|---|---|
| Characteristic | T (%) | C (%) | StdDiff | T (%) | C (%) | StdDiff | T (%) | C (%) | StdDiff |
| Peripheral vascular disease | 5.5 | 6.5 | -0.04 | 5.5 | 5.4 | 0.01 | 8.3 | 6.4 | 0.07 |
| Pulmonary embolism | 1.1 | 0.8 | 0.03 | 1.1 | 0.5 | 0.07 | 0.6 | 0.7 | -0.02 |
| Venous thrombosis | 0.9 | 1.0 | -0.01 | 0.8 | 0.9 | 0.00 | 0.8 | 1.0 | -0.03 |
| **Medical history: Neoplasms** | | | | | | | | | |
| Hematologic neoplasm | 1.2 | 1.1 | 0.01 | 1.2 | 0.7 | 0.04 | 2.4 | 1.0 | 0.10 |
| Malignant lymphoma | 0.7 | 0.8 | -0.01 | 0.8 | 0.7 | 0.02 | 1.7 | 0.8 | 0.08 |
| Malignant neoplastic disease | 7.1 | 8.9 | -0.07 | 6.8 | 7.2 | -0.02 | 10.5 | 8.7 | 0.06 |
| Malignant tumor of colon | <0.7 | 0.3 | -0.01 | <0.8 | 0.4 | -0.03 | <0.3 | 0.3 | -0.06 |
| Malignant tumor of lung | <0.7 | 0.4 | -0.07 | <0.8 | 0.2 | -0.03 | <0.3 | 0.4 | -0.08 |
| Malignant tumor of urinary bladder | <0.7 | 0.6 | -0.02 | <0.8 | 0.6 | -0.04 | <0.3 | 0.6 | -0.08 |
| Primary malignant neoplasm of prostate | 2.6 | 3.2 | -0.04 | 2.3 | 2.2 | 0.00 | 5.9 | 3.1 | 0.14 |
| **Medication use** | | | | | | | | | |
| Agents acting on the renin-angiotensin system | 61.4 | 69.1 | -0.16 | 61.5 | 58.6 | 0.06 | 65.6 | 68.1 | -0.05 |
| Antibacterials for systemic use | 23.4 | 25.0 | -0.04 | 22.2 | 21.2 | 0.02 | 20.9 | 24.5 | -0.09 |
| Antidepressants | 41.2 | 34.0 | 0.15 | 39.5 | 41.1 | -0.03 | 30.0 | 34.7 | -0.10 |
| Antiepileptics | 29.9 | 25.4 | 0.10 | 27.7 | 29.2 | -0.03 | 21.4 | 25.7 | -0.10 |
| Antiinflammatory and antirheumatic products | 47.5 | 51.6 | -0.08 | 46.3 | 47.4 | -0.02 | 48.5 | 51.2 | -0.05 |
| Antineoplastic agents | 5.3 | 4.6 | 0.03 | 5.4 | 5.0 | 0.02 | 4.4 | 4.6 | -0.01 |
| Antipsoriatics | 0.9 | 0.8 | 0.01 | 0.8 | 0.6 | 0.03 | 0.9 | 0.7 | 0.02 |
| Antithrombotic agents | 36.1 | 45.4 | -0.19 | 37.0 | 35.7 | 0.03 | 47.4 | 44.4 | 0.06 |
| Beta blocking agents | 37.0 | 48.8 | -0.24 | 38.1 | 35.4 | 0.06 | 47.7 | 47.4 | 0.00 |
| Calcium channel blockers | 24.5 | 29.1 | -0.10 | 24.5 | 23.5 | 0.02 | 24.8 | 28.5 | -0.08 |
| Diuretics | 39.1 | 41.4 | -0.05 | 38.6 | 37.5 | 0.02 | 43.9 | 41.0 | 0.06 |
| Drugs for acid related disorders | 40.3 | 41.8 | -0.03 | 39.4 | 38.0 | 0.03 | 38.4 | 41.3 | -0.06 |
| Drugs for obstructive airway diseases | 39.1 | 39.8 | -0.01 | 38.3 | 38.9 | -0.01 | 39.0 | 39.8 | -0.02 |
| Immunosuppressants | 3.6 | 2.8 | 0.04 | 3.5 | 3.6 | -0.01 | 3.4 | 2.9 | 0.03 |
| Lipid modifying agents | 74.3 | 83.5 | -0.22 | 75.3 | 71.8 | 0.08 | 82.5 | 82.3 | 0.01 |
| Opioids | 16.0 | 14.5 | 0.04 | 14.5 | 14.4 | 0.00 | 13.9 | 14.5 | -0.02 |
| Psycholeptics | 23.7 | 22.7 | 0.02 | 22.6 | 23.3 | -0.02 | 18.2 | 22.8 | -0.11 |
| Psychostimulants | 3.0 | 1.5 | 0.10 | 2.8 | 2.7 | 0.01 | 1.1 | 1.7 | -0.05 |

#### 2.2 SGLT2I (male) vs DPP4I (male)

##### 2.2.1 CCAE

**Supplementary Table 163:** Baseline patient characteristics for SGLT2I (male) (T) and DPP4I (male) (C) new-users in the CCAE data source. We report proportion of initiators satisfying selected base-line characteristics and the standardized difference of population proportions (StdDiff) before and after propensity score adjustment. Less extreme StdDiffs through matching and stratification suggest improved balance between patient cohorts through adjustment.

| | Before adjustment | | | After matching | | | After stratification | | |
|---|---|---|---|---|---|---|---|---|---|
| Characteristic | T (%) | C (%) | StdDiff | T (%) | C (%) | StdDiff | T (%) | C (%) | StdDiff |
| **Age group** | | | | | | | | | |
| 20 - 24 | 0.3 | 0.2 | -0.02 | 0.3 | 0.2 | -0.02 | 0.3 | 0.2 | -0.02 |
| 25 - 29 | 0.5 | 0.5 | 0.00 | 0.5 | 0.5 | 0.00 | 0.4 | 0.5 | 0.00 |
| 30 - 34 | 1.9 | 1.8 | 0.00 | 1.9 | 1.7 | -0.01 | 1.8 | 1.7 | -0.01 |
| 35 - 39 | 4.6 | 4.4 | -0.01 | 4.7 | 4.5 | -0.01 | 4.6 | 4.4 | -0.01 |
| 40 - 44 | 9.1 | 9.0 | 0.00 | 9.2 | 8.9 | -0.01 | 9.0 | 8.8 | 0.00 |
| 45 - 49 | 15.4 | 14.5 | -0.02 | 15.4 | 15.2 | 0.00 | 15.1 | 14.9 | -0.01 |
| 50 - 54 | 20.3 | 20.2 | 0.00 | 20.4 | 20.5 | 0.00 | 20.6 | 20.5 | 0.00 |
| 55 - 59 | 23.8 | 23.5 | -0.01 | 23.4 | 23.9 | 0.01 | 23.2 | 23.8 | 0.01 |
| 60 - 64 | 22.2 | 23.7 | 0.03 | 22.5 | 22.4 | 0.00 | 23.2 | 23.1 | 0.00 |
| 65 - 69 | 1.9 | 2.2 | 0.02 | 1.8 | 2.1 | 0.02 | 1.8 | 2.1 | 0.02 |
| **Medical history: General** | | | | | | | | | |
| Acute respiratory disease | 23.1 | 22.4 | -0.02 | 23.1 | 23.7 | 0.02 | 22.8 | 23.1 | 0.01 |
| Attention deficit hyperactivity disorder | 1.2 | 0.8 | -0.05 | 1.2 | 1.1 | -0.01 | 1.1 | 1.0 | -0.01 |
| Chronic liver disease | 1.6 | 2.7 | 0.08 | 1.8 | 1.8 | 0.00 | 2.1 | 2.0 | -0.01 |
| Chronic obstructive lung disease | 2.3 | 2.6 | 0.02 | 2.3 | 2.6 | 0.02 | 2.4 | 2.6 | 0.02 |
| Crohn’s disease | 0.1 | 0.2 | 0.02 | 0.1 | 0.2 | 0.02 | 0.1 | 0.2 | 0.02 |
| Dementia | 0.1 | 0.1 | 0.00 | 0.1 | 0.2 | 0.02 | 0.1 | 0.2 | 0.01 |
| Depressive disorder | 7.0 | 5.9 | -0.05 | 6.8 | 7.2 | 0.02 | 6.7 | 6.9 | 0.01 |
| Gastroesophageal reflux disease | 11.3 | 8.8 | -0.08 | 11.1 | 11.3 | 0.01 | 10.8 | 10.9 | 0.00 |
| Gastrointestinal hemorrhage | 1.4 | 1.8 | 0.03 | 1.4 | 1.6 | 0.01 | 1.4 | 1.6 | 0.01 |
| Human immunodeficiency virus infection | 0.5 | 0.4 | -0.01 | 0.5 | 0.6 | 0.01 | 0.5 | 0.5 | 0.01 |
| Hyperlipidemia | 75.7 | 70.1 | -0.13 | 75.1 | 75.2 | 0.00 | 75.0 | 75.0 | 0.00 |
| Hypertensive disorder | 72.2 | 66.3 | -0.13 | 71.5 | 71.6 | 0.00 | 71.2 | 71.1 | 0.00 |
| Lesion of liver | 0.6 | 0.6 | -0.01 | 0.6 | 0.6 | -0.01 | 0.6 | 0.6 | 0.00 |
| Obesity | 25.3 | 14.8 | -0.26 | 23.2 | 23.3 | 0.00 | 22.1 | 22.2 | 0.00 |
| Osteoarthritis | 15.8 | 15.1 | -0.02 | 15.8 | 16.1 | 0.01 | 15.8 | 16.1 | 0.01 |
| Pneumonia | 1.9 | 1.8 | -0.01 | 1.8 | 2.1 | 0.02 | 1.8 | 2.0 | 0.01 |
| Psoriasis | 1.7 | 1.5 | -0.02 | 1.6 | 1.6 | 0.00 | 1.6 | 1.6 | 0.00 |
| Renal impairment | 3.6 | 3.4 | -0.01 | 3.5 | 3.8 | 0.01 | 3.7 | 3.8 | 0.01 |
| Schizophrenia | 0.1 | 0.1 | 0.01 | 0.1 | 0.1 | 0.01 | 0.1 | 0.1 | 0.01 |
| Ulcerative colitis | 0.3 | 0.3 | 0.01 | 0.3 | 0.4 | 0.01 | 0.3 | 0.4 | 0.01 |
| Urinary tract infectious disease | 2.1 | 3.0 | 0.06 | 2.3 | 2.3 | 0.01 | 2.5 | 2.6 | 0.00 |
| Viral hepatitis C | 0.3 | 0.4 | 0.02 | 0.3 | 0.3 | 0.00 | 0.4 | 0.4 | 0.00 |
| Visual system disorder | 22.7 | 21.8 | -0.02 | 22.9 | 23.1 | 0.00 | 23.1 | 23.0 | 0.00 |
| **Medical history: Cardiovascular disease** | | | | | | | | | |
| Atrial fibrillation | 2.7 | 2.4 | -0.02 | 2.5 | 2.7 | 0.01 | 2.6 | 2.7 | 0.00 |
| Cerebrovascular disease | 2.5 | 2.5 | 0.00 | 2.4 | 2.2 | -0.01 | 2.6 | 2.3 | -0.02 |
| Coronary arteriosclerosis | 11.3 | 10.1 | -0.04 | 9.7 | 9.8 | 0.00 | 10.2 | 10.0 | -0.01 |
| Heart disease | 20.3 | 18.9 | -0.04 | 18.6 | 18.5 | 0.00 | 19.1 | 18.7 | -0.01 |
| Heart failure | 2.5 | 2.0 | -0.03 | 2.1 | 2.2 | 0.00 | 2.2 | 2.2 | 0.00 |
| Ischemic heart disease | 5.4 | 4.9 | -0.03 | 4.7 | 4.7 | 0.00 | 5.0 | 4.7 | -0.01 |
| Peripheral vascular disease | 4.2 | 3.3 | -0.04 | 3.8 | 3.6 | -0.01 | 3.9 | 3.6 | -0.01 |
| Pulmonary embolism | 0.4 | 0.4 | 0.00 | 0.4 | 0.5 | 0.01 | 0.4 | 0.5 | 0.01 |
| Venous thrombosis | 0.9 | 1.1 | 0.02 | 1.0 | 1.1 | 0.01 | 1.1 | 1.1 | 0.01 |
| **Medical history: Neoplasms** | | | | | | | | | |
| Hematologic neoplasm | 0.7 | 0.8 | 0.01 | 0.7 | 0.8 | 0.02 | 0.7 | 0.8 | 0.01 |
| Malignant neoplasm of anorectum | 0.1 | 0.1 | 0.01 | 0.1 | 0.1 | 0.00 | 0.2 | 0.1 | -0.01 |
| Malignant neoplastic disease | 4.7 | 5.3 | 0.03 | 4.7 | 4.9 | 0.01 | 4.7 | 4.9 | 0.01 |
| Malignant tumor of colon | 0.3 | 0.3 | 0.00 | 0.3 | 0.3 | -0.01 | 0.3 | 0.3 | -0.01 |
| Malignant tumor of lung | 0.1 | 0.1 | 0.01 | 0.1 | 0.1 | 0.01 | 0.1 | 0.1 | 0.02 |
| Malignant tumor of urinary bladder | 0.2 | 0.3 | 0.03 | 0.2 | 0.2 | 0.01 | 0.2 | 0.3 | 0.01 |
| Primary malignant neoplasm of prostate | 1.0 | 1.4 | 0.03 | 1.1 | 1.2 | 0.01 | 1.2 | 1.3 | 0.01 |
| **Medication use** | | | | | | | | | |
| Agents acting on the renin-angiotensin system | 71.2 | 70.0 | -0.03 | 70.6 | 71.0 | 0.01 | 70.3 | 70.5 | 0.00 |

| (Continued) | Before ajustment | | | After matching | | | After stratification | | |
|---|---|---|---|---|---|---|---|---|---|
| Characteristic | T (%) | C (%) | StdDiff | T (%) | C (%) | StdDiff | T (%) | C (%) | StdDiff |
| Antibacterials for systemic use | 50.9 | 51.7 | 0.01 | 51.0 | 50.9 | 0.00 | 51.0 | 50.9 | 0.00 |
| Antidepressants | 18.8 | 17.3 | -0.04 | 18.3 | 18.3 | 0.00 | 17.9 | 18.0 | 0.00 |
| Antiepileptics | 9.9 | 8.5 | -0.05 | 9.8 | 9.9 | 0.00 | 9.8 | 9.8 | 0.00 |
| Antiinflammatory and antirheumatic products | 30.2 | 27.4 | -0.06 | 29.8 | 29.7 | 0.00 | 29.7 | 29.5 | 0.00 |
| Antineoplastic agents | 3.1 | 3.4 | 0.01 | 3.1 | 3.0 | 0.00 | 3.1 | 3.2 | 0.00 |
| Antipsoriatics | 0.5 | 0.5 | 0.00 | 0.5 | 0.5 | -0.01 | 0.5 | 0.4 | -0.01 |
| Antithrombotic agents | 12.1 | 10.5 | -0.05 | 11.2 | 11.3 | 0.00 | 11.6 | 11.2 | -0.01 |
| Beta blocking agents | 26.0 | 25.3 | -0.02 | 24.8 | 24.6 | 0.00 | 25.1 | 24.9 | 0.00 |
| Calcium channel blockers | 22.8 | 21.3 | -0.04 | 22.1 | 22.4 | 0.01 | 22.0 | 22.1 | 0.00 |
| Diuretics | 32.5 | 31.9 | -0.01 | 31.8 | 32.1 | 0.01 | 31.8 | 31.7 | 0.00 |
| Drugs for acid related disorders | 20.6 | 19.9 | -0.02 | 20.4 | 20.2 | 0.00 | 20.4 | 20.2 | 0.00 |
| Drugs for obstructive airway diseases | 31.8 | 30.0 | -0.04 | 31.6 | 31.4 | 0.00 | 31.7 | 31.3 | -0.01 |
| Immunosuppressants | 1.7 | 1.7 | -0.01 | 1.7 | 1.7 | 0.00 | 1.7 | 1.9 | 0.01 |
| Lipid modifying agents | 72.9 | 71.7 | -0.03 | 71.7 | 71.5 | 0.00 | 71.7 | 71.5 | 0.00 |
| Opioids | 21.4 | 21.9 | 0.01 | 21.8 | 21.5 | -0.01 | 22.1 | 21.6 | -0.01 |
| Psycholeptics | 17.6 | 17.7 | 0.00 | 17.4 | 17.6 | 0.00 | 17.8 | 17.5 | -0.01 |
| Psychostimulants | 2.7 | 2.2 | -0.03 | 2.6 | 2.4 | -0.01 | 2.6 | 2.4 | -0.01 |

##### 2.2.2 DAG

**Supplementary Table 164:** Baseline patient characteristics for SGLT2I (male) (T) and DPP4I (male) (C) new-users in the DAG data source. We report proportion of initiators satisfying selected base-line characteristics and the standardized difference of population proportions (StdDiff) before and after propensity score adjustment. Less extreme StdDiffs through matching and stratification suggest improved balance between patient cohorts through adjustment.

| | Before adjustment | | | After matching | | | After stratification | | |
|---|---|---|---|---|---|---|---|---|---|
| Characteristic | T (%) | C (%) | StdDiff | T (%) | C (%) | StdDiff | T (%) | C (%) | StdDiff |
| **Age group** | | | | | | | | | |
| 20 - 24 | <0.1 | 0.1 | 0.01 | <0.1 | 0.1 | 0.02 | <0.1 | 0.1 | 0.00 |
| 25 - 29 | 0.1 | 0.2 | 0.01 | 0.1 | 0.2 | 0.02 | 0.1 | 0.2 | 0.01 |
| 30 - 34 | 0.6 | 0.4 | -0.02 | 0.7 | 0.4 | -0.04 | 0.7 | 0.5 | -0.02 |
| 35 - 39 | 1.5 | 1.3 | -0.01 | 1.6 | 1.5 | -0.01 | 1.8 | 1.4 | -0.04 |
| 40 - 44 | 3.1 | 2.9 | -0.01 | 3.4 | 3.0 | -0.02 | 3.7 | 2.9 | -0.05 |
| 45 - 49 | 6.0 | 5.9 | 0.00 | 6.3 | 5.6 | -0.03 | 6.3 | 5.6 | -0.03 |
| 50 - 54 | 10.9 | 10.4 | -0.02 | 11.0 | 11.1 | 0.00 | 11.3 | 10.4 | -0.03 |
| 55 - 59 | 15.4 | 14.9 | -0.02 | 15.7 | 15.8 | 0.00 | 15.5 | 15.2 | -0.01 |
| 60 - 64 | 17.5 | 15.9 | -0.04 | 16.7 | 16.9 | 0.01 | 16.1 | 15.9 | -0.01 |
| 65 - 69 | 14.3 | 14.2 | 0.00 | 14.3 | 14.5 | 0.01 | 14.1 | 14.6 | 0.01 |
| 70 - 74 | 12.6 | 12.9 | 0.01 | 12.0 | 12.9 | 0.03 | 12.0 | 12.4 | 0.01 |
| 75 - 79 | 9.5 | 10.8 | 0.04 | 9.5 | 9.3 | -0.01 | 9.7 | 10.6 | 0.03 |
| 80 - 84 | 6.0 | 6.9 | 0.04 | 6.2 | 5.9 | -0.01 | 6.9 | 7.0 | 0.00 |
| 85 - 89 | 2.0 | 2.6 | 0.04 | 2.1 | 2.1 | 0.00 | 2.1 | 2.7 | 0.04 |
| 90 - 94 | 0.2 | 0.6 | 0.06 | 0.2 | 0.7 | 0.07 | 0.2 | 0.6 | 0.06 |
| **Medical history: General** | | | | | | | | | |
| Acute respiratory disease | 15.1 | 17.7 | 0.07 | 15.7 | 15.6 | 0.00 | 16.3 | 16.6 | 0.01 |
| Attention deficit hyperactivity disorder | 0.1 | 0.1 | 0.02 | 0.1 | 0.1 | 0.01 | 0.1 | 0.2 | 0.03 |
| Chronic liver disease | 0.6 | 0.5 | -0.01 | 0.5 | 0.3 | -0.03 | 0.5 | 0.5 | 0.00 |
| Chronic obstructive lung disease | 6.2 | 6.3 | 0.00 | 5.8 | 5.9 | 0.00 | 6.2 | 6.2 | 0.00 |
| Crohn’s disease | 0.1 | 0.2 | 0.02 | 0.1 | 0.2 | 0.01 | 0.1 | 0.2 | 0.03 |
| Dementia | 0.8 | 1.8 | 0.08 | 0.9 | 1.1 | 0.02 | 1.2 | 1.7 | 0.04 |
| Depressive disorder | 6.4 | 6.7 | 0.01 | 6.1 | 7.0 | 0.03 | 6.3 | 6.8 | 0.02 |
| Gastroesophageal reflux disease | 1.9 | 2.1 | 0.01 | 1.8 | 1.9 | 0.01 | 1.9 | 2.0 | 0.00 |
| Gastrointestinal hemorrhage | 0.7 | 0.8 | 0.01 | 0.7 | 0.9 | 0.02 | 0.8 | 0.8 | 0.00 |
| Hyperlipidemia | 24.3 | 24.8 | 0.01 | 22.8 | 22.8 | 0.00 | 23.2 | 23.4 | 0.00 |
| Hypertensive disorder | 42.6 | 47.1 | 0.09 | 42.7 | 43.9 | 0.02 | 42.7 | 43.9 | 0.02 |
| Lesion of liver | 0.6 | 0.9 | 0.03 | 0.6 | 0.7 | 0.01 | 0.5 | 0.9 | 0.04 |
| Obesity | 10.0 | 7.9 | -0.07 | 9.1 | 8.7 | -0.01 | 9.1 | 8.7 | -0.01 |
| Osteoarthritis | 9.0 | 10.2 | 0.04 | 9.0 | 9.1 | 0.00 | 9.3 | 9.9 | 0.02 |
| Pneumonia | 1.5 | 2.4 | 0.06 | 1.5 | 1.6 | 0.00 | 1.6 | 2.1 | 0.04 |
| Psoriasis | 1.2 | 1.3 | 0.00 | 1.1 | 1.4 | 0.03 | 1.1 | 1.4 | 0.02 |
| Renal impairment | 5.2 | 5.9 | 0.03 | 5.0 | 4.9 | -0.01 | 5.2 | 5.7 | 0.02 |
| Rheumatoid arthritis | 0.9 | 0.9 | 0.01 | 0.8 | 0.6 | -0.02 | 0.8 | 0.8 | 0.00 |
| Schizophrenia | 0.2 | 0.3 | 0.01 | 0.3 | 0.2 | -0.02 | 0.3 | 0.2 | -0.03 |
| Ulcerative colitis | 0.2 | 0.2 | 0.00 | 0.2 | 0.2 | 0.01 | 0.1 | 0.2 | 0.02 |
| Urinary tract infectious disease | 2.5 | 2.8 | 0.02 | 2.5 | 2.5 | 0.00 | 2.4 | 2.8 | 0.03 |
| Viral hepatitis C | <0.1 | 0.1 | 0.02 | <0.1 | <0.1 | 0.01 | <0.1 | 0.1 | 0.02 |
| Visual system disorder | 6.5 | 8.2 | 0.06 | 6.5 | 6.2 | -0.01 | 7.1 | 7.2 | 0.00 |
| **Medical history: Cardiovascular disease** | | | | | | | | | |
| Atrial fibrillation | 3.5 | 2.9 | -0.03 | 3.1 | 3.0 | 0.00 | 3.0 | 3.2 | 0.01 |
| Cerebrovascular disease | 4.4 | 4.5 | 0.00 | 4.1 | 3.9 | -0.01 | 4.2 | 4.4 | 0.01 |
| Coronary arteriosclerosis | 9.0 | 5.6 | -0.13 | 6.8 | 6.6 | -0.01 | 6.3 | 6.6 | 0.01 |
| Heart disease | 31.5 | 26.8 | -0.10 | 26.2 | 26.7 | 0.01 | 25.8 | 27.3 | 0.03 |
| Heart failure | 8.8 | 6.4 | -0.09 | 6.6 | 6.6 | 0.00 | 6.7 | 6.8 | 0.01 |
| Ischemic heart disease | 15.8 | 12.9 | -0.08 | 12.4 | 12.7 | 0.01 | 12.0 | 12.8 | 0.02 |
| Peripheral vascular disease | 6.6 | 6.0 | -0.03 | 6.1 | 5.7 | -0.02 | 6.0 | 6.2 | 0.01 |
| Pulmonary embolism | 0.6 | 0.6 | 0.01 | 0.5 | 0.7 | 0.02 | 0.4 | 0.7 | 0.03 |
| Venous thrombosis | 1.3 | 1.3 | 0.00 | 1.3 | 1.4 | 0.01 | 1.4 | 1.4 | 0.00 |
| **Medical history: Neoplasms** | | | | | | | | | |
| Hematologic neoplasm | 0.3 | 0.4 | 0.02 | 0.2 | 0.3 | 0.01 | 0.3 | 0.3 | 0.01 |
| Malignant lymphoma | 0.2 | 0.3 | 0.02 | 0.2 | 0.2 | 0.01 | 0.3 | 0.3 | 0.00 |
| Malignant neoplasm of anorectum | 0.1 | 0.2 | 0.03 | 0.1 | 0.2 | 0.02 | 0.1 | 0.2 | 0.02 |
| Malignant neoplastic disease | 3.5 | 5.4 | 0.09 | 3.7 | 3.6 | 0.00 | 4.2 | 4.8 | 0.03 |
| Malignant tumor of colon | 0.3 | 0.4 | 0.01 | 0.3 | 0.3 | -0.01 | 0.3 | 0.4 | 0.01 |
| Malignant tumor of lung | <0.1 | 0.2 | 0.03 | <0.1 | 0.1 | 0.03 | <0.1 | 0.1 | 0.03 |
| Malignant tumor of urinary bladder | 0.2 | 0.3 | 0.02 | 0.2 | 0.2 | 0.00 | 0.3 | 0.2 | -0.02 |

| (Continued) | Before ajustment | | | After matching | | | After stratification | | |
|---|---|---|---|---|---|---|---|---|---|
| Characteristic | T (%) | C (%) | StdDiff | T (%) | C (%) | StdDiff | T (%) | C (%) | StdDiff |
| Primary malignant neoplasm of prostate | 1.0 | 1.3 | 0.03 | 1.0 | 0.7 | -0.03 | 1.4 | 1.1 | -0.02 |
| **Medication use** | | | | | | | | | |
| Agents acting on the renin-angiotensin system | 71.7 | 68.1 | -0.08 | 68.8 | 69.6 | 0.02 | 68.1 | 69.2 | 0.03 |
| Antibacterials for systemic use | 19.5 | 23.7 | 0.10 | 20.0 | 19.1 | -0.02 | 20.0 | 19.1 | -0.02 |
| Antidepressants | 7.4 | 7.5 | 0.01 | 7.5 | 7.3 | -0.01 | 7.7 | 7.8 | 0.01 |
| Antiepileptics | 3.9 | 4.3 | 0.02 | 3.9 | 4.1 | 0.01 | 4.1 | 4.4 | 0.01 |
| Antiinflammatory and antirheumatic products | 43.7 | 43.9 | 0.00 | 41.6 | 42.7 | 0.02 | 41.6 | 43.6 | 0.04 |
| Antineoplastic agents | 1.1 | 1.0 | -0.01 | 1.1 | 0.8 | -0.03 | 1.2 | 0.9 | -0.03 |
| Antipsoriatics | 0.6 | 0.4 | -0.02 | 0.5 | 0.6 | 0.01 | 0.6 | 0.5 | -0.01 |
| Antithrombotic agents | 39.1 | 34.3 | -0.10 | 34.0 | 34.8 | 0.02 | 33.5 | 35.4 | 0.04 |
| Beta blocking agents | 48.2 | 44.6 | -0.07 | 44.4 | 43.9 | -0.01 | 44.0 | 45.2 | 0.02 |
| Calcium channel blockers | 34.8 | 31.9 | -0.06 | 33.6 | 34.2 | 0.01 | 33.8 | 33.4 | -0.01 |
| Diuretics | 46.1 | 45.6 | -0.01 | 43.2 | 43.4 | 0.00 | 43.6 | 44.9 | 0.03 |
| Drugs for acid related disorders | 32.3 | 31.9 | -0.01 | 30.7 | 31.1 | 0.01 | 31.8 | 32.5 | 0.02 |
| Drugs for obstructive airway diseases | 16.6 | 16.3 | -0.01 | 16.3 | 16.2 | 0.00 | 15.6 | 16.4 | 0.02 |
| Immunosuppressants | 0.9 | 0.6 | -0.03 | 0.8 | 0.4 | -0.04 | 0.7 | 0.5 | -0.03 |
| Lipid modifying agents | 56.3 | 48.1 | -0.17 | 51.1 | 52.7 | 0.03 | 48.9 | 50.9 | 0.04 |
| Opioids | 9.0 | 10.6 | 0.05 | 9.2 | 9.0 | -0.01 | 9.7 | 10.3 | 0.02 |
| Psycholeptics | 6.3 | 7.0 | 0.03 | 6.3 | 6.7 | 0.01 | 6.9 | 6.9 | 0.00 |
| Psychostimulants | 0.1 | 0.2 | 0.02 | 0.1 | 0.2 | 0.00 | 0.1 | 0.2 | 0.01 |

##### 2.2.3 ODOD

**Supplementary Table 165:** Baseline patient characteristics for SGLT2I (male) (T) and DPP4I (male) (C) new-users in the ODOD data source. We report proportion of initiators satisfying selected base-line characteristics and the standardized difference of population proportions (StdDiff) before and after propensity score adjustment. Less extreme StdDiffs through matching and stratification suggest improved balance between patient cohorts through adjustment.

| | Before adjustment | | | After matching | | | After stratification | | |
|---|---|---|---|---|---|---|---|---|---|
| Characteristic | T (%) | C (%) | StdDiff | T (%) | C (%) | StdDiff | T (%) | C (%) | StdDiff |
| **Age group** | | | | | | | | | |
| 25 - 29 | 0.4 | 0.3 | -0.01 | 0.4 | 0.3 | -0.03 | 0.4 | 0.3 | -0.03 |
| 30 - 34 | 1.3 | 1.2 | -0.01 | 1.4 | 1.2 | -0.02 | 1.3 | 1.1 | -0.02 |
| 35 - 39 | 3.2 | 3.2 | 0.00 | 3.2 | 2.9 | -0.02 | 3.0 | 2.8 | -0.01 |
| 40 - 44 | 6.1 | 6.0 | 0.00 | 6.0 | 5.9 | 0.00 | 5.6 | 5.4 | -0.01 |
| 45 - 49 | 9.5 | 8.8 | -0.02 | 9.1 | 9.0 | 0.00 | 8.7 | 8.5 | -0.01 |
| 50 - 54 | 13.3 | 11.9 | -0.04 | 12.5 | 12.4 | 0.00 | 12.1 | 11.9 | -0.01 |
| 55 - 59 | 15.0 | 14.0 | -0.03 | 14.7 | 14.7 | 0.00 | 14.5 | 14.0 | -0.01 |
| 60 - 64 | 14.6 | 13.2 | -0.04 | 13.9 | 13.9 | 0.00 | 13.6 | 13.4 | -0.01 |
| 65 - 69 | 14.7 | 13.9 | -0.02 | 14.4 | 15.2 | 0.02 | 14.5 | 15.3 | 0.02 |
| 70 - 74 | 12.1 | 12.5 | 0.01 | 12.8 | 13.0 | 0.00 | 13.0 | 13.2 | 0.01 |
| 75 - 79 | 6.4 | 8.1 | 0.06 | 7.3 | 7.2 | 0.00 | 7.6 | 7.9 | 0.01 |
| 80 - 84 | 2.3 | 4.5 | 0.12 | 3.0 | 2.9 | 0.00 | 3.7 | 3.9 | 0.01 |
| 85 - 89 | 0.9 | 2.0 | 0.10 | 1.1 | 1.2 | 0.01 | 1.7 | 1.8 | 0.01 |
| 90 - 94 | 0.1 | 0.2 | 0.02 | 0.1 | 0.2 | 0.03 | 0.1 | 0.2 | 0.03 |
| **Medical history: General** | | | | | | | | | |
| Acute respiratory disease | 21.6 | 23.2 | 0.04 | 21.9 | 21.8 | 0.00 | 22.6 | 22.1 | -0.01 |
| Attention deficit hyperactivity disorder | 0.9 | 0.8 | -0.01 | 0.9 | 1.1 | 0.02 | 0.8 | 1.0 | 0.02 |
| Chronic liver disease | 1.7 | 3.3 | 0.10 | 2.0 | 1.8 | -0.01 | 2.7 | 2.3 | -0.02 |
| Chronic obstructive lung disease | 7.4 | 8.2 | 0.03 | 7.5 | 7.6 | 0.00 | 8.3 | 8.4 | 0.00 |
| Crohn’s disease | 0.3 | 0.3 | 0.00 | 0.3 | 0.3 | -0.01 | 0.4 | 0.3 | -0.02 |
| Dementia | 0.8 | 1.8 | 0.09 | 0.9 | 1.2 | 0.03 | 1.3 | 1.6 | 0.03 |
| Depressive disorder | 9.8 | 9.1 | -0.03 | 9.5 | 9.7 | 0.01 | 9.8 | 10.0 | 0.01 |
| Gastroesophageal reflux disease | 16.0 | 14.4 | -0.04 | 15.6 | 15.1 | -0.01 | 16.3 | 15.7 | -0.01 |
| Gastrointestinal hemorrhage | 2.0 | 2.6 | 0.04 | 2.0 | 1.9 | -0.01 | 2.0 | 1.9 | -0.01 |
| Human immunodeficiency virus infection | 0.5 | 0.5 | 0.00 | 0.5 | 0.6 | 0.02 | 0.5 | 0.6 | 0.02 |
| Hyperlipidemia | 81.8 | 82.3 | 0.01 | 81.1 | 81.3 | 0.00 | 81.1 | 81.3 | 0.00 |
| Hypertensive disorder | 78.8 | 78.6 | 0.00 | 78.1 | 77.7 | -0.01 | 79.0 | 78.7 | -0.01 |
| Lesion of liver | 1.2 | 1.1 | -0.01 | 1.2 | 1.0 | -0.03 | 1.4 | 1.1 | -0.03 |
| Obesity | 28.9 | 19.5 | -0.22 | 25.5 | 26.2 | 0.01 | 25.4 | 25.2 | 0.00 |
| Osteoarthritis | 22.2 | 22.1 | 0.00 | 22.4 | 22.0 | -0.01 | 23.1 | 22.8 | -0.01 |
| Pneumonia | 3.0 | 3.6 | 0.03 | 2.9 | 2.9 | 0.00 | 3.4 | 3.5 | 0.00 |
| Psoriasis | 2.0 | 1.6 | -0.03 | 1.9 | 1.6 | -0.02 | 1.9 | 1.7 | -0.02 |
| Renal impairment | 10.3 | 13.5 | 0.10 | 11.4 | 10.8 | -0.02 | 13.0 | 13.3 | 0.01 |
| Rheumatoid arthritis | 0.9 | 1.0 | 0.01 | 0.9 | 0.9 | 0.00 | 1.1 | 0.9 | -0.01 |
| Schizophrenia | 0.3 | 0.4 | 0.02 | 0.3 | 0.5 | 0.03 | 0.3 | 0.5 | 0.04 |
| Ulcerative colitis | 0.4 | 0.4 | 0.00 | 0.4 | 0.3 | 0.00 | 0.4 | 0.3 | -0.02 |
| Urinary tract infectious disease | 3.4 | 5.9 | 0.12 | 4.0 | 3.9 | -0.01 | 5.1 | 5.0 | 0.00 |
| Viral hepatitis C | 0.5 | 0.7 | 0.02 | 0.5 | 0.6 | 0.01 | 0.6 | 0.6 | 0.01 |
| Visual system disorder | 33.7 | 35.8 | 0.04 | 35.1 | 35.0 | 0.00 | 36.1 | 36.1 | 0.00 |
| **Medical history: Cardiovascular disease** | | | | | | | | | |
| Atrial fibrillation | 7.4 | 7.3 | -0.01 | 6.9 | 7.0 | 0.00 | 7.2 | 7.7 | 0.02 |
| Cerebrovascular disease | 5.4 | 6.5 | 0.04 | 5.4 | 5.2 | -0.01 | 5.9 | 6.0 | 0.00 |
| Coronary arteriosclerosis | 21.5 | 18.9 | -0.06 | 18.6 | 18.5 | 0.00 | 19.7 | 20.0 | 0.01 |
| Heart disease | 33.4 | 32.1 | -0.03 | 30.8 | 30.4 | -0.01 | 32.8 | 32.7 | 0.00 |
| Heart failure | 8.2 | 6.7 | -0.06 | 6.7 | 6.4 | -0.01 | 7.6 | 7.4 | -0.01 |
| Ischemic heart disease | 11.2 | 9.8 | -0.05 | 9.6 | 9.4 | -0.01 | 10.6 | 10.4 | -0.01 |
| Peripheral vascular disease | 10.8 | 10.4 | -0.01 | 10.5 | 10.4 | 0.00 | 11.4 | 11.4 | 0.00 |
| Pulmonary embolism | 0.7 | 0.7 | 0.00 | 0.7 | 0.6 | -0.01 | 0.7 | 0.8 | 0.00 |
| Venous thrombosis | 1.5 | 1.6 | 0.01 | 1.5 | 1.2 | -0.02 | 1.7 | 1.5 | -0.01 |
| **Medical history: Neoplasms** | | | | | | | | | |
| Hematologic neoplasm | 0.9 | 1.2 | 0.02 | 1.0 | 1.0 | 0.01 | 1.0 | 1.1 | 0.01 |
| Malignant lymphoma | 0.5 | 0.6 | 0.01 | 0.5 | 0.6 | 0.02 | 0.5 | 0.6 | 0.02 |
| Malignant neoplasm of anorectum | 0.2 | 0.2 | 0.01 | 0.2 | 0.2 | -0.01 | 0.3 | 0.2 | -0.01 |
| Malignant neoplastic disease | 8.7 | 10.9 | 0.07 | 9.4 | 9.0 | -0.01 | 10.1 | 10.4 | 0.01 |
| Malignant tumor of colon | 0.5 | 0.6 | 0.01 | 0.6 | 0.3 | -0.04 | 0.7 | 0.5 | -0.02 |
| Malignant tumor of lung | 0.2 | 0.4 | 0.04 | 0.2 | 0.3 | 0.03 | 0.2 | 0.4 | 0.02 |
| Malignant tumor of urinary bladder | 0.4 | 0.7 | 0.04 | 0.5 | 0.5 | 0.01 | 0.6 | 0.7 | 0.01 |

| (Continued) | Before ajustment | | | After matching | | | After stratification | | |
|---|---|---|---|---|---|---|---|---|---|
| Characteristic | T (%) | C (%) | StdDiff | T (%) | C (%) | StdDiff | T (%) | C (%) | StdDiff |
| Primary malignant neoplasm of prostate | 3.0 | 4.2 | 0.06 | 3.4 | 3.1 | -0.02 | 3.7 | 3.8 | 0.00 |
| **Medication use** | | | | | | | | | |
| Agents acting on the renin-angiotensin system | 73.1 | 71.4 | -0.04 | 72.2 | 71.9 | -0.01 | 72.2 | 72.4 | 0.00 |
| Antibacterials for systemic use | 49.7 | 52.8 | 0.06 | 50.6 | 50.2 | -0.01 | 51.8 | 51.2 | -0.01 |
| Antidepressants | 19.9 | 18.8 | -0.03 | 19.5 | 19.3 | 0.00 | 19.9 | 19.7 | 0.00 |
| Antiepileptics | 13.8 | 12.8 | -0.03 | 14.0 | 14.3 | 0.01 | 14.5 | 14.5 | 0.00 |
| Antiinflammatory and antirheumatic products | 27.6 | 26.7 | -0.02 | 27.4 | 27.5 | 0.00 | 28.2 | 27.6 | -0.01 |
| Antineoplastic agents | 4.2 | 4.8 | 0.03 | 4.3 | 4.6 | 0.02 | 4.5 | 4.7 | 0.01 |
| Antipsoriatics | 0.4 | 0.6 | 0.02 | 0.4 | 0.5 | 0.01 | 0.4 | 0.6 | 0.02 |
| Antithrombotic agents | 19.5 | 17.3 | -0.06 | 17.7 | 17.5 | -0.01 | 18.8 | 18.8 | 0.00 |
| Beta blocking agents | 35.3 | 33.6 | -0.03 | 33.4 | 33.4 | 0.00 | 34.6 | 34.6 | 0.00 |
| Calcium channel blockers | 25.8 | 25.7 | 0.00 | 25.9 | 26.1 | 0.00 | 26.4 | 26.5 | 0.00 |
| Diuretics | 36.9 | 36.3 | -0.01 | 35.8 | 35.3 | -0.01 | 36.6 | 36.4 | 0.00 |
| Drugs for acid related disorders | 22.2 | 22.6 | 0.01 | 22.5 | 22.1 | -0.01 | 23.7 | 23.4 | -0.01 |
| Drugs for obstructive airway diseases | 33.2 | 32.9 | -0.01 | 33.3 | 33.0 | 0.00 | 34.1 | 33.6 | -0.01 |
| Immunosuppressants | 1.8 | 1.8 | 0.00 | 1.8 | 1.8 | 0.00 | 1.9 | 1.8 | -0.01 |
| Lipid modifying agents | 78.4 | 76.3 | -0.05 | 77.1 | 77.4 | 0.01 | 77.3 | 77.7 | 0.01 |
| Opioids | 23.9 | 24.2 | 0.01 | 23.7 | 23.4 | -0.01 | 24.7 | 24.1 | -0.01 |
| Psycholeptics | 19.8 | 20.0 | 0.00 | 19.6 | 19.8 | 0.01 | 20.2 | 20.3 | 0.00 |
| Psychostimulants | 1.9 | 1.8 | -0.01 | 1.9 | 2.1 | 0.01 | 1.9 | 2.0 | 0.01 |

##### 2.2.4 MDCR

**Supplementary Table 166:** Baseline patient characteristics for SGLT2I (male) (T) and DPP4I (male) (C) new-users in the MDCR data source. We report proportion of initiators satisfying selected base-line characteristics and the standardized difference of population proportions (StdDiff) before and after propensity score adjustment. Less extreme StdDiffs through matching and stratification suggest improved balance between patient cohorts through adjustment.

| | Before adjustment | | | After matching | | | After stratification | | |
|---|---|---|---|---|---|---|---|---|---|
| Characteristic | T (%) | C (%) | StdDiff | T (%) | C (%) | StdDiff | T (%) | C (%) | StdDiff |
| **Age group** | | | | | | | | | |
| 45 - 49 | <0.2 | <0.1 | -0.02 | <0.2 | <0.3 | -0.01 | 0.3 | <0.1 | -0.06 |
| 50 - 54 | 0.3 | 0.2 | -0.03 | 0.4 | <0.3 | -0.05 | 0.2 | 0.2 | 0.00 |
| 55 - 59 | 1.0 | 0.5 | -0.05 | 1.1 | 0.5 | -0.06 | 1.1 | 0.5 | -0.06 |
| 60 - 64 | 2.4 | 1.9 | -0.04 | 2.5 | 1.9 | -0.04 | 3.3 | 2.0 | -0.08 |
| 65 - 69 | 36.1 | 31.6 | -0.10 | 36.7 | 34.9 | -0.04 | 35.3 | 33.0 | -0.05 |
| 70 - 74 | 35.8 | 29.9 | -0.12 | 34.2 | 34.5 | 0.01 | 32.5 | 31.3 | -0.03 |
| 75 - 79 | 16.8 | 19.2 | 0.06 | 16.7 | 17.0 | 0.01 | 16.0 | 18.2 | 0.06 |
| 80 - 84 | 5.6 | 10.5 | 0.18 | 6.0 | 6.1 | 0.00 | 8.2 | 8.9 | 0.02 |
| 85 - 89 | 1.6 | 4.6 | 0.17 | 2.0 | 3.0 | 0.06 | 2.5 | 4.1 | 0.09 |
| 90 - 94 | 0.3 | 1.3 | 0.12 | 0.3 | 1.7 | 0.14 | 0.7 | 1.5 | 0.08 |
| 95 - 99 | <0.2 | 0.2 | 0.04 | <0.2 | <0.3 | 0.06 | <0.2 | 0.2 | 0.05 |
| **Medical history: General** | | | | | | | | | |
| Acute respiratory disease | 21.2 | 21.1 | 0.00 | 22.3 | 21.6 | -0.02 | 24.2 | 21.8 | -0.06 |
| Attention deficit hyperactivity disorder | 0.2 | 0.2 | -0.01 | 0.2 | 0.2 | 0.01 | 0.2 | 0.2 | 0.01 |
| Chronic liver disease | 1.8 | 1.9 | 0.01 | 1.9 | 1.7 | -0.01 | 1.7 | 1.9 | 0.01 |
| Chronic obstructive lung disease | 10.2 | 10.8 | 0.02 | 10.2 | 10.7 | 0.02 | 10.9 | 11.3 | 0.01 |
| Crohn’s disease | 0.4 | 0.3 | -0.01 | 0.4 | <0.3 | -0.06 | 0.4 | 0.3 | -0.01 |
| Dementia | 1.1 | 2.4 | 0.10 | 1.1 | 2.2 | 0.08 | 0.9 | 2.6 | 0.14 |
| Depressive disorder | 6.5 | 6.0 | -0.02 | 6.4 | 6.9 | 0.02 | 7.1 | 7.3 | 0.00 |
| Gastroesophageal reflux disease | 15.2 | 11.0 | -0.12 | 14.8 | 14.1 | -0.02 | 13.8 | 14.6 | 0.02 |
| Gastrointestinal hemorrhage | 2.6 | 2.9 | 0.02 | 2.6 | 2.3 | -0.02 | 2.5 | 2.6 | 0.01 |
| Human immunodeficiency virus infection | <0.2 | 0.2 | 0.00 | <0.2 | <0.3 | 0.01 | <0.2 | 0.2 | 0.04 |
| Hyperlipidemia | 83.0 | 64.5 | -0.43 | 81.8 | 83.1 | 0.03 | 80.5 | 79.9 | -0.01 |
| Hypertensive disorder | 84.2 | 73.9 | -0.26 | 83.2 | 84.5 | 0.04 | 83.6 | 82.8 | -0.02 |
| Lesion of liver | 1.3 | 0.7 | -0.06 | 1.1 | 0.8 | -0.03 | 1.4 | 0.7 | -0.07 |
| Obesity | 22.9 | 9.7 | -0.36 | 20.5 | 19.5 | -0.02 | 17.7 | 17.0 | -0.02 |
| Osteoarthritis | 29.4 | 27.0 | -0.05 | 29.8 | 29.7 | 0.00 | 29.5 | 29.6 | 0.00 |
| Pneumonia | 4.0 | 5.1 | 0.05 | 3.9 | 4.4 | 0.03 | 4.5 | 4.8 | 0.01 |
| Psoriasis | 2.1 | 1.7 | -0.03 | 1.9 | 1.9 | 0.00 | 1.9 | 1.9 | 0.00 |
| Renal impairment | 13.1 | 14.5 | 0.04 | 13.1 | 12.1 | -0.03 | 13.2 | 16.2 | 0.09 |
| Rheumatoid arthritis | 1.2 | 1.3 | 0.01 | 1.2 | 1.6 | 0.04 | 1.1 | 1.4 | 0.03 |
| Schizophrenia | <0.2 | 0.1 | 0.00 | <0.2 | <0.3 | -0.01 | <0.2 | 0.1 | 0.02 |
| Ulcerative colitis | 0.3 | 0.5 | 0.03 | 0.3 | 0.6 | 0.05 | 0.3 | 0.6 | 0.04 |
| Urinary tract infectious disease | 4.1 | 6.3 | 0.10 | 4.3 | 5.5 | 0.06 | 5.7 | 6.3 | 0.02 |
| Viral hepatitis C | 0.7 | 0.3 | -0.06 | 0.7 | 0.5 | -0.02 | 0.5 | 0.5 | 0.00 |
| Visual system disorder | 52.9 | 49.5 | -0.07 | 51.7 | 51.7 | 0.00 | 51.8 | 51.3 | -0.01 |
| **Medical history: Cardiovascular disease** | | | | | | | | | |
| Atrial fibrillation | 15.0 | 13.2 | -0.05 | 13.9 | 14.4 | 0.01 | 13.0 | 14.5 | 0.05 |
| Cerebrovascular disease | 11.2 | 13.0 | 0.06 | 10.7 | 11.0 | 0.01 | 12.3 | 12.2 | -0.01 |
| Coronary arteriosclerosis | 36.4 | 32.2 | -0.09 | 33.2 | 33.6 | 0.01 | 31.3 | 32.7 | 0.03 |
| Heart disease | 54.2 | 50.5 | -0.07 | 50.9 | 50.1 | -0.02 | 50.3 | 50.3 | 0.00 |
| Heart failure | 10.6 | 9.4 | -0.04 | 9.1 | 9.4 | 0.01 | 8.1 | 9.5 | 0.05 |
| Ischemic heart disease | 13.8 | 13.6 | -0.01 | 13.6 | 13.8 | 0.01 | 13.8 | 13.5 | -0.01 |
| Peripheral vascular disease | 15.0 | 13.1 | -0.06 | 13.9 | 13.4 | -0.01 | 15.1 | 14.6 | -0.01 |
| Pulmonary embolism | 0.9 | 0.9 | -0.01 | 1.0 | 0.9 | -0.01 | 1.1 | 1.0 | -0.01 |
| Venous thrombosis | 1.9 | 2.2 | 0.03 | 2.0 | 2.2 | 0.01 | 2.4 | 2.2 | -0.01 |
| **Medical history: Neoplasms** | | | | | | | | | |
| Hematologic neoplasm | 2.0 | 2.1 | 0.00 | 1.9 | 2.3 | 0.02 | 2.1 | 2.4 | 0.02 |
| Malignant lymphoma | 1.4 | 1.1 | -0.03 | 1.3 | 1.2 | -0.01 | 1.0 | 1.3 | 0.03 |
| Malignant neoplasm of anorectum | 0.2 | 0.4 | 0.03 | 0.2 | <0.3 | 0.01 | <0.2 | 0.3 | 0.04 |
| Malignant neoplastic disease | 18.4 | 22.0 | 0.09 | 18.7 | 18.5 | 0.00 | 19.6 | 21.2 | 0.04 |
| Malignant tumor of breast | <0.2 | 0.1 | 0.02 | <0.2 | <0.3 | 0.04 | <0.2 | <0.1 | 0.02 |
| Malignant tumor of colon | 0.5 | 1.0 | 0.06 | 0.4 | 0.8 | 0.05 | 0.3 | 1.0 | 0.09 |
| Malignant tumor of lung | 0.3 | 0.7 | 0.05 | 0.4 | 0.3 | 0.00 | 0.3 | 0.7 | 0.06 |
| Malignant tumor of urinary bladder | 1.4 | 2.0 | 0.04 | 1.5 | 1.9 | 0.03 | 1.5 | 2.1 | 0.05 |
| Primary malignant neoplasm of prostate | 7.1 | 8.4 | 0.05 | 7.4 | 7.5 | 0.00 | 8.1 | 8.4 | 0.01 |
| **Medication use** | | | | | | | | | |

| (Continued) | Before ajustment | | | After matching | | | After stratification | | |
|---|---|---|---|---|---|---|---|---|---|
| Characteristic | T (%) | C (%) | StdDiff | T (%) | C (%) | StdDiff | T (%) | C (%) | StdDiff |
| Agents acting on the renin-angiotensin system | 77.6 | 76.3 | -0.03 | 77.3 | 76.4 | -0.02 | 76.5 | 76.4 | 0.00 |
| Antibacterials for systemic use | 59.8 | 58.7 | -0.02 | 60.3 | 57.9 | -0.05 | 61.4 | 58.7 | -0.05 |
| Antidepressants | 18.4 | 18.8 | 0.01 | 18.5 | 18.9 | 0.01 | 20.9 | 19.0 | -0.05 |
| Antiepileptics | 14.3 | 12.8 | -0.04 | 14.2 | 13.7 | -0.02 | 14.2 | 13.7 | -0.01 |
| Antiinflammatory and antirheumatic products | 26.0 | 25.5 | -0.01 | 26.6 | 26.5 | 0.00 | 25.8 | 26.9 | 0.03 |
| Antineoplastic agents | 7.7 | 8.2 | 0.02 | 8.0 | 7.6 | -0.01 | 8.9 | 8.0 | -0.03 |
| Antipsoriatics | 0.5 | 0.8 | 0.03 | 0.5 | 0.5 | 0.00 | 0.3 | 0.6 | 0.04 |
| Antithrombotic agents | 30.2 | 28.4 | -0.04 | 28.7 | 27.2 | -0.03 | 27.2 | 28.3 | 0.02 |
| Beta blocking agents | 51.1 | 50.3 | -0.02 | 49.0 | 49.7 | 0.01 | 49.4 | 50.2 | 0.01 |
| Calcium channel blockers | 32.9 | 32.8 | 0.00 | 32.1 | 32.8 | 0.01 | 31.0 | 33.6 | 0.06 |
| Diuretics | 45.1 | 45.7 | 0.01 | 44.6 | 43.7 | -0.02 | 42.4 | 43.7 | 0.03 |
| Drugs for acid related disorders | 30.5 | 29.8 | -0.01 | 30.2 | 30.0 | 0.00 | 31.3 | 30.4 | -0.02 |
| Drugs for obstructive airway diseases | 41.1 | 38.5 | -0.06 | 41.0 | 40.2 | -0.02 | 40.6 | 40.8 | 0.00 |
| Immunosuppressants | 2.7 | 2.4 | -0.01 | 2.5 | 3.0 | 0.03 | 2.3 | 2.7 | 0.03 |
| Lipid modifying agents | 85.8 | 82.7 | -0.09 | 84.4 | 85.8 | 0.04 | 82.8 | 83.9 | 0.03 |
| Opioids | 24.1 | 25.4 | 0.03 | 24.5 | 23.7 | -0.02 | 26.6 | 25.2 | -0.03 |
| Psycholeptics | 20.5 | 22.4 | 0.05 | 20.8 | 20.6 | 0.00 | 24.7 | 21.3 | -0.08 |
| Psychostimulants | 1.3 | 1.1 | -0.02 | 1.4 | 1.0 | -0.03 | 1.3 | 0.9 | -0.04 |

##### 2.2.5 MDCD

**Supplementary Table 167:** Baseline patient characteristics for SGLT2I (male) (T) and DPP4I (male) (C) new-users in the MDCD data source. We report proportion of initiators satisfying selected base-line characteristics and the standardized difference of population proportions (StdDiff) before and after propensity score adjustment. Less extreme StdDiffs through matching and stratification suggest improved balance between patient cohorts through adjustment.

| | Before adjustment | | | After matching | | | After stratification | | |
|---|---|---|---|---|---|---|---|---|---|
| Characteristic | T (%) | C (%) | StdDiff | T (%) | C (%) | StdDiff | T (%) | C (%) | StdDiff |
| **Age group** | | | | | | | | | |
| 15 - 19 | 0.7 | 0.7 | 0.01 | 0.7 | 0.8 | 0.02 | 0.7 | 0.8 | 0.01 |
| 20 - 24 | 1.1 | 1.2 | 0.01 | 1.0 | 1.3 | 0.03 | 1.1 | 1.2 | 0.01 |
| 25 - 29 | 2.5 | 2.3 | -0.02 | 2.4 | 2.2 | -0.01 | 2.2 | 2.2 | 0.00 |
| 30 - 34 | 5.3 | 4.0 | -0.06 | 5.2 | 3.9 | -0.06 | 5.3 | 4.1 | -0.06 |
| 35 - 39 | 8.6 | 7.0 | -0.06 | 8.6 | 7.8 | -0.03 | 8.5 | 7.3 | -0.04 |
| 40 - 44 | 11.1 | 10.7 | -0.01 | 10.9 | 11.6 | 0.03 | 10.7 | 11.1 | 0.02 |
| 45 - 49 | 16.3 | 14.5 | -0.05 | 16.1 | 15.4 | -0.02 | 15.8 | 14.7 | -0.03 |
| 50 - 54 | 15.9 | 16.6 | 0.02 | 16.1 | 15.6 | -0.02 | 16.2 | 16.3 | 0.00 |
| 55 - 59 | 20.0 | 18.3 | -0.04 | 20.3 | 18.0 | -0.06 | 20.8 | 18.8 | -0.05 |
| 60 - 64 | 13.9 | 15.8 | 0.05 | 13.9 | 15.8 | 0.05 | 14.0 | 15.8 | 0.05 |
| 65 - 69 | 2.7 | 3.9 | 0.06 | 2.8 | 3.5 | 0.04 | 3.0 | 3.6 | 0.04 |
| 70 - 74 | 1.3 | 2.5 | 0.09 | 1.3 | 2.0 | 0.06 | 1.0 | 2.1 | 0.09 |
| 75 - 79 | 0.3 | 1.4 | 0.11 | 0.4 | 1.1 | 0.08 | <0.3 | 0.9 | 0.09 |
| 80 - 84 | 0.4 | 1.0 | 0.07 | 0.4 | 0.8 | 0.05 | 0.6 | 0.8 | 0.02 |
| **Race** | | | | | | | | | |
| Black or African American | 19.5 | 19.6 | 0.00 | 19.7 | 18.4 | -0.03 | 22.2 | 19.8 | -0.06 |
| White | 65.2 | 61.7 | -0.07 | 64.6 | 64.4 | 0.00 | 61.5 | 61.8 | 0.01 |
| **Ethnicity** | | | | | | | | | |
| Hispanic or Latino | 1.9 | 2.8 | 0.06 | 1.9 | 2.8 | 0.06 | 2.1 | 3.0 | 0.06 |
| **Medical history: General** | | | | | | | | | |
| Acute respiratory disease | 27.3 | 27.7 | 0.01 | 27.0 | 26.8 | 0.00 | 27.5 | 27.2 | -0.01 |
| Attention deficit hyperactivity disorder | 2.9 | 1.9 | -0.07 | 2.8 | 2.2 | -0.04 | 3.2 | 2.1 | -0.07 |
| Chronic liver disease | 3.5 | 5.9 | 0.12 | 3.5 | 3.7 | 0.01 | 4.2 | 4.8 | 0.03 |
| Chronic obstructive lung disease | 17.7 | 19.4 | 0.04 | 17.5 | 18.0 | 0.01 | 17.3 | 18.3 | 0.03 |
| Crohn’s disease | 0.3 | 0.1 | -0.04 | 0.4 | <0.4 | -0.04 | <0.3 | <0.1 | -0.03 |
| Dementia | 0.9 | 2.3 | 0.11 | 1.0 | 1.9 | 0.08 | 0.9 | 2.1 | 0.10 |
| Depressive disorder | 27.0 | 25.5 | -0.03 | 26.6 | 27.1 | 0.01 | 25.9 | 26.1 | 0.00 |
| Gastroesophageal reflux disease | 23.0 | 22.7 | -0.01 | 22.9 | 23.6 | 0.02 | 22.9 | 23.5 | 0.01 |
| Gastrointestinal hemorrhage | 2.9 | 3.7 | 0.04 | 3.0 | 3.4 | 0.02 | 3.6 | 3.5 | 0.00 |
| Human immunodeficiency virus infection | 0.9 | 1.0 | 0.01 | 0.9 | 1.3 | 0.03 | 1.1 | 1.1 | 0.00 |
| Hyperlipidemia | 69.7 | 69.6 | 0.00 | 69.8 | 69.7 | 0.00 | 69.5 | 70.1 | 0.01 |
| Hypertensive disorder | 77.7 | 78.5 | 0.02 | 77.6 | 77.3 | -0.01 | 77.9 | 78.6 | 0.02 |
| Lesion of liver | 1.3 | 2.3 | 0.07 | 1.3 | 2.0 | 0.06 | 1.6 | 2.3 | 0.05 |
| Obesity | 36.2 | 32.2 | -0.09 | 35.2 | 39.1 | 0.08 | 32.7 | 34.7 | 0.04 |
| Osteoarthritis | 31.1 | 32.9 | 0.04 | 30.8 | 31.5 | 0.01 | 32.2 | 32.8 | 0.01 |
| Pneumonia | 4.1 | 6.1 | 0.09 | 4.1 | 6.0 | 0.09 | 4.8 | 5.6 | 0.04 |
| Psoriasis | 1.1 | 1.6 | 0.04 | 1.2 | 1.3 | 0.01 | 1.0 | 1.6 | 0.05 |
| Renal impairment | 6.7 | 10.2 | 0.13 | 7.0 | 7.6 | 0.02 | 9.2 | 9.8 | 0.02 |
| Rheumatoid arthritis | 0.8 | 1.3 | 0.05 | 0.8 | 1.5 | 0.07 | 0.8 | 1.3 | 0.06 |
| Schizophrenia | 4.4 | 5.4 | 0.04 | 4.4 | 4.8 | 0.02 | 5.2 | 5.1 | -0.01 |
| Ulcerative colitis | 0.3 | 0.3 | 0.00 | 0.4 | <0.4 | -0.06 | <0.3 | 0.2 | -0.01 |
| Urinary tract infectious disease | 2.5 | 4.9 | 0.13 | 2.5 | 4.1 | 0.09 | 3.3 | 4.4 | 0.06 |
| Viral hepatitis C | 2.0 | 3.4 | 0.09 | 2.1 | 2.5 | 0.03 | 2.0 | 3.1 | 0.07 |
| Visual system disorder | 41.1 | 40.8 | -0.01 | 41.0 | 40.9 | 0.00 | 42.2 | 40.9 | -0.03 |
| **Medical history: Cardiovascular disease** | | | | | | | | | |
| Atrial fibrillation | 4.1 | 4.6 | 0.02 | 4.1 | 4.1 | 0.00 | 3.7 | 4.4 | 0.03 |
| Cerebrovascular disease | 3.8 | 5.3 | 0.07 | 3.8 | 4.1 | 0.02 | 3.5 | 4.7 | 0.06 |
| Coronary arteriosclerosis | 15.7 | 16.7 | 0.03 | 15.6 | 14.3 | -0.04 | 15.5 | 15.7 | 0.00 |
| Heart disease | 28.9 | 32.0 | 0.07 | 28.8 | 28.9 | 0.00 | 28.1 | 30.5 | 0.05 |
| Heart failure | 9.6 | 9.9 | 0.01 | 9.5 | 8.9 | -0.02 | 9.7 | 9.4 | -0.01 |
| Ischemic heart disease | 10.2 | 11.4 | 0.04 | 10.1 | 10.2 | 0.00 | 9.9 | 10.9 | 0.03 |
| Peripheral vascular disease | 9.0 | 10.1 | 0.04 | 8.9 | 8.6 | -0.01 | 7.8 | 9.3 | 0.06 |
| Pulmonary embolism | 0.9 | 1.2 | 0.03 | 0.9 | 1.1 | 0.01 | 0.9 | 1.2 | 0.03 |
| Venous thrombosis | 1.7 | 1.6 | 0.00 | 1.7 | 1.4 | -0.02 | 1.9 | 1.4 | -0.04 |
| **Medical history: Neoplasms** | | | | | | | | | |
| Hematologic neoplasm | 0.7 | 1.1 | 0.04 | 0.8 | 1.1 | 0.03 | 0.8 | 1.1 | 0.02 |
| Malignant lymphoma | <0.3 | 0.7 | 0.06 | <0.3 | <0.4 | -0.01 | <0.3 | 0.6 | 0.05 |

| (Continued) | Before ajustment | | | After matching | | | After stratification | | |
|---|---|---|---|---|---|---|---|---|---|
| Characteristic | T (%) | C (%) | StdDiff | T (%) | C (%) | StdDiff | T (%) | C (%) | StdDiff |
| Malignant neoplasm of anorectum | <0.3 | 0.2 | 0.02 | <0.3 | <0.4 | 0.03 | <0.3 | 0.2 | 0.03 |
| Malignant neoplastic disease | 3.5 | 5.6 | 0.10 | 3.6 | 4.6 | 0.05 | 3.7 | 5.1 | 0.07 |
| Malignant tumor of breast | <0.3 | <0.1 | -0.02 | <0.3 | <0.1 | -0.01 | <0.3 | <0.1 | -0.01 |
| Malignant tumor of colon | 0.3 | 0.5 | 0.03 | <0.3 | 0.8 | 0.06 | 0.3 | 0.5 | 0.03 |
| Malignant tumor of lung | <0.3 | 0.5 | 0.06 | <0.3 | 0.4 | 0.05 | <0.3 | 0.4 | 0.06 |
| Malignant tumor of urinary bladder | <0.3 | 0.3 | 0.01 | <0.3 | 0.3 | 0.03 | <0.3 | 0.3 | 0.03 |
| Primary malignant neoplasm of prostate | 0.9 | 1.3 | 0.03 | 0.9 | 1.0 | 0.00 | 1.1 | 1.2 | 0.00 |
| **Medication use** | | | | | | | | | |
| Agents acting on the renin-angiotensin system | 67.9 | 70.0 | 0.04 | 67.6 | 67.0 | -0.01 | 68.6 | 69.0 | 0.01 |
| Antibacterials for systemic use | 57.3 | 59.2 | 0.04 | 57.0 | 56.1 | -0.02 | 58.7 | 58.1 | -0.01 |
| Antidepressants | 42.1 | 43.7 | 0.03 | 42.0 | 43.8 | 0.04 | 42.5 | 43.5 | 0.02 |
| Antiepileptics | 37.3 | 37.1 | 0.00 | 37.1 | 37.2 | 0.00 | 36.0 | 37.5 | 0.03 |
| Antiinflammatory and antirheumatic products | 55.7 | 53.9 | -0.04 | 55.8 | 56.3 | 0.01 | 56.4 | 54.7 | -0.04 |
| Antineoplastic agents | 2.6 | 4.3 | 0.09 | 2.6 | 3.9 | 0.07 | 2.7 | 4.1 | 0.07 |
| Antipsoriatics | <0.3 | 0.4 | 0.03 | <0.3 | <0.4 | 0.02 | <0.3 | 0.4 | 0.03 |
| Antithrombotic agents | 26.5 | 29.7 | 0.07 | 26.7 | 27.8 | 0.03 | 26.5 | 29.0 | 0.06 |
| Beta blocking agents | 35.5 | 38.0 | 0.05 | 35.7 | 34.4 | -0.03 | 36.4 | 37.0 | 0.01 |
| Calcium channel blockers | 24.4 | 26.2 | 0.04 | 24.4 | 25.2 | 0.02 | 25.6 | 25.7 | 0.00 |
| Diuretics | 39.8 | 41.2 | 0.03 | 39.6 | 38.0 | -0.03 | 40.3 | 39.8 | -0.01 |
| Drugs for acid related disorders | 41.2 | 41.7 | 0.01 | 41.2 | 39.1 | -0.04 | 41.6 | 40.9 | -0.01 |
| Drugs for obstructive airway diseases | 48.5 | 50.2 | 0.03 | 48.5 | 48.7 | 0.00 | 46.8 | 49.5 | 0.05 |
| Immunosuppressants | 1.7 | 1.8 | 0.01 | 1.8 | 2.1 | 0.03 | 1.5 | 1.9 | 0.03 |
| Lipid modifying agents | 70.9 | 71.2 | 0.01 | 70.8 | 71.1 | 0.01 | 69.1 | 71.1 | 0.04 |
| Opioids | 39.2 | 41.4 | 0.04 | 39.1 | 38.6 | -0.01 | 41.1 | 39.7 | -0.03 |
| Psycholeptics | 42.1 | 42.6 | 0.01 | 41.6 | 41.0 | -0.01 | 42.4 | 41.3 | -0.02 |
| Psychostimulants | 4.2 | 3.7 | -0.03 | 4.3 | 3.1 | -0.06 | 4.6 | 3.5 | -0.06 |

##### 2.2.6 Open Claims

**Supplementary Table 168:** Baseline patient characteristics for SGLT2I (male) (T) and DPP4I (male) (C) new-users in the Open Claims data source. We report proportion of initiators satisfying selected base-line characteristics and the standardized difference of population proportions (StdDiff) before and after propensity score adjustment. Less extreme StdDiffs through matching and stratification suggest improved balance between patient cohorts through adjustment.

| | Before adjustment | | | After matching | | | After stratification | | |
|---|---|---|---|---|---|---|---|---|---|
| Characteristic | T (%) | C (%) | StdDiff | T (%) | C (%) | StdDiff | T (%) | C (%) | StdDiff |
| **Age group** | | | | | | | | | |
| 15 - 19 | 0.1 | 0.1 | 0.00 | 0.1 | 0.1 | 0.00 | 0.1 | 0.1 | 0.00 |
| 20 - 24 | 0.3 | 0.2 | 0.00 | 0.3 | 0.3 | 0.00 | 0.3 | 0.3 | 0.00 |
| 25 - 29 | 0.6 | 0.6 | 0.00 | 0.6 | 0.6 | 0.00 | 0.5 | 0.6 | 0.00 |
| 30 - 34 | 1.5 | 1.3 | -0.01 | 1.5 | 1.5 | 0.00 | 1.5 | 1.4 | -0.01 |
| 35 - 39 | 3.3 | 3.0 | -0.02 | 3.4 | 3.4 | 0.00 | 3.2 | 3.2 | 0.00 |
| 40 - 44 | 6.1 | 5.5 | -0.03 | 6.1 | 6.3 | 0.01 | 5.8 | 6.0 | 0.01 |
| 45 - 49 | 10.0 | 8.9 | -0.04 | 10.2 | 10.1 | 0.00 | 9.6 | 9.5 | 0.00 |
| 50 - 54 | 13.9 | 12.5 | -0.04 | 13.3 | 13.1 | -0.01 | 13.3 | 13.1 | -0.01 |
| 55 - 59 | 16.5 | 15.2 | -0.04 | 16.5 | 16.6 | 0.00 | 16.1 | 16.0 | 0.00 |
| 60 - 64 | 17.1 | 15.5 | -0.04 | 16.5 | 16.8 | 0.01 | 16.4 | 16.3 | 0.00 |
| 65 - 69 | 13.3 | 13.9 | 0.02 | 13.2 | 13.2 | 0.00 | 13.6 | 13.5 | 0.00 |
| 70 - 74 | 8.9 | 10.4 | 0.05 | 8.9 | 8.9 | 0.00 | 8.9 | 8.9 | 0.00 |
| 75 - 79 | 5.4 | 9.3 | 0.15 | 5.8 | 5.3 | -0.02 | 7.0 | 7.3 | 0.01 |
| 80 - 84 | 2.9 | 3.6 | 0.04 | 3.1 | 3.3 | 0.01 | 3.1 | 3.3 | 0.01 |
| 85 - 89 | 0.2 | 0.1 | -0.04 | 0.1 | 0.1 | 0.01 | 0.1 | 0.2 | 0.01 |
| **Medical history: General** | | | | | | | | | |
| Acute respiratory disease | 11.0 | 12.3 | 0.04 | 11.4 | 11.3 | 0.00 | 11.9 | 11.7 | -0.01 |
| Attention deficit hyperactivity disorder | 0.6 | 0.4 | -0.02 | 0.5 | 0.5 | 0.00 | 0.5 | 0.5 | 0.00 |
| Chronic liver disease | 1.0 | 1.5 | 0.05 | 1.1 | 1.1 | 0.00 | 1.3 | 1.3 | 0.00 |
| Chronic obstructive lung disease | 4.2 | 5.0 | 0.04 | 4.1 | 4.1 | 0.00 | 4.6 | 4.6 | 0.00 |
| Crohn’s disease | 0.1 | 0.1 | 0.00 | 0.1 | 0.1 | 0.00 | 0.2 | 0.1 | 0.00 |
| Dementia | 0.4 | 0.8 | 0.06 | 0.4 | 0.4 | 0.00 | 0.6 | 0.6 | 0.01 |
| Depressive disorder | 4.7 | 4.9 | 0.01 | 5.0 | 4.8 | -0.01 | 5.0 | 4.8 | -0.01 |
| Gastroesophageal reflux disease | 6.9 | 7.2 | 0.01 | 7.3 | 7.0 | -0.01 | 7.3 | 7.0 | -0.01 |
| Gastrointestinal hemorrhage | 1.1 | 1.4 | 0.03 | 1.2 | 1.1 | -0.01 | 1.4 | 1.3 | -0.01 |
| Human immunodeficiency virus infection | 0.4 | 0.5 | 0.01 | 0.4 | 0.4 | 0.01 | 0.4 | 0.4 | 0.01 |
| Hyperlipidemia | 46.0 | 47.3 | 0.03 | 45.8 | 46.0 | 0.00 | 46.7 | 46.8 | 0.00 |
| Hypertensive disorder | 49.9 | 51.6 | 0.03 | 49.6 | 49.6 | 0.00 | 50.8 | 50.6 | 0.00 |
| Lesion of liver | 0.6 | 0.7 | 0.02 | 0.6 | 0.6 | 0.00 | 0.7 | 0.7 | 0.00 |
| Obesity | 11.3 | 9.4 | -0.06 | 10.6 | 10.8 | 0.01 | 10.6 | 10.8 | 0.01 |
| Osteoarthritis | 11.7 | 13.1 | 0.04 | 11.9 | 12.0 | 0.00 | 12.8 | 12.6 | -0.01 |
| Pneumonia | 2.0 | 2.3 | 0.02 | 1.9 | 1.8 | -0.01 | 2.2 | 2.1 | -0.01 |
| Psoriasis | 1.0 | 0.9 | -0.01 | 1.0 | 1.0 | 0.00 | 1.0 | 1.0 | 0.00 |
| Renal impairment | 5.3 | 7.3 | 0.08 | 5.1 | 4.9 | -0.01 | 6.1 | 6.2 | 0.00 |
| Rheumatoid arthritis | 0.5 | 0.6 | 0.01 | 0.5 | 0.5 | 0.00 | 0.6 | 0.6 | 0.00 |
| Schizophrenia | 0.3 | 0.4 | 0.03 | 0.3 | 0.4 | 0.01 | 0.4 | 0.4 | 0.01 |
| Ulcerative colitis | 0.2 | 0.2 | 0.00 | 0.2 | 0.2 | 0.00 | 0.2 | 0.2 | 0.00 |
| Urinary tract infectious disease | 1.8 | 2.9 | 0.08 | 2.0 | 2.0 | 0.00 | 2.6 | 2.5 | -0.01 |
| Viral hepatitis C | 0.3 | 0.5 | 0.03 | 0.3 | 0.4 | 0.00 | 0.4 | 0.4 | 0.01 |
| Visual system disorder | 15.5 | 17.9 | 0.06 | 16.0 | 15.9 | 0.00 | 16.9 | 16.8 | 0.00 |
| **Medical history: Cardiovascular disease** | | | | | | | | | |
| Atrial fibrillation | 4.5 | 4.7 | 0.01 | 3.9 | 4.0 | 0.00 | 3.9 | 4.0 | 0.00 |
| Coronary arteriosclerosis | 11.9 | 11.2 | -0.02 | 10.2 | 9.9 | -0.01 | 11.4 | 10.8 | -0.02 |
| Heart disease | 21.1 | 20.8 | -0.01 | 18.6 | 18.4 | -0.01 | 20.7 | 19.9 | -0.02 |
| Heart failure | 5.2 | 4.2 | -0.05 | 3.6 | 3.5 | 0.00 | 4.5 | 4.1 | -0.02 |
| Ischemic heart disease | 5.5 | 5.0 | -0.02 | 4.5 | 4.4 | -0.01 | 5.2 | 4.8 | -0.01 |
| Peripheral vascular disease | 4.7 | 4.9 | 0.01 | 4.3 | 4.3 | 0.00 | 4.8 | 4.7 | 0.00 |
| Pulmonary embolism | 0.4 | 0.5 | 0.00 | 0.4 | 0.4 | 0.00 | 0.5 | 0.4 | -0.01 |
| Venous thrombosis | 0.8 | 1.0 | 0.02 | 0.8 | 0.8 | 0.00 | 0.9 | 0.9 | -0.01 |
| **Medical history: Neoplasms** | | | | | | | | | |
| Hematologic neoplasm | 0.6 | 0.8 | 0.03 | 0.6 | 0.6 | 0.00 | 0.7 | 0.7 | 0.00 |
| Malignant lymphoma | 0.4 | 0.5 | 0.02 | 0.4 | 0.3 | 0.00 | 0.4 | 0.4 | 0.00 |
| Malignant neoplasm of anorectum | 0.1 | 0.2 | 0.01 | 0.1 | 0.1 | 0.00 | 0.2 | 0.1 | -0.01 |
| Malignant neoplastic disease | 4.9 | 6.7 | 0.08 | 5.2 | 5.0 | -0.01 | 5.9 | 5.8 | -0.01 |
| Malignant tumor of colon | 0.2 | 0.4 | 0.02 | 0.3 | 0.3 | 0.00 | 0.3 | 0.3 | 0.00 |
| Malignant tumor of lung | 0.2 | 0.2 | 0.02 | 0.2 | 0.2 | 0.00 | 0.2 | 0.2 | 0.01 |
| Malignant tumor of urinary bladder | 0.3 | 0.5 | 0.04 | 0.3 | 0.3 | 0.00 | 0.4 | 0.4 | 0.00 |

| (Continued) | Before ajustment | | | After matching | | | After stratification | | |
|---|---|---|---|---|---|---|---|---|---|
| Characteristic | T (%) | C (%) | StdDiff | T (%) | C (%) | StdDiff | T (%) | C (%) | StdDiff |
| Primary malignant neoplasm of prostate | 1.7 | 2.4 | 0.05 | 1.8 | 1.7 | 0.00 | 2.0 | 2.0 | 0.00 |
| **Medication use** | | | | | | | | | |
| Agents acting on the renin-angiotensin system | 71.6 | 71.0 | -0.01 | 70.7 | 70.6 | 0.00 | 70.9 | 70.8 | 0.00 |
| Antibacterials for systemic use | 48.9 | 51.3 | 0.05 | 49.9 | 49.8 | 0.00 | 50.9 | 50.2 | -0.01 |
| Antidepressants | 21.1 | 20.6 | -0.01 | 20.7 | 20.7 | 0.00 | 21.0 | 20.9 | 0.00 |
| Antiepileptics | 14.7 | 15.6 | 0.03 | 14.7 | 15.0 | 0.01 | 15.4 | 15.4 | 0.00 |
| Antiinflammatory and antirheumatic products | 32.1 | 33.0 | 0.02 | 32.5 | 32.6 | 0.00 | 33.0 | 32.7 | -0.01 |
| Antineoplastic agents | 4.0 | 4.2 | 0.01 | 3.9 | 4.0 | 0.00 | 4.1 | 4.2 | 0.00 |
| Antipsoriatics | 0.5 | 0.6 | 0.01 | 0.5 | 0.5 | 0.00 | 0.6 | 0.5 | 0.00 |
| Antithrombotic agents | 21.2 | 21.3 | 0.00 | 19.5 | 19.1 | -0.01 | 21.2 | 20.4 | -0.02 |
| Beta blocking agents | 35.1 | 33.8 | -0.03 | 32.4 | 32.5 | 0.00 | 33.8 | 33.5 | -0.01 |
| Calcium channel blockers | 25.8 | 26.0 | 0.00 | 25.3 | 25.1 | 0.00 | 25.7 | 25.7 | 0.00 |
| Diuretics | 36.3 | 35.3 | -0.02 | 34.5 | 34.7 | 0.00 | 34.5 | 34.7 | 0.00 |
| Drugs for acid related disorders | 27.4 | 28.7 | 0.03 | 27.1 | 27.1 | 0.00 | 28.4 | 28.0 | -0.01 |
| Drugs for obstructive airway diseases | 31.5 | 32.0 | 0.01 | 31.4 | 31.4 | 0.00 | 32.1 | 31.7 | -0.01 |
| Immunosuppressants | 1.9 | 1.9 | 0.00 | 1.9 | 1.9 | 0.00 | 2.0 | 2.0 | 0.00 |
| Lipid modifying agents | 75.3 | 73.6 | -0.04 | 73.7 | 73.6 | 0.00 | 73.9 | 73.9 | 0.00 |
| Opioids | 20.7 | 23.0 | 0.06 | 21.5 | 21.3 | 0.00 | 22.5 | 21.9 | -0.01 |
| Psycholeptics | 17.4 | 18.8 | 0.04 | 17.5 | 17.5 | 0.00 | 18.5 | 18.1 | -0.01 |
| Psychostimulants | 2.3 | 1.9 | -0.03 | 2.2 | 2.2 | 0.00 | 2.2 | 2.1 | -0.01 |

##### 2.2.7 OEHR

**Supplementary Table 169:** Baseline patient characteristics for SGLT2I (male) (T) and DPP4I (male) (C) new-users in the OEHR data source. We report proportion of initiators satisfying selected base-line characteristics and the standardized difference of population proportions (StdDiff) before and after propensity score adjustment. Less extreme StdDiffs through matching and stratification suggest improved balance between patient cohorts through adjustment.

| | Before adjustment | | | After matching | | | After stratification | | |
|---|---|---|---|---|---|---|---|---|---|
| Characteristic | T (%) | C (%) | StdDiff | T (%) | C (%) | StdDiff | T (%) | C (%) | StdDiff |
| **Age group** | | | | | | | | | |
| 15 - 19 | 0.1 | 0.1 | -0.01 | 0.1 | 0.1 | 0.00 | 0.1 | 0.1 | -0.01 |
| 20 - 24 | 0.2 | 0.2 | 0.00 | 0.2 | 0.3 | 0.01 | 0.2 | 0.3 | 0.01 |
| 25 - 29 | 0.7 | 0.6 | 0.00 | 0.7 | 0.7 | 0.00 | 0.7 | 0.7 | 0.00 |
| 30 - 34 | 1.9 | 1.6 | -0.02 | 1.9 | 1.8 | 0.00 | 1.7 | 1.7 | 0.00 |
| 35 - 39 | 4.3 | 3.8 | -0.03 | 4.3 | 4.2 | 0.00 | 4.2 | 3.9 | -0.01 |
| 40 - 44 | 8.0 | 6.9 | -0.04 | 7.7 | 7.8 | 0.01 | 7.0 | 7.1 | 0.00 |
| 45 - 49 | 11.9 | 11.1 | -0.03 | 11.9 | 11.8 | 0.00 | 11.4 | 11.3 | 0.00 |
| 50 - 54 | 15.7 | 14.4 | -0.04 | 15.7 | 15.6 | 0.00 | 14.8 | 14.8 | 0.00 |
| 55 - 59 | 18.1 | 16.5 | -0.04 | 17.3 | 17.1 | -0.01 | 17.3 | 17.1 | -0.01 |
| 60 - 64 | 17.3 | 15.5 | -0.05 | 16.8 | 17.2 | 0.01 | 16.8 | 17.2 | 0.01 |
| 65 - 69 | 10.5 | 12.1 | 0.05 | 10.7 | 10.7 | 0.00 | 11.4 | 11.4 | 0.00 |
| 70 - 74 | 6.5 | 8.2 | 0.06 | 6.8 | 6.7 | 0.00 | 7.4 | 7.5 | 0.00 |
| 75 - 79 | 3.2 | 5.3 | 0.10 | 3.5 | 3.3 | -0.01 | 4.3 | 4.3 | 0.00 |
| 80 - 84 | 1.4 | 3.0 | 0.11 | 1.6 | 1.6 | 0.00 | 2.4 | 2.5 | 0.01 |
| 85 - 89 | 0.4 | 0.8 | 0.06 | 0.4 | 0.5 | 0.01 | 0.6 | 0.7 | 0.02 |
| **Race** | | | | | | | | | |
| Asian | 2.3 | 2.9 | 0.04 | 2.5 | 2.5 | 0.00 | 2.6 | 2.7 | 0.01 |
| Black or African American | 8.2 | 9.5 | 0.04 | 8.6 | 8.6 | 0.00 | 8.6 | 8.6 | 0.00 |
| White | 81.8 | 80.1 | -0.04 | 81.1 | 81.4 | 0.01 | 80.6 | 80.5 | 0.00 |
| **Ethnicity** | | | | | | | | | |
| Hispanic or Latino | 5.2 | 6.0 | 0.03 | 5.5 | 5.7 | 0.01 | 5.7 | 5.8 | 0.01 |
| Not Hispanic or Latino | 85.6 | 85.9 | 0.01 | 85.4 | 85.2 | -0.01 | 85.7 | 85.3 | -0.01 |
| **Medical history: General** | | | | | | | | | |
| Acute respiratory disease | 15.0 | 15.0 | 0.00 | 14.8 | 14.8 | 0.00 | 15.0 | 15.1 | 0.00 |
| Attention deficit hyperactivity disorder | 1.0 | 0.8 | -0.02 | 0.9 | 1.0 | 0.01 | 0.9 | 0.9 | 0.01 |
| Chronic liver disease | 1.9 | 2.6 | 0.05 | 2.0 | 2.1 | 0.01 | 2.2 | 2.3 | 0.00 |
| Chronic obstructive lung disease | 4.3 | 4.9 | 0.03 | 4.3 | 4.3 | 0.00 | 4.6 | 4.8 | 0.01 |
| Crohn’s disease | 0.2 | 0.2 | -0.01 | 0.2 | 0.1 | -0.02 | 0.2 | 0.2 | -0.01 |
| Dementia | 0.3 | 0.7 | 0.06 | 0.4 | 0.5 | 0.01 | 0.6 | 0.7 | 0.01 |
| Depressive disorder | 9.3 | 8.6 | -0.03 | 9.1 | 9.0 | 0.00 | 9.5 | 9.0 | -0.01 |
| Gastroesophageal reflux disease | 14.9 | 13.7 | -0.03 | 14.4 | 14.3 | 0.00 | 15.0 | 14.4 | -0.02 |
| Gastrointestinal hemorrhage | 1.2 | 1.5 | 0.03 | 1.2 | 1.2 | 0.01 | 1.3 | 1.4 | 0.01 |
| Human immunodeficiency virus infection | 0.3 | 0.3 | 0.01 | 0.3 | 0.4 | 0.02 | 0.3 | 0.4 | 0.00 |
| Hyperlipidemia | 69.9 | 70.1 | 0.00 | 68.9 | 69.2 | 0.01 | 68.9 | 69.2 | 0.01 |
| Hypertensive disorder | 68.7 | 67.1 | -0.03 | 67.6 | 67.5 | 0.00 | 68.1 | 67.8 | 0.00 |
| Lesion of liver | 0.7 | 0.8 | 0.01 | 0.7 | 0.7 | 0.00 | 0.7 | 0.8 | 0.01 |
| Obesity | 31.3 | 23.3 | -0.18 | 28.5 | 29.1 | 0.01 | 27.2 | 27.1 | 0.00 |
| Osteoarthritis | 13.4 | 13.8 | 0.01 | 13.3 | 13.4 | 0.00 | 14.0 | 13.9 | 0.00 |
| Pneumonia | 1.5 | 1.7 | 0.02 | 1.4 | 1.5 | 0.00 | 1.7 | 1.7 | 0.00 |
| Psoriasis | 1.4 | 1.4 | 0.00 | 1.4 | 1.6 | 0.02 | 1.5 | 1.5 | 0.00 |
| Renal impairment | 4.8 | 6.4 | 0.07 | 5.0 | 4.8 | -0.01 | 5.8 | 5.8 | 0.00 |
| Rheumatoid arthritis | 0.6 | 0.6 | 0.00 | 0.6 | 0.5 | -0.02 | 0.7 | 0.5 | -0.02 |
| Schizophrenia | 0.2 | 0.3 | 0.02 | 0.2 | 0.3 | 0.02 | 0.3 | 0.3 | 0.01 |
| Ulcerative colitis | 0.3 | 0.2 | -0.01 | 0.3 | 0.2 | -0.02 | 0.3 | 0.2 | -0.02 |
| Urinary tract infectious disease | 1.1 | 2.1 | 0.08 | 1.2 | 1.2 | 0.00 | 1.6 | 1.7 | 0.01 |
| Viral hepatitis C | 0.4 | 0.6 | 0.03 | 0.4 | 0.5 | 0.01 | 0.4 | 0.6 | 0.02 |
| Visual system disorder | 10.1 | 11.7 | 0.05 | 10.2 | 10.4 | 0.01 | 11.3 | 11.2 | -0.01 |
| **Medical history: Cardiovascular disease** | | | | | | | | | |
| Atrial fibrillation | 4.6 | 4.9 | 0.02 | 4.3 | 4.4 | 0.00 | 4.8 | 4.9 | 0.00 |
| Cerebrovascular disease | 2.6 | 2.8 | 0.01 | 2.5 | 2.3 | -0.01 | 2.7 | 2.6 | 0.00 |
| Coronary arteriosclerosis | 15.7 | 13.6 | -0.06 | 13.9 | 13.7 | -0.01 | 13.8 | 14.1 | 0.01 |
| Heart disease | 24.7 | 23.3 | -0.03 | 22.8 | 22.5 | -0.01 | 23.6 | 23.5 | 0.00 |
| Heart failure | 4.0 | 3.5 | -0.03 | 3.3 | 3.2 | 0.00 | 3.6 | 3.5 | -0.01 |
| Ischemic heart disease | 7.0 | 5.7 | -0.05 | 6.1 | 5.8 | -0.01 | 6.2 | 6.0 | 0.00 |
| Peripheral vascular disease | 5.8 | 4.7 | -0.05 | 5.0 | 5.0 | 0.00 | 5.2 | 5.1 | 0.00 |
| Pulmonary embolism | 0.4 | 0.5 | 0.01 | 0.4 | 0.4 | 0.01 | 0.4 | 0.5 | 0.01 |
| Venous thrombosis | 1.0 | 1.2 | 0.01 | 1.0 | 1.0 | 0.00 | 1.1 | 1.1 | 0.00 |

| (Continued) | Before ajustment | | | After matching | | | After stratification | | |
|---|---|---|---|---|---|---|---|---|---|
| Characteristic | T (%) | C (%) | StdDiff | T (%) | C (%) | StdDiff | T (%) | C (%) | StdDiff |
| **Medical history: Neoplasms** | | | | | | | | | |
| Hematologic neoplasm | 0.6 | 0.8 | 0.02 | 0.7 | 0.7 | 0.01 | 0.7 | 0.8 | 0.00 |
| Malignant lymphoma | 0.3 | 0.4 | 0.02 | 0.3 | 0.3 | 0.00 | 0.4 | 0.4 | 0.00 |
| Malignant neoplasm of anorectum | 0.1 | 0.1 | 0.00 | 0.1 | 0.1 | 0.00 | 0.2 | 0.1 | -0.02 |
| Malignant neoplastic disease | 4.7 | 6.5 | 0.08 | 5.0 | 5.1 | 0.01 | 5.8 | 5.9 | 0.00 |
| Malignant tumor of colon | 0.3 | 0.3 | 0.01 | 0.3 | 0.3 | 0.00 | 0.3 | 0.3 | -0.01 |
| Malignant tumor of lung | 0.1 | 0.2 | 0.03 | 0.1 | 0.2 | 0.01 | 0.1 | 0.2 | 0.01 |
| Malignant tumor of urinary bladder | 0.3 | 0.4 | 0.03 | 0.3 | 0.3 | 0.00 | 0.4 | 0.4 | 0.00 |
| Primary malignant neoplasm of prostate | 1.6 | 2.3 | 0.06 | 1.7 | 1.6 | 0.00 | 1.9 | 2.0 | 0.00 |
| **Medication use** | | | | | | | | | |
| Agents acting on the renin-angiotensin system | 69.5 | 68.0 | -0.03 | 68.7 | 68.6 | 0.00 | 68.6 | 68.5 | 0.00 |
| Antibacterials for systemic use | 36.4 | 36.9 | 0.01 | 36.3 | 36.0 | -0.01 | 37.1 | 36.7 | -0.01 |
| Antidepressants | 21.4 | 20.3 | -0.03 | 21.1 | 21.2 | 0.00 | 21.1 | 20.9 | -0.01 |
| Antiepileptics | 13.2 | 13.1 | 0.00 | 13.2 | 13.3 | 0.00 | 13.8 | 13.6 | 0.00 |
| Antiinflammatory and antirheumatic products | 54.6 | 53.7 | -0.02 | 53.9 | 53.6 | 0.00 | 54.3 | 54.1 | 0.00 |
| Antineoplastic agents | 3.0 | 3.5 | 0.02 | 3.0 | 3.0 | 0.00 | 3.5 | 3.2 | -0.01 |
| Antipsoriatics | 0.3 | 0.5 | 0.03 | 0.3 | 0.5 | 0.02 | 0.4 | 0.5 | 0.01 |
| Antithrombotic agents | 41.3 | 42.3 | 0.02 | 40.5 | 40.4 | 0.00 | 41.6 | 41.8 | 0.00 |
| Beta blocking agents | 32.6 | 31.8 | -0.02 | 31.3 | 31.0 | -0.01 | 31.3 | 31.0 | -0.01 |
| Calcium channel blockers | 23.2 | 22.4 | -0.02 | 22.6 | 22.4 | 0.00 | 22.4 | 22.9 | 0.01 |
| Diuretics | 34.3 | 34.2 | 0.00 | 33.5 | 33.2 | -0.01 | 33.8 | 33.7 | 0.00 |
| Drugs for acid related disorders | 29.7 | 29.8 | 0.00 | 29.2 | 29.4 | 0.00 | 30.0 | 30.0 | 0.00 |
| Immunosuppressants | 1.8 | 1.6 | -0.01 | 1.8 | 1.6 | -0.01 | 2.0 | 1.7 | -0.02 |
| Lipid modifying agents | 75.2 | 73.3 | -0.04 | 74.1 | 73.9 | 0.00 | 74.1 | 73.9 | 0.00 |
| Opioids | 21.4 | 23.0 | 0.04 | 21.4 | 21.3 | 0.00 | 22.9 | 22.4 | -0.01 |
| Psycholeptics | 21.6 | 22.9 | 0.03 | 21.5 | 21.7 | 0.01 | 22.7 | 22.5 | 0.00 |
| Psychostimulants | 2.8 | 2.3 | -0.04 | 2.7 | 2.7 | 0.00 | 2.6 | 2.5 | -0.01 |

### 2.2.8 VA

**Supplementary Table 170:** Baseline patient characteristics for SGLT2I (male) (T) and DPP4I (male) (C) new-users in the VA data source. We report proportion of initiators satisfying selected base-line characteristics and the standardized difference of population proportions (StdDiff) before and after propensity score adjustment. Less extreme StdDiffs through matching and stratification suggest improved balance between patient cohorts through adjustment.

| | Before adjustment | | | After matching | | | After stratification | | |
|---|---|---|---|---|---|---|---|---|---|
| Characteristic | T (%) | C (%) | StdDiff | T (%) | C (%) | StdDiff | T (%) | C (%) | StdDiff |
| **Age group** | | | | | | | | | |
| 25 - 29 | <0.1 | 0.1 | 0.01 | <0.1 | 0.1 | 0.02 | <0.1 | 0.1 | 0.02 |
| 30 - 34 | 0.4 | 0.7 | 0.03 | 0.5 | 0.6 | 0.02 | 0.4 | 0.6 | 0.03 |
| 35 - 39 | 1.5 | 1.7 | 0.01 | 1.7 | 1.7 | 0.00 | 1.9 | 1.7 | -0.01 |
| 40 - 44 | 2.5 | 2.8 | 0.01 | 2.8 | 3.0 | 0.01 | 2.7 | 2.8 | 0.01 |
| 45 - 49 | 4.5 | 5.0 | 0.02 | 4.8 | 4.4 | -0.02 | 5.0 | 4.8 | -0.01 |
| 50 - 54 | 8.3 | 8.4 | 0.01 | 8.7 | 8.6 | 0.00 | 9.1 | 8.3 | -0.03 |
| 55 - 59 | 10.8 | 10.5 | -0.01 | 11.3 | 11.0 | -0.01 | 10.8 | 10.6 | -0.01 |
| 60 - 64 | 13.3 | 12.7 | -0.02 | 13.2 | 13.5 | 0.01 | 13.4 | 12.7 | -0.02 |
| 65 - 69 | 15.5 | 18.8 | 0.09 | 15.7 | 15.4 | -0.01 | 18.0 | 17.7 | -0.01 |
| 70 - 74 | 26.7 | 22.6 | -0.10 | 25.8 | 25.2 | -0.01 | 24.3 | 23.9 | -0.01 |
| 75 - 79 | 11.6 | 8.8 | -0.09 | 10.4 | 10.6 | 0.00 | 8.5 | 9.8 | 0.04 |
| 80 - 84 | 3.1 | 4.4 | 0.07 | 3.3 | 3.3 | 0.00 | 3.6 | 4.0 | 0.02 |
| 85 - 89 | 1.3 | 2.4 | 0.08 | 1.5 | 1.8 | 0.02 | 1.8 | 2.1 | 0.02 |
| 90 - 94 | 0.3 | 0.9 | 0.07 | 0.4 | 0.6 | 0.03 | 0.4 | 0.7 | 0.05 |
| 95 - 99 | <0.1 | 0.2 | 0.05 | <0.1 | 0.2 | 0.06 | <0.1 | 0.2 | 0.06 |
| **Race** | | | | | | | | | |
| Asian | 1.3 | 1.6 | 0.03 | 1.2 | 1.4 | 0.02 | 0.9 | 1.4 | 0.05 |
| Black or African American | 15.5 | 16.0 | 0.01 | 17.0 | 15.4 | -0.04 | 17.0 | 15.4 | -0.04 |
| White | 75.3 | 74.3 | -0.02 | 75.6 | 75.4 | 0.00 | 75.0 | 75.2 | 0.00 |
| Unknown | 6.0 | 6.1 | 0.00 | 5.3 | 6.0 | 0.03 | 5.3 | 6.0 | 0.03 |
| Native Hawaiian or Other Pacific Islander | 1.0 | 1.2 | 0.01 | 1.1 | 1.1 | -0.01 | 1.1 | 1.1 | 0.01 |
| American Indian or Alaska Native | 0.9 | 0.8 | -0.01 | 0.9 | 0.9 | 0.00 | 0.8 | 0.9 | 0.01 |
| **Ethnicity** | | | | | | | | | |
| Hispanic or Latino | 6.6 | 6.2 | -0.01 | 6.5 | 6.4 | 0.00 | 5.6 | 6.3 | 0.03 |
| Not Hispanic or Latino | 90.8 | 91.4 | 0.02 | 90.8 | 91.2 | 0.01 | 92.0 | 91.5 | -0.02 |
| **Medical history: General** | | | | | | | | | |
| Acute respiratory disease | 8.1 | 8.1 | 0.00 | 7.8 | 7.5 | -0.01 | 8.0 | 8.1 | 0.00 |
| Attention deficit hyperactivity disorder | 0.7 | 0.7 | 0.00 | 0.7 | 0.8 | 0.01 | 0.6 | 0.7 | 0.02 |
| Chronic liver disease | 2.2 | 2.0 | -0.01 | 2.0 | 1.6 | -0.03 | 1.7 | 1.9 | 0.02 |
| Chronic obstructive lung disease | 13.2 | 11.4 | -0.06 | 11.6 | 12.1 | 0.01 | 11.2 | 12.1 | 0.03 |
| Crohn’s disease | 0.2 | 0.2 | 0.00 | 0.2 | <0.1 | -0.02 | 0.1 | 0.1 | 0.00 |
| Dementia | 1.2 | 1.7 | 0.04 | 1.2 | 1.5 | 0.02 | 1.3 | 1.6 | 0.02 |
| Depressive disorder | 21.7 | 20.3 | -0.04 | 20.9 | 21.2 | 0.01 | 20.4 | 20.6 | 0.01 |
| Gastroesophageal reflux disease | 20.8 | 19.4 | -0.04 | 19.7 | 19.9 | 0.01 | 19.1 | 19.9 | 0.02 |
| Gastrointestinal hemorrhage | 1.4 | 1.2 | -0.02 | 1.3 | 1.1 | -0.02 | 1.6 | 1.2 | -0.03 |
| Human immunodeficiency virus infection | 0.6 | 0.5 | 0.00 | 0.6 | 0.5 | 0.00 | 0.8 | 0.5 | -0.04 |
| Hyperlipidemia | 74.2 | 73.1 | -0.03 | 72.4 | 73.2 | 0.02 | 72.9 | 73.7 | 0.02 |
| Hypertensive disorder | 77.7 | 74.6 | -0.07 | 75.0 | 76.0 | 0.02 | 74.4 | 75.7 | 0.03 |
| Lesion of liver | 2.0 | 1.4 | -0.05 | 1.8 | 1.6 | -0.01 | 2.3 | 1.5 | -0.06 |
| Obesity | 32.4 | 28.1 | -0.10 | 30.0 | 30.5 | 0.01 | 29.0 | 29.4 | 0.01 |
| Osteoarthritis | 20.7 | 20.8 | 0.00 | 20.5 | 20.5 | 0.00 | 21.2 | 20.8 | -0.01 |
| Pneumonia | 2.0 | 1.4 | -0.04 | 1.6 | 1.5 | -0.01 | 1.5 | 1.4 | -0.01 |
| Psoriasis | 1.9 | 1.8 | -0.01 | 1.7 | 1.8 | 0.01 | 1.7 | 1.8 | 0.01 |
| Renal impairment | 10.1 | 8.4 | -0.06 | 8.3 | 9.0 | 0.03 | 7.6 | 8.7 | 0.04 |
| Rheumatoid arthritis | 0.8 | 0.7 | -0.01 | 0.8 | 0.7 | -0.01 | 0.7 | 0.7 | 0.00 |
| Schizophrenia | 0.6 | 0.9 | 0.04 | 0.6 | 0.6 | 0.00 | 0.6 | 0.8 | 0.01 |
| Ulcerative colitis | 0.4 | 0.4 | 0.00 | 0.4 | 0.3 | -0.01 | 0.4 | 0.4 | -0.01 |
| Urinary tract infectious disease | 1.3 | 2.1 | 0.06 | 1.2 | 1.3 | 0.01 | 1.6 | 1.8 | 0.01 |
| Viral hepatitis C | 1.0 | 1.2 | 0.01 | 1.0 | 0.7 | -0.04 | 0.8 | 1.0 | 0.02 |
| Visual system disorder | 42.7 | 44.1 | 0.03 | 42.3 | 42.9 | 0.01 | 42.6 | 43.3 | 0.01 |
| **Medical history: Cardiovascular disease** | | | | | | | | | |
| Atrial fibrillation | 12.1 | 7.4 | -0.16 | 8.8 | 9.8 | 0.03 | 8.8 | 9.0 | 0.01 |
| Cerebrovascular disease | 4.6 | 3.2 | -0.07 | 3.6 | 3.5 | -0.01 | 3.6 | 3.5 | -0.01 |
| Coronary arteriosclerosis | 32.7 | 15.4 | -0.41 | 21.8 | 24.4 | 0.06 | 19.6 | 21.3 | 0.04 |
| Heart disease | 48.3 | 28.7 | -0.41 | 35.9 | 38.7 | 0.06 | 33.1 | 35.2 | 0.04 |
| Heart failure | 13.6 | 4.2 | -0.33 | 6.1 | 6.7 | 0.03 | 6.9 | 6.3 | -0.03 |
| Ischemic heart disease | 14.3 | 6.6 | -0.25 | 8.3 | 9.0 | 0.03 | 8.5 | 8.5 | 0.00 |

| (Continued) | Before ajustment | | | After matching | | | After stratification | | |
|---|---|---|---|---|---|---|---|---|---|
| Characteristic | T (%) | C (%) | StdDiff | T (%) | C (%) | StdDiff | T (%) | C (%) | StdDiff |
| Peripheral vascular disease | 6.5 | 4.6 | -0.09 | 5.3 | 5.6 | 0.01 | 5.0 | 5.3 | 0.02 |
| Pulmonary embolism | 0.8 | 0.6 | -0.02 | 0.7 | 0.6 | -0.01 | 0.7 | 0.6 | -0.01 |
| Venous thrombosis | 1.0 | 1.2 | 0.01 | 1.1 | 1.4 | 0.03 | 0.9 | 1.3 | 0.03 |
| **Medical history: Neoplasms** | | | | | | | | | |
| Hematologic neoplasm | 1.1 | 1.1 | 0.01 | 1.0 | 1.1 | 0.01 | 1.1 | 1.1 | 0.00 |
| Malignant lymphoma | 0.8 | 0.7 | -0.01 | 0.8 | 0.7 | -0.01 | 0.7 | 0.6 | -0.01 |
| Malignant neoplasm of anorectum | <0.1 | 0.1 | 0.00 | 0.1 | <0.1 | 0.01 | 0.1 | 0.1 | 0.01 |
| Malignant neoplastic disease | 8.9 | 9.1 | 0.01 | 8.6 | 8.5 | 0.00 | 8.2 | 8.8 | 0.02 |
| Malignant tumor of colon | 0.3 | 0.5 | 0.02 | 0.4 | 0.4 | 0.00 | 0.3 | 0.4 | 0.01 |
| Malignant tumor of lung | 0.4 | 0.3 | -0.01 | 0.4 | 0.4 | -0.01 | 0.3 | 0.4 | 0.01 |
| Malignant tumor of urinary bladder | 0.6 | 0.6 | 0.00 | 0.5 | 0.6 | 0.01 | 0.5 | 0.6 | 0.01 |
| Primary malignant neoplasm of prostate | 3.2 | 3.6 | 0.03 | 3.1 | 3.5 | 0.02 | 3.1 | 3.4 | 0.01 |
| **Medication use** | | | | | | | | | |
| Agents acting on the renin-angiotensin system | 69.1 | 63.9 | -0.11 | 64.9 | 65.5 | 0.01 | 64.5 | 65.0 | 0.01 |
| Antibacterials for systemic use | 25.0 | 26.0 | 0.02 | 23.7 | 23.8 | 0.00 | 24.6 | 25.2 | 0.01 |
| Antidepressants | 34.0 | 33.0 | -0.02 | 33.4 | 33.9 | 0.01 | 35.4 | 33.5 | -0.04 |
| Antiepileptics | 25.4 | 25.0 | -0.01 | 24.8 | 25.0 | 0.00 | 26.4 | 25.2 | -0.03 |
| Antiinflammatory and antirheumatic products | 51.6 | 46.8 | -0.10 | 48.2 | 49.3 | 0.02 | 48.7 | 48.4 | -0.01 |
| Antineoplastic agents | 4.6 | 4.1 | -0.03 | 4.5 | 4.4 | -0.01 | 4.4 | 4.2 | -0.01 |
| Antipsoriatics | 0.8 | 0.9 | 0.01 | 0.7 | 0.9 | 0.03 | 0.5 | 0.9 | 0.05 |
| Antithrombotic agents | 45.4 | 32.9 | -0.26 | 37.2 | 38.7 | 0.03 | 36.1 | 37.0 | 0.02 |
| Beta blocking agents | 48.8 | 35.0 | -0.28 | 39.1 | 41.3 | 0.04 | 38.4 | 39.1 | 0.01 |
| Calcium channel blockers | 29.1 | 27.1 | -0.04 | 27.7 | 27.9 | 0.01 | 26.9 | 27.4 | 0.01 |
| Diuretics | 41.4 | 35.2 | -0.13 | 36.1 | 36.8 | 0.01 | 38.2 | 36.7 | -0.03 |
| Drugs for acid related disorders | 41.8 | 40.3 | -0.03 | 39.5 | 39.9 | 0.01 | 40.1 | 40.5 | 0.01 |
| Drugs for obstructive airway diseases | 39.8 | 37.1 | -0.06 | 37.9 | 37.8 | 0.00 | 37.2 | 38.4 | 0.03 |
| Immunosuppressants | 2.8 | 2.2 | -0.04 | 2.6 | 2.5 | -0.01 | 2.3 | 2.3 | 0.00 |
| Lipid modifying agents | 83.5 | 78.8 | -0.12 | 80.7 | 81.4 | 0.02 | 80.7 | 81.4 | 0.02 |
| Opioids | 14.5 | 15.5 | 0.03 | 13.3 | 13.1 | -0.01 | 15.6 | 15.0 | -0.02 |
| Psycholeptics | 22.7 | 21.6 | -0.03 | 21.4 | 21.9 | 0.01 | 21.2 | 21.6 | 0.01 |
| Psychostimulants | 1.5 | 1.7 | 0.01 | 1.6 | 1.6 | 0.00 | 1.5 | 1.7 | 0.01 |

##### 2.2.9 SIDIAP

**Supplementary Table 171:** Baseline patient characteristics for SGLT2I (male) (T) and DPP4I (male) (C) new-users in the SIDIAP data source. We report proportion of initiators satisfying selected base-line characteristics and the standardized difference of population proportions (StdDiff) before and after propensity score adjustment. Less extreme StdDiffs through matching and stratification suggest improved balance between patient cohorts through adjustment.

| | Before adjustment | | | After matching | | | After stratification | | |
|---|---|---|---|---|---|---|---|---|---|
| Characteristic | T (%) | C (%) | StdDiff | T (%) | C (%) | StdDiff | T (%) | C (%) | StdDiff |
| **Age group** | | | | | | | | | |
| 25 - 29 | 0.1 | 0.1 | -0.02 | 0.1 | <0.1 | -0.02 | 0.2 | 0.1 | -0.03 |
| 30 - 34 | 0.4 | 0.3 | -0.02 | 0.5 | 0.2 | -0.04 | 0.5 | 0.3 | -0.03 |
| 35 - 39 | 1.1 | 1.0 | -0.01 | 1.2 | 0.8 | -0.04 | 1.2 | 0.9 | -0.03 |
| 40 - 44 | 3.1 | 2.6 | -0.03 | 2.9 | 2.5 | -0.03 | 2.9 | 2.5 | -0.03 |
| 45 - 49 | 6.3 | 5.7 | -0.03 | 6.7 | 6.8 | 0.00 | 6.5 | 5.4 | -0.04 |
| 50 - 54 | 11.1 | 9.5 | -0.05 | 11.6 | 11.2 | -0.01 | 10.9 | 9.6 | -0.04 |
| 55 - 59 | 15.2 | 13.2 | -0.06 | 15.3 | 15.4 | 0.00 | 13.8 | 13.4 | -0.01 |
| 60 - 64 | 17.5 | 15.6 | -0.05 | 17.1 | 16.5 | -0.02 | 15.9 | 15.6 | -0.01 |
| 65 - 69 | 17.2 | 15.4 | -0.05 | 16.7 | 17.8 | 0.03 | 16.0 | 15.7 | -0.01 |
| 70 - 74 | 12.6 | 13.9 | 0.04 | 12.5 | 12.1 | -0.01 | 13.3 | 13.9 | 0.01 |
| 75 - 79 | 9.0 | 10.7 | 0.06 | 8.6 | 8.5 | 0.00 | 10.2 | 10.6 | 0.01 |
| 80 - 84 | 4.0 | 6.9 | 0.13 | 3.9 | 4.2 | 0.01 | 5.1 | 6.6 | 0.06 |
| 85 - 89 | 1.8 | 4.0 | 0.13 | 1.9 | 2.1 | 0.01 | 2.6 | 4.0 | 0.08 |
| 90 - 94 | 0.3 | 1.0 | 0.08 | 0.4 | 0.5 | 0.02 | 0.7 | 1.0 | 0.03 |
| 95 - 99 | <0.1 | 0.1 | 0.04 | <0.1 | 0.3 | 0.06 | <0.1 | 0.2 | 0.05 |
| **Medical history: General** | | | | | | | | | |
| Acute respiratory disease | 10.6 | 10.9 | 0.01 | 10.3 | 10.2 | 0.00 | 11.5 | 11.8 | 0.01 |
| Chronic liver disease | 0.3 | 0.4 | 0.01 | 0.3 | 0.3 | -0.01 | 0.2 | 0.4 | 0.02 |
| Chronic obstructive lung disease | 1.7 | 1.9 | 0.01 | 1.5 | 1.8 | 0.02 | 1.3 | 1.8 | 0.05 |
| Dementia | 0.3 | 0.5 | 0.04 | 0.3 | 0.2 | -0.02 | 0.3 | 0.4 | 0.02 |
| Depressive disorder | 1.1 | 1.7 | 0.05 | 1.1 | 1.1 | 0.00 | 1.5 | 1.4 | 0.00 |
| Gastroesophageal reflux disease | 0.9 | 1.1 | 0.01 | 0.9 | 1.2 | 0.02 | 0.7 | 1.3 | 0.06 |
| Gastrointestinal hemorrhage | 1.1 | 1.8 | 0.06 | 1.1 | 1.2 | 0.01 | 1.6 | 1.7 | 0.01 |
| Hyperlipidemia | 2.8 | 3.8 | 0.06 | 3.0 | 3.1 | 0.01 | 2.9 | 3.3 | 0.02 |
| Hypertensive disorder | 3.8 | 4.5 | 0.04 | 3.9 | 4.0 | 0.00 | 3.9 | 3.8 | 0.00 |
| Lesion of liver | 0.3 | 0.5 | 0.03 | 0.3 | 0.4 | 0.01 | 0.3 | 0.4 | 0.03 |
| Obesity | 7.1 | 6.9 | -0.01 | 7.3 | 7.4 | 0.00 | 8.0 | 7.0 | -0.04 |
| Osteoarthritis | 2.8 | 3.5 | 0.04 | 2.9 | 2.4 | -0.03 | 3.7 | 3.1 | -0.03 |
| Pneumonia | 1.9 | 1.7 | -0.01 | 2.3 | 1.8 | -0.04 | 2.3 | 1.8 | -0.04 |
| Psoriasis | 0.4 | 0.4 | 0.00 | 0.4 | 0.4 | 0.00 | 0.4 | 0.4 | -0.01 |
| Renal impairment | 1.5 | 3.7 | 0.14 | 1.5 | 1.8 | 0.02 | 2.7 | 3.6 | 0.05 |
| Rheumatoid arthritis | <0.1 | 0.1 | 0.01 | <0.1 | <0.1 | -0.01 | <0.1 | 0.1 | 0.02 |
| Urinary tract infectious disease | 3.2 | 4.1 | 0.05 | 3.1 | 3.5 | 0.02 | 4.0 | 4.4 | 0.02 |
| Viral hepatitis C | <0.1 | 0.2 | 0.03 | 0.1 | 0.1 | 0.01 | 0.1 | 0.2 | 0.02 |
| Visual system disorder | 12.2 | 14.3 | 0.06 | 12.7 | 12.5 | 0.00 | 15.9 | 14.7 | -0.03 |
| **Medical history: Cardiovascular disease** | | | | | | | | | |
| Atrial fibrillation | 2.5 | 1.5 | -0.07 | 1.8 | 1.9 | 0.01 | 1.6 | 1.7 | 0.01 |
| Cerebrovascular disease | 0.9 | 1.2 | 0.03 | 0.8 | 0.9 | 0.01 | 1.1 | 1.2 | 0.01 |
| Heart disease | 16.2 | 9.8 | -0.19 | 10.7 | 11.1 | 0.01 | 9.4 | 10.1 | 0.02 |
| Heart failure | 3.4 | 1.7 | -0.11 | 1.9 | 1.7 | -0.01 | 1.7 | 1.8 | 0.01 |
| Ischemic heart disease | 8.0 | 3.3 | -0.20 | 4.3 | 4.6 | 0.01 | 3.7 | 3.4 | -0.02 |
| Peripheral vascular disease | 1.2 | 1.2 | 0.00 | 1.1 | 1.0 | -0.01 | 0.8 | 1.2 | 0.04 |
| Pulmonary embolism | 0.1 | 0.2 | 0.02 | 0.1 | 0.2 | 0.04 | 0.1 | 0.2 | 0.02 |
| **Medical history: Neoplasms** | | | | | | | | | |
| Hematologic neoplasm | 0.1 | 0.1 | 0.01 | 0.1 | <0.1 | -0.01 | 0.4 | 0.1 | -0.05 |
| Malignant neoplasm of anorectum | <0.1 | 0.1 | 0.02 | 0.1 | 0.1 | -0.01 | 0.1 | 0.1 | -0.01 |
| Malignant neoplastic disease | 1.7 | 3.1 | 0.09 | 1.8 | 2.2 | 0.03 | 2.5 | 3.0 | 0.03 |
| Malignant tumor of colon | 0.1 | 0.4 | 0.05 | 0.1 | 0.3 | 0.03 | 0.2 | 0.3 | 0.02 |
| Malignant tumor of lung | <0.1 | 0.1 | 0.02 | <0.1 | <0.1 | 0.02 | <0.1 | 0.1 | 0.03 |
| Malignant tumor of urinary bladder | 0.1 | 0.4 | 0.06 | 0.1 | 0.1 | 0.01 | 0.4 | 0.4 | -0.01 |
| Primary malignant neoplasm of prostate | 0.2 | 0.6 | 0.06 | 0.3 | 0.4 | 0.03 | 0.2 | 0.5 | 0.04 |
| **Medication use** | | | | | | | | | |
| Agents acting on the renin-angiotensin system | 69.7 | 61.2 | -0.18 | 65.5 | 66.9 | 0.03 | 59.8 | 63.2 | 0.07 |
| Antibacterials for systemic use | 33.6 | 34.9 | 0.03 | 33.3 | 32.5 | -0.02 | 33.3 | 32.5 | -0.02 |
| Antidepressants | 12.8 | 12.9 | 0.00 | 13.0 | 12.9 | 0.00 | 13.2 | 13.3 | 0.00 |
| Antiepileptics | 8.4 | 7.9 | -0.02 | 8.7 | 8.2 | -0.02 | 9.1 | 8.5 | -0.02 |
| Antiinflammatory and antirheumatic products | 55.2 | 51.4 | -0.07 | 51.7 | 51.5 | 0.00 | 51.1 | 50.6 | -0.01 |

| (Continued) | Before ajustment | | | After matching | | | After stratification | | |
|---|---|---|---|---|---|---|---|---|---|
| Characteristic | T (%) | C (%) | StdDiff | T (%) | C (%) | StdDiff | T (%) | C (%) | StdDiff |
| Antineoplastic agents | 1.4 | 1.6 | 0.02 | 1.4 | 1.7 | 0.02 | 1.4 | 1.6 | 0.02 |
| Antipsoriatics | 2.2 | 1.9 | -0.02 | 2.2 | 2.2 | 0.00 | 2.1 | 2.0 | -0.01 |
| Antithrombotic agents | 49.6 | 39.1 | -0.21 | 42.6 | 44.0 | 0.03 | 37.5 | 40.7 | 0.06 |
| Beta blocking agents | 42.0 | 26.3 | -0.34 | 33.8 | 34.5 | 0.01 | 27.0 | 29.3 | 0.05 |
| Calcium channel blockers | 26.6 | 24.7 | -0.04 | 26.1 | 25.9 | -0.01 | 23.5 | 25.4 | 0.04 |
| Diuretics | 45.8 | 40.2 | -0.11 | 41.8 | 41.8 | 0.00 | 40.1 | 41.8 | 0.03 |
| Drugs for acid related disorders | 50.2 | 46.0 | -0.09 | 45.9 | 45.8 | 0.00 | 44.8 | 46.6 | 0.04 |
| Drugs for obstructive airway diseases | 27.7 | 25.9 | -0.04 | 26.8 | 26.7 | 0.00 | 27.0 | 27.3 | 0.01 |
| Immunosuppressants | 1.4 | 1.2 | -0.02 | 1.4 | 1.5 | 0.01 | 1.3 | 1.4 | 0.01 |
| Lipid modifying agents | 70.6 | 64.7 | -0.13 | 67.0 | 67.1 | 0.00 | 62.9 | 65.4 | 0.05 |
| Opioids | 14.4 | 13.6 | -0.02 | 14.1 | 13.9 | -0.01 | 14.6 | 14.5 | 0.00 |
| Psycholeptics | 22.3 | 23.1 | 0.02 | 21.8 | 21.5 | -0.01 | 22.1 | 23.0 | 0.02 |
| Psychostimulants | 9.5 | 4.0 | -0.22 | 6.3 | 6.4 | 0.01 | 5.2 | 4.7 | -0.02 |

#### 2.3 SGLT2I (male) vs SU (male)

##### 2.3.1 CCAE

**Supplementary Table 172:** Baseline patient characteristics for SGLT2I (male) (T) and SU (male) (C) new-users in the CCAE data source. We report proportion of initiators satisfying selected base-line characteristics and the standardized difference of population proportions (StdDiff) before and after propensity score adjustment. Less extreme StdDiffs through matching and stratification suggest improved balance between patient cohorts through adjustment.

| | Before adjustment | | | After matching | | | After stratification | | |
|---|---|---|---|---|---|---|---|---|---|
| Characteristic | T (%) | C (%) | StdDiff | T (%) | C (%) | StdDiff | T (%) | C (%) | StdDiff |
| **Age group** | | | | | | | | | |
| 20 - 24 | 0.3 | 0.2 | 0.02 | 0.3 | 0.2 | 0.02 | 0.3 | 0.2 | 0.01 |
| 25 - 29 | 0.5 | 0.6 | -0.01 | 0.5 | 0.5 | 0.00 | 0.5 | 0.5 | 0.00 |
| 30 - 34 | 1.9 | 2.0 | -0.01 | 2.0 | 1.9 | 0.01 | 1.9 | 1.9 | 0.01 |
| 35 - 39 | 4.6 | 4.6 | 0.00 | 4.8 | 4.7 | 0.00 | 4.8 | 4.7 | 0.00 |
| 40 - 44 | 9.1 | 8.9 | 0.00 | 9.3 | 9.0 | 0.01 | 9.3 | 9.0 | 0.01 |
| 45 - 49 | 15.4 | 14.2 | 0.03 | 15.3 | 15.4 | 0.00 | 14.9 | 14.7 | 0.01 |
| 50 - 54 | 20.3 | 19.9 | 0.01 | 20.1 | 20.2 | 0.00 | 20.6 | 20.1 | 0.01 |
| 55 - 59 | 23.8 | 23.4 | 0.01 | 23.6 | 23.4 | 0.00 | 23.6 | 23.4 | 0.00 |
| 60 - 64 | 22.2 | 23.9 | -0.04 | 22.2 | 22.5 | 0.00 | 22.5 | 23.4 | -0.02 |
| 65 - 69 | 1.9 | 2.3 | -0.03 | 1.9 | 1.9 | 0.00 | 2.0 | 2.1 | -0.01 |
| **Medical history: General** | | | | | | | | | |
| Acute respiratory disease | 23.1 | 20.5 | 0.06 | 22.5 | 22.5 | 0.00 | 22.1 | 21.8 | 0.01 |
| Attention deficit hyperactivity disorder | 1.2 | 0.6 | 0.06 | 1.1 | 1.1 | 0.00 | 1.1 | 1.0 | 0.02 |
| Chronic liver disease | 1.6 | 2.7 | -0.07 | 1.7 | 1.6 | 0.01 | 2.1 | 2.1 | 0.00 |
| Chronic obstructive lung disease | 2.3 | 2.7 | -0.03 | 2.3 | 2.3 | 0.00 | 2.6 | 2.6 | 0.00 |
| Crohn's disease | 0.1 | 0.2 | -0.02 | 0.1 | 0.3 | -0.02 | 0.2 | 0.3 | -0.02 |
| Dementia | 0.1 | 0.1 | -0.01 | 0.1 | 0.2 | 0.00 | 0.1 | 0.2 | 0.00 |
| Depressive disorder | 7.0 | 6.0 | 0.04 | 6.9 | 7.0 | 0.00 | 7.0 | 7.2 | -0.01 |
| Gastroesophageal reflux disease | 11.3 | 7.5 | 0.13 | 10.9 | 11.0 | 0.00 | 10.1 | 10.4 | -0.01 |
| Gastrointestinal hemorrhage | 1.4 | 1.9 | -0.04 | 1.4 | 1.5 | -0.01 | 1.4 | 1.6 | -0.02 |
| Human immunodeficiency virus infection | 0.5 | 0.3 | 0.02 | 0.4 | 0.5 | -0.01 | 0.4 | 0.5 | -0.02 |
| Hyperlipidemia | 75.7 | 62.3 | 0.29 | 74.0 | 74.4 | -0.01 | 71.8 | 72.2 | -0.01 |
| Hypertensive disorder | 72.2 | 61.6 | 0.23 | 71.3 | 71.3 | 0.00 | 70.1 | 69.9 | 0.00 |
| Lesion of liver | 0.6 | 0.8 | -0.02 | 0.6 | 0.7 | 0.00 | 0.7 | 0.8 | -0.01 |
| Obesity | 25.3 | 13.7 | 0.30 | 23.9 | 24.2 | -0.01 | 21.5 | 22.3 | -0.02 |
| Osteoarthritis | 15.8 | 13.9 | 0.06 | 15.5 | 15.6 | 0.00 | 15.8 | 15.7 | 0.00 |
| Pneumonia | 1.9 | 2.2 | -0.02 | 2.1 | 2.0 | 0.00 | 2.1 | 2.0 | 0.00 |
| Psoriasis | 1.7 | 1.2 | 0.04 | 1.6 | 1.7 | 0.00 | 1.5 | 1.6 | -0.01 |
| Renal impairment | 3.6 | 3.7 | -0.01 | 3.6 | 3.5 | 0.01 | 4.3 | 4.4 | -0.01 |
| Rheumatoid arthritis | 0.6 | 0.5 | 0.00 | 0.5 | 0.5 | -0.01 | 0.5 | 0.5 | -0.01 |
| Schizophrenia | 0.1 | 0.1 | -0.01 | 0.1 | 0.1 | 0.01 | 0.1 | 0.1 | -0.01 |
| Ulcerative colitis | 0.3 | 0.3 | -0.01 | 0.3 | 0.3 | -0.01 | 0.3 | 0.4 | -0.01 |
| Urinary tract infectious disease | 2.1 | 2.9 | -0.05 | 2.5 | 2.5 | 0.00 | 2.5 | 2.5 | 0.00 |
| Viral hepatitis C | 0.3 | 0.5 | -0.04 | 0.3 | 0.3 | 0.01 | 0.4 | 0.4 | 0.00 |
| Visual system disorder | 22.7 | 20.7 | 0.05 | 22.3 | 21.9 | 0.01 | 22.3 | 21.9 | 0.01 |
| **Medical history: Cardiovascular disease** | | | | | | | | | |
| Atrial fibrillation | 2.7 | 2.4 | 0.02 | 2.5 | 2.5 | 0.00 | 2.6 | 2.5 | 0.00 |
| Cerebrovascular disease | 2.5 | 2.5 | 0.00 | 2.3 | 2.2 | 0.00 | 2.5 | 2.3 | 0.01 |
| Coronary arteriosclerosis | 11.3 | 9.7 | 0.05 | 9.8 | 9.9 | 0.00 | 9.7 | 9.5 | 0.01 |
| Heart disease | 20.3 | 18.1 | 0.06 | 18.2 | 18.1 | 0.00 | 18.1 | 17.8 | 0.01 |
| Heart failure | 2.5 | 2.3 | 0.01 | 2.2 | 2.1 | 0.01 | 2.2 | 2.2 | 0.00 |
| Peripheral vascular disease | 4.2 | 3.0 | 0.07 | 3.9 | 3.8 | 0.00 | 3.7 | 3.5 | 0.01 |
| Venous thrombosis | 0.9 | 1.0 | -0.01 | 1.0 | 0.9 | 0.00 | 1.1 | 1.1 | 0.00 |
| **Medical history: Neoplasms** | | | | | | | | | |
| Hematologic neoplasm | 0.7 | 0.7 | -0.01 | 0.7 | 0.7 | 0.00 | 0.8 | 0.7 | 0.00 |
| Malignant lymphoma | 0.4 | 0.4 | -0.01 | 0.3 | 0.4 | 0.00 | 0.4 | 0.4 | 0.00 |
| Malignant neoplasm of anorectum | 0.1 | 0.2 | -0.02 | 0.1 | 0.1 | 0.01 | 0.2 | 0.1 | 0.01 |
| Malignant neoplastic disease | 4.7 | 5.0 | -0.02 | 4.9 | 4.8 | 0.00 | 4.9 | 4.8 | 0.00 |
| Malignant tumor of colon | 0.3 | 0.4 | -0.01 | 0.3 | 0.2 | 0.01 | 0.4 | 0.3 | 0.02 |
| Malignant tumor of lung | 0.1 | 0.2 | -0.03 | 0.1 | 0.1 | -0.01 | 0.1 | 0.2 | -0.01 |
| Malignant tumor of urinary bladder | 0.2 | 0.3 | -0.02 | 0.2 | 0.1 | 0.01 | 0.2 | 0.2 | 0.00 |
| Primary malignant neoplasm of prostate | 1.0 | 1.3 | -0.03 | 1.0 | 1.1 | 0.00 | 1.2 | 1.2 | 0.00 |
| **Medication use** | | | | | | | | | |
| Agents acting on the renin-angiotensin system | 71.2 | 68.2 | 0.07 | 70.3 | 70.2 | 0.00 | 69.6 | 69.8 | 0.00 |
| Antibacterials for systemic use | 50.9 | 50.3 | 0.01 | 50.4 | 50.1 | 0.01 | 50.4 | 50.1 | 0.01 |
| Antidepressants | 18.8 | 17.4 | 0.04 | 18.2 | 18.3 | 0.00 | 18.2 | 18.1 | 0.00 |

| (Continued) | Before ajustment | | | After matching | | | After stratification | | |
|---|---|---|---|---|---|---|---|---|---|
| Characteristic | T (%) | C (%) | StdDiff | T (%) | C (%) | StdDiff | T (%) | C (%) | StdDiff |
| Antiepileptics | 9.9 | 8.7 | 0.04 | 9.7 | 9.6 | 0.00 | 10.2 | 10.0 | 0.01 |
| Antiinflammatory and antirheumatic products | 30.2 | 27.2 | 0.07 | 29.5 | 29.8 | -0.01 | 29.7 | 29.7 | 0.00 |
| Antineoplastic agents | 3.1 | 3.2 | 0.00 | 2.9 | 2.9 | 0.00 | 2.9 | 2.9 | 0.00 |
| Antipsoriatics | 0.5 | 0.5 | 0.01 | 0.5 | 0.4 | 0.01 | 0.5 | 0.4 | 0.01 |
| Antithrombotic agents | 12.1 | 10.7 | 0.04 | 11.5 | 11.2 | 0.01 | 11.7 | 11.7 | 0.00 |
| Beta blocking agents | 26.0 | 26.0 | 0.00 | 24.7 | 24.5 | 0.01 | 25.1 | 25.0 | 0.00 |
| Calcium channel blockers | 22.8 | 20.4 | 0.06 | 22.3 | 22.3 | 0.00 | 21.8 | 22.0 | 0.00 |
| Diuretics | 32.5 | 32.9 | -0.01 | 32.0 | 31.8 | 0.00 | 32.2 | 32.2 | 0.00 |
| Drugs for acid related disorders | 20.6 | 19.0 | 0.04 | 19.9 | 19.9 | 0.00 | 19.5 | 19.5 | 0.00 |
| Drugs for obstructive airway diseases | 31.8 | 27.8 | 0.09 | 30.6 | 30.7 | 0.00 | 30.5 | 30.0 | 0.01 |
| Immunosuppressants | 1.7 | 1.4 | 0.03 | 1.7 | 1.7 | 0.00 | 1.7 | 1.7 | 0.00 |
| Lipid modifying agents | 72.9 | 68.3 | 0.10 | 71.1 | 71.1 | 0.00 | 69.8 | 70.6 | -0.02 |
| Opioids | 21.4 | 22.4 | -0.02 | 21.3 | 21.1 | 0.00 | 22.7 | 22.4 | 0.01 |
| Psycholeptics | 17.6 | 16.9 | 0.02 | 17.0 | 17.0 | 0.00 | 17.6 | 17.2 | 0.01 |
| Psychostimulants | 2.7 | 1.9 | 0.05 | 2.4 | 2.4 | 0.00 | 2.5 | 2.2 | 0.02 |

##### 2.3.2 DAG

**Supplementary Table 173:** Baseline patient characteristics for SGLT2I (male) (T) and SU (male) (C) new-users in the DAG data source. We report proportion of initiators satisfying selected base-line characteristics and the standardized difference of population proportions (StdDiff) before and after propensity score adjustment. Less extreme StdDiffs through matching and stratification suggest improved balance between patient cohorts through adjustment.

| | Before adjustment | | | After matching | | | After stratification | | |
|---|---|---|---|---|---|---|---|---|---|
| Characteristic | T (%) | C (%) | StdDiff | T (%) | C (%) | StdDiff | T (%) | C (%) | StdDiff |
| **Age group** | | | | | | | | | |
| 30 - 34 | 0.6 | 0.4 | 0.03 | <0.5 | 0.7 | -0.02 | 0.6 | 0.4 | 0.04 |
| 35 - 39 | 1.5 | 1.2 | 0.03 | 2.2 | 1.4 | 0.06 | 1.6 | 0.8 | 0.07 |
| 40 - 44 | 3.1 | 2.2 | 0.06 | 4.1 | 2.7 | 0.07 | 3.3 | 3.1 | 0.01 |
| 45 - 49 | 6.0 | 5.3 | 0.03 | 5.4 | 4.6 | 0.04 | 5.8 | 5.9 | -0.01 |
| 50 - 54 | 10.9 | 8.8 | 0.07 | 10.6 | 10.2 | 0.01 | 11.3 | 12.6 | -0.04 |
| 55 - 59 | 15.4 | 11.9 | 0.10 | 12.8 | 12.6 | 0.01 | 15.0 | 14.3 | 0.02 |
| 60 - 64 | 17.5 | 15.5 | 0.06 | 15.2 | 15.7 | -0.01 | 17.1 | 15.1 | 0.06 |
| 65 - 69 | 14.3 | 16.2 | -0.05 | 15.3 | 14.3 | 0.03 | 14.1 | 17.4 | -0.09 |
| 70 - 74 | 12.6 | 16.7 | -0.12 | 12.8 | 12.9 | 0.00 | 12.6 | 12.5 | 0.00 |
| 75 - 79 | 9.5 | 12.7 | -0.10 | 11.6 | 12.5 | -0.03 | 10.0 | 9.8 | 0.00 |
| 80 - 84 | 6.0 | 6.8 | -0.03 | 7.1 | 8.5 | -0.05 | 6.4 | 5.5 | 0.04 |
| 85 - 89 | 2.0 | 1.8 | 0.02 | 1.6 | 2.7 | -0.07 | 1.8 | 1.8 | 0.00 |
| 90 - 94 | 0.2 | 0.3 | -0.02 | <0.5 | 1.1 | -0.13 | 0.2 | 0.8 | -0.09 |
| **Medical history: General** | | | | | | | | | |
| Acute respiratory disease | 15.1 | 17.2 | -0.06 | 16.9 | 15.3 | 0.04 | 15.5 | 16.2 | -0.02 |
| Attention deficit hyperactivity disorder | 0.1 | 0.4 | -0.06 | <0.1 | 0.4 | -0.07 | <0.1 | 0.4 | -0.07 |
| Chronic liver disease | 0.6 | 0.6 | 0.00 | <0.5 | 0.7 | -0.05 | 0.6 | 0.4 | 0.03 |
| Chronic obstructive lung disease | 6.2 | 6.7 | -0.02 | 6.4 | 5.8 | 0.02 | 6.1 | 6.2 | 0.00 |
| Crohn's disease | 0.1 | 0.1 | -0.01 | 0.1 | <0.2 | 0.00 | 0.1 | <0.2 | 0.00 |
| Dementia | 0.8 | 2.1 | -0.11 | 1.0 | 1.7 | -0.06 | 0.9 | 1.0 | -0.01 |
| Depressive disorder | 6.4 | 7.2 | -0.03 | 6.9 | 7.2 | -0.01 | 6.6 | 10.4 | -0.14 |
| Gastroesophageal reflux disease | 1.9 | 2.2 | -0.02 | 2.3 | 1.8 | 0.03 | 2.0 | 1.5 | 0.04 |
| Gastrointestinal hemorrhage | 0.7 | 1.0 | -0.03 | 1.0 | 0.7 | 0.03 | 0.8 | 0.4 | 0.06 |
| Hyperlipidemia | 24.3 | 29.2 | -0.11 | 23.0 | 24.4 | -0.03 | 24.2 | 22.2 | 0.05 |
| Hypertensive disorder | 42.6 | 55.3 | -0.26 | 44.3 | 43.7 | 0.01 | 43.5 | 39.8 | 0.07 |
| Lesion of liver | 0.6 | 1.1 | -0.05 | <0.5 | 1.2 | -0.12 | 0.6 | 0.7 | -0.02 |
| Obesity | 10.0 | 7.1 | 0.11 | 9.7 | 8.0 | 0.06 | 9.8 | 9.4 | 0.01 |
| Osteoarthritis | 9.0 | 12.9 | -0.13 | 11.1 | 11.1 | 0.00 | 9.6 | 9.3 | 0.01 |
| Pneumonia | 1.5 | 2.0 | -0.03 | 2.0 | 1.7 | 0.02 | 1.5 | 1.2 | 0.02 |
| Psoriasis | 1.2 | 1.3 | -0.01 | 0.7 | 1.0 | -0.03 | 1.2 | 0.8 | 0.04 |
| Renal impairment | 5.2 | 5.1 | 0.00 | 4.5 | 5.2 | -0.03 | 4.8 | 3.5 | 0.06 |
| Rheumatoid arthritis | 0.9 | 0.8 | 0.01 | 0.7 | 0.7 | 0.00 | 0.8 | 1.0 | -0.01 |
| Schizophrenia | 0.2 | 0.3 | -0.01 | <0.5 | 0.4 | -0.02 | 0.2 | 0.5 | -0.04 |
| Ulcerative colitis | 0.2 | 0.2 | -0.01 | 0.1 | 0.4 | -0.05 | 0.1 | 0.4 | -0.05 |
| Urinary tract infectious disease | 2.5 | 2.5 | 0.00 | 2.2 | 2.4 | -0.02 | 2.4 | 2.1 | 0.02 |
| Viral hepatitis C | <0.1 | <0.1 | -0.01 | <0.1 | <0.2 | -0.01 | <0.1 | <0.2 | -0.01 |
| Visual system disorder | 6.5 | 9.7 | -0.12 | 8.2 | 8.6 | -0.01 | 7.0 | 7.9 | -0.03 |
| **Medical history: Cardiovascular disease** | | | | | | | | | |
| Atrial fibrillation | 3.5 | 2.0 | 0.09 | 2.7 | 2.9 | -0.01 | 3.2 | 2.2 | 0.06 |
| Cerebrovascular disease | 4.4 | 5.5 | -0.05 | 4.5 | 4.4 | 0.00 | 4.5 | 5.3 | -0.04 |
| Coronary arteriosclerosis | 9.0 | 4.2 | 0.19 | 3.6 | 5.0 | -0.07 | 8.1 | 8.5 | -0.01 |
| Heart disease | 31.5 | 30.1 | 0.03 | 21.2 | 23.8 | -0.06 | 29.6 | 26.2 | 0.08 |
| Heart failure | 8.8 | 7.5 | 0.05 | 5.7 | 5.3 | 0.02 | 8.1 | 4.9 | 0.13 |
| Ischemic heart disease | 15.8 | 16.8 | -0.03 | 9.9 | 11.1 | -0.04 | 14.7 | 13.1 | 0.05 |
| Peripheral vascular disease | 6.6 | 6.5 | 0.00 | 6.6 | 5.7 | 0.04 | 6.5 | 4.6 | 0.08 |
| Pulmonary embolism | 0.6 | 0.2 | 0.06 | <0.5 | 0.3 | 0.01 | 0.5 | <0.2 | 0.07 |
| Venous thrombosis | 1.3 | 1.0 | 0.03 | 1.5 | 0.9 | 0.06 | 1.3 | 1.1 | 0.02 |
| **Medical history: Neoplasms** | | | | | | | | | |
| Hematologic neoplasm | 0.3 | 0.3 | 0.00 | <0.5 | 0.3 | 0.00 | 0.3 | <0.2 | 0.02 |
| Malignant lymphoma | 0.2 | 0.4 | -0.04 | <0.5 | <0.2 | 0.04 | 0.2 | <0.2 | 0.04 |
| Malignant neoplasm of anorectum | 0.1 | 0.2 | -0.02 | 0.1 | <0.2 | 0.03 | 0.1 | <0.2 | 0.03 |
| Malignant neoplastic disease | 3.5 | 6.5 | -0.14 | 4.0 | 4.8 | -0.04 | 3.8 | 4.2 | -0.02 |
| Malignant tumor of breast | <0.1 | <0.1 | -0.01 | <0.1 | <0.2 | -0.01 | <0.1 | <0.2 | -0.01 |
| Malignant tumor of colon | 0.3 | 0.5 | -0.03 | <0.5 | 0.4 | 0.00 | 0.3 | 1.0 | -0.09 |
| Malignant tumor of lung | <0.1 | 0.2 | -0.04 | <0.5 | <0.2 | 0.00 | <0.1 | <0.2 | -0.01 |
| Malignant tumor of urinary bladder | 0.2 | 0.4 | -0.03 | <0.5 | <0.2 | 0.09 | 0.3 | <0.2 | 0.06 |
| Primary malignant neoplasm of prostate | 1.0 | 1.9 | -0.08 | 1.2 | 1.3 | -0.01 | 1.1 | 1.6 | -0.04 |
| **Medication use** | | | | | | | | | |

| (Continued) | Before ajustment | | | After matching | | | After stratification | | |
|---|---|---|---|---|---|---|---|---|---|
| Characteristic | T (%) | C (%) | StdDiff | T (%) | C (%) | StdDiff | T (%) | C (%) | StdDiff |
| Agents acting on the renin-angiotensin system | 71.7 | 65.8 | 0.13 | 67.2 | 67.5 | -0.01 | 70.8 | 69.9 | 0.02 |
| Antibacterials for systemic use | 19.5 | 25.4 | -0.14 | 21.5 | 21.8 | -0.01 | 20.4 | 18.8 | 0.04 |
| Antidepressants | 7.4 | 7.1 | 0.01 | 7.3 | 7.0 | 0.01 | 7.4 | 8.1 | -0.03 |
| Antiepileptics | 3.9 | 3.5 | 0.03 | 4.3 | 2.8 | 0.08 | 3.9 | 4.2 | -0.02 |
| Antiinflammatory and antirheumatic products | 43.7 | 47.9 | -0.08 | 40.9 | 42.1 | -0.02 | 43.4 | 41.6 | 0.04 |
| Antineoplastic agents | 1.1 | 0.8 | 0.04 | 0.9 | 1.0 | -0.01 | 1.1 | 0.9 | 0.02 |
| Antipsoriatics | 0.6 | 0.4 | 0.03 | 0.7 | 0.2 | 0.07 | 0.5 | 0.2 | 0.05 |
| Antithrombotic agents | 39.1 | 34.3 | 0.10 | 29.8 | 31.2 | -0.03 | 36.9 | 34.9 | 0.04 |
| Beta blocking agents | 48.2 | 45.1 | 0.06 | 41.2 | 43.3 | -0.04 | 46.9 | 44.2 | 0.06 |
| Calcium channel blockers | 34.8 | 29.7 | 0.11 | 34.1 | 33.0 | 0.02 | 34.6 | 37.0 | -0.05 |
| Diuretics | 46.1 | 47.7 | -0.03 | 43.1 | 42.3 | 0.01 | 45.3 | 45.0 | 0.00 |
| Drugs for acid related disorders | 32.3 | 28.4 | 0.09 | 30.5 | 31.0 | -0.01 | 31.8 | 32.5 | -0.01 |
| Drugs for obstructive airway diseases | 16.6 | 16.1 | 0.01 | 13.1 | 14.7 | -0.05 | 15.7 | 15.6 | 0.00 |
| Immunosuppressants | 0.9 | 0.3 | 0.07 | 0.6 | 0.3 | 0.04 | 0.7 | 0.4 | 0.04 |
| Lipid modifying agents | 56.3 | 42.2 | 0.28 | 42.1 | 45.5 | -0.07 | 53.4 | 55.6 | -0.04 |
| Opioids | 9.0 | 11.9 | -0.10 | 9.9 | 10.5 | -0.02 | 9.7 | 10.3 | -0.02 |
| Psycholeptics | 6.3 | 7.6 | -0.05 | 6.7 | 6.2 | 0.02 | 6.5 | 7.7 | -0.05 |
| Psychostimulants | 0.1 | 0.6 | -0.07 | <0.5 | <0.2 | 0.06 | 0.1 | <0.2 | 0.01 |

##### 2.3.3 ODOD

**Supplementary Table 174:** Baseline patient characteristics for SGLT2I (male) (T) and SU (male) (C) new-users in the ODOD data source. We report proportion of initiators satisfying selected base-line characteristics and the standardized difference of population proportions (StdDiff) before and after propensity score adjustment. Less extreme StdDiffs through matching and stratification suggest improved balance between patient cohorts through adjustment.

| | Before adjustment | | | After matching | | | After stratification | | |
|---|---|---|---|---|---|---|---|---|---|
| Characteristic | T (%) | C (%) | StdDiff | T (%) | C (%) | StdDiff | T (%) | C (%) | StdDiff |
| **Age group** | | | | | | | | | |
| 20 - 24 | 0.1 | 0.1 | 0.00 | 0.1 | 0.1 | 0.01 | 0.1 | 0.1 | 0.00 |
| 25 - 29 | 0.4 | 0.3 | 0.01 | 0.4 | 0.3 | 0.03 | 0.4 | 0.3 | 0.03 |
| 30 - 34 | 1.3 | 1.1 | 0.01 | 1.3 | 1.2 | 0.01 | 1.3 | 1.1 | 0.03 |
| 35 - 39 | 3.2 | 2.7 | 0.03 | 3.3 | 3.2 | 0.01 | 2.7 | 2.6 | 0.01 |
| 40 - 44 | 6.1 | 5.0 | 0.05 | 6.1 | 6.3 | -0.01 | 5.0 | 4.9 | 0.00 |
| 45 - 49 | 9.5 | 7.9 | 0.06 | 9.5 | 9.6 | 0.00 | 7.9 | 7.8 | 0.00 |
| 50 - 54 | 13.3 | 10.4 | 0.09 | 12.9 | 13.3 | -0.01 | 10.8 | 10.7 | 0.00 |
| 55 - 59 | 15.0 | 12.5 | 0.07 | 14.6 | 14.9 | -0.01 | 13.0 | 12.7 | 0.01 |
| 60 - 64 | 14.6 | 12.7 | 0.06 | 14.2 | 14.3 | 0.00 | 13.0 | 13.0 | 0.00 |
| 65 - 69 | 14.7 | 15.8 | -0.03 | 14.8 | 15.2 | -0.01 | 16.1 | 16.5 | -0.01 |
| 70 - 74 | 12.1 | 14.8 | -0.08 | 12.5 | 12.0 | 0.01 | 15.0 | 15.1 | 0.00 |
| 75 - 79 | 6.4 | 9.5 | -0.11 | 6.7 | 6.3 | 0.02 | 8.6 | 8.9 | -0.01 |
| 80 - 84 | 2.3 | 4.9 | -0.14 | 2.5 | 2.3 | 0.01 | 4.0 | 4.3 | -0.01 |
| 85 - 89 | 0.9 | 2.0 | -0.10 | 0.9 | 1.0 | 0.00 | 1.8 | 1.9 | -0.01 |
| 90 - 94 | 0.1 | 0.1 | -0.01 | 0.1 | 0.1 | -0.01 | 0.1 | 0.1 | -0.01 |
| **Medical history: General** | | | | | | | | | |
| Acute respiratory disease | 21.6 | 21.7 | 0.00 | 21.0 | 21.0 | 0.00 | 22.5 | 21.2 | 0.03 |
| Attention deficit hyperactivity disorder | 0.9 | 0.6 | 0.04 | 0.9 | 0.9 | 0.00 | 0.8 | 0.7 | 0.00 |
| Chronic liver disease | 1.7 | 3.0 | -0.09 | 1.8 | 1.6 | 0.01 | 2.8 | 2.3 | 0.04 |
| Chronic obstructive lung disease | 7.4 | 9.6 | -0.08 | 7.2 | 7.1 | 0.00 | 9.6 | 9.3 | 0.01 |
| Crohn’s disease | 0.3 | 0.2 | 0.01 | 0.3 | 0.2 | 0.01 | 0.3 | 0.2 | 0.02 |
| Dementia | 0.8 | 1.8 | -0.09 | 0.8 | 0.8 | 0.00 | 1.6 | 1.7 | -0.01 |
| Depressive disorder | 9.8 | 9.6 | 0.01 | 10.5 | 10.3 | 0.01 | 10.5 | 10.3 | 0.01 |
| Gastroesophageal reflux disease | 16.0 | 14.1 | 0.05 | 15.3 | 15.4 | 0.00 | 16.3 | 15.6 | 0.02 |
| Gastrointestinal hemorrhage | 2.0 | 2.6 | -0.04 | 1.9 | 1.9 | 0.00 | 2.5 | 2.3 | 0.01 |
| Human immunodeficiency virus infection | 0.5 | 0.3 | 0.03 | 0.5 | 0.5 | 0.00 | 0.4 | 0.4 | 0.00 |
| Hyperlipidemia | 81.8 | 79.2 | 0.07 | 80.8 | 81.3 | -0.01 | 80.1 | 79.9 | 0.00 |
| Hypertensive disorder | 78.8 | 78.4 | 0.01 | 78.0 | 77.6 | 0.01 | 79.0 | 78.7 | 0.01 |
| Lesion of liver | 1.2 | 1.4 | -0.02 | 1.2 | 1.0 | 0.02 | 1.6 | 1.4 | 0.02 |
| Obesity | 28.9 | 20.0 | 0.21 | 27.6 | 28.1 | -0.01 | 25.4 | 24.6 | 0.02 |
| Osteoarthritis | 22.2 | 22.6 | -0.01 | 21.9 | 21.7 | 0.00 | 24.3 | 23.2 | 0.03 |
| Pneumonia | 3.0 | 4.2 | -0.06 | 2.9 | 2.9 | 0.00 | 3.9 | 3.8 | 0.00 |
| Psoriasis | 2.0 | 1.5 | 0.04 | 1.9 | 1.9 | 0.00 | 1.8 | 1.7 | 0.01 |
| Renal impairment | 10.3 | 14.7 | -0.13 | 10.4 | 9.8 | 0.02 | 14.4 | 14.6 | 0.00 |
| Rheumatoid arthritis | 0.9 | 1.1 | -0.02 | 0.9 | 0.9 | -0.01 | 1.1 | 1.1 | 0.01 |
| Schizophrenia | 0.3 | 0.4 | -0.02 | 0.3 | 0.3 | 0.00 | 0.3 | 0.4 | -0.01 |
| Ulcerative colitis | 0.4 | 0.3 | 0.01 | 0.4 | 0.3 | 0.01 | 0.4 | 0.3 | 0.02 |
| Urinary tract infectious disease | 3.4 | 5.8 | -0.12 | 3.4 | 3.4 | 0.00 | 5.4 | 5.1 | 0.01 |
| Viral hepatitis C | 0.5 | 0.8 | -0.04 | 0.5 | 0.6 | -0.01 | 0.7 | 0.8 | -0.01 |
| Visual system disorder | 33.7 | 36.0 | -0.05 | 33.8 | 33.4 | 0.01 | 36.3 | 36.4 | 0.00 |
| **Medical history: Cardiovascular disease** | | | | | | | | | |
| Atrial fibrillation | 7.4 | 7.6 | -0.01 | 6.9 | 6.8 | 0.00 | 7.4 | 7.8 | -0.02 |
| Cerebrovascular disease | 5.4 | 6.7 | -0.05 | 5.2 | 5.1 | 0.00 | 5.9 | 6.3 | -0.01 |
| Coronary arteriosclerosis | 21.5 | 19.8 | 0.04 | 19.5 | 19.4 | 0.00 | 19.3 | 20.0 | -0.02 |
| Heart disease | 33.4 | 33.0 | 0.01 | 31.5 | 30.9 | 0.01 | 31.5 | 30.9 | 0.01 |
| Heart failure | 8.2 | 7.7 | 0.02 | 7.0 | 6.8 | 0.00 | 7.9 | 7.8 | 0.01 |
| Ischemic heart disease | 11.2 | 10.6 | 0.02 | 10.0 | 9.9 | 0.00 | 10.5 | 10.4 | 0.00 |
| Peripheral vascular disease | 10.8 | 10.6 | 0.01 | 10.3 | 10.4 | 0.00 | 11.8 | 11.4 | 0.01 |
| Pulmonary embolism | 0.7 | 0.8 | -0.01 | 0.7 | 0.7 | 0.01 | 0.8 | 0.8 | -0.01 |
| Venous thrombosis | 1.5 | 1.7 | -0.02 | 1.4 | 1.3 | 0.01 | 1.8 | 1.7 | 0.01 |
| **Medical history: Neoplasms** | | | | | | | | | |
| Hematologic neoplasm | 0.9 | 1.4 | -0.04 | 0.9 | 1.0 | 0.00 | 1.0 | 1.3 | -0.03 |
| Malignant lymphoma | 0.5 | 0.7 | -0.02 | 0.5 | 0.5 | -0.01 | 0.5 | 0.7 | -0.03 |
| Malignant neoplasm of anorectum | 0.2 | 0.3 | -0.02 | 0.3 | 0.3 | 0.01 | 0.3 | 0.3 | 0.01 |
| Malignant neoplastic disease | 8.7 | 11.4 | -0.09 | 8.7 | 8.5 | 0.01 | 10.3 | 10.8 | -0.02 |
| Malignant tumor of colon | 0.5 | 0.7 | -0.03 | 0.5 | 0.4 | 0.02 | 0.8 | 0.7 | 0.02 |
| Malignant tumor of lung | 0.2 | 0.5 | -0.06 | 0.2 | 0.3 | -0.02 | 0.3 | 0.5 | -0.03 |
| Malignant tumor of urinary bladder | 0.4 | 0.8 | -0.05 | 0.4 | 0.5 | -0.01 | 0.5 | 0.8 | -0.04 |

| (Continued) | Before ajustment | | | After matching | | | After stratification | | |
|---|---|---|---|---|---|---|---|---|---|
| Characteristic | T (%) | C (%) | StdDiff | T (%) | C (%) | StdDiff | T (%) | C (%) | StdDiff |
| Primary malignant neoplasm of prostate | 3.0 | 4.1 | -0.06 | 3.1 | 2.9 | 0.01 | 3.6 | 3.8 | -0.01 |
| **Medication use** | | | | | | | | | |
| Agents acting on the renin-angiotensin system | 73.1 | 71.6 | 0.03 | 72.3 | 72.1 | 0.01 | 72.4 | 72.5 | 0.00 |
| Antibacterials for systemic use | 49.7 | 51.7 | -0.04 | 49.3 | 48.9 | 0.01 | 51.3 | 50.5 | 0.02 |
| Antidepressants | 19.9 | 19.0 | 0.02 | 19.5 | 19.4 | 0.00 | 20.3 | 19.7 | 0.01 |
| Antiepileptics | 13.8 | 13.6 | 0.01 | 15.7 | 15.0 | 0.02 | 15.7 | 15.0 | 0.02 |
| Antiinflammatory and antirheumatic products | 27.6 | 26.2 | 0.03 | 27.2 | 27.7 | -0.01 | 28.3 | 27.2 | 0.02 |
| Antineoplastic agents | 4.2 | 4.4 | -0.01 | 4.1 | 4.0 | 0.01 | 4.1 | 4.0 | 0.01 |
| Antipsoriatics | 0.4 | 0.4 | 0.00 | 0.4 | 0.4 | 0.00 | 0.4 | 0.4 | -0.01 |
| Antithrombotic agents | 19.5 | 18.0 | 0.04 | 18.2 | 18.0 | 0.00 | 19.1 | 18.9 | 0.00 |
| Beta blocking agents | 35.3 | 35.7 | -0.01 | 33.6 | 33.2 | 0.01 | 35.9 | 35.5 | 0.01 |
| Calcium channel blockers | 25.8 | 25.9 | 0.00 | 25.6 | 25.3 | 0.01 | 26.4 | 26.6 | 0.00 |
| Diuretics | 36.9 | 38.1 | -0.03 | 35.9 | 35.7 | 0.00 | 38.0 | 37.4 | 0.01 |
| Drugs for acid related disorders | 22.2 | 22.4 | 0.00 | 21.6 | 21.5 | 0.00 | 24.3 | 23.2 | 0.03 |
| Drugs for obstructive airway diseases | 33.2 | 31.3 | 0.04 | 32.5 | 32.8 | -0.01 | 33.5 | 32.4 | 0.02 |
| Immunosuppressants | 1.8 | 1.6 | 0.01 | 1.7 | 1.7 | 0.00 | 1.7 | 1.7 | 0.00 |
| Lipid modifying agents | 78.4 | 73.8 | 0.11 | 77.3 | 77.5 | -0.01 | 75.5 | 76.0 | -0.01 |
| Opioids | 23.9 | 25.1 | -0.03 | 23.3 | 23.3 | 0.00 | 25.4 | 24.7 | 0.01 |
| Psychostimulants | 1.9 | 1.2 | 0.05 | 1.8 | 1.7 | 0.00 | 1.7 | 1.4 | 0.02 |

##### 2.3.4 MDCR

**Supplementary Table 175:** Baseline patient characteristics for SGLT2I (male) (T) and SU (male) (C) new-users in the MDCR data source. We report proportion of initiators satisfying selected base-line characteristics and the standardized difference of population proportions (StdDiff) before and after propensity score adjustment. Less extreme StdDiffs through matching and stratification suggest improved balance between patient cohorts through adjustment.

| | Before adjustment | | | After matching | | | After stratification | | |
|---|---|---|---|---|---|---|---|---|---|
| Characteristic | T (%) | C (%) | StdDiff | T (%) | C (%) | StdDiff | T (%) | C (%) | StdDiff |
| **Age group** | | | | | | | | | |
| 45 - 49 | <0.2 | 0.1 | 0.01 | 0.5 | <0.1 | 0.09 | 0.5 | <0.1 | 0.09 |
| 50 - 54 | 0.3 | 0.2 | 0.02 | 0.3 | 0.3 | 0.01 | 0.2 | 0.2 | -0.01 |
| 55 - 59 | 1.0 | 0.6 | 0.05 | 1.1 | 0.6 | 0.06 | 2.2 | 0.6 | 0.14 |
| 60 - 64 | 2.4 | 1.6 | 0.06 | 2.4 | 1.9 | 0.03 | 3.0 | 2.0 | 0.07 |
| 65 - 69 | 36.1 | 28.3 | 0.17 | 35.9 | 35.6 | 0.01 | 31.9 | 31.7 | 0.00 |
| 70 - 74 | 35.8 | 30.7 | 0.11 | 35.1 | 34.8 | 0.01 | 32.8 | 31.8 | 0.02 |
| 75 - 79 | 16.8 | 20.6 | -0.10 | 16.8 | 17.7 | -0.02 | 15.9 | 18.6 | -0.07 |
| 80 - 84 | 5.6 | 12.1 | -0.23 | 6.0 | 6.1 | -0.01 | 11.2 | 9.8 | 0.05 |
| 85 - 89 | 1.6 | 4.7 | -0.17 | 1.9 | 2.2 | -0.03 | 2.0 | 4.0 | -0.12 |
| 90 - 94 | 0.3 | 1.2 | -0.10 | 0.3 | 0.7 | -0.05 | 0.3 | 1.1 | -0.10 |
| 95 - 99 | <0.2 | 0.1 | -0.03 | <0.3 | 0.1 | -0.02 | <0.2 | 0.2 | -0.04 |
| **Medical history: General** | | | | | | | | | |
| Acute respiratory disease | 21.2 | 19.6 | 0.04 | 21.3 | 21.0 | 0.01 | 24.9 | 21.6 | 0.08 |
| Attention deficit hyperactivity disorder | 0.2 | 0.1 | 0.03 | 0.3 | 0.4 | -0.02 | 0.2 | 0.3 | -0.01 |
| Chronic liver disease | 1.8 | 1.9 | -0.01 | 1.9 | 1.5 | 0.03 | 1.5 | 2.2 | -0.06 |
| Chronic obstructive lung disease | 10.2 | 11.8 | -0.05 | 10.3 | 11.1 | -0.03 | 13.1 | 12.4 | 0.02 |
| Crohn’s disease | 0.4 | 0.2 | 0.04 | 0.4 | 0.2 | 0.03 | 0.4 | 0.2 | 0.03 |
| Dementia | 1.1 | 2.5 | -0.10 | 1.2 | 1.7 | -0.05 | 1.0 | 3.0 | -0.14 |
| Depressive disorder | 6.5 | 5.2 | 0.06 | 6.6 | 7.5 | -0.03 | 9.2 | 7.8 | 0.05 |
| Gastroesophageal reflux disease | 15.2 | 8.9 | 0.19 | 12.6 | 14.5 | -0.06 | 12.6 | 14.5 | -0.06 |
| Gastrointestinal hemorrhage | 2.6 | 3.4 | -0.04 | 2.5 | 2.1 | 0.03 | 2.2 | 2.9 | -0.04 |
| Human immunodeficiency virus infection | <0.2 | 0.1 | 0.03 | <0.3 | 0.2 | -0.03 | <0.2 | 0.1 | -0.02 |
| Hyperlipidemia | 83.0 | 50.4 | 0.74 | 81.5 | 83.2 | -0.04 | 75.7 | 76.5 | -0.02 |
| Hypertensive disorder | 84.2 | 64.6 | 0.46 | 82.7 | 83.3 | -0.02 | 80.8 | 81.2 | -0.01 |
| Lesion of liver | 1.3 | 1.1 | 0.01 | 1.1 | 1.3 | -0.02 | 1.6 | 1.4 | 0.01 |
| Obesity | 22.9 | 7.3 | 0.45 | 21.3 | 22.4 | -0.03 | 16.1 | 16.7 | -0.02 |
| Osteoarthritis | 29.4 | 23.3 | 0.14 | 28.4 | 28.7 | 0.00 | 27.0 | 29.0 | -0.04 |
| Pneumonia | 4.0 | 6.0 | -0.09 | 4.1 | 4.6 | -0.03 | 5.3 | 5.6 | -0.01 |
| Psoriasis | 2.1 | 1.3 | 0.06 | 1.7 | 2.0 | -0.02 | 2.3 | 1.8 | 0.03 |
| Renal impairment | 13.1 | 12.9 | 0.01 | 13.2 | 12.7 | 0.01 | 12.8 | 17.5 | -0.13 |
| Rheumatoid arthritis | 1.2 | 1.0 | 0.02 | 1.2 | 1.2 | 0.00 | 1.0 | 1.1 | -0.01 |
| Schizophrenia | <0.2 | 0.1 | 0.00 | <0.3 | 0.1 | 0.01 | <0.2 | 0.1 | -0.02 |
| Ulcerative colitis | 0.3 | 0.4 | 0.00 | 0.3 | 0.4 | -0.03 | 0.2 | 0.5 | -0.04 |
| Urinary tract infectious disease | 4.1 | 6.2 | -0.09 | 4.2 | 4.7 | -0.02 | 5.3 | 6.1 | -0.04 |
| Viral hepatitis C | 0.7 | 0.2 | 0.07 | 0.7 | 0.3 | 0.06 | 0.5 | 0.3 | 0.03 |
| Visual system disorder | 52.9 | 45.6 | 0.15 | 51.1 | 51.8 | -0.01 | 50.7 | 50.5 | 0.00 |
| **Medical history: Cardiovascular disease** | | | | | | | | | |
| Atrial fibrillation | 15.0 | 12.8 | 0.06 | 13.7 | 13.7 | 0.00 | 10.7 | 14.2 | -0.11 |
| Cerebrovascular disease | 11.2 | 12.5 | -0.04 | 10.4 | 10.0 | 0.01 | 10.9 | 11.6 | -0.02 |
| Coronary arteriosclerosis | 36.4 | 30.1 | 0.13 | 32.6 | 34.6 | -0.04 | 28.0 | 32.0 | -0.09 |
| Heart disease | 54.2 | 48.6 | 0.11 | 49.6 | 50.8 | -0.02 | 45.7 | 49.7 | -0.08 |
| Heart failure | 10.6 | 11.2 | -0.02 | 9.0 | 9.0 | 0.00 | 7.4 | 10.5 | -0.11 |
| Ischemic heart disease | 13.8 | 14.4 | -0.02 | 13.3 | 13.3 | 0.00 | 11.7 | 13.9 | -0.07 |
| Peripheral vascular disease | 15.0 | 11.4 | 0.11 | 14.2 | 13.6 | 0.02 | 14.4 | 13.7 | 0.02 |
| Pulmonary embolism | 0.9 | 0.8 | 0.01 | 0.9 | 1.0 | -0.01 | 1.0 | 1.1 | -0.01 |
| Venous thrombosis | 1.9 | 2.1 | -0.02 | 2.0 | 1.9 | 0.01 | 2.9 | 2.3 | 0.04 |
| **Medical history: Neoplasms** | | | | | | | | | |
| Hematologic neoplasm | 2.0 | 1.9 | 0.01 | 1.8 | 1.8 | 0.00 | 1.5 | 2.3 | -0.05 |
| Malignant lymphoma | 1.4 | 0.9 | 0.04 | 1.1 | 1.3 | -0.02 | 0.7 | 1.3 | -0.06 |
| Malignant neoplasm of anorectum | 0.2 | 0.5 | -0.04 | <0.3 | 0.3 | -0.02 | <0.2 | 0.4 | -0.06 |
| Malignant neoplastic disease | 18.4 | 21.1 | -0.07 | 17.7 | 18.7 | -0.03 | 16.2 | 21.1 | -0.13 |
| Malignant tumor of breast | <0.2 | 0.1 | -0.02 | <0.3 | 0.1 | -0.02 | <0.2 | 0.1 | -0.04 |
| Malignant tumor of colon | 0.5 | 1.1 | -0.06 | 0.5 | 0.7 | -0.03 | 0.2 | 1.0 | -0.10 |
| Malignant tumor of lung | 0.3 | 0.9 | -0.08 | 0.3 | 0.6 | -0.04 | 0.4 | 1.0 | -0.08 |
| Malignant tumor of urinary bladder | 1.4 | 1.7 | -0.03 | 1.4 | 1.2 | 0.01 | 1.6 | 1.7 | -0.01 |
| Primary malignant neoplasm of prostate | 7.1 | 8.3 | -0.04 | 7.0 | 7.2 | -0.01 | 6.4 | 8.1 | -0.06 |
| **Medication use** | | | | | | | | | |
| Agents acting on the renin-angiotensin system | 77.6 | 72.4 | 0.12 | 76.5 | 75.9 | 0.01 | 77.2 | 75.3 | 0.04 |

| (Continued) | Before ajustment | | | After matching | | | After stratification | | |
|---|---|---|---|---|---|---|---|---|---|
| Characteristic | T (%) | C (%) | StdDiff | T (%) | C (%) | StdDiff | T (%) | C (%) | StdDiff |
| Antibacterials for systemic use | 59.8 | 57.7 | 0.04 | 58.4 | 58.6 | 0.00 | 59.2 | 58.0 | 0.02 |
| Antidepressants | 18.4 | 18.6 | -0.01 | 18.1 | 19.4 | -0.04 | 21.9 | 19.6 | 0.06 |
| Antiepileptics | 14.3 | 12.1 | 0.06 | 14.4 | 14.6 | 0.00 | 13.5 | 14.7 | -0.03 |
| Antiinflammatory and antirheumatic products | 26.0 | 25.0 | 0.02 | 25.9 | 26.2 | -0.01 | 28.1 | 25.9 | 0.05 |
| Antineoplastic agents | 7.7 | 8.3 | -0.02 | 7.5 | 7.2 | 0.01 | 7.9 | 7.6 | 0.01 |
| Antipsoriatics | 0.5 | 0.6 | -0.02 | 0.5 | 0.5 | 0.00 | 0.3 | 0.5 | -0.04 |
| Antithrombotic agents | 30.2 | 28.2 | 0.04 | 28.2 | 28.2 | 0.00 | 24.1 | 28.9 | -0.11 |
| Beta blocking agents | 51.1 | 49.5 | 0.03 | 48.9 | 49.7 | -0.02 | 48.2 | 50.4 | -0.04 |
| Calcium channel blockers | 32.9 | 31.7 | 0.03 | 31.9 | 32.4 | -0.01 | 31.6 | 33.8 | -0.05 |
| Diuretics | 45.1 | 47.7 | -0.05 | 43.6 | 42.6 | 0.02 | 45.9 | 45.1 | 0.02 |
| Drugs for acid related disorders | 30.5 | 28.9 | 0.04 | 29.8 | 29.7 | 0.00 | 30.0 | 29.3 | 0.01 |
| Drugs for obstructive airway diseases | 41.1 | 35.8 | 0.11 | 39.3 | 40.3 | -0.02 | 38.7 | 39.0 | -0.01 |
| Immunosuppressants | 2.7 | 2.0 | 0.04 | 2.6 | 2.8 | -0.01 | 2.0 | 2.6 | -0.04 |
| Lipid modifying agents | 85.8 | 76.9 | 0.23 | 84.1 | 85.3 | -0.04 | 81.7 | 82.0 | -0.01 |
| Opioids | 24.1 | 26.0 | -0.04 | 23.9 | 24.5 | -0.01 | 29.7 | 27.3 | 0.05 |
| Psycholeptics | 20.5 | 21.8 | -0.03 | 20.5 | 19.8 | 0.02 | 25.6 | 21.8 | 0.09 |
| Psychostimulants | 1.3 | 0.8 | 0.04 | 1.4 | 1.1 | 0.03 | 1.1 | 1.0 | 0.01 |

##### 2.3.5 MDCD

**Supplementary Table 176:** Baseline patient characteristics for SGLT2I (male) (T) and SU (male) (C) new-users in the MDCD data source. We report proportion of initiators satisfying selected base-line characteristics and the standardized difference of population proportions (StdDiff) before and after propensity score adjustment. Less extreme StdDiffs through matching and stratification suggest improved balance between patient cohorts through adjustment.

| | Before adjustment | | | After matching | | | After stratification | | |
|---|---|---|---|---|---|---|---|---|---|
| Characteristic | T (%) | C (%) | StdDiff | T (%) | C (%) | StdDiff | T (%) | C (%) | StdDiff |
| **Age group** | | | | | | | | | |
| 15 - 19 | 0.7 | 0.6 | 0.01 | 0.7 | 0.8 | -0.01 | 0.7 | 0.6 | 0.01 |
| 20 - 24 | 1.1 | 0.9 | 0.01 | 1.1 | 1.1 | 0.00 | 1.9 | 1.0 | 0.08 |
| 25 - 29 | 2.5 | 2.0 | 0.04 | 2.5 | 1.9 | 0.04 | 1.6 | 1.9 | -0.02 |
| 30 - 34 | 5.3 | 4.1 | 0.06 | 5.2 | 4.7 | 0.02 | 5.3 | 4.3 | 0.05 |
| 35 - 39 | 8.6 | 7.5 | 0.04 | 8.5 | 8.4 | 0.01 | 8.1 | 7.8 | 0.01 |
| 40 - 44 | 11.1 | 10.4 | 0.02 | 11.0 | 11.6 | -0.02 | 11.3 | 10.7 | 0.02 |
| 45 - 49 | 16.3 | 13.7 | 0.07 | 16.1 | 15.3 | 0.02 | 13.7 | 13.8 | 0.00 |
| 50 - 54 | 15.9 | 17.1 | -0.04 | 16.1 | 16.9 | -0.02 | 16.5 | 17.2 | -0.02 |
| 55 - 59 | 20.0 | 18.9 | 0.03 | 20.0 | 19.0 | 0.03 | 22.7 | 19.3 | 0.08 |
| 60 - 64 | 13.9 | 15.9 | -0.06 | 14.0 | 14.3 | -0.01 | 14.0 | 16.2 | -0.06 |
| 65 - 69 | 2.7 | 4.6 | -0.10 | 2.7 | 3.4 | -0.04 | 2.4 | 4.0 | -0.10 |
| 70 - 74 | 1.3 | 2.1 | -0.07 | 1.3 | 1.5 | -0.02 | 0.9 | 1.7 | -0.06 |
| 75 - 79 | 0.3 | 1.1 | -0.09 | 0.3 | 0.7 | -0.05 | <0.2 | 0.8 | -0.08 |
| 80 - 84 | 0.4 | 0.7 | -0.04 | 0.4 | 0.3 | 0.02 | 0.6 | 0.4 | 0.02 |
| **Race** | | | | | | | | | |
| Black or African American | 19.5 | 23.3 | -0.09 | 19.5 | 16.2 | 0.09 | 27.9 | 22.4 | 0.12 |
| White | 65.2 | 56.8 | 0.17 | 64.9 | 67.7 | -0.06 | 56.1 | 57.7 | -0.03 |
| **Ethnicity** | | | | | | | | | |
| Hispanic or Latino | 1.9 | 2.6 | -0.05 | 1.8 | 2.2 | -0.03 | 1.6 | 2.6 | -0.07 |
| **Medical history: General** | | | | | | | | | |
| Acute respiratory disease | 27.3 | 27.8 | -0.01 | 27.3 | 27.5 | 0.00 | 26.5 | 27.6 | -0.02 |
| Attention deficit hyperactivity disorder | 2.9 | 1.9 | 0.07 | 2.9 | 2.8 | 0.00 | 2.0 | 2.3 | -0.02 |
| Chronic liver disease | 3.5 | 5.9 | -0.12 | 3.5 | 3.5 | 0.00 | 6.8 | 5.1 | 0.07 |
| Chronic obstructive lung disease | 17.7 | 18.4 | -0.02 | 17.4 | 16.8 | 0.01 | 19.0 | 17.7 | 0.03 |
| Crohn's disease | 0.3 | 0.3 | 0.01 | 0.3 | 0.2 | 0.02 | <0.2 | 0.3 | -0.01 |
| Dementia | 0.9 | 1.8 | -0.07 | 1.0 | 1.1 | -0.01 | 0.9 | 1.6 | -0.06 |
| Depressive disorder | 27.0 | 25.7 | 0.03 | 26.9 | 28.2 | -0.03 | 26.8 | 27.0 | 0.00 |
| Gastroesophageal reflux disease | 23.0 | 22.3 | 0.01 | 23.3 | 24.3 | -0.02 | 24.1 | 23.8 | 0.01 |
| Gastrointestinal hemorrhage | 2.9 | 4.0 | -0.06 | 3.0 | 3.1 | 0.00 | 3.3 | 3.6 | -0.02 |
| Human immunodeficiency virus infection | 0.9 | 1.0 | -0.01 | 0.9 | 0.8 | 0.01 | 1.3 | 1.0 | 0.03 |
| Hyperlipidemia | 69.7 | 66.1 | 0.08 | 69.6 | 70.2 | -0.01 | 67.6 | 67.3 | 0.01 |
| Hypertensive disorder | 77.7 | 78.2 | -0.01 | 77.5 | 78.0 | -0.01 | 80.2 | 78.7 | 0.04 |
| Lesion of liver | 1.3 | 2.6 | -0.09 | 1.3 | 2.0 | -0.05 | 1.1 | 2.4 | -0.10 |
| Obesity | 36.2 | 29.3 | 0.15 | 35.6 | 37.8 | -0.05 | 33.3 | 33.4 | 0.00 |
| Osteoarthritis | 31.1 | 31.1 | 0.00 | 31.2 | 32.2 | -0.02 | 34.9 | 31.6 | 0.07 |
| Pneumonia | 4.1 | 5.8 | -0.08 | 4.2 | 4.5 | -0.02 | 4.7 | 5.3 | -0.03 |
| Psoriasis | 1.1 | 1.2 | -0.01 | 1.0 | 1.5 | -0.05 | 0.6 | 1.3 | -0.06 |
| Renal impairment | 6.7 | 10.2 | -0.12 | 6.7 | 6.6 | 0.01 | 8.4 | 10.0 | -0.06 |
| Rheumatoid arthritis | 0.8 | 1.1 | -0.03 | 0.8 | 1.0 | -0.02 | 0.5 | 1.1 | -0.07 |
| Schizophrenia | 4.4 | 6.0 | -0.07 | 4.4 | 4.9 | -0.02 | 4.6 | 5.6 | -0.04 |
| Ulcerative colitis | 0.3 | 0.3 | 0.01 | 0.3 | 0.2 | 0.02 | 0.3 | 0.3 | 0.00 |
| Urinary tract infectious disease | 2.5 | 5.0 | -0.13 | 2.5 | 2.9 | -0.02 | 3.5 | 4.0 | -0.03 |
| Viral hepatitis C | 2.0 | 4.0 | -0.12 | 2.1 | 2.4 | -0.03 | 3.8 | 3.6 | 0.01 |
| Visual system disorder | 41.1 | 38.3 | 0.06 | 40.9 | 42.4 | -0.03 | 39.5 | 39.7 | 0.00 |
| **Medical history: Cardiovascular disease** | | | | | | | | | |
| Atrial fibrillation | 4.1 | 4.6 | -0.03 | 3.8 | 4.1 | -0.01 | 3.4 | 4.5 | -0.06 |
| Cerebrovascular disease | 3.8 | 5.8 | -0.10 | 3.8 | 4.1 | -0.02 | 3.8 | 4.9 | -0.05 |
| Coronary arteriosclerosis | 15.7 | 16.1 | -0.01 | 15.3 | 14.5 | 0.02 | 15.1 | 15.4 | -0.01 |
| Heart disease | 28.9 | 32.0 | -0.07 | 28.5 | 28.2 | 0.01 | 28.0 | 30.5 | -0.05 |
| Heart failure | 9.6 | 10.2 | -0.02 | 9.4 | 8.5 | 0.03 | 9.6 | 9.5 | 0.01 |
| Ischemic heart disease | 10.2 | 11.3 | -0.04 | 9.9 | 10.5 | -0.02 | 11.0 | 10.9 | 0.00 |
| Peripheral vascular disease | 9.0 | 9.0 | 0.00 | 8.7 | 8.2 | 0.02 | 6.9 | 8.8 | -0.07 |
| Pulmonary embolism | 0.9 | 1.1 | -0.03 | 0.9 | 0.9 | 0.00 | 1.0 | 1.1 | -0.02 |
| Venous thrombosis | 1.7 | 2.1 | -0.04 | 1.7 | 1.6 | 0.01 | 2.7 | 2.0 | 0.04 |
| **Medical history: Neoplasms** | | | | | | | | | |
| Hematologic neoplasm | 0.7 | 1.0 | -0.03 | 0.8 | 0.7 | 0.00 | 0.6 | 0.9 | -0.04 |
| Malignant lymphoma | <0.3 | 0.4 | -0.02 | <0.3 | 0.4 | -0.02 | 0.2 | 0.4 | -0.03 |
| Malignant neoplasm of anorectum | <0.3 | 0.4 | -0.04 | <0.3 | 0.2 | -0.02 | <0.2 | 0.3 | -0.05 |

| (Continued) | Before ajustment | | | After matching | | | After stratification | | |
|---|---|---|---|---|---|---|---|---|---|
| Characteristic | T (%) | C (%) | StdDiff | T (%) | C (%) | StdDiff | T (%) | C (%) | StdDiff |
| Malignant neoplastic disease | 3.5 | 5.5 | -0.10 | 3.6 | 4.0 | -0.03 | 3.4 | 5.1 | -0.09 |
| Malignant tumor of breast | <0.3 | <0.1 | 0.03 | <0.2 | <0.1 | 0.02 | <0.2 | <0.1 | 0.02 |
| Malignant tumor of colon | 0.3 | 0.5 | -0.03 | 0.3 | 0.3 | 0.01 | <0.2 | 0.5 | -0.04 |
| Malignant tumor of lung | <0.3 | 0.4 | -0.05 | <0.3 | 0.3 | -0.03 | <0.2 | 0.4 | -0.07 |
| Malignant tumor of urinary bladder | <0.3 | 0.3 | -0.02 | <0.3 | 0.3 | -0.01 | <0.2 | 0.3 | -0.02 |
| Primary malignant neoplasm of prostate | 0.9 | 1.3 | -0.03 | 1.0 | 0.8 | 0.02 | 1.2 | 1.1 | 0.01 |
| **Medication use** | | | | | | | | | |
| Agents acting on the renin-angiotensin system | 67.9 | 70.0 | -0.04 | 67.9 | 68.4 | -0.01 | 68.5 | 69.9 | -0.03 |
| Antibacterials for systemic use | 57.3 | 58.5 | -0.02 | 57.1 | 57.4 | 0.00 | 58.0 | 57.7 | 0.00 |
| Antidepressants | 42.1 | 40.5 | 0.03 | 41.9 | 42.6 | -0.01 | 42.4 | 40.8 | 0.03 |
| Antiepileptics | 37.3 | 34.4 | 0.06 | 37.3 | 36.0 | 0.03 | 35.8 | 35.7 | 0.00 |
| Antiinflammatory and antirheumatic products | 55.7 | 53.3 | 0.05 | 55.8 | 56.0 | 0.00 | 60.4 | 55.4 | 0.10 |
| Antineoplastic agents | 2.6 | 3.0 | -0.03 | 2.5 | 2.7 | -0.01 | 2.3 | 2.8 | -0.03 |
| Antipsoriatics | <0.3 | 0.4 | -0.02 | <0.3 | 0.3 | -0.03 | <0.2 | 0.4 | -0.05 |
| Antithrombotic agents | 26.5 | 28.0 | -0.03 | 26.5 | 26.4 | 0.00 | 25.5 | 28.5 | -0.07 |
| Beta blocking agents | 35.5 | 36.7 | -0.02 | 35.1 | 35.8 | -0.01 | 34.0 | 36.1 | -0.04 |
| Calcium channel blockers | 24.4 | 26.7 | -0.05 | 24.3 | 23.6 | 0.02 | 27.2 | 26.4 | 0.02 |
| Diuretics | 39.8 | 42.8 | -0.06 | 39.6 | 39.5 | 0.00 | 42.0 | 41.9 | 0.00 |
| Drugs for acid related disorders | 41.2 | 39.1 | 0.04 | 41.6 | 40.8 | 0.02 | 41.1 | 39.1 | 0.04 |
| Drugs for obstructive airway diseases | 48.5 | 46.7 | 0.04 | 48.0 | 48.9 | -0.02 | 48.1 | 47.4 | 0.01 |
| Immunosuppressants | 1.7 | 1.7 | 0.00 | 1.5 | 2.1 | -0.04 | 1.0 | 1.9 | -0.07 |
| Lipid modifying agents | 70.9 | 67.8 | 0.07 | 70.5 | 71.7 | -0.03 | 64.3 | 69.0 | -0.10 |
| Opioids | 39.2 | 42.9 | -0.07 | 39.4 | 38.2 | 0.03 | 43.6 | 41.4 | 0.04 |
| Psycholeptics | 42.1 | 41.2 | 0.02 | 41.9 | 42.4 | -0.01 | 40.0 | 40.9 | -0.02 |
| Psychostimulants | 4.2 | 2.8 | 0.07 | 4.2 | 3.6 | 0.03 | 3.5 | 3.1 | 0.02 |

##### 2.3.6 Open Claims

**Supplementary Table 177:** Baseline patient characteristics for SGLT2I (male) (T) and SU (male) (C) new-users in the Open Claims data source. We report proportion of initiators satisfying selected base-line characteristics and the standardized difference of population proportions (StdDiff) before and after propensity score adjustment. Less extreme StdDiffs through matching and stratification suggest improved balance between patient cohorts through adjustment.

| | Before adjustment | | | After matching | | | After stratification | | |
|---|---|---|---|---|---|---|---|---|---|
| Characteristic | T (%) | C (%) | StdDiff | T (%) | C (%) | StdDiff | T (%) | C (%) | StdDiff |
| **Age group** | | | | | | | | | |
| 15 - 19 | 0.1 | 0.1 | 0.00 | 0.1 | 0.1 | 0.00 | 0.1 | 0.1 | 0.00 |
| 20 - 24 | 0.3 | 0.2 | 0.00 | 0.3 | 0.3 | 0.01 | 0.3 | 0.3 | 0.00 |
| 25 - 29 | 0.6 | 0.6 | 0.00 | 0.6 | 0.6 | 0.00 | 0.6 | 0.6 | 0.00 |
| 30 - 34 | 1.5 | 1.4 | 0.00 | 1.5 | 1.5 | 0.00 | 1.5 | 1.5 | 0.00 |
| 35 - 39 | 3.4 | 3.0 | 0.02 | 3.5 | 3.6 | -0.01 | 3.1 | 3.2 | 0.00 |
| 40 - 44 | 6.1 | 5.3 | 0.03 | 6.2 | 6.4 | -0.01 | 5.7 | 5.7 | 0.00 |
| 45 - 49 | 9.9 | 8.6 | 0.05 | 10.1 | 10.0 | 0.00 | 9.1 | 9.0 | 0.01 |
| 50 - 54 | 13.9 | 11.8 | 0.06 | 13.8 | 14.0 | -0.01 | 12.6 | 12.5 | 0.00 |
| 55 - 59 | 16.6 | 14.4 | 0.06 | 15.2 | 14.9 | 0.01 | 15.2 | 14.9 | 0.01 |
| 60 - 64 | 17.0 | 14.9 | 0.06 | 16.7 | 16.9 | 0.00 | 16.0 | 15.3 | 0.02 |
| 65 - 69 | 13.3 | 14.8 | -0.04 | 13.3 | 13.5 | 0.00 | 14.2 | 14.4 | -0.01 |
| 70 - 74 | 9.0 | 11.3 | -0.08 | 9.0 | 8.7 | 0.01 | 10.4 | 10.5 | -0.01 |
| 75 - 79 | 5.4 | 9.9 | -0.17 | 5.5 | 5.1 | 0.02 | 7.8 | 8.6 | -0.03 |
| 80 - 84 | 2.9 | 3.6 | -0.04 | 2.9 | 2.8 | 0.00 | 3.3 | 3.4 | -0.01 |
| 85 - 89 | 0.2 | 0.1 | 0.03 | 0.2 | 0.2 | 0.00 | 0.1 | 0.1 | 0.00 |
| **Medical history: General** | | | | | | | | | |
| Acute respiratory disease | 11.0 | 11.0 | 0.00 | 10.8 | 10.6 | 0.01 | 11.5 | 10.9 | 0.02 |
| Attention deficit hyperactivity disorder | 0.6 | 0.4 | 0.03 | 0.6 | 0.5 | 0.00 | 0.5 | 0.4 | 0.01 |
| Chronic liver disease | 1.0 | 1.4 | -0.04 | 1.0 | 1.0 | 0.00 | 1.3 | 1.3 | 0.00 |
| Chronic obstructive lung disease | 4.2 | 5.4 | -0.06 | 4.1 | 4.1 | 0.00 | 5.2 | 5.1 | 0.01 |
| Crohn’s disease | 0.1 | 0.1 | 0.00 | 0.1 | 0.1 | 0.00 | 0.1 | 0.2 | 0.00 |
| Dementia | 0.4 | 0.8 | -0.06 | 0.4 | 0.4 | 0.00 | 0.6 | 0.7 | 0.00 |
| Depressive disorder | 4.6 | 5.0 | -0.02 | 4.6 | 4.7 | -0.01 | 5.1 | 5.0 | 0.01 |
| Gastroesophageal reflux disease | 6.9 | 6.5 | 0.02 | 6.7 | 6.8 | 0.00 | 6.8 | 6.6 | 0.01 |
| Gastrointestinal hemorrhage | 1.1 | 1.4 | -0.02 | 1.1 | 1.0 | 0.01 | 1.4 | 1.3 | 0.01 |
| Human immunodeficiency virus infection | 0.4 | 0.3 | 0.00 | 0.3 | 0.3 | 0.00 | 0.3 | 0.3 | 0.00 |
| Hyperlipidemia | 45.8 | 41.8 | 0.08 | 44.6 | 44.6 | 0.00 | 43.6 | 42.8 | 0.02 |
| Hypertensive disorder | 49.8 | 48.5 | 0.03 | 48.8 | 48.4 | 0.01 | 49.2 | 48.8 | 0.01 |
| Lesion of liver | 0.6 | 0.8 | -0.03 | 0.6 | 0.6 | 0.00 | 0.8 | 0.8 | 0.00 |
| Obesity | 11.4 | 9.1 | 0.07 | 10.1 | 9.8 | 0.01 | 10.1 | 9.8 | 0.01 |
| Osteoarthritis | 11.8 | 12.3 | -0.02 | 11.4 | 11.9 | -0.01 | 12.4 | 12.2 | 0.00 |
| Pneumonia | 2.0 | 2.6 | -0.04 | 1.9 | 1.8 | 0.01 | 2.5 | 2.4 | 0.01 |
| Psoriasis | 1.0 | 0.8 | 0.02 | 0.9 | 1.0 | 0.00 | 0.9 | 0.9 | 0.00 |
| Renal impairment | 5.4 | 7.4 | -0.08 | 5.0 | 4.8 | 0.01 | 6.6 | 6.8 | -0.01 |
| Rheumatoid arthritis | 0.5 | 0.6 | 0.00 | 0.5 | 0.5 | 0.00 | 0.6 | 0.6 | 0.00 |
| Schizophrenia | 0.3 | 0.4 | -0.03 | 0.3 | 0.3 | -0.01 | 0.4 | 0.4 | 0.00 |
| Ulcerative colitis | 0.2 | 0.2 | 0.00 | 0.2 | 0.2 | 0.01 | 0.2 | 0.2 | 0.00 |
| Urinary tract infectious disease | 1.8 | 2.7 | -0.06 | 1.8 | 1.9 | 0.00 | 2.6 | 2.4 | 0.01 |
| Viral hepatitis C | 0.3 | 0.5 | -0.04 | 0.3 | 0.3 | 0.00 | 0.4 | 0.5 | 0.00 |
| Visual system disorder | 15.5 | 16.8 | -0.04 | 16.6 | 16.4 | 0.00 | 16.6 | 16.4 | 0.00 |
| **Medical history: Cardiovascular disease** | | | | | | | | | |
| Atrial fibrillation | 4.6 | 4.6 | 0.00 | 4.1 | 3.9 | 0.01 | 4.5 | 4.5 | 0.00 |
| Cerebrovascular disease | 2.6 | 3.3 | -0.04 | 2.5 | 2.5 | 0.00 | 3.1 | 3.1 | 0.00 |
| Coronary arteriosclerosis | 11.9 | 10.4 | 0.05 | 10.5 | 10.2 | 0.01 | 10.8 | 10.7 | 0.00 |
| Heart disease | 21.1 | 19.7 | 0.04 | 19.1 | 18.5 | 0.02 | 20.0 | 19.8 | 0.01 |
| Heart failure | 5.2 | 4.4 | 0.04 | 4.1 | 3.9 | 0.01 | 4.5 | 4.4 | 0.00 |
| Ischemic heart disease | 5.5 | 4.8 | 0.03 | 4.8 | 4.6 | 0.01 | 5.0 | 4.9 | 0.00 |
| Peripheral vascular disease | 4.6 | 4.8 | -0.01 | 4.4 | 4.4 | 0.00 | 4.8 | 4.8 | 0.00 |
| Pulmonary embolism | 0.4 | 0.5 | -0.01 | 0.4 | 0.5 | -0.01 | 0.5 | 0.5 | 0.01 |
| Venous thrombosis | 0.8 | 0.9 | -0.01 | 0.7 | 0.7 | 0.00 | 0.9 | 0.9 | 0.00 |
| **Medical history: Neoplasms** | | | | | | | | | |
| Hematologic neoplasm | 0.6 | 0.8 | -0.03 | 0.6 | 0.6 | -0.01 | 0.7 | 0.7 | 0.00 |
| Malignant lymphoma | 0.3 | 0.4 | -0.02 | 0.3 | 0.4 | -0.01 | 0.4 | 0.4 | 0.00 |
| Malignant neoplasm of anorectum | 0.1 | 0.2 | -0.01 | 0.1 | 0.1 | 0.00 | 0.2 | 0.1 | 0.01 |
| Malignant neoplastic disease | 4.9 | 6.5 | -0.07 | 4.9 | 4.8 | 0.00 | 6.2 | 6.1 | 0.00 |
| Malignant tumor of colon | 0.3 | 0.4 | -0.02 | 0.3 | 0.2 | 0.00 | 0.4 | 0.4 | 0.01 |
| Malignant tumor of lung | 0.2 | 0.3 | -0.04 | 0.2 | 0.2 | 0.00 | 0.3 | 0.3 | -0.01 |

| (Continued) | Before ajustment | | | After matching | | | After stratification | | |
|---|---|---|---|---|---|---|---|---|---|
| Characteristic | T (%) | C (%) | StdDiff | T (%) | C (%) | StdDiff | T (%) | C (%) | StdDiff |
| Malignant tumor of urinary bladder | 0.3 | 0.5 | -0.03 | 0.3 | 0.3 | -0.01 | 0.4 | 0.4 | -0.01 |
| Primary malignant neoplasm of prostate | 1.7 | 2.2 | -0.04 | 1.7 | 1.6 | 0.01 | 2.1 | 2.0 | 0.00 |
| **Medication use** | | | | | | | | | |
| Agents acting on the renin-angiotensin system | 71.5 | 70.5 | 0.02 | 70.7 | 70.4 | 0.01 | 70.3 | 70.7 | -0.01 |
| Antibacterials for systemic use | 48.9 | 49.2 | 0.00 | 48.5 | 48.5 | 0.00 | 50.3 | 49.2 | 0.02 |
| Antidepressants | 21.1 | 20.9 | 0.00 | 20.7 | 20.9 | 0.00 | 21.4 | 21.0 | 0.01 |
| Antiepileptics | 14.8 | 16.4 | -0.05 | 14.6 | 14.8 | 0.00 | 16.5 | 16.1 | 0.01 |
| Antiinflammatory and antirheumatic products | 32.1 | 32.0 | 0.00 | 31.9 | 31.7 | 0.00 | 32.9 | 32.0 | 0.02 |
| Antineoplastic agents | 4.0 | 3.6 | 0.02 | 3.8 | 3.9 | 0.00 | 3.8 | 3.7 | 0.00 |
| Antipsoriatics | 0.5 | 0.4 | 0.01 | 0.5 | 0.5 | 0.00 | 0.5 | 0.5 | 0.00 |
| Antithrombotic agents | 21.3 | 20.9 | 0.01 | 19.8 | 19.2 | 0.02 | 20.9 | 20.8 | 0.00 |
| Beta blocking agents | 35.1 | 34.3 | 0.02 | 33.3 | 32.4 | 0.02 | 34.1 | 34.2 | 0.00 |
| Calcium channel blockers | 25.7 | 25.4 | 0.01 | 25.3 | 24.9 | 0.01 | 25.3 | 25.5 | -0.01 |
| Diuretics | 36.3 | 36.3 | 0.00 | 35.1 | 34.9 | 0.00 | 35.9 | 36.1 | 0.00 |
| Drugs for acid related disorders | 27.4 | 27.0 | 0.01 | 26.5 | 26.6 | 0.00 | 27.4 | 27.1 | 0.01 |
| Drugs for obstructive airway diseases | 31.6 | 29.7 | 0.04 | 30.7 | 30.5 | 0.00 | 30.8 | 30.1 | 0.01 |
| Immunosuppressants | 1.9 | 1.6 | 0.02 | 1.8 | 1.8 | 0.00 | 1.7 | 1.7 | 0.00 |
| Lipid modifying agents | 75.3 | 70.6 | 0.11 | 74.0 | 73.3 | 0.01 | 71.7 | 71.7 | 0.00 |
| Opioids | 20.7 | 22.9 | -0.05 | 20.5 | 20.5 | 0.00 | 23.0 | 22.5 | 0.01 |
| Psycholeptics | 17.4 | 18.2 | -0.02 | 16.9 | 17.1 | 0.00 | 18.5 | 18.0 | 0.01 |
| Psychostimulants | 2.3 | 1.6 | 0.05 | 2.1 | 2.2 | 0.00 | 2.0 | 1.9 | 0.01 |

##### 2.3.7 OEHR

**Supplementary Table 178:** Baseline patient characteristics for SGLT2I (male) (T) and SU (male) (C) new-users in the OEHR data source. We report proportion of initiators satisfying selected base-line characteristics and the standardized difference of population proportions (StdDiff) before and after propensity score adjustment. Less extreme StdDiffs through matching and stratification suggest improved balance between patient cohorts through adjustment.

| | Before adjustment | | | After matching | | | After stratification | | |
|---|---|---|---|---|---|---|---|---|---|
| Characteristic | T (%) | C (%) | StdDiff | T (%) | C (%) | StdDiff | T (%) | C (%) | StdDiff |
| **Age group** | | | | | | | | | |
| 25 - 29 | 0.7 | 0.6 | 0.00 | 0.7 | 0.6 | 0.01 | 0.7 | 0.6 | 0.01 |
| 30 - 34 | 1.9 | 1.8 | 0.00 | 1.9 | 1.8 | 0.01 | 1.7 | 1.8 | -0.01 |
| 35 - 39 | 4.3 | 3.9 | 0.02 | 4.4 | 4.4 | 0.00 | 4.4 | 4.0 | 0.02 |
| 40 - 44 | 8.0 | 6.7 | 0.05 | 6.8 | 6.9 | 0.00 | 6.8 | 6.9 | 0.00 |
| 45 - 49 | 11.9 | 10.4 | 0.05 | 12.0 | 12.0 | 0.00 | 10.2 | 10.5 | -0.01 |
| 50 - 54 | 15.7 | 13.8 | 0.06 | 15.6 | 15.7 | 0.00 | 13.7 | 14.0 | -0.01 |
| 55 - 59 | 18.1 | 15.7 | 0.06 | 17.7 | 18.0 | -0.01 | 16.9 | 16.3 | 0.01 |
| 60 - 64 | 17.3 | 14.8 | 0.07 | 16.9 | 17.4 | -0.01 | 16.4 | 15.6 | 0.02 |
| 65 - 69 | 10.5 | 12.4 | -0.06 | 10.6 | 10.5 | 0.00 | 12.4 | 12.0 | 0.01 |
| 70 - 74 | 6.5 | 9.0 | -0.10 | 6.7 | 6.4 | 0.01 | 7.6 | 8.4 | -0.03 |
| 75 - 79 | 3.2 | 6.6 | -0.16 | 3.3 | 3.2 | 0.01 | 5.1 | 5.3 | -0.01 |
| 80 - 84 | 1.4 | 3.4 | -0.13 | 1.5 | 1.4 | 0.01 | 3.1 | 3.3 | -0.01 |
| 85 - 89 | 0.4 | 0.7 | -0.05 | 0.4 | 0.4 | 0.00 | 0.8 | 0.8 | 0.00 |
| **Race** | | | | | | | | | |
| Asian | 2.3 | 2.4 | 0.00 | 2.2 | 2.4 | -0.02 | 2.2 | 2.4 | -0.02 |
| Black or African American | 8.2 | 8.6 | -0.01 | 8.2 | 8.3 | 0.00 | 8.4 | 8.7 | -0.01 |
| White | 81.8 | 81.0 | 0.02 | 81.6 | 81.7 | 0.00 | 81.6 | 80.7 | 0.02 |
| **Ethnicity** | | | | | | | | | |
| Hispanic or Latino | 5.2 | 6.6 | -0.06 | 6.1 | 6.4 | -0.01 | 6.1 | 6.4 | -0.01 |
| Not Hispanic or Latino | 85.6 | 85.7 | 0.00 | 85.9 | 86.0 | 0.00 | 86.3 | 85.5 | 0.02 |
| **Medical history: General** | | | | | | | | | |
| Acute respiratory disease | 15.0 | 13.9 | 0.03 | 14.7 | 14.9 | 0.00 | 14.4 | 14.5 | 0.00 |
| Attention deficit hyperactivity disorder | 1.0 | 0.7 | 0.03 | 1.0 | 1.1 | -0.01 | 1.0 | 1.1 | -0.01 |
| Chronic liver disease | 1.9 | 2.7 | -0.06 | 2.0 | 2.0 | 0.00 | 2.4 | 2.5 | -0.01 |
| Chronic obstructive lung disease | 4.3 | 5.3 | -0.05 | 4.4 | 4.3 | 0.00 | 5.6 | 5.3 | 0.01 |
| Crohn's disease | 0.2 | 0.2 | 0.00 | 0.2 | 0.2 | 0.01 | 0.3 | 0.2 | 0.01 |
| Dementia | 0.3 | 0.8 | -0.06 | 0.4 | 0.4 | -0.01 | 0.6 | 0.8 | -0.02 |
| Depressive disorder | 9.3 | 9.4 | 0.00 | 9.3 | 9.2 | 0.00 | 10.5 | 9.6 | 0.03 |
| Gastroesophageal reflux disease | 14.9 | 12.8 | 0.06 | 14.5 | 14.6 | 0.00 | 15.1 | 13.9 | 0.03 |
| Gastrointestinal hemorrhage | 1.2 | 1.5 | -0.03 | 1.1 | 1.2 | 0.00 | 1.4 | 1.4 | 0.00 |
| Human immunodeficiency virus infection | 0.3 | 0.3 | 0.00 | 0.3 | 0.3 | 0.00 | 0.3 | 0.3 | 0.00 |
| Hyperlipidemia | 69.9 | 65.6 | 0.09 | 68.2 | 68.6 | -0.01 | 66.4 | 66.0 | 0.01 |
| Hypertensive disorder | 68.7 | 65.4 | 0.07 | 67.4 | 67.7 | -0.01 | 66.9 | 66.2 | 0.01 |
| Lesion of liver | 0.7 | 0.9 | -0.03 | 0.7 | 0.7 | 0.00 | 1.1 | 0.9 | 0.02 |
| Obesity | 31.3 | 21.9 | 0.22 | 29.9 | 30.6 | -0.01 | 26.2 | 25.4 | 0.02 |
| Osteoarthritis | 13.4 | 14.0 | -0.02 | 13.3 | 13.3 | 0.00 | 15.0 | 14.4 | 0.02 |
| Pneumonia | 1.5 | 2.0 | -0.04 | 1.5 | 1.5 | -0.01 | 2.2 | 1.9 | 0.02 |
| Psoriasis | 1.4 | 1.3 | 0.01 | 1.4 | 1.4 | 0.00 | 1.4 | 1.4 | 0.00 |
| Renal impairment | 4.8 | 6.9 | -0.09 | 4.9 | 4.6 | 0.01 | 6.8 | 6.8 | 0.00 |
| Rheumatoid arthritis | 0.6 | 0.6 | -0.01 | 0.8 | 0.6 | 0.02 | 0.8 | 0.6 | 0.02 |
| Schizophrenia | 0.2 | 0.4 | -0.03 | 0.2 | 0.3 | -0.01 | 0.4 | 0.4 | 0.00 |
| Ulcerative colitis | 0.3 | 0.3 | 0.00 | 0.3 | 0.3 | 0.00 | 0.3 | 0.3 | 0.01 |
| Urinary tract infectious disease | 1.1 | 2.1 | -0.08 | 2.1 | 1.8 | 0.01 | 2.1 | 1.8 | 0.01 |
| Viral hepatitis C | 0.4 | 0.8 | -0.05 | 0.4 | 0.5 | 0.00 | 0.4 | 0.7 | -0.04 |
| Visual system disorder | 10.1 | 13.5 | -0.11 | 10.2 | 9.9 | 0.01 | 12.8 | 12.7 | 0.00 |
| **Medical history: Cardiovascular disease** | | | | | | | | | |
| Atrial fibrillation | 4.6 | 5.1 | -0.03 | 4.4 | 4.3 | 0.01 | 5.7 | 5.1 | 0.03 |
| Cerebrovascular disease | 2.6 | 2.8 | -0.01 | 2.5 | 2.4 | 0.00 | 2.8 | 2.7 | 0.00 |
| Coronary arteriosclerosis | 15.7 | 13.4 | 0.06 | 14.2 | 14.2 | 0.00 | 13.4 | 13.5 | 0.00 |
| Heart disease | 24.7 | 23.0 | 0.04 | 23.0 | 23.0 | 0.00 | 24.2 | 23.2 | 0.02 |
| Heart failure | 4.0 | 3.8 | 0.01 | 3.5 | 3.6 | 0.00 | 4.3 | 3.8 | 0.02 |
| Ischemic heart disease | 7.0 | 5.8 | 0.05 | 6.2 | 6.3 | 0.00 | 5.8 | 6.1 | -0.01 |
| Peripheral vascular disease | 5.8 | 4.7 | 0.05 | 5.2 | 5.1 | 0.00 | 5.1 | 5.0 | 0.00 |
| Pulmonary embolism | 0.4 | 0.6 | -0.03 | 0.4 | 0.5 | -0.01 | 0.5 | 0.6 | -0.02 |
| Venous thrombosis | 1.0 | 1.2 | -0.02 | 1.0 | 0.9 | 0.01 | 1.2 | 1.1 | 0.00 |
| **Medical history: Neoplasms** | | | | | | | | | |
| Hematologic neoplasm | 0.6 | 0.8 | -0.02 | 0.6 | 0.6 | 0.00 | 0.8 | 0.8 | 0.00 |
| Malignant lymphoma | 0.3 | 0.5 | -0.03 | 0.3 | 0.4 | -0.01 | 0.5 | 0.5 | 0.00 |

| (Continued) | Before ajustment | | | After matching | | | After stratification | | |
|---|---|---|---|---|---|---|---|---|---|
| Characteristic | T (%) | C (%) | StdDiff | T (%) | C (%) | StdDiff | T (%) | C (%) | StdDiff |
| Malignant neoplasm of anorectum | 0.1 | 0.1 | -0.01 | 0.2 | 0.1 | 0.02 | 0.2 | 0.1 | 0.02 |
| Malignant neoplastic disease | 4.7 | 6.7 | -0.09 | 4.8 | 4.7 | 0.00 | 6.5 | 6.2 | 0.01 |
| Malignant tumor of colon | 0.3 | 0.4 | -0.02 | 0.3 | 0.2 | 0.01 | 0.4 | 0.4 | 0.01 |
| Malignant tumor of lung | 0.1 | 0.2 | -0.03 | 0.1 | 0.1 | -0.01 | 0.3 | 0.2 | 0.01 |
| Malignant tumor of urinary bladder | 0.3 | 0.5 | -0.03 | 0.3 | 0.3 | 0.00 | 0.6 | 0.4 | 0.02 |
| Primary malignant neoplasm of prostate | 1.6 | 2.4 | -0.06 | 1.8 | 2.1 | -0.03 | 1.8 | 2.1 | -0.03 |
| **Medication use** | | | | | | | | | |
| Agents acting on the renin-angiotensin system | 69.5 | 66.7 | 0.06 | 68.6 | 68.6 | 0.00 | 68.6 | 68.6 | 0.00 |
| Antibacterials for systemic use | 36.4 | 36.2 | 0.00 | 35.8 | 35.9 | 0.00 | 37.6 | 36.2 | 0.03 |
| Antidepressants | 21.4 | 20.9 | 0.01 | 21.3 | 21.2 | 0.00 | 22.0 | 21.3 | 0.02 |
| Antiepileptics | 13.2 | 13.7 | -0.02 | 13.3 | 13.2 | 0.00 | 15.4 | 14.4 | 0.03 |
| Antiinflammatory and antirheumatic products | 54.6 | 55.1 | -0.01 | 53.9 | 53.8 | 0.00 | 55.1 | 55.0 | 0.00 |
| Antineoplastic agents | 3.0 | 3.1 | 0.00 | 3.0 | 2.9 | 0.00 | 3.7 | 3.1 | 0.03 |
| Antipsoriatics | 0.3 | 0.4 | -0.01 | 0.3 | 0.3 | 0.00 | 0.4 | 0.4 | 0.00 |
| Antithrombotic agents | 41.3 | 44.1 | -0.06 | 40.6 | 40.3 | 0.01 | 40.6 | 40.3 | 0.01 |
| Beta blocking agents | 32.6 | 33.4 | -0.02 | 31.5 | 31.6 | 0.00 | 33.7 | 32.9 | 0.02 |
| Calcium channel blockers | 23.2 | 21.4 | 0.04 | 22.5 | 22.6 | 0.00 | 21.5 | 22.2 | -0.02 |
| Diuretics | 34.3 | 35.1 | -0.02 | 33.6 | 33.4 | 0.00 | 35.1 | 34.5 | 0.01 |
| Drugs for acid related disorders | 29.7 | 30.0 | -0.01 | 30.9 | 30.3 | 0.01 | 30.9 | 30.3 | 0.01 |
| Drugs for obstructive airway diseases | 35.0 | 34.4 | 0.01 | 34.3 | 34.1 | 0.00 | 35.7 | 35.0 | 0.01 |
| Immunosuppressants | 1.8 | 1.6 | 0.01 | 1.7 | 1.7 | 0.00 | 2.1 | 1.7 | 0.03 |
| Lipid modifying agents | 75.2 | 71.0 | 0.10 | 73.9 | 74.0 | 0.00 | 71.5 | 71.8 | -0.01 |
| Opioids | 21.4 | 24.6 | -0.08 | 25.9 | 24.2 | 0.04 | 25.9 | 24.2 | 0.04 |
| Psycholeptics | 21.6 | 22.4 | -0.02 | 21.1 | 21.4 | -0.01 | 23.6 | 22.3 | 0.03 |
| Psychostimulants | 2.8 | 1.9 | 0.06 | 2.7 | 2.7 | 0.00 | 2.4 | 2.2 | 0.02 |

### 2.3.8 VA

##### 2.3.9 SIDIAP

**Supplementary Table 179:** Baseline patient characteristics for SGLT2I (male) (T) and SU (male) (C) new-users in the SIDIAP data source. We report proportion of initiators satisfying selected base-line characteristics and the standardized difference of population proportions (StdDiff) before and after propensity score adjustment. Less extreme StdDiffs through matching and stratification suggest improved balance between patient cohorts through adjustment.

| | Before adjustment | | | After matching | | | After stratification | | |
|---|---|---|---|---|---|---|---|---|---|
| Characteristic | T (%) | C (%) | StdDiff | T (%) | C (%) | StdDiff | T (%) | C (%) | StdDiff |
| **Age group** | | | | | | | | | |
| 25 - 29 | 0.1 | 0.1 | 0.01 | <0.4 | <0.1 | 0.06 | 0.1 | <0.1 | 0.03 |
| 30 - 34 | 0.4 | 0.3 | 0.01 | 0.7 | 0.3 | 0.05 | 0.4 | 0.3 | 0.02 |
| 35 - 39 | 1.1 | 1.2 | -0.01 | 1.6 | 1.3 | 0.03 | 1.5 | 1.4 | 0.01 |
| 40 - 44 | 3.1 | 3.7 | -0.03 | 4.7 | 4.9 | -0.01 | 4.5 | 4.5 | 0.00 |
| 45 - 49 | 6.3 | 7.3 | -0.04 | 9.3 | 7.8 | 0.05 | 7.7 | 9.1 | -0.05 |
| 50 - 54 | 11.1 | 11.1 | 0.00 | 12.6 | 11.9 | 0.02 | 11.9 | 11.1 | 0.03 |
| 55 - 59 | 15.2 | 15.1 | 0.00 | 16.5 | 16.1 | 0.01 | 15.1 | 15.5 | -0.01 |
| 60 - 64 | 17.5 | 17.4 | 0.00 | 16.0 | 15.8 | 0.01 | 16.7 | 17.2 | -0.01 |
| 65 - 69 | 17.2 | 15.5 | 0.04 | 18.0 | 16.7 | 0.03 | 16.4 | 17.5 | -0.03 |
| 70 - 74 | 12.6 | 12.5 | 0.00 | 10.1 | 11.4 | -0.04 | 12.6 | 10.0 | 0.08 |
| 75 - 79 | 9.0 | 8.6 | 0.01 | 5.9 | 7.4 | -0.06 | 7.8 | 7.1 | 0.03 |
| 80 - 84 | 4.0 | 4.7 | -0.04 | 2.7 | 3.8 | -0.06 | 3.4 | 4.1 | -0.04 |
| 85 - 89 | 1.8 | 2.0 | -0.01 | 1.3 | 2.1 | -0.06 | 1.4 | 1.8 | -0.03 |
| 90 - 94 | 0.3 | 0.5 | -0.02 | <0.4 | 0.4 | -0.02 | 0.3 | 0.4 | -0.01 |
| 95 - 99 | <0.1 | <0.1 | 0.00 | <0.4 | 0.1 | 0.00 | <0.1 | <0.1 | -0.01 |
| **Medical history: General** | | | | | | | | | |
| Acute respiratory disease | 10.6 | 9.3 | 0.05 | 12.4 | 11.9 | 0.01 | 10.8 | 11.2 | -0.01 |
| Attention deficit hyperactivity disorder | <0.1 | <0.1 | 0.02 | <0.4 | <0.1 | 0.06 | <0.1 | <0.1 | 0.02 |
| Chronic liver disease | 0.3 | 0.5 | -0.03 | 0.5 | 0.3 | 0.03 | 0.3 | 0.2 | 0.01 |
| Chronic obstructive lung disease | 1.7 | 2.2 | -0.03 | 1.6 | 1.7 | 0.00 | 1.3 | 1.5 | -0.01 |
| Crohn’s disease | <0.1 | <0.1 | 0.01 | <0.1 | <0.1 | 0.00 | <0.1 | <0.1 | 0.00 |
| Dementia | 0.3 | 0.5 | -0.05 | <0.4 | 0.4 | -0.03 | 0.2 | 0.3 | -0.01 |
| Depressive disorder | 1.1 | 2.1 | -0.08 | 1.7 | 1.9 | -0.01 | 1.4 | 1.6 | -0.02 |
| Gastroesophageal reflux disease | 0.9 | 0.6 | 0.04 | 1.0 | 1.4 | -0.04 | 0.8 | 2.8 | -0.15 |
| Gastrointestinal hemorrhage | 1.1 | 1.5 | -0.04 | 1.8 | 2.0 | -0.01 | 1.5 | 1.4 | 0.01 |
| Hyperlipidemia | 2.8 | 5.0 | -0.11 | 3.8 | 3.9 | -0.01 | 3.3 | 4.1 | -0.04 |
| Hypertensive disorder | 3.8 | 6.7 | -0.13 | 4.2 | 4.9 | -0.03 | 3.8 | 6.0 | -0.10 |
| Lesion of liver | 0.3 | 0.6 | -0.04 | 0.5 | 0.6 | -0.01 | 0.3 | 0.4 | -0.02 |
| Obesity | 7.1 | 7.7 | -0.02 | 10.2 | 9.1 | 0.04 | 8.9 | 8.3 | 0.02 |
| Osteoarthritis | 2.8 | 3.8 | -0.05 | 3.1 | 3.3 | -0.01 | 3.4 | 2.6 | 0.04 |
| Pneumonia | 1.9 | 1.4 | 0.04 | 1.4 | 1.3 | 0.01 | 1.8 | 1.2 | 0.04 |
| Psoriasis | 0.4 | 0.6 | -0.04 | 0.5 | 0.6 | -0.02 | 0.4 | 0.4 | 0.00 |
| Renal impairment | 1.5 | 1.6 | -0.01 | 1.1 | 1.4 | -0.03 | 1.6 | 1.5 | 0.01 |
| Rheumatoid arthritis | <0.1 | 0.1 | -0.03 | <0.1 | <0.1 | -0.01 | <0.1 | <0.1 | -0.01 |
| Schizophrenia | <0.1 | 0.1 | -0.03 | <0.1 | <0.1 | 0.00 | <0.1 | <0.1 | 0.00 |
| Urinary tract infectious disease | 3.2 | 2.1 | 0.07 | 3.4 | 2.8 | 0.03 | 3.1 | 3.5 | -0.03 |
| Viral hepatitis C | <0.1 | 0.2 | -0.03 | <0.4 | 0.2 | 0.00 | <0.1 | <0.1 | -0.01 |
| Visual system disorder | 12.2 | 14.3 | -0.06 | 15.5 | 16.0 | -0.01 | 15.1 | 15.0 | 0.00 |
| **Medical history: Cardiovascular disease** | | | | | | | | | |
| Atrial fibrillation | 2.5 | 1.2 | 0.10 | 1.1 | 1.2 | -0.01 | 1.9 | 1.3 | 0.05 |
| Cerebrovascular disease | 0.9 | 1.2 | -0.03 | 1.0 | 1.1 | -0.01 | 1.1 | 0.7 | 0.04 |
| Heart disease | 16.2 | 8.7 | 0.23 | 7.0 | 8.3 | -0.05 | 12.8 | 9.4 | 0.11 |
| Heart failure | 3.4 | 1.3 | 0.14 | 0.6 | 1.1 | -0.05 | 2.4 | 0.9 | 0.12 |
| Ischemic heart disease | 8.0 | 3.5 | 0.19 | 2.7 | 3.0 | -0.02 | 6.2 | 4.1 | 0.10 |
| Peripheral vascular disease | 1.2 | 1.1 | 0.01 | 0.9 | 0.6 | 0.04 | 0.9 | 0.6 | 0.04 |
| Pulmonary embolism | 0.1 | 0.1 | -0.01 | <0.4 | 0.3 | -0.04 | 0.1 | 0.2 | -0.03 |
| Venous thrombosis | 0.8 | 0.8 | 0.00 | 1.6 | 1.0 | 0.05 | 1.3 | 0.9 | 0.04 |
| **Medical history: Neoplasms** | | | | | | | | | |
| Hematologic neoplasm | 0.1 | 0.1 | -0.01 | <0.4 | <0.1 | 0.07 | 0.2 | <0.1 | 0.05 |
| Malignant neoplasm of anorectum | <0.1 | 0.1 | -0.02 | <0.4 | <0.1 | 0.03 | <0.1 | <0.1 | 0.01 |
| Malignant neoplastic disease | 1.7 | 2.9 | -0.08 | 2.1 | 2.7 | -0.04 | 2.0 | 2.4 | -0.02 |
| Malignant tumor of colon | 0.1 | 0.4 | -0.06 | <0.4 | 0.2 | -0.03 | 0.1 | 0.2 | -0.03 |
| Malignant tumor of lung | <0.1 | <0.1 | -0.01 | <0.1 | <0.1 | -0.01 | <0.1 | <0.1 | -0.01 |
| Malignant tumor of urinary bladder | 0.1 | 0.4 | -0.06 | <0.4 | 0.4 | -0.04 | 0.5 | 0.5 | 0.00 |
| Primary malignant neoplasm of prostate | 0.2 | 0.5 | -0.04 | <0.4 | 0.4 | -0.01 | 0.2 | 0.3 | -0.03 |
| **Medication use** | | | | | | | | | |
| Agents acting on the renin-angiotensin system | 69.7 | 54.4 | 0.32 | 57.3 | 59.2 | -0.04 | 64.9 | 61.4 | 0.07 |
| Antibacterials for systemic use | 33.6 | 28.9 | 0.10 | 37.4 | 35.9 | 0.03 | 35.0 | 34.0 | 0.02 |

| (Continued) | Before ajustment | | | After matching | | | After stratification | | |
|---|---|---|---|---|---|---|---|---|---|
| Characteristic | T (%) | C (%) | StdDiff | T (%) | C (%) | StdDiff | T (%) | C (%) | StdDiff |
| Antidepressants | 12.8 | 11.6 | 0.04 | 13.7 | 14.0 | -0.01 | 13.4 | 13.9 | -0.02 |
| Antiepileptics | 8.4 | 6.1 | 0.09 | 10.6 | 8.9 | 0.06 | 9.8 | 9.9 | 0.00 |
| Antiinflammatory and antirheumatic products | 55.2 | 51.5 | 0.07 | 49.6 | 49.3 | 0.00 | 54.2 | 50.3 | 0.08 |
| Antineoplastic agents | 1.4 | 1.2 | 0.02 | 1.1 | 1.4 | -0.02 | 1.2 | 1.1 | 0.01 |
| Antipsoriatics | 2.2 | 1.4 | 0.06 | 2.0 | 1.9 | 0.01 | 2.5 | 1.5 | 0.07 |
| Antithrombotic agents | 49.6 | 34.5 | 0.31 | 28.9 | 32.3 | -0.07 | 41.5 | 34.7 | 0.14 |
| Beta blocking agents | 42.0 | 21.1 | 0.46 | 22.1 | 25.0 | -0.07 | 34.5 | 26.6 | 0.17 |
| Calcium channel blockers | 26.6 | 17.9 | 0.21 | 19.4 | 21.2 | -0.04 | 23.7 | 19.9 | 0.09 |
| Diuretics | 45.8 | 35.5 | 0.21 | 36.5 | 37.3 | -0.02 | 42.7 | 37.4 | 0.11 |
| Drugs for acid related disorders | 50.2 | 40.5 | 0.20 | 42.0 | 41.2 | 0.02 | 46.6 | 42.6 | 0.08 |
| Drugs for obstructive airway diseases | 27.7 | 21.2 | 0.15 | 26.5 | 25.6 | 0.02 | 27.3 | 26.0 | 0.03 |
| Immunosuppressants | 1.4 | 0.8 | 0.06 | 0.8 | 1.2 | -0.04 | 1.1 | 1.0 | 0.02 |
| Lipid modifying agents | 70.6 | 57.7 | 0.27 | 59.6 | 61.1 | -0.03 | 65.8 | 63.1 | 0.06 |
| Opioids | 14.4 | 10.9 | 0.11 | 15.6 | 15.9 | -0.01 | 14.7 | 14.6 | 0.00 |
| Psycholeptics | 22.3 | 21.2 | 0.03 | 22.4 | 22.8 | -0.01 | 21.8 | 23.5 | -0.04 |
| Psychostimulants | 9.5 | 3.8 | 0.23 | 3.2 | 4.0 | -0.04 | 6.9 | 5.1 | 0.07 |

#### 2.4 GLP1RA (male) vs DPP4I (male)

##### 2.4.1 CCAE

**Supplementary Table 180:** Baseline patient characteristics for GLP1RA (male) (T) and DPP4I (male) (C) new-users in the CCAE data source. We report proportion of initiators satisfying selected base-line characteristics and the standardized difference of population proportions (StdDiff) before and after propensity score adjustment. Less extreme StdDiffs through matching and stratification suggest improved balance between patient cohorts through adjustment.

| | Before adjustment | | | After matching | | | After stratification | | |
|---|---|---|---|---|---|---|---|---|---|
| Characteristic | T (%) | C (%) | StdDiff | T (%) | C (%) | StdDiff | T (%) | C (%) | StdDiff |
| **Age group** | | | | | | | | | |
| 20 - 24 | 0.4 | 0.2 | -0.03 | 0.3 | 0.2 | -0.02 | 0.3 | 0.2 | -0.03 |
| 25 - 29 | 0.8 | 0.5 | -0.05 | 0.8 | 0.5 | -0.04 | 0.6 | 0.5 | -0.02 |
| 30 - 34 | 2.6 | 1.8 | -0.05 | 2.4 | 2.3 | 0.00 | 2.0 | 1.9 | -0.01 |
| 35 - 39 | 5.7 | 4.4 | -0.06 | 5.6 | 5.4 | -0.01 | 5.0 | 4.6 | -0.02 |
| 40 - 44 | 10.8 | 9.0 | -0.06 | 10.3 | 10.4 | 0.00 | 9.6 | 9.3 | -0.01 |
| 45 - 49 | 16.1 | 14.5 | -0.04 | 15.9 | 15.8 | 0.00 | 14.9 | 14.8 | 0.00 |
| 50 - 54 | 20.8 | 20.2 | -0.01 | 20.7 | 20.5 | 0.00 | 20.5 | 20.3 | 0.00 |
| 55 - 59 | 21.8 | 23.5 | 0.04 | 21.9 | 22.8 | 0.02 | 22.6 | 23.4 | 0.02 |
| 60 - 64 | 19.3 | 23.7 | 0.11 | 20.2 | 20.1 | 0.00 | 22.1 | 22.9 | 0.02 |
| 65 - 69 | 1.7 | 2.2 | 0.03 | 1.7 | 1.8 | 0.00 | 2.2 | 2.1 | -0.01 |
| **Medical history: General** | | | | | | | | | |
| Acute respiratory disease | 24.7 | 22.4 | -0.05 | 24.2 | 24.2 | 0.00 | 22.5 | 22.7 | 0.00 |
| Attention deficit hyperactivity disorder | 1.6 | 0.8 | -0.07 | 1.4 | 1.3 | 0.00 | 0.9 | 0.9 | -0.01 |
| Chronic liver disease | 1.5 | 2.7 | 0.09 | 1.6 | 1.8 | 0.01 | 2.7 | 2.5 | -0.01 |
| Chronic obstructive lung disease | 2.6 | 2.6 | 0.00 | 2.5 | 2.6 | 0.01 | 2.6 | 2.6 | 0.00 |
| Crohn's disease | 0.2 | 0.2 | 0.00 | 0.2 | 0.3 | 0.01 | 0.2 | 0.2 | -0.01 |
| Depressive disorder | 9.0 | 5.9 | -0.12 | 8.2 | 9.1 | 0.03 | 7.0 | 6.5 | -0.02 |
| Gastroesophageal reflux disease | 11.2 | 8.8 | -0.08 | 10.8 | 10.9 | 0.00 | 9.7 | 9.3 | -0.01 |
| Gastrointestinal hemorrhage | 1.9 | 1.8 | -0.01 | 1.8 | 1.7 | -0.01 | 1.9 | 1.8 | -0.01 |
| Human immunodeficiency virus infection | 0.4 | 0.4 | 0.00 | 0.3 | 0.5 | 0.02 | 0.2 | 0.4 | 0.04 |
| Hyperlipidemia | 71.3 | 70.1 | -0.03 | 71.8 | 72.0 | 0.00 | 70.6 | 70.5 | 0.00 |
| Hypertensive disorder | 70.8 | 66.3 | -0.10 | 70.6 | 71.3 | 0.01 | 68.1 | 67.3 | -0.02 |
| Lesion of liver | 0.6 | 0.6 | -0.01 | 0.6 | 0.6 | 0.01 | 0.4 | 0.6 | 0.02 |
| Obesity | 33.0 | 14.8 | -0.44 | 28.3 | 29.4 | 0.03 | 19.4 | 18.2 | -0.03 |
| Osteoarthritis | 18.5 | 15.1 | -0.09 | 17.7 | 18.3 | 0.02 | 16.0 | 15.8 | 0.00 |
| Pneumonia | 2.1 | 1.8 | -0.02 | 1.9 | 2.0 | 0.01 | 1.9 | 2.0 | 0.01 |
| Psoriasis | 2.0 | 1.5 | -0.04 | 1.8 | 1.8 | 0.00 | 1.5 | 1.5 | 0.00 |
| Renal impairment | 4.1 | 3.4 | -0.04 | 4.0 | 4.3 | 0.01 | 3.4 | 3.6 | 0.01 |
| Rheumatoid arthritis | 0.5 | 0.6 | 0.00 | 0.5 | 0.7 | 0.03 | 0.5 | 0.6 | 0.01 |
| Schizophrenia | 0.1 | 0.1 | 0.01 | 0.1 | 0.1 | 0.02 | 0.1 | 0.1 | 0.02 |
| Ulcerative colitis | 0.2 | 0.3 | 0.02 | 0.2 | 0.4 | 0.03 | 0.3 | 0.3 | 0.01 |
| Urinary tract infectious disease | 2.8 | 3.0 | 0.01 | 2.7 | 2.9 | 0.01 | 3.0 | 2.9 | 0.00 |
| Viral hepatitis C | 0.2 | 0.4 | 0.03 | 0.3 | 0.4 | 0.02 | 0.3 | 0.4 | 0.01 |
| Visual system disorder | 22.0 | 21.8 | -0.01 | 21.9 | 22.0 | 0.00 | 20.5 | 21.8 | 0.03 |
| **Medical history: Cardiovascular disease** | | | | | | | | | |
| Atrial fibrillation | 2.8 | 2.4 | -0.03 | 2.7 | 2.9 | 0.02 | 2.5 | 2.5 | 0.00 |
| Cerebrovascular disease | 2.2 | 2.5 | 0.02 | 2.2 | 2.1 | 0.00 | 2.2 | 2.5 | 0.01 |
| Coronary arteriosclerosis | 10.2 | 10.1 | 0.00 | 10.1 | 10.0 | 0.00 | 10.4 | 10.1 | -0.01 |
| Heart disease | 20.4 | 18.9 | -0.04 | 19.4 | 19.0 | -0.01 | 19.4 | 19.0 | -0.01 |
| Heart failure | 2.5 | 2.0 | -0.03 | 2.4 | 2.4 | 0.00 | 2.0 | 2.1 | 0.01 |
| Ischemic heart disease | 5.0 | 4.9 | -0.01 | 4.8 | 5.2 | 0.02 | 5.2 | 4.9 | -0.01 |
| Peripheral vascular disease | 4.1 | 3.3 | -0.04 | 4.0 | 3.8 | -0.01 | 3.6 | 3.5 | -0.01 |
| Pulmonary embolism | 0.7 | 0.4 | -0.04 | 0.6 | 0.6 | -0.01 | 0.5 | 0.4 | -0.01 |
| Venous thrombosis | 1.2 | 1.1 | -0.01 | 1.1 | 1.2 | 0.01 | 1.0 | 1.1 | 0.01 |
| **Medical history: Neoplasms** | | | | | | | | | |
| Hematologic neoplasm | 0.7 | 0.8 | 0.01 | 0.7 | 0.9 | 0.01 | 0.8 | 0.8 | 0.00 |
| Malignant lymphoma | 0.3 | 0.4 | 0.00 | 0.4 | 0.4 | 0.00 | 0.3 | 0.4 | 0.01 |
| Malignant neoplasm of anorectum | 0.1 | 0.1 | 0.01 | 0.1 | 0.1 | 0.00 | 0.2 | 0.1 | -0.01 |
| Malignant neoplastic disease | 4.6 | 5.3 | 0.03 | 4.7 | 5.2 | 0.02 | 5.5 | 5.2 | -0.02 |
| Malignant tumor of colon | 0.2 | 0.3 | 0.02 | 0.2 | 0.3 | 0.01 | 0.3 | 0.3 | 0.00 |
| Malignant tumor of lung | 0.1 | 0.1 | 0.02 | 0.1 | 0.1 | 0.02 | 0.1 | 0.1 | 0.02 |
| Malignant tumor of urinary bladder | 0.2 | 0.3 | 0.02 | 0.2 | 0.2 | 0.00 | 0.4 | 0.3 | -0.02 |
| Primary malignant neoplasm of prostate | 1.0 | 1.4 | 0.04 | 1.1 | 1.2 | 0.01 | 1.4 | 1.3 | -0.01 |
| **Medication use** | | | | | | | | | |

| (Continued) | Before ajustment | | | After matching | | | After stratification | | |
|---|---|---|---|---|---|---|---|---|---|
| Characteristic | T (%) | C (%) | StdDiff | T (%) | C (%) | StdDiff | T (%) | C (%) | StdDiff |
| Agents acting on the renin-angiotensin system | 71.3 | 70.0 | -0.03 | 71.4 | 71.6 | 0.00 | 71.6 | 70.3 | -0.03 |
| Antibacterials for systemic use | 53.6 | 51.7 | -0.04 | 53.1 | 52.6 | -0.01 | 51.5 | 51.9 | 0.01 |
| Antidepressants | 24.9 | 17.3 | -0.19 | 23.1 | 23.7 | 0.01 | 19.1 | 18.7 | -0.01 |
| Antiepileptics | 11.8 | 8.5 | -0.11 | 11.0 | 11.6 | 0.02 | 9.3 | 9.2 | 0.00 |
| Antiinflammatory and antirheumatic products | 31.7 | 27.4 | -0.09 | 31.0 | 31.3 | 0.00 | 28.9 | 28.2 | -0.01 |
| Antineoplastic agents | 3.9 | 3.4 | -0.03 | 3.6 | 3.8 | 0.01 | 3.3 | 3.4 | 0.01 |
| Antipsoriatics | 0.8 | 0.5 | -0.03 | 0.7 | 0.5 | -0.03 | 0.6 | 0.5 | -0.01 |
| Antithrombotic agents | 11.5 | 10.5 | -0.03 | 11.2 | 11.2 | 0.00 | 10.8 | 10.6 | -0.01 |
| Beta blocking agents | 26.0 | 25.3 | -0.02 | 25.7 | 26.4 | 0.02 | 25.5 | 25.4 | 0.00 |
| Calcium channel blockers | 23.5 | 21.3 | -0.05 | 23.0 | 23.7 | 0.02 | 21.4 | 21.7 | 0.01 |
| Diuretics | 36.7 | 31.9 | -0.10 | 35.2 | 35.6 | 0.01 | 33.5 | 32.7 | -0.02 |
| Drugs for acid related disorders | 22.7 | 19.9 | -0.07 | 21.9 | 21.5 | -0.01 | 20.4 | 20.3 | 0.00 |
| Drugs for obstructive airway diseases | 35.3 | 30.0 | -0.11 | 34.0 | 34.2 | 0.00 | 31.8 | 30.9 | -0.02 |
| Immunosuppressants | 2.0 | 1.7 | -0.03 | 1.9 | 2.1 | 0.01 | 1.6 | 1.8 | 0.01 |
| Lipid modifying agents | 71.1 | 71.7 | 0.01 | 71.4 | 71.3 | 0.00 | 71.6 | 71.6 | 0.00 |
| Opioids | 22.9 | 21.9 | -0.03 | 22.8 | 22.9 | 0.00 | 21.5 | 22.0 | 0.01 |
| Psycholeptics | 19.9 | 17.7 | -0.06 | 17.8 | 18.0 | 0.01 | 17.8 | 18.0 | 0.01 |
| Psychostimulants | 4.0 | 2.2 | -0.10 | 3.5 | 3.4 | 0.00 | 2.6 | 2.4 | -0.01 |

##### 2.4.2 ODOD

**Supplementary Table 181:** Baseline patient characteristics for GLP1RA (male) (T) and DPP4I (male) (C) new-users in the ODOD data source. We report proportion of initiators satisfying selected base-line characteristics and the standardized difference of population proportions (StdDiff) before and after propensity score adjustment. Less extreme StdDiffs through matching and stratification suggest improved balance between patient cohorts through adjustment.

| | Before adjustment | | | After matching | | | After stratification | | |
|---|---|---|---|---|---|---|---|---|---|
| Characteristic | T (%) | C (%) | StdDiff | T (%) | C (%) | StdDiff | T (%) | C (%) | StdDiff |
| **Age group** | | | | | | | | | |
| 20 - 24 | 0.3 | 0.1 | -0.05 | 0.3 | 0.1 | -0.03 | 0.2 | 0.1 | -0.02 |
| 25 - 29 | 0.4 | 0.3 | -0.01 | 0.4 | 0.4 | 0.01 | 0.3 | 0.4 | 0.01 |
| 30 - 34 | 2.1 | 1.2 | -0.07 | 1.8 | 1.5 | -0.02 | 1.6 | 1.3 | -0.02 |
| 35 - 39 | 4.2 | 3.2 | -0.05 | 3.9 | 3.6 | -0.01 | 3.0 | 3.3 | 0.02 |
| 40 - 44 | 7.9 | 6.0 | -0.07 | 7.2 | 7.1 | 0.00 | 6.1 | 6.3 | 0.01 |
| 45 - 49 | 12.1 | 8.8 | -0.11 | 11.2 | 10.5 | -0.02 | 9.6 | 9.5 | 0.00 |
| 50 - 54 | 14.4 | 11.9 | -0.07 | 13.4 | 13.6 | 0.01 | 12.5 | 12.4 | 0.00 |
| 55 - 59 | 16.4 | 14.0 | -0.07 | 16.1 | 16.0 | 0.00 | 15.0 | 14.5 | -0.01 |
| 60 - 64 | 14.3 | 13.2 | -0.03 | 14.2 | 14.6 | 0.01 | 13.2 | 13.5 | 0.01 |
| 65 - 69 | 12.8 | 13.9 | 0.03 | 13.8 | 13.6 | -0.01 | 13.8 | 13.9 | 0.00 |
| 70 - 74 | 9.1 | 12.5 | 0.11 | 10.5 | 10.5 | 0.00 | 12.0 | 11.9 | 0.00 |
| 75 - 79 | 4.3 | 8.1 | 0.16 | 5.2 | 5.7 | 0.02 | 7.4 | 7.3 | 0.00 |
| 80 - 84 | 1.4 | 4.5 | 0.18 | 1.7 | 1.6 | 0.00 | 4.3 | 3.8 | -0.03 |
| 85 - 89 | 0.3 | 2.0 | 0.16 | 0.4 | 0.6 | 0.03 | 1.1 | 1.7 | 0.05 |
| **Medical history: General** | | | | | | | | | |
| Acute respiratory disease | 22.6 | 23.2 | 0.01 | 22.5 | 22.6 | 0.00 | 22.6 | 22.9 | 0.01 |
| Attention deficit hyperactivity disorder | 1.8 | 0.8 | -0.08 | 1.3 | 1.3 | 0.00 | 1.0 | 1.0 | 0.00 |
| Chronic liver disease | 1.7 | 3.3 | 0.10 | 2.0 | 1.8 | -0.01 | 3.1 | 3.0 | -0.01 |
| Chronic obstructive lung disease | 7.3 | 8.2 | 0.03 | 7.4 | 7.9 | 0.02 | 8.6 | 8.1 | -0.02 |
| Crohn’s disease | 0.2 | 0.3 | 0.01 | 0.2 | 0.3 | 0.01 | 0.3 | 0.3 | 0.00 |
| Dementia | 0.6 | 1.8 | 0.10 | 0.7 | 1.0 | 0.04 | 1.1 | 1.6 | 0.04 |
| Depressive disorder | 12.7 | 9.1 | -0.12 | 11.3 | 11.9 | 0.02 | 9.6 | 10.0 | 0.01 |
| Gastroesophageal reflux disease | 16.4 | 14.4 | -0.06 | 15.6 | 15.7 | 0.00 | 15.9 | 14.8 | -0.03 |
| Gastrointestinal hemorrhage | 2.1 | 2.6 | 0.03 | 2.2 | 2.3 | 0.00 | 3.2 | 2.5 | -0.04 |
| Human immunodeficiency virus infection | 0.6 | 0.5 | -0.01 | 0.6 | 0.7 | 0.01 | 0.6 | 0.6 | -0.01 |
| Hyperlipidemia | 79.8 | 82.3 | 0.06 | 80.0 | 80.4 | 0.01 | 82.3 | 81.7 | -0.01 |
| Hypertensive disorder | 79.1 | 78.6 | -0.01 | 78.5 | 78.3 | 0.00 | 80.6 | 78.7 | -0.05 |
| Lesion of liver | 1.0 | 1.1 | 0.01 | 1.0 | 1.2 | 0.02 | 1.0 | 1.1 | 0.01 |
| Obesity | 41.6 | 19.5 | -0.50 | 33.1 | 35.0 | 0.04 | 24.0 | 24.3 | 0.01 |
| Osteoarthritis | 25.3 | 22.1 | -0.07 | 24.1 | 24.4 | 0.01 | 24.2 | 22.8 | -0.03 |
| Pneumonia | 3.2 | 3.6 | 0.02 | 3.0 | 2.8 | -0.01 | 3.2 | 3.5 | 0.01 |
| Psoriasis | 2.2 | 1.6 | -0.04 | 2.1 | 1.9 | -0.01 | 1.7 | 1.6 | -0.01 |
| Renal impairment | 9.9 | 13.5 | 0.11 | 10.6 | 10.9 | 0.01 | 13.5 | 12.8 | -0.02 |
| Rheumatoid arthritis | 0.9 | 1.0 | 0.01 | 0.8 | 0.9 | 0.01 | 1.0 | 1.0 | -0.01 |
| Schizophrenia | 0.4 | 0.4 | -0.01 | 0.4 | 0.5 | 0.02 | 0.5 | 0.4 | -0.01 |
| Ulcerative colitis | 0.3 | 0.4 | 0.02 | 0.3 | 0.3 | 0.01 | 0.4 | 0.3 | -0.01 |
| Urinary tract infectious disease | 4.3 | 5.9 | 0.07 | 4.5 | 4.5 | 0.00 | 6.8 | 5.6 | -0.05 |
| Viral hepatitis C | 0.6 | 0.7 | 0.01 | 0.7 | 0.7 | 0.00 | 0.8 | 0.6 | -0.03 |
| Visual system disorder | 31.6 | 35.8 | 0.09 | 32.8 | 32.7 | 0.00 | 36.7 | 35.1 | -0.03 |
| **Medical history: Cardiovascular disease** | | | | | | | | | |
| Atrial fibrillation | 6.1 | 7.3 | 0.05 | 6.1 | 6.3 | 0.01 | 6.9 | 7.0 | 0.00 |
| Cerebrovascular disease | 4.0 | 6.5 | 0.11 | 4.2 | 4.1 | 0.00 | 5.6 | 6.0 | 0.02 |
| Coronary arteriosclerosis | 15.5 | 18.9 | 0.09 | 16.1 | 16.0 | 0.00 | 18.0 | 18.2 | 0.00 |
| Heart disease | 28.3 | 32.1 | 0.08 | 28.6 | 28.5 | 0.00 | 32.4 | 31.3 | -0.02 |
| Heart failure | 5.9 | 6.7 | 0.04 | 5.8 | 5.9 | 0.00 | 6.9 | 6.5 | -0.01 |
| Ischemic heart disease | 7.8 | 9.8 | 0.07 | 8.0 | 8.0 | 0.00 | 9.8 | 9.4 | -0.01 |
| Peripheral vascular disease | 9.9 | 10.4 | 0.01 | 9.9 | 10.2 | 0.01 | 9.4 | 10.4 | 0.03 |
| Pulmonary embolism | 0.8 | 0.7 | -0.02 | 0.9 | 0.7 | -0.03 | 0.9 | 0.7 | -0.03 |
| Venous thrombosis | 1.4 | 1.6 | 0.01 | 1.4 | 1.2 | -0.02 | 1.5 | 1.5 | 0.00 |
| **Medical history: Neoplasms** | | | | | | | | | |
| Hematologic neoplasm | 0.8 | 1.2 | 0.04 | 0.8 | 1.1 | 0.04 | 1.0 | 1.1 | 0.02 |
| Malignant lymphoma | 0.4 | 0.6 | 0.03 | 0.4 | 0.6 | 0.03 | 0.3 | 0.6 | 0.04 |
| Malignant neoplasm of anorectum | 0.2 | 0.2 | 0.01 | 0.2 | 0.2 | 0.01 | 0.2 | 0.2 | 0.01 |
| Malignant neoplastic disease | 7.9 | 10.9 | 0.10 | 8.6 | 8.7 | 0.00 | 9.8 | 10.4 | 0.02 |
| Malignant tumor of colon | 0.5 | 0.6 | 0.01 | 0.5 | 0.3 | -0.03 | 0.4 | 0.5 | 0.01 |
| Malignant tumor of lung | 0.2 | 0.4 | 0.03 | 0.3 | 0.3 | 0.01 | 0.6 | 0.3 | -0.04 |
| Malignant tumor of urinary bladder | 0.4 | 0.7 | 0.05 | 0.4 | 0.5 | 0.01 | 0.8 | 0.7 | -0.01 |

| (Continued) | Before ajustment | | | After matching | | | After stratification | | |
|---|---|---|---|---|---|---|---|---|---|
| Characteristic | T (%) | C (%) | StdDiff | T (%) | C (%) | StdDiff | T (%) | C (%) | StdDiff |
| Primary malignant neoplasm of prostate | 2.6 | 4.2 | 0.09 | 3.0 | 2.9 | 0.00 | 4.4 | 3.8 | -0.03 |
| **Medication use** | | | | | | | | | |
| Agents acting on the renin-angiotensin system | 72.9 | 71.4 | -0.03 | 72.5 | 72.3 | 0.00 | 72.5 | 71.7 | -0.02 |
| Antibacterials for systemic use | 53.5 | 52.8 | -0.01 | 52.8 | 52.6 | 0.00 | 53.9 | 52.9 | -0.02 |
| Antidepressants | 25.9 | 18.8 | -0.17 | 23.4 | 23.5 | 0.00 | 20.0 | 20.3 | 0.01 |
| Antiepileptics | 16.6 | 12.8 | -0.11 | 15.4 | 16.2 | 0.02 | 14.0 | 13.8 | -0.01 |
| Antiinflammatory and antirheumatic products | 30.9 | 26.7 | -0.09 | 29.4 | 29.8 | 0.01 | 27.6 | 27.4 | 0.00 |
| Antineoplastic agents | 4.7 | 4.8 | 0.00 | 4.7 | 4.9 | 0.01 | 4.7 | 4.8 | 0.00 |
| Antipsoriatics | 0.6 | 0.6 | -0.01 | 0.7 | 0.6 | -0.01 | 0.6 | 0.6 | 0.00 |
| Antithrombotic agents | 15.7 | 17.3 | 0.04 | 15.8 | 16.1 | 0.01 | 17.1 | 17.0 | 0.00 |
| Beta blocking agents | 31.3 | 33.6 | 0.05 | 31.4 | 31.7 | 0.01 | 34.8 | 33.3 | -0.03 |
| Calcium channel blockers | 25.6 | 25.7 | 0.00 | 25.6 | 25.9 | 0.01 | 25.0 | 25.7 | 0.02 |
| Diuretics | 40.5 | 36.3 | -0.09 | 38.6 | 39.5 | 0.02 | 40.2 | 37.1 | -0.06 |
| Drugs for acid related disorders | 22.2 | 22.6 | 0.01 | 21.8 | 22.2 | 0.01 | 23.5 | 22.5 | -0.02 |
| Drugs for obstructive airway diseases | 36.5 | 32.9 | -0.07 | 35.1 | 34.9 | 0.00 | 35.3 | 33.6 | -0.04 |
| Immunosuppressants | 2.0 | 1.8 | -0.02 | 1.9 | 1.8 | -0.01 | 1.7 | 1.8 | 0.01 |
| Lipid modifying agents | 74.7 | 76.3 | 0.04 | 75.4 | 75.6 | 0.00 | 77.0 | 76.1 | -0.02 |
| Opioids | 24.9 | 24.2 | -0.02 | 24.8 | 24.4 | -0.01 | 24.8 | 24.4 | -0.01 |
| Psycholeptics | 22.0 | 20.0 | -0.05 | 21.1 | 21.2 | 0.00 | 20.3 | 20.5 | 0.00 |
| Psychostimulants | 3.1 | 1.8 | -0.09 | 2.5 | 2.6 | 0.01 | 2.0 | 2.0 | 0.00 |

##### 2.4.3 MDCR

**Supplementary Table 182:** Baseline patient characteristics for GLP1RA (male) (T) and DPP4I (male) (C) new-users in the MDCR data source. We report proportion of initiators satisfying selected base-line characteristics and the standardized difference of population proportions (StdDiff) before and after propensity score adjustment. Less extreme StdDiffs through matching and stratification suggest improved balance between patient cohorts through adjustment.

| | Before adjustment | | | After matching | | | After stratification | | |
|---|---|---|---|---|---|---|---|---|---|
| Characteristic | T (%) | C (%) | StdDiff | T (%) | C (%) | StdDiff | T (%) | C (%) | StdDiff |
| **Age group** | | | | | | | | | |
| 45 - 49 | <0.5 | <0.1 | -0.04 | <0.5 | <0.6 | 0.04 | <0.2 | <0.1 | 0.01 |
| 50 - 54 | 0.6 | 0.2 | -0.07 | 0.3 | 0.2 | -0.03 | 0.3 | 0.2 | -0.03 |
| 55 - 59 | 1.5 | 0.5 | -0.09 | 1.4 | 0.7 | -0.07 | 1.0 | 0.6 | -0.05 |
| 60 - 64 | 2.8 | 1.9 | -0.06 | 3.0 | 2.5 | -0.03 | 2.3 | 1.9 | -0.02 |
| 65 - 69 | 40.4 | 31.6 | -0.18 | 39.7 | 36.9 | -0.06 | 35.7 | 32.1 | -0.08 |
| 70 - 74 | 35.1 | 29.9 | -0.11 | 34.8 | 33.8 | -0.02 | 27.9 | 30.4 | 0.06 |
| 75 - 79 | 12.9 | 19.2 | 0.17 | 13.3 | 15.3 | 0.06 | 13.3 | 15.3 | 0.06 |
| 80 - 84 | 4.9 | 10.5 | 0.21 | 5.3 | 6.2 | 0.04 | 10.0 | 10.1 | 0.00 |
| 85 - 89 | 1.3 | 4.6 | 0.20 | 1.5 | 3.0 | 0.10 | 4.0 | 4.5 | 0.02 |
| 90 - 94 | <0.5 | 1.3 | 0.10 | <0.5 | 1.4 | 0.10 | <0.2 | 1.3 | 0.14 |
| **Medical history: General** | | | | | | | | | |
| Acute respiratory disease | 21.6 | 21.1 | -0.01 | 21.2 | 22.0 | 0.02 | 21.2 | 22.0 | 0.02 |
| Attention deficit hyperactivity disorder | <0.5 | 0.2 | 0.00 | <0.5 | 0.6 | 0.06 | <0.2 | 0.2 | 0.03 |
| Chronic liver disease | 1.7 | 1.9 | 0.02 | 1.7 | 1.6 | -0.01 | 1.4 | 2.0 | 0.05 |
| Chronic obstructive lung disease | 13.0 | 10.8 | -0.07 | 12.6 | 13.3 | 0.02 | 14.4 | 10.9 | -0.11 |
| Crohn’s disease | <0.5 | 0.3 | 0.03 | <0.5 | <0.6 | 0.04 | 0.4 | 0.3 | -0.01 |
| Dementia | 1.4 | 2.4 | 0.07 | 1.5 | 2.8 | 0.09 | 0.8 | 2.4 | 0.13 |
| Depressive disorder | 8.8 | 6.0 | -0.11 | 7.7 | 9.7 | 0.07 | 6.8 | 6.2 | -0.02 |
| Gastroesophageal reflux disease | 11.9 | 11.0 | -0.03 | 11.9 | 14.0 | 0.06 | 8.5 | 11.3 | 0.09 |
| Gastrointestinal hemorrhage | 2.0 | 2.9 | 0.06 | 1.7 | 1.9 | 0.01 | 1.5 | 2.8 | 0.10 |
| Human immunodeficiency virus infection | <0.5 | 0.2 | 0.02 | <0.2 | 0.2 | 0.05 | <0.2 | 0.2 | 0.05 |
| Hyperlipidemia | 70.7 | 64.5 | -0.13 | 71.4 | 73.6 | 0.05 | 60.2 | 65.5 | 0.11 |
| Hypertensive disorder | 80.2 | 73.9 | -0.15 | 80.9 | 84.7 | 0.10 | 78.8 | 74.7 | -0.10 |
| Lesion of liver | 0.9 | 0.7 | -0.02 | 0.9 | 0.7 | -0.02 | 0.9 | 0.7 | -0.02 |
| Obesity | 27.7 | 9.7 | -0.48 | 23.6 | 27.5 | 0.09 | 10.4 | 11.2 | 0.03 |
| Osteoarthritis | 33.3 | 27.0 | -0.14 | 32.9 | 30.8 | -0.04 | 24.5 | 27.5 | 0.07 |
| Pneumonia | 4.1 | 5.1 | 0.05 | 4.3 | 4.6 | 0.01 | 2.6 | 5.1 | 0.13 |
| Psoriasis | 3.3 | 1.7 | -0.10 | 3.1 | 2.5 | -0.04 | 2.0 | 1.7 | -0.02 |
| Renal impairment | 11.1 | 14.5 | 0.10 | 11.3 | 13.6 | 0.07 | 10.2 | 14.4 | 0.13 |
| Rheumatoid arthritis | 1.2 | 1.3 | 0.01 | 1.0 | 1.6 | 0.06 | 0.8 | 1.3 | 0.05 |
| Schizophrenia | <0.5 | 0.1 | -0.02 | <0.5 | <0.6 | -0.01 | <0.2 | 0.1 | 0.01 |
| Ulcerative colitis | <0.5 | 0.5 | 0.05 | <0.5 | <0.6 | 0.02 | 0.3 | 0.5 | 0.03 |
| Urinary tract infectious disease | 4.9 | 6.3 | 0.06 | 4.8 | 4.2 | -0.03 | 3.6 | 6.1 | 0.12 |
| Viral hepatitis C | <0.5 | 0.3 | 0.00 | <0.5 | <0.6 | 0.02 | <0.2 | 0.3 | 0.04 |
| Visual system disorder | 49.9 | 49.5 | -0.01 | 53.7 | 49.5 | -0.08 | 53.7 | 49.5 | -0.08 |
| **Medical history: Cardiovascular disease** | | | | | | | | | |
| Atrial fibrillation | 11.4 | 13.2 | 0.05 | 11.0 | 11.7 | 0.02 | 12.4 | 13.1 | 0.02 |
| Cerebrovascular disease | 10.1 | 13.0 | 0.09 | 10.3 | 9.7 | -0.02 | 11.2 | 12.9 | 0.05 |
| Coronary arteriosclerosis | 30.3 | 32.2 | 0.04 | 29.9 | 30.0 | 0.00 | 31.5 | 32.0 | 0.01 |
| Heart disease | 47.7 | 50.5 | 0.06 | 47.6 | 47.7 | 0.00 | 49.3 | 50.4 | 0.02 |
| Heart failure | 8.1 | 9.4 | 0.04 | 7.5 | 9.0 | 0.05 | 8.0 | 9.4 | 0.05 |
| Ischemic heart disease | 11.4 | 13.6 | 0.07 | 11.2 | 13.6 | 0.07 | 11.2 | 13.5 | 0.07 |
| Peripheral vascular disease | 12.5 | 13.1 | 0.02 | 12.8 | 14.3 | 0.04 | 12.4 | 13.2 | 0.03 |
| Pulmonary embolism | 1.3 | 0.9 | -0.04 | 1.3 | 1.2 | -0.01 | 3.5 | 0.9 | -0.18 |
| Venous thrombosis | 1.7 | 2.2 | 0.04 | 1.7 | 1.9 | 0.01 | 1.0 | 2.2 | 0.09 |
| **Medical history: Neoplasms** | | | | | | | | | |
| Hematologic neoplasm | 2.0 | 2.1 | 0.01 | 1.9 | 2.9 | 0.07 | 0.8 | 2.1 | 0.11 |
| Malignant lymphoma | 1.2 | 1.1 | -0.01 | 0.9 | 1.3 | 0.03 | 0.4 | 1.1 | 0.08 |
| Malignant neoplastic disease | 21.4 | 22.0 | 0.01 | 22.0 | 22.3 | 0.01 | 20.8 | 21.9 | 0.03 |
| Malignant tumor of breast | <0.5 | 0.1 | -0.02 | <0.2 | 0.1 | 0.02 | <0.2 | 0.1 | 0.02 |
| Malignant tumor of colon | <0.5 | 1.0 | 0.07 | <0.5 | 0.7 | 0.05 | <0.2 | 1.0 | 0.12 |
| Malignant tumor of lung | 0.8 | 0.7 | -0.01 | 0.9 | 0.8 | -0.01 | 0.4 | 0.7 | 0.04 |
| Malignant tumor of urinary bladder | 1.9 | 2.0 | 0.00 | 1.9 | 1.6 | -0.02 | 1.3 | 1.9 | 0.05 |
| Primary malignant neoplasm of prostate | 8.4 | 8.4 | 0.00 | 8.5 | 8.6 | 0.01 | 11.1 | 8.4 | -0.09 |
| **Medication use** | | | | | | | | | |
| Agents acting on the renin-angiotensin system | 77.4 | 76.3 | -0.03 | 78.0 | 77.8 | 0.00 | 74.3 | 76.4 | 0.05 |
| Antibacterials for systemic use | 59.4 | 58.7 | -0.01 | 59.1 | 57.6 | -0.03 | 53.2 | 58.6 | 0.11 |

| (Continued) | Before ajustment | | | After matching | | | After stratification | | |
|---|---|---|---|---|---|---|---|---|---|
| Characteristic | T (%) | C (%) | StdDiff | T (%) | C (%) | StdDiff | T (%) | C (%) | StdDiff |
| Antidepressants | 27.0 | 18.8 | -0.20 | 24.9 | 26.1 | 0.03 | 19.4 | 19.3 | 0.00 |
| Antiepileptics | 17.1 | 12.8 | -0.12 | 16.7 | 16.5 | -0.01 | 13.5 | 13.1 | -0.01 |
| Antiinflammatory and antirheumatic products | 28.3 | 25.5 | -0.06 | 27.1 | 28.4 | 0.03 | 20.9 | 25.8 | 0.12 |
| Antineoplastic agents | 8.7 | 8.2 | -0.02 | 8.5 | 8.5 | 0.00 | 7.1 | 8.2 | 0.04 |
| Antipsoriatics | 0.9 | 0.8 | -0.01 | 0.6 | 0.8 | 0.02 | 0.5 | 0.8 | 0.03 |
| Antithrombotic agents | 27.8 | 28.4 | 0.01 | 28.4 | 25.7 | -0.06 | 29.9 | 28.4 | -0.03 |
| Beta blocking agents | 48.6 | 50.3 | 0.03 | 49.1 | 50.5 | 0.03 | 52.3 | 50.2 | -0.04 |
| Calcium channel blockers | 32.7 | 32.8 | 0.00 | 32.9 | 34.6 | 0.04 | 31.2 | 32.9 | 0.04 |
| Diuretics | 50.7 | 45.7 | -0.10 | 49.5 | 49.5 | 0.00 | 42.1 | 46.0 | 0.08 |
| Drugs for acid related disorders | 31.2 | 29.8 | -0.03 | 31.5 | 29.8 | -0.04 | 24.5 | 30.1 | 0.13 |
| Drugs for obstructive airway diseases | 45.1 | 38.5 | -0.14 | 44.1 | 44.4 | 0.00 | 35.7 | 39.0 | 0.07 |
| Immunosuppressants | 3.5 | 2.4 | -0.06 | 3.3 | 2.7 | -0.04 | 2.4 | 2.5 | 0.00 |
| Lipid modifying agents | 85.0 | 82.7 | -0.06 | 84.2 | 83.3 | -0.03 | 82.5 | 82.9 | 0.01 |
| Opioids | 27.3 | 25.4 | -0.04 | 27.3 | 25.7 | -0.04 | 25.2 | 25.5 | 0.01 |
| Psycholeptics | 23.0 | 22.4 | -0.01 | 22.4 | 22.2 | 0.00 | 22.2 | 22.4 | 0.01 |
| Psychostimulants | 1.5 | 1.1 | -0.04 | 1.6 | 1.1 | -0.05 | 0.9 | 1.1 | 0.01 |

##### 2.4.4 Open Claims

**Supplementary Table 183:** Baseline patient characteristics for GLP1RA (male) (T) and DPP4I (male) (C) new-users in the Open Claims data source. We report proportion of initiators satisfying selected base-line characteristics and the standardized difference of population proportions (StdDiff) before and after propensity score adjustment. Less extreme StdDiffs through matching and stratification suggest improved balance between patient cohorts through adjustment.

| | Before adjustment | | | After matching | | | After stratification | | |
|---|---|---|---|---|---|---|---|---|---|
| Characteristic | T (%) | C (%) | StdDiff | T (%) | C (%) | StdDiff | T (%) | C (%) | StdDiff |
| **Age group** | | | | | | | | | |
| 15 - 19 | 0.2 | 0.1 | -0.03 | 0.1 | 0.1 | -0.01 | 0.1 | 0.1 | -0.01 |
| 20 - 24 | 0.5 | 0.2 | -0.05 | 0.4 | 0.4 | -0.01 | 0.4 | 0.3 | -0.01 |
| 25 - 29 | 1.0 | 0.5 | -0.05 | 0.9 | 0.8 | -0.01 | 0.7 | 0.7 | -0.01 |
| 30 - 34 | 2.3 | 1.3 | -0.07 | 1.6 | 1.6 | 0.00 | 1.6 | 1.6 | 0.00 |
| 35 - 39 | 4.7 | 3.0 | -0.09 | 4.1 | 4.1 | 0.00 | 3.6 | 3.4 | -0.01 |
| 40 - 44 | 8.1 | 5.4 | -0.11 | 7.1 | 7.2 | 0.00 | 6.2 | 6.1 | 0.00 |
| 45 - 49 | 11.8 | 8.9 | -0.10 | 11.1 | 11.0 | 0.00 | 9.7 | 9.7 | 0.00 |
| 50 - 54 | 15.1 | 12.5 | -0.08 | 14.3 | 14.2 | 0.00 | 13.1 | 13.0 | 0.00 |
| 55 - 59 | 16.8 | 15.3 | -0.04 | 16.6 | 16.8 | 0.01 | 15.7 | 15.8 | 0.00 |
| 60 - 64 | 15.5 | 15.6 | 0.00 | 16.0 | 15.9 | 0.00 | 16.1 | 15.6 | -0.01 |
| 65 - 69 | 11.6 | 13.8 | 0.07 | 12.6 | 12.5 | 0.00 | 13.1 | 13.3 | 0.00 |
| 70 - 74 | 7.1 | 10.4 | 0.12 | 8.2 | 8.2 | 0.00 | 9.5 | 9.6 | 0.00 |
| 75 - 79 | 3.6 | 9.2 | 0.23 | 4.5 | 4.3 | -0.01 | 7.0 | 7.7 | 0.03 |
| 80 - 84 | 1.6 | 3.6 | 0.12 | 2.1 | 2.2 | 0.01 | 3.1 | 3.1 | 0.00 |
| 85 - 89 | 0.1 | 0.1 | -0.01 | 0.1 | 0.2 | 0.02 | 0.1 | 0.1 | 0.01 |
| **Medical history: General** | | | | | | | | | |
| Acute respiratory disease | 11.3 | 12.2 | 0.03 | 11.1 | 11.4 | 0.01 | 12.0 | 12.0 | 0.00 |
| Attention deficit hyperactivity disorder | 1.0 | 0.4 | -0.07 | 0.7 | 0.7 | 0.00 | 0.6 | 0.6 | 0.00 |
| Chronic liver disease | 0.8 | 1.6 | 0.07 | 0.9 | 0.8 | 0.00 | 1.4 | 1.3 | -0.01 |
| Chronic obstructive lung disease | 3.9 | 5.0 | 0.06 | 4.0 | 4.2 | 0.01 | 4.6 | 4.8 | 0.01 |
| Crohn’s disease | 0.1 | 0.1 | 0.00 | 0.1 | 0.1 | 0.01 | 0.1 | 0.1 | 0.00 |
| Dementia | 0.3 | 0.8 | 0.07 | 0.3 | 0.4 | 0.00 | 0.7 | 0.7 | 0.00 |
| Depressive disorder | 6.6 | 4.9 | -0.07 | 5.7 | 5.9 | 0.01 | 5.6 | 5.3 | -0.01 |
| Gastroesophageal reflux disease | 7.5 | 7.2 | -0.01 | 7.1 | 7.2 | 0.00 | 7.3 | 7.3 | 0.00 |
| Gastrointestinal hemorrhage | 1.1 | 1.5 | 0.03 | 1.1 | 1.1 | 0.00 | 1.3 | 1.3 | 0.00 |
| Human immunodeficiency virus infection | 0.5 | 0.5 | 0.00 | 0.5 | 0.5 | 0.01 | 0.5 | 0.5 | 0.00 |
| Hyperlipidemia | 42.8 | 47.3 | 0.09 | 42.6 | 42.8 | 0.00 | 46.0 | 46.1 | 0.00 |
| Hypertensive disorder | 49.1 | 51.5 | 0.05 | 48.0 | 48.1 | 0.00 | 50.8 | 50.8 | 0.00 |
| Lesion of liver | 0.6 | 0.8 | 0.02 | 0.6 | 0.6 | 0.00 | 0.6 | 0.7 | 0.01 |
| Obesity | 18.0 | 9.5 | -0.25 | 13.7 | 14.1 | 0.01 | 12.2 | 11.4 | -0.03 |
| Osteoarthritis | 13.3 | 13.1 | 0.00 | 12.7 | 12.7 | 0.00 | 13.4 | 13.2 | 0.00 |
| Pneumonia | 2.0 | 2.3 | 0.02 | 1.9 | 2.0 | 0.00 | 2.3 | 2.2 | 0.00 |
| Psoriasis | 1.3 | 0.9 | -0.03 | 1.1 | 1.1 | 0.00 | 1.1 | 1.0 | -0.01 |
| Renal impairment | 4.7 | 7.3 | 0.11 | 5.1 | 5.2 | 0.01 | 6.3 | 6.7 | 0.02 |
| Rheumatoid arthritis | 0.6 | 0.6 | 0.01 | 0.5 | 0.6 | 0.01 | 0.6 | 0.6 | 0.01 |
| Schizophrenia | 0.3 | 0.4 | 0.02 | 0.3 | 0.4 | 0.01 | 0.4 | 0.4 | 0.00 |
| Ulcerative colitis | 0.2 | 0.2 | 0.01 | 0.2 | 0.2 | 0.00 | 0.2 | 0.2 | 0.00 |
| Urinary tract infectious disease | 1.9 | 2.9 | 0.06 | 2.0 | 2.1 | 0.01 | 2.6 | 2.7 | 0.00 |
| Viral hepatitis C | 0.3 | 0.5 | 0.04 | 0.3 | 0.3 | 0.00 | 0.5 | 0.4 | -0.01 |
| Visual system disorder | 14.5 | 17.8 | 0.09 | 14.9 | 15.1 | 0.00 | 16.6 | 17.1 | 0.01 |
| **Medical history: Cardiovascular disease** | | | | | | | | | |
| Atrial fibrillation | 3.6 | 4.7 | 0.05 | 3.7 | 3.5 | -0.01 | 4.4 | 4.3 | 0.00 |
| Cerebrovascular disease | 1.9 | 3.6 | 0.10 | 2.1 | 2.1 | 0.00 | 3.0 | 3.1 | 0.00 |
| Coronary arteriosclerosis | 8.2 | 11.3 | 0.10 | 8.5 | 8.3 | 0.00 | 10.0 | 10.3 | 0.01 |
| Heart disease | 16.3 | 20.7 | 0.11 | 16.5 | 16.3 | -0.01 | 19.2 | 19.5 | 0.01 |
| Heart failure | 2.9 | 4.2 | 0.06 | 3.0 | 3.0 | 0.00 | 3.7 | 3.8 | 0.00 |
| Ischemic heart disease | 3.5 | 5.0 | 0.07 | 3.7 | 3.5 | -0.01 | 4.5 | 4.6 | 0.00 |
| Peripheral vascular disease | 4.2 | 5.0 | 0.04 | 4.1 | 4.1 | 0.00 | 4.6 | 4.7 | 0.00 |
| Pulmonary embolism | 0.5 | 0.5 | -0.01 | 0.5 | 0.5 | 0.00 | 0.5 | 0.5 | 0.00 |
| Venous thrombosis | 0.7 | 1.0 | 0.03 | 0.7 | 0.8 | 0.01 | 0.9 | 0.9 | 0.00 |
| **Medical history: Neoplasms** | | | | | | | | | |
| Hematologic neoplasm | 0.5 | 0.8 | 0.03 | 0.6 | 0.6 | 0.01 | 0.7 | 0.8 | 0.01 |
| Malignant lymphoma | 0.3 | 0.4 | 0.02 | 0.3 | 0.4 | 0.00 | 0.4 | 0.4 | 0.00 |
| Malignant neoplasm of anorectum | 0.1 | 0.2 | 0.02 | 0.1 | 0.1 | 0.01 | 0.1 | 0.2 | 0.00 |
| Malignant neoplastic disease | 4.4 | 6.7 | 0.10 | 4.7 | 4.9 | 0.01 | 6.0 | 6.2 | 0.01 |
| Malignant tumor of breast | 0.1 | 0.1 | 0.00 | 0.1 | 0.1 | 0.00 | 0.1 | 0.1 | 0.00 |
| Malignant tumor of colon | 0.2 | 0.4 | 0.03 | 0.2 | 0.3 | 0.00 | 0.3 | 0.3 | 0.00 |

| (Continued) | Before ajustment | | | After matching | | | After stratification | | |
|---|---|---|---|---|---|---|---|---|---|
| Characteristic | T (%) | C (%) | StdDiff | T (%) | C (%) | StdDiff | T (%) | C (%) | StdDiff |
| Malignant tumor of lung | 0.1 | 0.2 | 0.03 | 0.1 | 0.1 | 0.00 | 0.2 | 0.2 | 0.00 |
| Malignant tumor of urinary bladder | 0.3 | 0.5 | 0.04 | 0.3 | 0.3 | 0.00 | 0.5 | 0.4 | -0.01 |
| Primary malignant neoplasm of prostate | 1.5 | 2.4 | 0.06 | 1.7 | 1.7 | 0.00 | 2.1 | 2.2 | 0.00 |
| **Medication use** | | | | | | | | | |
| Agents acting on the renin-angiotensin system | 69.0 | 70.9 | 0.04 | 69.1 | 69.2 | 0.00 | 70.3 | 70.4 | 0.00 |
| Antibacterials for systemic use | 50.5 | 51.3 | 0.02 | 50.0 | 50.2 | 0.00 | 50.9 | 51.1 | 0.00 |
| Antidepressants | 27.4 | 20.6 | -0.16 | 24.6 | 24.7 | 0.00 | 23.2 | 22.5 | -0.02 |
| Antiepileptics | 17.7 | 15.6 | -0.06 | 16.7 | 17.1 | 0.01 | 16.4 | 16.3 | 0.00 |
| Antiinflammatory and antirheumatic products | 33.2 | 33.2 | 0.00 | 32.8 | 33.3 | 0.01 | 33.0 | 33.1 | 0.00 |
| Antineoplastic agents | 4.5 | 4.3 | -0.01 | 4.2 | 4.3 | 0.01 | 4.3 | 4.3 | 0.00 |
| Antipsoriatics | 0.6 | 0.5 | -0.01 | 0.6 | 0.5 | -0.01 | 0.6 | 0.5 | -0.01 |
| Antithrombotic agents | 16.8 | 21.4 | 0.12 | 17.7 | 17.3 | -0.01 | 19.8 | 20.1 | 0.01 |
| Beta blocking agents | 30.3 | 33.8 | 0.07 | 30.7 | 30.6 | 0.00 | 33.0 | 33.0 | 0.00 |
| Calcium channel blockers | 25.6 | 26.0 | 0.01 | 25.4 | 25.4 | 0.00 | 25.5 | 25.8 | 0.01 |
| Diuretics | 37.3 | 35.4 | -0.04 | 35.9 | 36.1 | 0.00 | 35.9 | 35.8 | 0.00 |
| Drugs for acid related disorders | 28.3 | 28.7 | 0.01 | 27.8 | 28.1 | 0.01 | 28.4 | 28.9 | 0.01 |
| Drugs for obstructive airway diseases | 34.4 | 32.2 | -0.05 | 33.1 | 33.6 | 0.01 | 32.6 | 32.9 | 0.01 |
| Immunosuppressants | 2.2 | 1.9 | -0.02 | 2.1 | 2.2 | 0.00 | 2.0 | 2.1 | 0.00 |
| Lipid modifying agents | 71.1 | 73.6 | 0.06 | 72.0 | 72.3 | 0.01 | 72.9 | 73.1 | 0.01 |
| Opioids | 21.7 | 23.0 | 0.03 | 21.6 | 21.5 | 0.00 | 22.8 | 22.7 | 0.00 |
| Psycholeptics | 20.0 | 18.7 | -0.03 | 18.9 | 19.0 | 0.00 | 19.5 | 19.1 | -0.01 |
| Psychostimulants | 3.7 | 2.0 | -0.11 | 2.5 | 2.4 | -0.01 | 2.5 | 2.4 | -0.01 |

##### 2.4.5 OEHR

**Supplementary Table 184:** Baseline patient characteristics for GLP1RA (male) (T) and DPP4I (male) (C) new-users in the OEHR data source. We report proportion of initiators satisfying selected base-line characteristics and the standardized difference of population proportions (StdDiff) before and after propensity score adjustment. Less extreme StdDiffs through matching and stratification suggest improved balance between patient cohorts through adjustment.

| | Before adjustment | | | After matching | | | After stratification | | |
|---|---|---|---|---|---|---|---|---|---|
| Characteristic | T (%) | C (%) | StdDiff | T (%) | C (%) | StdDiff | T (%) | C (%) | StdDiff |
| **Age group** | | | | | | | | | |
| 15 - 19 | 0.1 | 0.1 | -0.03 | 0.1 | 0.1 | 0.00 | 0.1 | 0.1 | 0.00 |
| 20 - 24 | 0.5 | 0.2 | -0.04 | 0.5 | 0.4 | 0.00 | 0.3 | 0.3 | -0.01 |
| 25 - 29 | 1.2 | 0.6 | -0.06 | 1.1 | 0.7 | -0.04 | 0.9 | 0.7 | -0.03 |
| 30 - 34 | 2.8 | 1.6 | -0.08 | 2.6 | 2.5 | -0.01 | 2.1 | 1.8 | -0.02 |
| 35 - 39 | 5.7 | 3.8 | -0.09 | 5.3 | 4.9 | -0.01 | 4.7 | 4.1 | -0.03 |
| 40 - 44 | 9.1 | 6.9 | -0.08 | 7.6 | 7.3 | -0.01 | 7.6 | 7.3 | -0.01 |
| 45 - 49 | 13.3 | 11.1 | -0.07 | 13.0 | 12.8 | 0.00 | 11.5 | 11.5 | 0.00 |
| 50 - 54 | 16.7 | 14.4 | -0.06 | 16.3 | 16.1 | 0.00 | 15.1 | 14.8 | -0.01 |
| 55 - 59 | 17.5 | 16.5 | -0.03 | 17.6 | 18.0 | 0.01 | 16.3 | 16.8 | 0.01 |
| 60 - 64 | 15.7 | 15.5 | 0.00 | 16.0 | 16.0 | 0.00 | 15.2 | 15.6 | 0.01 |
| 65 - 69 | 9.4 | 12.1 | 0.09 | 10.0 | 10.3 | 0.01 | 11.5 | 11.6 | 0.00 |
| 70 - 74 | 5.0 | 8.2 | 0.13 | 5.5 | 5.8 | 0.01 | 7.8 | 7.6 | -0.01 |
| 75 - 79 | 2.2 | 5.3 | 0.16 | 2.5 | 2.5 | 0.00 | 4.3 | 4.7 | 0.02 |
| 80 - 84 | 0.6 | 3.0 | 0.18 | 0.7 | 0.8 | 0.00 | 2.1 | 2.5 | 0.03 |
| 85 - 89 | 0.2 | 0.8 | 0.09 | 0.2 | 0.4 | 0.04 | 0.4 | 0.7 | 0.04 |
| **Race** | | | | | | | | | |
| Asian | 1.6 | 2.9 | 0.08 | 1.8 | 2.0 | 0.01 | 1.6 | 2.6 | 0.07 |
| Black or African American | 7.4 | 9.5 | 0.07 | 7.8 | 7.9 | 0.01 | 9.2 | 9.1 | 0.00 |
| White | 83.4 | 80.1 | -0.09 | 82.9 | 82.8 | 0.00 | 81.9 | 80.8 | -0.03 |
| **Ethnicity** | | | | | | | | | |
| Hispanic or Latino | 5.0 | 6.0 | 0.04 | 5.3 | 5.5 | 0.01 | 6.2 | 5.9 | -0.01 |
| Not Hispanic or Latino | 85.8 | 85.9 | 0.00 | 85.5 | 85.2 | -0.01 | 85.7 | 85.8 | 0.00 |
| **Medical history: General** | | | | | | | | | |
| Acute respiratory disease | 15.6 | 15.0 | -0.02 | 15.1 | 14.5 | -0.02 | 14.4 | 15.0 | 0.02 |
| Attention deficit hyperactivity disorder | 1.4 | 0.8 | -0.06 | 1.3 | 1.3 | 0.00 | 1.3 | 1.3 | 0.00 |
| Chronic liver disease | 2.3 | 2.6 | 0.02 | 2.4 | 2.2 | -0.01 | 2.7 | 2.5 | -0.01 |
| Chronic obstructive lung disease | 4.3 | 4.9 | 0.03 | 4.2 | 4.7 | 0.02 | 4.7 | 4.8 | 0.00 |
| Crohn's disease | 0.2 | 0.2 | 0.00 | 0.2 | 0.2 | 0.01 | 0.1 | 0.2 | 0.02 |
| Dementia | 0.3 | 0.7 | 0.07 | 0.3 | 0.5 | 0.03 | 0.2 | 0.7 | 0.08 |
| Depressive disorder | 12.6 | 8.6 | -0.13 | 11.4 | 11.9 | 0.01 | 9.9 | 9.3 | -0.02 |
| Gastroesophageal reflux disease | 15.5 | 13.7 | -0.05 | 14.6 | 15.7 | 0.03 | 14.2 | 14.1 | 0.00 |
| Gastrointestinal hemorrhage | 1.4 | 1.5 | 0.00 | 1.4 | 1.6 | 0.02 | 1.7 | 1.5 | -0.02 |
| Human immunodeficiency virus infection | 0.4 | 0.3 | -0.01 | 0.4 | 0.4 | 0.01 | 0.3 | 0.4 | 0.02 |
| Hyperlipidemia | 67.8 | 70.1 | 0.05 | 67.2 | 67.4 | 0.00 | 69.9 | 69.5 | -0.01 |
| Hypertensive disorder | 68.4 | 67.1 | -0.03 | 67.4 | 67.5 | 0.00 | 69.0 | 67.3 | -0.04 |
| Lesion of liver | 0.7 | 0.8 | 0.00 | 0.7 | 0.9 | 0.02 | 0.5 | 0.8 | 0.03 |
| Obesity | 41.8 | 23.3 | -0.40 | 36.4 | 38.8 | 0.05 | 27.4 | 26.8 | -0.02 |
| Osteoarthritis | 15.5 | 13.8 | -0.05 | 14.7 | 15.3 | 0.02 | 14.6 | 14.1 | -0.01 |
| Pneumonia | 1.8 | 1.7 | 0.00 | 1.6 | 1.7 | 0.00 | 1.9 | 1.7 | -0.01 |
| Psoriasis | 1.7 | 1.4 | -0.02 | 1.6 | 1.8 | 0.02 | 1.5 | 1.5 | 0.00 |
| Renal impairment | 5.4 | 6.4 | 0.04 | 5.5 | 5.7 | 0.01 | 5.8 | 6.3 | 0.02 |
| Rheumatoid arthritis | 0.6 | 0.6 | 0.00 | 0.6 | 0.6 | -0.01 | 0.6 | 0.6 | 0.00 |
| Schizophrenia | 0.3 | 0.3 | 0.01 | 0.2 | 0.4 | 0.04 | 0.3 | 0.3 | 0.00 |
| Ulcerative colitis | 0.3 | 0.2 | -0.01 | 0.2 | 0.3 | 0.01 | 0.2 | 0.2 | 0.00 |
| Urinary tract infectious disease | 1.4 | 2.1 | 0.06 | 1.4 | 1.5 | 0.01 | 1.8 | 2.0 | 0.02 |
| Visual system disorder | 11.1 | 11.7 | 0.02 | 11.0 | 11.1 | 0.00 | 10.4 | 11.6 | 0.04 |
| **Medical history: Cardiovascular disease** | | | | | | | | | |
| Atrial fibrillation | 4.2 | 4.9 | 0.04 | 4.1 | 4.3 | 0.01 | 5.0 | 4.8 | -0.01 |
| Cerebrovascular disease | 2.1 | 2.8 | 0.05 | 2.1 | 2.3 | 0.01 | 2.2 | 2.7 | 0.04 |
| Coronary arteriosclerosis | 11.0 | 13.6 | 0.08 | 11.2 | 11.1 | 0.00 | 13.3 | 13.1 | -0.01 |
| Heart disease | 20.8 | 23.3 | 0.06 | 20.8 | 20.7 | 0.00 | 23.4 | 22.7 | -0.02 |
| Heart failure | 2.9 | 3.5 | 0.03 | 2.8 | 3.2 | 0.02 | 3.1 | 3.4 | 0.02 |
| Ischemic heart disease | 5.1 | 5.7 | 0.03 | 5.1 | 4.8 | -0.01 | 6.0 | 5.5 | -0.02 |
| Peripheral vascular disease | 5.1 | 4.7 | -0.02 | 4.8 | 5.1 | 0.01 | 4.5 | 4.8 | 0.01 |
| Pulmonary embolism | 0.5 | 0.5 | 0.00 | 0.5 | 0.6 | 0.01 | 0.6 | 0.5 | -0.01 |
| Venous thrombosis | 1.0 | 1.2 | 0.02 | 1.0 | 1.2 | 0.02 | 1.1 | 1.1 | 0.01 |
| **Medical history: Neoplasms** | | | | | | | | | |

| (Continued) | Before ajustment | | | After matching | | | After stratification | | |
|---|---|---|---|---|---|---|---|---|---|
| Characteristic | T (%) | C (%) | StdDiff | T (%) | C (%) | StdDiff | T (%) | C (%) | StdDiff |
| Hematologic neoplasm | 0.6 | 0.8 | 0.02 | 0.6 | 0.8 | 0.02 | 0.8 | 0.8 | 0.00 |
| Malignant lymphoma | 0.4 | 0.4 | 0.00 | 0.4 | 0.3 | -0.03 | 0.5 | 0.4 | -0.01 |
| Malignant neoplasm of anorectum | 0.1 | 0.1 | 0.00 | 0.1 | <0.1 | -0.02 | 0.2 | 0.1 | -0.03 |
| Malignant neoplastic disease | 4.8 | 6.5 | 0.07 | 5.1 | 5.0 | 0.00 | 6.9 | 6.2 | -0.03 |
| Malignant tumor of colon | 0.2 | 0.3 | 0.02 | 0.2 | 0.2 | -0.01 | 0.2 | 0.3 | 0.01 |
| Malignant tumor of lung | 0.1 | 0.2 | 0.04 | 0.1 | 0.2 | 0.02 | 0.1 | 0.2 | 0.04 |
| Malignant tumor of urinary bladder | 0.3 | 0.4 | 0.02 | 0.3 | 0.4 | 0.01 | 0.5 | 0.4 | -0.02 |
| Primary malignant neoplasm of prostate | 1.3 | 2.3 | 0.08 | 1.4 | 1.5 | 0.01 | 2.5 | 2.1 | -0.03 |
| **Medication use** | | | | | | | | | |
| Agents acting on the renin-angiotensin system | 69.3 | 68.0 | -0.03 | 69.2 | 69.5 | 0.01 | 69.9 | 68.3 | -0.03 |
| Antibacterials for systemic use | 38.4 | 36.9 | -0.03 | 37.3 | 37.8 | 0.01 | 37.1 | 37.1 | 0.00 |
| Antidepressants | 27.3 | 20.3 | -0.17 | 25.4 | 25.5 | 0.00 | 21.8 | 21.5 | -0.01 |
| Antiepileptics | 15.7 | 13.1 | -0.07 | 15.1 | 15.5 | 0.01 | 15.1 | 15.5 | 0.01 |
| Antiinflammatory and antirheumatic products | 54.5 | 53.7 | -0.02 | 54.0 | 54.6 | 0.01 | 54.6 | 53.9 | -0.01 |
| Antineoplastic agents | 3.9 | 3.5 | -0.02 | 3.7 | 3.8 | 0.00 | 3.8 | 3.5 | -0.02 |
| Antipsoriatics | 0.5 | 0.5 | 0.00 | 0.4 | 0.6 | 0.02 | 0.4 | 0.5 | 0.03 |
| Antithrombotic agents | 38.4 | 42.3 | 0.08 | 39.0 | 39.7 | 0.01 | 42.6 | 41.7 | -0.02 |
| Beta blocking agents | 30.0 | 31.8 | 0.04 | 30.1 | 30.7 | 0.01 | 31.6 | 31.5 | 0.00 |
| Calcium channel blockers | 23.0 | 22.4 | -0.01 | 22.8 | 22.5 | -0.01 | 22.9 | 22.6 | -0.01 |
| Diuretics | 37.1 | 34.2 | -0.06 | 35.8 | 36.3 | 0.01 | 36.1 | 34.7 | -0.03 |
| Drugs for acid related disorders | 31.3 | 29.8 | -0.03 | 30.4 | 31.7 | 0.03 | 30.5 | 30.2 | -0.01 |
| Drugs for obstructive airway diseases | 39.4 | 34.9 | -0.09 | 37.8 | 38.6 | 0.02 | 35.6 | 35.9 | 0.01 |
| Immunosuppressants | 2.5 | 1.6 | -0.06 | 2.4 | 2.1 | -0.02 | 1.9 | 1.8 | -0.01 |
| Lipid modifying agents | 72.1 | 73.3 | 0.03 | 72.3 | 72.4 | 0.00 | 74.4 | 73.1 | -0.03 |
| Opioids | 23.4 | 23.0 | -0.01 | 23.0 | 23.4 | 0.01 | 24.6 | 23.0 | -0.04 |
| Psycholeptics | 24.6 | 22.9 | -0.04 | 23.6 | 24.0 | 0.01 | 23.9 | 23.2 | -0.02 |
| Psychostimulants | 4.1 | 2.3 | -0.10 | 3.6 | 3.5 | 0.00 | 2.7 | 2.5 | -0.01 |

### 2.4.6 VA

**Supplementary Table 185:** Baseline patient characteristics for GLP1RA (male) (T) and DPP4I (male) (C) new-users in the VA data source. We report proportion of initiators satisfying selected base-line characteristics and the standardized difference of population proportions (StdDiff) before and after propensity score adjustment. Less extreme StdDiffs through matching and stratification suggest improved balance between patient cohorts through adjustment.

| | Before adjustment | | | After matching | | | After stratification | | |
|---|---|---|---|---|---|---|---|---|---|
| Characteristic | T (%) | C (%) | StdDiff | T (%) | C (%) | StdDiff | T (%) | C (%) | StdDiff |
| **Age group** | | | | | | | | | |
| 25 - 29 | <0.7 | 0.1 | -0.02 | <0.6 | <0.8 | 0.03 | <0.2 | 0.1 | 0.03 |
| 30 - 34 | 0.8 | 0.7 | -0.01 | 0.7 | 1.3 | 0.06 | 0.3 | 0.7 | 0.05 |
| 35 - 39 | 1.8 | 1.7 | -0.01 | 1.7 | 2.7 | 0.07 | 0.6 | 1.7 | 0.10 |
| 40 - 44 | 4.1 | 2.8 | -0.07 | 3.9 | 4.3 | 0.02 | 4.0 | 2.8 | -0.06 |
| 45 - 49 | 7.1 | 5.0 | -0.09 | 6.6 | 7.1 | 0.02 | 4.9 | 5.1 | 0.01 |
| 50 - 54 | 10.6 | 8.4 | -0.07 | 10.0 | 9.1 | -0.03 | 18.4 | 8.5 | -0.29 |
| 55 - 59 | 12.0 | 10.5 | -0.04 | 11.9 | 11.0 | -0.03 | 9.5 | 10.6 | 0.04 |
| 60 - 64 | 12.4 | 12.7 | 0.01 | 12.4 | 12.9 | 0.01 | 11.9 | 12.7 | 0.03 |
| 65 - 69 | 15.2 | 18.8 | 0.10 | 16.2 | 15.7 | -0.01 | 11.5 | 18.7 | 0.20 |
| 70 - 74 | 22.6 | 22.6 | 0.00 | 23.0 | 19.9 | -0.08 | 27.7 | 22.5 | -0.12 |
| 75 - 79 | 8.8 | 8.8 | 0.00 | 8.5 | 9.0 | 0.02 | 6.3 | 8.9 | 0.10 |
| 80 - 84 | 3.0 | 4.4 | 0.07 | 3.4 | 4.3 | 0.04 | 4.3 | 4.4 | 0.00 |
| 85 - 89 | 1.1 | 2.4 | 0.10 | 1.2 | 1.4 | 0.01 | 0.4 | 2.4 | 0.17 |
| 90 - 94 | <0.7 | 0.9 | 0.08 | <0.6 | 1.2 | 0.10 | <0.2 | 0.9 | 0.10 |
| 95 - 99 | <0.7 | 0.2 | 0.03 | <0.6 | <0.8 | 0.02 | <0.2 | 0.2 | 0.05 |
| **Race** | | | | | | | | | |
| Asian | 1.4 | 1.6 | 0.01 | 1.5 | 1.2 | -0.02 | 1.1 | 1.6 | 0.05 |
| Black or African American | 12.2 | 16.0 | 0.11 | 12.2 | 13.1 | 0.03 | 10.0 | 15.8 | 0.18 |
| White | 77.3 | 74.3 | -0.07 | 77.6 | 77.5 | 0.00 | 81.6 | 74.4 | -0.17 |
| Unknown | 7.2 | 6.1 | -0.04 | 6.7 | 5.5 | -0.05 | 4.7 | 6.1 | 0.06 |
| Native Hawaiian or Other Pacific Islander | 1.4 | 1.2 | -0.02 | 1.4 | 1.2 | -0.01 | 2.5 | 1.2 | -0.10 |
| American Indian or Alaska Native | <0.7 | 0.8 | 0.03 | <0.6 | 1.4 | 0.09 | <0.2 | 0.8 | 0.09 |
| **Ethnicity** | | | | | | | | | |
| Hispanic or Latino | 6.2 | 6.2 | 0.00 | 6.4 | 6.3 | 0.00 | 5.9 | 6.2 | 0.01 |
| Not Hispanic or Latino | 90.5 | 91.4 | 0.03 | 90.4 | 91.2 | 0.03 | 91.6 | 91.4 | -0.01 |
| **Medical history: General** | | | | | | | | | |
| Acute respiratory disease | 6.9 | 8.1 | 0.04 | 6.5 | 6.8 | 0.01 | 16.6 | 8.1 | -0.26 |
| Attention deficit hyperactivity disorder | 1.3 | 0.7 | -0.06 | 1.0 | 1.4 | 0.04 | 1.9 | 0.7 | -0.11 |
| Chronic liver disease | 2.4 | 2.0 | -0.03 | 2.2 | 1.9 | -0.02 | 1.8 | 2.0 | 0.02 |
| Chronic obstructive lung disease | 11.2 | 11.4 | 0.01 | 10.8 | 11.3 | 0.02 | 5.5 | 11.4 | 0.21 |
| Crohn’s disease | <0.7 | 0.2 | -0.03 | <0.6 | <0.8 | -0.07 | <0.2 | 0.2 | 0.01 |
| Dementia | 1.8 | 1.7 | 0.00 | 1.7 | 2.1 | 0.03 | 1.1 | 1.7 | 0.05 |
| Depressive disorder | 26.3 | 20.3 | -0.14 | 24.6 | 24.0 | -0.01 | 14.2 | 20.6 | 0.17 |
| Gastroesophageal reflux disease | 21.3 | 19.4 | -0.05 | 20.5 | 18.5 | -0.05 | 15.0 | 19.4 | 0.12 |
| Gastrointestinal hemorrhage | 1.1 | 1.2 | 0.01 | 1.0 | 1.7 | 0.06 | 10.3 | 1.2 | -0.40 |
| Human immunodeficiency virus infection | 0.7 | 0.5 | -0.02 | 0.8 | <0.8 | -0.04 | 0.9 | 0.5 | -0.04 |
| Hyperlipidemia | 71.1 | 73.1 | 0.04 | 71.1 | 70.2 | -0.02 | 61.4 | 73.0 | 0.25 |
| Hypertensive disorder | 75.0 | 74.6 | -0.01 | 74.4 | 70.7 | -0.08 | 74.9 | 74.5 | -0.01 |
| Lesion of liver | 2.2 | 1.4 | -0.06 | 2.3 | 1.4 | -0.07 | 2.3 | 1.4 | -0.07 |
| Obesity | 46.9 | 28.1 | -0.40 | 41.7 | 48.1 | 0.13 | 26.1 | 28.9 | 0.06 |
| Osteoarthritis | 24.9 | 20.8 | -0.10 | 23.6 | 22.8 | -0.02 | 16.7 | 20.9 | 0.11 |
| Pneumonia | 1.0 | 1.4 | 0.04 | 0.9 | 1.2 | 0.03 | 0.4 | 1.4 | 0.11 |
| Psoriasis | 2.0 | 1.8 | -0.02 | 2.0 | 3.3 | 0.08 | 2.7 | 1.8 | -0.06 |
| Renal impairment | 10.5 | 8.4 | -0.07 | 9.9 | 10.9 | 0.03 | 7.3 | 8.5 | 0.04 |
| Rheumatoid arthritis | 0.7 | 0.7 | 0.00 | 0.6 | <0.8 | -0.03 | 0.3 | 0.7 | 0.05 |
| Schizophrenia | <0.7 | 0.9 | 0.06 | <0.6 | <0.8 | 0.00 | <0.2 | 0.9 | 0.11 |
| Ulcerative colitis | <0.7 | 0.4 | 0.04 | <0.6 | <0.8 | 0.01 | <0.2 | 0.4 | 0.08 |
| Urinary tract infectious disease | 2.3 | 2.1 | -0.01 | 2.2 | 1.7 | -0.03 | 1.6 | 2.1 | 0.04 |
| Viral hepatitis C | 0.9 | 1.2 | 0.03 | 0.9 | <0.8 | -0.04 | 1.4 | 1.1 | -0.02 |
| Visual system disorder | 40.8 | 44.1 | 0.07 | 39.8 | 37.5 | -0.05 | 42.6 | 43.8 | 0.03 |
| **Medical history: Cardiovascular disease** | | | | | | | | | |
| Atrial fibrillation | 9.9 | 7.4 | -0.09 | 9.7 | 9.2 | -0.02 | 7.2 | 7.5 | 0.01 |
| Cerebrovascular disease | 3.1 | 3.2 | 0.00 | 3.0 | 2.4 | -0.04 | 1.3 | 3.2 | 0.13 |
| Coronary arteriosclerosis | 23.2 | 15.4 | -0.20 | 22.1 | 22.8 | 0.02 | 14.0 | 15.8 | 0.05 |
| Heart disease | 35.9 | 28.7 | -0.16 | 34.7 | 35.1 | 0.01 | 25.5 | 29.1 | 0.08 |
| Heart failure | 6.5 | 4.2 | -0.10 | 5.7 | 5.5 | -0.01 | 3.4 | 4.3 | 0.04 |
| Ischemic heart disease | 8.2 | 6.6 | -0.06 | 7.7 | 7.2 | -0.02 | 8.6 | 6.7 | -0.07 |

| (Continued) | Before ajustment | | | After matching | | | After stratification | | |
|---|---|---|---|---|---|---|---|---|---|
| Characteristic | T (%) | C (%) | StdDiff | T (%) | C (%) | StdDiff | T (%) | C (%) | StdDiff |
| Peripheral vascular disease | 5.5 | 4.6 | -0.04 | 5.2 | 5.1 | -0.01 | 2.2 | 4.6 | 0.13 |
| Pulmonary embolism | 1.1 | 0.6 | -0.05 | 1.1 | <0.8 | -0.03 | 0.4 | 0.6 | 0.02 |
| Venous thrombosis | 0.9 | 1.2 | 0.02 | 0.7 | 1.2 | 0.05 | <0.2 | 1.2 | 0.13 |
| **Medical history: Neoplasms** | | | | | | | | | |
| Hematologic neoplasm | 1.2 | 1.1 | -0.01 | 1.1 | <0.8 | -0.05 | 1.3 | 1.1 | -0.01 |
| Malignant lymphoma | 0.7 | 0.7 | 0.00 | 0.8 | <0.8 | -0.09 | 0.9 | 0.7 | -0.03 |
| Malignant neoplastic disease | 7.1 | 9.1 | 0.07 | 7.3 | 7.2 | 0.00 | 8.6 | 9.0 | 0.01 |
| Malignant tumor of colon | <0.7 | 0.5 | 0.03 | <0.6 | <0.8 | 0.01 | <0.2 | 0.4 | 0.08 |
| Malignant tumor of lung | <0.7 | 0.3 | 0.06 | <0.6 | <0.8 | 0.02 | <0.2 | 0.3 | 0.08 |
| Malignant tumor of urinary bladder | <0.7 | 0.6 | 0.02 | <0.6 | <0.8 | 0.01 | <0.2 | 0.6 | 0.07 |
| Primary malignant neoplasm of prostate | 2.6 | 3.6 | 0.06 | 2.7 | 3.5 | 0.04 | 5.1 | 3.6 | -0.07 |
| **Medication use** | | | | | | | | | |
| Agents acting on the renin-angiotensin system | 61.4 | 63.9 | 0.05 | 61.0 | 57.1 | -0.08 | 55.9 | 63.7 | 0.16 |
| Antibacterials for systemic use | 23.4 | 26.0 | 0.06 | 22.6 | 20.5 | -0.05 | 27.8 | 25.8 | -0.04 |
| Antidepressants | 41.2 | 33.0 | -0.17 | 39.5 | 38.1 | -0.03 | 34.7 | 33.4 | -0.03 |
| Antiepileptics | 29.9 | 25.0 | -0.11 | 27.8 | 29.7 | 0.04 | 34.4 | 25.1 | -0.20 |
| Antiinflammatory and antirheumatic products | 47.5 | 46.8 | -0.01 | 46.2 | 46.7 | 0.01 | 47.6 | 46.8 | -0.02 |
| Antineoplastic agents | 5.3 | 4.1 | -0.06 | 5.1 | 4.0 | -0.05 | 3.3 | 4.1 | 0.04 |
| Antipsoriatics | 0.9 | 0.9 | 0.00 | 0.8 | 1.5 | 0.07 | 1.9 | 0.9 | -0.09 |
| Antithrombotic agents | 36.1 | 32.9 | -0.07 | 35.3 | 33.5 | -0.04 | 25.9 | 33.1 | 0.16 |
| Beta blocking agents | 37.0 | 35.0 | -0.04 | 36.5 | 35.5 | -0.02 | 28.4 | 35.0 | 0.14 |
| Calcium channel blockers | 24.5 | 27.1 | 0.06 | 23.9 | 21.7 | -0.05 | 16.9 | 27.0 | 0.25 |
| Diuretics | 39.1 | 35.2 | -0.08 | 37.7 | 36.4 | -0.03 | 33.9 | 35.3 | 0.03 |
| Drugs for acid related disorders | 40.3 | 40.3 | 0.00 | 39.5 | 38.9 | -0.01 | 40.4 | 40.2 | 0.00 |
| Drugs for obstructive airway diseases | 39.1 | 37.1 | -0.04 | 37.9 | 37.7 | 0.00 | 40.4 | 37.2 | -0.07 |
| Immunosuppressants | 3.6 | 2.2 | -0.08 | 3.5 | 3.2 | -0.02 | 2.3 | 2.3 | 0.00 |
| Lipid modifying agents | 74.3 | 78.8 | 0.10 | 74.0 | 72.8 | -0.03 | 75.1 | 78.5 | 0.08 |
| Opioids | 16.0 | 15.5 | -0.01 | 15.3 | 14.8 | -0.01 | 26.7 | 15.4 | -0.28 |
| Psycholeptics | 23.7 | 21.6 | -0.05 | 22.8 | 21.8 | -0.02 | 28.6 | 21.7 | -0.16 |
| Psychostimulants | 3.0 | 1.7 | -0.09 | 2.8 | 1.7 | -0.07 | 2.8 | 1.7 | -0.07 |

#### 2.5 GLP1RA (male) vs SU (male)

##### 2.5.1 CCAE

**Supplementary Table 186:** Baseline patient characteristics for GLP1RA (male) (T) and SU (male) (C) new-users in the CCAE data source. We report proportion of initiators satisfying selected base-line characteristics and the standardized difference of population proportions (StdDiff) before and after propensity score adjustment. Less extreme StdDiffs through matching and stratification suggest improved balance between patient cohorts through adjustment.

| | Before adjustment | | | After matching | | | After stratification | | |
|---|---|---|---|---|---|---|---|---|---|
| Characteristic | T (%) | C (%) | StdDiff | T (%) | C (%) | StdDiff | T (%) | C (%) | StdDiff |
| **Age group** | | | | | | | | | |
| 20 - 24 | 0.4 | 0.2 | 0.04 | 0.4 | 0.2 | 0.02 | 0.3 | 0.2 | 0.02 |
| 25 - 29 | 0.8 | 0.6 | 0.03 | 0.8 | 0.7 | 0.01 | 0.7 | 0.6 | 0.01 |
| 30 - 34 | 2.6 | 2.0 | 0.04 | 2.5 | 2.4 | 0.00 | 2.1 | 2.1 | 0.00 |
| 35 - 39 | 5.7 | 4.6 | 0.05 | 5.6 | 5.6 | 0.00 | 4.9 | 4.8 | 0.00 |
| 40 - 44 | 10.8 | 8.9 | 0.06 | 10.7 | 10.5 | 0.00 | 9.5 | 9.3 | 0.01 |
| 45 - 49 | 16.1 | 14.2 | 0.05 | 16.0 | 15.9 | 0.00 | 14.7 | 14.6 | 0.00 |
| 50 - 54 | 20.8 | 19.9 | 0.02 | 20.7 | 20.6 | 0.00 | 20.3 | 20.0 | 0.01 |
| 55 - 59 | 21.8 | 23.4 | -0.04 | 21.9 | 21.8 | 0.00 | 23.4 | 23.0 | 0.01 |
| 60 - 64 | 19.3 | 23.9 | -0.11 | 19.7 | 20.2 | -0.01 | 21.6 | 23.4 | -0.04 |
| 65 - 69 | 1.7 | 2.3 | -0.04 | 1.7 | 1.9 | -0.01 | 2.5 | 2.2 | 0.02 |
| **Medical history: General** | | | | | | | | | |
| Acute respiratory disease | 24.7 | 20.5 | 0.10 | 23.8 | 23.7 | 0.00 | 21.0 | 21.2 | 0.00 |
| Attention deficit hyperactivity disorder | 1.6 | 0.6 | 0.09 | 1.4 | 1.3 | 0.00 | 0.8 | 0.8 | 0.00 |
| Chronic liver disease | 1.5 | 2.7 | -0.08 | 1.6 | 1.6 | 0.00 | 3.3 | 2.6 | 0.04 |
| Chronic obstructive lung disease | 2.6 | 2.7 | -0.01 | 2.5 | 2.6 | -0.01 | 3.0 | 2.7 | 0.01 |
| Crohn’s disease | 0.2 | 0.2 | 0.00 | 0.2 | 0.2 | -0.01 | 0.4 | 0.2 | 0.03 |
| Dementia | 0.1 | 0.1 | 0.00 | 0.1 | 0.1 | 0.01 | 0.1 | 0.1 | -0.01 |
| Depressive disorder | 9.0 | 6.0 | 0.11 | 8.3 | 8.8 | -0.02 | 7.1 | 6.6 | 0.02 |
| Gastroesophageal reflux disease | 11.2 | 7.5 | 0.13 | 10.5 | 10.9 | -0.01 | 8.8 | 8.3 | 0.02 |
| Gastrointestinal hemorrhage | 1.9 | 1.9 | 0.00 | 1.8 | 1.9 | -0.01 | 2.0 | 1.8 | 0.01 |
| Human immunodeficiency virus infection | 0.4 | 0.3 | 0.00 | 0.3 | 0.5 | -0.03 | 0.3 | 0.4 | -0.02 |
| Hyperlipidemia | 71.3 | 62.3 | 0.19 | 70.3 | 71.0 | -0.01 | 64.9 | 64.9 | 0.00 |
| Hypertensive disorder | 70.8 | 61.6 | 0.20 | 69.9 | 70.5 | -0.01 | 65.9 | 64.0 | 0.04 |
| Lesion of liver | 0.6 | 0.8 | -0.02 | 0.6 | 0.7 | -0.01 | 0.8 | 0.8 | 0.00 |
| Obesity | 33.0 | 13.7 | 0.47 | 29.3 | 30.3 | -0.02 | 17.0 | 16.7 | 0.01 |
| Osteoarthritis | 18.5 | 13.9 | 0.13 | 17.7 | 18.0 | -0.01 | 14.7 | 14.7 | 0.00 |
| Pneumonia | 2.1 | 2.2 | 0.00 | 2.0 | 2.0 | 0.00 | 2.4 | 2.1 | 0.02 |
| Psoriasis | 2.0 | 1.2 | 0.06 | 1.9 | 1.8 | 0.00 | 1.2 | 1.3 | -0.01 |
| Renal impairment | 4.1 | 3.7 | 0.02 | 3.2 | 3.9 | -0.04 | 3.2 | 3.9 | -0.04 |
| Rheumatoid arthritis | 0.5 | 0.5 | 0.00 | 0.6 | 0.5 | 0.01 | 0.6 | 0.5 | 0.01 |
| Schizophrenia | 0.1 | 0.1 | -0.01 | 0.1 | 0.1 | 0.00 | 0.1 | 0.1 | 0.00 |
| Ulcerative colitis | 0.2 | 0.3 | -0.02 | 0.2 | 0.3 | -0.01 | 0.5 | 0.3 | 0.03 |
| Urinary tract infectious disease | 2.8 | 2.9 | -0.01 | 2.7 | 2.8 | -0.01 | 3.1 | 2.8 | 0.02 |
| Viral hepatitis C | 0.2 | 0.5 | -0.05 | 0.2 | 0.2 | 0.00 | 0.3 | 0.5 | -0.03 |
| Visual system disorder | 22.0 | 20.7 | 0.03 | 21.7 | 21.6 | 0.00 | 19.7 | 20.9 | -0.03 |
| **Medical history: Cardiovascular disease** | | | | | | | | | |
| Atrial fibrillation | 2.8 | 2.4 | 0.03 | 2.6 | 2.7 | 0.00 | 2.1 | 2.4 | -0.02 |
| Cerebrovascular disease | 2.2 | 2.5 | -0.02 | 2.1 | 2.3 | -0.01 | 2.1 | 2.3 | -0.01 |
| Coronary arteriosclerosis | 10.2 | 9.7 | 0.02 | 9.9 | 10.1 | -0.01 | 10.3 | 9.6 | 0.03 |
| Heart disease | 20.4 | 18.1 | 0.06 | 19.6 | 19.7 | 0.00 | 18.4 | 18.0 | 0.01 |
| Heart failure | 2.5 | 2.3 | 0.01 | 2.4 | 2.5 | 0.00 | 1.9 | 2.3 | -0.03 |
| Ischemic heart disease | 5.0 | 5.2 | -0.01 | 4.8 | 4.9 | 0.00 | 5.9 | 5.0 | 0.04 |
| Peripheral vascular disease | 4.1 | 3.0 | 0.06 | 3.9 | 3.8 | 0.00 | 3.5 | 3.1 | 0.02 |
| Pulmonary embolism | 0.7 | 0.4 | 0.03 | 0.7 | 0.6 | 0.01 | 0.4 | 0.5 | 0.00 |
| Venous thrombosis | 1.2 | 1.0 | 0.01 | 1.1 | 0.9 | 0.02 | 0.9 | 1.0 | -0.01 |
| **Medical history: Neoplasms** | | | | | | | | | |
| Hematologic neoplasm | 0.7 | 0.7 | 0.00 | 0.7 | 0.7 | 0.00 | 0.8 | 0.7 | 0.00 |
| Malignant lymphoma | 0.3 | 0.4 | -0.01 | 0.4 | 0.4 | 0.00 | 0.3 | 0.4 | -0.02 |
| Malignant neoplasm of anorectum | 0.1 | 0.2 | -0.02 | 0.3 | 0.2 | 0.03 | 0.3 | 0.2 | 0.03 |
| Malignant neoplastic disease | 4.6 | 5.0 | -0.02 | 4.6 | 4.8 | -0.01 | 5.6 | 4.9 | 0.03 |
| Malignant tumor of colon | 0.2 | 0.4 | -0.03 | 0.2 | 0.3 | -0.01 | 0.5 | 0.3 | 0.03 |
| Malignant tumor of lung | 0.1 | 0.2 | -0.04 | 0.1 | 0.1 | -0.01 | 0.1 | 0.2 | -0.02 |
| Malignant tumor of urinary bladder | 0.2 | 0.3 | -0.01 | 0.2 | 0.2 | 0.00 | 0.4 | 0.3 | 0.02 |
| Primary malignant neoplasm of prostate | 1.0 | 1.3 | -0.03 | 1.0 | 1.1 | -0.01 | 1.4 | 1.3 | 0.01 |
| **Medication use** | | | | | | | | | |
| Agents acting on the renin-angiotensin system | 71.3 | 68.2 | 0.07 | 71.1 | 71.1 | 0.00 | 71.3 | 69.4 | 0.04 |

| (Continued) | Before ajustment | | | After matching | | | After stratification | | |
|---|---|---|---|---|---|---|---|---|---|
| Characteristic | T (%) | C (%) | StdDiff | T (%) | C (%) | StdDiff | T (%) | C (%) | StdDiff |
| Antibacterials for systemic use | 53.6 | 50.3 | 0.06 | 52.7 | 52.8 | 0.00 | 51.7 | 50.7 | 0.02 |
| Antidepressants | 24.9 | 17.4 | 0.18 | 23.3 | 24.1 | -0.02 | 19.6 | 18.3 | 0.03 |
| Antiepileptics | 11.8 | 8.7 | 0.10 | 10.9 | 11.5 | -0.02 | 10.1 | 9.3 | 0.03 |
| Antiinflammatory and antirheumatic products | 31.7 | 27.2 | 0.10 | 30.7 | 31.1 | -0.01 | 28.4 | 28.0 | 0.01 |
| Antineoplastic agents | 3.9 | 3.2 | 0.04 | 3.7 | 3.6 | 0.00 | 2.9 | 3.1 | -0.01 |
| Antipsoriatics | 0.8 | 0.5 | 0.04 | 0.7 | 0.6 | 0.02 | 0.4 | 0.5 | -0.01 |
| Antithrombotic agents | 11.5 | 10.7 | 0.02 | 11.2 | 11.3 | 0.00 | 10.9 | 10.9 | 0.00 |
| Beta blocking agents | 26.0 | 26.0 | 0.00 | 25.7 | 25.8 | 0.00 | 27.1 | 26.1 | 0.02 |
| Calcium channel blockers | 23.5 | 20.4 | 0.07 | 22.9 | 23.4 | -0.01 | 21.1 | 21.0 | 0.00 |
| Diuretics | 36.7 | 32.9 | 0.08 | 36.0 | 36.1 | 0.00 | 35.0 | 33.6 | 0.03 |
| Drugs for acid related disorders | 22.7 | 19.0 | 0.09 | 21.7 | 21.8 | 0.00 | 19.0 | 19.4 | -0.01 |
| Drugs for obstructive airway diseases | 35.3 | 27.8 | 0.16 | 33.8 | 33.9 | 0.00 | 30.2 | 29.0 | 0.03 |
| Immunosuppressants | 2.0 | 1.4 | 0.05 | 1.9 | 1.8 | 0.01 | 1.5 | 1.5 | 0.00 |
| Lipid modifying agents | 71.1 | 68.3 | 0.06 | 70.8 | 70.9 | 0.00 | 69.9 | 69.6 | 0.00 |
| Opioids | 22.9 | 22.4 | 0.01 | 22.7 | 22.6 | 0.00 | 21.7 | 22.6 | -0.02 |
| Psycholeptics | 19.9 | 16.9 | 0.08 | 19.2 | 19.2 | 0.00 | 17.5 | 17.4 | 0.00 |
| Psychostimulants | 4.0 | 1.9 | 0.12 | 3.4 | 3.4 | 0.00 | 2.4 | 2.1 | 0.02 |

##### 2.5.2 ODOD

**Supplementary Table 187:** Baseline patient characteristics for GLP1RA (male) (T) and SU (male) (C) new-users in the ODOD data source. We report proportion of initiators satisfying selected base-line characteristics and the standardized difference of population proportions (StdDiff) before and after propensity score adjustment. Less extreme StdDiffs through matching and stratification suggest improved balance between patient cohorts through adjustment.

| | Before adjustment | | | After matching | | | After stratification | | |
|---|---|---|---|---|---|---|---|---|---|
| Characteristic | T (%) | C (%) | StdDiff | T (%) | C (%) | StdDiff | T (%) | C (%) | StdDiff |
| **Age group** | | | | | | | | | |
| 20 - 24 | 0.3 | 0.1 | 0.05 | 0.3 | 0.1 | 0.05 | 0.2 | 0.1 | 0.03 |
| 25 - 29 | 0.4 | 0.3 | 0.01 | 0.4 | 0.3 | 0.03 | 0.4 | 0.3 | 0.01 |
| 30 - 34 | 2.1 | 1.1 | 0.07 | 2.0 | 1.8 | 0.01 | 1.2 | 1.2 | 0.00 |
| 35 - 39 | 4.2 | 2.7 | 0.08 | 4.2 | 4.0 | 0.01 | 2.8 | 2.9 | 0.00 |
| 40 - 44 | 7.9 | 5.0 | 0.12 | 4.8 | 5.3 | -0.02 | 4.8 | 5.3 | -0.02 |
| 45 - 49 | 12.1 | 7.9 | 0.14 | 11.7 | 11.8 | 0.00 | 7.9 | 8.3 | -0.02 |
| 50 - 54 | 14.4 | 10.4 | 0.12 | 13.7 | 13.9 | 0.00 | 10.5 | 10.8 | -0.01 |
| 55 - 59 | 16.4 | 12.5 | 0.11 | 16.2 | 16.2 | 0.00 | 12.4 | 12.9 | -0.01 |
| 60 - 64 | 14.3 | 12.7 | 0.05 | 14.2 | 14.3 | 0.00 | 13.5 | 12.9 | 0.02 |
| 65 - 69 | 12.8 | 15.8 | -0.09 | 13.4 | 14.2 | -0.02 | 15.9 | 15.6 | 0.01 |
| 70 - 74 | 9.1 | 14.8 | -0.18 | 9.7 | 9.4 | 0.01 | 14.7 | 14.2 | 0.01 |
| 75 - 79 | 4.3 | 9.5 | -0.20 | 4.7 | 4.5 | 0.01 | 8.5 | 8.9 | -0.01 |
| 80 - 84 | 1.4 | 4.9 | -0.20 | 1.5 | 1.4 | 0.01 | 5.8 | 4.5 | 0.06 |
| 85 - 89 | 0.3 | 2.0 | -0.16 | 0.4 | 0.5 | -0.01 | 1.5 | 1.8 | -0.03 |
| 90 - 94 | <0.1 | 0.1 | -0.04 | <0.1 | 0.1 | -0.03 | <0.1 | 0.1 | -0.03 |
| **Medical history: General** | | | | | | | | | |
| Acute respiratory disease | 22.6 | 21.7 | 0.02 | 22.1 | 22.1 | 0.00 | 21.2 | 21.8 | -0.01 |
| Attention deficit hyperactivity disorder | 1.8 | 0.6 | 0.12 | 1.5 | 1.4 | 0.00 | 0.8 | 0.6 | 0.01 |
| Chronic liver disease | 1.7 | 3.0 | -0.08 | 1.8 | 1.7 | 0.01 | 2.6 | 2.9 | -0.02 |
| Chronic obstructive lung disease | 7.3 | 9.6 | -0.08 | 7.3 | 7.5 | -0.01 | 9.4 | 9.4 | 0.00 |
| Crohn’s disease | 0.2 | 0.2 | 0.00 | 0.2 | 0.3 | -0.01 | 0.3 | 0.2 | 0.01 |
| Dementia | 0.6 | 1.8 | -0.11 | 0.6 | 0.8 | -0.02 | 1.6 | 1.7 | 0.00 |
| Depressive disorder | 12.7 | 9.6 | 0.10 | 12.0 | 12.7 | -0.02 | 9.1 | 10.0 | -0.03 |
| Gastroesophageal reflux disease | 16.4 | 14.1 | 0.06 | 15.7 | 16.5 | -0.02 | 15.3 | 14.4 | 0.03 |
| Gastrointestinal hemorrhage | 2.1 | 2.6 | -0.03 | 2.1 | 2.1 | 0.00 | 2.7 | 2.5 | 0.01 |
| Human immunodeficiency virus infection | 0.6 | 0.3 | 0.04 | 0.6 | 0.4 | 0.02 | 0.4 | 0.4 | 0.00 |
| Hyperlipidemia | 79.8 | 79.2 | 0.01 | 79.3 | 79.6 | -0.01 | 81.7 | 79.2 | 0.06 |
| Hypertensive disorder | 79.1 | 78.4 | 0.02 | 78.8 | 78.7 | 0.00 | 81.4 | 78.4 | 0.07 |
| Lesion of liver | 1.0 | 1.4 | -0.04 | 1.0 | 1.2 | -0.02 | 1.0 | 1.4 | -0.04 |
| Obesity | 41.6 | 20.0 | 0.48 | 37.7 | 39.7 | -0.04 | 22.3 | 22.2 | 0.00 |
| Osteoarthritis | 25.3 | 22.6 | 0.06 | 24.3 | 24.7 | -0.01 | 24.9 | 22.8 | 0.05 |
| Pneumonia | 3.2 | 4.2 | -0.05 | 3.0 | 3.1 | -0.01 | 3.0 | 3.1 | -0.01 |
| Psoriasis | 2.2 | 1.5 | 0.05 | 2.2 | 2.2 | 0.00 | 1.4 | 1.6 | -0.02 |
| Renal impairment | 9.9 | 14.7 | -0.15 | 15.5 | 14.2 | 0.04 | 15.5 | 14.2 | 0.04 |
| Rheumatoid arthritis | 0.9 | 1.1 | -0.02 | 0.8 | 1.1 | -0.03 | 1.9 | 1.1 | 0.07 |
| Schizophrenia | 0.4 | 0.4 | 0.01 | 0.4 | 0.4 | 0.01 | 0.4 | 0.4 | 0.00 |
| Ulcerative colitis | 0.3 | 0.3 | -0.01 | 0.3 | 0.3 | 0.00 | 0.4 | 0.3 | 0.02 |
| Urinary tract infectious disease | 4.3 | 5.8 | -0.07 | 4.1 | 4.2 | -0.01 | 8.6 | 5.6 | 0.11 |
| Viral hepatitis C | 0.6 | 0.8 | -0.02 | 0.6 | 0.6 | 0.00 | 0.8 | 0.8 | 0.00 |
| Visual system disorder | 31.6 | 36.0 | -0.09 | 32.0 | 31.7 | 0.01 | 35.7 | 35.5 | 0.00 |
| **Medical history: Cardiovascular disease** | | | | | | | | | |
| Atrial fibrillation | 6.1 | 7.6 | -0.06 | 6.1 | 5.9 | 0.01 | 7.7 | 7.4 | 0.01 |
| Cerebrovascular disease | 4.0 | 6.7 | -0.12 | 4.0 | 4.0 | 0.00 | 6.0 | 6.4 | -0.02 |
| Coronary arteriosclerosis | 15.5 | 19.8 | -0.11 | 15.5 | 15.4 | 0.00 | 20.4 | 19.3 | 0.03 |
| Heart disease | 28.3 | 33.0 | -0.10 | 28.0 | 27.9 | 0.00 | 35.3 | 32.5 | 0.06 |
| Heart failure | 5.9 | 7.7 | -0.07 | 5.8 | 5.9 | 0.00 | 7.4 | 7.5 | 0.00 |
| Ischemic heart disease | 7.8 | 10.6 | -0.10 | 7.8 | 7.8 | 0.00 | 11.8 | 10.3 | 0.05 |
| Peripheral vascular disease | 9.9 | 10.6 | -0.02 | 9.9 | 10.1 | -0.01 | 10.6 | 10.5 | 0.00 |
| Pulmonary embolism | 0.8 | 0.8 | 0.01 | 0.9 | 0.7 | 0.01 | 0.8 | 0.8 | 0.00 |
| Venous thrombosis | 1.4 | 1.7 | -0.03 | 1.4 | 1.4 | 0.00 | 1.6 | 1.7 | 0.00 |
| **Medical history: Neoplasms** | | | | | | | | | |
| Hematologic neoplasm | 0.8 | 1.4 | -0.06 | 0.7 | 1.0 | -0.02 | 1.1 | 1.3 | -0.02 |
| Malignant lymphoma | 0.4 | 0.7 | -0.04 | 0.4 | 0.5 | -0.02 | 0.4 | 0.7 | -0.04 |
| Malignant neoplasm of anorectum | 0.2 | 0.3 | -0.02 | 0.3 | 0.3 | 0.00 | 0.3 | 0.3 | 0.00 |
| Malignant neoplastic disease | 7.9 | 11.4 | -0.12 | 8.1 | 8.0 | 0.00 | 10.6 | 11.0 | -0.01 |
| Malignant tumor of colon | 0.5 | 0.7 | -0.03 | 0.5 | 0.4 | 0.01 | 0.6 | 0.7 | -0.02 |
| Malignant tumor of lung | 0.2 | 0.5 | -0.05 | 0.3 | 0.2 | 0.01 | 1.0 | 0.5 | 0.06 |
| Malignant tumor of urinary bladder | 0.4 | 0.8 | -0.06 | 0.4 | 0.5 | -0.02 | 0.6 | 0.8 | -0.03 |

| (Continued) | Before ajustment | | | After matching | | | After stratification | | |
|---|---|---|---|---|---|---|---|---|---|
| Characteristic | T (%) | C (%) | StdDiff | T (%) | C (%) | StdDiff | T (%) | C (%) | StdDiff |
| Primary malignant neoplasm of prostate | 2.6 | 4.1 | -0.08 | 2.7 | 2.5 | 0.01 | 5.1 | 3.9 | 0.06 |
| **Medication use** | | | | | | | | | |
| Agents acting on the renin-angiotensin system | 72.9 | 71.6 | 0.03 | 72.8 | 72.5 | 0.01 | 74.9 | 71.7 | 0.07 |
| Antibacterials for systemic use | 53.5 | 51.7 | 0.04 | 52.5 | 52.5 | 0.00 | 51.9 | 51.7 | 0.00 |
| Antidepressants | 25.9 | 19.0 | 0.17 | 24.5 | 25.1 | -0.01 | 20.6 | 19.6 | 0.03 |
| Antiepileptics | 16.6 | 13.6 | 0.09 | 15.7 | 16.2 | -0.01 | 13.6 | 13.9 | -0.01 |
| Antiinflammatory and antirheumatic products | 30.9 | 26.2 | 0.10 | 29.7 | 30.3 | -0.01 | 25.2 | 26.7 | -0.03 |
| Antineoplastic agents | 4.7 | 4.4 | 0.01 | 4.6 | 4.4 | 0.01 | 4.3 | 4.4 | 0.00 |
| Antipsoriatics | 0.6 | 0.4 | 0.03 | 0.6 | 0.5 | 0.01 | 0.5 | 0.4 | 0.01 |
| Antithrombotic agents | 15.7 | 18.0 | -0.06 | 15.7 | 15.7 | 0.00 | 18.3 | 17.8 | 0.01 |
| Beta blocking agents | 31.3 | 35.7 | -0.09 | 38.9 | 35.2 | 0.08 | 38.9 | 35.2 | 0.08 |
| Calcium channel blockers | 25.6 | 25.9 | -0.01 | 25.3 | 25.6 | -0.01 | 25.7 | 25.9 | 0.00 |
| Diuretics | 40.5 | 38.1 | 0.05 | 39.6 | 39.5 | 0.00 | 42.2 | 38.3 | 0.08 |
| Drugs for acid related disorders | 22.2 | 22.4 | 0.00 | 21.6 | 22.0 | -0.01 | 23.9 | 22.3 | 0.04 |
| Drugs for obstructive airway diseases | 36.5 | 31.3 | 0.11 | 35.1 | 35.8 | -0.01 | 33.5 | 31.8 | 0.04 |
| Immunosuppressants | 2.0 | 1.6 | 0.03 | 2.0 | 2.0 | 0.00 | 1.5 | 1.7 | -0.01 |
| Lipid modifying agents | 74.7 | 73.8 | 0.02 | 74.7 | 74.8 | 0.00 | 77.3 | 73.9 | 0.08 |
| Opioids | 24.9 | 25.1 | 0.00 | 24.3 | 24.8 | -0.01 | 24.3 | 25.0 | -0.02 |
| Psycholeptics | 22.0 | 19.8 | 0.05 | 21.1 | 21.7 | -0.01 | 19.6 | 20.0 | -0.01 |
| Psychostimulants | 3.1 | 1.2 | 0.13 | 2.5 | 2.6 | -0.01 | 1.4 | 1.4 | 0.00 |

##### 2.5.3 MDCR

**Supplementary Table 188:** Baseline patient characteristics for GLP1RA (male) (T) and SU (male) (C) new-users in the MDCR data source. We report proportion of initiators satisfying selected base-line characteristics and the standardized difference of population proportions (StdDiff) before and after propensity score adjustment. Less extreme StdDiffs through matching and stratification suggest improved balance between patient cohorts through adjustment.

| | Before adjustment | | | After matching | | | After stratification | | |
|---|---|---|---|---|---|---|---|---|---|
| Characteristic | T (%) | C (%) | StdDiff | T (%) | C (%) | StdDiff | T (%) | C (%) | StdDiff |
| **Age group** | | | | | | | | | |
| 45 - 49 | <0.5 | 0.1 | 0.03 | 0.2 | 0.1 | 0.04 | 0.2 | 0.1 | 0.04 |
| 50 - 54 | 0.6 | 0.2 | 0.06 | <0.5 | 0.4 | -0.01 | 0.8 | 0.2 | 0.07 |
| 55 - 59 | 1.5 | 0.6 | 0.09 | 1.3 | 0.6 | 0.07 | 1.4 | 0.6 | 0.08 |
| 60 - 64 | 2.8 | 1.6 | 0.08 | 2.9 | 2.0 | 0.06 | 2.9 | 1.7 | 0.08 |
| 65 - 69 | 40.4 | 28.3 | 0.26 | 40.0 | 40.7 | -0.01 | 28.0 | 29.6 | -0.04 |
| 70 - 74 | 35.1 | 30.7 | 0.10 | 35.1 | 35.5 | -0.01 | 35.8 | 30.7 | 0.11 |
| 75 - 79 | 12.9 | 20.6 | -0.21 | 13.5 | 13.1 | 0.01 | 19.0 | 19.7 | -0.02 |
| 80 - 84 | 4.9 | 12.1 | -0.26 | 4.9 | 4.9 | 0.00 | 7.2 | 11.5 | -0.15 |
| 85 - 89 | 1.3 | 4.7 | -0.20 | 1.3 | 2.1 | -0.06 | 4.6 | 4.6 | 0.00 |
| 90 - 94 | <0.5 | 1.2 | -0.09 | <0.5 | 0.6 | -0.03 | <0.2 | 1.1 | -0.13 |
| **Medical history: General** | | | | | | | | | |
| Acute respiratory disease | 21.6 | 19.6 | 0.05 | 21.4 | 20.7 | 0.01 | 20.3 | 20.4 | 0.00 |
| Attention deficit hyperactivity disorder | <0.5 | 0.1 | 0.03 | <0.5 | 0.5 | -0.04 | <0.2 | 0.1 | -0.02 |
| Chronic liver disease | 1.7 | 1.9 | -0.02 | 1.8 | 1.9 | 0.00 | 1.2 | 2.1 | -0.08 |
| Chronic obstructive lung disease | 13.0 | 11.8 | 0.04 | 12.8 | 12.2 | 0.02 | 12.9 | 12.1 | 0.02 |
| Dementia | 1.4 | 2.5 | -0.08 | 1.5 | 1.6 | -0.01 | 1.9 | 2.6 | -0.05 |
| Depressive disorder | 8.8 | 5.2 | 0.14 | 7.6 | 9.5 | -0.07 | 7.2 | 5.9 | 0.06 |
| Gastroesophageal reflux disease | 11.9 | 8.9 | 0.10 | 11.7 | 13.4 | -0.05 | 7.6 | 10.0 | -0.09 |
| Gastrointestinal hemorrhage | 2.0 | 3.4 | -0.09 | 1.7 | 2.0 | -0.02 | 1.4 | 3.2 | -0.12 |
| Human immunodeficiency virus infection | <0.5 | 0.1 | 0.01 | <0.5 | 0.1 | 0.00 | <0.2 | 0.1 | -0.03 |
| Hyperlipidemia | 70.7 | 50.4 | 0.42 | 69.3 | 72.7 | -0.07 | 52.2 | 55.6 | -0.07 |
| Hypertensive disorder | 80.2 | 64.6 | 0.35 | 79.3 | 81.8 | -0.06 | 71.5 | 68.6 | 0.06 |
| Lesion of liver | 0.9 | 1.1 | -0.02 | 1.0 | 1.6 | -0.05 | 0.7 | 1.2 | -0.06 |
| Obesity | 27.7 | 7.3 | 0.56 | 23.8 | 27.0 | -0.07 | 9.7 | 9.0 | 0.03 |
| Osteoarthritis | 33.3 | 23.3 | 0.22 | 32.1 | 32.3 | 0.00 | 24.4 | 24.8 | -0.01 |
| Pneumonia | 4.1 | 6.0 | -0.09 | 4.3 | 4.7 | -0.02 | 2.7 | 5.9 | -0.16 |
| Psoriasis | 3.3 | 1.3 | 0.14 | 3.3 | 2.5 | 0.05 | 1.4 | 1.4 | 0.00 |
| Renal impairment | 11.1 | 12.9 | -0.05 | 11.2 | 11.5 | -0.01 | 12.2 | 14.1 | -0.06 |
| Rheumatoid arthritis | 1.2 | 1.0 | 0.01 | 1.3 | 1.1 | 0.02 | 0.6 | 1.1 | -0.05 |
| Schizophrenia | <0.5 | 0.1 | 0.02 | <0.5 | 0.1 | 0.04 | <0.2 | 0.1 | -0.03 |
| Ulcerative colitis | <0.5 | 0.4 | -0.03 | <0.5 | 0.4 | -0.03 | 0.4 | 0.4 | 0.00 |
| Urinary tract infectious disease | 4.9 | 6.2 | -0.06 | 4.7 | 4.5 | 0.01 | 3.1 | 6.2 | -0.15 |
| Viral hepatitis C | <0.5 | 0.2 | 0.01 | <0.5 | 0.4 | -0.01 | 0.2 | 0.3 | -0.01 |
| Visual system disorder | 49.9 | 45.6 | 0.09 | 49.7 | 50.7 | -0.02 | 53.0 | 46.3 | 0.13 |
| **Medical history: Cardiovascular disease** | | | | | | | | | |
| Atrial fibrillation | 11.4 | 12.8 | -0.04 | 11.4 | 11.3 | 0.00 | 13.3 | 13.0 | 0.01 |
| Cerebrovascular disease | 10.1 | 12.5 | -0.07 | 10.5 | 10.8 | -0.01 | 10.9 | 12.4 | -0.05 |
| Coronary arteriosclerosis | 30.3 | 30.1 | 0.00 | 30.0 | 30.5 | -0.01 | 26.7 | 30.2 | -0.08 |
| Heart disease | 47.7 | 48.6 | -0.02 | 47.7 | 48.2 | -0.01 | 45.4 | 48.7 | -0.06 |
| Heart failure | 8.1 | 11.2 | -0.10 | 7.8 | 9.0 | -0.04 | 7.3 | 10.8 | -0.12 |
| Ischemic heart disease | 11.4 | 14.4 | -0.09 | 11.7 | 12.5 | -0.02 | 9.4 | 14.0 | -0.14 |
| Peripheral vascular disease | 12.5 | 11.4 | 0.04 | 12.8 | 13.9 | -0.03 | 12.6 | 11.7 | 0.03 |
| Pulmonary embolism | 1.3 | 0.8 | 0.05 | 1.2 | 1.0 | 0.02 | 2.8 | 0.8 | 0.15 |
| Venous thrombosis | 1.7 | 2.1 | -0.03 | 1.6 | 1.8 | -0.02 | 1.0 | 2.2 | -0.09 |
| **Medical history: Neoplasms** | | | | | | | | | |
| Hematologic neoplasm | 2.0 | 1.9 | 0.00 | 1.8 | 2.2 | -0.03 | 1.9 | 2.0 | -0.01 |
| Malignant lymphoma | 1.2 | 0.9 | 0.03 | 1.1 | 1.6 | -0.04 | 1.6 | 1.0 | 0.06 |
| Malignant neoplastic disease | 21.4 | 21.1 | 0.01 | 21.7 | 21.3 | 0.01 | 18.8 | 21.5 | -0.07 |
| Malignant tumor of breast | <0.5 | 0.1 | 0.02 | <0.5 | 0.1 | -0.01 | <0.2 | 0.1 | -0.03 |
| Malignant tumor of colon | <0.5 | 1.1 | -0.08 | <0.5 | 0.7 | -0.03 | <0.2 | 1.0 | -0.12 |
| Malignant tumor of lung | 0.8 | 0.9 | -0.01 | 0.9 | 0.8 | 0.00 | 0.4 | 0.9 | -0.07 |
| Malignant tumor of urinary bladder | 1.9 | 1.7 | 0.01 | 1.9 | 1.8 | 0.01 | 1.3 | 1.8 | -0.04 |
| Primary malignant neoplasm of prostate | 8.4 | 8.3 | 0.00 | 8.4 | 8.1 | 0.01 | 9.1 | 8.4 | 0.03 |
| **Medication use** | | | | | | | | | |
| Agents acting on the renin-angiotensin system | 77.4 | 72.4 | 0.11 | 77.5 | 78.5 | -0.03 | 70.8 | 74.6 | -0.09 |
| Antibacterials for systemic use | 59.4 | 57.7 | 0.04 | 58.8 | 58.3 | 0.01 | 48.3 | 58.3 | -0.20 |
| Antidepressants | 27.0 | 18.6 | 0.20 | 25.2 | 25.9 | -0.02 | 20.4 | 19.4 | 0.03 |
| Antiepileptics | 17.1 | 12.1 | 0.14 | 16.4 | 16.0 | 0.01 | 11.9 | 13.0 | -0.03 |

| (Continued) | Before ajustment | | | After matching | | | After stratification | | |
|---|---|---|---|---|---|---|---|---|---|
| Characteristic | T (%) | C (%) | StdDiff | T (%) | C (%) | StdDiff | T (%) | C (%) | StdDiff |
| Antiinflammatory and antirheumatic products | 28.3 | 25.0 | 0.07 | 28.2 | 27.5 | 0.02 | 24.8 | 25.5 | -0.01 |
| Antineoplastic agents | 8.7 | 8.3 | 0.02 | 8.6 | 8.3 | 0.01 | 7.7 | 8.0 | -0.01 |
| Antipsoriatics | 0.9 | 0.6 | 0.03 | 1.0 | 0.7 | 0.04 | 0.8 | 0.6 | 0.02 |
| Antithrombotic agents | 27.8 | 28.2 | -0.01 | 28.4 | 27.6 | 0.02 | 27.6 | 28.8 | -0.03 |
| Beta blocking agents | 48.6 | 49.5 | -0.02 | 48.6 | 49.3 | -0.01 | 52.5 | 50.4 | 0.04 |
| Calcium channel blockers | 32.7 | 31.7 | 0.02 | 32.4 | 33.4 | -0.02 | 27.2 | 32.8 | -0.12 |
| Diuretics | 50.7 | 47.7 | 0.06 | 50.2 | 50.0 | 0.00 | 44.0 | 48.1 | -0.08 |
| Drugs for acid related disorders | 31.2 | 28.9 | 0.05 | 31.1 | 30.1 | 0.02 | 21.9 | 29.2 | -0.17 |
| Drugs for obstructive airway diseases | 45.1 | 35.8 | 0.19 | 43.2 | 44.2 | -0.02 | 34.9 | 37.5 | -0.05 |
| Immunosuppressants | 3.5 | 2.0 | 0.09 | 3.5 | 3.2 | 0.02 | 2.3 | 2.2 | 0.00 |
| Lipid modifying agents | 85.0 | 76.9 | 0.21 | 84.2 | 85.2 | -0.03 | 77.8 | 79.9 | -0.05 |
| Opioids | 27.3 | 26.0 | 0.03 | 27.5 | 27.6 | 0.00 | 26.1 | 26.9 | -0.02 |
| Psycholeptics | 23.0 | 21.8 | 0.03 | 22.3 | 22.9 | -0.01 | 22.7 | 22.3 | 0.01 |
| Psychostimulants | 1.5 | 0.8 | 0.06 | 1.5 | 1.4 | 0.01 | 1.1 | 0.9 | 0.02 |

##### 2.5.4 Open Claims

**Supplementary Table 189:** Baseline patient characteristics for GLP1RA (male) (T) and SU (male) (C) new-users in the Open Claims data source. We report proportion of initiators satisfying selected base-line characteristics and the standardized difference of population proportions (StdDiff) before and after propensity score adjustment. Less extreme StdDiffs through matching and stratification suggest improved balance between patient cohorts through adjustment.

| | Before adjustment | | | After matching | | | After stratification | | |
|---|---|---|---|---|---|---|---|---|---|
| Characteristic | T (%) | C (%) | StdDiff | T (%) | C (%) | StdDiff | T (%) | C (%) | StdDiff |
| **Age group** | | | | | | | | | |
| 15 - 19 | 0.2 | 0.1 | 0.03 | 0.2 | 0.1 | 0.01 | 0.1 | 0.1 | 0.01 |
| 20 - 24 | 0.5 | 0.3 | 0.04 | 0.5 | 0.4 | 0.02 | 0.3 | 0.3 | 0.01 |
| 25 - 29 | 1.0 | 0.6 | 0.05 | 0.9 | 0.9 | 0.00 | 0.7 | 0.7 | 0.00 |
| 30 - 34 | 2.3 | 1.5 | 0.06 | 2.2 | 2.3 | 0.00 | 1.5 | 1.5 | -0.01 |
| 35 - 39 | 4.7 | 2.9 | 0.09 | 4.5 | 4.5 | 0.00 | 3.3 | 3.2 | 0.00 |
| 40 - 44 | 8.1 | 5.3 | 0.11 | 7.8 | 7.6 | 0.01 | 5.8 | 5.7 | 0.00 |
| 45 - 49 | 11.8 | 8.6 | 0.11 | 11.4 | 11.7 | -0.01 | 8.8 | 9.0 | -0.01 |
| 50 - 54 | 15.1 | 11.9 | 0.09 | 14.8 | 14.6 | 0.00 | 12.4 | 12.3 | 0.00 |
| 55 - 59 | 16.8 | 14.4 | 0.06 | 16.6 | 16.6 | 0.00 | 14.7 | 14.8 | 0.00 |
| 60 - 64 | 15.5 | 15.0 | 0.01 | 15.7 | 15.9 | 0.00 | 15.2 | 15.1 | 0.00 |
| 65 - 69 | 11.6 | 14.8 | -0.09 | 12.1 | 12.0 | 0.00 | 14.4 | 14.3 | 0.00 |
| 70 - 74 | 7.1 | 11.3 | -0.15 | 7.5 | 7.7 | -0.01 | 10.8 | 10.7 | 0.00 |
| 75 - 79 | 3.6 | 9.8 | -0.25 | 3.9 | 3.8 | 0.00 | 8.5 | 8.9 | -0.02 |
| 80 - 84 | 1.6 | 3.5 | -0.12 | 1.7 | 1.8 | 0.00 | 3.4 | 3.3 | 0.01 |
| 85 - 89 | 0.1 | 0.1 | 0.01 | 0.1 | 0.1 | -0.01 | 0.1 | 0.1 | 0.00 |
| **Medical history: General** | | | | | | | | | |
| Acute respiratory disease | 11.3 | 11.1 | 0.01 | 10.8 | 11.2 | -0.01 | 11.4 | 11.1 | 0.01 |
| Attention deficit hyperactivity disorder | 1.0 | 0.4 | 0.08 | 0.8 | 0.8 | 0.00 | 0.5 | 0.5 | 0.01 |
| Chronic liver disease | 0.8 | 1.4 | -0.06 | 0.8 | 0.8 | 0.00 | 1.5 | 1.4 | 0.01 |
| Chronic obstructive lung disease | 3.9 | 5.4 | -0.07 | 3.9 | 4.2 | -0.02 | 5.3 | 5.3 | 0.00 |
| Crohn’s disease | 0.1 | 0.1 | -0.01 | 0.1 | 0.2 | -0.01 | 0.1 | 0.2 | -0.01 |
| Dementia | 0.3 | 0.8 | -0.07 | 0.3 | 0.4 | -0.02 | 0.3 | 0.4 | -0.02 |
| Depressive disorder | 6.6 | 5.0 | 0.07 | 6.0 | 6.2 | 0.00 | 5.5 | 5.2 | 0.01 |
| Gastroesophageal reflux disease | 7.5 | 6.5 | 0.04 | 7.1 | 7.4 | -0.01 | 6.8 | 6.6 | 0.01 |
| Gastrointestinal hemorrhage | 1.1 | 1.4 | -0.03 | 1.1 | 1.0 | 0.00 | 1.5 | 1.4 | 0.00 |
| Human immunodeficiency virus infection | 0.5 | 0.3 | 0.03 | 0.5 | 0.5 | 0.00 | 0.4 | 0.4 | 0.00 |
| Hyperlipidemia | 42.8 | 41.9 | 0.02 | 41.6 | 42.1 | -0.01 | 42.6 | 42.0 | 0.01 |
| Hypertensive disorder | 49.1 | 48.6 | 0.01 | 47.8 | 48.0 | 0.00 | 48.8 | 48.5 | 0.01 |
| Lesion of liver | 0.6 | 0.8 | -0.03 | 0.6 | 0.6 | -0.01 | 0.8 | 0.8 | -0.01 |
| Obesity | 18.0 | 9.1 | 0.26 | 15.5 | 15.5 | 0.00 | 11.1 | 10.2 | 0.03 |
| Osteoarthritis | 13.3 | 12.4 | 0.03 | 12.6 | 13.1 | -0.01 | 12.8 | 12.4 | 0.01 |
| Pneumonia | 2.0 | 2.5 | -0.04 | 1.9 | 2.0 | 0.00 | 2.6 | 2.5 | 0.00 |
| Psoriasis | 1.3 | 0.8 | 0.04 | 1.1 | 1.2 | -0.01 | 1.0 | 0.8 | 0.01 |
| Renal impairment | 4.7 | 7.4 | -0.11 | 4.7 | 5.1 | -0.02 | 6.8 | 7.1 | -0.01 |
| Rheumatoid arthritis | 0.6 | 0.6 | 0.00 | 0.5 | 0.6 | -0.01 | 0.5 | 0.6 | 0.00 |
| Schizophrenia | 0.3 | 0.4 | -0.02 | 0.3 | 0.3 | 0.00 | 0.5 | 0.5 | 0.00 |
| Ulcerative colitis | 0.2 | 0.2 | 0.00 | 0.2 | 0.1 | 0.01 | 0.2 | 0.2 | 0.01 |
| Urinary tract infectious disease | 1.9 | 2.7 | -0.05 | 1.9 | 2.0 | -0.01 | 2.8 | 2.6 | 0.01 |
| Viral hepatitis C | 0.3 | 0.5 | -0.04 | 0.3 | 0.3 | 0.00 | 0.5 | 0.5 | 0.00 |
| Visual system disorder | 14.5 | 16.9 | -0.07 | 14.4 | 14.6 | 0.00 | 16.1 | 16.5 | -0.01 |
| **Medical history: Cardiovascular disease** | | | | | | | | | |
| Atrial fibrillation | 3.6 | 4.6 | -0.05 | 3.5 | 3.5 | 0.00 | 4.7 | 4.4 | 0.01 |
| Cerebrovascular disease | 1.9 | 3.3 | -0.09 | 1.9 | 2.0 | -0.01 | 2.8 | 3.1 | -0.02 |
| Coronary arteriosclerosis | 8.2 | 10.5 | -0.08 | 8.1 | 8.3 | -0.01 | 10.2 | 10.2 | 0.00 |
| Heart disease | 16.3 | 19.8 | -0.09 | 16.0 | 16.2 | 0.00 | 19.6 | 19.3 | 0.01 |
| Heart failure | 2.9 | 4.5 | -0.08 | 2.9 | 3.0 | 0.00 | 4.4 | 4.3 | 0.01 |
| Ischemic heart disease | 3.5 | 4.9 | -0.07 | 3.5 | 3.7 | -0.01 | 4.5 | 4.7 | -0.01 |
| Peripheral vascular disease | 4.2 | 4.9 | -0.04 | 4.1 | 4.2 | -0.01 | 4.9 | 4.8 | 0.00 |
| Pulmonary embolism | 0.5 | 0.5 | 0.00 | 0.6 | 0.5 | 0.01 | 0.6 | 0.5 | 0.01 |
| Venous thrombosis | 0.7 | 0.9 | -0.03 | 0.7 | 0.8 | -0.01 | 0.9 | 0.9 | 0.00 |
| **Medical history: Neoplasms** | | | | | | | | | |
| Hematologic neoplasm | 0.5 | 0.8 | -0.04 | 0.5 | 0.5 | 0.01 | 0.8 | 0.8 | 0.00 |
| Malignant lymphoma | 0.3 | 0.5 | -0.03 | 0.3 | 0.3 | 0.00 | 0.5 | 0.4 | 0.01 |
| Malignant neoplasm of anorectum | 0.1 | 0.2 | -0.02 | 0.1 | 0.1 | -0.01 | 0.2 | 0.2 | 0.00 |
| Malignant neoplastic disease | 4.4 | 6.5 | -0.10 | 4.4 | 4.5 | -0.01 | 6.4 | 6.2 | 0.01 |
| Malignant tumor of breast | 0.1 | 0.1 | 0.00 | 0.1 | 0.1 | 0.01 | 0.1 | 0.1 | 0.01 |
| Malignant tumor of colon | 0.2 | 0.4 | -0.04 | 0.2 | 0.2 | 0.00 | 0.2 | 0.2 | 0.00 |

| (Continued) | Before ajustment | | | After matching | | | After stratification | | |
|---|---|---|---|---|---|---|---|---|---|
| Characteristic | T (%) | C (%) | StdDiff | T (%) | C (%) | StdDiff | T (%) | C (%) | StdDiff |
| Malignant tumor of lung | 0.1 | 0.3 | -0.04 | 0.1 | 0.2 | -0.01 | 0.3 | 0.3 | 0.00 |
| Malignant tumor of urinary bladder | 0.3 | 0.5 | -0.03 | 0.3 | 0.3 | -0.01 | 0.5 | 0.4 | 0.01 |
| Primary malignant neoplasm of prostate | 1.5 | 2.2 | -0.05 | 1.5 | 1.6 | -0.01 | 2.2 | 2.1 | 0.00 |
| **Medication use** | | | | | | | | | |
| Agents acting on the renin-angiotensin system | 69.0 | 70.4 | -0.03 | 68.7 | 69.1 | -0.01 | 70.2 | 70.2 | 0.00 |
| Antibacterials for systemic use | 50.5 | 49.3 | 0.03 | 49.6 | 50.5 | -0.02 | 49.9 | 49.5 | 0.01 |
| Antidepressants | 27.4 | 20.9 | 0.15 | 25.6 | 26.3 | -0.01 | 22.5 | 21.8 | 0.02 |
| Antiepileptics | 17.7 | 16.4 | 0.04 | 17.0 | 17.6 | -0.02 | 17.3 | 16.7 | 0.02 |
| Antiinflammatory and antirheumatic products | 33.2 | 32.0 | 0.03 | 32.5 | 33.3 | -0.02 | 32.1 | 32.1 | 0.00 |
| Antineoplastic agents | 4.5 | 3.6 | 0.04 | 4.1 | 4.3 | -0.01 | 4.0 | 3.7 | 0.01 |
| Antipsoriatics | 0.6 | 0.4 | 0.03 | 0.6 | 0.5 | 0.00 | 0.5 | 0.4 | 0.01 |
| Antithrombotic agents | 16.8 | 21.0 | -0.11 | 16.9 | 16.9 | 0.00 | 20.5 | 20.4 | 0.00 |
| Beta blocking agents | 30.3 | 34.3 | -0.09 | 30.0 | 30.5 | -0.01 | 34.1 | 33.8 | 0.00 |
| Calcium channel blockers | 25.6 | 25.5 | 0.00 | 25.3 | 25.3 | 0.00 | 25.1 | 25.4 | -0.01 |
| Diuretics | 37.3 | 36.3 | 0.02 | 36.5 | 36.7 | 0.00 | 36.7 | 36.4 | 0.01 |
| Drugs for acid related disorders | 28.3 | 27.1 | 0.03 | 27.3 | 28.1 | -0.02 | 27.4 | 27.3 | 0.00 |
| Drugs for obstructive airway diseases | 34.4 | 29.7 | 0.10 | 33.0 | 33.7 | -0.01 | 30.9 | 30.4 | 0.01 |
| Immunosuppressants | 2.2 | 1.7 | 0.04 | 2.1 | 2.1 | 0.00 | 1.8 | 1.7 | 0.01 |
| Lipid modifying agents | 71.1 | 70.7 | 0.01 | 70.8 | 71.6 | -0.02 | 71.4 | 70.8 | 0.01 |
| Opioids | 21.7 | 23.0 | -0.03 | 21.2 | 21.9 | -0.02 | 23.1 | 22.8 | 0.01 |
| Psycholeptics | 20.0 | 18.2 | 0.04 | 19.0 | 19.3 | -0.01 | 19.5 | 18.6 | 0.02 |
| Psychostimulants | 3.7 | 1.6 | 0.13 | 3.1 | 3.1 | 0.00 | 2.1 | 1.9 | 0.01 |

##### 2.5.5 OEHR

**Supplementary Table 190:** Baseline patient characteristics for GLP1RA (male) (T) and SU (male) (C) new-users in the OEHR data source. We report proportion of initiators satisfying selected base-line characteristics and the standardized difference of population proportions (StdDiff) before and after propensity score adjustment. Less extreme StdDiffs through matching and stratification suggest improved balance between patient cohorts through adjustment.

| | Before adjustment | | | After matching | | | After stratification | | |
|---|---|---|---|---|---|---|---|---|---|
| Characteristic | T (%) | C (%) | StdDiff | T (%) | C (%) | StdDiff | T (%) | C (%) | StdDiff |
| **Age group** | | | | | | | | | |
| 20 - 24 | 0.5 | 0.2 | 0.04 | 0.5 | 0.3 | 0.02 | 0.3 | 0.2 | 0.01 |
| 25 - 29 | 1.2 | 0.6 | 0.06 | 1.1 | 1.0 | 0.02 | 0.8 | 0.7 | 0.01 |
| 30 - 34 | 2.8 | 1.8 | 0.07 | 2.7 | 2.6 | 0.01 | 1.8 | 1.9 | 0.00 |
| 35 - 39 | 5.7 | 3.9 | 0.08 | 5.5 | 5.6 | 0.00 | 4.9 | 4.0 | 0.04 |
| 40 - 44 | 9.1 | 6.7 | 0.09 | 8.9 | 9.0 | 0.00 | 7.1 | 6.9 | 0.01 |
| 45 - 49 | 13.3 | 10.4 | 0.09 | 13.1 | 13.0 | 0.00 | 10.7 | 10.6 | 0.00 |
| 50 - 54 | 16.7 | 13.8 | 0.08 | 16.5 | 16.3 | 0.01 | 13.6 | 14.0 | -0.01 |
| 55 - 59 | 17.5 | 15.7 | 0.05 | 17.5 | 18.0 | -0.01 | 15.6 | 15.9 | -0.01 |
| 60 - 64 | 15.7 | 14.8 | 0.02 | 15.9 | 16.0 | 0.00 | 13.4 | 14.9 | -0.04 |
| 65 - 69 | 9.4 | 12.4 | -0.09 | 12.9 | 12.2 | 0.02 | 12.9 | 12.2 | 0.02 |
| 70 - 74 | 5.0 | 9.0 | -0.16 | 5.1 | 5.1 | 0.00 | 9.4 | 8.7 | 0.03 |
| 75 - 79 | 2.2 | 6.6 | -0.22 | 2.4 | 2.3 | 0.00 | 5.8 | 6.2 | -0.01 |
| 80 - 84 | 0.6 | 3.4 | -0.20 | 0.7 | 0.7 | -0.01 | 3.3 | 3.2 | 0.01 |
| 85 - 89 | 0.2 | 0.7 | -0.08 | 0.2 | 0.3 | -0.03 | 0.3 | 0.7 | -0.05 |
| **Race** | | | | | | | | | |
| Asian | 1.6 | 2.4 | -0.05 | 1.7 | 1.8 | -0.01 | 1.2 | 2.3 | -0.09 |
| Black or African American | 7.4 | 8.6 | -0.04 | 7.6 | 7.6 | 0.00 | 8.4 | 8.5 | 0.00 |
| White | 83.4 | 81.0 | 0.06 | 83.2 | 83.4 | -0.01 | 83.0 | 81.2 | 0.05 |
| **Ethnicity** | | | | | | | | | |
| Hispanic or Latino | 5.0 | 6.6 | -0.07 | 5.2 | 5.2 | 0.00 | 6.4 | 6.5 | -0.01 |
| Not Hispanic or Latino | 85.8 | 85.7 | 0.00 | 85.8 | 86.0 | -0.01 | 85.9 | 85.7 | 0.01 |
| **Medical history: General** | | | | | | | | | |
| Acute respiratory disease | 15.6 | 13.9 | 0.05 | 15.1 | 15.2 | 0.00 | 12.9 | 14.0 | -0.03 |
| Attention deficit hyperactivity disorder | 1.4 | 0.7 | 0.07 | 1.3 | 1.5 | -0.02 | 0.7 | 0.8 | -0.01 |
| Chronic liver disease | 2.3 | 2.7 | -0.03 | 2.4 | 2.1 | 0.02 | 2.5 | 2.7 | -0.01 |
| Chronic obstructive lung disease | 4.3 | 5.3 | -0.05 | 4.2 | 4.4 | -0.01 | 5.5 | 5.3 | 0.01 |
| Crohn’s disease | 0.2 | 0.2 | 0.00 | 0.2 | 0.2 | -0.01 | 0.1 | 0.2 | -0.03 |
| Dementia | 0.3 | 0.8 | -0.08 | 0.3 | 0.4 | -0.02 | 0.2 | 0.8 | -0.09 |
| Depressive disorder | 12.6 | 9.4 | 0.10 | 11.9 | 12.3 | -0.01 | 10.0 | 9.6 | 0.01 |
| Gastroesophageal reflux disease | 15.5 | 12.8 | 0.08 | 14.9 | 15.3 | -0.01 | 13.7 | 13.0 | 0.02 |
| Gastrointestinal hemorrhage | 1.4 | 1.5 | -0.01 | 1.4 | 1.3 | 0.00 | 1.5 | 1.5 | 0.00 |
| Human immunodeficiency virus infection | 0.4 | 0.3 | 0.03 | 0.4 | 0.4 | 0.00 | 0.2 | 0.3 | -0.01 |
| Hyperlipidemia | 67.8 | 65.6 | 0.05 | 66.8 | 67.2 | -0.01 | 67.3 | 65.8 | 0.03 |
| Hypertensive disorder | 68.4 | 65.4 | 0.06 | 67.5 | 68.1 | -0.01 | 68.8 | 65.6 | 0.07 |
| Lesion of liver | 0.7 | 0.9 | -0.02 | 0.8 | 0.9 | -0.01 | 0.6 | 0.9 | -0.04 |
| Obesity | 41.8 | 21.9 | 0.44 | 38.8 | 40.4 | -0.03 | 24.1 | 23.4 | 0.01 |
| Osteoarthritis | 15.5 | 14.0 | 0.04 | 14.9 | 15.2 | -0.01 | 13.9 | 14.1 | 0.00 |
| Pneumonia | 1.8 | 2.0 | -0.02 | 1.8 | 1.7 | 0.01 | 1.7 | 2.0 | -0.02 |
| Psoriasis | 1.7 | 1.3 | 0.04 | 1.7 | 1.6 | 0.00 | 1.7 | 1.3 | 0.03 |
| Renal impairment | 5.4 | 6.9 | -0.06 | 5.4 | 5.6 | -0.01 | 6.1 | 6.8 | -0.03 |
| Rheumatoid arthritis | 0.6 | 0.6 | 0.00 | 0.6 | 0.7 | -0.01 | 0.9 | 0.6 | 0.03 |
| Schizophrenia | 0.3 | 0.4 | -0.02 | 0.3 | 0.4 | -0.02 | 0.3 | 0.4 | -0.02 |
| Ulcerative colitis | 0.3 | 0.3 | 0.00 | 0.3 | 0.3 | 0.00 | 0.2 | 0.3 | -0.02 |
| Urinary tract infectious disease | 1.4 | 2.1 | -0.06 | 1.4 | 1.5 | -0.01 | 2.2 | 2.0 | 0.01 |
| Viral hepatitis C | 0.6 | 0.8 | -0.02 | 0.6 | 0.5 | 0.01 | 0.6 | 0.7 | -0.02 |
| Visual system disorder | 11.1 | 13.5 | -0.07 | 11.0 | 11.1 | 0.00 | 12.6 | 13.3 | -0.02 |
| **Medical history: Cardiovascular disease** | | | | | | | | | |
| Atrial fibrillation | 4.2 | 5.1 | -0.05 | 4.1 | 4.1 | 0.00 | 5.9 | 5.1 | 0.04 |
| Cerebrovascular disease | 2.1 | 2.8 | -0.05 | 2.0 | 2.0 | 0.00 | 2.3 | 2.8 | -0.03 |
| Coronary arteriosclerosis | 11.0 | 13.4 | -0.07 | 11.1 | 10.8 | 0.01 | 13.6 | 13.2 | 0.01 |
| Heart disease | 20.8 | 23.0 | -0.05 | 20.6 | 20.3 | 0.01 | 24.8 | 22.8 | 0.05 |
| Heart failure | 2.9 | 3.8 | -0.05 | 2.9 | 3.1 | -0.01 | 3.8 | 3.7 | 0.00 |
| Ischemic heart disease | 5.1 | 5.8 | -0.03 | 5.0 | 4.8 | 0.01 | 6.5 | 5.7 | 0.03 |
| Peripheral vascular disease | 5.1 | 4.7 | 0.02 | 4.9 | 5.1 | 0.00 | 4.3 | 4.7 | -0.02 |
| Pulmonary embolism | 0.5 | 0.6 | -0.01 | 0.5 | 0.5 | 0.00 | 1.0 | 0.6 | 0.04 |
| Venous thrombosis | 1.0 | 1.2 | -0.02 | 1.0 | 1.0 | 0.00 | 0.9 | 1.2 | -0.02 |
| **Medical history: Neoplasms** | | | | | | | | | |
| Hematologic neoplasm | 0.6 | 0.8 | -0.02 | 0.5 | 0.6 | -0.01 | 0.6 | 0.8 | -0.02 |

| (Continued) | Before ajustment | | | After matching | | | After stratification | | |
|---|---|---|---|---|---|---|---|---|---|
| Characteristic | T (%) | C (%) | StdDiff | T (%) | C (%) | StdDiff | T (%) | C (%) | StdDiff |
| Malignant lymphoma | 0.4 | 0.5 | -0.01 | 0.4 | 0.4 | -0.01 | 0.4 | 0.5 | -0.01 |
| Malignant neoplasm of anorectum | 0.1 | 0.1 | -0.01 | 0.1 | 0.1 | 0.00 | 0.2 | 0.1 | 0.02 |
| Malignant neoplastic disease | 4.8 | 6.7 | -0.08 | 4.9 | 4.8 | 0.00 | 7.5 | 6.5 | 0.04 |
| Malignant tumor of colon | 0.2 | 0.4 | -0.03 | 0.2 | 0.3 | -0.01 | 0.4 | 0.4 | 0.00 |
| Malignant tumor of lung | 0.1 | 0.2 | -0.04 | 0.1 | 0.2 | -0.02 | 0.1 | 0.2 | -0.04 |
| Malignant tumor of urinary bladder | 0.3 | 0.5 | -0.03 | 0.3 | 0.3 | 0.01 | 0.5 | 0.4 | 0.01 |
| Primary malignant neoplasm of prostate | 1.3 | 2.4 | -0.08 | 1.3 | 1.5 | -0.01 | 3.4 | 2.3 | 0.06 |
| **Medication use** | | | | | | | | | |
| Agents acting on the renin-angiotensin system | 69.3 | 66.7 | 0.06 | 69.1 | 69.2 | 0.00 | 69.9 | 66.9 | 0.06 |
| Antibacterials for systemic use | 38.4 | 36.2 | 0.05 | 37.6 | 38.0 | -0.01 | 37.1 | 36.3 | 0.02 |
| Antidepressants | 27.3 | 20.9 | 0.15 | 26.1 | 26.5 | -0.01 | 22.2 | 21.4 | 0.02 |
| Antiepileptics | 15.7 | 13.7 | 0.06 | 15.1 | 15.8 | -0.02 | 14.2 | 13.9 | 0.01 |
| Antiinflammatory and antirheumatic products | 54.5 | 55.1 | -0.01 | 54.2 | 54.1 | 0.00 | 56.2 | 55.0 | 0.02 |
| Antineoplastic agents | 3.9 | 3.1 | 0.04 | 3.7 | 3.7 | 0.00 | 3.8 | 3.2 | 0.04 |
| Antipsoriatics | 0.5 | 0.4 | 0.01 | 0.5 | 0.4 | 0.01 | 0.4 | 0.4 | 0.00 |
| Antithrombotic agents | 38.4 | 44.1 | -0.12 | 38.8 | 38.4 | 0.01 | 45.6 | 43.6 | 0.04 |
| Beta blocking agents | 30.0 | 33.4 | -0.07 | 30.1 | 29.8 | 0.01 | 34.1 | 33.1 | 0.02 |
| Calcium channel blockers | 23.0 | 21.4 | 0.04 | 22.9 | 23.1 | 0.00 | 23.7 | 21.5 | 0.05 |
| Diuretics | 37.1 | 35.1 | 0.04 | 36.5 | 36.4 | 0.00 | 38.5 | 35.2 | 0.07 |
| Drugs for acid related disorders | 31.3 | 30.0 | 0.03 | 30.6 | 31.0 | -0.01 | 32.1 | 30.1 | 0.04 |
| Drugs for obstructive airway diseases | 39.4 | 34.4 | 0.10 | 38.1 | 38.6 | -0.01 | 34.9 | 34.8 | 0.00 |
| Immunosuppressants | 2.5 | 1.6 | 0.06 | 2.4 | 2.3 | 0.01 | 1.9 | 1.7 | 0.02 |
| Lipid modifying agents | 72.1 | 71.0 | 0.02 | 71.8 | 71.6 | 0.00 | 73.9 | 71.0 | 0.06 |
| Opioids | 23.4 | 24.6 | -0.03 | 23.1 | 23.2 | 0.00 | 26.6 | 24.5 | 0.05 |
| Psycholeptics | 24.6 | 22.4 | 0.05 | 23.8 | 24.0 | -0.01 | 23.1 | 22.5 | 0.01 |
| Psychostimulants | 4.1 | 1.9 | 0.13 | 3.6 | 3.6 | 0.00 | 2.1 | 2.0 | 0.01 |

### 2.5.6 VA

**Supplementary Table 191:** Baseline patient characteristics for GLP1RA (male) (T) and SU (male) (C) new-users in the VA data source. We report proportion of initiators satisfying selected base-line characteristics and the standardized difference of population proportions (StdDiff) before and after propensity score adjustment. Less extreme StdDiffs through matching and stratification suggest improved balance between patient cohorts through adjustment.

| | Before adjustment | | | After matching | | | After stratification | | |
|---|---|---|---|---|---|---|---|---|---|
| Characteristic | T (%) | C (%) | StdDiff | T (%) | C (%) | StdDiff | T (%) | C (%) | StdDiff |
| **Age group** | | | | | | | | | |
| 25 - 29 | <0.7 | 0.1 | 0.01 | <0.8 | 0.3 | -0.03 | <0.1 | 0.1 | -0.04 |
| 30 - 34 | 0.8 | 0.5 | 0.04 | 0.8 | 0.8 | -0.01 | 0.3 | 0.6 | -0.04 |
| 35 - 39 | 1.8 | 1.3 | 0.04 | 1.7 | 2.2 | -0.04 | 0.2 | 1.4 | -0.13 |
| 40 - 44 | 4.1 | 2.8 | 0.07 | 4.1 | 4.1 | 0.00 | 3.8 | 3.0 | 0.05 |
| 45 - 49 | 7.1 | 5.4 | 0.07 | 6.8 | 7.0 | -0.01 | 9.0 | 5.6 | 0.13 |
| 50 - 54 | 10.6 | 8.9 | 0.06 | 10.4 | 10.1 | 0.01 | 8.8 | 8.7 | 0.00 |
| 55 - 59 | 12.0 | 14.1 | -0.06 | 11.9 | 11.2 | 0.02 | 5.5 | 12.6 | -0.25 |
| 60 - 64 | 12.4 | 19.6 | -0.20 | 12.8 | 12.0 | 0.03 | 39.7 | 20.8 | 0.42 |
| 65 - 69 | 15.2 | 19.2 | -0.11 | 15.9 | 14.3 | 0.05 | 12.7 | 20.6 | -0.21 |
| 70 - 74 | 22.6 | 13.6 | 0.24 | 22.5 | 22.4 | 0.00 | 14.6 | 13.5 | 0.03 |
| 75 - 79 | 8.8 | 7.8 | 0.03 | 8.4 | 8.8 | -0.01 | 3.4 | 6.8 | -0.15 |
| 80 - 84 | 3.0 | 4.5 | -0.08 | 3.1 | 3.9 | -0.04 | 1.7 | 4.0 | -0.14 |
| 85 - 89 | 1.1 | 1.8 | -0.05 | 1.1 | 1.8 | -0.05 | 0.2 | 1.8 | -0.17 |
| 90 - 94 | <0.7 | 0.4 | -0.02 | <0.8 | 0.8 | -0.07 | <0.1 | 0.4 | -0.08 |
| **Race** | | | | | | | | | |
| Asian | 1.4 | 0.8 | 0.06 | 1.4 | 1.2 | 0.03 | 1.1 | 0.9 | 0.02 |
| Black or African American | 12.2 | 14.2 | -0.06 | 12.2 | 12.1 | 0.00 | 18.0 | 15.2 | 0.07 |
| White | 77.3 | 74.4 | 0.07 | 77.3 | 77.5 | -0.01 | 74.7 | 75.1 | -0.01 |
| Unknown | 7.2 | 8.8 | -0.06 | 7.2 | 7.1 | 0.00 | 4.9 | 6.9 | -0.08 |
| Native Hawaiian or Other Pacific Islander | 1.4 | 1.0 | 0.04 | 1.0 | 1.0 | 0.00 | 1.0 | 1.0 | 0.00 |
| American Indian or Alaska Native | <0.7 | 0.8 | -0.03 | <0.8 | 0.8 | -0.03 | 0.3 | 0.8 | -0.08 |
| **Ethnicity** | | | | | | | | | |
| Hispanic or Latino | 6.2 | 6.1 | 0.00 | 6.3 | 7.2 | -0.03 | 2.6 | 6.6 | -0.19 |
| Not Hispanic or Latino | 90.5 | 89.1 | 0.05 | 90.4 | 90.1 | 0.01 | 95.2 | 90.4 | 0.18 |
| **Medical history: General** | | | | | | | | | |
| Acute respiratory disease | 6.9 | 8.8 | -0.07 | 7.0 | 7.2 | -0.01 | 5.7 | 8.9 | -0.13 |
| Attention deficit hyperactivity disorder | 1.3 | 0.4 | 0.10 | 1.1 | 0.9 | 0.02 | 1.0 | 0.5 | 0.06 |
| Chronic liver disease | 2.4 | 2.5 | -0.01 | 2.3 | 2.3 | 0.00 | 5.0 | 2.8 | 0.11 |
| Chronic obstructive lung disease | 11.2 | 11.1 | 0.00 | 10.9 | 12.0 | -0.03 | 7.1 | 11.2 | -0.14 |
| Crohn's disease | <0.7 | 0.2 | 0.03 | <0.8 | 0.2 | 0.03 | <0.1 | 0.2 | -0.04 |
| Dementia | 1.8 | 1.0 | 0.07 | 1.7 | 1.9 | -0.02 | 0.9 | 1.0 | -0.01 |
| Depressive disorder | 26.3 | 20.3 | 0.14 | 25.5 | 26.2 | -0.02 | 14.3 | 21.6 | -0.19 |
| Gastroesophageal reflux disease | 21.3 | 19.2 | 0.05 | 20.9 | 21.3 | -0.01 | 18.4 | 20.0 | -0.04 |
| Gastrointestinal hemorrhage | 1.1 | 1.4 | -0.03 | 1.0 | 1.5 | -0.04 | 1.1 | 1.4 | -0.02 |
| Human immunodeficiency virus infection | 0.7 | 0.3 | 0.06 | 0.8 | 0.4 | 0.04 | <0.1 | 0.3 | -0.05 |
| Hyperlipidemia | 71.1 | 73.9 | -0.06 | 70.7 | 70.8 | 0.00 | 80.4 | 75.5 | 0.12 |
| Hypertensive disorder | 75.0 | 77.8 | -0.06 | 74.3 | 73.6 | 0.02 | 85.8 | 77.8 | 0.21 |
| Lesion of liver | 2.2 | 1.1 | 0.08 | 2.1 | 2.2 | -0.01 | 1.5 | 1.3 | 0.02 |
| Obesity | 46.9 | 29.6 | 0.36 | 45.2 | 44.2 | 0.02 | 43.2 | 30.9 | 0.25 |
| Osteoarthritis | 24.9 | 22.2 | 0.07 | 24.8 | 23.7 | 0.02 | 17.0 | 22.5 | -0.14 |
| Pneumonia | 1.0 | 1.4 | -0.04 | 1.0 | 1.5 | -0.05 | <0.1 | 1.4 | -0.15 |
| Psoriasis | 2.0 | 1.6 | 0.03 | 1.9 | 1.9 | 0.00 | 0.6 | 1.6 | -0.10 |
| Renal impairment | 10.5 | 5.5 | 0.18 | 10.1 | 10.6 | -0.01 | 14.8 | 6.3 | 0.28 |
| Rheumatoid arthritis | 0.7 | 0.8 | -0.01 | <0.8 | 0.7 | -0.01 | <0.1 | 0.8 | -0.10 |
| Schizophrenia | <0.7 | 1.3 | -0.09 | <0.8 | 0.8 | -0.07 | <0.1 | 1.2 | -0.14 |
| Ulcerative colitis | <0.7 | 0.3 | -0.04 | <0.8 | 0.3 | -0.03 | <0.1 | 0.3 | -0.07 |
| Urinary tract infectious disease | 2.3 | 2.2 | 0.00 | 2.4 | 2.3 | 0.01 | 0.7 | 2.2 | -0.13 |
| Viral hepatitis C | 0.9 | 2.1 | -0.10 | 0.8 | 1.2 | -0.04 | 2.5 | 2.2 | 0.02 |
| Visual system disorder | 40.8 | 44.1 | -0.07 | 41.0 | 37.2 | 0.08 | 28.7 | 44.8 | -0.34 |
| **Medical history: Cardiovascular disease** | | | | | | | | | |
| Atrial fibrillation | 9.9 | 5.6 | 0.16 | 10.2 | 10.5 | -0.01 | 4.7 | 5.6 | -0.04 |
| Cerebrovascular disease | 3.1 | 4.5 | -0.07 | 3.2 | 3.1 | 0.01 | 2.5 | 4.2 | -0.10 |
| Coronary arteriosclerosis | 23.2 | 16.0 | 0.18 | 22.6 | 23.0 | -0.01 | 12.0 | 14.7 | -0.08 |
| Heart disease | 35.9 | 31.5 | 0.09 | 35.8 | 36.4 | -0.01 | 26.2 | 30.1 | -0.09 |
| Heart failure | 6.5 | 4.9 | 0.07 | 6.3 | 7.7 | -0.05 | 4.8 | 4.7 | 0.00 |
| Ischemic heart disease | 8.2 | 12.6 | -0.15 | 8.1 | 7.7 | 0.01 | 5.0 | 12.0 | -0.26 |
| Peripheral vascular disease | 5.5 | 5.1 | 0.02 | 5.5 | 5.7 | -0.01 | 1.6 | 4.8 | -0.18 |
| Pulmonary embolism | 1.1 | 0.5 | 0.07 | 1.1 | 0.8 | 0.03 | 0.3 | 0.5 | -0.03 |

| (Continued) | Before ajustment | | | After matching | | | After stratification | | |
|---|---|---|---|---|---|---|---|---|---|
| Characteristic | T (%) | C (%) | StdDiff | T (%) | C (%) | StdDiff | T (%) | C (%) | StdDiff |
| Venous thrombosis | 0.9 | 1.0 | 0.00 | 0.9 | 1.2 | -0.03 | <0.1 | 1.1 | -0.12 |
| **Medical history: Neoplasms** | | | | | | | | | |
| Hematologic neoplasm | 1.2 | 0.8 | 0.04 | 1.1 | 0.9 | 0.02 | 0.9 | 0.8 | 0.01 |
| Malignant lymphoma | 0.7 | 0.5 | 0.03 | 0.8 | 0.7 | 0.01 | 0.8 | 0.5 | 0.04 |
| Malignant neoplastic disease | 7.1 | 9.5 | -0.09 | 7.2 | 8.2 | -0.04 | 18.3 | 9.6 | 0.25 |
| Malignant tumor of colon | <0.7 | 0.7 | -0.06 | <0.8 | 0.4 | -0.02 | <0.1 | 0.6 | -0.10 |
| Malignant tumor of lung | <0.7 | 0.2 | -0.04 | <0.8 | 0.4 | -0.06 | <0.1 | 0.2 | -0.06 |
| Malignant tumor of urinary bladder | <0.7 | 0.6 | -0.03 | <0.8 | 0.6 | -0.03 | 0.2 | 0.7 | -0.06 |
| Primary malignant neoplasm of prostate | 2.6 | 4.1 | -0.08 | 2.7 | 3.1 | -0.03 | 3.1 | 4.0 | -0.05 |
| **Medication use** | | | | | | | | | |
| Agents acting on the renin-angiotensin system | 61.4 | 72.1 | -0.23 | 62.1 | 58.8 | 0.07 | 65.2 | 71.2 | -0.13 |
| Antibacterials for systemic use | 23.4 | 29.6 | -0.14 | 23.3 | 22.7 | 0.01 | 31.4 | 29.7 | 0.04 |
| Antidepressants | 41.2 | 33.2 | 0.17 | 41.1 | 41.0 | 0.00 | 30.4 | 34.2 | -0.08 |
| Antiepileptics | 29.9 | 20.1 | 0.23 | 29.3 | 29.9 | -0.01 | 22.2 | 22.4 | 0.00 |
| Antiinflammatory and antirheumatic products | 47.5 | 46.3 | 0.02 | 47.3 | 47.7 | -0.01 | 43.2 | 46.4 | -0.06 |
| Antineoplastic agents | 5.3 | 2.7 | 0.13 | 5.1 | 4.4 | 0.03 | 2.1 | 2.8 | -0.05 |
| Antipsoriatics | 0.9 | 0.7 | 0.02 | 0.8 | 0.8 | 0.00 | 0.3 | 0.7 | -0.06 |
| Antithrombotic agents | 36.1 | 33.4 | 0.06 | 36.1 | 37.3 | -0.02 | 29.2 | 32.8 | -0.08 |
| Beta blocking agents | 37.0 | 40.5 | -0.07 | 26.0 | 39.9 | -0.30 | 26.0 | 39.9 | -0.30 |
| Calcium channel blockers | 24.5 | 26.5 | -0.04 | 24.9 | 23.2 | 0.04 | 20.9 | 26.5 | -0.13 |
| Diuretics | 39.1 | 43.1 | -0.08 | 39.4 | 38.3 | 0.02 | 36.3 | 43.0 | -0.14 |
| Drugs for acid related disorders | 40.3 | 40.9 | -0.01 | 40.1 | 40.8 | -0.01 | 26.3 | 41.5 | -0.32 |
| Drugs for obstructive airway diseases | 39.1 | 33.2 | 0.12 | 38.6 | 39.1 | -0.01 | 27.1 | 34.3 | -0.16 |
| Immunosuppressants | 3.6 | 1.4 | 0.14 | 3.4 | 2.8 | 0.03 | 0.8 | 1.6 | -0.07 |
| Lipid modifying agents | 74.3 | 78.7 | -0.10 | 74.7 | 73.8 | 0.02 | 84.7 | 80.1 | 0.12 |
| Opioids | 16.0 | 20.5 | -0.12 | 16.2 | 15.3 | 0.03 | 18.3 | 21.1 | -0.07 |
| Psycholeptics | 23.7 | 21.3 | 0.06 | 23.5 | 24.1 | -0.01 | 17.3 | 22.3 | -0.12 |
| Psychostimulants | 3.0 | 1.4 | 0.11 | 2.9 | 2.0 | 0.06 | 1.4 | 1.5 | 0.00 |

#### 2.6 DPP4I (male) vs SU (male)

##### 2.6.1 CCAE

**Supplementary Table 192:** Baseline patient characteristics for DPP4I (male) (T) and SU (male) (C) new-users in the CCAE data source. We report proportion of initiators satisfying selected base-line characteristics and the standardized difference of population proportions (StdDiff) before and after propensity score adjustment. Less extreme StdDiffs through matching and stratification suggest improved balance between patient cohorts through adjustment.

| | Before adjustment | | | After matching | | | After stratification | | |
|---|---|---|---|---|---|---|---|---|---|
| Characteristic | T (%) | C (%) | StdDiff | T (%) | C (%) | StdDiff | T (%) | C (%) | StdDiff |
| **Age group** | | | | | | | | | |
| 20 - 24 | 0.2 | 0.2 | 0.00 | 0.2 | 0.2 | 0.01 | 0.2 | 0.2 | 0.01 |
| 25 - 29 | 0.5 | 0.6 | -0.01 | 0.5 | 0.5 | -0.01 | 0.5 | 0.5 | -0.01 |
| 30 - 34 | 1.8 | 2.0 | -0.01 | 1.9 | 1.8 | 0.01 | 1.9 | 1.9 | 0.00 |
| 35 - 39 | 4.4 | 4.6 | -0.01 | 4.5 | 4.5 | 0.00 | 4.7 | 4.6 | 0.00 |
| 40 - 44 | 9.0 | 8.9 | 0.00 | 9.1 | 9.0 | 0.00 | 9.1 | 9.0 | 0.00 |
| 45 - 49 | 14.5 | 14.2 | 0.01 | 14.7 | 14.5 | 0.00 | 14.4 | 14.4 | 0.00 |
| 50 - 54 | 20.2 | 19.9 | 0.01 | 20.0 | 20.1 | 0.00 | 20.0 | 20.0 | 0.00 |
| 55 - 59 | 23.5 | 23.4 | 0.00 | 23.4 | 23.4 | 0.00 | 23.4 | 23.3 | 0.00 |
| 60 - 64 | 23.7 | 23.9 | -0.01 | 23.5 | 23.8 | -0.01 | 23.7 | 23.9 | 0.00 |
| 65 - 69 | 2.2 | 2.3 | -0.01 | 2.1 | 2.2 | 0.00 | 2.2 | 2.2 | 0.00 |
| **Medical history: General** | | | | | | | | | |
| Acute respiratory disease | 22.4 | 20.5 | 0.04 | 21.7 | 21.9 | -0.01 | 21.8 | 21.5 | 0.01 |
| Attention deficit hyperactivity disorder | 0.8 | 0.6 | 0.02 | 0.8 | 0.7 | 0.00 | 0.7 | 0.7 | 0.00 |
| Chronic liver disease | 2.7 | 2.7 | 0.00 | 2.7 | 2.7 | 0.00 | 2.8 | 2.8 | 0.00 |
| Chronic obstructive lung disease | 2.6 | 2.7 | -0.01 | 2.8 | 2.7 | 0.01 | 2.8 | 2.7 | 0.01 |
| Crohn's disease | 0.2 | 0.2 | 0.00 | 0.2 | 0.2 | -0.01 | 0.2 | 0.2 | 0.00 |
| Dementia | 0.1 | 0.1 | 0.00 | 0.1 | 0.1 | 0.00 | 0.1 | 0.1 | 0.00 |
| Depressive disorder | 5.9 | 6.0 | -0.01 | 5.9 | 5.9 | 0.00 | 6.3 | 6.1 | 0.01 |
| Gastroesophageal reflux disease | 8.8 | 7.5 | 0.05 | 8.5 | 8.7 | -0.01 | 8.5 | 8.4 | 0.00 |
| Gastrointestinal hemorrhage | 1.8 | 1.9 | -0.01 | 1.8 | 1.8 | 0.00 | 1.8 | 1.8 | 0.00 |
| Human immunodeficiency virus infection | 0.4 | 0.3 | 0.01 | 0.4 | 0.4 | 0.00 | 0.4 | 0.4 | 0.00 |
| Hyperlipidemia | 70.1 | 62.3 | 0.17 | 68.6 | 69.3 | -0.01 | 66.9 | 67.0 | 0.00 |
| Hypertensive disorder | 66.3 | 61.6 | 0.10 | 65.5 | 65.8 | -0.01 | 65.2 | 64.8 | 0.01 |
| Lesion of liver | 0.6 | 0.8 | -0.03 | 0.6 | 0.6 | 0.00 | 0.7 | 0.7 | 0.00 |
| Obesity | 14.8 | 13.7 | 0.03 | 14.7 | 14.8 | 0.00 | 14.8 | 15.0 | 0.00 |
| Osteoarthritis | 15.1 | 13.9 | 0.04 | 14.7 | 14.9 | 0.00 | 15.0 | 14.6 | 0.01 |
| Pneumonia | 1.8 | 2.2 | -0.02 | 1.9 | 1.8 | 0.00 | 2.1 | 2.0 | 0.00 |
| Psoriasis | 1.5 | 1.2 | 0.02 | 1.4 | 1.4 | 0.00 | 1.4 | 1.4 | 0.00 |
| Renal impairment | 3.4 | 3.7 | -0.02 | 3.4 | 3.5 | 0.00 | 4.0 | 3.8 | 0.01 |
| Rheumatoid arthritis | 0.6 | 0.5 | 0.00 | 0.6 | 0.5 | 0.00 | 0.6 | 0.5 | 0.01 |
| Schizophrenia | 0.1 | 0.1 | 0.00 | 0.1 | 0.1 | 0.00 | 0.1 | 0.1 | 0.01 |
| Ulcerative colitis | 0.3 | 0.3 | 0.00 | 0.3 | 0.3 | 0.00 | 0.3 | 0.3 | 0.00 |
| Urinary tract infectious disease | 3.0 | 2.9 | 0.01 | 2.8 | 2.9 | 0.00 | 2.9 | 2.9 | 0.00 |
| Viral hepatitis C | 0.4 | 0.5 | -0.02 | 0.4 | 0.4 | 0.00 | 0.5 | 0.5 | 0.00 |
| Visual system disorder | 21.8 | 20.7 | 0.03 | 20.9 | 21.2 | -0.01 | 20.9 | 21.2 | -0.01 |
| **Medical history: Cardiovascular disease** | | | | | | | | | |
| Atrial fibrillation | 2.4 | 2.4 | 0.00 | 2.3 | 2.3 | 0.00 | 2.5 | 2.4 | 0.01 |
| Cerebrovascular disease | 2.5 | 2.5 | 0.00 | 2.4 | 2.4 | 0.00 | 2.6 | 2.5 | 0.01 |
| Coronary arteriosclerosis | 10.1 | 9.7 | 0.01 | 9.7 | 9.8 | 0.00 | 10.0 | 9.7 | 0.01 |
| Heart disease | 18.9 | 18.1 | 0.02 | 18.2 | 18.3 | 0.00 | 18.7 | 18.2 | 0.01 |
| Heart failure | 2.0 | 2.3 | -0.02 | 2.0 | 2.0 | 0.00 | 2.3 | 2.2 | 0.01 |
| Ischemic heart disease | 4.9 | 5.2 | -0.01 | 4.7 | 4.8 | 0.00 | 5.1 | 4.9 | 0.01 |
| Peripheral vascular disease | 3.3 | 3.0 | 0.02 | 3.2 | 3.2 | 0.00 | 3.3 | 3.1 | 0.01 |
| Pulmonary embolism | 0.4 | 0.4 | 0.00 | 0.4 | 0.4 | 0.00 | 0.5 | 0.4 | 0.00 |
| Venous thrombosis | 1.1 | 1.0 | 0.01 | 1.1 | 1.0 | 0.01 | 1.1 | 1.1 | 0.00 |
| **Medical history: Neoplasms** | | | | | | | | | |
| Hematologic neoplasm | 0.8 | 0.7 | 0.01 | 0.8 | 0.7 | 0.01 | 0.8 | 0.8 | 0.01 |
| Malignant lymphoma | 0.4 | 0.4 | -0.01 | 0.4 | 0.4 | 0.00 | 0.3 | 0.4 | -0.01 |
| Malignant neoplasm of anorectum | 0.1 | 0.2 | -0.01 | 0.2 | 0.1 | 0.00 | 0.2 | 0.2 | 0.00 |
| Malignant neoplastic disease | 5.3 | 5.0 | 0.01 | 5.1 | 5.1 | 0.00 | 5.1 | 5.1 | 0.00 |
| Malignant tumor of colon | 0.3 | 0.4 | -0.01 | 0.3 | 0.3 | 0.01 | 0.4 | 0.3 | 0.01 |
| Malignant tumor of lung | 0.1 | 0.2 | -0.02 | 0.2 | 0.2 | 0.00 | 0.2 | 0.2 | 0.00 |
| Malignant tumor of urinary bladder | 0.3 | 0.3 | 0.01 | 0.3 | 0.3 | 0.01 | 0.3 | 0.3 | 0.01 |
| Primary malignant neoplasm of prostate | 1.4 | 1.3 | 0.01 | 1.4 | 1.4 | -0.01 | 1.3 | 1.4 | 0.00 |
| **Medication use** | | | | | | | | | |
| Agents acting on the renin-angiotensin system | 70.0 | 68.2 | 0.04 | 69.4 | 69.5 | 0.00 | 69.7 | 69.6 | 0.00 |

| (Continued) | Before ajustment | | | After matching | | | After stratification | | |
|---|---|---|---|---|---|---|---|---|---|
| Characteristic | T (%) | C (%) | StdDiff | T (%) | C (%) | StdDiff | T (%) | C (%) | StdDiff |
| Antibacterials for systemic use | 51.7 | 50.3 | 0.03 | 50.9 | 51.0 | 0.00 | 51.3 | 50.8 | 0.01 |
| Antidepressants | 17.3 | 17.4 | 0.00 | 17.5 | 17.3 | 0.00 | 17.5 | 17.3 | 0.00 |
| Antiepileptics | 8.5 | 8.7 | -0.01 | 8.5 | 8.4 | 0.00 | 9.1 | 8.8 | 0.01 |
| Antiinflammatory and antirheumatic products | 27.4 | 27.2 | 0.00 | 27.6 | 27.5 | 0.00 | 27.6 | 27.5 | 0.00 |
| Antineoplastic agents | 3.4 | 3.2 | 0.01 | 3.2 | 3.1 | 0.01 | 3.2 | 3.1 | 0.01 |
| Antipsoriatics | 0.5 | 0.5 | 0.01 | 0.5 | 0.5 | 0.00 | 0.5 | 0.5 | 0.01 |
| Antithrombotic agents | 10.5 | 10.7 | -0.01 | 10.4 | 10.5 | 0.00 | 11.0 | 10.8 | 0.01 |
| Beta blocking agents | 25.3 | 26.0 | -0.02 | 26.1 | 25.8 | 0.01 | 26.1 | 25.8 | 0.01 |
| Calcium channel blockers | 21.3 | 20.4 | 0.02 | 20.9 | 21.0 | 0.00 | 21.2 | 20.9 | 0.01 |
| Diuretics | 31.9 | 32.9 | -0.02 | 31.8 | 31.6 | 0.00 | 33.0 | 32.6 | 0.01 |
| Drugs for acid related disorders | 19.9 | 19.0 | 0.02 | 19.3 | 19.4 | 0.00 | 19.5 | 19.3 | 0.00 |
| Drugs for obstructive airway diseases | 30.0 | 27.8 | 0.05 | 29.2 | 29.3 | 0.00 | 29.2 | 29.0 | 0.00 |
| Immunosuppressants | 1.7 | 1.4 | 0.02 | 1.6 | 1.6 | 0.00 | 1.6 | 1.5 | 0.01 |
| Lipid modifying agents | 71.7 | 68.3 | 0.07 | 70.4 | 70.6 | 0.00 | 70.1 | 70.4 | -0.01 |
| Opioids | 21.9 | 22.4 | -0.01 | 21.8 | 21.7 | 0.00 | 22.9 | 22.4 | 0.01 |
| Psycholeptics | 17.7 | 16.9 | 0.02 | 17.3 | 17.3 | 0.00 | 17.7 | 17.4 | 0.01 |
| Psychostimulants | 2.2 | 1.9 | 0.02 | 2.0 | 2.0 | 0.00 | 2.0 | 2.0 | 0.00 |

##### 2.6.2 DAG

**Supplementary Table 193:** Baseline patient characteristics for DPP4I (male) (T) and SU (male) (C) new-users in the DAG data source. We report proportion of initiators satisfying selected base-line characteristics and the standardized difference of population proportions (StdDiff) before and after propensity score adjustment. Less extreme StdDiffs through matching and stratification suggest improved balance between patient cohorts through adjustment.

| | Before adjustment | | | After matching | | | After stratification | | |
|---|---|---|---|---|---|---|---|---|---|
| Characteristic | T (%) | C (%) | StdDiff | T (%) | C (%) | StdDiff | T (%) | C (%) | StdDiff |
| **Age group** | | | | | | | | | |
| 30 - 34 | 0.4 | 0.4 | 0.01 | 0.4 | 0.5 | -0.02 | 0.4 | 0.6 | -0.03 |
| 35 - 39 | 1.3 | 1.2 | 0.01 | 1.2 | 1.3 | -0.01 | 1.3 | 1.4 | -0.01 |
| 40 - 44 | 2.9 | 2.2 | 0.04 | 3.2 | 2.3 | 0.05 | 2.9 | 2.8 | 0.01 |
| 45 - 49 | 5.9 | 5.3 | 0.03 | 5.8 | 5.3 | 0.03 | 5.8 | 5.2 | 0.03 |
| 50 - 54 | 10.4 | 8.8 | 0.05 | 10.2 | 9.6 | 0.02 | 10.3 | 10.4 | 0.00 |
| 55 - 59 | 14.9 | 11.9 | 0.09 | 12.2 | 12.2 | 0.00 | 14.4 | 13.6 | 0.02 |
| 60 - 64 | 15.9 | 15.5 | 0.01 | 15.8 | 14.9 | 0.03 | 15.7 | 15.0 | 0.02 |
| 65 - 69 | 14.2 | 16.2 | -0.06 | 13.6 | 13.9 | -0.01 | 14.1 | 14.6 | -0.01 |
| 70 - 74 | 12.9 | 16.7 | -0.11 | 15.5 | 16.2 | -0.02 | 15.5 | 16.2 | -0.02 |
| 75 - 79 | 10.8 | 12.7 | -0.06 | 11.8 | 13.2 | -0.04 | 11.2 | 11.0 | 0.01 |
| 80 - 84 | 6.9 | 6.8 | 0.00 | 7.8 | 7.8 | 0.00 | 7.0 | 7.4 | -0.02 |
| 85 - 89 | 2.6 | 1.8 | 0.06 | 1.7 | 2.4 | -0.05 | 2.5 | 3.0 | -0.03 |
| 90 - 94 | 0.6 | 0.3 | 0.04 | 0.5 | 0.5 | 0.01 | 0.5 | 1.5 | -0.09 |
| **Medical history: General** | | | | | | | | | |
| Acute respiratory disease | 17.7 | 17.2 | 0.01 | 17.2 | 17.7 | -0.01 | 17.8 | 16.9 | 0.02 |
| Attention deficit hyperactivity disorder | 0.1 | 0.4 | -0.05 | <0.2 | 0.5 | -0.05 | 0.1 | 0.4 | -0.06 |
| Chronic liver disease | 0.5 | 0.6 | -0.01 | 0.3 | 0.6 | -0.04 | 0.5 | 0.6 | -0.01 |
| Chronic obstructive lung disease | 6.3 | 6.7 | -0.02 | 6.3 | 6.9 | -0.02 | 6.3 | 6.0 | 0.01 |
| Crohn’s disease | 0.2 | 0.1 | 0.01 | <0.2 | 0.2 | -0.01 | 0.2 | 0.2 | 0.00 |
| Dementia | 1.8 | 2.1 | -0.03 | 1.5 | 2.0 | -0.03 | 1.7 | 2.3 | -0.04 |
| Depressive disorder | 6.7 | 7.2 | -0.02 | 7.7 | 8.0 | -0.01 | 6.8 | 7.0 | -0.01 |
| Gastroesophageal reflux disease | 2.1 | 2.2 | -0.01 | 2.0 | 2.2 | -0.02 | 2.1 | 2.1 | 0.00 |
| Gastrointestinal hemorrhage | 0.8 | 1.0 | -0.03 | 0.6 | 0.9 | -0.03 | 0.7 | 0.8 | 0.00 |
| Hyperlipidemia | 24.8 | 29.2 | -0.10 | 28.7 | 28.1 | 0.01 | 25.6 | 26.5 | -0.02 |
| Hypertensive disorder | 47.1 | 55.3 | -0.16 | 53.2 | 51.9 | 0.03 | 48.5 | 46.6 | 0.04 |
| Lesion of liver | 0.9 | 1.1 | -0.02 | 0.5 | 1.1 | -0.06 | 0.9 | 1.1 | -0.03 |
| Obesity | 7.9 | 7.1 | 0.03 | 7.7 | 7.1 | 0.02 | 7.8 | 7.4 | 0.02 |
| Osteoarthritis | 10.2 | 12.9 | -0.09 | 10.4 | 10.9 | -0.02 | 10.4 | 10.9 | -0.02 |
| Pneumonia | 2.4 | 2.0 | 0.03 | 2.4 | 2.0 | 0.03 | 2.4 | 1.9 | 0.03 |
| Psoriasis | 1.3 | 1.3 | 0.00 | 1.4 | 1.4 | 0.00 | 1.3 | 1.1 | 0.02 |
| Renal impairment | 5.9 | 5.1 | 0.04 | 5.0 | 5.5 | -0.02 | 5.8 | 5.5 | 0.01 |
| Rheumatoid arthritis | 0.9 | 0.8 | 0.01 | 0.8 | 0.8 | 0.00 | 0.9 | 0.8 | 0.02 |
| Schizophrenia | 0.3 | 0.3 | 0.00 | 0.4 | 0.3 | 0.01 | 0.3 | 0.4 | -0.02 |
| Ulcerative colitis | 0.2 | 0.2 | -0.01 | <0.2 | 0.3 | -0.02 | 0.2 | 0.5 | -0.06 |
| Urinary tract infectious disease | 2.8 | 2.5 | 0.02 | 2.4 | 2.6 | -0.01 | 2.8 | 2.8 | 0.00 |
| Viral hepatitis C | 0.1 | <0.1 | 0.01 | 0.3 | <0.2 | 0.04 | 0.1 | <0.1 | 0.02 |
| Visual system disorder | 8.2 | 9.7 | -0.05 | 9.3 | 9.9 | -0.02 | 8.5 | 9.4 | -0.03 |
| **Medical history: Cardiovascular disease** | | | | | | | | | |
| Atrial fibrillation | 2.9 | 2.0 | 0.06 | 2.7 | 2.2 | 0.03 | 2.9 | 2.5 | 0.02 |
| Cerebrovascular disease | 4.5 | 5.5 | -0.05 | 4.6 | 4.9 | -0.01 | 4.5 | 5.1 | -0.03 |
| Coronary arteriosclerosis | 5.6 | 4.2 | 0.06 | 4.0 | 4.8 | -0.04 | 5.4 | 5.2 | 0.01 |
| Heart disease | 26.8 | 30.1 | -0.07 | 27.5 | 27.4 | 0.00 | 27.1 | 25.5 | 0.04 |
| Heart failure | 6.4 | 7.5 | -0.04 | 6.3 | 6.8 | -0.02 | 6.5 | 6.2 | 0.01 |
| Ischemic heart disease | 12.9 | 16.8 | -0.11 | 14.1 | 13.9 | 0.00 | 13.2 | 12.3 | 0.03 |
| Peripheral vascular disease | 6.0 | 6.5 | -0.02 | 6.1 | 6.2 | -0.01 | 6.0 | 5.9 | 0.01 |
| Pulmonary embolism | 0.6 | 0.2 | 0.07 | 0.4 | 0.3 | 0.02 | 0.6 | 0.3 | 0.04 |
| Venous thrombosis | 1.3 | 1.0 | 0.03 | 0.9 | 0.7 | 0.02 | 1.3 | 1.1 | 0.02 |
| **Medical history: Neoplasms** | | | | | | | | | |
| Hematologic neoplasm | 0.4 | 0.3 | 0.02 | 0.5 | 0.3 | 0.02 | 0.4 | 0.2 | 0.03 |
| Malignant lymphoma | 0.3 | 0.4 | -0.02 | 0.4 | 0.4 | -0.01 | 0.3 | 0.3 | 0.01 |
| Malignant neoplasm of anorectum | 0.2 | 0.2 | 0.01 | <0.2 | <0.2 | 0.02 | 0.2 | <0.1 | 0.04 |
| Malignant neoplastic disease | 5.4 | 6.5 | -0.05 | 6.1 | 6.3 | -0.01 | 5.5 | 6.0 | -0.02 |
| Malignant tumor of colon | 0.4 | 0.5 | -0.02 | 0.4 | 0.4 | 0.00 | 0.4 | 0.4 | -0.01 |
| Malignant tumor of lung | 0.2 | 0.2 | 0.00 | <0.2 | <0.2 | 0.00 | 0.2 | 0.1 | 0.01 |
| Malignant tumor of urinary bladder | 0.3 | 0.4 | -0.01 | 0.4 | 0.3 | 0.02 | 0.3 | 0.2 | 0.02 |
| Primary malignant neoplasm of prostate | 1.3 | 1.9 | -0.05 | 1.7 | 1.8 | 0.00 | 1.4 | 1.9 | -0.04 |
| **Medication use** | | | | | | | | | |
| Agents acting on the renin-angiotensin system | 68.1 | 65.8 | 0.05 | 67.5 | 66.6 | 0.02 | 68.0 | 68.9 | -0.02 |

| (Continued) | Before ajustment | | | After matching | | | After stratification | | |
|---|---|---|---|---|---|---|---|---|---|
| Characteristic | T (%) | C (%) | StdDiff | T (%) | C (%) | StdDiff | T (%) | C (%) | StdDiff |
| Antibacterials for systemic use | 23.7 | 25.4 | -0.04 | 23.8 | 25.4 | -0.04 | 23.8 | 25.4 | -0.04 |
| Antidepressants | 7.5 | 7.1 | 0.02 | 7.7 | 7.6 | 0.00 | 7.4 | 7.2 | 0.01 |
| Antiepileptics | 4.3 | 3.5 | 0.04 | 3.4 | 3.7 | -0.01 | 4.2 | 3.7 | 0.03 |
| Antiinflammatory and antirheumatic products | 43.9 | 47.9 | -0.08 | 46.6 | 46.1 | 0.01 | 44.5 | 43.1 | 0.03 |
| Antineoplastic agents | 1.0 | 0.8 | 0.02 | 1.0 | 0.8 | 0.01 | 1.0 | 1.2 | -0.02 |
| Antipsoriatics | 0.4 | 0.4 | 0.01 | 0.3 | 0.3 | 0.00 | 0.4 | 0.3 | 0.01 |
| Antithrombotic agents | 34.3 | 34.3 | 0.00 | 32.4 | 32.7 | -0.01 | 34.1 | 33.0 | 0.02 |
| Beta blocking agents | 44.6 | 45.1 | -0.01 | 45.5 | 45.0 | 0.01 | 44.7 | 43.6 | 0.02 |
| Calcium channel blockers | 31.9 | 29.7 | 0.05 | 30.9 | 30.5 | 0.01 | 31.7 | 32.0 | -0.01 |
| Diuretics | 45.6 | 47.7 | -0.04 | 47.6 | 46.8 | 0.01 | 46.1 | 45.9 | 0.00 |
| Drugs for acid related disorders | 31.9 | 28.4 | 0.08 | 28.9 | 29.6 | -0.01 | 31.5 | 31.4 | 0.00 |
| Drugs for obstructive airway diseases | 16.3 | 16.1 | 0.00 | 14.9 | 15.9 | -0.03 | 16.1 | 16.1 | 0.00 |
| Immunosuppressants | 0.6 | 0.3 | 0.04 | 0.6 | 0.4 | 0.03 | 0.6 | 0.4 | 0.02 |
| Lipid modifying agents | 48.1 | 42.2 | 0.12 | 43.0 | 44.0 | -0.02 | 47.4 | 48.2 | -0.02 |
| Opioids | 10.6 | 11.9 | -0.04 | 10.8 | 11.3 | -0.02 | 10.7 | 10.6 | 0.00 |
| Psycholeptics | 7.0 | 7.6 | -0.02 | 6.8 | 6.8 | 0.00 | 7.0 | 7.1 | 0.00 |
| Psychostimulants | 0.2 | 0.6 | -0.06 | <0.2 | 0.3 | -0.01 | 0.2 | 0.2 | 0.00 |

##### 2.6.3 ODOD

**Supplementary Table 194:** Baseline patient characteristics for DPP4I (male) (T) and SU (male) (C) new-users in the ODOD data source. We report proportion of initiators satisfying selected base-line characteristics and the standardized difference of population proportions (StdDiff) before and after propensity score adjustment. Less extreme StdDiffs through matching and stratification suggest improved balance between patient cohorts through adjustment.

| | Before adjustment | | | After matching | | | After stratification | | |
|---|---|---|---|---|---|---|---|---|---|
| Characteristic | T (%) | C (%) | StdDiff | T (%) | C (%) | StdDiff | T (%) | C (%) | StdDiff |
| **Age group** | | | | | | | | | |
| 20 - 24 | 0.1 | 0.1 | 0.00 | 0.1 | 0.1 | 0.01 | 0.1 | 0.1 | 0.00 |
| 25 - 29 | 0.3 | 0.3 | 0.00 | 0.3 | 0.3 | 0.01 | 0.4 | 0.3 | 0.01 |
| 30 - 34 | 1.2 | 1.1 | 0.01 | 1.2 | 1.1 | 0.01 | 1.2 | 1.1 | 0.01 |
| 35 - 39 | 3.2 | 2.7 | 0.03 | 3.1 | 3.1 | 0.00 | 2.8 | 2.8 | 0.00 |
| 40 - 44 | 6.0 | 5.0 | 0.05 | 5.8 | 5.9 | 0.00 | 5.2 | 5.3 | 0.00 |
| 45 - 49 | 8.8 | 7.9 | 0.03 | 8.7 | 8.6 | 0.00 | 8.1 | 8.1 | 0.00 |
| 50 - 54 | 11.9 | 10.4 | 0.05 | 11.7 | 11.7 | 0.00 | 10.6 | 10.9 | -0.01 |
| 55 - 59 | 14.0 | 12.5 | 0.04 | 13.8 | 14.0 | -0.01 | 12.9 | 13.0 | 0.00 |
| 60 - 64 | 13.2 | 12.7 | 0.01 | 13.1 | 12.9 | 0.00 | 13.1 | 12.9 | 0.00 |
| 65 - 69 | 13.9 | 15.8 | -0.05 | 14.2 | 14.8 | -0.02 | 14.9 | 15.5 | -0.01 |
| 70 - 74 | 12.5 | 14.8 | -0.07 | 12.8 | 12.7 | 0.01 | 14.4 | 14.1 | 0.01 |
| 75 - 79 | 8.1 | 9.5 | -0.05 | 9.2 | 9.1 | 0.00 | 9.2 | 9.1 | 0.00 |
| 80 - 84 | 4.5 | 4.9 | -0.02 | 4.9 | 4.8 | 0.00 | 4.9 | 4.8 | 0.00 |
| 85 - 89 | 2.0 | 2.0 | 0.00 | 2.0 | 1.9 | 0.01 | 2.2 | 2.0 | 0.01 |
| 90 - 94 | 0.2 | 0.1 | 0.01 | 0.2 | 0.1 | 0.01 | 0.2 | 0.1 | 0.02 |
| **Medical history: General** | | | | | | | | | |
| Acute respiratory disease | 23.2 | 21.7 | 0.03 | 22.7 | 23.0 | -0.01 | 22.7 | 23.0 | -0.01 |
| Attention deficit hyperactivity disorder | 0.8 | 0.6 | 0.04 | 0.6 | 0.6 | 0.00 | 0.6 | 0.6 | 0.00 |
| Chronic liver disease | 3.3 | 3.0 | 0.02 | 3.2 | 3.1 | 0.01 | 3.2 | 3.1 | 0.01 |
| Chronic obstructive lung disease | 8.2 | 9.6 | -0.05 | 8.3 | 8.3 | 0.00 | 9.3 | 9.2 | 0.00 |
| Crohn's disease | 0.3 | 0.2 | 0.01 | 0.3 | 0.2 | 0.01 | 0.3 | 0.2 | 0.01 |
| Dementia | 1.8 | 1.8 | 0.00 | 1.8 | 1.7 | 0.01 | 2.0 | 1.8 | 0.01 |
| Depressive disorder | 9.1 | 9.6 | -0.02 | 9.0 | 9.1 | 0.00 | 9.5 | 9.5 | 0.00 |
| Gastroesophageal reflux disease | 14.4 | 14.1 | 0.01 | 14.3 | 14.4 | 0.00 | 14.5 | 14.3 | 0.00 |
| Human immunodeficiency virus infection | 0.5 | 0.3 | 0.03 | 0.5 | 0.5 | 0.00 | 0.5 | 0.5 | 0.00 |
| Hyperlipidemia | 82.3 | 79.2 | 0.08 | 81.7 | 82.3 | -0.02 | 79.8 | 80.2 | -0.01 |
| Hypertensive disorder | 78.6 | 78.4 | 0.00 | 78.3 | 78.4 | 0.00 | 78.9 | 78.5 | 0.01 |
| Lesion of liver | 1.1 | 1.4 | -0.03 | 1.1 | 1.2 | 0.00 | 1.3 | 1.3 | -0.01 |
| Obesity | 19.5 | 20.0 | -0.01 | 19.4 | 19.7 | -0.01 | 20.1 | 19.9 | 0.00 |
| Osteoarthritis | 22.1 | 22.6 | -0.01 | 22.0 | 22.1 | 0.00 | 22.6 | 22.5 | 0.00 |
| Pneumonia | 3.6 | 4.2 | -0.03 | 3.6 | 3.7 | 0.00 | 4.0 | 4.0 | 0.00 |
| Psoriasis | 1.6 | 1.5 | 0.01 | 1.5 | 1.7 | -0.01 | 1.5 | 1.6 | -0.01 |
| Renal impairment | 13.5 | 14.7 | -0.03 | 14.8 | 14.4 | 0.01 | 14.8 | 14.4 | 0.01 |
| Rheumatoid arthritis | 1.0 | 1.1 | -0.01 | 1.0 | 1.0 | -0.01 | 1.0 | 1.1 | 0.00 |
| Schizophrenia | 0.4 | 0.4 | 0.00 | 0.4 | 0.3 | 0.01 | 0.5 | 0.4 | 0.02 |
| Ulcerative colitis | 0.4 | 0.3 | 0.01 | 0.4 | 0.3 | 0.01 | 0.4 | 0.3 | 0.01 |
| Urinary tract infectious disease | 5.9 | 5.8 | 0.00 | 5.8 | 6.0 | -0.01 | 5.8 | 6.0 | -0.01 |
| Viral hepatitis C | 0.7 | 0.8 | -0.02 | 0.7 | 0.7 | -0.01 | 0.8 | 0.8 | 0.00 |
| Visual system disorder | 35.8 | 36.0 | 0.00 | 35.8 | 35.9 | 0.00 | 35.8 | 35.9 | 0.00 |
| **Medical history: Cardiovascular disease** | | | | | | | | | |
| Atrial fibrillation | 7.3 | 7.6 | -0.01 | 7.3 | 7.2 | 0.00 | 7.7 | 7.5 | 0.01 |
| Cerebrovascular disease | 6.5 | 6.7 | -0.01 | 6.4 | 6.5 | -0.01 | 6.6 | 6.7 | 0.00 |
| Coronary arteriosclerosis | 18.9 | 19.8 | -0.02 | 18.8 | 18.9 | 0.00 | 18.8 | 18.9 | 0.00 |
| Heart disease | 32.1 | 33.0 | -0.02 | 31.9 | 32.0 | 0.00 | 32.8 | 32.8 | 0.00 |
| Heart failure | 6.7 | 7.7 | -0.04 | 6.9 | 6.9 | 0.00 | 6.9 | 6.9 | 0.00 |
| Ischemic heart disease | 9.8 | 10.6 | -0.02 | 9.8 | 9.9 | 0.00 | 10.4 | 10.4 | 0.00 |
| Peripheral vascular disease | 10.4 | 10.6 | -0.01 | 10.7 | 10.6 | 0.00 | 10.7 | 10.6 | 0.00 |
| Pulmonary embolism | 0.7 | 0.8 | -0.01 | 0.7 | 0.7 | 0.00 | 0.8 | 0.7 | 0.00 |
| Venous thrombosis | 1.6 | 1.7 | -0.01 | 1.7 | 1.7 | 0.00 | 1.7 | 1.7 | 0.00 |
| **Medical history: Neoplasms** | | | | | | | | | |
| Hematologic neoplasm | 1.2 | 1.4 | -0.02 | 1.2 | 1.3 | -0.01 | 1.3 | 1.4 | -0.01 |
| Malignant lymphoma | 0.6 | 0.7 | -0.01 | 0.6 | 0.6 | 0.00 | 0.6 | 0.7 | -0.01 |
| Malignant neoplasm of anorectum | 0.2 | 0.3 | -0.01 | 0.3 | 0.2 | 0.00 | 0.3 | 0.3 | 0.00 |
| Malignant neoplastic disease | 10.9 | 11.4 | -0.01 | 11.0 | 11.1 | 0.00 | 11.3 | 11.3 | 0.00 |
| Malignant tumor of colon | 0.6 | 0.7 | -0.02 | 0.6 | 0.6 | 0.00 | 0.7 | 0.7 | 0.00 |
| Malignant tumor of lung | 0.4 | 0.5 | -0.02 | 0.4 | 0.4 | -0.01 | 0.5 | 0.5 | 0.00 |
| Malignant tumor of urinary bladder | 0.7 | 0.8 | -0.01 | 0.8 | 0.8 | 0.00 | 0.8 | 0.8 | 0.00 |
| Primary malignant neoplasm of prostate | 4.2 | 4.1 | 0.00 | 4.1 | 4.1 | 0.00 | 4.1 | 4.1 | 0.00 |

| (Continued) | Before ajustment | | | After matching | | | After stratification | | |
|---|---|---|---|---|---|---|---|---|---|
| Characteristic | T (%) | C (%) | StdDiff | T (%) | C (%) | StdDiff | T (%) | C (%) | StdDiff |
| **Medication use** | | | | | | | | | |
| Agents acting on the renin-angiotensin system | 71.4 | 71.6 | -0.01 | 71.2 | 71.0 | 0.00 | 71.9 | 71.5 | 0.01 |
| Antibacterials for systemic use | 52.8 | 51.7 | 0.02 | 52.5 | 52.9 | -0.01 | 52.1 | 52.2 | 0.00 |
| Antidepressants | 18.8 | 19.0 | 0.00 | 18.6 | 18.8 | 0.00 | 19.2 | 19.0 | 0.00 |
| Antiepileptics | 12.8 | 13.6 | -0.02 | 12.8 | 13.0 | -0.01 | 13.7 | 13.4 | 0.01 |
| Antiinflammatory and antirheumatic products | 26.7 | 26.2 | 0.01 | 26.4 | 26.6 | -0.01 | 26.4 | 26.6 | -0.01 |
| Antineoplastic agents | 4.8 | 4.4 | 0.02 | 4.6 | 4.7 | 0.00 | 4.6 | 4.5 | 0.00 |
| Antipsoriatics | 0.6 | 0.4 | 0.02 | 0.5 | 0.5 | 0.00 | 0.5 | 0.5 | 0.00 |
| Antithrombotic agents | 17.3 | 18.0 | -0.02 | 17.4 | 17.5 | 0.00 | 18.1 | 17.9 | 0.01 |
| Beta blocking agents | 33.6 | 35.7 | -0.04 | 33.6 | 33.4 | 0.00 | 35.4 | 35.0 | 0.01 |
| Calcium channel blockers | 25.7 | 25.9 | 0.00 | 26.2 | 25.8 | 0.01 | 26.2 | 25.8 | 0.01 |
| Diuretics | 36.3 | 38.1 | -0.04 | 36.3 | 36.0 | 0.00 | 38.0 | 37.5 | 0.01 |
| Drugs for acid related disorders | 22.6 | 22.4 | 0.01 | 22.4 | 22.6 | -0.01 | 22.8 | 22.5 | 0.01 |
| Drugs for obstructive airway diseases | 32.9 | 31.3 | 0.03 | 32.3 | 32.9 | -0.01 | 32.3 | 32.9 | -0.01 |
| Immunosuppressants | 1.8 | 1.6 | 0.01 | 1.7 | 1.7 | 0.00 | 1.7 | 1.6 | 0.00 |
| Lipid modifying agents | 76.3 | 73.8 | 0.06 | 75.7 | 75.9 | 0.00 | 74.5 | 74.6 | 0.00 |
| Opioids | 24.2 | 25.1 | -0.02 | 24.1 | 24.2 | 0.00 | 24.1 | 24.2 | 0.00 |
| Psycholeptics | 20.0 | 19.8 | 0.00 | 19.8 | 19.9 | 0.00 | 20.2 | 19.9 | 0.01 |
| Psychostimulants | 1.8 | 1.2 | 0.05 | 1.6 | 1.7 | 0.00 | 1.5 | 1.4 | 0.01 |

##### 2.6.4 MDCR

**Supplementary Table 195:** Baseline patient characteristics for DPP4I (male) (T) and SU (male) (C) new-users in the MDCR data source. We report proportion of initiators satisfying selected base-line characteristics and the standardized difference of population proportions (StdDiff) before and after propensity score adjustment. Less extreme StdDiffs through matching and stratification suggest improved balance between patient cohorts through adjustment.

| | Before adjustment | | | After matching | | | After stratification | | |
|---|---|---|---|---|---|---|---|---|---|
| Characteristic | T (%) | C (%) | StdDiff | T (%) | C (%) | StdDiff | T (%) | C (%) | StdDiff |
| **Age group** | | | | | | | | | |
| 50 - 54 | 0.2 | 0.2 | -0.01 | 0.2 | 0.2 | 0.00 | 0.2 | 0.2 | -0.01 |
| 55 - 59 | 0.5 | 0.6 | 0.00 | 0.5 | 0.6 | -0.01 | 0.6 | 0.6 | 0.00 |
| 60 - 64 | 1.9 | 1.6 | 0.02 | 1.8 | 1.8 | 0.00 | 1.8 | 1.8 | 0.00 |
| 65 - 69 | 31.6 | 28.3 | 0.07 | 31.3 | 31.4 | 0.00 | 29.6 | 30.1 | -0.01 |
| 70 - 74 | 29.9 | 30.7 | -0.01 | 30.0 | 30.2 | 0.00 | 29.9 | 30.4 | -0.01 |
| 75 - 79 | 19.2 | 20.6 | -0.03 | 19.2 | 19.3 | 0.00 | 19.6 | 19.6 | 0.00 |
| 80 - 84 | 10.5 | 12.1 | -0.05 | 10.7 | 10.8 | 0.00 | 11.4 | 11.3 | 0.00 |
| 85 - 89 | 4.6 | 4.7 | 0.00 | 4.7 | 4.5 | 0.01 | 5.0 | 4.6 | 0.02 |
| 90 - 94 | 1.3 | 1.2 | 0.01 | 1.4 | 1.0 | 0.03 | 1.6 | 1.1 | 0.05 |
| 95 - 99 | 0.2 | 0.1 | 0.01 | 0.2 | 0.1 | 0.01 | 0.2 | 0.1 | 0.02 |
| **Medical history: General** | | | | | | | | | |
| Acute respiratory disease | 21.1 | 19.6 | 0.04 | 20.7 | 21.0 | -0.01 | 21.2 | 20.8 | 0.01 |
| Attention deficit hyperactivity disorder | 0.2 | 0.1 | 0.02 | 0.2 | 0.2 | 0.00 | 0.2 | 0.1 | 0.01 |
| Chronic liver disease | 1.9 | 1.9 | 0.00 | 1.9 | 2.1 | -0.01 | 2.0 | 2.2 | -0.01 |
| Chronic obstructive lung disease | 10.8 | 11.8 | -0.03 | 10.9 | 11.0 | 0.00 | 12.4 | 11.8 | 0.02 |
| Crohn's disease | 0.3 | 0.2 | 0.03 | 0.4 | 0.2 | 0.03 | 0.3 | 0.2 | 0.02 |
| Dementia | 2.4 | 2.5 | -0.01 | 2.4 | 2.6 | -0.01 | 2.6 | 2.7 | 0.00 |
| Depressive disorder | 6.0 | 5.2 | 0.03 | 6.0 | 6.1 | 0.00 | 6.2 | 6.0 | 0.01 |
| Gastroesophageal reflux disease | 11.0 | 8.9 | 0.07 | 10.7 | 11.1 | -0.01 | 10.2 | 10.7 | -0.01 |
| Gastrointestinal hemorrhage | 2.9 | 3.4 | -0.03 | 2.9 | 3.1 | -0.01 | 3.0 | 3.1 | -0.01 |
| Human immunodeficiency virus infection | 0.2 | 0.1 | 0.03 | 0.2 | 0.1 | 0.02 | 0.1 | 0.1 | 0.01 |
| Hyperlipidemia | 64.5 | 50.4 | 0.29 | 63.3 | 65.1 | -0.04 | 58.8 | 60.3 | -0.03 |
| Hypertensive disorder | 73.9 | 64.6 | 0.20 | 73.1 | 73.8 | -0.01 | 71.2 | 71.3 | 0.00 |
| Lesion of liver | 0.7 | 1.1 | -0.04 | 0.7 | 1.0 | -0.04 | 0.9 | 1.2 | -0.03 |
| Obesity | 9.7 | 7.3 | 0.09 | 9.5 | 10.2 | -0.02 | 8.9 | 9.4 | -0.02 |
| Osteoarthritis | 27.0 | 23.3 | 0.09 | 26.5 | 27.0 | -0.01 | 26.2 | 25.8 | 0.01 |
| Pneumonia | 5.1 | 6.0 | -0.04 | 5.3 | 5.0 | 0.01 | 6.1 | 5.6 | 0.02 |
| Psoriasis | 1.7 | 1.3 | 0.03 | 1.6 | 1.6 | 0.00 | 1.6 | 1.5 | 0.01 |
| Renal impairment | 14.5 | 12.9 | 0.05 | 14.5 | 14.3 | 0.00 | 15.1 | 14.6 | 0.01 |
| Rheumatoid arthritis | 1.3 | 1.0 | 0.02 | 1.3 | 1.1 | 0.02 | 1.4 | 1.1 | 0.03 |
| Schizophrenia | 0.1 | 0.1 | 0.00 | 0.1 | 0.1 | 0.00 | 0.1 | 0.1 | 0.00 |
| Ulcerative colitis | 0.5 | 0.4 | 0.02 | 0.5 | 0.4 | 0.01 | 0.5 | 0.4 | 0.01 |
| Urinary tract infectious disease | 6.3 | 6.2 | 0.00 | 6.2 | 6.1 | 0.00 | 6.4 | 6.2 | 0.01 |
| Viral hepatitis C | 0.3 | 0.2 | 0.01 | 0.3 | 0.2 | 0.01 | 0.2 | 0.3 | 0.00 |
| Visual system disorder | 49.5 | 45.6 | 0.08 | 48.7 | 49.6 | -0.02 | 47.1 | 47.7 | -0.01 |
| **Medical history: Cardiovascular disease** | | | | | | | | | |
| Atrial fibrillation | 13.2 | 12.8 | 0.01 | 13.0 | 13.1 | 0.00 | 13.9 | 13.2 | 0.02 |
| Cerebrovascular disease | 13.0 | 12.5 | 0.02 | 12.8 | 12.8 | 0.00 | 12.9 | 12.5 | 0.01 |
| Coronary arteriosclerosis | 32.2 | 30.1 | 0.04 | 31.5 | 32.5 | -0.02 | 31.5 | 31.2 | 0.01 |
| Heart disease | 50.5 | 48.6 | 0.04 | 49.8 | 50.6 | -0.02 | 50.1 | 49.6 | 0.01 |
| Heart failure | 9.4 | 11.2 | -0.06 | 9.5 | 9.6 | 0.00 | 11.1 | 10.4 | 0.02 |
| Ischemic heart disease | 13.6 | 14.4 | -0.02 | 13.5 | 13.9 | -0.01 | 14.2 | 14.1 | 0.00 |
| Peripheral vascular disease | 13.1 | 11.4 | 0.05 | 12.7 | 12.9 | -0.01 | 12.7 | 12.4 | 0.01 |
| Pulmonary embolism | 0.9 | 0.8 | 0.01 | 0.8 | 0.9 | 0.00 | 1.1 | 0.9 | 0.02 |
| Venous thrombosis | 2.2 | 2.1 | 0.01 | 2.2 | 2.3 | 0.00 | 2.5 | 2.3 | 0.01 |
| **Medical history: Neoplasms** | | | | | | | | | |
| Hematologic neoplasm | 2.1 | 1.9 | 0.01 | 2.1 | 2.0 | 0.00 | 2.2 | 2.1 | 0.01 |
| Malignant lymphoma | 1.1 | 0.9 | 0.02 | 1.1 | 1.0 | 0.01 | 1.1 | 1.0 | 0.01 |
| Malignant neoplasm of anorectum | 0.4 | 0.5 | -0.02 | 0.4 | 0.4 | 0.00 | 0.4 | 0.4 | 0.00 |
| Malignant neoplastic disease | 22.0 | 21.1 | 0.02 | 22.0 | 22.0 | 0.00 | 21.6 | 21.8 | 0.00 |
| Malignant tumor of colon | 1.0 | 1.1 | -0.01 | 1.0 | 0.9 | 0.01 | 1.1 | 1.0 | 0.01 |
| Malignant tumor of lung | 0.7 | 0.9 | -0.03 | 0.7 | 0.8 | -0.01 | 0.8 | 0.9 | -0.01 |
| Malignant tumor of urinary bladder | 2.0 | 1.7 | 0.02 | 1.9 | 1.8 | 0.01 | 1.9 | 1.8 | 0.00 |
| Primary malignant neoplasm of prostate | 8.4 | 8.3 | 0.00 | 8.4 | 8.6 | -0.01 | 8.3 | 8.5 | -0.01 |
| **Medication use** | | | | | | | | | |
| Agents acting on the renin-angiotensin system | 76.3 | 72.4 | 0.09 | 75.8 | 76.0 | 0.00 | 74.9 | 75.5 | -0.01 |
| Antibacterials for systemic use | 58.7 | 57.7 | 0.02 | 58.0 | 58.5 | -0.01 | 58.6 | 58.4 | 0.00 |
| Antidepressants | 18.8 | 18.6 | 0.00 | 18.7 | 18.9 | 0.00 | 19.0 | 19.1 | 0.00 |

| (Continued) | Before ajustment | | | After matching | | | After stratification | | |
|---|---|---|---|---|---|---|---|---|---|
| Characteristic | T (%) | C (%) | StdDiff | T (%) | C (%) | StdDiff | T (%) | C (%) | StdDiff |
| Antiepileptics | 12.8 | 12.1 | 0.02 | 12.9 | 13.1 | -0.01 | 13.3 | 13.2 | 0.00 |
| Antiinflammatory and antirheumatic products | 25.5 | 25.0 | 0.01 | 25.3 | 25.6 | -0.01 | 25.0 | 25.5 | -0.01 |
| Antineoplastic agents | 8.2 | 8.3 | 0.00 | 8.1 | 7.9 | 0.01 | 8.1 | 7.9 | 0.01 |
| Antipsoriatics | 0.8 | 0.6 | 0.02 | 0.8 | 0.7 | 0.00 | 0.7 | 0.7 | 0.00 |
| Antithrombotic agents | 28.4 | 28.2 | 0.00 | 28.2 | 28.6 | -0.01 | 29.7 | 28.8 | 0.02 |
| Beta blocking agents | 50.3 | 49.5 | 0.02 | 50.0 | 50.2 | 0.00 | 51.0 | 50.8 | 0.00 |
| Calcium channel blockers | 32.8 | 31.7 | 0.02 | 32.7 | 33.1 | -0.01 | 32.6 | 33.1 | -0.01 |
| Diuretics | 45.7 | 47.7 | -0.04 | 45.9 | 45.8 | 0.00 | 47.8 | 47.2 | 0.01 |
| Drugs for acid related disorders | 29.8 | 28.9 | 0.02 | 29.4 | 29.3 | 0.00 | 29.4 | 29.3 | 0.00 |
| Drugs for obstructive airway diseases | 38.5 | 35.8 | 0.06 | 38.1 | 38.2 | 0.00 | 38.3 | 37.9 | 0.01 |
| Immunosuppressants | 2.4 | 2.0 | 0.03 | 2.5 | 2.3 | 0.01 | 2.5 | 2.3 | 0.01 |
| Lipid modifying agents | 82.7 | 76.9 | 0.15 | 82.1 | 82.3 | 0.00 | 80.4 | 81.3 | -0.02 |
| Opioids | 25.4 | 26.0 | -0.01 | 25.5 | 25.6 | 0.00 | 27.1 | 26.5 | 0.01 |
| Psycholeptics | 22.4 | 21.8 | 0.02 | 22.3 | 22.2 | 0.00 | 22.6 | 22.4 | 0.00 |
| Psychostimulants | 1.1 | 0.8 | 0.02 | 1.0 | 1.0 | 0.00 | 1.1 | 0.9 | 0.01 |

##### 2.6.5 MDCD

**Supplementary Table 196:** Baseline patient characteristics for DPP4I (male) (T) and SU (male) (C) new-users in the MDCD data source. We report proportion of initiators satisfying selected base-line characteristics and the standardized difference of population proportions (StdDiff) before and after propensity score adjustment. Less extreme StdDiffs through matching and stratification suggest improved balance between patient cohorts through adjustment.

| | Before adjustment | | | After matching | | | After stratification | | |
|---|---|---|---|---|---|---|---|---|---|
| Characteristic | T (%) | C (%) | StdDiff | T (%) | C (%) | StdDiff | T (%) | C (%) | StdDiff |
| **Age group** | | | | | | | | | |
| 15 - 19 | 0.7 | 0.6 | 0.02 | 0.7 | 0.6 | 0.02 | 0.7 | 0.6 | 0.02 |
| 20 - 24 | 1.2 | 0.9 | 0.03 | 1.2 | 1.0 | 0.02 | 1.3 | 0.9 | 0.03 |
| 25 - 29 | 2.3 | 2.0 | 0.02 | 2.3 | 1.8 | 0.04 | 2.4 | 1.9 | 0.03 |
| 30 - 34 | 4.0 | 4.1 | 0.00 | 4.1 | 4.0 | 0.01 | 4.2 | 4.1 | 0.00 |
| 35 - 39 | 7.0 | 7.5 | -0.02 | 7.0 | 7.3 | -0.01 | 7.7 | 7.5 | 0.01 |
| 40 - 44 | 10.7 | 10.4 | 0.01 | 10.8 | 10.5 | 0.01 | 10.8 | 10.4 | 0.01 |
| 45 - 49 | 14.5 | 13.7 | 0.02 | 14.6 | 13.9 | 0.02 | 14.2 | 13.8 | 0.01 |
| 50 - 54 | 16.6 | 17.1 | -0.01 | 16.6 | 17.4 | -0.02 | 15.9 | 17.2 | -0.03 |
| 55 - 59 | 18.3 | 18.9 | -0.02 | 18.2 | 18.8 | -0.02 | 18.5 | 18.9 | -0.01 |
| 60 - 64 | 15.8 | 15.9 | 0.00 | 15.6 | 15.6 | 0.00 | 15.6 | 15.6 | 0.00 |
| 65 - 69 | 3.9 | 4.6 | -0.04 | 3.9 | 4.7 | -0.04 | 3.9 | 4.6 | -0.04 |
| 70 - 74 | 2.5 | 2.1 | 0.02 | 2.5 | 2.1 | 0.02 | 2.3 | 2.2 | 0.01 |
| 75 - 79 | 1.4 | 1.1 | 0.03 | 1.3 | 1.1 | 0.03 | 1.3 | 1.1 | 0.02 |
| 80 - 84 | 1.0 | 0.7 | 0.04 | 1.0 | 0.8 | 0.02 | 1.0 | 0.7 | 0.03 |
| 85 - 89 | 0.1 | 0.3 | -0.03 | 0.1 | 0.2 | -0.02 | 0.1 | 0.2 | -0.03 |
| 90 - 94 | <0.1 | <0.1 | 0.00 | <0.1 | <0.1 | 0.00 | <0.1 | <0.1 | 0.00 |
| **Race** | | | | | | | | | |
| Black or African American | 19.6 | 23.3 | -0.09 | 19.9 | 19.1 | 0.02 | 22.1 | 22.0 | 0.00 |
| White | 61.7 | 56.8 | 0.10 | 61.4 | 62.3 | -0.02 | 57.4 | 58.5 | -0.02 |
| **Ethnicity** | | | | | | | | | |
| Hispanic or Latino | 2.8 | 2.6 | 0.01 | 2.8 | 2.6 | 0.01 | 2.9 | 2.6 | 0.02 |
| **Medical history: General** | | | | | | | | | |
| Acute respiratory disease | 27.7 | 27.8 | 0.00 | 27.6 | 28.4 | -0.02 | 27.3 | 28.0 | -0.01 |
| Attention deficit hyperactivity disorder | 1.9 | 1.9 | 0.00 | 1.8 | 2.3 | -0.03 | 1.6 | 2.0 | -0.03 |
| Chronic obstructive lung disease | 19.4 | 18.4 | 0.03 | 19.3 | 19.2 | 0.00 | 18.7 | 18.6 | 0.00 |
| Crohn’s disease | 0.1 | 0.3 | -0.03 | 0.1 | 0.3 | -0.03 | 0.2 | 0.3 | -0.02 |
| Dementia | 2.3 | 1.8 | 0.04 | 2.3 | 1.9 | 0.02 | 2.1 | 1.8 | 0.02 |
| Depressive disorder | 25.5 | 25.7 | 0.00 | 25.3 | 26.5 | -0.03 | 25.0 | 26.0 | -0.02 |
| Gastroesophageal reflux disease | 22.7 | 22.3 | 0.01 | 22.3 | 23.8 | -0.04 | 21.7 | 22.7 | -0.02 |
| Gastrointestinal hemorrhage | 3.7 | 4.0 | -0.02 | 3.7 | 3.8 | 0.00 | 3.7 | 4.0 | -0.02 |
| Hyperlipidemia | 69.6 | 66.1 | 0.07 | 69.1 | 69.9 | -0.02 | 66.6 | 67.4 | -0.02 |
| Hypertensive disorder | 78.5 | 78.2 | 0.01 | 78.3 | 77.7 | 0.01 | 78.4 | 78.1 | 0.01 |
| Lesion of liver | 2.3 | 2.6 | -0.02 | 2.3 | 2.4 | -0.01 | 2.3 | 2.4 | -0.01 |
| Obesity | 32.2 | 29.3 | 0.06 | 31.8 | 32.4 | -0.01 | 30.4 | 30.3 | 0.00 |
| Osteoarthritis | 32.9 | 31.1 | 0.04 | 32.5 | 32.8 | -0.01 | 31.7 | 31.7 | 0.00 |
| Pneumonia | 6.1 | 5.8 | 0.01 | 6.0 | 5.6 | 0.02 | 6.2 | 5.7 | 0.02 |
| Psoriasis | 1.6 | 1.2 | 0.03 | 1.6 | 1.3 | 0.02 | 1.6 | 1.3 | 0.03 |
| Renal impairment | 10.2 | 10.2 | 0.00 | 10.1 | 9.9 | 0.01 | 10.3 | 10.1 | 0.01 |
| Rheumatoid arthritis | 1.3 | 1.1 | 0.02 | 1.3 | 1.1 | 0.02 | 1.3 | 1.1 | 0.02 |
| Schizophrenia | 5.4 | 6.0 | -0.03 | 5.5 | 5.7 | -0.01 | 5.7 | 5.9 | -0.01 |
| Ulcerative colitis | 0.3 | 0.3 | 0.00 | 0.3 | 0.3 | 0.00 | 0.3 | 0.3 | 0.00 |
| Urinary tract infectious disease | 4.9 | 5.0 | 0.00 | 4.8 | 4.5 | 0.02 | 5.0 | 4.9 | 0.01 |
| Viral hepatitis C | 3.4 | 4.0 | -0.03 | 3.9 | 3.8 | 0.00 | 3.9 | 3.8 | 0.00 |
| Visual system disorder | 40.8 | 38.3 | 0.05 | 40.5 | 41.0 | -0.01 | 38.5 | 39.1 | -0.01 |
| **Medical history: Cardiovascular disease** | | | | | | | | | |
| Atrial fibrillation | 4.6 | 4.6 | 0.00 | 4.6 | 4.6 | 0.00 | 4.2 | 4.6 | -0.02 |
| Cerebrovascular disease | 5.3 | 5.8 | -0.02 | 5.3 | 5.8 | -0.02 | 5.0 | 5.8 | -0.04 |
| Coronary arteriosclerosis | 16.7 | 16.1 | 0.02 | 16.3 | 17.4 | -0.03 | 15.5 | 16.5 | -0.03 |
| Heart failure | 9.9 | 10.2 | -0.01 | 9.8 | 10.1 | -0.01 | 9.7 | 10.2 | -0.01 |
| Ischemic heart disease | 11.4 | 11.3 | 0.00 | 11.2 | 12.0 | -0.03 | 10.9 | 11.5 | -0.02 |
| Peripheral vascular disease | 10.1 | 9.0 | 0.04 | 9.9 | 9.5 | 0.01 | 9.6 | 9.2 | 0.01 |
| Pulmonary embolism | 1.2 | 1.1 | 0.01 | 1.1 | 1.0 | 0.01 | 1.1 | 1.1 | 0.00 |
| Venous thrombosis | 1.6 | 2.1 | -0.04 | 1.6 | 2.0 | -0.03 | 1.7 | 2.1 | -0.03 |
| **Medical history: Neoplasms** | | | | | | | | | |
| Hematologic neoplasm | 1.1 | 1.0 | 0.01 | 1.1 | 1.0 | 0.02 | 1.1 | 1.0 | 0.01 |
| Malignant lymphoma | 0.7 | 0.4 | 0.04 | 0.7 | 0.4 | 0.04 | 0.6 | 0.4 | 0.03 |
| Malignant neoplasm of anorectum | 0.2 | 0.4 | -0.03 | 0.2 | 0.4 | -0.03 | 0.2 | 0.4 | -0.03 |
| Malignant neoplastic disease | 5.6 | 5.5 | 0.00 | 5.5 | 5.8 | -0.01 | 5.4 | 5.6 | -0.01 |

| (Continued) | Before ajustment | | | After matching | | | After stratification | | |
|---|---|---|---|---|---|---|---|---|---|
| Characteristic | T (%) | C (%) | StdDiff | T (%) | C (%) | StdDiff | T (%) | C (%) | StdDiff |
| Malignant tumor of breast | <0.1 | <0.1 | 0.01 | <0.1 | <0.1 | 0.03 | <0.1 | <0.1 | 0.03 |
| Malignant tumor of colon | 0.5 | 0.5 | -0.01 | 0.5 | 0.6 | -0.01 | 0.5 | 0.6 | -0.01 |
| Malignant tumor of lung | 0.5 | 0.4 | 0.01 | 0.5 | 0.4 | 0.01 | 0.5 | 0.4 | 0.01 |
| Malignant tumor of urinary bladder | 0.3 | 0.3 | -0.01 | 0.2 | 0.3 | -0.01 | 0.3 | 0.3 | -0.01 |
| Primary malignant neoplasm of prostate | 1.3 | 1.3 | 0.00 | 1.2 | 1.2 | 0.01 | 1.3 | 1.2 | 0.01 |
| **Medication use** | | | | | | | | | |
| Antibacterials for systemic use | 59.2 | 58.5 | 0.02 | 58.9 | 58.6 | 0.01 | 59.4 | 58.5 | 0.02 |
| Antidepressants | 43.7 | 40.5 | 0.06 | 43.3 | 44.7 | -0.03 | 40.9 | 41.8 | -0.02 |
| Antiepileptics | 37.1 | 34.4 | 0.06 | 36.7 | 37.2 | -0.01 | 35.9 | 35.3 | 0.01 |
| Antiinflammatory and antirheumatic products | 53.9 | 53.3 | 0.01 | 53.9 | 54.3 | -0.01 | 53.5 | 53.7 | 0.00 |
| Antineoplastic agents | 4.3 | 3.0 | 0.07 | 4.2 | 3.4 | 0.04 | 4.0 | 3.2 | 0.04 |
| Antipsoriatics | 0.4 | 0.4 | 0.01 | 0.4 | 0.5 | 0.00 | 0.4 | 0.4 | 0.00 |
| Antithrombotic agents | 29.7 | 28.0 | 0.04 | 29.4 | 30.1 | -0.02 | 27.7 | 28.7 | -0.02 |
| Beta blocking agents | 38.0 | 36.7 | 0.03 | 37.6 | 38.3 | -0.01 | 36.1 | 37.2 | -0.02 |
| Calcium channel blockers | 26.2 | 26.7 | -0.01 | 26.1 | 26.4 | -0.01 | 27.1 | 26.7 | 0.01 |
| Diuretics | 41.2 | 42.8 | -0.03 | 41.3 | 40.6 | 0.01 | 42.6 | 42.2 | 0.01 |
| Drugs for acid related disorders | 41.7 | 39.1 | 0.05 | 41.0 | 42.1 | -0.02 | 39.4 | 40.0 | -0.01 |
| Drugs for obstructive airway diseases | 50.2 | 46.7 | 0.07 | 49.5 | 50.9 | -0.03 | 47.8 | 48.0 | 0.00 |
| Immunosuppressants | 1.8 | 1.7 | 0.01 | 1.8 | 1.9 | -0.01 | 1.7 | 1.8 | -0.01 |
| Lipid modifying agents | 71.2 | 67.8 | 0.07 | 70.8 | 72.4 | -0.04 | 67.4 | 69.2 | -0.04 |
| Opioids | 41.4 | 42.9 | -0.03 | 41.1 | 41.4 | -0.01 | 41.8 | 42.6 | -0.02 |
| Psycholeptics | 42.6 | 41.2 | 0.03 | 42.4 | 42.9 | -0.01 | 42.4 | 42.9 | -0.01 |
| Psychostimulants | 3.7 | 2.8 | 0.05 | 3.7 | 3.2 | 0.02 | 3.3 | 2.9 | 0.02 |

##### 2.6.6 Open Claims

**Supplementary Table 197:** Baseline patient characteristics for DPP4I (male) (T) and SU (male) (C) new-users in the Open Claims data source. We report proportion of initiators satisfying selected base-line characteristics and the standardized difference of population proportions (StdDiff) before and after propensity score adjustment. Less extreme StdDiffs through matching and stratification suggest improved balance between patient cohorts through adjustment.

| | Before adjustment | | | After matching | | | After stratification | | |
|---|---|---|---|---|---|---|---|---|---|
| Characteristic | T (%) | C (%) | StdDiff | T (%) | C (%) | StdDiff | T (%) | C (%) | StdDiff |
| **Age group** | | | | | | | | | |
| 15 - 19 | 0.1 | 0.1 | 0.00 | 0.1 | 0.1 | 0.00 | 0.1 | 0.1 | 0.00 |
| 20 - 24 | 0.2 | 0.3 | 0.00 | 0.2 | 0.2 | 0.00 | 0.2 | 0.2 | 0.00 |
| 25 - 29 | 0.5 | 0.6 | 0.00 | 0.6 | 0.5 | 0.00 | 0.6 | 0.6 | 0.00 |
| 30 - 34 | 1.3 | 1.4 | -0.01 | 1.4 | 1.3 | 0.01 | 1.4 | 1.4 | 0.01 |
| 35 - 39 | 3.0 | 3.0 | 0.00 | 3.0 | 2.9 | 0.01 | 3.0 | 3.0 | 0.00 |
| 40 - 44 | 5.4 | 5.4 | 0.00 | 5.5 | 5.4 | 0.00 | 5.4 | 5.3 | 0.00 |
| 45 - 49 | 8.9 | 8.5 | 0.01 | 8.9 | 8.9 | 0.00 | 8.9 | 8.9 | 0.00 |
| 50 - 54 | 12.4 | 12.0 | 0.01 | 12.4 | 12.3 | 0.00 | 12.0 | 12.0 | 0.00 |
| 55 - 59 | 15.2 | 14.3 | 0.03 | 15.2 | 15.1 | 0.00 | 14.7 | 14.6 | 0.00 |
| 60 - 64 | 15.5 | 15.0 | 0.02 | 15.5 | 15.6 | 0.00 | 15.1 | 15.2 | 0.00 |
| 65 - 69 | 13.9 | 14.7 | -0.02 | 13.9 | 14.1 | -0.01 | 14.4 | 14.6 | -0.01 |
| 70 - 74 | 10.5 | 11.4 | -0.03 | 10.4 | 10.6 | -0.01 | 11.0 | 11.2 | 0.00 |
| 75 - 79 | 9.2 | 9.8 | -0.02 | 9.2 | 9.2 | 0.00 | 9.7 | 9.7 | 0.00 |
| 80 - 84 | 3.6 | 3.5 | 0.00 | 3.6 | 3.6 | 0.00 | 3.6 | 3.6 | 0.00 |
| 85 - 89 | 0.1 | 0.1 | 0.00 | 0.1 | 0.1 | 0.00 | 0.1 | 0.1 | 0.01 |
| **Medical history: General** | | | | | | | | | |
| Acute respiratory disease | 12.2 | 11.1 | 0.03 | 12.1 | 11.9 | 0.00 | 11.6 | 11.3 | 0.01 |
| Attention deficit hyperactivity disorder | 0.4 | 0.4 | 0.01 | 0.4 | 0.5 | 0.00 | 0.4 | 0.4 | 0.00 |
| Chronic liver disease | 1.5 | 1.4 | 0.01 | 1.6 | 1.5 | 0.01 | 1.5 | 1.5 | 0.00 |
| Chronic obstructive lung disease | 5.0 | 5.4 | -0.02 | 5.0 | 5.1 | -0.01 | 5.2 | 5.4 | -0.01 |
| Crohn’s disease | 0.1 | 0.1 | 0.00 | 0.1 | 0.2 | 0.00 | 0.1 | 0.2 | 0.00 |
| Dementia | 0.8 | 0.8 | 0.00 | 0.8 | 0.8 | 0.00 | 0.8 | 0.8 | 0.00 |
| Depressive disorder | 4.9 | 5.0 | -0.01 | 5.0 | 5.1 | 0.00 | 5.0 | 5.1 | 0.00 |
| Gastroesophageal reflux disease | 7.1 | 6.4 | 0.03 | 7.1 | 7.0 | 0.00 | 6.8 | 6.7 | 0.00 |
| Gastrointestinal hemorrhage | 1.5 | 1.4 | 0.00 | 1.4 | 1.4 | 0.00 | 1.4 | 1.5 | 0.00 |
| Human immunodeficiency virus infection | 0.5 | 0.3 | 0.02 | 0.5 | 0.4 | 0.00 | 0.4 | 0.4 | 0.00 |
| Hyperlipidemia | 47.3 | 41.9 | 0.11 | 46.9 | 46.9 | 0.00 | 43.8 | 43.7 | 0.00 |
| Hypertensive disorder | 51.6 | 48.6 | 0.06 | 49.9 | 49.7 | 0.00 | 49.9 | 49.7 | 0.00 |
| Lesion of liver | 0.8 | 0.8 | -0.01 | 0.8 | 0.7 | 0.00 | 0.8 | 0.8 | 0.00 |
| Obesity | 9.4 | 9.1 | 0.01 | 9.4 | 9.6 | -0.01 | 9.2 | 9.4 | -0.01 |
| Osteoarthritis | 13.1 | 12.3 | 0.03 | 13.0 | 13.1 | 0.00 | 12.8 | 12.6 | 0.01 |
| Pneumonia | 2.3 | 2.5 | -0.01 | 2.5 | 2.5 | 0.00 | 2.5 | 2.5 | 0.00 |
| Psoriasis | 0.9 | 0.8 | 0.01 | 0.9 | 0.9 | 0.00 | 0.8 | 0.8 | 0.00 |
| Renal impairment | 7.2 | 7.4 | -0.01 | 7.2 | 7.4 | -0.01 | 7.4 | 7.5 | 0.00 |
| Rheumatoid arthritis | 0.6 | 0.6 | 0.01 | 0.6 | 0.7 | -0.01 | 0.6 | 0.6 | 0.00 |
| Schizophrenia | 0.4 | 0.5 | -0.01 | 0.5 | 0.5 | 0.00 | 0.5 | 0.5 | 0.00 |
| Ulcerative colitis | 0.2 | 0.2 | 0.00 | 0.2 | 0.2 | 0.00 | 0.2 | 0.2 | 0.00 |
| Urinary tract infectious disease | 2.9 | 2.7 | 0.01 | 2.9 | 2.9 | 0.00 | 2.8 | 2.8 | 0.00 |
| Viral hepatitis C | 0.5 | 0.5 | -0.01 | 0.5 | 0.5 | 0.00 | 0.5 | 0.5 | 0.00 |
| Visual system disorder | 18.0 | 16.9 | 0.03 | 17.8 | 17.7 | 0.00 | 17.3 | 17.2 | 0.00 |
| **Medical history: Cardiovascular disease** | | | | | | | | | |
| Atrial fibrillation | 4.7 | 4.6 | 0.00 | 4.7 | 4.7 | 0.00 | 4.7 | 4.7 | 0.00 |
| Cerebrovascular disease | 3.5 | 3.3 | 0.02 | 3.5 | 3.5 | 0.00 | 3.5 | 3.5 | 0.00 |
| Coronary arteriosclerosis | 11.1 | 10.5 | 0.02 | 11.0 | 11.3 | -0.01 | 10.8 | 10.8 | 0.00 |
| Heart disease | 20.7 | 19.9 | 0.02 | 20.4 | 20.6 | 0.00 | 20.3 | 20.3 | 0.00 |
| Heart failure | 4.2 | 4.5 | -0.02 | 4.2 | 4.3 | 0.00 | 4.4 | 4.4 | 0.00 |
| Ischemic heart disease | 5.0 | 4.8 | 0.01 | 4.9 | 5.1 | -0.01 | 4.9 | 5.0 | 0.00 |
| Peripheral vascular disease | 5.0 | 4.9 | 0.00 | 4.9 | 4.9 | 0.00 | 4.9 | 4.9 | 0.00 |
| Pulmonary embolism | 0.5 | 0.5 | 0.00 | 0.5 | 0.5 | -0.01 | 0.5 | 0.5 | -0.01 |
| Venous thrombosis | 1.0 | 0.9 | 0.00 | 1.0 | 1.0 | 0.00 | 1.0 | 1.0 | 0.00 |
| **Medical history: Neoplasms** | | | | | | | | | |
| Hematologic neoplasm | 0.8 | 0.8 | 0.00 | 0.8 | 0.8 | 0.00 | 0.8 | 0.8 | 0.00 |
| Malignant lymphoma | 0.4 | 0.5 | 0.00 | 0.4 | 0.5 | -0.01 | 0.4 | 0.5 | 0.00 |
| Malignant neoplasm of anorectum | 0.2 | 0.2 | 0.00 | 0.2 | 0.2 | 0.00 | 0.2 | 0.2 | 0.00 |
| Malignant neoplastic disease | 6.7 | 6.5 | 0.01 | 6.7 | 6.8 | -0.01 | 6.6 | 6.7 | 0.00 |
| Malignant tumor of breast | 0.1 | 0.1 | 0.00 | 0.1 | 0.1 | 0.00 | 0.1 | 0.1 | 0.00 |
| Malignant tumor of colon | 0.4 | 0.4 | 0.00 | 0.4 | 0.4 | 0.00 | 0.4 | 0.4 | 0.00 |

| (Continued) | Before ajustment | | | After matching | | | After stratification | | |
|---|---|---|---|---|---|---|---|---|---|
| Characteristic | T (%) | C (%) | StdDiff | T (%) | C (%) | StdDiff | T (%) | C (%) | StdDiff |
| Malignant tumor of lung | 0.2 | 0.3 | -0.02 | 0.2 | 0.2 | 0.00 | 0.3 | 0.3 | 0.00 |
| Malignant tumor of urinary bladder | 0.5 | 0.5 | 0.00 | 0.5 | 0.5 | 0.00 | 0.5 | 0.5 | 0.00 |
| Primary malignant neoplasm of prostate | 2.4 | 2.1 | 0.01 | 2.4 | 2.3 | 0.00 | 2.3 | 2.3 | 0.00 |
| **Medication use** | | | | | | | | | |
| Agents acting on the renin-angiotensin system | 71.0 | 70.3 | 0.01 | 70.6 | 70.8 | 0.00 | 70.7 | 70.6 | 0.00 |
| Antibacterials for systemic use | 51.2 | 49.2 | 0.04 | 51.1 | 51.3 | 0.00 | 50.0 | 49.9 | 0.00 |
| Antidepressants | 20.5 | 20.9 | -0.01 | 20.6 | 20.8 | -0.01 | 20.8 | 21.0 | -0.01 |
| Antiepileptics | 15.7 | 16.4 | -0.02 | 15.7 | 15.9 | -0.01 | 16.3 | 16.2 | 0.00 |
| Antiinflammatory and antirheumatic products | 33.1 | 32.0 | 0.02 | 32.7 | 32.3 | 0.01 | 32.7 | 32.3 | 0.01 |
| Antineoplastic agents | 4.2 | 3.6 | 0.03 | 4.1 | 4.2 | -0.01 | 3.8 | 3.8 | 0.00 |
| Antipsoriatics | 0.6 | 0.4 | 0.02 | 0.5 | 0.5 | 0.00 | 0.5 | 0.5 | 0.01 |
| Antithrombotic agents | 21.3 | 21.0 | 0.01 | 21.2 | 21.6 | -0.01 | 21.3 | 21.4 | 0.00 |
| Beta blocking agents | 33.9 | 34.3 | -0.01 | 33.7 | 34.0 | -0.01 | 34.5 | 34.4 | 0.00 |
| Calcium channel blockers | 26.0 | 25.5 | 0.01 | 25.8 | 26.1 | -0.01 | 25.9 | 25.6 | 0.00 |
| Diuretics | 35.4 | 36.4 | -0.02 | 35.2 | 35.6 | -0.01 | 36.1 | 36.0 | 0.00 |
| Drugs for acid related disorders | 28.7 | 26.9 | 0.04 | 28.5 | 28.7 | 0.00 | 27.8 | 27.7 | 0.00 |
| Drugs for obstructive airway diseases | 32.1 | 29.8 | 0.05 | 31.8 | 32.3 | -0.01 | 30.8 | 30.6 | 0.00 |
| Immunosuppressants | 2.0 | 1.6 | 0.03 | 1.9 | 1.9 | 0.00 | 1.8 | 1.7 | 0.00 |
| Lipid modifying agents | 73.7 | 70.7 | 0.07 | 71.8 | 71.6 | 0.00 | 71.8 | 71.6 | 0.00 |
| Opioids | 23.1 | 22.9 | 0.00 | 22.9 | 23.4 | -0.01 | 22.8 | 23.1 | -0.01 |
| Psycholeptics | 18.7 | 18.3 | 0.01 | 18.6 | 18.8 | 0.00 | 18.7 | 18.5 | 0.00 |
| Psychostimulants | 1.9 | 1.7 | 0.02 | 1.9 | 2.0 | 0.00 | 1.7 | 1.8 | 0.00 |

##### 2.6.7 OEHR

**Supplementary Table 198:** Baseline patient characteristics for DPP4I (male) (T) and SU (male) (C) new-users in the OEHR data source. We report proportion of initiators satisfying selected base-line characteristics and the standardized difference of population proportions (StdDiff) before and after propensity score adjustment. Less extreme StdDiffs through matching and stratification suggest improved balance between patient cohorts through adjustment.

| | Before adjustment | | | After matching | | | After stratification | | |
|---|---|---|---|---|---|---|---|---|---|
| Characteristic | T (%) | C (%) | StdDiff | T (%) | C (%) | StdDiff | T (%) | C (%) | StdDiff |
| **Age group** | | | | | | | | | |
| 20 - 24 | 0.2 | 0.2 | 0.00 | 0.2 | 0.2 | 0.01 | 0.2 | 0.2 | 0.00 |
| 25 - 29 | 0.6 | 0.6 | 0.00 | 0.6 | 0.6 | 0.01 | 0.6 | 0.6 | 0.00 |
| 30 - 34 | 1.6 | 1.8 | -0.01 | 1.7 | 1.7 | 0.00 | 1.7 | 1.8 | -0.01 |
| 35 - 39 | 3.8 | 3.9 | -0.01 | 3.8 | 3.8 | 0.00 | 3.9 | 3.9 | 0.00 |
| 40 - 44 | 6.9 | 6.7 | 0.01 | 7.0 | 6.9 | 0.00 | 6.7 | 6.7 | 0.00 |
| 45 - 49 | 11.1 | 10.4 | 0.02 | 10.5 | 10.6 | 0.00 | 10.5 | 10.6 | 0.00 |
| 50 - 54 | 14.4 | 13.8 | 0.02 | 14.3 | 14.4 | 0.00 | 13.9 | 13.9 | 0.00 |
| 55 - 59 | 16.5 | 15.7 | 0.02 | 16.4 | 16.5 | 0.00 | 15.9 | 16.0 | 0.00 |
| 60 - 64 | 15.5 | 14.8 | 0.02 | 15.3 | 15.4 | 0.00 | 15.0 | 15.0 | 0.00 |
| 65 - 69 | 12.1 | 12.4 | -0.01 | 12.1 | 12.2 | 0.00 | 12.3 | 12.3 | 0.00 |
| 70 - 74 | 8.2 | 9.0 | -0.03 | 8.2 | 8.2 | 0.00 | 8.9 | 8.7 | 0.00 |
| 75 - 79 | 5.3 | 6.6 | -0.05 | 5.4 | 5.3 | 0.00 | 6.3 | 6.2 | 0.01 |
| 80 - 84 | 3.0 | 3.4 | -0.03 | 3.0 | 3.0 | 0.00 | 3.4 | 3.3 | 0.00 |
| 85 - 89 | 0.8 | 0.7 | 0.01 | 0.8 | 0.7 | 0.01 | 0.8 | 0.7 | 0.01 |
| **Race** | | | | | | | | | |
| Asian | 2.9 | 2.4 | 0.03 | 2.8 | 2.7 | 0.00 | 2.5 | 2.5 | 0.00 |
| Black or African American | 9.5 | 8.6 | 0.03 | 9.5 | 9.5 | 0.00 | 8.9 | 8.9 | 0.00 |
| White | 80.1 | 81.0 | -0.02 | 80.2 | 80.1 | 0.00 | 80.9 | 80.7 | 0.00 |
| **Ethnicity** | | | | | | | | | |
| Hispanic or Latino | 6.0 | 6.6 | -0.03 | 6.1 | 6.0 | 0.00 | 6.3 | 6.4 | -0.01 |
| Not Hispanic or Latino | 85.9 | 85.7 | 0.01 | 85.9 | 86.0 | 0.00 | 86.1 | 85.8 | 0.01 |
| **Medical history: General** | | | | | | | | | |
| Acute respiratory disease | 15.0 | 13.9 | 0.03 | 14.7 | 14.9 | 0.00 | 14.4 | 14.2 | 0.00 |
| Attention deficit hyperactivity disorder | 0.8 | 0.7 | 0.01 | 0.8 | 0.8 | 0.00 | 0.7 | 0.8 | -0.01 |
| Chronic liver disease | 2.6 | 2.7 | 0.00 | 2.6 | 2.6 | 0.00 | 2.7 | 2.7 | 0.00 |
| Chronic obstructive lung disease | 4.9 | 5.3 | -0.02 | 5.4 | 5.2 | 0.01 | 5.4 | 5.2 | 0.01 |
| Crohn’s disease | 0.2 | 0.2 | 0.00 | 0.2 | 0.2 | 0.00 | 0.2 | 0.2 | 0.00 |
| Dementia | 0.7 | 0.8 | -0.01 | 0.8 | 0.8 | 0.00 | 0.8 | 0.8 | 0.00 |
| Depressive disorder | 8.6 | 9.4 | -0.03 | 9.3 | 9.2 | 0.00 | 9.3 | 9.2 | 0.00 |
| Gastroesophageal reflux disease | 13.7 | 12.8 | 0.03 | 13.5 | 13.5 | 0.00 | 13.4 | 13.1 | 0.01 |
| Gastrointestinal hemorrhage | 1.5 | 1.5 | 0.00 | 1.5 | 1.5 | 0.00 | 1.5 | 1.5 | 0.00 |
| Human immunodeficiency virus infection | 0.3 | 0.3 | 0.01 | 0.3 | 0.3 | 0.00 | 0.3 | 0.3 | 0.00 |
| Hyperlipidemia | 70.1 | 65.6 | 0.10 | 69.1 | 69.9 | -0.02 | 66.9 | 67.0 | 0.00 |
| Hypertensive disorder | 67.1 | 65.4 | 0.04 | 66.5 | 66.9 | -0.01 | 65.9 | 65.9 | 0.00 |
| Lesion of liver | 0.8 | 0.9 | -0.01 | 0.8 | 0.7 | 0.00 | 0.9 | 0.9 | 0.00 |
| Obesity | 23.3 | 21.9 | 0.03 | 22.9 | 23.3 | -0.01 | 22.6 | 22.3 | 0.01 |
| Osteoarthritis | 13.8 | 14.0 | 0.00 | 13.6 | 13.8 | -0.01 | 14.2 | 13.9 | 0.01 |
| Pneumonia | 1.7 | 2.0 | -0.02 | 2.0 | 2.0 | 0.00 | 2.0 | 2.0 | 0.00 |
| Psoriasis | 1.4 | 1.3 | 0.01 | 1.4 | 1.4 | 0.00 | 1.4 | 1.3 | 0.01 |
| Renal impairment | 6.4 | 6.9 | -0.02 | 6.4 | 6.4 | 0.00 | 6.9 | 6.8 | 0.00 |
| Rheumatoid arthritis | 0.6 | 0.6 | -0.01 | 0.6 | 0.6 | -0.01 | 0.6 | 0.6 | 0.00 |
| Schizophrenia | 0.3 | 0.4 | -0.01 | 0.3 | 0.3 | 0.00 | 0.4 | 0.4 | 0.00 |
| Ulcerative colitis | 0.2 | 0.3 | -0.01 | 0.2 | 0.2 | -0.01 | 0.2 | 0.3 | -0.01 |
| Urinary tract infectious disease | 2.1 | 2.1 | 0.00 | 2.1 | 2.1 | 0.00 | 2.2 | 2.1 | 0.01 |
| Viral hepatitis C | 0.6 | 0.8 | -0.02 | 0.6 | 0.6 | 0.00 | 0.7 | 0.7 | -0.01 |
| Visual system disorder | 11.7 | 13.5 | -0.05 | 11.7 | 11.7 | 0.00 | 13.2 | 13.0 | 0.00 |
| **Medical history: Cardiovascular disease** | | | | | | | | | |
| Atrial fibrillation | 4.9 | 5.1 | -0.01 | 4.9 | 4.8 | 0.00 | 5.3 | 5.1 | 0.01 |
| Cerebrovascular disease | 2.8 | 2.8 | 0.00 | 2.8 | 2.9 | -0.01 | 2.9 | 2.9 | 0.00 |
| Coronary arteriosclerosis | 13.6 | 13.4 | 0.01 | 13.4 | 13.5 | 0.00 | 13.5 | 13.5 | 0.00 |
| Heart disease | 23.3 | 23.0 | 0.01 | 22.9 | 23.0 | 0.00 | 23.5 | 23.1 | 0.01 |
| Heart failure | 3.5 | 3.8 | -0.02 | 3.4 | 3.4 | 0.00 | 3.8 | 3.7 | 0.01 |
| Ischemic heart disease | 5.7 | 5.8 | 0.00 | 5.6 | 5.7 | 0.00 | 5.9 | 5.8 | 0.00 |
| Peripheral vascular disease | 4.7 | 4.7 | 0.00 | 4.6 | 4.6 | 0.00 | 4.8 | 4.7 | 0.01 |
| Pulmonary embolism | 0.5 | 0.6 | -0.02 | 0.5 | 0.5 | -0.01 | 0.6 | 0.6 | 0.00 |
| Venous thrombosis | 1.2 | 1.2 | 0.00 | 1.1 | 1.1 | 0.00 | 1.2 | 1.2 | 0.00 |
| **Medical history: Neoplasms** | | | | | | | | | |
| Hematologic neoplasm | 0.8 | 0.8 | 0.00 | 0.8 | 0.8 | -0.01 | 0.8 | 0.8 | 0.00 |

| (Continued) | Before ajustment | | | After matching | | | After stratification | | |
|---|---|---|---|---|---|---|---|---|---|
| Characteristic | T (%) | C (%) | StdDiff | T (%) | C (%) | StdDiff | T (%) | C (%) | StdDiff |
| Malignant lymphoma | 0.4 | 0.5 | -0.01 | 0.4 | 0.5 | -0.01 | 0.5 | 0.5 | -0.01 |
| Malignant neoplasm of anorectum | 0.1 | 0.1 | -0.01 | 0.1 | 0.1 | 0.00 | 0.1 | 0.1 | 0.00 |
| Malignant neoplastic disease | 6.5 | 6.7 | -0.01 | 6.5 | 6.5 | 0.00 | 6.7 | 6.7 | 0.00 |
| Malignant tumor of colon | 0.3 | 0.4 | -0.01 | 0.3 | 0.4 | -0.01 | 0.4 | 0.4 | -0.01 |
| Malignant tumor of urinary bladder | 0.4 | 0.5 | 0.00 | 0.4 | 0.4 | 0.00 | 0.4 | 0.5 | 0.00 |
| Primary malignant neoplasm of prostate | 2.3 | 2.4 | 0.00 | 2.4 | 2.4 | 0.00 | 2.4 | 2.4 | 0.00 |
| **Medication use** | | | | | | | | | |
| Agents acting on the renin-angiotensin system | 68.0 | 66.7 | 0.03 | 67.6 | 67.7 | 0.00 | 67.3 | 67.1 | 0.00 |
| Antibacterials for systemic use | 36.9 | 36.2 | 0.01 | 36.3 | 36.6 | 0.00 | 36.4 | 36.4 | 0.00 |
| Antidepressants | 20.3 | 20.9 | -0.02 | 20.1 | 20.1 | 0.00 | 21.1 | 20.7 | 0.01 |
| Antiepileptics | 13.1 | 13.7 | -0.02 | 13.1 | 13.1 | 0.00 | 14.0 | 13.5 | 0.01 |
| Antiinflammatory and antirheumatic products | 53.7 | 55.1 | -0.03 | 53.5 | 53.3 | 0.00 | 55.0 | 54.7 | 0.01 |
| Antineoplastic agents | 3.5 | 3.1 | 0.02 | 3.3 | 3.4 | -0.01 | 3.3 | 3.2 | 0.00 |
| Antipsoriatics | 0.5 | 0.4 | 0.02 | 0.5 | 0.4 | 0.01 | 0.5 | 0.4 | 0.01 |
| Antithrombotic agents | 42.3 | 44.1 | -0.04 | 42.2 | 42.0 | 0.00 | 44.2 | 43.5 | 0.01 |
| Beta blocking agents | 31.8 | 33.4 | -0.04 | 31.6 | 31.4 | 0.00 | 33.6 | 32.9 | 0.01 |
| Calcium channel blockers | 22.4 | 21.4 | 0.03 | 22.1 | 22.3 | -0.01 | 22.2 | 21.7 | 0.01 |
| Diuretics | 34.2 | 35.1 | -0.02 | 35.5 | 34.8 | 0.01 | 35.5 | 34.8 | 0.01 |
| Drugs for acid related disorders | 29.8 | 30.0 | 0.00 | 30.3 | 29.9 | 0.01 | 30.3 | 29.9 | 0.01 |
| Drugs for obstructive airway diseases | 34.9 | 34.4 | 0.01 | 34.5 | 34.6 | 0.00 | 34.8 | 34.6 | 0.00 |
| Immunosuppressants | 1.6 | 1.6 | 0.00 | 1.6 | 1.6 | 0.00 | 1.7 | 1.6 | 0.00 |
| Lipid modifying agents | 73.3 | 71.0 | 0.05 | 72.7 | 73.0 | -0.01 | 71.7 | 71.7 | 0.00 |
| Opioids | 23.0 | 24.6 | -0.04 | 22.9 | 22.8 | 0.00 | 24.5 | 24.1 | 0.01 |
| Psycholeptics | 22.9 | 22.4 | 0.01 | 22.6 | 22.7 | 0.00 | 22.8 | 22.6 | 0.01 |
| Psychostimulants | 2.3 | 1.9 | 0.03 | 2.2 | 2.1 | 0.00 | 2.0 | 2.0 | 0.00 |

### 2.6.8 VA

##### 2.6.9 SIDIAP

**Supplementary Table 199:** Baseline patient characteristics for DPP4I (male) (T) and SU (male) (C) new-users in the SIDIAP data source. We report proportion of initiators satisfying selected base-line characteristics and the standardized difference of population proportions (StdDiff) before and after propensity score adjustment. Less extreme StdDiffs through matching and stratification suggest improved balance between patient cohorts through adjustment.

| | Before adjustment | | | After matching | | | After stratification | | |
|---|---|---|---|---|---|---|---|---|---|
| Characteristic | T (%) | C (%) | StdDiff | T (%) | C (%) | StdDiff | T (%) | C (%) | StdDiff |
| **Age group** | | | | | | | | | |
| 25 - 29 | 0.1 | 0.1 | -0.01 | <0.1 | 0.1 | -0.01 | <0.1 | 0.1 | -0.01 |
| 30 - 34 | 0.3 | 0.3 | -0.01 | 0.4 | 0.4 | 0.01 | 0.4 | 0.3 | 0.01 |
| 35 - 39 | 1.0 | 1.2 | -0.02 | 1.4 | 1.3 | 0.01 | 1.1 | 1.1 | 0.00 |
| 40 - 44 | 2.6 | 3.7 | -0.07 | 3.5 | 3.6 | -0.01 | 2.9 | 3.5 | -0.03 |
| 45 - 49 | 5.7 | 7.3 | -0.06 | 7.6 | 7.3 | 0.01 | 6.2 | 6.9 | -0.03 |
| 50 - 54 | 9.5 | 11.1 | -0.05 | 10.8 | 11.0 | -0.01 | 10.0 | 10.0 | 0.00 |
| 55 - 59 | 13.2 | 15.1 | -0.05 | 15.0 | 15.1 | 0.00 | 13.7 | 14.1 | -0.01 |
| 60 - 64 | 15.6 | 17.4 | -0.05 | 17.2 | 17.0 | 0.00 | 15.9 | 15.8 | 0.00 |
| 65 - 69 | 15.4 | 15.5 | 0.00 | 15.3 | 15.8 | -0.01 | 15.5 | 16.2 | -0.02 |
| 70 - 74 | 13.9 | 12.5 | 0.04 | 12.4 | 12.2 | 0.00 | 13.4 | 13.7 | -0.01 |
| 75 - 79 | 10.7 | 8.6 | 0.07 | 8.5 | 8.6 | 0.00 | 10.1 | 9.4 | 0.02 |
| 80 - 84 | 6.9 | 4.7 | 0.09 | 4.9 | 4.9 | 0.00 | 6.3 | 5.6 | 0.03 |
| 85 - 89 | 4.0 | 2.0 | 0.12 | 2.4 | 2.1 | 0.02 | 3.4 | 2.7 | 0.04 |
| 90 - 94 | 1.0 | 0.5 | 0.06 | 0.6 | 0.5 | 0.01 | 0.9 | 0.6 | 0.04 |
| **Medical history: General** | | | | | | | | | |
| Acute respiratory disease | 10.9 | 9.3 | 0.06 | 9.4 | 9.7 | -0.01 | 10.6 | 10.9 | -0.01 |
| Chronic liver disease | 0.4 | 0.5 | -0.01 | 0.5 | 0.4 | 0.01 | 0.4 | 0.3 | 0.01 |
| Chronic obstructive lung disease | 1.9 | 2.2 | -0.02 | 2.0 | 2.2 | -0.01 | 2.0 | 2.0 | 0.00 |
| Dementia | 0.5 | 0.5 | -0.01 | 0.4 | 0.6 | -0.02 | 0.4 | 0.5 | -0.01 |
| Depressive disorder | 1.7 | 2.1 | -0.03 | 2.0 | 2.1 | -0.01 | 1.8 | 2.0 | -0.01 |
| Gastroesophageal reflux disease | 1.1 | 0.6 | 0.05 | 0.8 | 0.7 | 0.01 | 0.9 | 1.1 | -0.02 |
| Gastrointestinal hemorrhage | 1.8 | 1.5 | 0.03 | 1.9 | 1.7 | 0.01 | 1.8 | 1.6 | 0.02 |
| Hyperlipidemia | 3.8 | 5.0 | -0.06 | 5.1 | 5.0 | 0.00 | 4.1 | 4.5 | -0.02 |
| Hypertensive disorder | 4.5 | 6.7 | -0.10 | 6.2 | 6.2 | 0.00 | 5.0 | 5.5 | -0.02 |
| Lesion of liver | 0.5 | 0.6 | -0.01 | 0.6 | 0.6 | 0.00 | 0.5 | 0.7 | -0.02 |
| Obesity | 6.9 | 7.7 | -0.03 | 7.7 | 8.0 | -0.01 | 7.2 | 7.8 | -0.02 |
| Osteoarthritis | 3.5 | 3.8 | -0.02 | 3.6 | 4.0 | -0.02 | 3.6 | 4.0 | -0.02 |
| Pneumonia | 1.7 | 1.4 | 0.02 | 1.7 | 1.5 | 0.01 | 1.7 | 1.5 | 0.01 |
| Psoriasis | 0.4 | 0.6 | -0.03 | 0.5 | 0.7 | -0.03 | 0.4 | 0.6 | -0.03 |
| Renal impairment | 3.7 | 1.6 | 0.13 | 2.0 | 1.9 | 0.01 | 3.2 | 2.6 | 0.03 |
| Rheumatoid arthritis | 0.1 | 0.1 | -0.01 | <0.1 | 0.1 | -0.02 | 0.1 | 0.1 | 0.00 |
| Urinary tract infectious disease | 4.1 | 2.1 | 0.11 | 2.8 | 2.6 | 0.01 | 3.6 | 3.3 | 0.01 |
| Viral hepatitis C | 0.2 | 0.2 | 0.00 | 0.2 | 0.2 | 0.01 | 0.2 | 0.1 | 0.01 |
| Visual system disorder | 14.3 | 14.3 | 0.00 | 14.4 | 14.8 | -0.01 | 14.4 | 14.1 | 0.01 |
| **Medical history: Cardiovascular disease** | | | | | | | | | |
| Atrial fibrillation | 1.5 | 1.2 | 0.03 | 1.1 | 1.1 | 0.00 | 1.4 | 1.2 | 0.02 |
| Cerebrovascular disease | 1.2 | 1.2 | 0.00 | 1.1 | 1.3 | -0.01 | 1.2 | 1.2 | 0.00 |
| Heart disease | 9.8 | 8.7 | 0.04 | 9.5 | 9.1 | 0.01 | 9.5 | 9.1 | 0.01 |
| Heart failure | 1.7 | 1.3 | 0.03 | 1.5 | 1.5 | 0.00 | 1.6 | 1.4 | 0.01 |
| Ischemic heart disease | 3.3 | 3.5 | -0.01 | 3.5 | 3.5 | 0.00 | 3.3 | 3.4 | -0.01 |
| Peripheral vascular disease | 1.2 | 1.1 | 0.01 | 1.1 | 1.0 | 0.01 | 1.2 | 0.9 | 0.03 |
| Pulmonary embolism | 0.2 | 0.1 | 0.01 | 0.2 | 0.1 | 0.00 | 0.2 | 0.3 | -0.02 |
| Venous thrombosis | 0.8 | 0.8 | 0.00 | 0.9 | 0.8 | 0.00 | 0.8 | 0.9 | -0.01 |
| **Medical history: Neoplasms** | | | | | | | | | |
| Hematologic neoplasm | 0.1 | 0.1 | 0.00 | 0.1 | 0.1 | -0.02 | 0.1 | 0.1 | 0.01 |
| Malignant lymphoma | 0.1 | 0.1 | -0.01 | <0.1 | 0.1 | -0.02 | 0.1 | 0.1 | -0.01 |
| Malignant neoplasm of anorectum | 0.1 | 0.1 | 0.00 | 0.1 | 0.1 | -0.02 | 0.1 | 0.1 | 0.01 |
| Malignant neoplastic disease | 3.1 | 2.9 | 0.01 | 2.8 | 3.0 | -0.01 | 3.1 | 3.5 | -0.02 |
| Malignant tumor of colon | 0.4 | 0.4 | -0.01 | 0.4 | 0.4 | -0.01 | 0.4 | 0.3 | 0.01 |
| Malignant tumor of urinary bladder | 0.4 | 0.4 | 0.00 | 0.5 | 0.4 | 0.01 | 0.4 | 0.5 | -0.01 |
| Primary malignant neoplasm of prostate | 0.6 | 0.5 | 0.02 | 0.5 | 0.5 | 0.01 | 0.6 | 0.6 | 0.00 |
| **Medication use** | | | | | | | | | |
| Agents acting on the renin-angiotensin system | 61.2 | 54.4 | 0.14 | 56.5 | 56.7 | 0.00 | 59.7 | 58.1 | 0.03 |
| Antibacterials for systemic use | 34.9 | 28.9 | 0.13 | 32.0 | 31.7 | 0.01 | 33.8 | 34.4 | -0.01 |
| Antidepressants | 12.9 | 11.6 | 0.04 | 12.3 | 12.2 | 0.00 | 12.8 | 13.6 | -0.03 |
| Antiepileptics | 7.9 | 6.1 | 0.07 | 6.4 | 6.7 | -0.01 | 7.5 | 8.1 | -0.02 |
| Antiinflammatory and antirheumatic products | 51.4 | 51.5 | 0.00 | 52.6 | 53.0 | -0.01 | 51.7 | 51.6 | 0.00 |
| Antineoplastic agents | 1.6 | 1.2 | 0.04 | 1.5 | 1.6 | -0.01 | 1.5 | 1.6 | -0.01 |

| (Continued) | Before ajustment | | | After matching | | | After stratification | | |
|---|---|---|---|---|---|---|---|---|---|
| Characteristic | T (%) | C (%) | StdDiff | T (%) | C (%) | StdDiff | T (%) | C (%) | StdDiff |
| Antipsoriatics | 1.9 | 1.4 | 0.04 | 1.4 | 1.5 | -0.01 | 1.8 | 1.4 | 0.03 |
| Antithrombotic agents | 39.1 | 34.5 | 0.10 | 34.7 | 35.5 | -0.02 | 37.9 | 36.7 | 0.02 |
| Beta blocking agents | 26.3 | 21.1 | 0.12 | 21.6 | 22.3 | -0.02 | 24.9 | 24.6 | 0.01 |
| Calcium channel blockers | 24.7 | 17.9 | 0.17 | 19.4 | 19.7 | -0.01 | 23.0 | 22.5 | 0.01 |
| Diuretics | 40.2 | 35.5 | 0.10 | 35.7 | 36.3 | -0.01 | 39.0 | 37.7 | 0.03 |
| Drugs for acid related disorders | 46.0 | 40.5 | 0.11 | 42.3 | 42.8 | -0.01 | 44.6 | 44.5 | 0.00 |
| Drugs for obstructive airway diseases | 25.9 | 21.2 | 0.11 | 22.1 | 22.6 | -0.01 | 24.8 | 24.6 | 0.00 |
| Immunosuppressants | 1.2 | 0.8 | 0.04 | 0.8 | 0.9 | -0.01 | 1.1 | 1.0 | 0.01 |
| Lipid modifying agents | 64.7 | 57.7 | 0.14 | 60.6 | 61.3 | -0.01 | 63.3 | 63.2 | 0.00 |
| Opioids | 13.6 | 10.9 | 0.08 | 11.7 | 11.7 | 0.00 | 13.2 | 14.0 | -0.02 |
| Psycholeptics | 23.1 | 21.2 | 0.05 | 22.1 | 22.0 | 0.00 | 22.8 | 24.1 | -0.03 |
| Psychostimulants | 4.0 | 3.8 | 0.01 | 3.9 | 4.0 | 0.00 | 4.0 | 4.0 | 0.00 |

### 3 Cohort balance and systematic error diagnostics across data sources

#### 3.1 GLP1RA (male) vs SGLT2I (male)

##### 3.1.1 CCAE

**Supplementary Figure 1:** Preference score distributions, propensity score stratified (middle column) and matched (right column) covariate balance and negative control error distributions (bottom row) for GLP1RA (male) vs SGLT2I (male) in the CCAE data source.

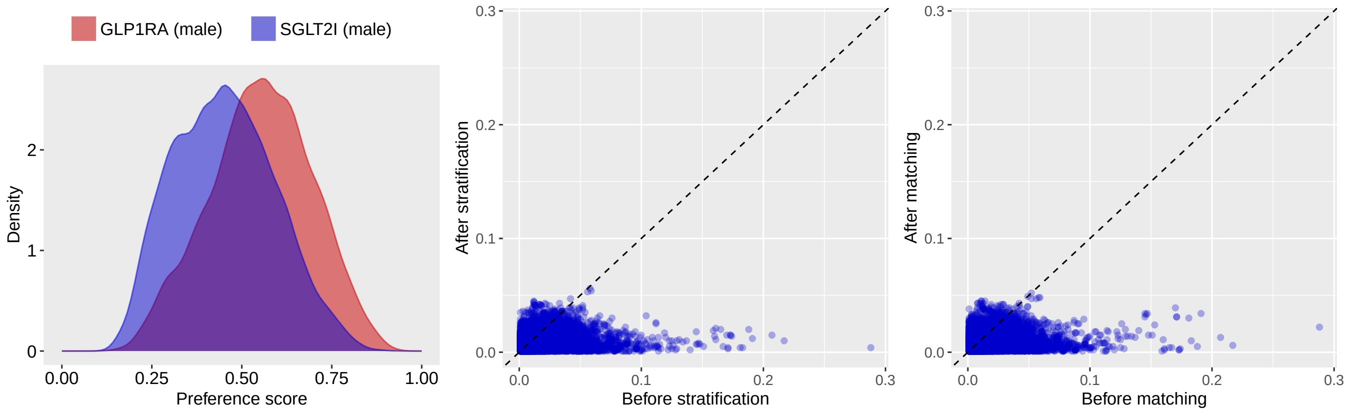

**Supplementary Table 200:** Most extreme patient characteristics balance diagnostics after stratification for GLP1RA (male) vs SGLT2I (male) in the CCAE data source.

| Characteristic (total count = 10415) | Before stratification | | | After stratification | | |
|---|---|---|---|---|---|---|
| | T (%) | C (%) | SDf | T (%) | C (%) | SDf |
| drug_era group during day -30 through 0 days relative to index: sacubitril | 0.1 | 0.4 | -0.06 | 0.1 | 0.4 | -0.06 |
| drug_era group during day -365 through 0 days relative to index: sacubitril | 0.2 | 0.5 | -0.06 | 0.2 | 0.5 | -0.05 |
| drug_era group during day 0 through 0 days relative to index: sacubitril | 0.1 | 0.4 | -0.06 | 0.1 | 0.4 | -0.05 |
| condition_era group during day -365 through 0 days relative to index: Mass of pancreas | 0.1 | 0.3 | -0.04 | 0.1 | 0.3 | -0.05 |
| measurement during day -30 through 0 days relative to index: Monocytes/100 leukocytes in Blood by Automated count | 0.6 | 0.7 | -0.01 | 1.0 | 0.6 | 0.04 |
| measurement during day -30 through 0 days relative to index: Lymphocytes/100 leukocytes in Blood by Automated count | 0.6 | 0.7 | -0.01 | 1.0 | 0.6 | 0.04 |

| (Continued) | T (%) | C (%) | SDf | T (%) | C (%) | SDf |
|---|---|---|---|---|---|---|
| measurement during day -30 through 0 days relative to index: Neutrophils/100 leukocytes in Blood by Automated count | 0.6 | 0.7 | -0.01 | 1.0 | 0.6 | 0.04 |
| measurement below normal range during day -365 through 0 days relative to index: Cholesterol [Mass/volume] in Serum or Plasma | 0.2 | 0.1 | 0.03 | 0.3 | 0.1 | 0.04 |
| measurement during day -30 through 0 days relative to index: Leukocytes [#/volume] in Blood by Automated count | 0.7 | 0.9 | -0.02 | 1.2 | 0.8 | 0.04 |
| condition_era group during day -365 through 0 days relative to index: Pelvic abscess | 0.2 | 0.3 | -0.02 | 0.1 | 0.3 | -0.04 |

**Supplementary Table 201:** Most extreme patient characteristics balance diagnostics after matching for GLP1RA (male) vs SGLT2I (male) in the CCAE data source.

| | Before matching | | | After matching | | |
|---|---|---|---|---|---|---|
| Characteristic (total count = 10547) | T (%) | C (%) | SDf | T (%) | C (%) | SDf |
| condition_era group during day -365 through 0 days relative to index: Acute pancreatitis | 0.3 | 0.6 | -0.05 | 0.2 | 0.6 | -0.05 |
| condition_era group during day -365 through 0 days relative to index: Combined systolic and diastolic dysfunction | 0.1 | 0.4 | -0.05 | 0.2 | 0.5 | -0.05 |
| drug_era group during day -365 through 0 days relative to index: sacubitril | 0.2 | 0.5 | -0.06 | 0.2 | 0.5 | -0.05 |
| drug_era group during day -30 through 0 days relative to index: sacubitril | 0.1 | 0.4 | -0.06 | 0.2 | 0.4 | -0.05 |
| drug_era group during day 0 through 0 days relative to index: sacubitril | 0.1 | 0.4 | -0.06 | 0.2 | 0.4 | -0.05 |
| condition_era group during day -365 through 0 days relative to index: Pancreatitis | 0.3 | 0.7 | -0.05 | 0.3 | 0.6 | -0.05 |
| condition_era group during day -365 through 0 days relative to index: Acute hypoxemic respiratory failure | 0.3 | 0.4 | -0.01 | 0.3 | 0.6 | -0.04 |
| condition_era group during day -365 through 0 days relative to index: Amnesia | 0.4 | 0.5 | -0.01 | 0.3 | 0.6 | -0.04 |
| condition_era group during day -365 through 0 days relative to index: Memory impairment | 0.4 | 0.5 | -0.01 | 0.3 | 0.6 | -0.04 |
| condition_era group during day -365 through 0 days relative to index: Mass of pancreas | 0.1 | 0.3 | -0.04 | 0.1 | 0.3 | -0.04 |

##### 3.1.2 ODOD

**Supplementary Figure 2:** Preference score distributions, propensity score stratified (middle column) and matched (right column) covariate balance and negative control error distributions (bottom row) for GLP1RA (male) vs SGLT2I (male) in the ODOD data source.

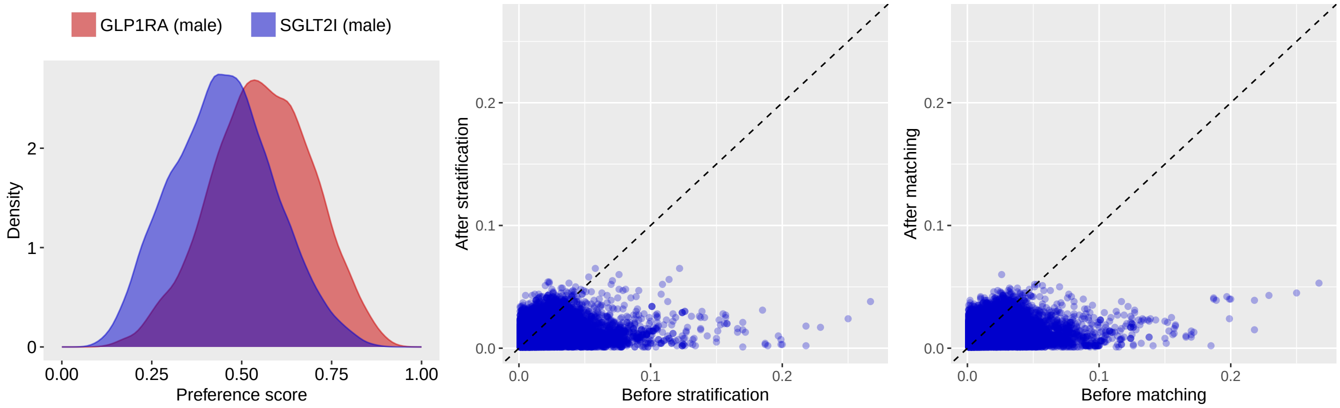

**Supplementary Table 202:** Most extreme patient characteristics balance diagnostics after stratification for GLP1RA (male) vs SGLT2I (male) in the ODOD data source.

| | Before stratification | | | After stratification | | |
|---|---|---|---|---|---|---|
| Characteristic (total count = 16349) | T (%) | C (%) | SDf | T (%) | C (%) | SDf |
| drug_era group during day -365 through 0 days relative to index: sacubitril | 0.4 | 1.7 | -0.12 | 0.7 | 1.4 | -0.07 |
| condition_era group during day -30 through 0 days relative to index: Atelectasis | 0.2 | 0.6 | -0.06 | 0.2 | 0.5 | -0.07 |
| drug_era group during day -30 through 0 days relative to index: prasugrel | 0.2 | 0.6 | -0.08 | 0.2 | 0.5 | -0.06 |
| condition_era group during day -30 through 0 days relative to index: Pancreatitis | -0.1 | 0.2 | -0.05 | -0.1 | 0.2 | -0.06 |
| drug_era group during day -30 through 0 days relative to index: sacubitril | 0.4 | 1.5 | -0.11 | 0.7 | 1.2 | -0.06 |
| drug_era group during day 0 through 0 days relative to index: prasugrel | 0.2 | 0.6 | -0.07 | 0.2 | 0.5 | -0.06 |

| (Continued) | T (%) | C (%) | SDf | T (%) | C (%) | SDf |
|---|---|---|---|---|---|---|
| procedure_occurrence during day -30 through 0 days relative to index: Transitional Care Management Services with the following required elements: Communication (direct contact, telephone, electronic) with the patient and/or caregiver within 2 business days of discharge Medical decision making of at least moderate complexity | 0.3 | 0.2 | 0.02 | 0.5 | 0.2 | 0.05 |
| observation during day -365 through 0 days relative to index: Ambulance (als or bls) oxygen and oxygen supplies, life sustaining situation | 0.2 | 0.1 | 0.02 | 0.4 | 0.1 | 0.05 |
| measurement during day -365 through 0 days relative to index: Infectious agent antigen detection by immunoassay technique, (eg, enzyme immunoassay [EIA], enzyme-linked immunosorbent assay [ELISA], immunochemiluminometric assay [IMCA]), qualitative or semiquantitative; multiple-step method, not otherwise specified, e | 0.4 | 0.2 | 0.04 | 0.5 | 0.2 | 0.05 |
| condition_era group during day -30 through 0 days relative to index: Infection of skin and/or subcutaneous tissue | 0.8 | 1.0 | -0.02 | 0.6 | 1.1 | -0.05 |

**Supplementary Table 203:** Most extreme patient characteristics balance diagnostics after matching for GLP1RA (male) vs SGLT2I (male) in the ODOD data source.

| | Before matching | | | After matching | | |
|---|---|---|---|---|---|---|
| Characteristic (total count = 15880) | T (%) | C (%) | SDf | T (%) | C (%) | SDf |
| procedure_occurrence during day -365 through 0 days relative to index: Magnetic resonance (eg, proton) imaging, abdomen; without contrast material(s), followed by with contrast material(s) and further sequences | 0.4 | 0.6 | -0.03 | 0.3 | 0.8 | -0.06 |
| condition_era group during day -365 through 0 days relative to index: Obesity | 41.6 | 28.9 | 0.27 | 38.9 | 41.5 | -0.05 |
| condition_era group during day -30 through 0 days relative to index: Pancreatitis | -0.1 | 0.2 | -0.05 | -0.1 | 0.2 | -0.05 |
| condition_era group during day -365 through 0 days relative to index: Pancreatitis | 0.3 | 0.7 | -0.05 | 0.4 | 0.8 | -0.05 |
| condition_era group during day -30 through 0 days relative to index: Diarrhea | 0.7 | 1.2 | -0.06 | 0.7 | 1.2 | -0.05 |
| age group: 85 - 89 | 0.3 | 0.9 | -0.07 | 0.4 | 0.7 | -0.05 |
| procedure_occurrence during day -365 through 0 days relative to index: Closure of the lacrimal punctum; by plug, each | -0.1 | 0.2 | -0.05 | -0.1 | 0.2 | -0.05 |
| measurement during day -365 through 0 days relative to index: Infectious agent antigen detection by immunoassay technique, (eg, enzyme immunoassay [EIA], enzyme-linked immunosorbent assay [ELISA], immunochemiluminometric assay [IMCA]), qualitative or semiquantitative; multiple-step method, not otherwise specified, e | 0.4 | 0.2 | 0.04 | 0.4 | 0.2 | 0.05 |
| condition_era group during day -30 through 0 days relative to index: Infection of foot | 0.3 | 0.5 | -0.03 | 0.3 | 0.6 | -0.05 |
| condition_era group during day -365 through 0 days relative to index: Chronic pancreatitis | 0.1 | 0.3 | -0.05 | 0.1 | 0.3 | -0.05 |

##### 3.1.3 MDCR

**Supplementary Figure 3:** Preference score distributions, propensity score stratified (middle column) and matched (right column) covariate balance and negative control error distributions (bottom row) for GLP1RA (male) vs SGLT2I (male) in the MDCR data source.

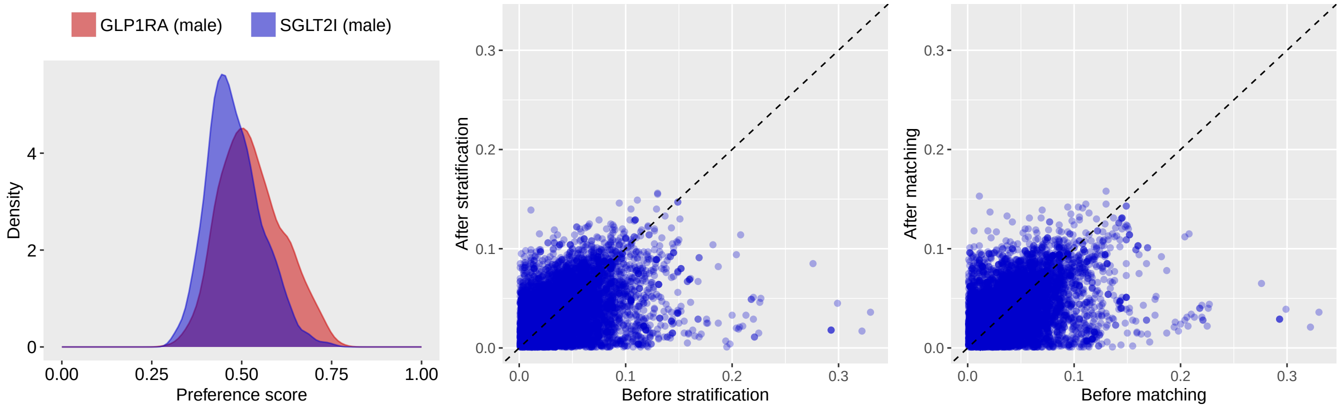

**Supplementary Table 204:** Most extreme patient characteristics balance diagnostics after stratification for GLP1RA (male) vs SGLT2I (male) in the MDCR data source.

| | Before stratification | | | After stratification | | |
|---|---|---|---|---|---|---|
| Characteristic (total count = 11997) | T (%) | C (%) | SDf | T (%) | C (%) | SDf |
| drug_era group during day -365 through 0 days relative to index: Other drugs for constipation | 1.6 | 0.3 | 0.13 | 2.0 | 0.3 | 0.16 |
| condition_era group during day -30 through 0 days relative to index: Metabolic syndrome X | 1.7 | 0.4 | 0.13 | 2.1 | 0.4 | 0.15 |
| condition_era group during day -365 through 0 days relative to index: Lesion of eyelid | 1.0 | 2.4 | -0.11 | -0.6 | 2.4 | -0.15 |
| condition_era group during day -365 through 0 days relative to index: Proteinuria | 1.4 | 3.7 | -0.15 | 1.5 | 3.9 | -0.15 |
| condition_era group during day -365 through 0 days relative to index: Abnormal presence of protein | 1.4 | 3.7 | -0.15 | 1.5 | 3.9 | -0.15 |
| condition_era group during day -365 through 0 days relative to index: Chronic fatigue syndrome | 1.3 | 0.4 | 0.09 | 2.0 | 0.4 | 0.15 |
| measurement during day -365 through 0 days relative to index: Uric acid; blood | 4.4 | 7.4 | -0.13 | 4.3 | 7.6 | -0.14 |
| drug_era group during day -365 through 0 days relative to index: linaclotide | 0.9 | -0.2 | 0.10 | 1.3 | -0.2 | 0.14 |

| (Continued) | T (%) | C (%) | SDf | T (%) | C (%) | SDf |
|---|---|---|---|---|---|---|
| index year: 2021 | 21.7 | 22.2 | -0.01 | 28.4 | 22.3 | 0.14 |
| observation during day -365 through 0 days relative to index: Bmi documented outside normal parameters, no follow-up plan documented, no reason given | -0.5 | 1.8 | -0.14 | -0.6 | 1.9 | -0.13 |

**Supplementary Table 205:** Most extreme patient characteristics balance diagnostics after matching for GLP1RA (male) vs SGLT2I (male) in the MDCR data source.

| | Before matching | | | After matching | | |
|---|---|---|---|---|---|---|
| Characteristic (total count = 11663) | T (%) | C (%) | SDf | T (%) | C (%) | SDf |
| condition_era group during day -30 through 0 days relative to index: Metabolic syndrome X | 1.7 | 0.4 | 0.13 | 2.3 | 0.5 | 0.16 |
| index year: 2021 | 21.7 | 22.2 | -0.01 | 29.2 | 22.6 | 0.15 |
| drug_era group during day -365 through 0 days relative to index: Other drugs for constipation | 1.6 | 0.3 | 0.13 | 1.9 | 0.3 | 0.14 |
| condition_era group during day -365 through 0 days relative to index: Lesion of eyelid | 1.0 | 2.4 | -0.11 | -0.7 | 2.3 | -0.14 |
| condition_era group during day -365 through 0 days relative to index: Proteinuria | 1.4 | 3.7 | -0.15 | 1.7 | 4.1 | -0.14 |
| condition_era group during day -365 through 0 days relative to index: Abnormal presence of protein | 1.4 | 3.7 | -0.15 | 1.7 | 4.1 | -0.14 |
| condition_era group during day -365 through 0 days relative to index: Metabolic syndrome X | 2.8 | 1.3 | 0.10 | 3.6 | 1.4 | 0.14 |
| condition_era group during day -365 through 0 days relative to index: Chronic fatigue syndrome | 1.3 | 0.4 | 0.09 | 1.9 | 0.4 | 0.14 |
| condition_era group during day -365 through 0 days relative to index: Left sided abdominal pain | 3.2 | 1.5 | 0.11 | 4.1 | 1.8 | 0.14 |
| condition_era group during day -365 through 0 days relative to index: Sleep related movement disorder | 1.7 | 2.7 | -0.07 | 1.4 | 3.6 | -0.14 |

##### 3.1.4 Open Claims

**Supplementary Figure 4:** Preference score distributions, propensity score stratified (middle column) and matched (right column) covariate balance and negative control error distributions (bottom row) for GLP1RA (male) vs SGLT2I (male) in the Open Claims data source.

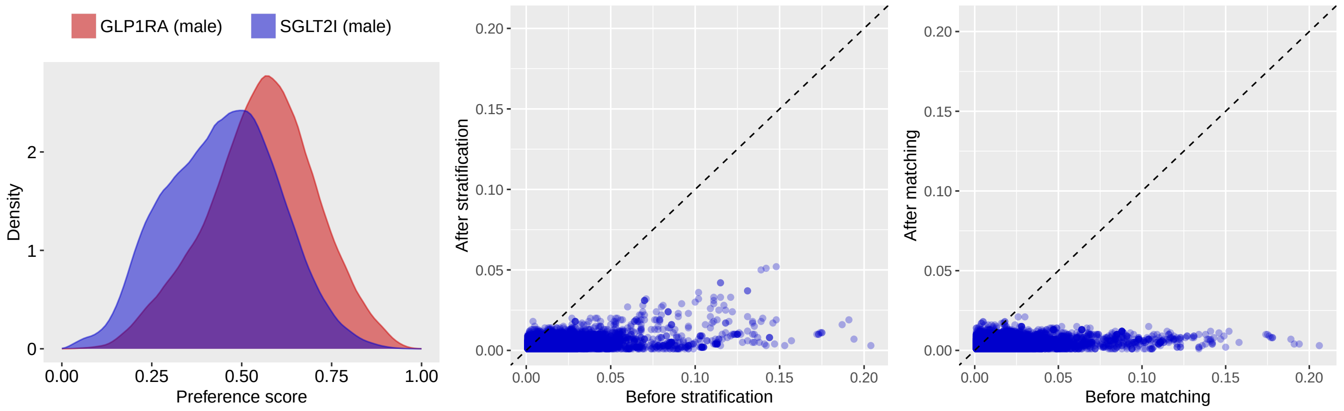

**Supplementary Table 206:** Most extreme patient characteristics balance diagnostics after stratification for GLP1RA (male) vs SGLT2I (male) in the Open Claims data source.

| | Before stratification | | | After stratification | | |
|---|---|---|---|---|---|---|
| Characteristic (total count = 9270) | T (%) | C (%) | SDf | T (%) | C (%) | SDf |
| drug_era group during day -365 through 0 days relative to index: sacubitril | 0.4 | 2.0 | -0.15 | 1.0 | 1.6 | -0.05 |
| drug_era group during day -30 through 0 days relative to index: sacubitril | 0.3 | 1.8 | -0.14 | 0.8 | 1.4 | -0.05 |
| drug_era group during day 0 through 0 days relative to index: sacubitril | 0.3 | 1.7 | -0.14 | 0.8 | 1.3 | -0.05 |
| condition_era group during day -30 through 0 days relative to index: Systolic dysfunction | 0.3 | 1.4 | -0.12 | 0.7 | 1.1 | -0.04 |
| condition_era group during day -30 through 0 days relative to index: Systolic heart failure | 0.3 | 1.4 | -0.12 | 0.7 | 1.1 | -0.04 |
| condition_era group during day -365 through 0 days relative to index: Systolic dysfunction | 1.0 | 2.8 | -0.13 | 1.8 | 2.3 | -0.04 |
| condition_era group during day -365 through 0 days relative to index: Systolic heart failure | 1.0 | 2.8 | -0.13 | 1.7 | 2.3 | -0.04 |
| condition_era group during day -30 through 0 days relative to index: Chronic systolic heart failure | 0.3 | 1.1 | -0.10 | 0.6 | 0.9 | -0.04 |

| (Continued) | T (%) | C (%) | SDf | T (%) | C (%) | SDf |
|---|---|---|---|---|---|---|
| condition_era group during day -365 through 0 days relative to index: Chronic systolic heart failure | 0.8 | 2.3 | -0.12 | 1.4 | 1.8 | -0.03 |
| condition_era group during day -30 through 0 days relative to index: Heart failure | 1.0 | 2.5 | -0.12 | 1.7 | 2.1 | -0.03 |

**Supplementary Table 207:** Most extreme patient characteristics balance diagnostics after matching for GLP1RA (male) vs SGLT2I (male) in the Open Claims data source.

| | Before matching | | | After matching | | |
|---|---|---|---|---|---|---|
| Characteristic (total count = 9162) | T (%) | C (%) | SDf | T (%) | C (%) | SDf |
| age group: 15 - 19 | 0.2 | 0.1 | 0.03 | 0.2 | 0.1 | 0.02 |
| observation during day -365 through 0 days relative to index: Coordinated care fee, maintenance rate | 0.1 | 0.1 | 0.03 | 0.1 | 0.1 | 0.02 |
| drug_era group during day -365 through 0 days relative to index: VITAMIN A AND D, INCL. COMBINATIONS OF THE TWO | 28.4 | 27.6 | 0.02 | 27.6 | 28.4 | -0.02 |
| drug_era group during day -365 through 0 days relative to index: Vitamin D and analogues | 28.4 | 27.6 | 0.02 | 27.6 | 28.4 | -0.02 |
| drug_era group during day -365 through 0 days relative to index: Electrolyte solutions | 13.7 | 13.9 | -0.01 | 13.4 | 14.0 | -0.02 |
| drug_era group during day -365 through 0 days relative to index: I.V. SOLUTION ADDITIVES | 13.7 | 13.9 | -0.01 | 13.4 | 14.0 | -0.02 |
| drug_era group during day -365 through 0 days relative to index: DIURETICS AND POTASSIUM-SPARING AGENTS IN COMBINATION | 40.8 | 39.9 | 0.02 | 40.1 | 40.9 | -0.02 |
| device_exposure during day -365 through 0 days relative to index: EASY COMFORT 30G LANCETS | 0.1 | 0.1 | -0.02 | 0.1 | 0.1 | -0.02 |
| drug_era group during day -365 through 0 days relative to index: POTASSIUM | 9.2 | 9.4 | -0.00 | 9.0 | 9.5 | -0.02 |
| drug_era group during day -365 through 0 days relative to index: Potassium | 9.2 | 9.4 | -0.00 | 9.0 | 9.5 | -0.02 |

##### 3.1.5 OEHR

**Supplementary Figure 5:** Preference score distributions, propensity score stratified (middle column) and matched (right column) covariate balance and negative control error distributions (bottom row) for GLP1RA (male) vs SGLT2I (male) in the OEHR data source.

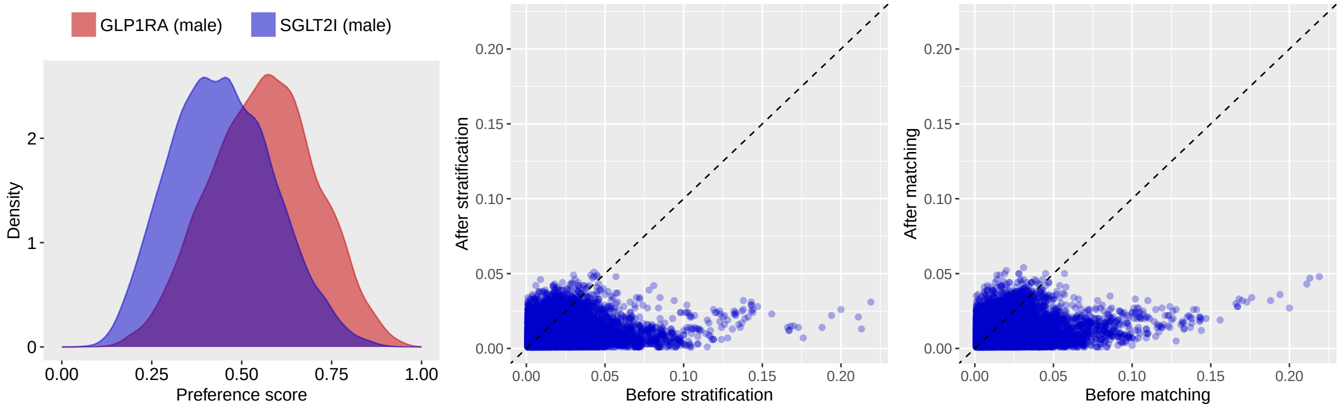

**Supplementary Table 208:** Most extreme patient characteristics balance diagnostics after stratification for GLP1RA (male) vs SGLT2I (male) in the OEHR data source.

| | Before stratification | | | After stratification | | |
|---|---|---|---|---|---|---|
| Characteristic (total count = 11289) | T (%) | C (%) | SDf | T (%) | C (%) | SDf |
| measurement above normal range during day -365 through 0 days relative to index: Fibrin D-dimer FEU [Mass/volume] in Platelet poor plasma | 0.1 | 0.4 | -0.04 | 0.1 | 0.4 | -0.05 |
| observation during day -365 through 0 days relative to index: Most recent LDL-C greater than or equal to 130 mg/dL (CAD) (DM) | 0.1 | 0.3 | -0.04 | 0.1 | 0.3 | -0.05 |
| condition_era group during day -365 through 0 days relative to index: Mass of pancreas | 0.1 | 0.2 | -0.04 | 0.1 | 0.2 | -0.05 |
| procedure_occurrence during day -30 through 0 days relative to index: Noninvasive ear or pulse oximetry for oxygen saturation; single determination | 1.0 | 0.7 | 0.03 | 1.2 | 0.7 | 0.05 |
| procedure_occurrence during day -365 through 0 days relative to index: Ambulatory continuous glucose monitoring of interstitial tissue fluid via a subcutaneous sensor for a minimum of 72 hours; analysis, interpretation and report | 0.3 | 0.1 | 0.06 | 0.3 | 0.1 | 0.05 |
| condition_era group during day -365 through 0 days relative to index: Idiopathic peripheral neuropathy | 0.4 | 0.1 | 0.04 | 0.4 | 0.2 | 0.05 |

| (Continued) | T (%) | C (%) | SDf | T (%) | C (%) | SDf |
|---|---|---|---|---|---|---|
| measurement during day -365 through 0 days relative to index: Serum uric acid measurement | 0.3 | 0.6 | -0.04 | 0.3 | 0.6 | -0.05 |
| condition_era group during day -30 through 0 days relative to index: Urinary tract infectious disease | 0.5 | 0.2 | 0.04 | 0.5 | 0.2 | 0.05 |
| condition_era group during day -30 through 0 days relative to index: Constipation | 0.6 | 0.4 | 0.03 | 0.7 | 0.4 | 0.05 |
| device_exposure during day -365 through 0 days relative to index: ONE TOUCH DELICA 33G LANCETS | 0.4 | 0.3 | 0.01 | 0.6 | 0.3 | 0.05 |

**Supplementary Table 209:** Most extreme patient characteristics balance diagnostics after matching for GLP1RA (male) vs SGLT2I (male) in the OEHR data source.

| | Before matching | | | After matching | | |
|---|---|---|---|---|---|---|
| Characteristic (total count = 11982) | T (%) | C (%) | SDf | T (%) | C (%) | SDf |
| condition_era group during day -365 through 0 days relative to index: Mass of axilla | -0.1 | 0.1 | -0.03 | -0.1 | 0.2 | -0.05 |
| condition_era group during day -365 through 0 days relative to index: Respiratory failure | 0.4 | 0.6 | -0.02 | 0.4 | 0.7 | -0.05 |
| procedure_occurrence during day -365 through 0 days relative to index: Ambulatory continuous glucose monitoring of interstitial tissue fluid via a subcutaneous sensor for a minimum of 72 hours; analysis, interpretation and report | 0.3 | 0.1 | 0.06 | 0.3 | 0.1 | 0.05 |
| observation during day -365 through 0 days relative to index: Most recent LDL-C greater than or equal to 130 mg/dL (CAD) (DM) | 0.1 | 0.3 | -0.04 | 0.1 | 0.4 | -0.05 |
| condition_era group during day -365 through 0 days relative to index: Abscess of axilla | -0.1 | 0.1 | -0.03 | -0.1 | 0.2 | -0.05 |
| measurement during day -30 through 0 days relative to index: Gases, blood, any combination of pH, pCO2, pO2, CO2, HCO3 (including calculated O2 saturation) | 0.1 | 0.2 | -0.03 | 0.1 | 0.3 | -0.05 |
| condition_era group during day -365 through 0 days relative to index: Respiratory insufficiency | 0.4 | 0.6 | -0.02 | 0.4 | 0.7 | -0.05 |
| condition_era group during day -365 through 0 days relative to index: Acute respiratory failure | 0.3 | 0.4 | -0.02 | 0.2 | 0.5 | -0.05 |
| condition_era group during day -365 through 0 days relative to index: Hypoxemic respiratory failure | 0.4 | 0.4 | -0.01 | 0.3 | 0.6 | -0.05 |
| condition_era group during day -365 through 0 days relative to index: Obesity | 41.8 | 31.3 | 0.22 | 39.0 | 41.4 | -0.05 |

### 3.1.6 VA

**Supplementary Figure 6:** Preference score distributions, propensity score stratified (middle column) and matched (right column) covariate balance and negative control error distributions (bottom row) for GLP1RA (male) vs SGLT2I (male) in the VA data source.

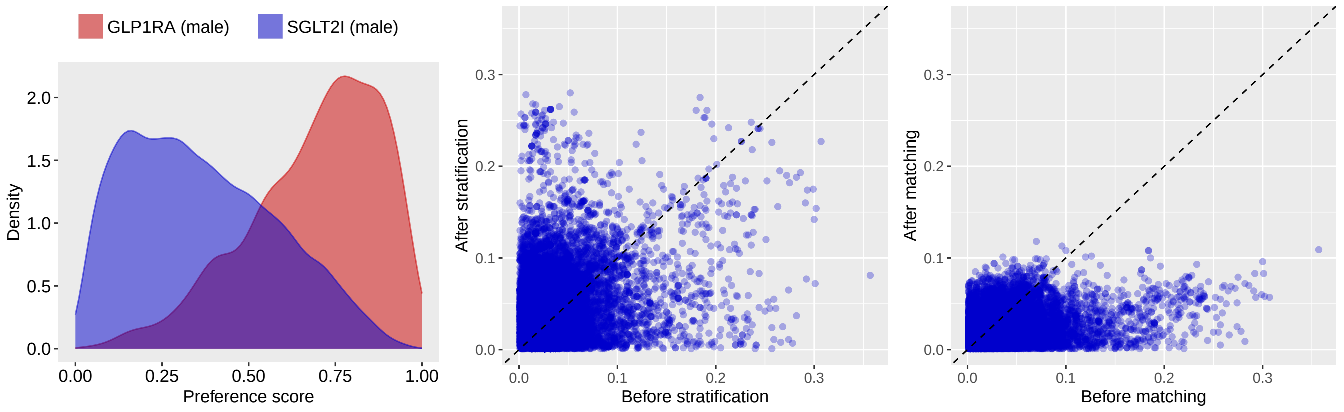

**Supplementary Table 210:** Most extreme patient characteristics balance diagnostics after stratification for GLP1RA (male) vs SGLT2I (male) in the VA data source.

| Characteristic (total count = 16665) | Before stratification | | | After stratification | | |
|---|---|---|---|---|---|---|
| | T (%) | C (%) | SDf | T (%) | C (%) | SDf |
| measurement during day -30 through 0 days relative to index: General health panel This panel must include the following: Comprehensive metabolic panel (80053) Blood count, complete (CBC), automated and automated differential WBC count (85025 or 85027 and 85004) OR Blood count, complete (CBC), automated (85027) and | 1.9 | 2.7 | -0.05 | 9.2 | 2.6 | 0.28 |
| measurement during day -30 through 0 days relative to index: Segmented neutrophils/100 leukocytes in Blood by Automated count | -0.7 | 0.6 | 0.01 | 5.1 | 0.6 | 0.28 |
| condition_era group during day -365 through 0 days relative to index: Atherosclerosis of coronary artery without angina pectoris | 20.6 | 28.4 | -0.18 | 40.5 | 27.6 | 0.28 |
| observation during day -30 through 0 days relative to index: Educational problem | -0.7 | 0.2 | -0.01 | 4.0 | 0.2 | 0.27 |
| procedure_occurrence during day -365 through 0 days relative to index: Brief communication technology-based service, e.g. virtual check-in, by a physician or other qualified health care professional who can report evaluation and management services, provided to an established patient, not originating from a related e/m se... | -0.7 | 0.2 | -0.02 | 4.0 | 0.2 | 0.27 |

| (Continued) | T (%) | C (%) | SDf | T (%) | C (%) | SDf |
|---|---|---|---|---|---|---|
| measurement above normal range during day -365 through 0 days relative to index: Calcium [Mass/volume] in Body fluid | -0.7 | -0.1 | 0.04 | 3.5 | -0.1 | 0.27 |
| condition_era group during day -30 through 0 days relative to index: Preputial adhesions | -0.7 | -0.1 | 0.03 | 3.3 | -0.1 | 0.26 |
| condition_era group during day -30 through 0 days relative to index: Lesion of penis | -0.7 | -0.1 | 0.03 | 3.3 | -0.1 | 0.26 |
| condition_era group during day -30 through 0 days relative to index: Lesion of prepuce | -0.7 | -0.1 | 0.03 | 3.3 | -0.1 | 0.26 |
| condition_era group during day -30 through 0 days relative to index: Phimosis | -0.7 | -0.1 | 0.03 | 3.3 | -0.1 | 0.26 |

**Supplementary Table 211:** Most extreme patient characteristics balance diagnostics after matching for GLP1RA (male) vs SGLT2I (male) in the VA data source.

| | Before matching | | | After matching | | |
|---|---|---|---|---|---|---|
| Characteristic (total count = 12830) | T (%) | C (%) | SDf | T (%) | C (%) | SDf |
| measurement below normal range during day -365 through 0 days relative to index: Anion gap 3 in Serum or Plasma | 1.1 | 0.5 | 0.07 | 1.3 | 0.2 | 0.12 |
| measurement during day -30 through 0 days relative to index: Globulin [Mass/volume] in Serum | -0.7 | 0.8 | -0.10 | -0.8 | 0.9 | -0.11 |
| condition_era group during day -30 through 0 days relative to index: Obesity | 30.8 | 15.9 | 0.36 | 24.6 | 29.5 | -0.11 |
| drug_era group during day -30 through 0 days relative to index: LIPID MODIFYING AGENTS | 67.3 | 75.6 | -0.18 | 68.4 | 63.3 | 0.11 |
| drug_era group during day -30 through 0 days relative to index: LIPID MODIFYING AGENTS, PLAIN | 67.3 | 75.6 | -0.18 | 68.4 | 63.3 | 0.11 |
| device_exposure during day -30 through 0 days relative to index: BD HOME SHARPS CONTAINER | 1.2 | 0.3 | 0.10 | 1.3 | 0.3 | 0.11 |
| device_exposure during day -365 through 0 days relative to index: ACCU-CHEK SOFTCLIX LANCETS | 1.3 | 0.2 | 0.13 | 1.1 | 0.3 | 0.10 |
| measurement during day -365 through 0 days relative to index: Calcium.ionized [Moles/volume] in Arterial blood | -0.7 | 0.9 | -0.12 | -0.8 | 0.7 | -0.10 |
| procedure_occurrence during day -30 through 0 days relative to index: Online digital evaluation and management service, for an established patient, for up to 7 days, cumulative time during the 7 days; 5-10 minutes | -0.7 | 0.4 | -0.04 | -0.8 | 0.7 | -0.10 |
| drug_era group during day -30 through 0 days relative to index: HMG CoA reductase inhibitors in combination with other lipid modifying agents | 67.3 | 75.7 | -0.19 | 68.3 | 63.6 | 0.10 |

#### 3.2 SGLT2I (male) vs DPP4I (male)

##### 3.2.1 CCAE

**Supplementary Figure 7:** Preference score distributions, propensity score stratified (middle column) and matched (right column) covariate balance and negative control error distributions (bottom row) for SGLT2I (male) vs DPP4I (male) in the CCAE data source.

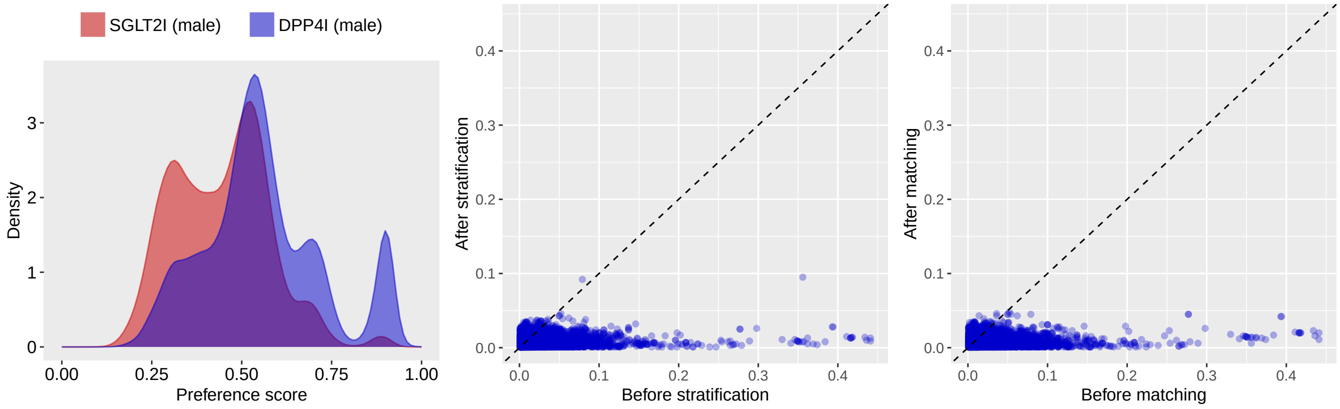

**Supplementary Table 212:** Most extreme patient characteristics balance diagnostics after stratification for SGLT2I (male) vs DPP4I (male) in the CCAE data source.

| | Before stratification | | | After stratification | | |
|---|---|---|---|---|---|---|
| Characteristic (total count = 10249) | T (%) | C (%) | SDf | T (%) | C (%) | SDf |
| index year: 2013 | 1.0 | 8.5 | 0.36 | 4.2 | 6.3 | 0.10 |
| index year: 2014 | 7.5 | 9.7 | 0.08 | 15.1 | 12.0 | -0.09 |
| condition_era group during day -30 through 0 days relative to index: Acute nephropathy | 0.2 | 0.5 | 0.05 | 0.2 | 0.5 | 0.05 |
| condition_era group during day -30 through 0 days relative to index: Acute renal failure syndrome | 0.2 | 0.5 | 0.05 | 0.2 | 0.5 | 0.04 |
| condition_era group during day -30 through 0 days relative to index: Acute renal impairment | 0.2 | 0.5 | 0.05 | 0.2 | 0.5 | 0.04 |
| condition_era group during day -30 through 0 days relative to index: Renal failure syndrome | 0.2 | 0.6 | 0.06 | 0.3 | 0.6 | 0.04 |
| condition_era group during day -30 through 0 days relative to index: Acute genitourinary disorder | 0.3 | 0.6 | 0.05 | 0.3 | 0.6 | 0.04 |
| condition_era group during day -365 through 0 days relative to index: Primary cardiomyopathy | 0.2 | 0.8 | 0.08 | 0.3 | 0.5 | 0.04 |

| (Continued) | T (%) | C (%) | SDf | T (%) | C (%) | SDf |
|---|---|---|---|---|---|---|
| condition_era group during day -365 through 0 days relative to index: Acute pancreatitis | 0.6 | 0.3 | -0.04 | 0.5 | 0.3 | -0.04 |
| condition_era group during day -30 through 0 days relative to index: Sepsis | 0.1 | 0.2 | 0.02 | 0.1 | 0.2 | 0.04 |

**Supplementary Table 213:** Most extreme patient characteristics balance diagnostics after matching for SGLT2I (male) vs DPP4I (male) in the CCAE data source.

| | Before matching | | | After matching | | |
|---|---|---|---|---|---|---|
| Characteristic (total count = 10289) | T (%) | C (%) | SDf | T (%) | C (%) | SDf |
| condition_era group during day -30 through 0 days relative to index: Acute nephropathy | 0.2 | 0.5 | 0.05 | 0.2 | 0.5 | 0.05 |
| condition_era group during day -365 through 0 days relative to index: Acute pancreatitis | 0.6 | 0.3 | -0.04 | 0.6 | 0.3 | -0.05 |
| condition_era group during day -30 through 0 days relative to index: Benign hypertension | 4.0 | 11.4 | 0.28 | 4.9 | 4.0 | -0.04 |
| condition_era group during day -30 through 0 days relative to index: Benign essential hypertension | 4.0 | 11.4 | 0.28 | 4.9 | 4.0 | -0.04 |
| index year: 2014 | 7.5 | 9.7 | 0.08 | 9.1 | 7.9 | -0.04 |
| condition_era group during day -30 through 0 days relative to index: Acute genitourinary disorder | 0.3 | 0.6 | 0.05 | 0.3 | 0.6 | 0.04 |
| condition_era group during day -30 through 0 days relative to index: Acute renal failure syndrome | 0.2 | 0.5 | 0.05 | 0.2 | 0.5 | 0.04 |
| condition_era group during day -30 through 0 days relative to index: Acute renal impairment | 0.2 | 0.5 | 0.05 | 0.2 | 0.5 | 0.04 |
| condition_era group during day -30 through 0 days relative to index: Renal failure syndrome | 0.2 | 0.6 | 0.06 | 0.3 | 0.5 | 0.04 |
| condition_era group during day -365 through 0 days relative to index: Pancreatitis | 0.7 | 0.4 | -0.04 | 0.7 | 0.4 | -0.04 |

##### 3.2.2 DAG

**Supplementary Figure 8:** Preference score distributions, propensity score stratified (middle column) and matched (right column) covariate balance and negative control error distributions (bottom row) for SGLT2I (male) vs DPP4I (male) in the DAG data source.

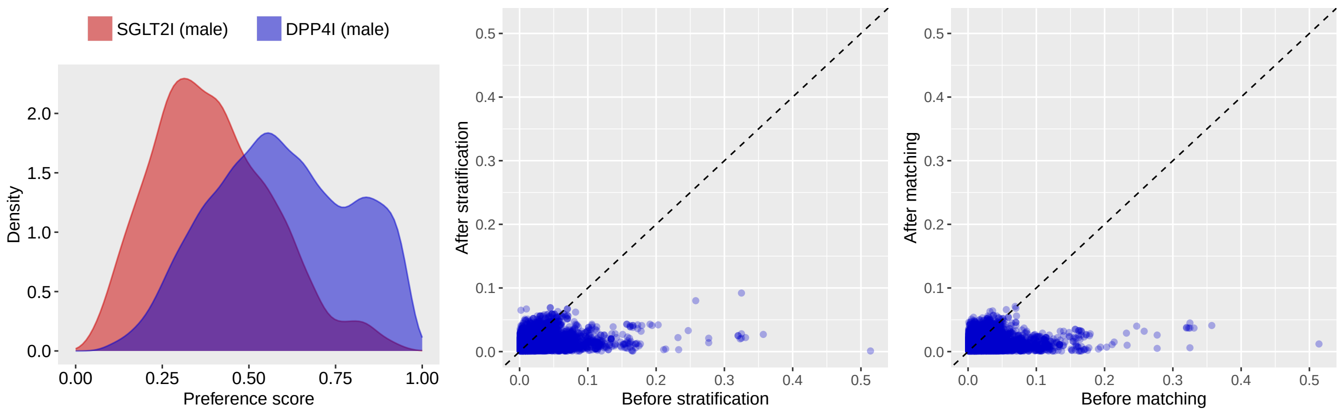

**Supplementary Table 214:** Most extreme patient characteristics balance diagnostics after stratification for SGLT2I (male) vs DPP4I (male) in the DAG data source.

| Characteristic (total count = 6426) | Before stratification | | | After stratification | | |
|---|---|---|---|---|---|---|
| | T (%) | C (%) | SDf | T (%) | C (%) | SDf |
| index year: 2013 | 0.4 | 6.1 | 0.33 | 2.9 | 4.7 | 0.09 |
| index year: 2015 | 2.4 | 8.0 | 0.26 | 9.3 | 7.1 | -0.08 |
| drug_era group during day -30 through 0 days relative to index: OTHER DRUGS FOR DISORDERS OF THE MUSCULO-SKELETAL SYSTEM | 0.7 | 0.3 | -0.04 | 0.9 | 0.3 | -0.07 |
| drug_era group during day -30 through 0 days relative to index: OTHER DRUGS FOR DISORDERS OF THE MUSCULO-SKELETAL SYSTEM | 0.7 | 0.3 | -0.04 | 0.9 | 0.3 | -0.07 |
| condition_era group during day -365 through 0 days relative to index: Anal fissure | -0.1 | 0.3 | 0.07 | -0.1 | 0.3 | 0.07 |
| drug_era group during day -30 through 0 days relative to index: enoxaparin | 0.2 | 0.6 | 0.07 | 0.2 | 0.6 | 0.07 |
| device_exposure during day -365 through 0 days relative to index: HEILMITTEL SONST. SONSTIGE L | 0.2 | 0.1 | -0.01 | 0.5 | 0.1 | -0.07 |
| device_exposure during day -365 through 0 days relative to index: HILFSMITTEL SONST. SONSTIGE T | 0.3 | 0.3 | 0.00 | 0.8 | 0.3 | -0.07 |

| (Continued) | T (%) | C (%) | SDf | T (%) | C (%) | SDf |
|---|---|---|---|---|---|---|
| age group: 90 - 94 | 0.2 | 0.6 | 0.06 | 0.2 | 0.6 | 0.06 |
| drug_era group during day -365 through 0 days relative to index: cefixime | -0.1 | 0.4 | 0.08 | -0.1 | 0.3 | 0.06 |

**Supplementary Table 215:** Most extreme patient characteristics balance diagnostics after matching for SGLT2I (male) vs DPP4I (male) in the DAG data source.

| | Before matching | | | After matching | | |
|---|---|---|---|---|---|---|
| Characteristic (total count = 6496) | T (%) | C (%) | SDf | T (%) | C (%) | SDf |
| device_exposure during day -365 through 0 days relative to index: HILFSMITTEL SONST. SONSTIGE I | 0.2 | 0.7 | 0.07 | 0.3 | 0.8 | 0.07 |
| condition_era group during day -365 through 0 days relative to index: Anal fissure | -0.1 | 0.3 | 0.07 | -0.1 | 0.3 | 0.07 |
| age group: 90 - 94 | 0.2 | 0.6 | 0.06 | 0.2 | 0.7 | 0.07 |
| condition_era group during day -365 through 0 days relative to index: Cerebral hemorrhage | 0.2 | 0.1 | -0.04 | 0.3 | -0.1 | -0.06 |
| condition_era group during day -365 through 0 days relative to index: Spontaneous cerebral hemorrhage | 0.2 | 0.1 | -0.03 | 0.3 | -0.1 | -0.06 |
| drug_era group during day -30 through 0 days relative to index: SURGICAL AIDS | 0.2 | 0.1 | -0.04 | 0.2 | -0.1 | -0.06 |
| drug_era group during day -30 through 0 days relative to index: Viscoelastic substances | 0.2 | 0.1 | -0.04 | 0.2 | -0.1 | -0.06 |
| condition_era group during day -365 through 0 days relative to index: Inborn error of metabolism | -0.1 | 0.3 | 0.07 | -0.1 | 0.2 | 0.06 |
| device_exposure during day -365 through 0 days relative to index: ACCUCH.AVIV.II RDC GLU.PLAS.STR 50 | 0.3 | 0.1 | -0.04 | 0.3 | -0.1 | -0.06 |
| condition_era group during day -365 through 0 days relative to index: Valvular endocarditis | 0.3 | 0.1 | -0.03 | 0.3 | -0.1 | -0.06 |

##### 3.2.3 ODOD

**Supplementary Figure 9:** Preference score distributions, propensity score stratified (middle column) and matched (right column) covariate balance and negative control error distributions (bottom row) for SGLT2I (male) vs DPP4I (male) in the ODOD data source.

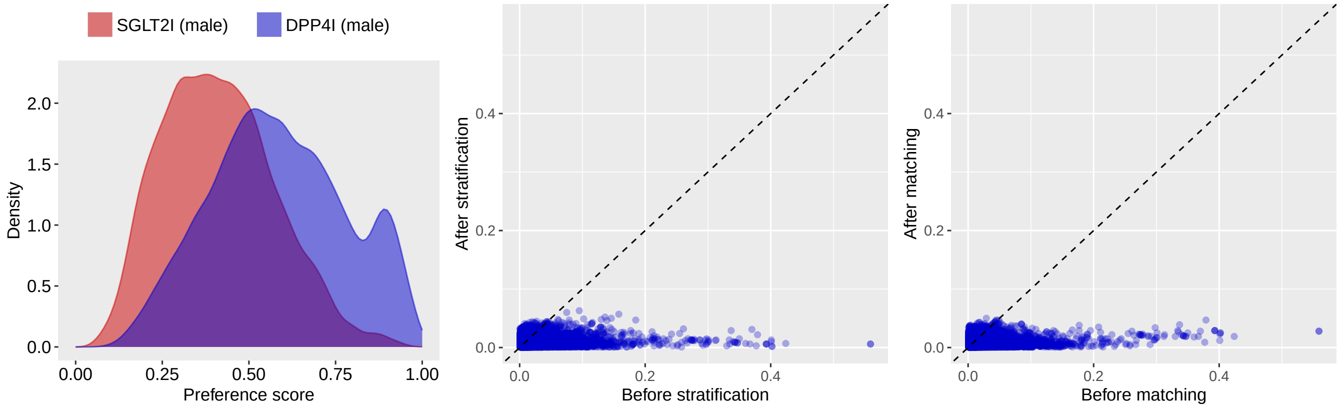

**Supplementary Table 216:** Most extreme patient characteristics balance diagnostics after stratification for SGLT2I (male) vs DPP4I (male) in the ODOD data source.

| | Before stratification | | | After stratification | | |
|---|---|---|---|---|---|---|
| Characteristic (total count = 14536) | T (%) | C (%) | SDf | T (%) | C (%) | SDf |
| procedure_occurrence during day -30 through 0 days relative to index: Ultrasound, retroperitoneal (eg, renal, aorta, nodes), real time with image documentation; complete | 0.2 | 1.0 | 0.10 | 0.3 | 0.7 | 0.06 |
| condition_era group during day -30 through 0 days relative to index: Chronic kidney disease stage 4 | 0.1 | 0.5 | 0.07 | 0.1 | 0.5 | 0.06 |
| procedure_occurrence during day -30 through 0 days relative to index: Radiologic examination, chest; single view, frontal | 0.2 | 1.8 | 0.16 | 0.6 | 1.1 | 0.06 |
| drug_era group during day -30 through 0 days relative to index: sacubitril | 1.5 | 0.2 | -0.14 | 0.9 | 0.5 | -0.05 |
| condition_era group during day -30 through 0 days relative to index: Renal failure syndrome | 0.7 | 2.6 | 0.15 | 1.4 | 2.0 | 0.05 |
| condition_era group during day -30 through 0 days relative to index: Abnormal breathing | 0.2 | 1.0 | 0.10 | 0.3 | 0.6 | 0.05 |
| drug_era group during day 0 through 0 days relative to index: sacubitril | 1.4 | 0.2 | -0.13 | 0.8 | 0.5 | -0.05 |

| (Continued) | T (%) | C (%) | SDf | T (%) | C (%) | SDf |
|---|---|---|---|---|---|---|
| observation during day -30 through 0 days relative to index: Blood Storage and Processing - General Classification | 0.1 | 0.5 | 0.06 | 0.1 | 0.4 | 0.05 |
| observation during day -365 through 0 days relative to index: Prescription(s) generated and transmitted via a qualified erx system | 0.2 | 2.1 | 0.18 | 0.6 | 1.0 | 0.05 |
| condition_era group during day -365 through 0 days relative to index: Chronic kidney disease stage 4 | 0.2 | 0.7 | 0.08 | 0.3 | 0.7 | 0.05 |

**Supplementary Table 217:** Most extreme patient characteristics balance diagnostics after matching for SGLT2I (male) vs DPP4I (male) in the ODOD data source.

| | Before matching | | | After matching | | |
|---|---|---|---|---|---|---|
| Characteristic (total count = 14241) | T (%) | C (%) | SDf | T (%) | C (%) | SDf |
| measurement during day -365 through 0 days relative to index: Thyroglobulin | 0.3 | 0.2 | -0.03 | 0.3 | 0.1 | -0.05 |
| procedure_occurrence during day -365 through 0 days relative to index: General examination of patient | 5.0 | 16.6 | 0.38 | 6.9 | 5.8 | -0.05 |
| condition_era group during day -365 through 0 days relative to index: Neoplasm of thorax | 0.3 | 0.6 | 0.05 | 0.3 | 0.6 | 0.05 |
| condition_era group during day -30 through 0 days relative to index: Neoplasm of thorax | 0.1 | 0.3 | 0.04 | 0.1 | 0.3 | 0.05 |
| condition_era group during day -30 through 0 days relative to index: Neoplasm of lower respiratory tract | 0.1 | 0.2 | 0.04 | 0.1 | 0.2 | 0.05 |
| condition_era group during day -30 through 0 days relative to index: Neoplasm of intrathoracic organs | 0.1 | 0.2 | 0.04 | 0.1 | 0.2 | 0.04 |
| condition_era group during day -30 through 0 days relative to index: Malignant neoplasm of lower respiratory tract | 0.1 | 0.2 | 0.04 | 0.1 | 0.2 | 0.04 |
| condition_era group during day -30 through 0 days relative to index: Tumor of respiratory system | 0.2 | 0.4 | 0.05 | 0.2 | 0.4 | 0.04 |
| condition_era group during day -30 through 0 days relative to index: Neoplasm of lung | 0.1 | 0.2 | 0.04 | 0.1 | 0.2 | 0.04 |
| measurement during day -365 through 0 days relative to index: Heterophile antibodies; screening | 0.2 | 0.1 | -0.02 | 0.2 | 0.1 | -0.04 |

##### 3.2.4 MDCR

**Supplementary Figure 10:** Preference score distributions, propensity score stratified (middle column) and matched (right column) covariate balance and negative control error distributions (bottom row) for SGLT2I (male) vs DPP4I (male) in the MDCR data source.

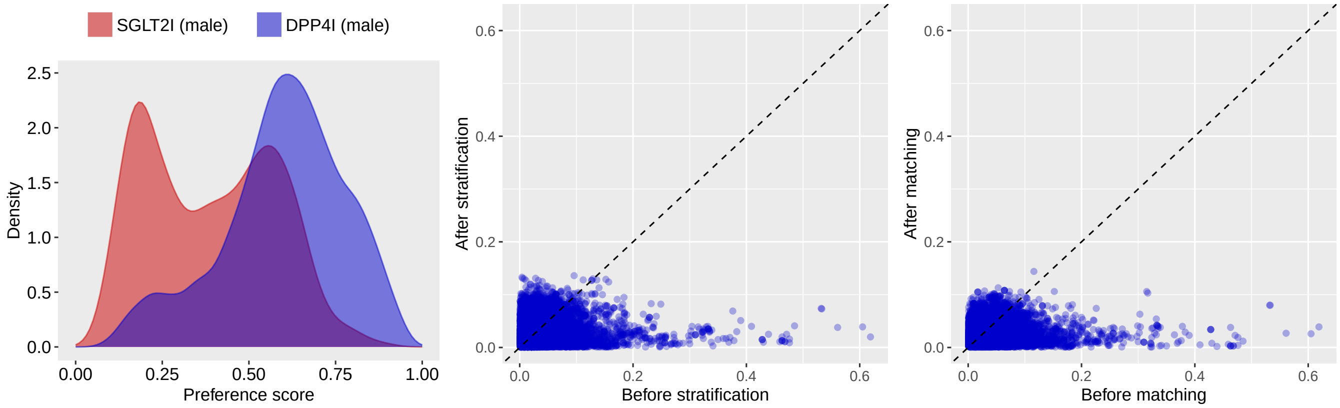

**Supplementary Table 218:** Most extreme patient characteristics balance diagnostics after stratification for SGLT2I (male) vs DPP4I (male) in the MDCR data source.

| Characteristic (total count = 16727) | Before stratification | | | After stratification | | |
|---|---|---|---|---|---|---|
| | T (%) | C (%) | SDf | T (%) | C (%) | SDf |
| condition_era group during day -365 through 0 days relative to index: Dementia | 1.1 | 2.4 | 0.10 | 0.9 | 2.6 | 0.14 |
| drug_era group during day -365 through 0 days relative to index: Belladonna alkaloids, tertiary amines | 1.7 | 1.7 | 0.00 | 3.6 | 1.5 | -0.13 |
| condition_era group during day -365 through 0 days relative to index: Herpes zoster without complication | 1.2 | 0.9 | -0.03 | 2.1 | 0.6 | -0.13 |
| drug_era group during day -30 through 0 days relative to index: Belladonna alkaloids, tertiary amines | 0.5 | 0.5 | 0.01 | 1.8 | 0.4 | -0.13 |
| condition_era group during day -30 through 0 days relative to index: Acute nephropathy | 0.9 | 2.6 | 0.13 | 1.0 | 2.7 | 0.13 |
| drug_era group during day -365 through 0 days relative to index: BELLADONNA AND DERIVATIVES, PLAIN | 1.7 | 1.8 | 0.01 | 3.6 | 1.5 | -0.13 |
| measurement during day -365 through 0 days relative to index: Screening for malignant neoplasm of prostate | 2.1 | 4.9 | 0.15 | 6.1 | 3.4 | -0.13 |
| condition_era group during day -30 through 0 days relative to index: Acute genitourinary disorder | 1.0 | 2.9 | 0.14 | 1.2 | 3.1 | 0.13 |

| (Continued) | T (%) | C (%) | SDf | T (%) | C (%) | SDf |
|---|---|---|---|---|---|---|
| observation during day -365 through 0 days relative to index: Ambulance service, basic life support, non-emergency transport, (bls) | 0.4 | 1.5 | 0.11 | 0.3 | 1.6 | 0.13 |
| condition_era group during day -30 through 0 days relative to index: Acute renal failure syndrome | 0.9 | 2.6 | 0.13 | 1.0 | 2.6 | 0.13 |

**Supplementary Table 219:** Most extreme patient characteristics balance diagnostics after matching for SGLT2I (male) vs DPP4I (male) in the MDCR data source.

| | Before matching | | | After matching | | |
|---|---|---|---|---|---|---|
| Characteristic (total count = 12800) | T (%) | C (%) | SDf | T (%) | C (%) | SDf |
| age group: 90 - 94 | 0.3 | 1.3 | 0.12 | 0.3 | 1.7 | 0.14 |
| condition_era group during day -365 through 0 days relative to index: Chronic kidney disease stage 2 | 2.2 | 1.5 | -0.05 | 2.3 | 0.9 | -0.11 |
| measurement during day -365 through 0 days relative to index: Epithelial cells.squamous [#/area] in Urine sediment by Microscopy high power field | 0.3 | 0.7 | 0.06 | 0.3 | 1.3 | 0.11 |
| measurement during day -365 through 0 days relative to index: Hyaline casts [#/area] in Urine sediment by Microscopy low power field | 0.3 | 0.7 | 0.06 | 0.3 | 1.3 | 0.11 |
| measurement within normal range during day -365 through 0 days relative to index: Epithelial cells.squamous [#/area] in Urine sediment by Microscopy high power field | 0.3 | 0.7 | 0.06 | 0.3 | 1.3 | 0.11 |
| measurement within normal range during day -365 through 0 days relative to index: Hyaline casts [#/area] in Urine sediment by Microscopy low power field | 0.3 | 0.7 | 0.06 | 0.3 | 1.3 | 0.11 |
| condition_era group during day -365 through 0 days relative to index: Disease due to Alphaherpesvirinae | 1.7 | 1.3 | -0.03 | 1.8 | 0.6 | -0.11 |
| condition_era group during day -30 through 0 days relative to index: Benign hypertension | 3.5 | 11.8 | 0.32 | 4.4 | 2.4 | -0.11 |
| condition_era group during day -365 through 0 days relative to index: Chronic kidney disease stage 3A | 1.3 | 0.1 | -0.15 | 1.0 | -0.3 | -0.11 |
| measurement during day -365 through 0 days relative to index: C-reactive protein; high sensitivity (hsCRP) | 2.1 | 1.2 | -0.07 | 2.2 | 0.9 | -0.11 |

##### 3.2.5 MDCD

**Supplementary Figure 11:** Preference score distributions, propensity score stratified (middle column) and matched (right column) covariate balance and negative control error distributions (bottom row) for SGLT2I (male) vs DPP4I (male) in the MDCD data source.

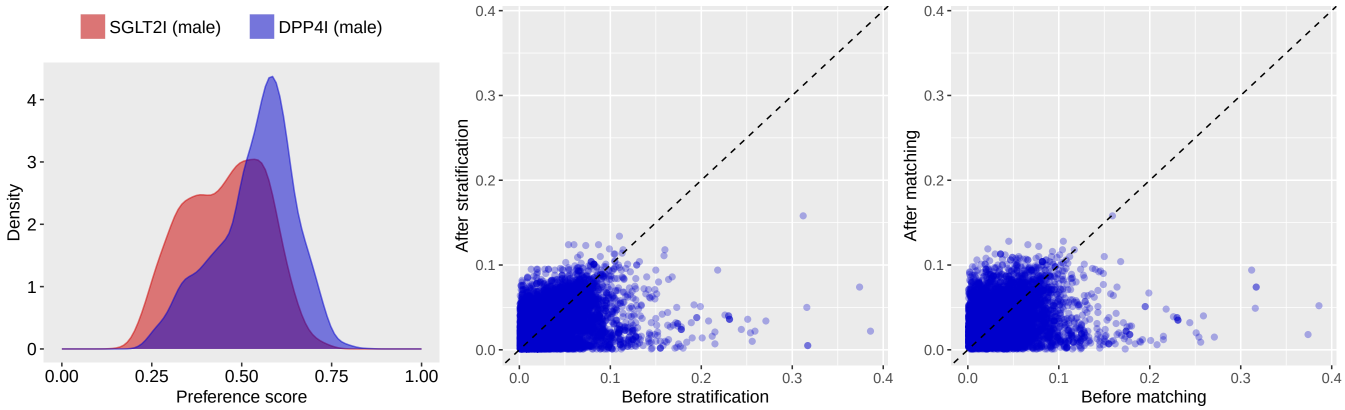

**Supplementary Table 220:** Most extreme patient characteristics balance diagnostics after stratification for SGLT2I (male) vs DPP4I (male) in the MDCD data source.

| Characteristic (total count = 15002) | Before stratification | | | After stratification | | |
|---|---|---|---|---|---|---|
| | T (%) | C (%) | SDf | T (%) | C (%) | SDf |
| index year: 2013 | -0.3 | 5.2 | 0.31 | 0.6 | 2.6 | 0.16 |
| condition_era group during day -30 through 0 days relative to index: Acute infectious disease | 0.8 | 2.1 | 0.11 | 0.6 | 2.2 | 0.13 |
| condition_era group during day -30 through 0 days relative to index: Acute respiratory infections | 0.7 | 1.7 | 0.09 | 0.5 | 1.8 | 0.12 |
| measurement during day -365 through 0 days relative to index: Lipoprotein, direct measurement; high density cholesterol (HDL cholesterol) | 2.5 | 3.5 | 0.06 | 1.7 | 3.8 | 0.12 |
| Charlson index - Romano adaptation | 394.8 | 408.1 | 0.05 | 383.9 | 414.6 | 0.12 |
| measurement during day -365 through 0 days relative to index: Mass spectrometry and tandem mass spectrometry (MS, MS/MS), analyte not elsewhere specified; qualitative, each specimen | 0.3 | -0.1 | -0.07 | 0.8 | -0.1 | -0.12 |
| procedure_occurrence during day -365 through 0 days relative to index: Scanning computerized ophthalmic diagnostic imaging, posterior segment, with interpretation and report, unilateral or bilateral; optic nerve | 4.2 | 2.5 | -0.10 | 4.8 | 2.5 | -0.12 |

| (Continued) | T (%) | C (%) | SDf | T (%) | C (%) | SDf |
|---|---|---|---|---|---|---|
| condition_era group during day -30 through 0 days relative to index: Renal failure syndrome | 0.5 | 2.3 | 0.16 | 0.7 | 2.0 | 0.12 |
| observation during day -365 through 0 days relative to index: Ambulance service, basic life support, non-emergency transport, (bls) | 0.7 | 2.0 | 0.11 | 0.6 | 1.9 | 0.12 |
| condition_era group during day -365 through 0 days relative to index: Anemia of chronic disease | -0.3 | 0.8 | 0.11 | -0.3 | 0.8 | 0.11 |

**Supplementary Table 221:** Most extreme patient characteristics balance diagnostics after matching for SGLT2I (male) vs DPP4I (male) in the MDCD data source.

| | Before matching | | | After matching | | |
|---|---|---|---|---|---|---|
| Characteristic (total count = 11140) | T (%) | C (%) | SDf | T (%) | C (%) | SDf |
| observation during day -30 through 0 days relative to index: Most recent systolic blood pressure $<$ 140 mmhg | 2.3 | 0.4 | -0.16 | 2.2 | 0.4 | -0.16 |
| observation during day -365 through 0 days relative to index: History of fall | -0.3 | 1.1 | 0.10 | -0.3 | 1.5 | 0.13 |
| condition_era group during day -365 through 0 days relative to index: Melanocytic tumor of skin | 0.3 | 0.6 | 0.04 | -0.3 | 1.5 | 0.13 |
| observation during day -365 through 0 days relative to index: Mobility: walking & moving around functional limitation, current status, at therapy episode outset and at reporting intervals | 0.4 | 0.9 | 0.07 | 0.4 | 1.7 | 0.12 |
| drug_era group during day -365 through 0 days relative to index: ceftriaxone | 2.7 | 4.1 | 0.08 | 2.5 | 4.8 | 0.12 |
| condition_era group during day -365 through 0 days relative to index: Melanocytic neoplasm | 0.7 | 0.9 | 0.02 | 0.7 | 2.1 | 0.12 |
| observation during day -365 through 0 days relative to index: Patient receiving angiotensin converting enzyme (ace) inhibitor or angiotensin receptor blocker (arb) therapy | 1.2 | 0.2 | -0.12 | 1.2 | -0.4 | -0.12 |
| condition_era group during day -365 through 0 days relative to index: Disorders of both mitral and tricuspid valves | 1.1 | 0.3 | -0.10 | 1.2 | -0.4 | -0.12 |
| condition_era group during day -365 through 0 days relative to index: Chronic systolic heart failure | 4.6 | 2.8 | -0.09 | 4.5 | 2.4 | -0.12 |
| condition_era group during day -365 through 0 days relative to index: Hamartoma of skin | 0.3 | 0.6 | 0.04 | -0.3 | 1.3 | 0.11 |

##### 3.2.6 Open Claims

**Supplementary Figure 12:** Preference score distributions, propensity score stratified (middle column) and matched (right column) covariate balance and negative control error distributions (bottom row) for SGLT2I (male) vs DPP4I (male) in the Open Claims data source.

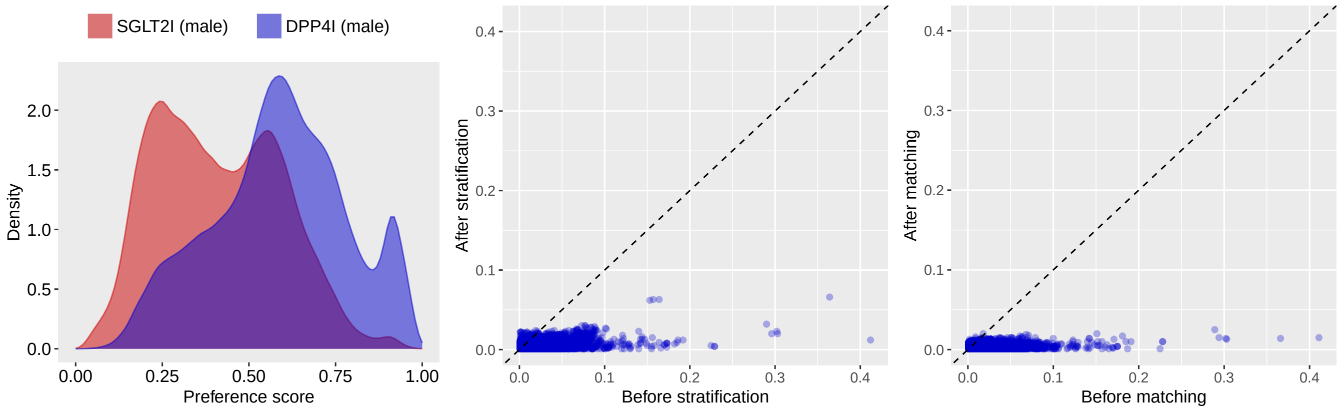

**Supplementary Table 222:** Most extreme patient characteristics balance diagnostics after stratification for SGLT2I (male) vs DPP4I (male) in the Open Claims data source.

| | Before stratification | | | After stratification | | |
|---|---|---|---|---|---|---|
| Characteristic (total count = 9661) | T (%) | C (%) | SDf | T (%) | C (%) | SDf |
| index year: 2013 | 0.6 | 7.8 | 0.36 | 3.1 | 4.4 | 0.07 |
| drug_era group during day -365 through 0 days relative to index: sacubitril | 2.0 | 0.3 | -0.16 | 1.2 | 0.6 | -0.06 |
| drug_era group during day -30 through 0 days relative to index: sacubitril | 1.8 | 0.2 | -0.16 | 1.0 | 0.5 | -0.06 |
| drug_era group during day 0 through 0 days relative to index: sacubitril | 1.7 | 0.2 | -0.15 | 1.0 | 0.5 | -0.06 |
| index year: 2014 | 4.3 | 12.2 | 0.29 | 9.5 | 8.6 | -0.03 |
| condition_era group during day -30 through 0 days relative to index: Systolic dysfunction | 1.4 | 0.6 | -0.08 | 1.0 | 0.7 | -0.03 |
| condition_era group during day -30 through 0 days relative to index: Systolic heart failure | 1.4 | 0.6 | -0.08 | 1.0 | 0.7 | -0.03 |
| condition_era group during day -30 through 0 days relative to index: Abnormal cardiovascular function | 1.8 | 0.9 | -0.07 | 1.3 | 1.0 | -0.03 |
| condition_era group during day -365 through 0 days relative to index: Chronic congestive heart failure | 1.6 | 0.6 | -0.09 | 1.1 | 0.8 | -0.03 |

| (Continued) | T (%) | C (%) | SDf | T (%) | C (%) | SDf |
|---|---|---|---|---|---|---|
| condition_era group during day -365 through 0 days relative to index: Systolic heart failure | 2.7 | 1.5 | -0.08 | 2.1 | 1.7 | -0.03 |

**Supplementary Table 223:** Most extreme patient characteristics balance diagnostics after matching for SGLT2I (male) vs DPP4I (male) in the Open Claims data source.

| | Before matching | | | After matching | | |
|---|---|---|---|---|---|---|
| Characteristic (total count = 9373) | T (%) | C (%) | SDf | T (%) | C (%) | SDf |
| index year: 2014 | 4.3 | 12.2 | 0.29 | 5.9 | 5.3 | -0.03 |
| age group: 75 - 79 | 5.4 | 9.3 | 0.15 | 5.8 | 5.3 | -0.02 |
| drug_era group during day -365 through 0 days relative to index: lorcaserin | 0.1 | 0.1 | -0.02 | 0.1 | 0.1 | -0.02 |
| drug_era group during day 0 through 0 days relative to index: DRUGS USED IN BENIGN PROSTATIC HYPERTROPHY | 9.2 | 11.3 | 0.07 | 9.6 | 9.0 | -0.02 |
| drug_era group during day -30 through 0 days relative to index: DRUGS USED IN BENIGN PROSTATIC HYPERTROPHY | 9.7 | 11.9 | 0.07 | 10.1 | 9.6 | -0.02 |
| index year: 2016 | 7.7 | 13.2 | 0.18 | 10.3 | 10.8 | 0.02 |
| drug_era group during day 0 through 0 days relative to index: Alpha-adrenoreceptor antagonists | 8.0 | 9.7 | 0.06 | 8.3 | 7.8 | -0.02 |
| drug_era group during day -30 through 0 days relative to index: Alpha-adrenoreceptor antagonists | 8.5 | 10.4 | 0.06 | 8.8 | 8.4 | -0.02 |
| device_exposure during day -365 through 0 days relative to index: TRUETEST GLUCOSE TEST STRIPS | 0.1 | 0.5 | 0.06 | 0.2 | 0.3 | 0.02 |
| condition_era group during day -365 through 0 days relative to index: Mass of pancreas | 0.2 | 0.2 | -0.01 | 0.2 | 0.1 | -0.02 |

##### 3.2.7 OEHR

**Supplementary Figure 13:** Preference score distributions, propensity score stratified (middle column) and matched (right column) covariate balance and negative control error distributions (bottom row) for SGLT2I (male) vs DPP4I (male) in the OEHR data source.

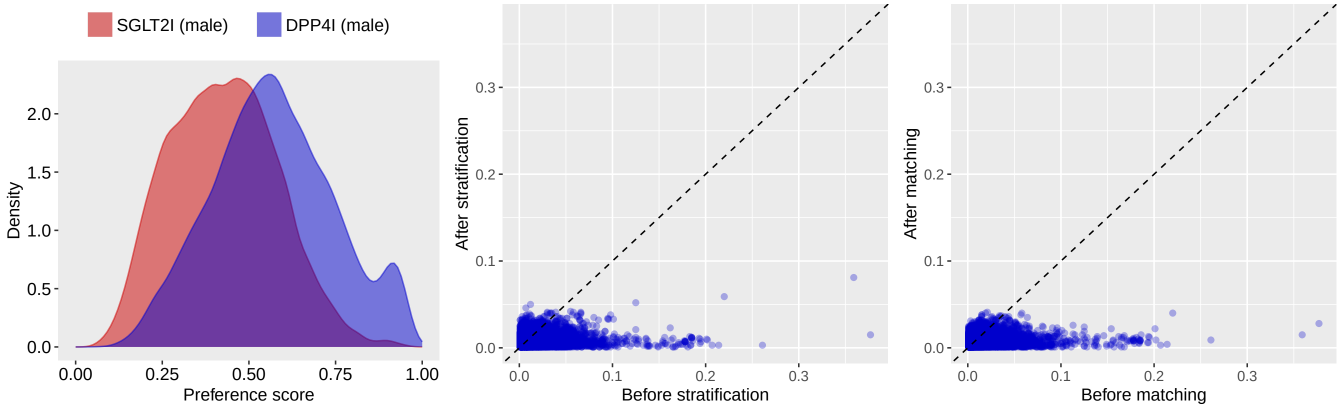

**Supplementary Table 224:** Most extreme patient characteristics balance diagnostics after stratification for SGLT2I (male) vs DPP4I (male) in the OEHR data source.

| Characteristic (total count = 11251) | Before stratification | | | After stratification | | |
|---|---|---|---|---|---|---|
| | T (%) | C (%) | SDf | T (%) | C (%) | SDf |
| index year: 2013 | 0.4 | 7.1 | 0.36 | 2.3 | 3.7 | 0.08 |
| index year: 2014 | 3.9 | 9.3 | 0.22 | 9.7 | 8.0 | -0.06 |
| observation during day -30 through 0 days relative to index: Prescription(s) generated and transmitted via a qualified erx system | 0.2 | 1.3 | 0.12 | 0.3 | 0.6 | 0.05 |
| drug_era group during day 0 through 0 days relative to index: ANTIMETABOLITES | 0.5 | 0.5 | -0.01 | 0.8 | 0.4 | -0.05 |
| drug_era group during day -30 through 0 days relative to index: ANTIMETABOLITES | 0.6 | 0.5 | -0.01 | 0.9 | 0.5 | -0.05 |
| observation during day -30 through 0 days relative to index: Pneumococcal vaccine administered or previously received (COPD) (PV), (IBD) | 0.1 | 0.4 | 0.06 | 0.1 | 0.3 | 0.04 |
| condition_era group during day -365 through 0 days relative to index: Acute exacerbation of chronic bronchitis | 0.1 | 0.5 | 0.07 | 0.2 | 0.4 | 0.04 |
| drug_era group during day 0 through 0 days relative to index: icosapent ethyl | 0.3 | 0.1 | -0.06 | 0.2 | 0.1 | -0.04 |

| (Continued) | T (%) | C (%) | SDf | T (%) | C (%) | SDf |
|---|---|---|---|---|---|---|
| observation during day -365 through 0 days relative to index: Pharmacy - Other Pharmacy | 0.1 | 0.3 | 0.04 | 0.1 | 0.3 | 0.04 |
| condition_era group during day -365 through 0 days relative to index: Chronic pancreatitis | 0.2 | 0.1 | -0.03 | 0.2 | 0.1 | -0.04 |

**Supplementary Table 225:** Most extreme patient characteristics balance diagnostics after matching for SGLT2I (male) vs DPP4I (male) in the OEHR data source.

| | Before matching | | | After matching | | |
|---|---|---|---|---|---|---|
| Characteristic (total count = 11113) | T (%) | C (%) | SDf | T (%) | C (%) | SDf |
| observation during day -30 through 0 days relative to index: Vehicle accident | 0.1 | 0.2 | 0.02 | 0.1 | 0.3 | 0.04 |
| index year: 2014 | 3.9 | 9.3 | 0.22 | 4.7 | 3.9 | -0.04 |
| drug_era group during day 0 through 0 days relative to index: icosapent ethyl | 0.3 | 0.1 | -0.06 | 0.2 | 0.1 | -0.04 |
| observation during day -365 through 0 days relative to index: Vehicle accident | 0.2 | 0.3 | 0.02 | 0.2 | 0.4 | 0.04 |
| drug_era group during day -30 through 0 days relative to index: icosapent ethyl | 0.3 | 0.1 | -0.06 | 0.3 | 0.1 | -0.04 |
| condition_era group during day -365 through 0 days relative to index: Deviated nasal septum | 0.4 | 0.5 | 0.01 | 0.4 | 0.7 | 0.04 |
| condition_era group during day -365 through 0 days relative to index: Nasal deviation | 0.4 | 0.5 | 0.01 | 0.4 | 0.7 | 0.04 |
| drug_era group during day -365 through 0 days relative to index: dexmedetomidine | 0.2 | 0.1 | -0.04 | 0.2 | 0.1 | -0.04 |
| condition_era group during day -365 through 0 days relative to index: Left inguinal hernia | 0.1 | 0.2 | 0.03 | 0.1 | 0.3 | 0.04 |
| observation during day -365 through 0 days relative to index: Tobacco use assessed (CAD, CAP, COPD, PV) (DM) | 0.1 | 0.2 | 0.04 | 0.1 | 0.2 | 0.04 |

### 3.2.8 VA

**Supplementary Figure 14:** Preference score distributions, propensity score stratified (middle column) and matched (right column) covariate balance and negative control error distributions (bottom row) for SGLT2I (male) vs DPP4I (male) in the VA data source.

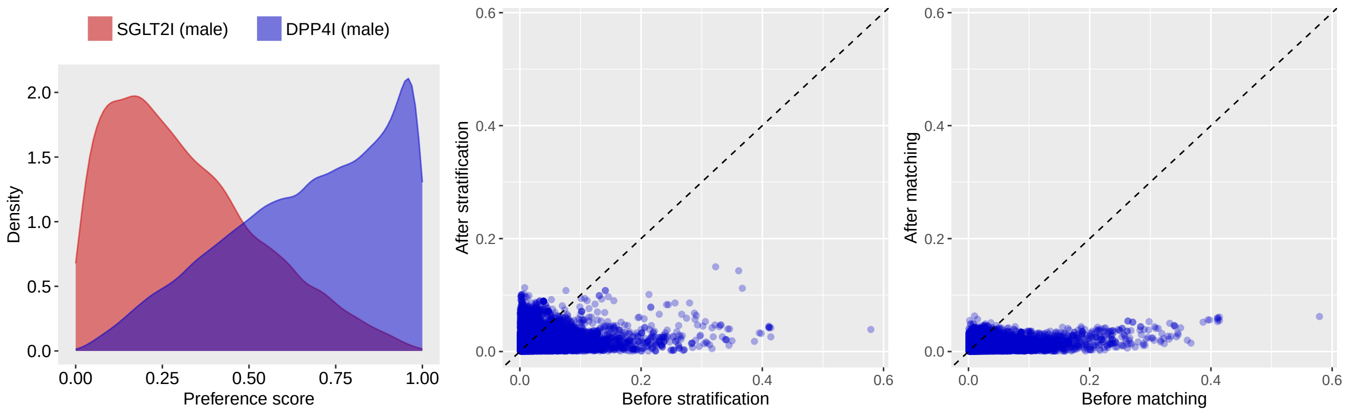

**Supplementary Table 226:** Most extreme patient characteristics balance diagnostics after stratification for SGLT2I (male) vs DPP4I (male) in the VA data source.

| | Before stratification | | | After stratification | | |
|---|---|---|---|---|---|---|
| Characteristic (total count = 14976) | T (%) | C (%) | SDf | T (%) | C (%) | SDf |
| index year: 2015 | -0.1 | 5.1 | 0.32 | 1.2 | 3.5 | 0.15 |
| index year: 2016 | 0.5 | 7.5 | 0.36 | 9.0 | 5.3 | -0.14 |
| device_exposure during day -365 through 0 days relative to index: BD 3 ML SYRINGE WITH NEEDLE | 0.1 | 0.1 | -0.01 | 0.9 | 0.1 | -0.11 |
| procedure_occurrence during day -365 through 0 days relative to index: Counseling | -0.1 | 6.5 | 0.37 | 1.7 | 3.5 | 0.11 |
| measurement during day -365 through 0 days relative to index: Glomerular filtration rate/1.73 sq M.predicted among non-blacks [Volume Rate/Area] in Serum, Plasma or Blood by Creatinine-based formula (CKD-EPI) | -0.1 | 1.2 | 0.14 | 0.1 | 0.8 | 0.11 |
| measurement during day -365 through 0 days relative to index: Glomerular filtration rate/1.73 sq M.predicted among blacks [Volume Rate/Area] in Serum, Plasma or Blood by Creatinine-based formula (CKD-EPI) | -0.1 | 1.2 | 0.14 | 0.1 | 0.8 | 0.11 |
| device_exposure during day -30 through 0 days relative to index: ACCU-CHEK AVIVA SOLUTION | 0.5 | 1.2 | 0.08 | 2.2 | 0.9 | -0.11 |

| (Continued) | T (%) | C (%) | SDf | T (%) | C (%) | SDf |
|---|---|---|---|---|---|---|
| measurement within normal range during day -365 through 0 days relative to index: Thyroglobulin Ab [Units/volume] in Serum or Plasma | 0.2 | 0.2 | 0.01 | 0.9 | 0.2 | -0.10 |
| measurement during day -365 through 0 days relative to index: Anion gap in Serum or Plasma | 52.9 | 42.4 | -0.21 | 41.2 | 46.2 | 0.10 |
| drug_era group during day 0 through 0 days relative to index: ANDROGENS AND FEMALE SEX HORMONES IN COMBINATION | 1.8 | 1.7 | -0.00 | 3.3 | 1.7 | -0.10 |

**Supplementary Table 227:** Most extreme patient characteristics balance diagnostics after matching for SGLT2I (male) vs DPP4I (male) in the VA data source.

| | Before matching | | | After matching | | |
|---|---|---|---|---|---|---|
| Characteristic (total count = 13830) | T (%) | C (%) | SDf | T (%) | C (%) | SDf |
| measurement during day -30 through 0 days relative to index: Creatinine [Mass/time] in 24 hour Urine | 0.4 | 0.3 | -0.01 | 0.4 | -0.1 | -0.06 |
| index year: 2021 | 39.8 | 15.0 | -0.58 | 29.6 | 32.5 | 0.06 |
| condition_era group during day -365 through 0 days relative to index: Coronary arteriosclerosis | 32.7 | 15.4 | -0.41 | 21.8 | 24.4 | 0.06 |
| condition_era group during day -365 through 0 days relative to index: Vascular degeneration | 33.6 | 16.2 | -0.41 | 22.7 | 25.3 | 0.06 |
| condition_era group during day -365 through 0 days relative to index: Arteriosclerotic vascular disease | 33.6 | 16.2 | -0.41 | 22.7 | 25.2 | 0.06 |
| condition_era group during day -365 through 0 days relative to index: Heart disease | 48.3 | 28.7 | -0.41 | 35.9 | 38.7 | 0.06 |
| observation during day -365 through 0 days relative to index: Screening tests for diabetes mellitus reviewed, requested, or ordered (DSP) | 0.2 | 0.1 | -0.01 | 0.2 | -0.1 | -0.06 |
| condition_era group during day -365 through 0 days relative to index: Coronary atherosclerosis | 29.5 | 13.5 | -0.40 | 19.8 | 22.1 | 0.06 |
| condition_era group during day -365 through 0 days relative to index: Atherosclerosis of artery | 30.4 | 14.3 | -0.40 | 20.6 | 23.0 | 0.06 |
| condition_era group during day -365 through 0 days relative to index: Atherosclerosis of coronary artery without angina pectoris | 28.4 | 12.1 | -0.41 | 19.1 | 21.3 | 0.06 |

##### 3.2.9 SIDIAP

**Supplementary Figure 15:** Preference score distributions, propensity score stratified (middle column) and matched (right column) covariate balance and negative control error distributions (bottom row) for SGLT2I (male) vs DPP4I (male) in the SIDIAP data source.

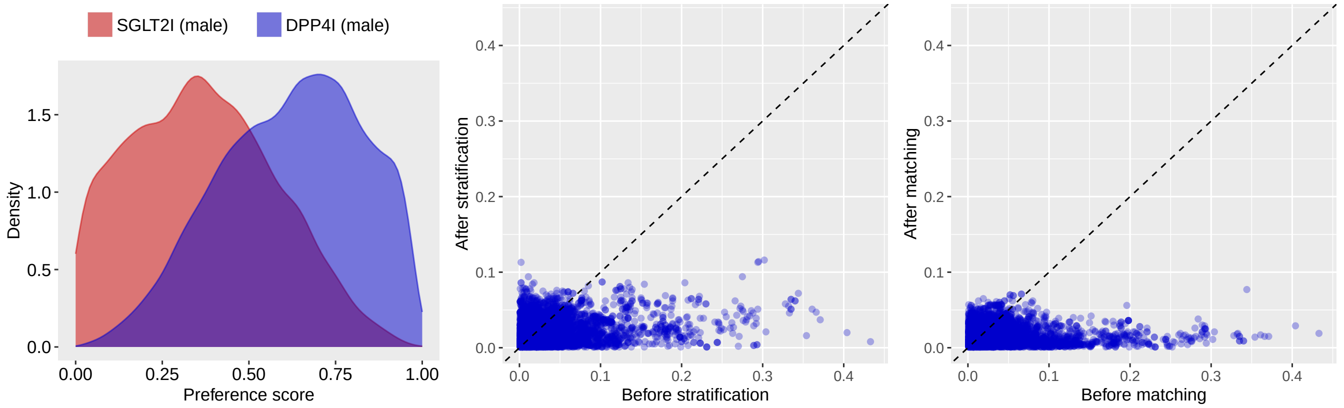

**Supplementary Table 228:** Most extreme patient characteristics balance diagnostics after stratification for SGLT2I (male) vs DPP4I (male) in the SIDIAP data source.

| | Before stratification | | | After stratification | | |
|---|---|---|---|---|---|---|
| Characteristic (total count = 6558) | T (%) | C (%) | SDf | T (%) | C (%) | SDf |
| drug_era group during day -365 through 0 days relative to index: sacubitril | 4.8 | 0.1 | -0.30 | 1.5 | 0.4 | -0.12 |
| drug_era group during day -30 through 0 days relative to index: sacubitril | 4.5 | 0.1 | -0.29 | 1.4 | 0.4 | -0.11 |
| drug_era group during day 0 through 0 days relative to index: sacubitril | 4.5 | 0.1 | -0.29 | 1.4 | 0.4 | -0.11 |
| drug_era group during day 0 through 0 days relative to index: dexketoprofen | 0.7 | 0.7 | 0.00 | 1.9 | 0.6 | -0.11 |
| procedure_occurrence during day -365 through 0 days relative to index: Fluoroscopy of Multiple Coronary Arteries using Low Osmolar Contrast | 4.1 | 0.2 | -0.28 | 1.3 | 0.4 | -0.09 |
| Charlson index - Romano adaptation | 222.6 | 220.8 | -0.01 | 213.9 | 229.7 | 0.09 |
| drug_era group during day 0 through 0 days relative to index: HIGH-CEILING DIURETICS | 12.3 | 9.1 | -0.10 | 7.2 | 9.7 | 0.09 |
| drug_era group during day 0 through 0 days relative to index: Sulfonamides, plain | 12.3 | 9.1 | -0.10 | 7.2 | 9.7 | 0.09 |

| (Continued) | T (%) | C (%) | SDf | T (%) | C (%) | SDf |
|---|---|---|---|---|---|---|
| procedure_occurrence during day -30 through 0 days relative to index: Fluoroscopy of Multiple Coronary Arteries using Low Osmolar Contrast | 2.2 | 0.1 | -0.20 | 0.7 | 0.1 | -0.09 |
| drug_era group during day -30 through 0 days relative to index: HMG CoA reductase inhibitors, other combinations | 76.5 | 70.7 | -0.13 | 67.7 | 71.6 | 0.09 |

**Supplementary Table 229:** Most extreme patient characteristics balance diagnostics after matching for SGLT2I (male) vs DPP4I (male) in the SIDIAP data source.

| | Before matching | | | After matching | | |
|---|---|---|---|---|---|---|
| Characteristic (total count = 6357) | T (%) | C (%) | SDf | T (%) | C (%) | SDf |
| index year: 2013 | -0.1 | 5.7 | 0.34 | -0.1 | 0.4 | 0.08 |
| condition_era group during day -365 through 0 days relative to index: Fracture of lower leg | 0.1 | 0.4 | 0.07 | 0.1 | 0.5 | 0.07 |
| condition_era group during day -365 through 0 days relative to index: Closed fracture of lower leg | 0.1 | 0.4 | 0.07 | 0.1 | 0.5 | 0.07 |
| condition_era group during day -30 through 0 days relative to index: Psychoactive substance use disorder | 0.9 | 1.5 | 0.05 | 0.9 | 0.4 | -0.07 |
| condition_era group during day -30 through 0 days relative to index: Mental disorder due to drug | 0.9 | 1.5 | 0.05 | 0.9 | 0.4 | -0.07 |
| condition_era group during day -30 through 0 days relative to index: Psychoactive substance dependence | 0.8 | 1.4 | 0.06 | 0.9 | 0.3 | -0.07 |
| condition_era group during day -30 through 0 days relative to index: Alcoholism | 0.8 | 1.4 | 0.06 | 0.9 | 0.3 | -0.07 |
| condition_era group during day -30 through 0 days relative to index: Psychoactive substance abuse | 0.6 | 1.1 | 0.05 | 0.6 | 0.2 | -0.07 |
| procedure_occurrence during day -365 through 0 days relative to index: Inspection of Upper Intestinal Tract, Via Natural or Artificial Opening Endoscopic | -0.1 | 0.1 | 0.02 | -0.1 | 0.3 | 0.07 |
| drug_era group during day 0 through 0 days relative to index: Anticholinesterases | 0.2 | 0.7 | 0.08 | 0.2 | 0.5 | 0.06 |

#### 3.3 SGLT2I (male) vs SU (male)

##### 3.3.1 CCAE

**Supplementary Figure 16:** Preference score distributions, propensity score stratified (middle column) and matched (right column) covariate balance and negative control error distributions (bottom row) for SGLT2I (male) vs SU (male) in the CCAE data source.

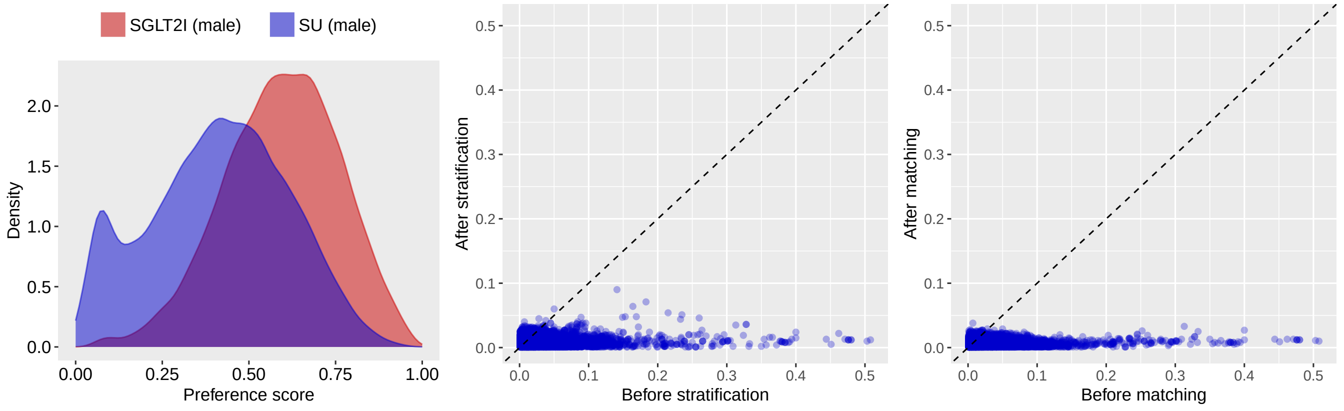

**Supplementary Table 230:** Most extreme patient characteristics balance diagnostics after stratification for SGLT2I (male) vs SU (male) in the CCAE data source.

| | Before stratification | | | After stratification | | |
|---|---|---|---|---|---|---|
| Characteristic (total count = 9977) | T (%) | C (%) | SDf | T (%) | C (%) | SDf |
| measurement during day -365 through 0 days relative to index: Immunoassay for infectious agent antibody, qualitative or semiquantitative, single step method (eg, reagent strip) | 0.1 | 1.3 | -0.14 | 0.1 | 0.6 | -0.09 |
| measurement during day -30 through 0 days relative to index: Potassium; serum, plasma or whole blood | 0.3 | 2.3 | -0.18 | 0.5 | 1.1 | -0.07 |
| measurement during day -30 through 0 days relative to index: Cholesterol, serum or whole blood, total | 0.1 | 1.7 | -0.16 | 0.2 | 0.5 | -0.06 |
| procedure_occurrence during day -365 through 0 days relative to index: Remote imaging for detection of retinal disease (eg, retinopathy in a patient with diabetes) with analysis and report under physician supervision, unilateral or bilateral | 0.1 | 0.3 | -0.05 | 0.1 | 0.4 | -0.06 |
| measurement during day -365 through 0 days relative to index: Cholesterol, serum or whole blood, total | 0.6 | 3.7 | -0.21 | 0.8 | 1.3 | -0.05 |

| (Continued) | T (%) | C (%) | SDf | T (%) | C (%) | SDf |
|---|---|---|---|---|---|---|
| measurement during day -365 through 0 days relative to index: Potassium; serum, plasma or whole blood | 1.4 | 5.7 | -0.23 | 2.2 | 3.0 | -0.05 |
| drug_era group during day -365 through 0 days relative to index: icosapent ethyl | 1.8 | 0.2 | 0.17 | 1.0 | 0.6 | 0.05 |
| drug_era group during day -30 through 0 days relative to index: icosapent ethyl | 1.5 | 0.1 | 0.15 | 0.8 | 0.4 | 0.05 |
| drug_era group during day 0 through 0 days relative to index: icosapent ethyl | 1.4 | 0.1 | 0.15 | 0.8 | 0.4 | 0.05 |
| measurement during day -365 through 0 days relative to index: Blood count; blood smear, microscopic examination without manual differential WBC count | 0.1 | 0.6 | -0.09 | 0.1 | 0.2 | -0.05 |

**Supplementary Table 231:** Most extreme patient characteristics balance diagnostics after matching for SGLT2I (male) vs SU (male) in the CCAE data source.

| | Before matching | | | After matching | | |
|---|---|---|---|---|---|---|
| Characteristic (total count = 9710) | T (%) | C (%) | SDf | T (%) | C (%) | SDf |
| index year: 2014 | 7.5 | 7.7 | -0.01 | 8.5 | 7.4 | 0.04 |
| index year: 2013 | 1.0 | 7.1 | -0.31 | 1.2 | 0.9 | 0.03 |
| drug_era group during day -365 through 0 days relative to index: vilazodone | 0.3 | 0.1 | 0.05 | 0.2 | 0.1 | 0.03 |
| observation during day -365 through 0 days relative to index: Sales tax | 0.1 | 0.2 | -0.03 | 0.1 | 0.2 | -0.03 |
| measurement during day -365 through 0 days relative to index: Benzodiazepine measurement | 0.1 | 0.1 | 0.00 | 0.1 | 0.2 | -0.03 |
| index year: 2019 | 17.6 | 5.1 | 0.40 | 16.2 | 17.2 | -0.03 |
| procedure_occurrence during day -365 through 0 days relative to index: Transcranial Doppler study of the intracranial arteries; complete study | 0.2 | 0.1 | 0.04 | 0.2 | 0.1 | 0.03 |
| condition_era group during day -365 through 0 days relative to index: Inflammatory bowel disease | 0.1 | 0.1 | -0.02 | 0.1 | 0.2 | -0.03 |
| condition_era group during day -365 through 0 days relative to index: Crohn’s disease of intestine | 0.1 | 0.1 | -0.02 | 0.1 | 0.2 | -0.03 |
| condition_era group during day -365 through 0 days relative to index: Gastrointestinal Crohn’s disease | 0.1 | 0.1 | -0.02 | 0.1 | 0.2 | -0.03 |

##### 3.3.2 DAG

**Supplementary Figure 17:** Preference score distributions, propensity score stratified (middle column) and matched (right column) covariate balance and negative control error distributions (bottom row) for SGLT2I (male) vs SU (male) in the DAG data source.

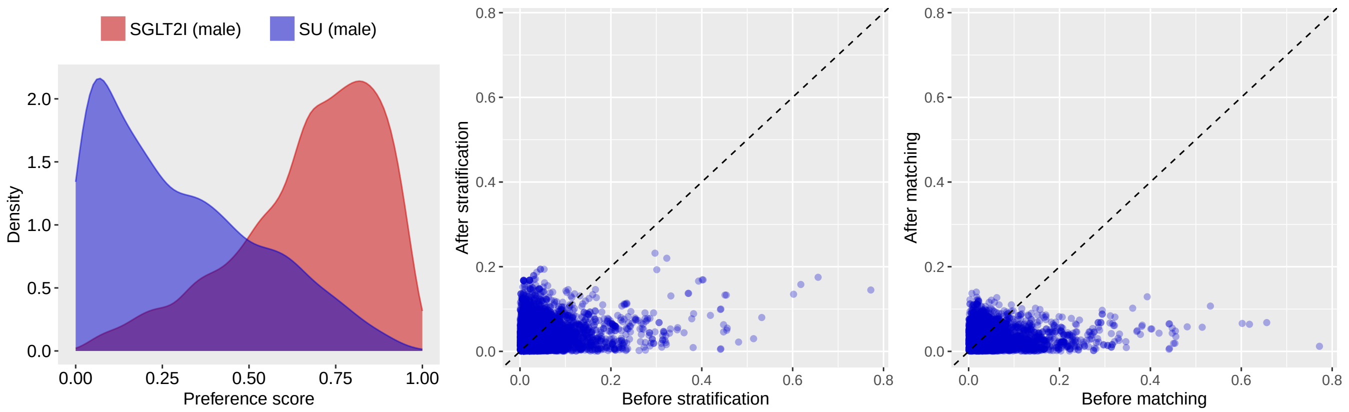

**Supplementary Table 232:** Most extreme patient characteristics balance diagnostics after stratification for SGLT2I (male) vs SU (male) in the DAG data source.

| Characteristic (total count = 6752) | Before stratification | | | After stratification | | |
|---|---|---|---|---|---|---|
| | T (%) | C (%) | SDf | T (%) | C (%) | SDf |
| drug_era group during day 0 through 0 days relative to index: rosuvastatin | 4.6 | -0.1 | 0.30 | 3.7 | 0.4 | 0.23 |
| drug_era group during day -365 through 0 days relative to index: rosuvastatin | 5.4 | 0.2 | 0.32 | 4.4 | 0.9 | 0.22 |
| condition_era group during day -30 through 0 days relative to index: Biliary calculus | 0.2 | 0.6 | -0.05 | 0.2 | 2.4 | -0.19 |
| condition_era group during day -30 through 0 days relative to index: Gallstone | 0.2 | 0.5 | -0.04 | 0.2 | 2.4 | -0.19 |
| condition_era group during day -30 through 0 days relative to index: Lesion of gallbladder | 0.2 | 0.5 | -0.04 | 0.2 | 2.4 | -0.19 |
| drug_era group during day -30 through 0 days relative to index: rosuvastatin | 4.7 | 0.1 | 0.30 | 3.8 | 0.9 | 0.19 |
| condition_era group during day -30 through 0 days relative to index: Calculus of gallbladder without cholecystitis or cholangitis | 0.2 | 0.4 | -0.04 | 0.2 | 2.3 | -0.19 |
| condition_era group during day -365 through 0 days relative to index: Gallstone | 1.2 | 1.5 | -0.03 | 1.2 | 4.1 | -0.18 |

| (Continued) | T (%) | C (%) | SDf | T (%) | C (%) | SDf |
|---|---|---|---|---|---|---|
| condition_era group during day -365 through 0 days relative to index: Lesion of gallbladder | 1.2 | 1.5 | -0.03 | 1.2 | 4.1 | -0.18 |
| drug_era group during day -365 through 0 days relative to index: atorvastatin | 28.6 | 5.2 | 0.66 | 24.5 | 32.4 | -0.17 |

**Supplementary Table 233:** Most extreme patient characteristics balance diagnostics after matching for SGLT2I (male) vs SU (male) in the DAG data source.

| | Before matching | | | After matching | | |
|---|---|---|---|---|---|---|
| Characteristic (total count = 5003) | T (%) | C (%) | SDf | T (%) | C (%) | SDf |
| drug_era group during day -365 through 0 days relative to index: urapidil | 0.8 | 0.7 | 0.02 | -0.5 | 1.2 | -0.14 |
| condition_era group during day -365 through 0 days relative to index: Mitral valve disorder | 1.1 | 1.1 | 0.01 | -0.5 | 1.2 | -0.14 |
| index year: 2019 | 14.5 | 3.5 | 0.39 | 8.0 | 11.9 | -0.13 |
| age group: 90 - 94 | 0.2 | 0.3 | -0.02 | -0.5 | 1.1 | -0.13 |
| condition_era group during day -365 through 0 days relative to index: Extrapyramidal disease | 0.3 | 0.8 | -0.06 | -0.5 | 1.1 | -0.13 |
| condition_era group during day -365 through 0 days relative to index: Intertrigo | 0.4 | 0.3 | 0.02 | 0.8 | -0.2 | 0.12 |
| drug_era group during day -365 through 0 days relative to index: bromazepam | 0.5 | 0.5 | -0.00 | 1.2 | -0.2 | 0.12 |
| condition_era group during day -365 through 0 days relative to index: Lesion of liver | 0.6 | 1.1 | -0.05 | -0.5 | 1.2 | -0.12 |
| condition_era group during day -365 through 0 days relative to index: Mitral valve regurgitation | 1.0 | 0.8 | 0.02 | -0.5 | 1.0 | -0.12 |
| drug_era group during day -30 through 0 days relative to index: ANTIADRENERGIC AGENTS, PERIPHERALLY ACTING | 1.7 | 2.0 | -0.02 | 0.7 | 2.1 | -0.12 |

##### 3.3.3 ODOD

**Supplementary Figure 18:** Preference score distributions, propensity score stratified (middle column) and matched (right column) covariate balance and negative control error distributions (bottom row) for SGLT2I (male) vs SU (male) in the ODOD data source.

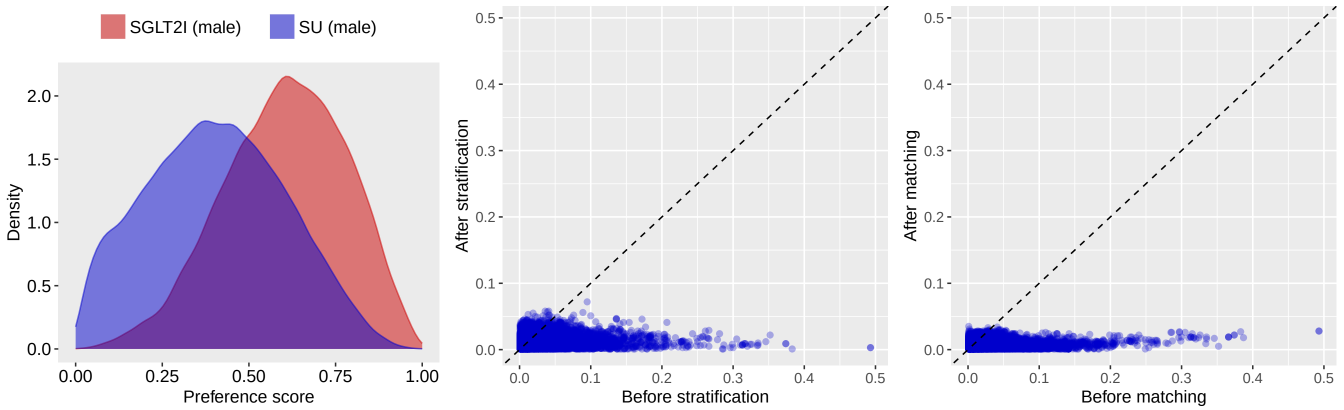

**Supplementary Table 234:** Most extreme patient characteristics balance diagnostics after stratification for SGLT2I (male) vs SU (male) in the ODOD data source.

| | Before stratification | | | After stratification | | |
|---|---|---|---|---|---|---|
| Characteristic (total count = 14433) | T (%) | C (%) | SDf | T (%) | C (%) | SDf |
| procedure_occurrence during day -30 through 0 days relative to index: Ultrasound, retroperitoneal (eg, renal, aorta, nodes), real time with image documentation; complete | 0.2 | 1.0 | -0.10 | 0.3 | 0.8 | -0.07 |
| measurement during day -365 through 0 days relative to index: Neutrophils [#/volume] in Blood | 0.5 | 0.3 | 0.04 | 0.8 | 0.3 | 0.06 |
| measurement within normal range during day -365 through 0 days relative to index: Monocytes [#/volume] in Blood | 0.4 | 0.2 | 0.04 | 0.6 | 0.2 | 0.06 |
| measurement within normal range during day -365 through 0 days relative to index: Lymphocytes [#/volume] in Blood | 0.4 | 0.2 | 0.04 | 0.6 | 0.3 | 0.06 |
| condition_era group during day -365 through 0 days relative to index: Atherosclerosis of arteries of the extremities | 0.6 | 1.6 | -0.09 | 1.7 | 1.0 | 0.06 |
| observation during day -365 through 0 days relative to index: Plan of care to address pain documented (COA) (ONC) | 0.5 | 0.4 | 0.02 | 0.9 | 0.4 | 0.06 |
| measurement within normal range during day -365 through 0 days relative to index: Eosinophils [#/volume] in Blood | 0.3 | 0.2 | 0.01 | 0.6 | 0.3 | 0.05 |

| (Continued) | T (%) | C (%) | SDf | T (%) | C (%) | SDf |
|---|---|---|---|---|---|---|
| observation during day -30 through 0 days relative to index: Blood Storage and Processing - General Classification | 0.1 | 0.6 | -0.08 | 0.1 | 0.4 | -0.05 |
| measurement during day -365 through 0 days relative to index: Lymphocytes [#/volume] in Blood | 0.7 | 0.4 | 0.04 | 1.0 | 0.5 | 0.05 |
| measurement within normal range during day -365 through 0 days relative to index: Basophils [#/volume] in Blood | 0.6 | 0.3 | 0.04 | 0.9 | 0.5 | 0.05 |

**Supplementary Table 235:** Most extreme patient characteristics balance diagnostics after matching for SGLT2I (male) vs SU (male) in the ODOD data source.

| | Before matching | | | After matching | | |
|---|---|---|---|---|---|---|
| Characteristic (total count = 14088) | T (%) | C (%) | SDf | T (%) | C (%) | SDf |
| observation during day -30 through 0 days relative to index: Other Therapeutic Services - Education/Training | 0.5 | 0.5 | -0.00 | 0.5 | 0.3 | 0.04 |
| observation during day -30 through 0 days relative to index: H/O: risk factor | 0.2 | 0.1 | 0.04 | 0.2 | 0.1 | 0.03 |
| procedure_occurrence during day -30 through 0 days relative to index: Transitional Care Management Services with the following required elements: Communication (direct contact, telephone, electronic) with the patient and/or caregiver within 2 business days of discharge Medical decision making of high complexity during the s | 0.3 | 0.2 | 0.03 | 0.3 | 0.1 | 0.03 |
| condition_era group during day -365 through 0 days relative to index: Tendon injury - upper limb | 0.1 | 0.1 | -0.00 | 0.1 | 0.2 | -0.03 |
| measurement during day -365 through 0 days relative to index: Molecular pathology procedure, Level 2 | 0.4 | 0.2 | 0.05 | 0.4 | 0.3 | 0.03 |
| measurement during day -365 through 0 days relative to index: Molecular pathology procedure, Level 1 | 0.2 | 0.1 | 0.04 | 0.2 | 0.1 | 0.03 |
| procedure_occurrence during day -365 through 0 days relative to index: Computed tomography, abdomen; without contrast material, followed by contrast material(s) and further sections | 0.4 | 0.7 | -0.04 | 0.4 | 0.2 | 0.03 |
| measurement during day -30 through 0 days relative to index: Culture, presumptive, pathogenic organisms, screening only | 0.1 | 0.2 | -0.03 | 0.1 | 0.2 | -0.03 |
| condition_era group during day -365 through 0 days relative to index: Longstanding persistent atrial fibrillation | 0.3 | 0.1 | 0.05 | 0.2 | 0.1 | 0.03 |
| measurement within normal range during day -365 through 0 days relative to index: Immunoassay for analyte other than infectious agent antibody or infectious agent antigen; quantitative, not otherwise specified | 0.3 | 0.1 | 0.04 | 0.2 | 0.1 | 0.03 |

##### 3.3.4 MDCR

**Supplementary Figure 19:** Preference score distributions, propensity score stratified (middle column) and matched (right column) covariate balance and negative control error distributions (bottom row) for SGLT2I (male) vs SU (male) in the MDCR data source.

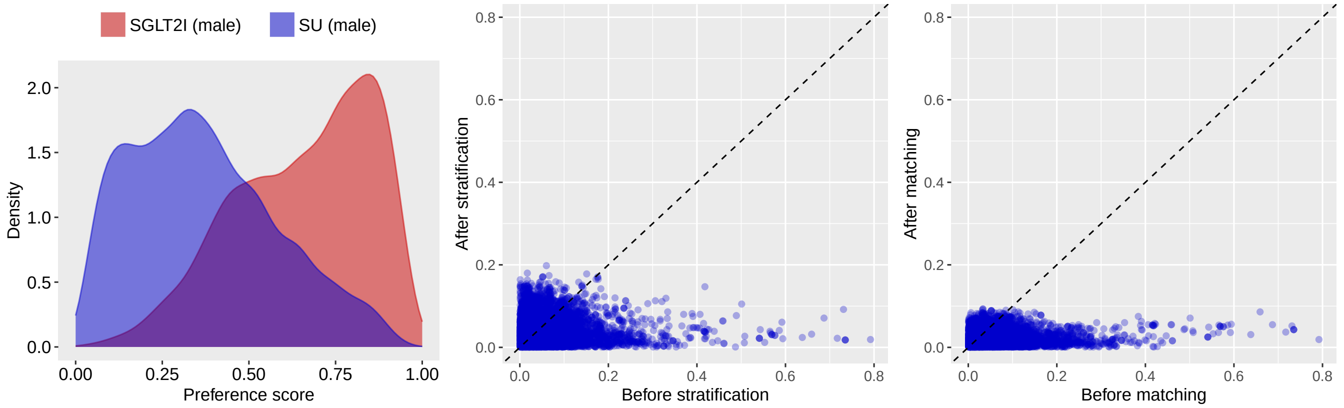

**Supplementary Table 236:** Most extreme patient characteristics balance diagnostics after stratification for SGLT2I (male) vs SU (male) in the MDCR data source.

| Characteristic (total count = 12544) | Before stratification | | | After stratification | | |
|---|---|---|---|---|---|---|
| | T (%) | C (%) | SDf | T (%) | C (%) | SDf |
| condition_era group during day -365 through 0 days relative to index: Abnormal chest sounds | -0.2 | 0.4 | -0.06 | 2.5 | 0.2 | 0.20 |
| measurement during day -365 through 0 days relative to index: Hepatic function panel This panel must include the following: Albumin (82040) Bilirubin, total (82247) Bilirubin, direct (82248) Phosphatase, alkaline (84075) Protein, total (84155) Transferase, alanine amino (ALT) (SGPT) (84460) Transferase, aspartate am | 4.8 | 4.4 | 0.02 | 9.1 | 4.6 | 0.18 |
| condition_era group during day -365 through 0 days relative to index: Renal failure syndrome | 3.8 | 7.2 | -0.15 | 3.9 | 8.1 | -0.18 |
| condition_era group during day -365 through 0 days relative to index: Genetic predisposition | 1.4 | 0.7 | 0.07 | 3.4 | 0.9 | 0.17 |
| condition_era group during day -365 through 0 days relative to index: Hypertensive renal failure | 0.3 | 2.3 | -0.18 | 0.4 | 2.4 | -0.17 |
| condition_era group during day -365 through 0 days relative to index: Cutaneous hypersensitivity | 1.2 | 0.7 | 0.05 | 3.3 | 0.9 | 0.17 |

| (Continued) | T (%) | C (%) | SDf | T (%) | C (%) | SDf |
|---|---|---|---|---|---|---|
| condition_era group during day -365 through 0 days relative to index: Allergic dermatitis | 1.2 | 0.7 | 0.05 | 3.3 | 0.9 | 0.17 |
| condition_era group during day -365 through 0 days relative to index: Atopic dermatitis | 1.1 | 0.6 | 0.05 | 3.2 | 0.9 | 0.17 |
| condition_era group during day -365 through 0 days relative to index: Benign hypertensive renal disease | 0.3 | 2.3 | -0.18 | 0.4 | 2.3 | -0.17 |
| condition_era group during day -30 through 0 days relative to index: Renal failure syndrome | 1.1 | 3.8 | -0.18 | 1.3 | 3.9 | -0.17 |

**Supplementary Table 237:** Most extreme patient characteristics balance diagnostics after matching for SGLT2I (male) vs SU (male) in the MDCR data source.

| | Before matching | | | After matching | | |
|---|---|---|---|---|---|---|
| Characteristic (total count = 11336) | T (%) | C (%) | SDf | T (%) | C (%) | SDf |
| condition_era group during day -365 through 0 days relative to index: Movement disorder | 1.2 | 1.6 | -0.03 | 1.1 | 2.3 | -0.09 |
| condition_era group during day -365 through 0 days relative to index: System disorder of the nervous system | 1.2 | 1.6 | -0.03 | 1.1 | 2.3 | -0.09 |
| condition_era group during day -365 through 0 days relative to index: Illness | 1.0 | 0.2 | 0.10 | 0.8 | 1.8 | -0.09 |
| condition_era group during day -365 through 0 days relative to index: Pruritic rash | 1.1 | 0.4 | 0.09 | 1.0 | 2.1 | -0.09 |
| condition_era group during day -365 through 0 days relative to index: Essential tremor | 0.4 | 0.2 | 0.03 | 0.3 | 1.1 | -0.09 |
| measurement during day -365 through 0 days relative to index: Syphilis test, non-treponemal antibody; qualitative (eg, VDRL, RPR, ART) | -0.2 | 0.5 | -0.05 | -0.3 | 0.8 | -0.09 |
| condition_era group during day -365 through 0 days relative to index: Absence of sensation | 0.7 | 0.3 | 0.05 | 0.5 | 1.4 | -0.09 |
| condition_era group during day -365 through 0 days relative to index: Anesthesia of skin | 0.7 | 0.3 | 0.05 | 0.5 | 1.4 | -0.09 |
| procedure_occurrence during day -365 through 0 days relative to index: Multi-leaf collimator (MLC) device(s) for intensity modulated radiation therapy (IMRT), design and construction per IMRT plan | -0.2 | 0.4 | -0.05 | -0.3 | 0.8 | -0.09 |
| index year: 2021 | 22.2 | 1.8 | 0.66 | 15.9 | 19.2 | -0.09 |

##### 3.3.5 MDCD

**Supplementary Figure 20:** Preference score distributions, propensity score stratified (middle column) and matched (right column) covariate balance and negative control error distributions (bottom row) for SGLT2I (male) vs SU (male) in the MDCD data source.

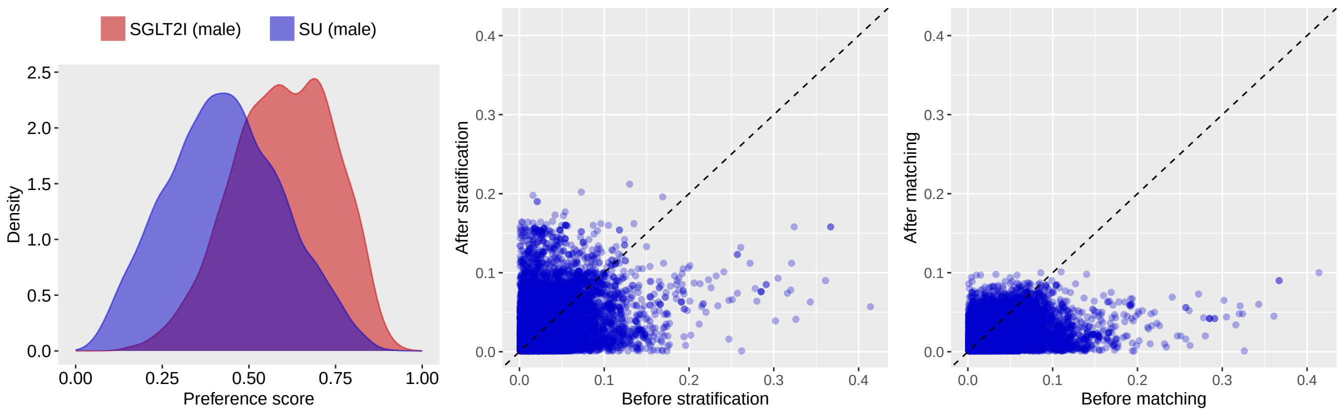

**Supplementary Table 238:** Most extreme patient characteristics balance diagnostics after stratification for SGLT2I (male) vs SU (male) in the MDCD data source.

| | Before stratification | | | After stratification | | |
|---|---|---|---|---|---|---|
| Characteristic (total count = 15620) | T (%) | C (%) | SDf | T (%) | C (%) | SDf |
| condition_era group during day -30 through 0 days relative to index: Tobacco dependence syndrome | 1.5 | 3.5 | -0.13 | 6.8 | 2.4 | 0.21 |
| condition_era group during day -30 through 0 days relative to index: Inflammatory disorder of genitourinary system | 0.5 | 1.1 | -0.07 | 4.3 | 1.0 | 0.20 |
| drug_era group during day -365 through 0 days relative to index: morphine | 6.4 | 6.8 | -0.02 | 12.5 | 6.7 | 0.20 |
| measurement during day -365 through 0 days relative to index: Drug screen, qualitative; single drug class method (eg, immunoassay, enzyme assay), each drug class | 0.8 | 3.1 | -0.17 | 4.5 | 1.2 | 0.20 |
| drug_era group during day -365 through 0 days relative to index: ANTIPROPULSIVES | 7.9 | 8.5 | -0.02 | 14.2 | 8.2 | 0.19 |
| drug_era group during day -365 through 0 days relative to index: Antipropulsives | 7.9 | 8.5 | -0.02 | 14.2 | 8.2 | 0.19 |
| drug_era group during day -30 through 0 days relative to index: morphine | 1.3 | 2.0 | -0.05 | 5.0 | 1.8 | 0.18 |

| (Continued) | T (%) | C (%) | SDf | T (%) | C (%) | SDf |
|---|---|---|---|---|---|---|
| procedure_occurrence during day -30 through 0 days relative to index: Smoking and tobacco use cessation counseling visit; intensive, greater than 10 minutes | 0.3 | 0.1 | 0.04 | 1.9 | 0.2 | 0.17 |
| measurement during day -30 through 0 days relative to index: Drug screen, qualitative; single drug class method (eg, immunoassay, enzyme assay), each drug class | -0.3 | 0.6 | -0.05 | 1.9 | 0.2 | 0.17 |
| measurement during day -30 through 0 days relative to index: Alcohol and/or drug screening; laboratory analysis of specimens for presence of alcohol and/or drugs | 0.3 | 0.1 | 0.04 | 1.8 | 0.2 | 0.17 |

**Supplementary Table 239:** Most extreme patient characteristics balance diagnostics after matching for SGLT2I (male) vs SU (male) in the MDCD data source.

| | Before matching | | | After matching | | |
|---|---|---|---|---|---|---|
| Characteristic (total count = 12499) | T (%) | C (%) | SDf | T (%) | C (%) | SDf |
| condition_era group during day -30 through 0 days relative to index: Diverticula of intestine | -0.3 | 0.9 | -0.11 | -0.3 | 0.8 | -0.10 |
| index year: 2020 | 17.9 | 5.0 | 0.41 | 16.1 | 19.9 | -0.10 |
| procedure_occurrence during day -365 through 0 days relative to index: Radiologic examination, spine, single view, specify level | 1.2 | 0.4 | 0.09 | 1.2 | 0.3 | 0.10 |
| condition_era group during day -30 through 0 days relative to index: Infectious disease of genitourinary system | -0.3 | 1.5 | -0.14 | -0.3 | 0.8 | -0.10 |
| observation during day -365 through 0 days relative to index: Functional status assessed (COA) (RA) | -0.3 | 0.5 | -0.03 | -0.3 | 1.1 | -0.10 |
| procedure_occurrence during day -365 through 0 days relative to index: Radiologic examination, elbow; complete, minimum of 3 views | 0.9 | 1.6 | -0.06 | 0.8 | 2.0 | -0.10 |
| observation during day -365 through 0 days relative to index: Inadequate immune status | 0.4 | 0.5 | -0.01 | 0.3 | 1.2 | -0.10 |
| observation during day -365 through 0 days relative to index: Pain severity quantified; no pain present (COA) (ONC) | 0.4 | 0.5 | -0.01 | 0.4 | 1.3 | -0.10 |
| condition_era group during day -365 through 0 days relative to index: Hordeolum | 0.3 | 0.7 | -0.05 | -0.3 | 0.9 | -0.09 |
| observation during day -30 through 0 days relative to index: Most recent systolic blood pressure less than 130 mm Hg (DM), (HTN, CKD, CAD) | 1.4 | 1.3 | 0.00 | 1.4 | 2.7 | -0.09 |

##### 3.3.6 Open Claims

**Supplementary Figure 21:** Preference score distributions, propensity score stratified (middle column) and matched (right column) covariate balance and negative control error distributions (bottom row) for SGLT2I (male) vs SU (male) in the Open Claims data source.

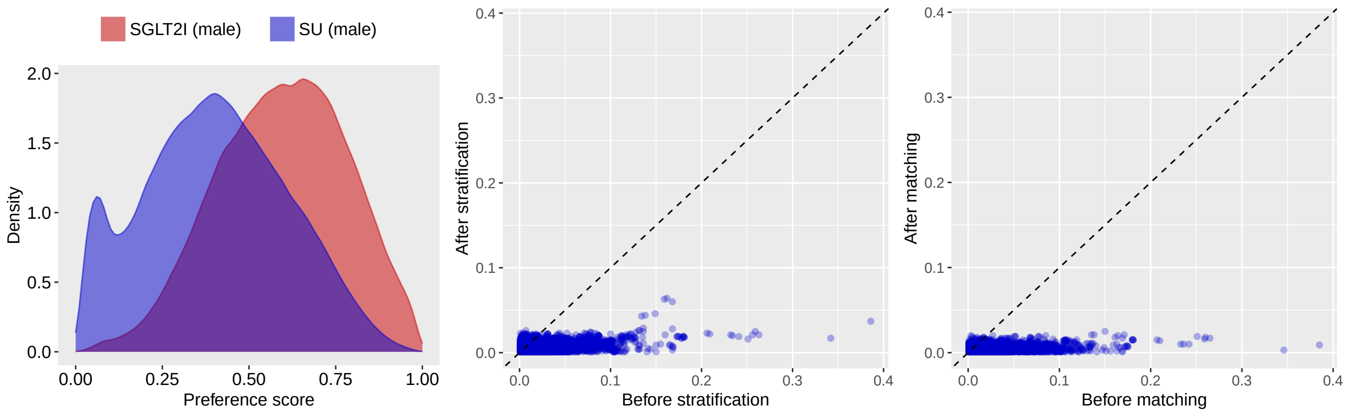

**Supplementary Table 240:** Most extreme patient characteristics balance diagnostics after stratification for SGLT2I (male) vs SU (male) in the Open Claims data source.

| | Before stratification | | | After stratification | | |
|---|---|---|---|---|---|---|
| Characteristic (total count = 9596) | T (%) | C (%) | SDf | T (%) | C (%) | SDf |
| drug_era group during day -30 through 0 days relative to index: sacubitril | 1.8 | 0.2 | 0.16 | 0.8 | 0.3 | 0.06 |
| drug_era group during day 0 through 0 days relative to index: sacubitril | 1.7 | 0.2 | 0.16 | 0.8 | 0.3 | 0.06 |
| drug_era group during day -365 through 0 days relative to index: sacubitril | 2.0 | 0.3 | 0.17 | 0.9 | 0.4 | 0.06 |
| drug_era group during day -365 through 0 days relative to index: icosapent ethyl | 1.9 | 0.3 | 0.15 | 0.9 | 0.5 | 0.05 |
| drug_era group during day -30 through 0 days relative to index: icosapent ethyl | 1.5 | 0.2 | 0.14 | 0.7 | 0.4 | 0.04 |
| drug_era group during day 0 through 0 days relative to index: icosapent ethyl | 1.4 | 0.2 | 0.13 | 0.7 | 0.4 | 0.04 |
| index year: 2013 | 0.6 | 8.5 | -0.39 | 4.9 | 5.7 | -0.04 |
| observation during day -365 through 0 days relative to index: Prescription(s) generated and transmitted via a qualified erx system | 0.2 | 1.3 | -0.14 | 0.7 | 1.0 | -0.03 |
| age group: 75 - 79 | 5.4 | 9.8 | -0.17 | 7.8 | 8.6 | -0.03 |

| (Continued) | T (%) | C (%) | SDf | T (%) | C (%) | SDf |
|---|---|---|---|---|---|---|
| observation during day -365 through 0 days relative to index: Exposure to SARS-CoV-2 | 2.7 | 0.9 | 0.13 | 1.6 | 1.3 | 0.03 |

**Supplementary Table 241:** Most extreme patient characteristics balance diagnostics after matching for SGLT2I (male) vs SU (male) in the Open Claims data source.

| | Before matching | | | After matching | | |
|---|---|---|---|---|---|---|
| Characteristic (total count = 9253) | T (%) | C (%) | SDf | T (%) | C (%) | SDf |
| drug_era group during day -365 through 0 days relative to index: icosapent ethyl | 1.9 | 0.3 | 0.15 | 1.0 | 0.8 | 0.03 |
| drug_era group during day -365 through 0 days relative to index: sacubitril | 2.0 | 0.2 | 0.17 | 0.8 | 0.6 | 0.02 |
| drug_era group during day -30 through 0 days relative to index: icosapent ethyl | 1.5 | 0.2 | 0.14 | 0.7 | 0.6 | 0.02 |
| drug_era group during day 0 through 0 days relative to index: icosapent ethyl | 1.4 | 0.2 | 0.14 | 0.7 | 0.5 | 0.02 |
| drug_era group during day -30 through 0 days relative to index: sacubitril | 1.8 | 0.2 | 0.16 | 0.7 | 0.5 | 0.02 |
| CHADS2VASc | 251.1 | 261.7 | -0.09 | 247.6 | 245.2 | 0.02 |
| procedure_occurrence during day -365 through 0 days relative to index: Ambulatory continuous glucose monitoring of interstitial tissue fluid via a subcutaneous sensor for a minimum of 72 hours; physician or other qualified health care professional (office) provided equipment, sensor placement, hook-up, calibration of monitor | 0.1 | 0.1 | 0.03 | 0.1 | 0.1 | 0.02 |
| condition_era group during day -365 through 0 days relative to index: Chronic pain | 4.5 | 4.5 | 0.00 | 4.3 | 4.8 | -0.02 |
| index year: 2014 | 4.3 | 10.8 | -0.25 | 4.7 | 4.3 | 0.02 |
| drug_era group during day -30 through 0 days relative to index: Aldosterone antagonists | 3.8 | 2.4 | 0.08 | 2.9 | 2.6 | 0.02 |

##### 3.3.7 OEHR

**Supplementary Figure 22:** Preference score distributions, propensity score stratified (middle column) and matched (right column) covariate balance and negative control error distributions (bottom row) for SGLT2I (male) vs SU (male) in the OEHR data source.

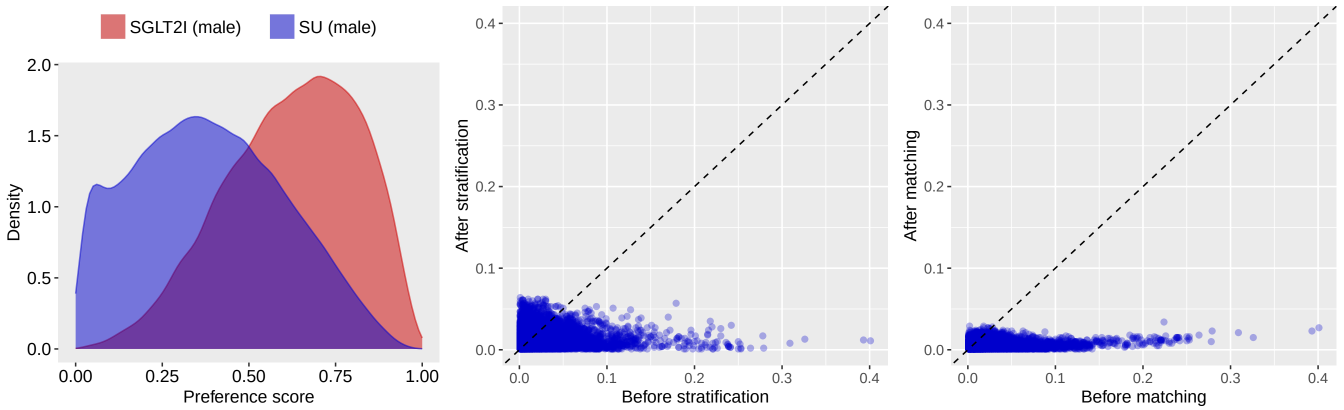

**Supplementary Table 242:** Most extreme patient characteristics balance diagnostics after stratification for SGLT2I (male) vs SU (male) in the OEHR data source.

| | Before stratification | | | After stratification | | |
|---|---|---|---|---|---|---|
| Characteristic (total count = 11257) | T (%) | C (%) | SDf | T (%) | C (%) | SDf |
| drug_era group during day -365 through 0 days relative to index: Other drugs for disorders of the musculo-skeletal system | 0.4 | 0.4 | 0.00 | 1.0 | 0.4 | 0.06 |
| drug_era group during day -365 through 0 days relative to index: Anesthetics for topical use | 10.8 | 9.9 | 0.03 | 12.7 | 10.7 | 0.06 |
| drug_era group during day -365 through 0 days relative to index: Local anesthetics | 10.6 | 9.8 | 0.03 | 12.6 | 10.6 | 0.06 |
| drug_era group during day -365 through 0 days relative to index: ANTIPRURITICS, INCL. ANTIHISTAMINES, ANESTHETICS, ETC. | 14.9 | 14.1 | 0.02 | 17.2 | 14.9 | 0.06 |
| drug_era group during day -365 through 0 days relative to index: ANTIPRURITICS, INCL. ANTIHISTAMINES, ANESTHETICS, ETC. | 14.9 | 14.1 | 0.02 | 17.2 | 14.9 | 0.06 |
| measurement during day -365 through 0 days relative to index: Blood, occult, by peroxidase activity (eg, guaiac), qualitative, feces, 1-3 simultaneous determinations, performed for other than colorectal neoplasm screening | 0.5 | 0.4 | 0.01 | 0.8 | 0.3 | 0.06 |

| (Continued) | T (%) | C (%) | SDf | T (%) | C (%) | SDf |
|---|---|---|---|---|---|---|
| drug_era group during day -365 through 0 days relative to index: OTHER DRUGS FOR DISORDERS OF THE MUSCULO-SKELETAL SYSTEM | 0.4 | 0.5 | -0.00 | 1.0 | 0.5 | 0.06 |
| drug_era group during day -365 through 0 days relative to index: OTHER DRUGS FOR DISORDERS OF THE MUSCULO-SKELETAL SYSTEM | 0.4 | 0.5 | -0.00 | 1.0 | 0.5 | 0.06 |
| drug_era group during day -365 through 0 days relative to index: MAGNETIC RESONANCE IMAGING CONTRAST MEDIA | 1.1 | 1.0 | 0.00 | 1.8 | 1.1 | 0.06 |
| drug_era group during day 0 through 0 days relative to index: ANTIMETABOLITES | 0.5 | 0.5 | 0.00 | 1.1 | 0.5 | 0.06 |

**Supplementary Table 243:** Most extreme patient characteristics balance diagnostics after matching for SGLT2I (male) vs SU (male) in the OEHR data source.

| | Before matching | | | After matching | | |
|---|---|---|---|---|---|---|
| Characteristic (total count = 10883) | T (%) | C (%) | SDf | T (%) | C (%) | SDf |
| index year: 2014 | 3.9 | 9.5 | -0.22 | 4.3 | 3.6 | 0.03 |
| condition_era group during day -365 through 0 days relative to index: Left inguinal hernia | 0.1 | 0.2 | -0.02 | 0.1 | 0.2 | -0.03 |
| observation during day -365 through 0 days relative to index: Problem situation relating to social and personal history | 0.1 | 0.1 | -0.00 | 0.1 | 0.2 | -0.03 |
| observation during day -365 through 0 days relative to index: Professional Fees - Anesthetist (CRNA) | 0.1 | 0.2 | -0.03 | 0.1 | 0.2 | -0.03 |
| index year: 2020 | 20.8 | 7.2 | 0.40 | 19.0 | 20.0 | -0.03 |
| procedure_occurrence during day -365 through 0 days relative to index: Medical nutrition therapy; group (2 or more individual(s)), each 30 minutes | 0.1 | 0.1 | -0.02 | 0.1 | 0.2 | -0.03 |
| drug_era group during day -365 through 0 days relative to index: zaleplon | 0.2 | 0.1 | 0.02 | 0.2 | 0.1 | 0.03 |
| condition_era group during day -365 through 0 days relative to index: Bilateral inguinal hernia | 0.1 | 0.2 | -0.02 | 0.1 | 0.2 | -0.03 |
| procedure_occurrence during day -365 through 0 days relative to index: Counseling about disease | 0.1 | 0.1 | -0.03 | 0.1 | 0.2 | -0.03 |
| measurement during day -30 through 0 days relative to index: Rheumatoid factor; quantitative | 0.1 | 0.2 | -0.03 | 0.1 | 0.2 | -0.03 |

### 3.3.8 VA

**Supplementary Figure 23:** Preference score distributions, propensity score stratified (middle column) and matched (right column) covariate balance and negative control error distributions (bottom row) for SGLT2I (male) vs SU (male) in the VA data source.

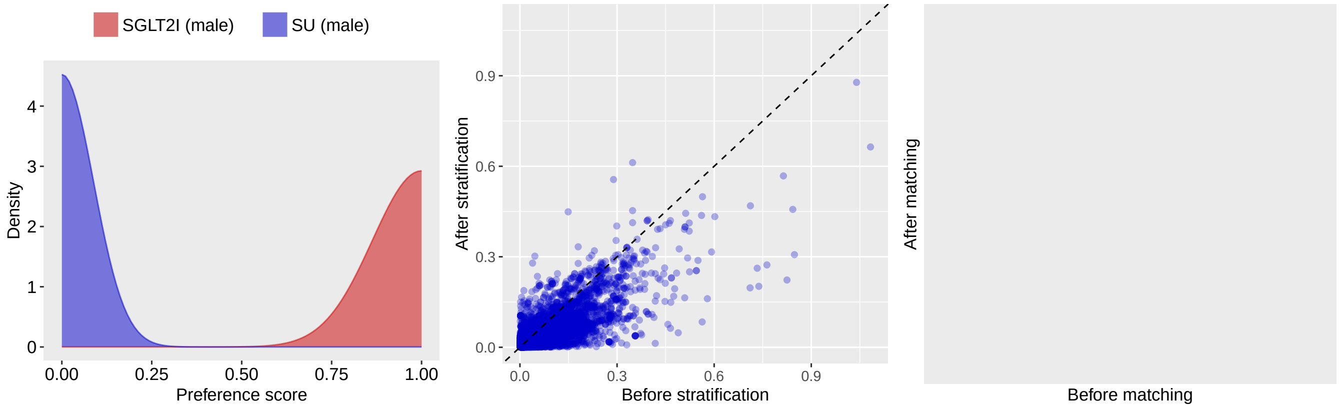

**Supplementary Table 244:** Most extreme patient characteristics balance diagnostics after stratification for SGLT2I (male) vs SU (male) in the VA data source.

| Characteristic (total count = 13324) | Before stratification | | | After stratification | | |
|---|---|---|---|---|---|---|
| | T (%) | C (%) | SDf | T (%) | C (%) | SDf |
| index year: 2021 | 39.8 | 2.2 | 1.04 | 39.8 | 6.0 | 0.88 |
| procedure_occurrence during day -365 through 0 days relative to index: Counseling | -0.1 | 37.1 | -1.08 | -0.1 | 18.2 | -0.66 |
| index year: 2015 | -0.1 | 5.8 | -0.35 | -0.1 | 15.8 | -0.61 |
| index year: 2020 | 33.1 | 3.9 | 0.81 | 33.1 | 10.5 | 0.57 |
| index year: 2016 | 0.5 | 5.4 | -0.29 | 0.5 | 14.8 | -0.56 |
| drug_era group during day -365 through 0 days relative to index: SARS-CoV-2 (COVID-19) vaccine, mRNA spike protein | 15.4 | 0.7 | 0.56 | 15.4 | 1.8 | 0.50 |
| condition_era group during day -365 through 0 days relative to index: Atherosclerosis of coronary artery without angina pectoris | 28.4 | 3.8 | 0.71 | 28.4 | 10.4 | 0.47 |
| observation during day -365 through 0 days relative to index: Requires influenza virus vaccination | -0.1 | 26.3 | -0.84 | -0.1 | 9.5 | -0.46 |
| index year: 2014 | -0.1 | 5.8 | -0.35 | -0.1 | 9.4 | -0.45 |
| index year: 2017 | 2.6 | 5.5 | -0.15 | 2.6 | 14.9 | -0.45 |

**Supplementary Table 245:** Most extreme patient characteristics balance diagnostics after matching for SGLT2I (male) vs SU (male) in the VA data source.

| Characteristic (total count = 0) | Before matching | | | After matching | | |
|---|---|---|---|---|---|---|
| | T (%) | C (%) | SDf | T (%) | C (%) | SDf |

##### 3.3.9 SIDIAP

**Supplementary Figure 24:** Preference score distributions, propensity score stratified (middle column) and matched (right column) covariate balance and negative control error distributions (bottom row) for SGLT2I (male) vs SU (male) in the SIDIAP data source.

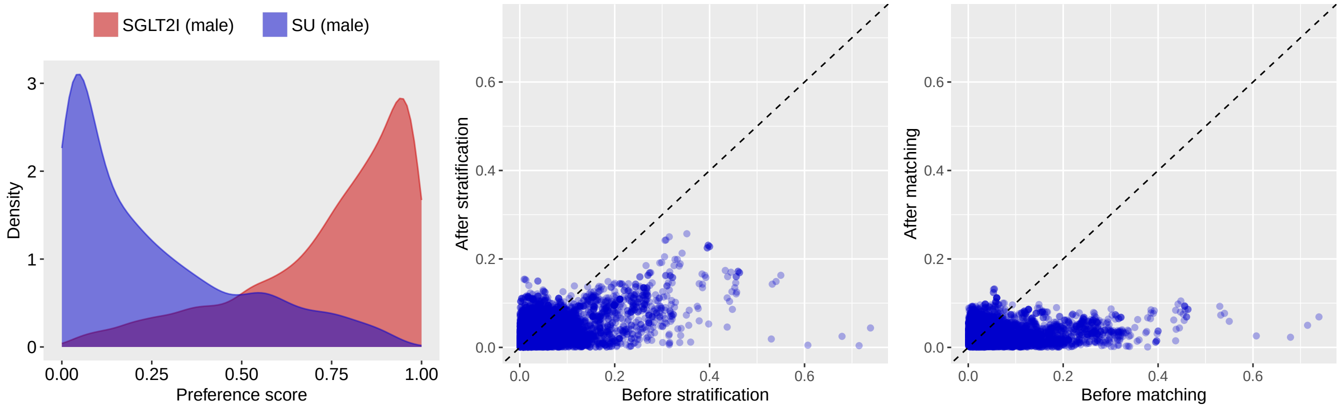

**Supplementary Table 246:** Most extreme patient characteristics balance diagnostics after stratification for SGLT2I (male) vs SU (male) in the SIDIAP data source.

| | Before stratification | | | After stratification | | |
|---|---|---|---|---|---|---|
| Characteristic (total count = 7569) | T (%) | C (%) | SDf | T (%) | C (%) | SDf |
| procedure_occurrence during day -365 through 0 days relative to index: Ultrasonography of Right and Left Heart | 5.9 | -0.1 | 0.35 | 3.9 | 0.3 | 0.26 |
| drug_era group during day -365 through 0 days relative to index: sacubitril | 4.8 | -0.1 | 0.32 | 3.2 | -0.1 | 0.25 |
| drug_era group during day -30 through 0 days relative to index: sacubitril | 4.5 | -0.1 | 0.31 | 3.0 | -0.1 | 0.24 |
| drug_era group during day 0 through 0 days relative to index: sacubitril | 4.5 | -0.1 | 0.31 | 3.0 | -0.1 | 0.24 |
| drug_era group during day -30 through 0 days relative to index: ezetimibe | 10.5 | 1.3 | 0.40 | 7.2 | 2.3 | 0.23 |
| drug_era group during day -30 through 0 days relative to index: Other lipid modifying agents | 10.5 | 1.3 | 0.40 | 7.2 | 2.3 | 0.23 |
| drug_era group during day -365 through 0 days relative to index: ezetimibe | 10.9 | 1.5 | 0.40 | 7.5 | 2.5 | 0.23 |
| drug_era group during day -365 through 0 days relative to index: Other lipid modifying agents | 10.9 | 1.5 | 0.40 | 7.5 | 2.5 | 0.23 |

| (Continued) | T (%) | C (%) | SDf | T (%) | C (%) | SDf |
|---|---|---|---|---|---|---|
| drug_era group during day 0 through 0 days relative to index: ezetimibe | 10.2 | 1.2 | 0.40 | 7.0 | 2.3 | 0.23 |
| drug_era group during day 0 through 0 days relative to index: Other lipid modifying agents | 10.2 | 1.2 | 0.40 | 7.0 | 2.3 | 0.23 |

**Supplementary Table 247:** Most extreme patient characteristics balance diagnostics after matching for SGLT2I (male) vs SU (male) in the SIDIAP data source.

| | Before matching | | | After matching | | |
|---|---|---|---|---|---|---|
| Characteristic (total count = 6187) | T (%) | C (%) | SDf | T (%) | C (%) | SDf |
| drug_era group during day -365 through 0 days relative to index: testosterone | 0.7 | 0.3 | 0.06 | 1.0 | -0.1 | 0.13 |
| drug_era group during day -365 through 0 days relative to index: 3-oxoandrosten (4) derivatives | 0.7 | 0.3 | 0.06 | 1.0 | -0.1 | 0.13 |
| drug_era group during day -365 through 0 days relative to index: ANDROGENS | 0.7 | 0.3 | 0.05 | 1.0 | -0.1 | 0.13 |
| drug_era group during day -365 through 0 days relative to index: ANDROGENS AND FEMALE SEX HORMONES IN COMBINATION | 0.7 | 0.3 | 0.05 | 1.0 | 0.1 | 0.12 |
| drug_era group during day -365 through 0 days relative to index: Androgens and estrogens | 0.7 | 0.3 | 0.05 | 1.0 | 0.1 | 0.12 |
| drug_era group during day -30 through 0 days relative to index: testosterone | 0.5 | 0.2 | 0.06 | 0.7 | -0.1 | 0.11 |
| drug_era group during day 0 through 0 days relative to index: testosterone | 0.5 | 0.2 | 0.06 | 0.7 | -0.1 | 0.11 |
| drug_era group during day -30 through 0 days relative to index: ANDROGENS | 0.5 | 0.2 | 0.06 | 0.7 | -0.1 | 0.11 |
| drug_era group during day 0 through 0 days relative to index: ANDROGENS | 0.5 | 0.2 | 0.06 | 0.7 | -0.1 | 0.11 |
| drug_era group during day -30 through 0 days relative to index: 3-oxoandrosten (4) derivatives | 0.5 | 0.2 | 0.06 | 0.7 | -0.1 | 0.11 |

#### 3.4 GLP1RA (male) vs DPP4I (male)

##### 3.4.1 CCAE

**Supplementary Figure 25:** Preference score distributions, propensity score stratified (middle column) and matched (right column) covariate balance and negative control error distributions (bottom row) for GLP1RA (male) vs DPP4I (male) in the CCAE data source.

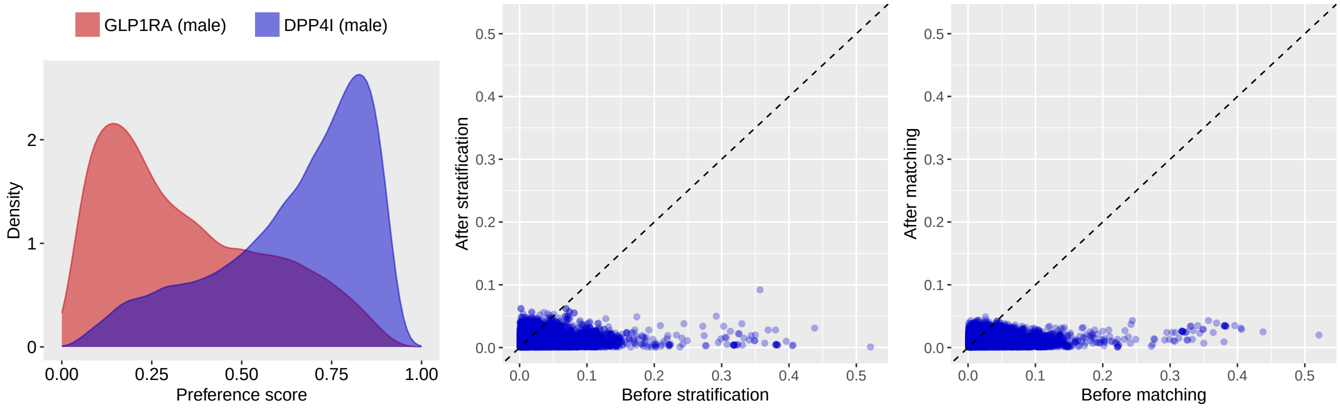

**Supplementary Table 248:** Most extreme patient characteristics balance diagnostics after stratification for GLP1RA (male) vs DPP4I (male) in the CCAE data source.

| Characteristic (total count = 10277) | Before stratification | | | After stratification | | |
|---|---|---|---|---|---|---|
| | T (%) | C (%) | SDf | T (%) | C (%) | SDf |
| index year: 2012 | 2.6 | 11.7 | 0.36 | 7.5 | 10.1 | 0.09 |
| drug_era group during day -365 through 0 days relative to index: niacin | 2.0 | 3.1 | 0.07 | 4.0 | 2.8 | -0.06 |
| drug_era group during day -365 through 0 days relative to index: Nicotinic acid and derivatives | 2.0 | 3.1 | 0.07 | 4.0 | 2.8 | -0.06 |
| condition_era group during day -30 through 0 days relative to index: Cramp | 0.1 | 0.1 | -0.00 | 0.3 | 0.1 | -0.06 |
| condition_era group during day -30 through 0 days relative to index: Cramp in limb | 0.1 | 0.1 | -0.00 | 0.3 | 0.1 | -0.06 |
| drug_era group during day -365 through 0 days relative to index: PERIPHERAL VASODILATORS | 2.1 | 3.2 | 0.07 | 4.0 | 3.0 | -0.06 |
| drug_era group during day -365 through 0 days relative to index: PERIPHERAL VASODILATORS | 2.1 | 3.2 | 0.07 | 4.0 | 3.0 | -0.06 |

| (Continued) | T (%) | C (%) | SDf | T (%) | C (%) | SDf |
|---|---|---|---|---|---|---|
| condition_era group during day -30 through 0 days relative to index: Idiopathic peripheral autonomic neuropathy | 0.1 | 0.1 | -0.02 | 0.3 | 0.1 | -0.06 |
| condition_era group during day -30 through 0 days relative to index: Cerebrovascular disease | 0.3 | 0.6 | 0.04 | 0.2 | 0.5 | 0.06 |
| measurement during day -365 through 0 days relative to index: Cell Fractions/Differential [Interpretation] in Blood | 0.1 | 0.5 | 0.08 | 0.2 | 0.5 | 0.06 |

**Supplementary Table 249:** Most extreme patient characteristics balance diagnostics after matching for GLP1RA (male) vs DPP4I (male) in the CCAE data source.

| | Before matching | | | After matching | | |
|---|---|---|---|---|---|---|
| Characteristic (total count = 10410) | T (%) | C (%) | SDf | T (%) | C (%) | SDf |
| procedure_occurrence during day -365 through 0 days relative to index: Electroencephalogram (EEG); including recording awake and drowsy | 0.1 | 0.2 | 0.03 | 0.1 | 0.3 | 0.05 |
| condition_era group during day -365 through 0 days relative to index: Chronic pancreatitis | 0.1 | 0.1 | 0.03 | 0.1 | 0.2 | 0.04 |
| condition_era group during day -365 through 0 days relative to index: Alopecia | 0.1 | 0.2 | 0.02 | 0.1 | 0.3 | 0.04 |
| index year: 2012 | 2.6 | 11.7 | 0.36 | 3.2 | 4.0 | 0.04 |
| observation during day -365 through 0 days relative to index: Requires influenza virus vaccination | 5.7 | 12.7 | 0.24 | 6.4 | 5.4 | -0.04 |
| condition_era group during day -30 through 0 days relative to index: Asthenia | 0.2 | 0.2 | 0.01 | 0.2 | 0.4 | 0.04 |
| measurement during day -365 through 0 days relative to index: HLA typing; A, B, or C (eg, A10, B7, B27), single antigen | 0.1 | 0.1 | 0.01 | 0.1 | 0.3 | 0.04 |
| observation during day -365 through 0 days relative to index: Body mass index 25-29 - overweight | 1.8 | 1.8 | -0.00 | 2.1 | 2.7 | 0.04 |
| drug_era group during day -365 through 0 days relative to index: ascorbic acid | 0.6 | 1.0 | 0.04 | 0.7 | 0.4 | -0.04 |
| drug_era group during day -365 through 0 days relative to index: Ascorbic acid (vitamin C), plain | 0.6 | 1.0 | 0.04 | 0.7 | 0.4 | -0.04 |

##### 3.4.2 ODOD

**Supplementary Figure 26:** Preference score distributions, propensity score stratified (middle column) and matched (right column) covariate balance and negative control error distributions (bottom row) for GLP1RA (male) vs DPP4I (male) in the ODOD data source.

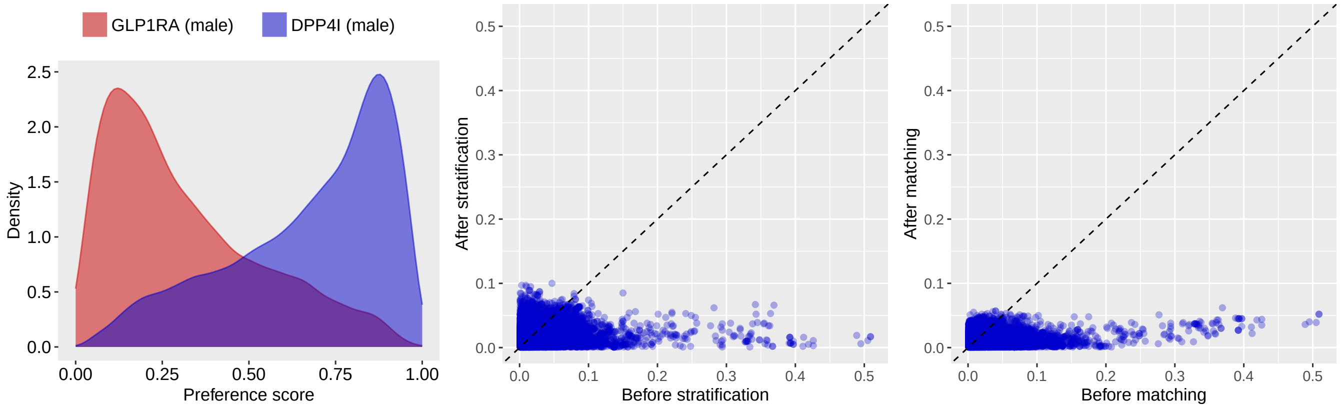

**Supplementary Table 250:** Most extreme patient characteristics balance diagnostics after stratification for GLP1RA (male) vs DPP4I (male) in the ODOD data source.

| | Before stratification | | | After stratification | | |
|---|---|---|---|---|---|---|
| Characteristic (total count = 14565) | T (%) | C (%) | SDf | T (%) | C (%) | SDf |
| procedure_occurrence during day -30 through 0 days relative to index: Transitional Care Management Services with the following required elements: Communication (direct contact, telephone, electronic) with the patient and/or caregiver within 2 business days of discharge Medical decision making of at least moderate complexity | 0.3 | 0.1 | -0.05 | 0.8 | 0.1 | -0.10 |
| measurement below normal range during day -365 through 0 days relative to index: Creatinine; clearance | 0.1 | 0.1 | 0.01 | 0.7 | 0.1 | -0.10 |
| measurement during day -365 through 0 days relative to index: Creatinine [Mass/time] in 24 hour Urine | 0.4 | 0.4 | 0.00 | 1.3 | 0.4 | -0.10 |
| measurement below normal range during day -365 through 0 days relative to index: Creatinine renal clearance in 24 hour Urine and Serum or Plasma | 0.1 | 0.1 | 0.02 | 0.7 | 0.1 | -0.10 |
| condition_era group during day -365 through 0 days relative to index: Corneal degeneration | 1.0 | 1.1 | 0.01 | 2.2 | 1.0 | -0.10 |
| condition_era group during day -30 through 0 days relative to index: Infective disorder of head | 0.3 | 0.2 | -0.01 | 0.9 | 0.2 | -0.09 |

| (Continued) | T (%) | C (%) | SDf | T (%) | C (%) | SDf |
|---|---|---|---|---|---|---|
| measurement during day -365 through 0 days relative to index: Creatinine renal clearance in 24 hour Urine and Serum or Plasma | 0.1 | 0.2 | 0.01 | 0.8 | 0.2 | -0.09 |
| measurement within normal range during day -365 through 0 days relative to index: Creatinine; clearance | 0.1 | 0.2 | 0.01 | 0.7 | 0.1 | -0.09 |
| condition_era group during day -365 through 0 days relative to index: Arcus senilis | 0.2 | 0.1 | -0.03 | 0.7 | 0.1 | -0.09 |
| condition_era group during day -365 through 0 days relative to index: Arcus of cornea | 0.2 | 0.1 | -0.03 | 0.7 | 0.1 | -0.09 |

**Supplementary Table 251:** Most extreme patient characteristics balance diagnostics after matching for GLP1RA (male) vs DPP4I (male) in the ODOD data source.

| | Before matching | | | After matching | | |
|---|---|---|---|---|---|---|
| Characteristic (total count = 14884) | T (%) | C (%) | SDf | T (%) | C (%) | SDf |
| observation during day -365 through 0 days relative to index: Requires influenza virus vaccination | 4.1 | 14.7 | 0.37 | 5.4 | 4.1 | -0.06 |
| condition_era group during day -30 through 0 days relative to index: Non-toxic uninodular goiter | 0.4 | 0.2 | -0.04 | 0.4 | 0.1 | -0.06 |
| condition_era group during day -30 through 0 days relative to index: Uninodular goiter | 0.4 | 0.2 | -0.04 | 0.4 | 0.1 | -0.06 |
| condition_era group during day -30 through 0 days relative to index: Varicose ulcer of lower extremity | 0.2 | 0.1 | -0.04 | 0.2 | -0.1 | -0.06 |
| observation during day -365 through 0 days relative to index: Medical/Surgical Supplies - Incident to radiology | 0.2 | 0.2 | 0.01 | 0.2 | -0.1 | -0.06 |
| procedure_occurrence during day -365 through 0 days relative to index: Counseling | 0.2 | 0.6 | 0.06 | 0.3 | 0.1 | -0.05 |
| condition_era group during day -365 through 0 days relative to index: Closed fracture of phalanx of foot | 0.1 | 0.2 | 0.03 | 0.1 | 0.3 | 0.05 |
| condition_era group during day -365 through 0 days relative to index: Benign hypertension | 7.1 | 25.2 | 0.51 | 9.2 | 7.8 | -0.05 |
| condition_era group during day -365 through 0 days relative to index: Benign essential hypertension | 7.1 | 25.2 | 0.51 | 9.2 | 7.8 | -0.05 |
| condition_era group during day -365 through 0 days relative to index: Parkinson’s disease | 0.2 | 0.6 | 0.06 | 0.2 | 0.6 | 0.05 |

##### 3.4.3 MDCR

**Supplementary Figure 27:** Preference score distributions, propensity score stratified (middle column) and matched (right column) covariate balance and negative control error distributions (bottom row) for GLP1RA (male) vs DPP4I (male) in the MDCR data source.

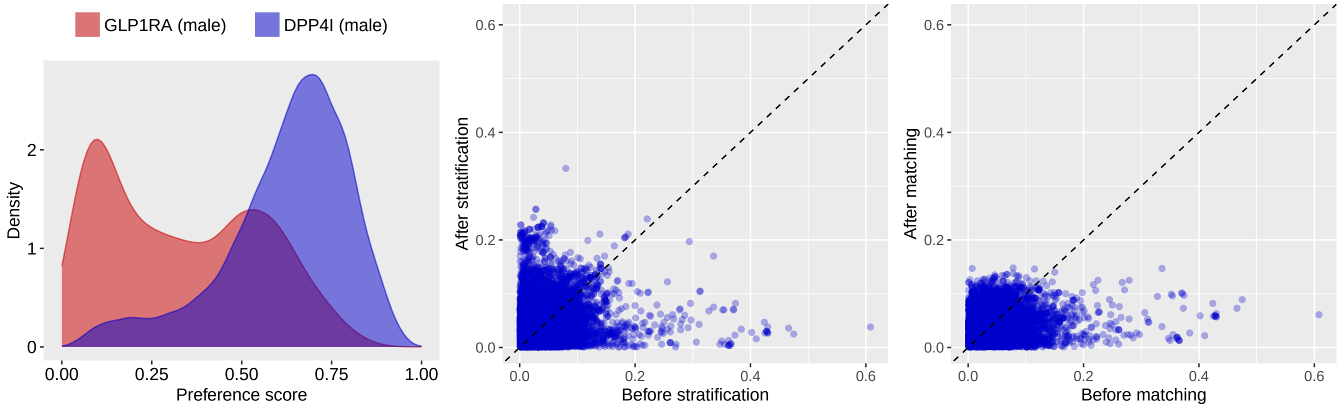

**Supplementary Table 252:** Most extreme patient characteristics balance diagnostics after stratification for GLP1RA (male) vs DPP4I (male) in the MDCR data source.

| | Before stratification | | | After stratification | | |
|---|---|---|---|---|---|---|
| Characteristic (total count = 13947) | T (%) | C (%) | SDf | T (%) | C (%) | SDf |
| device_exposure during day -365 through 0 days relative to index: Nebulizer, with compressor | 2.0 | 1.0 | -0.08 | 7.8 | 1.0 | -0.33 |
| condition_era group during day -365 through 0 days relative to index: Acute exacerbation of chronic bronchitis | 1.6 | 2.0 | 0.03 | 7.1 | 1.9 | -0.26 |
| condition_era group during day -365 through 0 days relative to index: Acute exacerbation of chronic obstructive bronchitis | 1.6 | 2.0 | 0.03 | 7.1 | 1.9 | -0.26 |
| condition_era group during day -365 through 0 days relative to index: Chronic bronchitis | 2.7 | 3.1 | 0.02 | 8.6 | 3.0 | -0.24 |
| condition_era group during day -30 through 0 days relative to index: Renal failure syndrome | -0.5 | 3.4 | 0.22 | -0.2 | 3.2 | 0.24 |
| condition_era group during day -365 through 0 days relative to index: Emphysematous bronchitis | 1.9 | 2.5 | 0.04 | 7.3 | 2.4 | -0.23 |
| condition_era group during day -365 through 0 days relative to index: Obstruction of bronchus | 1.9 | 2.5 | 0.04 | 7.3 | 2.4 | -0.23 |
| condition_era group during day -365 through 0 days relative to index: Obstruction of lower respiratory tract | 1.9 | 2.5 | 0.04 | 7.3 | 2.4 | -0.23 |

| (Continued) | T (%) | C (%) | SDf | T (%) | C (%) | SDf |
|---|---|---|---|---|---|---|
| device_exposure during day -365 through 0 days relative to index: TRUETRACK GLUCOSE TEST STRIPS | -0.5 | -0.1 | -0.05 | 2.6 | -0.1 | -0.23 |
| drug_era group during day 0 through 0 days relative to index: CORTICOSTEROIDS | 1.9 | 1.9 | -0.00 | 6.4 | 1.9 | -0.23 |

**Supplementary Table 253:** Most extreme patient characteristics balance diagnostics after matching for GLP1RA (male) vs DPP4I (male) in the MDCR data source.

| | Before matching | | | After matching | | |
|---|---|---|---|---|---|---|
| Characteristic (total count = 9490) | T (%) | C (%) | SDf | T (%) | C (%) | SDf |
| condition_era group during day -30 through 0 days relative to index: Ingrowing nail | 1.2 | 0.5 | -0.08 | 1.4 | -0.6 | -0.15 |
| index year: 2014 | 2.8 | 11.2 | 0.34 | 3.3 | 6.4 | 0.15 |
| condition_era group during day -365 through 0 days relative to index: Chronic metabolic disorder | 0.7 | 0.8 | 0.01 | -0.5 | 2.1 | 0.15 |
| observation during day -365 through 0 days relative to index: Bmi documented outside normal parameters, no follow-up plan documented, no reason given | -0.5 | 1.5 | 0.12 | 0.5 | 2.1 | 0.15 |
| condition_era group during day -365 through 0 days relative to index: Metabolic syndrome X | 2.8 | 0.8 | -0.15 | 2.6 | 0.8 | -0.14 |
| drug_era group during day -365 through 0 days relative to index: adenosine | 2.1 | 1.1 | -0.08 | 1.8 | -0.6 | -0.14 |
| condition_era group during day -365 through 0 days relative to index: Chronic respiratory failure | -0.5 | 0.8 | 0.07 | -0.5 | 1.4 | 0.14 |
| condition_era group during day -365 through 0 days relative to index: Chronic respiratory insufficiency | -0.5 | 0.8 | 0.07 | -0.5 | 1.4 | 0.14 |
| procedure_occurrence during day -365 through 0 days relative to index: Computed tomography, cervical spine; without contrast material | 0.9 | 1.4 | 0.05 | 0.8 | 2.6 | 0.14 |
| procedure_occurrence during day -365 through 0 days relative to index: Cystourethroscopy, with calibration and/or dilation of urethral stricture or stenosis, with or without meatotomy, with or without injection procedure for cystography, male or female | -0.5 | 0.6 | 0.08 | -0.5 | 1.1 | 0.13 |

##### 3.4.4 Open Claims

**Supplementary Figure 28:** Preference score distributions, propensity score stratified (middle column) and matched (right column) covariate balance and negative control error distributions (bottom row) for GLP1RA (male) vs DPP4I (male) in the Open Claims data source.

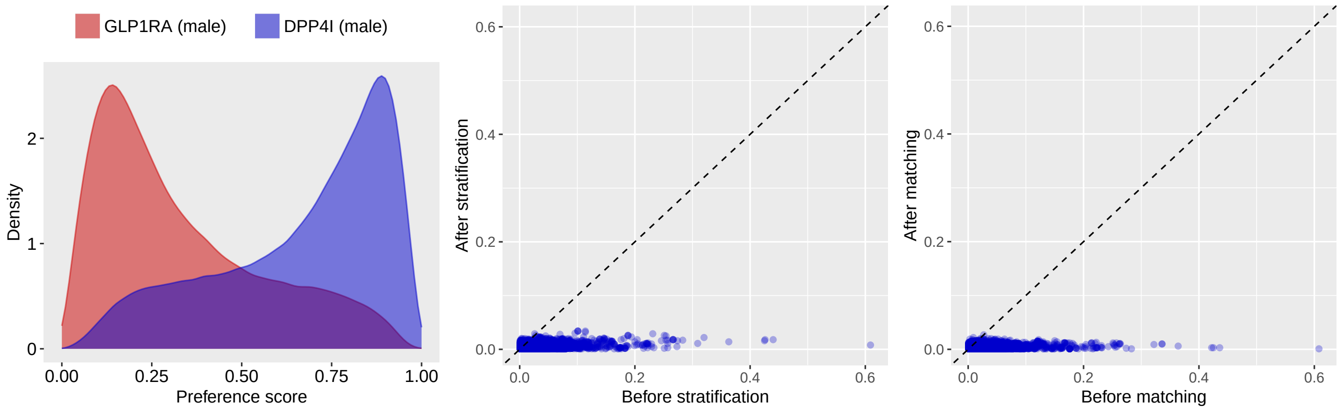

**Supplementary Table 254:** Most extreme patient characteristics balance diagnostics after stratification for GLP1RA (male) vs DPP4I (male) in the Open Claims data source.

| Characteristic (total count = 9685) | Before stratification | | | After stratification | | |
|---|---|---|---|---|---|---|
| | T (%) | C (%) | SDf | T (%) | C (%) | SDf |
| condition_era group during day -30 through 0 days relative to index: Renal failure syndrome | 0.3 | 1.3 | 0.11 | 0.7 | 1.1 | 0.03 |
| condition_era group during day -30 through 0 days relative to index: Acute nephropathy | 0.3 | 1.1 | 0.10 | 0.6 | 0.9 | 0.03 |
| condition_era group during day -30 through 0 days relative to index: Acute renal failure syndrome | 0.3 | 1.1 | 0.10 | 0.6 | 0.9 | 0.03 |
| condition_era group during day -30 through 0 days relative to index: Acute renal impairment | 0.3 | 1.1 | 0.10 | 0.6 | 0.9 | 0.03 |
| procedure_occurrence during day -30 through 0 days relative to index: Radiologic examination, chest; single view, frontal | 0.1 | 0.9 | 0.11 | 0.4 | 0.7 | 0.03 |
| condition_era group during day -30 through 0 days relative to index: Acute genitourinary disorder | 0.4 | 1.2 | 0.10 | 0.7 | 1.0 | 0.03 |
| age group: 75 - 79 | 3.6 | 9.2 | 0.23 | 7.0 | 7.7 | 0.03 |
| drug_era group during day -365 through 0 days relative to index: lorcaserin | 0.1 | 0.1 | -0.03 | 0.2 | 0.1 | -0.03 |

| (Continued) | T (%) | C (%) | SDf | T (%) | C (%) | SDf |
|---|---|---|---|---|---|---|
| observation during day -30 through 0 days relative to index: Principal physician of record | 0.1 | 0.5 | 0.07 | 0.3 | 0.5 | 0.03 |
| condition_era group during day -365 through 0 days relative to index: Obesity | 18.0 | 9.5 | -0.25 | 12.2 | 11.4 | -0.03 |

**Supplementary Table 255:** Most extreme patient characteristics balance diagnostics after matching for GLP1RA (male) vs DPP4I (male) in the Open Claims data source.

| | Before matching | | | After matching | | |
|---|---|---|---|---|---|---|
| Characteristic (total count = 9531) | T (%) | C (%) | SDf | T (%) | C (%) | SDf |
| drug_era group during day -365 through 0 days relative to index: lorcaserin | 0.1 | 0.1 | -0.03 | 0.2 | 0.1 | -0.03 |
| Charlson index - Romano adaptation | 328.7 | 330.7 | 0.01 | 328.7 | 333.6 | 0.02 |
| procedure_occurrence during day -365 through 0 days relative to index: Ambulatory continuous glucose monitoring of interstitial tissue fluid via a subcutaneous sensor for a minimum of 72 hours; analysis, interpretation and report | 0.3 | 0.1 | -0.04 | 0.2 | 0.1 | -0.02 |
| condition_era group during day -30 through 0 days relative to index: Acute respiratory failure | 0.2 | 0.4 | 0.04 | 0.2 | 0.3 | 0.02 |
| condition_era group during day -30 through 0 days relative to index: Acute metabolic disorder | 0.2 | 0.3 | 0.02 | 0.2 | 0.3 | 0.02 |
| index month: 3 | 11.5 | 9.6 | -0.06 | 10.3 | 10.9 | 0.02 |
| drug_era group during day -365 through 0 days relative to index: evolocumab | 0.3 | 0.1 | -0.05 | 0.3 | 0.2 | -0.02 |
| condition_era group during day -30 through 0 days relative to index: Malignant tumor of digestive organ | 0.1 | 0.3 | 0.05 | 0.1 | 0.2 | 0.02 |
| drug_era group during day 0 through 0 days relative to index: PROPULSIVES | 0.1 | 0.2 | 0.03 | 0.1 | 0.2 | 0.02 |
| drug_era group during day 0 through 0 days relative to index: Propulsives | 0.1 | 0.2 | 0.03 | 0.1 | 0.2 | 0.02 |

##### 3.4.5 OEHR

**Supplementary Figure 29:** Preference score distributions, propensity score stratified (middle column) and matched (right column) covariate balance and negative control error distributions (bottom row) for GLP1RA (male) vs DPP4I (male) in the OEHR data source.

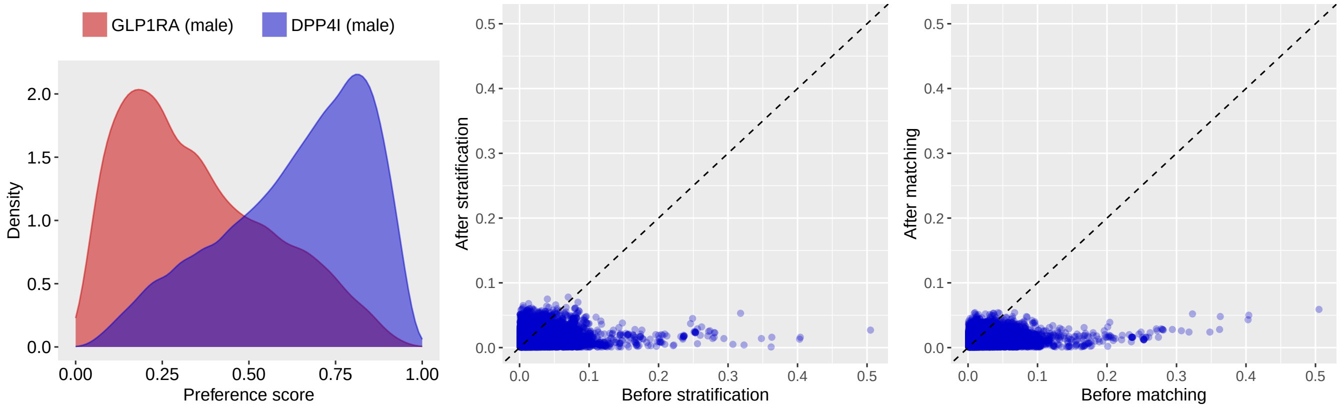

**Supplementary Table 256:** Most extreme patient characteristics balance diagnostics after stratification for GLP1RA (male) vs DPP4I (male) in the OEHR data source.

| | Before stratification | | | After stratification | | |
|---|---|---|---|---|---|---|
| Characteristic (total count = 11319) | T (%) | C (%) | SDf | T (%) | C (%) | SDf |
| condition_era group during day -365 through 0 days relative to index: Dementia | 0.3 | 0.7 | 0.07 | 0.2 | 0.7 | 0.08 |
| procedure_occurrence during day -365 through 0 days relative to index: Endoscopic polypectomy of large intestine | 0.2 | 0.4 | 0.04 | 1.0 | 0.4 | -0.07 |
| race = Asian | 1.6 | 2.9 | 0.08 | 1.6 | 2.6 | 0.07 |
| observation during day -30 through 0 days relative to index: History of urinary tract infection | 0.1 | 0.1 | -0.01 | 0.4 | 0.1 | -0.07 |
| measurement during day -30 through 0 days relative to index: Culture, bacterial; blood, aerobic, with isolation and presumptive identification of isolates (includes anaerobic culture, if appropriate) | 0.1 | 0.4 | 0.05 | 0.1 | 0.4 | 0.07 |
| drug_era group during day -30 through 0 days relative to index: enoxaparin | 0.7 | 0.7 | -0.01 | 1.3 | 0.7 | -0.07 |
| condition_era group during day -365 through 0 days relative to index: Acute respiratory failure | 0.3 | 0.5 | 0.04 | 0.1 | 0.5 | 0.06 |
| observation during day -30 through 0 days relative to index: Colorectal cancer screening results documented and reviewed (PV) | 0.2 | 0.6 | 0.07 | 0.2 | 0.5 | 0.06 |

| (Continued) | T (%) | C (%) | SDf | T (%) | C (%) | SDf |
|---|---|---|---|---|---|---|
| drug_era group during day -30 through 0 days relative to index: pravastatin | 5.0 | 6.0 | 0.04 | 7.2 | 5.7 | -0.06 |
| measurement during day -30 through 0 days relative to index: Complex medical conditions action plan | 0.1 | 0.1 | 0.01 | 0.4 | 0.1 | -0.06 |

**Supplementary Table 257:** Most extreme patient characteristics balance diagnostics after matching for GLP1RA (male) vs DPP4I (male) in the OEHR data source.

| | Before matching | | | After matching | | |
|---|---|---|---|---|---|---|
| Characteristic (total count = 11681) | T (%) | C (%) | SDf | T (%) | C (%) | SDf |
| index year: 2020 | 26.2 | 7.8 | -0.51 | 22.2 | 24.7 | 0.06 |
| drug_era group during day -365 through 0 days relative to index: icosapent ethyl | 0.5 | 0.1 | -0.07 | 0.5 | 0.2 | -0.05 |
| drug_era group during day -365 through 0 days relative to index: levetiracetam | 0.2 | 0.5 | 0.04 | 0.2 | 0.5 | 0.05 |
| measurement during day -365 through 0 days relative to index: Procalcitonin (PCT) | 0.3 | 0.3 | 0.01 | 0.2 | 0.6 | 0.05 |
| measurement during day -365 through 0 days relative to index: Thyroxine; total | 0.3 | 0.6 | 0.04 | 0.3 | 0.7 | 0.05 |
| measurement within normal range during day -365 through 0 days relative to index: Creatinine 24 hour urine panel - 24 hour Urine | 0.1 | 0.3 | 0.04 | 0.1 | 0.4 | 0.05 |
| observation during day -365 through 0 days relative to index: Most recent LDL-C greater than or equal to 130 mg/dL (CAD) (DM) | 0.1 | 0.3 | 0.04 | 0.1 | 0.3 | 0.05 |
| condition_era group during day -30 through 0 days relative to index: Malignant lymphoma | 0.3 | 0.2 | -0.01 | 0.3 | 0.1 | -0.05 |
| index year: 2019 | 23.5 | 11.4 | -0.32 | 23.9 | 21.7 | -0.05 |
| procedure_occurrence during day -365 through 0 days relative to index: Subsequent observation care, per day, for the evaluation and management of a patient, which requires at least 2 of these 3 key components: A detailed interval history; A detailed examination; Medical decision making of high complexity. Counseling and/or c | 0.1 | 0.1 | 0.03 | 0.1 | 0.3 | 0.05 |

### 3.4.6 VA

**Supplementary Figure 30:** Preference score distributions, propensity score stratified (middle column) and matched (right column) covariate balance and negative control error distributions (bottom row) for GLP1RA (male) vs DPP4I (male) in the VA data source.

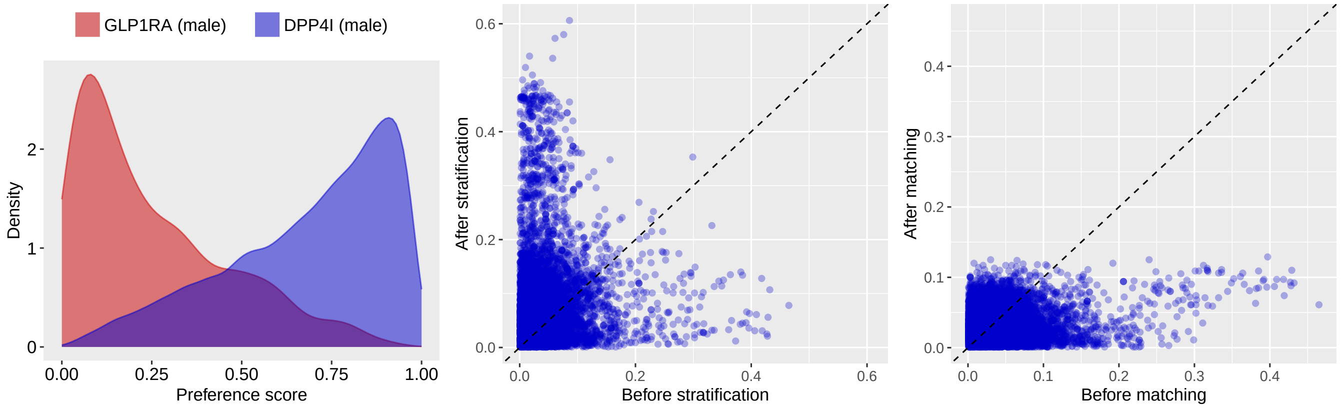

**Supplementary Table 258:** Most extreme patient characteristics balance diagnostics after stratification for GLP1RA (male) vs DPP4I (male) in the VA data source.

| Characteristic (total count = 13755) | Before stratification | | | After stratification | | |
|---|---|---|---|---|---|---|
| | T (%) | C (%) | SDf | T (%) | C (%) | SDf |
| measurement within normal range during day -365 through 0 days relative to index: Creatine kinase [Enzymatic activity/volume] in Serum or Plasma | 4.5 | 6.4 | 0.09 | 28.4 | 6.3 | -0.61 |
| measurement during day -365 through 0 days relative to index: Creatine kinase (CK), (CPK); total | 5.7 | 7.6 | 0.08 | 29.2 | 7.5 | -0.58 |
| measurement during day -365 through 0 days relative to index: Creatine kinase [Enzymatic activity/volume] in Serum or Plasma | 6.3 | 7.9 | 0.06 | 29.3 | 7.8 | -0.57 |
| procedure_occurrence during day -365 through 0 days relative to index: Intravenous infusion, for therapy, prophylaxis, or diagnosis (specify substance or drug); initial, up to 1 hour | 0.8 | 0.9 | 0.02 | 15.2 | 0.9 | -0.54 |
| procedure_occurrence during day -365 through 0 days relative to index: Intravenous infusion, hydration; initial, 31 minutes to 1 hour | -0.7 | 1.0 | 0.06 | 15.2 | 1.0 | -0.54 |
| condition_era group during day -365 through 0 days relative to index: Lesion of esophagus | 1.6 | 1.4 | -0.01 | 15.5 | 1.4 | -0.52 |

| (Continued) | T (%) | C (%) | SDf | T (%) | C (%) | SDf |
|---|---|---|---|---|---|---|
| procedure_occurrence during day -365 through 0 days relative to index: Instrument-based ocular screening (eg, photoscreening, automated-refraction), bilateral; with on-site analysis | -0.7 | 0.3 | 0.02 | 12.3 | 0.3 | -0.51 |
| procedure_occurrence during day -365 through 0 days relative to index: Sensorimotor examination with multiple measurements of ocular deviation (eg, restrictive or paretic muscle with diplopia) with interpretation and report (separate procedure) | -0.7 | 0.2 | 0.01 | 11.7 | 0.2 | -0.50 |
| procedure_occurrence during day -30 through 0 days relative to index: Radiologic examination, knee; complete, 4 or more views | -0.7 | 0.4 | -0.04 | 11.9 | 0.4 | -0.49 |
| drug_era group during day -365 through 0 days relative to index: Fentanyl | 1.8 | 2.2 | 0.03 | 15.9 | 2.2 | -0.49 |

**Supplementary Table 259:** Most extreme patient characteristics balance diagnostics after matching for GLP1RA (male) vs DPP4I (male) in the VA data source.

| | Before matching | | | After matching | | |
|---|---|---|---|---|---|---|
| Characteristic (total count = 11609) | T (%) | C (%) | SDf | T (%) | C (%) | SDf |
| condition_era group during day -365 through 0 days relative to index: Obesity | 46.9 | 28.1 | -0.40 | 41.7 | 48.1 | 0.13 |
| observation during day -30 through 0 days relative to index: Dietary management surveillance | 10.8 | 4.4 | -0.24 | 8.5 | 12.3 | 0.12 |
| procedure_occurrence during day -365 through 0 days relative to index: Oxycodone | 0.9 | 1.2 | 0.03 | 0.8 | 2.4 | 0.12 |
| condition_era group during day -365 through 0 days relative to index: Paralytic syndrome on one side of the body | 1.0 | 0.4 | -0.07 | 1.0 | -0.8 | -0.12 |
| measurement during day -30 through 0 days relative to index: Microalbumin/Creatinine [Mass Ratio] in Urine | 15.5 | 23.0 | 0.19 | 16.5 | 12.3 | -0.12 |
| procedure_occurrence during day -365 through 0 days relative to index: Direct skilled nursing services of a registered nurse (rn) in the home health or hospice setting, each 15 minutes | -0.7 | 0.5 | 0.01 | -0.6 | 1.6 | 0.12 |
| observation during day -30 through 0 days relative to index: Health status | 1.2 | 0.5 | -0.07 | 1.3 | -0.8 | -0.12 |
| drug_era group during day -30 through 0 days relative to index: Thiazides, combinations with other drugs | 67.8 | 72.5 | 0.10 | 67.3 | 61.6 | -0.12 |
| drug_era group during day 0 through 0 days relative to index: Thiazides, combinations with other drugs | 66.1 | 70.4 | 0.09 | 65.5 | 59.8 | -0.12 |
| measurement during day -30 through 0 days relative to index: Leukocytes [#/volume] in Blood by Automated count | 33.3 | 48.5 | 0.31 | 34.6 | 29.2 | -0.12 |

##### 3.5 GLP1RA (male) vs SU (male)

###### 3.5.1 CCAE

**Supplementary Figure 31:** Preference score distributions, propensity score stratified (middle column) and matched (right column) covariate balance and negative control error distributions (bottom row) for GLP1RA (male) vs SU (male) in the CCAE data source.

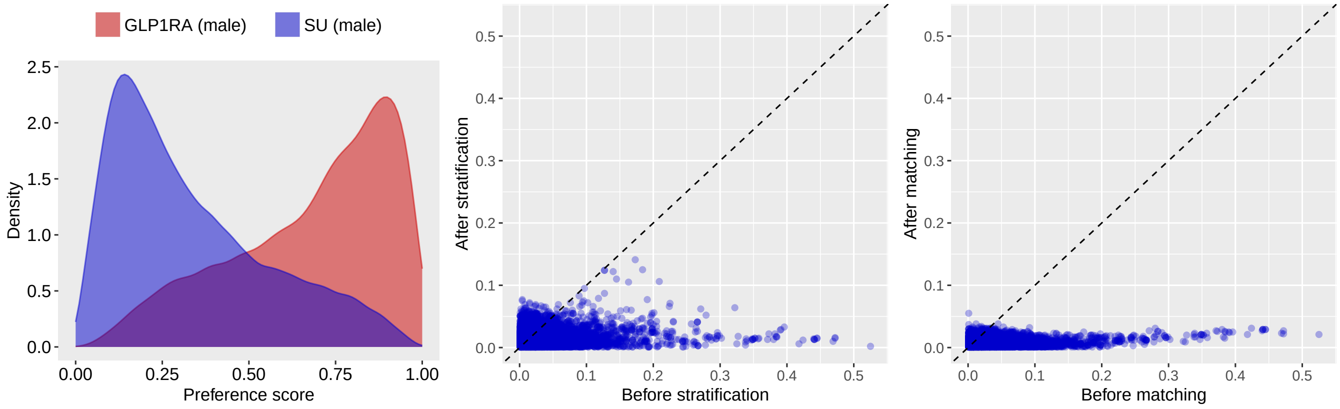

**Supplementary Table 260:** Most extreme patient characteristics balance diagnostics after stratification for GLP1RA (male) vs SU (male) in the CCAE data source.

| | Before stratification | | | After stratification | | |
|---|---|---|---|---|---|---|
| Characteristic (total count = 10083) | T (%) | C (%) | SDf | T (%) | C (%) | SDf |
| measurement during day -365 through 0 days relative to index: Cholesterol, serum or whole blood, total | 1.1 | 3.7 | -0.17 | 1.1 | 3.1 | -0.14 |
| measurement during day -365 through 0 days relative to index: Potassium; serum, plasma or whole blood | 2.1 | 5.7 | -0.18 | 2.7 | 5.1 | -0.12 |
| measurement during day -365 through 0 days relative to index: Hemoglobin A1c/Hemoglobin.total in Blood by calculation | 0.1 | 1.0 | -0.13 | 0.1 | 0.9 | -0.12 |
| measurement above normal range during day -365 through 0 days relative to index: Hemoglobin A1c/Hemoglobin.total in Blood by calculation | 0.1 | 1.0 | -0.13 | 0.1 | 0.9 | -0.12 |
| measurement during day -365 through 0 days relative to index: Immunoassay for infectious agent antibody, qualitative or semiquantitative, single step method (eg, reagent strip) | 0.1 | 1.3 | -0.14 | 0.2 | 1.2 | -0.12 |
| measurement during day -30 through 0 days relative to index: Cholesterol, serum or whole blood, total | 0.3 | 1.7 | -0.14 | 0.4 | 1.4 | -0.11 |

| (Continued) | T (%) | C (%) | SDf | T (%) | C (%) | SDf |
|---|---|---|---|---|---|---|
| measurement during day -365 through 0 days relative to index: Lipid panel This panel must include the following: Cholesterol, serum, total (82465) Lipoprotein, direct measurement, high density cholesterol (HDL cholesterol) (83718) Triglycerides (84478) | 85.3 | 77.2 | 0.21 | 83.4 | 79.3 | 0.11 |
| measurement during day -30 through 0 days relative to index: Potassium; serum, plasma or whole blood | 0.4 | 2.3 | -0.16 | 0.8 | 2.1 | -0.10 |
| device_exposure during day -365 through 0 days relative to index: ONE TOUCH ULTRA CONTROL SOLN | 0.1 | 0.7 | -0.10 | 0.1 | 0.6 | -0.10 |
| procedure_occurrence during day -365 through 0 days relative to index: Collection of venous blood by venipuncture | 75.4 | 69.7 | 0.13 | 75.1 | 71.2 | 0.09 |

**Supplementary Table 261:** Most extreme patient characteristics balance diagnostics after matching for GLP1RA (male) vs SU (male) in the CCAE data source.

| | Before matching | | | After matching | | |
|---|---|---|---|---|---|---|
| Characteristic (total count = 10260) | T (%) | C (%) | SDf | T (%) | C (%) | SDf |
| observation during day -30 through 0 days relative to index: Mental Health Substance Abuse Coverage Indicator | 99.9 | 99.9 | -0.00 | 99.9 | 99.7 | 0.06 |
| observation during day -365 through 0 days relative to index: Overexertion, strenuous movements and vibration | 0.3 | 0.1 | 0.04 | 0.3 | 0.1 | 0.04 |
| observation during day -365 through 0 days relative to index: Statin therapy prescribed or currently being taken (CAD) | 0.1 | 0.1 | 0.01 | 0.1 | 0.2 | -0.04 |
| observation during day -365 through 0 days relative to index: Advance care planning discussion documented in the medical record (COA) | 0.1 | 0.1 | 0.00 | 0.1 | 0.2 | -0.04 |
| procedure_occurrence during day -365 through 0 days relative to index: Cystourethroscopy, with insertion of indwelling ureteral stent (eg, Gibbons or double-J type) | 0.3 | 0.4 | -0.02 | 0.2 | 0.4 | -0.03 |
| procedure_occurrence during day -365 through 0 days relative to index: Basic radiation dosimetry calculation, central axis depth dose calculation, TDF, NSD, gap calculation, off axis factor, tissue inhomogeneity factors, calculation of non-ionizing radiation surface and depth dose, as required during course of treatment, onl | 0.1 | 0.3 | -0.05 | 0.1 | 0.2 | -0.03 |
| drug_era group during day 0 through 0 days relative to index: carbamazepine | 0.1 | 0.1 | -0.02 | 0.1 | 0.2 | -0.03 |
| condition_era group during day -365 through 0 days relative to index: Foreign body in hand | 0.1 | 0.1 | -0.01 | 0.1 | 0.2 | -0.03 |
| procedure_occurrence during day -365 through 0 days relative to index: Ambulatory continuous glucose monitoring of interstitial tissue fluid via a subcutaneous sensor for a minimum of 72 hours; analysis, interpretation and report | 0.6 | 0.1 | 0.09 | 0.4 | 0.2 | 0.03 |
| procedure_occurrence during day -30 through 0 days relative to index: Magnetic resonance (eg, proton) imaging, any joint of upper extremity; without contrast material(s) | 0.1 | 0.2 | -0.02 | 0.1 | 0.2 | -0.03 |

##### 3.5.2 ODOD

**Supplementary Figure 32:** Preference score distributions, propensity score stratified (middle column) and matched (right column) covariate balance and negative control error distributions (bottom row) for GLP1RA (male) vs SU (male) in the ODOD data source.

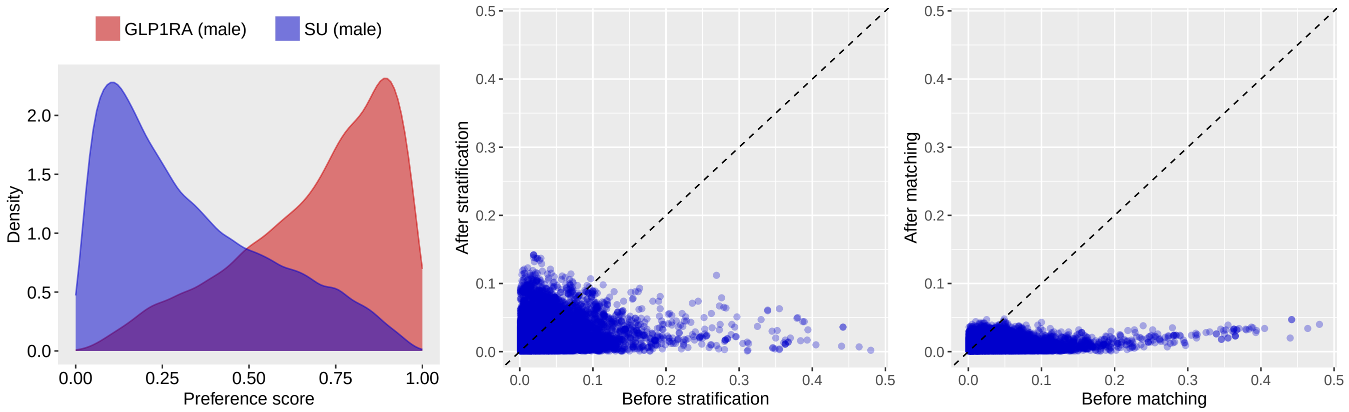

**Supplementary Table 262:** Most extreme patient characteristics balance diagnostics after stratification for GLP1RA (male) vs SU (male) in the ODOD data source.

| | Before stratification | | | After stratification | | |
|---|---|---|---|---|---|---|
| Characteristic (total count = 14296) | T (%) | C (%) | SDf | T (%) | C (%) | SDf |
| condition_era group during day -30 through 0 days relative to index: Benign prostatic hypertrophy with outflow obstruction | 2.0 | 2.2 | -0.02 | 4.9 | 2.2 | 0.14 |
| condition_era group during day -30 through 0 days relative to index: Prostatic obstruction | 2.0 | 2.2 | -0.02 | 4.9 | 2.2 | 0.14 |
| condition_era group during day -30 through 0 days relative to index: Obstruction of urethra | 2.0 | 2.2 | -0.02 | 4.9 | 2.2 | 0.14 |
| drug_era group during day -30 through 0 days relative to index: Second-generation cephalosporins | 0.2 | 0.3 | -0.02 | 1.7 | 0.3 | 0.14 |
| measurement during day -30 through 0 days relative to index: Lipase | 0.9 | 1.2 | -0.03 | 3.2 | 1.2 | 0.14 |
| condition_era group during day -30 through 0 days relative to index: Urinary outflow obstruction | 2.0 | 2.3 | -0.02 | 4.9 | 2.3 | 0.14 |
| condition_era group during day -30 through 0 days relative to index: Lower urinary tract obstructive syndrome | 2.0 | 2.3 | -0.02 | 4.9 | 2.3 | 0.14 |
| condition_era group during day -30 through 0 days relative to index: Orthostatic hypotension | 0.1 | 0.2 | -0.02 | 1.4 | 0.2 | 0.14 |

| (Continued) | T (%) | C (%) | SDf | T (%) | C (%) | SDf |
|---|---|---|---|---|---|---|
| drug_era group during day -30 through 0 days relative to index: cefuroxime | 0.2 | 0.3 | -0.03 | 1.6 | 0.3 | 0.13 |
| condition_era group during day -30 through 0 days relative to index: Urinary tract obstruction | 2.2 | 2.6 | -0.02 | 5.0 | 2.5 | 0.13 |

**Supplementary Table 263:** Most extreme patient characteristics balance diagnostics after matching for GLP1RA (male) vs SU (male) in the ODOD data source.

| | Before matching | | | After matching | | |
|---|---|---|---|---|---|---|
| Characteristic (total count = 14988) | T (%) | C (%) | SDf | T (%) | C (%) | SDf |
| age group: 20 - 24 | 0.3 | 0.1 | 0.05 | 0.3 | 0.1 | 0.05 |
| condition_era group during day -365 through 0 days relative to index: Benign hypertension | 7.1 | 22.3 | -0.44 | 7.7 | 6.5 | 0.05 |
| condition_era group during day -365 through 0 days relative to index: Benign essential hypertension | 7.1 | 22.3 | -0.44 | 7.7 | 6.5 | 0.05 |
| procedure_occurrence during day -365 through 0 days relative to index: Remote imaging for detection of retinal disease (eg, retinopathy in a patient with diabetes) with analysis and report under physician supervision, unilateral or bilateral | -0.1 | 0.1 | -0.03 | -0.1 | 0.2 | -0.05 |
| device_exposure during day -365 through 0 days relative to index: Deluxe frame | -0.1 | 0.1 | -0.05 | -0.1 | 0.1 | -0.04 |
| drug_era group during day 0 through 0 days relative to index: dutasteride | 0.4 | 0.5 | -0.01 | 0.5 | 0.2 | 0.04 |
| measurement during day -365 through 0 days relative to index: Dehydroepiandrosterone-sulfate (DHEA-S) | 1.1 | 0.3 | 0.10 | 0.6 | 1.0 | -0.04 |
| measurement during day -365 through 0 days relative to index: Microalbumin/Creatinine [Ratio] in Urine | 0.4 | 0.2 | 0.05 | 0.5 | 0.2 | 0.04 |
| measurement above normal range during day -365 through 0 days relative to index: Microalbumin/Creatinine [Ratio] in Urine | 0.3 | 0.1 | 0.05 | 0.4 | 0.2 | 0.04 |
| measurement within normal range during day -365 through 0 days relative to index: oxyMORphone [Mass/volume] in Urine by Confirmatory method | -0.1 | 0.1 | -0.02 | -0.1 | 0.2 | -0.04 |

##### 3.5.3 MDCR

**Supplementary Figure 33:** Preference score distributions, propensity score stratified (middle column) and matched (right column) covariate balance and negative control error distributions (bottom row) for GLP1RA (male) vs SU (male) in the MDCR data source.

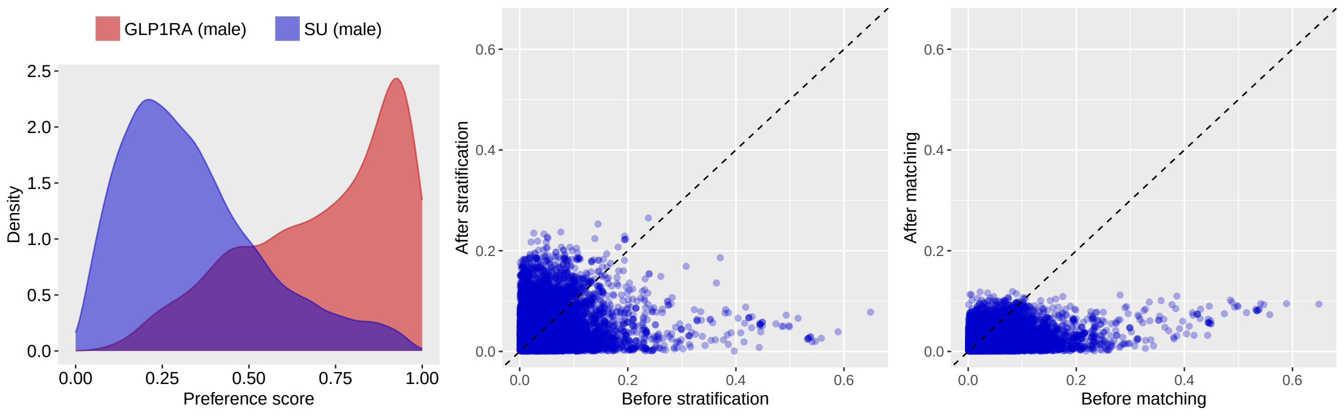

**Supplementary Table 264:** Most extreme patient characteristics balance diagnostics after stratification for GLP1RA (male) vs SU (male) in the MDCR data source.

| | Before stratification | | | After stratification | | |
|---|---|---|---|---|---|---|
| Characteristic (total count = 11430) | T (%) | C (%) | SDf | T (%) | C (%) | SDf |
| condition_era group during day -30 through 0 days relative to index: Renal failure syndrome | -0.5 | 3.8 | -0.24 | 0.2 | 4.0 | -0.27 |
| measurement during day -365 through 0 days relative to index: Potassium; serum, plasma or whole blood | 1.8 | 4.3 | -0.14 | 0.5 | 4.3 | -0.25 |
| condition_era group during day -365 through 0 days relative to index: Joint effusion of ankle AND/OR foot | 0.7 | 0.2 | 0.08 | 3.3 | 0.2 | 0.24 |
| condition_era group during day -365 through 0 days relative to index: Mature cataract | 0.6 | 0.8 | -0.03 | 4.4 | 0.7 | 0.23 |
| device_exposure during day -365 through 0 days relative to index: Nebulizer, with compressor | 2.0 | 1.4 | 0.04 | 5.7 | 1.4 | 0.23 |
| condition_era group during day -30 through 0 days relative to index: Acute genitourinary disorder | 0.5 | 3.0 | -0.19 | 0.2 | 3.2 | -0.23 |
| drug_era group during day -365 through 0 days relative to index: Corticosteroids, potent, combinations with antibiotics | 62.6 | 60.1 | 0.05 | 49.6 | 60.9 | -0.23 |
| drug_era group during day -365 through 0 days relative to index: Corticosteroids, very potent, combinations with antibiotics | 60.8 | 58.3 | 0.05 | 47.9 | 59.1 | -0.23 |

| (Continued) | T (%) | C (%) | SDf | T (%) | C (%) | SDf |
|---|---|---|---|---|---|---|
| condition_era group during day -30 through 0 days relative to index: Congestive heart failure | 2.1 | 4.6 | -0.14 | 0.8 | 4.4 | -0.22 |
| condition_era group during day -30 through 0 days relative to index: Acute nephropathy | -0.5 | 2.9 | -0.20 | 0.2 | 3.0 | -0.22 |

**Supplementary Table 265:** Most extreme patient characteristics balance diagnostics after matching for GLP1RA (male) vs SU (male) in the MDCR data source.

| | Before matching | | | After matching | | |
|---|---|---|---|---|---|---|
| Characteristic (total count = 10685) | T (%) | C (%) | SDf | T (%) | C (%) | SDf |
| observation during day -365 through 0 days relative to index: Bmi documented outside normal parameters, no follow-up plan documented, no reason given | -0.5 | 1.1 | -0.08 | -0.5 | 1.6 | -0.12 |
| condition_era group during day -365 through 0 days relative to index: Chronic metabolic disorder | 0.7 | 0.5 | 0.02 | -0.5 | 1.4 | -0.12 |
| condition_era group during day -365 through 0 days relative to index: Dependence on enabling machine or device | 1.1 | 0.7 | 0.04 | 1.0 | 2.5 | -0.12 |
| condition_era group during day -30 through 0 days relative to index: Neoplasm of hematopoietic cell type | -0.5 | 1.2 | -0.09 | -0.5 | 1.6 | -0.12 |
| observation during day -365 through 0 days relative to index: Patient screened for future fall risk; documentation of no falls in the past year or only 1 fall without injury in the past year (GER) | 1.2 | 1.3 | -0.01 | 1.0 | 2.4 | -0.11 |
| observation during day -365 through 0 days relative to index: FH: Cardiovascular disease | -0.5 | 0.3 | 0.00 | -0.5 | 1.1 | -0.11 |
| device_exposure during day -365 through 0 days relative to index: Water chamber for humidifier, used with positive airway pressure device, replacement, each | 8.4 | 2.2 | 0.28 | 6.9 | 10.0 | -0.11 |
| condition_era group during day -365 through 0 days relative to index: Measurement finding outside reference range | 78.1 | 60.6 | 0.39 | 76.7 | 81.2 | -0.11 |
| drug_era group during day -365 through 0 days relative to index: icosapent ethyl | 1.8 | 0.1 | 0.17 | 1.5 | 0.4 | 0.11 |
| drug_era group during day 0 through 0 days relative to index: naproxen | 0.6 | 1.0 | -0.05 | -0.5 | 1.3 | -0.11 |

##### 3.5.4 Open Claims

**Supplementary Figure 34:** Preference score distributions, propensity score stratified (middle column) and matched (right column) covariate balance and negative control error distributions (bottom row) for GLP1RA (male) vs SU (male) in the Open Claims data source.

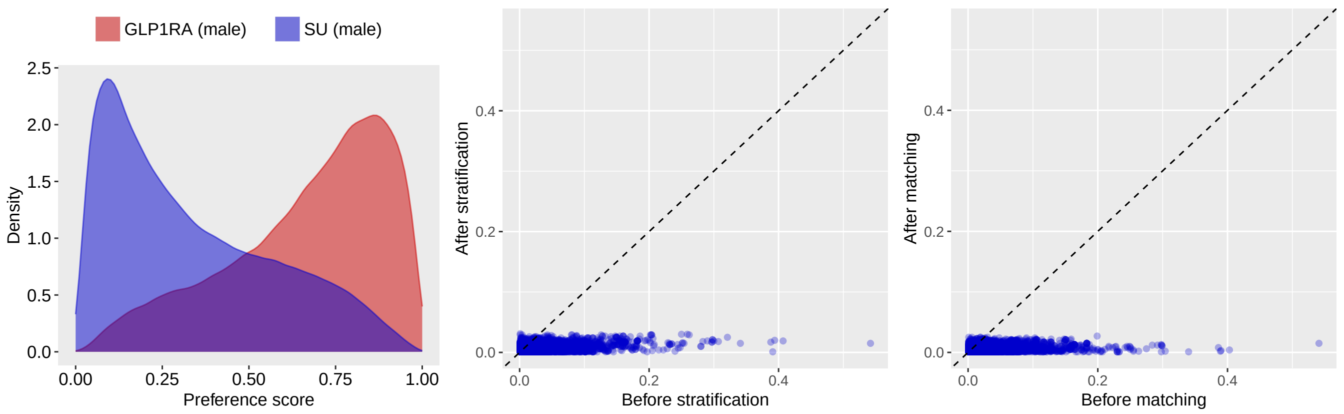

**Supplementary Table 266:** Most extreme patient characteristics balance diagnostics after stratification for GLP1RA (male) vs SU (male) in the Open Claims data source.

| | Before stratification | | | After stratification | | |
|---|---|---|---|---|---|---|
| Characteristic (total count = 9651) | T (%) | C (%) | SDf | T (%) | C (%) | SDf |
| drug_era group during day -365 through 0 days relative to index: fenofibrate | 6.7 | 6.8 | -0.00 | 7.5 | 6.7 | 0.03 |
| observation during day -365 through 0 days relative to index: Synchronous Telemedicine Service Rendered Via a Real-Time Interactive Audio and Video Telecommunications System | 7.7 | 2.2 | 0.26 | 3.4 | 2.9 | 0.03 |
| condition_era group during day -365 through 0 days relative to index: Morbid obesity | 9.8 | 3.6 | 0.25 | 5.0 | 4.3 | 0.03 |
| condition_era group during day -365 through 0 days relative to index: Obesity | 18.0 | 9.1 | 0.26 | 11.1 | 10.2 | 0.03 |
| drug_era group during day -365 through 0 days relative to index: ANTIOBESITY PREPARATIONS, EXCL. DIET PRODUCTS | 8.0 | 3.3 | 0.20 | 4.5 | 3.9 | 0.03 |
| drug_era group during day -365 through 0 days relative to index: ANTIOBESITY PREPARATIONS, EXCL. DIET PRODUCTS | 8.0 | 3.3 | 0.20 | 4.5 | 3.9 | 0.03 |
| drug_era group during day -365 through 0 days relative to index: Centrally acting antiobesity products | 8.0 | 3.3 | 0.20 | 4.5 | 3.9 | 0.03 |

| (Continued) | T (%) | C (%) | SDf | T (%) | C (%) | SDf |
|---|---|---|---|---|---|---|
| observation during day -365 through 0 days relative to index: Body mass index 40+ - severely obese | 6.0 | 2.0 | 0.20 | 3.0 | 2.5 | 0.03 |
| procedure_occurrence during day -30 through 0 days relative to index: Radiologic examination, chest; single view, frontal | 0.1 | 1.0 | -0.13 | 0.6 | 0.9 | -0.03 |
| condition_era group during day -365 through 0 days relative to index: Cerebral infarction due to thrombosis of cerebral arteries | 0.1 | 0.5 | -0.08 | 0.3 | 0.4 | -0.03 |

**Supplementary Table 267:** Most extreme patient characteristics balance diagnostics after matching for GLP1RA (male) vs SU (male) in the Open Claims data source.

| | Before matching | | | After matching | | |
|---|---|---|---|---|---|---|
| Characteristic (total count = 9387) | T (%) | C (%) | SDf | T (%) | C (%) | SDf |
| Diabetes Comorbidity Severity Index (DCSI) | 230.2 | 279.6 | -0.20 | 231.5 | 237.6 | -0.03 |
| drug_era group during day -365 through 0 days relative to index: Combinations for eradication of Helicobacter pylori | 39.2 | 37.8 | 0.03 | 38.3 | 39.5 | -0.03 |
| drug_era group during day -365 through 0 days relative to index: SULFONAMIDES AND TRIMETHOPRIM | 14.5 | 15.2 | -0.02 | 14.3 | 15.2 | -0.03 |
| drug_era group during day -365 through 0 days relative to index: Combinations of sulfonamides and trimethoprim, incl. derivatives | 14.5 | 15.2 | -0.02 | 14.3 | 15.2 | -0.03 |
| Charlson index - Romano adaptation | 328.7 | 330.1 | -0.01 | 326.2 | 331.8 | -0.03 |
| drug_era group during day -365 through 0 days relative to index: Non-selective monoamine reuptake inhibitors | 2.6 | 2.6 | 0.00 | 2.5 | 3.0 | -0.03 |
| condition_era group during day -30 through 0 days relative to index: Hyperglycemia | 13.7 | 10.4 | 0.10 | 13.5 | 14.3 | -0.02 |
| condition_era group during day -30 through 0 days relative to index: Prediabetes | 0.7 | 0.1 | 0.09 | 0.5 | 0.4 | 0.02 |
| procedure_occurrence during day -365 through 0 days relative to index: Office or other outpatient visit for the evaluation and management of an established patient, which requires at least 2 of these 3 key components: A detailed history; A detailed examination; Medical decision making of moderate complexity. Counseling and/o | 46.3 | 43.9 | 0.05 | 44.2 | 45.3 | -0.02 |
| condition_era group during day -30 through 0 days relative to index: Measurement finding above reference range | 30.0 | 28.8 | 0.03 | 29.5 | 30.6 | -0.02 |

##### 3.5.5 OEHR

**Supplementary Figure 35:** Preference score distributions, propensity score stratified (middle column) and matched (right column) covariate balance and negative control error distributions (bottom row) for GLP1RA (male) vs SU (male) in the OEHR data source.

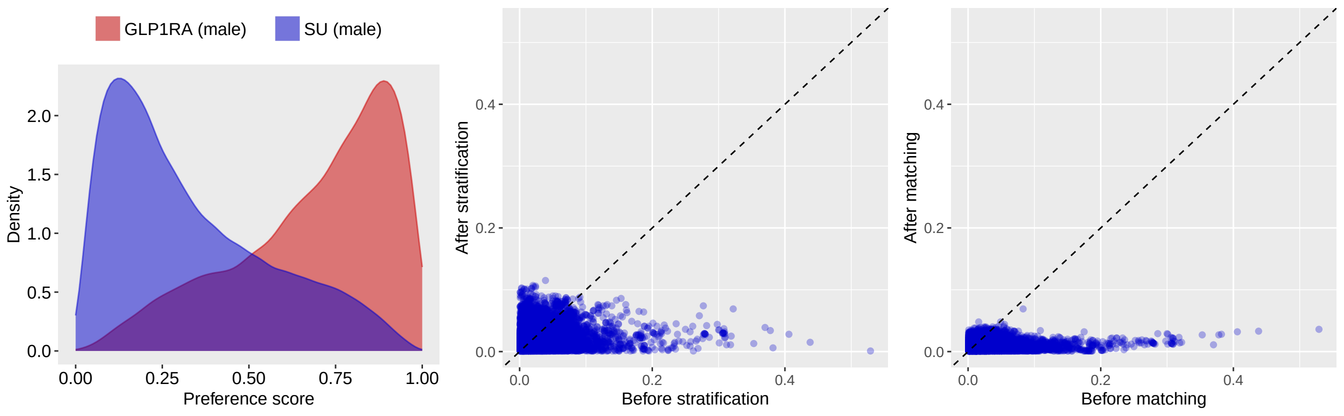

**Supplementary Table 268:** Most extreme patient characteristics balance diagnostics after stratification for GLP1RA (male) vs SU (male) in the OEHR data source.

| | Before stratification | | | After stratification | | |
|---|---|---|---|---|---|---|
| Characteristic (total count = 11076) | T (%) | C (%) | SDf | T (%) | C (%) | SDf |
| drug_era group during day 0 through 0 days relative to index: ceftriaxone | 0.3 | 0.5 | -0.04 | 1.7 | 0.5 | 0.12 |
| drug_era group during day -30 through 0 days relative to index: enoxaparin | 0.7 | 0.9 | -0.02 | 2.2 | 0.9 | 0.11 |
| drug_era group during day 0 through 0 days relative to index: Third-generation cephalosporins | 0.5 | 0.7 | -0.02 | 1.8 | 0.7 | 0.11 |
| drug_era group during day 0 through 0 days relative to index: enoxaparin | 0.5 | 0.7 | -0.02 | 1.8 | 0.6 | 0.10 |
| drug_era group during day -30 through 0 days relative to index: ceftriaxone | 0.5 | 0.7 | -0.02 | 1.8 | 0.7 | 0.10 |
| drug_era group during day 0 through 0 days relative to index: sodium phosphate, dibasic | 0.1 | 0.1 | -0.00 | 0.7 | 0.1 | 0.10 |
| condition_era group during day -365 through 0 days relative to index: Malaise and fatigue | 0.2 | 0.1 | 0.02 | 0.8 | 0.1 | 0.10 |
| condition_era group during day -365 through 0 days relative to index: Generally unwell | 0.2 | 0.1 | 0.02 | 0.8 | 0.1 | 0.10 |

| (Continued) | T (%) | C (%) | SDf | T (%) | C (%) | SDf |
|---|---|---|---|---|---|---|
| drug_era group during day 0 through 0 days relative to index: acetazolamide | 0.1 | 0.1 | 0.00 | 0.7 | 0.1 | 0.10 |
| drug_era group during day -30 through 0 days relative to index: acetazolamide | 0.1 | 0.1 | 0.01 | 0.7 | 0.1 | 0.10 |

**Supplementary Table 269:** Most extreme patient characteristics balance diagnostics after matching for GLP1RA (male) vs SU (male) in the OEHR data source.

| | Before matching | | | After matching | | |
|---|---|---|---|---|---|---|
| Characteristic (total count = 11828) | T (%) | C (%) | SDf | T (%) | C (%) | SDf |
| drug_era group during day -365 through 0 days relative to index: icosapent ethyl | 0.5 | 0.1 | 0.08 | 0.5 | 0.1 | 0.07 |
| condition_era group during day -365 through 0 days relative to index: Mass of pancreas | 0.1 | 0.3 | -0.05 | -0.1 | 0.2 | -0.05 |
| condition_era group during day -30 through 0 days relative to index: Idiopathic osteoarthritis | 0.1 | 0.2 | -0.02 | 0.1 | 0.3 | -0.05 |
| measurement during day -365 through 0 days relative to index: Lipoprotein, blood; quantitation of lipoprotein particle number(s) (eg, by nuclear magnetic resonance spectroscopy), includes lipoprotein particle subclass(es), when performed | 0.4 | 0.1 | 0.06 | 0.3 | 0.1 | 0.04 |
| observation during day -365 through 0 days relative to index: History of bariatric surgical procedure | 0.3 | 0.1 | 0.05 | 0.3 | 0.1 | 0.04 |
| measurement during day -30 through 0 days relative to index: Gases, blood, any combination of pH, pCO2, pO2, CO2, HCO3 (including calculated O2 saturation) | 0.1 | 0.3 | -0.05 | 0.1 | 0.3 | -0.04 |
| drug_era group during day 0 through 0 days relative to index: chlorpheniramine | -0.1 | 0.2 | -0.04 | -0.1 | 0.2 | -0.04 |
| drug_era group during day 0 through 0 days relative to index: Substituted alkylamines | 0.1 | 0.2 | -0.03 | 0.1 | 0.3 | -0.04 |
| condition_era group during day -365 through 0 days relative to index: Nonunion of fracture | -0.1 | 0.1 | -0.02 | -0.1 | 0.1 | -0.04 |
| procedure_occurrence during day -365 through 0 days relative to index: Transthoracic echocardiography, with contrast, or without contrast followed by with contrast, real-time with image documentation (2d), includes m-mode recording, when performed, during rest and cardiovascular stress test using treadmill, bicycle exerci... | 0.3 | 0.1 | 0.05 | 0.3 | 0.1 | 0.04 |

### 3.5.6 VA

**Supplementary Figure 36:** Preference score distributions, propensity score stratified (middle column) and matched (right column) covariate balance and negative control error distributions (bottom row) for GLP1RA (male) vs SU (male) in the VA data source.

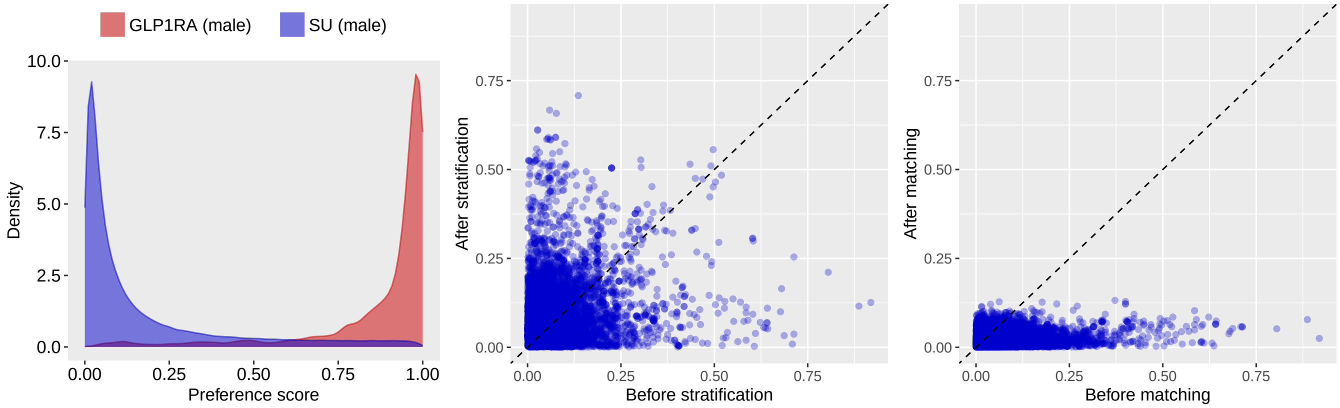

**Supplementary Table 270:** Most extreme patient characteristics balance diagnostics after stratification for GLP1RA (male) vs SU (male) in the VA data source.

| Characteristic (total count = 12644) | Before stratification T (%) | Before stratification C (%) | Before stratification SDf | After stratification T (%) | After stratification C (%) | After stratification SDf |
|---|---|---|---|---|---|---|
| measurement below normal range during day -365 through 0 days relative to index: Specific gravity of Urine by Test strip | 0.7 | 2.4 | -0.14 | 25.9 | 2.4 | 0.71 |
| condition_era group during day -365 through 0 days relative to index: Mass of lower limb | 4.5 | 3.4 | 0.06 | 26.5 | 3.6 | 0.67 |
| condition_era group during day -365 through 0 days relative to index: Lump on extremities | 5.7 | 4.1 | 0.08 | 27.5 | 4.4 | 0.66 |
| condition_era group during day -365 through 0 days relative to index: Acquired hallux malleus | -0.7 | 0.7 | -0.03 | 17.7 | 0.7 | 0.61 |
| condition_era group during day -365 through 0 days relative to index: Acquired dorsiflexion deformity of foot | -0.7 | 0.7 | -0.03 | 17.7 | 0.7 | 0.61 |
| condition_era group during day -365 through 0 days relative to index: Flexion deformity | 2.0 | 1.1 | 0.07 | 17.9 | 1.2 | 0.59 |
| condition_era group during day -365 through 0 days relative to index: Flexion deformity of toe | 2.0 | 1.1 | 0.07 | 17.9 | 1.2 | 0.59 |
| condition_era group during day -365 through 0 days relative to index: Acquired deformity of joint of big toe | 2.3 | 1.5 | 0.06 | 18.8 | 1.6 | 0.59 |

| (Continued) | T (%) | C (%) | SDf | T (%) | C (%) | SDf |
|---|---|---|---|---|---|---|
| condition_era group during day -365 through 0 days relative to index: Acquired deformity of joint of foot | 2.3 | 1.6 | 0.05 | 18.9 | 1.7 | 0.59 |
| condition_era group during day -365 through 0 days relative to index: Acquired deformity of limb | 4.6 | 3.6 | 0.05 | 23.1 | 3.8 | 0.58 |

**Supplementary Table 271:** Most extreme patient characteristics balance diagnostics after matching for GLP1RA (male) vs SU (male) in the VA data source.

| | Before matching | | | After matching | | |
|---|---|---|---|---|---|---|
| Characteristic (total count = 11841) | T (%) | C (%) | SDf | T (%) | C (%) | SDf |
| measurement during day -365 through 0 days relative to index: Body weight | 89.1 | 98.1 | -0.37 | 89.6 | 85.2 | 0.13 |
| measurement during day -365 through 0 days relative to index: Heart rate | 89.2 | 98.6 | -0.40 | 89.9 | 85.7 | 0.13 |
| procedure_occurrence during day -365 through 0 days relative to index: Direct skilled nursing services of a registered nurse (rn) in the home health or hospice setting, each 15 minutes | -0.7 | 0.1 | 0.06 | -0.8 | 1.7 | -0.13 |
| measurement during day -365 through 0 days relative to index: Hemoglobin A1c/Hemoglobin.total in Blood | 91.4 | 95.7 | -0.18 | 92.1 | 88.3 | 0.13 |
| measurement during day -365 through 0 days relative to index: Systolic blood pressure | 89.4 | 98.7 | -0.40 | 90.0 | 86.0 | 0.12 |
| measurement during day -365 through 0 days relative to index: Diastolic blood pressure | 89.4 | 98.7 | -0.40 | 90.0 | 86.0 | 0.12 |
| measurement during day -365 through 0 days relative to index: Hemoglobin; glycosylated (A1C) | 88.1 | 94.2 | -0.22 | 88.6 | 84.5 | 0.12 |
| procedure_occurrence during day -30 through 0 days relative to index: Direct skilled nursing services of a registered nurse (rn) in the home health or hospice setting, each 15 minutes | -0.7 | 0.1 | 0.04 | -0.8 | 1.1 | -0.12 |
| measurement during day -365 through 0 days relative to index: Respiratory rate | 87.6 | 95.1 | -0.27 | 88.3 | 84.3 | 0.12 |
| condition_era group during day -365 through 0 days relative to index: Adjustment disorder with anxious mood | -0.7 | 0.4 | -0.01 | -0.8 | 1.0 | -0.12 |

#### 3.6 DPP4I (male) vs SU (male)

##### 3.6.1 CCAE

**Supplementary Figure 37:** Preference score distributions, propensity score stratified (middle column) and matched (right column) covariate balance and negative control error distributions (bottom row) for DPP4I (male) vs SU (male) in the CCAE data source.

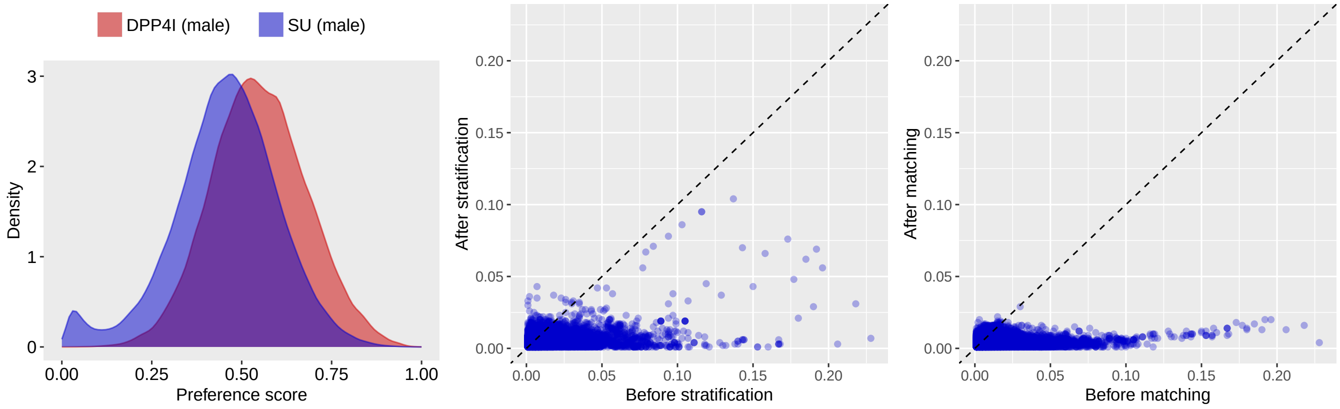

**Supplementary Table 272:** Most extreme patient characteristics balance diagnostics after stratification for DPP4I (male) vs SU (male) in the CCAE data source.

| Characteristic (total count = 10591) | Before stratification | | | After stratification | | |
|---|---|---|---|---|---|---|
| | T (%) | C (%) | SDf | T (%) | C (%) | SDf |
| measurement during day -365 through 0 days relative to index: Immunoassay for infectious agent antibody, qualitative or semiquantitative, single step method (eg, reagent strip) | 0.1 | 1.3 | -0.14 | 0.2 | 1.0 | -0.10 |
| measurement above normal range during day -365 through 0 days relative to index: Hemoglobin A1c/Hemoglobin.total in Blood by calculation | 0.1 | 1.0 | -0.12 | 0.1 | 0.8 | -0.10 |
| measurement during day -365 through 0 days relative to index: Hemoglobin A1c/Hemoglobin.total in Blood by calculation | 0.1 | 1.0 | -0.12 | 0.1 | 0.8 | -0.10 |
| measurement during day -30 through 0 days relative to index: Hemoglobin A1c/Hemoglobin.total in Blood by calculation | 0.1 | 0.7 | -0.10 | 0.1 | 0.6 | -0.09 |
| device_exposure during day -365 through 0 days relative to index: ONE TOUCH ULTRA CONTROL SOLN | 0.1 | 0.7 | -0.09 | 0.1 | 0.5 | -0.08 |
| measurement during day -365 through 0 days relative to index: Cholesterol, serum or whole blood, total | 1.1 | 3.7 | -0.17 | 1.4 | 2.5 | -0.08 |

| (Continued) | T (%) | C (%) | SDf | T (%) | C (%) | SDf |
|---|---|---|---|---|---|---|
| measurement within normal range during day -365 through 0 days relative to index: Hematocrit [Volume Fraction] of Blood | 0.1 | 0.6 | -0.08 | 0.1 | 0.5 | -0.07 |
| measurement during day -30 through 0 days relative to index: Cholesterol, serum or whole blood, total | 0.3 | 1.7 | -0.14 | 0.5 | 1.1 | -0.07 |
| measurement during day -365 through 0 days relative to index: Potassium; serum, plasma or whole blood | 2.0 | 5.7 | -0.19 | 2.9 | 4.2 | -0.07 |
| measurement during day -365 through 0 days relative to index: Hematocrit [Volume Fraction] of Blood | 0.2 | 0.7 | -0.08 | 0.2 | 0.6 | -0.07 |

**Supplementary Table 273:** Most extreme patient characteristics balance diagnostics after matching for DPP4I (male) vs SU (male) in the CCAE data source.

| | Before matching | | | After matching | | |
|---|---|---|---|---|---|---|
| Characteristic (total count = 10282) | T (%) | C (%) | SDf | T (%) | C (%) | SDf |
| observation during day -365 through 0 days relative to index: Diabetic management program, dietitian visit | 0.1 | 0.1 | 0.03 | 0.1 | 0.1 | 0.03 |
| device_exposure during day -365 through 0 days relative to index: TRUEPLUS 33G LANCETS | 0.1 | 0.1 | -0.01 | 0.1 | 0.1 | -0.02 |
| measurement during day -365 through 0 days relative to index: Creatinine; blood | 5.9 | 11.4 | -0.20 | 6.1 | 5.6 | 0.02 |
| measurement during day -365 through 0 days relative to index: Potassium; serum, plasma or whole blood | 2.0 | 5.7 | -0.19 | 2.1 | 1.8 | 0.02 |
| observation during day -365 through 0 days relative to index: Functional status assessed (COA) (RA) | 0.1 | 0.1 | -0.00 | 0.1 | 0.1 | -0.02 |
| measurement during day -30 through 0 days relative to index: Fluorescent noninfectious agent antibody; screen, each antibody | 0.1 | 0.1 | -0.02 | 0.1 | 0.1 | -0.02 |
| condition_era group during day -30 through 0 days relative to index: Enthesopathy of elbow region | 0.1 | 0.2 | -0.01 | 0.1 | 0.2 | -0.02 |
| measurement during day -365 through 0 days relative to index: Cholesterol, serum or whole blood, total | 1.1 | 3.7 | -0.17 | 1.1 | 0.9 | 0.02 |
| measurement during day -30 through 0 days relative to index: Antibody; herpes simplex, type 2 | 0.1 | 0.1 | 0.02 | 0.1 | 0.1 | 0.02 |
| measurement during day -30 through 0 days relative to index: Cystatin C | 0.1 | 0.1 | 0.02 | 0.1 | 0.1 | -0.02 |

##### 3.6.2 DAG

**Supplementary Figure 38:** Preference score distributions, propensity score stratified (middle column) and matched (right column) covariate balance and negative control error distributions (bottom row) for DPP4I (male) vs SU (male) in the DAG data source.

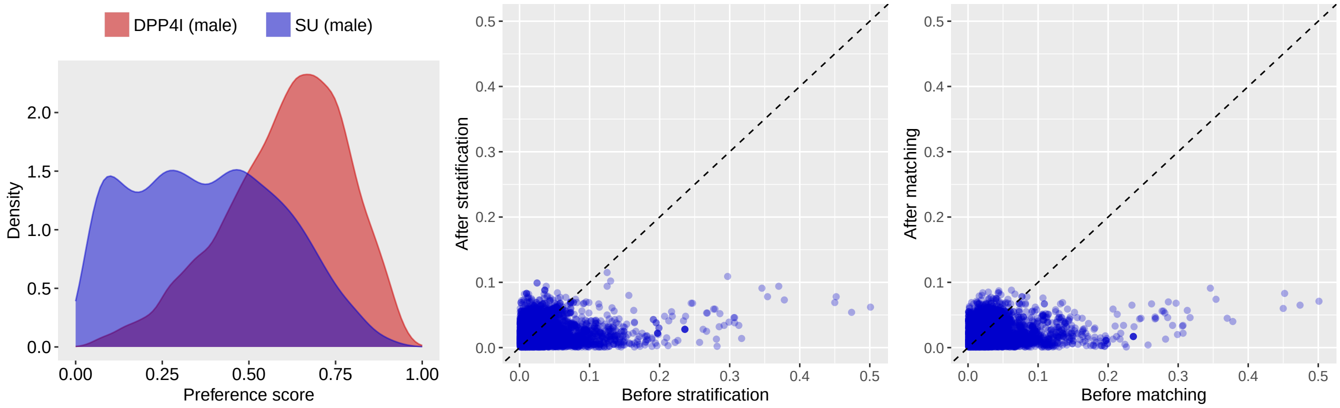

**Supplementary Table 274:** Most extreme patient characteristics balance diagnostics after stratification for DPP4I (male) vs SU (male) in the DAG data source.

| Characteristic (total count = 6274) | Before stratification | | | After stratification | | |
|---|---|---|---|---|---|---|
| | T (%) | C (%) | SDf | T (%) | C (%) | SDf |
| drug_era group during day 0 through 0 days relative to index: rosuvastatin | 1.1 | -0.1 | 0.12 | 1.0 | -0.1 | 0.12 |
| index year: 2020 | 11.2 | 3.5 | 0.30 | 10.4 | 7.3 | 0.11 |
| drug_era group during day -365 through 0 days relative to index: rosuvastatin | 1.3 | 0.2 | 0.13 | 1.2 | 0.3 | 0.10 |
| condition_era group during day -365 through 0 days relative to index: Cerebral atherosclerosis | 0.7 | 0.9 | -0.03 | 0.7 | 1.7 | -0.10 |
| condition_era group during day -365 through 0 days relative to index: Cerebral arteriosclerosis | 0.7 | 0.9 | -0.03 | 0.7 | 1.7 | -0.10 |
| index year: 2007 | 0.3 | 7.3 | -0.37 | 0.7 | 1.7 | -0.09 |
| drug_era group during day -30 through 0 days relative to index: rosuvastatin | 1.1 | 0.1 | 0.12 | 1.0 | 0.3 | 0.09 |
| age group: 90 - 94 | 0.6 | 0.3 | 0.04 | 0.5 | 1.5 | -0.09 |
| measurement during day -365 through 0 days relative to index: Hemoglobin [Mass/volume] in Body fluid | 34.3 | 19.2 | 0.35 | 32.4 | 36.8 | -0.09 |

| (Continued) | T (%) | C (%) | SDf | T (%) | C (%) | SDf |
|---|---|---|---|---|---|---|
| condition_era group during day -365 through 0 days relative to index: Presbycusis | 0.2 | 0.4 | -0.04 | 0.2 | 0.9 | -0.09 |

**Supplementary Table 275:** Most extreme patient characteristics balance diagnostics after matching for DPP4I (male) vs SU (male) in the DAG data source.

| | Before matching | | | After matching | | |
|---|---|---|---|---|---|---|
| Characteristic (total count = 5910) | T (%) | C (%) | SDf | T (%) | C (%) | SDf |
| measurement during day -365 through 0 days relative to index: Hemoglobin [Mass/volume] in Body fluid | 34.3 | 19.2 | 0.35 | 23.6 | 27.5 | -0.09 |
| condition_era group during day -30 through 0 days relative to index: Polyarthropathy | 0.1 | 0.5 | -0.06 | -0.2 | 0.6 | -0.09 |
| condition_era group during day -365 through 0 days relative to index: Potassium disorder | 0.9 | 1.4 | -0.04 | 0.5 | 1.3 | -0.09 |
| drug_era group during day -30 through 0 days relative to index: Other antiseptics and disinfectants | 0.5 | 0.3 | 0.03 | 0.8 | 0.2 | 0.09 |
| condition_era group during day -365 through 0 days relative to index: Candidiasis | 0.7 | 1.1 | -0.04 | 0.3 | 1.0 | -0.08 |
| condition_era group during day -365 through 0 days relative to index: Infection by Ascomycetes | 0.7 | 1.1 | -0.04 | 0.3 | 1.0 | -0.08 |
| condition_era group during day -365 through 0 days relative to index: Hepatic fibrosis | 0.5 | 0.6 | -0.02 | -0.2 | 0.6 | -0.08 |
| measurement during day -365 through 0 days relative to index: Leukocytes [#/volume] in Body fluid | 39.4 | 19.3 | 0.45 | 24.1 | 27.7 | -0.08 |
| condition_era group during day -365 through 0 days relative to index: Hypokalemia | 0.6 | 1.2 | -0.07 | 0.3 | 1.0 | -0.08 |
| drug_era group during day -365 through 0 days relative to index: bisacodyl | 0.2 | 0.5 | -0.05 | -0.2 | 0.6 | -0.08 |

##### 3.6.3 ODOD

**Supplementary Figure 39:** Preference score distributions, propensity score stratified (middle column) and matched (right column) covariate balance and negative control error distributions (bottom row) for DPP4I (male) vs SU (male) in the ODOD data source.

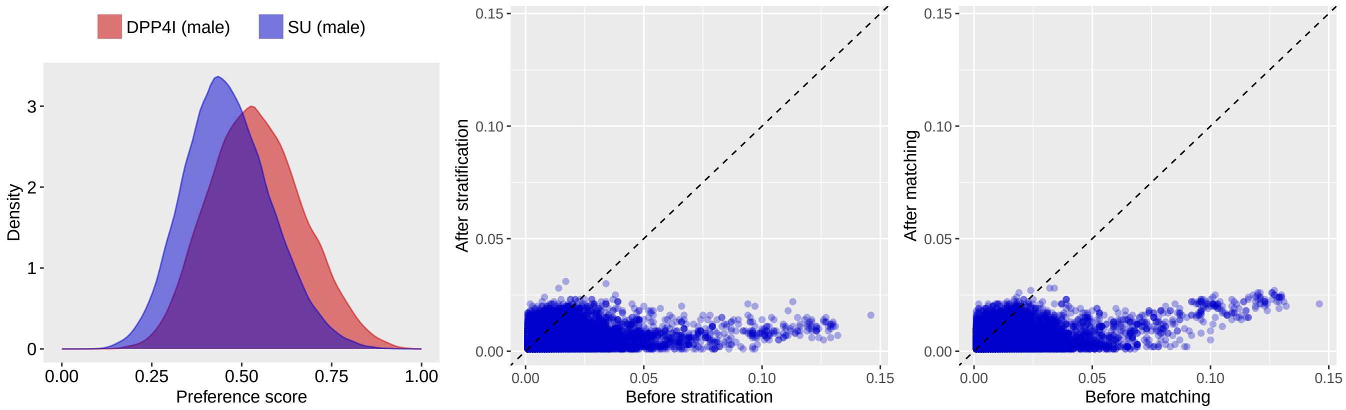

**Supplementary Table 276:** Most extreme patient characteristics balance diagnostics after stratification for DPP4I (male) vs SU (male) in the ODOD data source.

| | Before stratification | | | After stratification | | |
|---|---|---|---|---|---|---|
| Characteristic (total count = 15103) | T (%) | C (%) | SDf | T (%) | C (%) | SDf |
| procedure_occurrence during day -30 through 0 days relative to index: Hospital outpatient clinic visit specimen collection for severe acute respiratory syndrome coronavirus 2 (sars-cov-2) (coronavirus disease [covid-19]) | 0.1 | 0.1 | 0.02 | 0.2 | 0.1 | 0.03 |
| drug_era group during day -365 through 0 days relative to index: vilazodone | 0.2 | 0.1 | 0.03 | 0.2 | 0.1 | 0.03 |
| observation during day -365 through 0 days relative to index: Social work visit, in the home, per diem | 0.2 | 0.1 | 0.01 | 0.2 | 0.1 | 0.03 |
| condition_era group during day -365 through 0 days relative to index: Secondary malignant neoplasm of intra-abdominal organs | 0.1 | 0.3 | -0.04 | 0.2 | 0.3 | -0.03 |
| observation during day -365 through 0 days relative to index: Telehealth originating site facility fee | 0.2 | 0.2 | 0.01 | 0.3 | 0.2 | 0.02 |
| measurement during day -30 through 0 days relative to index: Ketone body(s) (eg, acetone, acetoacetic acid, beta-hydroxybutyrate); quantitative | 0.1 | 0.2 | -0.04 | 0.1 | 0.2 | -0.02 |

| (Continued) | T (%) | C (%) | SDf | T (%) | C (%) | SDf |
|---|---|---|---|---|---|---|
| measurement during day -365 through 0 days relative to index: Leukocyte esterase [Units/volume] in Urine | 0.1 | 0.1 | -0.03 | 0.1 | 0.1 | -0.02 |
| measurement within normal range during day -365 through 0 days relative to index: Leukocyte esterase [Units/volume] in Urine | 0.1 | 0.1 | -0.03 | 0.1 | 0.1 | -0.02 |
| drug_era group during day -365 through 0 days relative to index: suvorexant | 0.1 | 0.1 | 0.03 | 0.1 | 0.1 | 0.02 |
| measurement above normal range during day -365 through 0 days relative to index: Urobilinogen [Mass/volume] in Urine by Test strip | 0.2 | 0.1 | 0.02 | 0.2 | 0.1 | 0.02 |

**Supplementary Table 277:** Most extreme patient characteristics balance diagnostics after matching for DPP4I (male) vs SU (male) in the ODOD data source.

| | Before matching | | | After matching | | |
|---|---|---|---|---|---|---|
| Characteristic (total count = 14978) | T (%) | C (%) | SDf | T (%) | C (%) | SDf |
| drug_era group during day -365 through 0 days relative to index: vilazodone | 0.2 | 0.1 | 0.03 | 0.2 | 0.1 | 0.03 |
| drug_era group during day -365 through 0 days relative to index: insulin detemir | 0.2 | 0.1 | 0.03 | 0.2 | 0.1 | 0.03 |
| measurement within normal range during day -365 through 0 days relative to index: Albumin [Mass/volume] in Serum or Plasma | 40.6 | 34.4 | 0.13 | 39.5 | 40.9 | -0.03 |
| condition_era group during day -365 through 0 days relative to index: Organic anxiety disorder | 0.2 | 0.1 | 0.02 | 0.2 | 0.1 | 0.03 |
| measurement during day -365 through 0 days relative to index: Cholesterol [Mass/volume] in Serum or Plasma | 42.4 | 36.2 | 0.13 | 41.4 | 42.7 | -0.03 |
| measurement during day -365 through 0 days relative to index: Albumin [Mass/volume] in Serum or Plasma | 42.7 | 36.5 | 0.13 | 41.6 | 42.9 | -0.03 |
| measurement within normal range during day -365 through 0 days relative to index: Potassium [Moles/volume] in Serum or Plasma | 43.8 | 37.8 | 0.12 | 42.8 | 44.1 | -0.03 |
| measurement during day -365 through 0 days relative to index: Urea nitrogen/Creatinine [Mass Ratio] in Serum or Plasma | 42.9 | 37.1 | 0.12 | 42.0 | 43.3 | -0.03 |
| measurement during day -365 through 0 days relative to index: Dehydroepiandrosterone-sulfate (DHEA-S) | 0.4 | 0.3 | 0.02 | 0.3 | 0.5 | -0.03 |
| measurement during day -365 through 0 days relative to index: Triglyceride [Mass/volume] in Serum or Plasma | 42.2 | 35.9 | 0.13 | 41.2 | 42.4 | -0.03 |

##### 3.6.4 MDCR

**Supplementary Figure 40:** Preference score distributions, propensity score stratified (middle column) and matched (right column) covariate balance and negative control error distributions (bottom row) for DPP4I (male) vs SU (male) in the MDCR data source.

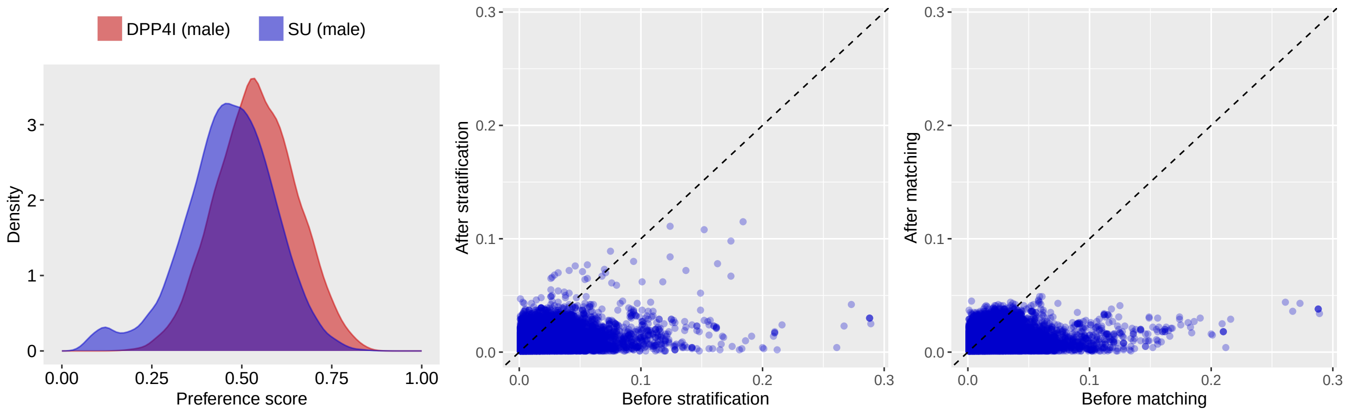

**Supplementary Table 278:** Most extreme patient characteristics balance diagnostics after stratification for DPP4I (male) vs SU (male) in the MDCR data source.

| | Before stratification | | | After stratification | | |
|---|---|---|---|---|---|---|
| Characteristic (total count = 12693) | T (%) | C (%) | SDf | T (%) | C (%) | SDf |
| measurement during day -365 through 0 days relative to index: Cholesterol, serum or whole blood, total | 0.5 | 2.8 | -0.18 | 0.8 | 2.2 | -0.12 |
| measurement during day -365 through 0 days relative to index: Immunoassay for infectious agent antibody, qualitative or semiquantitative, single step method (eg, reagent strip) | 0.1 | 0.9 | -0.12 | 0.1 | 0.8 | -0.11 |
| measurement during day -30 through 0 days relative to index: Potassium; serum, plasma or whole blood | 0.2 | 1.7 | -0.15 | 0.4 | 1.4 | -0.11 |
| measurement during day -365 through 0 days relative to index: Potassium; serum, plasma or whole blood | 1.4 | 4.3 | -0.17 | 1.9 | 3.5 | -0.10 |
| measurement above normal range during day -365 through 0 days relative to index: Cholesterol [Mass/volume] in Serum or Plasma | 1.3 | 2.3 | -0.07 | 1.3 | 2.5 | -0.09 |
| measurement during day -30 through 0 days relative to index: Cholesterol, serum or whole blood, total | 0.2 | 1.2 | -0.12 | 0.3 | 0.9 | -0.08 |
| measurement during day -365 through 0 days relative to index: Blood count; blood smear, microscopic examination without manual differential WBC count | 0.1 | 0.7 | -0.09 | 0.1 | 0.6 | -0.08 |

| (Continued) | T (%) | C (%) | SDf | T (%) | C (%) | SDf |
|---|---|---|---|---|---|---|
| measurement during day -30 through 0 days relative to index: Creatinine; blood | 1.0 | 3.3 | -0.16 | 1.6 | 2.8 | -0.08 |
| device_exposure during day -365 through 0 days relative to index: ONE TOUCH ULTRA 2 GLUCOSE SYST | 0.4 | 0.8 | -0.06 | 0.3 | 0.9 | -0.08 |
| measurement above normal range during day -365 through 0 days relative to index: Cholesterol in HDL [Mass/volume] in Serum or Plasma | 2.2 | 3.0 | -0.05 | 2.1 | 3.4 | -0.08 |

**Supplementary Table 279:** Most extreme patient characteristics balance diagnostics after matching for DPP4I (male) vs SU (male) in the MDCR data source.

| | Before matching | | | After matching | | |
|---|---|---|---|---|---|---|
| Characteristic (total count = 13334) | T (%) | C (%) | SDf | T (%) | C (%) | SDf |
| drug_era group during day -365 through 0 days relative to index: febuxostat | 0.6 | 0.2 | 0.06 | 0.5 | 0.2 | 0.05 |
| drug_era group during day 0 through 0 days relative to index: febuxostat | 0.5 | 0.1 | 0.06 | 0.4 | 0.2 | 0.05 |
| drug_era group during day -30 through 0 days relative to index: febuxostat | 0.5 | 0.2 | 0.06 | 0.5 | 0.2 | 0.05 |
| measurement during day -365 through 0 days relative to index: Vitamin D; 1, 25 dihydroxy, includes fraction(s), if performed | 0.5 | 0.2 | 0.06 | 0.4 | 0.2 | 0.04 |
| condition_era group during day -30 through 0 days relative to index: Neoplasm of skin of trunk | 0.5 | 0.5 | 0.00 | 0.4 | 0.7 | -0.04 |
| procedure_occurrence during day -365 through 0 days relative to index: Office or other outpatient visit for the evaluation and management of an established patient, which requires at least 2 of these 3 key components: A detailed history; A detailed examination; Medical decision making of moderate complexity. Counseling and/o | 87.8 | 78.1 | 0.26 | 87.2 | 88.6 | -0.04 |
| drug_era group during day 0 through 0 days relative to index: icosapent ethyl | 0.3 | 0.1 | 0.06 | 0.3 | 0.1 | 0.04 |
| condition_era group during day -365 through 0 days relative to index: Acute ethmoidal sinusitis | -0.1 | 0.1 | -0.04 | -0.1 | 0.1 | -0.04 |
| index year: 2006 | 0.4 | 4.6 | -0.27 | 0.4 | 0.7 | -0.04 |
| measurement during day -365 through 0 days relative to index: Urea nitrogen; quantitative | 3.4 | 4.9 | -0.07 | 3.5 | 2.8 | 0.04 |

##### 3.6.5 MDCD

**Supplementary Figure 41:** Preference score distributions, propensity score stratified (middle column) and matched (right column) covariate balance and negative control error distributions (bottom row) for DPP4I (male) vs SU (male) in the MDCD data source.

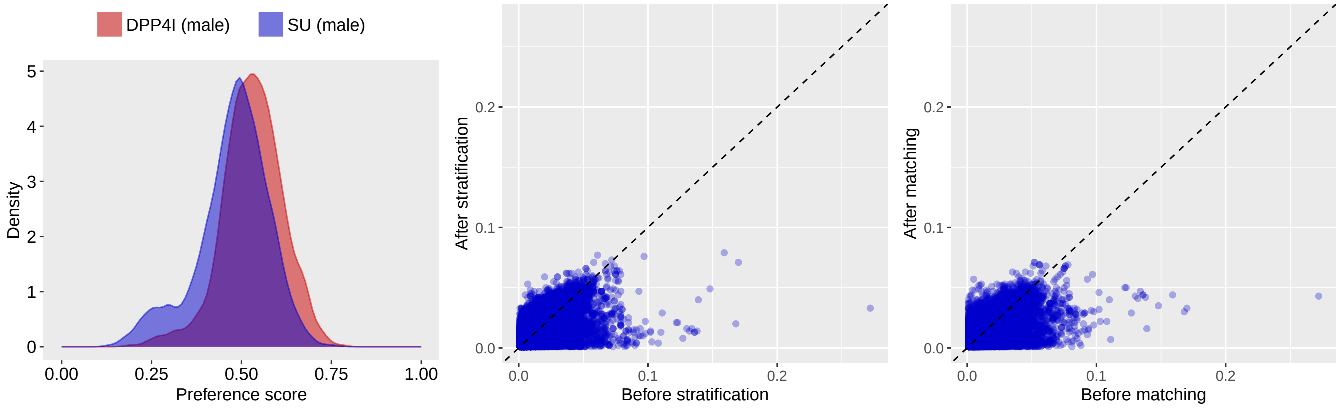

**Supplementary Table 280:** Most extreme patient characteristics balance diagnostics after stratification for DPP4I (male) vs SU (male) in the MDCD data source.

| | Before stratification | | | After stratification | | |
|---|---|---|---|---|---|---|
| Characteristic (total count = 22220) | T (%) | C (%) | SDf | T (%) | C (%) | SDf |
| condition_era group during day -365 through 0 days relative to index: Ill-defined disease | 0.4 | 2.1 | -0.16 | 0.8 | 1.7 | -0.08 |
| procedure_occurrence during day -365 through 0 days relative to index: Insertion of Infusion Device into Superior Vena Cava, Percutaneous Approach | 0.6 | 0.2 | 0.06 | 0.8 | 0.2 | 0.08 |
| device_exposure during day -365 through 0 days relative to index: UNILET COMFORTOUCH LANCET | -0.1 | 0.6 | -0.10 | -0.1 | 0.5 | -0.08 |
| condition_era group during day -365 through 0 days relative to index: Benign hypertensive heart disease without congestive heart failure | 0.3 | 0.8 | -0.07 | 0.2 | 0.7 | -0.07 |
| observation during day -365 through 0 days relative to index: Case management, per month | 0.6 | 2.9 | -0.17 | 1.4 | 2.3 | -0.07 |
| condition_era group during day -365 through 0 days relative to index: Absence of lower limb | 0.9 | 0.5 | 0.06 | 1.1 | 0.5 | 0.07 |
| condition_era group during day -365 through 0 days relative to index: Benign hypertensive heart disease | 0.3 | 0.8 | -0.07 | 0.3 | 0.8 | -0.07 |

| (Continued) | T (%) | C (%) | SDf | T (%) | C (%) | SDf |
|---|---|---|---|---|---|---|
| procedure_occurrence during day -365 through 0 days relative to index: Renewal of prescription | 0.4 | 1.0 | -0.07 | 0.3 | 0.9 | -0.07 |
| index year: 2017 | 16.6 | 14.0 | 0.07 | 16.7 | 14.3 | 0.07 |
| condition_era group during day -365 through 0 days relative to index: Chill | 0.6 | 0.3 | 0.05 | 0.7 | 0.3 | 0.07 |

**Supplementary Table 281:** Most extreme patient characteristics balance diagnostics after matching for DPP4I (male) vs SU (male) in the MDCD data source.

| | Before matching | | | After matching | | |
|---|---|---|---|---|---|---|
| Characteristic (total count = 14967) | T (%) | C (%) | SDf | T (%) | C (%) | SDf |
| condition_era group during day -365 through 0 days relative to index: Chill | 0.6 | 0.3 | 0.05 | 0.6 | 0.2 | 0.07 |
| condition_era group during day -365 through 0 days relative to index: Shivering or rigors | 0.6 | 0.3 | 0.05 | 0.6 | 0.2 | 0.07 |
| condition_era group during day -30 through 0 days relative to index: Dangerous and harmful thoughts | 0.1 | 0.6 | -0.08 | 0.1 | 0.6 | -0.07 |
| condition_era group during day -365 through 0 days relative to index: Internal hordeolum | -0.1 | 0.3 | -0.06 | -0.1 | 0.4 | -0.07 |
| condition_era group during day -365 through 0 days relative to index: Meibomianitis | -0.1 | 0.3 | -0.06 | -0.1 | 0.4 | -0.07 |
| condition_era group during day -365 through 0 days relative to index: Acute blepharitis | -0.1 | 0.3 | -0.06 | -0.1 | 0.4 | -0.07 |
| condition_era group during day -30 through 0 days relative to index: Disturbance in thinking | 0.2 | 0.7 | -0.08 | 0.2 | 0.6 | -0.07 |
| condition_era group during day -30 through 0 days relative to index: Disturbance in content of thought | 0.2 | 0.7 | -0.08 | 0.2 | 0.6 | -0.07 |
| procedure_occurrence during day -365 through 0 days relative to index: Insertion of Infusion Device into Superior Vena Cava, Percutaneous Approach | 0.6 | 0.2 | 0.06 | 0.7 | 0.2 | 0.07 |
| condition_era group during day -365 through 0 days relative to index: Melena | 0.7 | 1.2 | -0.05 | 0.7 | 1.4 | -0.07 |

##### 3.6.6 Open Claims

**Supplementary Figure 42:** Preference score distributions, propensity score stratified (middle column) and matched (right column) covariate balance and negative control error distributions (bottom row) for DPP4I (male) vs SU (male) in the Open Claims data source.

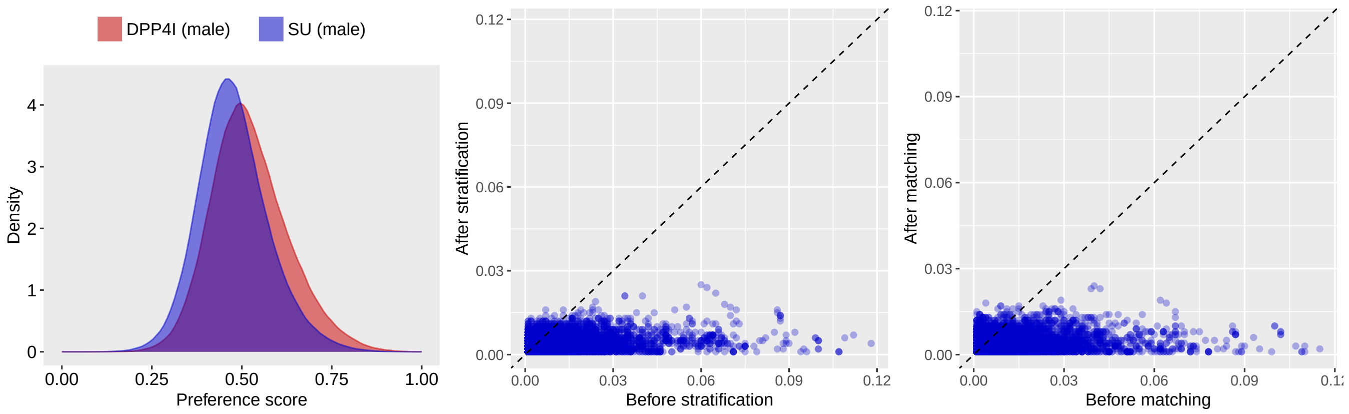

**Supplementary Table 282:** Most extreme patient characteristics balance diagnostics after stratification for DPP4I (male) vs SU (male) in the Open Claims data source.

| Characteristic (total count = 9749) | Before stratification | | | After stratification | | |
|---|---|---|---|---|---|---|
| | T (%) | C (%) | SDf | T (%) | C (%) | SDf |
| drug_era group during day 0 through 0 days relative to index: icosapent ethyl | 0.6 | 0.2 | 0.06 | 0.4 | 0.3 | 0.03 |
| drug_era group during day -30 through 0 days relative to index: icosapent ethyl | 0.6 | 0.2 | 0.06 | 0.4 | 0.3 | 0.02 |
| drug_era group during day -365 through 0 days relative to index: icosapent ethyl | 0.8 | 0.3 | 0.07 | 0.6 | 0.4 | 0.02 |
| drug_era group during day -365 through 0 days relative to index: pitavastatin | 0.3 | 0.1 | 0.04 | 0.2 | 0.1 | 0.02 |
| drug_era group during day 0 through 0 days relative to index: pitavastatin | 0.2 | 0.1 | 0.03 | 0.1 | 0.1 | 0.02 |
| drug_era group during day -30 through 0 days relative to index: pitavastatin | 0.2 | 0.1 | 0.03 | 0.2 | 0.1 | 0.02 |

| (Continued) | T (%) | C (%) | SDf | T (%) | C (%) | SDf |
|---|---|---|---|---|---|---|
| observation during day -365 through 0 days relative to index: Personal care services, per 15 minutes, not for an inpatient or resident of a hospital, nursing facility, icf/mr or imd, part of the individualized plan of treatment (code may not be used to identify services provided by home health aide or certified n... | 0.1 | 0.1 | 0.02 | 0.1 | 0.1 | 0.02 |
| drug_era group during day 0 through 0 days relative to index: nebivolol | 1.1 | 0.5 | 0.07 | 0.7 | 0.6 | 0.02 |
| drug_era group during day -30 through 0 days relative to index: nebivolol | 1.1 | 0.5 | 0.07 | 0.8 | 0.7 | 0.02 |
| measurement during day -365 through 0 days relative to index: C-reactive protein; high sensitivity (hsCRP) | 1.0 | 0.6 | 0.06 | 0.8 | 0.7 | 0.02 |

**Supplementary Table 283:** Most extreme patient characteristics balance diagnostics after matching for DPP4I (male) vs SU (male) in the Open Claims data source.

| | Before matching | | | After matching | | |
|---|---|---|---|---|---|---|
| Characteristic (total count = 9765) | T (%) | C (%) | SDf | T (%) | C (%) | SDf |
| drug_era group during day -30 through 0 days relative to index: pitavastatin | 0.2 | 0.1 | 0.04 | 0.2 | 0.1 | 0.02 |
| drug_era group during day -365 through 0 days relative to index: pitavastatin | 0.3 | 0.1 | 0.04 | 0.3 | 0.2 | 0.02 |
| drug_era group during day 0 through 0 days relative to index: pitavastatin | 0.2 | 0.1 | 0.04 | 0.2 | 0.1 | 0.02 |
| drug_era group during day 0 through 0 days relative to index: icosapent ethyl | 0.6 | 0.2 | 0.06 | 0.5 | 0.4 | 0.02 |
| observation during day -365 through 0 days relative to index: Personal care services, per 15 minutes, not for an inpatient or resident of a hospital, nursing facility, icf/mr or imd, part of the individualized plan of treatment (code may not be used to identify services provided by home health aide or certified n... | 0.2 | 0.1 | 0.03 | 0.1 | 0.1 | 0.02 |
| drug_era group during day -30 through 0 days relative to index: icosapent ethyl | 0.6 | 0.2 | 0.06 | 0.5 | 0.4 | 0.02 |
| drug_era group during day -365 through 0 days relative to index: nitroglycerin | 3.7 | 3.8 | -0.00 | 3.7 | 4.0 | -0.02 |
| condition_era group during day -30 through 0 days relative to index: Metabolic syndrome X | 0.2 | 0.1 | 0.02 | 0.2 | 0.1 | 0.02 |
| drug_era group during day 0 through 0 days relative to index: ACE INHIBITORS, COMBINATIONS | 71.3 | 72.0 | -0.01 | 71.2 | 72.0 | -0.02 |
| drug_era group during day -365 through 0 days relative to index: VASODILATORS USED IN CARDIAC DISEASES | 5.3 | 5.4 | -0.01 | 5.2 | 5.5 | -0.02 |

##### 3.6.7 OEHR

**Supplementary Figure 43:** Preference score distributions, propensity score stratified (middle column) and matched (right column) covariate balance and negative control error distributions (bottom row) for DPP4I (male) vs SU (male) in the OEHR data source.

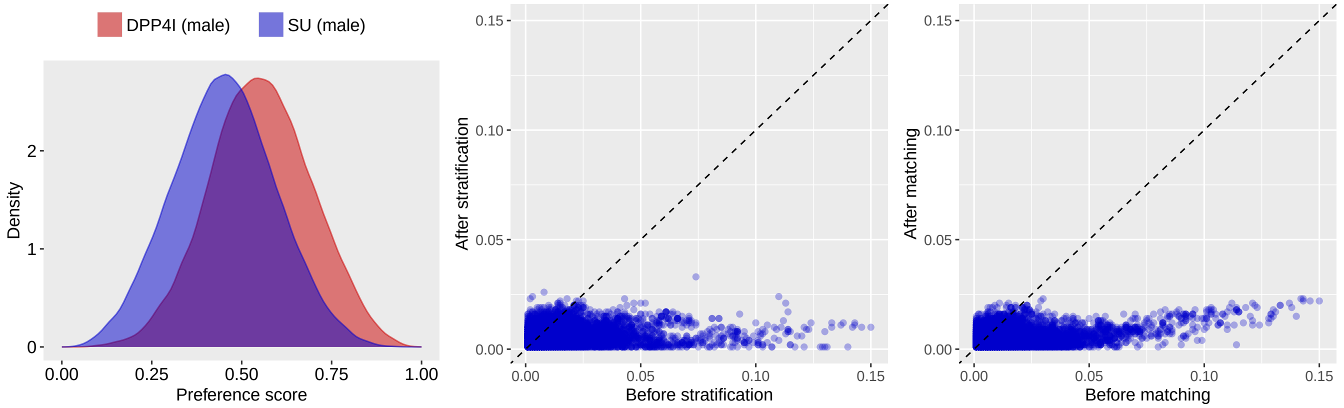

**Supplementary Table 284:** Most extreme patient characteristics balance diagnostics after stratification for DPP4I (male) vs SU (male) in the OEHR data source.

| | Before stratification | | | After stratification | | |
|---|---|---|---|---|---|---|
| Characteristic (total count = 11438) | T (%) | C (%) | SDf | T (%) | C (%) | SDf |
| measurement during day -365 through 0 days relative to index: Eye disorder screening | 0.1 | 0.6 | -0.07 | 0.3 | 0.5 | -0.03 |
| procedure_occurrence during day -365 through 0 days relative to index: Examination of research control | 0.1 | 0.1 | 0.01 | 0.2 | 0.1 | 0.03 |
| measurement during day -365 through 0 days relative to index: Basic metabolic panel (Calcium, total) This panel must include the following: Calcium, total (82310) Carbon dioxide (bicarbonate) (82374) Chloride (82435) Creatinine (82565) Glucose (82947) Potassium (84132) Sodium (84295) Urea nitrogen (BUN) (84520) | 20.3 | 24.9 | -0.11 | 24.5 | 23.5 | 0.02 |
| observation during day -365 through 0 days relative to index: Pain assessment documented as positive using a standardized tool and a follow-up plan is documented | 0.1 | 0.1 | -0.00 | 0.2 | 0.1 | 0.02 |
| measurement below normal range during day -365 through 0 days relative to index: Penicillin G potassium [Mass] of Dose | 6.1 | 7.2 | -0.04 | 7.5 | 6.9 | 0.02 |
| measurement during day -365 through 0 days relative to index: Anion gap in Blood | 51.7 | 52.6 | -0.02 | 53.6 | 52.5 | 0.02 |

| (Continued) | T (%) | C (%) | SDf | T (%) | C (%) | SDf |
|---|---|---|---|---|---|---|
| observation during day -30 through 0 days relative to index: Room & Board - Semi-private Two Bed (Medical or General) - General classification | 0.2 | 0.2 | 0.00 | 0.3 | 0.2 | 0.02 |
| condition_era group during day -30 through 0 days relative to index: Hypoglycemic disorder | 0.1 | 0.2 | -0.03 | 0.1 | 0.2 | -0.02 |
| measurement during day -365 through 0 days relative to index: Susceptibility studies, antimicrobial agent; disk method, per plate (12 or fewer agents) | 0.1 | 0.1 | -0.02 | 0.1 | 0.1 | -0.02 |
| measurement during day -365 through 0 days relative to index: Familial long QTS syndrome gene targeted mutation analysis in Blood or Tissue by Molecular genetics method | 0.1 | 0.1 | -0.02 | 0.1 | 0.1 | -0.02 |

**Supplementary Table 285:** Most extreme patient characteristics balance diagnostics after matching for DPP4I (male) vs SU (male) in the OEHR data source.

| | Before matching | | | After matching | | |
|---|---|---|---|---|---|---|
| Characteristic (total count = 11218) | T (%) | C (%) | SDf | T (%) | C (%) | SDf |
| measurement during day -365 through 0 days relative to index: Bilirubin.total [Presence] in Urine | 69.4 | 62.7 | 0.14 | 68.3 | 69.4 | -0.02 |
| measurement during day -365 through 0 days relative to index: Albumin [Presence] in Urine | 69.9 | 63.2 | 0.14 | 68.8 | 69.9 | -0.02 |
| measurement during day -365 through 0 days relative to index: Lipid panel | 0.1 | 0.1 | 0.03 | 0.1 | 0.1 | 0.02 |
| measurement during day -30 through 0 days relative to index: Body height | 71.5 | 64.6 | 0.15 | 70.8 | 71.8 | -0.02 |
| measurement during day -365 through 0 days relative to index: Albumin/Protein.total in Serum or Plasma | 69.5 | 62.6 | 0.15 | 68.4 | 69.4 | -0.02 |
| measurement during day -365 through 0 days relative to index: Alkaline phosphatase.renal/Alkaline phosphatase.total in Serum or Plasma | 69.5 | 62.8 | 0.14 | 68.4 | 69.5 | -0.02 |
| observation during day -30 through 0 days relative to index: Professional Fees - Clinic | 0.1 | 0.1 | -0.03 | 0.1 | 0.1 | -0.02 |
| measurement within normal range during day -365 through 0 days relative to index: Albumin/Protein.total in Serum or Plasma | 61.5 | 54.7 | 0.14 | 60.3 | 61.4 | -0.02 |
| measurement during day -365 through 0 days relative to index: Neutrophil cytoplasmic Ab.perinuclear [Presence] in Serum | 50.0 | 44.0 | 0.12 | 48.6 | 49.7 | -0.02 |
| measurement during day -365 through 0 days relative to index: Aspartate aminotransferase [Presence] in Body fluid | 71.7 | 66.5 | 0.11 | 70.6 | 71.6 | -0.02 |

### 3.6.8 VA

**Supplementary Figure 44:** Preference score distributions, propensity score stratified (middle column) and matched (right column) covariate balance and negative control error distributions (bottom row) for DPP4I (male) vs SU (male) in the VA data source.

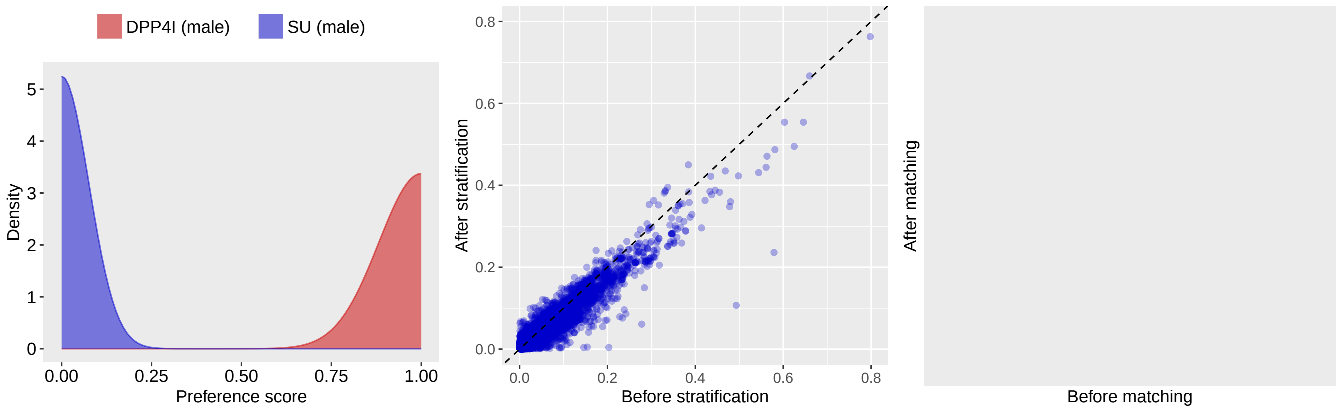

**Supplementary Table 286:** Most extreme patient characteristics balance diagnostics after stratification for DPP4I (male) vs SU (male) in the VA data source.

| Characteristic (total count = 13125) | Before stratification | | | After stratification | | |
|---|---|---|---|---|---|---|
| | T (%) | C (%) | SDf | T (%) | C (%) | SDf |
| procedure_occurrence during day -365 through 0 days relative to index: Counseling | 6.5 | 37.1 | -0.80 | 6.5 | 35.5 | -0.76 |
| observation during day -365 through 0 days relative to index: Requires influenza virus vaccination | 3.9 | 26.3 | -0.66 | 3.9 | 26.7 | -0.67 |
| drug_era group during day -365 through 0 days relative to index: Simvastatin | 16.5 | 44.8 | -0.65 | 16.5 | 40.6 | -0.55 |
| index year: 2020 | 23.8 | 3.9 | 0.60 | 23.8 | 5.0 | 0.55 |
| drug_era group during day -365 through 0 days relative to index: atorvastatin | 47.7 | 19.5 | 0.62 | 47.7 | 24.6 | 0.49 |
| drug_era group during day -30 through 0 days relative to index: Simvastatin | 13.2 | 37.5 | -0.58 | 13.2 | 33.2 | -0.49 |
| drug_era group during day 0 through 0 days relative to index: Simvastatin | 12.4 | 35.5 | -0.56 | 12.4 | 31.3 | -0.47 |
| measurement during day -365 through 0 days relative to index: Eye disorder screening | 2.4 | 12.1 | -0.38 | 2.4 | 14.6 | -0.45 |

| (Continued) | T (%) | C (%) | SDf | T (%) | C (%) | SDf |
|---|---|---|---|---|---|---|
| drug_era group during day -30 through 0 days relative to index: atorvastatin | 41.5 | 16.9 | 0.56 | 41.5 | 21.3 | 0.44 |
| index year: 2021 | 15.0 | 2.2 | 0.47 | 15.0 | 2.8 | 0.43 |

**Supplementary Table 287:** Most extreme patient characteristics balance diagnostics after matching for DPP4I (male) vs SU (male) in the VA data source.

| | Before matching | | | After matching | | |
|---|---|---|---|---|---|---|
| Characteristic (total count = 0) | T (%) | C (%) | SDf | T (%) | C (%) | SDf |

##### 3.6.9 SIDIAP

**Supplementary Figure 45:** Preference score distributions, propensity score stratified (middle column) and matched (right column) covariate balance and negative control error distributions (bottom row) for DPP4I (male) vs SU (male) in the SIDIAP data source.

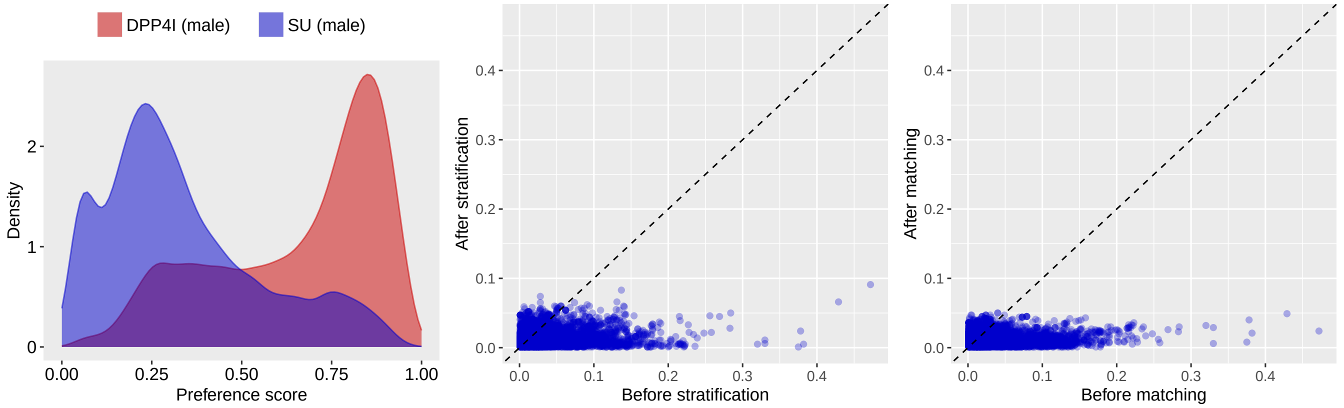

**Supplementary Table 288:** Most extreme patient characteristics balance diagnostics after stratification for DPP4I (male) vs SU (male) in the SIDIAP data source.

| | Before stratification | | | After stratification | | |
|---|---|---|---|---|---|---|
| Characteristic (total count = 6195) | T (%) | C (%) | SDf | T (%) | C (%) | SDf |
| index year: 2008 | 0.9 | 12.3 | -0.47 | 2.7 | 4.3 | -0.09 |
| procedure_occurrence during day -365 through 0 days relative to index: Plain Radiography of Chest | 1.0 | -0.1 | 0.14 | 0.8 | 0.2 | 0.08 |
| observation during day -365 through 0 days relative to index: Administrative reason for encounter | 0.7 | 0.5 | 0.03 | 0.6 | 1.4 | -0.07 |
| index year: 2019 | 13.1 | 2.0 | 0.43 | 10.5 | 8.6 | 0.07 |
| drug_era group during day 0 through 0 days relative to index: High-ceiling diuretics and potassium-sparing agents | 8.8 | 5.5 | 0.13 | 7.9 | 6.2 | 0.07 |
| drug_era group during day -365 through 0 days relative to index: Aldosterone antagonists | 3.1 | 1.9 | 0.07 | 2.8 | 1.8 | 0.07 |
| drug_era group during day -365 through 0 days relative to index: leuprolide | 0.5 | 0.3 | 0.03 | 0.4 | 1.0 | -0.07 |
| drug_era group during day -30 through 0 days relative to index: pitavastatin | 0.6 | 0.1 | 0.09 | 0.5 | 0.1 | 0.06 |
| drug_era group during day 0 through 0 days relative to index: furosemide | 7.2 | 4.0 | 0.14 | 6.4 | 5.0 | 0.06 |

| (Continued) | T (%) | C (%) | SDf | T (%) | C (%) | SDf |
|---|---|---|---|---|---|---|
| drug_era group during day -365 through 0 days relative to index: pitavastatin | 0.7 | 0.1 | 0.09 | 0.6 | 0.2 | 0.06 |

**Supplementary Table 289:** Most extreme patient characteristics balance diagnostics after matching for DPP4I (male) vs SU (male) in the SIDIAP data source.

| | Before matching | | | After matching | | |
|---|---|---|---|---|---|---|
| Characteristic (total count = 6282) | T (%) | C (%) | SDf | T (%) | C (%) | SDf |
| condition_era group during day -30 through 0 days relative to index: Closed fracture of femur | 0.1 | 0.3 | -0.04 | 0.1 | 0.3 | -0.05 |
| condition_era group during day -30 through 0 days relative to index: Fracture of femur | 0.1 | 0.3 | -0.04 | 0.1 | 0.3 | -0.05 |
| index year: 2019 | 13.1 | 2.0 | 0.43 | 3.7 | 2.8 | 0.05 |
| drug_era group during day 0 through 0 days relative to index: prednisone | 1.9 | 1.3 | 0.05 | 0.9 | 1.4 | -0.05 |
| drug_era group during day -365 through 0 days relative to index: perindopril | 0.5 | 0.3 | 0.03 | 0.6 | 0.3 | 0.05 |
| drug_era group during day -365 through 0 days relative to index: aliskiren | 0.3 | 0.2 | 0.02 | 0.5 | 0.2 | 0.05 |
| drug_era group during day -365 through 0 days relative to index: OTHER AGENTS ACTING ON THE RENIN-ANGIOTENSIN SYSTEM | 0.3 | 0.2 | 0.02 | 0.5 | 0.2 | 0.05 |
| drug_era group during day -365 through 0 days relative to index: Renin-inhibitors | 0.3 | 0.2 | 0.02 | 0.5 | 0.2 | 0.05 |
| drug_era group during day 0 through 0 days relative to index: trandolapril | 0.1 | 0.1 | 0.01 | 0.3 | 0.1 | 0.05 |
| condition_era group during day -365 through 0 days relative to index: Candidiasis of mouth | 0.3 | 0.3 | -0.00 | 0.2 | 0.4 | -0.05 |

### 4 Unadjusted incidence rates across data sources and outcomes

#### 4.1 GLP1RA (male) vs SGLT2I (male)

##### 4.1.1 3-point MACE

**Supplementary Table 290:** Exposures, outcomes and risk estimates for 3-point MACE in GLP1RA (male) (T) vs SGLT2I (male) (C) new-user cohorts under four different propensity score (PS) adjustment / time-at-risk (TAR) designs across data sources. We report total patient exposure time in 1,000 patient-year (KYr) units, uncalibrated and calibrated hazard ratios (HRs) and their 95% confidence intervals (CIs). Greyed out entries did not pass study diagnostics and are included only for completeness.

| | Patients | | Exposure (KYr) | | Outcomes | | Incidence (per KYr) | |
|---|---|---|---|---|---|---|---|---|
| | T | C | T | C | T | C | T | C |
| **PS-matching / on-treatment TAR** | | | | | | | | |
| CCAE | 9,303 | 23,130 | 6.3 | 19.3 | 50 | 140 | 7.93 (5.89 - 10.5) | 7.27 (6.11 - 8.58) |
| ODOD | 6,825 | 16,405 | 4.0 | 11.4 | 34 | 170 | 8.58 (5.94 - 12.0) | 15.0 (12.8 - 17.4) |
| MDCR | 653 | 1,972 | 0.4 | 1.6 | 9 | 35 | 21.7 (9.93 - 41.2) | 22.3 (15.5 - 31.0) |
| Open Claims | 137,357 | 308,574 | 96.7 | 270.1 | 615 | 2,092 | 6.36 (5.87 - 6.88) | 7.75 (7.42 - 8.08) |
| OEHR | 7,754 | 19,529 | 2.2 | 6.2 | 22 | 48 | 10.2 (6.36 - 15.4) | 7.70 (5.68 - 10.2) |
| VA | 1,119 | 10,349 | 0.8 | 6.3 | <10 | 38 | < 12.8 (6.14 - 23.5) | 6.07 (4.30 - 8.33) |
| Meta-analysis | 162,358 | 377,987 | 109.9 | 313.2 | <731 | 2,488 | < 6.65 (6.18 - 7.15) | 7.94 (7.63 - 8.26) |
| Meta-analysis (w/o USOC) | 25,001 | 69,413 | 13.2 | 43.1 | <116 | 396 | < 8.78 (7.26 - 10.5) | 9.18 (8.30 - 10.1) |
| **PS-stratification / on-treatment TAR** | | | | | | | | |
| CCAE | 9,931 | 23,133 | 6.6 | 19.3 | 52 | 140 | 7.87 (5.88 - 10.3) | 7.27 (6.11 - 8.57) |
| ODOD | 7,409 | 16,422 | 4.2 | 11.4 | 37 | 170 | 8.73 (6.15 - 12.0) | 14.9 (12.8 - 17.4) |
| MDCR | 685 | 1,977 | 0.4 | 1.6 | 9 | 36 | 20.8 (9.52 - 39.5) | 22.8 (15.9 - 31.5) |
| Open Claims | 147,406 | 308,612 | 101.6 | 270.1 | 644 | 2,093 | 6.34 (5.86 - 6.84) | 7.75 (7.42 - 8.09) |
| OEHR | 8,460 | 19,539 | 2.3 | 6.2 | 25 | 48 | 10.7 (6.91 - 15.8) | 7.69 (5.67 - 10.2) |
| VA | 1,281 | 11,375 | 0.9 | 6.9 | <10 | 50 | < 11.2 (5.36 - 20.6) | 7.26 (5.39 - 9.57) |
| Meta-analysis | 173,206 | 367,706 | 114.8 | 307.0 | 758 | 2,451 | 6.60 (6.14 - 7.09) | 7.98 (7.67 - 8.31) |
| Meta-analysis (w/o USOC) | 25,800 | 59,094 | 13.2 | 36.9 | 114 | 358 | 8.64 (7.13 - 10.4) | 9.71 (8.73 - 10.8) |
| **PS-matching / on-treatment TAR without escalation** | | | | | | | | |
| CCAE | 9,303 | 23,130 | 5.5 | 16.8 | 43 | 116 | 7.79 (5.64 - 10.5) | 6.89 (5.70 - 8.27) |
| ODOD | 6,825 | 16,405 | 3.5 | 10.1 | 28 | 150 | 7.92 (5.26 - 11.5) | 14.9 (12.6 - 17.5) |
| MDCR | 653 | 1,972 | 0.4 | 1.4 | 6 | 32 | 16.1 (5.90 - 35.0) | 22.8 (15.6 - 32.2) |
| Open Claims | 137,357 | 308,574 | 84.4 | 229.5 | 510 | 1,785 | 6.04 (5.53 - 6.59) | 7.78 (7.42 - 8.15) |
| OEHR | 7,754 | 19,529 | 1.9 | 5.5 | 18 | 38 | 9.52 (5.64 - 15.0) | 6.92 (4.89 - 9.49) |
| VA | 1,119 | 10,349 | 0.6 | 5.4 | <10 | 32 | < 16.5 (7.91 - 30.3) | 5.93 (4.06 - 8.38) |
| Meta-analysis | 161,239 | 367,638 | 95.3 | 261.9 | 599 | 2,089 | 6.28 (5.79 - 6.81) | 7.98 (7.64 - 8.33) |
| Meta-analysis (w/o USOC) | 23,882 | 59,064 | 10.9 | 32.4 | 89 | 304 | 8.13 (6.53 - 10.0) | 9.38 (8.36 - 10.5) |
| **PS-stratification / on-treatment TAR without escalation** | | | | | | | | |
| CCAE | 9,931 | 23,133 | 5.8 | 16.8 | 45 | 116 | 7.75 (5.65 - 10.4) | 6.89 (5.70 - 8.27) |
| ODOD | 7,409 | 16,422 | 3.8 | 10.1 | 31 | 150 | 8.21 (5.58 - 11.6) | 14.9 (12.6 - 17.4) |
| MDCR | 685 | 1,977 | 0.4 | 1.4 | 6 | 32 | 15.4 (5.64 - 33.4) | 22.7 (15.5 - 32.0) |
| Open Claims | 147,406 | 308,612 | 88.9 | 229.5 | 537 | 1,786 | 6.04 (5.54 - 6.57) | 7.78 (7.43 - 8.15) |
| OEHR | 8,460 | 19,539 | 2.0 | 5.5 | 21 | 38 | 10.3 (6.35 - 15.7) | 6.91 (4.89 - 9.49) |
| VA | 1,281 | 11,375 | 0.7 | 6.0 | <10 | 42 | < 14.4 (6.88 - 26.4) | 7.06 (5.09 - 9.54) |
| Meta-analysis | 173,206 | 367,706 | 100.5 | 261.9 | 634 | 2,090 | 6.31 (5.82 - 6.82) | 7.98 (7.64 - 8.33) |
| Meta-analysis (w/o USOC) | 25,800 | 59,094 | 11.6 | 32.4 | 97 | 304 | 8.34 (6.76 - 10.2) | 9.38 (8.35 - 10.5) |

##### 4.1.2 4-point MACE

**Supplementary Table 291:** Exposures, outcomes and risk estimates for 4-point MACE in GLP1RA (male) (T) vs SGLT2I (male) (C) new-user cohorts under four different propensity score (PS) adjustment / time-at-risk (TAR) designs across data sources. We report total patient exposure time in 1,000 patient-year (KYr) units, uncalibrated and calibrated hazard ratios (HRs) and their 95% confidence intervals (CIs). Greyed out entries did not pass study diagnostics and are included only for completeness.

| | Patients | | Exposure (KYr) | | Outcomes | | Incidence (per KYr) | |
|---|---|---|---|---|---|---|---|---|
| | T | C | T | C | T | C | T | C |
| **PS-matching / on-treatment TAR** | | | | | | | | |
| CCAE | 9,140 | 22,776 | 6.2 | 19.0 | 81 | 211 | 13.1 (10.4 - 16.2) | 11.1 (9.67 - 12.7) |
| ODOD | 6,578 | 15,701 | 3.8 | 10.9 | 66 | 257 | 17.3 (13.4 - 22.0) | 23.5 (20.7 - 26.5) |
| MDCR | 619 | 1,857 | 0.4 | 1.5 | 15 | 51 | 38.1 (21.3 - 62.8) | 34.3 (25.6 - 45.1) |
| Open Claims | 133,612 | 297,193 | 94.2 | 261.9 | 927 | 2,979 | 9.84 (9.22 - 10.5) | 11.4 (11.0 - 11.8) |
| OEHR | 7,624 | 19,116 | 2.1 | 6.1 | 27 | 92 | 12.7 (8.37 - 18.5) | 15.1 (12.2 - 18.5) |
| VA | 1,072 | 9,618 | 0.7 | 5.8 | <10 | 67 | < 13.4 (6.42 - 24.6) | 11.5 (8.88 - 14.6) |
| Meta-analysis | 158,026 | 364,404 | 107.1 | 303.8 | <1,111 | 3,606 | < 10.4 (9.78 - 11.0) | 11.9 (11.5 - 12.3) |
| Meta-analysis (w/o USOC) | 24,414 | 67,211 | 12.9 | 41.9 | <184 | 627 | < 14.3 (12.3 - 16.5) | 15.0 (13.8 - 16.2) |
| **PS-stratification / on-treatment TAR** | | | | | | | | |
| CCAE | 9,749 | 22,780 | 6.5 | 19.0 | 85 | 211 | 13.1 (10.5 - 16.2) | 11.1 (9.67 - 12.7) |
| ODOD | 7,135 | 15,713 | 4.1 | 10.9 | 70 | 257 | 17.1 (13.4 - 21.7) | 23.5 (20.7 - 26.5) |
| MDCR | 650 | 1,862 | 0.4 | 1.5 | 15 | 52 | 36.6 (20.5 - 60.4) | 34.7 (25.9 - 45.6) |
| Open Claims | 143,483 | 297,226 | 99.1 | 262.0 | 973 | 2,981 | 9.82 (9.22 - 10.5) | 11.4 (11.0 - 11.8) |
| OEHR | 8,332 | 19,146 | 2.3 | 6.1 | 31 | 93 | 13.5 (9.16 - 19.1) | 15.2 (12.3 - 18.6) |
| VA | 1,229 | 10,518 | 0.9 | 6.4 | <10 | 86 | < 11.6 (5.56 - 21.3) | 13.4 (10.7 - 16.6) |
| Meta-analysis | 168,699 | 354,865 | 111.9 | 298.0 | 1,159 | 3,542 | 10.4 (9.77 - 11.0) | 11.9 (11.5 - 12.3) |
| Meta-analysis (w/o USOC) | 25,216 | 57,639 | 12.9 | 36.0 | 186 | 561 | 14.4 (12.4 - 16.7) | 15.6 (14.3 - 16.9) |
| **PS-matching / on-treatment TAR without escalation** | | | | | | | | |
| CCAE | 9,140 | 22,776 | 5.4 | 16.6 | 70 | 176 | 12.9 (10.0 - 16.3) | 10.6 (9.11 - 12.3) |
| ODOD | 6,578 | 15,701 | 3.4 | 9.7 | 58 | 226 | 17.0 (12.9 - 22.0) | 23.3 (20.3 - 26.5) |
| MDCR | 619 | 1,857 | 0.4 | 1.3 | 12 | 46 | 33.9 (17.5 - 59.2) | 34.7 (25.4 - 46.3) |
| Open Claims | 133,612 | 297,193 | 82.2 | 222.6 | 784 | 2,506 | 9.54 (8.88 - 10.2) | 11.3 (10.8 - 11.7) |
| OEHR | 7,624 | 19,116 | 1.9 | 5.4 | 22 | 74 | 11.9 (7.44 - 18.0) | 13.8 (10.8 - 17.3) |
| VA | 1,072 | 9,618 | 0.6 | 5.0 | <10 | 55 | < 17.2 (8.24 - 31.6) | 10.9 (8.24 - 14.2) |
| Meta-analysis | 158,026 | 364,404 | 93.5 | 259.3 | <944 | 3,037 | < 10.1 (9.47 - 10.8) | 11.7 (11.3 - 12.1) |
| Meta-analysis (w/o USOC) | 24,414 | 67,211 | 11.3 | 36.7 | <160 | 531 | < 14.2 (12.1 - 16.6) | 14.5 (13.3 - 15.8) |
| **PS-stratification / on-treatment TAR without escalation** | | | | | | | | |
| CCAE | 9,749 | 22,780 | 5.7 | 16.6 | 74 | 176 | 13.0 (10.2 - 16.3) | 10.6 (9.11 - 12.3) |
| ODOD | 7,135 | 15,713 | 3.6 | 9.7 | 62 | 226 | 17.0 (13.1 - 21.9) | 23.2 (20.3 - 26.5) |
| MDCR | 650 | 1,862 | 0.4 | 1.3 | 12 | 46 | 32.5 (16.8 - 56.7) | 34.5 (25.3 - 46.1) |
| Open Claims | 143,483 | 297,226 | 86.7 | 222.7 | 828 | 2,508 | 9.55 (8.91 - 10.2) | 11.3 (10.8 - 11.7) |
| OEHR | 8,332 | 19,146 | 2.0 | 5.4 | 26 | 75 | 12.9 (8.45 - 18.9) | 13.9 (10.9 - 17.4) |
| VA | 1,229 | 10,518 | 0.7 | 5.5 | <10 | 71 | < 14.9 (7.16 - 27.5) | 12.8 (10.0 - 16.2) |
| Meta-analysis | 168,699 | 354,865 | 98.0 | 254.4 | 990 | 2,985 | 10.1 (9.48 - 10.7) | 11.7 (11.3 - 12.2) |
| Meta-analysis (w/o USOC) | 25,216 | 57,639 | 11.4 | 31.7 | 162 | 477 | 14.3 (12.2 - 16.6) | 15.1 (13.7 - 16.5) |

##### 4.1.3 Acute myocardial infarction

**Supplementary Table 292:** Exposures, outcomes and risk estimates for Acute myocardial infarction in GLP1RA (male) (T) vs SGLT2I (male) (C) new-user cohorts under four different propensity score (PS) adjustment / time-at-risk (TAR) designs across data sources. We report total patient exposure time in 1,000 patient-year (KYr) units, uncalibrated and calibrated hazard ratios (HRs) and their 95% confidence intervals (CIs). Greyed out entries did not pass study diagnostics and are included only for completeness.

| | Patients | | Exposure (KYr) | | Outcomes | | Incidence (per KYr) | |
|---|---|---|---|---|---|---|---|---|
| | T | C | T | C | T | C | T | C |
| **PS-matching / on-treatment TAR** | | | | | | | | |
| CCAE | 9,438 | 23,422 | 6.4 | 19.5 | 32 | 82 | 4.98 (3.41 - 7.04) | 4.20 (3.34 - 5.22) |
| ODOD | 6,953 | 16,763 | 4.0 | 11.7 | 16 | 101 | 3.97 (2.27 - 6.44) | 8.67 (7.06 - 10.5) |
| MDCR | 679 | 2,027 | 0.4 | 1.6 | 5 | 15 | 11.5 (3.73 - 26.8) | 9.30 (5.21 - 15.3) |
| Open Claims | 140,293 | 316,161 | 98.7 | 276.1 | 328 | 1,186 | 3.32 (2.97 - 3.70) | 4.30 (4.05 - 4.55) |
| OEHR | 7,822 | 19,712 | 2.2 | 6.3 | 9 | 26 | 4.11 (1.88 - 7.80) | 4.13 (2.70 - 6.05) |
| VA | 1,143 | 10,556 | 0.8 | 6.4 | <10 | 27 | < 12.6 (6.05 - 23.2) | 4.24 (2.80 - 6.17) |
| Meta-analysis | 164,506 | 376,058 | 111.3 | 313.5 | 385 | 1,395 | 3.46 (3.12 - 3.82) | 4.45 (4.22 - 4.69) |
| Meta-analysis (w/o USOC) | 24,213 | 59,897 | 12.6 | 37.5 | 57 | 209 | 4.51 (3.41 - 5.84) | 5.58 (4.85 - 6.39) |
| **PS-stratification / on-treatment TAR** | | | | | | | | |
| CCAE | 10,071 | 23,425 | 6.7 | 19.5 | 33 | 82 | 4.91 (3.38 - 6.89) | 4.20 (3.34 - 5.22) |
| ODOD | 7,546 | 16,779 | 4.3 | 11.7 | 17 | 101 | 3.94 (2.30 - 6.31) | 8.66 (7.05 - 10.5) |
| MDCR | 709 | 2,032 | 0.5 | 1.6 | 5 | 15 | 11.1 (3.60 - 25.9) | 9.23 (5.16 - 15.2) |
| Open Claims | 150,437 | 316,207 | 103.6 | 276.1 | 344 | 1,187 | 3.32 (2.98 - 3.69) | 4.30 (4.06 - 4.55) |
| OEHR | 8,530 | 19,723 | 2.4 | 6.3 | 10 | 26 | 4.23 (2.03 - 7.77) | 4.13 (2.70 - 6.05) |
| VA | 1,307 | 11,651 | 0.9 | 7.0 | <10 | 35 | < 11.0 (5.30 - 20.3) | 4.98 (3.47 - 6.92) |
| Meta-analysis | 176,584 | 376,134 | 117.0 | 313.6 | 404 | 1,396 | 3.45 (3.12 - 3.81) | 4.45 (4.22 - 4.69) |
| Meta-analysis (w/o USOC) | 26,147 | 59,927 | 13.4 | 37.5 | 60 | 209 | 4.48 (3.42 - 5.76) | 5.58 (4.85 - 6.39) |
| **PS-matching / on-treatment TAR without escalation** | | | | | | | | |
| CCAE | 9,438 | 23,422 | 5.6 | 17.0 | 29 | 70 | 5.16 (3.46 - 7.41) | 4.11 (3.21 - 5.19) |
| ODOD | 6,953 | 16,763 | 3.6 | 10.3 | 13 | 90 | 3.62 (1.93 - 6.18) | 8.72 (7.01 - 10.7) |
| MDCR | 679 | 2,027 | 0.4 | 1.4 | <5 | 14 | < 12.8 (4.14 - 29.8) | 9.69 (5.30 - 16.3) |
| Open Claims | 140,293 | 316,161 | 86.1 | 234.6 | 272 | 999 | 3.16 (2.79 - 3.56) | 4.26 (4.00 - 4.53) |
| OEHR | 7,822 | 19,712 | 1.9 | 5.5 | 8 | 25 | 4.19 (1.81 - 8.26) | 4.51 (2.92 - 6.66) |
| VA | 1,143 | 10,556 | 0.6 | 5.5 | <10 | 23 | < 16.2 (7.77 - 29.8) | 4.19 (2.66 - 6.29) |
| Meta-analysis | 164,506 | 376,058 | 97.2 | 267.5 | 322 | 1,184 | 3.31 (2.96 - 3.69) | 4.43 (4.18 - 4.69) |
| Meta-analysis (w/o USOC) | 24,213 | 59,897 | 11.1 | 32.9 | 50 | 185 | 4.50 (3.34 - 5.93) | 5.62 (4.84 - 6.50) |
| **PS-stratification / on-treatment TAR without escalation** | | | | | | | | |
| CCAE | 10,071 | 23,425 | 5.9 | 17.0 | 30 | 70 | 5.08 (3.43 - 7.26) | 4.11 (3.20 - 5.19) |
| ODOD | 7,546 | 16,779 | 3.8 | 10.3 | 14 | 90 | 3.64 (1.99 - 6.11) | 8.71 (7.00 - 10.7) |
| MDCR | 709 | 2,032 | 0.4 | 1.5 | <5 | 14 | < 12.3 (3.99 - 28.7) | 9.64 (5.27 - 16.2) |
| Open Claims | 150,437 | 316,207 | 90.7 | 234.6 | 288 | 1,000 | 3.18 (2.82 - 3.57) | 4.26 (4.00 - 4.54) |
| OEHR | 8,530 | 19,723 | 2.1 | 5.5 | 9 | 25 | 4.36 (1.99 - 8.27) | 4.51 (2.92 - 6.66) |
| VA | 1,307 | 11,651 | 0.7 | 6.1 | <10 | 31 | < 14.1 (6.78 - 26.0) | 5.10 (3.46 - 7.24) |
| Meta-analysis | 176,584 | 376,134 | 102.5 | 267.5 | 341 | 1,185 | 3.33 (2.98 - 3.70) | 4.43 (4.18 - 4.69) |
| Meta-analysis (w/o USOC) | 26,147 | 59,927 | 11.8 | 32.9 | 53 | 185 | 4.49 (3.36 - 5.87) | 5.62 (4.84 - 6.49) |

##### 4.1.4 Acute renal failure

**Supplementary Table 293:** Exposures, outcomes and risk estimates for Acute renal failure in GLP1RA (male) (T) vs SGLT2I (male) (C) new-user cohorts under four different propensity score (PS) adjustment / time-at-risk (TAR) designs across data sources. We report total patient exposure time in 1,000 patient-year (KYr) units, uncalibrated and calibrated hazard ratios (HRs) and their 95% confidence intervals (CIs). Greyed out entries did not pass study diagnostics and are included only for completeness.

| | Patients | | Exposure (KYr) | | Outcomes | | Incidence (per KYr) | |
|---|---|---|---|---|---|---|---|---|
| | T | C | T | C | T | C | T | C |
| **PS-matching / on-treatment TAR** | | | | | | | | |
| CCAE | 9,463 | 23,693 | 6.4 | 19.7 | 57 | 127 | 8.87 (6.72 - 11.5) | 6.44 (5.37 - 7.67) |
| ODOD | 6,861 | 16,732 | 4.0 | 11.7 | 79 | 198 | 19.9 (15.8 - 24.9) | 17.0 (14.7 - 19.5) |
| MDCR | 661 | 2,014 | 0.4 | 1.6 | 18 | 36 | 42.8 (25.3 - 67.6) | 22.3 (15.6 - 30.8) |
| Open Claims | 140,078 | 320,717 | 98.4 | 280.5 | 846 | 2,328 | 8.59 (8.02 - 9.19) | 8.30 (7.97 - 8.65) |
| OEHR | 7,820 | 19,901 | 2.2 | 6.4 | 28 | 66 | 12.8 (8.47 - 18.4) | 10.4 (8.03 - 13.2) |
| VA | 1,126 | 10,563 | 0.8 | 6.4 | <10 | 72 | < 12.7 (6.09 - 23.3) | 11.3 (8.80 - 14.2) |
| Meta-analysis | 165,348 | 391,606 | 111.8 | 324.6 | <1,020 | 2,791 | < 9.12 (8.57 - 9.70) | 8.60 (8.28 - 8.92) |
| Meta-analysis (w/o USOC) | 25,270 | 70,889 | 13.4 | 44.1 | <174 | 463 | < 13.0 (11.2 - 15.1) | 10.5 (9.56 - 11.5) |
| **PS-stratification / on-treatment TAR** | | | | | | | | |
| CCAE | 10,081 | 23,699 | 6.7 | 19.7 | 58 | 127 | 8.62 (6.55 - 11.1) | 6.44 (5.37 - 7.66) |
| ODOD | 7,422 | 16,747 | 4.2 | 11.7 | 87 | 199 | 20.6 (16.5 - 25.4) | 17.0 (14.7 - 19.6) |
| MDCR | 692 | 2,014 | 0.4 | 1.6 | 19 | 36 | 43.5 (26.2 - 68.0) | 22.3 (15.6 - 30.8) |
| Open Claims | 149,976 | 320,733 | 103.3 | 280.5 | 897 | 2,329 | 8.68 (8.12 - 9.27) | 8.30 (7.97 - 8.65) |
| OEHR | 8,532 | 19,913 | 2.4 | 6.4 | 32 | 66 | 13.5 (9.24 - 19.1) | 10.4 (8.03 - 13.2) |
| VA | 1,290 | 11,731 | 0.9 | 7.1 | <10 | 93 | < 11.2 (5.36 - 20.6) | 13.1 (10.6 - 16.0) |
| Meta-analysis | 176,011 | 381,092 | 116.6 | 318.2 | 1,074 | 2,721 | 9.21 (8.67 - 9.78) | 8.55 (8.23 - 8.88) |
| Meta-analysis (w/o USOC) | 26,035 | 60,359 | 13.3 | 37.8 | 177 | 392 | 13.3 (11.4 - 15.4) | 10.4 (9.38 - 11.5) |
| **PS-matching / on-treatment TAR without escalation** | | | | | | | | |
| CCAE | 9,463 | 23,693 | 5.6 | 17.2 | 49 | 102 | 8.71 (6.44 - 11.5) | 5.92 (4.82 - 7.18) |
| ODOD | 6,861 | 16,732 | 3.5 | 10.4 | 69 | 166 | 19.5 (15.2 - 24.7) | 16.0 (13.7 - 18.6) |
| MDCR | 661 | 2,014 | 0.4 | 1.5 | 14 | 32 | 36.9 (20.1 - 61.8) | 22.0 (15.1 - 31.1) |
| Open Claims | 140,078 | 320,717 | 85.9 | 238.6 | 707 | 1,919 | 8.23 (7.64 - 8.86) | 8.04 (7.69 - 8.41) |
| OEHR | 7,820 | 19,901 | 1.9 | 5.6 | 24 | 46 | 12.5 (8.03 - 18.6) | 8.21 (6.01 - 10.9) |
| VA | 1,126 | 10,563 | 0.6 | 5.5 | <10 | 61 | < 16.2 (7.79 - 29.9) | 11.0 (8.45 - 14.2) |
| Meta-analysis | 165,348 | 391,606 | 97.6 | 277.4 | <859 | 2,294 | < 8.80 (8.23 - 9.41) | 8.27 (7.94 - 8.62) |
| Meta-analysis (w/o USOC) | 25,270 | 70,889 | 11.7 | 38.7 | <152 | 375 | < 13.0 (11.0 - 15.2) | 9.68 (8.73 - 10.7) |
| **PS-stratification / on-treatment TAR without escalation** | | | | | | | | |
| CCAE | 10,081 | 23,699 | 5.9 | 17.3 | 50 | 102 | 8.47 (6.28 - 11.2) | 5.91 (4.82 - 7.18) |
| ODOD | 7,422 | 16,747 | 3.8 | 10.4 | 77 | 167 | 20.4 (16.1 - 25.5) | 16.1 (13.7 - 18.7) |
| MDCR | 692 | 2,014 | 0.4 | 1.5 | 15 | 32 | 38.1 (21.3 - 62.8) | 22.0 (15.1 - 31.1) |
| Open Claims | 149,976 | 320,733 | 90.3 | 238.7 | 752 | 1,920 | 8.32 (7.74 - 8.94) | 8.05 (7.69 - 8.41) |
| OEHR | 8,532 | 19,913 | 2.1 | 5.6 | 27 | 46 | 13.0 (8.59 - 19.0) | 8.20 (6.01 - 10.9) |
| VA | 1,290 | 11,731 | 0.7 | 6.2 | <10 | 80 | < 14.3 (6.84 - 26.2) | 13.0 (10.3 - 16.2) |
| Meta-analysis | 176,011 | 381,092 | 102.1 | 271.9 | 906 | 2,235 | 8.88 (8.31 - 9.47) | 8.22 (7.88 - 8.57) |
| Meta-analysis (w/o USOC) | 26,035 | 60,359 | 11.7 | 33.2 | 154 | 315 | 13.1 (11.1 - 15.4) | 9.48 (8.46 - 10.6) |

##### 4.1.5 Glycemic control

**Supplementary Table 294:** Exposures, outcomes and risk estimates for Glycemic control in GLP1RA (male) (T) vs SGLT2I (male) (C) new-user cohorts under four different propensity score (PS) adjustment / time-at-risk (TAR) designs across data sources. We report total patient exposure time in 1,000 patient-year (KYr) units, uncalibrated and calibrated hazard ratios (HRs) and their 95% confidence intervals (CIs). Greyed out entries did not pass study diagnostics and are included only for completeness.

| | Patients | | Exposure (KYr) | | Outcomes | | Incidence (per KYr) | |
|---|---|---|---|---|---|---|---|---|
| | T | C | T | C | T | C | T | C |
| **PS-matching / on-treatment TAR** | | | | | | | | |
| CCAE | 9,646 | 23,972 | 6.6 | 20.0 | <5 | 26 | < 0.76 (0.25 - 1.78) | 1.30 (0.85 - 1.91) |
| ODOD | 4,723 | 11,454 | 2.5 | 7.3 | 280 | 760 | 111 (98.1 - 124) | 105 (97.4 - 112) |
| MDCR | 700 | 2,128 | 0.4 | 1.7 | <5 | 0 | < 11.1 (3.61 - 26.0) | |
| Open Claims | 145,214 | 332,772 | 102.0 | 289.6 | 0 | 0 | | |
| OEHR | 7,951 | 20,219 | 2.2 | 6.5 | 0 | 0 | | |
| VA | 1,123 | 10,656 | 0.8 | 6.4 | 0 | 10 | | 1.56 (0.75 - 2.87) |
| Meta-analysis | 14,369 | 35,426 | 9.1 | 27.2 | <285 | 786 | < 31.4 (27.8 - 35.2) | 28.9 (26.9 - 31.0) |
| Meta-analysis (w/o USOC) | 14,369 | 35,426 | 9.1 | 27.2 | <285 | 786 | < 31.4 (27.8 - 35.2) | 28.9 (26.9 - 31.0) |
| **PS-stratification / on-treatment TAR** | | | | | | | | |
| CCAE | 10,271 | 23,976 | 6.9 | 20.0 | <5 | 26 | < 0.73 (0.24 - 1.70) | 1.30 (0.85 - 1.91) |
| ODOD | 5,040 | 11,471 | 2.7 | 7.3 | 299 | 761 | 112 (99.7 - 125) | 105 (97.3 - 112) |
| MDCR | 730 | 2,128 | 0.5 | 1.7 | <5 | 0 | < 10.8 (3.50 - 25.1) | |
| Open Claims | 155,418 | 332,801 | 107.0 | 289.7 | 0 | 0 | | |
| OEHR | 8,668 | 20,231 | 2.4 | 6.5 | 0 | 0 | | |
| VA | 1,288 | 11,946 | 0.9 | 7.2 | 0 | 12 | | 1.67 (0.86 - 2.92) |
| Meta-analysis | 15,311 | 35,447 | 9.5 | 27.2 | <304 | 787 | < 31.9 (28.4 - 35.7) | 28.9 (26.9 - 31.0) |
| Meta-analysis (w/o USOC) | 15,311 | 35,447 | 9.5 | 27.2 | <304 | 787 | < 31.9 (28.4 - 35.7) | 28.9 (26.9 - 31.0) |
| **PS-matching / on-treatment TAR without escalation** | | | | | | | | |
| CCAE | 9,646 | 23,972 | 5.7 | 17.4 | <5 | 25 | < 0.87 (0.28 - 2.04) | 1.44 (0.93 - 2.12) |
| ODOD | 4,723 | 11,454 | 2.2 | 6.5 | 251 | 668 | 113 (99.0 - 127) | 103 (95.5 - 111) |
| MDCR | 700 | 2,128 | 0.4 | 1.5 | <5 | 0 | < 12.4 (4.01 - 28.9) | |
| Open Claims | 145,214 | 332,772 | 88.9 | 246.2 | 0 | 0 | | |
| OEHR | 7,951 | 20,219 | 1.9 | 5.7 | 0 | 0 | | |
| VA | 1,123 | 10,656 | 0.6 | 5.5 | 0 | <10 | | < 1.81 (0.87 - 3.33) |
| Meta-analysis | 14,369 | 35,426 | 8.0 | 23.9 | <256 | 693 | < 32.2 (28.3 - 36.3) | 29.0 (26.9 - 31.3) |
| Meta-analysis (w/o USOC) | 14,369 | 35,426 | 8.0 | 23.9 | <256 | 693 | < 32.2 (28.3 - 36.3) | 29.0 (26.9 - 31.3) |
| **PS-stratification / on-treatment TAR without escalation** | | | | | | | | |
| CCAE | 10,271 | 23,976 | 6.0 | 17.4 | <5 | 25 | < 0.83 (0.27 - 1.94) | 1.44 (0.93 - 2.12) |
| ODOD | 5,040 | 11,471 | 2.4 | 6.5 | 269 | 669 | 114 (101 - 129) | 103 (95.4 - 111) |
| MDCR | 730 | 2,128 | 0.4 | 1.5 | <5 | 0 | < 12.0 (3.88 - 27.9) | |
| Open Claims | 155,418 | 332,801 | 93.5 | 246.2 | 0 | 0 | | |
| OEHR | 8,668 | 20,231 | 2.1 | 5.7 | 0 | 0 | | |
| VA | 1,288 | 11,946 | 0.7 | 6.2 | 0 | <10 | | < 1.61 (0.77 - 2.96) |
| Meta-analysis | 15,311 | 35,447 | 8.4 | 23.9 | <274 | 694 | < 32.7 (29.0 - 36.9) | 29.0 (26.9 - 31.3) |
| Meta-analysis (w/o USOC) | 15,311 | 35,447 | 8.4 | 23.9 | <274 | 694 | < 32.7 (29.0 - 36.9) | 29.0 (26.9 - 31.3) |

##### 4.1.6 Hospitalization with heart failure

**Supplementary Table 295:** Exposures, outcomes and risk estimates for Hospitalization with heart failure in GLP1RA (male) (T) vs SGLT2I (male) (C) new-user cohorts under four different propensity score (PS) adjustment / time-at-risk (TAR) designs across data sources. We report total patient exposure time in 1,000 patient-year (KYr) units, uncalibrated and calibrated hazard ratios (HRs) and their 95% confidence intervals (CIs). Greyed out entries did not pass study diagnostics and are included only for completeness.

| | Patients | | Exposure (KYr) | | Outcomes | | Incidence (per KYr) | |
|---|---|---|---|---|---|---|---|---|
| | T | C | T | C | T | C | T | C |
| **PS-matching / on-treatment TAR** | | | | | | | | |
| CCAE | 9,478 | 23,592 | 6.4 | 19.6 | 44 | 113 | 6.82 (4.96 - 9.16) | 5.75 (4.74 - 6.92) |
| ODOD | 6,804 | 16,405 | 3.9 | 11.5 | 47 | 163 | 11.9 (8.77 - 15.9) | 14.2 (12.1 - 16.5) |
| MDCR | 648 | 1,957 | 0.4 | 1.6 | 11 | 32 | 26.4 (13.2 - 47.2) | 20.4 (14.0 - 28.8) |
| Open Claims | 139,941 | 315,167 | 98.5 | 277.3 | 507 | 1,629 | 5.15 (4.71 - 5.62) | 5.87 (5.59 - 6.17) |
| OEHR | 7,782 | 19,667 | 2.2 | 6.3 | 12 | 63 | 5.50 (2.84 - 9.61) | 10.0 (7.70 - 12.8) |
| VA | 1,125 | 10,173 | 0.8 | 6.2 | <10 | 56 | < 12.8 (6.16 - 23.6) | 9.06 (6.84 - 11.8) |
| Meta-analysis | 165,130 | 385,004 | 111.8 | 320.9 | <620 | 2,024 | < 5.54 (5.12 - 6.00) | 6.31 (6.04 - 6.59) |
| Meta-analysis (w/o USOC) | 25,189 | 69,837 | 13.3 | 43.6 | <113 | 395 | < 8.47 (6.98 - 10.2) | 9.06 (8.19 - 10.00) |
| **PS-stratification / on-treatment TAR** | | | | | | | | |
| CCAE | 10,091 | 23,598 | 6.7 | 19.7 | 47 | 113 | 6.97 (5.12 - 9.27) | 5.75 (4.74 - 6.91) |
| ODOD | 7,372 | 16,421 | 4.2 | 11.5 | 51 | 163 | 12.1 (9.03 - 15.9) | 14.2 (12.1 - 16.5) |
| MDCR | 679 | 1,962 | 0.4 | 1.6 | 11 | 33 | 25.5 (12.7 - 45.6) | 20.9 (14.4 - 29.4) |
| Open Claims | 150,000 | 315,211 | 103.5 | 277.3 | 537 | 1,631 | 5.19 (4.76 - 5.65) | 5.88 (5.60 - 6.17) |
| OEHR | 8,499 | 19,700 | 2.4 | 6.3 | 16 | 64 | 6.80 (3.89 - 11.0) | 10.2 (7.83 - 13.0) |
| VA | 1,287 | 11,226 | 0.9 | 6.8 | <10 | 73 | < 11.2 (5.36 - 20.5) | 10.7 (8.37 - 13.4) |
| Meta-analysis | 175,962 | 374,930 | 116.8 | 314.8 | 651 | 1,971 | 5.57 (5.15 - 6.02) | 6.26 (5.99 - 6.54) |
| Meta-analysis (w/o USOC) | 25,962 | 59,719 | 13.3 | 37.5 | 114 | 340 | 8.57 (7.07 - 10.3) | 9.07 (8.14 - 10.1) |
| **PS-matching / on-treatment TAR without escalation** | | | | | | | | |
| CCAE | 9,478 | 23,592 | 5.6 | 17.2 | 39 | 93 | 6.91 (4.92 - 9.45) | 5.41 (4.37 - 6.63) |
| ODOD | 6,804 | 16,405 | 3.5 | 10.2 | 44 | 144 | 12.5 (9.11 - 16.8) | 14.1 (11.9 - 16.6) |
| MDCR | 648 | 1,957 | 0.4 | 1.4 | 9 | 28 | 23.9 (10.9 - 45.4) | 20.0 (13.3 - 28.9) |
| Open Claims | 139,941 | 315,167 | 85.9 | 235.8 | 434 | 1,367 | 5.05 (4.59 - 5.55) | 5.80 (5.49 - 6.11) |
| OEHR | 7,782 | 19,667 | 1.9 | 5.5 | 11 | 49 | 5.79 (2.89 - 10.4) | 8.86 (6.55 - 11.7) |
| VA | 1,125 | 10,173 | 0.6 | 5.3 | <10 | 51 | < 16.4 (7.88 - 30.2) | 9.57 (7.13 - 12.6) |
| Meta-analysis | 165,130 | 385,004 | 97.5 | 274.0 | <538 | 1,704 | < 5.52 (5.06 - 6.00) | 6.22 (5.93 - 6.52) |
| Meta-analysis (w/o USOC) | 25,189 | 69,837 | 11.7 | 38.2 | <104 | 337 | < 8.92 (7.29 - 10.8) | 8.81 (7.90 - 9.81) |
| **PS-stratification / on-treatment TAR without escalation** | | | | | | | | |
| CCAE | 10,091 | 23,598 | 5.9 | 17.2 | 42 | 93 | 7.10 (5.12 - 9.60) | 5.41 (4.37 - 6.63) |
| ODOD | 7,372 | 16,421 | 3.7 | 10.2 | 48 | 144 | 12.8 (9.45 - 17.0) | 14.1 (11.9 - 16.6) |
| MDCR | 679 | 1,962 | 0.4 | 1.4 | 9 | 28 | 23.1 (10.6 - 43.9) | 19.9 (13.2 - 28.7) |
| Open Claims | 150,000 | 315,211 | 90.4 | 235.8 | 462 | 1,369 | 5.11 (4.65 - 5.60) | 5.81 (5.50 - 6.12) |
| OEHR | 8,499 | 19,700 | 2.1 | 5.5 | 15 | 50 | 7.30 (4.09 - 12.0) | 9.02 (6.70 - 11.9) |
| VA | 1,287 | 11,226 | 0.7 | 5.9 | <10 | 66 | < 14.4 (6.88 - 26.4) | 11.2 (8.64 - 14.2) |
| Meta-analysis | 175,962 | 374,930 | 102.2 | 268.7 | 567 | 1,656 | 5.55 (5.10 - 6.03) | 6.16 (5.87 - 6.47) |
| Meta-analysis (w/o USOC) | 25,962 | 59,719 | 11.7 | 32.9 | 105 | 287 | 8.97 (7.33 - 10.9) | 8.71 (7.73 - 9.78) |

##### 4.1.7 Measured renal dysfunction

**Supplementary Table 296:** Exposures, outcomes and risk estimates for Measured renal dysfunction in GLP1RA (male) (T) vs SGLT2I (male) (C) new-user cohorts under four different propensity score (PS) adjustment / time-at-risk (TAR) designs across data sources. We report total patient exposure time in 1,000 patient-year (KYr) units, uncalibrated and calibrated hazard ratios (HRs) and their 95% confidence intervals (CIs). Greyed out entries did not pass study diagnostics and are included only for completeness.

| | Patients | | Exposure (KYr) | | Outcomes | | Incidence (per KYr) | |
|---|---|---|---|---|---|---|---|---|
| | T | C | T | C | T | C | T | C |
| **PS-matching / on-treatment TAR** | | | | | | | | |
| CCAE | 9,689 | 24,078 | 6.6 | 20.0 | 0 | 5 | | 0.25 (0.08 - 0.58) |
| ODOD | 7,134 | 17,452 | 4.1 | 12.1 | 6 | 6 | 1.45 (0.53 - 3.16) | 0.50 (0.18 - 1.08) |
| MDCR | 701 | 2,136 | 0.4 | 1.7 | 0 | 0 | | |
| Open Claims | 145,214 | 332,772 | 102.0 | 289.6 | 0 | 0 | | |
| OEHR | 7,951 | 20,219 | 2.2 | 6.5 | 0 | 0 | | |
| VA | 1,166 | 11,040 | 0.8 | 6.7 | <10 | 15 | < 12.3 (5.91 - 22.6) | 2.25 (1.26 - 3.71) |
| **PS-stratification / on-treatment TAR** | | | | | | | | |
| CCAE | 10,321 | 24,082 | 6.9 | 20.0 | 0 | 5 | | 0.25 (0.08 - 0.58) |
| ODOD | 7,725 | 17,471 | 4.4 | 12.1 | 6 | 7 | 1.36 (0.50 - 2.96) | 0.58 (0.23 - 1.19) |
| MDCR | 731 | 2,136 | 0.5 | 1.7 | 0 | 0 | | |
| Open Claims | 155,418 | 332,801 | 107.0 | 289.7 | 0 | 0 | | |
| OEHR | 8,668 | 20,231 | 2.4 | 6.5 | 0 | 0 | | |
| VA | 1,336 | 12,350 | 0.9 | 7.4 | <10 | 23 | < 10.8 (5.16 - 19.8) | 3.09 (1.96 - 4.64) |
| **PS-matching / on-treatment TAR without escalation** | | | | | | | | |
| CCAE | 9,689 | 24,078 | 5.8 | 17.5 | 0 | 5 | | 0.29 (0.09 - 0.67) |
| ODOD | 7,134 | 17,452 | 3.7 | 10.7 | <5 | 6 | < 1.36 (0.44 - 3.16) | 0.56 (0.21 - 1.22) |
| MDCR | 701 | 2,136 | 0.4 | 1.5 | 0 | 0 | | |
| Open Claims | 145,214 | 332,772 | 88.9 | 246.2 | 0 | 0 | | |
| OEHR | 7,951 | 20,219 | 1.9 | 5.7 | 0 | 0 | | |
| VA | 1,166 | 11,040 | 0.6 | 5.8 | <10 | 13 | < 15.8 (7.57 - 29.0) | 2.26 (1.20 - 3.87) |
| **PS-stratification / on-treatment TAR without escalation** | | | | | | | | |
| CCAE | 10,321 | 24,082 | 6.0 | 17.5 | 0 | 5 | | 0.29 (0.09 - 0.67) |
| ODOD | 7,725 | 17,471 | 3.9 | 10.8 | <5 | 7 | < 1.27 (0.41 - 2.97) | 0.65 (0.26 - 1.34) |
| MDCR | 731 | 2,136 | 0.4 | 1.5 | 0 | 0 | | |
| Open Claims | 155,418 | 332,801 | 93.5 | 246.2 | 0 | 0 | | |
| OEHR | 8,668 | 20,231 | 2.1 | 5.7 | 0 | 0 | | |
| VA | 1,336 | 12,350 | 0.7 | 6.4 | <10 | 18 | < 13.8 (6.60 - 25.3) | 2.79 (1.65 - 4.41) |

###### 4.1.8 Stroke

**Supplementary Table 297:** Exposures, outcomes and risk estimates for Stroke in GLP1RA (male) (T) vs SGLT2I (male) (C) new-user cohorts under four different propensity score (PS) adjustment / time-at-risk (TAR) designs across data sources. We report total patient exposure time in 1,000 patient-year (KYr) units, uncalibrated and calibrated hazard ratios (HRs) and their 95% confidence intervals (CIs). Greyed out entries did not pass study diagnostics and are included only for completeness.

| | Patients | | Exposure (KYr) | | Outcomes | | Incidence (per KYr) | |
|---|---|---|---|---|---|---|---|---|
| | T | C | T | C | T | C | T | C |
| **PS-matching / on-treatment TAR** | | | | | | | | |
| CCAE | 9,572 | 23,839 | 6.5 | 19.9 | 20 | 58 | 3.09 (1.89 - 4.77) | 2.92 (2.22 - 3.78) |
| ODOD | 7,003 | 17,071 | 4.1 | 11.8 | 19 | 81 | 4.68 (2.82 - 7.31) | 6.86 (5.45 - 8.52) |
| MDCR | 675 | 2,074 | 0.4 | 1.7 | <5 | 18 | < 11.8 (3.83 - 27.5) | 10.8 (6.43 - 17.1) |
| Open Claims | 142,136 | 324,458 | 99.9 | 283.2 | 277 | 934 | 2.77 (2.45 - 3.12) | 3.30 (3.09 - 3.52) |
| OEHR | 7,882 | 20,026 | 2.2 | 6.4 | 11 | 19 | 4.98 (2.48 - 8.90) | 2.97 (1.79 - 4.64) |
| VA | 1,158 | 10,944 | 0.8 | 6.6 | <10 | 12 | < 12.3 (5.91 - 22.7) | 1.81 (0.94 - 3.17) |
| Meta-analysis | 166,593 | 385,394 | 112.7 | 321.3 | 327 | 1,092 | 2.90 (2.60 - 3.23) | 3.40 (3.20 - 3.61) |
| Meta-analysis (w/o USOC) | 24,457 | 60,936 | 12.7 | 38.1 | 50 | 158 | 3.92 (2.91 - 5.17) | 4.15 (3.53 - 4.85) |
| **PS-stratification / on-treatment TAR** | | | | | | | | |
| CCAE | 10,198 | 23,843 | 6.8 | 19.9 | 21 | 58 | 3.10 (1.92 - 4.73) | 2.92 (2.22 - 3.77) |
| ODOD | 7,593 | 17,093 | 4.3 | 11.8 | 21 | 81 | 4.84 (3.00 - 7.40) | 6.85 (5.44 - 8.51) |
| MDCR | 707 | 2,074 | 0.4 | 1.7 | <5 | 18 | < 11.3 (3.67 - 26.4) | 10.8 (6.43 - 17.1) |
| Open Claims | 152,244 | 324,481 | 104.9 | 283.2 | 291 | 934 | 2.77 (2.46 - 3.11) | 3.30 (3.09 - 3.52) |
| OEHR | 8,594 | 20,037 | 2.4 | 6.4 | 11 | 19 | 4.61 (2.30 - 8.24) | 2.97 (1.79 - 4.63) |
| VA | 1,329 | 12,216 | 0.9 | 7.4 | <10 | 19 | < 10.8 (5.16 - 19.8) | 2.58 (1.55 - 4.02) |
| Meta-analysis | 178,629 | 385,454 | 118.4 | 321.3 | 344 | 1,092 | 2.90 (2.61 - 3.23) | 3.40 (3.20 - 3.61) |
| Meta-analysis (w/o USOC) | 26,385 | 60,973 | 13.5 | 38.1 | 53 | 158 | 3.92 (2.94 - 5.13) | 4.15 (3.53 - 4.85) |
| **PS-matching / on-treatment TAR without escalation** | | | | | | | | |
| CCAE | 9,572 | 23,839 | 5.7 | 17.3 | 17 | 46 | 3.00 (1.75 - 4.80) | 2.65 (1.94 - 3.54) |
| ODOD | 7,003 | 17,071 | 3.6 | 10.5 | 17 | 72 | 4.69 (2.73 - 7.52) | 6.87 (5.37 - 8.65) |
| MDCR | 675 | 2,074 | 0.4 | 1.5 | <5 | 15 | < 13.1 (4.25 - 30.5) | 10.1 (5.67 - 16.7) |
| Open Claims | 142,136 | 324,458 | 87.1 | 240.8 | 233 | 806 | 2.67 (2.34 - 3.04) | 3.35 (3.12 - 3.59) |
| OEHR | 7,882 | 20,026 | 1.9 | 5.6 | 9 | 12 | 4.67 (2.13 - 8.86) | 2.13 (1.10 - 3.72) |
| VA | 1,158 | 10,944 | 0.6 | 5.7 | <10 | 10 | < 15.8 (7.58 - 29.1) | 1.75 (0.84 - 3.23) |
| Meta-analysis | 166,593 | 385,394 | 98.3 | 274.2 | 276 | 936 | 2.81 (2.49 - 3.16) | 3.41 (3.20 - 3.64) |
| Meta-analysis (w/o USOC) | 24,457 | 60,936 | 11.2 | 33.5 | 43 | 130 | 3.83 (2.77 - 5.16) | 3.88 (3.25 - 4.61) |
| **PS-stratification / on-treatment TAR without escalation** | | | | | | | | |
| CCAE | 10,198 | 23,843 | 6.0 | 17.3 | 18 | 46 | 3.02 (1.79 - 4.78) | 2.65 (1.94 - 3.54) |
| ODOD | 7,593 | 17,093 | 3.9 | 10.5 | 19 | 72 | 4.91 (2.96 - 7.67) | 6.85 (5.36 - 8.63) |
| MDCR | 707 | 2,074 | 0.4 | 1.5 | <5 | 15 | < 12.5 (4.07 - 29.3) | 10.1 (5.67 - 16.7) |
| Open Claims | 152,244 | 324,481 | 91.7 | 240.8 | 245 | 806 | 2.67 (2.35 - 3.03) | 3.35 (3.12 - 3.59) |
| OEHR | 8,594 | 20,037 | 2.1 | 5.6 | 9 | 12 | 4.32 (1.97 - 8.19) | 2.13 (1.10 - 3.72) |
| VA | 1,329 | 12,216 | 0.7 | 6.4 | <10 | 14 | < 13.8 (6.60 - 25.3) | 2.19 (1.20 - 3.68) |
| Meta-analysis | 178,629 | 385,454 | 103.6 | 274.3 | 291 | 936 | 2.81 (2.50 - 3.15) | 3.41 (3.20 - 3.64) |
| Meta-analysis (w/o USOC) | 26,385 | 60,973 | 11.9 | 33.5 | 46 | 130 | 3.86 (2.83 - 5.15) | 3.88 (3.24 - 4.61) |

##### 4.1.9 Sudden cardiac death

**Supplementary Table 298:** Exposures, outcomes and risk estimates for Sudden cardiac death in GLP1RA (male) (T) vs SGLT2I (male) (C) new-user cohorts under four different propensity score (PS) adjustment / time-at-risk (TAR) designs across data sources. We report total patient exposure time in 1,000 patient-year (KYr) units, uncalibrated and calibrated hazard ratios (HRs) and their 95% confidence intervals (CIs). Greyed out entries did not pass study diagnostics and are included only for completeness.

| | Patients | | Exposure (KYr) | | Outcomes | | Incidence (per KYr) | |
|---|---|---|---|---|---|---|---|---|
| | T | C | T | C | T | C | T | C |
| **PS-matching / on-treatment TAR** | | | | | | | | |
| CCAE | 9,696 | 24,107 | 6.6 | 20.1 | <5 | 15 | < 0.76 (0.25 - 1.77) | 0.75 (0.42 - 1.23) |
| ODOD | 7,129 | 17,424 | 4.1 | 12.1 | 8 | 19 | 1.94 (0.84 - 3.81) | 1.57 (0.94 - 2.45) |
| MDCR | 698 | 2,130 | 0.4 | 1.7 | <5 | 11 | < 11.1 (3.62 - 26.0) | 6.47 (3.23 - 11.6) |
| Open Claims | 144,881 | 331,659 | 101.8 | 288.8 | 85 | 258 | 0.83 (0.67 - 1.03) | 0.89 (0.79 - 1.01) |
| OEHR | 7,945 | 20,193 | 2.2 | 6.5 | <5 | 5 | < 2.24 (0.73 - 5.22) | 0.77 (0.25 - 1.81) |
| VA | 1,181 | 11,164 | 0.8 | 6.7 | 0 | <10 | | < 1.49 (0.71 - 2.73) |
| Meta-analysis | 161,706 | 373,190 | 112.5 | 321.0 | <98 | 292 | < 0.87 (0.71 - 1.06) | 0.91 (0.81 - 1.02) |
| Meta-analysis (w/o USOC) | 16,825 | 41,531 | 10.7 | 32.2 | <13 | 34 | < 1.21 (0.65 - 2.07) | 1.06 (0.73 - 1.48) |
| **PS-stratification / on-treatment TAR** | | | | | | | | |
| CCAE | 10,325 | 24,111 | 6.9 | 20.1 | <5 | 15 | < 0.73 (0.24 - 1.69) | 0.75 (0.42 - 1.23) |
| ODOD | 7,720 | 17,442 | 4.4 | 12.1 | 9 | 19 | 2.04 (0.93 - 3.87) | 1.57 (0.94 - 2.45) |
| MDCR | 728 | 2,130 | 0.5 | 1.7 | <5 | 11 | < 10.8 (3.50 - 25.1) | 6.47 (3.23 - 11.6) |
| Open Claims | 155,084 | 331,685 | 106.8 | 288.8 | 87 | 259 | 0.81 (0.65 - 1.00) | 0.90 (0.79 - 1.01) |
| OEHR | 8,664 | 20,205 | 2.4 | 6.5 | 5 | 5 | 2.08 (0.67 - 4.85) | 0.77 (0.25 - 1.81) |
| VA | 1,352 | 12,496 | 0.9 | 7.5 | 0 | <10 | | < 1.33 (0.64 - 2.45) |
| Meta-analysis | 173,129 | 373,238 | 118.1 | 321.0 | <101 | 293 | < 0.86 (0.70 - 1.04) | 0.91 (0.81 - 1.02) |
| Meta-analysis (w/o USOC) | 18,045 | 41,553 | 11.3 | 32.2 | <14 | 34 | < 1.24 (0.68 - 2.08) | 1.06 (0.73 - 1.48) |
| **PS-matching / on-treatment TAR without escalation** | | | | | | | | |
| CCAE | 9,696 | 24,107 | 5.8 | 17.5 | <5 | 11 | < 0.87 (0.28 - 2.02) | 0.63 (0.31 - 1.12) |
| ODOD | 7,129 | 17,424 | 3.7 | 10.7 | 7 | 16 | 1.90 (0.76 - 3.92) | 1.49 (0.85 - 2.42) |
| MDCR | 698 | 2,130 | 0.4 | 1.5 | <5 | 10 | < 12.4 (4.02 - 28.9) | 6.59 (3.16 - 12.1) |
| Open Claims | 144,881 | 331,659 | 88.7 | 245.4 | 70 | 227 | 0.79 (0.62 - 1.00) | 0.92 (0.81 - 1.05) |
| OEHR | 7,945 | 20,193 | 1.9 | 5.7 | <5 | <5 | < 2.57 (0.83 - 6.00) | < 0.88 (0.29 - 2.05) |
| VA | 1,181 | 11,164 | 0.6 | 5.8 | 0 | <10 | | < 1.72 (0.83 - 3.17) |
| Meta-analysis | 152,010 | 349,083 | 92.4 | 256.2 | 77 | 243 | 0.83 (0.66 - 1.04) | 0.95 (0.83 - 1.08) |
| Meta-analysis (w/o USOC) | 7,129 | 17,424 | 3.7 | 10.7 | 7 | 16 | 1.90 (0.76 - 3.92) | 1.49 (0.85 - 2.42) |
| **PS-stratification / on-treatment TAR without escalation** | | | | | | | | |
| CCAE | 10,325 | 24,111 | 6.0 | 17.5 | <5 | 11 | < 0.83 (0.27 - 1.93) | 0.63 (0.31 - 1.12) |
| ODOD | 7,720 | 17,442 | 3.9 | 10.7 | 8 | 16 | 2.04 (0.88 - 4.01) | 1.49 (0.85 - 2.42) |
| MDCR | 728 | 2,130 | 0.4 | 1.5 | <5 | 10 | < 12.0 (3.89 - 27.9) | 6.59 (3.16 - 12.1) |
| Open Claims | 155,084 | 331,685 | 93.3 | 245.5 | 72 | 228 | 0.77 (0.60 - 0.97) | 0.93 (0.81 - 1.06) |
| OEHR | 8,664 | 20,205 | 2.1 | 5.7 | <5 | <5 | < 2.38 (0.77 - 5.56) | < 0.88 (0.29 - 2.05) |
| VA | 1,352 | 12,496 | 0.7 | 6.5 | 0 | <10 | | < 1.54 (0.74 - 2.83) |
| Meta-analysis | 162,804 | 349,127 | 97.2 | 256.2 | 80 | 244 | 0.82 (0.65 - 1.02) | 0.95 (0.84 - 1.08) |
| Meta-analysis (w/o USOC) | 7,720 | 17,442 | 3.9 | 10.7 | 8 | 16 | 2.04 (0.88 - 4.01) | 1.49 (0.85 - 2.42) |

##### 4.1.10 Abnormal weight gain

**Supplementary Table 299:** Exposures, outcomes and risk estimates for Abnormal weight gain in GLP1RA (male) (T) vs SGLT2I (male) (C) new-user cohorts under four different propensity score (PS) adjustment / time-at-risk (TAR) designs across data sources. We report total patient exposure time in 1,000 patient-year (KYr) units, uncalibrated and calibrated hazard ratios (HRs) and their 95% confidence intervals (CIs). Greyed out entries did not pass study diagnostics and are included only for completeness.

| | Patients | | Exposure (KYr) | | Outcomes | | Incidence (per KYr) | |
|---|---|---|---|---|---|---|---|---|
| | T | C | T | C | T | C | T | C |
| **PS-matching / on-treatment TAR** | | | | | | | | |
| CCAE | 9,199 | 23,296 | 6.3 | 19.3 | 30 | 68 | 4.80 (3.24 - 6.85) | 3.52 (2.74 - 4.47) |
| ODOD | 6,773 | 16,848 | 3.9 | 11.6 | 30 | 31 | 7.65 (5.16 - 10.9) | 2.66 (1.81 - 3.78) |
| MDCR | 682 | 2,099 | 0.4 | 1.7 | <5 | <5 | < 11.5 (3.75 - 26.9) | < 3.01 (0.98 - 7.02) |
| Open Claims | 138,102 | 321,351 | 96.9 | 279.0 | 209 | 317 | 2.16 (1.87 - 2.47) | 1.14 (1.01 - 1.27) |
| OEHR | 7,589 | 19,544 | 2.1 | 6.2 | 15 | 30 | 7.09 (3.97 - 11.7) | 4.81 (3.25 - 6.87) |
| VA | 1,144 | 10,914 | 0.8 | 6.6 | <10 | <10 | < 12.7 (6.08 - 23.3) | < 1.52 (0.73 - 2.79) |
| Meta-analysis | 161,663 | 381,039 | 109.2 | 316.2 | 284 | 446 | 2.60 (2.31 - 2.92) | 1.41 (1.28 - 1.55) |
| Meta-analysis (w/o USOC) | 23,561 | 59,688 | 12.3 | 37.2 | 75 | 129 | 6.10 (4.80 - 7.65) | 3.47 (2.90 - 4.12) |
| **PS-stratification / on-treatment TAR** | | | | | | | | |
| CCAE | 9,729 | 23,300 | 6.5 | 19.3 | 34 | 68 | 5.22 (3.61 - 7.29) | 3.52 (2.73 - 4.46) |
| ODOD | 7,282 | 16,867 | 4.2 | 11.7 | 38 | 32 | 9.14 (6.47 - 12.5) | 2.74 (1.88 - 3.87) |
| MDCR | 709 | 2,099 | 0.4 | 1.7 | <5 | <5 | < 11.2 (3.65 - 26.2) | < 3.01 (0.98 - 7.02) |
| Open Claims | 146,733 | 321,381 | 101.1 | 279.0 | 235 | 317 | 2.32 (2.04 - 2.64) | 1.14 (1.01 - 1.27) |
| OEHR | 8,164 | 19,556 | 2.3 | 6.2 | 20 | 30 | 8.87 (5.42 - 13.7) | 4.81 (3.24 - 6.86) |
| VA | 1,302 | 12,226 | 0.9 | 7.4 | <10 | <10 | < 11.1 (5.34 - 20.5) | < 1.36 (0.65 - 2.49) |
| Meta-analysis | 171,908 | 381,104 | 114.1 | 316.2 | 327 | 447 | 2.87 (2.56 - 3.20) | 1.41 (1.29 - 1.55) |
| Meta-analysis (w/o USOC) | 25,175 | 59,723 | 12.9 | 37.2 | 92 | 130 | 7.12 (5.74 - 8.73) | 3.49 (2.92 - 4.15) |
| **PS-matching / on-treatment TAR without escalation** | | | | | | | | |
| CCAE | 9,199 | 23,296 | 5.5 | 16.9 | 23 | 60 | 4.20 (2.66 - 6.30) | 3.56 (2.71 - 4.58) |
| ODOD | 6,773 | 16,848 | 3.5 | 10.4 | 23 | 24 | 6.58 (4.17 - 9.87) | 2.32 (1.49 - 3.45) |
| MDCR | 682 | 2,099 | 0.4 | 1.5 | <5 | <5 | < 12.8 (4.14 - 29.8) | < 3.36 (1.09 - 7.83) |
| Open Claims | 138,102 | 321,351 | 84.5 | 237.4 | 182 | 254 | 2.15 (1.85 - 2.49) | 1.07 (0.94 - 1.21) |
| OEHR | 7,589 | 19,544 | 1.8 | 5.5 | 14 | 24 | 7.60 (4.16 - 12.8) | 4.37 (2.80 - 6.51) |
| VA | 1,144 | 10,914 | 0.6 | 5.7 | <10 | <10 | < 16.2 (7.77 - 29.8) | < 1.76 (0.84 - 3.23) |
| Meta-analysis | 161,663 | 381,039 | 95.3 | 270.1 | 242 | 362 | 2.54 (2.23 - 2.88) | 1.34 (1.21 - 1.49) |
| Meta-analysis (w/o USOC) | 23,561 | 59,688 | 10.8 | 32.7 | 60 | 108 | 5.55 (4.23 - 7.14) | 3.30 (2.71 - 3.99) |
| **PS-stratification / on-treatment TAR without escalation** | | | | | | | | |
| CCAE | 9,729 | 23,300 | 5.7 | 16.9 | 27 | 60 | 4.72 (3.11 - 6.87) | 3.55 (2.71 - 4.57) |
| ODOD | 7,282 | 16,867 | 3.7 | 10.4 | 31 | 25 | 8.37 (5.68 - 11.9) | 2.41 (1.56 - 3.56) |
| MDCR | 709 | 2,099 | 0.4 | 1.5 | <5 | <5 | < 12.4 (4.04 - 29.0) | < 3.36 (1.09 - 7.83) |
| Open Claims | 146,733 | 321,381 | 88.3 | 237.4 | 206 | 254 | 2.33 (2.02 - 2.67) | 1.07 (0.94 - 1.21) |
| OEHR | 8,164 | 19,556 | 2.0 | 5.5 | 17 | 24 | 8.65 (5.04 - 13.8) | 4.37 (2.80 - 6.51) |
| VA | 1,302 | 12,226 | 0.7 | 6.4 | <10 | <10 | < 14.2 (6.79 - 26.0) | < 1.57 (0.75 - 2.88) |
| Meta-analysis | 171,908 | 381,104 | 99.7 | 270.1 | 281 | 363 | 2.82 (2.50 - 3.17) | 1.34 (1.21 - 1.49) |
| Meta-analysis (w/o USOC) | 25,175 | 59,723 | 11.4 | 32.7 | 75 | 109 | 6.59 (5.18 - 8.26) | 3.33 (2.73 - 4.02) |

##### 4.1.11 Abnormal weight loss

**Supplementary Table 300:** Exposures, outcomes and risk estimates for Abnormal weight loss in GLP1RA (male) (T) vs SGLT2I (male) (C) new-user cohorts under four different propensity score (PS) adjustment / time-at-risk (TAR) designs across data sources. We report total patient exposure time in 1,000 patient-year (KYr) units, uncalibrated and calibrated hazard ratios (HRs) and their 95% confidence intervals (CIs). Greyed out entries did not pass study diagnostics and are included only for completeness.

| | Patients | | Exposure (KYr) | | Outcomes | | Incidence (per KYr) | |
|---|---|---|---|---|---|---|---|---|
| | T | C | T | C | T | C | T | C |
| **PS-matching / on-treatment TAR** | | | | | | | | |
| CCAE | 9,478 | 23,564 | 6.4 | 19.6 | 58 | 145 | 9.07 (6.88 - 11.7) | 7.41 (6.25 - 8.72) |
| ODOD | 6,937 | 16,871 | 4.0 | 11.6 | 54 | 184 | 13.5 (10.2 - 17.6) | 15.8 (13.6 - 18.3) |
| MDCR | 684 | 2,062 | 0.4 | 1.6 | 5 | 35 | 11.5 (3.72 - 26.7) | 21.4 (14.9 - 29.7) |
| Open Claims | 141,191 | 322,477 | 99.1 | 280.1 | 435 | 1,594 | 4.39 (3.98 - 4.82) | 5.69 (5.42 - 5.98) |
| OEHR | 7,738 | 19,632 | 2.2 | 6.3 | 21 | 79 | 9.75 (6.04 - 14.9) | 12.6 (10.00 - 15.7) |
| VA | 1,144 | 10,816 | 0.8 | 6.5 | <10 | 82 | < 12.5 (6.00 - 23.0) | 12.6 (10.0 - 15.6) |
| Meta-analysis | 166,488 | 393,360 | 112.5 | 324.0 | <578 | 2,084 | < 5.14 (4.73 - 5.57) | 6.43 (6.16 - 6.71) |
| Meta-analysis (w/o USOC) | 25,297 | 70,883 | 13.3 | 43.9 | <143 | 490 | < 10.7 (9.03 - 12.6) | 11.2 (10.2 - 12.2) |
| **PS-stratification / on-treatment TAR** | | | | | | | | |
| CCAE | 10,096 | 23,568 | 6.7 | 19.6 | 63 | 145 | 9.41 (7.23 - 12.0) | 7.41 (6.25 - 8.72) |
| ODOD | 7,505 | 16,888 | 4.3 | 11.6 | 58 | 184 | 13.6 (10.3 - 17.6) | 15.8 (13.6 - 18.3) |
| MDCR | 714 | 2,062 | 0.5 | 1.6 | 5 | 35 | 11.1 (3.60 - 25.8) | 21.4 (14.9 - 29.7) |
| Open Claims | 151,068 | 322,505 | 104.0 | 280.1 | 462 | 1,595 | 4.44 (4.05 - 4.87) | 5.69 (5.42 - 5.98) |
| OEHR | 8,433 | 19,644 | 2.3 | 6.3 | 23 | 79 | 9.87 (6.26 - 14.8) | 12.6 (9.99 - 15.7) |
| VA | 1,315 | 12,060 | 0.9 | 7.2 | 11 | 100 | 12.0 (5.97 - 21.4) | 13.8 (11.2 - 16.8) |
| Meta-analysis | 177,102 | 382,605 | 117.3 | 317.5 | 606 | 2,003 | 5.17 (4.76 - 5.59) | 6.31 (6.03 - 6.59) |
| Meta-analysis (w/o USOC) | 26,034 | 60,100 | 13.3 | 37.5 | 144 | 408 | 10.8 (9.14 - 12.8) | 10.9 (9.86 - 12.0) |
| **PS-matching / on-treatment TAR without escalation** | | | | | | | | |
| CCAE | 9,478 | 23,564 | 5.6 | 17.1 | 52 | 126 | 9.27 (6.93 - 12.2) | 7.37 (6.14 - 8.77) |
| ODOD | 6,937 | 16,871 | 3.6 | 10.3 | 47 | 160 | 13.2 (9.69 - 17.5) | 15.5 (13.2 - 18.1) |
| MDCR | 684 | 2,062 | 0.4 | 1.5 | 5 | 31 | 12.8 (4.14 - 29.8) | 21.1 (14.4 - 30.0) |
| Open Claims | 141,191 | 322,477 | 86.4 | 238.3 | 368 | 1,385 | 4.26 (3.83 - 4.72) | 5.81 (5.51 - 6.13) |
| OEHR | 7,738 | 19,632 | 1.9 | 5.5 | 18 | 70 | 9.57 (5.67 - 15.1) | 12.7 (9.91 - 16.1) |
| VA | 1,144 | 10,816 | 0.6 | 5.6 | <10 | 73 | < 15.9 (7.62 - 29.2) | 13.0 (10.2 - 16.4) |
| Meta-analysis | 166,488 | 393,360 | 98.1 | 276.8 | <495 | 1,814 | < 5.04 (4.61 - 5.51) | 6.55 (6.26 - 6.86) |
| Meta-analysis (w/o USOC) | 25,297 | 70,883 | 11.7 | 38.5 | <127 | 429 | < 10.9 (9.06 - 12.9) | 11.1 (10.1 - 12.2) |
| **PS-stratification / on-treatment TAR without escalation** | | | | | | | | |
| CCAE | 10,096 | 23,568 | 5.9 | 17.1 | 57 | 126 | 9.69 (7.34 - 12.5) | 7.37 (6.14 - 8.77) |
| ODOD | 7,505 | 16,888 | 3.8 | 10.3 | 51 | 160 | 13.4 (9.98 - 17.6) | 15.5 (13.2 - 18.1) |
| MDCR | 714 | 2,062 | 0.4 | 1.5 | 5 | 31 | 12.3 (4.00 - 28.8) | 21.1 (14.4 - 30.0) |
| Open Claims | 151,068 | 322,505 | 90.9 | 238.3 | 394 | 1,386 | 4.33 (3.92 - 4.78) | 5.82 (5.51 - 6.13) |
| OEHR | 8,433 | 19,644 | 2.0 | 5.5 | 19 | 70 | 9.32 (5.61 - 14.5) | 12.7 (9.90 - 16.1) |
| VA | 1,315 | 12,060 | 0.7 | 6.3 | <10 | 87 | < 13.8 (6.64 - 25.5) | 13.9 (11.1 - 17.1) |
| Meta-analysis | 177,102 | 382,605 | 102.6 | 271.2 | 521 | 1,742 | 5.08 (4.65 - 5.53) | 6.42 (6.12 - 6.73) |
| Meta-analysis (w/o USOC) | 26,034 | 60,100 | 11.7 | 32.9 | 127 | 356 | 10.8 (9.03 - 12.9) | 10.8 (9.71 - 12.0) |

##### 4.1.12 Acute pancreatitis

**Supplementary Table 301:** Exposures, outcomes and risk estimates for Acute pancreatitis in GLP1RA (male) (T) vs SGLT2I (male) (C) new-user cohorts under four different propensity score (PS) adjustment / time-at-risk (TAR) designs across data sources. We report total patient exposure time in 1,000 patient-year (KYr) units, uncalibrated and calibrated hazard ratios (HRs) and their 95% confidence intervals (CIs). Greyed out entries did not pass study diagnostics and are included only for completeness.

| | Patients | | Exposure (KYr) | | Outcomes | | Incidence (per KYr) | |
|---|---|---|---|---|---|---|---|---|
| | T | C | T | C | T | C | T | C |
| **PS-matching / on-treatment TAR** | | | | | | | | |
| CCAE | 9,638 | 23,886 | 6.6 | 19.9 | 16 | 42 | 2.44 (1.40 - 3.97) | 2.11 (1.52 - 2.86) |
| ODOD | 7,100 | 17,288 | 4.1 | 12.0 | 8 | 29 | 1.94 (0.84 - 3.83) | 2.42 (1.62 - 3.47) |
| MDCR | 696 | 2,119 | 0.4 | 1.7 | 0 | <5 | | < 2.96 (0.96 - 6.91) |
| Open Claims | 143,625 | 327,916 | 101.0 | 285.4 | 138 | 377 | 1.37 (1.15 - 1.61) | 1.32 (1.19 - 1.46) |
| OEHR | 7,909 | 20,096 | 2.2 | 6.4 | 7 | 13 | 3.15 (1.27 - 6.48) | 2.02 (1.08 - 3.46) |
| VA | 1,172 | 11,059 | 0.8 | 6.7 | 0 | <10 | | < 1.50 (0.72 - 2.76) |
| Meta-analysis | 160,363 | 369,090 | 111.7 | 317.3 | 162 | 448 | 1.45 (1.24 - 1.69) | 1.41 (1.28 - 1.55) |
| Meta-analysis (w/o USOC) | 16,738 | 41,174 | 10.7 | 31.9 | 24 | 71 | 2.25 (1.44 - 3.35) | 2.23 (1.74 - 2.81) |
| **PS-stratification / on-treatment TAR** | | | | | | | | |
| CCAE | 10,266 | 23,890 | 6.9 | 19.9 | 17 | 42 | 2.48 (1.44 - 3.97) | 2.11 (1.52 - 2.86) |
| ODOD | 7,691 | 17,307 | 4.4 | 12.0 | 8 | 29 | 1.82 (0.79 - 3.58) | 2.41 (1.62 - 3.47) |
| MDCR | 727 | 2,119 | 0.5 | 1.7 | 0 | <5 | | < 2.96 (0.96 - 6.91) |
| Open Claims | 153,856 | 327,945 | 106.0 | 285.4 | 149 | 377 | 1.41 (1.19 - 1.65) | 1.32 (1.19 - 1.46) |
| OEHR | 8,626 | 20,108 | 2.4 | 6.4 | 7 | 13 | 2.92 (1.17 - 6.01) | 2.02 (1.08 - 3.46) |
| VA | 1,342 | 12,372 | 0.9 | 7.4 | 0 | <10 | | < 1.34 (0.64 - 2.47) |
| Meta-analysis | 180,439 | 389,250 | 119.7 | 323.7 | 181 | 461 | 1.51 (1.30 - 1.75) | 1.42 (1.30 - 1.56) |
| Meta-analysis (w/o USOC) | 26,583 | 61,305 | 13.7 | 38.3 | 32 | 84 | 2.34 (1.60 - 3.31) | 2.19 (1.75 - 2.71) |
| **PS-matching / on-treatment TAR without escalation** | | | | | | | | |
| CCAE | 9,638 | 23,886 | 5.7 | 17.4 | 11 | 34 | 1.92 (0.96 - 3.43) | 1.96 (1.36 - 2.74) |
| ODOD | 7,100 | 17,288 | 3.7 | 10.6 | 7 | 23 | 1.91 (0.77 - 3.93) | 2.16 (1.37 - 3.24) |
| MDCR | 696 | 2,119 | 0.4 | 1.5 | 0 | <5 | | < 3.32 (1.08 - 7.74) |
| Open Claims | 143,625 | 327,916 | 88.0 | 242.8 | 115 | 306 | 1.31 (1.08 - 1.57) | 1.26 (1.12 - 1.41) |
| OEHR | 7,909 | 20,096 | 1.9 | 5.7 | 6 | 11 | 3.10 (1.14 - 6.74) | 1.95 (0.97 - 3.48) |
| VA | 1,172 | 11,059 | 0.6 | 5.7 | 0 | <10 | | < 1.74 (0.83 - 3.20) |
| Meta-analysis | 160,363 | 369,090 | 97.5 | 270.8 | 133 | 363 | 1.36 (1.14 - 1.62) | 1.34 (1.21 - 1.49) |
| Meta-analysis (w/o USOC) | 16,738 | 41,174 | 9.4 | 28.0 | 18 | 57 | 1.91 (1.13 - 3.02) | 2.04 (1.54 - 2.64) |
| **PS-stratification / on-treatment TAR without escalation** | | | | | | | | |
| CCAE | 10,266 | 23,890 | 6.0 | 17.4 | 12 | 34 | 1.99 (1.03 - 3.48) | 1.96 (1.36 - 2.74) |
| ODOD | 7,691 | 17,307 | 3.9 | 10.7 | 7 | 23 | 1.78 (0.72 - 3.68) | 2.16 (1.37 - 3.24) |
| MDCR | 727 | 2,119 | 0.4 | 1.5 | 0 | <5 | | < 3.32 (1.08 - 7.74) |
| Open Claims | 153,856 | 327,945 | 92.7 | 242.8 | 123 | 306 | 1.33 (1.10 - 1.58) | 1.26 (1.12 - 1.41) |
| OEHR | 8,626 | 20,108 | 2.1 | 5.7 | 6 | 11 | 2.86 (1.05 - 6.23) | 1.95 (0.97 - 3.48) |
| VA | 1,342 | 12,372 | 0.7 | 6.4 | 0 | <10 | | < 1.55 (0.75 - 2.86) |
| Meta-analysis | 171,813 | 369,142 | 102.6 | 270.9 | 142 | 363 | 1.38 (1.17 - 1.63) | 1.34 (1.21 - 1.49) |
| Meta-analysis (w/o USOC) | 17,957 | 41,197 | 9.9 | 28.0 | 19 | 57 | 1.91 (1.15 - 2.98) | 2.03 (1.54 - 2.64) |

###### 4.1.13 Bone fracture

**Supplementary Table 302:** Exposures, outcomes and risk estimates for Bone fracture in GLP1RA (male) (T) vs SGLT2I (male) (C) new-user cohorts under four different propensity score (PS) adjustment / time-at-risk (TAR) designs across data sources. We report total patient exposure time in 1,000 patient-year (KYr) units, uncalibrated and calibrated hazard ratios (HRs) and their 95% confidence intervals (CIs). Greyed out entries did not pass study diagnostics and are included only for completeness.

| | Patients | | Exposure (KYr) | | Outcomes | | Incidence (per KYr) | |
|---|---|---|---|---|---|---|---|---|
| | T | C | T | C | T | C | T | C |
| **PS-matching / on-treatment TAR** | | | | | | | | |
| CCAE | 8,868 | 22,294 | 5.9 | 18.2 | 93 | 296 | 15.8 (12.7 - 19.3) | 16.3 (14.5 - 18.2) |
| ODOD | 6,617 | 16,177 | 3.8 | 11.0 | 94 | 211 | 24.9 (20.1 - 30.4) | 19.1 (16.6 - 21.9) |
| MDCR | 634 | 1,967 | 0.4 | 1.6 | 8 | 38 | 21.1 (9.09 - 41.5) | 24.5 (17.3 - 33.6) |
| Open Claims | 126,901 | 295,510 | 88.8 | 256.4 | 943 | 2,775 | 10.6 (9.96 - 11.3) | 10.8 (10.4 - 11.2) |
| OEHR | 7,399 | 18,967 | 2.1 | 6.1 | 21 | 70 | 10.1 (6.27 - 15.5) | 11.5 (8.99 - 14.6) |
| VA | 1,059 | 10,010 | 0.7 | 6.0 | <10 | 50 | < 13.6 (6.51 - 25.0) | 8.27 (6.14 - 10.9) |
| Meta-analysis | 150,844 | 362,958 | 101.2 | 297.7 | <1,161 | 3,402 | < 11.5 (10.8 - 12.1) | 11.4 (11.0 - 11.8) |
| Meta-analysis (w/o USOC) | 23,943 | 67,448 | 12.5 | 41.4 | <218 | 627 | < 17.5 (15.2 - 19.9) | 15.2 (14.0 - 16.4) |
| **PS-stratification / on-treatment TAR** | | | | | | | | |
| CCAE | 9,460 | 22,298 | 6.2 | 18.2 | 99 | 296 | 16.0 (13.0 - 19.5) | 16.2 (14.4 - 18.2) |
| ODOD | 7,151 | 16,194 | 4.0 | 11.1 | 103 | 211 | 25.5 (20.8 - 30.9) | 19.1 (16.6 - 21.8) |
| MDCR | 661 | 1,967 | 0.4 | 1.6 | 8 | 38 | 20.3 (8.77 - 40.0) | 24.5 (17.3 - 33.6) |
| Open Claims | 135,417 | 295,535 | 92.9 | 256.4 | 1,015 | 2,775 | 10.9 (10.3 - 11.6) | 10.8 (10.4 - 11.2) |
| OEHR | 8,094 | 18,979 | 2.2 | 6.1 | 25 | 70 | 11.1 (7.20 - 16.4) | 11.5 (8.98 - 14.6) |
| VA | 1,215 | 11,119 | 0.8 | 6.7 | <10 | 52 | < 11.8 (5.67 - 21.7) | 7.74 (5.78 - 10.1) |
| Meta-analysis | 160,122 | 353,006 | 105.4 | 291.7 | 1,242 | 3,352 | 11.8 (11.1 - 12.5) | 11.5 (11.1 - 11.9) |
| Meta-analysis (w/o USOC) | 24,705 | 57,471 | 12.5 | 35.4 | 227 | 577 | 18.2 (15.9 - 20.7) | 16.3 (15.0 - 17.7) |
| **PS-matching / on-treatment TAR without escalation** | | | | | | | | |
| CCAE | 8,868 | 22,294 | 5.2 | 15.9 | 77 | 254 | 14.9 (11.8 - 18.6) | 16.0 (14.1 - 18.0) |
| ODOD | 6,617 | 16,177 | 3.4 | 9.8 | 82 | 186 | 24.3 (19.3 - 30.2) | 19.0 (16.3 - 21.9) |
| MDCR | 634 | 1,967 | 0.3 | 1.4 | 8 | 32 | 23.0 (9.95 - 45.4) | 23.1 (15.8 - 32.7) |
| Open Claims | 126,901 | 295,510 | 77.4 | 218.3 | 805 | 2,332 | 10.4 (9.70 - 11.1) | 10.7 (10.3 - 11.1) |
| OEHR | 7,399 | 18,967 | 1.8 | 5.3 | 17 | 62 | 9.43 (5.49 - 15.1) | 11.6 (8.91 - 14.9) |
| VA | 1,059 | 10,010 | 0.6 | 5.2 | <10 | 45 | < 17.3 (8.30 - 31.8) | 8.61 (6.28 - 11.5) |
| Meta-analysis | 150,844 | 362,958 | 88.3 | 254.6 | <991 | 2,879 | < 11.2 (10.5 - 11.9) | 11.3 (10.9 - 11.7) |
| Meta-analysis (w/o USOC) | 23,943 | 67,448 | 10.9 | 36.3 | <186 | 547 | < 17.0 (14.7 - 19.7) | 15.1 (13.8 - 16.4) |
| **PS-stratification / on-treatment TAR without escalation** | | | | | | | | |
| CCAE | 9,460 | 22,298 | 5.4 | 15.9 | 82 | 254 | 15.1 (12.0 - 18.7) | 16.0 (14.1 - 18.0) |
| ODOD | 7,151 | 16,194 | 3.6 | 9.8 | 91 | 186 | 25.3 (20.3 - 31.0) | 18.9 (16.3 - 21.8) |
| MDCR | 661 | 1,967 | 0.4 | 1.4 | 8 | 32 | 22.3 (9.61 - 43.9) | 23.1 (15.8 - 32.7) |
| Open Claims | 135,417 | 295,535 | 81.2 | 218.3 | 868 | 2,332 | 10.7 (9.99 - 11.4) | 10.7 (10.3 - 11.1) |
| OEHR | 8,094 | 18,979 | 2.0 | 5.3 | 21 | 62 | 10.7 (6.63 - 16.4) | 11.6 (8.90 - 14.9) |
| VA | 1,215 | 11,119 | 0.7 | 5.8 | <10 | 46 | < 15.1 (7.23 - 27.7) | 7.89 (5.78 - 10.5) |
| Meta-analysis | 160,122 | 353,006 | 92.2 | 249.4 | 1,062 | 2,834 | 11.5 (10.8 - 12.2) | 11.4 (10.9 - 11.8) |
| Meta-analysis (w/o USOC) | 24,705 | 57,471 | 11.0 | 31.1 | 194 | 502 | 17.6 (15.2 - 20.3) | 16.1 (14.8 - 17.6) |

##### 4.1.14 Diabetic ketoacidosis

**Supplementary Table 303:** Exposures, outcomes and risk estimates for Diabetic ketoacidosis in GLP1RA (male) (T) vs SGLT2I (male) (C) new-user cohorts under four different propensity score (PS) adjustment / time-at-risk (TAR) designs across data sources. We report total patient exposure time in 1,000 patient-year (KYr) units, uncalibrated and calibrated hazard ratios (HRs) and their 95% confidence intervals (CIs). Greyed out entries did not pass study diagnostics and are included only for completeness.

| | Patients | | Exposure (KYr) | | Outcomes | | Incidence (per KYr) | |
|---|---|---|---|---|---|---|---|---|
| | T | C | T | C | T | C | T | C |
| **PS-matching / on-treatment TAR** | | | | | | | | |
| CCAE | 9,693 | 24,104 | 6.6 | 20.1 | 8 | 40 | 1.21 (0.52 - 2.39) | 1.99 (1.42 - 2.71) |
| ODOD | 7,133 | 17,440 | 4.1 | 12.1 | <5 | 23 | < 1.21 (0.39 - 2.82) | 1.90 (1.20 - 2.85) |
| MDCR | 700 | 2,135 | 0.4 | 1.7 | 0 | <5 | | < 2.93 (0.95 - 6.84) |
| Open Claims | 144,918 | 332,168 | 101.8 | 289.1 | 68 | 307 | 0.67 (0.52 - 0.85) | 1.06 (0.95 - 1.19) |
| OEHR | 7,945 | 20,206 | 2.2 | 6.5 | <5 | 14 | < 2.24 (0.73 - 5.23) | 2.17 (1.19 - 3.64) |
| VA | 1,185 | 11,176 | 0.8 | 6.7 | <10 | <10 | < 12.1 (5.82 - 22.3) | < 1.48 (0.71 - 2.73) |
| Meta-analysis | 161,744 | 373,712 | 112.6 | 321.3 | <81 | 370 | < 0.72 (0.57 - 0.89) | 1.15 (1.04 - 1.28) |
| Meta-analysis (w/o USOC) | 16,826 | 41,544 | 10.7 | 32.2 | <13 | 63 | < 1.21 (0.65 - 2.07) | 1.96 (1.50 - 2.50) |
| **PS-stratification / on-treatment TAR** | | | | | | | | |
| CCAE | 10,325 | 24,108 | 6.9 | 20.1 | 8 | 40 | 1.16 (0.50 - 2.29) | 1.99 (1.42 - 2.71) |
| ODOD | 7,721 | 17,459 | 4.4 | 12.1 | <5 | 23 | < 1.13 (0.37 - 2.64) | 1.90 (1.20 - 2.85) |
| MDCR | 730 | 2,135 | 0.5 | 1.7 | 0 | <5 | | < 2.93 (0.95 - 6.84) |
| Open Claims | 155,077 | 332,197 | 106.8 | 289.1 | 70 | 307 | 0.66 (0.51 - 0.83) | 1.06 (0.95 - 1.19) |
| OEHR | 8,665 | 20,218 | 2.4 | 6.5 | <5 | 14 | < 2.08 (0.67 - 4.84) | 2.17 (1.19 - 3.64) |
| VA | 1,357 | 12,523 | 0.9 | 7.5 | <10 | <10 | < 10.6 (5.09 - 19.5) | < 1.33 (0.64 - 2.44) |
| Meta-analysis | 173,123 | 373,764 | 118.1 | 321.3 | <83 | 370 | < 0.70 (0.56 - 0.87) | 1.15 (1.04 - 1.27) |
| Meta-analysis (w/o USOC) | 18,046 | 41,567 | 11.3 | 32.2 | <13 | 63 | < 1.15 (0.61 - 1.97) | 1.96 (1.50 - 2.50) |
| **PS-matching / on-treatment TAR without escalation** | | | | | | | | |
| CCAE | 9,693 | 24,104 | 5.8 | 17.5 | 7 | 33 | 1.21 (0.49 - 2.50) | 1.88 (1.30 - 2.64) |
| ODOD | 7,133 | 17,440 | 3.7 | 10.7 | <5 | 20 | < 1.36 (0.44 - 3.16) | 1.86 (1.14 - 2.88) |
| MDCR | 700 | 2,135 | 0.4 | 1.5 | 0 | 0 | | |
| Open Claims | 144,918 | 332,168 | 88.8 | 245.8 | 57 | 243 | 0.64 (0.49 - 0.83) | 0.99 (0.87 - 1.12) |
| OEHR | 7,945 | 20,206 | 1.9 | 5.7 | <5 | 13 | < 2.57 (0.84 - 6.00) | 2.29 (1.22 - 3.91) |
| VA | 1,185 | 11,176 | 0.6 | 5.8 | <10 | <10 | < 15.5 (7.44 - 28.5) | < 1.72 (0.83 - 3.16) |
| Meta-analysis | 161,744 | 373,712 | 98.2 | 274.1 | <69 | 296 | < 0.70 (0.55 - 0.89) | 1.08 (0.96 - 1.21) |
| Meta-analysis (w/o USOC) | 16,826 | 41,544 | 9.5 | 28.3 | <12 | 53 | < 1.27 (0.66 - 2.22) | 1.87 (1.40 - 2.45) |
| **PS-stratification / on-treatment TAR without escalation** | | | | | | | | |
| CCAE | 10,325 | 24,108 | 6.0 | 17.5 | 7 | 33 | 1.16 (0.47 - 2.38) | 1.88 (1.29 - 2.64) |
| ODOD | 7,721 | 17,459 | 3.9 | 10.8 | <5 | 20 | < 1.27 (0.41 - 2.97) | 1.86 (1.14 - 2.87) |
| MDCR | 730 | 2,135 | 0.4 | 1.5 | 0 | 0 | | |
| Open Claims | 155,077 | 332,197 | 93.4 | 245.8 | 58 | 243 | 0.62 (0.47 - 0.80) | 0.99 (0.87 - 1.12) |
| OEHR | 8,665 | 20,218 | 2.1 | 5.7 | <5 | 13 | < 2.38 (0.77 - 5.55) | 2.29 (1.22 - 3.91) |
| VA | 1,357 | 12,523 | 0.7 | 6.5 | <10 | <10 | < 13.6 (6.50 - 24.9) | < 1.53 (0.74 - 2.82) |
| Meta-analysis | 173,123 | 373,764 | 103.3 | 274.1 | <70 | 296 | < 0.68 (0.53 - 0.86) | 1.08 (0.96 - 1.21) |
| Meta-analysis (w/o USOC) | 18,046 | 41,567 | 10.0 | 28.3 | <12 | 53 | < 1.20 (0.62 - 2.10) | 1.87 (1.40 - 2.45) |

##### 4.1.15 Diarrhea

**Supplementary Table 304:** Exposures, outcomes and risk estimates for Diarrhea in GLP1RA (male) (T) vs SGLT2I (male) (C) new-user cohorts under four different propensity score (PS) adjustment / time-at-risk (TAR) designs across data sources. We report total patient exposure time in 1,000 patient-year (KYr) units, uncalibrated and calibrated hazard ratios (HRs) and their 95% confidence intervals (CIs). Greyed out entries did not pass study diagnostics and are included only for completeness.

| | Patients | | Exposure (KYr) | | Outcomes | | Incidence (per KYr) | |
|---|---|---|---|---|---|---|---|---|
| | T | C | T | C | T | C | T | C |
| **PS-matching / on-treatment TAR** | | | | | | | | |
| CCAE | 8,625 | 21,752 | 5.7 | 17.8 | 223 | 367 | 38.9 (34.0 - 44.4) | 20.6 (18.5 - 22.8) |
| ODOD | 6,363 | 15,584 | 3.6 | 10.6 | 163 | 284 | 45.2 (38.5 - 52.7) | 26.7 (23.7 - 30.0) |
| MDCR | 614 | 1,906 | 0.4 | 1.5 | 10 | 30 | 26.4 (12.7 - 48.6) | 20.1 (13.5 - 28.7) |
| Open Claims | 128,629 | 299,039 | 89.5 | 258.8 | 1,598 | 2,998 | 17.9 (17.0 - 18.8) | 11.6 (11.2 - 12.0) |
| OEHR | 7,231 | 18,494 | 2.0 | 5.8 | 81 | 134 | 40.6 (32.3 - 50.5) | 22.9 (19.2 - 27.2) |
| VA | 1,003 | 9,517 | 0.7 | 5.7 | 16 | 97 | 23.0 (13.1 - 37.3) | 17.0 (13.8 - 20.7) |
| Meta-analysis | 151,851 | 364,386 | 101.5 | 298.8 | 2,081 | 3,880 | 20.5 (19.6 - 21.4) | 13.0 (12.6 - 13.4) |
| Meta-analysis (w/o USOC) | 23,222 | 65,347 | 12.0 | 40.0 | 483 | 882 | 40.2 (36.7 - 43.9) | 22.0 (20.6 - 23.5) |
| **PS-stratification / on-treatment TAR** | | | | | | | | |
| CCAE | 9,160 | 21,758 | 6.0 | 17.9 | 243 | 367 | 40.6 (35.7 - 46.1) | 20.6 (18.5 - 22.8) |
| ODOD | 6,875 | 15,599 | 3.8 | 10.6 | 176 | 286 | 45.8 (39.3 - 53.1) | 26.9 (23.8 - 30.2) |
| MDCR | 649 | 1,906 | 0.4 | 1.5 | 10 | 30 | 25.1 (12.0 - 46.2) | 20.1 (13.5 - 28.7) |
| Open Claims | 137,191 | 299,156 | 93.6 | 258.9 | 1,776 | 3,000 | 19.0 (18.1 - 19.9) | 11.6 (11.2 - 12.0) |
| OEHR | 7,828 | 18,506 | 2.1 | 5.8 | 89 | 134 | 41.7 (33.5 - 51.3) | 22.9 (19.2 - 27.1) |
| VA | 1,146 | 10,635 | 0.8 | 6.4 | 21 | 107 | 26.7 (16.6 - 40.9) | 16.7 (13.7 - 20.2) |
| Meta-analysis | 161,054 | 355,019 | 105.6 | 293.2 | 2,284 | 3,787 | 21.6 (20.8 - 22.5) | 12.9 (12.5 - 13.3) |
| Meta-analysis (w/o USOC) | 23,863 | 55,863 | 12.0 | 34.3 | 508 | 787 | 42.5 (38.9 - 46.4) | 22.9 (21.3 - 24.6) |
| **PS-matching / on-treatment TAR without escalation** | | | | | | | | |
| CCAE | 8,625 | 21,752 | 5.0 | 15.6 | 200 | 307 | 39.7 (34.4 - 45.6) | 19.6 (17.5 - 21.9) |
| ODOD | 6,363 | 15,584 | 3.2 | 9.5 | 143 | 242 | 44.4 (37.4 - 52.3) | 25.5 (22.4 - 28.9) |
| MDCR | 614 | 1,906 | 0.3 | 1.3 | 9 | 27 | 26.3 (12.0 - 50.0) | 20.0 (13.2 - 29.2) |
| Open Claims | 128,629 | 299,039 | 78.2 | 220.9 | 1,385 | 2,471 | 17.7 (16.8 - 18.7) | 11.2 (10.7 - 11.6) |
| OEHR | 7,231 | 18,494 | 1.7 | 5.2 | 68 | 117 | 39.0 (30.3 - 49.5) | 22.7 (18.8 - 27.2) |
| VA | 1,003 | 9,517 | 0.5 | 4.9 | 13 | 87 | 23.9 (12.7 - 40.8) | 17.7 (14.2 - 21.8) |
| Meta-analysis | 151,851 | 364,386 | 88.7 | 256.1 | 1,809 | 3,224 | 20.4 (19.5 - 21.3) | 12.6 (12.2 - 13.0) |
| Meta-analysis (w/o USOC) | 23,222 | 65,347 | 10.5 | 35.2 | 424 | 753 | 40.2 (36.5 - 44.2) | 21.4 (19.9 - 23.0) |
| **PS-stratification / on-treatment TAR without escalation** | | | | | | | | |
| CCAE | 9,160 | 21,758 | 5.3 | 15.7 | 217 | 307 | 41.2 (35.9 - 47.1) | 19.6 (17.5 - 21.9) |
| ODOD | 6,875 | 15,599 | 3.4 | 9.5 | 155 | 244 | 45.1 (38.3 - 52.8) | 25.7 (22.6 - 29.1) |
| MDCR | 649 | 1,906 | 0.4 | 1.3 | 9 | 27 | 24.9 (11.4 - 47.3) | 20.0 (13.2 - 29.2) |
| Open Claims | 137,191 | 299,156 | 82.0 | 221.0 | 1,549 | 2,473 | 18.9 (18.0 - 19.9) | 11.2 (10.8 - 11.6) |
| OEHR | 7,828 | 18,506 | 1.9 | 5.2 | 76 | 117 | 40.6 (32.0 - 50.8) | 22.7 (18.8 - 27.2) |
| VA | 1,146 | 10,635 | 0.6 | 5.5 | 18 | 95 | 29.5 (17.5 - 46.6) | 17.2 (13.9 - 21.0) |
| Meta-analysis | 161,054 | 355,019 | 92.6 | 251.3 | 1,997 | 3,141 | 21.6 (20.6 - 22.5) | 12.5 (12.1 - 12.9) |
| Meta-analysis (w/o USOC) | 23,863 | 55,863 | 10.6 | 30.3 | 448 | 668 | 42.4 (38.5 - 46.5) | 22.0 (20.4 - 23.8) |

##### 4.1.16 Genitourinary infection

**Supplementary Table 305:** Exposures, outcomes and risk estimates for Genitourinary infection in GLP1RA (male) (T) vs SGLT2I (male) (C) new-user cohorts under four different propensity score (PS) adjustment / time-at-risk (TAR) designs across data sources. We report total patient exposure time in 1,000 patient-year (KYr) units, uncalibrated and calibrated hazard ratios (HRs) and their 95% confidence intervals (CIs). Greyed out entries did not pass study diagnostics and are included only for completeness.

| | Patients | | Exposure (KYr) | | Outcomes | | Incidence (per KYr) | |
|---|---|---|---|---|---|---|---|---|
| | T | C | T | C | T | C | T | C |
| **PS-matching / on-treatment TAR** | | | | | | | | |
| CCAE | 8,832 | 22,098 | 5.9 | 18.2 | 111 | 274 | 18.9 (15.5 - 22.7) | 15.0 (13.3 - 16.9) |
| ODOD | 6,369 | 15,688 | 3.6 | 10.8 | 105 | 287 | 29.0 (23.7 - 35.1) | 26.6 (23.6 - 29.9) |
| MDCR | 584 | 1,870 | 0.3 | 1.5 | 18 | 43 | 52.3 (31.0 - 82.7) | 29.5 (21.3 - 39.7) |
| Open Claims | 129,162 | 296,972 | 90.6 | 259.6 | 1,185 | 2,991 | 13.1 (12.3 - 13.9) | 11.5 (11.1 - 11.9) |
| OEHR | 7,520 | 19,160 | 2.1 | 6.1 | 41 | 109 | 19.5 (14.0 - 26.5) | 17.9 (14.7 - 21.6) |
| VA | 1,061 | 10,108 | 0.7 | 6.1 | 14 | 59 | 18.8 (10.3 - 31.6) | 9.63 (7.33 - 12.4) |
| Meta-analysis | 152,944 | 364,026 | 102.9 | 300.8 | 1,456 | 3,720 | 14.1 (13.4 - 14.9) | 12.4 (12.0 - 12.8) |
| Meta-analysis (w/o USOC) | 23,782 | 67,054 | 12.4 | 41.2 | 271 | 729 | 21.9 (19.4 - 24.7) | 17.7 (16.4 - 19.0) |
| **PS-stratification / on-treatment TAR** | | | | | | | | |
| CCAE | 9,362 | 22,104 | 6.1 | 18.3 | 115 | 274 | 18.8 (15.5 - 22.5) | 15.0 (13.3 - 16.9) |
| ODOD | 6,866 | 15,706 | 3.9 | 10.8 | 114 | 287 | 29.5 (24.4 - 35.5) | 26.6 (23.6 - 29.8) |
| MDCR | 621 | 1,870 | 0.4 | 1.5 | 18 | 43 | 48.5 (28.8 - 76.7) | 29.5 (21.3 - 39.7) |
| Open Claims | 137,954 | 297,034 | 94.9 | 259.6 | 1,276 | 2,993 | 13.4 (12.7 - 14.2) | 11.5 (11.1 - 11.9) |
| OEHR | 8,164 | 19,171 | 2.3 | 6.1 | 45 | 109 | 20.0 (14.6 - 26.7) | 17.9 (14.7 - 21.6) |
| VA | 1,210 | 11,310 | 0.8 | 6.8 | 14 | 68 | 16.5 (9.04 - 27.7) | 9.93 (7.71 - 12.6) |
| Meta-analysis | 162,346 | 354,015 | 107.1 | 294.8 | 1,550 | 3,663 | 14.5 (13.8 - 15.2) | 12.4 (12.0 - 12.8) |
| Meta-analysis (w/o USOC) | 24,392 | 56,981 | 12.2 | 35.1 | 274 | 670 | 22.4 (19.8 - 25.2) | 19.1 (17.7 - 20.6) |
| **PS-matching / on-treatment TAR without escalation** | | | | | | | | |
| CCAE | 8,832 | 22,098 | 5.2 | 16.0 | 95 | 233 | 18.4 (14.9 - 22.5) | 14.6 (12.8 - 16.6) |
| ODOD | 6,369 | 15,688 | 3.2 | 9.6 | 94 | 248 | 29.0 (23.4 - 35.5) | 25.9 (22.8 - 29.3) |
| MDCR | 584 | 1,870 | 0.3 | 1.3 | 16 | 40 | 52.4 (30.0 - 85.1) | 30.4 (21.7 - 41.4) |
| Open Claims | 129,162 | 296,972 | 79.0 | 220.9 | 1,033 | 2,548 | 13.1 (12.3 - 13.9) | 11.5 (11.1 - 12.0) |
| OEHR | 7,520 | 19,160 | 1.8 | 5.4 | 37 | 95 | 20.2 (14.3 - 27.9) | 17.7 (14.4 - 21.7) |
| VA | 1,061 | 10,108 | 0.6 | 5.3 | 11 | 53 | 19.1 (9.52 - 34.1) | 10.1 (7.53 - 13.1) |
| Meta-analysis | 152,944 | 364,026 | 89.9 | 257.1 | 1,270 | 3,177 | 14.1 (13.4 - 14.9) | 12.4 (11.9 - 12.8) |
| Meta-analysis (w/o USOC) | 23,782 | 67,054 | 10.8 | 36.2 | 237 | 629 | 21.9 (19.2 - 24.9) | 17.4 (16.0 - 18.8) |
| **PS-stratification / on-treatment TAR without escalation** | | | | | | | | |
| CCAE | 9,362 | 22,104 | 5.4 | 16.0 | 98 | 233 | 18.2 (14.8 - 22.1) | 14.6 (12.8 - 16.6) |
| ODOD | 6,866 | 15,706 | 3.5 | 9.6 | 103 | 248 | 29.8 (24.4 - 36.2) | 25.8 (22.7 - 29.3) |
| MDCR | 621 | 1,870 | 0.3 | 1.3 | 16 | 40 | 48.2 (27.5 - 78.2) | 30.4 (21.7 - 41.4) |
| Open Claims | 137,954 | 297,034 | 83.0 | 220.9 | 1,114 | 2,550 | 13.4 (12.6 - 14.2) | 11.5 (11.1 - 12.0) |
| OEHR | 8,164 | 19,171 | 2.0 | 5.4 | 41 | 95 | 20.8 (15.0 - 28.3) | 17.7 (14.3 - 21.7) |
| VA | 1,210 | 11,310 | 0.7 | 5.9 | 11 | 59 | 16.8 (8.37 - 30.0) | 9.97 (7.59 - 12.9) |
| Meta-analysis | 162,346 | 354,015 | 93.8 | 251.9 | 1,356 | 3,126 | 14.5 (13.7 - 15.2) | 12.4 (12.0 - 12.9) |
| Meta-analysis (w/o USOC) | 24,392 | 56,981 | 10.8 | 30.9 | 242 | 576 | 22.4 (19.7 - 25.4) | 18.6 (17.1 - 20.2) |

##### 4.1.17 Hyperkalemia

**Supplementary Table 306:** Exposures, outcomes and risk estimates for Hyperkalemia in GLP1RA (male) (T) vs SGLT2I (male) (C) new-user cohorts under four different propensity score (PS) adjustment / time-at-risk (TAR) designs across data sources. We report total patient exposure time in 1,000 patient-year (KYr) units, uncalibrated and calibrated hazard ratios (HRs) and their 95% confidence intervals (CIs). Greyed out entries did not pass study diagnostics and are included only for completeness.

| | Patients | | Exposure (KYr) | | Outcomes | | Incidence (per KYr) | |
|---|---|---|---|---|---|---|---|---|
| | T | C | T | C | T | C | T | C |
| **PS-matching / on-treatment TAR** | | | | | | | | |
| CCAE | 9,537 | 23,687 | 6.4 | 19.6 | 47 | 134 | 7.31 (5.37 - 9.72) | 6.84 (5.73 - 8.11) |
| ODOD | 6,873 | 16,781 | 3.9 | 11.6 | 62 | 191 | 15.7 (12.0 - 20.1) | 16.5 (14.3 - 19.0) |
| MDCR | 683 | 2,069 | 0.4 | 1.7 | 5 | 20 | 11.3 (3.67 - 26.4) | 12.1 (7.36 - 18.6) |
| Open Claims | 142,419 | 325,969 | 99.9 | 283.4 | 375 | 1,288 | 3.76 (3.38 - 4.16) | 4.54 (4.30 - 4.80) |
| OEHR | 7,810 | 19,852 | 2.2 | 6.3 | 16 | 62 | 7.30 (4.17 - 11.9) | 9.86 (7.56 - 12.6) |
| VA | 1,105 | 10,243 | 0.8 | 6.1 | <10 | 82 | < 12.9 (6.19 - 23.7) | 13.3 (10.6 - 16.6) |
| Meta-analysis | 167,744 | 396,532 | 113.2 | 327.0 | <510 | 1,757 | < 4.51 (4.12 - 4.91) | 5.37 (5.12 - 5.63) |
| Meta-analysis (w/o USOC) | 25,325 | 70,563 | 13.3 | 43.6 | <135 | 469 | < 10.1 (8.49 - 12.0) | 10.8 (9.81 - 11.8) |
| **PS-stratification / on-treatment TAR** | | | | | | | | |
| CCAE | 10,165 | 23,691 | 6.7 | 19.6 | 48 | 134 | 7.13 (5.26 - 9.45) | 6.84 (5.73 - 8.10) |
| ODOD | 7,438 | 16,805 | 4.2 | 11.6 | 66 | 192 | 15.7 (12.1 - 19.9) | 16.6 (14.3 - 19.1) |
| MDCR | 714 | 2,069 | 0.5 | 1.7 | 5 | 20 | 10.9 (3.56 - 25.6) | 12.1 (7.36 - 18.6) |
| Open Claims | 152,533 | 325,992 | 104.8 | 283.5 | 393 | 1,288 | 3.75 (3.39 - 4.14) | 4.54 (4.30 - 4.80) |
| OEHR | 8,502 | 19,863 | 2.4 | 6.3 | 18 | 62 | 7.63 (4.52 - 12.1) | 9.85 (7.55 - 12.6) |
| VA | 1,266 | 11,422 | 0.9 | 6.9 | <10 | 104 | < 11.4 (5.44 - 20.9) | 15.2 (12.4 - 18.4) |
| Meta-analysis | 178,638 | 386,351 | 118.1 | 320.9 | 525 | 1,676 | 4.45 (4.07 - 4.84) | 5.22 (4.98 - 5.48) |
| Meta-analysis (w/o USOC) | 26,105 | 60,359 | 13.3 | 37.5 | 132 | 388 | 9.92 (8.30 - 11.8) | 10.4 (9.35 - 11.4) |
| **PS-matching / on-treatment TAR without escalation** | | | | | | | | |
| CCAE | 9,537 | 23,687 | 5.6 | 17.1 | 36 | 109 | 6.38 (4.47 - 8.83) | 6.36 (5.22 - 7.68) |
| ODOD | 6,873 | 16,781 | 3.5 | 10.3 | 54 | 172 | 15.3 (11.5 - 20.0) | 16.7 (14.3 - 19.4) |
| MDCR | 683 | 2,069 | 0.4 | 1.5 | <5 | 19 | < 12.6 (4.08 - 29.4) | 12.8 (7.72 - 20.0) |
| Open Claims | 142,419 | 325,969 | 87.1 | 241.1 | 317 | 1,086 | 3.64 (3.25 - 4.06) | 4.50 (4.24 - 4.78) |
| OEHR | 7,810 | 19,852 | 1.9 | 5.6 | 12 | 47 | 6.29 (3.25 - 11.0) | 8.47 (6.22 - 11.3) |
| VA | 1,105 | 10,243 | 0.6 | 5.3 | <10 | 71 | < 16.6 (7.96 - 30.5) | 13.4 (10.4 - 16.9) |
| Meta-analysis | 167,744 | 396,532 | 98.7 | 279.3 | <429 | 1,485 | < 4.34 (3.94 - 4.78) | 5.32 (5.05 - 5.59) |
| Meta-analysis (w/o USOC) | 25,325 | 70,563 | 11.7 | 38.3 | <112 | 399 | < 9.59 (7.90 - 11.5) | 10.4 (9.43 - 11.5) |
| **PS-stratification / on-treatment TAR without escalation** | | | | | | | | |
| CCAE | 10,165 | 23,691 | 5.9 | 17.1 | 37 | 109 | 6.25 (4.40 - 8.61) | 6.36 (5.22 - 7.67) |
| ODOD | 7,438 | 16,805 | 3.8 | 10.3 | 58 | 173 | 15.4 (11.7 - 19.9) | 16.8 (14.4 - 19.5) |
| MDCR | 714 | 2,069 | 0.4 | 1.5 | <5 | 19 | < 12.2 (3.95 - 28.4) | 12.8 (7.72 - 20.0) |
| Open Claims | 152,533 | 325,992 | 91.6 | 241.1 | 333 | 1,086 | 3.63 (3.25 - 4.05) | 4.50 (4.24 - 4.78) |
| OEHR | 8,502 | 19,863 | 2.1 | 5.6 | 13 | 47 | 6.31 (3.36 - 10.8) | 8.46 (6.22 - 11.3) |
| VA | 1,266 | 11,422 | 0.7 | 6.0 | <10 | 89 | < 14.6 (6.99 - 26.8) | 15.0 (12.0 - 18.4) |
| Meta-analysis | 178,638 | 386,351 | 103.4 | 274.1 | 441 | 1,415 | 4.27 (3.88 - 4.68) | 5.16 (4.90 - 5.44) |
| Meta-analysis (w/o USOC) | 26,105 | 60,359 | 11.7 | 33.0 | 108 | 329 | 9.20 (7.54 - 11.1) | 9.97 (8.92 - 11.1) |

##### 4.1.18 Hypoglycemia

**Supplementary Table 307:** Exposures, outcomes and risk estimates for Hypoglycemia in GLP1RA (male) (T) vs SGLT2I (male) (C) new-user cohorts under four different propensity score (PS) adjustment / time-at-risk (TAR) designs across data sources. We report total patient exposure time in 1,000 patient-year (KYr) units, uncalibrated and calibrated hazard ratios (HRs) and their 95% confidence intervals (CIs). Greyed out entries did not pass study diagnostics and are included only for completeness.

| | Patients | | Exposure (KYr) | | Outcomes | | Incidence (per KYr) | |
|---|---|---|---|---|---|---|---|---|
| | T | C | T | C | T | C | T | C |
| **PS-matching / on-treatment TAR** | | | | | | | | |
| CCAE | 9,531 | 23,829 | 6.5 | 19.8 | 27 | 49 | 4.18 (2.75 - 6.08) | 2.47 (1.83 - 3.27) |
| ODOD | 7,046 | 17,242 | 4.1 | 12.0 | 21 | 51 | 5.15 (3.19 - 7.87) | 4.26 (3.17 - 5.61) |
| MDCR | 694 | 2,115 | 0.4 | 1.7 | <5 | 5 | < 11.3 (3.66 - 26.3) | 2.97 (0.96 - 6.92) |
| Open Claims | 143,099 | 328,434 | 100.5 | 285.7 | 193 | 421 | 1.92 (1.66 - 2.21) | 1.47 (1.34 - 1.62) |
| OEHR | 7,860 | 20,051 | 2.2 | 6.4 | 11 | 18 | 4.98 (2.49 - 8.91) | 2.81 (1.66 - 4.44) |
| VA | 1,170 | 11,030 | 0.8 | 6.6 | 0 | 12 | | 1.81 (0.93 - 3.15) |
| Meta-analysis | 167,536 | 389,556 | 113.2 | 323.9 | 252 | 539 | 2.23 (1.96 - 2.52) | 1.66 (1.53 - 1.81) |
| Meta-analysis (w/o USOC) | 24,437 | 61,122 | 12.8 | 38.2 | 59 | 118 | 4.63 (3.52 - 5.97) | 3.09 (2.56 - 3.70) |
| **PS-stratification / on-treatment TAR** | | | | | | | | |
| CCAE | 10,147 | 23,833 | 6.8 | 19.8 | 27 | 49 | 3.99 (2.63 - 5.81) | 2.47 (1.83 - 3.27) |
| ODOD | 7,628 | 17,261 | 4.4 | 12.0 | 24 | 51 | 5.52 (3.53 - 8.21) | 4.26 (3.17 - 5.60) |
| MDCR | 725 | 2,115 | 0.5 | 1.7 | <5 | 5 | < 10.8 (3.52 - 25.3) | 2.97 (0.96 - 6.92) |
| Open Claims | 152,956 | 328,462 | 105.3 | 285.7 | 219 | 421 | 2.08 (1.81 - 2.37) | 1.47 (1.34 - 1.62) |
| OEHR | 8,553 | 20,063 | 2.4 | 6.4 | 15 | 18 | 6.31 (3.53 - 10.4) | 2.81 (1.66 - 4.43) |
| VA | 1,336 | 12,356 | 0.9 | 7.4 | <10 | 14 | < 10.8 (5.18 - 19.9) | 1.88 (1.03 - 3.16) |
| Meta-analysis | 179,284 | 389,619 | 118.8 | 323.9 | 285 | 539 | 2.40 (2.13 - 2.69) | 1.66 (1.53 - 1.81) |
| Meta-analysis (w/o USOC) | 26,328 | 61,157 | 13.5 | 38.2 | 66 | 118 | 4.89 (3.78 - 6.22) | 3.09 (2.56 - 3.70) |
| **PS-matching / on-treatment TAR without escalation** | | | | | | | | |
| CCAE | 9,531 | 23,829 | 5.7 | 17.3 | 20 | 38 | 3.53 (2.16 - 5.46) | 2.20 (1.55 - 3.01) |
| ODOD | 7,046 | 17,242 | 3.6 | 10.6 | 15 | 36 | 4.12 (2.31 - 6.79) | 3.39 (2.37 - 4.69) |
| MDCR | 694 | 2,115 | 0.4 | 1.5 | 0 | <5 | | < 3.31 (1.07 - 7.72) |
| Open Claims | 143,099 | 328,434 | 87.6 | 243.0 | 149 | 289 | 1.70 (1.44 - 2.00) | 1.19 (1.06 - 1.33) |
| OEHR | 7,860 | 20,051 | 1.9 | 5.6 | 10 | 12 | 5.20 (2.50 - 9.57) | 2.13 (1.10 - 3.71) |
| VA | 1,170 | 11,030 | 0.6 | 5.7 | 0 | <10 | | < 1.75 (0.84 - 3.21) |
| Meta-analysis | 167,536 | 389,556 | 98.8 | 276.6 | 194 | 375 | 1.96 (1.70 - 2.26) | 1.36 (1.22 - 1.50) |
| Meta-analysis (w/o USOC) | 24,437 | 61,122 | 11.2 | 33.6 | 45 | 86 | 4.01 (2.92 - 5.36) | 2.56 (2.05 - 3.16) |
| **PS-stratification / on-treatment TAR without escalation** | | | | | | | | |
| CCAE | 10,147 | 23,833 | 5.9 | 17.3 | 20 | 38 | 3.37 (2.06 - 5.20) | 2.19 (1.55 - 3.01) |
| ODOD | 7,628 | 17,261 | 3.9 | 10.6 | 18 | 36 | 4.63 (2.75 - 7.32) | 3.39 (2.37 - 4.69) |
| MDCR | 725 | 2,115 | 0.4 | 1.5 | 0 | <5 | | < 3.31 (1.07 - 7.72) |
| Open Claims | 152,956 | 328,462 | 92.0 | 243.0 | 173 | 289 | 1.88 (1.61 - 2.18) | 1.19 (1.06 - 1.33) |
| OEHR | 8,553 | 20,063 | 2.1 | 5.6 | 12 | 12 | 5.79 (2.99 - 10.1) | 2.12 (1.10 - 3.71) |
| VA | 1,336 | 12,356 | 0.7 | 6.4 | 0 | <10 | | < 1.56 (0.75 - 2.86) |
| Meta-analysis | 179,284 | 389,619 | 103.9 | 276.6 | 223 | 375 | 2.15 (1.87 - 2.45) | 1.36 (1.22 - 1.50) |
| Meta-analysis (w/o USOC) | 26,328 | 61,157 | 11.9 | 33.6 | 50 | 86 | 4.20 (3.12 - 5.54) | 2.56 (2.05 - 3.16) |

##### 4.1.19 Hypotension

**Supplementary Table 308:** Exposures, outcomes and risk estimates for Hypotension in GLP1RA (male) (T) vs SGLT2I (male) (C) new-user cohorts under four different propensity score (PS) adjustment / time-at-risk (TAR) designs across data sources. We report total patient exposure time in 1,000 patient-year (KYr) units, uncalibrated and calibrated hazard ratios (HRs) and their 95% confidence intervals (CIs). Greyed out entries did not pass study diagnostics and are included only for completeness.

| | Patients | | Exposure (KYr) | | Outcomes | | Incidence (per KYr) | |
|---|---|---|---|---|---|---|---|---|
| | T | C | T | C | T | C | T | C |
| **PS-matching / on-treatment TAR** | | | | | | | | |
| CCAE | 9,511 | 23,650 | 6.4 | 19.6 | 59 | 175 | 9.18 (6.99 - 11.8) | 8.92 (7.65 - 10.3) |
| ODOD | 6,853 | 16,703 | 4.0 | 11.5 | 66 | 229 | 16.7 (12.9 - 21.2) | 19.8 (17.3 - 22.6) |
| MDCR | 665 | 2,017 | 0.4 | 1.6 | 11 | 35 | 25.8 (12.9 - 46.1) | 21.8 (15.2 - 30.3) |
| Open Claims | 140,648 | 321,358 | 98.7 | 279.9 | 585 | 1,986 | 5.93 (5.46 - 6.43) | 7.10 (6.79 - 7.41) |
| OEHR | 7,755 | 19,795 | 2.2 | 6.3 | 30 | 83 | 13.8 (9.32 - 19.7) | 13.2 (10.5 - 16.4) |
| VA | 1,105 | 10,372 | 0.8 | 6.2 | 12 | 129 | 15.5 (8.03 - 27.2) | 20.7 (17.3 - 24.6) |
| Meta-analysis | 165,872 | 391,878 | 112.0 | 323.6 | 752 | 2,602 | 6.71 (6.24 - 7.21) | 8.04 (7.74 - 8.36) |
| Meta-analysis (w/o USOC) | 25,224 | 70,520 | 13.3 | 43.7 | 167 | 616 | 12.5 (10.7 - 14.6) | 14.1 (13.0 - 15.3) |
| **PS-stratification / on-treatment TAR** | | | | | | | | |
| CCAE | 10,114 | 23,656 | 6.7 | 19.6 | 59 | 177 | 8.78 (6.69 - 11.3) | 9.02 (7.74 - 10.4) |
| ODOD | 7,418 | 16,736 | 4.2 | 11.6 | 71 | 229 | 16.8 (13.1 - 21.2) | 19.8 (17.3 - 22.5) |
| MDCR | 697 | 2,017 | 0.4 | 1.6 | 11 | 35 | 24.8 (12.4 - 44.5) | 21.8 (15.2 - 30.3) |
| Open Claims | 150,554 | 321,381 | 103.5 | 279.9 | 624 | 1,989 | 6.03 (5.56 - 6.52) | 7.11 (6.80 - 7.43) |
| OEHR | 8,467 | 19,804 | 2.3 | 6.3 | 32 | 83 | 13.7 (9.34 - 19.3) | 13.2 (10.5 - 16.3) |
| VA | 1,263 | 11,420 | 0.9 | 6.9 | 18 | 152 | 20.4 (12.1 - 32.3) | 22.2 (18.8 - 26.0) |
| Meta-analysis | 176,553 | 381,577 | 116.8 | 317.4 | 786 | 2,478 | 6.73 (6.27 - 7.22) | 7.81 (7.50 - 8.12) |
| Meta-analysis (w/o USOC) | 25,999 | 60,196 | 13.3 | 37.5 | 162 | 489 | 12.2 (10.4 - 14.2) | 13.0 (11.9 - 14.2) |
| **PS-matching / on-treatment TAR without escalation** | | | | | | | | |
| CCAE | 9,511 | 23,650 | 5.6 | 17.2 | 48 | 144 | 8.54 (6.30 - 11.3) | 8.39 (7.08 - 9.88) |
| ODOD | 6,853 | 16,703 | 3.5 | 10.2 | 57 | 204 | 16.2 (12.2 - 20.9) | 19.9 (17.3 - 22.9) |
| MDCR | 665 | 2,017 | 0.4 | 1.4 | 9 | 29 | 23.4 (10.7 - 44.4) | 20.2 (13.5 - 29.0) |
| Open Claims | 140,648 | 321,358 | 86.1 | 238.2 | 497 | 1,664 | 5.77 (5.28 - 6.31) | 6.99 (6.65 - 7.33) |
| OEHR | 7,755 | 19,795 | 1.9 | 5.5 | 30 | 65 | 15.9 (10.7 - 22.7) | 11.7 (9.04 - 14.9) |
| VA | 1,105 | 10,372 | 0.6 | 5.4 | 10 | 110 | 16.8 (8.04 - 30.8) | 20.5 (16.8 - 24.6) |
| Meta-analysis | 165,872 | 391,878 | 97.7 | 276.5 | 642 | 2,187 | 6.57 (6.07 - 7.10) | 7.91 (7.58 - 8.25) |
| Meta-analysis (w/o USOC) | 25,224 | 70,520 | 11.6 | 38.3 | 145 | 523 | 12.5 (10.5 - 14.7) | 13.6 (12.5 - 14.9) |
| **PS-stratification / on-treatment TAR without escalation** | | | | | | | | |
| CCAE | 10,114 | 23,656 | 5.9 | 17.2 | 48 | 146 | 8.15 (6.01 - 10.8) | 8.51 (7.18 - 10.0) |
| ODOD | 7,418 | 16,736 | 3.8 | 10.3 | 62 | 204 | 16.5 (12.6 - 21.1) | 19.9 (17.2 - 22.8) |
| MDCR | 697 | 2,017 | 0.4 | 1.4 | 9 | 29 | 22.5 (10.3 - 42.8) | 20.2 (13.5 - 29.0) |
| Open Claims | 150,554 | 321,381 | 90.5 | 238.2 | 534 | 1,667 | 5.90 (5.41 - 6.42) | 7.00 (6.67 - 7.34) |
| OEHR | 8,467 | 19,804 | 2.0 | 5.5 | 32 | 65 | 15.6 (10.7 - 22.1) | 11.7 (9.04 - 14.9) |
| VA | 1,263 | 11,420 | 0.7 | 5.9 | 15 | 129 | 22.0 (12.3 - 36.3) | 21.7 (18.1 - 25.8) |
| Meta-analysis | 176,553 | 381,577 | 102.2 | 271.2 | 676 | 2,082 | 6.62 (6.13 - 7.13) | 7.68 (7.35 - 8.01) |
| Meta-analysis (w/o USOC) | 25,999 | 60,196 | 11.7 | 33.0 | 142 | 415 | 12.1 (10.2 - 14.3) | 12.6 (11.4 - 13.9) |

##### 4.1.20 Joint pain

**Supplementary Table 309:** Exposures, outcomes and risk estimates for Joint pain in GLP1RA (male) (T) vs SGLT2I (male) (C) new-user cohorts under four different propensity score (PS) adjustment / time-at-risk (TAR) designs across data sources. We report total patient exposure time in 1,000 patient-year (KYr) units, uncalibrated and calibrated hazard ratios (HRs) and their 95% confidence intervals (CIs). Greyed out entries did not pass study diagnostics and are included only for completeness.

| | Patients | | Exposure (KYr) | | Outcomes | | Incidence (per KYr) | |
|---|---|---|---|---|---|---|---|---|
| | T | C | T | C | T | C | T | C |
| **PS-matching / on-treatment TAR** | | | | | | | | |
| CCAE | 9,238 | 23,071 | 6.2 | 19.1 | 49 | 137 | 7.90 (5.84 - 10.4) | 7.16 (6.01 - 8.47) |
| ODOD | 6,735 | 16,669 | 3.9 | 11.5 | 31 | 103 | 8.00 (5.43 - 11.4) | 8.92 (7.28 - 10.8) |
| MDCR | 674 | 2,062 | 0.4 | 1.6 | <5 | 11 | < 11.6 (3.76 - 27.0) | 6.70 (3.34 - 12.0) |
| Open Claims | 137,603 | 317,206 | 96.5 | 276.1 | 444 | 1,151 | 4.60 (4.18 - 5.05) | 4.17 (3.93 - 4.42) |
| OEHR | 7,589 | 19,447 | 2.1 | 6.2 | 14 | 51 | 6.61 (3.61 - 11.1) | 8.28 (6.16 - 10.9) |
| VA | 1,046 | 9,692 | 0.7 | 5.9 | <10 | 39 | < 13.7 (6.56 - 25.2) | 6.64 (4.72 - 9.08) |
| Meta-analysis | 162,211 | 386,085 | 109.5 | 318.8 | <548 | 1,481 | < 5.01 (4.60 - 5.44) | 4.64 (4.41 - 4.89) |
| Meta-analysis (w/o USOC) | 24,608 | 68,879 | 12.9 | 42.7 | <104 | 330 | < 8.04 (6.57 - 9.75) | 7.73 (6.92 - 8.61) |
| **PS-stratification / on-treatment TAR** | | | | | | | | |
| CCAE | 9,816 | 23,075 | 6.5 | 19.1 | 52 | 137 | 8.01 (5.98 - 10.5) | 7.16 (6.01 - 8.46) |
| ODOD | 7,306 | 16,688 | 4.2 | 11.6 | 38 | 103 | 9.15 (6.48 - 12.6) | 8.91 (7.27 - 10.8) |
| MDCR | 699 | 2,062 | 0.4 | 1.6 | <5 | 11 | < 11.3 (3.65 - 26.3) | 6.70 (3.34 - 12.0) |
| Open Claims | 146,963 | 317,231 | 101.2 | 276.1 | 492 | 1,151 | 4.86 (4.44 - 5.31) | 4.17 (3.93 - 4.42) |
| OEHR | 8,249 | 19,457 | 2.3 | 6.2 | 16 | 51 | 7.00 (4.00 - 11.4) | 8.28 (6.16 - 10.9) |
| VA | 1,201 | 10,977 | 0.8 | 6.6 | 11 | 46 | 13.2 (6.58 - 23.6) | 6.93 (5.08 - 9.25) |
| Meta-analysis | 172,334 | 376,451 | 114.1 | 313.0 | 598 | 1,442 | 5.24 (4.83 - 5.68) | 4.61 (4.37 - 4.85) |
| Meta-analysis (w/o USOC) | 25,371 | 59,220 | 12.9 | 36.9 | 106 | 291 | 8.20 (6.71 - 9.92) | 7.89 (7.01 - 8.86) |
| **PS-matching / on-treatment TAR without escalation** | | | | | | | | |
| CCAE | 9,238 | 23,071 | 5.4 | 16.7 | 42 | 115 | 7.72 (5.56 - 10.4) | 6.89 (5.69 - 8.27) |
| ODOD | 6,735 | 16,669 | 3.5 | 10.2 | 27 | 95 | 7.79 (5.14 - 11.3) | 9.28 (7.50 - 11.3) |
| MDCR | 674 | 2,062 | 0.4 | 1.5 | <5 | 10 | < 12.9 (4.19 - 30.1) | 6.82 (3.27 - 12.6) |
| Open Claims | 137,603 | 317,206 | 84.2 | 234.9 | 378 | 972 | 4.49 (4.05 - 4.97) | 4.14 (3.88 - 4.41) |
| OEHR | 7,589 | 19,447 | 1.8 | 5.4 | 14 | 45 | 7.58 (4.15 - 12.7) | 8.30 (6.06 - 11.1) |
| VA | 1,046 | 9,692 | 0.6 | 5.0 | <10 | 31 | < 17.6 (8.44 - 32.3) | 6.15 (4.18 - 8.73) |
| Meta-analysis | 161,165 | 376,393 | 94.9 | 267.2 | 461 | 1,227 | 4.86 (4.42 - 5.32) | 4.59 (4.34 - 4.86) |
| Meta-analysis (w/o USOC) | 23,562 | 59,187 | 10.8 | 32.4 | 83 | 255 | 7.72 (6.15 - 9.57) | 7.88 (6.94 - 8.91) |
| **PS-stratification / on-treatment TAR without escalation** | | | | | | | | |
| CCAE | 9,816 | 23,075 | 5.7 | 16.7 | 44 | 115 | 7.72 (5.61 - 10.4) | 6.88 (5.68 - 8.26) |
| ODOD | 7,306 | 16,688 | 3.7 | 10.3 | 33 | 95 | 8.90 (6.12 - 12.5) | 9.26 (7.49 - 11.3) |
| MDCR | 699 | 2,062 | 0.4 | 1.5 | <5 | 10 | < 12.5 (4.06 - 29.1) | 6.82 (3.27 - 12.6) |
| Open Claims | 146,963 | 317,231 | 88.4 | 234.9 | 423 | 972 | 4.79 (4.34 - 5.26) | 4.14 (3.88 - 4.41) |
| OEHR | 8,249 | 19,457 | 2.0 | 5.4 | 16 | 45 | 8.01 (4.58 - 13.0) | 8.30 (6.05 - 11.1) |
| VA | 1,201 | 10,977 | 0.7 | 5.7 | 10 | 36 | 15.3 (7.33 - 28.1) | 6.29 (4.40 - 8.71) |
| Meta-analysis | 172,334 | 376,451 | 99.8 | 267.3 | 516 | 1,227 | 5.17 (4.73 - 5.64) | 4.59 (4.34 - 4.85) |
| Meta-analysis (w/o USOC) | 25,371 | 59,220 | 11.4 | 32.4 | 93 | 255 | 8.15 (6.58 - 9.99) | 7.87 (6.94 - 8.90) |

##### 4.1.21 Lower extremity amputation

**Supplementary Table 310:** Exposures, outcomes and risk estimates for Lower extremity amputation in GLP1RA (male) (T) vs SGLT2I (male) (C) new-user cohorts under four different propensity score (PS) adjustment / time-at-risk (TAR) designs across data sources. We report total patient exposure time in 1,000 patient-year (KYr) units, uncalibrated and calibrated hazard ratios (HRs) and their 95% confidence intervals (CIs). Greyed out entries did not pass study diagnostics and are included only for completeness.

| | Patients | | Exposure (KYr) | | Outcomes | | Incidence (per KYr) | |
|---|---|---|---|---|---|---|---|---|
| | T | C | T | C | T | C | T | C |
| **PS-matching / on-treatment TAR** | | | | | | | | |
| CCAE | 9,688 | 24,118 | 6.6 | 20.1 | 5 | 19 | 0.76 (0.25 - 1.78) | 0.95 (0.57 - 1.48) |
| ODOD | 7,134 | 17,429 | 4.1 | 12.1 | 6 | 13 | 1.45 (0.53 - 3.16) | 1.07 (0.57 - 1.84) |
| MDCR | 701 | 2,129 | 0.4 | 1.7 | 0 | <5 | | < 2.94 (0.95 - 6.86) |
| Open Claims | 144,948 | 332,213 | 101.8 | 289.1 | 48 | 159 | 0.47 (0.35 - 0.63) | 0.55 (0.47 - 0.64) |
| OEHR | 7,943 | 20,209 | 2.2 | 6.5 | 0 | <5 | | < 0.77 (0.25 - 1.81) |
| VA | 1,179 | 11,159 | 0.8 | 6.7 | 0 | 11 | | 1.64 (0.82 - 2.93) |
| Meta-analysis | 154,636 | 356,331 | 108.4 | 309.1 | 53 | 178 | 0.49 (0.37 - 0.64) | 0.58 (0.49 - 0.67) |
| Meta-analysis (w/o USOC) | 9,688 | 24,118 | 6.6 | 20.1 | 5 | 19 | 0.76 (0.25 - 1.78) | 0.95 (0.57 - 1.48) |
| **PS-stratification / on-treatment TAR** | | | | | | | | |
| CCAE | 10,320 | 24,122 | 6.9 | 20.1 | 6 | 19 | 0.87 (0.32 - 1.90) | 0.95 (0.57 - 1.48) |
| ODOD | 7,717 | 17,448 | 4.4 | 12.1 | 6 | 13 | 1.36 (0.50 - 2.97) | 1.07 (0.57 - 1.83) |
| MDCR | 731 | 2,129 | 0.5 | 1.7 | 0 | <5 | | < 2.94 (0.95 - 6.86) |
| Open Claims | 155,107 | 332,242 | 106.8 | 289.1 | 49 | 159 | 0.46 (0.34 - 0.61) | 0.55 (0.47 - 0.64) |
| OEHR | 8,659 | 20,221 | 2.4 | 6.5 | 0 | <5 | | < 0.77 (0.25 - 1.81) |
| VA | 1,350 | 12,506 | 0.9 | 7.5 | 0 | 11 | | 1.46 (0.73 - 2.62) |
| Meta-analysis | 165,427 | 356,364 | 113.7 | 309.2 | 55 | 178 | 0.48 (0.36 - 0.63) | 0.58 (0.49 - 0.67) |
| Meta-analysis (w/o USOC) | 10,320 | 24,122 | 6.9 | 20.1 | 6 | 19 | 0.87 (0.32 - 1.90) | 0.95 (0.57 - 1.48) |
| **PS-matching / on-treatment TAR without escalation** | | | | | | | | |
| CCAE | 9,688 | 24,118 | 5.7 | 17.5 | 5 | 14 | 0.87 (0.28 - 2.03) | 0.80 (0.44 - 1.34) |
| ODOD | 7,134 | 17,429 | 3.7 | 10.7 | 5 | 10 | 1.36 (0.44 - 3.17) | 0.93 (0.45 - 1.71) |
| MDCR | 701 | 2,129 | 0.4 | 1.5 | 0 | <5 | | < 3.29 (1.07 - 7.67) |
| Open Claims | 144,948 | 332,213 | 88.7 | 245.8 | 38 | 116 | 0.43 (0.30 - 0.59) | 0.47 (0.39 - 0.57) |
| OEHR | 7,943 | 20,209 | 1.9 | 5.7 | 0 | <5 | | < 0.88 (0.29 - 2.05) |
| VA | 1,179 | 11,159 | 0.6 | 5.8 | 0 | 11 | | 1.90 (0.95 - 3.39) |
| Meta-analysis | 144,948 | 332,213 | 88.7 | 245.8 | 38 | 116 | 0.43 (0.30 - 0.59) | 0.47 (0.39 - 0.57) |
| **PS-stratification / on-treatment TAR without escalation** | | | | | | | | |
| CCAE | 10,320 | 24,122 | 6.0 | 17.5 | 5 | 14 | 0.83 (0.27 - 1.93) | 0.80 (0.44 - 1.34) |
| ODOD | 7,717 | 17,448 | 3.9 | 10.8 | 5 | 10 | 1.27 (0.41 - 2.97) | 0.93 (0.45 - 1.71) |
| MDCR | 731 | 2,129 | 0.4 | 1.5 | 0 | <5 | | < 3.29 (1.07 - 7.67) |
| Open Claims | 155,107 | 332,242 | 93.3 | 245.8 | 39 | 116 | 0.42 (0.30 - 0.57) | 0.47 (0.39 - 0.57) |
| OEHR | 8,659 | 20,221 | 2.1 | 5.7 | 0 | <5 | | < 0.88 (0.29 - 2.05) |
| VA | 1,350 | 12,506 | 0.7 | 6.5 | 0 | 11 | | 1.69 (0.84 - 3.02) |
| Meta-analysis | 155,107 | 332,242 | 93.3 | 245.8 | 39 | 116 | 0.42 (0.30 - 0.57) | 0.47 (0.39 - 0.57) |

##### 4.1.22 Nausea

**Supplementary Table 311:** Exposures, outcomes and risk estimates for Nausea in GLP1RA (male) (T) vs SGLT2I (male) (C) new-user cohorts under four different propensity score (PS) adjustment / time-at-risk (TAR) designs across data sources. We report total patient exposure time in 1,000 patient-year (KYr) units, uncalibrated and calibrated hazard ratios (HRs) and their 95% confidence intervals (CIs). Greyed out entries did not pass study diagnostics and are included only for completeness.

| | Patients | | Exposure (KYr) | | Outcomes | | Incidence (per KYr) | |
|---|---|---|---|---|---|---|---|---|
| | T | C | T | C | T | C | T | C |
| **PS-matching / on-treatment TAR** | | | | | | | | |
| CCAE | 8,595 | 21,816 | 5.8 | 17.8 | 213 | 432 | 36.9 (32.1 - 42.2) | 24.2 (22.0 - 26.6) |
| ODOD | 6,268 | 15,551 | 3.6 | 10.7 | 165 | 301 | 46.4 (39.6 - 54.1) | 28.1 (25.0 - 31.5) |
| MDCR | 633 | 1,958 | 0.4 | 1.5 | 9 | 38 | 22.3 (10.2 - 42.3) | 24.5 (17.3 - 33.7) |
| Open Claims | 128,477 | 299,122 | 90.1 | 261.1 | 1,849 | 3,189 | 20.5 (19.6 - 21.5) | 12.2 (11.8 - 12.6) |
| OEHR | 7,266 | 18,772 | 2.0 | 5.9 | 113 | 162 | 56.4 (46.5 - 67.8) | 27.3 (23.3 - 31.9) |
| VA | 1,104 | 10,357 | 0.8 | 6.3 | 20 | 45 | 26.5 (16.2 - 41.0) | 7.19 (5.24 - 9.62) |
| Meta-analysis | 151,710 | 365,618 | 102.2 | 301.8 | 2,360 | 4,129 | 23.1 (22.2 - 24.0) | 13.7 (13.3 - 14.1) |
| Meta-analysis (w/o USOC) | 23,233 | 66,496 | 12.1 | 40.7 | 511 | 940 | 42.3 (38.7 - 46.1) | 23.1 (21.6 - 24.6) |
| **PS-stratification / on-treatment TAR** | | | | | | | | |
| CCAE | 9,138 | 21,821 | 6.0 | 17.8 | 232 | 432 | 38.4 (33.7 - 43.7) | 24.2 (22.0 - 26.6) |
| ODOD | 6,794 | 15,574 | 3.8 | 10.7 | 182 | 302 | 48.0 (41.2 - 55.5) | 28.2 (25.1 - 31.5) |
| MDCR | 659 | 1,958 | 0.4 | 1.5 | 9 | 38 | 21.6 (9.85 - 40.9) | 24.5 (17.3 - 33.7) |
| Open Claims | 136,903 | 299,241 | 94.3 | 261.1 | 2,002 | 3,193 | 21.2 (20.3 - 22.2) | 12.2 (11.8 - 12.7) |
| OEHR | 7,906 | 18,784 | 2.2 | 5.9 | 122 | 162 | 56.5 (46.9 - 67.4) | 27.3 (23.3 - 31.8) |
| VA | 1,265 | 11,542 | 0.9 | 7.0 | 22 | 53 | 25.7 (16.1 - 39.0) | 7.61 (5.70 - 9.96) |
| Meta-analysis | 160,741 | 355,420 | 106.3 | 295.6 | 2,538 | 4,089 | 23.9 (23.0 - 24.8) | 13.8 (13.4 - 14.3) |
| Meta-analysis (w/o USOC) | 23,838 | 56,179 | 12.0 | 34.5 | 536 | 896 | 44.7 (41.0 - 48.7) | 26.0 (24.3 - 27.7) |
| **PS-matching / on-treatment TAR without escalation** | | | | | | | | |
| CCAE | 8,595 | 21,816 | 5.1 | 15.7 | 192 | 359 | 38.0 (32.8 - 43.7) | 22.9 (20.6 - 25.4) |
| ODOD | 6,268 | 15,551 | 3.2 | 9.6 | 151 | 255 | 47.5 (40.2 - 55.7) | 26.7 (23.5 - 30.2) |
| MDCR | 633 | 1,958 | 0.4 | 1.4 | 6 | 35 | 16.4 (6.00 - 35.6) | 25.2 (17.5 - 35.0) |
| Open Claims | 128,477 | 299,122 | 78.8 | 222.8 | 1,622 | 2,538 | 20.6 (19.6 - 21.6) | 11.4 (11.0 - 11.8) |
| OEHR | 7,266 | 18,772 | 1.8 | 5.2 | 98 | 130 | 55.7 (45.2 - 67.9) | 24.8 (20.7 - 29.5) |
| VA | 1,104 | 10,357 | 0.6 | 5.4 | 17 | 35 | 28.6 (16.6 - 45.7) | 6.48 (4.51 - 9.01) |
| Meta-analysis | 151,710 | 365,618 | 89.3 | 258.7 | 2,080 | 3,317 | 23.3 (22.3 - 24.3) | 12.8 (12.4 - 13.3) |
| Meta-analysis (w/o USOC) | 23,233 | 66,496 | 10.6 | 35.9 | 458 | 779 | 43.2 (39.4 - 47.4) | 21.7 (20.2 - 23.3) |
| **PS-stratification / on-treatment TAR without escalation** | | | | | | | | |
| CCAE | 9,138 | 21,821 | 5.3 | 15.7 | 207 | 359 | 39.1 (33.9 - 44.8) | 22.9 (20.6 - 25.4) |
| ODOD | 6,794 | 15,574 | 3.4 | 9.6 | 165 | 256 | 48.5 (41.4 - 56.5) | 26.8 (23.6 - 30.2) |
| MDCR | 659 | 1,958 | 0.4 | 1.4 | 6 | 35 | 15.8 (5.81 - 34.5) | 25.2 (17.5 - 35.0) |
| Open Claims | 136,903 | 299,241 | 82.5 | 222.9 | 1,763 | 2,542 | 21.4 (20.4 - 22.4) | 11.4 (11.0 - 11.9) |
| OEHR | 7,906 | 18,784 | 1.9 | 5.2 | 105 | 130 | 55.2 (45.2 - 66.9) | 24.8 (20.7 - 29.5) |
| VA | 1,265 | 11,542 | 0.7 | 6.0 | 19 | 41 | 28.2 (17.0 - 44.0) | 6.80 (4.88 - 9.22) |
| Meta-analysis | 160,741 | 355,420 | 93.1 | 253.3 | 2,240 | 3,287 | 24.1 (23.1 - 25.1) | 13.0 (12.5 - 13.4) |
| Meta-analysis (w/o USOC) | 23,838 | 56,179 | 10.6 | 30.5 | 477 | 745 | 45.0 (41.1 - 49.2) | 24.4 (22.7 - 26.3) |

##### 4.1.23 Peripheral edema

**Supplementary Table 312:** Exposures, outcomes and risk estimates for Peripheral edema in GLP1RA (male) (T) vs SGLT2I (male) (C) new-user cohorts under four different propensity score (PS) adjustment / time-at-risk (TAR) designs across data sources. We report total patient exposure time in 1,000 patient-year (KYr) units, uncalibrated and calibrated hazard ratios (HRs) and their 95% confidence intervals (CIs). Greyed out entries did not pass study diagnostics and are included only for completeness.

| | Patients | | Exposure (KYr) | | Outcomes | | Incidence (per KYr) | |
|---|---|---|---|---|---|---|---|---|
| | T | C | T | C | T | C | T | C |
| **PS-matching / on-treatment TAR** | | | | | | | | |
| CCAE | 7,982 | 20,325 | 5.3 | 16.4 | 189 | 516 | 35.7 (30.8 - 41.2) | 31.4 (28.7 - 34.2) |
| ODOD | 5,627 | 14,073 | 3.2 | 9.6 | 150 | 405 | 47.2 (39.9 - 55.3) | 42.2 (38.2 - 46.5) |
| MDCR | 526 | 1,632 | 0.3 | 1.3 | 14 | 56 | 43.2 (23.6 - 72.4) | 44.4 (33.5 - 57.7) |
| Open Claims | 114,996 | 269,857 | 80.2 | 235.1 | 1,568 | 4,193 | 19.5 (18.6 - 20.5) | 17.8 (17.3 - 18.4) |
| OEHR | 6,688 | 17,553 | 1.8 | 5.5 | 61 | 130 | 33.3 (25.5 - 42.8) | 23.5 (19.6 - 27.9) |
| VA | 872 | 8,466 | 0.6 | 5.1 | 15 | 161 | 24.8 (13.9 - 40.9) | 31.8 (27.1 - 37.1) |
| Meta-analysis | 136,165 | 330,274 | 91.2 | 271.7 | 1,983 | 5,405 | 21.8 (20.8 - 22.7) | 19.9 (19.4 - 20.4) |
| Meta-analysis (w/o USOC) | 21,169 | 60,417 | 10.9 | 36.7 | 415 | 1,212 | 38.0 (34.5 - 41.9) | 33.1 (31.2 - 35.0) |
| **PS-stratification / on-treatment TAR** | | | | | | | | |
| CCAE | 8,446 | 20,331 | 5.5 | 16.5 | 195 | 518 | 35.4 (30.6 - 40.7) | 31.5 (28.8 - 34.3) |
| ODOD | 6,057 | 14,095 | 3.4 | 9.6 | 160 | 406 | 47.2 (40.2 - 55.2) | 42.2 (38.2 - 46.5) |
| MDCR | 557 | 1,633 | 0.3 | 1.3 | 15 | 56 | 44.4 (24.9 - 73.3) | 44.4 (33.5 - 57.7) |
| Open Claims | 122,001 | 269,915 | 83.6 | 235.1 | 1,665 | 4,195 | 19.9 (19.0 - 20.9) | 17.8 (17.3 - 18.4) |
| OEHR | 7,265 | 17,562 | 2.0 | 5.5 | 68 | 130 | 34.6 (26.8 - 43.8) | 23.5 (19.6 - 27.9) |
| VA | 989 | 9,289 | 0.7 | 5.6 | 15 | 177 | 21.7 (12.2 - 35.8) | 31.8 (27.3 - 36.9) |
| Meta-analysis | 143,769 | 321,903 | 94.5 | 266.7 | 2,088 | 5,249 | 22.1 (21.2 - 23.1) | 19.7 (19.2 - 20.2) |
| Meta-analysis (w/o USOC) | 21,768 | 51,988 | 10.9 | 31.6 | 423 | 1,054 | 38.9 (35.3 - 42.8) | 33.3 (31.4 - 35.4) |
| **PS-matching / on-treatment TAR without escalation** | | | | | | | | |
| CCAE | 7,982 | 20,325 | 4.7 | 14.5 | 162 | 438 | 34.7 (29.5 - 40.4) | 30.3 (27.5 - 33.3) |
| ODOD | 5,627 | 14,073 | 2.9 | 8.6 | 137 | 336 | 48.0 (40.3 - 56.7) | 39.1 (35.0 - 43.5) |
| MDCR | 526 | 1,632 | 0.3 | 1.1 | 11 | 48 | 37.6 (18.8 - 67.3) | 42.4 (31.2 - 56.2) |
| Open Claims | 114,996 | 269,857 | 70.1 | 200.8 | 1,353 | 3,473 | 19.3 (18.3 - 20.3) | 17.3 (16.7 - 17.9) |
| OEHR | 6,688 | 17,553 | 1.6 | 4.9 | 52 | 105 | 32.6 (24.3 - 42.7) | 21.4 (17.5 - 25.9) |
| VA | 872 | 8,466 | 0.5 | 4.4 | 11 | 136 | 23.3 (11.6 - 41.7) | 31.0 (26.1 - 36.7) |
| Meta-analysis | 136,165 | 330,274 | 79.7 | 233.1 | 1,715 | 4,488 | 21.5 (20.5 - 22.6) | 19.3 (18.7 - 19.8) |
| Meta-analysis (w/o USOC) | 21,169 | 60,417 | 9.6 | 32.3 | 362 | 1,015 | 37.7 (33.9 - 41.8) | 31.4 (29.5 - 33.4) |
| **PS-stratification / on-treatment TAR without escalation** | | | | | | | | |
| CCAE | 8,446 | 20,331 | 4.9 | 14.5 | 167 | 440 | 34.3 (29.3 - 39.9) | 30.4 (27.6 - 33.4) |
| ODOD | 6,057 | 14,095 | 3.0 | 8.6 | 147 | 336 | 48.5 (41.0 - 57.0) | 39.0 (35.0 - 43.4) |
| MDCR | 557 | 1,633 | 0.3 | 1.1 | 12 | 48 | 39.4 (20.3 - 68.7) | 42.3 (31.2 - 56.1) |
| Open Claims | 122,001 | 269,915 | 73.2 | 200.8 | 1,445 | 3,475 | 19.7 (18.7 - 20.8) | 17.3 (16.7 - 17.9) |
| OEHR | 7,265 | 17,562 | 1.7 | 4.9 | 56 | 105 | 32.6 (24.6 - 42.3) | 21.4 (17.5 - 25.9) |
| VA | 989 | 9,289 | 0.5 | 4.8 | 11 | 151 | 20.5 (10.2 - 36.7) | 31.3 (26.5 - 36.7) |
| Meta-analysis | 143,769 | 321,903 | 82.9 | 228.8 | 1,815 | 4,356 | 21.9 (20.9 - 22.9) | 19.0 (18.5 - 19.6) |
| Meta-analysis (w/o USOC) | 21,768 | 51,988 | 9.6 | 28.0 | 370 | 881 | 38.4 (34.6 - 42.6) | 31.5 (29.4 - 33.6) |

###### 4.1.24 Photosensitivity

**Supplementary Table 313:** Exposures, outcomes and risk estimates for Photosensitivity in GLP1RA (male) (T) vs SGLT2I (male) (C) new-user cohorts under four different propensity score (PS) adjustment / time-at-risk (TAR) designs across data sources. We report total patient exposure time in 1,000 patient-year (KYr) units, uncalibrated and calibrated hazard ratios (HRs) and their 95% confidence intervals (CIs). Greyed out entries did not pass study diagnostics and are included only for completeness.

| | Patients | | Exposure (KYr) | | Outcomes | | Incidence (per KYr) | |
|---|---|---|---|---|---|---|---|---|
| | T | C | T | C | T | C | T | C |
| **PS-matching / on-treatment TAR** | | | | | | | | |
| CCAE | 9,624 | 23,941 | 6.5 | 19.9 | 6 | 11 | 0.92 (0.34 - 2.00) | 0.55 (0.28 - 0.99) |
| ODOD | 7,098 | 17,330 | 4.1 | 12.0 | <5 | 8 | < 1.22 (0.39 - 2.84) | 0.67 (0.29 - 1.31) |
| MDCR | 689 | 2,105 | 0.4 | 1.7 | <5 | <5 | < 11.3 (3.68 - 26.4) | < 2.99 (0.97 - 6.97) |
| Open Claims | 143,821 | 329,537 | 100.9 | 286.5 | 26 | 68 | 0.26 (0.17 - 0.38) | 0.24 (0.18 - 0.30) |
| OEHR | 7,905 | 20,095 | 2.2 | 6.4 | <5 | 5 | < 2.26 (0.73 - 5.27) | 0.78 (0.25 - 1.82) |
| VA | 1,167 | 11,033 | 0.8 | 6.6 | 0 | <10 | | < 1.51 (0.72 - 2.77) |
| Meta-analysis | 143,821 | 329,537 | 100.9 | 286.5 | 26 | 68 | 0.26 (0.17 - 0.38) | 0.24 (0.18 - 0.30) |
| **PS-stratification / on-treatment TAR** | | | | | | | | |
| CCAE | 10,254 | 23,945 | 6.8 | 19.9 | 6 | 11 | 0.88 (0.32 - 1.91) | 0.55 (0.28 - 0.99) |
| ODOD | 7,688 | 17,348 | 4.4 | 12.0 | <5 | 8 | < 1.14 (0.37 - 2.66) | 0.66 (0.29 - 1.31) |
| MDCR | 720 | 2,105 | 0.5 | 1.7 | <5 | <5 | < 10.9 (3.54 - 25.4) | < 2.99 (0.97 - 6.97) |
| Open Claims | 153,926 | 329,565 | 105.8 | 286.5 | 32 | 68 | 0.30 (0.21 - 0.43) | 0.24 (0.18 - 0.30) |
| OEHR | 8,610 | 20,107 | 2.4 | 6.4 | <5 | 5 | < 2.09 (0.68 - 4.89) | 0.78 (0.25 - 1.82) |
| VA | 1,337 | 12,352 | 0.9 | 7.4 | 0 | <10 | | < 1.35 (0.65 - 2.48) |
| Meta-analysis | 153,926 | 329,565 | 105.8 | 286.5 | 32 | 68 | 0.30 (0.21 - 0.43) | 0.24 (0.18 - 0.30) |
| **PS-matching / on-treatment TAR without escalation** | | | | | | | | |
| CCAE | 9,624 | 23,941 | 5.7 | 17.4 | 5 | 10 | 0.88 (0.28 - 2.04) | 0.58 (0.28 - 1.06) |
| ODOD | 7,098 | 17,330 | 3.7 | 10.7 | <5 | 7 | < 1.36 (0.44 - 3.18) | 0.66 (0.26 - 1.35) |
| MDCR | 689 | 2,105 | 0.4 | 1.5 | <5 | <5 | < 12.6 (4.09 - 29.4) | < 3.34 (1.08 - 7.80) |
| Open Claims | 143,821 | 329,537 | 87.9 | 243.6 | 24 | 55 | 0.27 (0.17 - 0.41) | 0.23 (0.17 - 0.29) |
| OEHR | 7,905 | 20,095 | 1.9 | 5.7 | <5 | <5 | < 2.60 (0.84 - 6.06) | < 0.88 (0.29 - 2.06) |
| VA | 1,167 | 11,033 | 0.6 | 5.7 | 0 | <10 | | < 1.75 (0.84 - 3.22) |
| Meta-analysis | 143,821 | 329,537 | 87.9 | 243.6 | 24 | 55 | 0.27 (0.17 - 0.41) | 0.23 (0.17 - 0.29) |
| **PS-stratification / on-treatment TAR without escalation** | | | | | | | | |
| CCAE | 10,254 | 23,945 | 6.0 | 17.4 | 5 | 10 | 0.83 (0.27 - 1.95) | 0.58 (0.28 - 1.06) |
| ODOD | 7,688 | 17,348 | 3.9 | 10.7 | <5 | 7 | < 1.28 (0.41 - 2.98) | 0.66 (0.26 - 1.35) |
| MDCR | 720 | 2,105 | 0.4 | 1.5 | <5 | <5 | < 12.1 (3.93 - 28.3) | < 3.34 (1.08 - 7.80) |
| Open Claims | 153,926 | 329,565 | 92.5 | 243.6 | 30 | 55 | 0.32 (0.22 - 0.46) | 0.23 (0.17 - 0.29) |
| OEHR | 8,610 | 20,107 | 2.1 | 5.7 | <5 | <5 | < 2.40 (0.78 - 5.60) | < 0.88 (0.29 - 2.06) |
| VA | 1,337 | 12,352 | 0.7 | 6.4 | 0 | <10 | | < 1.56 (0.75 - 2.87) |
| Meta-analysis | 153,926 | 329,565 | 92.5 | 243.6 | 30 | 55 | 0.32 (0.22 - 0.46) | 0.23 (0.17 - 0.29) |

##### 4.1.25 Venous thromboembolism

**Supplementary Table 314:** Exposures, outcomes and risk estimates for Venous thromboembolism in GLP1RA (male) (T) vs SGLT2I (male) (C) new-user cohorts under four different propensity score (PS) adjustment / time-at-risk (TAR) designs across data sources. We report total patient exposure time in 1,000 patient-year (KYr) units, uncalibrated and calibrated hazard ratios (HRs) and their 95% confidence intervals (CIs). Greyed out entries did not pass study diagnostics and are included only for completeness.

| | Patients | | Exposure (KYr) | | Outcomes | | Incidence (per KYr) | |
|---|---|---|---|---|---|---|---|---|
| | T | C | T | C | T | C | T | C |
| **PS-matching / on-treatment TAR** | | | | | | | | |
| CCAE | 9,448 | 23,616 | 6.4 | 19.6 | 32 | 77 | 5.00 (3.42 - 7.06) | 3.92 (3.10 - 4.90) |
| ODOD | 6,912 | 16,956 | 4.0 | 11.8 | 21 | 81 | 5.25 (3.25 - 8.02) | 6.89 (5.47 - 8.56) |
| MDCR | 669 | 2,048 | 0.4 | 1.6 | 6 | 15 | 14.1 (5.16 - 30.6) | 9.27 (5.19 - 15.3) |
| Open Claims | 139,783 | 321,841 | 98.2 | 280.3 | 346 | 985 | 3.52 (3.16 - 3.92) | 3.51 (3.30 - 3.74) |
| OEHR | 7,745 | 19,734 | 2.2 | 6.3 | 15 | 42 | 6.88 (3.85 - 11.3) | 6.71 (4.84 - 9.07) |
| VA | 1,133 | 10,718 | 0.8 | 6.5 | <10 | 36 | < 12.6 (6.04 - 23.2) | 5.58 (3.91 - 7.72) |
| Meta-analysis | 163,888 | 382,147 | 110.8 | 317.9 | 414 | 1,185 | 3.74 (3.39 - 4.12) | 3.73 (3.52 - 3.95) |
| Meta-analysis (w/o USOC) | 24,105 | 60,306 | 12.6 | 37.6 | 68 | 200 | 5.41 (4.20 - 6.85) | 5.31 (4.60 - 6.10) |
| **PS-stratification / on-treatment TAR** | | | | | | | | |
| CCAE | 10,036 | 23,622 | 6.7 | 19.6 | 35 | 77 | 5.24 (3.65 - 7.29) | 3.92 (3.09 - 4.90) |
| ODOD | 7,476 | 16,980 | 4.3 | 11.8 | 24 | 81 | 5.62 (3.60 - 8.36) | 6.88 (5.46 - 8.55) |
| MDCR | 697 | 2,048 | 0.4 | 1.6 | 6 | 15 | 13.7 (5.02 - 29.8) | 9.27 (5.19 - 15.3) |
| Open Claims | 149,433 | 321,867 | 102.9 | 280.3 | 369 | 985 | 3.59 (3.23 - 3.97) | 3.51 (3.30 - 3.74) |
| OEHR | 8,429 | 19,746 | 2.3 | 6.3 | 17 | 42 | 7.25 (4.23 - 11.6) | 6.71 (4.83 - 9.06) |
| VA | 1,292 | 11,951 | 0.9 | 7.2 | <10 | 44 | < 11.1 (5.34 - 20.5) | 6.13 (4.45 - 8.23) |
| Meta-analysis | 175,374 | 382,215 | 116.2 | 318.0 | 445 | 1,185 | 3.83 (3.48 - 4.20) | 3.73 (3.52 - 3.95) |
| Meta-analysis (w/o USOC) | 25,941 | 60,348 | 13.3 | 37.7 | 76 | 200 | 5.72 (4.51 - 7.16) | 5.31 (4.60 - 6.10) |
| **PS-matching / on-treatment TAR without escalation** | | | | | | | | |
| CCAE | 9,448 | 23,616 | 5.6 | 17.2 | 29 | 64 | 5.18 (3.47 - 7.44) | 3.73 (2.87 - 4.76) |
| ODOD | 6,912 | 16,956 | 3.6 | 10.4 | 20 | 66 | 5.60 (3.42 - 8.64) | 6.33 (4.89 - 8.05) |
| MDCR | 669 | 2,048 | 0.4 | 1.4 | <5 | 15 | < 13.0 (4.23 - 30.4) | 10.4 (5.81 - 17.1) |
| Open Claims | 139,783 | 321,841 | 85.6 | 238.4 | 302 | 805 | 3.53 (3.14 - 3.95) | 3.38 (3.15 - 3.62) |
| OEHR | 7,745 | 19,734 | 1.9 | 5.5 | 12 | 31 | 6.30 (3.26 - 11.0) | 5.62 (3.82 - 7.98) |
| VA | 1,133 | 10,718 | 0.6 | 5.6 | <10 | 29 | < 16.1 (7.70 - 29.5) | 5.21 (3.49 - 7.48) |
| Meta-analysis | 163,888 | 382,147 | 96.6 | 271.5 | 363 | 966 | 3.76 (3.38 - 4.16) | 3.56 (3.34 - 3.79) |
| Meta-analysis (w/o USOC) | 24,105 | 60,306 | 11.1 | 33.1 | 61 | 161 | 5.51 (4.21 - 7.07) | 4.86 (4.14 - 5.68) |
| **PS-stratification / on-treatment TAR without escalation** | | | | | | | | |
| CCAE | 10,036 | 23,622 | 5.9 | 17.2 | 32 | 64 | 5.46 (3.74 - 7.71) | 3.73 (2.87 - 4.76) |
| ODOD | 7,476 | 16,980 | 3.8 | 10.4 | 22 | 66 | 5.77 (3.62 - 8.74) | 6.32 (4.89 - 8.04) |
| MDCR | 697 | 2,048 | 0.4 | 1.4 | <5 | 15 | < 12.6 (4.11 - 29.5) | 10.4 (5.81 - 17.1) |
| Open Claims | 149,433 | 321,867 | 89.9 | 238.5 | 323 | 805 | 3.59 (3.21 - 4.01) | 3.38 (3.15 - 3.62) |
| OEHR | 8,429 | 19,746 | 2.0 | 5.5 | 14 | 31 | 6.83 (3.74 - 11.5) | 5.62 (3.82 - 7.97) |
| VA | 1,292 | 11,951 | 0.7 | 6.2 | <10 | 36 | < 14.2 (6.80 - 26.1) | 5.79 (4.06 - 8.02) |
| Meta-analysis | 175,374 | 382,215 | 101.7 | 271.6 | 391 | 966 | 3.85 (3.47 - 4.25) | 3.56 (3.34 - 3.79) |
| Meta-analysis (w/o USOC) | 25,941 | 60,348 | 11.7 | 33.1 | 68 | 161 | 5.80 (4.51 - 7.36) | 4.86 (4.14 - 5.67) |

##### 4.1.26 Vomiting

**Supplementary Table 315:** Exposures, outcomes and risk estimates for Vomiting in GLP1RA (male) (T) vs SGLT2I (male) (C) new-user cohorts under four different propensity score (PS) adjustment / time-at-risk (TAR) designs across data sources. We report total patient exposure time in 1,000 patient-year (KYr) units, uncalibrated and calibrated hazard ratios (HRs) and their 95% confidence intervals (CIs). Greyed out entries did not pass study diagnostics and are included only for completeness.

| | Patients | | Exposure (KYr) | | Outcomes | | Incidence (per KYr) | |
|---|---|---|---|---|---|---|---|---|
| | T | C | T | C | T | C | T | C |
| **PS-matching / on-treatment TAR** | | | | | | | | |
| CCAE | 8,995 | 22,524 | 6.1 | 18.5 | 142 | 308 | 23.4 (19.7 - 27.6) | 16.6 (14.8 - 18.6) |
| ODOD | 6,553 | 16,119 | 3.7 | 11.1 | 102 | 212 | 27.4 (22.4 - 33.3) | 19.0 (16.6 - 21.8) |
| MDCR | 664 | 2,025 | 0.4 | 1.6 | 8 | 29 | 19.2 (8.27 - 37.8) | 18.1 (12.1 - 26.0) |
| Open Claims | 132,807 | 307,109 | 93.4 | 268.3 | 1,364 | 2,347 | 14.6 (13.8 - 15.4) | 8.75 (8.40 - 9.11) |
| OEHR | 7,513 | 19,267 | 2.1 | 6.1 | 82 | 99 | 39.1 (31.1 - 48.6) | 16.2 (13.1 - 19.7) |
| VA | 1,128 | 10,553 | 0.8 | 6.4 | 12 | 30 | 15.3 (7.91 - 26.8) | 4.70 (3.17 - 6.71) |
| Meta-analysis | 155,868 | 365,019 | 105.3 | 304.1 | 1,690 | 2,966 | 16.1 (15.3 - 16.8) | 9.75 (9.41 - 10.1) |
| Meta-analysis (w/o USOC) | 23,061 | 57,910 | 11.9 | 35.8 | 326 | 619 | 27.5 (24.6 - 30.6) | 17.3 (16.0 - 18.7) |
| **PS-stratification / on-treatment TAR** | | | | | | | | |
| CCAE | 9,558 | 22,530 | 6.3 | 18.5 | 151 | 308 | 23.8 (20.2 - 28.0) | 16.6 (14.8 - 18.6) |
| ODOD | 7,096 | 16,142 | 4.0 | 11.2 | 111 | 213 | 27.9 (23.0 - 33.6) | 19.1 (16.6 - 21.8) |
| MDCR | 691 | 2,025 | 0.4 | 1.6 | 8 | 29 | 18.5 (7.99 - 36.5) | 18.1 (12.1 - 26.0) |
| Open Claims | 141,776 | 307,189 | 97.8 | 268.3 | 1,490 | 2,351 | 15.2 (14.5 - 16.0) | 8.76 (8.41 - 9.12) |
| OEHR | 8,172 | 19,279 | 2.3 | 6.1 | 88 | 99 | 39.0 (31.3 - 48.0) | 16.2 (13.1 - 19.7) |
| VA | 1,295 | 11,772 | 0.9 | 7.1 | 14 | 40 | 15.7 (8.56 - 26.3) | 5.62 (4.02 - 7.66) |
| Meta-analysis | 166,602 | 365,140 | 110.3 | 304.1 | 1,840 | 2,971 | 16.7 (15.9 - 17.5) | 9.77 (9.42 - 10.1) |
| Meta-analysis (w/o USOC) | 24,826 | 57,951 | 12.6 | 35.8 | 350 | 620 | 27.8 (25.0 - 30.9) | 17.3 (16.0 - 18.7) |
| **PS-matching / on-treatment TAR without escalation** | | | | | | | | |
| CCAE | 8,995 | 22,524 | 5.3 | 16.2 | 131 | 255 | 24.7 (20.6 - 29.3) | 15.7 (13.8 - 17.8) |
| ODOD | 6,553 | 16,119 | 3.3 | 9.9 | 95 | 178 | 28.5 (23.1 - 34.9) | 18.0 (15.4 - 20.8) |
| MDCR | 664 | 2,025 | 0.4 | 1.4 | 5 | 27 | 13.2 (4.30 - 30.9) | 18.8 (12.4 - 27.4) |
| Open Claims | 132,807 | 307,109 | 81.6 | 228.8 | 1,178 | 1,863 | 14.4 (13.6 - 15.3) | 8.14 (7.78 - 8.52) |
| OEHR | 7,513 | 19,267 | 1.8 | 5.4 | 67 | 84 | 36.5 (28.3 - 46.3) | 15.6 (12.4 - 19.3) |
| VA | 1,128 | 10,553 | 0.6 | 5.5 | 11 | 22 | 18.0 (8.97 - 32.1) | 3.99 (2.50 - 6.04) |
| Meta-analysis | 155,868 | 365,019 | 92.1 | 260.3 | 1,471 | 2,380 | 16.0 (15.2 - 16.8) | 9.14 (8.78 - 9.52) |
| Meta-analysis (w/o USOC) | 23,061 | 57,910 | 10.5 | 31.5 | 293 | 517 | 28.0 (24.8 - 31.4) | 16.4 (15.0 - 17.9) |
| **PS-stratification / on-treatment TAR without escalation** | | | | | | | | |
| CCAE | 9,558 | 22,530 | 5.6 | 16.2 | 139 | 255 | 25.0 (21.0 - 29.5) | 15.7 (13.8 - 17.8) |
| ODOD | 7,096 | 16,142 | 3.6 | 9.9 | 101 | 179 | 28.3 (23.1 - 34.4) | 18.0 (15.5 - 20.9) |
| MDCR | 691 | 2,025 | 0.4 | 1.4 | 5 | 27 | 12.8 (4.15 - 29.9) | 18.8 (12.4 - 27.4) |
| Open Claims | 141,776 | 307,189 | 85.6 | 228.8 | 1,291 | 1,867 | 15.1 (14.3 - 15.9) | 8.16 (7.79 - 8.54) |
| OEHR | 8,172 | 19,279 | 2.0 | 5.4 | 73 | 84 | 36.9 (28.9 - 46.4) | 15.5 (12.4 - 19.2) |
| VA | 1,295 | 11,772 | 0.7 | 6.2 | 13 | 31 | 18.6 (9.91 - 31.8) | 5.03 (3.42 - 7.14) |
| Meta-analysis | 166,602 | 365,140 | 96.7 | 260.4 | 1,604 | 2,385 | 16.6 (15.8 - 17.4) | 9.16 (8.80 - 9.54) |
| Meta-analysis (w/o USOC) | 24,826 | 57,951 | 11.1 | 31.6 | 313 | 518 | 28.2 (25.1 - 31.5) | 16.4 (15.0 - 17.9) |

#### 4.2 SGLT2I (male) vs DPP4I (male)

##### 4.2.1 3-point MACE

**Supplementary Table 316:** Exposures, outcomes and risk estimates for 3-point MACE in SGLT2I (male) (T) vs DPP4I (male) (C) new-user cohorts under four different propensity score (PS) adjustment / time-at-risk (TAR) designs across data sources. We report total patient exposure time in 1,000 patient-year (KYr) units, uncalibrated and calibrated hazard ratios (HRs) and their 95% confidence intervals (CIs). Greyed out entries did not pass study diagnostics and are included only for completeness.

| | Patients | | Exposure (KYr) | | Outcomes | | Incidence (per KYr) | |
|---|---|---|---|---|---|---|---|---|
| | T | C | T | C | T | C | T | C |
| **PS-matching / on-treatment TAR** | | | | | | | | |
| CCAE | 19,046 | 30,069 | 17.0 | 25.3 | 127 | 233 | 7.47 (6.23 - 8.89) | 9.20 (8.06 - 10.5) |
| DAG | 4,640 | 11,168 | 4.1 | 11.6 | 0 | 0 | | |
| ODOD | 11,893 | 20,135 | 9.1 | 15.8 | 133 | 331 | 14.7 (12.3 - 17.4) | 21.0 (18.8 - 23.3) |
| MDCR | 1,609 | 4,821 | 1.4 | 4.5 | 28 | 125 | 19.5 (13.0 - 28.2) | 27.5 (22.9 - 32.7) |
| MDCD | 1,303 | 3,149 | 0.8 | 2.3 | 17 | 48 | 21.3 (12.4 - 34.0) | 20.9 (15.4 - 27.7) |
| Open Claims | 230,169 | 399,585 | 223.7 | 382.0 | 1,716 | 4,171 | 7.67 (7.31 - 8.04) | 10.9 (10.6 - 11.3) |
| OEHR | 16,401 | 31,503 | 5.3 | 16.2 | 43 | 172 | 8.05 (5.82 - 10.8) | 10.6 (9.10 - 12.3) |
| VA | 7,323 | 23,032 | 5.0 | 20.8 | 23 | 112 | 4.62 (2.93 - 6.93) | 5.38 (4.43 - 6.47) |
| SIDIAP | 4,064 | 19,143 | 5.9 | 43.3 | 26 | 237 | 4.42 (2.88 - 6.47) | 5.47 (4.79 - 6.21) |
| Meta-analysis | 290,505 | 528,288 | 267.4 | 508.0 | 2,096 | 5,381 | 7.84 (7.51 - 8.18) | 10.6 (10.3 - 10.9) |
| Meta-analysis (w/o USOC) | 60,336 | 128,703 | 43.7 | 126.0 | 380 | 1,210 | 8.70 (7.85 - 9.62) | 9.60 (9.07 - 10.2) |
| **PS-stratification / on-treatment TAR** | | | | | | | | |
| CCAE | 23,133 | 30,091 | 19.3 | 25.3 | 140 | 233 | 7.27 (6.11 - 8.57) | 9.20 (8.06 - 10.5) |
| DAG | 5,763 | 11,200 | 4.7 | 11.7 | 0 | 0 | | |
| ODOD | 16,422 | 20,238 | 11.4 | 15.9 | 170 | 339 | 14.9 (12.8 - 17.4) | 21.3 (19.1 - 23.7) |
| MDCR | 1,977 | 4,866 | 1.6 | 4.6 | 36 | 127 | 22.8 (15.9 - 31.5) | 27.6 (23.0 - 32.8) |
| MDCD | 1,365 | 3,169 | 0.8 | 2.3 | 17 | 50 | 20.9 (12.2 - 33.5) | 21.5 (16.0 - 28.4) |
| Open Claims | 308,612 | 399,821 | 270.1 | 382.2 | 2,093 | 4,185 | 7.75 (7.42 - 8.09) | 11.0 (10.6 - 11.3) |
| OEHR | 19,539 | 31,569 | 6.2 | 16.2 | 48 | 175 | 7.69 (5.67 - 10.2) | 10.8 (9.26 - 12.5) |
| VA | 11,375 | 24,034 | 6.9 | 22.3 | 50 | 122 | 7.26 (5.39 - 9.57) | 5.47 (4.55 - 6.54) |
| SIDIAP | 4,643 | 19,567 | 6.4 | 44.9 | 29 | 248 | 4.55 (3.05 - 6.53) | 5.53 (4.86 - 6.26) |
| Meta-analysis | 374,326 | 506,152 | 315.0 | 489.1 | 2,516 | 5,307 | 7.99 (7.68 - 8.31) | 10.9 (10.6 - 11.1) |
| Meta-analysis (w/o USOC) | 65,714 | 106,331 | 44.8 | 106.9 | 423 | 1,122 | 9.43 (8.56 - 10.4) | 10.5 (9.89 - 11.1) |
| **PS-matching / on-treatment TAR without escalation** | | | | | | | | |
| CCAE | 19,046 | 30,069 | 14.7 | 20.5 | 104 | 187 | 7.06 (5.77 - 8.56) | 9.12 (7.86 - 10.5) |
| DAG | 4,640 | 11,168 | 3.7 | 10.5 | 0 | 0 | | |
| ODOD | 11,893 | 20,135 | 8.0 | 12.9 | 116 | 279 | 14.6 (12.0 - 17.5) | 21.6 (19.1 - 24.3) |
| MDCR | 1,609 | 4,821 | 1.3 | 3.9 | 24 | 108 | 18.9 (12.1 - 28.1) | 27.9 (22.9 - 33.7) |
| MDCD | 1,303 | 3,149 | 0.7 | 1.7 | 15 | 38 | 22.3 (12.5 - 36.8) | 22.1 (15.6 - 30.3) |
| Open Claims | 230,169 | 399,585 | 187.6 | 296.5 | 1,438 | 3,248 | 7.66 (7.27 - 8.07) | 11.0 (10.6 - 11.3) |
| OEHR | 16,401 | 31,503 | 4.7 | 13.1 | 33 | 123 | 7.05 (4.85 - 9.89) | 9.38 (7.80 - 11.2) |
| VA | 7,323 | 23,032 | 4.2 | 17.0 | 20 | 78 | 4.72 (2.88 - 7.29) | 4.60 (3.64 - 5.74) |
| SIDIAP | 4,064 | 19,143 | 5.2 | 34.5 | 24 | 180 | 4.59 (2.94 - 6.82) | 5.21 (4.48 - 6.03) |
| Meta-analysis | 290,505 | 528,288 | 225.7 | 398.4 | 1,759 | 4,203 | 7.79 (7.43 - 8.16) | 10.5 (10.2 - 10.9) |
| Meta-analysis (w/o USOC) | 60,336 | 128,703 | 38.1 | 101.9 | 321 | 955 | 8.42 (7.53 - 9.40) | 9.37 (8.79 - 9.99) |
| **PS-stratification / on-treatment TAR without escalation** | | | | | | | | |
| CCAE | 23,133 | 30,091 | 16.8 | 20.5 | 116 | 187 | 6.89 (5.70 - 8.27) | 9.11 (7.85 - 10.5) |
| DAG | 5,763 | 11,200 | 4.3 | 10.6 | 0 | 0 | | |
| ODOD | 16,422 | 20,238 | 10.1 | 13.0 | 150 | 285 | 14.9 (12.6 - 17.4) | 21.9 (19.4 - 24.6) |
| MDCR | 1,977 | 4,866 | 1.4 | 3.9 | 32 | 109 | 22.7 (15.5 - 32.0) | 27.9 (22.9 - 33.6) |
| MDCD | 1,365 | 3,169 | 0.7 | 1.7 | 15 | 39 | 21.9 (12.2 - 36.1) | 22.4 (16.0 - 30.7) |
| Open Claims | 308,612 | 399,821 | 229.5 | 296.7 | 1,786 | 3,260 | 7.78 (7.43 - 8.15) | 11.0 (10.6 - 11.4) |
| OEHR | 19,539 | 31,569 | 5.5 | 13.1 | 38 | 124 | 6.91 (4.89 - 9.49) | 9.44 (7.85 - 11.3) |
| VA | 11,375 | 24,034 | 6.0 | 18.1 | 42 | 85 | 7.06 (5.09 - 9.54) | 4.69 (3.75 - 5.81) |
| SIDIAP | 4,643 | 19,567 | 5.7 | 35.7 | 27 | 189 | 4.75 (3.13 - 6.91) | 5.30 (4.57 - 6.11) |
| Meta-analysis | 374,326 | 506,152 | 269.0 | 382.9 | 2,149 | 4,154 | 7.99 (7.65 - 8.33) | 10.8 (10.5 - 11.2) |
| Meta-analysis (w/o USOC) | 65,714 | 106,331 | 39.5 | 86.2 | 363 | 894 | 9.19 (8.26 - 10.2) | 10.4 (9.70 - 11.1) |

##### 4.2.2 4-point MACE

**Supplementary Table 317:** Exposures, outcomes and risk estimates for 4-point MACE in SGLT2I (male) (T) vs DPP4I (male) (C) new-user cohorts under four different propensity score (PS) adjustment / time-at-risk (TAR) designs across data sources. We report total patient exposure time in 1,000 patient-year (KYr) units, uncalibrated and calibrated hazard ratios (HRs) and their 95% confidence intervals (CIs). Greyed out entries did not pass study diagnostics and are included only for completeness.

| | Patients | | Exposure (KYr) | | Outcomes | | Incidence (per KYr) | |
|---|---|---|---|---|---|---|---|---|
| | T | C | T | C | T | C | T | C |
| **PS-matching / on-treatment TAR** | | | | | | | | |
| CCAE | 18,788 | 29,650 | 16.8 | 24.9 | 183 | 325 | 10.9 (9.40 - 12.6) | 13.0 (11.7 - 14.5) |
| DAG | 4,640 | 11,168 | 4.1 | 11.6 | 0 | 0 | | |
| ODOD | 11,493 | 19,247 | 8.8 | 15.0 | 191 | 491 | 21.8 (18.8 - 25.1) | 32.7 (29.8 - 35.7) |
| MDCR | 1,531 | 4,540 | 1.4 | 4.3 | 43 | 196 | 31.5 (22.8 - 42.4) | 46.1 (39.8 - 53.0) |
| MDCD | 1,212 | 2,924 | 0.7 | 2.1 | 20 | 60 | 27.2 (16.6 - 42.0) | 28.5 (21.8 - 36.7) |
| Open Claims | 224,326 | 386,854 | 218.7 | 369.1 | 2,394 | 6,506 | 10.9 (10.5 - 11.4) | 17.6 (17.2 - 18.1) |
| OEHR | 16,132 | 30,890 | 5.3 | 15.7 | 77 | 308 | 14.6 (11.5 - 18.3) | 19.6 (17.5 - 21.9) |
| VA | 7,057 | 22,438 | 4.8 | 20.2 | 43 | 244 | 9.00 (6.51 - 12.1) | 12.1 (10.6 - 13.7) |
| SIDIAP | 4,038 | 19,055 | 5.8 | 43.1 | 31 | 296 | 5.30 (3.60 - 7.52) | 6.86 (6.10 - 7.69) |
| Meta-analysis | 283,365 | 512,674 | 261.5 | 492.3 | 2,962 | 8,366 | 11.3 (10.9 - 11.7) | 17.0 (16.6 - 17.4) |
| Meta-analysis (w/o USOC) | 59,039 | 125,820 | 42.8 | 123.2 | 568 | 1,860 | 13.3 (12.2 - 14.4) | 15.1 (14.4 - 15.8) |
| **PS-stratification / on-treatment TAR** | | | | | | | | |
| CCAE | 22,780 | 29,670 | 19.0 | 24.9 | 211 | 325 | 11.1 (9.67 - 12.7) | 13.0 (11.7 - 14.5) |
| DAG | 5,763 | 11,200 | 4.7 | 11.7 | 0 | 0 | | |
| ODOD | 15,713 | 19,318 | 10.9 | 15.1 | 257 | 500 | 23.5 (20.7 - 26.5) | 33.1 (30.3 - 36.1) |
| MDCR | 1,862 | 4,574 | 1.5 | 4.3 | 52 | 198 | 34.7 (25.9 - 45.6) | 46.1 (39.9 - 53.0) |
| MDCD | 1,263 | 2,941 | 0.7 | 2.1 | 20 | 63 | 26.8 (16.3 - 41.3) | 29.7 (22.8 - 38.0) |
| Open Claims | 297,226 | 387,040 | 262.0 | 369.2 | 2,981 | 6,531 | 11.4 (11.0 - 11.8) | 17.7 (17.3 - 18.1) |
| OEHR | 19,146 | 30,946 | 6.1 | 15.7 | 93 | 312 | 15.2 (12.3 - 18.6) | 19.8 (17.7 - 22.2) |
| VA | 10,518 | 23,414 | 6.4 | 21.6 | 86 | 260 | 13.4 (10.7 - 16.6) | 12.0 (10.6 - 13.6) |
| SIDIAP | 4,579 | 19,474 | 6.3 | 44.6 | 33 | 314 | 5.24 (3.61 - 7.36) | 7.03 (6.28 - 7.86) |
| Meta-analysis | 361,306 | 491,022 | 305.8 | 473.9 | 3,627 | 8,180 | 11.9 (11.5 - 12.3) | 17.3 (16.9 - 17.6) |
| Meta-analysis (w/o USOC) | 64,080 | 103,982 | 43.8 | 104.7 | 646 | 1,649 | 14.7 (13.6 - 15.9) | 15.7 (15.0 - 16.5) |
| **PS-matching / on-treatment TAR without escalation** | | | | | | | | |
| CCAE | 18,788 | 29,650 | 14.5 | 20.2 | 152 | 261 | 10.5 (8.87 - 12.3) | 12.9 (11.4 - 14.6) |
| DAG | 4,640 | 11,168 | 3.7 | 10.5 | 0 | 0 | | |
| ODOD | 11,493 | 19,247 | 7.7 | 12.3 | 162 | 412 | 21.0 (17.9 - 24.5) | 33.5 (30.3 - 36.9) |
| MDCR | 1,531 | 4,540 | 1.2 | 3.6 | 37 | 174 | 30.6 (21.5 - 42.1) | 48.1 (41.3 - 55.8) |
| MDCD | 1,212 | 2,924 | 0.6 | 1.6 | 17 | 46 | 27.4 (16.0 - 43.9) | 29.1 (21.3 - 38.9) |
| Open Claims | 224,326 | 386,854 | 183.6 | 286.7 | 1,970 | 5,063 | 10.7 (10.3 - 11.2) | 17.7 (17.2 - 18.2) |
| OEHR | 16,132 | 30,890 | 4.6 | 12.8 | 60 | 222 | 13.0 (9.91 - 16.7) | 17.4 (15.2 - 19.8) |
| VA | 7,057 | 22,438 | 4.1 | 16.5 | 36 | 179 | 8.84 (6.19 - 12.2) | 10.9 (9.34 - 12.6) |
| SIDIAP | 4,038 | 19,055 | 5.2 | 34.4 | 27 | 224 | 5.19 (3.42 - 7.55) | 6.52 (5.69 - 7.43) |
| Meta-analysis | 283,365 | 512,674 | 220.9 | 386.4 | 2,444 | 6,535 | 11.1 (10.6 - 11.5) | 16.9 (16.5 - 17.3) |
| Meta-analysis (w/o USOC) | 59,039 | 125,820 | 37.4 | 99.7 | 474 | 1,472 | 12.7 (11.6 - 13.9) | 14.8 (14.0 - 15.5) |
| **PS-stratification / on-treatment TAR without escalation** | | | | | | | | |
| CCAE | 22,780 | 29,670 | 16.6 | 20.2 | 176 | 261 | 10.6 (9.11 - 12.3) | 12.9 (11.4 - 14.6) |
| DAG | 5,763 | 11,200 | 4.3 | 10.6 | 0 | 0 | | |
| ODOD | 15,713 | 19,318 | 9.7 | 12.4 | 226 | 419 | 23.2 (20.3 - 26.5) | 33.9 (30.7 - 37.3) |
| MDCR | 1,862 | 4,574 | 1.3 | 3.7 | 46 | 175 | 34.5 (25.3 - 46.1) | 47.9 (41.1 - 55.6) |
| MDCD | 1,263 | 2,941 | 0.6 | 1.6 | 17 | 48 | 27.0 (15.7 - 43.2) | 30.1 (22.2 - 39.9) |
| Open Claims | 297,226 | 387,040 | 222.7 | 286.8 | 2,508 | 5,085 | 11.3 (10.8 - 11.7) | 17.7 (17.2 - 18.2) |
| OEHR | 19,146 | 30,946 | 5.4 | 12.8 | 75 | 223 | 13.9 (10.9 - 17.4) | 17.4 (15.2 - 19.9) |
| VA | 10,518 | 23,414 | 5.5 | 17.6 | 71 | 192 | 12.8 (10.0 - 16.2) | 10.9 (9.43 - 12.6) |
| SIDIAP | 4,579 | 19,474 | 5.6 | 35.5 | 29 | 239 | 5.16 (3.46 - 7.41) | 6.73 (5.91 - 7.64) |
| Meta-analysis | 361,306 | 491,022 | 261.3 | 371.4 | 3,060 | 6,402 | 11.7 (11.3 - 12.1) | 17.2 (16.8 - 17.7) |
| Meta-analysis (w/o USOC) | 64,080 | 103,982 | 38.6 | 84.5 | 552 | 1,317 | 14.3 (13.1 - 15.5) | 15.6 (14.8 - 16.4) |

##### 4.2.3 Acute myocardial infarction

**Supplementary Table 318:** Exposures, outcomes and risk estimates for Acute myocardial infarction in SGLT2I (male) (T) vs DPP4I (male) (C) new-user cohorts under four different propensity score (PS) adjustment / time-at-risk (TAR) designs across data sources. We report total patient exposure time in 1,000 patient-year (KYr) units, uncalibrated and calibrated hazard ratios (HRs) and their 95% confidence intervals (CIs). Greyed out entries did not pass study diagnostics and are included only for completeness.

| | Patients | | Exposure (KYr) | | Outcomes | | Incidence (per KYr) | |
|---|---|---|---|---|---|---|---|---|
| | T | C | T | C | T | C | T | C |
| **PS-matching / on-treatment TAR** | | | | | | | | |
| CCAE | 19,282 | 30,461 | 17.2 | 25.7 | 75 | 137 | 4.36 (3.43 - 5.46) | 5.33 (4.48 - 6.31) |
| DAG | 4,640 | 11,168 | 4.1 | 11.6 | 0 | 0 | | |
| ODOD | 12,151 | 20,752 | 9.3 | 16.3 | 73 | 171 | 7.87 (6.17 - 9.89) | 10.5 (8.96 - 12.2) |
| MDCR | 1,652 | 5,026 | 1.5 | 4.8 | 9 | 64 | 6.11 (2.80 - 11.6) | 13.4 (10.3 - 17.1) |
| MDCD | 1,349 | 3,285 | 0.8 | 2.4 | 12 | 28 | 14.4 (7.45 - 25.2) | 11.6 (7.72 - 16.8) |
| Open Claims | 235,510 | 410,866 | 228.5 | 393.0 | 958 | 2,165 | 4.19 (3.93 - 4.47) | 5.51 (5.28 - 5.75) |
| OEHR | 16,549 | 31,819 | 5.4 | 16.4 | 22 | 99 | 4.08 (2.56 - 6.17) | 6.04 (4.91 - 7.36) |
| VA | 7,456 | 23,381 | 5.1 | 21.1 | 14 | 61 | 2.77 (1.51 - 4.64) | 2.89 (2.21 - 3.71) |
| SIDIAP | 4,136 | 19,402 | 6.0 | 44.0 | 14 | 124 | 2.34 (1.28 - 3.92) | 2.82 (2.34 - 3.36) |
| Meta-analysis | 296,736 | 541,707 | 272.9 | 521.3 | 1,165 | 2,821 | 4.27 (4.03 - 4.52) | 5.41 (5.21 - 5.61) |
| Meta-analysis (w/o USOC) | 61,226 | 130,841 | 44.4 | 128.3 | 207 | 656 | 4.66 (4.05 - 5.34) | 5.11 (4.73 - 5.52) |
| **PS-stratification / on-treatment TAR** | | | | | | | | |
| CCAE | 23,425 | 30,485 | 19.5 | 25.7 | 82 | 137 | 4.20 (3.34 - 5.22) | 5.33 (4.48 - 6.30) |
| DAG | 5,763 | 11,200 | 4.7 | 11.7 | 0 | 0 | | |
| ODOD | 16,779 | 20,863 | 11.7 | 16.4 | 101 | 174 | 8.66 (7.05 - 10.5) | 10.6 (9.07 - 12.3) |
| MDCR | 2,032 | 5,078 | 1.6 | 4.8 | 15 | 66 | 9.23 (5.16 - 15.2) | 13.7 (10.6 - 17.4) |
| MDCD | 1,420 | 3,306 | 0.8 | 2.4 | 12 | 29 | 14.1 (7.31 - 24.7) | 11.9 (7.99 - 17.1) |
| Open Claims | 316,207 | 411,152 | 276.1 | 393.2 | 1,187 | 2,173 | 4.30 (4.06 - 4.55) | 5.53 (5.30 - 5.76) |
| OEHR | 19,723 | 31,887 | 6.3 | 16.4 | 26 | 99 | 4.13 (2.70 - 6.05) | 6.03 (4.90 - 7.34) |
| VA | 11,651 | 24,407 | 7.0 | 22.7 | 35 | 63 | 4.98 (3.47 - 6.92) | 2.78 (2.14 - 3.56) |
| SIDIAP | 4,738 | 19,837 | 6.5 | 45.6 | 16 | 127 | 2.46 (1.41 - 4.00) | 2.79 (2.32 - 3.31) |
| Meta-analysis | 382,904 | 519,302 | 321.7 | 502.2 | 1,427 | 2,776 | 4.44 (4.21 - 4.67) | 5.53 (5.32 - 5.74) |
| Meta-analysis (w/o USOC) | 66,697 | 108,150 | 45.6 | 109.0 | 240 | 603 | 5.26 (4.62 - 5.97) | 5.53 (5.10 - 5.99) |
| **PS-matching / on-treatment TAR without escalation** | | | | | | | | |
| CCAE | 19,282 | 30,461 | 14.9 | 20.8 | 63 | 110 | 4.23 (3.25 - 5.41) | 5.29 (4.35 - 6.37) |
| DAG | 4,640 | 11,168 | 3.7 | 10.5 | 0 | 0 | | |
| ODOD | 12,151 | 20,752 | 8.1 | 13.4 | 65 | 138 | 7.98 (6.16 - 10.2) | 10.3 (8.67 - 12.2) |
| MDCR | 1,652 | 5,026 | 1.3 | 4.1 | 8 | 59 | 6.11 (2.64 - 12.0) | 14.6 (11.1 - 18.8) |
| MDCD | 1,349 | 3,285 | 0.7 | 1.8 | 12 | 26 | 17.1 (8.82 - 29.8) | 14.4 (9.40 - 21.1) |
| Open Claims | 235,510 | 410,866 | 191.6 | 305.0 | 791 | 1,667 | 4.13 (3.85 - 4.43) | 5.47 (5.21 - 5.74) |
| OEHR | 16,549 | 31,819 | 4.7 | 13.3 | 21 | 67 | 4.44 (2.75 - 6.79) | 5.05 (3.91 - 6.41) |
| VA | 7,456 | 23,381 | 4.3 | 17.2 | 13 | 46 | 3.02 (1.61 - 5.16) | 2.67 (1.96 - 3.56) |
| SIDIAP | 4,136 | 19,402 | 5.3 | 35.0 | 13 | 100 | 2.44 (1.30 - 4.18) | 2.86 (2.32 - 3.47) |
| Meta-analysis | 296,736 | 541,707 | 230.3 | 408.7 | 974 | 2,187 | 4.23 (3.97 - 4.50) | 5.35 (5.13 - 5.58) |
| Meta-analysis (w/o USOC) | 61,226 | 130,841 | 38.7 | 103.7 | 183 | 520 | 4.73 (4.07 - 5.46) | 5.01 (4.59 - 5.46) |
| **PS-stratification / on-treatment TAR without escalation** | | | | | | | | |
| CCAE | 23,425 | 30,485 | 17.0 | 20.8 | 70 | 110 | 4.11 (3.20 - 5.19) | 5.29 (4.34 - 6.37) |
| DAG | 5,763 | 11,200 | 4.3 | 10.6 | 0 | 0 | | |
| ODOD | 16,779 | 20,863 | 10.3 | 13.5 | 90 | 139 | 8.71 (7.00 - 10.7) | 10.3 (8.68 - 12.2) |
| MDCR | 2,032 | 5,078 | 1.5 | 4.1 | 14 | 60 | 9.64 (5.27 - 16.2) | 14.6 (11.2 - 18.8) |
| MDCD | 1,420 | 3,306 | 0.7 | 1.8 | 12 | 26 | 16.7 (8.65 - 29.2) | 14.3 (9.31 - 20.9) |
| Open Claims | 316,207 | 411,152 | 234.6 | 305.1 | 1,000 | 1,674 | 4.26 (4.00 - 4.54) | 5.49 (5.23 - 5.76) |
| OEHR | 19,723 | 31,887 | 5.5 | 13.3 | 25 | 67 | 4.51 (2.92 - 6.66) | 5.04 (3.91 - 6.40) |
| VA | 11,651 | 24,407 | 6.1 | 18.4 | 31 | 48 | 5.10 (3.46 - 7.24) | 2.61 (1.92 - 3.46) |
| SIDIAP | 4,738 | 19,837 | 5.8 | 36.2 | 15 | 101 | 2.59 (1.45 - 4.27) | 2.79 (2.27 - 3.39) |
| Meta-analysis | 382,904 | 519,302 | 274.8 | 393.0 | 1,214 | 2,151 | 4.42 (4.17 - 4.67) | 5.47 (5.24 - 5.71) |
| Meta-analysis (w/o USOC) | 66,697 | 108,150 | 40.2 | 87.9 | 214 | 477 | 5.33 (4.64 - 6.09) | 5.43 (4.95 - 5.94) |

##### 4.2.4 Acute renal failure

**Supplementary Table 319:** Exposures, outcomes and risk estimates for Acute renal failure in SGLT2I (male) (T) vs DPP4I (male) (C) new-user cohorts under four different propensity score (PS) adjustment / time-at-risk (TAR) designs across data sources. We report total patient exposure time in 1,000 patient-year (KYr) units, uncalibrated and calibrated hazard ratios (HRs) and their 95% confidence intervals (CIs). Greyed out entries did not pass study diagnostics and are included only for completeness.

| | Patients | | Exposure (KYr) | | Outcomes | | Incidence (per KYr) | |
|---|---|---|---|---|---|---|---|---|
| | T | C | T | C | T | C | T | C |
| **PS-matching / on-treatment TAR** | | | | | | | | |
| CCAE | 19,381 | 30,494 | 17.3 | 25.7 | 111 | 260 | 6.41 (5.27 - 7.72) | 10.1 (8.92 - 11.4) |
| DAG | 4,640 | 11,168 | 4.1 | 11.6 | 0 | 0 | | |
| ODOD | 11,987 | 20,055 | 9.2 | 15.7 | 152 | 513 | 16.4 (13.9 - 19.3) | 32.7 (30.0 - 35.7) |
| MDCR | 1,627 | 4,845 | 1.5 | 4.5 | 29 | 172 | 19.9 (13.3 - 28.6) | 38.1 (32.6 - 44.2) |
| MDCD | 1,337 | 3,140 | 0.8 | 2.3 | 17 | 81 | 20.2 (11.8 - 32.4) | 35.6 (28.3 - 44.3) |
| Open Claims | 236,488 | 408,696 | 230.3 | 390.0 | 1,787 | 5,910 | 7.76 (7.40 - 8.13) | 15.2 (14.8 - 15.5) |
| OEHR | 16,609 | 31,733 | 5.4 | 16.2 | 58 | 292 | 10.7 (8.10 - 13.8) | 18.0 (16.0 - 20.2) |
| VA | 7,454 | 23,043 | 5.1 | 20.8 | 53 | 265 | 10.4 (7.82 - 13.7) | 12.8 (11.3 - 14.4) |
| SIDIAP | 4,334 | 19,811 | 6.3 | 45.1 | <5 | 19 | < 0.79 (0.26 - 1.84) | 0.42 (0.25 - 0.66) |
| Meta-analysis | 293,546 | 518,866 | 268.8 | 472.9 | 2,190 | 7,412 | 8.15 (7.81 - 8.50) | 15.7 (15.3 - 16.0) |
| Meta-analysis (w/o USOC) | 57,058 | 110,170 | 38.5 | 82.9 | 403 | 1,502 | 10.5 (9.46 - 11.5) | 18.1 (17.2 - 19.1) |
| **PS-stratification / on-treatment TAR** | | | | | | | | |
| CCAE | 23,699 | 30,511 | 19.7 | 25.7 | 127 | 261 | 6.44 (5.37 - 7.66) | 10.1 (8.95 - 11.5) |
| DAG | 5,763 | 11,200 | 4.7 | 11.7 | 0 | 0 | | |
| ODOD | 16,747 | 20,115 | 11.7 | 15.7 | 199 | 521 | 17.0 (14.7 - 19.6) | 33.1 (30.3 - 36.1) |
| MDCR | 2,014 | 4,884 | 1.6 | 4.6 | 36 | 177 | 22.3 (15.6 - 30.8) | 38.8 (33.3 - 45.0) |
| MDCD | 1,409 | 3,155 | 0.9 | 2.3 | 17 | 83 | 19.9 (11.6 - 31.8) | 36.2 (28.9 - 44.9) |
| Open Claims | 320,733 | 408,870 | 280.5 | 390.1 | 2,329 | 5,925 | 8.30 (7.97 - 8.65) | 15.2 (14.8 - 15.6) |
| OEHR | 19,913 | 31,795 | 6.4 | 16.3 | 66 | 300 | 10.4 (8.03 - 13.2) | 18.5 (16.4 - 20.7) |
| VA | 11,731 | 24,015 | 7.1 | 22.2 | 93 | 284 | 13.1 (10.6 - 16.0) | 12.8 (11.4 - 14.4) |
| SIDIAP | 5,159 | 20,247 | 7.1 | 46.7 | <5 | 21 | < 0.71 (0.23 - 1.65) | 0.45 (0.28 - 0.69) |
| Meta-analysis | 388,265 | 516,422 | 326.9 | 499.1 | <2,762 | 7,205 | < 8.45 (8.14 - 8.77) | 14.4 (14.1 - 14.8) |
| Meta-analysis (w/o USOC) | 67,532 | 107,552 | 46.4 | 108.9 | <433 | 1,280 | < 9.32 (8.46 - 10.2) | 11.8 (11.1 - 12.4) |
| **PS-matching / on-treatment TAR without escalation** | | | | | | | | |
| CCAE | 19,381 | 30,494 | 15.0 | 20.8 | 89 | 214 | 5.92 (4.76 - 7.29) | 10.3 (8.94 - 11.7) |
| DAG | 4,640 | 11,168 | 3.7 | 10.5 | 0 | 0 | | |
| ODOD | 11,987 | 20,055 | 8.1 | 12.8 | 124 | 419 | 15.3 (12.7 - 18.2) | 32.7 (29.6 - 35.9) |
| MDCR | 1,627 | 4,845 | 1.3 | 3.8 | 25 | 146 | 19.2 (12.4 - 28.3) | 38.0 (32.1 - 44.7) |
| MDCD | 1,337 | 3,140 | 0.7 | 1.7 | 16 | 64 | 22.6 (12.9 - 36.6) | 37.7 (29.0 - 48.1) |
| Open Claims | 236,488 | 408,696 | 193.3 | 302.9 | 1,420 | 4,596 | 7.35 (6.97 - 7.74) | 15.2 (14.7 - 15.6) |
| OEHR | 16,609 | 31,733 | 4.8 | 13.2 | 40 | 206 | 8.39 (5.99 - 11.4) | 15.6 (13.5 - 17.9) |
| VA | 7,454 | 23,043 | 4.3 | 16.9 | 43 | 187 | 9.93 (7.18 - 13.4) | 11.0 (9.51 - 12.7) |
| SIDIAP | 4,334 | 19,811 | 5.6 | 35.8 | <5 | 13 | < 0.89 (0.29 - 2.08) | 0.36 (0.19 - 0.62) |
| Meta-analysis | 293,546 | 518,866 | 226.8 | 370.6 | 1,741 | 5,768 | 7.67 (7.32 - 8.04) | 15.6 (15.2 - 16.0) |
| Meta-analysis (w/o USOC) | 57,058 | 110,170 | 33.6 | 67.7 | 321 | 1,172 | 9.56 (8.55 - 10.7) | 17.3 (16.3 - 18.3) |
| **PS-stratification / on-treatment TAR without escalation** | | | | | | | | |
| CCAE | 23,699 | 30,511 | 17.3 | 20.8 | 102 | 215 | 5.91 (4.82 - 7.18) | 10.3 (8.98 - 11.8) |
| DAG | 5,763 | 11,200 | 4.3 | 10.6 | 0 | 0 | | |
| ODOD | 16,747 | 20,115 | 10.4 | 12.9 | 167 | 427 | 16.1 (13.7 - 18.7) | 33.2 (30.1 - 36.4) |
| MDCR | 2,014 | 4,884 | 1.5 | 3.9 | 32 | 150 | 22.0 (15.1 - 31.1) | 38.7 (32.7 - 45.4) |
| MDCD | 1,409 | 3,155 | 0.7 | 1.7 | 16 | 65 | 22.1 (12.7 - 36.0) | 37.9 (29.3 - 48.3) |
| Open Claims | 320,733 | 408,870 | 238.7 | 303.0 | 1,920 | 4,609 | 8.05 (7.69 - 8.41) | 15.2 (14.8 - 15.7) |
| OEHR | 19,913 | 31,795 | 5.6 | 13.2 | 46 | 211 | 8.20 (6.01 - 10.9) | 16.0 (13.9 - 18.3) |
| VA | 11,731 | 24,015 | 6.2 | 18.0 | 80 | 201 | 13.0 (10.3 - 16.2) | 11.1 (9.65 - 12.8) |
| SIDIAP | 5,159 | 20,247 | 6.3 | 37.0 | <5 | 13 | < 0.79 (0.26 - 1.85) | 0.35 (0.19 - 0.60) |
| Meta-analysis | 383,106 | 496,175 | 273.3 | 353.9 | 2,267 | 5,612 | 8.29 (7.96 - 8.64) | 15.9 (15.4 - 16.3) |
| Meta-analysis (w/o USOC) | 62,373 | 87,305 | 34.7 | 50.8 | 347 | 1,003 | 10.0 (8.98 - 11.1) | 19.7 (18.5 - 21.0) |

##### 4.2.5 Glycemic control

**Supplementary Table 320:** Exposures, outcomes and risk estimates for Glycemic control in SGLT2I (male) (T) vs DPP4I (male) (C) new-user cohorts under four different propensity score (PS) adjustment / time-at-risk (TAR) designs across data sources. We report total patient exposure time in 1,000 patient-year (KYr) units, uncalibrated and calibrated hazard ratios (HRs) and their 95% confidence intervals (CIs). Greyed out entries did not pass study diagnostics and are included only for completeness.

| | Patients | | Exposure (KYr) | | Outcomes | | Incidence (per KYr) | |
|---|---|---|---|---|---|---|---|---|
| | T | C | T | C | T | C | T | C |
| **PS-matching / on-treatment TAR** | | | | | | | | |
| CCAE | 19,610 | 30,810 | 17.5 | 26.1 | 26 | 40 | 1.48 (0.97 - 2.17) | 1.53 (1.10 - 2.09) |
| DAG | 1,654 | 4,152 | 1.2 | 3.7 | 180 | 560 | 146 (125 - 169) | 153 (141 - 167) |
| ODOD | 8,133 | 13,765 | 5.6 | 9.6 | 611 | 1,079 | 109 (100 - 117) | 112 (105 - 119) |
| MDCR | 1,714 | 5,229 | 1.5 | 5.0 | 0 | 5 | | 1.01 (0.33 - 2.35) |
| MDCD | 1,426 | 3,470 | 0.9 | 2.6 | 0 | 0 | | |
| Open Claims | 244,206 | 426,386 | 237.2 | 409.0 | 0 | 0 | | |
| OEHR | 16,869 | 32,344 | 5.5 | 16.7 | 0 | 0 | | |
| VA | 7,441 | 23,209 | 5.1 | 21.0 | <10 | 21 | < 1.98 (0.95 - 3.64) | 1.00 (0.62 - 1.53) |
| SIDIAP | 496 | 2,248 | 0.5 | 3.2 | 162 | 832 | 333 (284 - 388) | 258 (241 - 276) |
| Meta-analysis | 37,334 | 74,184 | 29.9 | 63.6 | <989 | 2,532 | < 33.0 (31.0 - 35.2) | 39.8 (38.3 - 41.4) |
| Meta-analysis (w/o USOC) | 37,334 | 74,184 | 29.9 | 63.6 | <989 | 2,532 | < 33.0 (31.0 - 35.2) | 39.8 (38.3 - 41.4) |
| **PS-stratification / on-treatment TAR** | | | | | | | | |
| CCAE | 23,976 | 30,829 | 20.0 | 26.1 | 26 | 40 | 1.30 (0.85 - 1.91) | 1.53 (1.10 - 2.09) |
| DAG | 1,994 | 4,184 | 1.4 | 3.7 | 210 | 560 | 148 (128 - 169) | 152 (140 - 165) |
| ODOD | 11,471 | 13,903 | 7.3 | 9.7 | 761 | 1,088 | 105 (97.3 - 112) | 112 (105 - 118) |
| MDCR | 2,128 | 5,285 | 1.7 | 5.0 | 0 | 5 | | 1.00 (0.32 - 2.33) |
| MDCD | 1,507 | 3,484 | 0.9 | 2.6 | 0 | 0 | | |
| Open Claims | 332,801 | 426,715 | 289.7 | 409.2 | 0 | 0 | | |
| OEHR | 20,231 | 32,415 | 6.5 | 16.8 | 0 | 0 | | |
| VA | 11,946 | 24,227 | 7.2 | 22.5 | 12 | 21 | 1.67 (0.86 - 2.92) | 0.93 (0.58 - 1.42) |
| SIDIAP | 541 | 2,295 | 0.5 | 3.3 | 180 | 857 | 348 (299 - 403) | 259 (242 - 277) |
| Meta-analysis | 37,982 | 51,211 | 29.2 | 42.8 | 1,177 | 2,545 | 40.3 (38.1 - 42.7) | 59.4 (57.2 - 61.8) |
| Meta-analysis (w/o USOC) | 37,982 | 51,211 | 29.2 | 42.8 | 1,177 | 2,545 | 40.3 (38.1 - 42.7) | 59.4 (57.2 - 61.8) |
| **PS-matching / on-treatment TAR without escalation** | | | | | | | | |
| CCAE | 19,610 | 30,810 | 15.2 | 21.1 | 25 | 33 | 1.65 (1.07 - 2.43) | 1.57 (1.08 - 2.20) |
| DAG | 1,654 | 4,152 | 1.1 | 3.3 | 157 | 507 | 137 (117 - 160) | 153 (140 - 167) |
| ODOD | 8,133 | 13,765 | 5.0 | 7.9 | 532 | 871 | 107 (97.9 - 116) | 111 (103 - 118) |
| MDCR | 1,714 | 5,229 | 1.4 | 4.2 | 0 | 5 | | 1.19 (0.39 - 2.77) |
| MDCD | 1,426 | 3,470 | 0.7 | 1.9 | 0 | 0 | | |
| Open Claims | 244,206 | 426,386 | 198.9 | 317.1 | 0 | 0 | | |
| OEHR | 16,869 | 32,344 | 4.8 | 13.5 | 0 | 0 | | |
| VA | 7,441 | 23,209 | 4.3 | 17.2 | <10 | 18 | < 2.32 (1.11 - 4.27) | 1.05 (0.62 - 1.66) |
| SIDIAP | 496 | 2,248 | 0.4 | 2.5 | 150 | 656 | 357 (302 - 419) | 266 (246 - 287) |
| Meta-analysis | 37,334 | 74,184 | 26.0 | 51.9 | <874 | 2,085 | < 33.6 (31.4 - 35.9) | 40.2 (38.5 - 41.9) |
| Meta-analysis (w/o USOC) | 37,334 | 74,184 | 26.0 | 51.9 | <874 | 2,085 | < 33.6 (31.4 - 35.9) | 40.2 (38.5 - 41.9) |
| **PS-stratification / on-treatment TAR without escalation** | | | | | | | | |
| CCAE | 23,976 | 30,829 | 17.4 | 21.1 | 25 | 33 | 1.44 (0.93 - 2.12) | 1.56 (1.08 - 2.20) |
| DAG | 1,994 | 4,184 | 1.3 | 3.3 | 185 | 507 | 140 (121 - 162) | 151 (139 - 165) |
| ODOD | 11,471 | 13,903 | 6.5 | 8.0 | 669 | 878 | 103 (95.4 - 111) | 110 (103 - 118) |
| MDCR | 2,128 | 5,285 | 1.5 | 4.3 | 0 | 5 | | 1.17 (0.38 - 2.74) |
| MDCD | 1,507 | 3,484 | 0.8 | 1.9 | 0 | 0 | | |
| Open Claims | 332,801 | 426,715 | 246.2 | 317.3 | 0 | 0 | | |
| OEHR | 20,231 | 32,415 | 5.7 | 13.6 | 0 | 0 | | |
| VA | 11,946 | 24,227 | 6.2 | 18.3 | <10 | 18 | < 1.61 (0.77 - 2.96) | 0.98 (0.58 - 1.55) |
| SIDIAP | 541 | 2,295 | 0.4 | 2.5 | 165 | 677 | 371 (317 - 432) | 268 (248 - 289) |
| Meta-analysis | 37,982 | 51,211 | 25.7 | 34.9 | 1,044 | 2,095 | 40.7 (38.2 - 43.2) | 60.0 (57.4 - 62.6) |
| Meta-analysis (w/o USOC) | 37,982 | 51,211 | 25.7 | 34.9 | 1,044 | 2,095 | 40.7 (38.2 - 43.2) | 60.0 (57.4 - 62.6) |

##### 4.2.6 Hospitalization with heart failure

**Supplementary Table 321:** Exposures, outcomes and risk estimates for Hospitalization with heart failure in SGLT2I (male) (T) vs DPP4I (male) (C) new-user cohorts under four different propensity score (PS) adjustment / time-at-risk (TAR) designs across data sources. We report total patient exposure time in 1,000 patient-year (KYr) units, uncalibrated and calibrated hazard ratios (HRs) and their 95% confidence intervals (CIs). Greyed out entries did not pass study diagnostics and are included only for completeness.

| | Patients | | Exposure (KYr) | | Outcomes | | Incidence (per KYr) | |
|---|---|---|---|---|---|---|---|---|
| | T | C | T | C | T | C | T | C |
| **PS-matching / on-treatment TAR** | | | | | | | | |
| CCAE | 19,363 | 30,576 | 17.3 | 25.8 | 96 | 151 | 5.55 (4.50 - 6.78) | 5.85 (4.95 - 6.86) |
| DAG | 4,640 | 11,168 | 4.1 | 11.6 | 0 | 0 | | |
| ODOD | 11,932 | 20,118 | 9.2 | 15.8 | 117 | 344 | 12.7 (10.5 - 15.3) | 21.8 (19.5 - 24.2) |
| MDCR | 1,605 | 4,825 | 1.4 | 4.6 | 28 | 137 | 19.6 (13.0 - 28.4) | 30.1 (25.3 - 35.6) |
| MDCD | 1,287 | 3,127 | 0.8 | 2.3 | 11 | 41 | 14.0 (6.99 - 25.1) | 18.1 (13.0 - 24.6) |
| Open Claims | 235,537 | 407,798 | 229.8 | 390.2 | 1,198 | 4,114 | 5.21 (4.92 - 5.52) | 10.5 (10.2 - 10.9) |
| OEHR | 16,513 | 31,579 | 5.4 | 16.1 | 50 | 202 | 9.25 (6.86 - 12.2) | 12.5 (10.9 - 14.4) |
| VA | 7,403 | 23,147 | 5.0 | 20.9 | 31 | 188 | 6.16 (4.18 - 8.74) | 9.01 (7.77 - 10.4) |
| SIDIAP | 4,309 | 19,733 | 6.3 | 44.9 | 5 | 70 | 0.79 (0.26 - 1.85) | 1.56 (1.22 - 1.97) |
| Meta-analysis | 296,662 | 537,776 | 274.4 | 518.3 | 1,525 | 5,206 | 5.56 (5.28 - 5.84) | 10.0 (9.77 - 10.3) |
| Meta-analysis (w/o USOC) | 61,125 | 129,978 | 44.6 | 128.1 | 327 | 1,092 | 7.33 (6.55 - 8.16) | 8.53 (8.03 - 9.05) |
| **PS-stratification / on-treatment TAR** | | | | | | | | |
| CCAE | 23,598 | 30,597 | 19.7 | 25.8 | 113 | 151 | 5.75 (4.74 - 6.91) | 5.84 (4.95 - 6.85) |
| DAG | 5,763 | 11,200 | 4.7 | 11.7 | 0 | 0 | | |
| ODOD | 16,421 | 20,195 | 11.5 | 15.9 | 163 | 349 | 14.2 (12.1 - 16.5) | 22.0 (19.7 - 24.4) |
| MDCR | 1,962 | 4,867 | 1.6 | 4.6 | 33 | 140 | 20.9 (14.4 - 29.4) | 30.4 (25.6 - 35.9) |
| MDCD | 1,350 | 3,145 | 0.8 | 2.3 | 11 | 42 | 13.7 (6.86 - 24.6) | 18.4 (13.3 - 24.9) |
| Open Claims | 315,211 | 408,030 | 277.3 | 390.3 | 1,631 | 4,137 | 5.88 (5.60 - 6.17) | 10.6 (10.3 - 10.9) |
| OEHR | 19,700 | 31,638 | 6.3 | 16.2 | 64 | 205 | 10.2 (7.83 - 13.0) | 12.7 (11.0 - 14.5) |
| VA | 11,226 | 24,158 | 6.8 | 22.3 | 73 | 194 | 10.7 (8.37 - 13.4) | 8.68 (7.50 - 9.99) |
| SIDIAP | 5,095 | 20,166 | 7.0 | 46.5 | 5 | 77 | 0.72 (0.23 - 1.67) | 1.66 (1.31 - 2.07) |
| Meta-analysis | 381,987 | 515,493 | 323.4 | 499.3 | 2,009 | 5,059 | 6.21 (5.94 - 6.49) | 10.1 (9.85 - 10.4) |
| Meta-analysis (w/o USOC) | 66,776 | 107,463 | 46.0 | 109.0 | 378 | 922 | 8.21 (7.40 - 9.08) | 8.46 (7.92 - 9.03) |
| **PS-matching / on-treatment TAR without escalation** | | | | | | | | |
| CCAE | 19,363 | 30,576 | 15.0 | 20.9 | 79 | 125 | 5.27 (4.17 - 6.57) | 5.98 (4.98 - 7.12) |
| DAG | 4,640 | 11,168 | 3.7 | 10.5 | 0 | 0 | | |
| ODOD | 11,932 | 20,118 | 8.1 | 12.9 | 99 | 291 | 12.3 (9.97 - 14.9) | 22.5 (20.0 - 25.2) |
| MDCR | 1,605 | 4,825 | 1.3 | 3.9 | 24 | 123 | 18.9 (12.1 - 28.1) | 31.8 (26.5 - 38.0) |
| MDCD | 1,287 | 3,127 | 0.7 | 1.7 | 10 | 33 | 15.0 (7.22 - 27.7) | 19.4 (13.4 - 27.2) |
| Open Claims | 235,537 | 407,798 | 192.8 | 302.8 | 969 | 3,212 | 5.03 (4.71 - 5.35) | 10.6 (10.2 - 11.0) |
| OEHR | 16,513 | 31,579 | 4.7 | 13.1 | 37 | 147 | 7.81 (5.50 - 10.8) | 11.2 (9.47 - 13.2) |
| VA | 7,403 | 23,147 | 4.3 | 17.0 | 27 | 147 | 6.28 (4.14 - 9.14) | 8.64 (7.30 - 10.2) |
| SIDIAP | 4,309 | 19,733 | 5.6 | 35.7 | <5 | 52 | < 0.89 (0.29 - 2.08) | 1.46 (1.09 - 1.91) |
| Meta-analysis | 296,662 | 537,776 | 231.8 | 406.3 | <1,240 | 4,097 | < 5.35 (5.06 - 5.66) | 10.1 (9.78 - 10.4) |
| Meta-analysis (w/o USOC) | 61,125 | 129,978 | 39.0 | 103.5 | <271 | 885 | < 6.96 (6.15 - 7.84) | 8.55 (8.00 - 9.14) |
| **PS-stratification / on-treatment TAR without escalation** | | | | | | | | |
| CCAE | 23,598 | 30,597 | 17.2 | 20.9 | 93 | 125 | 5.41 (4.37 - 6.63) | 5.98 (4.98 - 7.12) |
| DAG | 5,763 | 11,200 | 4.3 | 10.6 | 0 | 0 | | |
| ODOD | 16,421 | 20,195 | 10.2 | 13.0 | 144 | 295 | 14.1 (11.9 - 16.6) | 22.7 (20.2 - 25.4) |
| MDCR | 1,962 | 4,867 | 1.4 | 3.9 | 28 | 126 | 19.9 (13.2 - 28.7) | 32.3 (26.9 - 38.4) |
| MDCD | 1,350 | 3,145 | 0.7 | 1.7 | 10 | 34 | 14.8 (7.07 - 27.1) | 19.8 (13.7 - 27.7) |
| Open Claims | 315,211 | 408,030 | 235.8 | 302.9 | 1,369 | 3,233 | 5.81 (5.50 - 6.12) | 10.7 (10.3 - 11.0) |
| OEHR | 19,700 | 31,638 | 5.5 | 13.1 | 50 | 148 | 9.02 (6.70 - 11.9) | 11.3 (9.53 - 13.2) |
| VA | 11,226 | 24,158 | 5.9 | 18.2 | 66 | 153 | 11.2 (8.64 - 14.2) | 8.42 (7.14 - 9.87) |
| SIDIAP | 5,095 | 20,166 | 6.2 | 36.8 | <5 | 58 | < 0.80 (0.26 - 1.87) | 1.57 (1.20 - 2.03) |
| Meta-analysis | 381,987 | 515,493 | 276.4 | 390.7 | <1,689 | 3,985 | < 6.11 (5.82 - 6.41) | 10.2 (9.88 - 10.5) |
| Meta-analysis (w/o USOC) | 66,776 | 107,463 | 40.6 | 87.8 | <320 | 752 | < 7.88 (7.04 - 8.80) | 8.57 (7.96 - 9.20) |

##### 4.2.7 Measured renal dysfunction

**Supplementary Table 322:** Exposures, outcomes and risk estimates for Measured renal dysfunction in SGLT2I (male) (T) vs DPP4I (male) (C) new-user cohorts under four different propensity score (PS) adjustment / time-at-risk (TAR) designs across data sources. We report total patient exposure time in 1,000 patient-year (KYr) units, uncalibrated and calibrated hazard ratios (HRs) and their 95% confidence intervals (CIs). Greyed out entries did not pass study diagnostics and are included only for completeness.

| | Patients | | Exposure (KYr) | | Outcomes | | Incidence (per KYr) | |
|---|---|---|---|---|---|---|---|---|
| | T | C | T | C | T | C | T | C |
| **PS-matching / on-treatment TAR** | | | | | | | | |
| CCAE | 19,702 | 31,115 | 17.6 | 26.3 | 5 | 11 | 0.28 (0.09 - 0.66) | 0.42 (0.21 - 0.75) |
| DAG | 4,640 | 11,168 | 4.1 | 11.6 | 0 | 0 | | |
| ODOD | 12,499 | 21,418 | 9.5 | 16.9 | 6 | 42 | 0.63 (0.23 - 1.37) | 2.49 (1.79 - 3.37) |
| MDCR | 1,721 | 5,267 | 1.5 | 5.0 | 0 | <5 | | < 1.00 (0.33 - 2.34) |
| MDCD | 1,426 | 3,470 | 0.9 | 2.6 | 0 | 0 | | |
| Open Claims | 244,206 | 426,386 | 237.2 | 409.0 | 0 | 0 | | |
| OEHR | 16,869 | 32,344 | 5.5 | 16.7 | 0 | 0 | | |
| VA | 7,710 | 23,670 | 5.3 | 21.4 | 10 | 89 | 1.90 (0.91 - 3.49) | 4.16 (3.34 - 5.12) |
| SIDIAP | 4,322 | 19,658 | 6.3 | 44.5 | 13 | 276 | 2.06 (1.10 - 3.52) | 6.20 (5.49 - 6.98) |
| Meta-analysis | 24,531 | 64,746 | 21.1 | 82.8 | 29 | 407 | 1.37 (0.92 - 1.97) | 4.92 (4.45 - 5.42) |
| Meta-analysis (w/o USOC) | 24,531 | 64,746 | 21.1 | 82.8 | 29 | 407 | 1.37 (0.92 - 1.97) | 4.92 (4.45 - 5.42) |
| **PS-stratification / on-treatment TAR** | | | | | | | | |
| CCAE | 24,082 | 31,138 | 20.0 | 26.3 | 5 | 11 | 0.25 (0.08 - 0.58) | 0.42 (0.21 - 0.75) |
| DAG | 5,763 | 11,200 | 4.7 | 11.7 | 0 | 0 | | |
| ODOD | 17,471 | 21,538 | 12.1 | 17.0 | 7 | 44 | 0.58 (0.23 - 1.19) | 2.59 (1.88 - 3.48) |
| MDCR | 2,136 | 5,326 | 1.7 | 5.0 | 0 | <5 | | < 0.99 (0.32 - 2.31) |
| MDCD | 1,507 | 3,484 | 0.9 | 2.6 | 0 | 0 | | |
| Open Claims | 332,801 | 426,715 | 289.7 | 409.2 | 0 | 0 | | |
| OEHR | 20,231 | 32,415 | 6.5 | 16.8 | 0 | 0 | | |
| VA | 12,350 | 24,682 | 7.4 | 22.9 | 23 | 95 | 3.09 (1.96 - 4.64) | 4.15 (3.36 - 5.08) |
| SIDIAP | 5,147 | 20,075 | 7.0 | 46.0 | 16 | 299 | 2.27 (1.30 - 3.69) | 6.50 (5.79 - 7.28) |
| Meta-analysis | 22,618 | 41,613 | 19.2 | 62.9 | 23 | 343 | 1.20 (0.76 - 1.80) | 5.45 (4.89 - 6.06) |
| Meta-analysis (w/o USOC) | 22,618 | 41,613 | 19.2 | 62.9 | 23 | 343 | 1.20 (0.76 - 1.80) | 5.45 (4.89 - 6.06) |
| **PS-matching / on-treatment TAR without escalation** | | | | | | | | |
| CCAE | 19,702 | 31,115 | 15.2 | 21.3 | 5 | 10 | 0.33 (0.11 - 0.77) | 0.47 (0.23 - 0.86) |
| DAG | 4,640 | 11,168 | 3.7 | 10.5 | 0 | 0 | | |
| ODOD | 12,499 | 21,418 | 8.4 | 13.8 | 6 | 37 | 0.72 (0.26 - 1.56) | 2.68 (1.89 - 3.70) |
| MDCR | 1,721 | 5,267 | 1.4 | 4.2 | 0 | 0 | | |
| MDCD | 1,426 | 3,470 | 0.7 | 1.9 | 0 | 0 | | |
| Open Claims | 244,206 | 426,386 | 198.9 | 317.1 | 0 | 0 | | |
| OEHR | 16,869 | 32,344 | 4.8 | 13.5 | 0 | 0 | | |
| VA | 7,710 | 23,670 | 4.5 | 17.4 | <10 | 69 | < 2.23 (1.07 - 4.10) | 3.96 (3.08 - 5.01) |
| SIDIAP | 4,322 | 19,658 | 5.6 | 35.3 | 11 | 230 | 1.96 (0.98 - 3.51) | 6.51 (5.70 - 7.41) |
| Meta-analysis | 24,531 | 64,746 | 18.5 | 66.5 | <27 | 336 | < 1.46 (0.96 - 2.13) | 5.05 (4.52 - 5.62) |
| Meta-analysis (w/o USOC) | 24,531 | 64,746 | 18.5 | 66.5 | <27 | 336 | < 1.46 (0.96 - 2.13) | 5.05 (4.52 - 5.62) |
| **PS-stratification / on-treatment TAR without escalation** | | | | | | | | |
| CCAE | 24,082 | 31,138 | 17.5 | 21.3 | 5 | 10 | 0.29 (0.09 - 0.67) | 0.47 (0.23 - 0.86) |
| DAG | 5,763 | 11,200 | 4.3 | 10.6 | 0 | 0 | | |
| ODOD | 17,471 | 21,538 | 10.8 | 13.9 | 7 | 39 | 0.65 (0.26 - 1.34) | 2.81 (2.00 - 3.84) |
| MDCR | 2,136 | 5,326 | 1.5 | 4.3 | 0 | 0 | | |
| MDCD | 1,507 | 3,484 | 0.8 | 1.9 | 0 | 0 | | |
| Open Claims | 332,801 | 426,715 | 246.2 | 317.3 | 0 | 0 | | |
| OEHR | 20,231 | 32,415 | 5.7 | 13.6 | 0 | 0 | | |
| VA | 12,350 | 24,682 | 6.4 | 18.6 | 18 | 75 | 2.79 (1.65 - 4.41) | 4.04 (3.18 - 5.06) |
| SIDIAP | 5,147 | 20,075 | 6.3 | 36.4 | 14 | 246 | 2.23 (1.22 - 3.74) | 6.75 (5.94 - 7.65) |
| Meta-analysis | 22,618 | 41,613 | 17.0 | 50.3 | 21 | 285 | 1.23 (0.76 - 1.88) | 5.67 (5.03 - 6.36) |
| Meta-analysis (w/o USOC) | 22,618 | 41,613 | 17.0 | 50.3 | 21 | 285 | 1.23 (0.76 - 1.88) | 5.67 (5.03 - 6.36) |

###### 4.2.8 Stroke

**Supplementary Table 323:** Exposures, outcomes and risk estimates for Stroke in SGLT2I (male) (T) vs DPP4I (male) (C) new-user cohorts under four different propensity score (PS) adjustment / time-at-risk (TAR) designs across data sources. We report total patient exposure time in 1,000 patient-year (KYr) units, uncalibrated and calibrated hazard ratios (HRs) and their 95% confidence intervals (CIs). Greyed out entries did not pass study diagnostics and are included only for completeness.

| | Patients | | Exposure (KYr) | | Outcomes | | Incidence (per KYr) | |
|---|---|---|---|---|---|---|---|---|
| | T | C | T | C | T | C | T | C |
| **PS-matching / on-treatment TAR** | | | | | | | | |
| CCAE | 19,517 | 30,782 | 17.4 | 26.0 | 51 | 93 | 2.92 (2.18 - 3.84) | 3.57 (2.88 - 4.38) |
| DAG | 4,640 | 11,168 | 4.1 | 11.6 | 0 | 0 | | |
| ODOD | 12,233 | 20,802 | 9.3 | 16.4 | 67 | 161 | 7.20 (5.58 - 9.15) | 9.83 (8.37 - 11.5) |
| MDCR | 1,670 | 5,029 | 1.5 | 4.7 | 16 | 60 | 10.8 (6.16 - 17.5) | 12.7 (9.68 - 16.3) |
| MDCD | 1,375 | 3,307 | 0.9 | 2.4 | 6 | 26 | 7.06 (2.59 - 15.4) | 10.8 (7.02 - 15.8) |
| Open Claims | 238,515 | 414,277 | 232.2 | 397.2 | 781 | 2,049 | 3.36 (3.13 - 3.61) | 5.16 (4.94 - 5.39) |
| OEHR | 16,716 | 32,022 | 5.5 | 16.5 | 18 | 72 | 3.30 (1.95 - 5.21) | 4.36 (3.41 - 5.49) |
| VA | 7,643 | 23,577 | 5.2 | 21.4 | 11 | 55 | 2.10 (1.05 - 3.77) | 2.57 (1.94 - 3.35) |
| SIDIAP | 4,251 | 19,534 | 6.2 | 44.4 | 13 | 131 | 2.09 (1.11 - 3.58) | 2.95 (2.47 - 3.50) |
| Meta-analysis | 300,545 | 546,023 | 277.3 | 526.6 | 957 | 2,621 | 3.45 (3.24 - 3.68) | 4.98 (4.79 - 5.17) |
| Meta-analysis (w/o USOC) | 62,030 | 131,746 | 45.1 | 129.4 | 176 | 572 | 3.90 (3.34 - 4.52) | 4.42 (4.07 - 4.80) |
| **PS-stratification / on-treatment TAR** | | | | | | | | |
| CCAE | 23,843 | 30,803 | 19.9 | 26.0 | 58 | 93 | 2.92 (2.22 - 3.77) | 3.57 (2.88 - 4.37) |
| DAG | 5,763 | 11,200 | 4.7 | 11.7 | 0 | 0 | | |
| ODOD | 17,093 | 20,912 | 11.8 | 16.5 | 81 | 167 | 6.85 (5.44 - 8.51) | 10.1 (8.66 - 11.8) |
| MDCR | 2,074 | 5,079 | 1.7 | 4.8 | 18 | 60 | 10.8 (6.43 - 17.1) | 12.5 (9.57 - 16.1) |
| MDCD | 1,446 | 3,329 | 0.9 | 2.4 | 6 | 27 | 6.93 (2.54 - 15.1) | 11.1 (7.29 - 16.1) |
| Open Claims | 324,481 | 414,551 | 283.2 | 397.4 | 934 | 2,055 | 3.30 (3.09 - 3.52) | 5.17 (4.95 - 5.40) |
| OEHR | 20,037 | 32,091 | 6.4 | 16.6 | 19 | 73 | 2.97 (1.79 - 4.63) | 4.41 (3.45 - 5.54) |
| VA | 12,216 | 24,595 | 7.4 | 22.9 | 19 | 63 | 2.58 (1.55 - 4.02) | 2.76 (2.12 - 3.53) |
| SIDIAP | 5,058 | 19,960 | 6.9 | 45.9 | 14 | 141 | 2.02 (1.10 - 3.39) | 3.07 (2.59 - 3.62) |
| Meta-analysis | 392,586 | 523,396 | 329.9 | 507.2 | 1,124 | 2,589 | 3.41 (3.21 - 3.61) | 5.10 (4.91 - 5.31) |
| Meta-analysis (w/o USOC) | 68,105 | 108,845 | 46.7 | 109.8 | 190 | 534 | 4.07 (3.51 - 4.69) | 4.86 (4.46 - 5.30) |
| **PS-matching / on-treatment TAR without escalation** | | | | | | | | |
| CCAE | 19,517 | 30,782 | 15.1 | 21.1 | 41 | 73 | 2.71 (1.95 - 3.68) | 3.46 (2.72 - 4.36) |
| DAG | 4,640 | 11,168 | 3.7 | 10.5 | 0 | 0 | | |
| ODOD | 12,233 | 20,802 | 8.2 | 13.4 | 59 | 138 | 7.21 (5.49 - 9.31) | 10.3 (8.66 - 12.2) |
| MDCR | 1,670 | 5,029 | 1.3 | 4.0 | 13 | 50 | 9.86 (5.25 - 16.9) | 12.4 (9.23 - 16.4) |
| MDCD | 1,375 | 3,307 | 0.7 | 1.8 | <5 | 19 | < 7.00 (2.27 - 16.3) | 10.5 (6.33 - 16.4) |
| Open Claims | 238,515 | 414,277 | 194.7 | 308.1 | 664 | 1,629 | 3.41 (3.16 - 3.68) | 5.29 (5.03 - 5.55) |
| OEHR | 16,716 | 32,022 | 4.8 | 13.4 | 11 | 53 | 2.30 (1.15 - 4.12) | 3.97 (2.97 - 5.19) |
| VA | 7,643 | 23,577 | 4.5 | 17.4 | <10 | 36 | < 2.25 (1.08 - 4.13) | 2.07 (1.45 - 2.86) |
| SIDIAP | 4,251 | 19,534 | 5.5 | 35.2 | 12 | 96 | 2.17 (1.12 - 3.80) | 2.72 (2.21 - 3.33) |
| Meta-analysis | 300,545 | 546,023 | 234.1 | 412.6 | <810 | 2,075 | < 3.46 (3.23 - 3.71) | 5.03 (4.81 - 5.25) |
| Meta-analysis (w/o USOC) | 62,030 | 131,746 | 39.4 | 104.5 | <146 | 446 | < 3.71 (3.13 - 4.36) | 4.27 (3.88 - 4.68) |
| **PS-stratification / on-treatment TAR without escalation** | | | | | | | | |
| CCAE | 23,843 | 30,803 | 17.3 | 21.1 | 46 | 73 | 2.65 (1.94 - 3.54) | 3.46 (2.71 - 4.35) |
| DAG | 5,763 | 11,200 | 4.3 | 10.6 | 0 | 0 | | |
| ODOD | 17,093 | 20,912 | 10.5 | 13.5 | 72 | 144 | 6.85 (5.36 - 8.63) | 10.7 (9.01 - 12.6) |
| MDCR | 2,074 | 5,079 | 1.5 | 4.1 | 15 | 50 | 10.1 (5.67 - 16.7) | 12.3 (9.12 - 16.2) |
| MDCD | 1,446 | 3,329 | 0.7 | 1.8 | <5 | 19 | < 6.85 (2.22 - 16.0) | 10.4 (6.25 - 16.2) |
| Open Claims | 324,481 | 414,551 | 240.8 | 308.3 | 806 | 1,634 | 3.35 (3.12 - 3.59) | 5.30 (5.05 - 5.56) |
| OEHR | 20,037 | 32,091 | 5.6 | 13.4 | 12 | 53 | 2.13 (1.10 - 3.72) | 3.96 (2.96 - 5.18) |
| VA | 12,216 | 24,595 | 6.4 | 18.6 | 14 | 41 | 2.19 (1.20 - 3.68) | 2.21 (1.58 - 2.99) |
| SIDIAP | 5,058 | 19,960 | 6.2 | 36.4 | 13 | 104 | 2.11 (1.12 - 3.60) | 2.86 (2.33 - 3.46) |
| Meta-analysis | 392,586 | 523,396 | 281.9 | 396.7 | 964 | 2,058 | 3.42 (3.21 - 3.64) | 5.19 (4.97 - 5.42) |
| Meta-analysis (w/o USOC) | 68,105 | 108,845 | 41.2 | 88.4 | 158 | 424 | 3.84 (3.26 - 4.49) | 4.80 (4.35 - 5.27) |

##### 4.2.9 Sudden cardiac death

**Supplementary Table 324:** Exposures, outcomes and risk estimates for Sudden cardiac death in SGLT2I (male) (T) vs DPP4I (male) (C) new-user cohorts under four different propensity score (PS) adjustment / time-at-risk (TAR) designs across data sources. We report total patient exposure time in 1,000 patient-year (KYr) units, uncalibrated and calibrated hazard ratios (HRs) and their 95% confidence intervals (CIs). Greyed out entries did not pass study diagnostics and are included only for completeness.

| | Patients | | Exposure (KYr) | | Outcomes | | Incidence (per KYr) | |
|---|---|---|---|---|---|---|---|---|
| | T | C | T | C | T | C | T | C |
| **PS-matching / on-treatment TAR** | | | | | | | | |
| CCAE | 19,730 | 31,143 | 17.6 | 26.4 | 15 | 27 | 0.85 (0.48 - 1.40) | 1.02 (0.67 - 1.49) |
| DAG | 4,640 | 11,168 | 4.1 | 11.6 | 0 | 0 | | |
| ODOD | 12,497 | 21,421 | 9.6 | 16.9 | 15 | 70 | 1.57 (0.88 - 2.59) | 4.15 (3.23 - 5.24) |
| MDCR | 1,715 | 5,246 | 1.5 | 5.0 | 9 | 21 | 5.91 (2.70 - 11.2) | 4.23 (2.62 - 6.47) |
| MDCD | 1,419 | 3,454 | 0.9 | 2.5 | <5 | 8 | < 5.67 (1.84 - 13.2) | 3.14 (1.36 - 6.19) |
| Open Claims | 243,645 | 425,371 | 236.7 | 407.8 | 188 | 643 | 0.79 (0.68 - 0.92) | 1.58 (1.46 - 1.70) |
| OEHR | 16,851 | 32,316 | 5.5 | 16.7 | <5 | 22 | < 0.91 (0.29 - 2.12) | 1.32 (0.82 - 1.99) |
| VA | 7,788 | 23,941 | 5.3 | 21.7 | 0 | <10 | | < 0.46 (0.22 - 0.85) |
| SIDIAP | 4,338 | 19,828 | 6.3 | 45.1 | 0 | <5 | | < 0.11 (0.04 - 0.26) |
| Meta-analysis | 294,438 | 515,497 | 270.9 | 472.7 | <232 | 783 | < 0.86 (0.75 - 0.97) | 1.66 (1.54 - 1.78) |
| Meta-analysis (w/o USOC) | 50,793 | 90,126 | 34.2 | 64.9 | <44 | 140 | < 1.29 (0.93 - 1.73) | 2.16 (1.81 - 2.54) |
| **PS-stratification / on-treatment TAR** | | | | | | | | |
| CCAE | 24,111 | 31,166 | 20.1 | 26.4 | 15 | 27 | 0.75 (0.42 - 1.23) | 1.02 (0.67 - 1.49) |
| DAG | 5,763 | 11,200 | 4.7 | 11.7 | 0 | 0 | | |
| ODOD | 17,442 | 21,543 | 12.1 | 17.0 | 19 | 70 | 1.57 (0.94 - 2.45) | 4.12 (3.21 - 5.20) |
| MDCR | 2,130 | 5,305 | 1.7 | 5.0 | 11 | 21 | 6.47 (3.23 - 11.6) | 4.18 (2.59 - 6.39) |
| MDCD | 1,499 | 3,468 | 0.9 | 2.6 | <5 | 8 | < 5.55 (1.80 - 13.0) | 3.13 (1.35 - 6.16) |
| Open Claims | 331,685 | 425,694 | 288.8 | 408.0 | 259 | 648 | 0.90 (0.79 - 1.01) | 1.59 (1.47 - 1.72) |
| OEHR | 20,205 | 32,387 | 6.5 | 16.8 | 5 | 24 | 0.77 (0.25 - 1.81) | 1.43 (0.92 - 2.13) |
| VA | 12,496 | 24,984 | 7.5 | 23.2 | <10 | <10 | < 1.33 (0.64 - 2.45) | < 0.43 (0.21 - 0.79) |
| SIDIAP | 5,160 | 20,267 | 7.1 | 46.7 | 0 | <5 | | < 0.11 (0.03 - 0.25) |
| Meta-analysis | 395,573 | 516,095 | 329.2 | 473.2 | 309 | 790 | 0.94 (0.84 - 1.05) | 1.67 (1.56 - 1.79) |
| Meta-analysis (w/o USOC) | 63,888 | 90,401 | 40.4 | 65.1 | 50 | 142 | 1.24 (0.92 - 1.63) | 2.18 (1.84 - 2.57) |
| **PS-matching / on-treatment TAR without escalation** | | | | | | | | |
| CCAE | 19,730 | 31,143 | 15.3 | 21.3 | 11 | 22 | 0.72 (0.36 - 1.29) | 1.03 (0.65 - 1.56) |
| DAG | 4,640 | 11,168 | 3.7 | 10.5 | 0 | 0 | | |
| ODOD | 12,497 | 21,421 | 8.4 | 13.8 | 12 | 64 | 1.43 (0.74 - 2.50) | 4.63 (3.57 - 5.92) |
| MDCR | 1,715 | 5,246 | 1.4 | 4.2 | 8 | 13 | 5.90 (2.55 - 11.6) | 3.08 (1.64 - 5.27) |
| MDCD | 1,419 | 3,454 | 0.7 | 1.9 | <5 | 7 | < 6.71 (2.18 - 15.6) | 3.66 (1.47 - 7.54) |
| Open Claims | 243,645 | 425,371 | 198.4 | 316.3 | 160 | 495 | 0.81 (0.69 - 0.94) | 1.57 (1.43 - 1.71) |
| OEHR | 16,851 | 32,316 | 4.8 | 13.5 | <5 | 16 | < 1.04 (0.34 - 2.42) | 1.18 (0.68 - 1.92) |
| VA | 7,788 | 23,941 | 4.5 | 17.7 | 0 | <10 | | < 0.57 (0.27 - 1.04) |
| SIDIAP | 4,338 | 19,828 | 5.6 | 35.8 | 0 | <5 | | < 0.14 (0.05 - 0.33) |
| Meta-analysis | 292,723 | 510,251 | 226.9 | 364.9 | <188 | 597 | < 0.83 (0.71 - 0.96) | 1.64 (1.51 - 1.77) |
| Meta-analysis (w/o USOC) | 49,078 | 84,880 | 28.5 | 48.7 | <28 | 102 | < 0.98 (0.65 - 1.42) | 2.10 (1.71 - 2.55) |
| **PS-stratification / on-treatment TAR without escalation** | | | | | | | | |
| CCAE | 24,111 | 31,166 | 17.5 | 21.3 | 11 | 22 | 0.63 (0.31 - 1.12) | 1.03 (0.65 - 1.56) |
| DAG | 5,763 | 11,200 | 4.3 | 10.6 | 0 | 0 | | |
| ODOD | 17,442 | 21,543 | 10.7 | 13.9 | 16 | 64 | 1.49 (0.85 - 2.42) | 4.60 (3.54 - 5.87) |
| MDCR | 2,130 | 5,305 | 1.5 | 4.3 | 10 | 13 | 6.59 (3.16 - 12.1) | 3.05 (1.62 - 5.21) |
| MDCD | 1,499 | 3,468 | 0.8 | 1.9 | <5 | 7 | < 6.55 (2.13 - 15.3) | 3.64 (1.46 - 7.50) |
| Open Claims | 331,685 | 425,694 | 245.5 | 316.4 | 228 | 500 | 0.93 (0.81 - 1.06) | 1.58 (1.44 - 1.72) |
| OEHR | 20,205 | 32,387 | 5.7 | 13.5 | <5 | 17 | < 0.88 (0.29 - 2.05) | 1.26 (0.73 - 2.01) |
| VA | 12,496 | 24,984 | 6.5 | 18.9 | <10 | <10 | < 1.54 (0.74 - 2.83) | < 0.53 (0.25 - 0.98) |
| SIDIAP | 5,160 | 20,267 | 6.3 | 37.0 | 0 | <5 | | < 0.13 (0.04 - 0.32) |
| Meta-analysis | 395,573 | 516,095 | 280.9 | 369.5 | <270 | 616 | < 0.96 (0.85 - 1.08) | 1.67 (1.54 - 1.80) |
| Meta-analysis (w/o USOC) | 63,888 | 90,401 | 35.5 | 53.1 | <42 | 116 | < 1.18 (0.85 - 1.60) | 2.19 (1.81 - 2.62) |

##### 4.2.10 Abnormal weight gain

**Supplementary Table 325:** Exposures, outcomes and risk estimates for Abnormal weight gain in SGLT2I (male) (T) vs DPP4I (male) (C) new-user cohorts under four different propensity score (PS) adjustment / time-at-risk (TAR) designs across data sources. We report total patient exposure time in 1,000 patient-year (KYr) units, uncalibrated and calibrated hazard ratios (HRs) and their 95% confidence intervals (CIs). Greyed out entries did not pass study diagnostics and are included only for completeness.

| | Patients | | Exposure (KYr) | | Outcomes | | Incidence (per KYr) | |
|---|---|---|---|---|---|---|---|---|
| | T | C | T | C | T | C | T | C |
| **PS-matching / on-treatment TAR** | | | | | | | | |
| CCAE | 19,056 | 30,297 | 17.0 | 25.6 | 59 | 85 | 3.48 (2.65 - 4.48) | 3.32 (2.65 - 4.11) |
| DAG | 4,609 | 11,102 | 4.0 | 11.6 | 0 | 13 | | 1.12 (0.60 - 1.92) |
| ODOD | 12,108 | 20,883 | 9.2 | 16.4 | 28 | 92 | 3.03 (2.01 - 4.38) | 5.62 (4.53 - 6.89) |
| MDCR | 1,687 | 5,199 | 1.5 | 4.9 | <5 | 10 | < 3.36 (1.09 - 7.85) | 2.03 (0.97 - 3.73) |
| MDCD | 1,334 | 3,301 | 0.8 | 2.4 | <5 | 16 | < 6.05 (1.97 - 14.1) | 6.65 (3.80 - 10.8) |
| Open Claims | 236,160 | 414,950 | 228.8 | 397.2 | 269 | 700 | 1.18 (1.04 - 1.33) | 1.76 (1.63 - 1.90) |
| OEHR | 16,309 | 31,521 | 5.3 | 16.2 | 26 | 67 | 4.89 (3.19 - 7.16) | 4.12 (3.20 - 5.24) |
| VA | 7,634 | 23,500 | 5.2 | 21.3 | <10 | 11 | < 1.92 (0.92 - 3.53) | 0.52 (0.26 - 0.92) |
| SIDIAP | 4,284 | 19,581 | 6.2 | 44.6 | 8 | 53 | 1.28 (0.55 - 2.52) | 1.19 (0.89 - 1.56) |
| Meta-analysis | 287,917 | 517,232 | 266.6 | 500.0 | 390 | 997 | 1.46 (1.32 - 1.62) | 1.99 (1.87 - 2.12) |
| Meta-analysis (w/o USOC) | 51,757 | 102,282 | 37.8 | 102.8 | 121 | 297 | 3.20 (2.66 - 3.83) | 2.89 (2.57 - 3.24) |
| **PS-stratification / on-treatment TAR** | | | | | | | | |
| CCAE | 23,300 | 30,318 | 19.3 | 25.6 | 68 | 86 | 3.52 (2.73 - 4.46) | 3.36 (2.69 - 4.15) |
| DAG | 5,722 | 11,134 | 4.7 | 11.6 | 0 | 13 | | 1.12 (0.60 - 1.92) |
| ODOD | 16,867 | 21,000 | 11.7 | 16.5 | 32 | 92 | 2.74 (1.88 - 3.87) | 5.58 (4.50 - 6.85) |
| MDCR | 2,099 | 5,258 | 1.7 | 5.0 | <5 | 10 | < 3.01 (0.98 - 7.02) | 2.01 (0.96 - 3.69) |
| MDCD | 1,408 | 3,314 | 0.8 | 2.4 | <5 | 16 | < 5.93 (1.92 - 13.8) | 6.61 (3.78 - 10.7) |
| Open Claims | 321,381 | 415,274 | 279.0 | 397.5 | 317 | 701 | 1.14 (1.01 - 1.27) | 1.76 (1.64 - 1.90) |
| OEHR | 19,556 | 31,592 | 6.2 | 16.3 | 30 | 67 | 4.81 (3.24 - 6.86) | 4.12 (3.19 - 5.23) |
| VA | 12,226 | 24,501 | 7.4 | 22.8 | <10 | 11 | < 1.36 (0.65 - 2.49) | 0.48 (0.24 - 0.86) |
| SIDIAP | 5,107 | 20,006 | 7.0 | 46.1 | 8 | 60 | 1.15 (0.50 - 2.26) | 1.30 (0.99 - 1.68) |
| Meta-analysis | 386,211 | 518,190 | 323.2 | 501.9 | 455 | 1,006 | 1.41 (1.28 - 1.54) | 2.00 (1.88 - 2.13) |
| Meta-analysis (w/o USOC) | 64,830 | 102,916 | 44.2 | 104.5 | 138 | 305 | 3.12 (2.62 - 3.69) | 2.92 (2.60 - 3.27) |
| **PS-matching / on-treatment TAR without escalation** | | | | | | | | |
| CCAE | 19,056 | 30,297 | 14.7 | 20.7 | 51 | 69 | 3.47 (2.58 - 4.56) | 3.33 (2.59 - 4.21) |
| DAG | 4,609 | 11,102 | 3.7 | 10.4 | 0 | 11 | | 1.05 (0.53 - 1.89) |
| ODOD | 12,108 | 20,883 | 8.1 | 13.5 | 23 | 70 | 2.83 (1.79 - 4.24) | 5.20 (4.06 - 6.57) |
| MDCR | 1,687 | 5,199 | 1.3 | 4.2 | <5 | 7 | < 3.77 (1.23 - 8.81) | 1.67 (0.67 - 3.44) |
| MDCD | 1,334 | 3,301 | 0.7 | 1.8 | <5 | 10 | < 7.19 (2.33 - 16.8) | 5.53 (2.65 - 10.2) |
| Open Claims | 236,160 | 414,950 | 192.0 | 308.7 | 213 | 526 | 1.11 (0.97 - 1.27) | 1.70 (1.56 - 1.86) |
| OEHR | 16,309 | 31,521 | 4.7 | 13.1 | 21 | 52 | 4.51 (2.79 - 6.90) | 3.96 (2.95 - 5.19) |
| VA | 7,634 | 23,500 | 4.4 | 17.4 | <10 | <10 | < 2.26 (1.08 - 4.15) | < 0.58 (0.28 - 1.06) |
| SIDIAP | 4,284 | 19,581 | 5.6 | 35.4 | 8 | 45 | 1.44 (0.62 - 2.84) | 1.27 (0.93 - 1.70) |
| Meta-analysis | 287,917 | 517,232 | 225.0 | 391.4 | 316 | 762 | 1.40 (1.25 - 1.57) | 1.95 (1.81 - 2.09) |
| Meta-analysis (w/o USOC) | 51,757 | 102,282 | 33.0 | 82.7 | 103 | 236 | 3.12 (2.54 - 3.78) | 2.85 (2.50 - 3.24) |
| **PS-stratification / on-treatment TAR without escalation** | | | | | | | | |
| CCAE | 23,300 | 30,318 | 16.9 | 20.7 | 60 | 70 | 3.55 (2.71 - 4.57) | 3.38 (2.63 - 4.27) |
| DAG | 5,722 | 11,134 | 4.3 | 10.5 | 0 | 11 | | 1.05 (0.52 - 1.88) |
| ODOD | 16,867 | 21,000 | 10.4 | 13.5 | 25 | 70 | 2.41 (1.56 - 3.56) | 5.17 (4.03 - 6.53) |
| MDCR | 2,099 | 5,258 | 1.5 | 4.2 | <5 | 7 | < 3.36 (1.09 - 7.83) | 1.65 (0.66 - 3.40) |
| MDCD | 1,408 | 3,314 | 0.7 | 1.8 | <5 | 10 | < 7.03 (2.28 - 16.4) | 5.50 (2.64 - 10.1) |
| Open Claims | 321,381 | 415,274 | 237.4 | 308.9 | 254 | 527 | 1.07 (0.94 - 1.21) | 1.71 (1.56 - 1.86) |
| OEHR | 19,556 | 31,592 | 5.5 | 13.2 | 24 | 52 | 4.37 (2.80 - 6.51) | 3.95 (2.95 - 5.18) |
| VA | 12,226 | 24,501 | 6.4 | 18.5 | <10 | <10 | < 1.57 (0.75 - 2.88) | < 0.54 (0.26 - 0.99) |
| SIDIAP | 5,107 | 20,006 | 6.2 | 36.5 | 8 | 50 | 1.29 (0.56 - 2.53) | 1.37 (1.02 - 1.80) |
| Meta-analysis | 386,211 | 518,190 | 276.4 | 392.8 | 371 | 769 | 1.34 (1.21 - 1.49) | 1.96 (1.82 - 2.10) |
| Meta-analysis (w/o USOC) | 64,830 | 102,916 | 39.0 | 84.0 | 117 | 242 | 3.00 (2.48 - 3.60) | 2.88 (2.53 - 3.27) |

###### 4.2.11 Abnormal weight loss

**Supplementary Table 326:** Exposures, outcomes and risk estimates for Abnormal weight loss in SGLT2I (male) (T) vs DPP4I (male) (C) new-user cohorts under four different propensity score (PS) adjustment / time-at-risk (TAR) designs across data sources. We report total patient exposure time in 1,000 patient-year (KYr) units, uncalibrated and calibrated hazard ratios (HRs) and their 95% confidence intervals (CIs). Greyed out entries did not pass study diagnostics and are included only for completeness.

| | Patients | | Exposure (KYr) | | Outcomes | | Incidence (per KYr) | |
|---|---|---|---|---|---|---|---|---|
| | T | C | T | C | T | C | T | C |
| **PS-matching / on-treatment TAR** | | | | | | | | |
| CCAE | 19,278 | 30,384 | 17.2 | 25.6 | 123 | 205 | 7.15 (5.94 - 8.53) | 8.01 (6.95 - 9.18) |
| DAG | 4,550 | 10,926 | 4.0 | 11.4 | 33 | 79 | 8.33 (5.73 - 11.7) | 6.96 (5.51 - 8.67) |
| ODOD | 12,051 | 20,464 | 9.1 | 16.0 | 158 | 265 | 17.3 (14.7 - 20.2) | 16.6 (14.6 - 18.7) |
| MDCR | 1,655 | 5,015 | 1.5 | 4.7 | 33 | 72 | 22.5 (15.5 - 31.6) | 15.2 (11.9 - 19.1) |
| MDCD | 1,361 | 3,296 | 0.8 | 2.4 | 14 | 38 | 16.7 (9.12 - 28.0) | 15.7 (11.1 - 21.6) |
| Open Claims | 236,402 | 410,998 | 229.1 | 393.1 | 1,313 | 2,502 | 5.73 (5.42 - 6.05) | 6.37 (6.12 - 6.62) |
| OEHR | 16,368 | 31,327 | 5.3 | 16.1 | 74 | 193 | 13.9 (10.9 - 17.4) | 12.0 (10.4 - 13.8) |
| VA | 7,516 | 23,081 | 5.1 | 20.9 | 69 | 188 | 13.5 (10.5 - 17.1) | 9.00 (7.76 - 10.4) |
| SIDIAP | 4,288 | 19,572 | 6.3 | 44.5 | 29 | 186 | 4.62 (3.09 - 6.63) | 4.18 (3.60 - 4.83) |
| Meta-analysis | 304,043 | 559,339 | 279.5 | 541.8 | 1,839 | 3,720 | 6.58 (6.28 - 6.89) | 6.87 (6.65 - 7.09) |
| Meta-analysis (w/o USOC) | 67,641 | 148,341 | 50.4 | 148.8 | 526 | 1,218 | 10.4 (9.57 - 11.4) | 8.19 (7.73 - 8.66) |
| **PS-stratification / on-treatment TAR** | | | | | | | | |
| CCAE | 23,568 | 30,406 | 19.6 | 25.6 | 145 | 206 | 7.41 (6.25 - 8.72) | 8.05 (6.98 - 9.22) |
| DAG | 5,651 | 10,957 | 4.6 | 11.4 | 39 | 79 | 8.45 (6.01 - 11.5) | 6.93 (5.49 - 8.64) |
| ODOD | 16,888 | 20,567 | 11.6 | 16.1 | 184 | 267 | 15.8 (13.6 - 18.3) | 16.6 (14.7 - 18.7) |
| MDCR | 2,062 | 5,063 | 1.6 | 4.8 | 35 | 73 | 21.4 (14.9 - 29.7) | 15.2 (11.9 - 19.2) |
| MDCD | 1,443 | 3,308 | 0.9 | 2.4 | 14 | 38 | 16.3 (8.91 - 27.3) | 15.7 (11.1 - 21.5) |
| Open Claims | 322,505 | 411,312 | 280.1 | 393.3 | 1,595 | 2,511 | 5.69 (5.42 - 5.98) | 6.38 (6.14 - 6.64) |
| OEHR | 19,644 | 31,392 | 6.3 | 16.1 | 79 | 195 | 12.6 (9.99 - 15.7) | 12.1 (10.5 - 13.9) |
| VA | 12,060 | 24,052 | 7.2 | 22.3 | 100 | 206 | 13.8 (11.2 - 16.8) | 9.24 (8.02 - 10.6) |
| SIDIAP | 5,093 | 19,998 | 7.0 | 46.0 | 36 | 192 | 5.15 (3.61 - 7.13) | 4.18 (3.61 - 4.81) |
| Meta-analysis | 395,411 | 529,695 | 330.8 | 513.2 | 2,113 | 3,523 | 6.39 (6.12 - 6.67) | 6.86 (6.64 - 7.09) |
| Meta-analysis (w/o USOC) | 72,906 | 118,383 | 50.7 | 120.0 | 518 | 1,012 | 10.2 (9.35 - 11.1) | 8.44 (7.92 - 8.97) |
| **PS-matching / on-treatment TAR without escalation** | | | | | | | | |
| CCAE | 19,278 | 30,384 | 14.9 | 20.7 | 106 | 159 | 7.11 (5.83 - 8.61) | 7.67 (6.52 - 8.96) |
| DAG | 4,550 | 10,926 | 3.6 | 10.3 | 32 | 68 | 8.80 (6.02 - 12.4) | 6.63 (5.15 - 8.41) |
| ODOD | 12,051 | 20,464 | 8.0 | 13.1 | 135 | 216 | 16.8 (14.1 - 19.9) | 16.5 (14.4 - 18.8) |
| MDCR | 1,655 | 5,015 | 1.3 | 4.0 | 30 | 65 | 23.0 (15.5 - 32.8) | 16.1 (12.5 - 20.6) |
| MDCD | 1,361 | 3,296 | 0.7 | 1.8 | 13 | 24 | 18.2 (9.70 - 31.2) | 13.2 (8.47 - 19.7) |
| Open Claims | 236,402 | 410,998 | 192.3 | 304.9 | 1,128 | 1,972 | 5.87 (5.53 - 6.22) | 6.47 (6.19 - 6.76) |
| OEHR | 16,368 | 31,327 | 4.7 | 13.0 | 66 | 144 | 14.1 (10.9 - 18.0) | 11.1 (9.32 - 13.0) |
| VA | 7,516 | 23,081 | 4.4 | 17.0 | 58 | 156 | 13.3 (10.1 - 17.2) | 9.17 (7.79 - 10.7) |
| SIDIAP | 4,288 | 19,572 | 5.6 | 35.3 | 25 | 152 | 4.48 (2.90 - 6.62) | 4.31 (3.65 - 5.05) |
| Meta-analysis | 304,043 | 559,339 | 236.4 | 426.4 | 1,587 | 2,962 | 6.71 (6.39 - 7.05) | 6.95 (6.70 - 7.20) |
| Meta-analysis (w/o USOC) | 67,641 | 148,341 | 44.2 | 121.5 | 459 | 990 | 10.4 (9.46 - 11.4) | 8.15 (7.65 - 8.67) |
| **PS-stratification / on-treatment TAR without escalation** | | | | | | | | |
| CCAE | 23,568 | 30,406 | 17.1 | 20.7 | 126 | 159 | 7.37 (6.14 - 8.77) | 7.66 (6.52 - 8.95) |
| DAG | 5,651 | 10,957 | 4.3 | 10.3 | 36 | 68 | 8.47 (5.93 - 11.7) | 6.60 (5.13 - 8.37) |
| ODOD | 16,888 | 20,567 | 10.3 | 13.2 | 160 | 218 | 15.5 (13.2 - 18.1) | 16.5 (14.4 - 18.9) |
| MDCR | 2,062 | 5,063 | 1.5 | 4.1 | 31 | 66 | 21.1 (14.4 - 30.0) | 16.2 (12.5 - 20.6) |
| MDCD | 1,443 | 3,308 | 0.7 | 1.8 | 13 | 24 | 17.8 (9.47 - 30.4) | 13.2 (8.44 - 19.6) |
| Open Claims | 322,505 | 411,312 | 238.3 | 305.1 | 1,386 | 1,979 | 5.82 (5.51 - 6.13) | 6.49 (6.20 - 6.78) |
| OEHR | 19,644 | 31,392 | 5.5 | 13.1 | 70 | 145 | 12.7 (9.90 - 16.1) | 11.1 (9.37 - 13.1) |
| VA | 12,060 | 24,052 | 6.3 | 18.1 | 87 | 170 | 13.9 (11.1 - 17.1) | 9.38 (8.03 - 10.9) |
| SIDIAP | 5,093 | 19,998 | 6.2 | 36.4 | 32 | 157 | 5.14 (3.52 - 7.26) | 4.31 (3.66 - 5.04) |
| Meta-analysis | 395,411 | 529,695 | 283.2 | 402.9 | 1,841 | 2,792 | 6.50 (6.21 - 6.81) | 6.93 (6.68 - 7.19) |
| Meta-analysis (w/o USOC) | 72,906 | 118,383 | 44.9 | 97.8 | 455 | 813 | 10.1 (9.23 - 11.1) | 8.31 (7.75 - 8.91) |

###### 4.2.12 Acute pancreatitis

**Supplementary Table 327:** Exposures, outcomes and risk estimates for Acute pancreatitis in SGLT2I (male) (T) vs DPP4I (male) (C) new-user cohorts under four different propensity score (PS) adjustment / time-at-risk (TAR) designs across data sources. We report total patient exposure time in 1,000 patient-year (KYr) units, uncalibrated and calibrated hazard ratios (HRs) and their 95% confidence intervals (CIs). Greyed out entries did not pass study diagnostics and are included only for completeness.

| | Patients | | Exposure (KYr) | | Outcomes | | Incidence (per KYr) | |
|---|---|---|---|---|---|---|---|---|
| | T | C | T | C | T | C | T | C |
| **PS-matching / on-treatment TAR** | | | | | | | | |
| CCAE | 19,555 | 30,931 | 17.5 | 26.2 | 36 | 61 | 2.06 (1.44 - 2.85) | 2.33 (1.78 - 2.99) |
| DAG | 4,640 | 11,168 | 4.1 | 11.6 | 0 | 0 | | |
| ODOD | 12,395 | 21,291 | 9.5 | 16.8 | 25 | 47 | 2.64 (1.71 - 3.89) | 2.80 (2.06 - 3.72) |
| MDCR | 1,708 | 5,225 | 1.5 | 4.9 | <5 | 17 | < 3.31 (1.07 - 7.71) | 3.44 (2.00 - 5.51) |
| MDCD | 1,404 | 3,403 | 0.9 | 2.5 | 6 | 15 | 7.00 (2.57 - 15.2) | 5.98 (3.35 - 9.86) |
| Open Claims | 240,896 | 421,406 | 233.9 | 404.3 | 305 | 803 | 1.30 (1.16 - 1.46) | 1.99 (1.85 - 2.13) |
| OEHR | 16,778 | 32,198 | 5.5 | 16.6 | 12 | 38 | 2.19 (1.13 - 3.83) | 2.28 (1.62 - 3.13) |
| VA | 7,703 | 23,745 | 5.3 | 21.5 | <10 | 38 | < 1.90 (0.91 - 3.50) | 1.76 (1.25 - 2.42) |
| SIDIAP | 4,320 | 19,781 | 6.3 | 45.0 | <5 | 18 | < 0.79 (0.26 - 1.85) | 0.40 (0.24 - 0.63) |
| Meta-analysis | 299,035 | 534,796 | 273.1 | 490.4 | <393 | 1,004 | < 1.44 (1.30 - 1.59) | 2.05 (1.92 - 2.18) |
| Meta-analysis (w/o USOC) | 58,139 | 113,390 | 39.2 | 86.1 | <88 | 201 | < 2.25 (1.80 - 2.77) | 2.33 (2.02 - 2.68) |
| **PS-stratification / on-treatment TAR** | | | | | | | | |
| CCAE | 23,890 | 30,952 | 19.9 | 26.2 | 42 | 61 | 2.11 (1.52 - 2.86) | 2.33 (1.78 - 2.99) |
| DAG | 5,763 | 11,200 | 4.7 | 11.7 | 0 | 0 | | |
| ODOD | 17,307 | 21,412 | 12.0 | 16.9 | 29 | 47 | 2.41 (1.62 - 3.47) | 2.78 (2.04 - 3.70) |
| MDCR | 2,119 | 5,282 | 1.7 | 5.0 | <5 | 17 | < 2.96 (0.96 - 6.91) | 3.40 (1.98 - 5.44) |
| MDCD | 1,481 | 3,416 | 0.9 | 2.5 | 6 | 16 | 6.86 (2.52 - 14.9) | 6.35 (3.63 - 10.3) |
| Open Claims | 327,945 | 421,728 | 285.4 | 404.5 | 377 | 804 | 1.32 (1.19 - 1.46) | 1.99 (1.85 - 2.13) |
| OEHR | 20,108 | 32,269 | 6.4 | 16.7 | 13 | 38 | 2.02 (1.08 - 3.46) | 2.28 (1.61 - 3.13) |
| VA | 12,372 | 24,770 | 7.4 | 23.0 | <10 | 40 | < 1.34 (0.64 - 2.47) | 1.74 (1.24 - 2.36) |
| SIDIAP | 5,142 | 20,218 | 7.0 | 46.6 | <5 | 20 | < 0.71 (0.23 - 1.66) | 0.43 (0.26 - 0.66) |
| Meta-analysis | 391,369 | 511,643 | 325.4 | 469.2 | <466 | 967 | < 1.43 (1.30 - 1.57) | 2.06 (1.93 - 2.19) |
| Meta-analysis (w/o USOC) | 63,424 | 89,915 | 40.0 | 64.8 | <89 | 163 | < 2.22 (1.79 - 2.74) | 2.52 (2.15 - 2.93) |
| **PS-matching / on-treatment TAR without escalation** | | | | | | | | |
| CCAE | 19,555 | 30,931 | 15.1 | 21.2 | 29 | 45 | 1.92 (1.28 - 2.75) | 2.12 (1.55 - 2.84) |
| DAG | 4,640 | 11,168 | 3.7 | 10.5 | 0 | 0 | | |
| ODOD | 12,395 | 21,291 | 8.3 | 13.7 | 19 | 33 | 2.28 (1.37 - 3.57) | 2.40 (1.65 - 3.37) |
| MDCR | 1,708 | 5,225 | 1.3 | 4.2 | <5 | 16 | < 3.72 (1.21 - 8.68) | 3.81 (2.18 - 6.19) |
| MDCD | 1,404 | 3,403 | 0.7 | 1.9 | 5 | 10 | 6.91 (2.24 - 16.1) | 5.31 (2.55 - 9.76) |
| Open Claims | 240,896 | 421,406 | 196.3 | 313.7 | 239 | 587 | 1.22 (1.07 - 1.38) | 1.87 (1.72 - 2.03) |
| OEHR | 16,778 | 32,198 | 4.8 | 13.5 | 10 | 26 | 2.09 (1.00 - 3.84) | 1.93 (1.26 - 2.83) |
| VA | 7,703 | 23,745 | 4.5 | 17.5 | <10 | 27 | < 2.23 (1.07 - 4.11) | 1.54 (1.01 - 2.24) |
| SIDIAP | 4,320 | 19,781 | 5.6 | 35.8 | <5 | 16 | < 0.89 (0.29 - 2.08) | 0.45 (0.26 - 0.73) |
| Meta-analysis | 297,327 | 529,571 | 229.0 | 379.6 | <307 | 718 | < 1.34 (1.19 - 1.50) | 1.89 (1.76 - 2.04) |
| Meta-analysis (w/o USOC) | 56,431 | 108,165 | 32.7 | 65.9 | <68 | 131 | < 2.08 (1.61 - 2.63) | 1.99 (1.66 - 2.36) |
| **PS-stratification / on-treatment TAR without escalation** | | | | | | | | |
| CCAE | 23,890 | 30,952 | 17.4 | 21.2 | 34 | 45 | 1.96 (1.36 - 2.74) | 2.12 (1.55 - 2.84) |
| DAG | 5,763 | 11,200 | 4.3 | 10.6 | 0 | 0 | | |
| ODOD | 17,307 | 21,412 | 10.7 | 13.8 | 23 | 33 | 2.16 (1.37 - 3.24) | 2.39 (1.64 - 3.35) |
| MDCR | 2,119 | 5,282 | 1.5 | 4.2 | <5 | 16 | < 3.32 (1.08 - 7.74) | 3.77 (2.15 - 6.12) |
| MDCD | 1,481 | 3,416 | 0.7 | 1.9 | 5 | 11 | 6.76 (2.20 - 15.8) | 5.81 (2.90 - 10.4) |
| Open Claims | 327,945 | 421,728 | 242.8 | 313.8 | 306 | 588 | 1.26 (1.12 - 1.41) | 1.87 (1.73 - 2.03) |
| OEHR | 20,108 | 32,269 | 5.7 | 13.5 | 11 | 26 | 1.95 (0.97 - 3.48) | 1.93 (1.26 - 2.82) |
| VA | 12,372 | 24,770 | 6.4 | 18.7 | <10 | 29 | < 1.55 (0.75 - 2.86) | 1.55 (1.04 - 2.23) |
| SIDIAP | 5,142 | 20,218 | 6.3 | 37.0 | <5 | 18 | < 0.80 (0.26 - 1.86) | 0.49 (0.29 - 0.77) |
| Meta-analysis | 391,369 | 511,643 | 278.0 | 366.6 | <379 | 708 | < 1.36 (1.23 - 1.51) | 1.93 (1.79 - 2.08) |
| Meta-analysis (w/o USOC) | 63,424 | 89,915 | 35.2 | 52.8 | <73 | 120 | < 2.08 (1.63 - 2.61) | 2.27 (1.88 - 2.72) |

##### 4.2.13 Bone fracture

**Supplementary Table 328:** Exposures, outcomes and risk estimates for Bone fracture in SGLT2I (male) (T) vs DPP4I (male) (C) new-user cohorts under four different propensity score (PS) adjustment / time-at-risk (TAR) designs across data sources. We report total patient exposure time in 1,000 patient-year (KYr) units, uncalibrated and calibrated hazard ratios (HRs) and their 95% confidence intervals (CIs). Greyed out entries did not pass study diagnostics and are included only for completeness.

| | Patients | | Exposure (KYr) | | Outcomes | | Incidence (per KYr) | |
|---|---|---|---|---|---|---|---|---|
| | T | C | T | C | T | C | T | C |
| **PS-matching / on-treatment TAR** | | | | | | | | |
| CCAE | 18,252 | 28,862 | 16.0 | 24.1 | 258 | 342 | 16.1 (14.2 - 18.2) | 14.2 (12.7 - 15.8) |
| DAG | 4,145 | 9,946 | 3.6 | 10.2 | 51 | 174 | 14.3 (10.7 - 18.8) | 17.0 (14.6 - 19.8) |
| ODOD | 11,627 | 19,777 | 8.7 | 15.3 | 173 | 313 | 19.8 (16.9 - 23.0) | 20.4 (18.2 - 22.8) |
| MDCR | 1,574 | 4,723 | 1.4 | 4.4 | 37 | 111 | 26.7 (18.8 - 36.8) | 25.2 (20.7 - 30.4) |
| MDCD | 1,250 | 3,021 | 0.8 | 2.2 | 35 | 61 | 46.1 (32.1 - 64.2) | 27.6 (21.1 - 35.5) |
| Open Claims | 218,226 | 382,506 | 210.7 | 363.0 | 2,328 | 4,935 | 11.0 (10.6 - 11.5) | 13.6 (13.2 - 14.0) |
| OEHR | 15,884 | 30,462 | 5.2 | 15.6 | 62 | 240 | 11.9 (9.14 - 15.3) | 15.4 (13.5 - 17.5) |
| VA | 6,972 | 21,479 | 4.8 | 19.5 | 34 | 183 | 7.12 (4.93 - 9.95) | 9.41 (8.09 - 10.9) |
| SIDIAP | 3,903 | 18,011 | 5.6 | 40.8 | 51 | 394 | 9.03 (6.72 - 11.9) | 9.67 (8.73 - 10.7) |
| Meta-analysis | 282,383 | 522,911 | 257.8 | 501.9 | 3,004 | 6,747 | 11.7 (11.2 - 12.1) | 13.4 (13.1 - 13.8) |
| Meta-analysis (w/o USOC) | 64,157 | 140,405 | 47.1 | 138.9 | 676 | 1,812 | 14.3 (13.3 - 15.5) | 13.0 (12.4 - 13.7) |
| **PS-stratification / on-treatment TAR** | | | | | | | | |
| CCAE | 22,298 | 28,882 | 18.2 | 24.1 | 296 | 342 | 16.2 (14.4 - 18.2) | 14.2 (12.7 - 15.8) |
| DAG | 5,164 | 9,967 | 4.2 | 10.2 | 56 | 175 | 13.4 (10.1 - 17.4) | 17.1 (14.7 - 19.8) |
| ODOD | 16,194 | 19,885 | 11.1 | 15.4 | 211 | 317 | 19.1 (16.6 - 21.8) | 20.6 (18.4 - 23.0) |
| MDCR | 1,967 | 4,786 | 1.6 | 4.5 | 38 | 113 | 24.5 (17.3 - 33.6) | 25.3 (20.9 - 30.5) |
| MDCD | 1,321 | 3,032 | 0.8 | 2.2 | 35 | 62 | 45.1 (31.4 - 62.7) | 27.9 (21.4 - 35.8) |
| Open Claims | 295,535 | 382,793 | 256.4 | 363.2 | 2,775 | 4,949 | 10.8 (10.4 - 11.2) | 13.6 (13.2 - 14.0) |
| OEHR | 18,979 | 30,527 | 6.1 | 15.6 | 70 | 242 | 11.5 (8.98 - 14.6) | 15.5 (13.6 - 17.6) |
| VA | 11,119 | 22,469 | 6.7 | 20.9 | 52 | 209 | 7.74 (5.78 - 10.1) | 10.0 (8.69 - 11.5) |
| SIDIAP | 4,634 | 18,465 | 6.3 | 42.4 | 55 | 417 | 8.72 (6.57 - 11.3) | 9.85 (8.92 - 10.8) |
| Meta-analysis | 364,771 | 495,305 | 303.8 | 475.4 | 3,501 | 6,555 | 11.5 (11.1 - 11.9) | 13.8 (13.5 - 14.1) |
| Meta-analysis (w/o USOC) | 69,236 | 112,512 | 47.4 | 112.2 | 726 | 1,606 | 15.3 (14.2 - 16.5) | 14.3 (13.6 - 15.0) |
| **PS-matching / on-treatment TAR without escalation** | | | | | | | | |
| CCAE | 18,252 | 28,862 | 13.9 | 19.6 | 218 | 279 | 15.7 (13.7 - 17.9) | 14.3 (12.6 - 16.0) |
| DAG | 4,145 | 9,946 | 3.3 | 9.2 | 47 | 161 | 14.4 (10.6 - 19.1) | 17.4 (14.8 - 20.3) |
| ODOD | 11,627 | 19,777 | 7.7 | 12.6 | 150 | 251 | 19.5 (16.5 - 22.8) | 19.9 (17.5 - 22.5) |
| MDCR | 1,574 | 4,723 | 1.2 | 3.8 | 31 | 95 | 25.2 (17.1 - 35.8) | 25.2 (20.4 - 30.8) |
| MDCD | 1,250 | 3,021 | 0.6 | 1.7 | 31 | 44 | 47.9 (32.6 - 68.0) | 26.6 (19.3 - 35.7) |
| Open Claims | 218,226 | 382,506 | 177.0 | 282.3 | 1,930 | 3,805 | 10.9 (10.4 - 11.4) | 13.5 (13.1 - 13.9) |
| OEHR | 15,884 | 30,462 | 4.5 | 12.6 | 55 | 181 | 12.1 (9.11 - 15.7) | 14.3 (12.3 - 16.6) |
| VA | 6,972 | 21,479 | 4.1 | 15.9 | 30 | 140 | 7.37 (4.97 - 10.5) | 8.82 (7.42 - 10.4) |
| SIDIAP | 3,903 | 18,011 | 5.0 | 32.5 | 46 | 311 | 9.18 (6.72 - 12.2) | 9.58 (8.55 - 10.7) |
| Meta-analysis | 282,383 | 522,911 | 218.3 | 396.0 | 2,516 | 5,266 | 11.5 (11.1 - 12.0) | 13.3 (12.9 - 13.7) |
| Meta-analysis (w/o USOC) | 64,157 | 140,405 | 41.3 | 113.7 | 586 | 1,461 | 14.2 (13.1 - 15.4) | 12.8 (12.2 - 13.5) |
| **PS-stratification / on-treatment TAR without escalation** | | | | | | | | |
| CCAE | 22,298 | 28,882 | 15.9 | 19.6 | 254 | 279 | 16.0 (14.1 - 18.0) | 14.3 (12.6 - 16.0) |
| DAG | 5,164 | 9,967 | 3.8 | 9.3 | 52 | 162 | 13.5 (10.1 - 17.8) | 17.5 (14.9 - 20.4) |
| ODOD | 16,194 | 19,885 | 9.8 | 12.7 | 186 | 254 | 18.9 (16.3 - 21.8) | 20.0 (17.6 - 22.6) |
| MDCR | 1,967 | 4,786 | 1.4 | 3.8 | 32 | 97 | 23.1 (15.8 - 32.7) | 25.4 (20.6 - 31.0) |
| MDCD | 1,321 | 3,032 | 0.7 | 1.7 | 31 | 45 | 46.8 (31.8 - 66.4) | 27.0 (19.7 - 36.2) |
| Open Claims | 295,535 | 382,793 | 218.3 | 282.5 | 2,332 | 3,817 | 10.7 (10.3 - 11.1) | 13.5 (13.1 - 13.9) |
| OEHR | 18,979 | 30,527 | 5.3 | 12.7 | 62 | 181 | 11.6 (8.90 - 14.9) | 14.3 (12.3 - 16.5) |
| VA | 11,119 | 22,469 | 5.8 | 17.0 | 46 | 161 | 7.89 (5.78 - 10.5) | 9.47 (8.07 - 11.1) |
| SIDIAP | 4,634 | 18,465 | 5.6 | 33.7 | 50 | 326 | 8.89 (6.60 - 11.7) | 9.69 (8.66 - 10.8) |
| Meta-analysis | 364,771 | 495,305 | 260.3 | 374.1 | 2,968 | 5,116 | 11.4 (11.0 - 11.8) | 13.7 (13.3 - 14.1) |
| Meta-analysis (w/o USOC) | 69,236 | 112,512 | 41.9 | 91.7 | 636 | 1,299 | 15.2 (14.0 - 16.4) | 14.2 (13.4 - 15.0) |

##### 4.2.14 Diabetic ketoacidosis

**Supplementary Table 329:** Exposures, outcomes and risk estimates for Diabetic ketoacidosis in SGLT2I (male) (T) vs DPP4I (male) (C) new-user cohorts under four different propensity score (PS) adjustment / time-at-risk (TAR) designs across data sources. We report total patient exposure time in 1,000 patient-year (KYr) units, uncalibrated and calibrated hazard ratios (HRs) and their 95% confidence intervals (CIs). Greyed out entries did not pass study diagnostics and are included only for completeness.

| | Patients | | Exposure (KYr) | | Outcomes | | Incidence (per KYr) | |
|---|---|---|---|---|---|---|---|---|
| | T | C | T | C | T | C | T | C |
| **PS-matching / on-treatment TAR** | | | | | | | | |
| CCAE | 19,732 | 31,147 | 17.6 | 26.4 | 36 | 24 | 2.04 (1.43 - 2.82) | 0.91 (0.58 - 1.35) |
| DAG | 4,640 | 11,168 | 4.1 | 11.6 | 0 | 0 | | |
| ODOD | 12,497 | 21,443 | 9.6 | 16.9 | 16 | 28 | 1.67 (0.96 - 2.72) | 1.66 (1.10 - 2.39) |
| MDCR | 1,717 | 5,257 | 1.5 | 5.0 | <5 | 5 | < 3.27 (1.06 - 7.64) | 1.00 (0.33 - 2.34) |
| MDCD | 1,418 | 3,456 | 0.9 | 2.5 | <5 | 5 | < 5.70 (1.85 - 13.3) | 1.97 (0.64 - 4.59) |
| Open Claims | 243,735 | 425,574 | 236.8 | 408.1 | 233 | 302 | 0.98 (0.86 - 1.12) | 0.74 (0.66 - 0.83) |
| OEHR | 16,856 | 32,318 | 5.5 | 16.7 | 13 | 23 | 2.36 (1.26 - 4.04) | 1.38 (0.87 - 2.06) |
| VA | 7,793 | 23,947 | 5.3 | 21.7 | <10 | 10 | < 1.88 (0.90 - 3.46) | 0.46 (0.22 - 0.85) |
| SIDIAP | 4,340 | 19,836 | 6.3 | 45.1 | 0 | 0 | | |
| Meta-analysis | 292,820 | 510,482 | 269.5 | 468.1 | 298 | 377 | 1.11 (0.98 - 1.24) | 0.81 (0.73 - 0.89) |
| Meta-analysis (w/o USOC) | 49,085 | 84,908 | 32.7 | 60.0 | 65 | 75 | 1.99 (1.53 - 2.53) | 1.25 (0.98 - 1.57) |
| **PS-stratification / on-treatment TAR** | | | | | | | | |
| CCAE | 24,108 | 31,170 | 20.1 | 26.4 | 40 | 24 | 1.99 (1.42 - 2.71) | 0.91 (0.58 - 1.35) |
| DAG | 5,763 | 11,200 | 4.7 | 11.7 | 0 | 0 | | |
| ODOD | 17,459 | 21,565 | 12.1 | 17.0 | 23 | 28 | 1.90 (1.20 - 2.85) | 1.65 (1.09 - 2.38) |
| MDCR | 2,135 | 5,316 | 1.7 | 5.0 | <5 | 5 | < 2.93 (0.95 - 6.84) | 0.99 (0.32 - 2.32) |
| MDCD | 1,499 | 3,470 | 0.9 | 2.6 | <5 | 5 | < 5.58 (1.81 - 13.0) | 1.96 (0.64 - 4.57) |
| Open Claims | 332,197 | 425,900 | 289.1 | 408.4 | 307 | 302 | 1.06 (0.95 - 1.19) | 0.74 (0.66 - 0.83) |
| OEHR | 20,218 | 32,389 | 6.5 | 16.8 | 14 | 23 | 2.17 (1.19 - 3.64) | 1.37 (0.87 - 2.06) |
| VA | 12,523 | 24,988 | 7.5 | 23.2 | <10 | 11 | < 1.33 (0.64 - 2.44) | 0.47 (0.24 - 0.85) |
| SIDIAP | 5,168 | 20,274 | 7.1 | 46.8 | 0 | 0 | | |
| Meta-analysis | 393,982 | 511,024 | 327.8 | 468.5 | 384 | 377 | 1.17 (1.06 - 1.29) | 0.80 (0.73 - 0.89) |
| Meta-analysis (w/o USOC) | 61,785 | 85,124 | 38.7 | 60.2 | 77 | 75 | 1.99 (1.57 - 2.49) | 1.25 (0.98 - 1.56) |
| **PS-matching / on-treatment TAR without escalation** | | | | | | | | |
| CCAE | 19,732 | 31,147 | 15.3 | 21.3 | 29 | 19 | 1.90 (1.27 - 2.72) | 0.89 (0.54 - 1.39) |
| DAG | 4,640 | 11,168 | 3.7 | 10.5 | 0 | 0 | | |
| ODOD | 12,497 | 21,443 | 8.4 | 13.8 | 13 | 21 | 1.55 (0.83 - 2.65) | 1.52 (0.94 - 2.32) |
| MDCR | 1,717 | 5,257 | 1.4 | 4.2 | 0 | <5 | | < 1.18 (0.38 - 2.76) |
| MDCD | 1,418 | 3,456 | 0.7 | 1.9 | <5 | 5 | < 6.72 (2.18 - 15.7) | 2.62 (0.85 - 6.11) |
| Open Claims | 243,735 | 425,574 | 198.5 | 316.6 | 176 | 217 | 0.89 (0.76 - 1.03) | 0.69 (0.60 - 0.78) |
| OEHR | 16,856 | 32,318 | 4.8 | 13.5 | 12 | 16 | 2.49 (1.29 - 4.35) | 1.18 (0.68 - 1.92) |
| VA | 7,793 | 23,947 | 4.5 | 17.7 | <10 | <10 | < 2.21 (1.06 - 4.06) | < 0.57 (0.27 - 1.04) |
| SIDIAP | 4,340 | 19,836 | 5.6 | 35.8 | 0 | 0 | | |
| Meta-analysis | 292,820 | 510,482 | 227.0 | 365.3 | 230 | 273 | 1.01 (0.89 - 1.15) | 0.75 (0.66 - 0.84) |
| Meta-analysis (w/o USOC) | 49,085 | 84,908 | 28.5 | 48.7 | 54 | 56 | 1.89 (1.42 - 2.47) | 1.15 (0.87 - 1.49) |
| **PS-stratification / on-treatment TAR without escalation** | | | | | | | | |
| CCAE | 24,108 | 31,170 | 17.5 | 21.4 | 33 | 19 | 1.88 (1.29 - 2.64) | 0.89 (0.54 - 1.39) |
| DAG | 5,763 | 11,200 | 4.3 | 10.6 | 0 | 0 | | |
| ODOD | 17,459 | 21,565 | 10.8 | 13.9 | 20 | 21 | 1.86 (1.14 - 2.87) | 1.51 (0.93 - 2.30) |
| MDCR | 2,135 | 5,316 | 1.5 | 4.3 | 0 | <5 | | < 1.17 (0.38 - 2.73) |
| MDCD | 1,499 | 3,470 | 0.8 | 1.9 | <5 | 5 | < 6.57 (2.13 - 15.3) | 2.60 (0.85 - 6.08) |
| Open Claims | 332,197 | 425,900 | 245.8 | 316.8 | 243 | 217 | 0.99 (0.87 - 1.12) | 0.69 (0.60 - 0.78) |
| OEHR | 20,218 | 32,389 | 5.7 | 13.5 | 13 | 16 | 2.29 (1.22 - 3.91) | 1.18 (0.68 - 1.92) |
| VA | 12,523 | 24,988 | 6.5 | 18.9 | <10 | <10 | < 1.53 (0.74 - 2.82) | < 0.53 (0.25 - 0.97) |
| SIDIAP | 5,168 | 20,274 | 6.3 | 37.1 | 0 | 0 | | |
| Meta-analysis | 393,982 | 511,024 | 279.8 | 365.6 | 309 | 273 | 1.10 (0.98 - 1.23) | 0.75 (0.66 - 0.84) |
| Meta-analysis (w/o USOC) | 61,785 | 85,124 | 34.0 | 48.8 | 66 | 56 | 1.94 (1.50 - 2.47) | 1.15 (0.87 - 1.49) |

##### 4.2.15 Diarrhea

**Supplementary Table 330:** Exposures, outcomes and risk estimates for Diarrhea in SGLT2I (male) (T) vs DPP4I (male) (C) new-user cohorts under four different propensity score (PS) adjustment / time-at-risk (TAR) designs across data sources. We report total patient exposure time in 1,000 patient-year (KYr) units, uncalibrated and calibrated hazard ratios (HRs) and their 95% confidence intervals (CIs). Greyed out entries did not pass study diagnostics and are included only for completeness.

| | Patients | | Exposure (KYr) | | Outcomes | | Incidence (per KYr) | |
|---|---|---|---|---|---|---|---|---|
| | T | C | T | C | T | C | T | C |
| **PS-matching / on-treatment TAR** | | | | | | | | |
| CCAE | 17,875 | 28,266 | 15.7 | 23.4 | 315 | 540 | 20.0 (17.9 - 22.3) | 23.0 (21.1 - 25.1) |
| DAG | 4,563 | 11,024 | 4.0 | 11.5 | 6 | 20 | 1.50 (0.55 - 3.27) | 1.74 (1.07 - 2.69) |
| ODOD | 11,129 | 18,887 | 8.4 | 14.5 | 222 | 452 | 26.5 (23.2 - 30.3) | 31.3 (28.5 - 34.3) |
| MDCR | 1,531 | 4,626 | 1.3 | 4.3 | 27 | 149 | 20.2 (13.3 - 29.3) | 34.7 (29.3 - 40.7) |
| MDCD | 1,215 | 2,921 | 0.7 | 2.1 | 34 | 94 | 46.0 (31.9 - 64.3) | 44.3 (35.8 - 54.2) |
| Open Claims | 219,670 | 383,846 | 211.8 | 363.1 | 2,498 | 5,465 | 11.8 (11.3 - 12.3) | 15.1 (14.7 - 15.5) |
| OEHR | 15,443 | 29,633 | 5.0 | 14.9 | 116 | 400 | 23.2 (19.2 - 27.9) | 26.8 (24.2 - 29.6) |
| VA | 6,699 | 20,641 | 4.5 | 18.5 | 69 | 369 | 15.2 (11.8 - 19.3) | 20.0 (18.0 - 22.1) |
| SIDIAP | 4,182 | 18,936 | 6.1 | 42.8 | 36 | 412 | 5.87 (4.11 - 8.13) | 9.63 (8.73 - 10.6) |
| Meta-analysis | 282,764 | 522,125 | 258.6 | 500.7 | 3,311 | 7,952 | 12.8 (12.4 - 13.2) | 15.9 (15.5 - 16.2) |
| Meta-analysis (w/o USOC) | 63,094 | 138,279 | 46.7 | 137.7 | 813 | 2,487 | 17.4 (16.2 - 18.6) | 18.1 (17.4 - 18.8) |
| **PS-stratification / on-treatment TAR** | | | | | | | | |
| CCAE | 21,758 | 28,286 | 17.9 | 23.4 | 367 | 540 | 20.6 (18.5 - 22.8) | 23.0 (21.1 - 25.1) |
| DAG | 5,660 | 11,056 | 4.6 | 11.5 | 6 | 20 | 1.29 (0.47 - 2.81) | 1.74 (1.06 - 2.68) |
| ODOD | 15,599 | 19,076 | 10.6 | 14.6 | 286 | 462 | 26.9 (23.8 - 30.2) | 31.6 (28.8 - 34.6) |
| MDCR | 1,906 | 4,679 | 1.5 | 4.3 | 30 | 149 | 20.1 (13.5 - 28.7) | 34.3 (29.0 - 40.2) |
| MDCD | 1,283 | 2,930 | 0.8 | 2.1 | 34 | 95 | 45.0 (31.2 - 62.9) | 44.6 (36.1 - 54.5) |
| Open Claims | 299,156 | 384,186 | 258.9 | 363.3 | 3,000 | 5,481 | 11.6 (11.2 - 12.0) | 15.1 (14.7 - 15.5) |
| OEHR | 18,506 | 29,693 | 5.8 | 15.0 | 134 | 402 | 22.9 (19.2 - 27.1) | 26.9 (24.3 - 29.6) |
| VA | 10,635 | 21,651 | 6.4 | 20.0 | 107 | 398 | 16.7 (13.7 - 20.2) | 19.9 (18.0 - 22.0) |
| SIDIAP | 4,981 | 19,435 | 6.8 | 44.5 | 46 | 436 | 6.73 (4.93 - 8.98) | 9.80 (8.90 - 10.8) |
| Meta-analysis | 367,566 | 496,411 | 306.2 | 476.7 | 3,869 | 7,490 | 12.6 (12.2 - 13.0) | 15.7 (15.4 - 16.1) |
| Meta-analysis (w/o USOC) | 68,410 | 112,225 | 47.3 | 113.4 | 869 | 2,009 | 18.4 (17.2 - 19.6) | 17.7 (17.0 - 18.5) |
| **PS-matching / on-treatment TAR without escalation** | | | | | | | | |
| CCAE | 17,875 | 28,266 | 13.7 | 19.1 | 263 | 435 | 19.2 (16.9 - 21.7) | 22.8 (20.7 - 25.0) |
| DAG | 4,563 | 11,024 | 3.7 | 10.4 | 6 | 18 | 1.64 (0.60 - 3.57) | 1.74 (1.03 - 2.74) |
| ODOD | 11,129 | 18,887 | 7.4 | 11.9 | 188 | 373 | 25.4 (21.9 - 29.3) | 31.3 (28.2 - 34.6) |
| MDCR | 1,531 | 4,626 | 1.2 | 3.7 | 24 | 129 | 19.9 (12.8 - 29.7) | 35.2 (29.4 - 41.8) |
| MDCD | 1,215 | 2,921 | 0.6 | 1.6 | 29 | 67 | 45.9 (30.7 - 65.9) | 41.5 (32.2 - 52.8) |
| Open Claims | 219,670 | 383,846 | 178.3 | 283.1 | 2,022 | 4,233 | 11.3 (10.9 - 11.8) | 15.0 (14.5 - 15.4) |
| OEHR | 15,443 | 29,633 | 4.4 | 12.2 | 102 | 290 | 23.3 (19.0 - 28.3) | 23.8 (21.1 - 26.7) |
| VA | 6,699 | 20,641 | 3.9 | 15.2 | 60 | 303 | 15.5 (11.9 - 20.0) | 20.0 (17.8 - 22.4) |
| SIDIAP | 4,182 | 18,936 | 5.4 | 34.0 | 31 | 336 | 5.70 (3.88 - 8.10) | 9.88 (8.85 - 11.0) |
| Meta-analysis | 282,764 | 522,125 | 219.5 | 396.1 | 2,717 | 6,237 | 12.4 (11.9 - 12.9) | 15.7 (15.4 - 16.1) |
| Meta-analysis (w/o USOC) | 63,094 | 138,279 | 41.1 | 113.0 | 695 | 2,004 | 16.9 (15.7 - 18.2) | 17.7 (17.0 - 18.5) |
| **PS-stratification / on-treatment TAR without escalation** | | | | | | | | |
| CCAE | 21,758 | 28,286 | 15.7 | 19.1 | 307 | 435 | 19.6 (17.5 - 21.9) | 22.8 (20.7 - 25.0) |
| DAG | 5,660 | 11,056 | 4.3 | 10.4 | 6 | 18 | 1.40 (0.52 - 3.06) | 1.73 (1.02 - 2.73) |
| ODOD | 15,599 | 19,076 | 9.5 | 12.1 | 244 | 382 | 25.7 (22.6 - 29.1) | 31.7 (28.6 - 35.0) |
| MDCR | 1,906 | 4,679 | 1.3 | 3.7 | 27 | 129 | 20.0 (13.2 - 29.2) | 34.8 (29.0 - 41.3) |
| MDCD | 1,283 | 2,930 | 0.6 | 1.6 | 29 | 68 | 44.9 (30.1 - 64.5) | 42.0 (32.6 - 53.2) |
| Open Claims | 299,156 | 384,186 | 221.0 | 283.3 | 2,473 | 4,246 | 11.2 (10.8 - 11.6) | 15.0 (14.5 - 15.4) |
| OEHR | 18,506 | 29,693 | 5.2 | 12.2 | 117 | 290 | 22.7 (18.8 - 27.2) | 23.7 (21.1 - 26.6) |
| VA | 10,635 | 21,651 | 5.5 | 16.3 | 95 | 321 | 17.2 (13.9 - 21.0) | 19.7 (17.6 - 21.9) |
| SIDIAP | 4,981 | 19,435 | 6.1 | 35.3 | 41 | 353 | 6.74 (4.83 - 9.14) | 9.99 (8.98 - 11.1) |
| Meta-analysis | 367,566 | 496,411 | 263.0 | 376.2 | 3,215 | 5,853 | 12.2 (11.8 - 12.7) | 15.6 (15.2 - 16.0) |
| Meta-analysis (w/o USOC) | 68,410 | 112,225 | 42.0 | 92.8 | 742 | 1,607 | 17.7 (16.4 - 19.0) | 17.3 (16.5 - 18.2) |

##### 4.2.16 Genitourinary infection

**Supplementary Table 331:** Exposures, outcomes and risk estimates for Genitourinary infection in SGLT2I (male) (T) vs DPP4I (male) (C) new-user cohorts under four different propensity score (PS) adjustment / time-at-risk (TAR) designs across data sources. We report total patient exposure time in 1,000 patient-year (KYr) units, uncalibrated and calibrated hazard ratios (HRs) and their 95% confidence intervals (CIs). Greyed out entries did not pass study diagnostics and are included only for completeness.

| | Patients | | Exposure (KYr) | | Outcomes | | Incidence (per KYr) | |
|---|---|---|---|---|---|---|---|---|
| | T | C | T | C | T | C | T | C |
| **PS-matching / on-treatment TAR** | | | | | | | | |
| CCAE | 18,063 | 28,312 | 16.0 | 23.6 | 243 | 468 | 15.2 (13.3 - 17.2) | 19.8 (18.1 - 21.7) |
| DAG | 4,031 | 9,692 | 3.5 | 9.8 | 79 | 233 | 22.7 (18.0 - 28.3) | 23.8 (20.8 - 27.0) |
| ODOD | 11,059 | 18,283 | 8.4 | 14.0 | 243 | 556 | 28.9 (25.4 - 32.8) | 39.7 (36.4 - 43.1) |
| MDCR | 1,499 | 4,364 | 1.3 | 4.1 | 41 | 166 | 31.5 (22.6 - 42.7) | 40.6 (34.7 - 47.3) |
| MDCD | 1,306 | 3,061 | 0.8 | 2.2 | 15 | 62 | 18.5 (10.4 - 30.6) | 28.3 (21.7 - 36.3) |
| Open Claims | 217,569 | 374,695 | 212.3 | 354.2 | 2,496 | 6,880 | 11.8 (11.3 - 12.2) | 19.4 (19.0 - 19.9) |
| OEHR | 15,971 | 30,272 | 5.2 | 15.4 | 95 | 329 | 18.3 (14.8 - 22.4) | 21.4 (19.1 - 23.8) |
| VA | 7,042 | 21,383 | 4.8 | 19.3 | 39 | 268 | 8.05 (5.72 - 11.0) | 13.9 (12.3 - 15.7) |
| SIDIAP | 3,828 | 17,257 | 5.7 | 38.5 | 119 | 976 | 21.0 (17.4 - 25.2) | 25.3 (23.8 - 27.0) |
| Meta-analysis | 280,892 | 511,290 | 259.0 | 487.8 | 3,365 | 9,956 | 13.0 (12.6 - 13.4) | 20.4 (20.0 - 20.8) |
| Meta-analysis (w/o USOC) | 63,323 | 136,595 | 46.7 | 133.6 | 869 | 3,076 | 18.6 (17.4 - 19.9) | 23.0 (22.2 - 23.9) |
| **PS-stratification / on-treatment TAR** | | | | | | | | |
| CCAE | 22,104 | 28,325 | 18.3 | 23.6 | 274 | 470 | 15.0 (13.3 - 16.9) | 19.9 (18.2 - 21.8) |
| DAG | 4,973 | 9,717 | 4.0 | 9.8 | 96 | 236 | 23.8 (19.3 - 29.1) | 24.0 (21.0 - 27.3) |
| ODOD | 15,706 | 18,423 | 10.8 | 14.1 | 287 | 568 | 26.6 (23.6 - 29.8) | 40.1 (36.9 - 43.6) |
| MDCR | 1,870 | 4,406 | 1.5 | 4.1 | 43 | 168 | 29.5 (21.3 - 39.7) | 40.7 (34.8 - 47.3) |
| MDCD | 1,377 | 3,076 | 0.8 | 2.2 | 15 | 64 | 18.2 (10.2 - 30.0) | 29.1 (22.4 - 37.2) |
| Open Claims | 297,034 | 374,885 | 259.6 | 354.3 | 2,993 | 6,901 | 11.5 (11.1 - 11.9) | 19.5 (19.0 - 19.9) |
| OEHR | 19,171 | 30,328 | 6.1 | 15.4 | 109 | 336 | 17.9 (14.7 - 21.6) | 21.8 (19.5 - 24.2) |
| VA | 11,310 | 22,289 | 6.8 | 20.6 | 68 | 289 | 9.93 (7.71 - 12.6) | 14.0 (12.5 - 15.7) |
| SIDIAP | 4,522 | 17,643 | 6.3 | 39.8 | 131 | 1,029 | 20.9 (17.5 - 24.8) | 25.8 (24.3 - 27.5) |
| Meta-analysis | 365,380 | 483,727 | 306.5 | 461.3 | 3,933 | 9,708 | 12.8 (12.4 - 13.2) | 21.0 (20.6 - 21.5) |
| Meta-analysis (w/o USOC) | 68,346 | 108,842 | 46.9 | 106.9 | 940 | 2,807 | 20.0 (18.8 - 21.4) | 26.2 (25.3 - 27.2) |
| **PS-matching / on-treatment TAR without escalation** | | | | | | | | |
| CCAE | 18,063 | 28,312 | 13.9 | 19.2 | 206 | 381 | 14.8 (12.9 - 17.0) | 19.9 (17.9 - 22.0) |
| DAG | 4,031 | 9,692 | 3.2 | 8.9 | 71 | 207 | 22.2 (17.4 - 28.0) | 23.3 (20.3 - 26.7) |
| ODOD | 11,059 | 18,283 | 7.4 | 11.5 | 208 | 465 | 28.1 (24.4 - 32.2) | 40.5 (36.9 - 44.4) |
| MDCR | 1,499 | 4,364 | 1.2 | 3.5 | 38 | 144 | 32.4 (22.9 - 44.4) | 41.1 (34.6 - 48.4) |
| MDCD | 1,306 | 3,061 | 0.7 | 1.6 | 12 | 49 | 17.6 (9.09 - 30.7) | 29.9 (22.1 - 39.5) |
| Open Claims | 217,569 | 374,695 | 178.2 | 274.9 | 2,090 | 5,470 | 11.7 (11.2 - 12.2) | 19.9 (19.4 - 20.4) |
| OEHR | 15,971 | 30,272 | 4.5 | 12.5 | 82 | 254 | 18.0 (14.3 - 22.4) | 20.3 (17.9 - 23.0) |
| VA | 7,042 | 21,383 | 4.1 | 15.7 | 31 | 220 | 7.51 (5.10 - 10.7) | 14.0 (12.2 - 16.0) |
| SIDIAP | 3,828 | 17,257 | 5.0 | 30.7 | 103 | 771 | 20.4 (16.7 - 24.8) | 25.1 (23.4 - 27.0) |
| Meta-analysis | 280,892 | 511,290 | 219.2 | 384.2 | 2,839 | 7,978 | 12.9 (12.5 - 13.4) | 20.8 (20.3 - 21.2) |
| Meta-analysis (w/o USOC) | 63,323 | 136,595 | 41.0 | 109.3 | 749 | 2,508 | 18.3 (17.0 - 19.6) | 22.9 (22.0 - 23.9) |
| **PS-stratification / on-treatment TAR without escalation** | | | | | | | | |
| CCAE | 22,104 | 28,325 | 16.0 | 19.2 | 233 | 383 | 14.6 (12.8 - 16.6) | 20.0 (18.0 - 22.1) |
| DAG | 4,973 | 9,717 | 3.7 | 8.9 | 88 | 210 | 23.7 (19.0 - 29.2) | 23.6 (20.5 - 27.0) |
| ODOD | 15,706 | 18,423 | 9.6 | 11.6 | 248 | 475 | 25.8 (22.7 - 29.3) | 40.9 (37.3 - 44.8) |
| MDCR | 1,870 | 4,406 | 1.3 | 3.5 | 40 | 145 | 30.4 (21.7 - 41.4) | 41.0 (34.6 - 48.2) |
| MDCD | 1,377 | 3,076 | 0.7 | 1.7 | 12 | 50 | 17.2 (8.91 - 30.1) | 30.3 (22.5 - 39.9) |
| Open Claims | 297,034 | 374,885 | 220.9 | 275.0 | 2,550 | 5,488 | 11.5 (11.1 - 12.0) | 20.0 (19.4 - 20.5) |
| OEHR | 19,171 | 30,328 | 5.4 | 12.5 | 95 | 261 | 17.7 (14.3 - 21.7) | 20.8 (18.4 - 23.5) |
| VA | 11,310 | 22,289 | 5.9 | 16.7 | 59 | 238 | 9.97 (7.59 - 12.9) | 14.2 (12.5 - 16.2) |
| SIDIAP | 4,522 | 17,643 | 5.6 | 31.7 | 115 | 809 | 20.6 (17.0 - 24.7) | 25.5 (23.8 - 27.4) |
| Meta-analysis | 365,380 | 483,727 | 262.5 | 362.5 | 3,369 | 7,771 | 12.8 (12.4 - 13.3) | 21.4 (21.0 - 21.9) |
| Meta-analysis (w/o USOC) | 68,346 | 108,842 | 41.6 | 87.4 | 819 | 2,283 | 19.7 (18.4 - 21.1) | 26.1 (25.1 - 27.2) |

##### 4.2.17 Hyperkalemia

**Supplementary Table 332:** Exposures, outcomes and risk estimates for Hyperkalemia in SGLT2I (male) (T) vs DPP4I (male) (C) new-user cohorts under four different propensity score (PS) adjustment / time-at-risk (TAR) designs across data sources. We report total patient exposure time in 1,000 patient-year (KYr) units, uncalibrated and calibrated hazard ratios (HRs) and their 95% confidence intervals (CIs). Greyed out entries did not pass study diagnostics and are included only for completeness.

| | Patients | | Exposure (KYr) | | Outcomes | | Incidence (per KYr) | |
|---|---|---|---|---|---|---|---|---|
| | T | C | T | C | T | C | T | C |
| **PS-matching / on-treatment TAR** | | | | | | | | |
| CCAE | 19,405 | 30,586 | 17.2 | 25.6 | 118 | 218 | 6.86 (5.68 - 8.22) | 8.51 (7.41 - 9.71) |
| DAG | 4,358 | 10,400 | 3.8 | 10.7 | 44 | 162 | 11.6 (8.45 - 15.6) | 15.2 (12.9 - 17.7) |
| ODOD | 12,017 | 20,373 | 9.1 | 15.8 | 152 | 394 | 16.7 (14.1 - 19.6) | 25.0 (22.6 - 27.6) |
| MDCR | 1,664 | 5,089 | 1.5 | 4.7 | 18 | 103 | 12.1 (7.18 - 19.1) | 21.7 (17.7 - 26.3) |
| MDCD | 1,384 | 3,314 | 0.8 | 2.4 | 14 | 42 | 16.5 (9.04 - 27.7) | 17.5 (12.6 - 23.7) |
| Open Claims | 239,626 | 416,521 | 232.3 | 396.4 | 1,007 | 3,251 | 4.34 (4.07 - 4.61) | 8.20 (7.92 - 8.49) |
| OEHR | 16,549 | 31,585 | 5.4 | 16.1 | 51 | 263 | 9.52 (7.09 - 12.5) | 16.3 (14.4 - 18.4) |
| VA | 7,242 | 22,546 | 4.9 | 20.1 | 64 | 287 | 13.1 (10.1 - 16.7) | 14.3 (12.7 - 16.0) |
| SIDIAP | 4,139 | 18,612 | 6.0 | 41.8 | 60 | 528 | 9.92 (7.57 - 12.8) | 12.6 (11.6 - 13.7) |
| Meta-analysis | 305,000 | 555,712 | 280.1 | 531.3 | 1,514 | 5,206 | 5.40 (5.14 - 5.68) | 9.80 (9.53 - 10.1) |
| Meta-analysis (w/o USOC) | 65,374 | 139,191 | 47.9 | 134.9 | 507 | 1,955 | 10.6 (9.69 - 11.6) | 14.5 (13.9 - 15.2) |
| **PS-stratification / on-treatment TAR** | | | | | | | | |
| CCAE | 23,691 | 30,605 | 19.6 | 25.6 | 134 | 218 | 6.84 (5.73 - 8.10) | 8.50 (7.41 - 9.71) |
| DAG | 5,392 | 10,429 | 4.4 | 10.7 | 52 | 164 | 11.8 (8.81 - 15.5) | 15.3 (13.1 - 17.8) |
| ODOD | 16,805 | 20,471 | 11.6 | 15.8 | 192 | 401 | 16.6 (14.3 - 19.1) | 25.3 (22.9 - 27.9) |
| MDCR | 2,069 | 5,141 | 1.7 | 4.8 | 20 | 103 | 12.1 (7.36 - 18.6) | 21.5 (17.5 - 26.0) |
| MDCD | 1,462 | 3,327 | 0.9 | 2.4 | 15 | 42 | 17.3 (9.69 - 28.6) | 17.4 (12.5 - 23.5) |
| Open Claims | 325,992 | 416,779 | 283.5 | 396.6 | 1,288 | 3,267 | 4.54 (4.30 - 4.80) | 8.24 (7.96 - 8.53) |
| OEHR | 19,863 | 31,647 | 6.3 | 16.1 | 62 | 266 | 9.85 (7.55 - 12.6) | 16.5 (14.6 - 18.6) |
| VA | 11,422 | 23,467 | 6.9 | 21.4 | 104 | 307 | 15.2 (12.4 - 18.4) | 14.3 (12.8 - 16.0) |
| SIDIAP | 4,910 | 18,967 | 6.7 | 43.1 | 78 | 556 | 11.6 (9.19 - 14.5) | 12.9 (11.8 - 14.0) |
| Meta-analysis | 398,722 | 534,039 | 333.7 | 512.8 | 1,826 | 4,975 | 5.47 (5.22 - 5.73) | 9.70 (9.43 - 9.97) |
| Meta-analysis (w/o USOC) | 72,730 | 117,260 | 50.2 | 116.3 | 538 | 1,708 | 10.7 (9.82 - 11.7) | 14.7 (14.0 - 15.4) |
| **PS-matching / on-treatment TAR without escalation** | | | | | | | | |
| CCAE | 19,405 | 30,586 | 14.9 | 20.8 | 97 | 178 | 6.50 (5.27 - 7.93) | 8.56 (7.35 - 9.92) |
| DAG | 4,358 | 10,400 | 3.5 | 9.7 | 38 | 143 | 10.9 (7.73 - 15.0) | 14.8 (12.5 - 17.4) |
| ODOD | 12,017 | 20,373 | 8.0 | 12.9 | 136 | 319 | 17.0 (14.2 - 20.1) | 24.7 (22.1 - 27.6) |
| MDCR | 1,664 | 5,089 | 1.3 | 4.0 | 17 | 84 | 12.9 (7.49 - 20.6) | 20.8 (16.6 - 25.7) |
| MDCD | 1,384 | 3,314 | 0.7 | 1.8 | 13 | 30 | 18.1 (9.63 - 30.9) | 16.7 (11.2 - 23.8) |
| Open Claims | 239,626 | 416,521 | 194.9 | 307.8 | 829 | 2,555 | 4.25 (3.97 - 4.55) | 8.30 (7.98 - 8.63) |
| OEHR | 16,549 | 31,585 | 4.7 | 13.1 | 39 | 203 | 8.29 (5.90 - 11.3) | 15.5 (13.5 - 17.8) |
| VA | 7,242 | 22,546 | 4.2 | 16.4 | 56 | 220 | 13.4 (10.1 - 17.4) | 13.4 (11.7 - 15.3) |
| SIDIAP | 4,139 | 18,612 | 5.4 | 33.2 | 46 | 412 | 8.56 (6.27 - 11.4) | 12.4 (11.2 - 13.6) |
| Meta-analysis | 305,000 | 555,712 | 236.8 | 417.9 | 1,258 | 4,114 | 5.31 (5.02 - 5.61) | 9.84 (9.55 - 10.1) |
| Meta-analysis (w/o USOC) | 65,374 | 139,191 | 42.0 | 110.1 | 429 | 1,559 | 10.2 (9.27 - 11.2) | 14.2 (13.5 - 14.9) |
| **PS-stratification / on-treatment TAR without escalation** | | | | | | | | |
| CCAE | 23,691 | 30,605 | 17.1 | 20.8 | 109 | 178 | 6.36 (5.22 - 7.67) | 8.56 (7.35 - 9.92) |
| DAG | 5,392 | 10,429 | 4.1 | 9.7 | 46 | 145 | 11.3 (8.30 - 15.1) | 15.0 (12.6 - 17.6) |
| ODOD | 16,805 | 20,471 | 10.3 | 13.0 | 173 | 326 | 16.8 (14.4 - 19.5) | 25.1 (22.4 - 28.0) |
| MDCR | 2,069 | 5,141 | 1.5 | 4.1 | 19 | 84 | 12.8 (7.72 - 20.0) | 20.6 (16.4 - 25.5) |
| MDCD | 1,462 | 3,327 | 0.7 | 1.8 | 14 | 30 | 19.0 (10.4 - 31.9) | 16.5 (11.2 - 23.6) |
| Open Claims | 325,992 | 416,779 | 241.1 | 308.0 | 1,086 | 2,569 | 4.50 (4.24 - 4.78) | 8.34 (8.02 - 8.67) |
| OEHR | 19,863 | 31,647 | 5.6 | 13.1 | 47 | 205 | 8.46 (6.22 - 11.3) | 15.7 (13.6 - 18.0) |
| VA | 11,422 | 23,467 | 6.0 | 17.4 | 89 | 238 | 15.0 (12.0 - 18.4) | 13.7 (12.0 - 15.5) |
| SIDIAP | 4,910 | 18,967 | 6.0 | 34.2 | 62 | 436 | 10.4 (7.94 - 13.3) | 12.7 (11.6 - 14.0) |
| Meta-analysis | 398,722 | 534,039 | 285.6 | 402.9 | 1,542 | 3,943 | 5.40 (5.13 - 5.68) | 9.79 (9.48 - 10.1) |
| Meta-analysis (w/o USOC) | 72,730 | 117,260 | 44.5 | 94.9 | 456 | 1,374 | 10.2 (9.32 - 11.2) | 14.5 (13.7 - 15.3) |

##### 4.2.18 Hypoglycemia

**Supplementary Table 333:** Exposures, outcomes and risk estimates for Hypoglycemia in SGLT2I (male) (T) vs DPP4I (male) (C) new-user cohorts under four different propensity score (PS) adjustment / time-at-risk (TAR) designs across data sources. We report total patient exposure time in 1,000 patient-year (KYr) units, uncalibrated and calibrated hazard ratios (HRs) and their 95% confidence intervals (CIs). Greyed out entries did not pass study diagnostics and are included only for completeness.

| | Patients | | Exposure (KYr) | | Outcomes | | Incidence (per KYr) | |
|---|---|---|---|---|---|---|---|---|
| | T | C | T | C | T | C | T | C |
| **PS-matching / on-treatment TAR** | | | | | | | | |
| CCAE | 19,492 | 30,753 | 17.4 | 26.0 | 42 | 80 | 2.42 (1.74 - 3.26) | 3.08 (2.44 - 3.83) |
| DAG | 4,612 | 11,097 | 4.0 | 11.6 | <5 | 17 | < 1.24 (0.40 - 2.90) | 1.47 (0.86 - 2.35) |
| ODOD | 12,339 | 21,176 | 9.4 | 16.7 | 35 | 85 | 3.71 (2.59 - 5.16) | 5.10 (4.07 - 6.31) |
| MDCR | 1,702 | 5,209 | 1.5 | 4.9 | 5 | 18 | 3.32 (1.08 - 7.75) | 3.66 (2.17 - 5.78) |
| MDCD | 1,388 | 3,405 | 0.9 | 2.5 | 6 | 20 | 7.00 (2.57 - 15.2) | 8.02 (4.90 - 12.4) |
| Open Claims | 241,000 | 420,640 | 234.0 | 403.0 | 353 | 960 | 1.51 (1.36 - 1.67) | 2.38 (2.23 - 2.54) |
| OEHR | 16,739 | 32,099 | 5.5 | 16.5 | 15 | 93 | 2.74 (1.53 - 4.52) | 5.63 (4.54 - 6.89) |
| VA | 7,705 | 23,718 | 5.3 | 21.5 | 10 | 71 | 1.90 (0.91 - 3.50) | 3.31 (2.58 - 4.17) |
| SIDIAP | 4,333 | 19,814 | 6.3 | 45.1 | <5 | 22 | < 0.79 (0.26 - 1.84) | 0.49 (0.31 - 0.74) |
| Meta-analysis | 303,589 | 544,692 | 277.1 | 500.1 | <465 | 1,324 | < 1.68 (1.53 - 1.84) | 2.65 (2.51 - 2.79) |
| Meta-analysis (w/o USOC) | 62,589 | 124,052 | 43.1 | 97.1 | <112 | 364 | < 2.60 (2.14 - 3.13) | 3.75 (3.37 - 4.15) |
| **PS-stratification / on-treatment TAR** | | | | | | | | |
| CCAE | 23,833 | 30,776 | 19.8 | 26.0 | 49 | 80 | 2.47 (1.83 - 3.27) | 3.08 (2.44 - 3.83) |
| DAG | 5,735 | 11,129 | 4.7 | 11.6 | 5 | 17 | 1.06 (0.35 - 2.48) | 1.46 (0.85 - 2.34) |
| ODOD | 17,261 | 21,296 | 12.0 | 16.8 | 51 | 85 | 4.26 (3.17 - 5.60) | 5.07 (4.05 - 6.27) |
| MDCR | 2,115 | 5,268 | 1.7 | 5.0 | 5 | 18 | 2.97 (0.96 - 6.92) | 3.62 (2.14 - 5.72) |
| MDCD | 1,468 | 3,418 | 0.9 | 2.5 | 6 | 20 | 6.84 (2.51 - 14.9) | 7.98 (4.87 - 12.3) |
| Open Claims | 328,462 | 420,953 | 285.7 | 403.2 | 421 | 962 | 1.47 (1.34 - 1.62) | 2.39 (2.24 - 2.54) |
| OEHR | 20,063 | 32,169 | 6.4 | 16.6 | 18 | 93 | 2.81 (1.66 - 4.43) | 5.61 (4.53 - 6.88) |
| VA | 12,356 | 24,768 | 7.4 | 23.0 | 14 | 77 | 1.88 (1.03 - 3.16) | 3.35 (2.64 - 4.19) |
| SIDIAP | 5,159 | 20,250 | 7.1 | 46.7 | 5 | 23 | 0.71 (0.23 - 1.65) | 0.49 (0.31 - 0.74) |
| Meta-analysis | 402,628 | 541,841 | 337.4 | 525.8 | 554 | 1,278 | 1.64 (1.51 - 1.78) | 2.43 (2.30 - 2.57) |
| Meta-analysis (w/o USOC) | 74,166 | 120,888 | 51.7 | 122.6 | 133 | 316 | 2.57 (2.16 - 3.05) | 2.58 (2.30 - 2.88) |
| **PS-matching / on-treatment TAR without escalation** | | | | | | | | |
| CCAE | 19,492 | 30,753 | 15.1 | 21.1 | 32 | 53 | 2.12 (1.45 - 2.99) | 2.52 (1.89 - 3.29) |
| DAG | 4,612 | 11,097 | 3.7 | 10.4 | <5 | 14 | < 1.35 (0.44 - 3.16) | 1.34 (0.73 - 2.25) |
| ODOD | 12,339 | 21,176 | 8.3 | 13.7 | 23 | 48 | 2.78 (1.76 - 4.16) | 3.51 (2.59 - 4.66) |
| MDCR | 1,702 | 5,209 | 1.3 | 4.2 | <5 | 12 | < 3.72 (1.21 - 8.67) | 2.87 (1.48 - 5.01) |
| MDCD | 1,388 | 3,405 | 0.7 | 1.9 | <5 | 9 | < 6.92 (2.25 - 16.1) | 4.78 (2.19 - 9.08) |
| Open Claims | 241,000 | 420,640 | 196.3 | 312.9 | 240 | 532 | 1.22 (1.07 - 1.39) | 1.70 (1.56 - 1.85) |
| OEHR | 16,739 | 32,099 | 4.8 | 13.4 | 9 | 47 | 1.88 (0.86 - 3.57) | 3.51 (2.58 - 4.67) |
| VA | 7,705 | 23,718 | 4.5 | 17.5 | <10 | 40 | < 2.23 (1.07 - 4.10) | 2.29 (1.63 - 3.11) |
| SIDIAP | 4,333 | 19,814 | 5.6 | 35.8 | <5 | 12 | < 0.89 (0.29 - 2.07) | 0.34 (0.17 - 0.59) |
| Meta-analysis | 297,275 | 528,386 | 228.9 | 378.5 | <314 | 720 | < 1.37 (1.22 - 1.53) | 1.90 (1.77 - 2.05) |
| Meta-analysis (w/o USOC) | 56,275 | 107,746 | 32.6 | 65.6 | <74 | 188 | < 2.27 (1.78 - 2.85) | 2.87 (2.47 - 3.31) |
| **PS-stratification / on-treatment TAR without escalation** | | | | | | | | |
| CCAE | 23,833 | 30,776 | 17.3 | 21.1 | 38 | 53 | 2.19 (1.55 - 3.01) | 2.52 (1.88 - 3.29) |
| DAG | 5,735 | 11,129 | 4.3 | 10.5 | 5 | 14 | 1.16 (0.38 - 2.70) | 1.34 (0.73 - 2.24) |
| ODOD | 17,261 | 21,296 | 10.6 | 13.8 | 36 | 48 | 3.39 (2.37 - 4.69) | 3.49 (2.57 - 4.62) |
| MDCR | 2,115 | 5,268 | 1.5 | 4.2 | <5 | 12 | < 3.31 (1.07 - 7.72) | 2.83 (1.46 - 4.95) |
| MDCD | 1,468 | 3,418 | 0.7 | 1.9 | <5 | 9 | < 6.76 (2.19 - 15.8) | 4.76 (2.17 - 9.03) |
| Open Claims | 328,462 | 420,953 | 243.0 | 313.0 | 289 | 533 | 1.19 (1.06 - 1.33) | 1.70 (1.56 - 1.85) |
| OEHR | 20,063 | 32,169 | 5.6 | 13.4 | 12 | 47 | 2.12 (1.10 - 3.71) | 3.50 (2.57 - 4.66) |
| VA | 12,356 | 24,768 | 6.4 | 18.7 | <10 | 43 | < 1.56 (0.75 - 2.86) | 2.30 (1.66 - 3.10) |
| SIDIAP | 5,159 | 20,250 | 6.3 | 37.0 | <5 | 12 | < 0.79 (0.26 - 1.85) | 0.32 (0.17 - 0.57) |
| Meta-analysis | 395,354 | 516,323 | 281.0 | 371.8 | 380 | 695 | 1.35 (1.22 - 1.50) | 1.87 (1.73 - 2.01) |
| Meta-analysis (w/o USOC) | 66,892 | 95,370 | 37.9 | 58.7 | 91 | 162 | 2.40 (1.93 - 2.95) | 2.76 (2.35 - 3.22) |

##### 4.2.19 Hypotension

**Supplementary Table 334:** Exposures, outcomes and risk estimates for Hypotension in SGLT2I (male) (T) vs DPP4I (male) (C) new-user cohorts under four different propensity score (PS) adjustment / time-at-risk (TAR) designs across data sources. We report total patient exposure time in 1,000 patient-year (KYr) units, uncalibrated and calibrated hazard ratios (HRs) and their 95% confidence intervals (CIs). Greyed out entries did not pass study diagnostics and are included only for completeness.

| | Patients | | Exposure (KYr) | | Outcomes | | Incidence (per KYr) | |
|---|---|---|---|---|---|---|---|---|
| | T | C | T | C | T | C | T | C |
| **PS-matching / on-treatment TAR** | | | | | | | | |
| CCAE | 19,373 | 30,593 | 17.2 | 25.7 | 150 | 234 | 8.70 (7.36 - 10.2) | 9.10 (7.97 - 10.3) |
| DAG | 4,506 | 10,904 | 3.9 | 11.3 | 17 | 59 | 4.34 (2.53 - 6.95) | 5.21 (3.97 - 6.73) |
| ODOD | 11,998 | 20,309 | 9.1 | 15.8 | 187 | 360 | 20.5 (17.7 - 23.7) | 22.8 (20.5 - 25.3) |
| MDCR | 1,613 | 4,915 | 1.4 | 4.6 | 29 | 133 | 20.4 (13.6 - 29.2) | 29.0 (24.3 - 34.4) |
| MDCD | 1,343 | 3,198 | 0.8 | 2.3 | 20 | 75 | 24.3 (14.8 - 37.5) | 32.4 (25.5 - 40.6) |
| Open Claims | 236,828 | 411,272 | 229.8 | 392.3 | 1,604 | 3,732 | 6.98 (6.64 - 7.33) | 9.51 (9.21 - 9.82) |
| OEHR | 16,524 | 31,578 | 5.4 | 16.1 | 74 | 287 | 13.8 (10.8 - 17.3) | 17.8 (15.8 - 20.0) |
| VA | 7,264 | 22,448 | 4.9 | 20.1 | 91 | 259 | 18.4 (14.9 - 22.7) | 12.9 (11.4 - 14.6) |
| SIDIAP | 4,311 | 19,685 | 6.3 | 44.7 | 19 | 93 | 3.01 (1.81 - 4.71) | 2.08 (1.68 - 2.55) |
| Meta-analysis | 302,417 | 551,704 | 278.1 | 530.6 | 2,171 | 5,157 | 7.81 (7.48 - 8.14) | 9.72 (9.46 - 9.99) |
| Meta-analysis (w/o USOC) | 65,589 | 140,432 | 48.3 | 138.4 | 567 | 1,425 | 11.7 (10.8 - 12.7) | 10.3 (9.77 - 10.8) |
| **PS-stratification / on-treatment TAR** | | | | | | | | |
| CCAE | 23,656 | 30,615 | 19.6 | 25.7 | 177 | 234 | 9.02 (7.74 - 10.4) | 9.10 (7.97 - 10.3) |
| DAG | 5,584 | 10,935 | 4.6 | 11.4 | 25 | 60 | 5.48 (3.55 - 8.09) | 5.28 (4.03 - 6.80) |
| ODOD | 16,736 | 20,394 | 11.6 | 15.9 | 229 | 369 | 19.8 (17.3 - 22.5) | 23.2 (20.9 - 25.7) |
| MDCR | 2,017 | 4,966 | 1.6 | 4.6 | 35 | 138 | 21.8 (15.2 - 30.3) | 29.8 (25.0 - 35.2) |
| MDCD | 1,415 | 3,209 | 0.8 | 2.3 | 21 | 77 | 25.0 (15.4 - 38.1) | 33.1 (26.2 - 41.4) |
| Open Claims | 321,381 | 411,524 | 279.9 | 392.4 | 1,989 | 3,746 | 7.11 (6.80 - 7.43) | 9.55 (9.24 - 9.86) |
| OEHR | 19,804 | 31,642 | 6.3 | 16.2 | 83 | 295 | 13.2 (10.5 - 16.3) | 18.3 (16.2 - 20.5) |
| VA | 11,420 | 23,485 | 6.9 | 21.6 | 152 | 277 | 22.2 (18.8 - 26.0) | 12.8 (11.4 - 14.4) |
| SIDIAP | 5,126 | 20,114 | 7.0 | 46.3 | 23 | 96 | 3.28 (2.08 - 4.92) | 2.07 (1.68 - 2.53) |
| Meta-analysis | 394,304 | 530,190 | 330.6 | 512.5 | 2,561 | 4,938 | 7.75 (7.45 - 8.05) | 9.64 (9.37 - 9.91) |
| Meta-analysis (w/o USOC) | 72,923 | 118,666 | 50.7 | 120.0 | 572 | 1,192 | 11.3 (10.4 - 12.2) | 9.93 (9.37 - 10.5) |
| **PS-matching / on-treatment TAR without escalation** | | | | | | | | |
| CCAE | 19,373 | 30,593 | 14.9 | 20.8 | 121 | 191 | 8.09 (6.72 - 9.67) | 9.17 (7.92 - 10.6) |
| DAG | 4,506 | 10,904 | 3.6 | 10.2 | 14 | 53 | 3.90 (2.13 - 6.55) | 5.19 (3.88 - 6.78) |
| ODOD | 11,998 | 20,309 | 8.0 | 13.0 | 167 | 274 | 20.9 (17.8 - 24.3) | 21.1 (18.7 - 23.8) |
| MDCR | 1,613 | 4,915 | 1.3 | 3.9 | 24 | 115 | 18.9 (12.1 - 28.1) | 29.5 (24.4 - 35.5) |
| MDCD | 1,343 | 3,198 | 0.7 | 1.7 | 16 | 58 | 22.9 (13.1 - 37.2) | 33.3 (25.3 - 43.1) |
| Open Claims | 236,828 | 411,272 | 192.9 | 304.4 | 1,321 | 2,896 | 6.85 (6.48 - 7.23) | 9.51 (9.17 - 9.87) |
| OEHR | 16,524 | 31,578 | 4.7 | 13.1 | 58 | 204 | 12.3 (9.36 - 15.9) | 15.6 (13.5 - 17.8) |
| VA | 7,264 | 22,448 | 4.2 | 16.4 | 73 | 189 | 17.4 (13.6 - 21.8) | 11.5 (9.94 - 13.3) |
| SIDIAP | 4,311 | 19,685 | 5.6 | 35.5 | 17 | 79 | 3.04 (1.77 - 4.87) | 2.22 (1.76 - 2.77) |
| Meta-analysis | 302,417 | 551,704 | 235.3 | 417.3 | 1,795 | 4,001 | 7.63 (7.28 - 7.99) | 9.59 (9.29 - 9.89) |
| Meta-analysis (w/o USOC) | 65,589 | 140,432 | 42.3 | 112.9 | 474 | 1,105 | 11.2 (10.2 - 12.3) | 9.79 (9.22 - 10.4) |
| **PS-stratification / on-treatment TAR without escalation** | | | | | | | | |
| CCAE | 23,656 | 30,615 | 17.2 | 20.8 | 146 | 191 | 8.51 (7.18 - 10.0) | 9.17 (7.92 - 10.6) |
| DAG | 5,584 | 10,935 | 4.2 | 10.3 | 22 | 54 | 5.25 (3.29 - 7.96) | 5.26 (3.95 - 6.87) |
| ODOD | 16,736 | 20,394 | 10.3 | 13.0 | 204 | 282 | 19.9 (17.2 - 22.8) | 21.7 (19.2 - 24.3) |
| MDCR | 2,017 | 4,966 | 1.4 | 3.9 | 29 | 120 | 20.2 (13.5 - 29.0) | 30.5 (25.3 - 36.4) |
| MDCD | 1,415 | 3,209 | 0.7 | 1.8 | 17 | 59 | 23.8 (13.8 - 38.1) | 33.7 (25.7 - 43.5) |
| Open Claims | 321,381 | 411,524 | 238.2 | 304.6 | 1,667 | 2,908 | 7.00 (6.67 - 7.34) | 9.55 (9.20 - 9.90) |
| OEHR | 19,804 | 31,642 | 5.5 | 13.1 | 65 | 208 | 11.7 (9.04 - 14.9) | 15.8 (13.8 - 18.1) |
| VA | 11,420 | 23,485 | 5.9 | 17.6 | 129 | 203 | 21.7 (18.1 - 25.8) | 11.6 (10.0 - 13.3) |
| SIDIAP | 5,126 | 20,114 | 6.3 | 36.7 | 20 | 82 | 3.20 (1.95 - 4.94) | 2.24 (1.78 - 2.77) |
| Meta-analysis | 394,304 | 530,190 | 283.1 | 402.4 | 2,153 | 3,845 | 7.61 (7.29 - 7.93) | 9.55 (9.25 - 9.86) |
| Meta-analysis (w/o USOC) | 72,923 | 118,666 | 44.9 | 97.9 | 486 | 937 | 10.8 (9.89 - 11.8) | 9.57 (8.97 - 10.2) |

##### 4.2.20 Joint pain

**Supplementary Table 335:** Exposures, outcomes and risk estimates for Joint pain in SGLT2I (male) (T) vs DPP4I (male) (C) new-user cohorts under four different propensity score (PS) adjustment / time-at-risk (TAR) designs across data sources. We report total patient exposure time in 1,000 patient-year (KYr) units, uncalibrated and calibrated hazard ratios (HRs) and their 95% confidence intervals (CIs). Greyed out entries did not pass study diagnostics and are included only for completeness.

| | Patients | | Exposure (KYr) | | Outcomes | | Incidence (per KYr) | |
|---|---|---|---|---|---|---|---|---|
| | T | C | T | C | T | C | T | C |
| **PS-matching / on-treatment TAR** | | | | | | | | |
| CCAE | 18,888 | 29,889 | 16.9 | 25.1 | 118 | 229 | 7.00 (5.80 - 8.39) | 9.11 (7.97 - 10.4) |
| DAG | 3,957 | 9,569 | 3.4 | 9.8 | 58 | 213 | 17.1 (13.0 - 22.1) | 21.8 (19.0 - 24.9) |
| ODOD | 11,934 | 20,464 | 9.1 | 16.0 | 83 | 177 | 9.12 (7.26 - 11.3) | 11.1 (9.51 - 12.8) |
| MDCR | 1,669 | 5,087 | 1.5 | 4.8 | 10 | 49 | 6.78 (3.25 - 12.5) | 10.2 (7.58 - 13.5) |
| MDCD | 1,311 | 3,225 | 0.8 | 2.3 | 8 | 43 | 9.96 (4.30 - 19.6) | 18.4 (13.3 - 24.8) |
| Open Claims | 232,773 | 407,388 | 226.1 | 389.3 | 977 | 2,059 | 4.32 (4.05 - 4.60) | 5.29 (5.06 - 5.52) |
| OEHR | 16,259 | 31,055 | 5.3 | 15.9 | 47 | 120 | 8.94 (6.57 - 11.9) | 7.54 (6.25 - 9.02) |
| VA | 6,862 | 21,136 | 4.7 | 19.3 | 27 | 92 | 5.74 (3.79 - 8.36) | 4.76 (3.84 - 5.84) |
| SIDIAP | 3,476 | 16,242 | 5.1 | 36.3 | 81 | 640 | 15.8 (12.5 - 19.6) | 17.6 (16.3 - 19.0) |
| Meta-analysis | 297,656 | 548,054 | 273.8 | 525.7 | 1,411 | 3,619 | 5.15 (4.89 - 5.43) | 6.88 (6.66 - 7.11) |
| Meta-analysis (w/o USOC) | 64,883 | 140,666 | 47.7 | 136.5 | 434 | 1,560 | 9.09 (8.26 - 9.99) | 11.4 (10.9 - 12.0) |
| **PS-stratification / on-treatment TAR** | | | | | | | | |
| CCAE | 23,075 | 29,911 | 19.1 | 25.1 | 137 | 229 | 7.16 (6.01 - 8.46) | 9.11 (7.97 - 10.4) |
| DAG | 4,896 | 9,596 | 4.0 | 9.8 | 60 | 213 | 15.2 (11.6 - 19.5) | 21.7 (18.9 - 24.8) |
| ODOD | 16,688 | 20,572 | 11.6 | 16.1 | 103 | 177 | 8.91 (7.27 - 10.8) | 11.0 (9.45 - 12.8) |
| MDCR | 2,062 | 5,143 | 1.6 | 4.8 | 11 | 49 | 6.70 (3.34 - 12.0) | 10.1 (7.49 - 13.4) |
| MDCD | 1,397 | 3,238 | 0.8 | 2.3 | 9 | 43 | 10.9 (5.00 - 20.7) | 18.3 (13.3 - 24.7) |
| Open Claims | 317,231 | 407,695 | 276.1 | 389.5 | 1,151 | 2,059 | 4.17 (3.93 - 4.42) | 5.29 (5.06 - 5.52) |
| OEHR | 19,457 | 31,120 | 6.2 | 15.9 | 51 | 120 | 8.28 (6.16 - 10.9) | 7.53 (6.24 - 9.00) |
| VA | 10,977 | 21,929 | 6.6 | 20.5 | 46 | 99 | 6.93 (5.08 - 9.25) | 4.83 (3.93 - 5.88) |
| SIDIAP | 4,147 | 16,590 | 5.7 | 37.6 | 87 | 668 | 15.2 (12.2 - 18.7) | 17.8 (16.5 - 19.2) |
| Meta-analysis | 387,556 | 520,627 | 324.3 | 498.9 | 1,600 | 3,515 | 4.93 (4.69 - 5.18) | 7.05 (6.82 - 7.28) |
| Meta-analysis (w/o USOC) | 70,325 | 112,932 | 48.2 | 109.4 | 449 | 1,456 | 9.32 (8.48 - 10.2) | 13.3 (12.6 - 14.0) |
| **PS-matching / on-treatment TAR without escalation** | | | | | | | | |
| CCAE | 18,888 | 29,889 | 14.6 | 20.4 | 100 | 183 | 6.86 (5.58 - 8.34) | 8.98 (7.72 - 10.4) |
| DAG | 3,957 | 9,569 | 3.1 | 8.8 | 52 | 201 | 16.7 (12.5 - 21.9) | 22.7 (19.7 - 26.1) |
| ODOD | 11,934 | 20,464 | 8.0 | 13.1 | 77 | 152 | 9.63 (7.60 - 12.0) | 11.6 (9.82 - 13.6) |
| MDCR | 1,669 | 5,087 | 1.3 | 4.1 | 9 | 43 | 6.86 (3.13 - 13.0) | 10.6 (7.66 - 14.3) |
| MDCD | 1,311 | 3,225 | 0.7 | 1.8 | 7 | 29 | 10.3 (4.14 - 21.2) | 16.3 (10.9 - 23.4) |
| Open Claims | 232,773 | 407,388 | 189.7 | 302.3 | 813 | 1,564 | 4.29 (4.00 - 4.59) | 5.17 (4.92 - 5.44) |
| OEHR | 16,259 | 31,055 | 4.6 | 12.9 | 41 | 91 | 8.91 (6.40 - 12.1) | 7.06 (5.68 - 8.66) |
| VA | 6,862 | 21,136 | 4.0 | 15.8 | 20 | 71 | 5.01 (3.06 - 7.74) | 4.51 (3.52 - 5.68) |
| SIDIAP | 3,476 | 16,242 | 4.6 | 28.9 | 71 | 516 | 15.6 (12.2 - 19.7) | 17.8 (16.3 - 19.5) |
| Meta-analysis | 297,656 | 548,054 | 231.5 | 413.9 | 1,193 | 2,851 | 5.15 (4.87 - 5.45) | 6.89 (6.64 - 7.15) |
| Meta-analysis (w/o USOC) | 64,883 | 140,666 | 41.8 | 111.6 | 380 | 1,287 | 9.10 (8.20 - 10.1) | 11.5 (10.9 - 12.2) |
| **PS-stratification / on-treatment TAR without escalation** | | | | | | | | |
| CCAE | 23,075 | 29,911 | 16.7 | 20.4 | 115 | 183 | 6.88 (5.68 - 8.26) | 8.97 (7.72 - 10.4) |
| DAG | 4,896 | 9,596 | 3.6 | 8.9 | 54 | 201 | 14.8 (11.2 - 19.4) | 22.7 (19.6 - 26.0) |
| ODOD | 16,688 | 20,572 | 10.3 | 13.2 | 95 | 152 | 9.26 (7.49 - 11.3) | 11.5 (9.75 - 13.5) |
| MDCR | 2,062 | 5,143 | 1.5 | 4.1 | 10 | 43 | 6.82 (3.27 - 12.6) | 10.5 (7.57 - 14.1) |
| MDCD | 1,397 | 3,238 | 0.7 | 1.8 | 8 | 29 | 11.5 (4.95 - 22.6) | 16.2 (10.8 - 23.2) |
| Open Claims | 317,231 | 407,695 | 234.9 | 302.4 | 972 | 1,564 | 4.14 (3.88 - 4.41) | 5.17 (4.92 - 5.43) |
| OEHR | 19,457 | 31,120 | 5.4 | 12.9 | 45 | 91 | 8.30 (6.05 - 11.1) | 7.04 (5.67 - 8.65) |
| VA | 10,977 | 21,929 | 5.7 | 16.7 | 36 | 78 | 6.29 (4.40 - 8.71) | 4.68 (3.70 - 5.85) |
| SIDIAP | 4,147 | 16,590 | 5.1 | 29.8 | 76 | 536 | 14.9 (11.7 - 18.6) | 18.0 (16.5 - 19.5) |
| Meta-analysis | 387,556 | 520,627 | 277.5 | 391.8 | 1,367 | 2,770 | 4.93 (4.67 - 5.19) | 7.07 (6.81 - 7.34) |
| Meta-analysis (w/o USOC) | 70,325 | 112,932 | 42.6 | 89.3 | 395 | 1,206 | 9.27 (8.38 - 10.2) | 13.5 (12.7 - 14.3) |

##### 4.2.21 Lower extremity amputation

**Supplementary Table 336:** Exposures, outcomes and risk estimates for Lower extremity amputation in SGLT2I (male) (T) vs DPP4I (male) (C) new-user cohorts under four different propensity score (PS) adjustment / time-at-risk (TAR) designs across data sources. We report total patient exposure time in 1,000 patient-year (KYr) units, uncalibrated and calibrated hazard ratios (HRs) and their 95% confidence intervals (CIs). Greyed out entries did not pass study diagnostics and are included only for completeness.

| | Patients | | Exposure (KYr) | | Outcomes | | Incidence (per KYr) | |
|---|---|---|---|---|---|---|---|---|
| | T | C | T | C | T | C | T | C |
| **PS-matching / on-treatment TAR** | | | | | | | | |
| CCAE | 19,732 | 31,141 | 17.6 | 26.4 | 19 | 20 | 1.08 (0.65 - 1.68) | 0.76 (0.46 - 1.17) |
| DAG | 4,640 | 11,168 | 4.1 | 11.6 | 0 | 0 | | |
| ODOD | 12,486 | 21,414 | 9.6 | 16.9 | 10 | 22 | 1.05 (0.50 - 1.93) | 1.30 (0.82 - 1.97) |
| MDCR | 1,717 | 5,253 | 1.5 | 5.0 | <5 | <5 | < 3.29 (1.07 - 7.67) | < 1.01 (0.33 - 2.35) |
| MDCD | 1,423 | 3,450 | 0.9 | 2.5 | <5 | 9 | < 5.67 (1.84 - 13.2) | 3.56 (1.63 - 6.75) |
| Open Claims | 243,789 | 425,547 | 236.7 | 408.0 | 135 | 242 | 0.57 (0.48 - 0.67) | 0.59 (0.52 - 0.67) |
| OEHR | 16,859 | 32,311 | 5.5 | 16.7 | <5 | 13 | < 0.91 (0.29 - 2.12) | 0.78 (0.41 - 1.33) |
| VA | 7,780 | 23,914 | 5.3 | 21.7 | <10 | 13 | < 1.88 (0.90 - 3.47) | 0.60 (0.32 - 1.03) |
| SIDIAP | 4,337 | 19,807 | 6.3 | 45.0 | 0 | 14 | | 0.31 (0.17 - 0.52) |
| Meta-analysis | 283,787 | 502,016 | 269.2 | 472.9 | <174 | 297 | < 0.65 (0.55 - 0.75) | 0.63 (0.56 - 0.70) |
| Meta-analysis (w/o USOC) | 39,998 | 76,469 | 32.5 | 64.9 | <39 | 55 | < 1.20 (0.85 - 1.64) | 0.85 (0.64 - 1.10) |
| **PS-stratification / on-treatment TAR** | | | | | | | | |
| CCAE | 24,122 | 31,164 | 20.1 | 26.4 | 19 | 20 | 0.95 (0.57 - 1.48) | 0.76 (0.46 - 1.17) |
| DAG | 5,763 | 11,200 | 4.7 | 11.7 | 0 | 0 | | |
| ODOD | 17,448 | 21,537 | 12.1 | 17.0 | 13 | 22 | 1.07 (0.57 - 1.83) | 1.29 (0.81 - 1.96) |
| MDCR | 2,129 | 5,312 | 1.7 | 5.0 | <5 | <5 | < 2.94 (0.95 - 6.86) | < 0.99 (0.32 - 2.32) |
| MDCD | 1,503 | 3,464 | 0.9 | 2.5 | <5 | 9 | < 5.55 (1.80 - 12.9) | 3.54 (1.62 - 6.72) |
| Open Claims | 332,242 | 425,871 | 289.1 | 408.2 | 159 | 243 | 0.55 (0.47 - 0.64) | 0.60 (0.52 - 0.67) |
| OEHR | 20,221 | 32,382 | 6.5 | 16.8 | <5 | 13 | < 0.77 (0.25 - 1.81) | 0.78 (0.41 - 1.33) |
| VA | 12,506 | 24,952 | 7.5 | 23.2 | 11 | 13 | 1.46 (0.73 - 2.62) | 0.56 (0.30 - 0.96) |
| SIDIAP | 5,166 | 20,241 | 7.1 | 46.6 | 0 | 16 | | 0.34 (0.20 - 0.56) |
| Meta-analysis | 394,033 | 510,954 | 327.7 | 468.4 | <196 | 298 | < 0.60 (0.52 - 0.69) | 0.64 (0.57 - 0.71) |
| Meta-analysis (w/o USOC) | 61,791 | 85,083 | 38.7 | 60.1 | <37 | 55 | < 0.96 (0.67 - 1.32) | 0.91 (0.69 - 1.19) |
| **PS-matching / on-treatment TAR without escalation** | | | | | | | | |
| CCAE | 19,732 | 31,141 | 15.3 | 21.3 | 14 | 17 | 0.92 (0.50 - 1.54) | 0.80 (0.46 - 1.28) |
| DAG | 4,640 | 11,168 | 3.7 | 10.5 | 0 | 0 | | |
| ODOD | 12,486 | 21,414 | 8.4 | 13.8 | 8 | 16 | 0.95 (0.41 - 1.88) | 1.16 (0.66 - 1.88) |
| MDCR | 1,717 | 5,253 | 1.4 | 4.2 | <5 | <5 | < 3.69 (1.20 - 8.62) | < 1.19 (0.38 - 2.77) |
| MDCD | 1,423 | 3,450 | 0.7 | 1.9 | <5 | 7 | < 6.71 (2.18 - 15.7) | 3.68 (1.48 - 7.58) |
| Open Claims | 243,789 | 425,547 | 198.5 | 316.5 | 97 | 183 | 0.49 (0.40 - 0.60) | 0.58 (0.50 - 0.67) |
| OEHR | 16,859 | 32,311 | 4.8 | 13.5 | <5 | 10 | < 1.04 (0.34 - 2.42) | 0.74 (0.35 - 1.36) |
| VA | 7,780 | 23,914 | 4.5 | 17.6 | <10 | 11 | < 2.21 (1.06 - 4.07) | 0.62 (0.31 - 1.12) |
| SIDIAP | 4,337 | 19,807 | 5.6 | 35.8 | 0 | 11 | | 0.31 (0.15 - 0.55) |
| Meta-analysis | 276,007 | 478,102 | 222.2 | 351.6 | 119 | 216 | 0.54 (0.44 - 0.64) | 0.61 (0.54 - 0.70) |
| Meta-analysis (w/o USOC) | 32,218 | 52,555 | 23.7 | 35.2 | 22 | 33 | 0.93 (0.58 - 1.41) | 0.94 (0.65 - 1.32) |
| **PS-stratification / on-treatment TAR without escalation** | | | | | | | | |
| CCAE | 24,122 | 31,164 | 17.5 | 21.3 | 14 | 17 | 0.80 (0.44 - 1.34) | 0.80 (0.46 - 1.28) |
| DAG | 5,763 | 11,200 | 4.3 | 10.6 | 0 | 0 | | |
| ODOD | 17,448 | 21,537 | 10.8 | 13.9 | 10 | 16 | 0.93 (0.45 - 1.71) | 1.15 (0.66 - 1.87) |
| MDCR | 2,129 | 5,312 | 1.5 | 4.3 | <5 | <5 | < 3.29 (1.07 - 7.67) | < 1.17 (0.38 - 2.73) |
| MDCD | 1,503 | 3,464 | 0.8 | 1.9 | <5 | 7 | < 6.56 (2.13 - 15.3) | 3.66 (1.47 - 7.53) |
| Open Claims | 332,242 | 425,871 | 245.8 | 316.7 | 116 | 184 | 0.47 (0.39 - 0.57) | 0.58 (0.50 - 0.67) |
| OEHR | 20,221 | 32,382 | 5.7 | 13.5 | <5 | 10 | < 0.88 (0.29 - 2.05) | 0.74 (0.35 - 1.36) |
| VA | 12,506 | 24,952 | 6.5 | 18.8 | 11 | 11 | 1.69 (0.84 - 3.02) | 0.58 (0.29 - 1.05) |
| SIDIAP | 5,166 | 20,241 | 6.3 | 37.0 | 0 | 13 | | 0.35 (0.19 - 0.60) |
| Meta-analysis | 373,812 | 478,572 | 274.1 | 351.9 | 140 | 217 | 0.51 (0.43 - 0.60) | 0.62 (0.54 - 0.70) |
| Meta-analysis (w/o USOC) | 41,570 | 52,701 | 28.3 | 35.3 | 24 | 33 | 0.85 (0.54 - 1.26) | 0.94 (0.64 - 1.31) |

###### 4.2.22 Nausea

**Supplementary Table 337:** Exposures, outcomes and risk estimates for Nausea in SGLT2I (male) (T) vs DPP4I (male) (C) new-user cohorts under four different propensity score (PS) adjustment / time-at-risk (TAR) designs across data sources. We report total patient exposure time in 1,000 patient-year (KYr) units, uncalibrated and calibrated hazard ratios (HRs) and their 95% confidence intervals (CIs). Greyed out entries did not pass study diagnostics and are included only for completeness.

| | Patients | | Exposure (KYr) | | Outcomes | | Incidence (per KYr) | |
|---|---|---|---|---|---|---|---|---|
| | T | C | T | C | T | C | T | C |
| **PS-matching / on-treatment TAR** | | | | | | | | |
| CCAE | 17,863 | 28,402 | 15.7 | 23.6 | 384 | 640 | 24.5 (22.1 - 27.1) | 27.1 (25.0 - 29.3) |
| DAG | 4,388 | 10,521 | 3.8 | 10.9 | 28 | 94 | 7.28 (4.84 - 10.5) | 8.61 (6.96 - 10.5) |
| ODOD | 11,162 | 19,099 | 8.5 | 14.8 | 238 | 494 | 28.1 (24.6 - 31.9) | 33.5 (30.6 - 36.6) |
| MDCR | 1,574 | 4,783 | 1.4 | 4.5 | 35 | 117 | 25.1 (17.5 - 34.9) | 26.1 (21.6 - 31.2) |
| MDCD | 1,144 | 2,754 | 0.7 | 2.0 | 43 | 122 | 62.4 (45.1 - 84.0) | 62.0 (51.5 - 74.0) |
| Open Claims | 220,752 | 387,306 | 214.5 | 369.1 | 2,628 | 5,718 | 12.3 (11.8 - 12.7) | 15.5 (15.1 - 15.9) |
| OEHR | 15,676 | 30,166 | 5.1 | 15.3 | 142 | 398 | 28.0 (23.6 - 33.0) | 26.1 (23.6 - 28.7) |
| VA | 7,229 | 22,387 | 4.9 | 20.3 | 31 | 168 | 6.27 (4.26 - 8.90) | 8.26 (7.06 - 9.61) |
| SIDIAP | 4,206 | 19,292 | 6.2 | 43.8 | 16 | 224 | 2.58 (1.48 - 4.19) | 5.12 (4.47 - 5.84) |
| Meta-analysis | 282,850 | 521,956 | 260.1 | 502.3 | 3,502 | 7,853 | 13.5 (13.0 - 13.9) | 15.6 (15.3 - 16.0) |
| Meta-analysis (w/o USOC) | 62,098 | 134,650 | 45.6 | 133.2 | 874 | 2,135 | 19.2 (17.9 - 20.5) | 16.0 (15.4 - 16.7) |
| **PS-stratification / on-treatment TAR** | | | | | | | | |
| CCAE | 21,821 | 28,422 | 17.8 | 23.6 | 432 | 640 | 24.2 (22.0 - 26.6) | 27.1 (25.0 - 29.3) |
| DAG | 5,453 | 10,548 | 4.5 | 11.0 | 34 | 94 | 7.61 (5.27 - 10.6) | 8.58 (6.93 - 10.5) |
| ODOD | 15,574 | 19,195 | 10.7 | 14.8 | 302 | 502 | 28.2 (25.1 - 31.5) | 33.8 (30.9 - 36.9) |
| MDCR | 1,958 | 4,835 | 1.5 | 4.5 | 38 | 120 | 24.5 (17.3 - 33.7) | 26.4 (21.9 - 31.6) |
| MDCD | 1,195 | 2,764 | 0.7 | 2.0 | 43 | 122 | 61.5 (44.5 - 82.9) | 61.7 (51.2 - 73.6) |
| Open Claims | 299,241 | 387,604 | 261.1 | 369.3 | 3,193 | 5,727 | 12.2 (11.8 - 12.7) | 15.5 (15.1 - 15.9) |
| OEHR | 18,784 | 30,230 | 5.9 | 15.3 | 162 | 402 | 27.3 (23.3 - 31.8) | 26.3 (23.8 - 29.0) |
| VA | 11,542 | 23,328 | 7.0 | 21.7 | 53 | 181 | 7.61 (5.70 - 9.96) | 8.34 (7.17 - 9.64) |
| SIDIAP | 5,004 | 19,705 | 6.9 | 45.2 | 20 | 236 | 2.90 (1.77 - 4.48) | 5.22 (4.57 - 5.93) |
| Meta-analysis | 367,835 | 500,539 | 308.5 | 483.8 | 4,181 | 7,721 | 13.6 (13.1 - 14.0) | 16.0 (15.6 - 16.3) |
| Meta-analysis (w/o USOC) | 68,594 | 112,935 | 47.4 | 114.5 | 988 | 1,994 | 20.8 (19.6 - 22.2) | 17.4 (16.7 - 18.2) |
| **PS-matching / on-treatment TAR without escalation** | | | | | | | | |
| CCAE | 17,863 | 28,402 | 13.7 | 19.2 | 318 | 526 | 23.3 (20.8 - 26.0) | 27.4 (25.1 - 29.8) |
| DAG | 4,388 | 10,521 | 3.5 | 9.9 | 26 | 84 | 7.38 (4.82 - 10.8) | 8.51 (6.79 - 10.5) |
| ODOD | 11,162 | 19,099 | 7.5 | 12.2 | 201 | 399 | 26.8 (23.2 - 30.8) | 32.8 (29.7 - 36.2) |
| MDCR | 1,574 | 4,783 | 1.2 | 3.8 | 32 | 105 | 25.6 (17.5 - 36.2) | 27.5 (22.5 - 33.3) |
| MDCD | 1,144 | 2,754 | 0.6 | 1.5 | 37 | 97 | 63.2 (44.5 - 87.1) | 64.8 (52.6 - 79.1) |
| Open Claims | 220,752 | 387,306 | 180.6 | 287.5 | 2,065 | 4,359 | 11.4 (10.9 - 11.9) | 15.2 (14.7 - 15.6) |
| OEHR | 15,676 | 30,166 | 4.5 | 12.5 | 114 | 291 | 25.6 (21.1 - 30.7) | 23.3 (20.7 - 26.2) |
| VA | 7,229 | 22,387 | 4.2 | 16.6 | 22 | 137 | 5.21 (3.26 - 7.89) | 8.25 (6.92 - 9.75) |
| SIDIAP | 4,206 | 19,292 | 5.5 | 34.8 | 14 | 186 | 2.55 (1.39 - 4.28) | 5.34 (4.60 - 6.17) |
| Meta-analysis | 282,850 | 521,956 | 220.7 | 396.5 | 2,792 | 6,087 | 12.6 (12.2 - 13.1) | 15.4 (15.0 - 15.7) |
| Meta-analysis (w/o USOC) | 62,098 | 134,650 | 40.1 | 108.9 | 727 | 1,728 | 18.1 (16.8 - 19.5) | 15.9 (15.1 - 16.6) |
| **PS-stratification / on-treatment TAR without escalation** | | | | | | | | |
| CCAE | 21,821 | 28,422 | 15.7 | 19.2 | 359 | 526 | 22.9 (20.6 - 25.4) | 27.4 (25.1 - 29.8) |
| DAG | 5,453 | 10,548 | 4.1 | 9.9 | 31 | 84 | 7.55 (5.13 - 10.7) | 8.48 (6.76 - 10.5) |
| ODOD | 15,574 | 19,195 | 9.6 | 12.2 | 256 | 407 | 26.8 (23.6 - 30.2) | 33.3 (30.1 - 36.7) |
| MDCR | 1,958 | 4,835 | 1.4 | 3.9 | 35 | 108 | 25.2 (17.5 - 35.0) | 28.0 (23.0 - 33.8) |
| MDCD | 1,195 | 2,764 | 0.6 | 1.5 | 37 | 97 | 62.3 (43.9 - 85.9) | 64.5 (52.3 - 78.6) |
| Open Claims | 299,241 | 387,604 | 222.9 | 287.7 | 2,542 | 4,368 | 11.4 (11.0 - 11.9) | 15.2 (14.7 - 15.6) |
| OEHR | 18,784 | 30,230 | 5.2 | 12.5 | 130 | 294 | 24.8 (20.7 - 29.5) | 23.5 (20.9 - 26.4) |
| VA | 11,542 | 23,328 | 6.0 | 17.7 | 41 | 148 | 6.80 (4.88 - 9.22) | 8.37 (7.07 - 9.83) |
| SIDIAP | 5,004 | 19,705 | 6.1 | 35.9 | 18 | 196 | 2.93 (1.74 - 4.63) | 5.46 (4.72 - 6.28) |
| Meta-analysis | 367,835 | 500,539 | 265.0 | 381.4 | 3,371 | 5,983 | 12.7 (12.3 - 13.2) | 15.7 (15.3 - 16.1) |
| Meta-analysis (w/o USOC) | 68,594 | 112,935 | 42.1 | 93.6 | 829 | 1,615 | 19.7 (18.4 - 21.1) | 17.2 (16.4 - 18.1) |

##### 4.2.23 Peripheral edema

**Supplementary Table 338:** Exposures, outcomes and risk estimates for Peripheral edema in SGLT2I (male) (T) vs DPP4I (male) (C) new-user cohorts under four different propensity score (PS) adjustment / time-at-risk (TAR) designs across data sources. We report total patient exposure time in 1,000 patient-year (KYr) units, uncalibrated and calibrated hazard ratios (HRs) and their 95% confidence intervals (CIs). Greyed out entries did not pass study diagnostics and are included only for completeness.

| | Patients | | Exposure (KYr) | | Outcomes | | Incidence (per KYr) | |
|---|---|---|---|---|---|---|---|---|
| | T | C | T | C | T | C | T | C |
| **PS-matching / on-treatment TAR** | | | | | | | | |
| CCAE | 16,678 | 26,547 | 14.5 | 21.8 | 457 | 817 | 31.5 (28.7 - 34.6) | 37.5 (35.0 - 40.2) |
| DAG | 4,104 | 9,849 | 3.5 | 10.1 | 48 | 235 | 13.6 (10.0 - 18.0) | 23.3 (20.4 - 26.5) |
| ODOD | 10,187 | 16,998 | 7.7 | 12.9 | 338 | 813 | 44.1 (39.5 - 49.1) | 63.2 (58.9 - 67.7) |
| MDCR | 1,301 | 3,926 | 1.1 | 3.6 | 50 | 285 | 44.6 (33.1 - 58.8) | 80.2 (71.1 - 90.0) |
| MDCD | 1,057 | 2,501 | 0.6 | 1.7 | 37 | 135 | 59.1 (41.6 - 81.5) | 78.9 (66.2 - 93.4) |
| Open Claims | 200,483 | 347,984 | 193.9 | 325.3 | 3,470 | 9,842 | 17.9 (17.3 - 18.5) | 30.3 (29.7 - 30.9) |
| OEHR | 14,800 | 28,212 | 4.8 | 14.0 | 107 | 540 | 22.4 (18.4 - 27.1) | 38.5 (35.4 - 41.9) |
| VA | 5,997 | 18,905 | 4.0 | 16.8 | 108 | 570 | 26.7 (21.9 - 32.2) | 34.0 (31.3 - 36.9) |
| SIDIAP | 3,910 | 18,105 | 5.7 | 40.6 | 59 | 694 | 10.3 (7.84 - 13.3) | 17.1 (15.8 - 18.4) |
| Meta-analysis | 259,139 | 476,856 | 236.9 | 452.9 | 4,652 | 13,933 | 19.6 (19.1 - 20.2) | 30.8 (30.3 - 31.3) |
| Meta-analysis (w/o USOC) | 58,656 | 128,872 | 43.0 | 127.6 | 1,182 | 4,091 | 27.5 (25.9 - 29.1) | 32.1 (31.1 - 33.1) |
| **PS-stratification / on-treatment TAR** | | | | | | | | |
| CCAE | 20,331 | 26,564 | 16.5 | 21.8 | 518 | 820 | 31.5 (28.8 - 34.3) | 37.7 (35.1 - 40.3) |
| DAG | 5,059 | 9,872 | 4.1 | 10.1 | 57 | 236 | 13.8 (10.5 - 17.9) | 23.3 (20.5 - 26.5) |
| ODOD | 14,095 | 17,061 | 9.6 | 12.9 | 406 | 820 | 42.2 (38.2 - 46.5) | 63.5 (59.2 - 68.0) |
| MDCR | 1,633 | 3,973 | 1.3 | 3.6 | 56 | 294 | 44.4 (33.5 - 57.7) | 82.0 (72.9 - 91.9) |
| MDCD | 1,095 | 2,510 | 0.6 | 1.7 | 37 | 138 | 58.6 (41.2 - 80.7) | 80.5 (67.6 - 95.1) |
| Open Claims | 269,915 | 348,152 | 235.1 | 325.4 | 4,195 | 9,863 | 17.8 (17.3 - 18.4) | 30.3 (29.7 - 30.9) |
| OEHR | 17,562 | 28,266 | 5.5 | 14.0 | 130 | 544 | 23.5 (19.6 - 27.9) | 38.8 (35.6 - 42.2) |
| VA | 9,289 | 19,618 | 5.6 | 17.8 | 177 | 603 | 31.8 (27.3 - 36.9) | 33.9 (31.3 - 36.7) |
| SIDIAP | 4,631 | 18,482 | 6.4 | 41.9 | 66 | 729 | 10.3 (8.00 - 13.2) | 17.4 (16.1 - 18.7) |
| Meta-analysis | 333,226 | 452,370 | 278.5 | 429.8 | 5,428 | 13,306 | 19.5 (19.0 - 20.0) | 31.0 (30.4 - 31.5) |
| Meta-analysis (w/o USOC) | 63,311 | 104,218 | 43.4 | 104.3 | 1,233 | 3,443 | 28.4 (26.9 - 30.1) | 33.0 (31.9 - 34.1) |
| **PS-matching / on-treatment TAR without escalation** | | | | | | | | |
| CCAE | 16,678 | 26,547 | 12.6 | 17.7 | 381 | 664 | 30.2 (27.2 - 33.3) | 37.4 (34.6 - 40.4) |
| DAG | 4,104 | 9,849 | 3.2 | 9.1 | 45 | 222 | 13.9 (10.1 - 18.6) | 24.4 (21.3 - 27.8) |
| ODOD | 10,187 | 16,998 | 6.8 | 10.6 | 271 | 674 | 39.8 (35.2 - 44.9) | 63.3 (58.6 - 68.3) |
| MDCR | 1,301 | 3,926 | 1.0 | 3.1 | 42 | 244 | 41.7 (30.0 - 56.4) | 79.6 (70.0 - 90.3) |
| MDCD | 1,057 | 2,501 | 0.5 | 1.3 | 30 | 117 | 55.9 (37.7 - 79.8) | 90.2 (74.6 - 108) |
| Open Claims | 200,483 | 347,984 | 163.5 | 253.6 | 2,832 | 7,676 | 17.3 (16.7 - 18.0) | 30.3 (29.6 - 31.0) |
| OEHR | 14,800 | 28,212 | 4.2 | 11.5 | 88 | 399 | 20.9 (16.8 - 25.8) | 34.8 (31.4 - 38.3) |
| VA | 5,997 | 18,905 | 3.5 | 13.7 | 89 | 450 | 25.7 (20.6 - 31.6) | 32.8 (29.8 - 36.0) |
| SIDIAP | 3,910 | 18,105 | 5.1 | 32.3 | 53 | 551 | 10.4 (7.79 - 13.6) | 17.0 (15.7 - 18.5) |
| Meta-analysis | 259,139 | 476,856 | 201.4 | 358.2 | 3,816 | 10,997 | 18.9 (18.3 - 19.6) | 30.7 (30.1 - 31.3) |
| Meta-analysis (w/o USOC) | 58,656 | 128,872 | 37.9 | 104.6 | 984 | 3,321 | 25.9 (24.3 - 27.6) | 31.7 (30.7 - 32.8) |
| **PS-stratification / on-treatment TAR without escalation** | | | | | | | | |
| CCAE | 20,331 | 26,564 | 14.5 | 17.8 | 440 | 667 | 30.4 (27.6 - 33.4) | 37.6 (34.8 - 40.5) |
| DAG | 5,059 | 9,872 | 3.8 | 9.1 | 53 | 223 | 14.0 (10.5 - 18.3) | 24.4 (21.3 - 27.9) |
| ODOD | 14,095 | 17,061 | 8.6 | 10.7 | 336 | 680 | 39.0 (35.0 - 43.4) | 63.6 (58.9 - 68.5) |
| MDCR | 1,633 | 3,973 | 1.1 | 3.1 | 48 | 252 | 42.3 (31.2 - 56.1) | 81.5 (71.8 - 92.2) |
| MDCD | 1,095 | 2,510 | 0.5 | 1.3 | 30 | 120 | 55.3 (37.3 - 79.0) | 92.2 (76.5 - 110) |
| Open Claims | 269,915 | 348,152 | 200.8 | 253.7 | 3,475 | 7,695 | 17.3 (16.7 - 17.9) | 30.3 (29.7 - 31.0) |
| OEHR | 17,562 | 28,266 | 4.9 | 11.5 | 105 | 399 | 21.4 (17.5 - 25.9) | 34.7 (31.4 - 38.3) |
| VA | 9,289 | 19,618 | 4.8 | 14.5 | 151 | 478 | 31.3 (26.5 - 36.7) | 32.9 (30.1 - 36.0) |
| SIDIAP | 4,631 | 18,482 | 5.7 | 33.3 | 60 | 578 | 10.5 (8.03 - 13.6) | 17.3 (16.0 - 18.8) |
| Meta-analysis | 333,226 | 452,370 | 239.4 | 339.2 | 4,517 | 10,494 | 18.9 (18.3 - 19.4) | 30.9 (30.4 - 31.5) |
| Meta-analysis (w/o USOC) | 63,311 | 104,218 | 38.6 | 85.5 | 1,042 | 2,799 | 27.0 (25.4 - 28.7) | 32.7 (31.5 - 34.0) |

##### 4.2.24 Photosensitivity

**Supplementary Table 339:** Exposures, outcomes and risk estimates for Photosensitivity in SGLT2I (male) (T) vs DPP4I (male) (C) new-user cohorts under four different propensity score (PS) adjustment / time-at-risk (TAR) designs across data sources. We report total patient exposure time in 1,000 patient-year (KYr) units, uncalibrated and calibrated hazard ratios (HRs) and their 95% confidence intervals (CIs). Greyed out entries did not pass study diagnostics and are included only for completeness.

| | Patients | | Exposure (KYr) | | Outcomes | | Incidence (per KYr) | |
|---|---|---|---|---|---|---|---|---|
| | T | C | T | C | T | C | T | C |
| **PS-matching / on-treatment TAR** | | | | | | | | |
| CCAE | 19,583 | 30,894 | 17.5 | 26.1 | 10 | 23 | 0.57 (0.27 - 1.05) | 0.88 (0.56 - 1.32) |
| DAG | 4,625 | 11,133 | 4.0 | 11.6 | 0 | <5 | | < 0.43 (0.14 - 1.00) |
| ODOD | 12,399 | 21,234 | 9.5 | 16.7 | 6 | 23 | 0.63 (0.23 - 1.38) | 1.38 (0.87 - 2.07) |
| MDCR | 1,690 | 5,139 | 1.5 | 4.8 | <5 | 14 | < 3.34 (1.08 - 7.79) | 2.90 (1.58 - 4.86) |
| MDCD | 1,413 | 3,447 | 0.9 | 2.5 | 0 | 0 | | |
| Open Claims | 241,800 | 421,909 | 234.6 | 403.9 | 65 | 213 | 0.28 (0.21 - 0.35) | 0.53 (0.46 - 0.60) |
| OEHR | 16,774 | 32,082 | 5.5 | 16.6 | 5 | 21 | 0.91 (0.30 - 2.13) | 1.27 (0.78 - 1.94) |
| VA | 7,694 | 23,539 | 5.2 | 21.2 | <10 | <10 | < 1.91 (0.92 - 3.52) | < 0.47 (0.23 - 0.87) |
| SIDIAP | 4,334 | 19,796 | 6.3 | 45.0 | <5 | <5 | < 0.79 (0.26 - 1.84) | < 0.11 (0.04 - 0.26) |
| Meta-analysis | 290,556 | 506,119 | 267.0 | 463.3 | 86 | 280 | 0.32 (0.26 - 0.40) | 0.60 (0.54 - 0.68) |
| Meta-analysis (w/o USOC) | 48,756 | 84,210 | 32.4 | 59.4 | 21 | 67 | 0.65 (0.40 - 0.99) | 1.13 (0.87 - 1.43) |
| **PS-stratification / on-treatment TAR** | | | | | | | | |
| CCAE | 23,945 | 30,916 | 19.9 | 26.1 | 11 | 23 | 0.55 (0.28 - 0.99) | 0.88 (0.56 - 1.32) |
| DAG | 5,747 | 11,165 | 4.7 | 11.7 | 0 | <5 | | < 0.43 (0.14 - 1.00) |
| ODOD | 17,348 | 21,351 | 12.0 | 16.8 | 8 | 23 | 0.66 (0.29 - 1.31) | 1.37 (0.87 - 2.05) |
| MDCR | 2,105 | 5,194 | 1.7 | 4.9 | <5 | 15 | < 2.99 (0.97 - 6.97) | 3.07 (1.72 - 5.06) |
| MDCD | 1,495 | 3,460 | 0.9 | 2.5 | 0 | 0 | | |
| Open Claims | 329,565 | 422,234 | 286.5 | 404.2 | 68 | 214 | 0.24 (0.18 - 0.30) | 0.53 (0.46 - 0.61) |
| OEHR | 20,107 | 32,216 | 6.4 | 16.6 | 5 | 21 | 0.78 (0.25 - 1.82) | 1.26 (0.78 - 1.93) |
| VA | 12,352 | 24,648 | 7.4 | 22.9 | <10 | <10 | < 1.35 (0.65 - 2.48) | < 0.44 (0.21 - 0.80) |
| SIDIAP | 5,159 | 20,233 | 7.1 | 46.6 | <5 | <5 | < 0.71 (0.23 - 1.65) | < 0.11 (0.03 - 0.25) |
| Meta-analysis | 393,070 | 511,911 | 326.6 | 468.6 | <97 | 296 | < 0.30 (0.24 - 0.36) | 0.63 (0.56 - 0.71) |
| Meta-analysis (w/o USOC) | 63,505 | 89,677 | 40.0 | 64.4 | <29 | 82 | < 0.72 (0.49 - 1.04) | 1.27 (1.01 - 1.58) |
| **PS-matching / on-treatment TAR without escalation** | | | | | | | | |
| CCAE | 19,583 | 30,894 | 15.1 | 21.1 | 9 | 19 | 0.60 (0.27 - 1.13) | 0.90 (0.54 - 1.41) |
| DAG | 4,625 | 11,133 | 3.7 | 10.5 | 0 | <5 | | < 0.48 (0.15 - 1.11) |
| ODOD | 12,399 | 21,234 | 8.3 | 13.7 | 5 | 18 | 0.60 (0.20 - 1.40) | 1.32 (0.78 - 2.08) |
| MDCR | 1,690 | 5,139 | 1.3 | 4.1 | <5 | 13 | < 3.75 (1.22 - 8.76) | 3.16 (1.68 - 5.40) |
| MDCD | 1,413 | 3,447 | 0.7 | 1.9 | 0 | 0 | | |
| Open Claims | 241,800 | 421,909 | 196.7 | 313.3 | 52 | 180 | 0.26 (0.20 - 0.35) | 0.57 (0.49 - 0.66) |
| OEHR | 16,774 | 32,082 | 4.8 | 13.4 | <5 | 19 | < 1.04 (0.34 - 2.43) | 1.42 (0.85 - 2.22) |
| VA | 7,694 | 23,539 | 4.4 | 17.3 | <10 | <10 | < 2.25 (1.08 - 4.14) | < 0.58 (0.28 - 1.06) |
| SIDIAP | 4,334 | 19,796 | 5.6 | 35.8 | <5 | <5 | < 0.89 (0.29 - 2.08) | < 0.14 (0.05 - 0.33) |
| Meta-analysis | 290,556 | 506,119 | 224.9 | 361.4 | <71 | 236 | < 0.32 (0.25 - 0.40) | 0.65 (0.57 - 0.74) |
| Meta-analysis (w/o USOC) | 48,756 | 84,210 | 28.2 | 48.2 | <19 | 56 | < 0.67 (0.41 - 1.05) | 1.16 (0.88 - 1.51) |
| **PS-stratification / on-treatment TAR without escalation** | | | | | | | | |
| CCAE | 23,945 | 30,916 | 17.4 | 21.1 | 10 | 19 | 0.58 (0.28 - 1.06) | 0.90 (0.54 - 1.40) |
| DAG | 5,747 | 11,165 | 4.3 | 10.5 | 0 | <5 | | < 0.48 (0.15 - 1.11) |
| ODOD | 17,348 | 21,351 | 10.7 | 13.8 | 7 | 18 | 0.66 (0.26 - 1.35) | 1.31 (0.77 - 2.07) |
| MDCR | 2,105 | 5,194 | 1.5 | 4.2 | <5 | 14 | < 3.34 (1.08 - 7.80) | 3.37 (1.84 - 5.65) |
| MDCD | 1,495 | 3,460 | 0.8 | 1.9 | 0 | 0 | | |
| Open Claims | 329,565 | 422,234 | 243.6 | 313.5 | 55 | 181 | 0.23 (0.17 - 0.29) | 0.58 (0.50 - 0.67) |
| OEHR | 20,107 | 32,216 | 5.7 | 13.4 | <5 | 19 | < 0.88 (0.29 - 2.06) | 1.41 (0.85 - 2.21) |
| VA | 12,352 | 24,648 | 6.4 | 18.6 | <10 | <10 | < 1.56 (0.75 - 2.87) | < 0.54 (0.26 - 0.99) |
| SIDIAP | 5,159 | 20,233 | 6.3 | 37.0 | <5 | <5 | < 0.79 (0.26 - 1.85) | < 0.14 (0.04 - 0.32) |
| Meta-analysis | 390,965 | 506,717 | 277.3 | 361.8 | <77 | 237 | < 0.28 (0.22 - 0.35) | 0.66 (0.57 - 0.74) |
| Meta-analysis (w/o USOC) | 61,400 | 84,483 | 33.7 | 48.3 | <22 | 56 | < 0.65 (0.41 - 0.99) | 1.16 (0.88 - 1.50) |

##### 4.2.25 Venous thromboembolism

**Supplementary Table 340:** Exposures, outcomes and risk estimates for Venous thromboembolism in SGLT2I (male) (T) vs DPP4I (male) (C) new-user cohorts under four different propensity score (PS) adjustment / time-at-risk (TAR) designs across data sources. We report total patient exposure time in 1,000 patient-year (KYr) units, uncalibrated and calibrated hazard ratios (HRs) and their 95% confidence intervals (CIs). Greyed out entries did not pass study diagnostics and are included only for completeness.

| | Patients | | Exposure (KYr) | | Outcomes | | Incidence (per KYr) | |
|---|---|---|---|---|---|---|---|---|
| | T | C | T | C | T | C | T | C |
| **PS-matching / on-treatment TAR** | | | | | | | | |
| CCAE | 19,330 | 30,468 | 17.2 | 25.7 | 66 | 124 | 3.83 (2.96 - 4.87) | 4.82 (4.01 - 5.75) |
| DAG | 4,400 | 10,540 | 3.8 | 10.9 | 24 | 71 | 6.27 (4.02 - 9.33) | 6.51 (5.09 - 8.21) |
| ODOD | 12,183 | 20,779 | 9.3 | 16.3 | 59 | 147 | 6.34 (4.83 - 8.18) | 9.00 (7.60 - 10.6) |
| MDCR | 1,645 | 5,019 | 1.4 | 4.7 | 12 | 57 | 8.30 (4.29 - 14.5) | 12.0 (9.11 - 15.6) |
| MDCD | 1,375 | 3,292 | 0.8 | 2.4 | 10 | 33 | 11.8 (5.65 - 21.7) | 13.6 (9.39 - 19.2) |
| Open Claims | 236,728 | 412,478 | 229.8 | 394.5 | 783 | 1,936 | 3.41 (3.17 - 3.65) | 4.91 (4.69 - 5.13) |
| OEHR | 16,473 | 31,512 | 5.3 | 16.2 | 36 | 102 | 6.76 (4.73 - 9.35) | 6.30 (5.14 - 7.65) |
| VA | 7,470 | 23,118 | 5.1 | 20.9 | 25 | 97 | 4.92 (3.18 - 7.26) | 4.64 (3.76 - 5.66) |
| SIDIAP | 4,220 | 19,368 | 6.1 | 44.0 | 22 | 170 | 3.58 (2.24 - 5.42) | 3.86 (3.31 - 4.49) |
| Meta-analysis | 304,347 | 560,749 | 280.1 | 542.7 | 1,035 | 2,736 | 3.70 (3.47 - 3.93) | 5.04 (4.85 - 5.23) |
| Meta-analysis (w/o USOC) | 67,619 | 148,271 | 50.2 | 148.3 | 252 | 800 | 5.02 (4.42 - 5.67) | 5.40 (5.03 - 5.78) |
| **PS-stratification / on-treatment TAR** | | | | | | | | |
| CCAE | 23,622 | 30,488 | 19.6 | 25.7 | 77 | 126 | 3.92 (3.09 - 4.90) | 4.90 (4.08 - 5.83) |
| DAG | 5,471 | 10,567 | 4.5 | 10.9 | 27 | 72 | 6.04 (3.98 - 8.79) | 6.58 (5.15 - 8.29) |
| ODOD | 16,980 | 20,888 | 11.8 | 16.4 | 81 | 148 | 6.88 (5.46 - 8.55) | 9.00 (7.61 - 10.6) |
| MDCR | 2,048 | 5,074 | 1.6 | 4.8 | 15 | 57 | 9.27 (5.19 - 15.3) | 11.9 (9.00 - 15.4) |
| MDCD | 1,452 | 3,305 | 0.9 | 2.4 | 10 | 33 | 11.6 (5.55 - 21.3) | 13.6 (9.34 - 19.1) |
| Open Claims | 321,867 | 412,766 | 280.3 | 394.7 | 985 | 1,944 | 3.51 (3.30 - 3.74) | 4.93 (4.71 - 5.15) |
| OEHR | 19,746 | 31,577 | 6.3 | 16.2 | 42 | 102 | 6.71 (4.83 - 9.06) | 6.29 (5.13 - 7.64) |
| VA | 11,951 | 24,130 | 7.2 | 22.4 | 44 | 99 | 6.13 (4.45 - 8.23) | 4.42 (3.59 - 5.38) |
| SIDIAP | 5,024 | 19,780 | 6.9 | 45.5 | 26 | 179 | 3.79 (2.48 - 5.56) | 3.94 (3.38 - 4.56) |
| Meta-analysis | 394,758 | 531,140 | 330.9 | 514.3 | 1,253 | 2,628 | 3.79 (3.58 - 4.00) | 5.11 (4.92 - 5.31) |
| Meta-analysis (w/o USOC) | 72,891 | 118,374 | 50.6 | 119.6 | 268 | 684 | 5.29 (4.68 - 5.97) | 5.72 (5.30 - 6.17) |
| **PS-matching / on-treatment TAR without escalation** | | | | | | | | |
| CCAE | 19,330 | 30,468 | 14.9 | 20.8 | 53 | 106 | 3.55 (2.66 - 4.64) | 5.10 (4.17 - 6.16) |
| DAG | 4,400 | 10,540 | 3.5 | 9.8 | 23 | 66 | 6.55 (4.15 - 9.83) | 6.71 (5.19 - 8.53) |
| ODOD | 12,183 | 20,779 | 8.2 | 13.4 | 44 | 125 | 5.38 (3.91 - 7.22) | 9.35 (7.79 - 11.1) |
| MDCR | 1,645 | 5,019 | 1.3 | 4.0 | 12 | 45 | 9.32 (4.82 - 16.3) | 11.2 (8.15 - 15.0) |
| MDCD | 1,375 | 3,292 | 0.7 | 1.8 | 9 | 28 | 12.5 (5.73 - 23.8) | 15.4 (10.2 - 22.2) |
| Open Claims | 236,728 | 412,478 | 192.9 | 306.3 | 630 | 1,507 | 3.27 (3.02 - 3.53) | 4.92 (4.68 - 5.18) |
| OEHR | 16,473 | 31,512 | 4.7 | 13.1 | 26 | 78 | 5.57 (3.64 - 8.17) | 5.96 (4.71 - 7.43) |
| VA | 7,470 | 23,118 | 4.3 | 17.0 | 21 | 72 | 4.84 (3.00 - 7.40) | 4.23 (3.31 - 5.32) |
| SIDIAP | 4,220 | 19,368 | 5.5 | 34.9 | 19 | 120 | 3.47 (2.09 - 5.42) | 3.43 (2.85 - 4.11) |
| Meta-analysis | 304,347 | 560,749 | 237.0 | 427.3 | 835 | 2,147 | 3.52 (3.29 - 3.77) | 5.02 (4.81 - 5.24) |
| Meta-analysis (w/o USOC) | 67,619 | 148,271 | 44.1 | 121.0 | 205 | 640 | 4.65 (4.03 - 5.33) | 5.29 (4.89 - 5.72) |
| **PS-stratification / on-treatment TAR without escalation** | | | | | | | | |
| CCAE | 23,622 | 30,488 | 17.2 | 20.8 | 64 | 108 | 3.73 (2.87 - 4.76) | 5.19 (4.26 - 6.27) |
| DAG | 5,471 | 10,567 | 4.1 | 9.9 | 26 | 67 | 6.32 (4.13 - 9.26) | 6.78 (5.25 - 8.61) |
| ODOD | 16,980 | 20,888 | 10.4 | 13.5 | 66 | 126 | 6.32 (4.89 - 8.04) | 9.37 (7.80 - 11.2) |
| MDCR | 2,048 | 5,074 | 1.4 | 4.1 | 15 | 45 | 10.4 (5.81 - 17.1) | 11.0 (8.05 - 14.8) |
| MDCD | 1,452 | 3,305 | 0.7 | 1.8 | 9 | 28 | 12.3 (5.62 - 23.3) | 15.3 (10.2 - 22.1) |
| Open Claims | 321,867 | 412,766 | 238.5 | 306.5 | 805 | 1,514 | 3.38 (3.15 - 3.62) | 4.94 (4.69 - 5.20) |
| OEHR | 19,746 | 31,577 | 5.5 | 13.1 | 31 | 78 | 5.62 (3.82 - 7.97) | 5.95 (4.70 - 7.42) |
| VA | 11,951 | 24,130 | 6.2 | 18.2 | 36 | 72 | 5.79 (4.06 - 8.02) | 3.96 (3.10 - 4.98) |
| SIDIAP | 5,024 | 19,780 | 6.1 | 36.0 | 23 | 127 | 3.76 (2.38 - 5.64) | 3.52 (2.94 - 4.19) |
| Meta-analysis | 394,758 | 531,140 | 283.3 | 403.8 | 1,030 | 2,065 | 3.64 (3.42 - 3.87) | 5.11 (4.90 - 5.34) |
| Meta-analysis (w/o USOC) | 72,891 | 118,374 | 44.8 | 97.4 | 225 | 551 | 5.02 (4.39 - 5.72) | 5.66 (5.20 - 6.15) |

##### 4.2.26 Vomiting

**Supplementary Table 341:** Exposures, outcomes and risk estimates for Vomiting in SGLT2I (male) (T) vs DPP4I (male) (C) new-user cohorts under four different propensity score (PS) adjustment / time-at-risk (TAR) designs across data sources. We report total patient exposure time in 1,000 patient-year (KYr) units, uncalibrated and calibrated hazard ratios (HRs) and their 95% confidence intervals (CIs). Greyed out entries did not pass study diagnostics and are included only for completeness.

| | Patients | | Exposure (KYr) | | Outcomes | | Incidence (per KYr) | |
|---|---|---|---|---|---|---|---|---|
| | T | C | T | C | T | C | T | C |
| **PS-matching / on-treatment TAR** | | | | | | | | |
| CCAE | 18,420 | 29,193 | 16.3 | 24.5 | 269 | 451 | 16.5 (14.6 - 18.6) | 18.4 (16.8 - 20.2) |
| DAG | 4,388 | 10,512 | 3.8 | 10.9 | 28 | 94 | 7.28 (4.84 - 10.5) | 8.61 (6.96 - 10.5) |
| ODOD | 11,564 | 19,749 | 8.8 | 15.3 | 178 | 392 | 20.3 (17.4 - 23.5) | 25.6 (23.1 - 28.2) |
| MDCR | 1,614 | 4,910 | 1.4 | 4.6 | 26 | 101 | 18.1 (11.9 - 26.6) | 22.0 (17.9 - 26.7) |
| MDCD | 1,218 | 2,886 | 0.7 | 2.1 | 29 | 94 | 38.8 (26.0 - 55.8) | 44.9 (36.3 - 55.0) |
| Open Claims | 225,990 | 395,523 | 220.1 | 377.7 | 1,941 | 4,573 | 8.82 (8.43 - 9.22) | 12.1 (11.8 - 12.5) |
| OEHR | 16,094 | 30,889 | 5.2 | 15.8 | 85 | 254 | 16.3 (13.0 - 20.1) | 16.1 (14.2 - 18.2) |
| VA | 7,360 | 22,787 | 5.0 | 20.7 | 22 | 136 | 4.37 (2.74 - 6.62) | 6.57 (5.51 - 7.77) |
| SIDIAP | 4,195 | 19,235 | 6.2 | 43.6 | 18 | 238 | 2.91 (1.73 - 4.60) | 5.45 (4.78 - 6.19) |
| Meta-analysis | 289,625 | 532,798 | 266.8 | 513.1 | 2,567 | 6,239 | 9.62 (9.25 - 10.00) | 12.2 (11.9 - 12.5) |
| Meta-analysis (w/o USOC) | 63,635 | 137,275 | 46.8 | 135.4 | 626 | 1,666 | 13.4 (12.4 - 14.5) | 12.3 (11.7 - 12.9) |
| **PS-stratification / on-treatment TAR** | | | | | | | | |
| CCAE | 22,530 | 29,213 | 18.5 | 24.5 | 308 | 451 | 16.6 (14.8 - 18.6) | 18.4 (16.8 - 20.2) |
| DAG | 5,453 | 10,539 | 4.5 | 11.0 | 34 | 94 | 7.61 (5.27 - 10.6) | 8.58 (6.94 - 10.5) |
| ODOD | 16,142 | 19,845 | 11.2 | 15.4 | 213 | 397 | 19.1 (16.6 - 21.8) | 25.7 (23.3 - 28.4) |
| MDCR | 2,025 | 4,964 | 1.6 | 4.7 | 29 | 103 | 18.1 (12.1 - 26.0) | 22.1 (18.1 - 26.9) |
| MDCD | 1,271 | 2,896 | 0.8 | 2.1 | 29 | 94 | 38.3 (25.6 - 55.0) | 44.7 (36.1 - 54.7) |
| Open Claims | 307,189 | 395,776 | 268.3 | 377.9 | 2,351 | 4,583 | 8.76 (8.41 - 9.12) | 12.1 (11.8 - 12.5) |
| OEHR | 19,279 | 30,954 | 6.1 | 15.8 | 99 | 256 | 16.2 (13.1 - 19.7) | 16.2 (14.3 - 18.3) |
| VA | 11,772 | 23,743 | 7.1 | 22.1 | 40 | 149 | 5.62 (4.02 - 7.66) | 6.74 (5.70 - 7.91) |
| SIDIAP | 4,990 | 19,644 | 6.9 | 45.1 | 22 | 252 | 3.20 (2.00 - 4.84) | 5.59 (4.92 - 6.32) |
| Meta-analysis | 377,608 | 510,935 | 317.1 | 494.3 | 3,056 | 6,136 | 9.64 (9.30 - 9.99) | 12.4 (12.1 - 12.7) |
| Meta-analysis (w/o USOC) | 70,419 | 115,159 | 48.7 | 116.4 | 705 | 1,553 | 14.5 (13.4 - 15.6) | 13.3 (12.7 - 14.0) |
| **PS-matching / on-treatment TAR without escalation** | | | | | | | | |
| CCAE | 18,420 | 29,193 | 14.1 | 19.8 | 219 | 372 | 15.5 (13.5 - 17.7) | 18.7 (16.9 - 20.8) |
| DAG | 4,388 | 10,512 | 3.5 | 9.9 | 26 | 85 | 7.38 (4.82 - 10.8) | 8.62 (6.88 - 10.7) |
| ODOD | 11,564 | 19,749 | 7.8 | 12.6 | 149 | 323 | 19.2 (16.2 - 22.5) | 25.6 (22.9 - 28.5) |
| MDCR | 1,614 | 4,910 | 1.3 | 3.9 | 24 | 90 | 18.8 (12.0 - 28.0) | 23.0 (18.5 - 28.3) |
| MDCD | 1,218 | 2,886 | 0.6 | 1.6 | 27 | 78 | 42.8 (28.2 - 62.3) | 49.2 (38.9 - 61.4) |
| Open Claims | 225,990 | 395,523 | 185.1 | 294.0 | 1,518 | 3,461 | 8.20 (7.79 - 8.62) | 11.8 (11.4 - 12.2) |
| OEHR | 16,094 | 30,889 | 4.6 | 12.8 | 72 | 171 | 15.7 (12.3 - 19.8) | 13.3 (11.4 - 15.5) |
| VA | 7,360 | 22,787 | 4.3 | 16.9 | 15 | 111 | 3.49 (1.95 - 5.75) | 6.57 (5.41 - 7.92) |
| SIDIAP | 4,195 | 19,235 | 5.5 | 34.7 | 16 | 200 | 2.92 (1.67 - 4.74) | 5.76 (4.99 - 6.62) |
| Meta-analysis | 289,625 | 532,798 | 226.2 | 404.6 | 2,039 | 4,813 | 9.02 (8.63 - 9.42) | 11.9 (11.6 - 12.2) |
| Meta-analysis (w/o USOC) | 63,635 | 137,275 | 41.1 | 110.7 | 521 | 1,352 | 12.7 (11.6 - 13.8) | 12.2 (11.6 - 12.9) |
| **PS-stratification / on-treatment TAR without escalation** | | | | | | | | |
| CCAE | 22,530 | 29,213 | 16.2 | 19.9 | 255 | 372 | 15.7 (13.8 - 17.8) | 18.7 (16.9 - 20.7) |
| DAG | 5,453 | 10,539 | 4.1 | 9.9 | 31 | 85 | 7.55 (5.13 - 10.7) | 8.58 (6.85 - 10.6) |
| ODOD | 16,142 | 19,845 | 9.9 | 12.7 | 179 | 327 | 18.0 (15.5 - 20.9) | 25.8 (23.0 - 28.7) |
| MDCR | 2,025 | 4,964 | 1.4 | 4.0 | 27 | 92 | 18.8 (12.4 - 27.4) | 23.2 (18.7 - 28.5) |
| MDCD | 1,271 | 2,896 | 0.6 | 1.6 | 27 | 78 | 42.2 (27.8 - 61.3) | 49.0 (38.7 - 61.1) |
| Open Claims | 307,189 | 395,776 | 228.8 | 294.1 | 1,867 | 3,469 | 8.16 (7.79 - 8.54) | 11.8 (11.4 - 12.2) |
| OEHR | 19,279 | 30,954 | 5.4 | 12.9 | 84 | 172 | 15.5 (12.4 - 19.2) | 13.4 (11.5 - 15.5) |
| VA | 11,772 | 23,743 | 6.2 | 18.0 | 31 | 122 | 5.03 (3.42 - 7.14) | 6.78 (5.63 - 8.10) |
| SIDIAP | 4,990 | 19,644 | 6.1 | 35.8 | 20 | 212 | 3.26 (1.99 - 5.04) | 5.92 (5.15 - 6.77) |
| Meta-analysis | 377,608 | 510,935 | 272.0 | 389.2 | 2,463 | 4,729 | 9.05 (8.70 - 9.42) | 12.2 (11.8 - 12.5) |
| Meta-analysis (w/o USOC) | 70,419 | 115,159 | 43.3 | 95.1 | 596 | 1,260 | 13.8 (12.7 - 14.9) | 13.3 (12.5 - 14.0) |

#### 4.3 SGLT2I (male) vs SU (male)

##### 4.3.1 3-point MACE

**Supplementary Table 342:** Exposures, outcomes and risk estimates for 3-point MACE in SGLT2I (male) (T) vs SU (male) (C) new-user cohorts under four different propensity score (PS) adjustment / time-at-risk (TAR) designs across data sources. We report total patient exposure time in 1,000 patient-year (KYr) units, uncalibrated and calibrated hazard ratios (HRs) and their 95% confidence intervals (CIs). Greyed out entries did not pass study diagnostics and are included only for completeness.

| | Patients | | Exposure (KYr) | | Outcomes | | Incidence (per KYr) | |
|---|---|---|---|---|---|---|---|---|
| | T | C | T | C | T | C | T | C |
| **PS-matching / on-treatment TAR** | | | | | | | | |
| CCAE | 19,864 | 45,123 | 17.0 | 37.5 | 123 | 429 | 7.26 (6.03 - 8.66) | 11.4 (10.4 - 12.6) |
| DAG | 5,471 | 973 | 4.6 | 0.9 | 0 | 0 | | |
| ODOD | 14,875 | 45,890 | 10.6 | 41.3 | 155 | 1,052 | 14.7 (12.4 - 17.1) | 25.5 (23.9 - 27.0) |
| MDCR | 1,661 | 7,509 | 1.4 | 7.0 | 33 | 214 | 23.1 (15.9 - 32.5) | 30.5 (26.5 - 34.9) |
| MDCD | 1,331 | 6,621 | 0.8 | 4.6 | 17 | 132 | 21.2 (12.3 - 33.9) | 28.8 (24.1 - 34.2) |
| Open Claims | 285,069 | 830,545 | 254.9 | 893.3 | 1,943 | 12,369 | 7.62 (7.29 - 7.97) | 13.8 (13.6 - 14.1) |
| OEHR | 17,735 | 71,834 | 5.6 | 39.8 | 44 | 557 | 7.83 (5.69 - 10.5) | 14.0 (12.9 - 15.2) |
| VA | 0 | 0 | 0.0 | 0.0 | 0 | 0 | | |
| SIDIAP | 4,099 | 1,187 | 6.0 | 2.2 | 27 | 12 | 4.53 (2.99 - 6.59) | 5.55 (2.87 - 9.70) |
| Meta-analysis | 344,634 | 1,008,709 | 296.2 | 1025.6 | 2,342 | 14,765 | 7.91 (7.59 - 8.23) | 14.4 (14.2 - 14.6) |
| Meta-analysis (w/o USOC) | 59,565 | 178,164 | 41.3 | 132.4 | 399 | 2,396 | 9.65 (8.73 - 10.6) | 18.1 (17.4 - 18.8) |
| **PS-stratification / on-treatment TAR** | | | | | | | | |
| CCAE | 23,133 | 45,722 | 19.3 | 37.8 | 140 | 434 | 7.27 (6.11 - 8.57) | 11.5 (10.4 - 12.6) |
| DAG | 5,702 | 1,403 | 4.7 | 1.4 | 0 | 0 | | |
| ODOD | 16,422 | 46,390 | 11.4 | 41.8 | 170 | 1,084 | 14.9 (12.8 - 17.4) | 25.9 (24.4 - 27.5) |
| MDCR | 1,977 | 7,751 | 1.6 | 7.2 | 36 | 230 | 22.8 (15.9 - 31.5) | 31.8 (27.8 - 36.2) |
| MDCD | 1,365 | 6,824 | 0.8 | 4.8 | 17 | 140 | 20.9 (12.2 - 33.5) | 29.5 (24.8 - 34.8) |
| Open Claims | 308,612 | 835,362 | 270.1 | 898.3 | 2,093 | 12,566 | 7.75 (7.42 - 8.09) | 14.0 (13.7 - 14.2) |
| OEHR | 19,539 | 74,077 | 6.2 | 41.3 | 48 | 604 | 7.69 (5.67 - 10.2) | 14.6 (13.5 - 15.8) |
| VA | 11,375 | 105,972 | 6.9 | 96.9 | 50 | 842 | 7.26 (5.39 - 9.57) | 8.69 (8.11 - 9.30) |
| SIDIAP | 4,643 | 2,634 | 6.4 | 6.1 | 29 | 39 | 4.55 (3.05 - 6.53) | 6.43 (4.57 - 8.78) |
| Meta-analysis | 367,706 | 1,001,551 | 307.0 | 1019.2 | 2,451 | 14,688 | 7.98 (7.67 - 8.31) | 14.4 (14.2 - 14.6) |
| Meta-analysis (w/o USOC) | 59,094 | 166,189 | 36.9 | 120.9 | 358 | 2,122 | 9.71 (8.73 - 10.8) | 17.5 (16.8 - 18.3) |
| **PS-matching / on-treatment TAR without escalation** | | | | | | | | |
| CCAE | 19,864 | 45,123 | 14.8 | 32.6 | 100 | 373 | 6.77 (5.51 - 8.24) | 11.5 (10.3 - 12.7) |
| DAG | 5,471 | 973 | 4.2 | 0.8 | 0 | 0 | | |
| ODOD | 14,875 | 45,890 | 9.4 | 37.0 | 137 | 942 | 14.6 (12.3 - 17.3) | 25.4 (23.8 - 27.1) |
| MDCR | 1,661 | 7,509 | 1.3 | 6.4 | 30 | 193 | 23.6 (15.9 - 33.7) | 30.0 (26.0 - 34.6) |
| MDCD | 1,331 | 6,621 | 0.7 | 3.8 | 15 | 111 | 22.2 (12.4 - 36.6) | 29.1 (23.9 - 35.0) |
| Open Claims | 285,069 | 830,545 | 215.9 | 758.2 | 1,649 | 10,619 | 7.64 (7.27 - 8.01) | 14.0 (13.7 - 14.3) |
| OEHR | 17,735 | 71,834 | 4.9 | 34.7 | 34 | 405 | 6.89 (4.77 - 9.63) | 11.7 (10.6 - 12.9) |
| VA | 0 | 0 | 0.0 | 0.0 | 0 | 0 | | |
| SIDIAP | 4,099 | 1,187 | 5.3 | 1.9 | 25 | 11 | 4.72 (3.06 - 6.97) | 5.88 (2.93 - 10.5) |
| Meta-analysis | 344,634 | 1,008,709 | 252.3 | 874.6 | 1,990 | 12,654 | 7.89 (7.55 - 8.24) | 14.5 (14.2 - 14.7) |
| Meta-analysis (w/o USOC) | 59,565 | 178,164 | 36.3 | 116.4 | 341 | 2,035 | 9.39 (8.42 - 10.4) | 17.5 (16.7 - 18.3) |
| **PS-stratification / on-treatment TAR without escalation** | | | | | | | | |
| CCAE | 23,133 | 45,722 | 16.8 | 32.9 | 116 | 378 | 6.89 (5.70 - 8.27) | 11.5 (10.4 - 12.7) |
| DAG | 5,702 | 1,403 | 4.3 | 1.3 | 0 | 0 | | |
| ODOD | 16,422 | 46,390 | 10.1 | 37.4 | 150 | 971 | 14.9 (12.6 - 17.4) | 25.9 (24.3 - 27.6) |
| MDCR | 1,977 | 7,751 | 1.4 | 6.6 | 32 | 209 | 22.7 (15.5 - 32.0) | 31.5 (27.4 - 36.1) |
| MDCD | 1,365 | 6,824 | 0.7 | 4.0 | 15 | 116 | 21.9 (12.2 - 36.1) | 29.3 (24.2 - 35.1) |
| Open Claims | 308,612 | 835,362 | 229.5 | 762.6 | 1,786 | 10,794 | 7.78 (7.43 - 8.15) | 14.2 (13.9 - 14.4) |
| OEHR | 19,539 | 74,077 | 5.5 | 35.9 | 38 | 440 | 6.91 (4.89 - 9.49) | 12.2 (11.1 - 13.4) |
| VA | 11,375 | 105,972 | 6.0 | 85.9 | 42 | 727 | 7.06 (5.09 - 9.54) | 8.46 (7.86 - 9.10) |
| SIDIAP | 4,643 | 2,634 | 5.7 | 5.2 | 27 | 33 | 4.75 (3.13 - 6.91) | 6.35 (4.37 - 8.92) |
| Meta-analysis | 367,706 | 1,001,551 | 261.9 | 868.8 | 2,090 | 12,583 | 7.98 (7.64 - 8.33) | 14.5 (14.2 - 14.7) |
| Meta-analysis (w/o USOC) | 59,094 | 166,189 | 32.4 | 106.2 | 304 | 1,789 | 9.38 (8.35 - 10.5) | 16.8 (16.1 - 17.6) |

##### 4.3.2 4-point MACE

**Supplementary Table 343:** Exposures, outcomes and risk estimates for 4-point MACE in SGLT2I (male) (T) vs SU (male) (C) new-user cohorts under four different propensity score (PS) adjustment / time-at-risk (TAR) designs across data sources. We report total patient exposure time in 1,000 patient-year (KYr) units, uncalibrated and calibrated hazard ratios (HRs) and their 95% confidence intervals (CIs). Greyed out entries did not pass study diagnostics and are included only for completeness.

| | Patients | | Exposure (KYr) | | Outcomes | | Incidence (per KYr) | |
|---|---|---|---|---|---|---|---|---|
| | T | C | T | C | T | C | T | C |
| **PS-matching / on-treatment TAR** | | | | | | | | |
| CCAE | 19,596 | 44,420 | 16.7 | 36.8 | 181 | 597 | 10.8 (9.31 - 12.5) | 16.2 (14.9 - 17.6) |
| DAG | 5,471 | 973 | 4.6 | 0.9 | 0 | 0 | | |
| ODOD | 14,341 | 43,929 | 10.2 | 39.1 | 236 | 1,675 | 23.1 (20.2 - 26.2) | 42.9 (40.9 - 45.0) |
| MDCR | 1,571 | 6,993 | 1.4 | 6.5 | 46 | 326 | 34.0 (24.9 - 45.4) | 50.2 (44.9 - 55.9) |
| MDCD | 1,234 | 6,134 | 0.7 | 4.2 | 20 | 194 | 27.1 (16.5 - 41.8) | 46.0 (39.8 - 53.0) |
| Open Claims | 276,531 | 802,166 | 248.4 | 858.7 | 2,753 | 19,306 | 11.1 (10.7 - 11.5) | 22.5 (22.2 - 22.8) |
| OEHR | 17,413 | 70,491 | 5.5 | 38.6 | 84 | 988 | 15.2 (12.1 - 18.8) | 25.6 (24.1 - 27.3) |
| VA | 0 | 0 | 0.0 | 0.0 | 0 | 0 | | |
| SIDIAP | 4,065 | 1,186 | 5.9 | 2.2 | 32 | 12 | 5.42 (3.71 - 7.65) | 5.55 (2.87 - 9.70) |
| Meta-analysis | 334,751 | 975,319 | 288.8 | 986.0 | 3,352 | 23,098 | 11.6 (11.2 - 12.0) | 23.4 (23.1 - 23.7) |
| Meta-analysis (w/o USOC) | 58,220 | 173,153 | 40.4 | 127.3 | 599 | 3,792 | 14.8 (13.6 - 16.0) | 29.8 (28.8 - 30.8) |
| **PS-stratification / on-treatment TAR** | | | | | | | | |
| CCAE | 22,780 | 44,992 | 19.0 | 37.1 | 211 | 603 | 11.1 (9.67 - 12.7) | 16.2 (15.0 - 17.6) |
| DAG | 5,702 | 1,403 | 4.7 | 1.4 | 0 | 0 | | |
| ODOD | 15,713 | 44,282 | 10.9 | 39.4 | 257 | 1,716 | 23.5 (20.7 - 26.5) | 43.6 (41.5 - 45.7) |
| MDCR | 1,862 | 7,168 | 1.5 | 6.6 | 52 | 344 | 34.7 (25.9 - 45.6) | 51.8 (46.5 - 57.6) |
| MDCD | 1,263 | 6,287 | 0.7 | 4.4 | 20 | 204 | 26.8 (16.3 - 41.3) | 46.9 (40.7 - 53.8) |
| Open Claims | 297,226 | 806,034 | 262.0 | 862.9 | 2,981 | 19,609 | 11.4 (11.0 - 11.8) | 22.7 (22.4 - 23.0) |
| OEHR | 19,146 | 72,526 | 6.1 | 39.9 | 93 | 1,074 | 15.2 (12.3 - 18.6) | 26.9 (25.3 - 28.6) |
| VA | 10,518 | 102,634 | 6.4 | 93.0 | 86 | 1,665 | 13.4 (10.7 - 16.6) | 17.9 (17.1 - 18.8) |
| SIDIAP | 4,579 | 2,630 | 6.3 | 6.1 | 33 | 44 | 5.24 (3.61 - 7.36) | 7.26 (5.27 - 9.74) |
| Meta-analysis | 354,865 | 967,834 | 298.0 | 979.3 | 3,542 | 23,002 | 11.9 (11.5 - 12.3) | 23.5 (23.2 - 23.8) |
| Meta-analysis (w/o USOC) | 57,639 | 161,800 | 36.0 | 116.4 | 561 | 3,393 | 15.6 (14.3 - 16.9) | 29.1 (28.2 - 30.1) |
| **PS-matching / on-treatment TAR without escalation** | | | | | | | | |
| CCAE | 19,596 | 44,420 | 14.6 | 32.0 | 149 | 517 | 10.2 (8.66 - 12.0) | 16.1 (14.8 - 17.6) |
| DAG | 5,471 | 973 | 4.2 | 0.8 | 0 | 0 | | |
| ODOD | 14,341 | 43,929 | 9.1 | 35.0 | 207 | 1,494 | 22.8 (19.8 - 26.1) | 42.7 (40.5 - 44.9) |
| MDCR | 1,571 | 6,993 | 1.2 | 6.0 | 41 | 303 | 34.2 (24.5 - 46.4) | 50.7 (45.2 - 56.8) |
| MDCD | 1,234 | 6,134 | 0.6 | 3.5 | 17 | 161 | 27.3 (15.9 - 43.7) | 45.7 (38.9 - 53.3) |
| Open Claims | 276,531 | 802,166 | 210.6 | 729.7 | 2,307 | 16,514 | 11.0 (10.5 - 11.4) | 22.6 (22.3 - 23.0) |
| OEHR | 17,413 | 70,491 | 4.8 | 33.8 | 67 | 767 | 13.8 (10.7 - 17.6) | 22.7 (21.1 - 24.4) |
| VA | 0 | 0 | 0.0 | 0.0 | 0 | 0 | | |
| SIDIAP | 4,065 | 1,186 | 5.2 | 1.9 | 28 | 11 | 5.33 (3.54 - 7.71) | 5.88 (2.93 - 10.5) |
| Meta-analysis | 334,751 | 975,319 | 246.1 | 841.8 | 2,816 | 19,767 | 11.4 (11.0 - 11.9) | 23.5 (23.2 - 23.8) |
| Meta-analysis (w/o USOC) | 58,220 | 173,153 | 35.6 | 112.2 | 509 | 3,253 | 14.3 (13.1 - 15.6) | 29.0 (28.0 - 30.0) |
| **PS-stratification / on-treatment TAR without escalation** | | | | | | | | |
| CCAE | 22,780 | 44,992 | 16.6 | 32.3 | 176 | 523 | 10.6 (9.11 - 12.3) | 16.2 (14.8 - 17.6) |
| DAG | 5,702 | 1,403 | 4.3 | 1.3 | 0 | 0 | | |
| ODOD | 15,713 | 44,282 | 9.7 | 35.3 | 226 | 1,530 | 23.2 (20.3 - 26.5) | 43.3 (41.2 - 45.6) |
| MDCR | 1,862 | 7,168 | 1.3 | 6.1 | 46 | 321 | 34.5 (25.3 - 46.1) | 52.5 (46.9 - 58.6) |
| MDCD | 1,263 | 6,287 | 0.6 | 3.6 | 17 | 168 | 27.0 (15.7 - 43.2) | 46.2 (39.5 - 53.8) |
| Open Claims | 297,226 | 806,034 | 222.7 | 733.3 | 2,508 | 16,789 | 11.3 (10.8 - 11.7) | 22.9 (22.5 - 23.2) |
| OEHR | 19,146 | 72,526 | 5.4 | 34.9 | 75 | 836 | 13.9 (10.9 - 17.4) | 24.0 (22.4 - 25.6) |
| VA | 10,518 | 102,634 | 5.5 | 82.6 | 71 | 1,428 | 12.8 (10.0 - 16.2) | 17.3 (16.4 - 18.2) |
| SIDIAP | 4,579 | 2,630 | 5.6 | 5.2 | 29 | 38 | 5.16 (3.46 - 7.41) | 7.32 (5.18 - 10.0) |
| Meta-analysis | 354,865 | 967,834 | 254.4 | 835.8 | 2,985 | 19,678 | 11.7 (11.3 - 12.2) | 23.5 (23.2 - 23.9) |
| Meta-analysis (w/o USOC) | 57,639 | 161,800 | 31.7 | 102.5 | 477 | 2,889 | 15.1 (13.7 - 16.5) | 28.2 (27.2 - 29.2) |

##### 4.3.3 Acute myocardial infarction

**Supplementary Table 344:** Exposures, outcomes and risk estimates for Acute myocardial infarction in SGLT2I (male) (T) vs SU (male) (C) new-user cohorts under four different propensity score (PS) adjustment / time-at-risk (TAR) designs across data sources. We report total patient exposure time in 1,000 patient-year (KYr) units, uncalibrated and calibrated hazard ratios (HRs) and their 95% confidence intervals (CIs). Greyed out entries did not pass study diagnostics and are included only for completeness.

| | Patients | | Exposure (KYr) | | Outcomes | | Incidence (per KYr) | |
|---|---|---|---|---|---|---|---|---|
| | T | C | T | C | T | C | T | C |
| **PS-matching / on-treatment TAR** | | | | | | | | |
| CCAE | 20,117 | 45,808 | 17.1 | 38.1 | 72 | 255 | 4.20 (3.29 - 5.29) | 6.69 (5.90 - 7.57) |
| DAG | 5,471 | 973 | 4.6 | 0.9 | 0 | 0 | | |
| ODOD | 15,190 | 47,220 | 10.8 | 42.8 | 90 | 580 | 8.30 (6.67 - 10.2) | 13.6 (12.5 - 14.7) |
| MDCR | 1,705 | 7,915 | 1.5 | 7.4 | 12 | 112 | 8.18 (4.23 - 14.3) | 15.0 (12.4 - 18.1) |
| MDCD | 1,383 | 6,895 | 0.8 | 4.8 | 12 | 67 | 14.3 (7.41 - 25.1) | 13.9 (10.8 - 17.7) |
| Open Claims | 291,920 | 854,971 | 260.5 | 921.5 | 1,091 | 6,456 | 4.19 (3.94 - 4.44) | 7.01 (6.84 - 7.18) |
| OEHR | 17,905 | 72,582 | 5.7 | 40.4 | 21 | 318 | 3.70 (2.29 - 5.66) | 7.87 (7.02 - 8.78) |
| VA | 0 | 0 | 0.0 | 0.0 | 0 | 0 | | |
| SIDIAP | 4,168 | 1,203 | 6.1 | 2.2 | 15 | 8 | 2.47 (1.38 - 4.08) | 3.64 (1.57 - 7.18) |
| Meta-analysis | 348,220 | 1,035,391 | 296.4 | 1055.1 | 1,298 | 7,788 | 4.38 (4.14 - 4.62) | 7.38 (7.22 - 7.55) |
| Meta-analysis (w/o USOC) | 56,300 | 180,420 | 36.0 | 133.5 | 207 | 1,332 | 5.76 (5.00 - 6.60) | 9.97 (9.45 - 10.5) |
| **PS-stratification / on-treatment TAR** | | | | | | | | |
| CCAE | 23,425 | 46,429 | 19.5 | 38.5 | 82 | 256 | 4.20 (3.34 - 5.22) | 6.66 (5.87 - 7.52) |
| DAG | 5,702 | 1,403 | 4.7 | 1.4 | 0 | 0 | | |
| ODOD | 16,779 | 47,825 | 11.7 | 43.4 | 101 | 601 | 8.66 (7.05 - 10.5) | 13.9 (12.8 - 15.0) |
| MDCR | 2,032 | 8,121 | 1.6 | 7.6 | 15 | 121 | 9.23 (5.16 - 15.2) | 15.9 (13.2 - 19.0) |
| MDCD | 1,420 | 7,116 | 0.8 | 5.0 | 12 | 73 | 14.1 (7.31 - 24.7) | 14.6 (11.5 - 18.4) |
| Open Claims | 316,207 | 860,148 | 276.1 | 926.9 | 1,187 | 6,563 | 4.30 (4.06 - 4.55) | 7.08 (6.91 - 7.25) |
| OEHR | 19,723 | 74,941 | 6.3 | 42.0 | 26 | 347 | 4.13 (2.70 - 6.05) | 8.26 (7.41 - 9.18) |
| VA | 11,651 | 107,705 | 7.0 | 98.7 | 35 | 520 | 4.98 (3.47 - 6.92) | 5.27 (4.83 - 5.74) |
| SIDIAP | 4,738 | 2,664 | 6.5 | 6.2 | 16 | 23 | 2.46 (1.41 - 4.00) | 3.73 (2.36 - 5.59) |
| Meta-analysis | 376,134 | 1,029,343 | 313.6 | 1050.8 | 1,396 | 7,767 | 4.45 (4.22 - 4.69) | 7.39 (7.23 - 7.56) |
| Meta-analysis (w/o USOC) | 59,927 | 169,195 | 37.5 | 123.8 | 209 | 1,204 | 5.58 (4.85 - 6.39) | 9.72 (9.18 - 10.3) |
| **PS-matching / on-treatment TAR without escalation** | | | | | | | | |
| CCAE | 20,117 | 45,808 | 14.9 | 33.1 | 60 | 216 | 4.02 (3.07 - 5.17) | 6.53 (5.69 - 7.46) |
| DAG | 5,471 | 973 | 4.2 | 0.8 | 0 | 0 | | |
| ODOD | 15,190 | 47,220 | 9.6 | 38.3 | 80 | 524 | 8.34 (6.61 - 10.4) | 13.7 (12.5 - 14.9) |
| MDCR | 1,705 | 7,915 | 1.3 | 6.8 | 12 | 98 | 9.19 (4.75 - 16.0) | 14.4 (11.7 - 17.5) |
| MDCD | 1,383 | 6,895 | 0.7 | 4.0 | 12 | 57 | 17.0 (8.78 - 29.7) | 14.3 (10.8 - 18.5) |
| Open Claims | 291,920 | 854,971 | 220.6 | 782.1 | 913 | 5,507 | 4.14 (3.87 - 4.42) | 7.04 (6.86 - 7.23) |
| OEHR | 17,905 | 72,582 | 5.0 | 35.2 | 20 | 219 | 4.02 (2.46 - 6.21) | 6.23 (5.43 - 7.11) |
| VA | 0 | 0 | 0.0 | 0.0 | 0 | 0 | | |
| SIDIAP | 4,168 | 1,203 | 5.4 | 1.9 | 14 | 7 | 2.60 (1.42 - 4.36) | 3.67 (1.47 - 7.55) |
| Meta-analysis | 348,220 | 1,035,391 | 252.1 | 899.5 | 1,097 | 6,621 | 4.35 (4.10 - 4.62) | 7.36 (7.18 - 7.54) |
| Meta-analysis (w/o USOC) | 56,300 | 180,420 | 31.5 | 117.4 | 184 | 1,114 | 5.84 (5.03 - 6.75) | 9.49 (8.94 - 10.1) |
| **PS-stratification / on-treatment TAR without escalation** | | | | | | | | |
| CCAE | 23,425 | 46,429 | 17.0 | 33.4 | 70 | 217 | 4.11 (3.20 - 5.19) | 6.49 (5.66 - 7.42) |
| DAG | 5,702 | 1,403 | 4.3 | 1.3 | 0 | 0 | | |
| ODOD | 16,779 | 47,825 | 10.3 | 38.8 | 90 | 541 | 8.71 (7.00 - 10.7) | 13.9 (12.8 - 15.2) |
| MDCR | 2,032 | 8,121 | 1.5 | 7.0 | 14 | 107 | 9.64 (5.27 - 16.2) | 15.4 (12.6 - 18.6) |
| MDCD | 1,420 | 7,116 | 0.7 | 4.1 | 12 | 61 | 16.7 (8.65 - 29.2) | 14.7 (11.2 - 18.9) |
| Open Claims | 316,207 | 860,148 | 234.6 | 786.8 | 1,000 | 5,603 | 4.26 (4.00 - 4.54) | 7.12 (6.94 - 7.31) |
| OEHR | 19,723 | 74,941 | 5.5 | 36.5 | 25 | 240 | 4.51 (2.92 - 6.66) | 6.58 (5.77 - 7.47) |
| VA | 11,651 | 107,705 | 6.1 | 87.5 | 31 | 443 | 5.10 (3.46 - 7.24) | 5.07 (4.60 - 5.56) |
| SIDIAP | 4,738 | 2,664 | 5.8 | 5.3 | 15 | 18 | 2.59 (1.45 - 4.27) | 3.41 (2.02 - 5.38) |
| Meta-analysis | 376,134 | 1,029,343 | 267.5 | 895.5 | 1,185 | 6,601 | 4.43 (4.18 - 4.69) | 7.37 (7.19 - 7.55) |
| Meta-analysis (w/o USOC) | 59,927 | 169,195 | 32.9 | 108.7 | 185 | 998 | 5.62 (4.84 - 6.49) | 9.18 (8.62 - 9.77) |

##### 4.3.4 Acute renal failure

**Supplementary Table 345:** Exposures, outcomes and risk estimates for Acute renal failure in SGLT2I (male) (T) vs SU (male) (C) new-user cohorts under four different propensity score (PS) adjustment / time-at-risk (TAR) designs across data sources. We report total patient exposure time in 1,000 patient-year (KYr) units, uncalibrated and calibrated hazard ratios (HRs) and their 95% confidence intervals (CIs). Greyed out entries did not pass study diagnostics and are included only for completeness.

| | Patients | | Exposure (KYr) | | Outcomes | | Incidence (per KYr) | |
|---|---|---|---|---|---|---|---|---|
| | T | C | T | C | T | C | T | C |
| **PS-matching / on-treatment TAR** | | | | | | | | |
| CCAE | 20,248 | 45,772 | 17.3 | 38.2 | 112 | 452 | 6.49 (5.34 - 7.81) | 11.8 (10.8 - 13.0) |
| DAG | 5,471 | 973 | 4.6 | 0.9 | 0 | 0 | | |
| ODOD | 15,107 | 45,548 | 10.8 | 40.7 | 180 | 1,562 | 16.6 (14.3 - 19.2) | 38.4 (36.5 - 40.3) |
| MDCR | 1,684 | 7,647 | 1.4 | 7.1 | 30 | 293 | 20.8 (14.0 - 29.6) | 41.2 (36.6 - 46.1) |
| MDCD | 1,366 | 6,610 | 0.8 | 4.6 | 16 | 193 | 19.0 (10.9 - 30.8) | 42.1 (36.4 - 48.5) |
| Open Claims | 294,583 | 849,930 | 263.5 | 912.8 | 2,103 | 16,419 | 7.98 (7.64 - 8.33) | 18.0 (17.7 - 18.3) |
| OEHR | 17,982 | 72,411 | 5.7 | 40.0 | 60 | 853 | 10.5 (8.02 - 13.5) | 21.3 (19.9 - 22.8) |
| VA | 0 | 0 | 0.0 | 0.0 | 0 | 0 | | |
| SIDIAP | 4,546 | 1,216 | 6.6 | 2.2 | <5 | <5 | < 0.76 (0.25 - 1.77) | < 2.23 (0.72 - 5.21) |
| Meta-analysis | 350,970 | 1,027,918 | 299.6 | 1043.4 | 2,501 | 19,772 | 8.35 (8.02 - 8.68) | 18.9 (18.7 - 19.2) |
| Meta-analysis (w/o USOC) | 56,387 | 177,988 | 36.1 | 130.6 | 398 | 3,353 | 11.0 (9.97 - 12.2) | 25.7 (24.8 - 26.6) |
| **PS-stratification / on-treatment TAR** | | | | | | | | |
| CCAE | 23,699 | 46,349 | 19.7 | 38.5 | 127 | 459 | 6.44 (5.37 - 7.66) | 11.9 (10.9 - 13.1) |
| DAG | 5,702 | 1,403 | 4.7 | 1.4 | 0 | 0 | | |
| ODOD | 16,747 | 46,175 | 11.7 | 41.3 | 199 | 1,629 | 17.0 (14.7 - 19.6) | 39.4 (37.5 - 41.4) |
| MDCR | 2,014 | 7,786 | 1.6 | 7.2 | 36 | 301 | 22.3 (15.6 - 30.8) | 41.7 (37.1 - 46.6) |
| MDCD | 1,409 | 6,762 | 0.9 | 4.7 | 17 | 204 | 19.9 (11.6 - 31.8) | 43.2 (37.5 - 49.5) |
| Open Claims | 320,733 | 854,446 | 280.5 | 917.6 | 2,329 | 16,716 | 8.30 (7.97 - 8.65) | 18.2 (17.9 - 18.5) |
| OEHR | 19,913 | 74,597 | 6.4 | 41.4 | 66 | 934 | 10.4 (8.03 - 13.2) | 22.5 (21.1 - 24.0) |
| VA | 11,731 | 106,207 | 7.1 | 96.6 | 93 | 1,559 | 13.1 (10.6 - 16.0) | 16.1 (15.3 - 17.0) |
| SIDIAP | 5,159 | 2,688 | 7.1 | 6.3 | <5 | <5 | < 0.71 (0.23 - 1.65) | < 0.80 (0.26 - 1.86) |
| Meta-analysis | 381,092 | 1,021,567 | 318.2 | 1038.9 | 2,721 | 19,738 | 8.55 (8.23 - 8.88) | 19.0 (18.7 - 19.3) |
| Meta-analysis (w/o USOC) | 60,359 | 167,121 | 37.8 | 121.3 | 392 | 3,022 | 10.4 (9.38 - 11.5) | 24.9 (24.0 - 25.8) |
| **PS-matching / on-treatment TAR without escalation** | | | | | | | | |
| CCAE | 20,248 | 45,772 | 15.1 | 33.1 | 87 | 380 | 5.77 (4.62 - 7.12) | 11.5 (10.3 - 12.7) |
| DAG | 5,471 | 973 | 4.2 | 0.8 | 0 | 0 | | |
| ODOD | 15,107 | 45,548 | 9.6 | 36.5 | 151 | 1,403 | 15.7 (13.3 - 18.4) | 38.4 (36.4 - 40.5) |
| MDCR | 1,684 | 7,647 | 1.3 | 6.5 | 27 | 266 | 20.9 (13.8 - 30.4) | 40.7 (35.9 - 45.9) |
| MDCD | 1,366 | 6,610 | 0.7 | 3.8 | 15 | 157 | 21.1 (11.8 - 34.8) | 41.0 (34.8 - 47.9) |
| Open Claims | 294,583 | 849,930 | 223.5 | 775.7 | 1,709 | 13,780 | 7.65 (7.29 - 8.02) | 17.8 (17.5 - 18.1) |
| OEHR | 17,982 | 72,411 | 5.0 | 35.0 | 40 | 630 | 7.98 (5.70 - 10.9) | 18.0 (16.6 - 19.5) |
| VA | 0 | 0 | 0.0 | 0.0 | 0 | 0 | | |
| SIDIAP | 4,546 | 1,216 | 5.9 | 2.0 | <5 | 0 | < 0.85 (0.28 - 1.99) | |
| Meta-analysis | 350,970 | 1,027,918 | 255.2 | 890.7 | 2,029 | 16,616 | 7.95 (7.61 - 8.30) | 18.7 (18.4 - 18.9) |
| Meta-analysis (w/o USOC) | 56,387 | 177,988 | 31.7 | 115.0 | 320 | 2,836 | 10.1 (9.02 - 11.3) | 24.7 (23.8 - 25.6) |
| **PS-stratification / on-treatment TAR without escalation** | | | | | | | | |
| CCAE | 23,699 | 46,349 | 17.3 | 33.4 | 102 | 386 | 5.91 (4.82 - 7.18) | 11.5 (10.4 - 12.8) |
| DAG | 5,702 | 1,403 | 4.3 | 1.3 | 0 | 0 | | |
| ODOD | 16,747 | 46,175 | 10.4 | 37.1 | 167 | 1,464 | 16.1 (13.7 - 18.7) | 39.5 (37.5 - 41.6) |
| MDCR | 2,014 | 7,786 | 1.5 | 6.6 | 32 | 272 | 22.0 (15.1 - 31.1) | 41.0 (36.2 - 46.1) |
| MDCD | 1,409 | 6,762 | 0.7 | 3.9 | 16 | 165 | 22.1 (12.7 - 36.0) | 41.8 (35.7 - 48.7) |
| Open Claims | 320,733 | 854,446 | 238.7 | 779.9 | 1,920 | 14,041 | 8.05 (7.69 - 8.41) | 18.0 (17.7 - 18.3) |
| OEHR | 19,913 | 74,597 | 5.6 | 36.2 | 46 | 694 | 8.20 (6.01 - 10.9) | 19.2 (17.8 - 20.7) |
| VA | 11,731 | 106,207 | 6.2 | 85.7 | 80 | 1,275 | 13.0 (10.3 - 16.2) | 14.9 (14.1 - 15.7) |
| SIDIAP | 5,159 | 2,688 | 6.3 | 5.4 | <5 | 0 | < 0.79 (0.26 - 1.85) | |
| Meta-analysis | 381,092 | 1,021,567 | 271.9 | 886.6 | 2,235 | 16,585 | 8.22 (7.88 - 8.57) | 18.7 (18.4 - 19.0) |
| Meta-analysis (w/o USOC) | 60,359 | 167,121 | 33.2 | 106.7 | 315 | 2,544 | 9.48 (8.46 - 10.6) | 23.9 (22.9 - 24.8) |

##### 4.3.5 Glycemic control

**Supplementary Table 346:** Exposures, outcomes and risk estimates for Glycemic control in SGLT2I (male) (T) vs SU (male) (C) new-user cohorts under four different propensity score (PS) adjustment / time-at-risk (TAR) designs across data sources. We report total patient exposure time in 1,000 patient-year (KYr) units, uncalibrated and calibrated hazard ratios (HRs) and their 95% confidence intervals (CIs). Greyed out entries did not pass study diagnostics and are included only for completeness.

| | Patients | | Exposure (KYr) | | Outcomes | | Incidence (per KYr) | |
|---|---|---|---|---|---|---|---|---|
| | T | C | T | C | T | C | T | C |
| **PS-matching / on-treatment TAR** | | | | | | | | |
| CCAE | 20,489 | 46,544 | 17.5 | 38.9 | 22 | 60 | 1.26 (0.79 - 1.91) | 1.54 (1.18 - 1.98) |
| DAG | 1,909 | 406 | 1.4 | 0.3 | 199 | 54 | 145 (125 - 166) | 163 (122 - 212) |
| ODOD | 10,546 | 33,733 | 6.9 | 26.6 | 716 | 2,844 | 104 (96.7 - 112) | 107 (103 - 111) |
| MDCR | 1,772 | 8,248 | 1.5 | 7.8 | 0 | 11 | | 1.41 (0.70 - 2.52) |
| MDCD | 1,464 | 7,245 | 0.9 | 5.1 | 0 | 0 | | |
| Open Claims | 304,948 | 889,205 | 271.7 | 962.4 | 0 | 0 | | |
| OEHR | 18,285 | 73,829 | 5.8 | 41.4 | 0 | 0 | | |
| VA | 0 | 0 | 0.0 | 0.0 | 0 | 0 | | |
| SIDIAP | 487 | 169 | 0.5 | 0.2 | 161 | 46 | 331 (281 - 386) | 231 (169 - 308) |
| Meta-analysis | 33,431 | 80,852 | 26.2 | 66.1 | 1,098 | 3,004 | 41.9 (39.5 - 44.5) | 45.5 (43.8 - 47.1) |
| Meta-analysis (w/o USOC) | 33,431 | 80,852 | 26.2 | 66.1 | 1,098 | 3,004 | 41.9 (39.5 - 44.5) | 45.5 (43.8 - 47.1) |
| **PS-stratification / on-treatment TAR** | | | | | | | | |
| CCAE | 23,976 | 47,040 | 20.0 | 39.3 | 26 | 73 | 1.30 (0.85 - 1.91) | 1.86 (1.46 - 2.34) |
| DAG | 1,981 | 611 | 1.4 | 0.5 | 210 | 78 | 148 (129 - 169) | 154 (122 - 193) |
| ODOD | 11,471 | 34,473 | 7.3 | 27.2 | 761 | 2,931 | 105 (97.3 - 112) | 108 (104 - 112) |
| MDCR | 2,128 | 8,484 | 1.7 | 8.0 | 0 | 12 | | 1.50 (0.77 - 2.62) |
| MDCD | 1,507 | 7,493 | 0.9 | 5.3 | 0 | 0 | | |
| Open Claims | 332,801 | 894,802 | 289.7 | 968.3 | 0 | 0 | | |
| OEHR | 20,231 | 76,288 | 6.5 | 43.1 | 0 | 0 | | |
| VA | 11,946 | 105,730 | 7.2 | 96.4 | 12 | 146 | 1.67 (0.86 - 2.92) | 1.51 (1.28 - 1.78) |
| SIDIAP | 541 | 437 | 0.5 | 0.6 | 180 | 150 | 348 (299 - 403) | 232 (196 - 272) |
| Meta-analysis | 35,447 | 81,513 | 27.2 | 66.5 | 787 | 3,004 | 28.9 (26.9 - 31.0) | 45.2 (43.6 - 46.8) |
| Meta-analysis (w/o USOC) | 35,447 | 81,513 | 27.2 | 66.5 | 787 | 3,004 | 28.9 (26.9 - 31.0) | 45.2 (43.6 - 46.8) |
| **PS-matching / on-treatment TAR without escalation** | | | | | | | | |
| CCAE | 20,489 | 46,544 | 15.2 | 33.7 | 21 | 56 | 1.38 (0.86 - 2.11) | 1.66 (1.26 - 2.16) |
| DAG | 1,909 | 406 | 1.3 | 0.3 | 174 | 48 | 136 (117 - 158) | 156 (115 - 206) |
| ODOD | 10,546 | 33,733 | 6.1 | 23.9 | 628 | 2,598 | 103 (94.9 - 111) | 109 (105 - 113) |
| MDCR | 1,772 | 8,248 | 1.4 | 7.2 | 0 | 10 | | 1.40 (0.67 - 2.57) |
| MDCD | 1,464 | 7,245 | 0.8 | 4.2 | 0 | 0 | | |
| Open Claims | 304,948 | 889,205 | 230.2 | 816.1 | 0 | 0 | | |
| OEHR | 18,285 | 73,829 | 5.1 | 35.9 | 0 | 0 | | |
| VA | 0 | 0 | 0.0 | 0.0 | 0 | 0 | | |
| SIDIAP | 487 | 169 | 0.4 | 0.2 | 150 | 42 | 357 (302 - 419) | 251 (181 - 339) |
| Meta-analysis | 33,431 | 80,852 | 23.0 | 58.0 | 973 | 2,744 | 42.3 (39.7 - 45.0) | 47.3 (45.5 - 49.1) |
| Meta-analysis (w/o USOC) | 33,431 | 80,852 | 23.0 | 58.0 | 973 | 2,744 | 42.3 (39.7 - 45.0) | 47.3 (45.5 - 49.1) |
| **PS-stratification / on-treatment TAR without escalation** | | | | | | | | |
| CCAE | 23,976 | 47,040 | 17.4 | 34.0 | 25 | 68 | 1.44 (0.93 - 2.12) | 2.00 (1.55 - 2.54) |
| DAG | 1,981 | 611 | 1.3 | 0.5 | 185 | 71 | 141 (121 - 162) | 154 (120 - 194) |
| ODOD | 11,471 | 34,473 | 6.5 | 24.4 | 669 | 2,677 | 103 (95.4 - 111) | 110 (106 - 114) |
| MDCR | 2,128 | 8,484 | 1.5 | 7.3 | 0 | 11 | | 1.50 (0.75 - 2.68) |
| MDCD | 1,507 | 7,493 | 0.8 | 4.4 | 0 | 0 | | |
| Open Claims | 332,801 | 894,802 | 246.2 | 821.2 | 0 | 0 | | |
| OEHR | 20,231 | 76,288 | 5.7 | 37.3 | 0 | 0 | | |
| VA | 11,946 | 105,730 | 6.2 | 85.3 | <10 | 133 | < 1.61 (0.77 - 2.96) | 1.56 (1.30 - 1.85) |
| SIDIAP | 541 | 437 | 0.4 | 0.5 | 165 | 133 | 371 (317 - 432) | 253 (212 - 300) |
| Meta-analysis | 35,447 | 81,513 | 23.9 | 58.4 | 694 | 2,745 | 29.0 (26.9 - 31.3) | 47.0 (45.3 - 48.8) |
| Meta-analysis (w/o USOC) | 35,447 | 81,513 | 23.9 | 58.4 | 694 | 2,745 | 29.0 (26.9 - 31.3) | 47.0 (45.3 - 48.8) |

##### 4.3.6 Hospitalization with heart failure

**Supplementary Table 347:** Exposures, outcomes and risk estimates for Hospitalization with heart failure in SGLT2I (male) (T) vs SU (male) (C) new-user cohorts under four different propensity score (PS) adjustment / time-at-risk (TAR) designs across data sources. We report total patient exposure time in 1,000 patient-year (KYr) units, uncalibrated and calibrated hazard ratios (HRs) and their 95% confidence intervals (CIs). Greyed out entries did not pass study diagnostics and are included only for completeness.

| | Patients | | Exposure (KYr) | | Outcomes | | Incidence (per KYr) | |
|---|---|---|---|---|---|---|---|---|
| | T | C | T | C | T | C | T | C |
| **PS-matching / on-treatment TAR** | | | | | | | | |
| CCAE | 20,240 | 45,906 | 17.3 | 38.3 | 93 | 286 | 5.38 (4.34 - 6.59) | 7.47 (6.63 - 8.39) |
| DAG | 5,471 | 973 | 4.6 | 0.9 | 0 | 0 | | |
| ODOD | 14,941 | 45,955 | 10.7 | 41.2 | 145 | 1,161 | 13.5 (11.4 - 15.9) | 28.2 (26.6 - 29.8) |
| MDCR | 1,653 | 7,560 | 1.4 | 7.1 | 31 | 227 | 21.7 (14.8 - 30.9) | 32.2 (28.1 - 36.6) |
| MDCD | 1,314 | 6,533 | 0.8 | 4.5 | 11 | 122 | 13.9 (6.96 - 24.9) | 26.9 (22.3 - 32.1) |
| Open Claims | 292,046 | 847,603 | 262.1 | 912.3 | 1,442 | 12,311 | 5.50 (5.22 - 5.79) | 13.5 (13.3 - 13.7) |
| OEHR | 17,851 | 72,117 | 5.7 | 39.8 | 55 | 662 | 9.71 (7.31 - 12.6) | 16.6 (15.4 - 17.9) |
| VA | 0 | 0 | 0.0 | 0.0 | 0 | 0 | | |
| SIDIAP | 4,513 | 1,218 | 6.6 | 2.2 | 5 | 0 | 0.76 (0.25 - 1.78) | |
| Meta-analysis | 348,045 | 1,025,674 | 298.0 | 1043.2 | 1,777 | 14,769 | 5.96 (5.69 - 6.25) | 14.2 (13.9 - 14.4) |
| Meta-analysis (w/o USOC) | 55,999 | 178,071 | 35.9 | 131.0 | 335 | 2,458 | 9.33 (8.36 - 10.4) | 18.8 (18.0 - 19.5) |
| **PS-stratification / on-treatment TAR** | | | | | | | | |
| CCAE | 23,598 | 46,509 | 19.7 | 38.6 | 113 | 292 | 5.75 (4.74 - 6.91) | 7.56 (6.72 - 8.48) |
| DAG | 5,702 | 1,403 | 4.7 | 1.4 | 0 | 0 | | |
| ODOD | 16,421 | 46,384 | 11.5 | 41.6 | 163 | 1,194 | 14.2 (12.1 - 16.5) | 28.7 (27.1 - 30.3) |
| MDCR | 1,962 | 7,706 | 1.6 | 7.2 | 33 | 237 | 20.9 (14.4 - 29.4) | 33.0 (29.0 - 37.5) |
| MDCD | 1,350 | 6,710 | 0.8 | 4.7 | 11 | 129 | 13.7 (6.86 - 24.6) | 27.5 (23.0 - 32.7) |
| Open Claims | 315,211 | 852,161 | 277.3 | 917.1 | 1,631 | 12,575 | 5.88 (5.60 - 6.17) | 13.7 (13.5 - 14.0) |
| OEHR | 19,700 | 74,317 | 6.3 | 41.3 | 64 | 730 | 10.2 (7.83 - 13.0) | 17.7 (16.4 - 19.0) |
| VA | 11,226 | 106,553 | 6.8 | 97.0 | 73 | 1,224 | 10.7 (8.37 - 13.4) | 12.6 (11.9 - 13.3) |
| SIDIAP | 5,095 | 2,687 | 7.0 | 6.3 | 5 | 5 | 0.72 (0.23 - 1.67) | 0.80 (0.26 - 1.86) |
| Meta-analysis | 374,930 | 1,019,371 | 314.8 | 1038.6 | 1,971 | 14,791 | 6.26 (5.99 - 6.54) | 14.2 (14.0 - 14.5) |
| Meta-analysis (w/o USOC) | 59,719 | 167,210 | 37.5 | 121.5 | 340 | 2,216 | 9.07 (8.14 - 10.1) | 18.2 (17.5 - 19.0) |
| **PS-matching / on-treatment TAR without escalation** | | | | | | | | |
| CCAE | 20,240 | 45,906 | 15.1 | 33.2 | 75 | 242 | 4.98 (3.91 - 6.24) | 7.28 (6.39 - 8.26) |
| DAG | 5,471 | 973 | 4.2 | 0.8 | 0 | 0 | | |
| ODOD | 14,941 | 45,955 | 9.5 | 36.9 | 127 | 1,029 | 13.4 (11.1 - 15.9) | 27.8 (26.2 - 29.6) |
| MDCR | 1,653 | 7,560 | 1.3 | 6.5 | 26 | 212 | 20.5 (13.4 - 30.0) | 32.6 (28.4 - 37.3) |
| MDCD | 1,314 | 6,533 | 0.7 | 3.8 | 10 | 101 | 15.0 (7.19 - 27.6) | 26.8 (21.8 - 32.5) |
| Open Claims | 292,046 | 847,603 | 222.3 | 774.6 | 1,197 | 10,491 | 5.39 (5.08 - 5.70) | 13.5 (13.3 - 13.8) |
| OEHR | 17,851 | 72,117 | 5.0 | 34.8 | 42 | 511 | 8.46 (6.09 - 11.4) | 14.7 (13.4 - 16.0) |
| VA | 0 | 0 | 0.0 | 0.0 | 0 | 0 | | |
| SIDIAP | 4,513 | 1,218 | 5.8 | 1.9 | <5 | 0 | < 0.86 (0.28 - 2.00) | |
| Meta-analysis | 348,045 | 1,025,674 | 253.7 | 889.9 | 1,477 | 12,586 | 5.82 (5.53 - 6.13) | 14.1 (13.9 - 14.4) |
| Meta-analysis (w/o USOC) | 55,999 | 178,071 | 31.5 | 115.3 | 280 | 2,095 | 8.89 (7.88 - 10.00) | 18.2 (17.4 - 19.0) |
| **PS-stratification / on-treatment TAR without escalation** | | | | | | | | |
| CCAE | 23,598 | 46,509 | 17.2 | 33.5 | 93 | 248 | 5.41 (4.37 - 6.63) | 7.39 (6.50 - 8.37) |
| DAG | 5,702 | 1,403 | 4.3 | 1.3 | 0 | 0 | | |
| ODOD | 16,421 | 46,384 | 10.2 | 37.3 | 144 | 1,056 | 14.1 (11.9 - 16.6) | 28.3 (26.6 - 30.1) |
| MDCR | 1,962 | 7,706 | 1.4 | 6.6 | 28 | 221 | 19.9 (13.2 - 28.7) | 33.4 (29.1 - 38.1) |
| MDCD | 1,350 | 6,710 | 0.7 | 3.9 | 10 | 106 | 14.8 (7.07 - 27.1) | 27.2 (22.3 - 32.9) |
| Open Claims | 315,211 | 852,161 | 235.8 | 778.8 | 1,369 | 10,730 | 5.81 (5.50 - 6.12) | 13.8 (13.5 - 14.0) |
| OEHR | 19,700 | 74,317 | 5.5 | 36.0 | 50 | 561 | 9.02 (6.70 - 11.9) | 15.6 (14.3 - 16.9) |
| VA | 11,226 | 106,553 | 5.9 | 86.0 | 66 | 1,051 | 11.2 (8.64 - 14.2) | 12.2 (11.5 - 13.0) |
| SIDIAP | 5,095 | 2,687 | 6.2 | 5.4 | <5 | 5 | < 0.80 (0.26 - 1.87) | 0.93 (0.30 - 2.18) |
| Meta-analysis | 374,930 | 1,019,371 | 268.7 | 885.7 | 1,656 | 12,595 | 6.16 (5.87 - 6.47) | 14.2 (14.0 - 14.5) |
| Meta-analysis (w/o USOC) | 59,719 | 167,210 | 32.9 | 106.8 | 287 | 1,865 | 8.71 (7.73 - 9.78) | 17.5 (16.7 - 18.3) |

##### 4.3.7 Measured renal dysfunction

**Supplementary Table 348:** Exposures, outcomes and risk estimates for Measured renal dysfunction in SGLT2I (male) (T) vs SU (male) (C) new-user cohorts under four different propensity score (PS) adjustment / time-at-risk (TAR) designs across data sources. We report total patient exposure time in 1,000 patient-year (KYr) units, uncalibrated and calibrated hazard ratios (HRs) and their 95% confidence intervals (CIs). Greyed out entries did not pass study diagnostics and are included only for completeness.

| | Patients | | Exposure (KYr) | | Outcomes | | Incidence (per KYr) | |
|---|---|---|---|---|---|---|---|---|
| | T | C | T | C | T | C | T | C |
| **PS-matching / on-treatment TAR** | | | | | | | | |
| CCAE | 20,587 | 46,816 | 17.6 | 39.1 | <5 | 23 | < 0.28 (0.09 - 0.66) | 0.59 (0.37 - 0.88) |
| DAG | 5,471 | 973 | 4.6 | 0.9 | 0 | 0 | | |
| ODOD | 15,718 | 48,808 | 11.2 | 44.4 | 7 | 116 | 0.62 (0.25 - 1.29) | 2.61 (2.16 - 3.13) |
| MDCR | 1,777 | 8,314 | 1.5 | 7.9 | 0 | <5 | | < 0.64 (0.21 - 1.49) |
| MDCD | 1,464 | 7,245 | 0.9 | 5.1 | 0 | 0 | | |
| Open Claims | 304,948 | 889,205 | 271.7 | 962.4 | 0 | 0 | | |
| OEHR | 18,285 | 73,829 | 5.8 | 41.4 | 0 | 0 | | |
| VA | 0 | 0 | 0.0 | 0.0 | 0 | 0 | | |
| SIDIAP | 4,535 | 1,216 | 6.6 | 2.2 | 13 | <5 | 1.97 (1.05 - 3.38) | < 2.24 (0.73 - 5.23) |
| Meta-analysis | 36,305 | 95,624 | 28.8 | 83.5 | <12 | 139 | < 0.42 (0.22 - 0.73) | 1.66 (1.40 - 1.96) |
| Meta-analysis (w/o USOC) | 36,305 | 95,624 | 28.8 | 83.5 | <12 | 139 | < 0.42 (0.22 - 0.73) | 1.66 (1.40 - 1.96) |
| **PS-stratification / on-treatment TAR** | | | | | | | | |
| CCAE | 24,082 | 47,464 | 20.0 | 39.5 | 5 | 23 | 0.25 (0.08 - 0.58) | 0.58 (0.37 - 0.87) |
| DAG | 5,702 | 1,403 | 4.7 | 1.4 | 0 | 0 | | |
| ODOD | 17,471 | 49,460 | 12.1 | 45.0 | 7 | 117 | 0.58 (0.23 - 1.19) | 2.60 (2.15 - 3.11) |
| MDCR | 2,136 | 8,601 | 1.7 | 8.1 | 0 | <5 | | < 0.62 (0.20 - 1.44) |
| MDCD | 1,507 | 7,493 | 0.9 | 5.3 | 0 | 0 | | |
| Open Claims | 332,801 | 894,802 | 289.7 | 968.3 | 0 | 0 | | |
| OEHR | 20,231 | 76,288 | 6.5 | 43.1 | 0 | 0 | | |
| VA | 12,350 | 109,470 | 7.4 | 100.4 | 23 | 492 | 3.09 (1.96 - 4.64) | 4.90 (4.48 - 5.35) |
| SIDIAP | 5,147 | 2,685 | 7.0 | 6.3 | 16 | 7 | 2.27 (1.30 - 3.69) | 1.12 (0.45 - 2.31) |
| Meta-analysis | 41,553 | 96,924 | 32.2 | 84.5 | 12 | 140 | 0.37 (0.19 - 0.65) | 1.66 (1.39 - 1.95) |
| Meta-analysis (w/o USOC) | 41,553 | 96,924 | 32.2 | 84.5 | 12 | 140 | 0.37 (0.19 - 0.65) | 1.66 (1.39 - 1.95) |
| **PS-matching / on-treatment TAR without escalation** | | | | | | | | |
| CCAE | 20,587 | 46,816 | 15.3 | 33.9 | <5 | 21 | < 0.33 (0.11 - 0.76) | 0.62 (0.38 - 0.95) |
| DAG | 5,471 | 973 | 4.2 | 0.8 | 0 | 0 | | |
| ODOD | 15,718 | 48,808 | 9.9 | 39.7 | 7 | 102 | 0.70 (0.28 - 1.45) | 2.57 (2.09 - 3.12) |
| MDCR | 1,777 | 8,314 | 1.4 | 7.2 | 0 | <5 | | < 0.70 (0.23 - 1.62) |
| MDCD | 1,464 | 7,245 | 0.8 | 4.2 | 0 | 0 | | |
| Open Claims | 304,948 | 889,205 | 230.2 | 816.1 | 0 | 0 | | |
| OEHR | 18,285 | 73,829 | 5.1 | 35.9 | 0 | 0 | | |
| VA | 0 | 0 | 0.0 | 0.0 | 0 | 0 | | |
| SIDIAP | 4,535 | 1,216 | 5.9 | 1.9 | 11 | <5 | 1.88 (0.94 - 3.36) | < 2.58 (0.84 - 6.03) |
| Meta-analysis | 36,305 | 95,624 | 25.2 | 73.6 | <12 | 123 | < 0.48 (0.25 - 0.83) | 1.67 (1.39 - 1.99) |
| Meta-analysis (w/o USOC) | 36,305 | 95,624 | 25.2 | 73.6 | <12 | 123 | < 0.48 (0.25 - 0.83) | 1.67 (1.39 - 1.99) |
| **PS-stratification / on-treatment TAR without escalation** | | | | | | | | |
| CCAE | 24,082 | 47,464 | 17.5 | 34.2 | 5 | 21 | 0.29 (0.09 - 0.67) | 0.61 (0.38 - 0.94) |
| DAG | 5,702 | 1,403 | 4.3 | 1.3 | 0 | 0 | | |
| ODOD | 17,471 | 49,460 | 10.8 | 40.3 | 7 | 103 | 0.65 (0.26 - 1.34) | 2.56 (2.09 - 3.10) |
| MDCR | 2,136 | 8,601 | 1.5 | 7.4 | 0 | <5 | | < 0.68 (0.22 - 1.58) |
| MDCD | 1,507 | 7,493 | 0.8 | 4.4 | 0 | 0 | | |
| Open Claims | 332,801 | 894,802 | 246.2 | 821.2 | 0 | 0 | | |
| OEHR | 20,231 | 76,288 | 5.7 | 37.3 | 0 | 0 | | |
| VA | 12,350 | 109,470 | 6.4 | 88.9 | 18 | 407 | 2.79 (1.65 - 4.41) | 4.58 (4.14 - 5.04) |
| SIDIAP | 5,147 | 2,685 | 6.3 | 5.3 | 14 | <5 | 2.23 (1.22 - 3.74) | < 0.94 (0.30 - 2.18) |
| Meta-analysis | 41,553 | 96,924 | 28.3 | 74.5 | 12 | 124 | 0.42 (0.22 - 0.74) | 1.66 (1.38 - 1.98) |
| Meta-analysis (w/o USOC) | 41,553 | 96,924 | 28.3 | 74.5 | 12 | 124 | 0.42 (0.22 - 0.74) | 1.66 (1.38 - 1.98) |

##### 4.3.8 Stroke

**Supplementary Table 349:** Exposures, outcomes and risk estimates for Stroke in SGLT2I (male) (T) vs SU (male) (C) new-user cohorts under four different propensity score (PS) adjustment / time-at-risk (TAR) designs across data sources. We report total patient exposure time in 1,000 patient-year (KYr) units, uncalibrated and calibrated hazard ratios (HRs) and their 95% confidence intervals (CIs). Greyed out entries did not pass study diagnostics and are included only for completeness.

| | Patients | | Exposure (KYr) | | Outcomes | | Incidence (per KYr) | |
|---|---|---|---|---|---|---|---|---|
| | T | C | T | C | T | C | T | C |
| **PS-matching / on-treatment TAR** | | | | | | | | |
| CCAE | 20,375 | 46,202 | 17.4 | 38.7 | 50 | 157 | 2.87 (2.13 - 3.79) | 4.06 (3.45 - 4.75) |
| DAG | 5,471 | 973 | 4.6 | 0.9 | 0 | 0 | | |
| ODOD | 15,392 | 47,482 | 10.9 | 43.0 | 73 | 506 | 6.67 (5.23 - 8.39) | 11.8 (10.8 - 12.8) |
| MDCR | 1,731 | 7,871 | 1.5 | 7.4 | 18 | 115 | 12.2 (7.20 - 19.2) | 15.5 (12.8 - 18.7) |
| MDCD | 1,404 | 6,941 | 0.9 | 4.8 | 6 | 67 | 7.04 (2.58 - 15.3) | 13.9 (10.7 - 17.6) |
| Open Claims | 297,554 | 862,949 | 265.9 | 932.4 | 874 | 6,002 | 3.29 (3.07 - 3.51) | 6.44 (6.28 - 6.60) |
| OEHR | 18,104 | 73,055 | 5.8 | 40.8 | 19 | 220 | 3.30 (1.99 - 5.16) | 5.39 (4.70 - 6.16) |
| VA | 0 | 0 | 0.0 | 0.0 | 0 | 0 | | |
| SIDIAP | 4,471 | 1,203 | 6.5 | 2.2 | 13 | 5 | 2.01 (1.07 - 3.43) | 2.27 (0.74 - 5.29) |
| Meta-analysis | 354,560 | 1,044,500 | 302.3 | 1067.1 | 1,040 | 7,067 | 3.44 (3.23 - 3.66) | 6.62 (6.47 - 6.78) |
| Meta-analysis (w/o USOC) | 57,006 | 181,551 | 36.4 | 134.7 | 166 | 1,065 | 4.56 (3.89 - 5.30) | 7.91 (7.44 - 8.40) |
| **PS-stratification / on-treatment TAR** | | | | | | | | |
| CCAE | 23,843 | 46,828 | 19.9 | 39.0 | 58 | 161 | 2.92 (2.22 - 3.77) | 4.13 (3.51 - 4.82) |
| DAG | 5,702 | 1,403 | 4.7 | 1.4 | 0 | 0 | | |
| ODOD | 17,093 | 48,024 | 11.8 | 43.5 | 81 | 519 | 6.85 (5.44 - 8.51) | 11.9 (10.9 - 13.0) |
| MDCR | 2,074 | 8,165 | 1.7 | 7.7 | 18 | 127 | 10.8 (6.43 - 17.1) | 16.6 (13.8 - 19.7) |
| MDCD | 1,446 | 7,166 | 0.9 | 5.0 | 6 | 70 | 6.93 (2.54 - 15.1) | 14.0 (10.9 - 17.6) |
| Open Claims | 324,481 | 868,102 | 283.2 | 937.7 | 934 | 6,104 | 3.30 (3.09 - 3.52) | 6.51 (6.35 - 6.67) |
| OEHR | 20,037 | 75,392 | 6.4 | 42.4 | 19 | 239 | 2.97 (1.79 - 4.63) | 5.64 (4.95 - 6.40) |
| VA | 12,216 | 109,267 | 7.4 | 100.4 | 19 | 364 | 2.58 (1.55 - 4.02) | 3.63 (3.26 - 4.02) |
| SIDIAP | 5,058 | 2,662 | 6.9 | 6.2 | 14 | 17 | 2.02 (1.10 - 3.39) | 2.75 (1.60 - 4.41) |
| Meta-analysis | 385,454 | 1,038,346 | 321.3 | 1062.6 | 1,092 | 7,023 | 3.40 (3.20 - 3.61) | 6.61 (6.46 - 6.77) |
| Meta-analysis (w/o USOC) | 60,973 | 170,244 | 38.1 | 124.9 | 158 | 919 | 4.15 (3.53 - 4.85) | 7.36 (6.89 - 7.85) |
| **PS-matching / on-treatment TAR without escalation** | | | | | | | | |
| CCAE | 20,375 | 46,202 | 15.2 | 33.5 | 38 | 140 | 2.51 (1.77 - 3.44) | 4.18 (3.52 - 4.93) |
| DAG | 5,471 | 973 | 4.2 | 0.8 | 0 | 0 | | |
| ODOD | 15,392 | 47,482 | 9.7 | 38.5 | 65 | 452 | 6.70 (5.17 - 8.54) | 11.7 (10.7 - 12.9) |
| MDCR | 1,731 | 7,871 | 1.3 | 6.8 | 15 | 108 | 11.4 (6.37 - 18.8) | 16.0 (13.1 - 19.3) |
| MDCD | 1,404 | 6,941 | 0.7 | 4.0 | <5 | 52 | < 6.97 (2.26 - 16.3) | 13.0 (9.70 - 17.0) |
| Open Claims | 297,554 | 862,949 | 225.3 | 790.7 | 750 | 5,201 | 3.33 (3.10 - 3.58) | 6.58 (6.40 - 6.76) |
| OEHR | 18,104 | 73,055 | 5.0 | 35.4 | 12 | 169 | 2.38 (1.23 - 4.16) | 4.77 (4.08 - 5.55) |
| VA | 0 | 0 | 0.0 | 0.0 | 0 | 0 | | |
| SIDIAP | 4,471 | 1,203 | 5.8 | 1.9 | 12 | 5 | 2.08 (1.08 - 3.64) | 2.61 (0.85 - 6.10) |
| Meta-analysis | 354,560 | 1,044,500 | 257.2 | 908.9 | <885 | 6,122 | < 3.44 (3.22 - 3.67) | 6.74 (6.57 - 6.91) |
| Meta-analysis (w/o USOC) | 57,006 | 181,551 | 31.9 | 118.2 | <135 | 921 | < 4.23 (3.54 - 5.00) | 7.79 (7.30 - 8.31) |
| **PS-stratification / on-treatment TAR without escalation** | | | | | | | | |
| CCAE | 23,843 | 46,828 | 17.3 | 33.8 | 46 | 144 | 2.65 (1.94 - 3.54) | 4.26 (3.59 - 5.01) |
| DAG | 5,702 | 1,403 | 4.3 | 1.3 | 0 | 0 | | |
| ODOD | 17,093 | 48,024 | 10.5 | 38.9 | 72 | 463 | 6.85 (5.36 - 8.63) | 11.9 (10.8 - 13.0) |
| MDCR | 2,074 | 8,165 | 1.5 | 7.0 | 15 | 120 | 10.1 (5.67 - 16.7) | 17.1 (14.2 - 20.5) |
| MDCD | 1,446 | 7,166 | 0.7 | 4.2 | <5 | 53 | < 6.85 (2.22 - 16.0) | 12.7 (9.54 - 16.7) |
| Open Claims | 324,481 | 868,102 | 240.8 | 795.4 | 806 | 5,285 | 3.35 (3.12 - 3.59) | 6.64 (6.47 - 6.83) |
| OEHR | 20,037 | 75,392 | 5.6 | 36.7 | 12 | 183 | 2.13 (1.10 - 3.72) | 4.98 (4.29 - 5.76) |
| VA | 12,216 | 109,267 | 6.4 | 88.9 | 14 | 317 | 2.19 (1.20 - 3.68) | 3.57 (3.19 - 3.98) |
| SIDIAP | 5,058 | 2,662 | 6.2 | 5.3 | 13 | 17 | 2.11 (1.12 - 3.60) | 3.23 (1.88 - 5.17) |
| Meta-analysis | 385,454 | 1,038,346 | 274.3 | 904.9 | 936 | 6,075 | 3.41 (3.20 - 3.64) | 6.71 (6.55 - 6.88) |
| Meta-analysis (w/o USOC) | 60,973 | 170,244 | 33.5 | 109.5 | 130 | 790 | 3.88 (3.24 - 4.61) | 7.22 (6.72 - 7.74) |

##### 4.3.9 Sudden cardiac death

**Supplementary Table 350:** Exposures, outcomes and risk estimates for Sudden cardiac death in SGLT2I (male) (T) vs SU (male) (C) new-user cohorts under four different propensity score (PS) adjustment / time-at-risk (TAR) designs across data sources. We report total patient exposure time in 1,000 patient-year (KYr) units, uncalibrated and calibrated hazard ratios (HRs) and their 95% confidence intervals (CIs). Greyed out entries did not pass study diagnostics and are included only for completeness.

| | Patients | | Exposure (KYr) | | Outcomes | | Incidence (per KYr) | |
|---|---|---|---|---|---|---|---|---|
| | T | C | T | C | T | C | T | C |
| **PS-matching / on-treatment TAR** | | | | | | | | |
| CCAE | 20,619 | 46,876 | 17.6 | 39.2 | 13 | 72 | 0.74 (0.39 - 1.26) | 1.84 (1.44 - 2.31) |
| DAG | 5,471 | 973 | 4.6 | 0.9 | 0 | 0 | | |
| ODOD | 15,698 | 48,810 | 11.2 | 44.5 | 18 | 167 | 1.61 (0.95 - 2.54) | 3.76 (3.21 - 4.37) |
| MDCR | 1,774 | 8,290 | 1.5 | 7.8 | 10 | 25 | 6.55 (3.14 - 12.1) | 3.20 (2.07 - 4.72) |
| MDCD | 1,457 | 7,215 | 0.9 | 5.1 | <5 | 22 | < 5.63 (1.83 - 13.1) | 4.34 (2.72 - 6.58) |
| Open Claims | 304,126 | 887,080 | 271.0 | 959.7 | 231 | 1,802 | 0.85 (0.75 - 0.97) | 1.88 (1.79 - 1.97) |
| OEHR | 18,264 | 73,759 | 5.8 | 41.4 | 5 | 79 | 0.86 (0.28 - 2.01) | 1.91 (1.51 - 2.38) |
| VA | 0 | 0 | 0.0 | 0.0 | 0 | 0 | | |
| SIDIAP | 4,551 | 1,220 | 6.6 | 2.2 | 0 | 0 | | |
| Meta-analysis | 360,481 | 1,064,815 | 307.2 | 1092.6 | 277 | 2,145 | 0.90 (0.80 - 1.01) | 1.96 (1.88 - 2.05) |
| Meta-analysis (w/o USOC) | 56,355 | 177,735 | 36.2 | 132.9 | 46 | 343 | 1.27 (0.93 - 1.70) | 2.58 (2.32 - 2.87) |
| **PS-stratification / on-treatment TAR** | | | | | | | | |
| CCAE | 24,111 | 47,518 | 20.1 | 39.6 | 15 | 72 | 0.75 (0.42 - 1.23) | 1.82 (1.42 - 2.29) |
| DAG | 5,702 | 1,403 | 4.7 | 1.4 | 0 | 0 | | |
| ODOD | 17,442 | 49,468 | 12.1 | 45.1 | 19 | 176 | 1.57 (0.94 - 2.45) | 3.90 (3.35 - 4.52) |
| MDCR | 2,130 | 8,575 | 1.7 | 8.0 | 11 | 26 | 6.47 (3.23 - 11.6) | 3.23 (2.11 - 4.73) |
| MDCD | 1,499 | 7,461 | 0.9 | 5.3 | <5 | 24 | < 5.55 (1.80 - 13.0) | 4.56 (2.92 - 6.79) |
| Open Claims | 331,685 | 892,613 | 288.8 | 965.5 | 259 | 1,851 | 0.90 (0.79 - 1.01) | 1.92 (1.83 - 2.01) |
| OEHR | 20,205 | 76,222 | 6.5 | 43.0 | 5 | 85 | 0.77 (0.25 - 1.81) | 1.98 (1.58 - 2.44) |
| VA | 12,496 | 111,025 | 7.5 | 102.2 | <10 | 47 | < 1.33 (0.64 - 2.45) | 0.46 (0.34 - 0.61) |
| SIDIAP | 5,160 | 2,691 | 7.1 | 6.3 | 0 | <5 | | < 0.80 (0.26 - 1.86) |
| Meta-analysis | 393,443 | 1,065,821 | 327.5 | 1093.2 | 298 | 2,184 | 0.91 (0.81 - 1.02) | 2.00 (1.91 - 2.08) |
| Meta-analysis (w/o USOC) | 61,758 | 173,208 | 38.7 | 127.7 | 39 | 333 | 1.01 (0.72 - 1.38) | 2.61 (2.34 - 2.90) |
| **PS-matching / on-treatment TAR without escalation** | | | | | | | | |
| CCAE | 20,619 | 46,876 | 15.3 | 34.0 | 9 | 64 | 0.59 (0.27 - 1.11) | 1.88 (1.45 - 2.41) |
| DAG | 5,471 | 973 | 4.2 | 0.8 | 0 | 0 | | |
| ODOD | 15,698 | 48,810 | 9.9 | 39.8 | 15 | 149 | 1.51 (0.85 - 2.49) | 3.75 (3.17 - 4.40) |
| MDCR | 1,774 | 8,290 | 1.4 | 7.2 | 9 | 19 | 6.64 (3.03 - 12.6) | 2.65 (1.60 - 4.14) |
| MDCD | 1,457 | 7,215 | 0.8 | 4.2 | <5 | 20 | < 6.66 (2.16 - 15.5) | 4.76 (2.91 - 7.36) |
| Open Claims | 304,126 | 887,080 | 229.6 | 813.9 | 202 | 1,532 | 0.88 (0.76 - 1.01) | 1.88 (1.79 - 1.98) |
| OEHR | 18,264 | 73,759 | 5.1 | 35.9 | <5 | 55 | < 0.98 (0.32 - 2.29) | 1.53 (1.15 - 2.00) |
| VA | 0 | 0 | 0.0 | 0.0 | 0 | 0 | | |
| SIDIAP | 4,551 | 1,220 | 5.9 | 2.0 | 0 | 0 | | |
| Meta-analysis | 358,707 | 1,056,525 | 260.0 | 923.5 | <231 | 1,800 | < 0.89 (0.78 - 1.01) | 1.95 (1.86 - 2.04) |
| Meta-analysis (w/o USOC) | 54,581 | 169,445 | 30.4 | 109.6 | <29 | 268 | < 0.96 (0.64 - 1.37) | 2.44 (2.16 - 2.76) |
| **PS-stratification / on-treatment TAR without escalation** | | | | | | | | |
| CCAE | 24,111 | 47,518 | 17.5 | 34.3 | 11 | 64 | 0.63 (0.31 - 1.12) | 1.87 (1.44 - 2.38) |
| DAG | 5,702 | 1,403 | 4.3 | 1.3 | 0 | 0 | | |
| ODOD | 17,442 | 49,468 | 10.7 | 40.3 | 16 | 156 | 1.49 (0.85 - 2.42) | 3.87 (3.28 - 4.52) |
| MDCR | 2,130 | 8,575 | 1.5 | 7.4 | 10 | 20 | 6.59 (3.16 - 12.1) | 2.71 (1.66 - 4.19) |
| MDCD | 1,499 | 7,461 | 0.8 | 4.4 | <5 | 20 | < 6.55 (2.13 - 15.3) | 4.58 (2.80 - 7.08) |
| Open Claims | 331,685 | 892,613 | 245.5 | 818.9 | 228 | 1,575 | 0.93 (0.81 - 1.06) | 1.92 (1.83 - 2.02) |
| OEHR | 20,205 | 76,222 | 5.7 | 37.3 | <5 | 60 | < 0.88 (0.29 - 2.05) | 1.61 (1.23 - 2.07) |
| VA | 12,496 | 111,025 | 6.5 | 90.4 | <10 | 38 | < 1.54 (0.74 - 2.83) | 0.42 (0.30 - 0.58) |
| SIDIAP | 5,160 | 2,691 | 6.3 | 5.4 | 0 | <5 | | < 0.93 (0.30 - 2.18) |
| Meta-analysis | 393,443 | 1,065,821 | 279.4 | 930.8 | <260 | 1,855 | < 0.93 (0.82 - 1.05) | 1.99 (1.90 - 2.09) |
| Meta-analysis (w/o USOC) | 61,758 | 173,208 | 34.0 | 111.9 | <32 | 280 | < 0.94 (0.64 - 1.33) | 2.50 (2.22 - 2.81) |

##### 4.3.10 Abnormal weight gain

**Supplementary Table 351:** Exposures, outcomes and risk estimates for Abnormal weight gain in SGLT2I (male) (T) vs SU (male) (C) new-user cohorts under four different propensity score (PS) adjustment / time-at-risk (TAR) designs across data sources. We report total patient exposure time in 1,000 patient-year (KYr) units, uncalibrated and calibrated hazard ratios (HRs) and their 95% confidence intervals (CIs). Greyed out entries did not pass study diagnostics and are included only for completeness.

| | Patients | | Exposure (KYr) | | Outcomes | | Incidence (per KYr) | |
|---|---|---|---|---|---|---|---|---|
| | T | C | T | C | T | C | T | C |
| **PS-matching / on-treatment TAR** | | | | | | | | |
| CCAE | 19,985 | 45,765 | 17.0 | 38.2 | 53 | 175 | 3.12 (2.34 - 4.09) | 4.59 (3.93 - 5.32) |
| DAG | 5,432 | 968 | 4.5 | 0.9 | 0 | <5 | | < 5.50 (1.79 - 12.8) |
| ODOD | 15,208 | 47,704 | 10.8 | 43.3 | 32 | 244 | 2.96 (2.02 - 4.18) | 5.64 (4.95 - 6.39) |
| MDCR | 1,747 | 8,227 | 1.5 | 7.7 | <5 | 31 | < 3.36 (1.09 - 7.84) | 4.01 (2.72 - 5.69) |
| MDCD | 1,367 | 6,949 | 0.8 | 4.8 | <5 | 49 | < 6.02 (1.95 - 14.0) | 10.1 (7.50 - 13.4) |
| Open Claims | 294,899 | 868,913 | 262.1 | 939.0 | 290 | 1,795 | 1.11 (0.98 - 1.24) | 1.91 (1.82 - 2.00) |
| OEHR | 17,680 | 72,048 | 5.6 | 40.2 | 26 | 216 | 4.63 (3.02 - 6.78) | 5.37 (4.68 - 6.14) |
| VA | 0 | 0 | 0.0 | 0.0 | 0 | 0 | | |
| SIDIAP | 4,500 | 1,199 | 6.5 | 2.2 | 8 | <5 | 1.23 (0.53 - 2.42) | < 2.27 (0.74 - 5.30) |
| Meta-analysis | 350,886 | 1,049,606 | 297.8 | 1073.2 | <411 | 2,510 | < 1.38 (1.25 - 1.52) | 2.34 (2.25 - 2.43) |
| Meta-analysis (w/o USOC) | 55,987 | 180,693 | 35.7 | 134.2 | <121 | 715 | < 3.39 (2.81 - 4.05) | 5.33 (4.94 - 5.73) |
| **PS-stratification / on-treatment TAR** | | | | | | | | |
| CCAE | 23,300 | 46,404 | 19.3 | 38.5 | 68 | 176 | 3.52 (2.73 - 4.46) | 4.57 (3.92 - 5.30) |
| DAG | 5,661 | 1,392 | 4.7 | 1.4 | 0 | <5 | | < 3.64 (1.18 - 8.49) |
| ODOD | 16,867 | 48,341 | 11.7 | 43.9 | 32 | 250 | 2.74 (1.88 - 3.87) | 5.69 (5.01 - 6.45) |
| MDCR | 2,099 | 8,500 | 1.7 | 7.9 | <5 | 34 | < 3.01 (0.98 - 7.02) | 4.28 (2.96 - 5.98) |
| MDCD | 1,408 | 7,185 | 0.8 | 5.0 | <5 | 51 | < 5.93 (1.92 - 13.8) | 10.2 (7.56 - 13.4) |
| Open Claims | 321,381 | 874,572 | 279.0 | 944.9 | 317 | 1,820 | 1.14 (1.01 - 1.27) | 1.93 (1.84 - 2.02) |
| OEHR | 19,556 | 74,612 | 6.2 | 41.9 | 30 | 229 | 4.81 (3.24 - 6.86) | 5.46 (4.78 - 6.22) |
| VA | 12,226 | 109,285 | 7.4 | 100.4 | <10 | 172 | < 1.36 (0.65 - 2.49) | 1.71 (1.47 - 1.99) |
| SIDIAP | 5,107 | 2,666 | 7.0 | 6.2 | 8 | 8 | 1.15 (0.50 - 2.26) | 1.29 (0.55 - 2.53) |
| Meta-analysis | 381,104 | 1,043,929 | 316.2 | 1069.2 | 447 | 2,475 | 1.41 (1.29 - 1.55) | 2.31 (2.22 - 2.41) |
| Meta-analysis (w/o USOC) | 59,723 | 169,357 | 37.2 | 124.4 | 130 | 655 | 3.49 (2.92 - 4.15) | 5.27 (4.87 - 5.69) |
| **PS-matching / on-treatment TAR without escalation** | | | | | | | | |
| CCAE | 19,985 | 45,765 | 14.8 | 33.1 | 47 | 147 | 3.18 (2.34 - 4.23) | 4.44 (3.75 - 5.22) |
| DAG | 5,432 | 968 | 4.2 | 0.8 | 0 | <5 | | < 5.98 (1.94 - 14.0) |
| ODOD | 15,208 | 47,704 | 9.6 | 38.8 | 25 | 211 | 2.60 (1.69 - 3.84) | 5.44 (4.73 - 6.22) |
| MDCR | 1,747 | 8,227 | 1.3 | 7.1 | <5 | 28 | < 3.77 (1.22 - 8.79) | 3.95 (2.63 - 5.71) |
| MDCD | 1,367 | 6,949 | 0.7 | 4.0 | <5 | 35 | < 7.15 (2.32 - 16.7) | 8.71 (6.06 - 12.1) |
| Open Claims | 294,899 | 868,913 | 222.2 | 797.2 | 229 | 1,485 | 1.03 (0.90 - 1.17) | 1.86 (1.77 - 1.96) |
| OEHR | 17,680 | 72,048 | 4.9 | 34.9 | 21 | 178 | 4.27 (2.64 - 6.52) | 5.10 (4.37 - 5.90) |
| VA | 0 | 0 | 0.0 | 0.0 | 0 | 0 | | |
| SIDIAP | 4,500 | 1,199 | 5.8 | 1.9 | 8 | <5 | 1.38 (0.60 - 2.72) | < 2.62 (0.85 - 6.11) |
| Meta-analysis | 350,886 | 1,049,606 | 253.5 | 915.1 | <332 | 2,084 | < 1.31 (1.17 - 1.46) | 2.28 (2.18 - 2.38) |
| Meta-analysis (w/o USOC) | 55,987 | 180,693 | 31.3 | 118.0 | <103 | 599 | < 3.29 (2.68 - 3.99) | 5.08 (4.68 - 5.50) |
| **PS-stratification / on-treatment TAR without escalation** | | | | | | | | |
| CCAE | 23,300 | 46,404 | 16.9 | 33.4 | 60 | 148 | 3.55 (2.71 - 4.57) | 4.43 (3.74 - 5.20) |
| DAG | 5,661 | 1,392 | 4.3 | 1.3 | 0 | <5 | | < 3.98 (1.29 - 9.29) |
| ODOD | 16,867 | 48,341 | 10.4 | 39.3 | 25 | 217 | 2.41 (1.56 - 3.56) | 5.52 (4.81 - 6.30) |
| MDCR | 2,099 | 8,500 | 1.5 | 7.3 | <5 | 31 | < 3.36 (1.09 - 7.83) | 4.25 (2.89 - 6.04) |
| MDCD | 1,408 | 7,185 | 0.7 | 4.2 | <5 | 36 | < 7.03 (2.28 - 16.4) | 8.61 (6.03 - 11.9) |
| Open Claims | 321,381 | 874,572 | 237.4 | 802.3 | 254 | 1,507 | 1.07 (0.94 - 1.21) | 1.88 (1.78 - 1.98) |
| OEHR | 19,556 | 74,612 | 5.5 | 36.4 | 24 | 188 | 4.37 (2.80 - 6.51) | 5.17 (4.46 - 5.96) |
| VA | 12,226 | 109,285 | 6.4 | 88.9 | <10 | 131 | < 1.57 (0.75 - 2.88) | 1.47 (1.23 - 1.75) |
| SIDIAP | 5,107 | 2,666 | 6.2 | 5.3 | 8 | 6 | 1.29 (0.56 - 2.53) | 1.13 (0.41 - 2.46) |
| Meta-analysis | 381,104 | 1,043,929 | 270.1 | 911.5 | 363 | 2,060 | 1.34 (1.21 - 1.49) | 2.26 (2.16 - 2.36) |
| Meta-analysis (w/o USOC) | 59,723 | 169,357 | 32.7 | 109.2 | 109 | 553 | 3.33 (2.73 - 4.02) | 5.07 (4.65 - 5.51) |

##### 4.3.11 Abnormal weight loss

**Supplementary Table 352:** Exposures, outcomes and risk estimates for Abnormal weight loss in SGLT2I (male) (T) vs SU (male) (C) new-user cohorts under four different propensity score (PS) adjustment / time-at-risk (TAR) designs across data sources. We report total patient exposure time in 1,000 patient-year (KYr) units, uncalibrated and calibrated hazard ratios (HRs) and their 95% confidence intervals (CIs). Greyed out entries did not pass study diagnostics and are included only for completeness.

| | Patients | | Exposure (KYr) | | Outcomes | | Incidence (per KYr) | |
|---|---|---|---|---|---|---|---|---|
| | T | C | T | C | T | C | T | C |
| **PS-matching / on-treatment TAR** | | | | | | | | |
| CCAE | 20,137 | 45,662 | 17.1 | 38.2 | 124 | 211 | 7.24 (6.02 - 8.63) | 5.53 (4.81 - 6.32) |
| DAG | 5,363 | 957 | 4.5 | 0.9 | 39 | <5 | 8.74 (6.22 - 12.0) | < 5.56 (1.81 - 13.0) |
| ODOD | 15,188 | 46,639 | 10.8 | 42.3 | 173 | 569 | 16.1 (13.8 - 18.7) | 13.4 (12.4 - 14.6) |
| MDCR | 1,712 | 7,847 | 1.5 | 7.4 | 32 | 113 | 21.8 (14.9 - 30.8) | 15.2 (12.5 - 18.3) |
| MDCD | 1,400 | 6,848 | 0.8 | 4.8 | 14 | 50 | 16.6 (9.06 - 27.8) | 10.5 (7.76 - 13.8) |
| Open Claims | 295,525 | 858,080 | 262.7 | 927.2 | 1,500 | 4,775 | 5.71 (5.42 - 6.01) | 5.15 (5.00 - 5.30) |
| OEHR | 17,739 | 71,325 | 5.6 | 39.8 | 72 | 410 | 12.8 (10.0 - 16.1) | 10.3 (9.34 - 11.4) |
| VA | 0 | 0 | 0.0 | 0.0 | 0 | 0 | | |
| SIDIAP | 4,493 | 1,206 | 6.5 | 2.2 | 29 | 8 | 4.43 (2.97 - 6.36) | 3.62 (1.56 - 7.14) |
| Meta-analysis | 361,557 | 1,038,564 | 309.5 | 1062.8 | 1,983 | <6,141 | 6.41 (6.13 - 6.69) | < 5.78 (5.63 - 5.92) |
| Meta-analysis (w/o USOC) | 66,032 | 180,484 | 46.8 | 135.6 | 483 | <1,366 | 10.3 (9.41 - 11.3) | < 10.1 (9.55 - 10.6) |
| **PS-stratification / on-treatment TAR** | | | | | | | | |
| CCAE | 23,568 | 46,288 | 19.6 | 38.5 | 145 | 215 | 7.41 (6.25 - 8.72) | 5.58 (4.86 - 6.37) |
| DAG | 5,592 | 1,377 | 4.6 | 1.4 | 39 | 7 | 8.47 (6.02 - 11.6) | 5.17 (2.08 - 10.7) |
| ODOD | 16,888 | 47,195 | 11.6 | 42.9 | 184 | 582 | 15.8 (13.6 - 18.3) | 13.6 (12.5 - 14.7) |
| MDCR | 2,062 | 8,142 | 1.6 | 7.7 | 35 | 119 | 21.4 (14.9 - 29.7) | 15.5 (12.8 - 18.5) |
| MDCD | 1,443 | 7,139 | 0.9 | 5.0 | 14 | 55 | 16.3 (8.91 - 27.3) | 11.0 (8.28 - 14.3) |
| Open Claims | 322,505 | 863,615 | 280.1 | 933.0 | 1,595 | 4,867 | 5.69 (5.42 - 5.98) | 5.22 (5.07 - 5.37) |
| OEHR | 19,644 | 73,670 | 6.3 | 41.3 | 79 | 446 | 12.6 (9.99 - 15.7) | 10.8 (9.82 - 11.8) |
| VA | 12,060 | 106,937 | 7.2 | 98.2 | 100 | 668 | 13.8 (11.2 - 16.8) | 6.80 (6.30 - 7.34) |
| SIDIAP | 5,093 | 2,666 | 7.0 | 6.2 | 36 | 18 | 5.15 (3.61 - 7.13) | 2.89 (1.71 - 4.57) |
| Meta-analysis | 382,605 | 1,030,768 | 317.5 | 1055.8 | 2,003 | 6,110 | 6.31 (6.03 - 6.59) | 5.79 (5.64 - 5.93) |
| Meta-analysis (w/o USOC) | 60,100 | 167,153 | 37.5 | 122.7 | 408 | 1,243 | 10.9 (9.86 - 12.0) | 10.1 (9.57 - 10.7) |
| **PS-matching / on-treatment TAR without escalation** | | | | | | | | |
| CCAE | 20,137 | 45,662 | 14.9 | 33.1 | 109 | 184 | 7.30 (6.00 - 8.81) | 5.56 (4.78 - 6.42) |
| DAG | 5,363 | 957 | 4.1 | 0.8 | 36 | <5 | 8.76 (6.14 - 12.1) | < 6.05 (1.97 - 14.1) |
| ODOD | 15,188 | 46,639 | 9.5 | 37.9 | 150 | 497 | 15.7 (13.3 - 18.4) | 13.1 (12.0 - 14.3) |
| MDCR | 1,712 | 7,847 | 1.3 | 6.8 | 29 | 100 | 22.2 (14.9 - 31.9) | 14.7 (11.9 - 17.8) |
| MDCD | 1,400 | 6,848 | 0.7 | 4.0 | 13 | 36 | 18.1 (9.64 - 31.0) | 9.06 (6.35 - 12.5) |
| Open Claims | 295,525 | 858,080 | 222.8 | 786.7 | 1,301 | 4,071 | 5.84 (5.53 - 6.17) | 5.17 (5.02 - 5.34) |
| OEHR | 17,739 | 71,325 | 4.9 | 34.5 | 65 | 336 | 13.2 (10.2 - 16.8) | 9.73 (8.72 - 10.8) |
| VA | 0 | 0 | 0.0 | 0.0 | 0 | 0 | | |
| SIDIAP | 4,493 | 1,206 | 5.8 | 1.9 | 25 | 6 | 4.30 (2.78 - 6.35) | 3.13 (1.15 - 6.80) |
| Meta-analysis | 361,557 | 1,038,564 | 264.1 | 905.8 | 1,728 | <5,235 | 6.54 (6.24 - 6.86) | < 5.78 (5.62 - 5.94) |
| Meta-analysis (w/o USOC) | 66,032 | 180,484 | 41.3 | 119.1 | 427 | <1,164 | 10.3 (9.37 - 11.4) | < 9.78 (9.22 - 10.4) |
| **PS-stratification / on-treatment TAR without escalation** | | | | | | | | |
| CCAE | 23,568 | 46,288 | 17.1 | 33.4 | 126 | 188 | 7.37 (6.14 - 8.77) | 5.62 (4.85 - 6.49) |
| DAG | 5,592 | 1,377 | 4.2 | 1.2 | 36 | 5 | 8.49 (5.95 - 11.8) | 4.04 (1.31 - 9.43) |
| ODOD | 16,888 | 47,195 | 10.3 | 38.4 | 160 | 510 | 15.5 (13.2 - 18.1) | 13.3 (12.2 - 14.5) |
| MDCR | 2,062 | 8,142 | 1.5 | 7.1 | 31 | 105 | 21.1 (14.4 - 30.0) | 14.9 (12.2 - 18.0) |
| MDCD | 1,443 | 7,139 | 0.7 | 4.2 | 13 | 41 | 17.8 (9.47 - 30.4) | 9.87 (7.08 - 13.4) |
| Open Claims | 322,505 | 863,615 | 238.3 | 791.7 | 1,386 | 4,156 | 5.82 (5.51 - 6.13) | 5.25 (5.09 - 5.41) |
| OEHR | 19,644 | 73,670 | 5.5 | 35.8 | 70 | 368 | 12.7 (9.90 - 16.1) | 10.3 (9.25 - 11.4) |
| VA | 12,060 | 106,937 | 6.3 | 86.9 | 87 | 571 | 13.9 (11.1 - 17.1) | 6.57 (6.04 - 7.13) |
| SIDIAP | 5,093 | 2,666 | 6.2 | 5.3 | 32 | 15 | 5.14 (3.52 - 7.26) | 2.82 (1.58 - 4.66) |
| Meta-analysis | 382,605 | 1,030,768 | 271.2 | 899.4 | 1,742 | 5,222 | 6.42 (6.12 - 6.73) | 5.81 (5.65 - 5.97) |
| Meta-analysis (w/o USOC) | 60,100 | 167,153 | 32.9 | 107.6 | 356 | 1,066 | 10.8 (9.71 - 12.0) | 9.91 (9.32 - 10.5) |

##### 4.3.12 Acute pancreatitis

**Supplementary Table 353:** Exposures, outcomes and risk estimates for Acute pancreatitis in SGLT2I (male) (T) vs SU (male) (C) new-user cohorts under four different propensity score (PS) adjustment / time-at-risk (TAR) designs across data sources. We report total patient exposure time in 1,000 patient-year (KYr) units, uncalibrated and calibrated hazard ratios (HRs) and their 95% confidence intervals (CIs). Greyed out entries did not pass study diagnostics and are included only for completeness.

| | Patients | | Exposure (KYr) | | Outcomes | | Incidence (per KYr) | |
|---|---|---|---|---|---|---|---|---|
| | T | C | T | C | T | C | T | C |
| **PS-matching / on-treatment TAR** | | | | | | | | |
| CCAE | 20,418 | 46,370 | 17.4 | 38.8 | 36 | 108 | 2.07 (1.45 - 2.86) | 2.79 (2.28 - 3.36) |
| DAG | 5,471 | 973 | 4.6 | 0.9 | 0 | 0 | | |
| ODOD | 15,572 | 48,378 | 11.1 | 44.0 | 27 | 164 | 2.43 (1.60 - 3.54) | 3.73 (3.18 - 4.35) |
| MDCR | 1,762 | 8,208 | 1.5 | 7.7 | <5 | 27 | < 3.31 (1.07 - 7.72) | 3.49 (2.30 - 5.07) |
| MDCD | 1,436 | 7,035 | 0.9 | 4.9 | 6 | 38 | 6.97 (2.56 - 15.2) | 7.70 (5.45 - 10.6) |
| Open Claims | 300,492 | 875,565 | 267.7 | 947.3 | 357 | 2,128 | 1.33 (1.20 - 1.48) | 2.25 (2.15 - 2.34) |
| OEHR | 18,169 | 73,312 | 5.8 | 41.1 | 11 | 126 | 1.91 (0.95 - 3.41) | 3.07 (2.56 - 3.65) |
| VA | 0 | 0 | 0.0 | 0.0 | 0 | 0 | | |
| SIDIAP | 4,533 | 1,218 | 6.6 | 2.2 | <5 | 0 | < 0.76 (0.25 - 1.77) | |
| Meta-analysis | 357,849 | 1,058,868 | 304.4 | 1083.8 | <442 | 2,591 | < 1.45 (1.32 - 1.59) | 2.39 (2.30 - 2.48) |
| Meta-analysis (w/o USOC) | 57,357 | 183,303 | 36.7 | 136.5 | <85 | 463 | < 2.32 (1.85 - 2.87) | 3.39 (3.09 - 3.72) |
| **PS-stratification / on-treatment TAR** | | | | | | | | |
| CCAE | 23,890 | 47,008 | 19.9 | 39.1 | 42 | 109 | 2.11 (1.52 - 2.86) | 2.78 (2.29 - 3.36) |
| DAG | 5,702 | 1,403 | 4.7 | 1.4 | 0 | 0 | | |
| ODOD | 17,307 | 49,005 | 12.0 | 44.6 | 29 | 167 | 2.41 (1.62 - 3.47) | 3.75 (3.20 - 4.36) |
| MDCR | 2,119 | 8,479 | 1.7 | 8.0 | <5 | 27 | < 2.96 (0.96 - 6.91) | 3.40 (2.24 - 4.94) |
| MDCD | 1,481 | 7,259 | 0.9 | 5.1 | 6 | 41 | 6.86 (2.52 - 14.9) | 8.00 (5.74 - 10.9) |
| Open Claims | 327,945 | 880,953 | 285.4 | 952.9 | 377 | 2,155 | 1.32 (1.19 - 1.46) | 2.26 (2.17 - 2.36) |
| OEHR | 20,108 | 75,742 | 6.4 | 42.7 | 13 | 133 | 2.02 (1.08 - 3.46) | 3.11 (2.61 - 3.69) |
| VA | 12,372 | 109,694 | 7.4 | 100.9 | <10 | 214 | < 1.34 (0.64 - 2.47) | 2.12 (1.85 - 2.43) |
| SIDIAP | 5,142 | 2,685 | 7.0 | 6.3 | <5 | <5 | < 0.71 (0.23 - 1.66) | < 0.80 (0.26 - 1.86) |
| Meta-analysis | 389,250 | 1,052,708 | 323.7 | 1079.3 | 461 | 2,564 | 1.42 (1.30 - 1.56) | 2.38 (2.28 - 2.47) |
| Meta-analysis (w/o USOC) | 61,305 | 171,755 | 38.3 | 126.4 | 84 | 409 | 2.19 (1.75 - 2.71) | 3.23 (2.93 - 3.56) |
| **PS-matching / on-treatment TAR without escalation** | | | | | | | | |
| CCAE | 20,418 | 46,370 | 15.2 | 33.6 | 28 | 91 | 1.84 (1.23 - 2.67) | 2.71 (2.18 - 3.32) |
| DAG | 5,471 | 973 | 4.2 | 0.8 | 0 | 0 | | |
| ODOD | 15,572 | 48,378 | 9.8 | 39.4 | 21 | 146 | 2.14 (1.32 - 3.26) | 3.71 (3.13 - 4.36) |
| MDCR | 1,762 | 8,208 | 1.3 | 7.1 | <5 | 24 | < 3.72 (1.21 - 8.69) | 3.38 (2.17 - 5.03) |
| MDCD | 1,436 | 7,035 | 0.7 | 4.1 | 5 | 33 | 6.88 (2.23 - 16.1) | 8.07 (5.55 - 11.3) |
| Open Claims | 300,492 | 875,565 | 227.1 | 804.0 | 288 | 1,765 | 1.27 (1.13 - 1.42) | 2.20 (2.09 - 2.30) |
| OEHR | 18,169 | 73,312 | 5.1 | 35.7 | 9 | 90 | 1.78 (0.81 - 3.38) | 2.52 (2.03 - 3.10) |
| VA | 0 | 0 | 0.0 | 0.0 | 0 | 0 | | |
| SIDIAP | 4,533 | 1,218 | 5.8 | 2.0 | <5 | 0 | < 0.85 (0.28 - 1.99) | |
| Meta-analysis | 357,849 | 1,058,868 | 259.2 | 923.8 | <356 | 2,149 | < 1.37 (1.23 - 1.52) | 2.33 (2.23 - 2.43) |
| Meta-analysis (w/o USOC) | 57,357 | 183,303 | 32.1 | 119.8 | <68 | 384 | < 2.12 (1.64 - 2.68) | 3.20 (2.89 - 3.54) |
| **PS-stratification / on-treatment TAR without escalation** | | | | | | | | |
| CCAE | 23,890 | 47,008 | 17.4 | 34.0 | 34 | 92 | 1.96 (1.36 - 2.74) | 2.71 (2.18 - 3.32) |
| DAG | 5,702 | 1,403 | 4.3 | 1.3 | 0 | 0 | | |
| ODOD | 17,307 | 49,005 | 10.7 | 39.9 | 23 | 149 | 2.16 (1.37 - 3.24) | 3.74 (3.16 - 4.39) |
| MDCR | 2,119 | 8,479 | 1.5 | 7.3 | <5 | 24 | < 3.32 (1.08 - 7.74) | 3.29 (2.11 - 4.89) |
| MDCD | 1,481 | 7,259 | 0.7 | 4.2 | 5 | 36 | 6.76 (2.20 - 15.8) | 8.47 (5.93 - 11.7) |
| Open Claims | 327,945 | 880,953 | 242.8 | 808.8 | 306 | 1,785 | 1.26 (1.12 - 1.41) | 2.21 (2.11 - 2.31) |
| OEHR | 20,108 | 75,742 | 5.7 | 37.0 | 11 | 96 | 1.95 (0.97 - 3.48) | 2.59 (2.10 - 3.17) |
| VA | 12,372 | 109,694 | 6.4 | 89.3 | <10 | 175 | < 1.55 (0.75 - 2.86) | 1.96 (1.68 - 2.27) |
| SIDIAP | 5,142 | 2,685 | 6.3 | 5.4 | <5 | <5 | < 0.80 (0.26 - 1.86) | < 0.93 (0.30 - 2.18) |
| Meta-analysis | 389,250 | 1,052,708 | 276.5 | 919.7 | 374 | 2,122 | 1.35 (1.22 - 1.50) | 2.31 (2.21 - 2.41) |
| Meta-analysis (w/o USOC) | 61,305 | 171,755 | 33.7 | 110.9 | 68 | 337 | 2.02 (1.57 - 2.56) | 3.04 (2.72 - 3.38) |

##### 4.3.13 Bone fracture

**Supplementary Table 354:** Exposures, outcomes and risk estimates for Bone fracture in SGLT2I (male) (T) vs SU (male) (C) new-user cohorts under four different propensity score (PS) adjustment / time-at-risk (TAR) designs across data sources. We report total patient exposure time in 1,000 patient-year (KYr) units, uncalibrated and calibrated hazard ratios (HRs) and their 95% confidence intervals (CIs). Greyed out entries did not pass study diagnostics and are included only for completeness.

| | Patients | | Exposure (KYr) | | Outcomes | | Incidence (per KYr) | |
|---|---|---|---|---|---|---|---|---|
| | T | C | T | C | T | C | T | C |
| **PS-matching / on-treatment TAR** | | | | | | | | |
| CCAE | 19,098 | 43,539 | 16.0 | 35.9 | 249 | 567 | 15.6 (13.7 - 17.6) | 15.8 (14.5 - 17.2) |
| DAG | 4,918 | 876 | 4.0 | 0.8 | 56 | 16 | 13.9 (10.5 - 18.0) | 19.3 (11.1 - 31.4) |
| ODOD | 14,591 | 45,502 | 10.3 | 40.7 | 197 | 930 | 19.2 (16.6 - 22.1) | 22.8 (21.4 - 24.4) |
| MDCR | 1,640 | 7,527 | 1.4 | 7.0 | 33 | 185 | 23.7 (16.3 - 33.3) | 26.6 (22.9 - 30.7) |
| MDCD | 1,290 | 6,310 | 0.8 | 4.3 | 35 | 139 | 45.6 (31.8 - 63.4) | 32.2 (27.0 - 38.0) |
| Open Claims | 271,263 | 798,298 | 240.8 | 855.6 | 2,620 | 12,263 | 10.9 (10.5 - 11.3) | 14.3 (14.1 - 14.6) |
| OEHR | 17,159 | 68,847 | 5.5 | 38.0 | 64 | 636 | 11.7 (9.00 - 14.9) | 16.7 (15.5 - 18.1) |
| VA | 0 | 0 | 0.0 | 0.0 | 0 | 0 | | |
| SIDIAP | 4,092 | 1,110 | 5.9 | 2.0 | 51 | 23 | 8.65 (6.44 - 11.4) | 11.4 (7.21 - 17.1) |
| Meta-analysis | 334,051 | 972,009 | 284.6 | 984.3 | 3,305 | 14,759 | 11.6 (11.2 - 12.0) | 15.0 (14.8 - 15.2) |
| Meta-analysis (w/o USOC) | 62,788 | 173,711 | 43.8 | 128.7 | 685 | 2,496 | 15.6 (14.5 - 16.9) | 19.4 (18.6 - 20.2) |
| **PS-stratification / on-treatment TAR** | | | | | | | | |
| CCAE | 22,298 | 44,128 | 18.2 | 36.2 | 296 | 574 | 16.2 (14.4 - 18.2) | 15.8 (14.6 - 17.2) |
| DAG | 5,113 | 1,271 | 4.2 | 1.3 | 56 | 26 | 13.5 (10.2 - 17.5) | 20.7 (13.5 - 30.3) |
| ODOD | 16,194 | 45,989 | 11.1 | 41.2 | 211 | 946 | 19.1 (16.6 - 21.8) | 23.0 (21.5 - 24.5) |
| MDCR | 1,967 | 7,745 | 1.6 | 7.1 | 38 | 194 | 24.5 (17.3 - 33.6) | 27.2 (23.5 - 31.3) |
| MDCD | 1,321 | 6,508 | 0.8 | 4.5 | 35 | 149 | 45.1 (31.4 - 62.7) | 33.2 (28.1 - 39.0) |
| Open Claims | 295,535 | 803,838 | 256.4 | 861.3 | 2,775 | 12,461 | 10.8 (10.4 - 11.2) | 14.5 (14.2 - 14.7) |
| OEHR | 18,979 | 71,259 | 6.1 | 39.6 | 70 | 681 | 11.5 (8.98 - 14.6) | 17.2 (15.9 - 18.6) |
| VA | 11,119 | 99,994 | 6.7 | 91.5 | 52 | 926 | 7.74 (5.78 - 10.1) | 10.1 (9.48 - 10.8) |
| SIDIAP | 4,634 | 2,478 | 6.3 | 5.7 | 55 | 62 | 8.72 (6.57 - 11.3) | 10.8 (8.28 - 13.8) |
| Meta-analysis | 353,006 | 965,214 | 291.7 | 978.3 | 3,352 | 14,662 | 11.5 (11.1 - 11.9) | 15.0 (14.7 - 15.2) |
| Meta-analysis (w/o USOC) | 57,471 | 161,376 | 35.4 | 117.0 | 577 | 2,201 | 16.3 (15.0 - 17.7) | 18.8 (18.0 - 19.6) |
| **PS-matching / on-treatment TAR without escalation** | | | | | | | | |
| CCAE | 19,098 | 43,539 | 13.9 | 31.2 | 212 | 485 | 15.2 (13.2 - 17.4) | 15.6 (14.2 - 17.0) |
| DAG | 4,918 | 876 | 3.7 | 0.8 | 52 | 16 | 14.0 (10.5 - 18.4) | 21.0 (12.0 - 34.1) |
| ODOD | 14,591 | 45,502 | 9.1 | 36.5 | 174 | 838 | 19.1 (16.4 - 22.2) | 23.0 (21.4 - 24.6) |
| MDCR | 1,640 | 7,527 | 1.2 | 6.4 | 28 | 175 | 22.7 (15.1 - 32.8) | 27.4 (23.5 - 31.8) |
| MDCD | 1,290 | 6,310 | 0.7 | 3.6 | 31 | 115 | 47.4 (32.2 - 67.2) | 31.7 (26.2 - 38.1) |
| Open Claims | 271,263 | 798,298 | 204.4 | 727.4 | 2,196 | 10,360 | 10.7 (10.3 - 11.2) | 14.2 (14.0 - 14.5) |
| OEHR | 17,159 | 68,847 | 4.8 | 33.1 | 57 | 539 | 11.9 (9.01 - 15.4) | 16.3 (15.0 - 17.7) |
| VA | 0 | 0 | 0.0 | 0.0 | 0 | 0 | | |
| SIDIAP | 4,092 | 1,110 | 5.2 | 1.8 | 46 | 19 | 8.79 (6.43 - 11.7) | 10.7 (6.43 - 16.7) |
| Meta-analysis | 334,051 | 972,009 | 243.1 | 840.7 | 2,796 | 12,547 | 11.5 (11.1 - 11.9) | 14.9 (14.7 - 15.2) |
| Meta-analysis (w/o USOC) | 62,788 | 173,711 | 38.7 | 113.3 | 600 | 2,187 | 15.5 (14.3 - 16.8) | 19.3 (18.5 - 20.1) |
| **PS-stratification / on-treatment TAR without escalation** | | | | | | | | |
| CCAE | 22,298 | 44,128 | 15.9 | 31.5 | 254 | 492 | 16.0 (14.1 - 18.0) | 15.6 (14.3 - 17.1) |
| DAG | 5,113 | 1,271 | 3.8 | 1.1 | 52 | 26 | 13.6 (10.1 - 17.8) | 22.8 (14.9 - 33.4) |
| ODOD | 16,194 | 45,989 | 9.8 | 36.9 | 186 | 853 | 18.9 (16.3 - 21.8) | 23.1 (21.6 - 24.7) |
| MDCR | 1,967 | 7,745 | 1.4 | 6.5 | 32 | 183 | 23.1 (15.8 - 32.7) | 28.0 (24.1 - 32.4) |
| MDCD | 1,321 | 6,508 | 0.7 | 3.8 | 31 | 123 | 46.8 (31.8 - 66.4) | 32.7 (27.1 - 39.0) |
| Open Claims | 295,535 | 803,838 | 218.3 | 732.3 | 2,332 | 10,533 | 10.7 (10.3 - 11.1) | 14.4 (14.1 - 14.7) |
| OEHR | 18,979 | 71,259 | 5.3 | 34.4 | 62 | 577 | 11.6 (8.90 - 14.9) | 16.8 (15.4 - 18.2) |
| VA | 11,119 | 99,994 | 5.8 | 81.2 | 46 | 816 | 7.89 (5.78 - 10.5) | 10.1 (9.38 - 10.8) |
| SIDIAP | 4,634 | 2,478 | 5.6 | 4.9 | 50 | 55 | 8.89 (6.60 - 11.7) | 11.1 (8.39 - 14.5) |
| Meta-analysis | 353,006 | 965,214 | 249.4 | 835.1 | 2,834 | 12,455 | 11.4 (10.9 - 11.8) | 14.9 (14.7 - 15.2) |
| Meta-analysis (w/o USOC) | 57,471 | 161,376 | 31.1 | 102.8 | 502 | 1,922 | 16.1 (14.8 - 17.6) | 18.7 (17.9 - 19.6) |

##### 4.3.14 Diabetic ketoacidosis

**Supplementary Table 355:** Exposures, outcomes and risk estimates for Diabetic ketoacidosis in SGLT2I (male) (T) vs SU (male) (C) new-user cohorts under four different propensity score (PS) adjustment / time-at-risk (TAR) designs across data sources. We report total patient exposure time in 1,000 patient-year (KYr) units, uncalibrated and calibrated hazard ratios (HRs) and their 95% confidence intervals (CIs). Greyed out entries did not pass study diagnostics and are included only for completeness.

| | Patients | | Exposure (KYr) | | Outcomes | | Incidence (per KYr) | |
|---|---|---|---|---|---|---|---|---|
| | T | C | T | C | T | C | T | C |
| **PS-matching / on-treatment TAR** | | | | | | | | |
| CCAE | 20,611 | 46,816 | 17.6 | 39.2 | 38 | 28 | 2.16 (1.53 - 2.96) | 0.71 (0.47 - 1.03) |
| DAG | 5,471 | 973 | 4.6 | 0.9 | 0 | 0 | | |
| ODOD | 15,705 | 48,826 | 11.2 | 44.5 | 22 | 65 | 1.96 (1.23 - 2.97) | 1.46 (1.13 - 1.86) |
| MDCR | 1,776 | 8,308 | 1.5 | 7.8 | 0 | 11 | | 1.40 (0.70 - 2.51) |
| MDCD | 1,456 | 7,196 | 0.9 | 5.0 | <5 | 30 | < 5.66 (1.84 - 13.2) | 5.95 (4.01 - 8.49) |
| Open Claims | 304,375 | 886,843 | 271.2 | 960.3 | 287 | 771 | 1.06 (0.94 - 1.19) | 0.80 (0.75 - 0.86) |
| OEHR | 18,273 | 73,770 | 5.8 | 41.4 | 12 | 50 | 2.07 (1.07 - 3.61) | 1.21 (0.90 - 1.59) |
| VA | 0 | 0 | 0.0 | 0.0 | 0 | 0 | | |
| SIDIAP | 4,555 | 1,220 | 6.6 | 2.2 | 0 | <5 | | < 2.23 (0.72 - 5.20) |
| Meta-analysis | 360,420 | 1,063,451 | 306.7 | 1090.5 | <364 | 944 | < 1.19 (1.07 - 1.32) | 0.87 (0.81 - 0.92) |
| Meta-analysis (w/o USOC) | 56,045 | 176,608 | 35.5 | 130.1 | <77 | 173 | < 2.17 (1.71 - 2.71) | 1.33 (1.14 - 1.54) |
| **PS-stratification / on-treatment TAR** | | | | | | | | |
| CCAE | 24,108 | 47,460 | 20.1 | 39.6 | 40 | 29 | 1.99 (1.42 - 2.71) | 0.73 (0.49 - 1.05) |
| DAG | 5,702 | 1,403 | 4.7 | 1.4 | 0 | 0 | | |
| ODOD | 17,459 | 49,485 | 12.1 | 45.1 | 23 | 66 | 1.90 (1.20 - 2.85) | 1.46 (1.13 - 1.86) |
| MDCR | 2,135 | 8,594 | 1.7 | 8.1 | <5 | 11 | < 2.93 (0.95 - 6.84) | 1.36 (0.68 - 2.44) |
| MDCD | 1,499 | 7,439 | 0.9 | 5.2 | <5 | 32 | < 5.58 (1.81 - 13.0) | 6.11 (4.18 - 8.63) |
| Open Claims | 332,197 | 892,383 | 289.1 | 966.1 | 307 | 783 | 1.06 (0.95 - 1.19) | 0.81 (0.75 - 0.87) |
| OEHR | 20,218 | 76,232 | 6.5 | 43.1 | 14 | 52 | 2.17 (1.19 - 3.64) | 1.21 (0.90 - 1.58) |
| VA | 12,523 | 111,064 | 7.5 | 102.3 | <10 | 58 | < 1.33 (0.64 - 2.44) | 0.57 (0.43 - 0.73) |
| SIDIAP | 5,168 | 2,692 | 7.1 | 6.3 | 0 | <5 | | < 0.80 (0.26 - 1.86) |
| Meta-analysis | 393,982 | 1,065,560 | 327.8 | 1093.9 | 384 | 930 | 1.17 (1.06 - 1.29) | 0.85 (0.80 - 0.91) |
| Meta-analysis (w/o USOC) | 61,785 | 173,177 | 38.7 | 127.8 | 77 | 147 | 1.99 (1.57 - 2.49) | 1.15 (0.97 - 1.35) |
| **PS-matching / on-treatment TAR without escalation** | | | | | | | | |
| CCAE | 20,611 | 46,816 | 15.3 | 34.0 | 31 | 21 | 2.02 (1.37 - 2.87) | 0.62 (0.38 - 0.95) |
| DAG | 5,471 | 973 | 4.2 | 0.8 | 0 | 0 | | |
| ODOD | 15,705 | 48,826 | 9.9 | 39.8 | 19 | 50 | 1.91 (1.15 - 2.99) | 1.26 (0.93 - 1.65) |
| MDCR | 1,776 | 8,308 | 1.4 | 7.2 | 0 | 9 | | 1.25 (0.57 - 2.38) |
| MDCD | 1,456 | 7,196 | 0.7 | 4.2 | <5 | 23 | < 6.68 (2.17 - 15.6) | 5.50 (3.49 - 8.25) |
| Open Claims | 304,375 | 886,843 | 229.9 | 814.7 | 227 | 594 | 0.99 (0.86 - 1.12) | 0.73 (0.67 - 0.79) |
| OEHR | 18,273 | 73,770 | 5.1 | 35.9 | 11 | 29 | 2.16 (1.08 - 3.87) | 0.81 (0.54 - 1.16) |
| VA | 0 | 0 | 0.0 | 0.0 | 0 | 0 | | |
| SIDIAP | 4,555 | 1,220 | 5.9 | 2.0 | 0 | <5 | | < 2.56 (0.83 - 5.97) |
| Meta-analysis | 358,964 | 1,056,255 | 260.2 | 924.3 | 288 | 694 | 1.11 (0.98 - 1.24) | 0.75 (0.70 - 0.81) |
| Meta-analysis (w/o USOC) | 54,589 | 169,412 | 30.4 | 109.7 | 61 | 100 | 2.01 (1.54 - 2.58) | 0.91 (0.74 - 1.11) |
| **PS-stratification / on-treatment TAR without escalation** | | | | | | | | |
| CCAE | 24,108 | 47,460 | 17.5 | 34.3 | 33 | 22 | 1.88 (1.29 - 2.64) | 0.64 (0.40 - 0.97) |
| DAG | 5,702 | 1,403 | 4.3 | 1.3 | 0 | 0 | | |
| ODOD | 17,459 | 49,485 | 10.8 | 40.4 | 20 | 51 | 1.86 (1.14 - 2.87) | 1.26 (0.94 - 1.66) |
| MDCR | 2,135 | 8,594 | 1.5 | 7.4 | 0 | 9 | | 1.22 (0.56 - 2.31) |
| MDCD | 1,499 | 7,439 | 0.8 | 4.3 | <5 | 25 | < 6.57 (2.13 - 15.3) | 5.75 (3.72 - 8.49) |
| Open Claims | 332,197 | 892,383 | 245.8 | 819.7 | 243 | 603 | 0.99 (0.87 - 1.12) | 0.74 (0.68 - 0.80) |
| OEHR | 20,218 | 76,232 | 5.7 | 37.3 | 13 | 30 | 2.29 (1.22 - 3.91) | 0.80 (0.54 - 1.15) |
| VA | 12,523 | 111,064 | 6.5 | 90.5 | <10 | 38 | < 1.53 (0.74 - 2.82) | 0.42 (0.30 - 0.58) |
| SIDIAP | 5,168 | 2,692 | 6.3 | 5.4 | 0 | <5 | | < 0.93 (0.30 - 2.18) |
| Meta-analysis | 393,982 | 1,065,560 | 279.8 | 931.7 | 309 | 706 | 1.10 (0.98 - 1.23) | 0.76 (0.70 - 0.82) |
| Meta-analysis (w/o USOC) | 61,785 | 173,177 | 34.0 | 112.0 | 66 | 103 | 1.94 (1.50 - 2.47) | 0.92 (0.75 - 1.12) |

##### 4.3.15 Diarrhea

**Supplementary Table 356:** Exposures, outcomes and risk estimates for Diarrhea in SGLT2I (male) (T) vs SU (male) (C) new-user cohorts under four different propensity score (PS) adjustment / time-at-risk (TAR) designs across data sources. We report total patient exposure time in 1,000 patient-year (KYr) units, uncalibrated and calibrated hazard ratios (HRs) and their 95% confidence intervals (CIs). Greyed out entries did not pass study diagnostics and are included only for completeness.

| | Patients | | Exposure (KYr) | | Outcomes | | Incidence (per KYr) | |
|---|---|---|---|---|---|---|---|---|
| | T | C | T | C | T | C | T | C |
| **PS-matching / on-treatment TAR** | | | | | | | | |
| CCAE | 18,695 | 42,551 | 15.8 | 35.0 | 321 | 823 | 20.4 (18.2 - 22.7) | 23.5 (21.9 - 25.1) |
| DAG | 5,370 | 960 | 4.5 | 0.9 | 6 | <5 | 1.34 (0.49 - 2.91) | < 5.58 (1.81 - 13.0) |
| ODOD | 14,057 | 43,197 | 9.9 | 38.3 | 265 | 1,228 | 26.8 (23.7 - 30.3) | 32.1 (30.3 - 33.9) |
| MDCR | 1,592 | 7,280 | 1.3 | 6.7 | 27 | 198 | 20.1 (13.2 - 29.2) | 29.5 (25.5 - 33.9) |
| MDCD | 1,250 | 6,122 | 0.7 | 4.2 | 34 | 190 | 45.6 (31.6 - 63.8) | 45.5 (39.3 - 52.4) |
| Open Claims | 274,570 | 806,193 | 243.1 | 861.5 | 2,783 | 12,671 | 11.4 (11.0 - 11.9) | 14.7 (14.5 - 15.0) |
| OEHR | 16,714 | 67,327 | 5.2 | 36.4 | 122 | 1,112 | 23.3 (19.3 - 27.8) | 30.5 (28.8 - 32.4) |
| VA | 0 | 0 | 0.0 | 0.0 | 0 | 0 | | |
| SIDIAP | 4,396 | 1,173 | 6.4 | 2.1 | 39 | 20 | 6.10 (4.34 - 8.33) | 9.36 (5.71 - 14.4) |
| Meta-analysis | 331,274 | 973,843 | 282.4 | 984.2 | 3,591 | 16,242 | 12.7 (12.3 - 13.1) | 16.5 (16.2 - 16.8) |
| Meta-analysis (w/o USOC) | 56,704 | 167,650 | 39.4 | 122.7 | 808 | 3,571 | 20.5 (19.1 - 22.0) | 29.1 (28.1 - 30.1) |
| **PS-stratification / on-treatment TAR** | | | | | | | | |
| CCAE | 21,758 | 43,117 | 17.9 | 35.4 | 367 | 829 | 20.6 (18.5 - 22.8) | 23.4 (21.9 - 25.1) |
| DAG | 5,600 | 1,385 | 4.6 | 1.4 | 6 | <5 | 1.30 (0.48 - 2.82) | < 3.66 (1.19 - 8.55) |
| ODOD | 15,599 | 44,007 | 10.6 | 39.1 | 286 | 1,274 | 26.9 (23.8 - 30.2) | 32.6 (30.9 - 34.5) |
| MDCR | 1,906 | 7,505 | 1.5 | 6.9 | 30 | 207 | 20.1 (13.5 - 28.7) | 30.1 (26.1 - 34.5) |
| MDCD | 1,283 | 6,366 | 0.8 | 4.3 | 34 | 198 | 45.0 (31.2 - 62.9) | 45.5 (39.4 - 52.4) |
| Open Claims | 299,156 | 811,534 | 258.9 | 866.9 | 3,000 | 12,838 | 11.6 (11.2 - 12.0) | 14.8 (14.6 - 15.1) |
| OEHR | 18,506 | 69,602 | 5.8 | 37.8 | 134 | 1,196 | 22.9 (19.2 - 27.1) | 31.6 (29.9 - 33.5) |
| VA | 10,635 | 96,741 | 6.4 | 87.3 | 107 | 1,717 | 16.7 (13.7 - 20.2) | 19.7 (18.7 - 20.6) |
| SIDIAP | 4,981 | 2,590 | 6.8 | 6.0 | 46 | 47 | 6.73 (4.93 - 8.98) | 7.87 (5.78 - 10.5) |
| Meta-analysis | 355,019 | 968,260 | 293.2 | 979.1 | 3,787 | 16,137 | 12.9 (12.5 - 13.3) | 16.5 (16.2 - 16.7) |
| Meta-analysis (w/o USOC) | 55,863 | 156,726 | 34.3 | 112.2 | 787 | 3,299 | 22.9 (21.3 - 24.6) | 29.4 (28.4 - 30.4) |
| **PS-matching / on-treatment TAR without escalation** | | | | | | | | |
| CCAE | 18,695 | 42,551 | 13.8 | 30.5 | 266 | 709 | 19.3 (17.0 - 21.8) | 23.2 (21.6 - 25.0) |
| DAG | 5,370 | 960 | 4.1 | 0.8 | 6 | <5 | 1.45 (0.53 - 3.16) | < 6.07 (1.97 - 14.2) |
| ODOD | 14,057 | 43,197 | 8.8 | 34.4 | 225 | 1,088 | 25.6 (22.3 - 29.1) | 31.6 (29.8 - 33.6) |
| MDCR | 1,592 | 7,280 | 1.2 | 6.2 | 24 | 185 | 19.9 (12.7 - 29.6) | 29.9 (25.8 - 34.6) |
| MDCD | 1,250 | 6,122 | 0.6 | 3.5 | 29 | 158 | 45.5 (30.5 - 65.3) | 44.8 (38.1 - 52.3) |
| Open Claims | 274,570 | 806,193 | 206.8 | 734.1 | 2,284 | 10,660 | 11.0 (10.6 - 11.5) | 14.5 (14.2 - 14.8) |
| OEHR | 16,714 | 67,327 | 4.6 | 31.9 | 106 | 881 | 23.0 (18.9 - 27.9) | 27.6 (25.8 - 29.5) |
| VA | 0 | 0 | 0.0 | 0.0 | 0 | 0 | | |
| SIDIAP | 4,396 | 1,173 | 5.7 | 1.9 | 34 | 17 | 5.99 (4.15 - 8.37) | 9.14 (5.32 - 14.6) |
| Meta-analysis | 331,274 | 973,843 | 241.6 | 842.4 | 2,968 | 13,698 | 12.3 (11.8 - 12.7) | 16.3 (16.0 - 16.5) |
| Meta-analysis (w/o USOC) | 56,704 | 167,650 | 34.7 | 108.4 | 684 | 3,038 | 19.7 (18.3 - 21.2) | 28.0 (27.0 - 29.0) |
| **PS-stratification / on-treatment TAR without escalation** | | | | | | | | |
| CCAE | 21,758 | 43,117 | 15.7 | 30.8 | 307 | 715 | 19.6 (17.5 - 21.9) | 23.2 (21.5 - 25.0) |
| DAG | 5,600 | 1,385 | 4.3 | 1.2 | 6 | <5 | 1.41 (0.52 - 3.07) | < 4.02 (1.30 - 9.38) |
| ODOD | 15,599 | 44,007 | 9.5 | 35.1 | 244 | 1,130 | 25.7 (22.6 - 29.1) | 32.2 (30.4 - 34.1) |
| MDCR | 1,906 | 7,505 | 1.3 | 6.3 | 27 | 194 | 20.0 (13.2 - 29.2) | 30.6 (26.4 - 35.2) |
| MDCD | 1,283 | 6,366 | 0.6 | 3.7 | 29 | 166 | 44.9 (30.1 - 64.5) | 45.2 (38.6 - 52.6) |
| Open Claims | 299,156 | 811,534 | 221.0 | 738.8 | 2,473 | 10,811 | 11.2 (10.8 - 11.6) | 14.6 (14.4 - 14.9) |
| OEHR | 18,506 | 69,602 | 5.2 | 33.1 | 117 | 945 | 22.7 (18.8 - 27.2) | 28.6 (26.8 - 30.4) |
| VA | 10,635 | 96,741 | 5.5 | 77.8 | 95 | 1,496 | 17.2 (13.9 - 21.0) | 19.2 (18.3 - 20.2) |
| SIDIAP | 4,981 | 2,590 | 6.1 | 5.1 | 41 | 41 | 6.74 (4.83 - 9.14) | 8.01 (5.75 - 10.9) |
| Meta-analysis | 355,019 | 968,260 | 251.3 | 837.8 | 3,141 | 13,601 | 12.5 (12.1 - 12.9) | 16.2 (16.0 - 16.5) |
| Meta-analysis (w/o USOC) | 55,863 | 156,726 | 30.3 | 99.0 | 668 | 2,790 | 22.0 (20.4 - 23.8) | 28.2 (27.2 - 29.3) |

##### 4.3.16 Genitourinary infection

**Supplementary Table 357:** Exposures, outcomes and risk estimates for Genitourinary infection in SGLT2I (male) (T) vs SU (male) (C) new-user cohorts under four different propensity score (PS) adjustment / time-at-risk (TAR) designs across data sources. We report total patient exposure time in 1,000 patient-year (KYr) units, uncalibrated and calibrated hazard ratios (HRs) and their 95% confidence intervals (CIs). Greyed out entries did not pass study diagnostics and are included only for completeness.

| | Patients | | Exposure (KYr) | | Outcomes | | Incidence (per KYr) | |
|---|---|---|---|---|---|---|---|---|
| | T | C | T | C | T | C | T | C |
| **PS-matching / on-treatment TAR** | | | | | | | | |
| CCAE | 18,933 | 42,878 | 16.0 | 35.3 | 242 | 712 | 15.1 (13.2 - 17.1) | 20.2 (18.7 - 21.7) |
| DAG | 4,678 | 844 | 3.9 | 0.8 | 93 | 16 | 24.1 (19.4 - 29.5) | 19.8 (11.3 - 32.1) |
| ODOD | 14,139 | 42,339 | 10.0 | 37.3 | 266 | 1,516 | 26.6 (23.5 - 30.0) | 40.7 (38.7 - 42.8) |
| MDCR | 1,575 | 7,087 | 1.3 | 6.5 | 37 | 287 | 28.1 (19.8 - 38.8) | 44.1 (39.1 - 49.5) |
| MDCD | 1,333 | 6,465 | 0.8 | 4.4 | 15 | 126 | 18.5 (10.3 - 30.4) | 28.6 (23.8 - 34.0) |
| Open Claims | 272,767 | 789,830 | 244.0 | 841.1 | 2,747 | 16,133 | 11.3 (10.8 - 11.7) | 19.2 (18.9 - 19.5) |
| OEHR | 17,285 | 69,322 | 5.5 | 38.0 | 97 | 895 | 17.8 (14.4 - 21.7) | 23.6 (22.1 - 25.2) |
| VA | 0 | 0 | 0.0 | 0.0 | 0 | 0 | | |
| SIDIAP | 3,966 | 1,104 | 5.8 | 2.0 | 123 | 39 | 21.1 (17.5 - 25.1) | 19.7 (14.0 - 27.0) |
| Meta-analysis | 334,676 | 959,869 | 287.3 | 965.3 | 3,620 | 19,724 | 12.6 (12.2 - 13.0) | 20.4 (20.1 - 20.7) |
| Meta-analysis (w/o USOC) | 61,909 | 170,039 | 43.3 | 124.2 | 873 | 3,591 | 20.2 (18.8 - 21.5) | 28.9 (28.0 - 29.9) |
| **PS-stratification / on-treatment TAR** | | | | | | | | |
| CCAE | 22,104 | 43,443 | 18.3 | 35.6 | 274 | 724 | 15.0 (13.3 - 16.9) | 20.3 (18.9 - 21.9) |
| DAG | 4,928 | 1,221 | 4.0 | 1.2 | 96 | 20 | 23.9 (19.3 - 29.1) | 16.8 (10.3 - 26.0) |
| ODOD | 15,706 | 42,920 | 10.8 | 37.8 | 287 | 1,564 | 26.6 (23.6 - 29.8) | 41.4 (39.3 - 43.5) |
| MDCR | 1,870 | 7,208 | 1.5 | 6.6 | 43 | 293 | 29.5 (21.3 - 39.7) | 44.3 (39.4 - 49.7) |
| MDCD | 1,377 | 6,646 | 0.8 | 4.6 | 15 | 131 | 18.2 (10.2 - 30.0) | 28.7 (24.0 - 34.1) |
| Open Claims | 297,034 | 794,829 | 259.6 | 846.1 | 2,993 | 16,427 | 11.5 (11.1 - 11.9) | 19.4 (19.1 - 19.7) |
| OEHR | 19,171 | 71,370 | 6.1 | 39.3 | 109 | 961 | 17.9 (14.7 - 21.6) | 24.5 (22.9 - 26.0) |
| VA | 11,310 | 99,225 | 6.8 | 89.8 | 68 | 1,456 | 9.93 (7.71 - 12.6) | 16.2 (15.4 - 17.1) |
| SIDIAP | 4,522 | 2,420 | 6.3 | 5.5 | 131 | 111 | 20.9 (17.5 - 24.8) | 20.3 (16.7 - 24.4) |
| Meta-analysis | 354,015 | 952,562 | 294.8 | 958.8 | 3,663 | 19,676 | 12.4 (12.0 - 12.8) | 20.5 (20.2 - 20.8) |
| Meta-analysis (w/o USOC) | 56,981 | 157,733 | 35.1 | 112.7 | 670 | 3,249 | 19.1 (17.7 - 20.6) | 28.8 (27.8 - 29.8) |
| **PS-matching / on-treatment TAR without escalation** | | | | | | | | |
| CCAE | 18,933 | 42,878 | 14.0 | 30.6 | 205 | 627 | 14.6 (12.7 - 16.8) | 20.5 (18.9 - 22.1) |
| DAG | 4,678 | 844 | 3.6 | 0.7 | 86 | 15 | 24.2 (19.3 - 29.8) | 20.3 (11.4 - 33.5) |
| ODOD | 14,139 | 42,339 | 8.9 | 33.4 | 228 | 1,374 | 25.7 (22.5 - 29.3) | 41.2 (39.0 - 43.4) |
| MDCR | 1,575 | 7,087 | 1.2 | 6.0 | 34 | 263 | 28.8 (19.9 - 40.2) | 44.1 (38.9 - 49.7) |
| MDCD | 1,333 | 6,465 | 0.7 | 3.7 | 12 | 104 | 17.5 (9.06 - 30.6) | 28.2 (23.1 - 34.2) |
| Open Claims | 272,767 | 789,830 | 206.9 | 714.5 | 2,327 | 13,847 | 11.2 (10.8 - 11.7) | 19.4 (19.1 - 19.7) |
| OEHR | 17,285 | 69,322 | 4.8 | 33.1 | 84 | 712 | 17.6 (14.0 - 21.7) | 21.5 (20.0 - 23.1) |
| VA | 0 | 0 | 0.0 | 0.0 | 0 | 0 | | |
| SIDIAP | 3,966 | 1,104 | 5.2 | 1.7 | 107 | 35 | 20.6 (16.9 - 24.9) | 20.2 (14.1 - 28.1) |
| Meta-analysis | 334,676 | 959,869 | 245.2 | 823.8 | 3,083 | 16,977 | 12.6 (12.1 - 13.0) | 20.6 (20.3 - 20.9) |
| Meta-analysis (w/o USOC) | 61,909 | 170,039 | 38.3 | 109.2 | 756 | 3,130 | 19.7 (18.4 - 21.2) | 28.7 (27.7 - 29.7) |
| **PS-stratification / on-treatment TAR without escalation** | | | | | | | | |
| CCAE | 22,104 | 43,443 | 16.0 | 30.9 | 233 | 639 | 14.6 (12.8 - 16.6) | 20.7 (19.1 - 22.3) |
| DAG | 4,928 | 1,221 | 3.7 | 1.1 | 88 | 19 | 23.7 (19.0 - 29.3) | 17.6 (10.6 - 27.5) |
| ODOD | 15,706 | 42,920 | 9.6 | 33.9 | 248 | 1,417 | 25.8 (22.7 - 29.3) | 41.9 (39.7 - 44.1) |
| MDCR | 1,870 | 7,208 | 1.3 | 6.1 | 40 | 269 | 30.4 (21.7 - 41.4) | 44.4 (39.2 - 50.0) |
| MDCD | 1,377 | 6,646 | 0.7 | 3.8 | 12 | 108 | 17.2 (8.91 - 30.1) | 28.3 (23.2 - 34.2) |
| Open Claims | 297,034 | 794,829 | 220.9 | 718.9 | 2,550 | 14,105 | 11.5 (11.1 - 12.0) | 19.6 (19.3 - 19.9) |
| OEHR | 19,171 | 71,370 | 5.4 | 34.2 | 95 | 758 | 17.7 (14.3 - 21.7) | 22.1 (20.6 - 23.8) |
| VA | 11,310 | 99,225 | 5.9 | 79.6 | 59 | 1,250 | 9.97 (7.59 - 12.9) | 15.7 (14.9 - 16.6) |
| SIDIAP | 4,522 | 2,420 | 5.6 | 4.7 | 115 | 97 | 20.6 (17.0 - 24.7) | 20.6 (16.7 - 25.2) |
| Meta-analysis | 354,015 | 952,562 | 251.9 | 818.0 | 3,126 | 16,919 | 12.4 (12.0 - 12.9) | 20.7 (20.4 - 21.0) |
| Meta-analysis (w/o USOC) | 56,981 | 157,733 | 30.9 | 99.0 | 576 | 2,814 | 18.6 (17.1 - 20.2) | 28.4 (27.4 - 29.5) |

##### 4.3.17 Hyperkalemia

**Supplementary Table 358:** Exposures, outcomes and risk estimates for Hyperkalemia in SGLT2I (male) (T) vs SU (male) (C) new-user cohorts under four different propensity score (PS) adjustment / time-at-risk (TAR) designs across data sources. We report total patient exposure time in 1,000 patient-year (KYr) units, uncalibrated and calibrated hazard ratios (HRs) and their 95% confidence intervals (CIs). Greyed out entries did not pass study diagnostics and are included only for completeness.

| | Patients | | Exposure (KYr) | | Outcomes | | Incidence (per KYr) | |
|---|---|---|---|---|---|---|---|---|
| | T | C | T | C | T | C | T | C |
| **PS-matching / on-treatment TAR** | | | | | | | | |
| CCAE | 20,266 | 46,044 | 17.2 | 38.2 | 115 | 367 | 6.70 (5.53 - 8.04) | 9.60 (8.64 - 10.6) |
| DAG | 5,131 | 910 | 4.3 | 0.9 | 50 | 12 | 11.7 (8.71 - 15.5) | 14.0 (7.23 - 24.4) |
| ODOD | 15,150 | 46,307 | 10.7 | 41.1 | 178 | 1,101 | 16.6 (14.2 - 19.2) | 26.8 (25.2 - 28.4) |
| MDCR | 1,729 | 7,998 | 1.5 | 7.5 | 17 | 172 | 11.4 (6.62 - 18.2) | 23.1 (19.8 - 26.8) |
| MDCD | 1,418 | 6,953 | 0.9 | 4.8 | 15 | 112 | 17.6 (9.84 - 29.0) | 23.2 (19.1 - 27.9) |
| Open Claims | 299,090 | 868,896 | 266.1 | 931.6 | 1,170 | 7,952 | 4.40 (4.15 - 4.66) | 8.54 (8.35 - 8.73) |
| OEHR | 17,953 | 72,312 | 5.7 | 39.9 | 57 | 598 | 10.1 (7.64 - 13.1) | 15.0 (13.8 - 16.2) |
| VA | 0 | 0 | 0.0 | 0.0 | 0 | 0 | | |
| SIDIAP | 4,355 | 1,181 | 6.3 | 2.2 | 68 | 18 | 10.8 (8.38 - 13.7) | 8.32 (4.93 - 13.1) |
| Meta-analysis | 365,092 | 1,050,601 | 312.6 | 1066.1 | 1,670 | 10,332 | 5.34 (5.09 - 5.60) | 9.69 (9.51 - 9.88) |
| Meta-analysis (w/o USOC) | 66,002 | 181,705 | 46.5 | 134.5 | 500 | 2,380 | 10.8 (9.84 - 11.7) | 17.7 (17.0 - 18.4) |
| **PS-stratification / on-treatment TAR** | | | | | | | | |
| CCAE | 23,691 | 46,675 | 19.6 | 38.6 | 134 | 371 | 6.84 (5.73 - 8.10) | 9.61 (8.66 - 10.6) |
| DAG | 5,339 | 1,317 | 4.4 | 1.3 | 52 | 14 | 11.8 (8.83 - 15.5) | 10.8 (5.88 - 18.1) |
| ODOD | 16,805 | 46,980 | 11.6 | 41.7 | 192 | 1,138 | 16.6 (14.3 - 19.1) | 27.3 (25.7 - 28.9) |
| MDCR | 2,069 | 8,235 | 1.7 | 7.6 | 20 | 181 | 12.1 (7.36 - 18.6) | 23.7 (20.4 - 27.4) |
| MDCD | 1,462 | 7,170 | 0.9 | 5.0 | 15 | 117 | 17.3 (9.69 - 28.6) | 23.4 (19.4 - 28.1) |
| Open Claims | 325,992 | 874,103 | 283.5 | 937.0 | 1,288 | 8,117 | 4.54 (4.30 - 4.80) | 8.66 (8.48 - 8.85) |
| OEHR | 19,863 | 74,522 | 6.3 | 41.4 | 62 | 645 | 9.85 (7.55 - 12.6) | 15.6 (14.4 - 16.9) |
| VA | 11,422 | 104,088 | 6.9 | 94.0 | 104 | 1,465 | 15.2 (12.4 - 18.4) | 15.6 (14.8 - 16.4) |
| SIDIAP | 4,910 | 2,594 | 6.7 | 6.0 | 78 | 59 | 11.6 (9.19 - 14.5) | 9.83 (7.48 - 12.7) |
| Meta-analysis | 386,351 | 1,042,280 | 320.9 | 1058.7 | 1,676 | 10,271 | 5.22 (4.98 - 5.48) | 9.70 (9.51 - 9.89) |
| Meta-analysis (w/o USOC) | 60,359 | 168,177 | 37.5 | 121.7 | 388 | 2,154 | 10.4 (9.35 - 11.4) | 17.7 (17.0 - 18.5) |
| **PS-matching / on-treatment TAR without escalation** | | | | | | | | |
| CCAE | 20,266 | 46,044 | 15.0 | 33.2 | 93 | 316 | 6.20 (5.01 - 7.60) | 9.52 (8.50 - 10.6) |
| DAG | 5,131 | 910 | 3.9 | 0.8 | 45 | 12 | 11.5 (8.36 - 15.3) | 15.3 (7.90 - 26.7) |
| ODOD | 15,150 | 46,307 | 9.5 | 36.9 | 161 | 977 | 16.9 (14.4 - 19.7) | 26.5 (24.9 - 28.2) |
| MDCR | 1,729 | 7,998 | 1.3 | 6.8 | 17 | 159 | 12.8 (7.44 - 20.5) | 23.3 (19.8 - 27.2) |
| MDCD | 1,418 | 6,953 | 0.7 | 4.0 | 14 | 89 | 19.3 (10.6 - 32.4) | 22.2 (17.8 - 27.3) |
| Open Claims | 299,090 | 868,896 | 225.6 | 791.1 | 975 | 6,657 | 4.32 (4.05 - 4.60) | 8.41 (8.21 - 8.62) |
| OEHR | 17,953 | 72,312 | 5.0 | 34.7 | 43 | 449 | 8.66 (6.27 - 11.7) | 12.9 (11.8 - 14.2) |
| VA | 0 | 0 | 0.0 | 0.0 | 0 | 0 | | |
| SIDIAP | 4,355 | 1,181 | 5.6 | 1.9 | 53 | 14 | 9.45 (7.08 - 12.4) | 7.44 (4.07 - 12.5) |
| Meta-analysis | 365,092 | 1,050,601 | 266.7 | 909.4 | 1,401 | 8,673 | 5.25 (4.98 - 5.54) | 9.54 (9.34 - 9.74) |
| Meta-analysis (w/o USOC) | 66,002 | 181,705 | 41.1 | 118.3 | 426 | 2,016 | 10.4 (9.41 - 11.4) | 17.0 (16.3 - 17.8) |
| **PS-stratification / on-treatment TAR without escalation** | | | | | | | | |
| CCAE | 23,691 | 46,675 | 17.1 | 33.5 | 109 | 320 | 6.36 (5.22 - 7.67) | 9.55 (8.53 - 10.7) |
| DAG | 5,339 | 1,317 | 4.0 | 1.2 | 46 | 14 | 11.4 (8.32 - 15.2) | 11.9 (6.48 - 19.9) |
| ODOD | 16,805 | 46,980 | 10.3 | 37.4 | 173 | 1,008 | 16.8 (14.4 - 19.5) | 27.0 (25.3 - 28.7) |
| MDCR | 2,069 | 8,235 | 1.5 | 7.0 | 19 | 168 | 12.8 (7.72 - 20.0) | 24.0 (20.5 - 27.9) |
| MDCD | 1,462 | 7,170 | 0.7 | 4.2 | 14 | 92 | 19.0 (10.4 - 31.9) | 22.1 (17.8 - 27.1) |
| Open Claims | 325,992 | 874,103 | 241.1 | 795.8 | 1,086 | 6,803 | 4.50 (4.24 - 4.78) | 8.55 (8.35 - 8.75) |
| OEHR | 19,863 | 74,522 | 5.6 | 36.0 | 47 | 484 | 8.46 (6.22 - 11.3) | 13.5 (12.3 - 14.7) |
| VA | 11,422 | 104,088 | 6.0 | 83.4 | 89 | 1,239 | 15.0 (12.0 - 18.4) | 14.9 (14.0 - 15.7) |
| SIDIAP | 4,910 | 2,594 | 6.0 | 5.1 | 62 | 48 | 10.4 (7.94 - 13.3) | 9.34 (6.89 - 12.4) |
| Meta-analysis | 386,351 | 1,042,280 | 274.1 | 902.7 | 1,415 | 8,615 | 5.16 (4.90 - 5.44) | 9.54 (9.34 - 9.75) |
| Meta-analysis (w/o USOC) | 60,359 | 168,177 | 33.0 | 106.9 | 329 | 1,812 | 9.97 (8.92 - 11.1) | 17.0 (16.2 - 17.8) |

##### 4.3.18 Hypoglycemia

**Supplementary Table 359:** Exposures, outcomes and risk estimates for Hypoglycemia in SGLT2I (male) (T) vs SU (male) (C) new-user cohorts under four different propensity score (PS) adjustment / time-at-risk (TAR) designs across data sources. We report total patient exposure time in 1,000 patient-year (KYr) units, uncalibrated and calibrated hazard ratios (HRs) and their 95% confidence intervals (CIs). Greyed out entries did not pass study diagnostics and are included only for completeness.

| | Patients | | Exposure (KYr) | | Outcomes | | Incidence (per KYr) | |
|---|---|---|---|---|---|---|---|---|
| | T | C | T | C | T | C | T | C |
| **PS-matching / on-treatment TAR** | | | | | | | | |
| CCAE | 20,386 | 46,405 | 17.4 | 38.7 | 37 | 361 | 2.13 (1.50 - 2.93) | 9.34 (8.40 - 10.4) |
| DAG | 5,443 | 969 | 4.5 | 0.9 | 5 | <5 | 1.10 (0.36 - 2.57) | < 5.49 (1.78 - 12.8) |
| ODOD | 15,530 | 48,361 | 11.1 | 43.7 | 46 | 888 | 4.15 (3.04 - 5.54) | 20.3 (19.0 - 21.7) |
| MDCR | 1,756 | 8,229 | 1.5 | 7.7 | 5 | 149 | 3.32 (1.08 - 7.74) | 19.3 (16.4 - 22.7) |
| MDCD | 1,427 | 7,084 | 0.9 | 4.9 | 6 | 128 | 6.94 (2.55 - 15.1) | 26.0 (21.7 - 31.0) |
| Open Claims | 301,109 | 878,668 | 268.1 | 947.5 | 395 | 7,185 | 1.47 (1.33 - 1.63) | 7.58 (7.41 - 7.76) |
| OEHR | 18,137 | 73,263 | 5.8 | 40.8 | 15 | 616 | 2.60 (1.46 - 4.29) | 15.1 (13.9 - 16.4) |
| VA | 0 | 0 | 0.0 | 0.0 | 0 | 0 | | |
| SIDIAP | 4,547 | 1,220 | 6.6 | 2.2 | <5 | 10 | < 0.76 (0.25 - 1.77) | 4.48 (2.15 - 8.23) |
| Meta-analysis | 358,345 | 1,062,010 | 304.7 | 1083.2 | 504 | 9,327 | 1.65 (1.51 - 1.80) | 8.61 (8.44 - 8.79) |
| Meta-analysis (w/o USOC) | 57,236 | 183,342 | 36.6 | 135.7 | 109 | 2,142 | 2.98 (2.44 - 3.59) | 15.8 (15.1 - 16.5) |
| **PS-stratification / on-treatment TAR** | | | | | | | | |
| CCAE | 23,833 | 47,046 | 19.8 | 39.0 | 49 | 366 | 2.47 (1.83 - 3.27) | 9.38 (8.44 - 10.4) |
| DAG | 5,675 | 1,395 | 4.7 | 1.4 | 5 | 5 | 1.07 (0.35 - 2.49) | 3.63 (1.18 - 8.46) |
| ODOD | 17,261 | 48,998 | 12.0 | 44.3 | 51 | 915 | 4.26 (3.17 - 5.60) | 20.7 (19.3 - 22.0) |
| MDCR | 2,115 | 8,517 | 1.7 | 7.9 | 5 | 157 | 2.97 (0.96 - 6.92) | 19.8 (16.8 - 23.2) |
| MDCD | 1,468 | 7,319 | 0.9 | 5.1 | 6 | 136 | 6.84 (2.51 - 14.9) | 26.7 (22.4 - 31.5) |
| Open Claims | 328,462 | 883,945 | 285.7 | 953.0 | 421 | 7,333 | 1.47 (1.34 - 1.62) | 7.70 (7.52 - 7.87) |
| OEHR | 20,063 | 75,672 | 6.4 | 42.4 | 18 | 652 | 2.81 (1.66 - 4.43) | 15.4 (14.2 - 16.6) |
| VA | 12,356 | 110,156 | 7.4 | 100.9 | 14 | 1,038 | 1.88 (1.03 - 3.16) | 10.3 (9.67 - 10.9) |
| SIDIAP | 5,159 | 2,687 | 7.1 | 6.3 | 5 | 20 | 0.71 (0.23 - 1.65) | 3.20 (1.95 - 4.94) |
| Meta-analysis | 389,619 | 1,055,661 | 323.9 | 1078.7 | 539 | 9,266 | 1.66 (1.53 - 1.81) | 8.59 (8.42 - 8.77) |
| Meta-analysis (w/o USOC) | 61,157 | 171,716 | 38.2 | 125.7 | 118 | 1,933 | 3.09 (2.56 - 3.70) | 15.4 (14.7 - 16.1) |
| **PS-matching / on-treatment TAR without escalation** | | | | | | | | |
| CCAE | 20,386 | 46,405 | 15.2 | 33.5 | 27 | 303 | 1.78 (1.17 - 2.59) | 9.04 (8.05 - 10.1) |
| DAG | 5,443 | 969 | 4.2 | 0.8 | 5 | <5 | 1.20 (0.39 - 2.79) | < 5.98 (1.94 - 14.0) |
| ODOD | 15,530 | 48,361 | 9.8 | 39.1 | 32 | 765 | 3.26 (2.23 - 4.60) | 19.5 (18.2 - 21.0) |
| MDCR | 1,756 | 8,229 | 1.3 | 7.1 | <5 | 141 | < 3.71 (1.21 - 8.66) | 20.0 (16.8 - 23.5) |
| MDCD | 1,427 | 7,084 | 0.7 | 4.1 | <5 | 100 | < 6.87 (2.23 - 16.0) | 24.4 (19.9 - 29.7) |
| Open Claims | 301,109 | 878,668 | 227.3 | 804.4 | 269 | 5,922 | 1.18 (1.05 - 1.33) | 7.36 (7.18 - 7.55) |
| OEHR | 18,137 | 73,263 | 5.1 | 35.5 | 10 | 463 | 1.98 (0.95 - 3.64) | 13.1 (11.9 - 14.3) |
| VA | 0 | 0 | 0.0 | 0.0 | 0 | 0 | | |
| SIDIAP | 4,547 | 1,220 | 5.9 | 1.9 | <5 | 9 | < 0.85 (0.28 - 1.99) | 4.62 (2.11 - 8.77) |
| Meta-analysis | 358,345 | 1,062,010 | 259.4 | 923.7 | <348 | 7,694 | < 1.34 (1.20 - 1.49) | 8.33 (8.14 - 8.52) |
| Meta-analysis (w/o USOC) | 57,236 | 183,342 | 32.1 | 119.3 | <79 | 1,772 | < 2.46 (1.95 - 3.07) | 14.9 (14.2 - 15.6) |
| **PS-stratification / on-treatment TAR without escalation** | | | | | | | | |
| CCAE | 23,833 | 47,046 | 17.3 | 33.9 | 38 | 308 | 2.19 (1.55 - 3.01) | 9.10 (8.11 - 10.2) |
| DAG | 5,675 | 1,395 | 4.3 | 1.3 | 5 | 5 | 1.16 (0.38 - 2.71) | 3.98 (1.29 - 9.29) |
| ODOD | 17,261 | 48,998 | 10.6 | 39.7 | 36 | 789 | 3.39 (2.37 - 4.69) | 19.9 (18.5 - 21.3) |
| MDCR | 2,115 | 8,517 | 1.5 | 7.3 | <5 | 149 | < 3.31 (1.07 - 7.72) | 20.5 (17.3 - 24.0) |
| MDCD | 1,468 | 7,319 | 0.7 | 4.3 | <5 | 106 | < 6.76 (2.19 - 15.8) | 24.9 (20.4 - 30.1) |
| Open Claims | 328,462 | 883,945 | 243.0 | 809.2 | 289 | 6,052 | 1.19 (1.06 - 1.33) | 7.48 (7.29 - 7.67) |
| OEHR | 20,063 | 75,672 | 5.6 | 36.8 | 12 | 487 | 2.12 (1.10 - 3.71) | 13.2 (12.1 - 14.5) |
| VA | 12,356 | 110,156 | 6.4 | 89.4 | <10 | 842 | < 1.56 (0.75 - 2.86) | 9.42 (8.79 - 10.1) |
| SIDIAP | 5,159 | 2,687 | 6.3 | 5.4 | <5 | 16 | < 0.79 (0.26 - 1.85) | 2.99 (1.71 - 4.86) |
| Meta-analysis | 389,619 | 1,055,661 | 276.6 | 919.5 | 375 | 7,636 | 1.36 (1.22 - 1.50) | 8.30 (8.12 - 8.49) |
| Meta-analysis (w/o USOC) | 61,157 | 171,716 | 33.6 | 110.4 | 86 | 1,584 | 2.56 (2.05 - 3.16) | 14.4 (13.7 - 15.1) |

##### 4.3.19 Hypotension

**Supplementary Table 360:** Exposures, outcomes and risk estimates for Hypotension in SGLT2I (male) (T) vs SU (male) (C) new-user cohorts under four different propensity score (PS) adjustment / time-at-risk (TAR) designs across data sources. We report total patient exposure time in 1,000 patient-year (KYr) units, uncalibrated and calibrated hazard ratios (HRs) and their 95% confidence intervals (CIs). Greyed out entries did not pass study diagnostics and are included only for completeness.

| | Patients | | Exposure (KYr) | | Outcomes | | Incidence (per KYr) | |
|---|---|---|---|---|---|---|---|---|
| | T | C | T | C | T | C | T | C |
| **PS-matching / on-treatment TAR** | | | | | | | | |
| CCAE | 20,246 | 46,020 | 17.2 | 38.4 | 151 | 314 | 8.76 (7.42 - 10.3) | 8.18 (7.30 - 9.14) |
| DAG | 5,260 | 947 | 4.4 | 0.9 | 25 | 5 | 5.71 (3.70 - 8.43) | 5.63 (1.83 - 13.1) |
| ODOD | 15,100 | 46,353 | 10.7 | 41.6 | 208 | 1,056 | 19.4 (16.8 - 22.2) | 25.4 (23.9 - 27.0) |
| MDCR | 1,678 | 7,772 | 1.4 | 7.3 | 30 | 195 | 20.9 (14.1 - 29.8) | 26.7 (23.1 - 30.8) |
| MDCD | 1,375 | 6,741 | 0.8 | 4.6 | 21 | 123 | 25.4 (15.7 - 38.8) | 26.5 (22.0 - 31.6) |
| Open Claims | 295,340 | 857,867 | 263.1 | 923.3 | 1,790 | 8,573 | 6.80 (6.49 - 7.13) | 9.29 (9.09 - 9.48) |
| OEHR | 17,909 | 71,996 | 5.7 | 39.8 | 74 | 680 | 13.1 (10.3 - 16.4) | 17.1 (15.8 - 18.4) |
| VA | 0 | 0 | 0.0 | 0.0 | 0 | 0 | | |
| SIDIAP | 4,522 | 1,213 | 6.6 | 2.2 | 23 | <5 | 3.50 (2.22 - 5.26) | < 2.24 (0.73 - 5.23) |
| Meta-analysis | 356,170 | 1,037,962 | 305.6 | 1057.3 | 2,297 | <10,946 | 7.52 (7.21 - 7.83) | < 10.4 (10.2 - 10.5) |
| Meta-analysis (w/o USOC) | 60,830 | 180,095 | 42.5 | 134.0 | 507 | <2,373 | 11.9 (10.9 - 13.0) | < 17.7 (17.0 - 18.4) |
| **PS-stratification / on-treatment TAR** | | | | | | | | |
| CCAE | 23,656 | 46,632 | 19.6 | 38.7 | 177 | 320 | 9.02 (7.74 - 10.4) | 8.26 (7.38 - 9.22) |
| DAG | 5,526 | 1,358 | 4.5 | 1.3 | 25 | 7 | 5.50 (3.56 - 8.12) | 5.21 (2.10 - 10.7) |
| ODOD | 16,736 | 46,829 | 11.6 | 42.0 | 229 | 1,089 | 19.8 (17.3 - 22.5) | 25.9 (24.4 - 27.5) |
| MDCR | 2,017 | 7,968 | 1.6 | 7.4 | 35 | 205 | 21.8 (15.2 - 30.3) | 27.5 (23.9 - 31.6) |
| MDCD | 1,415 | 6,935 | 0.8 | 4.8 | 21 | 131 | 25.0 (15.4 - 38.1) | 27.3 (22.8 - 32.4) |
| Open Claims | 321,381 | 863,294 | 279.9 | 929.0 | 1,989 | 8,819 | 7.11 (6.80 - 7.43) | 9.49 (9.30 - 9.69) |
| OEHR | 19,804 | 74,383 | 6.3 | 41.3 | 83 | 748 | 13.2 (10.5 - 16.3) | 18.1 (16.8 - 19.4) |
| VA | 11,420 | 104,433 | 6.9 | 94.8 | 152 | 1,252 | 22.2 (18.8 - 26.0) | 13.2 (12.5 - 14.0) |
| SIDIAP | 5,126 | 2,678 | 7.0 | 6.2 | 23 | 7 | 3.28 (2.08 - 4.92) | 1.12 (0.45 - 2.31) |
| Meta-analysis | 381,577 | 1,031,138 | 317.4 | 1051.1 | 2,478 | 10,976 | 7.81 (7.50 - 8.12) | 10.4 (10.2 - 10.6) |
| Meta-analysis (w/o USOC) | 60,196 | 167,844 | 37.5 | 122.1 | 489 | 2,157 | 13.0 (11.9 - 14.2) | 17.7 (16.9 - 18.4) |
| **PS-matching / on-treatment TAR without escalation** | | | | | | | | |
| CCAE | 20,246 | 46,020 | 15.0 | 33.3 | 123 | 255 | 8.19 (6.81 - 9.77) | 7.66 (6.75 - 8.66) |
| DAG | 5,260 | 947 | 4.0 | 0.8 | 22 | <5 | 5.47 (3.43 - 8.28) | < 6.14 (1.99 - 14.3) |
| ODOD | 15,100 | 46,353 | 9.5 | 37.2 | 185 | 956 | 19.5 (16.7 - 22.5) | 25.7 (24.1 - 27.3) |
| MDCR | 1,678 | 7,772 | 1.3 | 6.7 | 24 | 173 | 18.7 (12.0 - 27.8) | 25.9 (22.2 - 30.1) |
| MDCD | 1,375 | 6,741 | 0.7 | 3.9 | 17 | 102 | 24.2 (14.1 - 38.8) | 26.4 (21.5 - 32.0) |
| Open Claims | 295,340 | 857,867 | 223.2 | 783.7 | 1,486 | 7,180 | 6.66 (6.32 - 7.00) | 9.16 (8.95 - 9.38) |
| OEHR | 17,909 | 71,996 | 5.0 | 34.7 | 56 | 474 | 11.3 (8.51 - 14.6) | 13.7 (12.5 - 14.9) |
| VA | 0 | 0 | 0.0 | 0.0 | 0 | 0 | | |
| SIDIAP | 4,522 | 1,213 | 5.8 | 1.9 | 20 | <5 | 3.43 (2.09 - 5.30) | < 2.57 (0.84 - 6.01) |
| Meta-analysis | 356,170 | 1,037,962 | 260.6 | 901.4 | 1,911 | <9,145 | 7.33 (7.01 - 7.67) | < 10.1 (9.94 - 10.4) |
| Meta-analysis (w/o USOC) | 60,830 | 180,095 | 37.3 | 117.7 | 425 | <1,965 | 11.4 (10.3 - 12.5) | < 16.7 (16.0 - 17.4) |
| **PS-stratification / on-treatment TAR without escalation** | | | | | | | | |
| CCAE | 23,656 | 46,632 | 17.2 | 33.6 | 146 | 259 | 8.51 (7.18 - 10.0) | 7.70 (6.79 - 8.70) |
| DAG | 5,526 | 1,358 | 4.2 | 1.2 | 22 | 6 | 5.27 (3.30 - 7.98) | 4.91 (1.80 - 10.7) |
| ODOD | 16,736 | 46,829 | 10.3 | 37.6 | 204 | 986 | 19.9 (17.2 - 22.8) | 26.2 (24.6 - 27.9) |
| MDCR | 2,017 | 7,968 | 1.4 | 6.8 | 29 | 181 | 20.2 (13.5 - 29.0) | 26.6 (22.8 - 30.7) |
| MDCD | 1,415 | 6,935 | 0.7 | 4.0 | 17 | 108 | 23.8 (13.8 - 38.1) | 27.0 (22.1 - 32.6) |
| Open Claims | 321,381 | 863,294 | 238.2 | 788.6 | 1,667 | 7,409 | 7.00 (6.67 - 7.34) | 9.39 (9.18 - 9.61) |
| OEHR | 19,804 | 74,383 | 5.5 | 36.0 | 65 | 520 | 11.7 (9.04 - 14.9) | 14.4 (13.2 - 15.7) |
| VA | 11,420 | 104,433 | 5.9 | 84.1 | 129 | 1,000 | 21.7 (18.1 - 25.8) | 11.9 (11.2 - 12.7) |
| SIDIAP | 5,126 | 2,678 | 6.3 | 5.3 | 20 | 6 | 3.20 (1.95 - 4.94) | 1.13 (0.41 - 2.45) |
| Meta-analysis | 381,577 | 1,031,138 | 271.2 | 895.9 | 2,082 | 9,174 | 7.68 (7.35 - 8.01) | 10.2 (10.0 - 10.5) |
| Meta-analysis (w/o USOC) | 60,196 | 167,844 | 33.0 | 107.2 | 415 | 1,765 | 12.6 (11.4 - 13.9) | 16.5 (15.7 - 17.2) |

##### 4.3.20 Joint pain

**Supplementary Table 361:** Exposures, outcomes and risk estimates for Joint pain in SGLT2I (male) (T) vs SU (male) (C) new-user cohorts under four different propensity score (PS) adjustment / time-at-risk (TAR) designs across data sources. We report total patient exposure time in 1,000 patient-year (KYr) units, uncalibrated and calibrated hazard ratios (HRs) and their 95% confidence intervals (CIs). Greyed out entries did not pass study diagnostics and are included only for completeness.

| | Patients | | Exposure (KYr) | | Outcomes | | Incidence (per KYr) | |
|---|---|---|---|---|---|---|---|---|
| | T | C | T | C | T | C | T | C |
| **PS-matching / on-treatment TAR** | | | | | | | | |
| CCAE | 19,768 | 44,945 | 16.8 | 37.2 | 116 | 324 | 6.91 (5.71 - 8.29) | 8.70 (7.78 - 9.70) |
| DAG | 4,662 | 846 | 3.8 | 0.8 | 59 | 13 | 15.4 (11.7 - 19.9) | 16.9 (8.98 - 28.8) |
| ODOD | 15,016 | 46,626 | 10.7 | 42.2 | 98 | 430 | 9.16 (7.43 - 11.2) | 10.2 (9.25 - 11.2) |
| MDCR | 1,723 | 7,929 | 1.5 | 7.4 | 11 | 66 | 7.45 (3.72 - 13.3) | 8.89 (6.87 - 11.3) |
| MDCD | 1,350 | 6,763 | 0.8 | 4.7 | 9 | 62 | 11.1 (5.08 - 21.1) | 13.2 (10.2 - 17.0) |
| Open Claims | 290,971 | 853,349 | 259.2 | 920.9 | 1,062 | 4,402 | 4.10 (3.85 - 4.35) | 4.78 (4.64 - 4.92) |
| OEHR | 17,579 | 71,027 | 5.5 | 39.5 | 45 | 319 | 8.14 (5.94 - 10.9) | 8.08 (7.22 - 9.02) |
| VA | 0 | 0 | 0.0 | 0.0 | 0 | 0 | | |
| SIDIAP | 3,648 | 1,010 | 5.3 | 1.8 | 82 | 46 | 15.3 (12.2 - 19.0) | 25.9 (19.0 - 34.5) |
| Meta-analysis | 354,717 | 1,032,495 | 303.7 | 1054.5 | 1,482 | 5,662 | 4.88 (4.63 - 5.13) | 5.37 (5.23 - 5.51) |
| Meta-analysis (w/o USOC) | 63,746 | 179,146 | 44.5 | 133.5 | 420 | 1,260 | 9.44 (8.56 - 10.4) | 9.44 (8.92 - 9.97) |
| **PS-stratification / on-treatment TAR** | | | | | | | | |
| CCAE | 23,075 | 45,566 | 19.1 | 37.6 | 137 | 326 | 7.16 (6.01 - 8.46) | 8.67 (7.76 - 9.67) |
| DAG | 4,851 | 1,219 | 3.9 | 1.2 | 60 | 19 | 15.2 (11.6 - 19.6) | 16.3 (9.81 - 25.4) |
| ODOD | 16,688 | 47,302 | 11.6 | 42.9 | 103 | 435 | 8.91 (7.27 - 10.8) | 10.2 (9.22 - 11.2) |
| MDCR | 2,062 | 8,283 | 1.6 | 7.7 | 11 | 68 | 6.70 (3.34 - 12.0) | 8.78 (6.82 - 11.1) |
| MDCD | 1,397 | 6,982 | 0.8 | 4.9 | 9 | 64 | 10.9 (5.00 - 20.7) | 13.2 (10.2 - 16.8) |
| Open Claims | 317,231 | 859,008 | 276.1 | 926.8 | 1,151 | 4,442 | 4.17 (3.93 - 4.42) | 4.79 (4.65 - 4.94) |
| OEHR | 19,457 | 73,365 | 6.2 | 41.0 | 51 | 338 | 8.28 (6.16 - 10.9) | 8.24 (7.38 - 9.17) |
| VA | 10,977 | 97,215 | 6.6 | 88.9 | 46 | 581 | 6.93 (5.08 - 9.25) | 6.54 (6.01 - 7.09) |
| SIDIAP | 4,147 | 2,263 | 5.7 | 5.0 | 87 | 118 | 15.2 (12.2 - 18.7) | 23.5 (19.4 - 28.1) |
| Meta-analysis | 376,451 | 1,025,241 | 313.0 | 1048.3 | 1,442 | 5,541 | 4.61 (4.37 - 4.85) | 5.29 (5.15 - 5.43) |
| Meta-analysis (w/o USOC) | 59,220 | 166,233 | 36.9 | 121.5 | 291 | 1,099 | 7.89 (7.01 - 8.86) | 9.05 (8.52 - 9.60) |
| **PS-matching / on-treatment TAR without escalation** | | | | | | | | |
| CCAE | 19,768 | 44,945 | 14.6 | 32.3 | 96 | 278 | 6.58 (5.33 - 8.03) | 8.60 (7.62 - 9.67) |
| DAG | 4,662 | 846 | 3.5 | 0.7 | 53 | 13 | 15.0 (11.3 - 19.7) | 18.3 (9.73 - 31.2) |
| ODOD | 15,016 | 46,626 | 9.5 | 37.8 | 90 | 383 | 9.49 (7.63 - 11.7) | 10.1 (9.14 - 11.2) |
| MDCR | 1,723 | 7,929 | 1.3 | 6.8 | 10 | 62 | 7.61 (3.65 - 14.0) | 9.11 (6.98 - 11.7) |
| MDCD | 1,350 | 6,763 | 0.7 | 3.9 | 8 | 51 | 11.7 (5.05 - 23.0) | 13.1 (9.75 - 17.2) |
| Open Claims | 290,971 | 853,349 | 219.8 | 781.7 | 892 | 3,728 | 4.06 (3.80 - 4.33) | 4.77 (4.62 - 4.92) |
| OEHR | 17,579 | 71,027 | 4.8 | 34.3 | 39 | 267 | 8.06 (5.73 - 11.0) | 7.79 (6.88 - 8.78) |
| VA | 0 | 0 | 0.0 | 0.0 | 0 | 0 | | |
| SIDIAP | 3,648 | 1,010 | 4.7 | 1.5 | 72 | 38 | 15.2 (11.9 - 19.1) | 24.6 (17.4 - 33.8) |
| Meta-analysis | 354,717 | 1,032,495 | 259.0 | 899.1 | 1,260 | 4,820 | 4.86 (4.60 - 5.14) | 5.36 (5.21 - 5.51) |
| Meta-analysis (w/o USOC) | 63,746 | 179,146 | 39.2 | 117.4 | 368 | 1,092 | 9.39 (8.45 - 10.4) | 9.31 (8.76 - 9.87) |
| **PS-stratification / on-treatment TAR without escalation** | | | | | | | | |
| CCAE | 23,075 | 45,566 | 16.7 | 32.6 | 115 | 280 | 6.88 (5.68 - 8.26) | 8.58 (7.60 - 9.64) |
| DAG | 4,851 | 1,219 | 3.6 | 1.1 | 54 | 19 | 14.9 (11.2 - 19.4) | 17.9 (10.8 - 28.0) |
| ODOD | 16,688 | 47,302 | 10.3 | 38.4 | 95 | 387 | 9.26 (7.49 - 11.3) | 10.1 (9.10 - 11.1) |
| MDCR | 2,062 | 8,283 | 1.5 | 7.1 | 10 | 64 | 6.82 (3.27 - 12.6) | 9.01 (6.94 - 11.5) |
| MDCD | 1,397 | 6,982 | 0.7 | 4.0 | 8 | 51 | 11.5 (4.95 - 22.6) | 12.6 (9.40 - 16.6) |
| Open Claims | 317,231 | 859,008 | 234.9 | 786.8 | 972 | 3,761 | 4.14 (3.88 - 4.41) | 4.78 (4.63 - 4.94) |
| OEHR | 19,457 | 73,365 | 5.4 | 35.6 | 45 | 283 | 8.30 (6.05 - 11.1) | 7.96 (7.06 - 8.94) |
| VA | 10,977 | 97,215 | 5.7 | 78.8 | 36 | 524 | 6.29 (4.40 - 8.71) | 6.65 (6.09 - 7.24) |
| SIDIAP | 4,147 | 2,263 | 5.1 | 4.3 | 76 | 104 | 14.9 (11.7 - 18.6) | 24.2 (19.7 - 29.3) |
| Meta-analysis | 376,451 | 1,025,241 | 267.3 | 893.4 | 1,227 | 4,711 | 4.59 (4.34 - 4.85) | 5.27 (5.12 - 5.43) |
| Meta-analysis (w/o USOC) | 59,220 | 166,233 | 32.4 | 106.6 | 255 | 950 | 7.87 (6.94 - 8.90) | 8.91 (8.35 - 9.50) |

###### 4.3.21 Lower extremity amputation

**Supplementary Table 362:** Exposures, outcomes and risk estimates for Lower extremity amputation in SGLT2I (male) (T) vs SU (male) (C) new-user cohorts under four different propensity score (PS) adjustment / time-at-risk (TAR) designs across data sources. We report total patient exposure time in 1,000 patient-year (KYr) units, uncalibrated and calibrated hazard ratios (HRs) and their 95% confidence intervals (CIs). Greyed out entries did not pass study diagnostics and are included only for completeness.

| | Patients | | Exposure (KYr) | | Outcomes | | Incidence (per KYr) | |
|---|---|---|---|---|---|---|---|---|
| | T | C | T | C | T | C | T | C |
| **PS-matching / on-treatment TAR** | | | | | | | | |
| CCAE | 20,621 | 46,859 | 17.6 | 39.2 | 19 | 43 | 1.08 (0.65 - 1.69) | 1.10 (0.79 - 1.48) |
| DAG | 5,471 | 973 | 4.6 | 0.9 | 0 | 0 | | |
| ODOD | 15,700 | 48,812 | 11.2 | 44.5 | 13 | 78 | 1.16 (0.62 - 1.98) | 1.75 (1.39 - 2.19) |
| MDCR | 1,774 | 8,300 | 1.5 | 7.8 | <5 | 13 | < 3.28 (1.06 - 7.65) | 1.66 (0.88 - 2.84) |
| MDCD | 1,460 | 7,201 | 0.9 | 5.1 | <5 | 11 | < 5.63 (1.83 - 13.1) | 2.18 (1.09 - 3.90) |
| Open Claims | 304,425 | 886,839 | 271.2 | 959.5 | 154 | 777 | 0.57 (0.48 - 0.67) | 0.81 (0.75 - 0.87) |
| OEHR | 18,275 | 73,767 | 5.8 | 41.4 | <5 | 53 | < 0.86 (0.28 - 2.01) | 1.28 (0.96 - 1.68) |
| VA | 0 | 0 | 0.0 | 0.0 | 0 | 0 | | |
| SIDIAP | 4,552 | 1,219 | 6.6 | 2.2 | 0 | 0 | | |
| Meta-analysis | 359,021 | 1,056,277 | 305.8 | 1084.5 | <191 | 951 | < 0.62 (0.54 - 0.72) | 0.88 (0.82 - 0.93) |
| Meta-analysis (w/o USOC) | 54,596 | 169,438 | 34.6 | 125.0 | <37 | 174 | < 1.07 (0.75 - 1.47) | 1.39 (1.19 - 1.61) |
| **PS-stratification / on-treatment TAR** | | | | | | | | |
| CCAE | 24,122 | 47,502 | 20.1 | 39.6 | 19 | 45 | 0.95 (0.57 - 1.48) | 1.14 (0.83 - 1.52) |
| DAG | 5,702 | 1,403 | 4.7 | 1.4 | 0 | 0 | | |
| ODOD | 17,448 | 49,467 | 12.1 | 45.1 | 13 | 80 | 1.07 (0.57 - 1.83) | 1.77 (1.41 - 2.21) |
| MDCR | 2,129 | 8,585 | 1.7 | 8.1 | <5 | 13 | < 2.94 (0.95 - 6.86) | 1.61 (0.86 - 2.76) |
| MDCD | 1,503 | 7,445 | 0.9 | 5.2 | <5 | 11 | < 5.55 (1.80 - 12.9) | 2.10 (1.05 - 3.75) |
| Open Claims | 332,242 | 892,529 | 289.1 | 965.4 | 159 | 807 | 0.55 (0.47 - 0.64) | 0.84 (0.78 - 0.90) |
| OEHR | 20,221 | 76,212 | 6.5 | 43.0 | <5 | 58 | < 0.77 (0.25 - 1.81) | 1.35 (1.02 - 1.74) |
| VA | 12,506 | 110,856 | 7.5 | 102.0 | 11 | 89 | 1.46 (0.73 - 2.62) | 0.87 (0.70 - 1.07) |
| SIDIAP | 5,166 | 2,688 | 7.1 | 6.3 | 0 | <5 | | < 0.80 (0.26 - 1.86) |
| Meta-analysis | 394,033 | 1,065,710 | 327.7 | 1093.1 | <196 | 990 | < 0.60 (0.52 - 0.69) | 0.91 (0.85 - 0.96) |
| Meta-analysis (w/o USOC) | 61,791 | 173,181 | 38.7 | 127.7 | <37 | 183 | < 0.96 (0.67 - 1.32) | 1.43 (1.23 - 1.66) |
| **PS-matching / on-treatment TAR without escalation** | | | | | | | | |
| CCAE | 20,621 | 46,859 | 15.3 | 33.9 | 14 | 33 | 0.91 (0.50 - 1.53) | 0.97 (0.67 - 1.37) |
| DAG | 5,471 | 973 | 4.2 | 0.8 | 0 | 0 | | |
| ODOD | 15,700 | 48,812 | 9.9 | 39.8 | 10 | 66 | 1.01 (0.48 - 1.85) | 1.66 (1.28 - 2.11) |
| MDCR | 1,774 | 8,300 | 1.4 | 7.2 | <5 | 12 | < 3.69 (1.20 - 8.60) | 1.67 (0.86 - 2.92) |
| MDCD | 1,460 | 7,201 | 0.7 | 4.2 | <5 | 8 | < 6.67 (2.17 - 15.6) | 1.91 (0.83 - 3.77) |
| Open Claims | 304,425 | 886,839 | 229.8 | 813.7 | 111 | 626 | 0.48 (0.40 - 0.58) | 0.77 (0.71 - 0.83) |
| OEHR | 18,275 | 73,767 | 5.1 | 35.9 | <5 | 29 | < 0.98 (0.32 - 2.29) | 0.81 (0.54 - 1.16) |
| VA | 0 | 0 | 0.0 | 0.0 | 0 | 0 | | |
| SIDIAP | 4,552 | 1,219 | 5.9 | 2.0 | 0 | 0 | | |
| Meta-analysis | 359,021 | 1,056,277 | 260.2 | 923.3 | <140 | 754 | < 0.54 (0.45 - 0.63) | 0.82 (0.76 - 0.88) |
| Meta-analysis (w/o USOC) | 54,596 | 169,438 | 30.3 | 109.6 | <29 | 128 | < 0.96 (0.64 - 1.37) | 1.17 (0.97 - 1.39) |
| **PS-stratification / on-treatment TAR without escalation** | | | | | | | | |
| CCAE | 24,122 | 47,502 | 17.5 | 34.3 | 14 | 35 | 0.80 (0.44 - 1.34) | 1.02 (0.71 - 1.42) |
| DAG | 5,702 | 1,403 | 4.3 | 1.3 | 0 | 0 | | |
| ODOD | 17,448 | 49,467 | 10.8 | 40.3 | 10 | 67 | 0.93 (0.45 - 1.71) | 1.66 (1.29 - 2.11) |
| MDCR | 2,129 | 8,585 | 1.5 | 7.4 | <5 | 12 | < 3.29 (1.07 - 7.67) | 1.62 (0.84 - 2.84) |
| MDCD | 1,503 | 7,445 | 0.8 | 4.3 | <5 | 8 | < 6.56 (2.13 - 15.3) | 1.84 (0.79 - 3.63) |
| Open Claims | 332,242 | 892,529 | 245.8 | 818.9 | 116 | 654 | 0.47 (0.39 - 0.57) | 0.80 (0.74 - 0.86) |
| OEHR | 20,221 | 76,212 | 5.7 | 37.3 | <5 | 31 | < 0.88 (0.29 - 2.05) | 0.83 (0.57 - 1.18) |
| VA | 12,506 | 110,856 | 6.5 | 90.3 | 11 | 71 | 1.69 (0.84 - 3.02) | 0.79 (0.61 - 0.99) |
| SIDIAP | 5,166 | 2,688 | 6.3 | 5.4 | 0 | 0 | | |
| Meta-analysis | 394,033 | 1,065,710 | 279.8 | 930.7 | <145 | 787 | < 0.52 (0.44 - 0.61) | 0.85 (0.79 - 0.91) |
| Meta-analysis (w/o USOC) | 61,791 | 173,181 | 34.0 | 111.9 | <29 | 133 | < 0.85 (0.57 - 1.23) | 1.19 (1.00 - 1.41) |

##### 4.3.22 Nausea

**Supplementary Table 363:** Exposures, outcomes and risk estimates for Nausea in SGLT2I (male) (T) vs SU (male) (C) new-user cohorts under four different propensity score (PS) adjustment / time-at-risk (TAR) designs across data sources. We report total patient exposure time in 1,000 patient-year (KYr) units, uncalibrated and calibrated hazard ratios (HRs) and their 95% confidence intervals (CIs). Greyed out entries did not pass study diagnostics and are included only for completeness.

| | Patients | | Exposure (KYr) | | Outcomes | | Incidence (per KYr) | |
|---|---|---|---|---|---|---|---|---|
| | T | C | T | C | T | C | T | C |
| **PS-matching / on-treatment TAR** | | | | | | | | |
| CCAE | 18,644 | 42,437 | 15.6 | 35.2 | 381 | 816 | 24.4 (22.1 - 27.0) | 23.2 (21.6 - 24.8) |
| DAG | 5,175 | 921 | 4.3 | 0.9 | 33 | 8 | 7.64 (5.26 - 10.7) | 9.22 (3.98 - 18.2) |
| ODOD | 14,037 | 43,462 | 9.9 | 39.0 | 281 | 1,284 | 28.3 (25.1 - 31.8) | 32.9 (31.2 - 34.8) |
| MDCR | 1,627 | 7,529 | 1.4 | 7.1 | 36 | 226 | 26.0 (18.2 - 36.0) | 32.0 (28.0 - 36.5) |
| MDCD | 1,157 | 5,722 | 0.7 | 4.0 | 43 | 252 | 62.6 (45.3 - 84.3) | 63.4 (55.8 - 71.7) |
| Open Claims | 274,852 | 807,270 | 245.4 | 871.4 | 2,980 | 13,407 | 12.1 (11.7 - 12.6) | 15.4 (15.1 - 15.6) |
| OEHR | 16,955 | 68,391 | 5.3 | 37.5 | 147 | 1,001 | 27.6 (23.3 - 32.4) | 26.7 (25.1 - 28.4) |
| VA | 0 | 0 | 0.0 | 0.0 | 0 | 0 | | |
| SIDIAP | 4,418 | 1,186 | 6.5 | 2.2 | 19 | 5 | 2.94 (1.77 - 4.59) | 2.29 (0.74 - 5.34) |
| Meta-analysis | 332,447 | 975,732 | 282.7 | 995.0 | 3,901 | 16,994 | 13.8 (13.4 - 14.2) | 17.1 (16.8 - 17.3) |
| Meta-analysis (w/o USOC) | 57,595 | 168,462 | 37.3 | 123.5 | 921 | 3,587 | 24.7 (23.2 - 26.4) | 29.0 (28.1 - 30.0) |
| **PS-stratification / on-treatment TAR** | | | | | | | | |
| CCAE | 21,821 | 43,009 | 17.8 | 35.5 | 432 | 829 | 24.2 (22.0 - 26.6) | 23.3 (21.8 - 25.0) |
| DAG | 5,395 | 1,334 | 4.5 | 1.3 | 34 | 12 | 7.63 (5.29 - 10.7) | 9.23 (4.77 - 16.1) |
| ODOD | 15,574 | 44,119 | 10.7 | 39.6 | 302 | 1,322 | 28.2 (25.1 - 31.5) | 33.4 (31.6 - 35.2) |
| MDCR | 1,958 | 7,697 | 1.5 | 7.2 | 38 | 233 | 24.5 (17.3 - 33.7) | 32.4 (28.4 - 36.8) |
| MDCD | 1,195 | 5,861 | 0.7 | 4.1 | 43 | 262 | 61.5 (44.5 - 82.9) | 64.2 (56.7 - 72.5) |
| Open Claims | 299,241 | 812,448 | 261.1 | 876.8 | 3,193 | 13,602 | 12.2 (11.8 - 12.7) | 15.5 (15.3 - 15.8) |
| OEHR | 18,784 | 70,594 | 5.9 | 38.9 | 162 | 1,064 | 27.3 (23.3 - 31.8) | 27.4 (25.7 - 29.1) |
| VA | 11,542 | 103,745 | 7.0 | 95.1 | 53 | 948 | 7.61 (5.70 - 9.96) | 9.97 (9.35 - 10.6) |
| SIDIAP | 5,004 | 2,630 | 6.9 | 6.1 | 20 | 25 | 2.90 (1.77 - 4.48) | 4.09 (2.65 - 6.04) |
| Meta-analysis | 355,420 | 970,170 | 295.6 | 990.8 | 4,089 | 16,817 | 13.8 (13.4 - 14.3) | 17.0 (16.7 - 17.2) |
| Meta-analysis (w/o USOC) | 56,179 | 157,722 | 34.5 | 114.0 | 896 | 3,215 | 26.0 (24.3 - 27.7) | 28.2 (27.2 - 29.2) |
| **PS-matching / on-treatment TAR without escalation** | | | | | | | | |
| CCAE | 18,644 | 42,437 | 13.7 | 30.6 | 316 | 707 | 23.1 (20.7 - 25.8) | 23.1 (21.4 - 24.8) |
| DAG | 5,175 | 921 | 4.0 | 0.8 | 30 | 8 | 7.55 (5.10 - 10.8) | 10.1 (4.34 - 19.8) |
| ODOD | 14,037 | 43,462 | 8.9 | 35.0 | 239 | 1,138 | 27.0 (23.7 - 30.6) | 32.5 (30.7 - 34.5) |
| MDCR | 1,627 | 7,529 | 1.2 | 6.5 | 33 | 200 | 26.6 (18.3 - 37.4) | 30.8 (26.7 - 35.4) |
| MDCD | 1,157 | 5,722 | 0.6 | 3.4 | 37 | 216 | 63.4 (44.7 - 87.5) | 64.4 (56.1 - 73.5) |
| Open Claims | 274,852 | 807,270 | 208.8 | 742.1 | 2,361 | 11,117 | 11.3 (10.9 - 11.8) | 15.0 (14.7 - 15.3) |
| OEHR | 16,955 | 68,391 | 4.7 | 32.8 | 117 | 772 | 25.0 (20.6 - 29.9) | 23.6 (21.9 - 25.3) |
| VA | 0 | 0 | 0.0 | 0.0 | 0 | 0 | | |
| SIDIAP | 4,418 | 1,186 | 5.7 | 1.9 | 17 | <5 | 2.96 (1.73 - 4.74) | < 2.63 (0.86 - 6.15) |
| Meta-analysis | 332,447 | 975,732 | 241.8 | 851.2 | 3,133 | 14,158 | 13.0 (12.5 - 13.4) | 16.6 (16.4 - 16.9) |
| Meta-analysis (w/o USOC) | 57,595 | 168,462 | 33.0 | 109.0 | 772 | 3,041 | 23.4 (21.8 - 25.1) | 27.9 (26.9 - 28.9) |
| **PS-stratification / on-treatment TAR without escalation** | | | | | | | | |
| CCAE | 21,821 | 43,009 | 15.7 | 30.9 | 359 | 720 | 22.9 (20.6 - 25.4) | 23.3 (21.6 - 25.0) |
| DAG | 5,395 | 1,334 | 4.1 | 1.2 | 31 | 12 | 7.57 (5.14 - 10.7) | 10.1 (5.22 - 17.6) |
| ODOD | 15,574 | 44,119 | 9.6 | 35.5 | 256 | 1,170 | 26.8 (23.6 - 30.2) | 32.9 (31.1 - 34.9) |
| MDCR | 1,958 | 7,697 | 1.4 | 6.6 | 35 | 206 | 25.2 (17.5 - 35.0) | 31.1 (27.0 - 35.7) |
| MDCD | 1,195 | 5,861 | 0.6 | 3.4 | 37 | 226 | 62.3 (43.9 - 85.9) | 65.6 (57.3 - 74.7) |
| Open Claims | 299,241 | 812,448 | 222.9 | 746.8 | 2,542 | 11,285 | 11.4 (11.0 - 11.9) | 15.1 (14.8 - 15.4) |
| OEHR | 18,784 | 70,594 | 5.2 | 34.0 | 130 | 821 | 24.8 (20.7 - 29.5) | 24.2 (22.5 - 25.9) |
| VA | 11,542 | 103,745 | 6.0 | 84.3 | 41 | 765 | 6.80 (4.88 - 9.22) | 9.07 (8.44 - 9.74) |
| SIDIAP | 5,004 | 2,630 | 6.1 | 5.2 | 18 | 22 | 2.93 (1.74 - 4.63) | 4.22 (2.64 - 6.38) |
| Meta-analysis | 355,420 | 970,170 | 253.3 | 847.2 | 3,287 | 13,996 | 13.0 (12.5 - 13.4) | 16.5 (16.2 - 16.8) |
| Meta-analysis (w/o USOC) | 56,179 | 157,722 | 30.5 | 100.5 | 745 | 2,711 | 24.4 (22.7 - 26.3) | 27.0 (26.0 - 28.0) |

##### 4.3.23 Peripheral edema

**Supplementary Table 364:** Exposures, outcomes and risk estimates for Peripheral edema in SGLT2I (male) (T) vs SU (male) (C) new-user cohorts under four different propensity score (PS) adjustment / time-at-risk (TAR) designs across data sources. We report total patient exposure time in 1,000 patient-year (KYr) units, uncalibrated and calibrated hazard ratios (HRs) and their 95% confidence intervals (CIs). Greyed out entries did not pass study diagnostics and are included only for completeness.

| | Patients | | Exposure (KYr) | | Outcomes | | Incidence (per KYr) | |
|---|---|---|---|---|---|---|---|---|
| | T | C | T | C | T | C | T | C |
| **PS-matching / on-treatment TAR** | | | | | | | | |
| CCAE | 17,523 | 39,972 | 14.5 | 32.3 | 451 | 1,346 | 31.0 (28.2 - 34.0) | 41.7 (39.5 - 44.0) |
| DAG | 4,810 | 854 | 4.0 | 0.8 | 57 | 21 | 14.3 (10.8 - 18.5) | 26.9 (16.6 - 41.1) |
| ODOD | 12,839 | 38,577 | 9.0 | 32.8 | 377 | 2,478 | 41.9 (37.8 - 46.4) | 75.5 (72.6 - 78.6) |
| MDCR | 1,383 | 6,216 | 1.1 | 5.4 | 52 | 475 | 45.7 (34.1 - 59.9) | 87.7 (79.9 - 95.9) |
| MDCD | 1,069 | 5,088 | 0.6 | 3.3 | 37 | 278 | 59.4 (41.8 - 81.9) | 83.4 (73.8 - 93.8) |
| Open Claims | 249,361 | 729,685 | 221.8 | 764.2 | 3,909 | 25,663 | 17.6 (17.1 - 18.2) | 33.6 (33.2 - 34.0) |
| OEHR | 15,892 | 63,880 | 5.0 | 33.8 | 116 | 1,739 | 23.3 (19.2 - 27.9) | 51.4 (49.0 - 53.8) |
| VA | 0 | 0 | 0.0 | 0.0 | 0 | 0 | | |
| SIDIAP | 4,070 | 1,122 | 6.0 | 2.0 | 60 | 29 | 10.1 (7.68 - 13.0) | 14.3 (9.57 - 20.5) |
| Meta-analysis | 306,947 | 885,394 | 262.0 | 874.7 | 5,059 | 32,029 | 19.3 (18.8 - 19.8) | 36.6 (36.2 - 37.0) |
| Meta-analysis (w/o USOC) | 57,586 | 155,709 | 40.2 | 110.5 | 1,150 | 6,366 | 28.6 (27.0 - 30.3) | 57.6 (56.2 - 59.0) |
| **PS-stratification / on-treatment TAR** | | | | | | | | |
| CCAE | 20,331 | 40,503 | 16.5 | 32.5 | 518 | 1,366 | 31.5 (28.8 - 34.3) | 42.0 (39.8 - 44.3) |
| DAG | 5,011 | 1,246 | 4.1 | 1.2 | 57 | 39 | 13.9 (10.5 - 18.0) | 32.9 (23.4 - 45.0) |
| ODOD | 14,095 | 39,119 | 9.6 | 33.3 | 406 | 2,552 | 42.2 (38.2 - 46.5) | 76.6 (73.7 - 79.7) |
| MDCR | 1,633 | 6,377 | 1.3 | 5.5 | 56 | 501 | 44.4 (33.5 - 57.7) | 90.7 (82.9 - 99.0) |
| MDCD | 1,095 | 5,322 | 0.6 | 3.5 | 37 | 296 | 58.6 (41.2 - 80.7) | 84.5 (75.2 - 94.7) |
| Open Claims | 269,915 | 733,872 | 235.1 | 768.3 | 4,195 | 26,074 | 17.8 (17.3 - 18.4) | 33.9 (33.5 - 34.4) |
| OEHR | 17,562 | 65,731 | 5.5 | 34.9 | 130 | 1,856 | 23.5 (19.6 - 27.9) | 53.2 (50.8 - 55.6) |
| VA | 9,289 | 87,849 | 5.6 | 77.1 | 177 | 3,099 | 31.8 (27.3 - 36.9) | 40.2 (38.8 - 41.6) |
| SIDIAP | 4,631 | 2,504 | 6.4 | 5.7 | 66 | 79 | 10.3 (8.00 - 13.2) | 13.7 (10.9 - 17.1) |
| Meta-analysis | 321,903 | 879,225 | 266.7 | 869.0 | 5,249 | 31,848 | 19.7 (19.2 - 20.2) | 36.6 (36.2 - 37.1) |
| Meta-analysis (w/o USOC) | 51,988 | 145,353 | 31.6 | 100.8 | 1,054 | 5,774 | 33.3 (31.4 - 35.4) | 57.3 (55.8 - 58.8) |
| **PS-matching / on-treatment TAR without escalation** | | | | | | | | |
| CCAE | 17,523 | 39,972 | 12.7 | 28.2 | 378 | 1,176 | 29.7 (26.7 - 32.8) | 41.7 (39.4 - 44.2) |
| DAG | 4,810 | 854 | 3.7 | 0.7 | 53 | 21 | 14.5 (10.8 - 18.9) | 29.1 (18.0 - 44.5) |
| ODOD | 12,839 | 38,577 | 8.0 | 29.6 | 307 | 2,209 | 38.2 (34.0 - 42.7) | 74.6 (71.5 - 77.8) |
| MDCR | 1,383 | 6,216 | 1.0 | 5.0 | 44 | 436 | 43.1 (31.3 - 57.9) | 87.0 (79.0 - 95.5) |
| MDCD | 1,069 | 5,088 | 0.5 | 2.8 | 30 | 229 | 56.2 (37.9 - 80.3) | 81.3 (71.1 - 92.6) |
| Open Claims | 249,361 | 729,685 | 188.8 | 652.7 | 3,222 | 21,880 | 17.1 (16.5 - 17.7) | 33.5 (33.1 - 34.0) |
| OEHR | 15,892 | 63,880 | 4.4 | 29.7 | 95 | 1,393 | 21.6 (17.5 - 26.4) | 46.8 (44.4 - 49.4) |
| VA | 0 | 0 | 0.0 | 0.0 | 0 | 0 | | |
| SIDIAP | 4,070 | 1,122 | 5.3 | 1.8 | 54 | 26 | 10.2 (7.65 - 13.3) | 14.7 (9.63 - 21.6) |
| Meta-analysis | 306,947 | 885,394 | 224.5 | 750.5 | 4,183 | 27,370 | 18.6 (18.1 - 19.2) | 36.5 (36.0 - 36.9) |
| Meta-analysis (w/o USOC) | 57,586 | 155,709 | 35.7 | 97.9 | 961 | 5,490 | 26.9 (25.2 - 28.7) | 56.1 (54.6 - 57.6) |
| **PS-stratification / on-treatment TAR without escalation** | | | | | | | | |
| CCAE | 20,331 | 40,503 | 14.5 | 28.4 | 440 | 1,195 | 30.4 (27.6 - 33.4) | 42.0 (39.7 - 44.5) |
| DAG | 5,011 | 1,246 | 3.8 | 1.1 | 53 | 39 | 14.0 (10.5 - 18.3) | 36.0 (25.6 - 49.3) |
| ODOD | 14,095 | 39,119 | 8.6 | 30.1 | 336 | 2,277 | 39.0 (35.0 - 43.4) | 75.7 (72.7 - 78.9) |
| MDCR | 1,633 | 6,377 | 1.1 | 5.1 | 48 | 460 | 42.3 (31.2 - 56.1) | 90.1 (82.0 - 98.7) |
| MDCD | 1,095 | 5,322 | 0.5 | 3.0 | 30 | 246 | 55.3 (37.3 - 79.0) | 83.1 (73.1 - 94.2) |
| Open Claims | 269,915 | 733,872 | 200.8 | 656.3 | 3,475 | 22,252 | 17.3 (16.7 - 17.9) | 33.9 (33.5 - 34.4) |
| OEHR | 17,562 | 65,731 | 4.9 | 30.7 | 105 | 1,472 | 21.4 (17.5 - 25.9) | 48.0 (45.6 - 50.5) |
| VA | 9,289 | 87,849 | 4.8 | 68.7 | 151 | 2,720 | 31.3 (26.5 - 36.7) | 39.6 (38.1 - 41.1) |
| SIDIAP | 4,631 | 2,504 | 5.7 | 4.9 | 60 | 67 | 10.5 (8.03 - 13.6) | 13.6 (10.5 - 17.3) |
| Meta-analysis | 321,903 | 879,225 | 228.8 | 745.4 | 4,356 | 27,196 | 19.0 (18.5 - 19.6) | 36.5 (36.1 - 36.9) |
| Meta-analysis (w/o USOC) | 51,988 | 145,353 | 28.0 | 89.2 | 881 | 4,944 | 31.5 (29.4 - 33.6) | 55.4 (53.9 - 57.0) |

###### 4.3.24 Photosensitivity

**Supplementary Table 365:** Exposures, outcomes and risk estimates for Photosensitivity in SGLT2I (male) (T) vs SU (male) (C) new-user cohorts under four different propensity score (PS) adjustment / time-at-risk (TAR) designs across data sources. We report total patient exposure time in 1,000 patient-year (KYr) units, uncalibrated and calibrated hazard ratios (HRs) and their 95% confidence intervals (CIs). Greyed out entries did not pass study diagnostics and are included only for completeness.

| | Patients | | Exposure (KYr) | | Outcomes | | Incidence (per KYr) | |
|---|---|---|---|---|---|---|---|---|
| | T | C | T | C | T | C | T | C |
| **PS-matching / on-treatment TAR** | | | | | | | | |
| CCAE | 20,476 | 46,543 | 17.4 | 38.9 | 10 | 42 | 0.57 (0.28 - 1.05) | 1.08 (0.78 - 1.46) |
| DAG | 5,457 | 970 | 4.6 | 0.9 | 0 | 0 | | |
| ODOD | 15,608 | 48,467 | 11.1 | 44.0 | 5 | 42 | 0.45 (0.15 - 1.05) | 0.95 (0.69 - 1.29) |
| MDCR | 1,749 | 8,143 | 1.5 | 7.6 | <5 | 25 | < 3.34 (1.08 - 7.78) | 3.27 (2.12 - 4.83) |
| MDCD | 1,453 | 7,219 | 0.9 | 5.1 | 0 | <5 | | < 0.99 (0.32 - 2.30) |
| Open Claims | 302,021 | 881,219 | 268.8 | 951.5 | 67 | 446 | 0.25 (0.19 - 0.32) | 0.47 (0.43 - 0.51) |
| OEHR | 18,182 | 73,261 | 5.8 | 41.1 | 5 | 68 | 0.87 (0.28 - 2.02) | 1.66 (1.29 - 2.10) |
| VA | 0 | 0 | 0.0 | 0.0 | 0 | 0 | | |
| SIDIAP | 4,549 | 1,219 | 6.6 | 2.2 | <5 | <5 | < 0.76 (0.25 - 1.77) | < 2.24 (0.73 - 5.22) |
| Meta-analysis | 358,036 | 1,057,633 | 304.7 | 1083.2 | <92 | 623 | < 0.30 (0.24 - 0.37) | 0.58 (0.53 - 0.62) |
| Meta-analysis (w/o USOC) | 56,015 | 176,414 | 35.9 | 131.6 | <25 | 177 | < 0.70 (0.45 - 1.03) | 1.34 (1.15 - 1.56) |
| **PS-stratification / on-treatment TAR** | | | | | | | | |
| CCAE | 23,945 | 47,187 | 19.9 | 39.3 | 11 | 42 | 0.55 (0.28 - 0.99) | 1.07 (0.77 - 1.45) |
| DAG | 5,686 | 1,398 | 4.7 | 1.4 | 0 | <5 | | < 3.62 (1.17 - 8.44) |
| ODOD | 17,348 | 49,106 | 12.0 | 44.7 | 8 | 43 | 0.66 (0.29 - 1.31) | 0.96 (0.70 - 1.30) |
| MDCR | 2,105 | 8,426 | 1.7 | 7.9 | <5 | 25 | < 2.99 (0.97 - 6.97) | 3.18 (2.06 - 4.69) |
| MDCD | 1,495 | 7,464 | 0.9 | 5.3 | 0 | <5 | | < 0.95 (0.31 - 2.22) |
| Open Claims | 329,565 | 886,842 | 286.5 | 957.4 | 68 | 465 | 0.24 (0.18 - 0.30) | 0.49 (0.44 - 0.53) |
| OEHR | 20,107 | 75,737 | 6.4 | 42.7 | 5 | 79 | 0.78 (0.25 - 1.82) | 1.85 (1.46 - 2.31) |
| VA | 12,352 | 109,597 | 7.4 | 100.6 | <10 | 100 | < 1.35 (0.65 - 2.48) | 0.99 (0.81 - 1.21) |
| SIDIAP | 5,159 | 2,688 | 7.1 | 6.3 | <5 | 6 | < 0.71 (0.23 - 1.65) | 0.96 (0.35 - 2.09) |
| Meta-analysis | 390,965 | 1,058,872 | 324.9 | 1084.0 | 92 | 629 | 0.28 (0.23 - 0.35) | 0.58 (0.54 - 0.63) |
| Meta-analysis (w/o USOC) | 61,400 | 172,030 | 38.4 | 126.6 | 24 | 164 | 0.63 (0.40 - 0.93) | 1.30 (1.10 - 1.51) |
| **PS-matching / on-treatment TAR without escalation** | | | | | | | | |
| CCAE | 20,476 | 46,543 | 15.2 | 33.7 | 9 | 37 | 0.59 (0.27 - 1.13) | 1.10 (0.77 - 1.51) |
| DAG | 5,457 | 970 | 4.2 | 0.8 | 0 | 0 | | |
| ODOD | 15,608 | 48,467 | 9.9 | 39.4 | <5 | 40 | < 0.51 (0.16 - 1.18) | 1.02 (0.73 - 1.38) |
| MDCR | 1,749 | 8,143 | 1.3 | 7.0 | <5 | 24 | < 3.75 (1.22 - 8.75) | 3.43 (2.20 - 5.10) |
| MDCD | 1,453 | 7,219 | 0.7 | 4.2 | 0 | <5 | | < 1.19 (0.39 - 2.78) |
| Open Claims | 302,021 | 881,219 | 227.8 | 806.9 | 54 | 407 | 0.24 (0.18 - 0.31) | 0.50 (0.46 - 0.56) |
| OEHR | 18,182 | 73,261 | 5.1 | 35.6 | <5 | 62 | < 0.99 (0.32 - 2.31) | 1.74 (1.34 - 2.23) |
| VA | 0 | 0 | 0.0 | 0.0 | 0 | 0 | | |
| SIDIAP | 4,549 | 1,219 | 5.9 | 1.9 | <5 | <5 | < 0.85 (0.28 - 1.99) | < 2.57 (0.83 - 5.99) |
| Meta-analysis | 358,036 | 1,057,633 | 259.2 | 922.5 | <78 | 570 | < 0.30 (0.24 - 0.38) | 0.62 (0.57 - 0.67) |
| Meta-analysis (w/o USOC) | 56,015 | 176,414 | 31.4 | 115.7 | <24 | 163 | < 0.76 (0.49 - 1.14) | 1.41 (1.20 - 1.64) |
| **PS-stratification / on-treatment TAR without escalation** | | | | | | | | |
| CCAE | 23,945 | 47,187 | 17.4 | 34.0 | 10 | 37 | 0.58 (0.28 - 1.06) | 1.09 (0.77 - 1.50) |
| DAG | 5,686 | 1,398 | 4.3 | 1.3 | 0 | <5 | | < 3.97 (1.29 - 9.27) |
| ODOD | 17,348 | 49,106 | 10.7 | 39.9 | 7 | 41 | 0.66 (0.26 - 1.35) | 1.03 (0.74 - 1.39) |
| MDCR | 2,105 | 8,426 | 1.5 | 7.2 | <5 | 24 | < 3.34 (1.08 - 7.80) | 3.33 (2.13 - 4.95) |
| MDCD | 1,495 | 7,464 | 0.8 | 4.4 | 0 | <5 | | < 1.14 (0.37 - 2.67) |
| Open Claims | 329,565 | 886,842 | 243.6 | 811.9 | 55 | 426 | 0.23 (0.17 - 0.29) | 0.52 (0.48 - 0.58) |
| OEHR | 20,107 | 75,737 | 5.7 | 37.0 | <5 | 73 | < 0.88 (0.29 - 2.06) | 1.98 (1.55 - 2.48) |
| VA | 12,352 | 109,597 | 6.4 | 89.1 | <10 | 92 | < 1.56 (0.75 - 2.87) | 1.03 (0.83 - 1.27) |
| SIDIAP | 5,159 | 2,688 | 6.3 | 5.3 | <5 | 5 | < 0.79 (0.26 - 1.85) | 0.93 (0.30 - 2.18) |
| Meta-analysis | 390,965 | 1,058,872 | 277.3 | 922.9 | <77 | 577 | < 0.28 (0.22 - 0.35) | 0.63 (0.58 - 0.68) |
| Meta-analysis (w/o USOC) | 61,400 | 172,030 | 33.7 | 110.9 | <22 | 151 | < 0.65 (0.41 - 0.99) | 1.36 (1.15 - 1.60) |

##### 4.3.25 Venous thromboembolism

**Supplementary Table 366:** Exposures, outcomes and risk estimates for Venous thromboembolism in SGLT2I (male) (T) vs SU (male) (C) new-user cohorts under four different propensity score (PS) adjustment / time-at-risk (TAR) designs across data sources. We report total patient exposure time in 1,000 patient-year (KYr) units, uncalibrated and calibrated hazard ratios (HRs) and their 95% confidence intervals (CIs). Greyed out entries did not pass study diagnostics and are included only for completeness.

| | Patients | | Exposure (KYr) | | Outcomes | | Incidence (per KYr) | |
|---|---|---|---|---|---|---|---|---|
| | T | C | T | C | T | C | T | C |
| **PS-matching / on-treatment TAR** | | | | | | | | |
| CCAE | 20,188 | 45,819 | 17.2 | 38.2 | 69 | 224 | 4.01 (3.12 - 5.08) | 5.87 (5.12 - 6.69) |
| DAG | 5,202 | 928 | 4.3 | 0.9 | 27 | 6 | 6.25 (4.12 - 9.09) | 6.83 (2.51 - 14.9) |
| ODOD | 15,300 | 47,283 | 10.9 | 42.7 | 72 | 511 | 6.61 (5.17 - 8.32) | 12.0 (11.0 - 13.1) |
| MDCR | 1,698 | 7,910 | 1.4 | 7.4 | 12 | 113 | 8.29 (4.28 - 14.5) | 15.3 (12.6 - 18.3) |
| MDCD | 1,409 | 6,928 | 0.9 | 4.8 | 10 | 50 | 11.8 (5.64 - 21.6) | 10.3 (7.68 - 13.6) |
| Open Claims | 295,267 | 859,629 | 263.2 | 926.0 | 921 | 5,559 | 3.50 (3.28 - 3.73) | 6.00 (5.85 - 6.16) |
| OEHR | 17,845 | 71,845 | 5.6 | 39.9 | 39 | 358 | 6.93 (4.92 - 9.47) | 8.97 (8.07 - 9.95) |
| VA | 0 | 0 | 0.0 | 0.0 | 0 | 0 | | |
| SIDIAP | 4,441 | 1,186 | 6.4 | 2.2 | 24 | 11 | 3.74 (2.40 - 5.56) | 5.06 (2.53 - 9.05) |
| Meta-analysis | 361,350 | 1,041,528 | 309.9 | 1062.1 | 1,174 | 6,832 | 3.79 (3.57 - 4.01) | 6.43 (6.28 - 6.59) |
| Meta-analysis (w/o USOC) | 66,083 | 181,899 | 46.8 | 136.0 | 253 | 1,273 | 5.41 (4.76 - 6.12) | 9.36 (8.85 - 9.89) |
| **PS-stratification / on-treatment TAR** | | | | | | | | |
| CCAE | 23,622 | 46,448 | 19.6 | 38.5 | 77 | 231 | 3.92 (3.09 - 4.90) | 5.99 (5.24 - 6.82) |
| DAG | 5,411 | 1,343 | 4.5 | 1.3 | 27 | 14 | 6.06 (3.99 - 8.82) | 10.6 (5.78 - 17.7) |
| ODOD | 16,980 | 47,874 | 11.8 | 43.2 | 81 | 533 | 6.88 (5.46 - 8.55) | 12.3 (11.3 - 13.4) |
| MDCR | 2,048 | 8,160 | 1.6 | 7.6 | 15 | 120 | 9.27 (5.19 - 15.3) | 15.8 (13.1 - 18.9) |
| MDCD | 1,452 | 7,144 | 0.9 | 5.0 | 10 | 51 | 11.6 (5.55 - 21.3) | 10.2 (7.59 - 13.4) |
| Open Claims | 321,867 | 865,397 | 280.3 | 932.0 | 985 | 5,672 | 3.51 (3.30 - 3.74) | 6.09 (5.93 - 6.25) |
| OEHR | 19,746 | 74,195 | 6.3 | 41.4 | 42 | 402 | 6.71 (4.83 - 9.06) | 9.70 (8.78 - 10.7) |
| VA | 11,951 | 107,388 | 7.2 | 98.3 | 44 | 537 | 6.13 (4.45 - 8.23) | 5.46 (5.01 - 5.95) |
| SIDIAP | 5,024 | 2,636 | 6.9 | 6.1 | 26 | 22 | 3.79 (2.48 - 5.56) | 3.59 (2.25 - 5.44) |
| Meta-analysis | 382,215 | 1,033,914 | 318.0 | 1055.3 | 1,185 | 6,838 | 3.73 (3.52 - 3.95) | 6.48 (6.33 - 6.64) |
| Meta-analysis (w/o USOC) | 60,348 | 168,517 | 37.7 | 123.2 | 200 | 1,166 | 5.31 (4.60 - 6.10) | 9.46 (8.93 - 10.0) |
| **PS-matching / on-treatment TAR without escalation** | | | | | | | | |
| CCAE | 20,188 | 45,819 | 15.0 | 33.1 | 57 | 181 | 3.80 (2.88 - 4.93) | 5.46 (4.70 - 6.32) |
| DAG | 5,202 | 928 | 4.0 | 0.8 | 26 | 6 | 6.54 (4.27 - 9.58) | 7.40 (2.72 - 16.1) |
| ODOD | 15,300 | 47,283 | 9.7 | 38.2 | 58 | 453 | 6.00 (4.56 - 7.76) | 11.9 (10.8 - 13.0) |
| MDCR | 1,698 | 7,910 | 1.3 | 6.8 | 12 | 104 | 9.30 (4.80 - 16.2) | 15.3 (12.5 - 18.6) |
| MDCD | 1,409 | 6,928 | 0.7 | 4.0 | 9 | 39 | 12.5 (5.72 - 23.7) | 9.72 (6.91 - 13.3) |
| Open Claims | 295,267 | 859,629 | 223.2 | 785.9 | 752 | 4,759 | 3.37 (3.13 - 3.62) | 6.06 (5.88 - 6.23) |
| OEHR | 17,845 | 71,845 | 4.9 | 34.7 | 28 | 270 | 5.67 (3.77 - 8.19) | 7.78 (6.88 - 8.77) |
| VA | 0 | 0 | 0.0 | 0.0 | 0 | 0 | | |
| SIDIAP | 4,441 | 1,186 | 5.7 | 1.9 | 21 | 9 | 3.68 (2.28 - 5.62) | 4.75 (2.17 - 9.02) |
| Meta-analysis | 361,350 | 1,041,528 | 264.5 | 905.5 | 963 | 5,821 | 3.64 (3.41 - 3.88) | 6.43 (6.26 - 6.60) |
| Meta-analysis (w/o USOC) | 66,083 | 181,899 | 41.3 | 119.5 | 211 | 1,062 | 5.11 (4.44 - 5.85) | 8.88 (8.36 - 9.43) |
| **PS-stratification / on-treatment TAR without escalation** | | | | | | | | |
| CCAE | 23,622 | 46,448 | 17.2 | 33.5 | 64 | 188 | 3.73 (2.87 - 4.76) | 5.62 (4.84 - 6.48) |
| DAG | 5,411 | 1,343 | 4.1 | 1.2 | 26 | 14 | 6.34 (4.14 - 9.29) | 11.6 (6.34 - 19.5) |
| ODOD | 16,980 | 47,874 | 10.4 | 38.7 | 66 | 474 | 6.32 (4.89 - 8.04) | 12.2 (11.2 - 13.4) |
| MDCR | 2,048 | 8,160 | 1.4 | 7.0 | 15 | 111 | 10.4 (5.81 - 17.1) | 15.9 (13.1 - 19.2) |
| MDCD | 1,452 | 7,144 | 0.7 | 4.2 | 9 | 40 | 12.3 (5.62 - 23.3) | 9.63 (6.88 - 13.1) |
| Open Claims | 321,867 | 865,397 | 238.5 | 791.1 | 805 | 4,864 | 3.38 (3.15 - 3.62) | 6.15 (5.98 - 6.32) |
| OEHR | 19,746 | 74,195 | 5.5 | 36.0 | 31 | 297 | 5.62 (3.82 - 7.97) | 8.25 (7.34 - 9.25) |
| VA | 11,951 | 107,388 | 6.2 | 87.1 | 36 | 447 | 5.79 (4.06 - 8.02) | 5.13 (4.67 - 5.63) |
| SIDIAP | 5,024 | 2,636 | 6.1 | 5.2 | 23 | 19 | 3.76 (2.38 - 5.64) | 3.63 (2.19 - 5.67) |
| Meta-analysis | 382,215 | 1,033,914 | 271.6 | 899.3 | 966 | 5,823 | 3.56 (3.34 - 3.79) | 6.48 (6.31 - 6.64) |
| Meta-analysis (w/o USOC) | 60,348 | 168,517 | 33.1 | 108.1 | 161 | 959 | 4.86 (4.14 - 5.67) | 8.87 (8.32 - 9.45) |

##### 4.3.26 Vomiting

**Supplementary Table 367:** Exposures, outcomes and risk estimates for Vomiting in SGLT2I (male) (T) vs SU (male) (C) new-user cohorts under four different propensity score (PS) adjustment / time-at-risk (TAR) designs across data sources. We report total patient exposure time in 1,000 patient-year (KYr) units, uncalibrated and calibrated hazard ratios (HRs) and their 95% confidence intervals (CIs). Greyed out entries did not pass study diagnostics and are included only for completeness.

| | Patients | | Exposure (KYr) | | Outcomes | | Incidence (per KYr) | |
|---|---|---|---|---|---|---|---|---|
| | T | C | T | C | T | C | T | C |
| **PS-matching / on-treatment TAR** | | | | | | | | |
| CCAE | 19,268 | 43,723 | 16.2 | 36.3 | 277 | 588 | 17.1 (15.1 - 19.2) | 16.2 (14.9 - 17.5) |
| DAG | 5,175 | 921 | 4.3 | 0.9 | 33 | 8 | 7.64 (5.26 - 10.7) | 9.22 (3.98 - 18.2) |
| ODOD | 14,548 | 44,916 | 10.3 | 40.5 | 202 | 924 | 19.5 (16.9 - 22.4) | 22.8 (21.3 - 24.3) |
| MDCR | 1,684 | 7,707 | 1.4 | 7.2 | 28 | 177 | 19.5 (13.0 - 28.2) | 24.5 (21.0 - 28.4) |
| MDCD | 1,233 | 6,000 | 0.7 | 4.2 | 29 | 206 | 38.9 (26.0 - 55.8) | 49.0 (42.5 - 56.2) |
| Open Claims | 281,769 | 822,887 | 252.0 | 891.1 | 2,192 | 10,560 | 8.70 (8.34 - 9.07) | 11.9 (11.6 - 12.1) |
| OEHR | 17,371 | 70,182 | 5.5 | 38.8 | 91 | 713 | 16.6 (13.4 - 20.4) | 18.4 (17.1 - 19.8) |
| VA | 0 | 0 | 0.0 | 0.0 | 0 | 0 | | |
| SIDIAP | 4,404 | 1,181 | 6.4 | 2.2 | 21 | 8 | 3.26 (2.02 - 4.98) | 3.67 (1.58 - 7.22) |
| Meta-analysis | 345,452 | 997,517 | 296.9 | 1021.2 | 2,873 | 13,184 | 9.68 (9.32 - 10.0) | 12.9 (12.7 - 13.1) |
| Meta-analysis (w/o USOC) | 63,683 | 174,630 | 45.0 | 130.1 | 681 | 2,624 | 15.1 (14.0 - 16.3) | 20.2 (19.4 - 21.0) |
| **PS-stratification / on-treatment TAR** | | | | | | | | |
| CCAE | 22,530 | 44,307 | 18.5 | 36.7 | 308 | 599 | 16.6 (14.8 - 18.6) | 16.3 (15.0 - 17.7) |
| DAG | 5,395 | 1,334 | 4.5 | 1.3 | 34 | 12 | 7.63 (5.29 - 10.7) | 9.23 (4.77 - 16.1) |
| ODOD | 16,142 | 45,579 | 11.2 | 41.2 | 213 | 954 | 19.1 (16.6 - 21.8) | 23.2 (21.7 - 24.7) |
| MDCR | 2,025 | 7,919 | 1.6 | 7.4 | 29 | 191 | 18.1 (12.1 - 26.0) | 25.8 (22.3 - 29.8) |
| MDCD | 1,271 | 6,147 | 0.8 | 4.3 | 29 | 215 | 38.3 (25.6 - 55.0) | 49.8 (43.3 - 56.9) |
| Open Claims | 307,189 | 827,933 | 268.3 | 896.3 | 2,351 | 10,719 | 8.76 (8.41 - 9.12) | 12.0 (11.7 - 12.2) |
| OEHR | 19,279 | 72,408 | 6.1 | 40.2 | 99 | 760 | 16.2 (13.1 - 19.7) | 18.9 (17.6 - 20.3) |
| VA | 11,772 | 105,490 | 7.1 | 96.9 | 40 | 713 | 5.62 (4.02 - 7.66) | 7.36 (6.83 - 7.92) |
| SIDIAP | 4,990 | 2,623 | 6.9 | 6.1 | 22 | 28 | 3.20 (2.00 - 4.84) | 4.59 (3.05 - 6.64) |
| Meta-analysis | 365,140 | 990,227 | 304.1 | 1014.4 | 2,971 | 13,032 | 9.77 (9.42 - 10.1) | 12.8 (12.6 - 13.1) |
| Meta-analysis (w/o USOC) | 57,951 | 162,294 | 35.8 | 118.1 | 620 | 2,313 | 17.3 (16.0 - 18.7) | 19.6 (18.8 - 20.4) |
| **PS-matching / on-treatment TAR without escalation** | | | | | | | | |
| CCAE | 19,268 | 43,723 | 14.2 | 31.6 | 228 | 517 | 16.1 (14.1 - 18.3) | 16.4 (15.0 - 17.8) |
| DAG | 5,175 | 921 | 4.0 | 0.8 | 30 | 8 | 7.55 (5.10 - 10.8) | 10.1 (4.34 - 19.8) |
| ODOD | 14,548 | 44,916 | 9.2 | 36.3 | 171 | 816 | 18.6 (15.9 - 21.6) | 22.4 (20.9 - 24.0) |
| MDCR | 1,684 | 7,707 | 1.3 | 6.6 | 26 | 150 | 20.3 (13.3 - 29.8) | 22.6 (19.2 - 26.6) |
| MDCD | 1,233 | 6,000 | 0.6 | 3.5 | 27 | 173 | 42.8 (28.2 - 62.3) | 49.0 (42.0 - 56.9) |
| Open Claims | 281,769 | 822,887 | 214.1 | 758.0 | 1,724 | 8,742 | 8.05 (7.68 - 8.44) | 11.5 (11.3 - 11.8) |
| OEHR | 17,371 | 70,182 | 4.8 | 33.8 | 79 | 543 | 16.4 (13.0 - 20.4) | 16.1 (14.7 - 17.5) |
| VA | 0 | 0 | 0.0 | 0.0 | 0 | 0 | | |
| SIDIAP | 4,404 | 1,181 | 5.7 | 1.9 | 19 | 6 | 3.32 (2.00 - 5.19) | 3.17 (1.16 - 6.89) |
| Meta-analysis | 345,452 | 997,517 | 253.9 | 872.6 | 2,304 | 10,955 | 9.07 (8.71 - 9.45) | 12.6 (12.3 - 12.8) |
| Meta-analysis (w/o USOC) | 63,683 | 174,630 | 39.8 | 114.6 | 580 | 2,213 | 14.6 (13.4 - 15.8) | 19.3 (18.5 - 20.1) |
| **PS-stratification / on-treatment TAR without escalation** | | | | | | | | |
| CCAE | 22,530 | 44,307 | 16.2 | 31.9 | 255 | 528 | 15.7 (13.8 - 17.8) | 16.5 (15.2 - 18.0) |
| DAG | 5,395 | 1,334 | 4.1 | 1.2 | 31 | 12 | 7.57 (5.14 - 10.7) | 10.1 (5.22 - 17.6) |
| ODOD | 16,142 | 45,579 | 9.9 | 36.9 | 179 | 841 | 18.0 (15.5 - 20.9) | 22.8 (21.3 - 24.4) |
| MDCR | 2,025 | 7,919 | 1.4 | 6.8 | 27 | 163 | 18.8 (12.4 - 27.4) | 24.0 (20.5 - 28.0) |
| MDCD | 1,271 | 6,147 | 0.6 | 3.6 | 27 | 181 | 42.2 (27.8 - 61.3) | 49.9 (42.9 - 57.7) |
| Open Claims | 307,189 | 827,933 | 228.8 | 762.5 | 1,867 | 8,881 | 8.16 (7.79 - 8.54) | 11.6 (11.4 - 11.9) |
| OEHR | 19,279 | 72,408 | 5.4 | 35.1 | 84 | 576 | 15.5 (12.4 - 19.2) | 16.4 (15.1 - 17.8) |
| VA | 11,772 | 105,490 | 6.2 | 85.9 | 31 | 587 | 5.03 (3.42 - 7.14) | 6.83 (6.29 - 7.41) |
| SIDIAP | 4,990 | 2,623 | 6.1 | 5.2 | 20 | 24 | 3.26 (1.99 - 5.04) | 4.61 (2.95 - 6.86) |
| Meta-analysis | 365,140 | 990,227 | 260.4 | 866.4 | 2,385 | 10,826 | 9.16 (8.80 - 9.54) | 12.5 (12.3 - 12.7) |
| Meta-analysis (w/o USOC) | 57,951 | 162,294 | 31.6 | 103.9 | 518 | 1,945 | 16.4 (15.0 - 17.9) | 18.7 (17.9 - 19.6) |

#### 4.4 GLP1RA (male) vs DPP4I (male)

##### 4.4.1 3-point MACE

**Supplementary Table 368:** Exposures, outcomes and risk estimates for 3-point MACE in GLP1RA (male) (T) vs DPP4I (male) (C) new-user cohorts under four different propensity score (PS) adjustment / time-at-risk (TAR) designs across data sources. We report total patient exposure time in 1,000 patient-year (KYr) units, uncalibrated and calibrated hazard ratios (HRs) and their 95% confidence intervals (CIs). Greyed out entries did not pass study diagnostics and are included only for completeness.

| | Patients | | Exposure (KYr) | | Outcomes | | Incidence (per KYr) | |
|---|---|---|---|---|---|---|---|---|
| | T | C | T | C | T | C | T | C |
| **PS-matching / on-treatment TAR** | | | | | | | | |
| CCAE | 10,198 | 50,982 | 7.0 | 44.6 | 56 | 383 | 7.95 (6.01 - 10.3) | 8.60 (7.76 - 9.50) |
| ODOD | 6,134 | 28,677 | 3.8 | 24.1 | 37 | 435 | 9.64 (6.79 - 13.3) | 18.0 (16.4 - 19.8) |
| MDCR | 800 | 8,308 | 0.5 | 8.7 | 11 | 233 | 20.1 (10.0 - 35.9) | 26.8 (23.4 - 30.4) |
| Open Claims | 109,355 | 401,048 | 84.8 | 384.3 | 561 | 4,126 | 6.62 (6.08 - 7.19) | 10.7 (10.4 - 11.1) |
| OEHR | 7,631 | 37,284 | 2.3 | 18.5 | 23 | 204 | 10.2 (6.47 - 15.3) | 11.0 (9.54 - 12.6) |
| VA | 1,153 | 17,084 | 0.9 | 14.5 | <10 | 67 | < 11.7 (5.60 - 21.5) | 4.62 (3.58 - 5.87) |
| Meta-analysis | 135,271 | 543,383 | 99.3 | 494.7 | <698 | 5,448 | < 7.03 (6.52 - 7.57) | 11.0 (10.7 - 11.3) |
| Meta-analysis (w/o USOC) | 25,916 | 142,335 | 14.5 | 110.4 | <137 | 1,322 | < 9.42 (7.91 - 11.1) | 12.0 (11.3 - 12.6) |
| **PS-stratification / on-treatment TAR** | | | | | | | | |
| CCAE | 12,059 | 51,105 | 7.9 | 44.7 | 60 | 383 | 7.63 (5.82 - 9.82) | 8.57 (7.74 - 9.48) |
| ODOD | 8,175 | 29,196 | 4.7 | 24.5 | 42 | 458 | 8.96 (6.46 - 12.1) | 18.7 (17.0 - 20.5) |
| MDCR | 893 | 8,780 | 0.6 | 9.2 | 11 | 259 | 19.0 (9.49 - 34.0) | 28.3 (24.9 - 31.9) |
| Open Claims | 147,591 | 402,784 | 101.8 | 385.9 | 647 | 4,214 | 6.36 (5.88 - 6.87) | 10.9 (10.6 - 11.3) |
| OEHR | 8,896 | 37,369 | 2.5 | 18.6 | 26 | 209 | 10.3 (6.71 - 15.0) | 11.2 (9.77 - 12.9) |
| VA | 1,334 | 24,642 | 0.9 | 23.0 | <10 | 124 | < 10.7 (5.15 - 19.8) | 5.38 (4.48 - 6.42) |
| Meta-analysis | 176,721 | 520,454 | 116.9 | 473.7 | 775 | 5,264 | 6.63 (6.17 - 7.12) | 11.1 (10.8 - 11.4) |
| Meta-analysis (w/o USOC) | 29,130 | 117,670 | 15.1 | 87.8 | 128 | 1,050 | 8.49 (7.08 - 10.1) | 12.0 (11.2 - 12.7) |
| **PS-matching / on-treatment TAR without escalation** | | | | | | | | |
| CCAE | 10,198 | 50,982 | 6.1 | 36.1 | 49 | 306 | 8.04 (5.94 - 10.6) | 8.49 (7.56 - 9.49) |
| ODOD | 6,134 | 28,677 | 3.4 | 19.7 | 30 | 369 | 8.86 (5.98 - 12.7) | 18.7 (16.9 - 20.8) |
| MDCR | 800 | 8,308 | 0.5 | 7.4 | 7 | 191 | 14.4 (5.79 - 29.7) | 25.7 (22.2 - 29.6) |
| Open Claims | 109,355 | 401,048 | 73.2 | 298.0 | 464 | 3,205 | 6.34 (5.78 - 6.95) | 10.8 (10.4 - 11.1) |
| OEHR | 7,631 | 37,284 | 1.9 | 15.1 | 19 | 144 | 9.76 (5.88 - 15.2) | 9.54 (8.04 - 11.2) |
| VA | 1,153 | 17,084 | 0.7 | 11.8 | <10 | 46 | < 15.0 (7.18 - 27.5) | 3.89 (2.85 - 5.19) |
| Meta-analysis | 134,118 | 526,299 | 85.1 | 376.3 | 569 | 4,215 | 6.69 (6.15 - 7.26) | 11.2 (10.9 - 11.5) |
| Meta-analysis (w/o USOC) | 24,763 | 125,251 | 11.9 | 78.3 | 105 | 1,010 | 8.81 (7.21 - 10.7) | 12.9 (12.1 - 13.7) |
| **PS-stratification / on-treatment TAR without escalation** | | | | | | | | |
| CCAE | 12,059 | 51,105 | 6.9 | 36.2 | 53 | 306 | 7.73 (5.79 - 10.1) | 8.46 (7.54 - 9.47) |
| ODOD | 8,175 | 29,196 | 4.2 | 20.0 | 35 | 390 | 8.40 (5.85 - 11.7) | 19.5 (17.6 - 21.5) |
| MDCR | 893 | 8,780 | 0.5 | 7.8 | 7 | 215 | 13.6 (5.47 - 28.0) | 27.4 (23.9 - 31.3) |
| Open Claims | 147,591 | 402,784 | 89.0 | 299.3 | 538 | 3,277 | 6.04 (5.54 - 6.58) | 10.9 (10.6 - 11.3) |
| OEHR | 8,896 | 37,369 | 2.2 | 15.1 | 22 | 147 | 9.98 (6.26 - 15.1) | 9.71 (8.21 - 11.4) |
| VA | 1,334 | 24,642 | 0.7 | 18.7 | <10 | 86 | < 13.6 (6.55 - 25.1) | 4.59 (3.67 - 5.67) |
| Meta-analysis | 176,721 | 520,454 | 102.2 | 370.6 | 648 | 4,120 | 6.34 (5.86 - 6.85) | 11.1 (10.8 - 11.5) |
| Meta-analysis (w/o USOC) | 29,130 | 117,670 | 13.2 | 71.3 | 110 | 843 | 8.32 (6.84 - 10.0) | 11.8 (11.0 - 12.6) |

##### 4.4.2 4-point MACE

**Supplementary Table 369:** Exposures, outcomes and risk estimates for 4-point MACE in GLP1RA (male) (T) vs DPP4I (male) (C) new-user cohorts under four different propensity score (PS) adjustment / time-at-risk (TAR) designs across data sources. We report total patient exposure time in 1,000 patient-year (KYr) units, uncalibrated and calibrated hazard ratios (HRs) and their 95% confidence intervals (CIs). Greyed out entries did not pass study diagnostics and are included only for completeness.

| | Patients | | Exposure (KYr) | | Outcomes | | Incidence (per KYr) | |
|---|---|---|---|---|---|---|---|---|
| | T | C | T | C | T | C | T | C |
| **PS-matching / on-treatment TAR** | | | | | | | | |
| CCAE | 10,031 | 50,301 | 6.9 | 43.9 | 87 | 553 | 12.6 (10.1 - 15.5) | 12.6 (11.6 - 13.7) |
| ODOD | 5,924 | 27,592 | 3.7 | 23.1 | 66 | 650 | 17.8 (13.8 - 22.7) | 28.1 (26.0 - 30.4) |
| MDCR | 758 | 7,793 | 0.5 | 8.1 | 21 | 365 | 40.3 (25.0 - 61.7) | 45.1 (40.6 - 50.0) |
| Open Claims | 106,379 | 388,374 | 82.6 | 371.4 | 852 | 6,449 | 10.3 (9.64 - 11.0) | 17.4 (16.9 - 17.8) |
| OEHR | 7,520 | 36,606 | 2.2 | 18.0 | 28 | 361 | 12.6 (8.40 - 18.3) | 20.1 (18.0 - 22.2) |
| VA | 1,114 | 16,661 | 0.8 | 14.1 | <10 | 158 | < 12.1 (5.80 - 22.3) | 11.2 (9.55 - 13.1) |
| Meta-analysis | 131,726 | 527,327 | 96.8 | 478.5 | <1,064 | 8,536 | < 11.0 (10.3 - 11.7) | 17.8 (17.5 - 18.2) |
| Meta-analysis (w/o USOC) | 25,347 | 138,953 | 14.2 | 107.1 | <212 | 2,087 | < 14.9 (13.0 - 17.1) | 19.5 (18.7 - 20.3) |
| **PS-stratification / on-treatment TAR** | | | | | | | | |
| CCAE | 11,840 | 50,414 | 7.7 | 44.0 | 96 | 553 | 12.4 (10.1 - 15.2) | 12.6 (11.6 - 13.7) |
| ODOD | 7,889 | 28,074 | 4.5 | 23.5 | 77 | 680 | 17.0 (13.4 - 21.3) | 28.9 (26.8 - 31.2) |
| MDCR | 847 | 8,212 | 0.5 | 8.5 | 22 | 407 | 40.2 (25.2 - 60.8) | 47.9 (43.3 - 52.7) |
| Open Claims | 143,665 | 389,938 | 99.2 | 372.9 | 977 | 6,586 | 9.85 (9.24 - 10.5) | 17.7 (17.2 - 18.1) |
| OEHR | 8,765 | 36,683 | 2.5 | 18.0 | 33 | 371 | 13.3 (9.13 - 18.6) | 20.6 (18.5 - 22.8) |
| VA | 1,281 | 24,006 | 0.9 | 22.3 | <10 | 268 | < 11.1 (5.33 - 20.5) | 12.0 (10.6 - 13.5) |
| Meta-analysis | 172,159 | 505,109 | 113.9 | 458.4 | 1,183 | 8,190 | 10.4 (9.80 - 11.0) | 17.9 (17.5 - 18.3) |
| Meta-analysis (w/o USOC) | 28,494 | 115,171 | 14.7 | 85.5 | 206 | 1,604 | 14.0 (12.1 - 16.0) | 18.8 (17.8 - 19.7) |
| **PS-matching / on-treatment TAR without escalation** | | | | | | | | |
| CCAE | 10,031 | 50,301 | 6.0 | 35.5 | 77 | 441 | 12.8 (10.1 - 16.0) | 12.4 (11.3 - 13.6) |
| ODOD | 5,924 | 27,592 | 3.3 | 18.9 | 57 | 545 | 17.5 (13.2 - 22.6) | 28.8 (26.5 - 31.4) |
| MDCR | 758 | 7,793 | 0.5 | 6.9 | 17 | 310 | 37.0 (21.6 - 59.3) | 44.9 (40.0 - 50.1) |
| Open Claims | 106,379 | 388,374 | 71.3 | 288.2 | 718 | 5,009 | 10.1 (9.35 - 10.8) | 17.4 (16.9 - 17.9) |
| OEHR | 7,520 | 36,606 | 1.9 | 14.7 | 23 | 261 | 12.0 (7.62 - 18.0) | 17.7 (15.6 - 20.0) |
| VA | 1,114 | 16,661 | 0.6 | 11.5 | <10 | 118 | < 15.6 (7.47 - 28.6) | 10.2 (8.48 - 12.3) |
| Meta-analysis | 131,726 | 527,327 | 83.6 | 375.7 | <902 | 6,684 | < 10.8 (10.1 - 11.5) | 17.8 (17.4 - 18.2) |
| Meta-analysis (w/o USOC) | 25,347 | 138,953 | 12.3 | 87.5 | <184 | 1,675 | < 15.0 (12.9 - 17.3) | 19.1 (18.2 - 20.1) |
| **PS-stratification / on-treatment TAR without escalation** | | | | | | | | |
| CCAE | 11,840 | 50,414 | 6.7 | 35.6 | 85 | 441 | 12.6 (10.1 - 15.6) | 12.4 (11.3 - 13.6) |
| ODOD | 7,889 | 28,074 | 4.0 | 19.2 | 67 | 573 | 16.7 (12.9 - 21.2) | 29.8 (27.4 - 32.3) |
| MDCR | 847 | 8,212 | 0.5 | 7.3 | 18 | 349 | 37.0 (21.9 - 58.5) | 48.0 (43.1 - 53.3) |
| Open Claims | 143,665 | 389,938 | 86.8 | 289.4 | 830 | 5,120 | 9.57 (8.93 - 10.2) | 17.7 (17.2 - 18.2) |
| OEHR | 8,765 | 36,683 | 2.2 | 14.8 | 28 | 267 | 12.9 (8.59 - 18.7) | 18.1 (16.0 - 20.4) |
| VA | 1,281 | 24,006 | 0.7 | 18.2 | <10 | 197 | < 14.2 (6.80 - 26.1) | 10.8 (9.38 - 12.5) |
| Meta-analysis | 172,159 | 505,109 | 99.7 | 359.0 | 1,010 | 6,401 | 10.1 (9.52 - 10.8) | 17.8 (17.4 - 18.3) |
| Meta-analysis (w/o USOC) | 28,494 | 115,171 | 12.9 | 69.6 | 180 | 1,281 | 13.9 (12.0 - 16.1) | 18.4 (17.4 - 19.4) |

##### 4.4.3 Acute myocardial infarction

**Supplementary Table 370:** Exposures, outcomes and risk estimates for Acute myocardial infarction in GLP1RA (male) (T) vs DPP4I (male) (C) new-user cohorts under four different propensity score (PS) adjustment / time-at-risk (TAR) designs across data sources. We report total patient exposure time in 1,000 patient-year (KYr) units, uncalibrated and calibrated hazard ratios (HRs) and their 95% confidence intervals (CIs). Greyed out entries did not pass study diagnostics and are included only for completeness.

| | Patients | | Exposure (KYr) | | Outcomes | | Incidence (per KYr) | |
|---|---|---|---|---|---|---|---|---|
| | T | C | T | C | T | C | T | C |
| **PS-matching / on-treatment TAR** | | | | | | | | |
| CCAE | 10,327 | 51,541 | 7.2 | 45.1 | 35 | 230 | 4.89 (3.41 - 6.80) | 5.10 (4.46 - 5.80) |
| ODOD | 6,255 | 29,323 | 3.9 | 24.7 | 17 | 220 | 4.34 (2.53 - 6.96) | 8.90 (7.76 - 10.2) |
| MDCR | 826 | 8,574 | 0.6 | 9.0 | 7 | 107 | 12.2 (4.90 - 25.1) | 11.8 (9.69 - 14.3) |
| Open Claims | 111,826 | 412,257 | 86.5 | 395.3 | 301 | 2,145 | 3.48 (3.10 - 3.89) | 5.43 (5.20 - 5.66) |
| OEHR | 7,690 | 37,619 | 2.3 | 18.8 | 7 | 113 | 3.07 (1.24 - 6.33) | 6.02 (4.96 - 7.24) |
| VA | 1,172 | 17,282 | 0.9 | 14.7 | <10 | 35 | < 11.5 (5.54 - 21.2) | 2.39 (1.66 - 3.32) |
| Meta-analysis | 136,924 | 539,314 | 100.5 | 492.9 | 367 | 2,815 | 3.65 (3.29 - 4.05) | 5.71 (5.50 - 5.93) |
| Meta-analysis (w/o USOC) | 25,098 | 127,057 | 13.9 | 97.6 | 66 | 670 | 4.74 (3.67 - 6.03) | 6.86 (6.35 - 7.40) |
| **PS-stratification / on-treatment TAR** | | | | | | | | |
| CCAE | 12,209 | 51,682 | 8.0 | 45.2 | 38 | 231 | 4.76 (3.37 - 6.53) | 5.11 (4.47 - 5.81) |
| ODOD | 8,318 | 29,940 | 4.8 | 25.2 | 19 | 235 | 3.99 (2.40 - 6.23) | 9.31 (8.16 - 10.6) |
| MDCR | 923 | 9,130 | 0.6 | 9.6 | 7 | 120 | 11.6 (4.67 - 23.9) | 12.5 (10.4 - 15.0) |
| Open Claims | 150,626 | 414,162 | 103.8 | 397.0 | 346 | 2,192 | 3.33 (2.99 - 3.70) | 5.52 (5.29 - 5.76) |
| OEHR | 8,966 | 37,708 | 2.6 | 18.8 | 10 | 116 | 3.91 (1.87 - 7.19) | 6.17 (5.09 - 7.39) |
| VA | 1,360 | 25,018 | 0.9 | 23.4 | <10 | 65 | < 10.6 (5.09 - 19.5) | 2.78 (2.14 - 3.54) |
| Meta-analysis | 180,119 | 533,492 | 119.1 | 486.3 | 413 | 2,774 | 3.47 (3.14 - 3.82) | 5.70 (5.49 - 5.92) |
| Meta-analysis (w/o USOC) | 29,493 | 119,330 | 15.3 | 89.3 | 67 | 582 | 4.38 (3.39 - 5.56) | 6.52 (6.00 - 7.07) |
| **PS-matching / on-treatment TAR without escalation** | | | | | | | | |
| CCAE | 10,327 | 51,541 | 6.2 | 36.5 | 32 | 181 | 5.17 (3.54 - 7.30) | 4.96 (4.26 - 5.74) |
| ODOD | 6,255 | 29,323 | 3.5 | 20.2 | 14 | 178 | 4.06 (2.22 - 6.80) | 8.81 (7.57 - 10.2) |
| MDCR | 826 | 8,574 | 0.5 | 7.7 | <5 | 88 | < 9.83 (3.19 - 22.9) | 11.4 (9.16 - 14.1) |
| Open Claims | 111,826 | 412,257 | 74.7 | 306.4 | 251 | 1,648 | 3.36 (2.96 - 3.80) | 5.38 (5.12 - 5.64) |
| OEHR | 7,690 | 37,619 | 2.0 | 15.3 | 6 | 76 | 3.06 (1.12 - 6.65) | 4.98 (3.92 - 6.23) |
| VA | 1,172 | 17,282 | 0.7 | 12.0 | <10 | 26 | < 14.8 (7.08 - 27.1) | 2.17 (1.42 - 3.18) |
| Meta-analysis | 136,924 | 539,314 | 86.8 | 386.1 | <308 | 2,171 | < 3.55 (3.16 - 3.97) | 5.62 (5.39 - 5.87) |
| Meta-analysis (w/o USOC) | 25,098 | 127,057 | 12.1 | 79.7 | <57 | 523 | < 4.70 (3.56 - 6.10) | 6.56 (6.01 - 7.15) |
| **PS-stratification / on-treatment TAR without escalation** | | | | | | | | |
| CCAE | 12,209 | 51,682 | 7.0 | 36.6 | 35 | 182 | 5.03 (3.51 - 7.00) | 4.97 (4.28 - 5.75) |
| ODOD | 8,318 | 29,940 | 4.2 | 20.6 | 16 | 190 | 3.78 (2.16 - 6.14) | 9.21 (7.95 - 10.6) |
| MDCR | 923 | 9,130 | 0.5 | 8.2 | <5 | 100 | < 9.32 (3.02 - 21.7) | 12.2 (9.93 - 14.9) |
| Open Claims | 150,626 | 414,162 | 90.8 | 307.8 | 289 | 1,685 | 3.18 (2.83 - 3.57) | 5.48 (5.22 - 5.74) |
| OEHR | 8,966 | 37,708 | 2.2 | 15.3 | 9 | 78 | 4.05 (1.85 - 7.69) | 5.10 (4.03 - 6.36) |
| VA | 1,360 | 25,018 | 0.7 | 19.0 | <10 | 50 | < 13.5 (6.45 - 24.7) | 2.63 (1.95 - 3.47) |
| Meta-analysis | 180,119 | 533,492 | 104.2 | 380.3 | 349 | 2,135 | 3.35 (3.01 - 3.72) | 5.61 (5.38 - 5.86) |
| Meta-analysis (w/o USOC) | 29,493 | 119,330 | 13.4 | 72.5 | 60 | 450 | 4.48 (3.41 - 5.76) | 6.20 (5.64 - 6.80) |

##### 4.4.4 Acute renal failure

**Supplementary Table 371:** Exposures, outcomes and risk estimates for Acute renal failure in GLP1RA (male) (T) vs DPP4I (male) (C) new-user cohorts under four different propensity score (PS) adjustment / time-at-risk (TAR) designs across data sources. We report total patient exposure time in 1,000 patient-year (KYr) units, uncalibrated and calibrated hazard ratios (HRs) and their 95% confidence intervals (CIs). Greyed out entries did not pass study diagnostics and are included only for completeness.

| | Patients | | Exposure (KYr) | | Outcomes | | Incidence (per KYr) | |
|---|---|---|---|---|---|---|---|---|
| | T | C | T | C | T | C | T | C |
| **PS-matching / on-treatment TAR** | | | | | | | | |
| CCAE | 10,390 | 51,877 | 7.2 | 45.5 | 54 | 385 | 7.54 (5.66 - 9.83) | 8.47 (7.64 - 9.36) |
| ODOD | 6,146 | 28,625 | 3.8 | 24.0 | 76 | 624 | 19.8 (15.6 - 24.8) | 26.0 (24.0 - 28.1) |
| MDCR | 816 | 8,402 | 0.6 | 8.7 | 23 | 301 | 40.9 (26.0 - 61.4) | 34.5 (30.7 - 38.6) |
| Open Claims | 111,443 | 410,208 | 86.2 | 392.3 | 773 | 5,862 | 8.97 (8.35 - 9.62) | 14.9 (14.6 - 15.3) |
| OEHR | 7,691 | 37,538 | 2.3 | 18.6 | 30 | 334 | 13.2 (8.89 - 18.8) | 18.0 (16.1 - 20.0) |
| VA | 1,157 | 17,032 | 0.9 | 14.4 | <10 | 181 | < 11.7 (5.63 - 21.6) | 12.6 (10.8 - 14.5) |
| Meta-analysis | 137,643 | 553,682 | 100.9 | 503.5 | <966 | 7,687 | < 9.58 (8.98 - 10.2) | 15.3 (14.9 - 15.6) |
| Meta-analysis (w/o USOC) | 26,200 | 143,474 | 14.7 | 111.2 | <193 | 1,825 | < 13.1 (11.4 - 15.1) | 16.4 (15.7 - 17.2) |
| **PS-stratification / on-treatment TAR** | | | | | | | | |
| CCAE | 12,261 | 51,990 | 8.0 | 45.6 | 61 | 388 | 7.62 (5.83 - 9.79) | 8.51 (7.69 - 9.40) |
| ODOD | 8,198 | 29,178 | 4.7 | 24.5 | 91 | 659 | 19.5 (15.7 - 23.9) | 26.9 (24.9 - 29.1) |
| MDCR | 910 | 8,907 | 0.6 | 9.2 | 23 | 339 | 38.8 (24.6 - 58.3) | 36.8 (33.0 - 40.9) |
| Open Claims | 150,166 | 411,910 | 103.4 | 394.0 | 898 | 5,966 | 8.68 (8.12 - 9.27) | 15.1 (14.8 - 15.5) |
| OEHR | 8,969 | 37,635 | 2.6 | 18.6 | 34 | 341 | 13.3 (9.20 - 18.6) | 18.3 (16.4 - 20.3) |
| VA | 1,342 | 24,628 | 0.9 | 22.9 | <10 | 295 | < 10.7 (5.15 - 19.7) | 12.9 (11.4 - 14.4) |
| Meta-analysis | 179,594 | 530,713 | 118.7 | 482.7 | 1,084 | 7,354 | 9.13 (8.60 - 9.70) | 15.2 (14.9 - 15.6) |
| Meta-analysis (w/o USOC) | 29,428 | 118,803 | 15.2 | 88.7 | 186 | 1,388 | 12.2 (10.5 - 14.1) | 15.7 (14.8 - 16.5) |
| **PS-matching / on-treatment TAR without escalation** | | | | | | | | |
| CCAE | 10,390 | 51,877 | 6.2 | 36.8 | 46 | 304 | 7.40 (5.42 - 9.87) | 8.26 (7.36 - 9.25) |
| ODOD | 6,146 | 28,625 | 3.4 | 19.6 | 68 | 499 | 20.1 (15.6 - 25.5) | 25.4 (23.2 - 27.7) |
| MDCR | 816 | 8,402 | 0.5 | 7.5 | 19 | 249 | 38.2 (23.0 - 59.7) | 33.3 (29.3 - 37.8) |
| Open Claims | 111,443 | 410,208 | 74.4 | 304.4 | 639 | 4,547 | 8.59 (7.94 - 9.28) | 14.9 (14.5 - 15.4) |
| OEHR | 7,691 | 37,538 | 2.0 | 15.2 | 25 | 229 | 12.7 (8.23 - 18.8) | 15.1 (13.2 - 17.2) |
| VA | 1,157 | 17,032 | 0.7 | 11.8 | <10 | 132 | < 15.0 (7.17 - 27.5) | 11.2 (9.39 - 13.3) |
| Meta-analysis | 137,643 | 553,682 | 87.1 | 395.2 | <807 | 5,960 | < 9.26 (8.64 - 9.93) | 15.1 (14.7 - 15.5) |
| Meta-analysis (w/o USOC) | 26,200 | 143,474 | 12.7 | 90.9 | <168 | 1,413 | < 13.2 (11.3 - 15.4) | 15.5 (14.7 - 16.4) |
| **PS-stratification / on-treatment TAR without escalation** | | | | | | | | |
| CCAE | 12,261 | 51,990 | 7.0 | 36.9 | 53 | 307 | 7.59 (5.69 - 9.93) | 8.32 (7.42 - 9.31) |
| ODOD | 8,198 | 29,178 | 4.2 | 20.0 | 80 | 531 | 19.3 (15.3 - 24.0) | 26.5 (24.3 - 28.9) |
| MDCR | 910 | 8,907 | 0.5 | 7.9 | 19 | 283 | 36.2 (21.8 - 56.5) | 35.9 (31.8 - 40.3) |
| Open Claims | 150,166 | 411,910 | 90.4 | 305.7 | 753 | 4,632 | 8.33 (7.74 - 8.94) | 15.2 (14.7 - 15.6) |
| OEHR | 8,969 | 37,635 | 2.2 | 15.2 | 29 | 234 | 13.0 (8.72 - 18.7) | 15.4 (13.4 - 17.4) |
| VA | 1,342 | 24,628 | 0.7 | 18.6 | <10 | 212 | < 13.6 (6.51 - 25.0) | 11.4 (9.90 - 13.0) |
| Meta-analysis | 179,594 | 530,713 | 103.8 | 377.8 | 915 | 5,704 | 8.82 (8.25 - 9.41) | 15.1 (14.7 - 15.5) |
| Meta-analysis (w/o USOC) | 29,428 | 118,803 | 13.4 | 72.1 | 162 | 1,072 | 12.1 (10.3 - 14.1) | 14.9 (14.0 - 15.8) |

##### 4.4.5 Glycemic control

**Supplementary Table 372:** Exposures, outcomes and risk estimates for Glycemic control in GLP1RA (male) (T) vs DPP4I (male) (C) new-user cohorts under four different propensity score (PS) adjustment / time-at-risk (TAR) designs across data sources. We report total patient exposure time in 1,000 patient-year (KYr) units, uncalibrated and calibrated hazard ratios (HRs) and their 95% confidence intervals (CIs). Greyed out entries did not pass study diagnostics and are included only for completeness.

| | Patients | | Exposure (KYr) | | Outcomes | | Incidence (per KYr) | |
|---|---|---|---|---|---|---|---|---|
| | T | C | T | C | T | C | T | C |
| **PS-matching / on-treatment TAR** | | | | | | | | |
| CCAE | 10,544 | 52,228 | 7.3 | 45.9 | 6 | 130 | 0.82 (0.30 - 1.79) | 2.84 (2.37 - 3.37) |
| ODOD | 4,318 | 20,609 | 2.4 | 15.5 | 277 | 1,651 | 113 (100 - 127) | 106 (101 - 112) |
| MDCR | 855 | 8,869 | 0.6 | 9.4 | <5 | 11 | < 8.39 (2.72 - 19.6) | 1.17 (0.59 - 2.10) |
| Open Claims | 115,660 | 427,485 | 89.4 | 411.0 | 0 | 0 | | |
| OEHR | 7,806 | 38,195 | 2.3 | 19.2 | 0 | 0 | | |
| VA | 1,158 | 16,768 | 0.9 | 14.2 | 0 | 12 | | 0.84 (0.44 - 1.47) |
| Meta-analysis | 14,862 | 72,837 | 9.7 | 61.4 | 283 | 1,781 | 29.1 (25.8 - 32.6) | 29.0 (27.7 - 30.4) |
| Meta-analysis (w/o USOC) | 14,862 | 72,837 | 9.7 | 61.4 | 283 | 1,781 | 29.1 (25.8 - 32.6) | 29.0 (27.7 - 30.4) |
| **PS-stratification / on-treatment TAR** | | | | | | | | |
| CCAE | 12,456 | 52,351 | 8.1 | 46.0 | 6 | 131 | 0.74 (0.27 - 1.61) | 2.85 (2.38 - 3.38) |
| ODOD | 5,649 | 20,906 | 3.0 | 15.7 | 329 | 1,677 | 111 (98.9 - 123) | 107 (102 - 112) |
| MDCR | 952 | 9,515 | 0.6 | 10.0 | <5 | 12 | < 8.01 (2.60 - 18.7) | 1.20 (0.62 - 2.09) |
| Open Claims | 155,612 | 429,807 | 107.2 | 413.1 | 0 | 0 | | |
| OEHR | 9,105 | 38,288 | 2.6 | 19.2 | 0 | 0 | | |
| VA | 1,341 | 24,844 | 0.9 | 23.3 | 0 | 21 | | 0.90 (0.56 - 1.38) |
| Meta-analysis | 18,105 | 73,257 | 11.1 | 61.7 | 335 | 1,808 | 30.2 (27.0 - 33.6) | 29.3 (28.0 - 30.7) |
| Meta-analysis (w/o USOC) | 18,105 | 73,257 | 11.1 | 61.7 | 335 | 1,808 | 30.2 (27.0 - 33.6) | 29.3 (28.0 - 30.7) |
| **PS-matching / on-treatment TAR without escalation** | | | | | | | | |
| CCAE | 10,544 | 52,228 | 6.3 | 37.0 | 5 | 107 | 0.79 (0.26 - 1.85) | 2.89 (2.37 - 3.49) |
| ODOD | 4,318 | 20,609 | 2.1 | 12.7 | 249 | 1,340 | 116 (102 - 131) | 106 (100.0 - 111) |
| MDCR | 855 | 8,869 | 0.5 | 8.0 | <5 | 10 | < 9.48 (3.08 - 22.1) | 1.25 (0.60 - 2.30) |
| Open Claims | 115,660 | 427,485 | 77.1 | 318.3 | 0 | 0 | | |
| OEHR | 7,806 | 38,195 | 2.0 | 15.5 | 0 | 0 | | |
| VA | 1,158 | 16,768 | 0.7 | 11.6 | 0 | 11 | | 0.95 (0.47 - 1.69) |
| Meta-analysis | 14,862 | 72,837 | 8.5 | 49.7 | 254 | 1,447 | 30.0 (26.5 - 34.0) | 29.1 (27.6 - 30.6) |
| Meta-analysis (w/o USOC) | 14,862 | 72,837 | 8.5 | 49.7 | 254 | 1,447 | 30.0 (26.5 - 34.0) | 29.1 (27.6 - 30.6) |
| **PS-stratification / on-treatment TAR without escalation** | | | | | | | | |
| CCAE | 12,456 | 52,351 | 7.1 | 37.1 | 5 | 108 | 0.71 (0.23 - 1.65) | 2.91 (2.39 - 3.51) |
| ODOD | 5,649 | 20,906 | 2.6 | 12.9 | 297 | 1,360 | 113 (101 - 127) | 106 (100 - 112) |
| MDCR | 952 | 9,515 | 0.6 | 8.5 | <5 | 11 | < 9.04 (2.93 - 21.1) | 1.29 (0.64 - 2.31) |
| Open Claims | 155,612 | 429,807 | 93.6 | 320.0 | 0 | 0 | | |
| OEHR | 9,105 | 38,288 | 2.3 | 15.6 | 0 | 0 | | |
| VA | 1,341 | 24,844 | 0.7 | 18.9 | 0 | 18 | | 0.95 (0.56 - 1.50) |
| Meta-analysis | 18,105 | 73,257 | 9.7 | 50.0 | 302 | 1,468 | 31.1 (27.7 - 34.8) | 29.4 (27.9 - 30.9) |
| Meta-analysis (w/o USOC) | 18,105 | 73,257 | 9.7 | 50.0 | 302 | 1,468 | 31.1 (27.7 - 34.8) | 29.4 (27.9 - 30.9) |

##### 4.4.6 Hospitalization with heart failure

**Supplementary Table 373:** Exposures, outcomes and risk estimates for Hospitalization with heart failure in GLP1RA (male) (T) vs DPP4I (male) (C) new-user cohorts under four different propensity score (PS) adjustment / time-at-risk (TAR) designs across data sources. We report total patient exposure time in 1,000 patient-year (KYr) units, uncalibrated and calibrated hazard ratios (HRs) and their 95% confidence intervals (CIs). Greyed out entries did not pass study diagnostics and are included only for completeness.

| | Patients | | Exposure (KYr) | | Outcomes | | Incidence (per KYr) | |
|---|---|---|---|---|---|---|---|---|
| | T | C | T | C | T | C | T | C |
| **PS-matching / on-treatment TAR** | | | | | | | | |
| CCAE | 10,377 | 51,798 | 7.2 | 45.4 | 44 | 254 | 6.14 (4.46 - 8.24) | 5.59 (4.92 - 6.32) |
| ODOD | 6,125 | 28,604 | 3.8 | 24.1 | 46 | 421 | 12.1 (8.83 - 16.1) | 17.5 (15.8 - 19.2) |
| MDCR | 798 | 8,221 | 0.6 | 8.6 | 16 | 247 | 28.9 (16.5 - 46.9) | 28.6 (25.2 - 32.4) |
| Open Claims | 111,489 | 409,329 | 86.4 | 392.5 | 472 | 4,073 | 5.46 (4.98 - 5.98) | 10.4 (10.1 - 10.7) |
| OEHR | 7,657 | 37,350 | 2.3 | 18.5 | 14 | 232 | 6.18 (3.38 - 10.4) | 12.6 (11.0 - 14.3) |
| VA | 1,160 | 17,097 | 0.9 | 14.5 | <10 | 117 | < 11.7 (5.62 - 21.5) | 8.09 (6.69 - 9.70) |
| Meta-analysis | 137,606 | 552,399 | 101.0 | 503.6 | <602 | 5,344 | < 5.96 (5.49 - 6.45) | 10.6 (10.3 - 10.9) |
| Meta-analysis (w/o USOC) | 26,117 | 143,070 | 14.7 | 111.1 | <130 | 1,271 | < 8.87 (7.41 - 10.5) | 11.4 (10.8 - 12.1) |
| **PS-stratification / on-treatment TAR** | | | | | | | | |
| CCAE | 12,239 | 51,915 | 8.0 | 45.6 | 52 | 256 | 6.50 (4.86 - 8.53) | 5.62 (4.95 - 6.35) |
| ODOD | 8,142 | 29,193 | 4.7 | 24.6 | 55 | 448 | 11.8 (8.90 - 15.4) | 18.2 (16.6 - 20.0) |
| MDCR | 889 | 8,741 | 0.6 | 9.1 | 17 | 284 | 29.2 (17.0 - 46.8) | 31.1 (27.6 - 35.0) |
| Open Claims | 150,190 | 411,035 | 103.6 | 394.2 | 538 | 4,180 | 5.19 (4.76 - 5.65) | 10.6 (10.3 - 10.9) |
| OEHR | 8,933 | 37,434 | 2.5 | 18.5 | 17 | 239 | 6.68 (3.89 - 10.7) | 12.9 (11.3 - 14.6) |
| VA | 1,339 | 24,760 | 0.9 | 23.1 | <10 | 201 | < 10.7 (5.15 - 19.7) | 8.71 (7.55 - 10.00) |
| Meta-analysis | 179,504 | 529,577 | 118.8 | 482.8 | 662 | 5,123 | 5.57 (5.16 - 6.01) | 10.6 (10.3 - 10.9) |
| Meta-analysis (w/o USOC) | 29,314 | 118,542 | 15.2 | 88.7 | 124 | 943 | 8.16 (6.79 - 9.73) | 10.6 (9.97 - 11.3) |
| **PS-matching / on-treatment TAR without escalation** | | | | | | | | |
| CCAE | 10,377 | 51,798 | 6.2 | 36.7 | 40 | 207 | 6.44 (4.60 - 8.77) | 5.63 (4.89 - 6.46) |
| ODOD | 6,125 | 28,604 | 3.4 | 19.7 | 43 | 349 | 12.8 (9.24 - 17.2) | 17.7 (15.9 - 19.7) |
| MDCR | 798 | 8,221 | 0.5 | 7.4 | 13 | 211 | 26.6 (14.1 - 45.4) | 28.7 (24.9 - 32.8) |
| Open Claims | 111,489 | 409,329 | 74.5 | 304.3 | 403 | 3,168 | 5.41 (4.89 - 5.96) | 10.4 (10.1 - 10.8) |
| OEHR | 7,657 | 37,350 | 2.0 | 15.1 | 13 | 172 | 6.66 (3.55 - 11.4) | 11.4 (9.77 - 13.2) |
| VA | 1,160 | 17,097 | 0.7 | 11.8 | <10 | 94 | < 15.0 (7.21 - 27.6) | 7.94 (6.42 - 9.72) |
| Meta-analysis | 137,606 | 552,399 | 87.2 | 395.0 | <522 | 4,201 | < 5.99 (5.48 - 6.52) | 10.6 (10.3 - 11.0) |
| Meta-analysis (w/o USOC) | 26,117 | 143,070 | 12.7 | 90.7 | <119 | 1,033 | < 9.38 (7.77 - 11.2) | 11.4 (10.7 - 12.1) |
| **PS-stratification / on-treatment TAR without escalation** | | | | | | | | |
| CCAE | 12,239 | 51,915 | 7.0 | 36.8 | 47 | 209 | 6.75 (4.96 - 8.98) | 5.67 (4.93 - 6.50) |
| ODOD | 8,142 | 29,193 | 4.1 | 20.1 | 51 | 374 | 12.3 (9.18 - 16.2) | 18.6 (16.8 - 20.6) |
| MDCR | 889 | 8,741 | 0.5 | 7.8 | 14 | 246 | 27.2 (14.9 - 45.6) | 31.6 (27.7 - 35.8) |
| Open Claims | 150,190 | 411,035 | 90.5 | 305.6 | 463 | 3,260 | 5.11 (4.66 - 5.60) | 10.7 (10.3 - 11.0) |
| OEHR | 8,933 | 37,434 | 2.2 | 15.1 | 16 | 176 | 7.24 (4.14 - 11.8) | 11.6 (9.99 - 13.5) |
| VA | 1,339 | 24,760 | 0.7 | 18.8 | <10 | 158 | < 13.7 (6.55 - 25.1) | 8.42 (7.16 - 9.84) |
| Meta-analysis | 179,504 | 529,577 | 103.9 | 377.6 | 577 | 4,019 | 5.56 (5.11 - 6.03) | 10.6 (10.3 - 11.0) |
| Meta-analysis (w/o USOC) | 29,314 | 118,542 | 13.3 | 72.0 | 114 | 759 | 8.57 (7.07 - 10.3) | 10.5 (9.80 - 11.3) |

##### 4.4.7 Measured renal dysfunction

**Supplementary Table 374:** Exposures, outcomes and risk estimates for Measured renal dysfunction in GLP1RA (male) (T) vs DPP4I (male) (C) new-user cohorts under four different propensity score (PS) adjustment / time-at-risk (TAR) designs across data sources. We report total patient exposure time in 1,000 patient-year (KYr) units, uncalibrated and calibrated hazard ratios (HRs) and their 95% confidence intervals (CIs). Greyed out entries did not pass study diagnostics and are included only for completeness.

| | Patients | | Exposure (KYr) | | Outcomes | | Incidence (per KYr) | |
|---|---|---|---|---|---|---|---|---|
| | T | C | T | C | T | C | T | C |
| **PS-matching / on-treatment TAR** | | | | | | | | |
| CCAE | 10,598 | 52,681 | 7.3 | 46.3 | 0 | 16 | | 0.35 (0.20 - 0.56) |
| ODOD | 6,403 | 30,170 | 4.0 | 25.5 | <5 | 46 | < 1.25 (0.41 - 2.91) | 1.81 (1.32 - 2.41) |
| MDCR | 856 | 8,910 | 0.6 | 9.4 | 0 | <5 | | < 0.53 (0.17 - 1.24) |
| Open Claims | 115,660 | 427,485 | 89.4 | 411.0 | 0 | 0 | | |
| OEHR | 7,806 | 38,195 | 2.3 | 19.2 | 0 | 0 | | |
| VA | 1,194 | 17,479 | 0.9 | 14.8 | <10 | 69 | < 11.3 (5.41 - 20.8) | 4.66 (3.62 - 5.89) |
| Meta-analysis | 7,597 | 47,649 | 4.9 | 40.3 | <15 | 115 | < 3.07 (1.72 - 5.06) | 2.85 (2.36 - 3.43) |
| Meta-analysis (w/o USOC) | 7,597 | 47,649 | 4.9 | 40.3 | <15 | 115 | < 3.07 (1.72 - 5.06) | 2.85 (2.36 - 3.43) |
| **PS-stratification / on-treatment TAR** | | | | | | | | |
| CCAE | 12,522 | 52,816 | 8.2 | 46.4 | 0 | 17 | | 0.37 (0.21 - 0.59) |
| ODOD | 8,508 | 30,791 | 4.9 | 26.0 | 6 | 54 | 1.23 (0.45 - 2.68) | 2.08 (1.56 - 2.71) |
| MDCR | 953 | 9,568 | 0.6 | 10.0 | 0 | <5 | | < 0.50 (0.16 - 1.16) |
| Open Claims | 155,612 | 429,807 | 107.2 | 413.1 | 0 | 0 | | |
| OEHR | 9,105 | 38,288 | 2.6 | 19.2 | 0 | 0 | | |
| VA | 1,388 | 25,292 | 1.0 | 23.6 | <10 | 98 | < 10.4 (4.97 - 19.1) | 4.15 (3.37 - 5.05) |
| Meta-analysis | 8,508 | 30,791 | 4.9 | 26.0 | 6 | 54 | 1.23 (0.45 - 2.68) | 2.08 (1.56 - 2.71) |
| Meta-analysis (w/o USOC) | 8,508 | 30,791 | 4.9 | 26.0 | 6 | 54 | 1.23 (0.45 - 2.68) | 2.08 (1.56 - 2.71) |
| **PS-matching / on-treatment TAR without escalation** | | | | | | | | |
| CCAE | 10,598 | 52,681 | 6.3 | 37.4 | 0 | 12 | | 0.32 (0.17 - 0.56) |
| ODOD | 6,403 | 30,170 | 3.5 | 20.8 | <5 | 40 | < 1.41 (0.46 - 3.30) | 1.92 (1.37 - 2.62) |
| MDCR | 856 | 8,910 | 0.5 | 8.0 | 0 | 0 | | |
| Open Claims | 115,660 | 427,485 | 77.1 | 318.3 | 0 | 0 | | |
| OEHR | 7,806 | 38,195 | 2.0 | 15.5 | 0 | 0 | | |
| VA | 1,194 | 17,479 | 0.7 | 12.1 | <10 | 54 | < 14.4 (6.91 - 26.5) | 4.46 (3.35 - 5.82) |
| Meta-analysis | 6,403 | 30,170 | 3.5 | 20.8 | <5 | 40 | < 1.41 (0.46 - 3.30) | 1.92 (1.37 - 2.62) |
| Meta-analysis (w/o USOC) | 6,403 | 30,170 | 3.5 | 20.8 | <5 | 40 | < 1.41 (0.46 - 3.30) | 1.92 (1.37 - 2.62) |
| **PS-stratification / on-treatment TAR without escalation** | | | | | | | | |
| CCAE | 12,522 | 52,816 | 7.1 | 37.5 | 0 | 13 | | 0.35 (0.18 - 0.59) |
| ODOD | 8,508 | 30,791 | 4.3 | 21.2 | <5 | 48 | < 1.15 (0.37 - 2.69) | 2.26 (1.67 - 3.00) |
| MDCR | 953 | 9,568 | 0.6 | 8.6 | 0 | 0 | | |
| Open Claims | 155,612 | 429,807 | 93.6 | 320.0 | 0 | 0 | | |
| OEHR | 9,105 | 38,288 | 2.3 | 15.6 | 0 | 0 | | |
| VA | 1,388 | 25,292 | 0.8 | 19.2 | <10 | 77 | < 13.1 (6.29 - 24.1) | 4.01 (3.17 - 5.02) |
| Meta-analysis | 8,508 | 30,791 | 4.3 | 21.2 | <5 | 48 | < 1.15 (0.37 - 2.69) | 2.26 (1.67 - 3.00) |
| Meta-analysis (w/o USOC) | 8,508 | 30,791 | 4.3 | 21.2 | <5 | 48 | < 1.15 (0.37 - 2.69) | 2.26 (1.67 - 3.00) |

##### 4.4.8 Stroke

**Supplementary Table 375:** Exposures, outcomes and risk estimates for Stroke in GLP1RA (male) (T) vs DPP4I (male) (C) new-user cohorts under four different propensity score (PS) adjustment / time-at-risk (TAR) designs across data sources. We report total patient exposure time in 1,000 patient-year (KYr) units, uncalibrated and calibrated hazard ratios (HRs) and their 95% confidence intervals (CIs). Greyed out entries did not pass study diagnostics and are included only for completeness.

| | Patients | | Exposure (KYr) | | Outcomes | | Incidence (per KYr) | |
|---|---|---|---|---|---|---|---|---|
| | T | C | T | C | T | C | T | C |
| **PS-matching / on-treatment TAR** | | | | | | | | |
| CCAE | 10,484 | 52,183 | 7.2 | 45.8 | 23 | 152 | 3.18 (2.02 - 4.78) | 3.32 (2.81 - 3.89) |
| ODOD | 6,282 | 29,513 | 3.9 | 24.9 | 20 | 213 | 5.09 (3.11 - 7.86) | 8.55 (7.44 - 9.78) |
| MDCR | 828 | 8,611 | 0.6 | 9.0 | 5 | 128 | 8.84 (2.87 - 20.6) | 14.2 (11.8 - 16.9) |
| Open Claims | 113,043 | 415,501 | 87.5 | 399.3 | 255 | 2,015 | 2.91 (2.57 - 3.29) | 5.05 (4.83 - 5.27) |
| OEHR | 7,745 | 37,856 | 2.3 | 18.9 | 11 | 85 | 4.81 (2.40 - 8.60) | 4.49 (3.58 - 5.55) |
| VA | 1,192 | 17,454 | 0.9 | 14.8 | <10 | 32 | < 11.3 (5.41 - 20.7) | 2.16 (1.48 - 3.05) |
| Meta-analysis | 138,382 | 543,664 | 101.5 | 498.1 | 314 | 2,593 | 3.09 (2.76 - 3.45) | 5.21 (5.01 - 5.41) |
| Meta-analysis (w/o USOC) | 25,339 | 128,163 | 14.0 | 98.7 | 59 | 578 | 4.21 (3.21 - 5.43) | 5.85 (5.39 - 6.35) |
| **PS-stratification / on-treatment TAR** | | | | | | | | |
| CCAE | 12,388 | 52,301 | 8.1 | 46.0 | 24 | 152 | 2.97 (1.91 - 4.43) | 3.31 (2.80 - 3.88) |
| ODOD | 8,371 | 30,043 | 4.8 | 25.4 | 24 | 223 | 5.00 (3.21 - 7.44) | 8.79 (7.68 - 10.0) |
| MDCR | 922 | 9,165 | 0.6 | 9.6 | 5 | 141 | 8.39 (2.72 - 19.6) | 14.7 (12.4 - 17.4) |
| Open Claims | 152,434 | 417,598 | 105.1 | 401.3 | 292 | 2,067 | 2.78 (2.47 - 3.12) | 5.15 (4.93 - 5.38) |
| OEHR | 9,031 | 37,945 | 2.6 | 19.0 | 12 | 87 | 4.66 (2.41 - 8.13) | 4.58 (3.67 - 5.65) |
| VA | 1,382 | 25,218 | 1.0 | 23.6 | <10 | 64 | < 10.4 (4.97 - 19.0) | 2.71 (2.09 - 3.46) |
| Meta-analysis | 182,224 | 537,887 | 120.5 | 491.6 | 352 | 2,529 | 2.92 (2.62 - 3.24) | 5.14 (4.95 - 5.35) |
| Meta-analysis (w/o USOC) | 29,790 | 120,289 | 15.4 | 90.3 | 60 | 462 | 3.89 (2.96 - 5.00) | 5.12 (4.66 - 5.60) |
| **PS-matching / on-treatment TAR without escalation** | | | | | | | | |
| CCAE | 10,484 | 52,183 | 6.3 | 37.0 | 20 | 121 | 3.20 (1.95 - 4.94) | 3.27 (2.71 - 3.90) |
| ODOD | 6,282 | 29,513 | 3.5 | 20.3 | 17 | 184 | 4.90 (2.86 - 7.85) | 9.05 (7.79 - 10.5) |
| MDCR | 828 | 8,611 | 0.5 | 7.7 | <5 | 107 | < 9.99 (3.24 - 23.3) | 13.9 (11.4 - 16.8) |
| Open Claims | 113,043 | 415,501 | 75.5 | 309.4 | 212 | 1,599 | 2.81 (2.44 - 3.21) | 5.17 (4.92 - 5.43) |
| OEHR | 7,745 | 37,856 | 2.0 | 15.4 | 9 | 63 | 4.55 (2.08 - 8.64) | 4.10 (3.15 - 5.24) |
| VA | 1,192 | 17,454 | 0.7 | 12.1 | <10 | 22 | < 14.4 (6.91 - 26.5) | 1.82 (1.14 - 2.75) |
| Meta-analysis | 138,382 | 543,664 | 87.7 | 389.9 | <263 | 2,074 | < 3.00 (2.65 - 3.39) | 5.32 (5.09 - 5.55) |
| Meta-analysis (w/o USOC) | 25,339 | 128,163 | 12.2 | 80.5 | <51 | 475 | < 4.18 (3.11 - 5.50) | 5.90 (5.38 - 6.46) |
| **PS-stratification / on-treatment TAR without escalation** | | | | | | | | |
| CCAE | 12,388 | 52,301 | 7.0 | 37.1 | 21 | 121 | 2.99 (1.85 - 4.57) | 3.26 (2.70 - 3.89) |
| ODOD | 8,371 | 30,043 | 4.3 | 20.7 | 21 | 194 | 4.93 (3.05 - 7.53) | 9.38 (8.10 - 10.8) |
| MDCR | 922 | 9,165 | 0.5 | 8.2 | <5 | 118 | < 9.47 (3.08 - 22.1) | 14.4 (11.9 - 17.3) |
| Open Claims | 152,434 | 417,598 | 91.8 | 311.0 | 245 | 1,640 | 2.67 (2.35 - 3.03) | 5.27 (5.02 - 5.54) |
| OEHR | 9,031 | 37,945 | 2.2 | 15.4 | 10 | 64 | 4.46 (2.14 - 8.20) | 4.15 (3.20 - 5.30) |
| VA | 1,382 | 25,218 | 0.8 | 19.2 | <10 | 41 | < 13.1 (6.29 - 24.1) | 2.14 (1.53 - 2.90) |
| Meta-analysis | 182,224 | 537,887 | 105.3 | 384.2 | 297 | 2,019 | 2.82 (2.51 - 3.16) | 5.25 (5.03 - 5.49) |
| Meta-analysis (w/o USOC) | 29,790 | 120,289 | 13.5 | 73.2 | 52 | 379 | 3.84 (2.87 - 5.04) | 5.17 (4.67 - 5.72) |

##### 4.4.9 Sudden cardiac death

**Supplementary Table 376:** Exposures, outcomes and risk estimates for Sudden cardiac death in GLP1RA (male) (T) vs DPP4I (male) (C) new-user cohorts under four different propensity score (PS) adjustment / time-at-risk (TAR) designs across data sources. We report total patient exposure time in 1,000 patient-year (KYr) units, uncalibrated and calibrated hazard ratios (HRs) and their 95% confidence intervals (CIs). Greyed out entries did not pass study diagnostics and are included only for completeness.

| | Patients | | Exposure (KYr) | | Outcomes | | Incidence (per KYr) | |
|---|---|---|---|---|---|---|---|---|
| | T | C | T | C | T | C | T | C |
| **PS-matching / on-treatment TAR** | | | | | | | | |
| CCAE | 10,601 | 52,701 | 7.3 | 46.4 | <5 | 36 | < 0.68 (0.22 - 1.59) | 0.78 (0.54 - 1.08) |
| ODOD | 6,400 | 30,099 | 4.0 | 25.4 | 9 | 81 | 2.25 (1.03 - 4.27) | 3.18 (2.53 - 3.96) |
| MDCR | 851 | 8,878 | 0.6 | 9.4 | <5 | 35 | < 8.42 (2.73 - 19.6) | 3.74 (2.60 - 5.20) |
| Open Claims | 115,407 | 426,516 | 89.2 | 409.8 | 76 | 621 | 0.85 (0.67 - 1.07) | 1.52 (1.40 - 1.64) |
| OEHR | 7,802 | 38,165 | 2.3 | 19.1 | 5 | 29 | 2.16 (0.70 - 5.05) | 1.52 (1.01 - 2.18) |
| VA | 1,212 | 17,668 | 0.9 | 15.0 | 0 | <10 | | < 0.67 (0.32 - 1.23) |
| Meta-analysis | 140,210 | 547,481 | 102.9 | 500.8 | <95 | 767 | < 0.92 (0.75 - 1.13) | 1.53 (1.43 - 1.64) |
| Meta-analysis (w/o USOC) | 24,803 | 120,965 | 13.6 | 90.9 | <19 | 146 | < 1.39 (0.84 - 2.17) | 1.61 (1.36 - 1.89) |
| **PS-stratification / on-treatment TAR** | | | | | | | | |
| CCAE | 12,525 | 52,834 | 8.2 | 46.5 | <5 | 36 | < 0.61 (0.20 - 1.43) | 0.77 (0.54 - 1.07) |
| ODOD | 8,502 | 30,796 | 4.9 | 26.0 | 10 | 87 | 2.05 (0.98 - 3.78) | 3.34 (2.68 - 4.12) |
| MDCR | 949 | 9,537 | 0.6 | 10.0 | <5 | 40 | < 8.02 (2.60 - 18.7) | 3.99 (2.85 - 5.44) |
| Open Claims | 155,278 | 428,778 | 106.9 | 412.0 | 87 | 650 | 0.81 (0.65 - 1.00) | 1.58 (1.46 - 1.70) |
| OEHR | 9,101 | 38,258 | 2.6 | 19.2 | 5 | 30 | 1.92 (0.62 - 4.49) | 1.56 (1.05 - 2.23) |
| VA | 1,405 | 25,610 | 1.0 | 24.0 | 0 | <10 | | < 0.42 (0.20 - 0.77) |
| Meta-analysis | 185,406 | 550,666 | 122.6 | 503.6 | <107 | 803 | < 0.87 (0.72 - 1.05) | 1.59 (1.49 - 1.71) |
| Meta-analysis (w/o USOC) | 30,128 | 121,888 | 15.7 | 91.7 | <20 | 153 | < 1.28 (0.78 - 1.97) | 1.67 (1.41 - 1.96) |
| **PS-matching / on-treatment TAR without escalation** | | | | | | | | |
| CCAE | 10,601 | 52,701 | 6.3 | 37.5 | <5 | 30 | < 0.79 (0.26 - 1.84) | 0.80 (0.54 - 1.14) |
| ODOD | 6,400 | 30,099 | 3.5 | 20.8 | 8 | 73 | 2.27 (0.98 - 4.47) | 3.52 (2.76 - 4.42) |
| MDCR | 851 | 8,878 | 0.5 | 8.0 | <5 | 23 | < 9.52 (3.09 - 22.2) | 2.89 (1.83 - 4.33) |
| Open Claims | 115,407 | 426,516 | 76.9 | 317.5 | 64 | 475 | 0.83 (0.64 - 1.06) | 1.50 (1.36 - 1.64) |
| OEHR | 7,802 | 38,165 | 2.0 | 15.5 | <5 | 19 | < 2.51 (0.82 - 5.86) | 1.22 (0.74 - 1.91) |
| VA | 1,212 | 17,668 | 0.7 | 12.3 | 0 | <10 | | < 0.82 (0.39 - 1.50) |
| Meta-analysis | 132,408 | 509,316 | 86.8 | 375.7 | <77 | 578 | < 0.89 (0.70 - 1.11) | 1.54 (1.42 - 1.67) |
| Meta-analysis (w/o USOC) | 17,001 | 82,800 | 9.9 | 58.2 | <13 | 103 | < 1.32 (0.70 - 2.25) | 1.77 (1.44 - 2.15) |
| **PS-stratification / on-treatment TAR without escalation** | | | | | | | | |
| CCAE | 12,525 | 52,834 | 7.1 | 37.6 | <5 | 30 | < 0.70 (0.23 - 1.64) | 0.80 (0.54 - 1.14) |
| ODOD | 8,502 | 30,796 | 4.3 | 21.2 | 9 | 78 | 2.08 (0.95 - 3.95) | 3.67 (2.90 - 4.58) |
| MDCR | 949 | 9,537 | 0.6 | 8.5 | <5 | 27 | < 9.05 (2.94 - 21.1) | 3.16 (2.08 - 4.60) |
| Open Claims | 155,278 | 428,778 | 93.4 | 319.2 | 72 | 501 | 0.77 (0.60 - 0.97) | 1.57 (1.44 - 1.71) |
| OEHR | 9,101 | 38,258 | 2.3 | 15.6 | <5 | 20 | < 2.21 (0.72 - 5.17) | 1.28 (0.78 - 1.98) |
| VA | 1,405 | 25,610 | 0.8 | 19.5 | 0 | <10 | | < 0.51 (0.25 - 0.94) |
| Meta-analysis | 176,305 | 512,408 | 104.9 | 378.0 | <86 | 609 | < 0.82 (0.66 - 1.01) | 1.61 (1.49 - 1.74) |
| Meta-analysis (w/o USOC) | 21,027 | 83,630 | 11.5 | 58.8 | <14 | 108 | < 1.22 (0.67 - 2.05) | 1.84 (1.51 - 2.22) |

###### 4.4.10 Abnormal weight gain

**Supplementary Table 377:** Exposures, outcomes and risk estimates for Abnormal weight gain in GLP1RA (male) (T) vs DPP4I (male) (C) new-user cohorts under four different propensity score (PS) adjustment / time-at-risk (TAR) designs across data sources. We report total patient exposure time in 1,000 patient-year (KYr) units, uncalibrated and calibrated hazard ratios (HRs) and their 95% confidence intervals (CIs). Greyed out entries did not pass study diagnostics and are included only for completeness.

| | Patients | | Exposure (KYr) | | Outcomes | | Incidence (per KYr) | |
|---|---|---|---|---|---|---|---|---|
| | T | C | T | C | T | C | T | C |
| **PS-matching / on-treatment TAR** | | | | | | | | |
| CCAE | 10,111 | 51,366 | 7.0 | 45.1 | 37 | 152 | 5.29 (3.72 - 7.29) | 3.37 (2.85 - 3.95) |
| ODOD | 6,116 | 29,345 | 3.8 | 24.6 | 28 | 156 | 7.34 (4.88 - 10.6) | 6.33 (5.38 - 7.41) |
| MDCR | 835 | 8,821 | 0.6 | 9.3 | 5 | 20 | 8.74 (2.84 - 20.4) | 2.15 (1.31 - 3.32) |
| Open Claims | 110,691 | 415,969 | 85.3 | 399.1 | 175 | 708 | 2.05 (1.76 - 2.38) | 1.77 (1.65 - 1.91) |
| OEHR | 7,454 | 37,259 | 2.2 | 18.6 | 20 | 84 | 9.13 (5.58 - 14.1) | 4.52 (3.60 - 5.59) |
| VA | 1,177 | 17,367 | 0.9 | 14.8 | <10 | <10 | < 11.6 (5.58 - 21.4) | < 0.68 (0.32 - 1.25) |
| Meta-analysis | 134,372 | 533,939 | 98.3 | 487.5 | 260 | 1,100 | 2.64 (2.33 - 2.99) | 2.26 (2.13 - 2.39) |
| Meta-analysis (w/o USOC) | 23,681 | 117,970 | 13.0 | 88.4 | 85 | 392 | 6.54 (5.22 - 8.08) | 4.44 (4.01 - 4.90) |
| **PS-stratification / on-treatment TAR** | | | | | | | | |
| CCAE | 11,829 | 51,500 | 7.8 | 45.3 | 41 | 152 | 5.29 (3.80 - 7.17) | 3.36 (2.85 - 3.94) |
| ODOD | 8,011 | 29,955 | 4.6 | 25.1 | 40 | 157 | 8.70 (6.22 - 11.9) | 6.25 (5.31 - 7.30) |
| MDCR | 928 | 9,468 | 0.6 | 10.0 | 5 | 22 | 8.32 (2.70 - 19.4) | 2.21 (1.38 - 3.35) |
| Open Claims | 146,922 | 418,287 | 101.3 | 401.3 | 235 | 710 | 2.32 (2.03 - 2.64) | 1.77 (1.64 - 1.90) |
| OEHR | 8,589 | 37,352 | 2.4 | 18.6 | 23 | 84 | 9.43 (5.98 - 14.1) | 4.50 (3.59 - 5.58) |
| VA | 1,353 | 25,119 | 0.9 | 23.5 | <10 | 11 | < 10.7 (5.14 - 19.7) | 0.47 (0.23 - 0.84) |
| Meta-analysis | 175,351 | 537,094 | 116.1 | 490.3 | 339 | 1,103 | 2.92 (2.62 - 3.25) | 2.25 (2.12 - 2.39) |
| Meta-analysis (w/o USOC) | 28,429 | 118,807 | 14.8 | 89.0 | 104 | 393 | 7.03 (5.75 - 8.52) | 4.41 (3.99 - 4.87) |
| **PS-matching / on-treatment TAR without escalation** | | | | | | | | |
| CCAE | 10,111 | 51,366 | 6.1 | 36.5 | 30 | 121 | 4.94 (3.34 - 7.06) | 3.32 (2.75 - 3.96) |
| ODOD | 6,116 | 29,345 | 3.4 | 20.2 | 21 | 121 | 6.24 (3.87 - 9.55) | 6.00 (4.97 - 7.16) |
| MDCR | 835 | 8,821 | 0.5 | 7.9 | <5 | 14 | < 9.83 (3.19 - 22.9) | 1.76 (0.96 - 2.96) |
| Open Claims | 110,691 | 415,969 | 73.6 | 309.8 | 149 | 532 | 2.03 (1.71 - 2.38) | 1.72 (1.57 - 1.87) |
| OEHR | 7,454 | 37,259 | 1.9 | 15.1 | 18 | 66 | 9.53 (5.65 - 15.1) | 4.37 (3.38 - 5.55) |
| VA | 1,177 | 17,367 | 0.7 | 12.1 | <10 | <10 | < 14.8 (7.08 - 27.2) | < 0.83 (0.40 - 1.52) |
| Meta-analysis | 134,372 | 533,939 | 84.9 | 381.6 | 218 | 840 | 2.57 (2.24 - 2.93) | 2.20 (2.05 - 2.36) |
| Meta-analysis (w/o USOC) | 23,681 | 117,970 | 11.3 | 71.8 | 69 | 308 | 6.09 (4.74 - 7.71) | 4.29 (3.82 - 4.80) |
| **PS-stratification / on-treatment TAR without escalation** | | | | | | | | |
| CCAE | 11,829 | 51,500 | 6.8 | 36.6 | 34 | 121 | 5.03 (3.49 - 7.03) | 3.31 (2.74 - 3.95) |
| ODOD | 8,011 | 29,955 | 4.1 | 20.6 | 33 | 122 | 8.09 (5.57 - 11.4) | 5.92 (4.92 - 7.07) |
| MDCR | 928 | 9,468 | 0.5 | 8.5 | <5 | 16 | < 9.34 (3.03 - 21.8) | 1.88 (1.08 - 3.06) |
| Open Claims | 146,922 | 418,287 | 88.4 | 311.5 | 206 | 534 | 2.33 (2.02 - 2.67) | 1.71 (1.57 - 1.87) |
| OEHR | 8,589 | 37,352 | 2.1 | 15.2 | 19 | 66 | 8.96 (5.40 - 14.0) | 4.35 (3.37 - 5.54) |
| VA | 1,353 | 25,119 | 0.7 | 19.1 | <10 | <10 | < 13.5 (6.48 - 24.8) | < 0.52 (0.25 - 0.96) |
| Meta-analysis | 175,351 | 537,094 | 101.4 | 383.9 | 292 | 843 | 2.88 (2.56 - 3.23) | 2.20 (2.05 - 2.35) |
| Meta-analysis (w/o USOC) | 28,429 | 118,807 | 13.0 | 72.4 | 86 | 309 | 6.64 (5.31 - 8.20) | 4.27 (3.81 - 4.77) |

##### 4.4.11 Abnormal weight loss

**Supplementary Table 378:** Exposures, outcomes and risk estimates for Abnormal weight loss in GLP1RA (male) (T) vs DPP4I (male) (C) new-user cohorts under four different propensity score (PS) adjustment / time-at-risk (TAR) designs across data sources. We report total patient exposure time in 1,000 patient-year (KYr) units, uncalibrated and calibrated hazard ratios (HRs) and their 95% confidence intervals (CIs). Greyed out entries did not pass study diagnostics and are included only for completeness.

| | Patients | | Exposure (KYr) | | Outcomes | | Incidence (per KYr) | |
|---|---|---|---|---|---|---|---|---|
| | T | C | T | C | T | C | T | C |
| **PS-matching / on-treatment TAR** | | | | | | | | |
| CCAE | 10,401 | 51,534 | 7.2 | 45.2 | 56 | 297 | 7.82 (5.91 - 10.2) | 6.57 (5.85 - 7.37) |
| ODOD | 6,222 | 29,041 | 3.9 | 24.3 | 50 | 368 | 13.0 (9.62 - 17.1) | 15.1 (13.6 - 16.8) |
| MDCR | 835 | 8,595 | 0.6 | 9.1 | 5 | 122 | 8.65 (2.81 - 20.2) | 13.5 (11.2 - 16.1) |
| Open Claims | 112,193 | 412,516 | 86.7 | 395.4 | 390 | 2,494 | 4.50 (4.06 - 4.97) | 6.31 (6.06 - 6.56) |
| OEHR | 7,600 | 37,056 | 2.2 | 18.4 | 24 | 221 | 10.7 (6.87 - 16.0) | 12.0 (10.5 - 13.7) |
| VA | 1,178 | 17,162 | 0.9 | 14.6 | 10 | 109 | 11.3 (5.44 - 20.9) | 7.47 (6.14 - 9.01) |
| Meta-analysis | 138,429 | 555,904 | 101.5 | 507.0 | 535 | 3,611 | 5.27 (4.84 - 5.74) | 7.12 (6.89 - 7.36) |
| Meta-analysis (w/o USOC) | 26,236 | 143,388 | 14.7 | 111.6 | 145 | 1,117 | 9.86 (8.32 - 11.6) | 10.0 (9.43 - 10.6) |
| **PS-stratification / on-treatment TAR** | | | | | | | | |
| CCAE | 12,276 | 51,674 | 8.0 | 45.3 | 65 | 298 | 8.15 (6.29 - 10.4) | 6.58 (5.85 - 7.37) |
| ODOD | 8,275 | 29,539 | 4.7 | 24.7 | 61 | 379 | 13.0 (9.91 - 16.6) | 15.3 (13.8 - 16.9) |
| MDCR | 935 | 9,144 | 0.6 | 9.6 | 5 | 137 | 8.22 (2.67 - 19.2) | 14.3 (12.0 - 16.9) |
| Open Claims | 151,256 | 414,312 | 104.1 | 397.1 | 462 | 2,535 | 4.44 (4.04 - 4.86) | 6.38 (6.14 - 6.64) |
| OEHR | 8,866 | 37,138 | 2.5 | 18.5 | 25 | 221 | 9.92 (6.42 - 14.6) | 12.0 (10.4 - 13.7) |
| VA | 1,367 | 24,656 | 1.0 | 23.0 | 11 | 212 | 11.5 (5.74 - 20.6) | 9.21 (8.01 - 10.5) |
| Meta-analysis | 180,673 | 532,663 | 119.3 | 485.6 | 613 | 3,433 | 5.14 (4.74 - 5.56) | 7.07 (6.84 - 7.31) |
| Meta-analysis (w/o USOC) | 29,417 | 118,351 | 15.2 | 88.5 | 151 | 898 | 9.93 (8.41 - 11.6) | 10.1 (9.49 - 10.8) |
| **PS-matching / on-treatment TAR without escalation** | | | | | | | | |
| CCAE | 10,401 | 51,534 | 6.2 | 36.5 | 51 | 231 | 8.23 (6.13 - 10.8) | 6.32 (5.53 - 7.19) |
| ODOD | 6,222 | 29,041 | 3.4 | 19.9 | 44 | 310 | 12.9 (9.37 - 17.3) | 15.6 (13.9 - 17.4) |
| MDCR | 835 | 8,595 | 0.5 | 7.7 | 5 | 109 | 9.81 (3.18 - 22.9) | 14.1 (11.6 - 17.0) |
| Open Claims | 112,193 | 412,516 | 74.8 | 306.4 | 325 | 1,964 | 4.35 (3.89 - 4.84) | 6.41 (6.13 - 6.70) |
| OEHR | 7,600 | 37,056 | 1.9 | 15.0 | 19 | 164 | 9.81 (5.91 - 15.3) | 10.9 (9.32 - 12.7) |
| VA | 1,178 | 17,162 | 0.7 | 11.9 | <10 | 92 | < 14.5 (6.93 - 26.6) | 7.72 (6.22 - 9.46) |
| Meta-analysis | 138,429 | 555,904 | 87.5 | 397.5 | <454 | 2,870 | < 5.19 (4.72 - 5.69) | 7.22 (6.96 - 7.49) |
| Meta-analysis (w/o USOC) | 26,236 | 143,388 | 12.7 | 91.1 | <129 | 906 | < 10.1 (8.45 - 12.0) | 9.95 (9.31 - 10.6) |
| **PS-stratification / on-treatment TAR without escalation** | | | | | | | | |
| CCAE | 12,276 | 51,674 | 7.0 | 36.6 | 59 | 232 | 8.48 (6.46 - 10.9) | 6.33 (5.54 - 7.20) |
| ODOD | 8,275 | 29,539 | 4.2 | 20.2 | 53 | 318 | 12.6 (9.47 - 16.5) | 15.7 (14.0 - 17.5) |
| MDCR | 935 | 9,144 | 0.5 | 8.2 | 5 | 121 | 9.30 (3.02 - 21.7) | 14.8 (12.3 - 17.7) |
| Open Claims | 151,256 | 414,312 | 91.0 | 307.7 | 394 | 1,998 | 4.33 (3.91 - 4.78) | 6.49 (6.21 - 6.78) |
| OEHR | 8,866 | 37,138 | 2.2 | 15.0 | 20 | 164 | 9.11 (5.57 - 14.1) | 10.9 (9.30 - 12.7) |
| VA | 1,367 | 24,656 | 0.8 | 18.7 | <10 | 174 | < 13.2 (6.32 - 24.3) | 9.30 (7.97 - 10.8) |
| Meta-analysis | 180,673 | 532,663 | 104.3 | 379.6 | 526 | 2,712 | 5.04 (4.62 - 5.49) | 7.14 (6.88 - 7.42) |
| Meta-analysis (w/o USOC) | 29,417 | 118,351 | 13.3 | 71.9 | 132 | 714 | 9.89 (8.28 - 11.7) | 9.93 (9.21 - 10.7) |

##### 4.4.12 Acute pancreatitis

**Supplementary Table 379:** Exposures, outcomes and risk estimates for Acute pancreatitis in GLP1RA (male) (T) vs DPP4I (male) (C) new-user cohorts under four different propensity score (PS) adjustment / time-at-risk (TAR) designs across data sources. We report total patient exposure time in 1,000 patient-year (KYr) units, uncalibrated and calibrated hazard ratios (HRs) and their 95% confidence intervals (CIs). Greyed out entries did not pass study diagnostics and are included only for completeness.

| | Patients | | Exposure (KYr) | | Outcomes | | Incidence (per KYr) | |
|---|---|---|---|---|---|---|---|---|
| | T | C | T | C | T | C | T | C |
| **PS-matching / on-treatment TAR** | | | | | | | | |
| CCAE | 10,542 | 52,351 | 7.3 | 46.0 | 16 | 112 | 2.19 (1.25 - 3.56) | 2.43 (2.00 - 2.93) |
| ODOD | 6,368 | 29,991 | 4.0 | 25.4 | 8 | 67 | 2.00 (0.86 - 3.95) | 2.64 (2.05 - 3.35) |
| MDCR | 851 | 8,848 | 0.6 | 9.3 | 0 | 35 | | 3.75 (2.61 - 5.22) |
| Open Claims | 114,402 | 422,380 | 88.5 | 406.1 | 124 | 802 | 1.40 (1.17 - 1.67) | 1.97 (1.84 - 2.12) |
| OEHR | 7,767 | 38,036 | 2.3 | 19.1 | 6 | 44 | 2.61 (0.96 - 5.67) | 2.31 (1.68 - 3.10) |
| VA | 1,203 | 17,550 | 0.9 | 14.9 | 0 | 21 | | 1.41 (0.87 - 2.15) |
| Meta-analysis | 139,079 | 542,758 | 102.1 | 496.6 | 154 | 1,025 | 1.51 (1.28 - 1.77) | 2.06 (1.94 - 2.19) |
| Meta-analysis (w/o USOC) | 24,677 | 120,378 | 13.6 | 90.5 | 30 | 223 | 2.21 (1.49 - 3.15) | 2.46 (2.15 - 2.81) |
| **PS-stratification / on-treatment TAR** | | | | | | | | |
| CCAE | 12,453 | 52,487 | 8.1 | 46.2 | 18 | 112 | 2.21 (1.31 - 3.49) | 2.43 (2.00 - 2.92) |
| ODOD | 8,472 | 30,602 | 4.9 | 25.9 | 9 | 67 | 1.85 (0.85 - 3.51) | 2.59 (2.01 - 3.29) |
| MDCR | 948 | 9,484 | 0.6 | 9.9 | 0 | 38 | | 3.82 (2.70 - 5.24) |
| Open Claims | 154,048 | 424,790 | 106.2 | 408.4 | 149 | 812 | 1.40 (1.19 - 1.65) | 1.99 (1.85 - 2.13) |
| OEHR | 9,062 | 38,128 | 2.6 | 19.1 | 7 | 44 | 2.70 (1.09 - 5.56) | 2.30 (1.67 - 3.09) |
| VA | 1,395 | 25,396 | 1.0 | 23.8 | 0 | 42 | | 1.76 (1.27 - 2.38) |
| Meta-analysis | 184,035 | 546,007 | 121.8 | 499.5 | 183 | 1,035 | 1.50 (1.29 - 1.74) | 2.07 (1.95 - 2.20) |
| Meta-analysis (w/o USOC) | 29,987 | 121,217 | 15.6 | 91.1 | 34 | 223 | 2.18 (1.51 - 3.05) | 2.45 (2.14 - 2.79) |
| **PS-matching / on-treatment TAR without escalation** | | | | | | | | |
| CCAE | 10,542 | 52,351 | 6.3 | 37.2 | 12 | 81 | 1.90 (0.98 - 3.31) | 2.18 (1.73 - 2.71) |
| ODOD | 6,368 | 29,991 | 3.5 | 20.7 | 7 | 52 | 1.99 (0.80 - 4.09) | 2.51 (1.87 - 3.29) |
| MDCR | 851 | 8,848 | 0.5 | 7.9 | 0 | 33 | | 4.16 (2.86 - 5.84) |
| Open Claims | 114,402 | 422,380 | 76.3 | 314.8 | 99 | 588 | 1.30 (1.05 - 1.58) | 1.87 (1.72 - 2.03) |
| OEHR | 7,767 | 38,036 | 2.0 | 15.5 | 5 | 32 | 2.52 (0.82 - 5.87) | 2.07 (1.41 - 2.92) |
| VA | 1,203 | 17,550 | 0.7 | 12.2 | 0 | 15 | | 1.23 (0.69 - 2.03) |
| Meta-analysis | 139,079 | 542,758 | 88.1 | 388.2 | 123 | 753 | 1.40 (1.16 - 1.66) | 1.94 (1.80 - 2.08) |
| Meta-analysis (w/o USOC) | 24,677 | 120,378 | 11.8 | 73.4 | 24 | 165 | 2.03 (1.30 - 3.02) | 2.25 (1.92 - 2.62) |
| **PS-stratification / on-treatment TAR without escalation** | | | | | | | | |
| CCAE | 12,453 | 52,487 | 7.1 | 37.3 | 13 | 81 | 1.83 (0.98 - 3.13) | 2.17 (1.72 - 2.70) |
| ODOD | 8,472 | 30,602 | 4.3 | 21.1 | 8 | 52 | 1.85 (0.80 - 3.65) | 2.46 (1.84 - 3.23) |
| MDCR | 948 | 9,484 | 0.6 | 8.5 | 0 | 36 | | 4.24 (2.97 - 5.87) |
| Open Claims | 154,048 | 424,790 | 92.8 | 316.6 | 123 | 595 | 1.33 (1.10 - 1.58) | 1.88 (1.73 - 2.04) |
| OEHR | 9,062 | 38,128 | 2.3 | 15.5 | 6 | 32 | 2.66 (0.98 - 5.80) | 2.06 (1.41 - 2.91) |
| VA | 1,395 | 25,396 | 0.8 | 19.3 | 0 | 31 | | 1.60 (1.09 - 2.28) |
| Meta-analysis | 184,035 | 546,007 | 106.4 | 390.5 | 150 | 760 | 1.41 (1.19 - 1.65) | 1.95 (1.81 - 2.09) |
| Meta-analysis (w/o USOC) | 29,987 | 121,217 | 13.7 | 74.0 | 27 | 165 | 1.98 (1.30 - 2.87) | 2.23 (1.90 - 2.60) |

##### 4.4.13 Bone fracture

**Supplementary Table 380:** Exposures, outcomes and risk estimates for Bone fracture in GLP1RA (male) (T) vs DPP4I (male) (C) new-user cohorts under four different propensity score (PS) adjustment / time-at-risk (TAR) designs across data sources. We report total patient exposure time in 1,000 patient-year (KYr) units, uncalibrated and calibrated hazard ratios (HRs) and their 95% confidence intervals (CIs). Greyed out entries did not pass study diagnostics and are included only for completeness.

| | Patients | | Exposure (KYr) | | Outcomes | | Incidence (per KYr) | |
|---|---|---|---|---|---|---|---|---|
| | T | C | T | C | T | C | T | C |
| **PS-matching / on-treatment TAR** | | | | | | | | |
| CCAE | 9,745 | 49,178 | 6.6 | 42.5 | 111 | 632 | 16.8 (13.8 - 20.2) | 14.9 (13.7 - 16.1) |
| ODOD | 5,938 | 28,138 | 3.7 | 23.4 | 85 | 449 | 23.2 (18.5 - 28.7) | 19.2 (17.5 - 21.1) |
| MDCR | 769 | 8,059 | 0.5 | 8.3 | 11 | 227 | 21.6 (10.8 - 38.7) | 27.4 (24.0 - 31.2) |
| Open Claims | 101,595 | 383,240 | 78.0 | 364.6 | 852 | 4,916 | 10.9 (10.2 - 11.7) | 13.5 (13.1 - 13.9) |
| OEHR | 7,309 | 36,130 | 2.2 | 17.9 | 27 | 281 | 12.5 (8.21 - 18.1) | 15.7 (13.9 - 17.7) |
| VA | 1,086 | 15,990 | 0.8 | 13.7 | <10 | 123 | < 12.4 (5.96 - 22.8) | 9.01 (7.49 - 10.7) |
| Meta-analysis | 126,442 | 520,735 | 91.8 | 470.3 | <1,096 | 6,628 | < 11.9 (11.2 - 12.7) | 14.1 (13.8 - 14.4) |
| Meta-analysis (w/o USOC) | 24,847 | 137,495 | 13.8 | 105.7 | <244 | 1,712 | < 17.7 (15.6 - 20.1) | 16.2 (15.4 - 17.0) |
| **PS-stratification / on-treatment TAR** | | | | | | | | |
| CCAE | 11,492 | 49,297 | 7.4 | 42.6 | 122 | 633 | 16.5 (13.7 - 19.7) | 14.9 (13.7 - 16.1) |
| ODOD | 7,882 | 28,638 | 4.5 | 23.8 | 111 | 462 | 24.9 (20.5 - 30.0) | 19.4 (17.7 - 21.3) |
| MDCR | 864 | 8,670 | 0.5 | 8.8 | 11 | 242 | 20.5 (10.2 - 36.7) | 27.4 (24.0 - 31.0) |
| Open Claims | 135,593 | 385,620 | 93.0 | 366.8 | 1,016 | 4,993 | 10.9 (10.3 - 11.6) | 13.6 (13.2 - 14.0) |
| OEHR | 8,521 | 36,214 | 2.4 | 17.9 | 30 | 284 | 12.3 (8.31 - 17.6) | 15.8 (14.1 - 17.8) |
| VA | 1,264 | 23,057 | 0.9 | 21.6 | <10 | 216 | < 11.4 (5.47 - 21.0) | 9.99 (8.70 - 11.4) |
| Meta-analysis | 163,488 | 499,769 | 107.3 | 451.1 | 1,279 | 6,372 | 11.9 (11.3 - 12.6) | 14.1 (13.8 - 14.5) |
| Meta-analysis (w/o USOC) | 27,895 | 114,149 | 14.3 | 84.3 | 263 | 1,379 | 18.4 (16.3 - 20.8) | 16.4 (15.5 - 17.2) |
| **PS-matching / on-treatment TAR without escalation** | | | | | | | | |
| CCAE | 9,745 | 49,178 | 5.7 | 34.5 | 89 | 510 | 15.5 (12.5 - 19.1) | 14.8 (13.5 - 16.1) |
| ODOD | 5,938 | 28,138 | 3.2 | 19.2 | 74 | 360 | 22.9 (17.9 - 28.7) | 18.8 (16.9 - 20.8) |
| MDCR | 769 | 8,059 | 0.5 | 7.1 | 10 | 197 | 22.0 (10.6 - 40.5) | 27.8 (24.1 - 32.0) |
| Open Claims | 101,595 | 383,240 | 67.3 | 283.1 | 724 | 3,775 | 10.8 (9.99 - 11.6) | 13.3 (12.9 - 13.8) |
| OEHR | 7,309 | 36,130 | 1.9 | 14.6 | 21 | 207 | 11.2 (6.96 - 17.2) | 14.2 (12.3 - 16.3) |
| VA | 1,086 | 15,990 | 0.6 | 11.2 | <10 | 95 | < 15.9 (7.63 - 29.3) | 8.51 (6.88 - 10.4) |
| Meta-analysis | 126,442 | 520,735 | 79.2 | 369.7 | <928 | 5,144 | < 11.7 (11.0 - 12.5) | 13.9 (13.5 - 14.3) |
| Meta-analysis (w/o USOC) | 24,847 | 137,495 | 11.9 | 86.5 | <204 | 1,369 | < 17.1 (14.8 - 19.6) | 15.8 (15.0 - 16.7) |
| **PS-stratification / on-treatment TAR without escalation** | | | | | | | | |
| CCAE | 11,492 | 49,297 | 6.4 | 34.6 | 99 | 511 | 15.4 (12.5 - 18.7) | 14.8 (13.5 - 16.1) |
| ODOD | 7,882 | 28,638 | 4.0 | 19.5 | 99 | 372 | 25.0 (20.3 - 30.4) | 19.0 (17.1 - 21.1) |
| MDCR | 864 | 8,670 | 0.5 | 7.6 | 10 | 211 | 20.8 (9.98 - 38.3) | 27.8 (24.2 - 31.8) |
| Open Claims | 135,593 | 385,620 | 81.3 | 284.9 | 869 | 3,843 | 10.7 (9.99 - 11.4) | 13.5 (13.1 - 13.9) |
| OEHR | 8,521 | 36,214 | 2.1 | 14.6 | 24 | 209 | 11.4 (7.27 - 16.9) | 14.3 (12.4 - 16.4) |
| VA | 1,264 | 23,057 | 0.7 | 17.6 | <10 | 166 | < 14.4 (6.93 - 26.6) | 9.44 (8.06 - 11.0) |
| Meta-analysis | 163,488 | 499,769 | 93.8 | 353.7 | 1,091 | 4,935 | 11.6 (11.0 - 12.3) | 14.0 (13.6 - 14.3) |
| Meta-analysis (w/o USOC) | 27,895 | 114,149 | 12.5 | 68.8 | 222 | 1,092 | 17.7 (15.5 - 20.2) | 15.9 (15.0 - 16.9) |

##### 4.4.14 Diabetic ketoacidosis

**Supplementary Table 381:** Exposures, outcomes and risk estimates for Diabetic ketoacidosis in GLP1RA (male) (T) vs DPP4I (male) (C) new-user cohorts under four different propensity score (PS) adjustment / time-at-risk (TAR) designs across data sources. We report total patient exposure time in 1,000 patient-year (KYr) units, uncalibrated and calibrated hazard ratios (HRs) and their 95% confidence intervals (CIs). Greyed out entries did not pass study diagnostics and are included only for completeness.

| | Patients | | Exposure (KYr) | | Outcomes | | Incidence (per KYr) | |
|---|---|---|---|---|---|---|---|---|
| | T | C | T | C | T | C | T | C |
| **PS-matching / on-treatment TAR** | | | | | | | | |
| CCAE | 10,603 | 52,701 | 7.3 | 46.4 | 7 | 36 | 0.95 (0.38 - 1.97) | 0.78 (0.54 - 1.07) |
| ODOD | 6,402 | 30,186 | 4.0 | 25.5 | <5 | 37 | < 1.25 (0.41 - 2.91) | 1.45 (1.02 - 2.00) |
| MDCR | 854 | 8,895 | 0.6 | 9.4 | 0 | <5 | | < 0.53 (0.17 - 1.24) |
| Open Claims | 115,399 | 426,651 | 89.2 | 410.1 | 60 | 303 | 0.67 (0.51 - 0.87) | 0.74 (0.66 - 0.83) |
| OEHR | 7,800 | 38,169 | 2.3 | 19.1 | <5 | 24 | < 2.16 (0.70 - 5.05) | 1.25 (0.80 - 1.87) |
| VA | 1,214 | 17,669 | 0.9 | 15.0 | <10 | <10 | < 11.1 (5.33 - 20.5) | < 0.67 (0.32 - 1.23) |
| Meta-analysis | 140,204 | 547,707 | 102.9 | 501.2 | <77 | 400 | < 0.75 (0.59 - 0.94) | 0.80 (0.72 - 0.88) |
| Meta-analysis (w/o USOC) | 24,805 | 121,056 | 13.7 | 91.1 | <17 | 97 | < 1.25 (0.73 - 1.99) | 1.07 (0.86 - 1.30) |
| **PS-stratification / on-treatment TAR** | | | | | | | | |
| CCAE | 12,523 | 52,836 | 8.2 | 46.5 | 8 | 36 | 0.98 (0.42 - 1.93) | 0.77 (0.54 - 1.07) |
| ODOD | 8,503 | 30,811 | 4.9 | 26.0 | <5 | 38 | < 1.03 (0.33 - 2.39) | 1.46 (1.03 - 2.00) |
| MDCR | 952 | 9,553 | 0.6 | 10.0 | 0 | 7 | | 0.70 (0.28 - 1.44) |
| Open Claims | 155,271 | 428,987 | 107.0 | 412.3 | 70 | 304 | 0.65 (0.51 - 0.83) | 0.74 (0.66 - 0.83) |
| OEHR | 9,102 | 38,262 | 2.6 | 19.2 | <5 | 24 | < 1.92 (0.62 - 4.48) | 1.25 (0.80 - 1.86) |
| VA | 1,410 | 25,614 | 1.0 | 24.0 | <10 | 11 | < 10.2 (4.90 - 18.8) | 0.46 (0.23 - 0.82) |
| Meta-analysis | 185,399 | 550,896 | 122.6 | 504.0 | <88 | 402 | < 0.72 (0.58 - 0.88) | 0.80 (0.72 - 0.88) |
| Meta-analysis (w/o USOC) | 30,128 | 121,909 | 15.7 | 91.7 | <18 | 98 | < 1.15 (0.68 - 1.82) | 1.07 (0.87 - 1.30) |
| **PS-matching / on-treatment TAR without escalation** | | | | | | | | |
| CCAE | 10,603 | 52,701 | 6.4 | 37.5 | 6 | 28 | 0.94 (0.35 - 2.06) | 0.75 (0.50 - 1.08) |
| ODOD | 6,402 | 30,186 | 3.5 | 20.8 | <5 | 25 | < 1.41 (0.46 - 3.30) | 1.20 (0.78 - 1.77) |
| MDCR | 854 | 8,895 | 0.5 | 8.0 | 0 | <5 | | < 0.63 (0.20 - 1.46) |
| Open Claims | 115,399 | 426,651 | 76.9 | 317.8 | 49 | 216 | 0.64 (0.47 - 0.84) | 0.68 (0.59 - 0.78) |
| OEHR | 7,800 | 38,169 | 2.0 | 15.5 | <5 | 17 | < 2.51 (0.81 - 5.86) | 1.09 (0.64 - 1.75) |
| VA | 1,214 | 17,669 | 0.7 | 12.3 | <10 | <10 | < 14.2 (6.81 - 26.1) | < 0.82 (0.39 - 1.50) |
| Meta-analysis | 132,404 | 509,538 | 86.8 | 376.1 | <60 | 269 | < 0.69 (0.53 - 0.89) | 0.72 (0.63 - 0.81) |
| Meta-analysis (w/o USOC) | 17,005 | 82,887 | 9.9 | 58.3 | <11 | 53 | < 1.11 (0.56 - 1.99) | 0.91 (0.68 - 1.19) |
| **PS-stratification / on-treatment TAR without escalation** | | | | | | | | |
| CCAE | 12,523 | 52,836 | 7.1 | 37.6 | 7 | 28 | 0.98 (0.39 - 2.02) | 0.75 (0.50 - 1.08) |
| ODOD | 8,503 | 30,811 | 4.3 | 21.3 | <5 | 26 | < 1.15 (0.38 - 2.70) | 1.22 (0.80 - 1.79) |
| MDCR | 952 | 9,553 | 0.6 | 8.6 | 0 | 6 | | 0.70 (0.26 - 1.53) |
| Open Claims | 155,271 | 428,987 | 93.5 | 319.5 | 58 | 217 | 0.62 (0.47 - 0.80) | 0.68 (0.59 - 0.78) |
| OEHR | 9,102 | 38,262 | 2.3 | 15.6 | <5 | 17 | < 2.21 (0.72 - 5.16) | 1.09 (0.64 - 1.75) |
| VA | 1,410 | 25,614 | 0.8 | 19.5 | <10 | <10 | < 12.9 (6.20 - 23.8) | < 0.51 (0.25 - 0.94) |
| Meta-analysis | 176,297 | 512,634 | 104.9 | 378.4 | <70 | 271 | < 0.67 (0.52 - 0.84) | 0.72 (0.63 - 0.81) |
| Meta-analysis (w/o USOC) | 21,026 | 83,647 | 11.5 | 58.9 | <12 | 54 | < 1.05 (0.54 - 1.83) | 0.92 (0.69 - 1.20) |

##### 4.4.15 Diarrhea

**Supplementary Table 382:** Exposures, outcomes and risk estimates for Diarrhea in GLP1RA (male) (T) vs DPP4I (male) (C) new-user cohorts under four different propensity score (PS) adjustment / time-at-risk (TAR) designs across data sources. We report total patient exposure time in 1,000 patient-year (KYr) units, uncalibrated and calibrated hazard ratios (HRs) and their 95% confidence intervals (CIs). Greyed out entries did not pass study diagnostics and are included only for completeness.

| | Patients | | Exposure (KYr) | | Outcomes | | Incidence (per KYr) | |
|---|---|---|---|---|---|---|---|---|
| | T | C | T | C | T | C | T | C |
| **PS-matching / on-treatment TAR** | | | | | | | | |
| CCAE | 9,532 | 48,344 | 6.4 | 41.7 | 237 | 869 | 36.8 (32.3 - 41.8) | 20.8 (19.5 - 22.3) |
| ODOD | 5,742 | 26,979 | 3.5 | 22.1 | 151 | 661 | 43.0 (36.4 - 50.5) | 29.8 (27.6 - 32.2) |
| MDCR | 764 | 7,959 | 0.5 | 8.3 | 17 | 237 | 32.7 (19.0 - 52.3) | 28.7 (25.2 - 32.6) |
| Open Claims | 102,627 | 385,298 | 78.6 | 365.3 | 1,387 | 5,462 | 17.6 (16.7 - 18.6) | 15.0 (14.6 - 15.4) |
| OEHR | 7,127 | 35,171 | 2.1 | 17.2 | 82 | 466 | 39.8 (31.6 - 49.4) | 27.2 (24.8 - 29.7) |
| VA | 1,041 | 15,382 | 0.8 | 13.0 | 21 | 258 | 27.8 (17.2 - 42.5) | 19.8 (17.5 - 22.4) |
| Meta-analysis | 126,833 | 519,133 | 91.9 | 467.6 | 1,895 | 7,953 | 20.6 (19.7 - 21.6) | 17.0 (16.6 - 17.4) |
| Meta-analysis (w/o USOC) | 24,206 | 133,835 | 13.3 | 102.3 | 508 | 2,491 | 38.3 (35.0 - 41.7) | 24.4 (23.4 - 25.3) |
| **PS-stratification / on-treatment TAR** | | | | | | | | |
| CCAE | 11,200 | 48,527 | 7.2 | 41.9 | 273 | 874 | 38.2 (33.8 - 43.0) | 20.9 (19.5 - 22.3) |
| ODOD | 7,599 | 27,566 | 4.3 | 22.6 | 191 | 676 | 44.8 (38.7 - 51.6) | 29.9 (27.7 - 32.2) |
| MDCR | 850 | 8,555 | 0.5 | 8.8 | 17 | 264 | 31.2 (18.2 - 50.0) | 30.0 (26.5 - 33.8) |
| Open Claims | 137,360 | 387,058 | 93.7 | 366.9 | 1,776 | 5,531 | 18.9 (18.1 - 19.8) | 15.1 (14.7 - 15.5) |
| OEHR | 8,239 | 35,251 | 2.3 | 17.2 | 99 | 470 | 43.0 (34.9 - 52.3) | 27.3 (24.9 - 29.9) |
| VA | 1,194 | 22,232 | 0.8 | 20.7 | 23 | 408 | 28.0 (17.8 - 42.1) | 19.7 (17.9 - 21.7) |
| Meta-analysis | 164,398 | 498,402 | 107.5 | 448.6 | 2,339 | 7,551 | 21.8 (20.9 - 22.7) | 16.8 (16.5 - 17.2) |
| Meta-analysis (w/o USOC) | 27,038 | 111,344 | 13.7 | 81.7 | 563 | 2,020 | 41.0 (37.7 - 44.6) | 24.7 (23.7 - 25.8) |
| **PS-matching / on-treatment TAR without escalation** | | | | | | | | |
| CCAE | 9,532 | 48,344 | 5.6 | 33.9 | 214 | 707 | 38.2 (33.3 - 43.7) | 20.8 (19.3 - 22.4) |
| ODOD | 5,742 | 26,979 | 3.1 | 18.2 | 132 | 546 | 42.4 (35.5 - 50.3) | 30.0 (27.5 - 32.6) |
| MDCR | 764 | 7,959 | 0.5 | 7.1 | 15 | 202 | 32.6 (18.2 - 53.7) | 28.6 (24.8 - 32.9) |
| Open Claims | 102,627 | 385,298 | 68.0 | 284.6 | 1,192 | 4,223 | 17.5 (16.6 - 18.6) | 14.8 (14.4 - 15.3) |
| OEHR | 7,127 | 35,171 | 1.8 | 14.1 | 70 | 330 | 39.3 (30.6 - 49.6) | 23.4 (21.0 - 26.1) |
| VA | 1,041 | 15,382 | 0.6 | 10.7 | 18 | 213 | 30.5 (18.1 - 48.2) | 19.9 (17.4 - 22.8) |
| Meta-analysis | 126,833 | 519,133 | 79.5 | 368.5 | 1,641 | 6,221 | 20.6 (19.7 - 21.7) | 16.9 (16.5 - 17.3) |
| Meta-analysis (w/o USOC) | 24,206 | 133,835 | 11.5 | 84.0 | 449 | 1,998 | 38.9 (35.4 - 42.7) | 23.8 (22.8 - 24.9) |
| **PS-stratification / on-treatment TAR without escalation** | | | | | | | | |
| CCAE | 11,200 | 48,527 | 6.3 | 34.1 | 246 | 711 | 39.3 (34.6 - 44.6) | 20.9 (19.4 - 22.5) |
| ODOD | 7,599 | 27,566 | 3.8 | 18.6 | 165 | 561 | 43.4 (37.0 - 50.6) | 30.2 (27.7 - 32.8) |
| MDCR | 850 | 8,555 | 0.5 | 7.6 | 15 | 227 | 31.0 (17.4 - 51.2) | 30.1 (26.3 - 34.2) |
| Open Claims | 137,360 | 387,058 | 82.1 | 285.9 | 1,549 | 4,278 | 18.9 (17.9 - 19.8) | 15.0 (14.5 - 15.4) |
| OEHR | 8,239 | 35,251 | 2.0 | 14.1 | 85 | 332 | 42.3 (33.8 - 52.4) | 23.5 (21.1 - 26.2) |
| VA | 1,194 | 22,232 | 0.6 | 16.9 | 20 | 329 | 31.0 (18.9 - 47.9) | 19.5 (17.4 - 21.7) |
| Meta-analysis | 164,398 | 498,402 | 94.2 | 352.6 | 2,045 | 5,882 | 21.7 (20.8 - 22.7) | 16.7 (16.3 - 17.1) |
| Meta-analysis (w/o USOC) | 27,038 | 111,344 | 12.1 | 66.8 | 496 | 1,604 | 41.1 (37.6 - 44.9) | 24.0 (22.9 - 25.2) |

##### 4.4.16 Genitourinary infection

**Supplementary Table 383:** Exposures, outcomes and risk estimates for Genitourinary infection in GLP1RA (male) (T) vs DPP4I (male) (C) new-user cohorts under four different propensity score (PS) adjustment / time-at-risk (TAR) designs across data sources. We report total patient exposure time in 1,000 patient-year (KYr) units, uncalibrated and calibrated hazard ratios (HRs) and their 95% confidence intervals (CIs). Greyed out entries did not pass study diagnostics and are included only for completeness.

| | Patients | | Exposure (KYr) | | Outcomes | | Incidence (per KYr) | |
|---|---|---|---|---|---|---|---|---|
| | T | C | T | C | T | C | T | C |
| **PS-matching / on-treatment TAR** | | | | | | | | |
| CCAE | 9,683 | 47,980 | 6.6 | 41.3 | 137 | 863 | 20.9 (17.5 - 24.7) | 20.9 (19.5 - 22.3) |
| ODOD | 5,664 | 26,092 | 3.5 | 21.4 | 95 | 762 | 27.2 (22.0 - 33.3) | 35.5 (33.1 - 38.1) |
| MDCR | 736 | 7,646 | 0.5 | 7.9 | 27 | 334 | 56.6 (37.3 - 82.3) | 42.4 (38.0 - 47.2) |
| Open Claims | 102,361 | 374,892 | 79.1 | 355.3 | 1,075 | 6,758 | 13.6 (12.8 - 14.4) | 19.0 (18.6 - 19.5) |
| OEHR | 7,365 | 35,861 | 2.2 | 17.6 | 44 | 395 | 20.3 (14.8 - 27.3) | 22.4 (20.2 - 24.7) |
| VA | 1,084 | 15,730 | 0.8 | 13.3 | 14 | 186 | 17.2 (9.39 - 28.8) | 14.0 (12.1 - 16.2) |
| Meta-analysis | 126,893 | 508,201 | 92.6 | 456.8 | 1,392 | 9,298 | 15.0 (14.3 - 15.8) | 20.4 (19.9 - 20.8) |
| Meta-analysis (w/o USOC) | 24,532 | 133,309 | 13.5 | 101.6 | 317 | 2,540 | 23.5 (21.0 - 26.2) | 25.0 (24.0 - 26.0) |
| **PS-stratification / on-treatment TAR** | | | | | | | | |
| CCAE | 11,380 | 48,125 | 7.3 | 41.4 | 145 | 868 | 19.9 (16.8 - 23.4) | 20.9 (19.6 - 22.4) |
| ODOD | 7,578 | 26,713 | 4.3 | 21.9 | 121 | 803 | 28.3 (23.5 - 33.8) | 36.6 (34.1 - 39.2) |
| MDCR | 819 | 8,073 | 0.5 | 8.3 | 27 | 365 | 53.9 (35.5 - 78.4) | 44.1 (39.7 - 48.9) |
| Open Claims | 138,123 | 377,644 | 95.0 | 357.7 | 1,277 | 6,968 | 13.4 (12.7 - 14.2) | 19.5 (19.0 - 19.9) |
| OEHR | 8,578 | 35,940 | 2.4 | 17.7 | 50 | 402 | 20.5 (15.2 - 27.1) | 22.7 (20.6 - 25.1) |
| VA | 1,260 | 22,854 | 0.9 | 21.3 | 15 | 298 | 17.0 (9.52 - 28.1) | 14.0 (12.5 - 15.7) |
| Meta-analysis | 165,659 | 488,422 | 109.0 | 438.8 | 1,593 | 9,041 | 14.6 (13.9 - 15.3) | 20.6 (20.2 - 21.0) |
| Meta-analysis (w/o USOC) | 27,536 | 110,778 | 14.0 | 81.1 | 316 | 2,073 | 22.6 (20.1 - 25.2) | 25.6 (24.5 - 26.7) |
| **PS-matching / on-treatment TAR without escalation** | | | | | | | | |
| CCAE | 9,683 | 47,980 | 5.7 | 33.6 | 118 | 696 | 20.7 (17.1 - 24.8) | 20.7 (19.2 - 22.3) |
| ODOD | 5,664 | 26,092 | 3.1 | 17.6 | 86 | 632 | 27.9 (22.3 - 34.4) | 36.0 (33.2 - 38.9) |
| MDCR | 736 | 7,646 | 0.4 | 6.7 | 21 | 283 | 49.8 (30.8 - 76.2) | 42.0 (37.2 - 47.1) |
| Open Claims | 102,361 | 374,892 | 68.3 | 275.4 | 925 | 5,348 | 13.5 (12.7 - 14.4) | 19.4 (18.9 - 19.9) |
| OEHR | 7,365 | 35,861 | 1.9 | 14.4 | 40 | 305 | 21.4 (15.3 - 29.2) | 21.2 (18.9 - 23.7) |
| VA | 1,084 | 15,730 | 0.6 | 10.9 | 11 | 152 | 17.4 (8.67 - 31.1) | 14.0 (11.9 - 16.4) |
| Meta-analysis | 126,893 | 508,201 | 80.0 | 358.6 | 1,201 | 7,416 | 15.0 (14.2 - 15.9) | 20.7 (20.2 - 21.2) |
| Meta-analysis (w/o USOC) | 24,532 | 133,309 | 11.7 | 83.2 | 276 | 2,068 | 23.6 (20.9 - 26.5) | 24.9 (23.8 - 26.0) |
| **PS-stratification / on-treatment TAR without escalation** | | | | | | | | |
| CCAE | 11,380 | 48,125 | 6.4 | 33.7 | 125 | 701 | 19.6 (16.3 - 23.4) | 20.8 (19.3 - 22.4) |
| ODOD | 7,578 | 26,713 | 3.8 | 18.0 | 110 | 670 | 28.9 (23.8 - 34.8) | 37.3 (34.5 - 40.2) |
| MDCR | 819 | 8,073 | 0.4 | 7.1 | 21 | 309 | 47.2 (29.2 - 72.1) | 43.5 (38.8 - 48.7) |
| Open Claims | 138,123 | 377,644 | 83.1 | 277.4 | 1,115 | 5,531 | 13.4 (12.6 - 14.2) | 19.9 (19.4 - 20.5) |
| OEHR | 8,578 | 35,940 | 2.1 | 14.4 | 45 | 311 | 21.3 (15.5 - 28.4) | 21.5 (19.2 - 24.1) |
| VA | 1,260 | 22,854 | 0.7 | 17.3 | 12 | 243 | 17.4 (8.98 - 30.4) | 14.1 (12.4 - 16.0) |
| Meta-analysis | 165,659 | 488,422 | 95.4 | 343.5 | 1,395 | 7,213 | 14.6 (13.9 - 15.4) | 21.0 (20.5 - 21.5) |
| Meta-analysis (w/o USOC) | 27,536 | 110,778 | 12.3 | 66.1 | 280 | 1,682 | 22.8 (20.2 - 25.6) | 25.4 (24.2 - 26.7) |

##### 4.4.17 Hyperkalemia

**Supplementary Table 384:** Exposures, outcomes and risk estimates for Hyperkalemia in GLP1RA (male) (T) vs DPP4I (male) (C) new-user cohorts under four different propensity score (PS) adjustment / time-at-risk (TAR) designs across data sources. We report total patient exposure time in 1,000 patient-year (KYr) units, uncalibrated and calibrated hazard ratios (HRs) and their 95% confidence intervals (CIs). Greyed out entries did not pass study diagnostics and are included only for completeness.

| | Patients | | Exposure (KYr) | | Outcomes | | Incidence (per KYr) | |
|---|---|---|---|---|---|---|---|---|
| | T | C | T | C | T | C | T | C |
| **PS-matching / on-treatment TAR** | | | | | | | | |
| CCAE | 10,450 | 51,940 | 7.2 | 45.3 | 47 | 321 | 6.55 (4.81 - 8.70) | 7.09 (6.34 - 7.91) |
| ODOD | 6,159 | 29,035 | 3.8 | 24.1 | 58 | 498 | 15.3 (11.6 - 19.7) | 20.6 (18.9 - 22.5) |
| MDCR | 834 | 8,711 | 0.6 | 9.1 | 7 | 134 | 12.0 (4.82 - 24.7) | 14.7 (12.3 - 17.4) |
| Open Claims | 113,360 | 417,653 | 87.4 | 398.4 | 351 | 3,208 | 4.02 (3.61 - 4.46) | 8.05 (7.78 - 8.34) |
| OEHR | 7,661 | 37,332 | 2.3 | 18.4 | 19 | 293 | 8.39 (5.05 - 13.1) | 15.9 (14.1 - 17.8) |
| VA | 1,134 | 16,675 | 0.8 | 14.0 | <10 | 189 | < 11.9 (5.72 - 22.0) | 13.5 (11.7 - 15.6) |
| Meta-analysis | 139,598 | 561,346 | 102.1 | 509.3 | <492 | 4,643 | < 4.82 (4.40 - 5.27) | 9.12 (8.86 - 9.38) |
| Meta-analysis (w/o USOC) | 26,238 | 143,693 | 14.7 | 110.9 | <141 | 1,435 | < 9.61 (8.09 - 11.3) | 12.9 (12.3 - 13.6) |
| **PS-stratification / on-treatment TAR** | | | | | | | | |
| CCAE | 12,345 | 52,064 | 8.0 | 45.4 | 52 | 323 | 6.49 (4.85 - 8.51) | 7.12 (6.36 - 7.94) |
| ODOD | 8,212 | 29,533 | 4.7 | 24.6 | 70 | 515 | 15.0 (11.7 - 19.0) | 21.0 (19.2 - 22.9) |
| MDCR | 932 | 9,303 | 0.6 | 9.7 | 7 | 160 | 11.4 (4.59 - 23.5) | 16.5 (14.1 - 19.3) |
| Open Claims | 152,722 | 419,830 | 104.9 | 400.4 | 394 | 3,291 | 3.75 (3.39 - 4.14) | 8.22 (7.94 - 8.50) |
| OEHR | 8,937 | 37,431 | 2.5 | 18.5 | 20 | 294 | 7.85 (4.79 - 12.1) | 15.9 (14.1 - 17.8) |
| VA | 1,317 | 24,038 | 0.9 | 22.1 | <10 | 315 | < 10.9 (5.24 - 20.1) | 14.2 (12.7 - 15.9) |
| Meta-analysis | 182,216 | 538,858 | 120.2 | 488.9 | 536 | 4,423 | 4.46 (4.09 - 4.86) | 9.05 (8.78 - 9.32) |
| Meta-analysis (w/o USOC) | 29,494 | 119,028 | 15.2 | 88.4 | 142 | 1,132 | 9.33 (7.86 - 11.0) | 12.8 (12.1 - 13.6) |
| **PS-matching / on-treatment TAR without escalation** | | | | | | | | |
| CCAE | 10,450 | 51,940 | 6.2 | 36.7 | 37 | 256 | 5.94 (4.18 - 8.18) | 6.98 (6.16 - 7.89) |
| ODOD | 6,159 | 29,035 | 3.4 | 19.8 | 51 | 406 | 15.1 (11.3 - 19.9) | 20.6 (18.6 - 22.7) |
| MDCR | 834 | 8,711 | 0.5 | 7.8 | 6 | 110 | 11.6 (4.27 - 25.3) | 14.1 (11.6 - 17.0) |
| Open Claims | 113,360 | 417,653 | 75.4 | 309.0 | 295 | 2,522 | 3.91 (3.48 - 4.38) | 8.16 (7.85 - 8.49) |
| OEHR | 7,661 | 37,332 | 2.0 | 15.0 | 14 | 222 | 7.16 (3.92 - 12.0) | 14.8 (12.9 - 16.8) |
| VA | 1,134 | 16,675 | 0.7 | 11.4 | <10 | 142 | < 15.3 (7.35 - 28.2) | 12.4 (10.5 - 14.7) |
| Meta-analysis | 139,598 | 561,346 | 88.1 | 399.6 | <413 | 3,658 | < 4.69 (4.25 - 5.16) | 9.15 (8.86 - 9.45) |
| Meta-analysis (w/o USOC) | 26,238 | 143,693 | 12.7 | 90.6 | <118 | 1,136 | < 9.28 (7.68 - 11.1) | 12.5 (11.8 - 13.3) |
| **PS-stratification / on-treatment TAR without escalation** | | | | | | | | |
| CCAE | 12,345 | 52,064 | 7.0 | 36.7 | 40 | 258 | 5.72 (4.09 - 7.79) | 7.02 (6.19 - 7.93) |
| ODOD | 8,212 | 29,533 | 4.2 | 20.1 | 61 | 420 | 14.7 (11.2 - 18.9) | 20.9 (19.0 - 23.0) |
| MDCR | 932 | 9,303 | 0.5 | 8.3 | 6 | 134 | 11.1 (4.06 - 24.1) | 16.2 (13.6 - 19.2) |
| Open Claims | 152,722 | 419,830 | 91.7 | 310.6 | 334 | 2,584 | 3.64 (3.26 - 4.05) | 8.32 (8.00 - 8.65) |
| OEHR | 8,937 | 37,431 | 2.2 | 15.1 | 15 | 223 | 6.77 (3.79 - 11.2) | 14.8 (12.9 - 16.9) |
| VA | 1,317 | 24,038 | 0.7 | 18.0 | <10 | 243 | < 13.9 (6.66 - 25.5) | 13.5 (11.9 - 15.3) |
| Meta-analysis | 182,216 | 538,858 | 105.1 | 382.6 | 450 | 3,485 | 4.28 (3.90 - 4.70) | 9.11 (8.81 - 9.42) |
| Meta-analysis (w/o USOC) | 29,494 | 119,028 | 13.4 | 71.9 | 116 | 901 | 8.68 (7.17 - 10.4) | 12.5 (11.7 - 13.4) |

##### 4.4.18 Hypoglycemia

**Supplementary Table 385:** Exposures, outcomes and risk estimates for Hypoglycemia in GLP1RA (male) (T) vs DPP4I (male) (C) new-user cohorts under four different propensity score (PS) adjustment / time-at-risk (TAR) designs across data sources. We report total patient exposure time in 1,000 patient-year (KYr) units, uncalibrated and calibrated hazard ratios (HRs) and their 95% confidence intervals (CIs). Greyed out entries did not pass study diagnostics and are included only for completeness.

| | Patients | | Exposure (KYr) | | Outcomes | | Incidence (per KYr) | |
|---|---|---|---|---|---|---|---|---|
| | T | C | T | C | T | C | T | C |
| **PS-matching / on-treatment TAR** | | | | | | | | |
| CCAE | 10,442 | 52,040 | 7.2 | 45.7 | 34 | 125 | 4.72 (3.27 - 6.59) | 2.74 (2.28 - 3.26) |
| ODOD | 6,324 | 29,836 | 3.9 | 25.1 | 22 | 124 | 5.57 (3.49 - 8.44) | 4.93 (4.10 - 5.88) |
| MDCR | 847 | 8,828 | 0.6 | 9.3 | <5 | 28 | < 8.48 (2.75 - 19.8) | 3.01 (2.00 - 4.34) |
| Open Claims | 113,979 | 421,712 | 88.0 | 405.0 | 177 | 959 | 2.01 (1.72 - 2.33) | 2.37 (2.22 - 2.52) |
| OEHR | 7,722 | 37,931 | 2.3 | 18.9 | 13 | 103 | 5.69 (3.03 - 9.73) | 5.44 (4.44 - 6.60) |
| VA | 1,200 | 17,540 | 0.9 | 14.9 | 0 | 50 | | 3.36 (2.50 - 4.43) |
| Meta-analysis | 138,467 | 541,519 | 101.5 | 494.7 | 246 | 1,311 | 2.42 (2.13 - 2.75) | 2.65 (2.51 - 2.80) |
| Meta-analysis (w/o USOC) | 24,488 | 119,807 | 13.4 | 89.7 | 69 | 352 | 5.13 (4.00 - 6.50) | 3.92 (3.52 - 4.35) |
| **PS-stratification / on-treatment TAR** | | | | | | | | |
| CCAE | 12,307 | 52,171 | 8.0 | 45.8 | 36 | 125 | 4.49 (3.14 - 6.21) | 2.73 (2.27 - 3.25) |
| ODOD | 8,400 | 30,447 | 4.8 | 25.6 | 25 | 127 | 5.20 (3.36 - 7.67) | 4.96 (4.13 - 5.90) |
| MDCR | 944 | 9,475 | 0.6 | 9.9 | <5 | 34 | < 8.09 (2.63 - 18.9) | 3.42 (2.37 - 4.78) |
| Open Claims | 153,146 | 424,002 | 105.4 | 407.1 | 219 | 972 | 2.08 (1.81 - 2.37) | 2.39 (2.24 - 2.54) |
| OEHR | 8,985 | 38,024 | 2.6 | 19.0 | 15 | 104 | 5.85 (3.27 - 9.64) | 5.48 (4.48 - 6.64) |
| VA | 1,388 | 25,385 | 1.0 | 23.8 | <10 | 78 | < 10.4 (4.99 - 19.1) | 3.28 (2.60 - 4.10) |
| Meta-analysis | 182,838 | 544,644 | 120.8 | 497.5 | 295 | 1,328 | 2.44 (2.17 - 2.74) | 2.67 (2.53 - 2.82) |
| Meta-analysis (w/o USOC) | 29,692 | 120,642 | 15.4 | 90.4 | 76 | 356 | 4.93 (3.89 - 6.18) | 3.94 (3.54 - 4.37) |
| **PS-matching / on-treatment TAR without escalation** | | | | | | | | |
| CCAE | 10,442 | 52,040 | 6.2 | 37.0 | 25 | 83 | 4.01 (2.59 - 5.92) | 2.25 (1.79 - 2.78) |
| ODOD | 6,324 | 29,836 | 3.5 | 20.6 | 16 | 65 | 4.58 (2.62 - 7.44) | 3.16 (2.44 - 4.03) |
| MDCR | 847 | 8,828 | 0.5 | 7.9 | 0 | 16 | | 2.01 (1.15 - 3.27) |
| Open Claims | 113,979 | 421,712 | 76.0 | 314.0 | 133 | 534 | 1.75 (1.47 - 2.07) | 1.70 (1.56 - 1.85) |
| OEHR | 7,722 | 37,931 | 2.0 | 15.4 | 11 | 50 | 5.59 (2.79 - 10.00) | 3.25 (2.41 - 4.28) |
| VA | 1,200 | 17,540 | 0.7 | 12.2 | 0 | 30 | | 2.47 (1.67 - 3.52) |
| Meta-analysis | 138,467 | 541,519 | 87.7 | 387.0 | 185 | 732 | 2.11 (1.82 - 2.44) | 1.89 (1.76 - 2.03) |
| Meta-analysis (w/o USOC) | 24,488 | 119,807 | 11.7 | 72.9 | 52 | 198 | 4.45 (3.32 - 5.83) | 2.71 (2.35 - 3.12) |
| **PS-stratification / on-treatment TAR without escalation** | | | | | | | | |
| CCAE | 12,307 | 52,171 | 7.0 | 37.1 | 27 | 83 | 3.86 (2.54 - 5.62) | 2.24 (1.78 - 2.78) |
| ODOD | 8,400 | 30,447 | 4.3 | 21.0 | 19 | 67 | 4.44 (2.67 - 6.94) | 3.19 (2.47 - 4.05) |
| MDCR | 944 | 9,475 | 0.5 | 8.5 | 0 | 20 | | 2.36 (1.44 - 3.64) |
| Open Claims | 153,146 | 424,002 | 92.1 | 315.7 | 173 | 537 | 1.88 (1.61 - 2.18) | 1.70 (1.56 - 1.85) |
| OEHR | 8,985 | 38,024 | 2.2 | 15.4 | 12 | 51 | 5.39 (2.78 - 9.41) | 3.30 (2.46 - 4.34) |
| VA | 1,388 | 25,385 | 0.8 | 19.3 | 0 | 44 | | 2.28 (1.66 - 3.06) |
| Meta-analysis | 182,838 | 544,644 | 105.6 | 389.2 | 231 | 738 | 2.19 (1.91 - 2.49) | 1.90 (1.76 - 2.04) |
| Meta-analysis (w/o USOC) | 29,692 | 120,642 | 13.5 | 73.5 | 58 | 201 | 4.30 (3.26 - 5.56) | 2.73 (2.37 - 3.14) |

##### 4.4.19 Hypotension

**Supplementary Table 386:** Exposures, outcomes and risk estimates for Hypotension in GLP1RA (male) (T) vs DPP4I (male) (C) new-user cohorts under four different propensity score (PS) adjustment / time-at-risk (TAR) designs across data sources. We report total patient exposure time in 1,000 patient-year (KYr) units, uncalibrated and calibrated hazard ratios (HRs) and their 95% confidence intervals (CIs). Greyed out entries did not pass study diagnostics and are included only for completeness.

| | Patients | | Exposure (KYr) | | Outcomes | | Incidence (per KYr) | |
|---|---|---|---|---|---|---|---|---|
| | T | C | T | C | T | C | T | C |
| **PS-matching / on-treatment TAR** | | | | | | | | |
| CCAE | 10,416 | 51,890 | 7.2 | 45.3 | 70 | 346 | 9.78 (7.63 - 12.4) | 7.63 (6.85 - 8.48) |
| ODOD | 6,168 | 28,752 | 3.8 | 24.0 | 66 | 487 | 17.2 (13.3 - 21.9) | 20.3 (18.5 - 22.1) |
| MDCR | 818 | 8,345 | 0.6 | 8.7 | 15 | 225 | 26.4 (14.8 - 43.6) | 25.9 (22.6 - 29.5) |
| Open Claims | 111,950 | 412,509 | 86.4 | 394.4 | 541 | 3,678 | 6.26 (5.74 - 6.81) | 9.33 (9.03 - 9.63) |
| OEHR | 7,635 | 37,341 | 2.3 | 18.4 | 37 | 338 | 16.4 (11.6 - 22.6) | 18.3 (16.4 - 20.4) |
| VA | 1,130 | 16,781 | 0.8 | 14.1 | 15 | 178 | 17.8 (9.97 - 29.4) | 12.6 (10.8 - 14.6) |
| Meta-analysis | 138,117 | 555,618 | 101.1 | 505.0 | 744 | 5,252 | 7.36 (6.84 - 7.91) | 10.4 (10.1 - 10.7) |
| Meta-analysis (w/o USOC) | 26,167 | 143,109 | 14.7 | 110.6 | 203 | 1,574 | 13.9 (12.0 - 15.9) | 14.2 (13.5 - 14.9) |
| **PS-stratification / on-treatment TAR** | | | | | | | | |
| CCAE | 12,285 | 52,015 | 8.0 | 45.5 | 75 | 348 | 9.40 (7.39 - 11.8) | 7.66 (6.87 - 8.50) |
| ODOD | 8,191 | 29,398 | 4.7 | 24.6 | 76 | 520 | 16.3 (12.8 - 20.4) | 21.2 (19.4 - 23.1) |
| MDCR | 914 | 8,981 | 0.6 | 9.3 | 16 | 263 | 26.7 (15.3 - 43.3) | 28.3 (25.0 - 32.0) |
| Open Claims | 150,746 | 414,545 | 103.6 | 396.2 | 624 | 3,770 | 6.02 (5.56 - 6.51) | 9.51 (9.21 - 9.82) |
| OEHR | 8,903 | 37,424 | 2.5 | 18.5 | 38 | 345 | 15.0 (10.6 - 20.6) | 18.7 (16.7 - 20.7) |
| VA | 1,316 | 24,071 | 0.9 | 22.3 | 18 | 285 | 19.6 (11.6 - 31.0) | 12.8 (11.4 - 14.4) |
| Meta-analysis | 180,125 | 533,382 | 118.8 | 484.7 | 813 | 4,983 | 6.84 (6.38 - 7.33) | 10.3 (10.00 - 10.6) |
| Meta-analysis (w/o USOC) | 29,379 | 118,837 | 15.2 | 88.5 | 189 | 1,213 | 12.4 (10.7 - 14.4) | 13.7 (12.9 - 14.5) |
| **PS-matching / on-treatment TAR without escalation** | | | | | | | | |
| CCAE | 10,416 | 51,890 | 6.2 | 36.7 | 57 | 276 | 9.21 (6.97 - 11.9) | 7.53 (6.66 - 8.47) |
| ODOD | 6,168 | 28,752 | 3.4 | 19.7 | 56 | 378 | 16.5 (12.5 - 21.5) | 19.2 (17.3 - 21.3) |
| MDCR | 818 | 8,345 | 0.5 | 7.4 | 12 | 189 | 23.9 (12.3 - 41.7) | 25.5 (22.0 - 29.4) |
| Open Claims | 111,950 | 412,509 | 74.5 | 305.7 | 458 | 2,844 | 6.15 (5.60 - 6.73) | 9.30 (8.96 - 9.65) |
| OEHR | 7,635 | 37,341 | 1.9 | 15.1 | 36 | 235 | 18.5 (13.0 - 25.6) | 15.6 (13.7 - 17.7) |
| VA | 1,130 | 16,781 | 0.7 | 11.6 | 13 | 134 | 19.9 (10.6 - 34.1) | 11.6 (9.71 - 13.7) |
| Meta-analysis | 138,117 | 555,618 | 87.2 | 396.1 | 632 | 4,056 | 7.25 (6.69 - 7.84) | 10.2 (9.93 - 10.6) |
| Meta-analysis (w/o USOC) | 26,167 | 143,109 | 12.7 | 90.4 | 174 | 1,212 | 13.7 (11.8 - 15.9) | 13.4 (12.7 - 14.2) |
| **PS-stratification / on-treatment TAR without escalation** | | | | | | | | |
| CCAE | 12,285 | 52,015 | 6.9 | 36.8 | 61 | 278 | 8.78 (6.72 - 11.3) | 7.56 (6.70 - 8.50) |
| ODOD | 8,191 | 29,398 | 4.1 | 20.1 | 65 | 407 | 15.7 (12.1 - 20.0) | 20.3 (18.3 - 22.3) |
| MDCR | 914 | 8,981 | 0.5 | 7.9 | 13 | 226 | 24.4 (13.0 - 41.8) | 28.5 (24.9 - 32.5) |
| Open Claims | 150,746 | 414,545 | 90.6 | 307.2 | 534 | 2,921 | 5.90 (5.41 - 6.42) | 9.51 (9.17 - 9.86) |
| OEHR | 8,903 | 37,424 | 2.2 | 15.1 | 37 | 240 | 16.8 (11.8 - 23.2) | 15.9 (13.9 - 18.0) |
| VA | 1,316 | 24,071 | 0.7 | 18.1 | 15 | 210 | 20.9 (11.7 - 34.5) | 11.6 (10.1 - 13.3) |
| Meta-analysis | 180,125 | 533,382 | 103.9 | 379.2 | 697 | 3,846 | 6.71 (6.22 - 7.23) | 10.1 (9.82 - 10.5) |
| Meta-analysis (w/o USOC) | 29,379 | 118,837 | 13.3 | 72.0 | 163 | 925 | 12.3 (10.4 - 14.3) | 12.9 (12.0 - 13.7) |

##### 4.4.20 Joint pain

**Supplementary Table 387:** Exposures, outcomes and risk estimates for Joint pain in GLP1RA (male) (T) vs DPP4I (male) (C) new-user cohorts under four different propensity score (PS) adjustment / time-at-risk (TAR) designs across data sources. We report total patient exposure time in 1,000 patient-year (KYr) units, uncalibrated and calibrated hazard ratios (HRs) and their 95% confidence intervals (CIs). Greyed out entries did not pass study diagnostics and are included only for completeness.

| | Patients | | Exposure (KYr) | | Outcomes | | Incidence (per KYr) | |
|---|---|---|---|---|---|---|---|---|
| | T | C | T | C | T | C | T | C |
| **PS-matching / on-treatment TAR** | | | | | | | | |
| CCAE | 10,133 | 50,771 | 6.9 | 44.3 | 55 | 376 | 7.92 (5.97 - 10.3) | 8.48 (7.65 - 9.38) |
| ODOD | 6,076 | 28,925 | 3.8 | 24.2 | 31 | 261 | 8.22 (5.59 - 11.7) | 10.8 (9.50 - 12.2) |
| MDCR | 823 | 8,563 | 0.6 | 9.0 | <5 | 83 | < 8.73 (2.84 - 20.4) | 9.21 (7.33 - 11.4) |
| Open Claims | 109,630 | 408,535 | 84.6 | 391.2 | 397 | 2,067 | 4.69 (4.24 - 5.18) | 5.28 (5.06 - 5.52) |
| OEHR | 7,460 | 36,748 | 2.2 | 18.2 | 13 | 151 | 5.91 (3.15 - 10.1) | 8.28 (7.01 - 9.71) |
| VA | 1,078 | 15,142 | 0.8 | 12.9 | <10 | 71 | < 12.5 (5.97 - 22.9) | 5.52 (4.31 - 6.97) |
| Meta-analysis | 135,200 | 548,684 | 98.9 | 499.9 | <511 | 3,009 | < 5.17 (4.73 - 5.63) | 6.02 (5.81 - 6.24) |
| Meta-analysis (w/o USOC) | 25,570 | 140,149 | 14.3 | 108.7 | <114 | 942 | < 7.98 (6.58 - 9.58) | 8.67 (8.12 - 9.24) |
| **PS-stratification / on-treatment TAR** | | | | | | | | |
| CCAE | 11,936 | 50,908 | 7.7 | 44.5 | 62 | 376 | 8.02 (6.15 - 10.3) | 8.46 (7.63 - 9.36) |
| ODOD | 8,069 | 29,493 | 4.6 | 24.7 | 44 | 265 | 9.57 (6.96 - 12.9) | 10.7 (9.47 - 12.1) |
| MDCR | 913 | 9,268 | 0.6 | 9.7 | <5 | 90 | < 8.34 (2.71 - 19.5) | 9.29 (7.47 - 11.4) |
| Open Claims | 147,146 | 410,683 | 101.3 | 393.3 | 492 | 2,082 | 4.86 (4.44 - 5.31) | 5.29 (5.07 - 5.53) |
| OEHR | 8,667 | 36,838 | 2.5 | 18.3 | 16 | 152 | 6.48 (3.70 - 10.5) | 8.31 (7.04 - 9.74) |
| VA | 1,247 | 22,503 | 0.9 | 21.2 | 12 | 108 | 13.8 (7.15 - 24.2) | 5.09 (4.18 - 6.15) |
| Meta-analysis | 175,818 | 527,922 | 116.1 | 480.7 | 614 | 2,875 | 5.29 (4.88 - 5.72) | 5.98 (5.76 - 6.20) |
| Meta-analysis (w/o USOC) | 28,672 | 117,239 | 14.8 | 87.4 | 122 | 793 | 8.24 (6.85 - 9.84) | 9.07 (8.45 - 9.72) |
| **PS-matching / on-treatment TAR without escalation** | | | | | | | | |
| CCAE | 10,133 | 50,771 | 6.0 | 35.9 | 46 | 310 | 7.63 (5.59 - 10.2) | 8.64 (7.70 - 9.65) |
| ODOD | 6,076 | 28,925 | 3.3 | 19.9 | 27 | 222 | 8.10 (5.34 - 11.8) | 11.2 (9.76 - 12.8) |
| MDCR | 823 | 8,563 | 0.5 | 7.7 | <5 | 70 | < 9.91 (3.22 - 23.1) | 9.12 (7.11 - 11.5) |
| Open Claims | 109,630 | 408,535 | 73.0 | 303.5 | 333 | 1,569 | 4.56 (4.09 - 5.08) | 5.17 (4.92 - 5.43) |
| OEHR | 7,460 | 36,748 | 1.9 | 14.8 | 13 | 119 | 6.85 (3.65 - 11.7) | 8.01 (6.64 - 9.59) |
| VA | 1,078 | 15,142 | 0.6 | 10.5 | <10 | 56 | < 15.9 (7.61 - 29.2) | 5.32 (4.02 - 6.90) |
| Meta-analysis | 135,200 | 548,684 | 85.4 | 392.3 | <434 | 2,346 | < 5.08 (4.62 - 5.58) | 5.98 (5.74 - 6.23) |
| Meta-analysis (w/o USOC) | 25,570 | 140,149 | 12.4 | 88.8 | <101 | 777 | < 8.15 (6.64 - 9.90) | 8.75 (8.14 - 9.39) |
| **PS-stratification / on-treatment TAR without escalation** | | | | | | | | |
| CCAE | 11,936 | 50,908 | 6.8 | 36.0 | 52 | 310 | 7.70 (5.75 - 10.1) | 8.61 (7.68 - 9.63) |
| ODOD | 8,069 | 29,493 | 4.1 | 20.2 | 39 | 226 | 9.53 (6.78 - 13.0) | 11.2 (9.75 - 12.7) |
| MDCR | 913 | 9,268 | 0.5 | 8.3 | <5 | 76 | < 9.43 (3.06 - 22.0) | 9.20 (7.25 - 11.5) |
| Open Claims | 147,146 | 410,683 | 88.5 | 305.0 | 423 | 1,578 | 4.78 (4.34 - 5.26) | 5.17 (4.92 - 5.43) |
| OEHR | 8,667 | 36,838 | 2.1 | 14.9 | 16 | 120 | 7.45 (4.26 - 12.1) | 8.06 (6.68 - 9.64) |
| VA | 1,247 | 22,503 | 0.7 | 17.2 | 11 | 85 | 16.0 (8.00 - 28.7) | 4.93 (3.94 - 6.10) |
| Meta-analysis | 175,818 | 527,922 | 101.5 | 376.2 | 530 | 2,234 | 5.22 (4.79 - 5.69) | 5.94 (5.69 - 6.19) |
| Meta-analysis (w/o USOC) | 28,672 | 117,239 | 13.0 | 71.1 | 107 | 656 | 8.24 (6.75 - 9.95) | 9.22 (8.53 - 9.96) |

###### 4.4.21 Lower extremity amputation

**Supplementary Table 388:** Exposures, outcomes and risk estimates for Lower extremity amputation in GLP1RA (male) (T) vs DPP4I (male) (C) new-user cohorts under four different propensity score (PS) adjustment / time-at-risk (TAR) designs across data sources. We report total patient exposure time in 1,000 patient-year (KYr) units, uncalibrated and calibrated hazard ratios (HRs) and their 95% confidence intervals (CIs). Greyed out entries did not pass study diagnostics and are included only for completeness.

| | Patients | | Exposure (KYr) | | Outcomes | | Incidence (per KYr) | |
|---|---|---|---|---|---|---|---|---|
| | T | C | T | C | T | C | T | C |
| **PS-matching / on-treatment TAR** | | | | | | | | |
| CCAE | 10,595 | 52,689 | 7.3 | 46.3 | 6 | 33 | 0.82 (0.30 - 1.78) | 0.71 (0.49 - 1.00) |
| ODOD | 6,392 | 30,161 | 4.0 | 25.5 | <5 | 26 | < 1.25 (0.41 - 2.92) | 1.02 (0.67 - 1.49) |
| MDCR | 854 | 8,887 | 0.6 | 9.4 | 0 | 8 | | 0.85 (0.37 - 1.68) |
| Open Claims | 115,432 | 426,667 | 89.2 | 410.0 | 46 | 239 | 0.52 (0.38 - 0.69) | 0.58 (0.51 - 0.66) |
| OEHR | 7,797 | 38,157 | 2.3 | 19.1 | 0 | 16 | | 0.84 (0.48 - 1.36) |
| VA | 1,210 | 17,651 | 0.9 | 15.0 | 0 | <10 | | < 0.67 (0.32 - 1.23) |
| Meta-analysis | 132,419 | 509,517 | 100.5 | 481.9 | <57 | 298 | < 0.57 (0.43 - 0.73) | 0.62 (0.55 - 0.69) |
| Meta-analysis (w/o USOC) | 16,987 | 82,850 | 11.3 | 71.8 | <11 | 59 | < 0.97 (0.49 - 1.74) | 0.82 (0.63 - 1.06) |
| **PS-stratification / on-treatment TAR** | | | | | | | | |
| CCAE | 12,517 | 52,823 | 8.2 | 46.5 | 7 | 33 | 0.86 (0.34 - 1.77) | 0.71 (0.49 - 1.00) |
| ODOD | 8,500 | 30,785 | 4.9 | 26.0 | 6 | 28 | 1.23 (0.45 - 2.68) | 1.08 (0.71 - 1.56) |
| MDCR | 952 | 9,542 | 0.6 | 10.0 | 0 | 10 | | 1.00 (0.48 - 1.84) |
| Open Claims | 155,300 | 428,959 | 107.0 | 412.2 | 49 | 244 | 0.46 (0.34 - 0.61) | 0.59 (0.52 - 0.67) |
| OEHR | 9,096 | 38,250 | 2.6 | 19.2 | 0 | 16 | | 0.83 (0.48 - 1.35) |
| VA | 1,403 | 25,577 | 1.0 | 24.0 | 0 | 13 | | 0.54 (0.29 - 0.93) |
| Meta-analysis | 176,317 | 512,567 | 120.0 | 484.7 | 62 | 305 | 0.52 (0.40 - 0.66) | 0.63 (0.56 - 0.70) |
| Meta-analysis (w/o USOC) | 21,017 | 83,608 | 13.0 | 72.5 | 13 | 61 | 1.00 (0.53 - 1.71) | 0.84 (0.64 - 1.08) |
| **PS-matching / on-treatment TAR without escalation** | | | | | | | | |
| CCAE | 10,595 | 52,689 | 6.3 | 37.5 | 6 | 25 | 0.95 (0.35 - 2.06) | 0.67 (0.43 - 0.99) |
| ODOD | 6,392 | 30,161 | 3.5 | 20.8 | <5 | 19 | < 1.42 (0.46 - 3.30) | 0.91 (0.55 - 1.42) |
| MDCR | 854 | 8,887 | 0.5 | 8.0 | 0 | 8 | | 1.00 (0.43 - 1.97) |
| Open Claims | 115,432 | 426,667 | 76.9 | 317.7 | 37 | 182 | 0.48 (0.34 - 0.66) | 0.57 (0.49 - 0.66) |
| OEHR | 7,797 | 38,157 | 2.0 | 15.5 | 0 | 11 | | 0.71 (0.35 - 1.27) |
| VA | 1,210 | 17,651 | 0.7 | 12.2 | 0 | <10 | | < 0.82 (0.39 - 1.50) |
| Meta-analysis | 126,027 | 479,356 | 83.3 | 355.2 | 43 | 207 | 0.52 (0.37 - 0.70) | 0.58 (0.51 - 0.67) |
| Meta-analysis (w/o USOC) | 10,595 | 52,689 | 6.3 | 37.5 | 6 | 25 | 0.95 (0.35 - 2.06) | 0.67 (0.43 - 0.99) |
| **PS-stratification / on-treatment TAR without escalation** | | | | | | | | |
| CCAE | 12,517 | 52,823 | 7.1 | 37.6 | 6 | 25 | 0.84 (0.31 - 1.84) | 0.67 (0.43 - 0.98) |
| ODOD | 8,500 | 30,785 | 4.3 | 21.3 | 5 | 21 | 1.16 (0.38 - 2.70) | 0.99 (0.61 - 1.51) |
| MDCR | 952 | 9,542 | 0.6 | 8.5 | 0 | 10 | | 1.17 (0.56 - 2.15) |
| Open Claims | 155,300 | 428,959 | 93.4 | 319.4 | 39 | 185 | 0.42 (0.30 - 0.57) | 0.58 (0.50 - 0.67) |
| OEHR | 9,096 | 38,250 | 2.3 | 15.6 | 0 | 11 | | 0.71 (0.35 - 1.26) |
| VA | 1,403 | 25,577 | 0.8 | 19.5 | 0 | 11 | | 0.57 (0.28 - 1.01) |
| Meta-analysis | 176,317 | 512,567 | 104.9 | 378.2 | 50 | 231 | 0.48 (0.35 - 0.63) | 0.61 (0.53 - 0.69) |
| Meta-analysis (w/o USOC) | 21,017 | 83,608 | 11.4 | 58.8 | 11 | 46 | 0.96 (0.48 - 1.72) | 0.78 (0.57 - 1.04) |

##### 4.4.22 Nausea

**Supplementary Table 389:** Exposures, outcomes and risk estimates for Nausea in GLP1RA (male) (T) vs DPP4I (male) (C) new-user cohorts under four different propensity score (PS) adjustment / time-at-risk (TAR) designs across data sources. We report total patient exposure time in 1,000 patient-year (KYr) units, uncalibrated and calibrated hazard ratios (HRs) and their 95% confidence intervals (CIs). Greyed out entries did not pass study diagnostics and are included only for completeness.

| | Patients | | Exposure (KYr) | | Outcomes | | Incidence (per KYr) | |
|---|---|---|---|---|---|---|---|---|
| | T | C | T | C | T | C | T | C |
| **PS-matching / on-treatment TAR** | | | | | | | | |
| CCAE | 9,578 | 48,902 | 6.5 | 42.4 | 221 | 971 | 33.7 (29.4 - 38.5) | 22.9 (21.5 - 24.4) |
| ODOD | 5,655 | 27,190 | 3.5 | 22.5 | 151 | 701 | 43.6 (37.0 - 51.2) | 31.1 (28.8 - 33.5) |
| MDCR | 783 | 8,162 | 0.5 | 8.5 | 12 | 209 | 22.3 (11.5 - 39.0) | 24.6 (21.3 - 28.1) |
| Open Claims | 102,841 | 388,233 | 79.3 | 370.9 | 1,572 | 5,699 | 19.8 (18.8 - 20.8) | 15.4 (15.0 - 15.8) |
| OEHR | 7,171 | 35,809 | 2.1 | 17.5 | 106 | 457 | 50.6 (41.4 - 61.2) | 26.0 (23.7 - 28.6) |
| VA | 1,133 | 16,631 | 0.8 | 14.2 | 22 | 121 | 27.1 (17.0 - 41.0) | 8.52 (7.07 - 10.2) |
| Meta-analysis | 127,161 | 524,927 | 92.8 | 476.1 | 2,084 | 8,158 | 22.5 (21.5 - 23.4) | 17.1 (16.8 - 17.5) |
| Meta-analysis (w/o USOC) | 24,320 | 136,694 | 13.5 | 105.2 | 512 | 2,459 | 38.1 (34.8 - 41.5) | 23.4 (22.5 - 24.3) |
| **PS-stratification / on-treatment TAR** | | | | | | | | |
| CCAE | 11,233 | 49,023 | 7.3 | 42.5 | 258 | 977 | 35.5 (31.3 - 40.1) | 23.0 (21.6 - 24.5) |
| ODOD | 7,529 | 27,806 | 4.2 | 23.0 | 197 | 724 | 46.7 (40.4 - 53.7) | 31.4 (29.2 - 33.8) |
| MDCR | 870 | 8,802 | 0.6 | 9.1 | 12 | 238 | 21.3 (11.0 - 37.1) | 26.1 (22.9 - 29.6) |
| Open Claims | 137,078 | 390,524 | 94.4 | 373.0 | 2,003 | 5,784 | 21.2 (20.3 - 22.2) | 15.5 (15.1 - 15.9) |
| OEHR | 8,322 | 35,895 | 2.3 | 17.6 | 125 | 462 | 53.3 (44.4 - 63.6) | 26.3 (23.9 - 28.8) |
| VA | 1,316 | 23,930 | 0.9 | 22.4 | 22 | 192 | 24.7 (15.5 - 37.4) | 8.56 (7.39 - 9.85) |
| Meta-analysis | 164,162 | 503,248 | 108.2 | 456.1 | 2,583 | 7,947 | 23.9 (23.0 - 24.8) | 17.4 (17.0 - 17.8) |
| Meta-analysis (w/o USOC) | 27,084 | 112,724 | 13.8 | 83.1 | 580 | 2,163 | 42.0 (38.6 - 45.5) | 26.0 (24.9 - 27.1) |
| **PS-matching / on-treatment TAR without escalation** | | | | | | | | |
| CCAE | 9,578 | 48,902 | 5.7 | 34.4 | 197 | 800 | 34.7 (30.0 - 39.9) | 23.2 (21.7 - 24.9) |
| ODOD | 5,655 | 27,190 | 3.1 | 18.5 | 140 | 576 | 45.6 (38.4 - 53.8) | 31.1 (28.6 - 33.7) |
| MDCR | 783 | 8,162 | 0.5 | 7.2 | 9 | 180 | 18.8 (8.61 - 35.8) | 24.8 (21.3 - 28.8) |
| Open Claims | 102,841 | 388,233 | 68.6 | 288.6 | 1,358 | 4,337 | 19.8 (18.8 - 20.9) | 15.0 (14.6 - 15.5) |
| OEHR | 7,171 | 35,809 | 1.8 | 14.4 | 88 | 336 | 48.3 (38.7 - 59.5) | 23.4 (20.9 - 26.0) |
| VA | 1,133 | 16,631 | 0.6 | 11.6 | 19 | 97 | 29.6 (17.8 - 46.2) | 8.34 (6.76 - 10.2) |
| Meta-analysis | 127,161 | 524,927 | 80.3 | 374.8 | 1,811 | 6,326 | 22.6 (21.5 - 23.6) | 16.9 (16.5 - 17.3) |
| Meta-analysis (w/o USOC) | 24,320 | 136,694 | 11.7 | 86.2 | 453 | 1,989 | 38.8 (35.3 - 42.5) | 23.1 (22.1 - 24.1) |
| **PS-stratification / on-treatment TAR without escalation** | | | | | | | | |
| CCAE | 11,233 | 49,023 | 6.3 | 34.5 | 230 | 801 | 36.3 (31.8 - 41.3) | 23.2 (21.6 - 24.9) |
| ODOD | 7,529 | 27,806 | 3.8 | 18.9 | 179 | 596 | 47.5 (40.8 - 54.9) | 31.5 (29.0 - 34.1) |
| MDCR | 870 | 8,802 | 0.5 | 7.8 | 9 | 207 | 17.9 (8.18 - 34.0) | 26.6 (23.1 - 30.5) |
| Open Claims | 137,078 | 390,524 | 82.6 | 290.3 | 1,764 | 4,408 | 21.3 (20.4 - 22.4) | 15.2 (14.7 - 15.6) |
| OEHR | 8,322 | 35,895 | 2.0 | 14.4 | 107 | 340 | 52.2 (42.8 - 63.1) | 23.6 (21.2 - 26.2) |
| VA | 1,316 | 23,930 | 0.7 | 18.3 | 19 | 158 | 26.8 (16.1 - 41.8) | 8.64 (7.35 - 10.1) |
| Meta-analysis | 164,162 | 503,248 | 94.8 | 358.1 | 2,280 | 6,145 | 24.1 (23.1 - 25.1) | 17.2 (16.7 - 17.6) |
| Meta-analysis (w/o USOC) | 27,084 | 112,724 | 12.2 | 67.9 | 516 | 1,737 | 42.4 (38.9 - 46.3) | 25.6 (24.4 - 26.8) |

##### 4.4.23 Peripheral edema

**Supplementary Table 390:** Exposures, outcomes and risk estimates for Peripheral edema in GLP1RA (male) (T) vs DPP4I (male) (C) new-user cohorts under four different propensity score (PS) adjustment / time-at-risk (TAR) designs across data sources. We report total patient exposure time in 1,000 patient-year (KYr) units, uncalibrated and calibrated hazard ratios (HRs) and their 95% confidence intervals (CIs). Greyed out entries did not pass study diagnostics and are included only for completeness.

| | Patients | | Exposure (KYr) | | Outcomes | | Incidence (per KYr) | |
|---|---|---|---|---|---|---|---|---|
| | T | C | T | C | T | C | T | C |
| **PS-matching / on-treatment TAR** | | | | | | | | |
| CCAE | 8,834 | 45,734 | 6.0 | 38.9 | 206 | 1,437 | 34.6 (30.0 - 39.6) | 36.9 (35.0 - 38.9) |
| ODOD | 5,129 | 24,462 | 3.1 | 19.8 | 155 | 1,155 | 49.4 (41.9 - 57.8) | 58.5 (55.1 - 61.9) |
| MDCR | 646 | 6,936 | 0.4 | 6.9 | 26 | 526 | 60.9 (39.8 - 89.2) | 75.8 (69.5 - 82.6) |
| Open Claims | 92,057 | 348,757 | 70.6 | 326.9 | 1,404 | 9,744 | 19.9 (18.9 - 21.0) | 29.8 (29.2 - 30.4) |
| OEHR | 6,629 | 33,499 | 1.9 | 16.1 | 73 | 624 | 38.2 (30.0 - 48.1) | 38.7 (35.7 - 41.9) |
| VA | 921 | 13,982 | 0.7 | 11.6 | 16 | 377 | 23.8 (13.6 - 38.6) | 32.5 (29.3 - 36.0) |
| Meta-analysis | 114,216 | 473,370 | 82.7 | 420.2 | 1,880 | 13,863 | 22.7 (21.7 - 23.8) | 33.0 (32.4 - 33.5) |
| Meta-analysis (w/o USOC) | 22,159 | 124,613 | 12.1 | 93.3 | 476 | 4,119 | 39.3 (35.9 - 43.0) | 44.1 (42.8 - 45.5) |
| **PS-stratification / on-treatment TAR** | | | | | | | | |
| CCAE | 10,344 | 45,839 | 6.6 | 39.0 | 229 | 1,443 | 34.6 (30.3 - 39.4) | 37.0 (35.1 - 38.9) |
| ODOD | 6,702 | 24,995 | 3.8 | 20.2 | 185 | 1,204 | 49.1 (42.3 - 56.7) | 59.7 (56.3 - 63.1) |
| MDCR | 730 | 7,379 | 0.5 | 7.3 | 28 | 597 | 61.5 (40.9 - 88.9) | 81.7 (75.3 - 88.6) |
| Open Claims | 122,153 | 350,794 | 83.8 | 328.6 | 1,666 | 9,964 | 19.9 (18.9 - 20.9) | 30.3 (29.7 - 30.9) |
| OEHR | 7,667 | 33,645 | 2.1 | 16.2 | 79 | 637 | 37.0 (29.3 - 46.2) | 39.4 (36.4 - 42.5) |
| VA | 1,029 | 20,139 | 0.7 | 18.4 | 16 | 620 | 22.2 (12.7 - 36.1) | 33.7 (31.1 - 36.5) |
| Meta-analysis | 146,866 | 455,273 | 96.3 | 404.0 | 2,159 | 13,248 | 22.4 (21.5 - 23.4) | 32.8 (32.2 - 33.4) |
| Meta-analysis (w/o USOC) | 24,713 | 104,479 | 12.5 | 75.4 | 493 | 3,284 | 39.4 (36.0 - 43.0) | 43.6 (42.1 - 45.1) |
| **PS-matching / on-treatment TAR without escalation** | | | | | | | | |
| CCAE | 8,834 | 45,734 | 5.2 | 31.7 | 172 | 1,155 | 33.0 (28.3 - 38.3) | 36.4 (34.3 - 38.5) |
| ODOD | 5,129 | 24,462 | 2.8 | 16.3 | 139 | 954 | 50.0 (42.0 - 59.0) | 58.4 (54.8 - 62.2) |
| MDCR | 646 | 6,936 | 0.4 | 6.0 | 22 | 440 | 58.5 (36.7 - 88.6) | 73.6 (66.9 - 80.8) |
| Open Claims | 92,057 | 348,757 | 61.0 | 254.5 | 1,203 | 7,568 | 19.7 (18.6 - 20.9) | 29.7 (29.1 - 30.4) |
| OEHR | 6,629 | 33,499 | 1.6 | 13.3 | 60 | 461 | 36.4 (27.8 - 46.8) | 34.7 (31.6 - 38.1) |
| VA | 921 | 13,982 | 0.5 | 9.5 | 12 | 303 | 22.8 (11.8 - 39.8) | 31.8 (28.3 - 35.6) |
| Meta-analysis | 114,216 | 473,370 | 71.6 | 331.3 | 1,608 | 10,881 | 22.5 (21.4 - 23.6) | 32.8 (32.2 - 33.5) |
| Meta-analysis (w/o USOC) | 22,159 | 124,613 | 10.5 | 76.8 | 405 | 3,313 | 38.4 (34.8 - 42.3) | 43.1 (41.7 - 44.6) |
| **PS-stratification / on-treatment TAR without escalation** | | | | | | | | |
| CCAE | 10,344 | 45,839 | 5.8 | 31.8 | 194 | 1,159 | 33.4 (28.9 - 38.5) | 36.4 (34.3 - 38.6) |
| ODOD | 6,702 | 24,995 | 3.4 | 16.7 | 168 | 994 | 49.9 (42.7 - 58.1) | 59.5 (55.9 - 63.3) |
| MDCR | 730 | 7,379 | 0.4 | 6.3 | 24 | 502 | 59.5 (38.1 - 88.6) | 79.6 (72.8 - 86.9) |
| Open Claims | 122,153 | 350,794 | 73.3 | 256.0 | 1,445 | 7,757 | 19.7 (18.7 - 20.8) | 30.3 (29.6 - 31.0) |
| OEHR | 7,667 | 33,645 | 1.9 | 13.3 | 65 | 467 | 35.0 (27.0 - 44.7) | 35.0 (31.9 - 38.4) |
| VA | 1,029 | 20,139 | 0.6 | 15.0 | 12 | 491 | 21.2 (11.0 - 37.0) | 32.8 (29.9 - 35.8) |
| Meta-analysis | 146,866 | 455,273 | 84.3 | 317.8 | 1,872 | 10,377 | 22.2 (21.2 - 23.2) | 32.7 (32.0 - 33.3) |
| Meta-analysis (w/o USOC) | 24,713 | 104,479 | 11.0 | 61.9 | 427 | 2,620 | 38.7 (35.1 - 42.6) | 42.4 (40.7 - 44.0) |

##### 4.4.24 Photosensitivity

**Supplementary Table 391:** Exposures, outcomes and risk estimates for Photosensitivity in GLP1RA (male) (T) vs DPP4I (male) (C) new-user cohorts under four different propensity score (PS) adjustment / time-at-risk (TAR) designs across data sources. We report total patient exposure time in 1,000 patient-year (KYr) units, uncalibrated and calibrated hazard ratios (HRs) and their 95% confidence intervals (CIs). Greyed out entries did not pass study diagnostics and are included only for completeness.

| | Patients | | Exposure (KYr) | | Outcomes | | Incidence (per KYr) | |
|---|---|---|---|---|---|---|---|---|
| | T | C | T | C | T | C | T | C |
| **PS-matching / on-treatment TAR** | | | | | | | | |
| CCAE | 10,525 | 52,283 | 7.3 | 45.9 | 9 | 64 | 1.24 (0.57 - 2.35) | 1.39 (1.07 - 1.78) |
| ODOD | 6,375 | 29,904 | 4.0 | 25.2 | <5 | 47 | < 1.26 (0.41 - 2.93) | 1.86 (1.37 - 2.48) |
| MDCR | 836 | 8,723 | 0.6 | 9.1 | <5 | 43 | < 8.54 (2.77 - 19.9) | 4.70 (3.40 - 6.34) |
| Open Claims | 114,514 | 423,044 | 88.4 | 406.0 | 27 | 220 | 0.31 (0.20 - 0.44) | 0.54 (0.47 - 0.62) |
| OEHR | 7,758 | 37,977 | 2.3 | 19.0 | 5 | 23 | 2.18 (0.71 - 5.09) | 1.21 (0.77 - 1.82) |
| VA | 1,195 | 17,491 | 0.9 | 14.9 | 0 | <10 | | < 0.67 (0.32 - 1.24) |
| Meta-analysis | 131,414 | 505,231 | 99.6 | 477.1 | <41 | 331 | < 0.41 (0.30 - 0.56) | 0.69 (0.62 - 0.77) |
| Meta-analysis (w/o USOC) | 16,900 | 82,187 | 11.3 | 71.1 | <14 | 111 | < 1.24 (0.68 - 2.09) | 1.56 (1.28 - 1.88) |
| **PS-stratification / on-treatment TAR** | | | | | | | | |
| CCAE | 12,437 | 52,413 | 8.1 | 46.0 | 9 | 65 | 1.11 (0.51 - 2.11) | 1.41 (1.09 - 1.80) |
| ODOD | 8,466 | 30,499 | 4.8 | 25.7 | <5 | 50 | < 1.03 (0.34 - 2.41) | 1.94 (1.44 - 2.56) |
| MDCR | 936 | 9,340 | 0.6 | 9.7 | <5 | 47 | < 8.11 (2.63 - 18.9) | 4.83 (3.55 - 6.42) |
| Open Claims | 154,117 | 425,286 | 106.0 | 408.1 | 32 | 221 | 0.30 (0.21 - 0.43) | 0.54 (0.47 - 0.62) |
| OEHR | 9,044 | 38,070 | 2.6 | 19.1 | 5 | 23 | 1.94 (0.63 - 4.52) | 1.21 (0.76 - 1.81) |
| VA | 1,389 | 25,264 | 1.0 | 23.7 | 0 | <10 | | < 0.42 (0.20 - 0.78) |
| Meta-analysis | 184,064 | 546,268 | 121.5 | 498.8 | <51 | 359 | < 0.42 (0.31 - 0.55) | 0.72 (0.65 - 0.80) |
| Meta-analysis (w/o USOC) | 29,947 | 120,982 | 15.5 | 90.8 | <19 | 138 | < 1.22 (0.74 - 1.91) | 1.52 (1.28 - 1.80) |
| **PS-matching / on-treatment TAR without escalation** | | | | | | | | |
| CCAE | 10,525 | 52,283 | 6.3 | 37.1 | 8 | 54 | 1.27 (0.55 - 2.50) | 1.46 (1.09 - 1.90) |
| ODOD | 6,375 | 29,904 | 3.5 | 20.6 | <5 | 40 | < 1.42 (0.46 - 3.32) | 1.94 (1.39 - 2.64) |
| MDCR | 836 | 8,723 | 0.5 | 7.8 | <5 | 36 | < 9.66 (3.14 - 22.6) | 4.62 (3.24 - 6.40) |
| Open Claims | 114,514 | 423,044 | 76.2 | 314.5 | 25 | 186 | 0.33 (0.21 - 0.48) | 0.59 (0.51 - 0.68) |
| OEHR | 7,758 | 37,977 | 2.0 | 15.4 | <5 | 20 | < 2.53 (0.82 - 5.91) | 1.30 (0.79 - 2.00) |
| VA | 1,195 | 17,491 | 0.7 | 12.1 | 0 | <10 | | < 0.82 (0.40 - 1.51) |
| Meta-analysis | 131,414 | 505,231 | 86.0 | 372.2 | <38 | 280 | < 0.44 (0.31 - 0.61) | 0.75 (0.67 - 0.85) |
| Meta-analysis (w/o USOC) | 16,900 | 82,187 | 9.8 | 57.7 | <13 | 94 | < 1.33 (0.71 - 2.27) | 1.63 (1.32 - 1.99) |
| **PS-stratification / on-treatment TAR without escalation** | | | | | | | | |
| CCAE | 12,437 | 52,413 | 7.1 | 37.2 | 8 | 54 | 1.13 (0.49 - 2.23) | 1.45 (1.09 - 1.90) |
| ODOD | 8,466 | 30,499 | 4.3 | 21.0 | <5 | 42 | < 1.16 (0.38 - 2.72) | 2.00 (1.44 - 2.70) |
| MDCR | 936 | 9,340 | 0.5 | 8.3 | <5 | 40 | < 9.16 (2.97 - 21.4) | 4.81 (3.44 - 6.56) |
| Open Claims | 154,117 | 425,286 | 92.6 | 316.1 | 30 | 187 | 0.32 (0.22 - 0.46) | 0.59 (0.51 - 0.68) |
| OEHR | 9,044 | 38,070 | 2.2 | 15.5 | <5 | 20 | < 2.23 (0.73 - 5.21) | 1.29 (0.79 - 2.00) |
| VA | 1,389 | 25,264 | 0.8 | 19.2 | 0 | <10 | | < 0.52 (0.25 - 0.96) |
| Meta-analysis | 175,020 | 508,198 | 103.9 | 374.3 | <43 | 283 | < 0.41 (0.30 - 0.56) | 0.76 (0.67 - 0.85) |
| Meta-analysis (w/o USOC) | 20,903 | 82,912 | 11.4 | 58.2 | <13 | 96 | < 1.14 (0.61 - 1.96) | 1.65 (1.34 - 2.01) |

###### 4.4.25 Venous thromboembolism

**Supplementary Table 392:** Exposures, outcomes and risk estimates for Venous thromboembolism in GLP1RA (male) (T) vs DPP4I (male) (C) new-user cohorts under four different propensity score (PS) adjustment / time-at-risk (TAR) designs across data sources. We report total patient exposure time in 1,000 patient-year (KYr) units, uncalibrated and calibrated hazard ratios (HRs) and their 95% confidence intervals (CIs). Greyed out entries did not pass study diagnostics and are included only for completeness.

| | Patients | | Exposure (KYr) | | Outcomes | | Incidence (per KYr) | |
|---|---|---|---|---|---|---|---|---|
| | T | C | T | C | T | C | T | C |
| **PS-matching / on-treatment TAR** | | | | | | | | |
| CCAE | 10,331 | 51,708 | 7.1 | 45.3 | 34 | 203 | 4.78 (3.31 - 6.68) | 4.48 (3.88 - 5.14) |
| ODOD | 6,230 | 29,301 | 3.9 | 24.7 | 24 | 214 | 6.18 (3.96 - 9.19) | 8.68 (7.56 - 9.92) |
| MDCR | 814 | 8,491 | 0.6 | 8.9 | 8 | 110 | 14.1 (6.11 - 27.9) | 12.3 (10.1 - 14.8) |
| Open Claims | 111,380 | 413,513 | 86.1 | 396.4 | 317 | 1,916 | 3.68 (3.29 - 4.11) | 4.83 (4.62 - 5.06) |
| OEHR | 7,598 | 37,256 | 2.3 | 18.5 | 19 | 122 | 8.42 (5.07 - 13.2) | 6.58 (5.46 - 7.86) |
| VA | 1,162 | 17,157 | 0.9 | 14.5 | <10 | 64 | < 11.6 (5.57 - 21.4) | 4.40 (3.39 - 5.62) |
| Meta-analysis | 137,515 | 557,426 | 100.7 | 508.4 | <412 | 2,629 | < 4.09 (3.70 - 4.50) | 5.17 (4.98 - 5.37) |
| Meta-analysis (w/o USOC) | 26,135 | 143,913 | 14.7 | 112.0 | <95 | 713 | < 6.47 (5.24 - 7.91) | 6.37 (5.91 - 6.85) |
| **PS-stratification / on-treatment TAR** | | | | | | | | |
| CCAE | 12,188 | 51,836 | 7.9 | 45.4 | 42 | 204 | 5.30 (3.82 - 7.17) | 4.49 (3.89 - 5.15) |
| ODOD | 8,239 | 29,963 | 4.7 | 25.2 | 27 | 223 | 5.72 (3.77 - 8.33) | 8.84 (7.71 - 10.1) |
| MDCR | 910 | 9,176 | 0.6 | 9.6 | 9 | 128 | 15.2 (6.94 - 28.8) | 13.4 (11.2 - 15.9) |
| Open Claims | 149,623 | 415,788 | 103.1 | 398.5 | 370 | 1,959 | 3.59 (3.23 - 3.98) | 4.92 (4.70 - 5.14) |
| OEHR | 8,858 | 37,344 | 2.5 | 18.6 | 19 | 124 | 7.50 (4.52 - 11.7) | 6.67 (5.55 - 7.96) |
| VA | 1,344 | 24,743 | 0.9 | 23.1 | <10 | 103 | < 10.7 (5.13 - 19.7) | 4.46 (3.64 - 5.40) |
| Meta-analysis | 178,908 | 534,931 | 118.2 | 487.8 | 458 | 2,510 | 3.87 (3.53 - 4.25) | 5.15 (4.95 - 5.35) |
| Meta-analysis (w/o USOC) | 29,285 | 119,143 | 15.2 | 89.3 | 88 | 551 | 5.80 (4.65 - 7.15) | 6.17 (5.67 - 6.71) |
| **PS-matching / on-treatment TAR without escalation** | | | | | | | | |
| CCAE | 10,331 | 51,708 | 6.2 | 36.6 | 31 | 167 | 5.03 (3.42 - 7.14) | 4.56 (3.89 - 5.30) |
| ODOD | 6,230 | 29,301 | 3.4 | 20.1 | 21 | 174 | 6.11 (3.78 - 9.34) | 8.64 (7.40 - 10.0) |
| MDCR | 814 | 8,491 | 0.5 | 7.6 | <5 | 90 | < 9.99 (3.24 - 23.3) | 11.8 (9.53 - 14.6) |
| Open Claims | 111,380 | 413,513 | 74.2 | 307.4 | 275 | 1,487 | 3.71 (3.28 - 4.17) | 4.84 (4.59 - 5.09) |
| OEHR | 7,598 | 37,256 | 1.9 | 15.1 | 15 | 94 | 7.71 (4.31 - 12.7) | 6.24 (5.04 - 7.63) |
| VA | 1,162 | 17,157 | 0.7 | 11.9 | <10 | 50 | < 14.8 (7.10 - 27.2) | 4.21 (3.12 - 5.55) |
| Meta-analysis | 136,353 | 540,269 | 86.3 | 386.8 | <347 | 2,012 | < 4.02 (3.61 - 4.47) | 5.20 (4.98 - 5.43) |
| Meta-analysis (w/o USOC) | 24,973 | 126,756 | 12.0 | 79.4 | <72 | 525 | < 5.98 (4.68 - 7.53) | 6.61 (6.05 - 7.20) |
| **PS-stratification / on-treatment TAR without escalation** | | | | | | | | |
| CCAE | 12,188 | 51,836 | 6.9 | 36.7 | 39 | 168 | 5.65 (4.02 - 7.72) | 4.57 (3.91 - 5.32) |
| ODOD | 8,239 | 29,963 | 4.2 | 20.6 | 24 | 183 | 5.71 (3.66 - 8.50) | 8.88 (7.64 - 10.3) |
| MDCR | 910 | 9,176 | 0.5 | 8.2 | 5 | 105 | 9.49 (3.08 - 22.1) | 12.9 (10.5 - 15.6) |
| Open Claims | 149,623 | 415,788 | 90.0 | 309.1 | 324 | 1,522 | 3.60 (3.22 - 4.01) | 4.92 (4.68 - 5.18) |
| OEHR | 8,858 | 37,344 | 2.2 | 15.1 | 15 | 96 | 6.81 (3.81 - 11.2) | 6.36 (5.15 - 7.76) |
| VA | 1,344 | 24,743 | 0.7 | 18.8 | <10 | 74 | < 13.5 (6.48 - 24.8) | 3.94 (3.09 - 4.94) |
| Meta-analysis | 178,908 | 534,931 | 103.4 | 381.6 | 402 | 1,969 | 3.89 (3.52 - 4.29) | 5.16 (4.93 - 5.39) |
| Meta-analysis (w/o USOC) | 29,285 | 119,143 | 13.3 | 72.5 | 78 | 447 | 5.86 (4.63 - 7.31) | 6.17 (5.61 - 6.77) |

##### 4.4.26 Vomiting

**Supplementary Table 393:** Exposures, outcomes and risk estimates for Vomiting in GLP1RA (male) (T) vs DPP4I (male) (C) new-user cohorts under four different propensity score (PS) adjustment / time-at-risk (TAR) designs across data sources. We report total patient exposure time in 1,000 patient-year (KYr) units, uncalibrated and calibrated hazard ratios (HRs) and their 95% confidence intervals (CIs). Greyed out entries did not pass study diagnostics and are included only for completeness.

| | Patients | | Exposure (KYr) | | Outcomes | | Incidence (per KYr) | |
|---|---|---|---|---|---|---|---|---|
| | T | C | T | C | T | C | T | C |
| **PS-matching / on-treatment TAR** | | | | | | | | |
| CCAE | 9,911 | 49,940 | 6.8 | 43.5 | 153 | 713 | 22.5 (19.1 - 26.4) | 16.4 (15.2 - 17.6) |
| ODOD | 5,885 | 28,044 | 3.6 | 23.4 | 97 | 533 | 26.8 (21.7 - 32.6) | 22.8 (20.9 - 24.8) |
| MDCR | 808 | 8,326 | 0.6 | 8.7 | 11 | 175 | 20.0 (9.97 - 35.7) | 20.1 (17.3 - 23.3) |
| Open Claims | 105,894 | 396,335 | 81.9 | 379.5 | 1,196 | 4,527 | 14.6 (13.8 - 15.5) | 11.9 (11.6 - 12.3) |
| OEHR | 7,405 | 36,590 | 2.2 | 18.1 | 79 | 290 | 36.2 (28.7 - 45.2) | 16.0 (14.3 - 18.0) |
| VA | 1,157 | 16,781 | 0.8 | 14.3 | 14 | 94 | 16.5 (9.01 - 27.7) | 6.58 (5.32 - 8.05) |
| Meta-analysis | 131,060 | 536,016 | 95.9 | 487.4 | 1,550 | 6,332 | 16.2 (15.4 - 17.0) | 13.0 (12.7 - 13.3) |
| Meta-analysis (w/o USOC) | 25,166 | 139,681 | 14.0 | 108.0 | 354 | 1,805 | 25.3 (22.7 - 28.1) | 16.7 (16.0 - 17.5) |
| **PS-stratification / on-treatment TAR** | | | | | | | | |
| CCAE | 11,678 | 50,059 | 7.6 | 43.6 | 176 | 717 | 23.2 (19.9 - 26.9) | 16.4 (15.2 - 17.7) |
| ODOD | 7,839 | 28,613 | 4.4 | 23.8 | 125 | 555 | 28.3 (23.6 - 33.7) | 23.3 (21.4 - 25.3) |
| MDCR | 905 | 8,966 | 0.6 | 9.3 | 11 | 202 | 18.9 (9.44 - 33.8) | 21.7 (18.8 - 24.9) |
| Open Claims | 141,956 | 398,725 | 97.9 | 381.6 | 1,492 | 4,622 | 15.2 (14.5 - 16.0) | 12.1 (11.8 - 12.5) |
| OEHR | 8,595 | 36,696 | 2.4 | 18.1 | 90 | 295 | 36.8 (29.6 - 45.3) | 16.3 (14.5 - 18.2) |
| VA | 1,348 | 24,357 | 0.9 | 22.9 | 14 | 156 | 15.0 (8.23 - 25.2) | 6.82 (5.79 - 7.98) |
| Meta-analysis | 170,068 | 514,093 | 112.3 | 467.2 | 1,883 | 6,189 | 16.8 (16.0 - 17.5) | 13.2 (12.9 - 13.6) |
| Meta-analysis (w/o USOC) | 28,112 | 115,368 | 14.4 | 85.6 | 391 | 1,567 | 27.1 (24.5 - 29.9) | 18.3 (17.4 - 19.2) |
| **PS-matching / on-treatment TAR without escalation** | | | | | | | | |
| CCAE | 9,911 | 49,940 | 5.9 | 35.3 | 138 | 593 | 23.4 (19.7 - 27.7) | 16.8 (15.5 - 18.2) |
| ODOD | 5,885 | 28,044 | 3.2 | 19.2 | 91 | 447 | 28.3 (22.8 - 34.8) | 23.3 (21.2 - 25.6) |
| MDCR | 808 | 8,326 | 0.5 | 7.4 | 8 | 147 | 16.4 (7.08 - 32.3) | 19.8 (16.8 - 23.3) |
| Open Claims | 105,894 | 396,335 | 70.8 | 295.0 | 1,016 | 3,420 | 14.4 (13.5 - 15.3) | 11.6 (11.2 - 12.0) |
| OEHR | 7,405 | 36,590 | 1.9 | 14.8 | 63 | 195 | 33.3 (25.6 - 42.6) | 13.2 (11.4 - 15.2) |
| VA | 1,157 | 16,781 | 0.7 | 11.7 | 13 | 74 | 19.5 (10.4 - 33.4) | 6.34 (4.98 - 7.96) |
| Meta-analysis | 131,060 | 536,016 | 82.9 | 383.3 | 1,329 | 4,876 | 16.0 (15.2 - 16.9) | 12.7 (12.4 - 13.1) |
| Meta-analysis (w/o USOC) | 25,166 | 139,681 | 12.1 | 88.3 | 313 | 1,456 | 25.8 (23.0 - 28.8) | 16.5 (15.7 - 17.4) |
| **PS-stratification / on-treatment TAR without escalation** | | | | | | | | |
| CCAE | 11,678 | 50,059 | 6.6 | 35.4 | 159 | 593 | 24.1 (20.5 - 28.1) | 16.8 (15.4 - 18.2) |
| ODOD | 7,839 | 28,613 | 3.9 | 19.6 | 114 | 466 | 28.9 (23.9 - 34.8) | 23.8 (21.7 - 26.1) |
| MDCR | 905 | 8,966 | 0.5 | 7.9 | 8 | 172 | 15.5 (6.68 - 30.5) | 21.7 (18.5 - 25.1) |
| Open Claims | 141,956 | 398,725 | 85.7 | 296.7 | 1,293 | 3,494 | 15.1 (14.3 - 15.9) | 11.8 (11.4 - 12.2) |
| OEHR | 8,595 | 36,696 | 2.1 | 14.8 | 74 | 198 | 34.7 (27.3 - 43.6) | 13.4 (11.6 - 15.4) |
| VA | 1,348 | 24,357 | 0.7 | 18.6 | 13 | 128 | 17.7 (9.42 - 30.3) | 6.89 (5.74 - 8.19) |
| Meta-analysis | 170,068 | 514,093 | 98.3 | 366.4 | 1,640 | 4,751 | 16.7 (15.9 - 17.5) | 13.0 (12.6 - 13.3) |
| Meta-analysis (w/o USOC) | 28,112 | 115,368 | 12.7 | 69.7 | 347 | 1,257 | 27.4 (24.6 - 30.4) | 18.0 (17.0 - 19.1) |

#### 4.5 GLP1RA (male) vs SU (male)

##### 4.5.1 3-point MACE

**Supplementary Table 394:** Exposures, outcomes and risk estimates for 3-point MACE in GLP1RA (male) (T) vs SU (male) (C) new-user cohorts under four different propensity score (PS) adjustment / time-at-risk (TAR) designs across data sources. We report total patient exposure time in 1,000 patient-year (KYr) units, uncalibrated and calibrated hazard ratios (HRs) and their 95% confidence intervals (CIs). Greyed out entries did not pass study diagnostics and are included only for completeness.

| | Patients | | Exposure (KYr) | | Outcomes | | Incidence (per KYr) | |
|---|---|---|---|---|---|---|---|---|
| | T | C | T | C | T | C | T | C |
| **PS-matching / on-treatment TAR** | | | | | | | | |
| CCAE | 11,015 | 82,939 | 7.4 | 71.1 | 55 | 722 | 7.45 (5.62 - 9.70) | 10.2 (9.43 - 10.9) |
| ODOD | 7,287 | 59,676 | 4.3 | 53.9 | 39 | 1,228 | 9.10 (6.47 - 12.4) | 22.8 (21.5 - 24.1) |
| MDCR | 871 | 14,448 | 0.6 | 15.5 | 11 | 458 | 18.4 (9.21 - 33.0) | 29.5 (26.8 - 32.3) |
| Open Claims | 132,622 | 817,979 | 94.6 | 881.7 | 604 | 11,673 | 6.38 (5.88 - 6.91) | 13.2 (13.0 - 13.5) |
| OEHR | 8,180 | 84,320 | 2.4 | 46.0 | 25 | 600 | 10.6 (6.83 - 15.6) | 13.1 (12.0 - 14.1) |
| VA | 1,240 | 31,321 | 0.9 | 23.6 | <10 | 152 | < 11.1 (5.32 - 20.4) | 6.45 (5.46 - 7.56) |
| Meta-analysis | 159,975 | 1,059,362 | 109.3 | 1068.1 | 734 | 14,681 | 6.72 (6.24 - 7.22) | 13.7 (13.5 - 14.0) |
| Meta-analysis (w/o USOC) | 27,353 | 241,383 | 14.6 | 186.4 | 130 | 3,008 | 8.89 (7.42 - 10.6) | 16.1 (15.6 - 16.7) |
| **PS-stratification / on-treatment TAR** | | | | | | | | |
| CCAE | 12,495 | 87,190 | 8.1 | 74.5 | 62 | 764 | 7.63 (5.85 - 9.78) | 10.3 (9.54 - 11.0) |
| ODOD | 8,175 | 66,607 | 4.7 | 61.8 | 42 | 1,568 | 8.96 (6.46 - 12.1) | 25.4 (24.1 - 26.7) |
| MDCR | 945 | 18,402 | 0.6 | 19.5 | 11 | 654 | 17.6 (8.80 - 31.5) | 33.5 (31.0 - 36.2) |
| Open Claims | 147,591 | 843,396 | 101.8 | 911.1 | 647 | 12,750 | 6.36 (5.88 - 6.87) | 14.0 (13.8 - 14.2) |
| OEHR | 8,896 | 93,416 | 2.5 | 51.3 | 26 | 750 | 10.3 (6.71 - 15.0) | 14.6 (13.6 - 15.7) |
| VA | 1,335 | 230,206 | 0.9 | 244.4 | <10 | 2,056 | < 10.7 (5.15 - 19.7) | 8.41 (8.05 - 8.78) |
| Meta-analysis | 177,157 | 1,090,609 | 117.1 | 1098.7 | 777 | 15,832 | 6.63 (6.18 - 7.12) | 14.4 (14.2 - 14.6) |
| Meta-analysis (w/o USOC) | 29,566 | 247,213 | 15.3 | 187.6 | 130 | 3,082 | 8.47 (7.08 - 10.1) | 16.4 (15.9 - 17.0) |
| **PS-matching / on-treatment TAR without escalation** | | | | | | | | |
| CCAE | 11,015 | 82,939 | 6.4 | 61.7 | 49 | 629 | 7.65 (5.66 - 10.1) | 10.2 (9.42 - 11.0) |
| ODOD | 7,287 | 59,676 | 3.8 | 47.9 | 32 | 1,095 | 8.40 (5.75 - 11.9) | 22.8 (21.5 - 24.2) |
| MDCR | 871 | 14,448 | 0.5 | 14.1 | 7 | 406 | 13.3 (5.33 - 27.3) | 28.9 (26.1 - 31.8) |
| Open Claims | 132,622 | 817,979 | 82.3 | 747.0 | 497 | 9,969 | 6.04 (5.52 - 6.59) | 13.3 (13.1 - 13.6) |
| OEHR | 8,180 | 84,320 | 2.0 | 40.1 | 21 | 442 | 10.3 (6.35 - 15.7) | 11.0 (10.0 - 12.1) |
| VA | 1,240 | 31,321 | 0.7 | 20.7 | <10 | 136 | < 14.1 (6.78 - 26.0) | 6.57 (5.51 - 7.78) |
| Meta-analysis | 159,975 | 1,059,362 | 95.1 | 910.8 | 606 | 12,541 | 6.37 (5.87 - 6.90) | 13.8 (13.5 - 14.0) |
| Meta-analysis (w/o USOC) | 27,353 | 241,383 | 12.8 | 163.8 | 109 | 2,572 | 8.52 (7.00 - 10.3) | 15.7 (15.1 - 16.3) |
| **PS-stratification / on-treatment TAR without escalation** | | | | | | | | |
| CCAE | 12,495 | 87,190 | 7.1 | 64.8 | 55 | 670 | 7.75 (5.84 - 10.1) | 10.3 (9.58 - 11.2) |
| ODOD | 8,175 | 66,607 | 4.2 | 55.2 | 35 | 1,402 | 8.40 (5.85 - 11.7) | 25.4 (24.1 - 26.8) |
| MDCR | 945 | 18,402 | 0.6 | 17.8 | 7 | 584 | 12.6 (5.08 - 26.0) | 32.9 (30.3 - 35.7) |
| Open Claims | 147,591 | 843,396 | 89.0 | 772.2 | 538 | 10,929 | 6.04 (5.54 - 6.58) | 14.2 (13.9 - 14.4) |
| OEHR | 8,896 | 93,416 | 2.2 | 44.7 | 22 | 548 | 9.98 (6.26 - 15.1) | 12.3 (11.2 - 13.3) |
| VA | 1,335 | 230,206 | 0.7 | 217.9 | <10 | 1,754 | < 13.6 (6.54 - 25.1) | 8.05 (7.68 - 8.44) |
| Meta-analysis | 177,157 | 1,090,609 | 102.5 | 936.9 | 650 | 13,549 | 6.34 (5.86 - 6.85) | 14.5 (14.2 - 14.7) |
| Meta-analysis (w/o USOC) | 29,566 | 247,213 | 13.5 | 164.7 | 112 | 2,620 | 8.32 (6.85 - 10.0) | 15.9 (15.3 - 16.5) |

##### 4.5.2 4-point MACE

**Supplementary Table 395:** Exposures, outcomes and risk estimates for 4-point MACE in GLP1RA (male) (T) vs SU (male) (C) new-user cohorts under four different propensity score (PS) adjustment / time-at-risk (TAR) designs across data sources. We report total patient exposure time in 1,000 patient-year (KYr) units, uncalibrated and calibrated hazard ratios (HRs) and their 95% confidence intervals (CIs). Greyed out entries did not pass study diagnostics and are included only for completeness.

| | Patients | | Exposure (KYr) | | Outcomes | | Incidence (per KYr) | |
|---|---|---|---|---|---|---|---|---|
| | T | C | T | C | T | C | T | C |
| **PS-matching / on-treatment TAR** | | | | | | | | |
| CCAE | 10,829 | 81,745 | 7.3 | 69.8 | 85 | 1,071 | 11.7 (9.36 - 14.5) | 15.3 (14.4 - 16.3) |
| ODOD | 7,033 | 57,505 | 4.1 | 51.3 | 71 | 1,953 | 17.2 (13.4 - 21.6) | 38.0 (36.4 - 39.8) |
| MDCR | 823 | 13,580 | 0.6 | 14.4 | 21 | 742 | 37.4 (23.2 - 57.2) | 51.4 (47.8 - 55.3) |
| Open Claims | 128,962 | 790,564 | 92.1 | 847.8 | 921 | 18,264 | 10.00 (9.36 - 10.7) | 21.5 (21.2 - 21.9) |
| OEHR | 8,057 | 82,802 | 2.3 | 44.5 | 31 | 1,099 | 13.3 (9.04 - 18.9) | 24.7 (23.2 - 26.2) |
| VA | 1,184 | 30,622 | 0.9 | 22.9 | <10 | 364 | < 11.5 (5.54 - 21.2) | 15.9 (14.3 - 17.7) |
| Meta-analysis | 155,704 | 1,026,196 | 106.4 | 1027.9 | 1,129 | 23,129 | 10.6 (10.00 - 11.2) | 22.5 (22.2 - 22.8) |
| Meta-analysis (w/o USOC) | 26,742 | 235,632 | 14.3 | 180.1 | 208 | 4,865 | 14.6 (12.6 - 16.7) | 27.0 (26.3 - 27.8) |
| **PS-stratification / on-treatment TAR** | | | | | | | | |
| CCAE | 12,270 | 85,777 | 8.0 | 73.1 | 98 | 1,132 | 12.3 (9.97 - 15.0) | 15.5 (14.6 - 16.4) |
| ODOD | 7,889 | 63,755 | 4.5 | 58.4 | 77 | 2,460 | 17.0 (13.4 - 21.3) | 42.1 (40.4 - 43.8) |
| MDCR | 895 | 17,003 | 0.6 | 17.9 | 23 | 1,035 | 39.1 (24.8 - 58.7) | 57.9 (54.4 - 61.5) |
| Open Claims | 143,665 | 813,841 | 99.2 | 875.2 | 977 | 19,937 | 9.85 (9.24 - 10.5) | 22.8 (22.5 - 23.1) |
| OEHR | 8,765 | 91,617 | 2.5 | 49.6 | 33 | 1,350 | 13.3 (9.13 - 18.6) | 27.2 (25.8 - 28.7) |
| VA | 1,282 | 223,743 | 0.9 | 235.5 | <10 | 4,077 | < 11.1 (5.33 - 20.4) | 17.3 (16.8 - 17.9) |
| Meta-analysis | 172,589 | 1,054,990 | 114.2 | 1056.3 | 1,185 | 24,879 | 10.4 (9.80 - 11.0) | 23.6 (23.3 - 23.8) |
| Meta-analysis (w/o USOC) | 28,924 | 241,149 | 15.0 | 181.1 | 208 | 4,942 | 13.9 (12.1 - 15.9) | 27.3 (26.5 - 28.1) |
| **PS-matching / on-treatment TAR without escalation** | | | | | | | | |
| CCAE | 10,829 | 81,745 | 6.3 | 60.6 | 77 | 925 | 12.2 (9.66 - 15.3) | 15.3 (14.3 - 16.3) |
| ODOD | 7,033 | 57,505 | 3.7 | 45.8 | 61 | 1,725 | 16.6 (12.7 - 21.3) | 37.7 (35.9 - 39.5) |
| MDCR | 823 | 13,580 | 0.5 | 13.1 | 16 | 663 | 32.2 (18.4 - 52.3) | 50.7 (46.9 - 54.7) |
| Open Claims | 128,962 | 790,564 | 80.2 | 719.1 | 778 | 15,559 | 9.70 (9.03 - 10.4) | 21.6 (21.3 - 22.0) |
| OEHR | 8,057 | 82,802 | 2.0 | 39.1 | 26 | 848 | 12.9 (8.43 - 18.9) | 21.7 (20.3 - 23.2) |
| VA | 1,184 | 30,622 | 0.7 | 20.1 | <10 | 298 | < 14.8 (7.09 - 27.2) | 14.8 (13.2 - 16.6) |
| Meta-analysis | 155,704 | 1,026,196 | 92.7 | 877.6 | 958 | 19,720 | 10.3 (9.69 - 11.0) | 22.5 (22.2 - 22.8) |
| Meta-analysis (w/o USOC) | 26,742 | 235,632 | 12.5 | 158.6 | 180 | 4,161 | 14.4 (12.4 - 16.7) | 26.2 (25.5 - 27.1) |
| **PS-stratification / on-treatment TAR without escalation** | | | | | | | | |
| CCAE | 12,270 | 85,777 | 7.0 | 63.6 | 87 | 979 | 12.5 (10.0 - 15.4) | 15.4 (14.4 - 16.4) |
| ODOD | 7,889 | 63,755 | 4.0 | 52.2 | 67 | 2,184 | 16.7 (12.9 - 21.2) | 41.8 (40.1 - 43.6) |
| MDCR | 895 | 17,003 | 0.5 | 16.3 | 18 | 937 | 34.5 (20.4 - 54.5) | 57.4 (53.8 - 61.2) |
| Open Claims | 143,665 | 813,841 | 86.8 | 742.6 | 830 | 17,034 | 9.57 (8.93 - 10.2) | 22.9 (22.6 - 23.3) |
| OEHR | 8,765 | 91,617 | 2.2 | 43.5 | 28 | 1,045 | 12.9 (8.59 - 18.7) | 24.0 (22.6 - 25.5) |
| VA | 1,282 | 223,743 | 0.7 | 210.3 | <10 | 3,467 | < 14.2 (6.79 - 26.1) | 16.5 (15.9 - 17.0) |
| Meta-analysis | 172,589 | 1,054,990 | 99.9 | 902.0 | 1,012 | 21,242 | 10.1 (9.51 - 10.8) | 23.6 (23.2 - 23.9) |
| Meta-analysis (w/o USOC) | 28,924 | 241,149 | 13.2 | 159.4 | 182 | 4,208 | 13.8 (11.9 - 16.0) | 26.4 (25.6 - 27.2) |

##### 4.5.3 Acute myocardial infarction

**Supplementary Table 396:** Exposures, outcomes and risk estimates for Acute myocardial infarction in GLP1RA (male) (T) vs SU (male) (C) new-user cohorts under four different propensity score (PS) adjustment / time-at-risk (TAR) designs across data sources. We report total patient exposure time in 1,000 patient-year (KYr) units, uncalibrated and calibrated hazard ratios (HRs) and their 95% confidence intervals (CIs). Greyed out entries did not pass study diagnostics and are included only for completeness.

| | Patients | | Exposure (KYr) | | Outcomes | | Incidence (per KYr) | |
|---|---|---|---|---|---|---|---|---|
| | T | C | T | C | T | C | T | C |
| **PS-matching / on-treatment TAR** | | | | | | | | |
| CCAE | 11,160 | 83,945 | 7.5 | 72.1 | 34 | 434 | 4.53 (3.14 - 6.34) | 6.02 (5.47 - 6.62) |
| ODOD | 7,417 | 61,088 | 4.4 | 55.5 | 17 | 658 | 3.90 (2.27 - 6.24) | 11.9 (11.0 - 12.8) |
| MDCR | 898 | 14,949 | 0.6 | 16.3 | 7 | 222 | 11.3 (4.55 - 23.3) | 13.7 (11.9 - 15.6) |
| Open Claims | 135,489 | 841,364 | 96.5 | 908.9 | 319 | 6,108 | 3.30 (2.95 - 3.69) | 6.72 (6.55 - 6.89) |
| OEHR | 8,240 | 85,150 | 2.4 | 46.6 | 9 | 344 | 3.76 (1.72 - 7.14) | 7.37 (6.62 - 8.20) |
| VA | 1,264 | 31,658 | 0.9 | 23.9 | <10 | 99 | < 11.0 (5.26 - 20.2) | 4.15 (3.37 - 5.05) |
| Meta-analysis | 163,204 | 1,086,496 | 111.4 | 1099.4 | 386 | 7,766 | 3.46 (3.13 - 3.83) | 7.06 (6.91 - 7.22) |
| Meta-analysis (w/o USOC) | 27,715 | 245,132 | 14.9 | 190.5 | 67 | 1,658 | 4.50 (3.49 - 5.72) | 8.70 (8.29 - 9.13) |
| **PS-stratification / on-treatment TAR** | | | | | | | | |
| CCAE | 12,650 | 88,414 | 8.3 | 75.7 | 40 | 456 | 4.85 (3.46 - 6.60) | 6.03 (5.49 - 6.61) |
| ODOD | 8,318 | 68,419 | 4.8 | 63.9 | 19 | 825 | 3.99 (2.40 - 6.23) | 12.9 (12.0 - 13.8) |
| MDCR | 975 | 19,284 | 0.6 | 20.6 | 7 | 323 | 10.8 (4.34 - 22.3) | 15.7 (14.0 - 17.5) |
| Open Claims | 150,626 | 868,394 | 103.8 | 940.1 | 346 | 6,663 | 3.33 (2.99 - 3.70) | 7.09 (6.92 - 7.26) |
| OEHR | 8,966 | 94,391 | 2.6 | 52.1 | 10 | 434 | 3.91 (1.87 - 7.19) | 8.32 (7.56 - 9.14) |
| VA | 1,361 | 233,198 | 0.9 | 248.0 | <10 | 1,249 | < 10.6 (5.09 - 19.5) | 5.04 (4.76 - 5.32) |
| Meta-analysis | 180,560 | 1,119,618 | 119.4 | 1131.9 | 415 | 8,378 | 3.48 (3.15 - 3.83) | 7.40 (7.24 - 7.56) |
| Meta-analysis (w/o USOC) | 29,934 | 251,224 | 15.6 | 191.8 | 69 | 1,715 | 4.43 (3.45 - 5.61) | 8.94 (8.53 - 9.38) |
| **PS-matching / on-treatment TAR without escalation** | | | | | | | | |
| CCAE | 11,160 | 83,945 | 6.5 | 62.5 | 32 | 372 | 4.92 (3.37 - 6.95) | 5.95 (5.36 - 6.59) |
| ODOD | 7,417 | 61,088 | 3.9 | 49.4 | 14 | 591 | 3.61 (1.97 - 6.06) | 12.0 (11.0 - 13.0) |
| MDCR | 898 | 14,949 | 0.5 | 14.7 | <5 | 196 | < 9.12 (2.96 - 21.3) | 13.3 (11.5 - 15.3) |
| Open Claims | 135,489 | 841,364 | 84.0 | 769.9 | 263 | 5,189 | 3.13 (2.76 - 3.53) | 6.74 (6.56 - 6.93) |
| OEHR | 8,240 | 85,150 | 2.1 | 40.7 | 8 | 246 | 3.87 (1.67 - 7.63) | 6.05 (5.32 - 6.86) |
| VA | 1,264 | 31,658 | 0.7 | 20.9 | <10 | 88 | < 13.9 (6.68 - 25.6) | 4.20 (3.37 - 5.18) |
| Meta-analysis | 163,204 | 1,086,496 | 97.0 | 937.2 | <322 | 6,594 | < 3.32 (2.97 - 3.70) | 7.04 (6.87 - 7.21) |
| Meta-analysis (w/o USOC) | 27,715 | 245,132 | 13.0 | 167.3 | <59 | 1,405 | < 4.54 (3.46 - 5.86) | 8.40 (7.97 - 8.85) |
| **PS-stratification / on-treatment TAR without escalation** | | | | | | | | |
| CCAE | 12,650 | 88,414 | 7.2 | 65.8 | 37 | 393 | 5.14 (3.62 - 7.08) | 5.98 (5.40 - 6.60) |
| ODOD | 8,318 | 68,419 | 4.2 | 57.1 | 16 | 737 | 3.78 (2.16 - 6.14) | 12.9 (12.0 - 13.9) |
| MDCR | 975 | 19,284 | 0.6 | 18.7 | <5 | 283 | < 8.68 (2.82 - 20.3) | 15.1 (13.4 - 17.0) |
| Open Claims | 150,626 | 868,394 | 90.8 | 796.7 | 289 | 5,680 | 3.18 (2.83 - 3.57) | 7.13 (6.94 - 7.32) |
| OEHR | 8,966 | 94,391 | 2.2 | 45.4 | 9 | 303 | 4.05 (1.85 - 7.69) | 6.68 (5.95 - 7.47) |
| VA | 1,361 | 233,198 | 0.7 | 221.0 | <10 | 1,055 | < 13.4 (6.44 - 24.7) | 4.77 (4.49 - 5.07) |
| Meta-analysis | 180,560 | 1,119,618 | 104.4 | 964.9 | 351 | 7,113 | 3.36 (3.02 - 3.73) | 7.37 (7.20 - 7.54) |
| Meta-analysis (w/o USOC) | 29,934 | 251,224 | 13.7 | 168.2 | 62 | 1,433 | 4.54 (3.48 - 5.82) | 8.52 (8.08 - 8.97) |

##### 4.5.4 Acute renal failure

**Supplementary Table 397:** Exposures, outcomes and risk estimates for Acute renal failure in GLP1RA (male) (T) vs SU (male) (C) new-user cohorts under four different propensity score (PS) adjustment / time-at-risk (TAR) designs across data sources. We report total patient exposure time in 1,000 patient-year (KYr) units, uncalibrated and calibrated hazard ratios (HRs) and their 95% confidence intervals (CIs). Greyed out entries did not pass study diagnostics and are included only for completeness.

| | Patients | | Exposure (KYr) | | Outcomes | | Incidence (per KYr) | |
|---|---|---|---|---|---|---|---|---|
| | T | C | T | C | T | C | T | C |
| **PS-matching / on-treatment TAR** | | | | | | | | |
| CCAE | 11,229 | 84,870 | 7.5 | 73.0 | 53 | 715 | 7.03 (5.26 - 9.19) | 9.79 (9.09 - 10.5) |
| ODOD | 7,315 | 60,157 | 4.3 | 54.2 | 83 | 1,773 | 19.4 (15.5 - 24.1) | 32.7 (31.2 - 34.3) |
| MDCR | 892 | 14,768 | 0.6 | 15.9 | 23 | 533 | 37.4 (23.7 - 56.2) | 33.5 (30.7 - 36.5) |
| Open Claims | 135,101 | 837,375 | 96.2 | 901.1 | 860 | 15,464 | 8.94 (8.35 - 9.56) | 17.2 (16.9 - 17.4) |
| OEHR | 8,242 | 85,033 | 2.4 | 46.2 | 34 | 891 | 14.2 (9.83 - 19.8) | 19.3 (18.0 - 20.6) |
| VA | 1,249 | 31,250 | 0.9 | 23.4 | <10 | 361 | < 11.1 (5.30 - 20.3) | 15.4 (13.9 - 17.1) |
| Meta-analysis | 162,779 | 1,082,203 | 111.0 | 1090.4 | 1,053 | 19,376 | 9.48 (8.92 - 10.1) | 17.8 (17.5 - 18.0) |
| Meta-analysis (w/o USOC) | 27,678 | 244,828 | 14.8 | 189.3 | 193 | 3,912 | 13.0 (11.2 - 15.0) | 20.7 (20.0 - 21.3) |
| **PS-stratification / on-treatment TAR** | | | | | | | | |
| CCAE | 12,718 | 88,812 | 8.3 | 76.2 | 61 | 777 | 7.36 (5.63 - 9.46) | 10.2 (9.49 - 10.9) |
| ODOD | 8,198 | 66,571 | 4.7 | 61.5 | 91 | 2,227 | 19.5 (15.7 - 23.9) | 36.2 (34.7 - 37.7) |
| MDCR | 963 | 18,886 | 0.6 | 20.0 | 23 | 791 | 35.9 (22.8 - 53.9) | 39.5 (36.8 - 42.3) |
| Open Claims | 150,166 | 862,791 | 103.4 | 930.8 | 898 | 16,968 | 8.68 (8.12 - 9.27) | 18.2 (18.0 - 18.5) |
| OEHR | 8,969 | 94,051 | 2.6 | 51.5 | 34 | 1,133 | 13.3 (9.20 - 18.6) | 22.0 (20.8 - 23.3) |
| VA | 1,343 | 232,074 | 0.9 | 244.9 | <10 | 3,743 | < 10.7 (5.14 - 19.7) | 15.3 (14.8 - 15.8) |
| Meta-analysis | 180,051 | 1,112,225 | 118.9 | 1119.9 | 1,084 | 21,105 | 9.11 (8.58 - 9.67) | 18.8 (18.6 - 19.1) |
| Meta-analysis (w/o USOC) | 29,885 | 249,434 | 15.5 | 189.2 | 186 | 4,137 | 12.0 (10.3 - 13.8) | 21.9 (21.2 - 22.5) |
| **PS-matching / on-treatment TAR without escalation** | | | | | | | | |
| CCAE | 11,229 | 84,870 | 6.5 | 63.3 | 47 | 609 | 7.18 (5.28 - 9.55) | 9.62 (8.87 - 10.4) |
| ODOD | 7,315 | 60,157 | 3.8 | 48.3 | 74 | 1,587 | 19.5 (15.3 - 24.5) | 32.9 (31.3 - 34.5) |
| MDCR | 892 | 14,768 | 0.5 | 14.4 | 19 | 467 | 35.0 (21.1 - 54.6) | 32.4 (29.5 - 35.4) |
| Open Claims | 135,101 | 837,375 | 83.7 | 764.2 | 717 | 12,926 | 8.57 (7.95 - 9.22) | 16.9 (16.6 - 17.2) |
| OEHR | 8,242 | 85,033 | 2.1 | 40.4 | 29 | 655 | 14.0 (9.37 - 20.1) | 16.2 (15.0 - 17.5) |
| VA | 1,249 | 31,250 | 0.7 | 20.5 | <10 | 293 | < 14.0 (6.72 - 25.8) | 14.3 (12.7 - 16.0) |
| Meta-analysis | 162,779 | 1,082,203 | 96.6 | 930.6 | 886 | 16,244 | 9.17 (8.57 - 9.79) | 17.5 (17.2 - 17.7) |
| Meta-analysis (w/o USOC) | 27,678 | 244,828 | 13.0 | 166.4 | 169 | 3,318 | 13.0 (11.2 - 15.2) | 19.9 (19.3 - 20.6) |
| **PS-stratification / on-treatment TAR without escalation** | | | | | | | | |
| CCAE | 12,718 | 88,812 | 7.2 | 66.2 | 53 | 663 | 7.32 (5.48 - 9.58) | 10.0 (9.27 - 10.8) |
| ODOD | 8,198 | 66,571 | 4.2 | 55.0 | 80 | 1,986 | 19.3 (15.3 - 24.0) | 36.1 (34.6 - 37.8) |
| MDCR | 963 | 18,886 | 0.6 | 18.3 | 19 | 704 | 33.5 (20.2 - 52.3) | 38.5 (35.7 - 41.5) |
| Open Claims | 150,166 | 862,791 | 90.4 | 789.9 | 753 | 14,219 | 8.33 (7.74 - 8.94) | 18.0 (17.7 - 18.3) |
| OEHR | 8,969 | 94,051 | 2.2 | 45.0 | 29 | 834 | 13.0 (8.72 - 18.7) | 18.5 (17.3 - 19.8) |
| VA | 1,343 | 232,074 | 0.7 | 218.6 | <10 | 3,030 | < 13.6 (6.50 - 24.9) | 13.9 (13.4 - 14.4) |
| Meta-analysis | 180,051 | 1,112,225 | 104.1 | 956.0 | 915 | 17,702 | 8.79 (8.23 - 9.38) | 18.5 (18.2 - 18.8) |
| Meta-analysis (w/o USOC) | 29,885 | 249,434 | 13.6 | 166.2 | 162 | 3,483 | 11.9 (10.1 - 13.9) | 21.0 (20.3 - 21.7) |

##### 4.5.5 Glycemic control

**Supplementary Table 398:** Exposures, outcomes and risk estimates for Glycemic control in GLP1RA (male) (T) vs SU (male) (C) new-user cohorts under four different propensity score (PS) adjustment / time-at-risk (TAR) designs across data sources. We report total patient exposure time in 1,000 patient-year (KYr) units, uncalibrated and calibrated hazard ratios (HRs) and their 95% confidence intervals (CIs). Greyed out entries did not pass study diagnostics and are included only for completeness.

| | Patients | | Exposure (KYr) | | Outcomes | | Incidence (per KYr) | |
|---|---|---|---|---|---|---|---|---|
| | T | C | T | C | T | C | T | C |
| **PS-matching / on-treatment TAR** | | | | | | | | |
| CCAE | 11,397 | 85,482 | 7.7 | 73.6 | 6 | 301 | 0.78 (0.29 - 1.71) | 4.09 (3.64 - 4.58) |
| ODOD | 5,201 | 45,690 | 2.8 | 36.9 | 311 | 3,538 | 112 (99.7 - 125) | 95.9 (92.7 - 99.1) |
| MDCR | 931 | 15,433 | 0.6 | 16.9 | <5 | 77 | < 7.77 (2.52 - 18.1) | 4.56 (3.60 - 5.70) |
| Open Claims | 140,133 | 873,951 | 99.8 | 948.1 | 0 | 0 | | |
| OEHR | 8,369 | 86,443 | 2.4 | 47.7 | 0 | 0 | | |
| VA | 1,245 | 30,995 | 0.9 | 23.1 | 0 | 45 | | 1.94 (1.42 - 2.60) |
| Meta-analysis | 17,529 | 146,605 | 11.1 | 127.3 | <322 | 3,916 | < 29.1 (26.0 - 32.4) | 30.8 (29.8 - 31.7) |
| Meta-analysis (w/o USOC) | 17,529 | 146,605 | 11.1 | 127.3 | <322 | 3,916 | < 29.1 (26.0 - 32.4) | 30.8 (29.8 - 31.7) |
| **PS-stratification / on-treatment TAR** | | | | | | | | |
| CCAE | 12,911 | 89,467 | 8.4 | 76.6 | 6 | 512 | 0.71 (0.26 - 1.55) | 6.68 (6.12 - 7.29) |
| ODOD | 5,649 | 52,045 | 3.0 | 43.2 | 329 | 4,236 | 111 (98.9 - 123) | 98.1 (95.2 - 101) |
| MDCR | 1,007 | 20,047 | 0.7 | 21.4 | <5 | 178 | < 7.42 (2.41 - 17.3) | 8.32 (7.14 - 9.63) |
| Open Claims | 155,612 | 903,312 | 107.2 | 981.9 | 0 | 0 | | |
| OEHR | 9,105 | 95,902 | 2.6 | 53.4 | 0 | 0 | | |
| VA | 1,342 | 227,080 | 0.9 | 239.3 | 0 | 536 | | 2.24 (2.05 - 2.44) |
| Meta-analysis | 18,560 | 141,512 | 11.4 | 119.8 | 335 | 4,748 | 29.4 (26.3 - 32.7) | 39.6 (38.5 - 40.8) |
| Meta-analysis (w/o USOC) | 18,560 | 141,512 | 11.4 | 119.8 | 335 | 4,748 | 29.4 (26.3 - 32.7) | 39.6 (38.5 - 40.8) |
| **PS-matching / on-treatment TAR without escalation** | | | | | | | | |
| CCAE | 11,397 | 85,482 | 6.6 | 63.6 | 5 | 270 | 0.75 (0.24 - 1.76) | 4.24 (3.75 - 4.78) |
| ODOD | 5,201 | 45,690 | 2.5 | 32.9 | 280 | 3,207 | 114 (101 - 128) | 97.5 (94.1 - 101) |
| MDCR | 931 | 15,433 | 0.6 | 15.2 | <5 | 70 | < 8.80 (2.86 - 20.5) | 4.59 (3.58 - 5.80) |
| Open Claims | 140,133 | 873,951 | 86.7 | 802.3 | 0 | 0 | | |
| OEHR | 8,369 | 86,443 | 2.1 | 41.5 | 0 | 0 | | |
| VA | 1,245 | 30,995 | 0.7 | 20.3 | 0 | 43 | | 2.12 (1.54 - 2.86) |
| Meta-analysis | 16,598 | 131,172 | 9.1 | 96.5 | 285 | 3,477 | 31.3 (27.8 - 35.2) | 36.0 (34.8 - 37.2) |
| Meta-analysis (w/o USOC) | 16,598 | 131,172 | 9.1 | 96.5 | 285 | 3,477 | 31.3 (27.8 - 35.2) | 36.0 (34.8 - 37.2) |
| **PS-stratification / on-treatment TAR without escalation** | | | | | | | | |
| CCAE | 12,911 | 89,467 | 7.3 | 66.4 | 5 | 467 | 0.68 (0.22 - 1.59) | 7.04 (6.41 - 7.70) |
| ODOD | 5,649 | 52,045 | 2.6 | 38.6 | 297 | 3,853 | 113 (101 - 127) | 99.8 (96.7 - 103) |
| MDCR | 1,007 | 20,047 | 0.6 | 19.4 | <5 | 168 | < 8.38 (2.72 - 19.6) | 8.65 (7.39 - 10.1) |
| Open Claims | 155,612 | 903,312 | 93.6 | 831.4 | 0 | 0 | | |
| OEHR | 9,105 | 95,902 | 2.3 | 46.3 | 0 | 0 | | |
| VA | 1,342 | 227,080 | 0.7 | 213.0 | 0 | 500 | | 2.35 (2.15 - 2.56) |
| Meta-analysis | 18,560 | 141,512 | 10.0 | 105.0 | 302 | 4,320 | 30.3 (27.0 - 33.9) | 41.2 (39.9 - 42.4) |
| Meta-analysis (w/o USOC) | 18,560 | 141,512 | 10.0 | 105.0 | 302 | 4,320 | 30.3 (27.0 - 33.9) | 41.2 (39.9 - 42.4) |

##### 4.5.6 Hospitalization with heart failure

**Supplementary Table 399:** Exposures, outcomes and risk estimates for Hospitalization with heart failure in GLP1RA (male) (T) vs SU (male) (C) new-user cohorts under four different propensity score (PS) adjustment / time-at-risk (TAR) designs across data sources. We report total patient exposure time in 1,000 patient-year (KYr) units, uncalibrated and calibrated hazard ratios (HRs) and their 95% confidence intervals (CIs). Greyed out entries did not pass study diagnostics and are included only for completeness.

| | Patients | | Exposure (KYr) | | Outcomes | | Incidence (per KYr) | |
|---|---|---|---|---|---|---|---|---|
| | T | C | T | C | T | C | T | C |
| **PS-matching / on-treatment TAR** | | | | | | | | |
| CCAE | 11,207 | 84,502 | 7.5 | 72.7 | 45 | 537 | 5.98 (4.36 - 8.00) | 7.39 (6.78 - 8.04) |
| ODOD | 7,267 | 59,738 | 4.3 | 53.9 | 51 | 1,333 | 12.0 (8.91 - 15.7) | 24.8 (23.4 - 26.1) |
| MDCR | 867 | 14,380 | 0.6 | 15.5 | 17 | 494 | 28.4 (16.5 - 45.4) | 31.9 (29.1 - 34.8) |
| Open Claims | 135,009 | 834,445 | 96.3 | 899.5 | 514 | 11,503 | 5.34 (4.89 - 5.82) | 12.8 (12.6 - 13.0) |
| OEHR | 8,208 | 84,581 | 2.4 | 45.9 | 16 | 734 | 6.72 (3.84 - 10.9) | 16.0 (14.8 - 17.2) |
| VA | 1,240 | 31,306 | 0.9 | 23.3 | <10 | 289 | < 11.1 (5.34 - 20.5) | 12.4 (11.0 - 13.9) |
| Meta-analysis | 162,558 | 1,077,646 | 111.1 | 1087.4 | 643 | 14,601 | 5.79 (5.35 - 6.25) | 13.4 (13.2 - 13.6) |
| Meta-analysis (w/o USOC) | 27,549 | 243,201 | 14.8 | 187.9 | 129 | 3,098 | 8.73 (7.29 - 10.4) | 16.5 (15.9 - 17.1) |
| **PS-stratification / on-treatment TAR** | | | | | | | | |
| CCAE | 12,687 | 88,628 | 8.3 | 76.0 | 53 | 573 | 6.41 (4.80 - 8.38) | 7.54 (6.93 - 8.18) |
| ODOD | 8,142 | 66,524 | 4.7 | 61.6 | 55 | 1,697 | 11.8 (8.90 - 15.4) | 27.5 (26.2 - 28.9) |
| MDCR | 939 | 18,277 | 0.6 | 19.5 | 18 | 738 | 28.7 (17.0 - 45.4) | 37.9 (35.2 - 40.8) |
| Open Claims | 150,190 | 860,350 | 103.6 | 930.0 | 538 | 12,800 | 5.19 (4.76 - 5.65) | 13.8 (13.5 - 14.0) |
| OEHR | 8,933 | 93,635 | 2.5 | 51.2 | 17 | 920 | 6.68 (3.89 - 10.7) | 18.0 (16.8 - 19.2) |
| VA | 1,340 | 231,003 | 0.9 | 244.2 | <10 | 2,973 | < 10.7 (5.14 - 19.7) | 12.2 (11.7 - 12.6) |
| Meta-analysis | 179,952 | 1,109,137 | 119.1 | 1118.8 | 663 | 15,990 | 5.57 (5.15 - 6.01) | 14.3 (14.1 - 14.5) |
| Meta-analysis (w/o USOC) | 29,762 | 248,787 | 15.5 | 188.8 | 125 | 3,190 | 8.08 (6.72 - 9.62) | 16.9 (16.3 - 17.5) |
| **PS-matching / on-treatment TAR without escalation** | | | | | | | | |
| CCAE | 11,207 | 84,502 | 6.5 | 63.0 | 42 | 457 | 6.43 (4.64 - 8.70) | 7.25 (6.60 - 7.95) |
| ODOD | 7,267 | 59,738 | 3.8 | 47.9 | 47 | 1,173 | 12.4 (9.13 - 16.5) | 24.5 (23.1 - 25.9) |
| MDCR | 867 | 14,380 | 0.5 | 14.1 | 13 | 443 | 24.5 (13.1 - 42.0) | 31.5 (28.6 - 34.6) |
| Open Claims | 135,009 | 834,445 | 83.7 | 762.3 | 441 | 9,760 | 5.27 (4.79 - 5.78) | 12.8 (12.5 - 13.1) |
| OEHR | 8,208 | 84,581 | 2.1 | 40.2 | 15 | 565 | 7.30 (4.09 - 12.0) | 14.1 (12.9 - 15.3) |
| VA | 1,240 | 31,306 | 0.7 | 20.5 | <10 | 238 | < 14.2 (6.82 - 26.1) | 11.6 (10.2 - 13.2) |
| Meta-analysis | 162,558 | 1,077,646 | 96.6 | 927.6 | 558 | 12,398 | 5.77 (5.30 - 6.27) | 13.4 (13.1 - 13.6) |
| Meta-analysis (w/o USOC) | 27,549 | 243,201 | 12.9 | 165.2 | 117 | 2,638 | 9.07 (7.50 - 10.9) | 16.0 (15.4 - 16.6) |
| **PS-stratification / on-treatment TAR without escalation** | | | | | | | | |
| CCAE | 12,687 | 88,628 | 7.2 | 66.0 | 48 | 487 | 6.65 (4.90 - 8.82) | 7.37 (6.73 - 8.06) |
| ODOD | 8,142 | 66,524 | 4.1 | 55.0 | 51 | 1,497 | 12.3 (9.18 - 16.2) | 27.2 (25.8 - 28.6) |
| MDCR | 939 | 18,277 | 0.6 | 17.7 | 14 | 673 | 25.2 (13.8 - 42.3) | 37.9 (35.1 - 40.9) |
| Open Claims | 150,190 | 860,350 | 90.5 | 788.6 | 463 | 10,898 | 5.11 (4.66 - 5.60) | 13.8 (13.6 - 14.1) |
| OEHR | 8,933 | 93,635 | 2.2 | 44.8 | 16 | 704 | 7.24 (4.14 - 11.8) | 15.7 (14.6 - 16.9) |
| VA | 1,340 | 231,003 | 0.7 | 217.9 | <10 | 2,550 | < 13.6 (6.54 - 25.1) | 11.7 (11.3 - 12.2) |
| Meta-analysis | 179,952 | 1,109,137 | 104.1 | 954.4 | 578 | 13,586 | 5.55 (5.11 - 6.02) | 14.2 (14.0 - 14.5) |
| Meta-analysis (w/o USOC) | 29,762 | 248,787 | 13.6 | 165.8 | 115 | 2,688 | 8.48 (7.00 - 10.2) | 16.2 (15.6 - 16.8) |

##### 4.5.7 Measured renal dysfunction

**Supplementary Table 400:** Exposures, outcomes and risk estimates for Measured renal dysfunction in GLP1RA (male) (T) vs SU (male) (C) new-user cohorts under four different propensity score (PS) adjustment / time-at-risk (TAR) designs across data sources. We report total patient exposure time in 1,000 patient-year (KYr) units, uncalibrated and calibrated hazard ratios (HRs) and their 95% confidence intervals (CIs). Greyed out entries did not pass study diagnostics and are included only for completeness.

| | Patients | | Exposure (KYr) | | Outcomes | | Incidence (per KYr) | |
|---|---|---|---|---|---|---|---|---|
| | T | C | T | C | T | C | T | C |
| **PS-matching / on-treatment TAR** | | | | | | | | |
| CCAE | 11,450 | 86,141 | 7.7 | 74.3 | 0 | 30 | | 0.40 (0.27 - 0.58) |
| ODOD | 7,591 | 62,908 | 4.5 | 57.5 | 6 | 109 | 1.34 (0.49 - 2.93) | 1.90 (1.56 - 2.29) |
| MDCR | 932 | 15,503 | 0.6 | 17.0 | 0 | 5 | | 0.29 (0.10 - 0.69) |
| Open Claims | 140,133 | 873,951 | 99.8 | 948.1 | 0 | 0 | | |
| OEHR | 8,369 | 86,443 | 2.4 | 47.7 | 0 | 0 | | |
| VA | 1,293 | 31,902 | 0.9 | 23.9 | <10 | 143 | < 10.7 (5.12 - 19.6) | 5.98 (5.04 - 7.04) |
| Meta-analysis | 7,591 | 62,908 | 4.5 | 57.5 | 6 | 109 | 1.34 (0.49 - 2.93) | 1.90 (1.56 - 2.29) |
| Meta-analysis (w/o USOC) | 7,591 | 62,908 | 4.5 | 57.5 | 6 | 109 | 1.34 (0.49 - 2.93) | 1.90 (1.56 - 2.29) |
| **PS-stratification / on-treatment TAR** | | | | | | | | |
| CCAE | 12,979 | 90,487 | 8.5 | 77.8 | 0 | 31 | | 0.40 (0.27 - 0.57) |
| ODOD | 8,508 | 70,635 | 4.9 | 66.4 | 6 | 146 | 1.23 (0.45 - 2.68) | 2.20 (1.86 - 2.59) |
| MDCR | 1,008 | 20,367 | 0.7 | 21.9 | 0 | 8 | | 0.37 (0.16 - 0.72) |
| Open Claims | 155,612 | 903,312 | 107.2 | 981.9 | 0 | 0 | | |
| OEHR | 9,105 | 95,902 | 2.6 | 53.4 | 0 | 0 | | |
| VA | 1,389 | 235,040 | 1.0 | 249.7 | <10 | 1,309 | < 10.4 (4.96 - 19.0) | 5.24 (4.96 - 5.53) |
| Meta-analysis | 8,508 | 70,635 | 4.9 | 66.4 | 6 | 146 | 1.23 (0.45 - 2.68) | 2.20 (1.86 - 2.59) |
| Meta-analysis (w/o USOC) | 8,508 | 70,635 | 4.9 | 66.4 | 6 | 146 | 1.23 (0.45 - 2.68) | 2.20 (1.86 - 2.59) |
| **PS-matching / on-treatment TAR without escalation** | | | | | | | | |
| CCAE | 11,450 | 86,141 | 6.7 | 64.3 | 0 | 26 | | 0.40 (0.26 - 0.59) |
| ODOD | 7,591 | 62,908 | 4.0 | 51.1 | <5 | 94 | < 1.26 (0.41 - 2.94) | 1.84 (1.49 - 2.25) |
| MDCR | 932 | 15,503 | 0.6 | 15.3 | 0 | <5 | | < 0.33 (0.11 - 0.76) |
| Open Claims | 140,133 | 873,951 | 86.7 | 802.3 | 0 | 0 | | |
| OEHR | 8,369 | 86,443 | 2.1 | 41.5 | 0 | 0 | | |
| VA | 1,293 | 31,902 | 0.7 | 21.0 | <10 | 119 | < 13.6 (6.50 - 24.9) | 5.67 (4.70 - 6.79) |
| Meta-analysis | 7,591 | 62,908 | 4.0 | 51.1 | <5 | 94 | < 1.26 (0.41 - 2.94) | 1.84 (1.49 - 2.25) |
| Meta-analysis (w/o USOC) | 7,591 | 62,908 | 4.0 | 51.1 | <5 | 94 | < 1.26 (0.41 - 2.94) | 1.84 (1.49 - 2.25) |
| **PS-stratification / on-treatment TAR without escalation** | | | | | | | | |
| CCAE | 12,979 | 90,487 | 7.4 | 67.5 | 0 | 27 | | 0.40 (0.26 - 0.58) |
| ODOD | 8,508 | 70,635 | 4.3 | 59.1 | <5 | 126 | < 1.15 (0.37 - 2.69) | 2.13 (1.78 - 2.54) |
| MDCR | 1,008 | 20,367 | 0.6 | 19.9 | 0 | 7 | | 0.35 (0.14 - 0.72) |
| Open Claims | 155,612 | 903,312 | 93.6 | 831.4 | 0 | 0 | | |
| OEHR | 9,105 | 95,902 | 2.3 | 46.3 | 0 | 0 | | |
| VA | 1,389 | 235,040 | 0.8 | 222.5 | <10 | 1,073 | < 13.1 (6.28 - 24.1) | 4.82 (4.54 - 5.12) |
| Meta-analysis | 8,508 | 70,635 | 4.3 | 59.1 | <5 | 126 | < 1.15 (0.37 - 2.69) | 2.13 (1.78 - 2.54) |
| Meta-analysis (w/o USOC) | 8,508 | 70,635 | 4.3 | 59.1 | <5 | 126 | < 1.15 (0.37 - 2.69) | 2.13 (1.78 - 2.54) |

##### 4.5.8 Stroke

**Supplementary Table 401:** Exposures, outcomes and risk estimates for Stroke in GLP1RA (male) (T) vs SU (male) (C) new-user cohorts under four different propensity score (PS) adjustment / time-at-risk (TAR) designs across data sources. We report total patient exposure time in 1,000 patient-year (KYr) units, uncalibrated and calibrated hazard ratios (HRs) and their 95% confidence intervals (CIs). Greyed out entries did not pass study diagnostics and are included only for completeness.

| | Patients | | Exposure (KYr) | | Outcomes | | Incidence (per KYr) | |
|---|---|---|---|---|---|---|---|---|
| | T | C | T | C | T | C | T | C |
| **PS-matching / on-treatment TAR** | | | | | | | | |
| CCAE | 11,318 | 85,192 | 7.6 | 73.5 | 23 | 259 | 3.03 (1.92 - 4.55) | 3.53 (3.11 - 3.98) |
| ODOD | 7,468 | 61,465 | 4.4 | 55.9 | 23 | 595 | 5.23 (3.32 - 7.85) | 10.6 (9.81 - 11.5) |
| MDCR | 905 | 14,960 | 0.6 | 16.2 | 5 | 251 | 8.09 (2.63 - 18.9) | 15.5 (13.6 - 17.5) |
| Open Claims | 137,113 | 848,723 | 97.7 | 919.0 | 279 | 5,620 | 2.85 (2.53 - 3.21) | 6.12 (5.96 - 6.28) |
| OEHR | 8,308 | 85,605 | 2.4 | 47.0 | 12 | 230 | 4.97 (2.57 - 8.68) | 4.89 (4.28 - 5.57) |
| VA | 1,283 | 31,903 | 0.9 | 24.0 | <10 | 61 | < 10.7 (5.13 - 19.7) | 2.54 (1.95 - 3.27) |
| Meta-analysis | 165,112 | 1,095,945 | 112.7 | 1111.6 | 342 | 6,955 | 3.03 (2.72 - 3.37) | 6.26 (6.11 - 6.41) |
| Meta-analysis (w/o USOC) | 27,999 | 247,222 | 15.0 | 192.6 | 63 | 1,335 | 4.20 (3.23 - 5.37) | 6.93 (6.56 - 7.31) |
| **PS-stratification / on-treatment TAR** | | | | | | | | |
| CCAE | 12,840 | 89,313 | 8.3 | 76.8 | 24 | 279 | 2.88 (1.84 - 4.28) | 3.63 (3.22 - 4.09) |
| ODOD | 8,371 | 68,804 | 4.8 | 64.3 | 24 | 770 | 5.00 (3.21 - 7.44) | 12.0 (11.1 - 12.8) |
| MDCR | 977 | 19,375 | 0.6 | 20.7 | 5 | 362 | 7.75 (2.52 - 18.1) | 17.5 (15.7 - 19.4) |
| Open Claims | 152,434 | 876,391 | 105.1 | 951.0 | 292 | 6,193 | 2.78 (2.47 - 3.12) | 6.51 (6.35 - 6.68) |
| OEHR | 9,031 | 94,894 | 2.6 | 52.5 | 12 | 296 | 4.66 (2.41 - 8.13) | 5.64 (5.01 - 6.32) |
| VA | 1,383 | 236,388 | 1.0 | 252.0 | <10 | 880 | < 10.3 (4.96 - 19.0) | 3.49 (3.27 - 3.73) |
| Meta-analysis | 182,676 | 1,129,402 | 120.8 | 1144.6 | 352 | 7,538 | 2.91 (2.62 - 3.24) | 6.59 (6.44 - 6.74) |
| Meta-analysis (w/o USOC) | 30,242 | 253,011 | 15.7 | 193.6 | 60 | 1,345 | 3.82 (2.91 - 4.91) | 6.95 (6.58 - 7.33) |
| **PS-matching / on-treatment TAR without escalation** | | | | | | | | |
| CCAE | 11,318 | 85,192 | 6.6 | 63.6 | 20 | 234 | 3.04 (1.86 - 4.70) | 3.68 (3.22 - 4.18) |
| ODOD | 7,468 | 61,465 | 3.9 | 49.7 | 20 | 528 | 5.12 (3.13 - 7.91) | 10.6 (9.74 - 11.6) |
| MDCR | 905 | 14,960 | 0.5 | 14.7 | <5 | 227 | < 9.17 (2.98 - 21.4) | 15.5 (13.5 - 17.6) |
| Open Claims | 137,113 | 848,723 | 84.9 | 777.9 | 232 | 4,844 | 2.73 (2.39 - 3.11) | 6.23 (6.05 - 6.41) |
| OEHR | 8,308 | 85,605 | 2.1 | 40.9 | 10 | 174 | 4.79 (2.30 - 8.81) | 4.25 (3.64 - 4.93) |
| VA | 1,283 | 31,903 | 0.7 | 21.0 | <10 | 51 | < 13.6 (6.51 - 25.0) | 2.43 (1.81 - 3.19) |
| Meta-analysis | 165,112 | 1,095,945 | 98.0 | 946.8 | <287 | 6,007 | < 2.93 (2.60 - 3.29) | 6.34 (6.19 - 6.51) |
| Meta-analysis (w/o USOC) | 27,999 | 247,222 | 13.1 | 168.9 | <55 | 1,163 | < 4.19 (3.16 - 5.46) | 6.89 (6.50 - 7.29) |
| **PS-stratification / on-treatment TAR without escalation** | | | | | | | | |
| CCAE | 12,840 | 89,313 | 7.3 | 66.6 | 21 | 253 | 2.88 (1.78 - 4.40) | 3.80 (3.34 - 4.30) |
| ODOD | 8,371 | 68,804 | 4.3 | 57.3 | 21 | 687 | 4.93 (3.05 - 7.53) | 12.0 (11.1 - 12.9) |
| MDCR | 977 | 19,375 | 0.6 | 18.8 | <5 | 331 | < 8.76 (2.84 - 20.4) | 17.6 (15.8 - 19.6) |
| Open Claims | 152,434 | 876,391 | 91.8 | 805.3 | 245 | 5,349 | 2.67 (2.35 - 3.03) | 6.64 (6.47 - 6.82) |
| OEHR | 9,031 | 94,894 | 2.2 | 45.6 | 10 | 225 | 4.46 (2.14 - 8.20) | 4.93 (4.31 - 5.62) |
| VA | 1,383 | 236,388 | 0.8 | 224.4 | <10 | 757 | < 13.1 (6.28 - 24.1) | 3.37 (3.14 - 3.62) |
| Meta-analysis | 182,676 | 1,129,402 | 105.6 | 974.9 | 297 | 6,514 | 2.81 (2.50 - 3.15) | 6.68 (6.52 - 6.85) |
| Meta-analysis (w/o USOC) | 30,242 | 253,011 | 13.8 | 169.6 | 52 | 1,165 | 3.77 (2.82 - 4.94) | 6.87 (6.48 - 7.28) |

##### 4.5.9 Sudden cardiac death

**Supplementary Table 402:** Exposures, outcomes and risk estimates for Sudden cardiac death in GLP1RA (male) (T) vs SU (male) (C) new-user cohorts under four different propensity score (PS) adjustment / time-at-risk (TAR) designs across data sources. We report total patient exposure time in 1,000 patient-year (KYr) units, uncalibrated and calibrated hazard ratios (HRs) and their 95% confidence intervals (CIs). Greyed out entries did not pass study diagnostics and are included only for completeness.

| | Patients | | Exposure (KYr) | | Outcomes | | Incidence (per KYr) | |
|---|---|---|---|---|---|---|---|---|
| | T | C | T | C | T | C | T | C |
| **PS-matching / on-treatment TAR** | | | | | | | | |
| CCAE | 11,460 | 86,179 | 7.7 | 74.4 | <5 | 109 | < 0.65 (0.21 - 1.52) | 1.47 (1.20 - 1.77) |
| ODOD | 7,587 | 62,755 | 4.5 | 57.4 | 10 | 177 | 2.24 (1.08 - 4.12) | 3.08 (2.65 - 3.57) |
| MDCR | 927 | 15,472 | 0.6 | 17.0 | <5 | 48 | < 7.78 (2.53 - 18.1) | 2.83 (2.09 - 3.75) |
| Open Claims | 139,827 | 871,935 | 99.5 | 945.5 | 80 | 1,647 | 0.80 (0.64 - 1.00) | 1.74 (1.66 - 1.83) |
| OEHR | 8,363 | 86,388 | 2.4 | 47.6 | 5 | 84 | 2.05 (0.67 - 4.79) | 1.76 (1.41 - 2.18) |
| VA | 1,309 | 32,260 | 0.9 | 24.3 | 0 | <10 | | < 0.41 (0.20 - 0.76) |
| Meta-analysis | 167,237 | 1,107,257 | 114.1 | 1124.9 | <100 | 2,017 | < 0.88 (0.71 - 1.07) | 1.79 (1.72 - 1.87) |
| Meta-analysis (w/o USOC) | 27,410 | 235,322 | 14.6 | 179.4 | <20 | 370 | < 1.37 (0.84 - 2.12) | 2.06 (1.86 - 2.28) |
| **PS-stratification / on-treatment TAR** | | | | | | | | |
| CCAE | 12,982 | 90,509 | 8.5 | 77.8 | <5 | 115 | < 0.59 (0.19 - 1.38) | 1.48 (1.22 - 1.77) |
| ODOD | 8,502 | 70,624 | 4.9 | 66.4 | 10 | 253 | 2.05 (0.98 - 3.78) | 3.81 (3.35 - 4.31) |
| MDCR | 1,004 | 20,293 | 0.7 | 21.8 | <5 | 77 | < 7.43 (2.41 - 17.3) | 3.53 (2.79 - 4.41) |
| Open Claims | 155,278 | 901,112 | 106.9 | 979.1 | 87 | 1,869 | 0.81 (0.65 - 1.00) | 1.91 (1.82 - 2.00) |
| OEHR | 9,101 | 95,825 | 2.6 | 53.3 | 5 | 104 | 1.92 (0.62 - 4.49) | 1.95 (1.59 - 2.36) |
| VA | 1,406 | 239,362 | 1.0 | 255.6 | 0 | 134 | | 0.52 (0.44 - 0.62) |
| Meta-analysis | 185,863 | 1,158,070 | 122.9 | 1176.7 | <107 | 2,341 | < 0.87 (0.71 - 1.05) | 1.99 (1.91 - 2.07) |
| Meta-analysis (w/o USOC) | 30,585 | 256,958 | 15.9 | 197.6 | <20 | 472 | < 1.25 (0.77 - 1.94) | 2.39 (2.18 - 2.61) |
| **PS-matching / on-treatment TAR without escalation** | | | | | | | | |
| CCAE | 11,460 | 86,179 | 6.7 | 64.4 | <5 | 94 | < 0.75 (0.24 - 1.75) | 1.46 (1.18 - 1.79) |
| ODOD | 7,587 | 62,755 | 4.0 | 51.0 | 9 | 157 | 2.27 (1.04 - 4.32) | 3.08 (2.62 - 3.60) |
| MDCR | 927 | 15,472 | 0.6 | 15.3 | <5 | 39 | < 8.81 (2.86 - 20.6) | 2.55 (1.81 - 3.48) |
| Open Claims | 139,827 | 871,935 | 86.5 | 800.2 | 66 | 1,383 | 0.76 (0.59 - 0.97) | 1.73 (1.64 - 1.82) |
| OEHR | 8,363 | 86,388 | 2.1 | 41.4 | <5 | 59 | < 2.38 (0.77 - 5.56) | 1.43 (1.08 - 1.84) |
| VA | 1,309 | 32,260 | 0.7 | 21.3 | 0 | <10 | | < 0.47 (0.23 - 0.86) |
| Meta-analysis | 167,237 | 1,107,257 | 99.2 | 957.0 | <85 | 1,693 | < 0.86 (0.68 - 1.06) | 1.77 (1.69 - 1.86) |
| Meta-analysis (w/o USOC) | 27,410 | 235,322 | 12.7 | 156.8 | <19 | 310 | < 1.49 (0.90 - 2.33) | 1.98 (1.76 - 2.21) |
| **PS-stratification / on-treatment TAR without escalation** | | | | | | | | |
| CCAE | 12,982 | 90,509 | 7.4 | 67.5 | <5 | 100 | < 0.68 (0.22 - 1.58) | 1.48 (1.21 - 1.80) |
| ODOD | 8,502 | 70,624 | 4.3 | 59.2 | 9 | 223 | 2.08 (0.95 - 3.95) | 3.77 (3.29 - 4.30) |
| MDCR | 1,004 | 20,293 | 0.6 | 19.8 | <5 | 66 | < 8.39 (2.73 - 19.6) | 3.33 (2.57 - 4.23) |
| Open Claims | 155,278 | 901,112 | 93.4 | 829.1 | 72 | 1,585 | 0.77 (0.60 - 0.97) | 1.91 (1.82 - 2.01) |
| OEHR | 9,101 | 95,825 | 2.3 | 46.3 | <5 | 73 | < 2.21 (0.72 - 5.17) | 1.58 (1.24 - 1.98) |
| VA | 1,406 | 239,362 | 0.8 | 227.5 | 0 | 102 | | 0.45 (0.37 - 0.54) |
| Meta-analysis | 185,863 | 1,158,070 | 107.4 | 1002.1 | <91 | 1,981 | < 0.85 (0.68 - 1.04) | 1.98 (1.89 - 2.07) |
| Meta-analysis (w/o USOC) | 30,585 | 256,958 | 14.0 | 173.0 | <19 | 396 | < 1.36 (0.82 - 2.12) | 2.29 (2.07 - 2.53) |

##### 4.5.10 Abnormal weight gain

**Supplementary Table 403:** Exposures, outcomes and risk estimates for Abnormal weight gain in GLP1RA (male) (T) vs SU (male) (C) new-user cohorts under four different propensity score (PS) adjustment / time-at-risk (TAR) designs across data sources. We report total patient exposure time in 1,000 patient-year (KYr) units, uncalibrated and calibrated hazard ratios (HRs) and their 95% confidence intervals (CIs). Greyed out entries did not pass study diagnostics and are included only for completeness.

| | Patients | | Exposure (KYr) | | Outcomes | | Incidence (per KYr) | |
|---|---|---|---|---|---|---|---|---|
| | T | C | T | C | T | C | T | C |
| **PS-matching / on-treatment TAR** | | | | | | | | |
| CCAE | 10,954 | 84,463 | 7.4 | 72.7 | 35 | 288 | 4.75 (3.31 - 6.61) | 3.96 (3.52 - 4.45) |
| ODOD | 7,230 | 61,302 | 4.2 | 55.8 | 36 | 305 | 8.48 (5.94 - 11.7) | 5.47 (4.87 - 6.12) |
| MDCR | 913 | 15,392 | 0.6 | 16.8 | 5 | 48 | 8.02 (2.60 - 18.7) | 2.85 (2.10 - 3.78) |
| Open Claims | 133,544 | 853,326 | 94.9 | 924.2 | 203 | 1,746 | 2.14 (1.86 - 2.46) | 1.89 (1.80 - 1.98) |
| OEHR | 7,961 | 84,542 | 2.3 | 46.3 | 18 | 259 | 7.85 (4.66 - 12.4) | 5.59 (4.93 - 6.31) |
| VA | 1,269 | 31,600 | 0.9 | 23.7 | <10 | 57 | < 11.0 (5.29 - 20.3) | 2.40 (1.82 - 3.11) |
| Meta-analysis | 159,689 | 1,083,633 | 108.8 | 1099.0 | 292 | 2,598 | 2.68 (2.39 - 3.01) | 2.36 (2.27 - 2.46) |
| Meta-analysis (w/o USOC) | 26,145 | 230,307 | 13.9 | 174.8 | 89 | 852 | 6.40 (5.14 - 7.88) | 4.87 (4.55 - 5.21) |
| **PS-stratification / on-treatment TAR** | | | | | | | | |
| CCAE | 12,272 | 88,789 | 8.0 | 76.2 | 42 | 301 | 5.23 (3.77 - 7.07) | 3.95 (3.52 - 4.42) |
| ODOD | 8,011 | 69,036 | 4.6 | 64.6 | 40 | 374 | 8.70 (6.22 - 11.9) | 5.79 (5.22 - 6.40) |
| MDCR | 983 | 20,211 | 0.7 | 21.7 | 5 | 73 | 7.68 (2.50 - 17.9) | 3.37 (2.64 - 4.24) |
| Open Claims | 146,922 | 882,934 | 101.3 | 958.2 | 235 | 1,841 | 2.32 (2.03 - 2.64) | 1.92 (1.83 - 2.01) |
| OEHR | 8,589 | 93,991 | 2.4 | 52.0 | 23 | 286 | 9.43 (5.98 - 14.1) | 5.50 (4.88 - 6.18) |
| VA | 1,354 | 235,922 | 0.9 | 251.3 | <10 | 415 | < 10.7 (5.14 - 19.7) | 1.65 (1.50 - 1.82) |
| Meta-analysis | 175,794 | 1,134,750 | 116.3 | 1151.0 | 340 | 2,802 | 2.92 (2.62 - 3.25) | 2.43 (2.35 - 2.53) |
| Meta-analysis (w/o USOC) | 28,872 | 251,816 | 15.1 | 192.8 | 105 | 961 | 6.97 (5.70 - 8.44) | 4.99 (4.68 - 5.31) |
| **PS-matching / on-treatment TAR without escalation** | | | | | | | | |
| CCAE | 10,954 | 84,463 | 6.4 | 63.0 | 28 | 243 | 4.38 (2.91 - 6.33) | 3.86 (3.39 - 4.37) |
| ODOD | 7,230 | 61,302 | 3.8 | 49.7 | 29 | 265 | 7.70 (5.16 - 11.1) | 5.33 (4.71 - 6.02) |
| MDCR | 913 | 15,392 | 0.6 | 15.2 | <5 | 44 | < 9.05 (2.94 - 21.1) | 2.89 (2.10 - 3.88) |
| Open Claims | 133,544 | 853,326 | 82.5 | 783.1 | 175 | 1,441 | 2.12 (1.82 - 2.46) | 1.84 (1.75 - 1.94) |
| OEHR | 7,961 | 84,542 | 2.0 | 40.4 | 15 | 217 | 7.58 (4.24 - 12.5) | 5.37 (4.68 - 6.14) |
| VA | 1,269 | 31,600 | 0.7 | 20.8 | <10 | 42 | < 13.9 (6.68 - 25.6) | 2.02 (1.45 - 2.73) |
| Meta-analysis | 159,689 | 1,083,633 | 94.6 | 936.2 | 247 | 2,166 | 2.61 (2.30 - 2.96) | 2.31 (2.22 - 2.41) |
| Meta-analysis (w/o USOC) | 26,145 | 230,307 | 12.1 | 153.1 | 72 | 725 | 5.93 (4.64 - 7.47) | 4.74 (4.40 - 5.09) |
| **PS-stratification / on-treatment TAR without escalation** | | | | | | | | |
| CCAE | 12,272 | 88,789 | 7.0 | 66.1 | 34 | 256 | 4.85 (3.36 - 6.78) | 3.87 (3.41 - 4.38) |
| ODOD | 8,011 | 69,036 | 4.1 | 57.7 | 33 | 326 | 8.09 (5.57 - 11.4) | 5.65 (5.05 - 6.30) |
| MDCR | 983 | 20,211 | 0.6 | 19.7 | <5 | 67 | < 8.65 (2.81 - 20.2) | 3.40 (2.64 - 4.32) |
| Open Claims | 146,922 | 882,934 | 88.4 | 812.4 | 206 | 1,524 | 2.33 (2.02 - 2.67) | 1.88 (1.78 - 1.97) |
| OEHR | 8,589 | 93,991 | 2.1 | 45.2 | 19 | 237 | 8.96 (5.40 - 14.0) | 5.24 (4.60 - 5.95) |
| VA | 1,354 | 235,922 | 0.7 | 223.8 | <10 | 339 | < 13.5 (6.47 - 24.8) | 1.51 (1.36 - 1.68) |
| Meta-analysis | 175,794 | 1,134,750 | 101.7 | 981.4 | 292 | 2,343 | 2.87 (2.55 - 3.22) | 2.39 (2.29 - 2.49) |
| Meta-analysis (w/o USOC) | 28,872 | 251,816 | 13.2 | 169.1 | 86 | 819 | 6.51 (5.21 - 8.04) | 4.84 (4.52 - 5.19) |

##### 4.5.11 Abnormal weight loss

**Supplementary Table 404:** Exposures, outcomes and risk estimates for Abnormal weight loss in GLP1RA (male) (T) vs SU (male) (C) new-user cohorts under four different propensity score (PS) adjustment / time-at-risk (TAR) designs across data sources. We report total patient exposure time in 1,000 patient-year (KYr) units, uncalibrated and calibrated hazard ratios (HRs) and their 95% confidence intervals (CIs). Greyed out entries did not pass study diagnostics and are included only for completeness.

| | Patients | | Exposure (KYr) | | Outcomes | | Incidence (per KYr) | |
|---|---|---|---|---|---|---|---|---|
| | T | C | T | C | T | C | T | C |
| **PS-matching / on-treatment TAR** | | | | | | | | |
| CCAE | 11,234 | 84,259 | 7.5 | 72.7 | 55 | 336 | 7.32 (5.51 - 9.52) | 4.62 (4.14 - 5.14) |
| ODOD | 7,392 | 60,690 | 4.3 | 55.3 | 55 | 667 | 12.7 (9.60 - 16.6) | 12.1 (11.2 - 13.0) |
| MDCR | 914 | 14,983 | 0.6 | 16.4 | 5 | 183 | 7.97 (2.59 - 18.6) | 11.2 (9.60 - 12.9) |
| Open Claims | 136,139 | 845,324 | 96.9 | 915.3 | 419 | 4,538 | 4.32 (3.92 - 4.76) | 4.96 (4.81 - 5.10) |
| OEHR | 8,138 | 84,145 | 2.4 | 46.1 | 23 | 464 | 9.77 (6.20 - 14.7) | 10.1 (9.17 - 11.0) |
| VA | 1,273 | 31,466 | 0.9 | 23.7 | <10 | 162 | < 10.8 (5.16 - 19.8) | 6.83 (5.82 - 7.97) |
| Meta-analysis | 163,817 | 1,089,401 | 111.7 | 1105.9 | 557 | 6,188 | 4.99 (4.58 - 5.42) | 5.60 (5.46 - 5.74) |
| Meta-analysis (w/o USOC) | 27,678 | 244,077 | 14.8 | 190.5 | 138 | 1,650 | 9.32 (7.83 - 11.0) | 8.66 (8.25 - 9.09) |
| **PS-stratification / on-treatment TAR** | | | | | | | | |
| CCAE | 12,729 | 88,509 | 8.3 | 76.1 | 65 | 361 | 7.87 (6.08 - 10.0) | 4.74 (4.27 - 5.26) |
| ODOD | 8,275 | 67,640 | 4.7 | 63.4 | 61 | 834 | 13.0 (9.91 - 16.6) | 13.2 (12.3 - 14.1) |
| MDCR | 989 | 19,461 | 0.7 | 20.9 | 5 | 256 | 7.61 (2.47 - 17.8) | 12.2 (10.8 - 13.8) |
| Open Claims | 151,256 | 871,889 | 104.1 | 946.2 | 462 | 4,959 | 4.44 (4.04 - 4.86) | 5.24 (5.10 - 5.39) |
| OEHR | 8,866 | 92,813 | 2.5 | 51.3 | 25 | 552 | 9.92 (6.42 - 14.6) | 10.8 (9.89 - 11.7) |
| VA | 1,368 | 231,885 | 1.0 | 247.0 | 11 | 1,510 | 11.5 (5.73 - 20.6) | 6.11 (5.81 - 6.43) |
| Meta-analysis | 181,126 | 1,120,851 | 119.6 | 1136.9 | 613 | 6,706 | 5.12 (4.73 - 5.55) | 5.90 (5.76 - 6.04) |
| Meta-analysis (w/o USOC) | 29,870 | 248,962 | 15.5 | 190.7 | 151 | 1,747 | 9.75 (8.26 - 11.4) | 9.16 (8.73 - 9.60) |
| **PS-matching / on-treatment TAR without escalation** | | | | | | | | |
| CCAE | 11,234 | 84,259 | 6.5 | 63.0 | 49 | 287 | 7.51 (5.56 - 9.93) | 4.56 (4.04 - 5.12) |
| ODOD | 7,392 | 60,690 | 3.8 | 49.2 | 48 | 581 | 12.5 (9.21 - 16.6) | 11.8 (10.9 - 12.8) |
| MDCR | 914 | 14,983 | 0.6 | 14.8 | 5 | 161 | 9.06 (2.94 - 21.1) | 10.8 (9.23 - 12.7) |
| Open Claims | 136,139 | 845,324 | 84.2 | 775.1 | 354 | 3,853 | 4.20 (3.78 - 4.66) | 4.97 (4.82 - 5.13) |
| OEHR | 8,138 | 84,145 | 2.0 | 40.2 | 19 | 374 | 9.33 (5.61 - 14.6) | 9.31 (8.39 - 10.3) |
| VA | 1,273 | 31,466 | 0.7 | 20.8 | <10 | 139 | < 13.6 (6.52 - 25.0) | 6.69 (5.62 - 7.90) |
| Meta-analysis | 163,817 | 1,089,401 | 97.2 | 942.3 | 475 | 5,256 | 4.89 (4.46 - 5.35) | 5.58 (5.43 - 5.73) |
| Meta-analysis (w/o USOC) | 27,678 | 244,077 | 13.0 | 167.2 | 121 | 1,403 | 9.34 (7.75 - 11.2) | 8.39 (7.96 - 8.84) |
| **PS-stratification / on-treatment TAR without escalation** | | | | | | | | |
| CCAE | 12,729 | 88,509 | 7.2 | 66.1 | 59 | 311 | 8.18 (6.23 - 10.6) | 4.71 (4.20 - 5.26) |
| ODOD | 8,275 | 67,640 | 4.2 | 56.5 | 53 | 733 | 12.6 (9.47 - 16.5) | 13.0 (12.1 - 13.9) |
| MDCR | 989 | 19,461 | 0.6 | 19.1 | 5 | 228 | 8.62 (2.80 - 20.1) | 12.0 (10.5 - 13.6) |
| Open Claims | 151,256 | 871,889 | 91.0 | 801.6 | 394 | 4,225 | 4.33 (3.91 - 4.78) | 5.27 (5.11 - 5.43) |
| OEHR | 8,866 | 92,813 | 2.2 | 44.6 | 20 | 448 | 9.11 (5.57 - 14.1) | 10.1 (9.14 - 11.0) |
| VA | 1,368 | 231,885 | 0.8 | 220.0 | <10 | 1,312 | < 13.2 (6.32 - 24.2) | 5.96 (5.65 - 6.30) |
| Meta-analysis | 181,126 | 1,120,851 | 104.6 | 968.7 | 526 | 5,717 | 5.03 (4.61 - 5.48) | 5.90 (5.75 - 6.06) |
| Meta-analysis (w/o USOC) | 29,870 | 248,962 | 13.6 | 167.1 | 132 | 1,492 | 9.70 (8.12 - 11.5) | 8.93 (8.48 - 9.39) |

##### 4.5.12 Acute pancreatitis

**Supplementary Table 405:** Exposures, outcomes and risk estimates for Acute pancreatitis in GLP1RA (male) (T) vs SU (male) (C) new-user cohorts under four different propensity score (PS) adjustment / time-at-risk (TAR) designs across data sources. We report total patient exposure time in 1,000 patient-year (KYr) units, uncalibrated and calibrated hazard ratios (HRs) and their 95% confidence intervals (CIs). Greyed out entries did not pass study diagnostics and are included only for completeness.

| | Patients | | Exposure (KYr) | | Outcomes | | Incidence (per KYr) | |
|---|---|---|---|---|---|---|---|---|
| | T | C | T | C | T | C | T | C |
| **PS-matching / on-treatment TAR** | | | | | | | | |
| CCAE | 11,384 | 85,387 | 7.7 | 73.6 | 14 | 200 | 1.83 (1.00 - 3.07) | 2.72 (2.35 - 3.12) |
| ODOD | 7,559 | 62,479 | 4.5 | 57.1 | 9 | 193 | 2.02 (0.92 - 3.83) | 3.38 (2.92 - 3.89) |
| MDCR | 927 | 15,310 | 0.6 | 16.7 | 0 | 52 | | 3.11 (2.32 - 4.07) |
| Open Claims | 138,619 | 861,514 | 98.7 | 934.2 | 139 | 2,056 | 1.41 (1.18 - 1.66) | 2.20 (2.11 - 2.30) |
| OEHR | 8,327 | 85,885 | 2.4 | 47.3 | 7 | 135 | 2.89 (1.16 - 5.95) | 2.85 (2.39 - 3.38) |
| VA | 1,297 | 32,059 | 0.9 | 24.1 | 0 | 44 | | 1.82 (1.33 - 2.45) |
| Meta-analysis | 165,889 | 1,095,265 | 113.3 | 1112.2 | 169 | 2,584 | 1.49 (1.28 - 1.73) | 2.32 (2.23 - 2.41) |
| Meta-analysis (w/o USOC) | 27,270 | 233,751 | 14.5 | 178.0 | 30 | 528 | 2.06 (1.39 - 2.94) | 2.97 (2.72 - 3.23) |
| **PS-stratification / on-treatment TAR** | | | | | | | | |
| CCAE | 12,906 | 89,619 | 8.4 | 77.0 | 18 | 213 | 2.14 (1.27 - 3.38) | 2.76 (2.41 - 3.16) |
| ODOD | 8,472 | 69,985 | 4.9 | 65.7 | 9 | 238 | 1.85 (0.85 - 3.51) | 3.62 (3.18 - 4.11) |
| MDCR | 1,003 | 20,114 | 0.7 | 21.6 | 0 | 77 | | 3.57 (2.82 - 4.46) |
| Open Claims | 154,048 | 889,363 | 106.2 | 966.4 | 149 | 2,182 | 1.40 (1.19 - 1.65) | 2.26 (2.16 - 2.35) |
| OEHR | 9,062 | 95,290 | 2.6 | 52.9 | 7 | 164 | 2.70 (1.09 - 5.56) | 3.10 (2.64 - 3.61) |
| VA | 1,396 | 237,077 | 1.0 | 252.9 | 0 | 521 | | 2.06 (1.89 - 2.25) |
| Meta-analysis | 184,488 | 1,144,257 | 122.0 | 1162.1 | 183 | 2,797 | 1.50 (1.29 - 1.73) | 2.41 (2.32 - 2.50) |
| Meta-analysis (w/o USOC) | 30,440 | 254,894 | 15.9 | 195.7 | 34 | 615 | 2.14 (1.48 - 2.99) | 3.14 (2.90 - 3.40) |
| **PS-matching / on-treatment TAR without escalation** | | | | | | | | |
| CCAE | 11,384 | 85,387 | 6.6 | 63.8 | 9 | 168 | 1.35 (0.62 - 2.57) | 2.63 (2.25 - 3.06) |
| ODOD | 7,559 | 62,479 | 4.0 | 50.7 | 8 | 171 | 2.02 (0.87 - 3.98) | 3.37 (2.88 - 3.91) |
| MDCR | 927 | 15,310 | 0.6 | 15.1 | 0 | 45 | | 2.97 (2.17 - 3.98) |
| Open Claims | 138,619 | 861,514 | 85.8 | 791.2 | 113 | 1,699 | 1.32 (1.08 - 1.58) | 2.15 (2.05 - 2.25) |
| OEHR | 8,327 | 85,885 | 2.1 | 41.2 | 6 | 99 | 2.86 (1.05 - 6.23) | 2.41 (1.95 - 2.93) |
| VA | 1,297 | 32,059 | 0.7 | 21.1 | 0 | 37 | | 1.75 (1.23 - 2.41) |
| Meta-analysis | 165,889 | 1,095,265 | 98.5 | 946.9 | 136 | 2,137 | 1.38 (1.16 - 1.63) | 2.26 (2.16 - 2.35) |
| Meta-analysis (w/o USOC) | 27,270 | 233,751 | 12.7 | 155.7 | 23 | 438 | 1.81 (1.15 - 2.72) | 2.81 (2.56 - 3.09) |
| **PS-stratification / on-treatment TAR without escalation** | | | | | | | | |
| CCAE | 12,906 | 89,619 | 7.4 | 66.9 | 13 | 181 | 1.77 (0.94 - 3.02) | 2.71 (2.33 - 3.13) |
| ODOD | 8,472 | 69,985 | 4.3 | 58.6 | 8 | 208 | 1.85 (0.80 - 3.65) | 3.55 (3.08 - 4.07) |
| MDCR | 1,003 | 20,114 | 0.6 | 19.6 | 0 | 67 | | 3.41 (2.65 - 4.34) |
| Open Claims | 154,048 | 889,363 | 92.8 | 819.0 | 123 | 1,807 | 1.33 (1.10 - 1.58) | 2.21 (2.11 - 2.31) |
| OEHR | 9,062 | 95,290 | 2.3 | 46.0 | 6 | 120 | 2.66 (0.98 - 5.80) | 2.61 (2.16 - 3.12) |
| VA | 1,396 | 237,077 | 0.8 | 225.3 | 0 | 431 | | 1.91 (1.74 - 2.10) |
| Meta-analysis | 184,488 | 1,144,257 | 106.7 | 990.4 | 150 | 2,316 | 1.41 (1.19 - 1.65) | 2.34 (2.24 - 2.44) |
| Meta-analysis (w/o USOC) | 30,440 | 254,894 | 13.9 | 171.5 | 27 | 509 | 1.94 (1.28 - 2.82) | 2.97 (2.72 - 3.24) |

###### 4.5.13 Bone fracture

**Supplementary Table 406:** Exposures, outcomes and risk estimates for Bone fracture in GLP1RA (male) (T) vs SU (male) (C) new-user cohorts under four different propensity score (PS) adjustment / time-at-risk (TAR) designs across data sources. We report total patient exposure time in 1,000 patient-year (KYr) units, uncalibrated and calibrated hazard ratios (HRs) and their 95% confidence intervals (CIs). Greyed out entries did not pass study diagnostics and are included only for completeness.

| | Patients | | Exposure (KYr) | | Outcomes | | Incidence (per KYr) | |
|---|---|---|---|---|---|---|---|---|
| | T | C | T | C | T | C | T | C |
| **PS-matching / on-treatment TAR** | | | | | | | | |
| CCAE | 10,570 | 80,684 | 7.0 | 68.6 | 114 | 1,068 | 16.3 (13.5 - 19.6) | 15.6 (14.6 - 16.5) |
| ODOD | 7,048 | 58,762 | 4.1 | 52.8 | 101 | 1,109 | 24.7 (20.1 - 30.0) | 21.0 (19.8 - 22.3) |
| MDCR | 850 | 14,248 | 0.6 | 15.3 | 11 | 380 | 19.7 (9.84 - 35.3) | 24.9 (22.5 - 27.5) |
| Open Claims | 122,443 | 783,604 | 86.8 | 841.6 | 932 | 11,844 | 10.7 (10.1 - 11.5) | 14.1 (13.8 - 14.3) |
| OEHR | 7,836 | 81,327 | 2.3 | 44.0 | 29 | 729 | 12.7 (8.51 - 18.2) | 16.6 (15.4 - 17.8) |
| VA | 1,173 | 29,601 | 0.8 | 22.2 | <10 | 248 | < 11.8 (5.67 - 21.8) | 11.2 (9.81 - 12.6) |
| Meta-analysis | 148,747 | 1,018,625 | 100.7 | 1022.4 | 1,187 | 15,130 | 11.8 (11.1 - 12.5) | 14.8 (14.6 - 15.0) |
| Meta-analysis (w/o USOC) | 26,304 | 235,021 | 13.9 | 180.8 | 255 | 3,286 | 18.3 (16.1 - 20.7) | 18.2 (17.6 - 18.8) |
| **PS-stratification / on-treatment TAR** | | | | | | | | |
| CCAE | 11,928 | 84,627 | 7.7 | 71.8 | 126 | 1,110 | 16.5 (13.7 - 19.6) | 15.5 (14.6 - 16.4) |
| ODOD | 7,882 | 66,029 | 4.5 | 61.0 | 111 | 1,355 | 24.9 (20.5 - 30.0) | 22.2 (21.1 - 23.4) |
| MDCR | 916 | 18,429 | 0.6 | 19.4 | 11 | 515 | 18.8 (9.39 - 33.7) | 26.5 (24.3 - 28.9) |
| Open Claims | 135,593 | 811,681 | 93.0 | 873.6 | 1,016 | 12,691 | 10.9 (10.3 - 11.6) | 14.5 (14.3 - 14.8) |
| OEHR | 8,521 | 90,116 | 2.4 | 49.2 | 30 | 894 | 12.3 (8.31 - 17.6) | 18.2 (17.0 - 19.4) |
| VA | 1,265 | 219,434 | 0.9 | 232.5 | <10 | 2,502 | < 11.4 (5.47 - 21.0) | 10.8 (10.3 - 11.2) |
| Meta-analysis | 163,924 | 1,052,453 | 107.6 | 1055.5 | 1,283 | 16,050 | 11.9 (11.3 - 12.6) | 15.2 (15.0 - 15.4) |
| Meta-analysis (w/o USOC) | 28,331 | 240,772 | 14.5 | 181.9 | 267 | 3,359 | 18.4 (16.2 - 20.7) | 18.5 (17.8 - 19.1) |
| **PS-matching / on-treatment TAR without escalation** | | | | | | | | |
| CCAE | 10,570 | 80,684 | 6.1 | 59.6 | 91 | 927 | 15.0 (12.1 - 18.4) | 15.6 (14.6 - 16.6) |
| ODOD | 7,048 | 58,762 | 3.6 | 47.1 | 90 | 991 | 24.8 (19.9 - 30.5) | 21.1 (19.8 - 22.4) |
| MDCR | 850 | 14,248 | 0.5 | 13.8 | 10 | 348 | 20.1 (9.64 - 37.0) | 25.1 (22.6 - 27.9) |
| Open Claims | 122,443 | 783,604 | 75.4 | 713.9 | 792 | 9,958 | 10.5 (9.78 - 11.3) | 13.9 (13.7 - 14.2) |
| OEHR | 7,836 | 81,327 | 2.0 | 38.5 | 23 | 611 | 11.7 (7.40 - 17.5) | 15.9 (14.7 - 17.2) |
| VA | 1,173 | 29,601 | 0.7 | 19.5 | <10 | 220 | < 15.0 (7.21 - 27.7) | 11.3 (9.85 - 12.9) |
| Meta-analysis | 148,747 | 1,018,625 | 87.6 | 872.9 | 1,006 | 12,835 | 11.5 (10.8 - 12.2) | 14.7 (14.5 - 15.0) |
| Meta-analysis (w/o USOC) | 26,304 | 235,021 | 12.2 | 159.0 | 214 | 2,877 | 17.6 (15.3 - 20.1) | 18.1 (17.4 - 18.8) |
| **PS-stratification / on-treatment TAR without escalation** | | | | | | | | |
| CCAE | 11,928 | 84,627 | 6.7 | 62.4 | 102 | 965 | 15.3 (12.5 - 18.5) | 15.5 (14.5 - 16.5) |
| ODOD | 7,882 | 66,029 | 4.0 | 54.5 | 99 | 1,222 | 25.0 (20.3 - 30.4) | 22.4 (21.2 - 23.7) |
| MDCR | 916 | 18,429 | 0.5 | 17.7 | 10 | 471 | 19.2 (9.19 - 35.2) | 26.6 (24.3 - 29.1) |
| Open Claims | 135,593 | 811,681 | 81.3 | 741.6 | 869 | 10,693 | 10.7 (9.99 - 11.4) | 14.4 (14.1 - 14.7) |
| OEHR | 8,521 | 90,116 | 2.1 | 42.9 | 24 | 745 | 11.4 (7.27 - 16.9) | 17.4 (16.1 - 18.7) |
| VA | 1,265 | 219,434 | 0.7 | 207.6 | <10 | 2,203 | < 14.4 (6.92 - 26.5) | 10.6 (10.2 - 11.1) |
| Meta-analysis | 163,924 | 1,052,453 | 94.0 | 901.4 | 1,094 | 13,625 | 11.6 (11.0 - 12.3) | 15.1 (14.9 - 15.4) |
| Meta-analysis (w/o USOC) | 28,331 | 240,772 | 12.8 | 159.8 | 225 | 2,932 | 17.6 (15.4 - 20.1) | 18.3 (17.7 - 19.0) |

##### 4.5.14 Diabetic ketoacidosis

**Supplementary Table 407:** Exposures, outcomes and risk estimates for Diabetic ketoacidosis in GLP1RA (male) (T) vs SU (male) (C) new-user cohorts under four different propensity score (PS) adjustment / time-at-risk (TAR) designs across data sources. We report total patient exposure time in 1,000 patient-year (KYr) units, uncalibrated and calibrated hazard ratios (HRs) and their 95% confidence intervals (CIs). Greyed out entries did not pass study diagnostics and are included only for completeness.

| | Patients | | Exposure (KYr) | | Outcomes | | Incidence (per KYr) | |
|---|---|---|---|---|---|---|---|---|
| | T | C | T | C | T | C | T | C |
| **PS-matching / on-treatment TAR** | | | | | | | | |
| CCAE | 11,456 | 86,109 | 7.7 | 74.3 | 8 | 52 | 1.04 (0.45 - 2.05) | 0.70 (0.52 - 0.92) |
| ODOD | 7,588 | 62,916 | 4.5 | 57.6 | <5 | 71 | < 1.12 (0.36 - 2.61) | 1.23 (0.96 - 1.55) |
| MDCR | 932 | 15,496 | 0.6 | 17.0 | 0 | 16 | | 0.94 (0.54 - 1.53) |
| Open Claims | 139,827 | 871,590 | 99.6 | 946.0 | 67 | 727 | 0.67 (0.52 - 0.85) | 0.77 (0.71 - 0.83) |
| OEHR | 8,366 | 86,386 | 2.4 | 47.6 | <5 | 56 | < 2.05 (0.67 - 4.79) | 1.18 (0.89 - 1.53) |
| VA | 1,312 | 32,262 | 0.9 | 24.3 | <10 | 19 | < 10.6 (5.06 - 19.4) | 0.78 (0.47 - 1.22) |
| Meta-analysis | 167,237 | 1,107,001 | 114.2 | 1125.5 | <85 | 906 | < 0.74 (0.59 - 0.92) | 0.80 (0.75 - 0.86) |
| Meta-analysis (w/o USOC) | 27,410 | 235,411 | 14.6 | 179.6 | <18 | 179 | < 1.23 (0.73 - 1.95) | 1.00 (0.86 - 1.15) |
| **PS-stratification / on-treatment TAR** | | | | | | | | |
| CCAE | 12,980 | 90,423 | 8.5 | 77.8 | 8 | 55 | 0.95 (0.41 - 1.86) | 0.71 (0.53 - 0.92) |
| ODOD | 8,503 | 70,638 | 4.9 | 66.5 | <5 | 87 | < 1.03 (0.33 - 2.39) | 1.31 (1.05 - 1.61) |
| MDCR | 1,007 | 20,353 | 0.7 | 21.9 | 0 | 23 | | 1.05 (0.67 - 1.58) |
| Open Claims | 155,271 | 900,860 | 107.0 | 979.7 | 70 | 788 | 0.65 (0.51 - 0.83) | 0.80 (0.75 - 0.86) |
| OEHR | 9,102 | 95,832 | 2.6 | 53.3 | <5 | 62 | < 1.92 (0.62 - 4.48) | 1.16 (0.89 - 1.49) |
| VA | 1,411 | 239,435 | 1.0 | 255.7 | <10 | 127 | < 10.2 (4.90 - 18.8) | 0.50 (0.41 - 0.59) |
| Meta-analysis | 185,856 | 1,157,753 | 122.9 | 1177.4 | <88 | 992 | < 0.72 (0.57 - 0.88) | 0.84 (0.79 - 0.90) |
| Meta-analysis (w/o USOC) | 30,585 | 256,893 | 15.9 | 197.6 | <18 | 204 | < 1.13 (0.67 - 1.78) | 1.03 (0.90 - 1.18) |
| **PS-matching / on-treatment TAR without escalation** | | | | | | | | |
| CCAE | 11,456 | 86,109 | 6.7 | 64.4 | 7 | 40 | 1.05 (0.42 - 2.16) | 0.62 (0.44 - 0.85) |
| ODOD | 7,588 | 62,916 | 4.0 | 51.2 | <5 | 52 | < 1.26 (0.41 - 2.94) | 1.02 (0.76 - 1.33) |
| MDCR | 932 | 15,496 | 0.6 | 15.3 | 0 | 12 | | 0.78 (0.40 - 1.37) |
| Open Claims | 139,827 | 871,590 | 86.5 | 800.8 | 56 | 557 | 0.65 (0.49 - 0.84) | 0.70 (0.64 - 0.76) |
| OEHR | 8,366 | 86,386 | 2.1 | 41.4 | <5 | 30 | < 2.38 (0.77 - 5.55) | 0.72 (0.49 - 1.03) |
| VA | 1,312 | 32,262 | 0.7 | 21.3 | <10 | 12 | < 13.4 (6.41 - 24.6) | 0.56 (0.29 - 0.98) |
| Meta-analysis | 158,871 | 1,020,615 | 97.2 | 916.4 | <68 | 649 | < 0.70 (0.54 - 0.89) | 0.71 (0.65 - 0.76) |
| Meta-analysis (w/o USOC) | 19,044 | 149,025 | 10.6 | 115.6 | <12 | 92 | < 1.13 (0.58 - 1.97) | 0.80 (0.64 - 0.98) |
| **PS-stratification / on-treatment TAR without escalation** | | | | | | | | |
| CCAE | 12,980 | 90,423 | 7.4 | 67.5 | 7 | 43 | 0.95 (0.38 - 1.95) | 0.64 (0.46 - 0.86) |
| ODOD | 8,503 | 70,638 | 4.3 | 59.2 | <5 | 65 | < 1.15 (0.38 - 2.70) | 1.10 (0.85 - 1.40) |
| MDCR | 1,007 | 20,353 | 0.6 | 19.9 | 0 | 19 | | 0.96 (0.58 - 1.49) |
| Open Claims | 155,271 | 900,860 | 93.5 | 829.9 | 58 | 606 | 0.62 (0.47 - 0.80) | 0.73 (0.67 - 0.79) |
| OEHR | 9,102 | 95,832 | 2.3 | 46.3 | <5 | 35 | < 2.21 (0.72 - 5.16) | 0.76 (0.53 - 1.05) |
| VA | 1,411 | 239,435 | 0.8 | 227.6 | <10 | 91 | < 12.9 (6.19 - 23.7) | 0.40 (0.32 - 0.49) |
| Meta-analysis | 176,754 | 1,061,921 | 105.2 | 956.7 | <70 | 714 | < 0.67 (0.52 - 0.84) | 0.75 (0.69 - 0.80) |
| Meta-analysis (w/o USOC) | 21,483 | 161,061 | 11.7 | 126.8 | <12 | 108 | < 1.02 (0.53 - 1.79) | 0.85 (0.70 - 1.03) |

##### 4.5.15 Diarrhea

**Supplementary Table 408:** Exposures, outcomes and risk estimates for Diarrhea in GLP1RA (male) (T) vs SU (male) (C) new-user cohorts under four different propensity score (PS) adjustment / time-at-risk (TAR) designs across data sources. We report total patient exposure time in 1,000 patient-year (KYr) units, uncalibrated and calibrated hazard ratios (HRs) and their 95% confidence intervals (CIs). Greyed out entries did not pass study diagnostics and are included only for completeness.

| | Patients | | Exposure (KYr) | | Outcomes | | Incidence (per KYr) | |
|---|---|---|---|---|---|---|---|---|
| | T | C | T | C | T | C | T | C |
| **PS-matching / on-treatment TAR** | | | | | | | | |
| CCAE | 10,329 | 79,209 | 6.8 | 67.3 | 243 | 1,390 | 35.9 (31.5 - 40.7) | 20.7 (19.6 - 21.8) |
| ODOD | 6,819 | 56,936 | 3.9 | 50.7 | 166 | 1,603 | 42.4 (36.2 - 49.4) | 31.6 (30.1 - 33.2) |
| MDCR | 836 | 13,964 | 0.6 | 14.9 | 18 | 393 | 31.8 (18.8 - 50.2) | 26.4 (23.8 - 29.1) |
| Open Claims | 124,238 | 792,429 | 87.6 | 848.8 | 1,593 | 12,255 | 18.2 (17.3 - 19.1) | 14.4 (14.2 - 14.7) |
| OEHR | 7,621 | 79,157 | 2.2 | 42.0 | 91 | 1,318 | 42.2 (34.0 - 51.8) | 31.4 (29.7 - 33.1) |
| VA | 1,109 | 27,701 | 0.8 | 20.7 | 22 | 389 | 27.7 (17.4 - 42.0) | 18.8 (17.0 - 20.8) |
| Meta-analysis | 149,843 | 1,021,695 | 101.0 | 1023.7 | 2,111 | 16,959 | 20.9 (20.0 - 21.8) | 16.6 (16.3 - 16.8) |
| Meta-analysis (w/o USOC) | 25,605 | 229,266 | 13.4 | 174.9 | 518 | 4,704 | 38.6 (35.4 - 42.1) | 26.9 (26.1 - 27.7) |
| **PS-stratification / on-treatment TAR** | | | | | | | | |
| CCAE | 11,634 | 83,498 | 7.4 | 70.7 | 278 | 1,463 | 37.5 (33.2 - 42.2) | 20.7 (19.6 - 21.8) |
| ODOD | 7,599 | 63,534 | 4.3 | 58.0 | 191 | 1,910 | 44.8 (38.7 - 51.6) | 32.9 (31.5 - 34.4) |
| MDCR | 901 | 18,340 | 0.6 | 19.2 | 18 | 543 | 30.6 (18.1 - 48.3) | 28.3 (26.0 - 30.8) |
| Open Claims | 137,360 | 819,429 | 93.7 | 879.3 | 1,776 | 13,027 | 18.9 (18.1 - 19.8) | 14.8 (14.6 - 15.1) |
| OEHR | 8,239 | 88,182 | 2.3 | 47.1 | 99 | 1,557 | 43.0 (34.9 - 52.3) | 33.1 (31.4 - 34.7) |
| VA | 1,195 | 214,202 | 0.8 | 224.2 | 23 | 4,038 | 28.0 (17.8 - 42.0) | 18.0 (17.5 - 18.6) |
| Meta-analysis | 164,832 | 1,054,643 | 107.7 | 1055.2 | 2,344 | 17,957 | 21.8 (20.9 - 22.7) | 17.0 (16.8 - 17.3) |
| Meta-analysis (w/o USOC) | 27,472 | 235,214 | 14.0 | 175.8 | 568 | 4,930 | 40.6 (37.4 - 44.1) | 28.0 (27.3 - 28.8) |
| **PS-matching / on-treatment TAR without escalation** | | | | | | | | |
| CCAE | 10,329 | 79,209 | 5.9 | 58.5 | 218 | 1,208 | 36.9 (32.2 - 42.2) | 20.6 (19.5 - 21.8) |
| ODOD | 6,819 | 56,936 | 3.5 | 45.3 | 145 | 1,424 | 41.5 (35.0 - 48.9) | 31.4 (29.8 - 33.1) |
| MDCR | 836 | 13,964 | 0.5 | 13.5 | 16 | 356 | 32.0 (18.3 - 51.9) | 26.3 (23.6 - 29.2) |
| Open Claims | 124,238 | 792,429 | 76.3 | 721.7 | 1,375 | 10,270 | 18.0 (17.1 - 19.0) | 14.2 (14.0 - 14.5) |
| OEHR | 7,621 | 79,157 | 1.9 | 36.9 | 78 | 1,057 | 41.8 (33.0 - 52.2) | 28.6 (26.9 - 30.4) |
| VA | 1,109 | 27,701 | 0.6 | 18.2 | 19 | 337 | 30.5 (18.4 - 47.6) | 18.5 (16.6 - 20.6) |
| Meta-analysis | 149,843 | 1,021,695 | 88.1 | 876.1 | 1,832 | 14,315 | 20.8 (19.9 - 21.8) | 16.3 (16.1 - 16.6) |
| Meta-analysis (w/o USOC) | 25,605 | 229,266 | 11.8 | 154.3 | 457 | 4,045 | 38.9 (35.4 - 42.6) | 26.2 (25.4 - 27.0) |
| **PS-stratification / on-treatment TAR without escalation** | | | | | | | | |
| CCAE | 11,634 | 83,498 | 6.5 | 61.7 | 250 | 1,273 | 38.5 (33.9 - 43.6) | 20.6 (19.5 - 21.8) |
| ODOD | 7,599 | 63,534 | 3.8 | 52.0 | 165 | 1,712 | 43.4 (37.0 - 50.6) | 33.0 (31.4 - 34.6) |
| MDCR | 901 | 18,340 | 0.5 | 17.5 | 16 | 498 | 30.7 (17.6 - 49.9) | 28.4 (26.0 - 31.0) |
| Open Claims | 137,360 | 819,429 | 82.1 | 748.1 | 1,549 | 10,955 | 18.9 (17.9 - 19.8) | 14.6 (14.4 - 14.9) |
| OEHR | 8,239 | 88,182 | 2.0 | 41.4 | 85 | 1,243 | 42.3 (33.8 - 52.4) | 30.0 (28.4 - 31.8) |
| VA | 1,195 | 214,202 | 0.6 | 200.8 | 20 | 3,535 | 30.9 (18.9 - 47.8) | 17.6 (17.0 - 18.2) |
| Meta-analysis | 164,832 | 1,054,643 | 94.4 | 903.1 | 2,049 | 15,183 | 21.7 (20.8 - 22.7) | 16.8 (16.5 - 17.1) |
| Meta-analysis (w/o USOC) | 27,472 | 235,214 | 12.3 | 155.0 | 500 | 4,228 | 40.6 (37.2 - 44.4) | 27.3 (26.5 - 28.1) |

##### 4.5.16 Genitourinary infection

**Supplementary Table 409:** Exposures, outcomes and risk estimates for Genitourinary infection in GLP1RA (male) (T) vs SU (male) (C) new-user cohorts under four different propensity score (PS) adjustment / time-at-risk (TAR) designs across data sources. We report total patient exposure time in 1,000 patient-year (KYr) units, uncalibrated and calibrated hazard ratios (HRs) and their 95% confidence intervals (CIs). Greyed out entries did not pass study diagnostics and are included only for completeness.

| | Patients | | Exposure (KYr) | | Outcomes | | Incidence (per KYr) | |
|---|---|---|---|---|---|---|---|---|
| | T | C | T | C | T | C | T | C |
| **PS-matching / on-treatment TAR** | | | | | | | | |
| CCAE | 10,455 | 78,994 | 6.9 | 67.0 | 144 | 1,346 | 20.9 (17.6 - 24.6) | 20.1 (19.0 - 21.2) |
| ODOD | 6,788 | 54,840 | 3.9 | 48.3 | 108 | 1,836 | 27.5 (22.6 - 33.2) | 38.0 (36.3 - 39.8) |
| MDCR | 814 | 13,541 | 0.5 | 14.4 | 28 | 555 | 53.8 (35.8 - 77.8) | 38.5 (35.4 - 41.9) |
| Open Claims | 124,593 | 776,506 | 88.5 | 828.9 | 1,171 | 15,299 | 13.2 (12.5 - 14.0) | 18.5 (18.2 - 18.8) |
| OEHR | 7,896 | 81,313 | 2.3 | 43.7 | 47 | 1,007 | 20.6 (15.2 - 27.5) | 23.0 (21.6 - 24.5) |
| VA | 1,170 | 28,977 | 0.9 | 21.6 | 15 | 325 | 17.5 (9.82 - 28.9) | 15.0 (13.5 - 16.8) |
| Meta-analysis | 150,546 | 1,005,194 | 102.1 | 1002.3 | 1,498 | 20,043 | 14.7 (13.9 - 15.4) | 20.0 (19.7 - 20.3) |
| Meta-analysis (w/o USOC) | 25,953 | 228,688 | 13.6 | 173.4 | 327 | 4,744 | 24.0 (21.5 - 26.8) | 27.4 (26.6 - 28.2) |
| **PS-stratification / on-treatment TAR** | | | | | | | | |
| CCAE | 11,792 | 83,057 | 7.5 | 70.2 | 153 | 1,414 | 20.3 (17.2 - 23.8) | 20.1 (19.1 - 21.2) |
| ODOD | 7,578 | 61,676 | 4.3 | 55.7 | 121 | 2,374 | 28.3 (23.5 - 33.8) | 42.7 (41.0 - 44.4) |
| MDCR | 868 | 17,360 | 0.5 | 18.2 | 28 | 795 | 51.6 (34.3 - 74.5) | 43.8 (40.8 - 46.9) |
| Open Claims | 138,123 | 802,465 | 95.0 | 857.9 | 1,277 | 16,698 | 13.4 (12.7 - 14.2) | 19.5 (19.2 - 19.8) |
| OEHR | 8,578 | 90,094 | 2.4 | 48.8 | 50 | 1,242 | 20.5 (15.2 - 27.1) | 25.5 (24.1 - 26.9) |
| VA | 1,261 | 216,571 | 0.9 | 227.0 | 15 | 3,830 | 17.0 (9.51 - 28.0) | 16.9 (16.3 - 17.4) |
| Meta-analysis | 166,071 | 1,037,292 | 109.3 | 1032.6 | 1,601 | 21,728 | 14.7 (13.9 - 15.4) | 21.0 (20.8 - 21.3) |
| Meta-analysis (w/o USOC) | 27,948 | 234,827 | 14.3 | 174.7 | 324 | 5,030 | 22.7 (20.3 - 25.3) | 28.8 (28.0 - 29.6) |
| **PS-matching / on-treatment TAR without escalation** | | | | | | | | |
| CCAE | 10,455 | 78,994 | 6.0 | 58.2 | 125 | 1,176 | 20.8 (17.4 - 24.8) | 20.2 (19.1 - 21.4) |
| ODOD | 6,788 | 54,840 | 3.5 | 43.0 | 97 | 1,658 | 27.8 (22.5 - 33.9) | 38.6 (36.7 - 40.5) |
| MDCR | 814 | 13,541 | 0.5 | 13.1 | 22 | 508 | 47.9 (30.0 - 72.5) | 38.9 (35.6 - 42.5) |
| Open Claims | 124,593 | 776,506 | 77.0 | 702.7 | 1,013 | 13,064 | 13.1 (12.4 - 14.0) | 18.6 (18.3 - 18.9) |
| OEHR | 7,896 | 81,313 | 2.0 | 38.2 | 42 | 798 | 21.3 (15.4 - 28.9) | 20.9 (19.4 - 22.4) |
| VA | 1,170 | 28,977 | 0.7 | 18.9 | 12 | 281 | 18.0 (9.29 - 31.4) | 14.9 (13.2 - 16.7) |
| Meta-analysis | 150,546 | 1,005,194 | 89.0 | 855.2 | 1,299 | 17,204 | 14.6 (13.8 - 15.4) | 20.1 (19.8 - 20.4) |
| Meta-analysis (w/o USOC) | 25,953 | 228,688 | 11.9 | 152.5 | 286 | 4,140 | 24.0 (21.3 - 27.0) | 27.2 (26.3 - 28.0) |
| **PS-stratification / on-treatment TAR without escalation** | | | | | | | | |
| CCAE | 11,792 | 83,057 | 6.6 | 61.1 | 133 | 1,238 | 20.1 (16.9 - 23.9) | 20.3 (19.1 - 21.4) |
| ODOD | 7,578 | 61,676 | 3.8 | 49.7 | 110 | 2,156 | 28.9 (23.8 - 34.8) | 43.4 (41.6 - 45.3) |
| MDCR | 868 | 17,360 | 0.5 | 16.6 | 22 | 718 | 45.7 (28.7 - 69.2) | 43.4 (40.3 - 46.7) |
| Open Claims | 138,123 | 802,465 | 83.1 | 727.8 | 1,115 | 14,311 | 13.4 (12.6 - 14.2) | 19.7 (19.3 - 20.0) |
| OEHR | 8,578 | 90,094 | 2.1 | 42.6 | 45 | 981 | 21.3 (15.5 - 28.4) | 23.0 (21.6 - 24.5) |
| VA | 1,261 | 216,571 | 0.7 | 202.7 | 12 | 3,300 | 17.4 (8.97 - 30.3) | 16.3 (15.7 - 16.8) |
| Meta-analysis | 166,071 | 1,037,292 | 95.6 | 881.3 | 1,403 | 18,686 | 14.7 (13.9 - 15.5) | 21.2 (20.9 - 21.5) |
| Meta-analysis (w/o USOC) | 27,948 | 234,827 | 12.5 | 153.5 | 288 | 4,375 | 23.0 (20.4 - 25.8) | 28.5 (27.7 - 29.4) |

##### 4.5.17 Hyperkalemia

**Supplementary Table 410:** Exposures, outcomes and risk estimates for Hyperkalemia in GLP1RA (male) (T) vs SU (male) (C) new-user cohorts under four different propensity score (PS) adjustment / time-at-risk (TAR) designs across data sources. We report total patient exposure time in 1,000 patient-year (KYr) units, uncalibrated and calibrated hazard ratios (HRs) and their 95% confidence intervals (CIs). Greyed out entries did not pass study diagnostics and are included only for completeness.

| | Patients | | Exposure (KYr) | | Outcomes | | Incidence (per KYr) | |
|---|---|---|---|---|---|---|---|---|
| | T | C | T | C | T | C | T | C |
| **PS-matching / on-treatment TAR** | | | | | | | | |
| CCAE | 11,300 | 84,787 | 7.5 | 72.7 | 49 | 564 | 6.50 (4.81 - 8.59) | 7.76 (7.13 - 8.43) |
| ODOD | 7,326 | 60,481 | 4.3 | 54.2 | 70 | 1,220 | 16.4 (12.8 - 20.7) | 22.5 (21.3 - 23.8) |
| MDCR | 911 | 15,191 | 0.6 | 16.5 | 7 | 249 | 11.0 (4.44 - 22.8) | 15.1 (13.3 - 17.1) |
| Open Claims | 137,448 | 854,895 | 97.6 | 919.0 | 378 | 7,452 | 3.87 (3.49 - 4.28) | 8.11 (7.93 - 8.29) |
| OEHR | 8,215 | 84,844 | 2.4 | 46.0 | 20 | 677 | 8.37 (5.11 - 12.9) | 14.7 (13.6 - 15.9) |
| VA | 1,219 | 30,847 | 0.9 | 23.1 | <10 | 347 | < 11.3 (5.42 - 20.8) | 15.0 (13.5 - 16.7) |
| Meta-analysis | 165,200 | 1,100,198 | 112.4 | 1108.4 | 524 | 10,162 | 4.66 (4.27 - 5.08) | 9.17 (8.99 - 9.35) |
| Meta-analysis (w/o USOC) | 27,752 | 245,303 | 14.8 | 189.4 | 146 | 2,710 | 9.84 (8.31 - 11.6) | 14.3 (13.8 - 14.9) |
| **PS-stratification / on-treatment TAR** | | | | | | | | |
| CCAE | 12,798 | 89,234 | 8.3 | 76.3 | 55 | 596 | 6.63 (5.00 - 8.63) | 7.82 (7.20 - 8.47) |
| ODOD | 8,212 | 67,531 | 4.7 | 62.0 | 70 | 1,534 | 15.0 (11.7 - 19.0) | 24.7 (23.5 - 26.0) |
| MDCR | 986 | 19,737 | 0.7 | 21.0 | 7 | 369 | 10.6 (4.26 - 21.8) | 17.6 (15.8 - 19.5) |
| Open Claims | 152,722 | 882,468 | 104.9 | 950.2 | 394 | 8,247 | 3.75 (3.39 - 4.14) | 8.68 (8.49 - 8.87) |
| OEHR | 8,937 | 93,845 | 2.5 | 51.2 | 20 | 841 | 7.85 (4.79 - 12.1) | 16.4 (15.3 - 17.6) |
| VA | 1,318 | 225,516 | 0.9 | 235.1 | <10 | 4,092 | < 10.9 (5.23 - 20.1) | 17.4 (16.9 - 17.9) |
| Meta-analysis | 182,669 | 1,133,078 | 120.4 | 1139.7 | 539 | 11,218 | 4.48 (4.11 - 4.87) | 9.84 (9.66 - 10.0) |
| Meta-analysis (w/o USOC) | 29,947 | 250,610 | 15.5 | 189.5 | 145 | 2,971 | 9.35 (7.89 - 11.0) | 15.7 (15.1 - 16.3) |
| **PS-matching / on-treatment TAR without escalation** | | | | | | | | |
| CCAE | 11,300 | 84,787 | 6.6 | 63.0 | 38 | 490 | 5.80 (4.10 - 7.96) | 7.78 (7.10 - 8.50) |
| ODOD | 7,326 | 60,481 | 3.8 | 48.2 | 61 | 1,081 | 16.1 (12.3 - 20.6) | 22.4 (21.1 - 23.8) |
| MDCR | 911 | 15,191 | 0.6 | 14.9 | 6 | 223 | 10.7 (3.94 - 23.4) | 15.0 (13.1 - 17.1) |
| Open Claims | 137,448 | 854,895 | 84.9 | 778.8 | 320 | 6,209 | 3.77 (3.37 - 4.21) | 7.97 (7.78 - 8.17) |
| OEHR | 8,215 | 84,844 | 2.1 | 40.2 | 15 | 517 | 7.26 (4.06 - 12.0) | 12.9 (11.8 - 14.0) |
| VA | 1,219 | 30,847 | 0.7 | 20.2 | <10 | 291 | < 14.4 (6.91 - 26.5) | 14.4 (12.8 - 16.1) |
| Meta-analysis | 165,200 | 1,100,198 | 97.8 | 945.1 | 440 | 8,520 | 4.50 (4.09 - 4.94) | 9.01 (8.82 - 9.21) |
| Meta-analysis (w/o USOC) | 27,752 | 245,303 | 13.0 | 166.3 | 120 | 2,311 | 9.25 (7.67 - 11.1) | 13.9 (13.3 - 14.5) |
| **PS-stratification / on-treatment TAR without escalation** | | | | | | | | |
| CCAE | 12,798 | 89,234 | 7.2 | 66.2 | 43 | 517 | 5.93 (4.29 - 7.99) | 7.81 (7.15 - 8.51) |
| ODOD | 8,212 | 67,531 | 4.2 | 55.4 | 61 | 1,356 | 14.7 (11.2 - 18.9) | 24.5 (23.2 - 25.8) |
| MDCR | 986 | 19,737 | 0.6 | 19.1 | 6 | 336 | 10.3 (3.77 - 22.3) | 17.6 (15.8 - 19.6) |
| Open Claims | 152,722 | 882,468 | 91.7 | 805.8 | 334 | 6,891 | 3.64 (3.26 - 4.05) | 8.55 (8.35 - 8.76) |
| OEHR | 8,937 | 93,845 | 2.2 | 44.7 | 15 | 626 | 6.77 (3.79 - 11.2) | 14.0 (12.9 - 15.1) |
| VA | 1,318 | 225,516 | 0.7 | 210.0 | <10 | 3,455 | < 13.9 (6.65 - 25.5) | 16.5 (15.9 - 17.0) |
| Meta-analysis | 182,669 | 1,133,078 | 105.3 | 972.1 | 453 | 9,390 | 4.30 (3.91 - 4.72) | 9.66 (9.47 - 9.86) |
| Meta-analysis (w/o USOC) | 29,947 | 250,610 | 13.6 | 166.3 | 119 | 2,499 | 8.74 (7.24 - 10.5) | 15.0 (14.4 - 15.6) |

##### 4.5.18 Hypoglycemia

**Supplementary Table 411:** Exposures, outcomes and risk estimates for Hypoglycemia in GLP1RA (male) (T) vs SU (male) (C) new-user cohorts under four different propensity score (PS) adjustment / time-at-risk (TAR) designs across data sources. We report total patient exposure time in 1,000 patient-year (KYr) units, uncalibrated and calibrated hazard ratios (HRs) and their 95% confidence intervals (CIs). Greyed out entries did not pass study diagnostics and are included only for completeness.

| | Patients | | Exposure (KYr) | | Outcomes | | Incidence (per KYr) | |
|---|---|---|---|---|---|---|---|---|
| | T | C | T | C | T | C | T | C |
| **PS-matching / on-treatment TAR** | | | | | | | | |
| CCAE | 11,282 | 85,231 | 7.6 | 73.3 | 34 | 537 | 4.49 (3.11 - 6.28) | 7.33 (6.72 - 7.97) |
| ODOD | 7,516 | 62,401 | 4.4 | 56.7 | 20 | 957 | 4.53 (2.77 - 7.00) | 16.9 (15.8 - 18.0) |
| MDCR | 924 | 15,389 | 0.6 | 16.8 | <5 | 233 | < 7.85 (2.55 - 18.3) | 13.9 (12.2 - 15.8) |
| Open Claims | 138,137 | 863,757 | 98.3 | 933.6 | 196 | 6,745 | 1.99 (1.73 - 2.29) | 7.22 (7.05 - 7.40) |
| OEHR | 8,267 | 85,803 | 2.4 | 46.9 | 12 | 648 | 4.99 (2.58 - 8.72) | 13.8 (12.8 - 14.9) |
| VA | 1,293 | 31,887 | 0.9 | 23.9 | 0 | 253 | | 10.6 (9.33 - 12.0) |
| Meta-analysis | 166,126 | 1,112,581 | 113.3 | 1127.2 | <267 | 9,120 | < 2.36 (2.08 - 2.66) | 8.09 (7.93 - 8.26) |
| Meta-analysis (w/o USOC) | 27,989 | 248,824 | 15.0 | 193.7 | <71 | 2,375 | < 4.73 (3.69 - 5.96) | 12.3 (11.8 - 12.8) |
| **PS-stratification / on-treatment TAR** | | | | | | | | |
| CCAE | 12,757 | 89,567 | 8.3 | 76.7 | 37 | 570 | 4.45 (3.14 - 6.14) | 7.43 (6.83 - 8.06) |
| ODOD | 8,400 | 69,983 | 4.8 | 65.3 | 25 | 1,202 | 5.20 (3.36 - 7.67) | 18.4 (17.4 - 19.5) |
| MDCR | 998 | 20,199 | 0.7 | 21.6 | <5 | 356 | < 7.50 (2.44 - 17.5) | 16.5 (14.8 - 18.3) |
| Open Claims | 153,146 | 892,316 | 105.4 | 966.3 | 219 | 7,417 | 2.08 (1.81 - 2.37) | 7.68 (7.50 - 7.85) |
| OEHR | 8,985 | 95,219 | 2.6 | 52.5 | 15 | 763 | 5.85 (3.27 - 9.64) | 14.5 (13.5 - 15.6) |
| VA | 1,389 | 237,705 | 1.0 | 252.5 | <10 | 2,179 | < 10.4 (4.98 - 19.1) | 8.63 (8.27 - 9.00) |
| Meta-analysis | 183,288 | 1,147,085 | 121.1 | 1160.9 | 296 | 9,952 | 2.44 (2.17 - 2.74) | 8.57 (8.40 - 8.74) |
| Meta-analysis (w/o USOC) | 30,142 | 254,769 | 15.7 | 194.6 | 77 | 2,535 | 4.91 (3.87 - 6.14) | 13.0 (12.5 - 13.5) |
| **PS-matching / on-treatment TAR without escalation** | | | | | | | | |
| CCAE | 11,282 | 85,231 | 6.6 | 63.5 | 25 | 445 | 3.81 (2.47 - 5.62) | 7.01 (6.37 - 7.69) |
| ODOD | 7,516 | 62,401 | 3.9 | 50.4 | 14 | 825 | 3.57 (1.95 - 5.99) | 16.4 (15.3 - 17.5) |
| MDCR | 924 | 15,389 | 0.6 | 15.2 | 0 | 207 | | 13.6 (11.8 - 15.6) |
| Open Claims | 138,137 | 863,757 | 85.4 | 791.0 | 151 | 5,522 | 1.77 (1.50 - 2.07) | 6.98 (6.80 - 7.17) |
| OEHR | 8,267 | 85,803 | 2.1 | 41.0 | 10 | 486 | 4.82 (2.31 - 8.86) | 11.9 (10.8 - 13.0) |
| VA | 1,293 | 31,887 | 0.7 | 20.9 | 0 | 207 | | 9.88 (8.58 - 11.3) |
| Meta-analysis | 165,202 | 1,097,192 | 98.0 | 945.9 | 200 | 7,278 | 2.04 (1.77 - 2.34) | 7.69 (7.52 - 7.87) |
| Meta-analysis (w/o USOC) | 27,065 | 233,435 | 12.6 | 154.9 | 49 | 1,756 | 3.90 (2.89 - 5.16) | 11.3 (10.8 - 11.9) |
| **PS-stratification / on-treatment TAR without escalation** | | | | | | | | |
| CCAE | 12,757 | 89,567 | 7.3 | 66.6 | 28 | 475 | 3.86 (2.57 - 5.58) | 7.13 (6.50 - 7.80) |
| ODOD | 8,400 | 69,983 | 4.3 | 58.3 | 19 | 1,045 | 4.44 (2.67 - 6.94) | 17.9 (16.9 - 19.1) |
| MDCR | 998 | 20,199 | 0.6 | 19.6 | 0 | 322 | | 16.4 (14.7 - 18.3) |
| Open Claims | 153,146 | 892,316 | 92.1 | 819.2 | 173 | 6,105 | 1.88 (1.61 - 2.18) | 7.45 (7.27 - 7.64) |
| OEHR | 8,985 | 95,219 | 2.2 | 45.7 | 12 | 569 | 5.39 (2.78 - 9.41) | 12.4 (11.4 - 13.5) |
| VA | 1,389 | 237,705 | 0.8 | 225.1 | 0 | 1,785 | | 7.93 (7.57 - 8.31) |
| Meta-analysis | 183,288 | 1,147,085 | 105.9 | 989.9 | 232 | 8,194 | 2.19 (1.92 - 2.49) | 8.28 (8.10 - 8.46) |
| Meta-analysis (w/o USOC) | 30,142 | 254,769 | 13.8 | 170.7 | 59 | 2,089 | 4.29 (3.27 - 5.53) | 12.2 (11.7 - 12.8) |

##### 4.5.19 Hypotension

**Supplementary Table 412:** Exposures, outcomes and risk estimates for Hypotension in GLP1RA (male) (T) vs SU (male) (C) new-user cohorts under four different propensity score (PS) adjustment / time-at-risk (TAR) designs across data sources. We report total patient exposure time in 1,000 patient-year (KYr) units, uncalibrated and calibrated hazard ratios (HRs) and their 95% confidence intervals (CIs). Greyed out entries did not pass study diagnostics and are included only for completeness.

| | Patients | | Exposure (KYr) | | Outcomes | | Incidence (per KYr) | |
|---|---|---|---|---|---|---|---|---|
| | T | C | T | C | T | C | T | C |
| **PS-matching / on-treatment TAR** | | | | | | | | |
| CCAE | 11,253 | 84,856 | 7.5 | 73.0 | 66 | 510 | 8.79 (6.80 - 11.2) | 6.98 (6.39 - 7.62) |
| ODOD | 7,303 | 60,032 | 4.3 | 54.1 | 71 | 1,178 | 16.6 (13.0 - 21.0) | 21.8 (20.5 - 23.1) |
| MDCR | 894 | 14,745 | 0.6 | 16.0 | 16 | 333 | 26.0 (14.8 - 42.2) | 20.8 (18.6 - 23.1) |
| Open Claims | 135,689 | 844,420 | 96.5 | 910.9 | 575 | 7,991 | 5.96 (5.48 - 6.47) | 8.77 (8.58 - 8.97) |
| OEHR | 8,178 | 84,514 | 2.4 | 45.9 | 37 | 752 | 15.6 (11.0 - 21.5) | 16.4 (15.2 - 17.6) |
| VA | 1,221 | 30,932 | 0.9 | 23.1 | 17 | 305 | 19.1 (11.2 - 30.6) | 13.2 (11.7 - 14.7) |
| Meta-analysis | 163,317 | 1,088,567 | 111.2 | 1099.9 | 765 | 10,764 | 6.88 (6.40 - 7.38) | 9.79 (9.60 - 9.97) |
| Meta-analysis (w/o USOC) | 27,628 | 244,147 | 14.8 | 189.1 | 190 | 2,773 | 12.9 (11.1 - 14.8) | 14.7 (14.1 - 15.2) |
| **PS-stratification / on-treatment TAR** | | | | | | | | |
| CCAE | 12,737 | 89,064 | 8.3 | 76.4 | 75 | 555 | 9.08 (7.15 - 11.4) | 7.26 (6.67 - 7.89) |
| ODOD | 8,191 | 67,236 | 4.7 | 62.2 | 76 | 1,548 | 16.3 (12.8 - 20.4) | 24.9 (23.7 - 26.2) |
| MDCR | 967 | 19,153 | 0.6 | 20.4 | 17 | 515 | 26.4 (15.4 - 42.3) | 25.2 (23.1 - 27.5) |
| Open Claims | 150,746 | 871,592 | 103.6 | 942.1 | 624 | 8,952 | 6.02 (5.56 - 6.51) | 9.50 (9.31 - 9.70) |
| OEHR | 8,903 | 93,699 | 2.5 | 51.2 | 38 | 957 | 15.0 (10.6 - 20.6) | 18.7 (17.5 - 19.9) |
| VA | 1,317 | 227,052 | 0.9 | 238.7 | 18 | 3,318 | 19.6 (11.6 - 31.0) | 13.9 (13.4 - 14.4) |
| Meta-analysis | 180,577 | 1,121,591 | 119.1 | 1132.0 | 813 | 12,012 | 6.83 (6.36 - 7.31) | 10.6 (10.4 - 10.8) |
| Meta-analysis (w/o USOC) | 29,831 | 249,999 | 15.5 | 189.8 | 189 | 3,060 | 12.2 (10.5 - 14.1) | 16.1 (15.6 - 16.7) |
| **PS-matching / on-treatment TAR without escalation** | | | | | | | | |
| CCAE | 11,253 | 84,856 | 6.5 | 63.3 | 55 | 419 | 8.44 (6.36 - 11.0) | 6.62 (6.00 - 7.28) |
| ODOD | 7,303 | 60,032 | 3.8 | 48.1 | 60 | 1,057 | 15.8 (12.1 - 20.4) | 22.0 (20.7 - 23.3) |
| MDCR | 894 | 14,745 | 0.5 | 14.5 | 13 | 291 | 23.9 (12.7 - 40.9) | 20.1 (17.9 - 22.6) |
| Open Claims | 135,689 | 844,420 | 83.9 | 771.6 | 488 | 6,645 | 5.82 (5.31 - 6.36) | 8.61 (8.41 - 8.82) |
| OEHR | 8,178 | 84,514 | 2.0 | 40.1 | 36 | 519 | 17.6 (12.3 - 24.4) | 12.9 (11.8 - 14.1) |
| VA | 1,221 | 30,932 | 0.7 | 20.3 | 15 | 249 | 21.7 (12.1 - 35.7) | 12.3 (10.8 - 13.9) |
| Meta-analysis | 163,317 | 1,088,567 | 96.8 | 937.6 | 652 | 8,931 | 6.74 (6.23 - 7.28) | 9.53 (9.33 - 9.72) |
| Meta-analysis (w/o USOC) | 27,628 | 244,147 | 12.9 | 166.0 | 164 | 2,286 | 12.7 (10.8 - 14.8) | 13.8 (13.2 - 14.3) |
| **PS-stratification / on-treatment TAR without escalation** | | | | | | | | |
| CCAE | 12,737 | 89,064 | 7.2 | 66.4 | 61 | 461 | 8.47 (6.48 - 10.9) | 6.95 (6.33 - 7.61) |
| ODOD | 8,191 | 67,236 | 4.1 | 55.5 | 65 | 1,397 | 15.7 (12.1 - 20.0) | 25.2 (23.9 - 26.5) |
| MDCR | 967 | 19,153 | 0.6 | 18.6 | 14 | 452 | 24.6 (13.4 - 41.2) | 24.4 (22.2 - 26.7) |
| Open Claims | 150,746 | 871,592 | 90.6 | 798.5 | 534 | 7,508 | 5.90 (5.41 - 6.42) | 9.40 (9.19 - 9.62) |
| OEHR | 8,903 | 93,699 | 2.2 | 44.7 | 37 | 667 | 16.8 (11.8 - 23.2) | 14.9 (13.8 - 16.1) |
| VA | 1,317 | 227,052 | 0.7 | 213.1 | 15 | 2,734 | 20.9 (11.7 - 34.4) | 12.8 (12.4 - 13.3) |
| Meta-analysis | 180,577 | 1,121,591 | 104.1 | 965.1 | 697 | 10,033 | 6.69 (6.21 - 7.21) | 10.4 (10.2 - 10.6) |
| Meta-analysis (w/o USOC) | 29,831 | 249,999 | 13.6 | 166.6 | 163 | 2,525 | 12.0 (10.2 - 14.0) | 15.2 (14.6 - 15.8) |

##### 4.5.20 Joint pain

**Supplementary Table 413:** Exposures, outcomes and risk estimates for Joint pain in GLP1RA (male) (T) vs SU (male) (C) new-user cohorts under four different propensity score (PS) adjustment / time-at-risk (TAR) designs across data sources. We report total patient exposure time in 1,000 patient-year (KYr) units, uncalibrated and calibrated hazard ratios (HRs) and their 95% confidence intervals (CIs). Greyed out entries did not pass study diagnostics and are included only for completeness.

| | Patients | | Exposure (KYr) | | Outcomes | | Incidence (per KYr) | |
|---|---|---|---|---|---|---|---|---|
| | T | C | T | C | T | C | T | C |
| **PS-matching / on-treatment TAR** | | | | | | | | |
| CCAE | 10,943 | 83,101 | 7.3 | 71.2 | 61 | 578 | 8.37 (6.40 - 10.8) | 8.12 (7.47 - 8.81) |
| ODOD | 7,211 | 60,514 | 4.2 | 54.9 | 37 | 571 | 8.77 (6.18 - 12.1) | 10.4 (9.56 - 11.3) |
| MDCR | 896 | 14,874 | 0.6 | 16.2 | <5 | 140 | < 8.04 (2.61 - 18.8) | 8.62 (7.26 - 10.2) |
| Open Claims | 132,805 | 838,401 | 94.4 | 906.9 | 441 | 4,276 | 4.67 (4.25 - 5.13) | 4.71 (4.57 - 4.86) |
| OEHR | 7,966 | 83,548 | 2.3 | 45.6 | 14 | 382 | 6.06 (3.31 - 10.2) | 8.38 (7.56 - 9.26) |
| VA | 1,157 | 28,186 | 0.8 | 20.9 | 11 | 137 | 13.1 (6.55 - 23.5) | 6.54 (5.49 - 7.73) |
| Meta-analysis | 159,821 | 1,080,438 | 108.9 | 1094.8 | <558 | 5,947 | < 5.13 (4.71 - 5.57) | 5.43 (5.29 - 5.57) |
| Meta-analysis (w/o USOC) | 27,016 | 242,037 | 14.4 | 187.9 | <117 | 1,671 | < 8.10 (6.70 - 9.71) | 8.89 (8.47 - 9.33) |
| **PS-stratification / on-treatment TAR** | | | | | | | | |
| CCAE | 12,378 | 87,314 | 8.0 | 74.5 | 64 | 600 | 7.99 (6.16 - 10.2) | 8.05 (7.42 - 8.73) |
| ODOD | 8,069 | 67,870 | 4.6 | 63.3 | 44 | 646 | 9.57 (6.96 - 12.9) | 10.2 (9.43 - 11.0) |
| MDCR | 967 | 19,773 | 0.6 | 21.2 | <5 | 176 | < 7.71 (2.50 - 18.0) | 8.32 (7.13 - 9.64) |
| Open Claims | 147,146 | 867,272 | 101.3 | 940.0 | 492 | 4,493 | 4.86 (4.44 - 5.31) | 4.78 (4.64 - 4.92) |
| OEHR | 8,667 | 92,518 | 2.5 | 50.9 | 16 | 431 | 6.48 (3.70 - 10.5) | 8.47 (7.69 - 9.31) |
| VA | 1,248 | 212,284 | 0.9 | 224.9 | 12 | 1,767 | 13.8 (7.14 - 24.1) | 7.86 (7.49 - 8.23) |
| Meta-analysis | 176,260 | 1,114,974 | 116.4 | 1128.7 | 616 | 6,170 | 5.29 (4.88 - 5.73) | 5.47 (5.33 - 5.60) |
| Meta-analysis (w/o USOC) | 29,114 | 247,702 | 15.1 | 188.7 | 124 | 1,677 | 8.23 (6.84 - 9.81) | 8.89 (8.47 - 9.32) |
| **PS-matching / on-treatment TAR without escalation** | | | | | | | | |
| CCAE | 10,943 | 83,101 | 6.3 | 61.7 | 51 | 504 | 8.05 (5.99 - 10.6) | 8.17 (7.47 - 8.91) |
| ODOD | 7,211 | 60,514 | 3.8 | 48.9 | 33 | 506 | 8.80 (6.05 - 12.4) | 10.3 (9.47 - 11.3) |
| MDCR | 896 | 14,874 | 0.5 | 14.7 | <5 | 127 | < 9.11 (2.96 - 21.3) | 8.66 (7.22 - 10.3) |
| Open Claims | 132,805 | 838,401 | 82.1 | 768.3 | 375 | 3,616 | 4.57 (4.12 - 5.06) | 4.71 (4.55 - 4.86) |
| OEHR | 7,966 | 83,548 | 2.0 | 39.7 | 14 | 318 | 7.01 (3.83 - 11.8) | 8.00 (7.15 - 8.93) |
| VA | 1,157 | 28,186 | 0.7 | 18.4 | 11 | 123 | 16.6 (8.30 - 29.7) | 6.67 (5.55 - 7.96) |
| Meta-analysis | 159,821 | 1,080,438 | 94.7 | 933.3 | <478 | 5,071 | < 5.05 (4.61 - 5.52) | 5.43 (5.28 - 5.59) |
| Meta-analysis (w/o USOC) | 27,016 | 242,037 | 12.6 | 165.0 | <103 | 1,455 | < 8.15 (6.66 - 9.89) | 8.82 (8.37 - 9.28) |
| **PS-stratification / on-treatment TAR without escalation** | | | | | | | | |
| CCAE | 12,378 | 87,314 | 7.0 | 64.7 | 54 | 525 | 7.71 (5.80 - 10.1) | 8.11 (7.43 - 8.84) |
| ODOD | 8,069 | 67,870 | 4.1 | 56.5 | 39 | 576 | 9.53 (6.78 - 13.0) | 10.2 (9.38 - 11.1) |
| MDCR | 967 | 19,773 | 0.6 | 19.3 | <5 | 161 | < 8.72 (2.83 - 20.3) | 8.36 (7.12 - 9.76) |
| Open Claims | 147,146 | 867,272 | 88.5 | 796.8 | 423 | 3,799 | 4.78 (4.34 - 5.26) | 4.77 (4.62 - 4.92) |
| OEHR | 8,667 | 92,518 | 2.1 | 44.3 | 16 | 354 | 7.45 (4.26 - 12.1) | 7.99 (7.18 - 8.87) |
| VA | 1,248 | 212,284 | 0.7 | 200.9 | 11 | 1,590 | 16.0 (7.99 - 28.6) | 7.92 (7.53 - 8.31) |
| Meta-analysis | 176,260 | 1,114,974 | 101.7 | 962.3 | 532 | 5,254 | 5.23 (4.79 - 5.69) | 5.46 (5.31 - 5.61) |
| Meta-analysis (w/o USOC) | 29,114 | 247,702 | 13.2 | 165.5 | 109 | 1,455 | 8.23 (6.76 - 9.93) | 8.79 (8.34 - 9.25) |

##### 4.5.21 Lower extremity amputation

**Supplementary Table 414:** Exposures, outcomes and risk estimates for Lower extremity amputation in GLP1RA (male) (T) vs SU (male) (C) new-user cohorts under four different propensity score (PS) adjustment / time-at-risk (TAR) designs across data sources. We report total patient exposure time in 1,000 patient-year (KYr) units, uncalibrated and calibrated hazard ratios (HRs) and their 95% confidence intervals (CIs). Greyed out entries did not pass study diagnostics and are included only for completeness.

| | Patients | | Exposure (KYr) | | Outcomes | | Incidence (per KYr) | |
|---|---|---|---|---|---|---|---|---|
| | T | C | T | C | T | C | T | C |
| **PS-matching / on-treatment TAR** | | | | | | | | |
| CCAE | 11,450 | 86,132 | 7.7 | 74.3 | 7 | 72 | 0.91 (0.37 - 1.88) | 0.97 (0.76 - 1.22) |
| ODOD | 7,586 | 62,895 | 4.5 | 57.5 | 6 | 88 | 1.35 (0.49 - 2.93) | 1.53 (1.23 - 1.88) |
| MDCR | 931 | 15,477 | 0.6 | 17.0 | 0 | 23 | | 1.36 (0.86 - 2.04) |
| Open Claims | 139,861 | 871,656 | 99.6 | 945.1 | 49 | 762 | 0.49 (0.36 - 0.65) | 0.81 (0.75 - 0.87) |
| OEHR | 8,358 | 86,380 | 2.4 | 47.6 | 0 | 68 | | 1.43 (1.11 - 1.81) |
| VA | 1,306 | 32,239 | 0.9 | 24.2 | 0 | 23 | | 0.95 (0.60 - 1.42) |
| Meta-analysis | 158,897 | 1,020,683 | 111.7 | 1076.9 | 62 | 922 | 0.56 (0.43 - 0.71) | 0.86 (0.80 - 0.91) |
| Meta-analysis (w/o USOC) | 19,036 | 149,027 | 12.1 | 131.8 | 13 | 160 | 1.07 (0.57 - 1.83) | 1.21 (1.03 - 1.42) |
| **PS-stratification / on-treatment TAR** | | | | | | | | |
| CCAE | 12,974 | 90,464 | 8.5 | 77.8 | 7 | 80 | 0.83 (0.33 - 1.71) | 1.03 (0.82 - 1.28) |
| ODOD | 8,500 | 70,613 | 4.9 | 66.4 | 6 | 102 | 1.23 (0.45 - 2.68) | 1.54 (1.25 - 1.86) |
| MDCR | 1,007 | 20,312 | 0.7 | 21.8 | 0 | 31 | | 1.42 (0.97 - 2.02) |
| Open Claims | 155,300 | 901,023 | 107.0 | 979.0 | 49 | 832 | 0.46 (0.34 - 0.61) | 0.85 (0.79 - 0.91) |
| OEHR | 9,096 | 95,815 | 2.6 | 53.3 | 0 | 83 | | 1.56 (1.24 - 1.93) |
| VA | 1,404 | 239,038 | 1.0 | 255.1 | 0 | 204 | | 0.80 (0.69 - 0.92) |
| Meta-analysis | 176,774 | 1,062,100 | 120.3 | 1123.2 | 62 | 1,014 | 0.52 (0.40 - 0.66) | 0.90 (0.85 - 0.96) |
| Meta-analysis (w/o USOC) | 21,474 | 161,077 | 13.3 | 144.2 | 13 | 182 | 0.98 (0.52 - 1.67) | 1.26 (1.09 - 1.46) |
| **PS-matching / on-treatment TAR without escalation** | | | | | | | | |
| CCAE | 11,450 | 86,132 | 6.7 | 64.3 | 6 | 57 | 0.90 (0.33 - 1.96) | 0.89 (0.67 - 1.15) |
| ODOD | 7,586 | 62,895 | 4.0 | 51.1 | 5 | 73 | 1.26 (0.41 - 2.94) | 1.43 (1.12 - 1.80) |
| MDCR | 931 | 15,477 | 0.6 | 15.3 | 0 | 19 | | 1.24 (0.75 - 1.94) |
| Open Claims | 139,861 | 871,656 | 86.5 | 799.9 | 39 | 612 | 0.45 (0.32 - 0.62) | 0.77 (0.71 - 0.83) |
| OEHR | 8,358 | 86,380 | 2.1 | 41.4 | 0 | 35 | | 0.85 (0.59 - 1.18) |
| VA | 1,306 | 32,239 | 0.7 | 21.3 | 0 | 16 | | 0.75 (0.43 - 1.22) |
| Meta-analysis | 158,897 | 1,020,683 | 97.1 | 915.4 | 50 | 742 | 0.51 (0.38 - 0.68) | 0.81 (0.75 - 0.87) |
| Meta-analysis (w/o USOC) | 19,036 | 149,027 | 10.6 | 115.5 | 11 | 130 | 1.04 (0.52 - 1.85) | 1.13 (0.94 - 1.34) |
| **PS-stratification / on-treatment TAR without escalation** | | | | | | | | |
| CCAE | 12,974 | 90,464 | 7.4 | 67.5 | 6 | 64 | 0.81 (0.30 - 1.77) | 0.95 (0.73 - 1.21) |
| ODOD | 8,500 | 70,613 | 4.3 | 59.2 | 5 | 86 | 1.16 (0.38 - 2.70) | 1.45 (1.16 - 1.79) |
| MDCR | 1,007 | 20,312 | 0.6 | 19.8 | 0 | 26 | | 1.31 (0.86 - 1.92) |
| Open Claims | 155,300 | 901,023 | 93.4 | 829.1 | 39 | 670 | 0.42 (0.30 - 0.57) | 0.81 (0.75 - 0.87) |
| OEHR | 9,096 | 95,815 | 2.3 | 46.3 | 0 | 44 | | 0.95 (0.69 - 1.28) |
| VA | 1,404 | 239,038 | 0.8 | 227.1 | 0 | 153 | | 0.67 (0.57 - 0.79) |
| Meta-analysis | 176,774 | 1,062,100 | 105.1 | 955.7 | 50 | 820 | 0.48 (0.35 - 0.63) | 0.86 (0.80 - 0.92) |
| Meta-analysis (w/o USOC) | 21,474 | 161,077 | 11.7 | 126.7 | 11 | 150 | 0.94 (0.47 - 1.68) | 1.18 (1.00 - 1.39) |

##### 4.5.22 Nausea

**Supplementary Table 415:** Exposures, outcomes and risk estimates for Nausea in GLP1RA (male) (T) vs SU (male) (C) new-user cohorts under four different propensity score (PS) adjustment / time-at-risk (TAR) designs across data sources. We report total patient exposure time in 1,000 patient-year (KYr) units, uncalibrated and calibrated hazard ratios (HRs) and their 95% confidence intervals (CIs). Greyed out entries did not pass study diagnostics and are included only for completeness.

| | Patients | | Exposure (KYr) | | Outcomes | | Incidence (per KYr) | |
|---|---|---|---|---|---|---|---|---|
| | T | C | T | C | T | C | T | C |
| **PS-matching / on-treatment TAR** | | | | | | | | |
| CCAE | 10,329 | 79,722 | 6.9 | 68.4 | 223 | 1,359 | 32.3 (28.2 - 36.9) | 19.9 (18.8 - 21.0) |
| ODOD | 6,728 | 56,420 | 3.9 | 50.6 | 178 | 1,601 | 46.1 (39.6 - 53.4) | 31.6 (30.1 - 33.2) |
| MDCR | 859 | 14,193 | 0.6 | 15.4 | 14 | 411 | 23.8 (13.0 - 40.0) | 26.7 (24.2 - 29.4) |
| Open Claims | 123,951 | 793,055 | 88.2 | 857.8 | 1,820 | 12,893 | 20.6 (19.7 - 21.6) | 15.0 (14.8 - 15.3) |
| OEHR | 7,668 | 80,799 | 2.2 | 43.4 | 118 | 1,161 | 53.7 (44.4 - 64.3) | 26.7 (25.2 - 28.3) |
| VA | 1,224 | 30,316 | 0.9 | 22.8 | 22 | 215 | 25.5 (16.0 - 38.6) | 9.45 (8.23 - 10.8) |
| Meta-analysis | 149,535 | 1,024,189 | 101.7 | 1035.6 | 2,353 | 17,425 | 23.1 (22.2 - 24.1) | 16.8 (16.6 - 17.1) |
| Meta-analysis (w/o USOC) | 25,584 | 231,134 | 13.5 | 177.8 | 533 | 4,532 | 39.3 (36.1 - 42.8) | 25.5 (24.8 - 26.2) |
| **PS-stratification / on-treatment TAR** | | | | | | | | |
| CCAE | 11,671 | 83,812 | 7.5 | 71.6 | 263 | 1,451 | 34.9 (30.8 - 39.4) | 20.3 (19.2 - 21.3) |
| ODOD | 7,529 | 63,655 | 4.2 | 58.8 | 197 | 1,934 | 46.7 (40.4 - 53.7) | 32.9 (31.4 - 34.4) |
| MDCR | 924 | 18,733 | 0.6 | 20.0 | 14 | 565 | 22.8 (12.5 - 38.3) | 28.3 (26.0 - 30.8) |
| Open Claims | 137,078 | 820,444 | 94.4 | 889.4 | 2,003 | 13,802 | 21.2 (20.3 - 22.2) | 15.5 (15.3 - 15.8) |
| OEHR | 8,322 | 89,549 | 2.3 | 48.5 | 125 | 1,328 | 53.3 (44.4 - 63.6) | 27.4 (25.9 - 28.9) |
| VA | 1,317 | 226,602 | 0.9 | 240.4 | 22 | 2,291 | 24.7 (15.5 - 37.3) | 9.53 (9.14 - 9.93) |
| Meta-analysis | 164,600 | 1,057,460 | 108.5 | 1068.4 | 2,588 | 18,515 | 23.9 (22.9 - 24.8) | 17.3 (17.1 - 17.6) |
| Meta-analysis (w/o USOC) | 27,522 | 237,016 | 14.1 | 178.9 | 585 | 4,713 | 41.5 (38.2 - 45.0) | 26.3 (25.6 - 27.1) |
| **PS-matching / on-treatment TAR without escalation** | | | | | | | | |
| CCAE | 10,329 | 79,722 | 6.0 | 59.4 | 199 | 1,170 | 33.2 (28.8 - 38.2) | 19.7 (18.6 - 20.8) |
| ODOD | 6,728 | 56,420 | 3.4 | 45.2 | 161 | 1,406 | 46.7 (39.8 - 54.5) | 31.1 (29.5 - 32.8) |
| MDCR | 859 | 14,193 | 0.5 | 14.0 | 10 | 364 | 19.2 (9.19 - 35.2) | 26.1 (23.5 - 28.9) |
| Open Claims | 123,951 | 793,055 | 76.8 | 729.0 | 1,588 | 10,636 | 20.7 (19.7 - 21.7) | 14.6 (14.3 - 14.9) |
| OEHR | 7,668 | 80,799 | 1.9 | 38.1 | 100 | 893 | 52.3 (42.5 - 63.6) | 23.4 (21.9 - 25.0) |
| VA | 1,224 | 30,316 | 0.7 | 20.0 | 19 | 169 | 27.7 (16.7 - 43.2) | 8.47 (7.24 - 9.84) |
| Meta-analysis | 149,535 | 1,024,189 | 88.7 | 885.6 | 2,058 | 14,469 | 23.2 (22.2 - 24.2) | 16.3 (16.1 - 16.6) |
| Meta-analysis (w/o USOC) | 25,584 | 231,134 | 11.9 | 156.7 | 470 | 3,833 | 39.6 (36.1 - 43.3) | 24.5 (23.7 - 25.3) |
| **PS-stratification / on-treatment TAR without escalation** | | | | | | | | |
| CCAE | 11,671 | 83,812 | 6.6 | 62.4 | 235 | 1,251 | 35.7 (31.3 - 40.6) | 20.0 (18.9 - 21.2) |
| ODOD | 7,529 | 63,655 | 3.8 | 52.6 | 179 | 1,706 | 47.5 (40.8 - 54.9) | 32.4 (30.9 - 34.0) |
| MDCR | 924 | 18,733 | 0.5 | 18.2 | 10 | 505 | 18.3 (8.78 - 33.7) | 27.8 (25.4 - 30.3) |
| Open Claims | 137,078 | 820,444 | 82.6 | 756.3 | 1,764 | 11,431 | 21.3 (20.4 - 22.4) | 15.1 (14.8 - 15.4) |
| OEHR | 8,322 | 89,549 | 2.0 | 42.5 | 107 | 1,021 | 52.2 (42.8 - 63.1) | 24.0 (22.6 - 25.5) |
| VA | 1,317 | 226,602 | 0.7 | 214.6 | 19 | 1,925 | 26.7 (16.1 - 41.7) | 8.97 (8.57 - 9.38) |
| Meta-analysis | 164,600 | 1,057,460 | 95.0 | 913.8 | 2,285 | 15,409 | 24.0 (23.1 - 25.1) | 16.9 (16.6 - 17.1) |
| Meta-analysis (w/o USOC) | 27,522 | 237,016 | 12.4 | 157.5 | 521 | 3,978 | 42.0 (38.5 - 45.8) | 25.2 (24.5 - 26.0) |

##### 4.5.23 Peripheral edema

**Supplementary Table 416:** Exposures, outcomes and risk estimates for Peripheral edema in GLP1RA (male) (T) vs SU (male) (C) new-user cohorts under four different propensity score (PS) adjustment / time-at-risk (TAR) designs across data sources. We report total patient exposure time in 1,000 patient-year (KYr) units, uncalibrated and calibrated hazard ratios (HRs) and their 95% confidence intervals (CIs). Greyed out entries did not pass study diagnostics and are included only for completeness.

| | Patients | | Exposure (KYr) | | Outcomes | | Incidence (per KYr) | |
|---|---|---|---|---|---|---|---|---|
| | T | C | T | C | T | C | T | C |
| **PS-matching / on-treatment TAR** | | | | | | | | |
| CCAE | 9,596 | 75,118 | 6.3 | 62.5 | 215 | 2,537 | 34.2 (29.8 - 39.1) | 40.6 (39.0 - 42.2) |
| ODOD | 6,069 | 50,874 | 3.5 | 43.4 | 168 | 3,085 | 47.9 (41.0 - 55.8) | 71.0 (68.5 - 73.6) |
| MDCR | 718 | 12,217 | 0.5 | 12.5 | 30 | 958 | 63.7 (43.0 - 91.0) | 76.4 (71.6 - 81.4) |
| Open Claims | 110,824 | 717,174 | 78.4 | 752.6 | 1,562 | 24,428 | 19.9 (18.9 - 20.9) | 32.5 (32.1 - 32.9) |
| OEHR | 7,080 | 75,134 | 2.0 | 39.0 | 76 | 2,006 | 38.1 (30.0 - 47.7) | 51.4 (49.2 - 53.7) |
| VA | 969 | 25,382 | 0.7 | 18.3 | 16 | 797 | 22.8 (13.1 - 37.1) | 43.5 (40.6 - 46.7) |
| Meta-analysis | 134,287 | 930,517 | 90.7 | 910.1 | 2,051 | 33,014 | 22.6 (21.7 - 23.6) | 36.3 (35.9 - 36.7) |
| Meta-analysis (w/o USOC) | 23,463 | 213,343 | 12.2 | 157.5 | 489 | 8,586 | 39.9 (36.5 - 43.6) | 54.5 (53.4 - 55.7) |
| **PS-stratification / on-treatment TAR** | | | | | | | | |
| CCAE | 10,734 | 78,888 | 6.8 | 65.5 | 244 | 2,707 | 35.7 (31.3 - 40.4) | 41.4 (39.8 - 42.9) |
| ODOD | 6,702 | 57,013 | 3.8 | 49.9 | 185 | 3,814 | 49.1 (42.3 - 56.7) | 76.4 (74.0 - 78.9) |
| MDCR | 772 | 15,931 | 0.5 | 16.0 | 32 | 1,343 | 65.1 (44.5 - 91.9) | 83.9 (79.5 - 88.5) |
| Open Claims | 122,153 | 741,055 | 83.8 | 779.2 | 1,666 | 26,510 | 19.9 (18.9 - 20.9) | 34.0 (33.6 - 34.4) |
| OEHR | 7,667 | 83,664 | 2.1 | 43.6 | 79 | 2,420 | 37.0 (29.3 - 46.2) | 55.6 (53.4 - 57.8) |
| VA | 1,030 | 195,946 | 0.7 | 198.7 | 16 | 7,866 | 22.2 (12.7 - 36.0) | 39.6 (38.7 - 40.5) |
| Meta-analysis | 147,256 | 960,620 | 96.5 | 938.1 | 2,174 | 35,451 | 22.5 (21.6 - 23.5) | 37.8 (37.4 - 38.2) |
| Meta-analysis (w/o USOC) | 25,103 | 219,565 | 12.7 | 158.9 | 508 | 8,941 | 39.9 (36.5 - 43.5) | 56.3 (55.1 - 57.4) |
| **PS-matching / on-treatment TAR without escalation** | | | | | | | | |
| CCAE | 9,596 | 75,118 | 5.5 | 54.5 | 179 | 2,212 | 32.5 (27.9 - 37.6) | 40.6 (38.9 - 42.3) |
| ODOD | 6,069 | 50,874 | 3.1 | 39.0 | 153 | 2,721 | 48.9 (41.5 - 57.3) | 69.8 (67.2 - 72.5) |
| MDCR | 718 | 12,217 | 0.4 | 11.4 | 26 | 864 | 62.8 (41.0 - 92.0) | 75.6 (70.6 - 80.8) |
| Open Claims | 110,824 | 717,174 | 68.3 | 641.4 | 1,346 | 20,754 | 19.7 (18.7 - 20.8) | 32.4 (31.9 - 32.8) |
| OEHR | 7,080 | 75,134 | 1.7 | 34.4 | 62 | 1,624 | 36.0 (27.6 - 46.1) | 47.2 (44.9 - 49.5) |
| VA | 969 | 25,382 | 0.5 | 16.2 | 12 | 697 | 21.8 (11.3 - 38.1) | 43.1 (40.0 - 46.4) |
| Meta-analysis | 134,287 | 930,517 | 79.1 | 780.8 | 1,766 | 28,175 | 22.3 (21.3 - 23.4) | 36.1 (35.7 - 36.5) |
| Meta-analysis (w/o USOC) | 23,463 | 213,343 | 10.8 | 139.4 | 420 | 7,421 | 39.0 (35.4 - 42.9) | 53.2 (52.0 - 54.5) |
| **PS-stratification / on-treatment TAR without escalation** | | | | | | | | |
| CCAE | 10,734 | 78,888 | 6.0 | 57.2 | 207 | 2,366 | 34.4 (29.8 - 39.4) | 41.3 (39.7 - 43.0) |
| ODOD | 6,702 | 57,013 | 3.4 | 44.9 | 168 | 3,392 | 49.9 (42.7 - 58.1) | 75.5 (73.0 - 78.1) |
| MDCR | 772 | 15,931 | 0.4 | 14.7 | 28 | 1,224 | 64.6 (42.9 - 93.4) | 83.5 (78.9 - 88.3) |
| Open Claims | 122,153 | 741,055 | 73.3 | 664.6 | 1,445 | 22,577 | 19.7 (18.7 - 20.8) | 34.0 (33.5 - 34.4) |
| OEHR | 7,667 | 83,664 | 1.9 | 38.4 | 65 | 1,940 | 35.0 (27.0 - 44.7) | 50.5 (48.3 - 52.8) |
| VA | 1,030 | 195,946 | 0.6 | 178.7 | 12 | 6,851 | 21.2 (10.9 - 37.0) | 38.3 (37.4 - 39.3) |
| Meta-analysis | 147,256 | 960,620 | 84.6 | 805.1 | 1,885 | 30,275 | 22.3 (21.3 - 23.3) | 37.6 (37.2 - 38.0) |
| Meta-analysis (w/o USOC) | 25,103 | 219,565 | 11.2 | 140.6 | 440 | 7,698 | 39.1 (35.6 - 43.0) | 54.8 (53.5 - 56.0) |

##### 4.5.24 Photosensitivity

**Supplementary Table 417:** Exposures, outcomes and risk estimates for Photosensitivity in GLP1RA (male) (T) vs SU (male) (C) new-user cohorts under four different propensity score (PS) adjustment / time-at-risk (TAR) designs across data sources. We report total patient exposure time in 1,000 patient-year (KYr) units, uncalibrated and calibrated hazard ratios (HRs) and their 95% confidence intervals (CIs). Greyed out entries did not pass study diagnostics and are included only for completeness.

| | Patients | | Exposure (KYr) | | Outcomes | | Incidence (per KYr) | |
|---|---|---|---|---|---|---|---|---|
| | T | C | T | C | T | C | T | C |
| **PS-matching / on-treatment TAR** | | | | | | | | |
| CCAE | 11,381 | 85,596 | 7.6 | 73.8 | 11 | 101 | 1.44 (0.72 - 2.58) | 1.37 (1.11 - 1.66) |
| ODOD | 7,555 | 62,423 | 4.4 | 56.9 | <5 | 96 | < 1.13 (0.37 - 2.63) | 1.69 (1.37 - 2.06) |
| MDCR | 915 | 15,256 | 0.6 | 16.7 | <5 | 57 | < 7.88 (2.56 - 18.4) | 3.42 (2.59 - 4.43) |
| Open Claims | 138,800 | 866,440 | 98.7 | 937.8 | 29 | 434 | 0.29 (0.20 - 0.42) | 0.46 (0.42 - 0.51) |
| OEHR | 8,315 | 85,941 | 2.4 | 47.3 | 5 | 86 | 2.07 (0.67 - 4.83) | 1.82 (1.45 - 2.24) |
| VA | 1,290 | 31,833 | 0.9 | 23.9 | 0 | 13 | | 0.54 (0.29 - 0.93) |
| Meta-analysis | 166,051 | 1,100,400 | 113.1 | 1115.8 | <50 | 717 | < 0.44 (0.33 - 0.58) | 0.64 (0.60 - 0.69) |
| Meta-analysis (w/o USOC) | 27,251 | 233,960 | 14.5 | 178.0 | <21 | 283 | < 1.45 (0.90 - 2.22) | 1.59 (1.41 - 1.79) |
| **PS-stratification / on-treatment TAR** | | | | | | | | |
| CCAE | 12,891 | 89,977 | 8.4 | 77.3 | 11 | 104 | 1.31 (0.65 - 2.35) | 1.35 (1.10 - 1.63) |
| ODOD | 8,466 | 70,086 | 4.8 | 65.7 | <5 | 120 | < 1.03 (0.34 - 2.41) | 1.83 (1.52 - 2.19) |
| MDCR | 991 | 19,987 | 0.7 | 21.4 | <5 | 83 | < 7.52 (2.44 - 17.6) | 3.88 (3.09 - 4.81) |
| Open Claims | 154,117 | 895,273 | 106.0 | 970.9 | 32 | 484 | 0.30 (0.21 - 0.43) | 0.50 (0.46 - 0.54) |
| OEHR | 9,044 | 95,253 | 2.6 | 52.9 | 5 | 110 | 1.94 (0.63 - 4.52) | 2.08 (1.71 - 2.51) |
| VA | 1,390 | 236,875 | 1.0 | 252.2 | 0 | 376 | | 1.49 (1.34 - 1.65) |
| Meta-analysis | 184,518 | 1,150,589 | 121.8 | 1166.7 | <53 | 818 | < 0.44 (0.33 - 0.57) | 0.70 (0.65 - 0.75) |
| Meta-analysis (w/o USOC) | 30,401 | 255,316 | 15.8 | 195.8 | <21 | 334 | < 1.33 (0.82 - 2.03) | 1.71 (1.53 - 1.90) |
| **PS-matching / on-treatment TAR without escalation** | | | | | | | | |
| CCAE | 11,381 | 85,596 | 6.6 | 63.9 | 10 | 92 | 1.51 (0.72 - 2.78) | 1.44 (1.16 - 1.77) |
| ODOD | 7,555 | 62,423 | 3.9 | 50.6 | <5 | 90 | < 1.27 (0.41 - 2.97) | 1.78 (1.43 - 2.19) |
| MDCR | 915 | 15,256 | 0.6 | 15.1 | <5 | 51 | < 8.93 (2.90 - 20.8) | 3.38 (2.52 - 4.45) |
| Open Claims | 138,800 | 866,440 | 85.7 | 793.6 | 27 | 393 | 0.31 (0.21 - 0.46) | 0.50 (0.45 - 0.55) |
| OEHR | 8,315 | 85,941 | 2.1 | 41.2 | <5 | 76 | < 2.40 (0.78 - 5.60) | 1.85 (1.45 - 2.31) |
| VA | 1,290 | 31,833 | 0.7 | 21.0 | 0 | <10 | | < 0.48 (0.23 - 0.88) |
| Meta-analysis | 166,051 | 1,100,400 | 98.4 | 949.2 | <47 | 651 | < 0.48 (0.35 - 0.64) | 0.69 (0.63 - 0.74) |
| Meta-analysis (w/o USOC) | 27,251 | 233,960 | 12.6 | 155.6 | <20 | 258 | < 1.58 (0.97 - 2.44) | 1.66 (1.46 - 1.87) |
| **PS-stratification / on-treatment TAR without escalation** | | | | | | | | |
| CCAE | 12,891 | 89,977 | 7.3 | 67.0 | 10 | 95 | 1.37 (0.65 - 2.51) | 1.42 (1.15 - 1.73) |
| ODOD | 8,466 | 70,086 | 4.3 | 58.5 | <5 | 112 | < 1.16 (0.38 - 2.72) | 1.91 (1.58 - 2.30) |
| MDCR | 991 | 19,987 | 0.6 | 19.5 | <5 | 77 | < 8.51 (2.76 - 19.9) | 3.96 (3.12 - 4.94) |
| Open Claims | 154,117 | 895,273 | 92.6 | 822.1 | 30 | 443 | 0.32 (0.22 - 0.46) | 0.54 (0.49 - 0.59) |
| OEHR | 9,044 | 95,253 | 2.2 | 45.9 | <5 | 98 | < 2.23 (0.73 - 5.21) | 2.14 (1.73 - 2.60) |
| VA | 1,390 | 236,875 | 0.8 | 224.6 | 0 | 344 | | 1.53 (1.37 - 1.70) |
| Meta-analysis | 184,518 | 1,150,589 | 106.4 | 993.5 | <50 | 748 | < 0.47 (0.35 - 0.62) | 0.75 (0.70 - 0.81) |
| Meta-analysis (w/o USOC) | 30,401 | 255,316 | 13.9 | 171.5 | <20 | 305 | < 1.44 (0.88 - 2.23) | 1.78 (1.58 - 1.99) |

##### 4.5.25 Venous thromboembolism

**Supplementary Table 418:** Exposures, outcomes and risk estimates for Venous thromboembolism in GLP1RA (male) (T) vs SU (male) (C) new-user cohorts under four different propensity score (PS) adjustment / time-at-risk (TAR) designs across data sources. We report total patient exposure time in 1,000 patient-year (KYr) units, uncalibrated and calibrated hazard ratios (HRs) and their 95% confidence intervals (CIs). Greyed out entries did not pass study diagnostics and are included only for completeness.

| | Patients | | Exposure (KYr) | | Outcomes | | Incidence (per KYr) | |
|---|---|---|---|---|---|---|---|---|
| | T | C | T | C | T | C | T | C |
| **PS-matching / on-treatment TAR** | | | | | | | | |
| CCAE | 11,172 | 84,495 | 7.5 | 72.6 | 38 | 408 | 5.10 (3.61 - 7.00) | 5.62 (5.08 - 6.19) |
| ODOD | 7,358 | 61,099 | 4.3 | 55.4 | 26 | 611 | 6.02 (3.93 - 8.82) | 11.0 (10.2 - 11.9) |
| MDCR | 892 | 14,897 | 0.6 | 16.2 | 9 | 226 | 14.6 (6.67 - 27.7) | 13.9 (12.2 - 15.9) |
| Open Claims | 134,797 | 845,418 | 95.9 | 912.8 | 348 | 5,254 | 3.63 (3.26 - 4.03) | 5.76 (5.60 - 5.91) |
| OEHR | 8,157 | 84,240 | 2.4 | 45.9 | 19 | 421 | 8.01 (4.82 - 12.5) | 9.17 (8.31 - 10.1) |
| VA | 1,255 | 31,500 | 0.9 | 23.6 | <10 | 142 | < 11.0 (5.28 - 20.2) | 6.01 (5.06 - 7.09) |
| Meta-analysis | 162,376 | 1,090,149 | 110.7 | 1103.0 | 440 | 6,920 | 3.97 (3.61 - 4.36) | 6.27 (6.13 - 6.42) |
| Meta-analysis (w/o USOC) | 27,579 | 244,731 | 14.8 | 190.2 | 92 | 1,666 | 6.23 (5.02 - 7.64) | 8.76 (8.34 - 9.19) |
| **PS-stratification / on-treatment TAR** | | | | | | | | |
| CCAE | 12,632 | 88,756 | 8.2 | 76.1 | 43 | 443 | 5.25 (3.80 - 7.07) | 5.82 (5.30 - 6.39) |
| ODOD | 8,239 | 68,583 | 4.7 | 63.9 | 27 | 787 | 5.72 (3.77 - 8.33) | 12.3 (11.5 - 13.2) |
| MDCR | 962 | 19,507 | 0.6 | 20.8 | 9 | 312 | 14.0 (6.41 - 26.6) | 15.0 (13.4 - 16.7) |
| Open Claims | 149,623 | 873,663 | 103.1 | 945.2 | 370 | 5,765 | 3.59 (3.23 - 3.98) | 6.10 (5.94 - 6.26) |
| OEHR | 8,858 | 93,438 | 2.5 | 51.4 | 19 | 501 | 7.50 (4.52 - 11.7) | 9.75 (8.92 - 10.6) |
| VA | 1,345 | 233,342 | 0.9 | 247.7 | <10 | 1,323 | < 10.7 (5.12 - 19.6) | 5.34 (5.06 - 5.64) |
| Meta-analysis | 179,352 | 1,124,440 | 118.5 | 1136.5 | 459 | 7,496 | 3.87 (3.53 - 4.24) | 6.60 (6.45 - 6.75) |
| Meta-analysis (w/o USOC) | 29,729 | 250,777 | 15.4 | 191.3 | 89 | 1,731 | 5.76 (4.63 - 7.09) | 9.05 (8.63 - 9.49) |
| **PS-matching / on-treatment TAR without escalation** | | | | | | | | |
| CCAE | 11,172 | 84,495 | 6.5 | 63.0 | 35 | 340 | 5.40 (3.76 - 7.51) | 5.40 (4.84 - 6.00) |
| ODOD | 7,358 | 61,099 | 3.8 | 49.3 | 23 | 529 | 5.98 (3.79 - 8.98) | 10.7 (9.84 - 11.7) |
| MDCR | 892 | 14,897 | 0.5 | 14.7 | 5 | 200 | 9.15 (2.97 - 21.4) | 13.6 (11.8 - 15.7) |
| Open Claims | 134,797 | 845,418 | 83.4 | 773.2 | 305 | 4,468 | 3.66 (3.26 - 4.09) | 5.78 (5.61 - 5.95) |
| OEHR | 8,157 | 84,240 | 2.1 | 40.1 | 15 | 316 | 7.31 (4.09 - 12.1) | 7.89 (7.04 - 8.81) |
| VA | 1,255 | 31,500 | 0.7 | 20.7 | <10 | 114 | < 13.9 (6.68 - 25.6) | 5.50 (4.54 - 6.61) |
| Meta-analysis | 162,376 | 1,090,149 | 96.3 | 940.2 | 383 | 5,853 | 3.98 (3.59 - 4.40) | 6.23 (6.07 - 6.39) |
| Meta-analysis (w/o USOC) | 27,579 | 244,731 | 12.9 | 167.0 | 78 | 1,385 | 6.04 (4.77 - 7.54) | 8.29 (7.86 - 8.74) |
| **PS-stratification / on-treatment TAR without escalation** | | | | | | | | |
| CCAE | 12,632 | 88,756 | 7.2 | 66.1 | 40 | 370 | 5.59 (3.99 - 7.61) | 5.60 (5.04 - 6.20) |
| ODOD | 8,239 | 68,583 | 4.2 | 57.0 | 24 | 691 | 5.71 (3.66 - 8.50) | 12.1 (11.2 - 13.1) |
| MDCR | 962 | 19,507 | 0.6 | 18.9 | 5 | 282 | 8.78 (2.85 - 20.5) | 14.9 (13.2 - 16.7) |
| Open Claims | 149,623 | 873,663 | 90.0 | 801.0 | 324 | 4,927 | 3.60 (3.22 - 4.01) | 6.15 (5.98 - 6.33) |
| OEHR | 8,858 | 93,438 | 2.2 | 44.7 | 15 | 370 | 6.81 (3.81 - 11.2) | 8.27 (7.45 - 9.16) |
| VA | 1,345 | 233,342 | 0.7 | 220.8 | <10 | 1,087 | < 13.5 (6.47 - 24.8) | 4.92 (4.64 - 5.23) |
| Meta-analysis | 179,352 | 1,124,440 | 103.6 | 968.8 | 403 | 6,358 | 3.89 (3.52 - 4.29) | 6.56 (6.40 - 6.73) |
| Meta-analysis (w/o USOC) | 29,729 | 250,777 | 13.6 | 167.8 | 79 | 1,431 | 5.82 (4.61 - 7.26) | 8.53 (8.09 - 8.98) |

##### 4.5.26 Vomiting

**Supplementary Table 419:** Exposures, outcomes and risk estimates for Vomiting in GLP1RA (male) (T) vs SU (male) (C) new-user cohorts under four different propensity score (PS) adjustment / time-at-risk (TAR) designs across data sources. We report total patient exposure time in 1,000 patient-year (KYr) units, uncalibrated and calibrated hazard ratios (HRs) and their 95% confidence intervals (CIs). Greyed out entries did not pass study diagnostics and are included only for completeness.

| | Patients | | Exposure (KYr) | | Outcomes | | Incidence (per KYr) | |
|---|---|---|---|---|---|---|---|---|
| | T | C | T | C | T | C | T | C |
| **PS-matching / on-treatment TAR** | | | | | | | | |
| CCAE | 10,715 | 81,419 | 7.2 | 69.9 | 161 | 1,000 | 22.5 (19.2 - 26.3) | 14.3 (13.4 - 15.2) |
| ODOD | 7,003 | 58,306 | 4.0 | 52.6 | 116 | 1,150 | 28.7 (23.7 - 34.5) | 21.8 (20.6 - 23.1) |
| MDCR | 889 | 14,632 | 0.6 | 15.9 | 12 | 356 | 19.9 (10.3 - 34.7) | 22.4 (20.2 - 24.9) |
| Open Claims | 128,040 | 808,320 | 91.3 | 877.2 | 1,378 | 10,087 | 15.1 (14.3 - 15.9) | 11.5 (11.3 - 11.7) |
| OEHR | 7,894 | 82,571 | 2.3 | 44.7 | 85 | 833 | 37.2 (29.7 - 46.0) | 18.6 (17.4 - 19.9) |
| VA | 1,250 | 30,824 | 0.9 | 23.1 | 14 | 167 | 15.6 (8.52 - 26.1) | 7.24 (6.18 - 8.43) |
| Meta-analysis | 154,541 | 1,045,248 | 105.3 | 1060.3 | 1,752 | 13,426 | 16.6 (15.9 - 17.4) | 12.7 (12.4 - 12.9) |
| Meta-analysis (w/o USOC) | 26,501 | 236,928 | 14.1 | 183.1 | 374 | 3,339 | 26.6 (23.9 - 29.4) | 18.2 (17.6 - 18.9) |
| **PS-stratification / on-treatment TAR** | | | | | | | | |
| CCAE | 12,118 | 85,512 | 7.8 | 73.2 | 180 | 1,068 | 23.0 (19.7 - 26.6) | 14.6 (13.7 - 15.5) |
| ODOD | 7,839 | 65,485 | 4.4 | 60.9 | 125 | 1,409 | 28.3 (23.6 - 33.7) | 23.1 (21.9 - 24.4) |
| MDCR | 960 | 19,051 | 0.6 | 20.3 | 12 | 494 | 19.0 (9.82 - 33.2) | 24.3 (22.2 - 26.6) |
| Open Claims | 141,956 | 835,989 | 97.9 | 909.2 | 1,492 | 10,876 | 15.2 (14.5 - 16.0) | 12.0 (11.7 - 12.2) |
| OEHR | 8,595 | 91,557 | 2.4 | 50.0 | 90 | 954 | 36.8 (29.6 - 45.3) | 19.1 (17.9 - 20.3) |
| VA | 1,349 | 229,643 | 0.9 | 244.3 | 14 | 1,751 | 15.0 (8.22 - 25.2) | 7.17 (6.84 - 7.51) |
| Meta-analysis | 170,508 | 1,078,543 | 112.6 | 1093.4 | 1,887 | 14,307 | 16.8 (16.0 - 17.5) | 13.1 (12.9 - 13.3) |
| Meta-analysis (w/o USOC) | 28,552 | 242,554 | 14.7 | 184.2 | 395 | 3,431 | 26.9 (24.3 - 29.7) | 18.6 (18.0 - 19.3) |
| **PS-matching / on-treatment TAR without escalation** | | | | | | | | |
| CCAE | 10,715 | 81,419 | 6.2 | 60.7 | 145 | 877 | 23.3 (19.7 - 27.4) | 14.4 (13.5 - 15.4) |
| ODOD | 7,003 | 58,306 | 3.6 | 46.9 | 106 | 1,002 | 29.4 (24.1 - 35.6) | 21.4 (20.1 - 22.7) |
| MDCR | 889 | 14,632 | 0.5 | 14.4 | 8 | 309 | 15.0 (6.46 - 29.5) | 21.5 (19.2 - 24.1) |
| Open Claims | 128,040 | 808,320 | 79.5 | 744.5 | 1,183 | 8,312 | 14.9 (14.0 - 15.8) | 11.2 (10.9 - 11.4) |
| OEHR | 7,894 | 82,571 | 2.0 | 39.1 | 69 | 632 | 34.8 (27.1 - 44.1) | 16.1 (14.9 - 17.5) |
| VA | 1,250 | 30,824 | 0.7 | 20.2 | 13 | 133 | 18.4 (9.80 - 31.5) | 6.57 (5.50 - 7.79) |
| Meta-analysis | 154,541 | 1,045,248 | 91.8 | 905.7 | 1,511 | 11,132 | 16.5 (15.6 - 17.3) | 12.3 (12.1 - 12.5) |
| Meta-analysis (w/o USOC) | 26,501 | 236,928 | 12.3 | 161.1 | 328 | 2,820 | 26.6 (23.8 - 29.6) | 17.5 (16.9 - 18.2) |
| **PS-stratification / on-treatment TAR without escalation** | | | | | | | | |
| CCAE | 12,118 | 85,512 | 6.9 | 63.7 | 163 | 938 | 23.8 (20.3 - 27.7) | 14.7 (13.8 - 15.7) |
| ODOD | 7,839 | 65,485 | 3.9 | 54.4 | 114 | 1,234 | 28.9 (23.9 - 34.8) | 22.7 (21.4 - 24.0) |
| MDCR | 960 | 19,051 | 0.6 | 18.5 | 8 | 433 | 14.3 (6.16 - 28.1) | 23.4 (21.3 - 25.7) |
| Open Claims | 141,956 | 835,989 | 85.7 | 772.2 | 1,293 | 8,987 | 15.1 (14.3 - 15.9) | 11.6 (11.4 - 11.9) |
| OEHR | 8,595 | 91,557 | 2.1 | 43.7 | 74 | 718 | 34.7 (27.3 - 43.6) | 16.4 (15.2 - 17.7) |
| VA | 1,349 | 229,643 | 0.7 | 217.9 | 13 | 1,469 | 17.7 (9.41 - 30.2) | 6.74 (6.40 - 7.09) |
| Meta-analysis | 170,508 | 1,078,543 | 98.6 | 934.1 | 1,644 | 11,877 | 16.7 (15.9 - 17.5) | 12.7 (12.5 - 12.9) |
| Meta-analysis (w/o USOC) | 28,552 | 242,554 | 12.9 | 161.9 | 351 | 2,890 | 27.2 (24.4 - 30.1) | 17.9 (17.2 - 18.5) |

#### 4.6 DPP4I (male) vs SU (male)

##### 4.6.1 3-point MACE

**Supplementary Table 420:** Exposures, outcomes and risk estimates for 3-point MACE in DPP4I (male) (T) vs SU (male) (C) new-user cohorts under four different propensity score (PS) adjustment / time-at-risk (TAR) designs across data sources. We report total patient exposure time in 1,000 patient-year (KYr) units, uncalibrated and calibrated hazard ratios (HRs) and their 95% confidence intervals (CIs). Greyed out entries did not pass study diagnostics and are included only for completeness.

| | Patients | | Exposure (KYr) | | Outcomes | | Incidence (per KYr) | |
|---|---|---|---|---|---|---|---|---|
| | T | C | T | C | T | C | T | C |
| **PS-matching / on-treatment TAR** | | | | | | | | |
| CCAE | 45,796 | 81,969 | 39.9 | 69.6 | 354 | 722 | 8.88 (7.98 - 9.85) | 10.4 (9.64 - 11.2) |
| DAG | 13,419 | 2,361 | 14.3 | 2.4 | 0 | 0 | | |
| ODOD | 27,622 | 66,613 | 23.1 | 61.8 | 442 | 1,567 | 19.1 (17.4 - 21.0) | 25.3 (24.1 - 26.6) |
| MDCR | 8,273 | 16,421 | 8.6 | 17.1 | 247 | 571 | 28.8 (25.3 - 32.6) | 33.4 (30.7 - 36.2) |
| MDCD | 3,727 | 8,994 | 2.8 | 6.4 | 65 | 195 | 22.9 (17.7 - 29.1) | 30.5 (26.3 - 35.1) |
| Open Claims | 395,491 | 843,428 | 376.8 | 911.1 | 4,098 | 12,752 | 10.9 (10.5 - 11.2) | 14.0 (13.8 - 14.2) |
| OEHR | 35,578 | 93,394 | 17.5 | 51.3 | 194 | 749 | 11.1 (9.57 - 12.7) | 14.6 (13.6 - 15.7) |
| VA | 0 | 0 | 0.0 | 0.0 | 0 | 0 | | |
| SIDIAP | 25,996 | 7,090 | 75.8 | 20.0 | 375 | 72 | 4.95 (4.46 - 5.47) | 3.59 (2.81 - 4.52) |
| Meta-analysis | 542,483 | 1,117,909 | 544.6 | 1137.4 | 5,775 | 16,628 | 10.6 (10.3 - 10.9) | 14.6 (14.4 - 14.8) |
| Meta-analysis (w/o USOC) | 146,992 | 274,481 | 167.7 | 226.2 | 1,677 | 3,876 | 10.00 (9.53 - 10.5) | 17.1 (16.6 - 17.7) |
| **PS-stratification / on-treatment TAR** | | | | | | | | |
| CCAE | 51,105 | 83,416 | 44.7 | 70.7 | 383 | 728 | 8.57 (7.74 - 9.48) | 10.3 (9.56 - 11.1) |
| DAG | 13,447 | 2,736 | 14.3 | 2.8 | 0 | 0 | | |
| ODOD | 29,200 | 66,618 | 24.5 | 61.8 | 458 | 1,568 | 18.7 (17.0 - 20.5) | 25.4 (24.1 - 26.7) |
| MDCR | 8,780 | 16,871 | 9.2 | 17.6 | 259 | 589 | 28.3 (24.9 - 31.9) | 33.4 (30.8 - 36.2) |
| MDCD | 3,809 | 9,012 | 2.9 | 6.4 | 67 | 195 | 23.0 (17.8 - 29.2) | 30.4 (26.3 - 35.0) |
| Open Claims | 402,801 | 843,441 | 386.0 | 911.2 | 4,214 | 12,752 | 10.9 (10.6 - 11.3) | 14.0 (13.8 - 14.2) |
| OEHR | 37,370 | 93,423 | 18.6 | 51.3 | 209 | 750 | 11.2 (9.77 - 12.9) | 14.6 (13.6 - 15.7) |
| VA | 24,642 | 224,563 | 23.0 | 237.8 | 124 | 2,007 | 5.38 (4.48 - 6.42) | 8.44 (8.07 - 8.82) |
| SIDIAP | 26,076 | 9,118 | 75.9 | 27.1 | 376 | 92 | 4.95 (4.47 - 5.48) | 3.40 (2.74 - 4.17) |
| Meta-analysis | 559,141 | 1,121,899 | 561.7 | 1146.1 | 5,966 | 16,674 | 10.6 (10.4 - 10.9) | 14.5 (14.3 - 14.8) |
| Meta-analysis (w/o USOC) | 156,340 | 278,458 | 175.8 | 235.0 | 1,752 | 3,922 | 9.97 (9.51 - 10.4) | 16.7 (16.2 - 17.2) |
| **PS-matching / on-treatment TAR without escalation** | | | | | | | | |
| CCAE | 45,796 | 81,969 | 32.2 | 60.5 | 281 | 632 | 8.72 (7.73 - 9.80) | 10.4 (9.64 - 11.3) |
| DAG | 13,419 | 2,361 | 12.9 | 2.2 | 0 | 0 | | |
| ODOD | 27,622 | 66,613 | 18.9 | 55.2 | 377 | 1,401 | 20.0 (18.0 - 22.1) | 25.4 (24.1 - 26.8) |
| MDCR | 8,273 | 16,421 | 7.3 | 15.6 | 206 | 509 | 28.1 (24.4 - 32.2) | 32.6 (29.9 - 35.6) |
| MDCD | 3,727 | 8,994 | 2.2 | 5.4 | 47 | 165 | 21.8 (16.0 - 29.0) | 30.7 (26.2 - 35.7) |
| Open Claims | 395,491 | 843,428 | 292.4 | 772.3 | 3,195 | 10,930 | 10.9 (10.6 - 11.3) | 14.2 (13.9 - 14.4) |
| OEHR | 35,578 | 93,394 | 14.2 | 44.7 | 135 | 547 | 9.48 (7.95 - 11.2) | 12.2 (11.2 - 13.3) |
| VA | 0 | 0 | 0.0 | 0.0 | 0 | 0 | | |
| SIDIAP | 25,996 | 7,090 | 55.7 | 16.8 | 260 | 60 | 4.67 (4.12 - 5.27) | 3.58 (2.73 - 4.61) |
| Meta-analysis | 542,483 | 1,117,909 | 422.9 | 970.4 | 4,501 | 14,244 | 10.6 (10.3 - 11.0) | 14.7 (14.4 - 14.9) |
| Meta-analysis (w/o USOC) | 146,992 | 274,481 | 130.5 | 198.2 | 1,306 | 3,314 | 10.0 (9.47 - 10.6) | 16.7 (16.2 - 17.3) |
| **PS-stratification / on-treatment TAR without escalation** | | | | | | | | |
| CCAE | 51,105 | 83,416 | 36.2 | 61.6 | 306 | 638 | 8.46 (7.54 - 9.47) | 10.3 (9.56 - 11.2) |
| DAG | 13,447 | 2,736 | 12.9 | 2.6 | 0 | 0 | | |
| ODOD | 29,200 | 66,618 | 20.0 | 55.2 | 390 | 1,402 | 19.5 (17.6 - 21.5) | 25.4 (24.1 - 26.8) |
| MDCR | 8,780 | 16,871 | 7.8 | 16.1 | 215 | 524 | 27.4 (23.9 - 31.3) | 32.6 (29.8 - 35.5) |
| MDCD | 3,809 | 9,012 | 2.2 | 5.4 | 49 | 165 | 22.2 (16.5 - 29.4) | 30.6 (26.1 - 35.7) |
| Open Claims | 402,801 | 843,441 | 299.3 | 772.3 | 3,277 | 10,930 | 10.9 (10.6 - 11.3) | 14.2 (13.9 - 14.4) |
| OEHR | 37,370 | 93,423 | 15.1 | 44.7 | 147 | 548 | 9.71 (8.21 - 11.4) | 12.2 (11.2 - 13.3) |
| VA | 24,642 | 224,563 | 18.7 | 212.0 | 86 | 1,719 | 4.59 (3.67 - 5.67) | 8.11 (7.73 - 8.50) |
| SIDIAP | 26,076 | 9,118 | 55.7 | 22.4 | 261 | 76 | 4.68 (4.13 - 5.29) | 3.39 (2.67 - 4.25) |
| Meta-analysis | 559,141 | 1,121,899 | 436.4 | 977.7 | 4,645 | 14,283 | 10.6 (10.3 - 11.0) | 14.6 (14.4 - 14.9) |
| Meta-analysis (w/o USOC) | 156,340 | 278,458 | 137.1 | 205.4 | 1,368 | 3,353 | 9.98 (9.45 - 10.5) | 16.3 (15.8 - 16.9) |

##### 4.6.2 4-point MACE

**Supplementary Table 421:** Exposures, outcomes and risk estimates for 4-point MACE in DPP4I (male) (T) vs SU (male) (C) new-user cohorts under four different propensity score (PS) adjustment / time-at-risk (TAR) designs across data sources. We report total patient exposure time in 1,000 patient-year (KYr) units, uncalibrated and calibrated hazard ratios (HRs) and their 95% confidence intervals (CIs). Greyed out entries did not pass study diagnostics and are included only for completeness.

| | Patients | | Exposure (KYr) | | Outcomes | | Incidence (per KYr) | |
|---|---|---|---|---|---|---|---|---|
| | T | C | T | C | T | C | T | C |
| **PS-matching / on-treatment TAR** | | | | | | | | |
| CCAE | 45,149 | 80,662 | 39.2 | 68.3 | 514 | 1,061 | 13.1 (12.0 - 14.3) | 15.5 (14.6 - 16.5) |
| DAG | 13,419 | 2,361 | 14.3 | 2.4 | 0 | 0 | | |
| ODOD | 26,531 | 63,762 | 22.1 | 58.5 | 649 | 2,459 | 29.3 (27.1 - 31.7) | 42.1 (40.4 - 43.8) |
| MDCR | 7,728 | 15,158 | 7.9 | 15.7 | 386 | 912 | 48.7 (43.9 - 53.8) | 58.2 (54.5 - 62.1) |
| MDCD | 3,462 | 8,289 | 2.6 | 5.8 | 91 | 286 | 34.9 (28.1 - 42.9) | 48.9 (43.4 - 54.9) |
| Open Claims | 382,594 | 813,874 | 363.8 | 875.2 | 6,389 | 19,940 | 17.6 (17.1 - 18.0) | 22.8 (22.5 - 23.1) |
| OEHR | 34,918 | 91,595 | 17.0 | 49.6 | 344 | 1,349 | 20.2 (18.2 - 22.5) | 27.2 (25.8 - 28.7) |
| VA | 0 | 0 | 0.0 | 0.0 | 0 | 0 | | |
| SIDIAP | 25,887 | 7,076 | 75.5 | 20.0 | 471 | 95 | 6.24 (5.69 - 6.83) | 4.75 (3.85 - 5.81) |
| Meta-analysis | 526,269 | 1,080,416 | 528.2 | 1093.1 | 8,844 | 26,102 | 16.7 (16.4 - 17.1) | 23.9 (23.6 - 24.2) |
| Meta-analysis (w/o USOC) | 143,675 | 266,542 | 164.4 | 217.8 | 2,455 | 6,162 | 14.9 (14.4 - 15.5) | 28.3 (27.6 - 29.0) |
| **PS-stratification / on-treatment TAR** | | | | | | | | |
| CCAE | 50,414 | 82,081 | 44.0 | 69.5 | 553 | 1,070 | 12.6 (11.6 - 13.7) | 15.4 (14.5 - 16.4) |
| DAG | 13,447 | 2,736 | 14.3 | 2.8 | 0 | 0 | | |
| ODOD | 28,078 | 63,766 | 23.5 | 58.5 | 680 | 2,460 | 28.9 (26.8 - 31.2) | 42.1 (40.4 - 43.8) |
| MDCR | 8,212 | 15,576 | 8.5 | 16.1 | 407 | 935 | 47.9 (43.3 - 52.7) | 57.9 (54.2 - 61.7) |
| MDCD | 3,536 | 8,304 | 2.7 | 5.9 | 92 | 286 | 34.5 (27.8 - 42.3) | 48.8 (43.3 - 54.8) |
| Open Claims | 389,953 | 813,885 | 372.9 | 875.2 | 6,586 | 19,940 | 17.7 (17.2 - 18.1) | 22.8 (22.5 - 23.1) |
| OEHR | 36,683 | 91,624 | 18.0 | 49.6 | 371 | 1,350 | 20.6 (18.5 - 22.8) | 27.2 (25.8 - 28.7) |
| VA | 24,006 | 218,190 | 22.3 | 229.1 | 268 | 3,973 | 12.0 (10.6 - 13.5) | 17.3 (16.8 - 17.9) |
| SIDIAP | 25,965 | 9,103 | 75.6 | 27.0 | 472 | 123 | 6.25 (5.70 - 6.84) | 4.55 (3.78 - 5.43) |
| Meta-analysis | 542,841 | 1,084,339 | 545.2 | 1101.8 | 9,161 | 26,164 | 16.8 (16.5 - 17.2) | 23.7 (23.5 - 24.0) |
| Meta-analysis (w/o USOC) | 152,888 | 270,454 | 172.3 | 226.5 | 2,575 | 6,224 | 14.9 (14.4 - 15.5) | 27.5 (26.8 - 28.2) |
| **PS-matching / on-treatment TAR without escalation** | | | | | | | | |
| CCAE | 45,149 | 80,662 | 31.7 | 59.5 | 409 | 919 | 12.9 (11.7 - 14.2) | 15.4 (14.5 - 16.5) |
| DAG | 13,419 | 2,361 | 12.9 | 2.2 | 0 | 0 | | |
| ODOD | 26,531 | 63,762 | 18.1 | 52.2 | 546 | 2,183 | 30.1 (27.7 - 32.8) | 41.8 (40.1 - 43.6) |
| MDCR | 7,728 | 15,158 | 6.8 | 14.3 | 332 | 826 | 48.9 (43.8 - 54.4) | 57.8 (53.9 - 61.8) |
| MDCD | 3,462 | 8,289 | 2.0 | 4.9 | 68 | 240 | 34.3 (26.6 - 43.5) | 48.8 (42.8 - 55.4) |
| Open Claims | 382,594 | 813,874 | 282.5 | 742.6 | 4,974 | 17,036 | 17.6 (17.1 - 18.1) | 22.9 (22.6 - 23.3) |
| OEHR | 34,918 | 91,595 | 13.9 | 43.5 | 247 | 1,044 | 17.8 (15.6 - 20.1) | 24.0 (22.6 - 25.5) |
| VA | 0 | 0 | 0.0 | 0.0 | 0 | 0 | | |
| SIDIAP | 25,887 | 7,076 | 55.4 | 16.7 | 330 | 80 | 5.95 (5.33 - 6.63) | 4.79 (3.79 - 5.96) |
| Meta-analysis | 526,269 | 1,080,416 | 410.4 | 933.8 | 6,906 | 22,328 | 16.8 (16.4 - 17.2) | 23.9 (23.6 - 24.2) |
| Meta-analysis (w/o USOC) | 143,675 | 266,542 | 127.9 | 191.2 | 1,932 | 5,292 | 15.1 (14.4 - 15.8) | 27.7 (26.9 - 28.4) |
| **PS-stratification / on-treatment TAR without escalation** | | | | | | | | |
| CCAE | 50,414 | 82,081 | 35.6 | 60.6 | 441 | 928 | 12.4 (11.3 - 13.6) | 15.3 (14.3 - 16.3) |
| DAG | 13,447 | 2,736 | 12.9 | 2.6 | 0 | 0 | | |
| ODOD | 28,078 | 63,766 | 19.2 | 52.2 | 573 | 2,184 | 29.8 (27.4 - 32.3) | 41.8 (40.1 - 43.6) |
| MDCR | 8,212 | 15,576 | 7.3 | 14.8 | 349 | 847 | 48.0 (43.1 - 53.3) | 57.3 (53.5 - 61.3) |
| MDCD | 3,536 | 8,304 | 2.0 | 4.9 | 69 | 240 | 34.2 (26.6 - 43.2) | 48.7 (42.7 - 55.3) |
| Open Claims | 389,953 | 813,885 | 289.4 | 742.6 | 5,120 | 17,036 | 17.7 (17.2 - 18.2) | 22.9 (22.6 - 23.3) |
| OEHR | 36,683 | 91,624 | 14.8 | 43.5 | 267 | 1,045 | 18.1 (16.0 - 20.4) | 24.0 (22.6 - 25.5) |
| VA | 24,006 | 218,190 | 18.2 | 204.6 | 197 | 3,386 | 10.8 (9.38 - 12.5) | 16.5 (16.0 - 17.1) |
| SIDIAP | 25,965 | 9,103 | 55.5 | 22.3 | 331 | 101 | 5.96 (5.34 - 6.64) | 4.52 (3.68 - 5.49) |
| Meta-analysis | 542,841 | 1,084,339 | 423.8 | 941.0 | 7,150 | 22,381 | 16.9 (16.5 - 17.3) | 23.8 (23.5 - 24.1) |
| Meta-analysis (w/o USOC) | 152,888 | 270,454 | 134.4 | 198.4 | 2,030 | 5,345 | 15.1 (14.5 - 15.8) | 26.9 (26.2 - 27.7) |

##### 4.6.3 Acute myocardial infarction

**Supplementary Table 422:** Exposures, outcomes and risk estimates for Acute myocardial infarction in DPP4I (male) (T) vs SU (male) (C) new-user cohorts under four different propensity score (PS) adjustment / time-at-risk (TAR) designs across data sources. We report total patient exposure time in 1,000 patient-year (KYr) units, uncalibrated and calibrated hazard ratios (HRs) and their 95% confidence intervals (CIs). Greyed out entries did not pass study diagnostics and are included only for completeness.

| | Patients | | Exposure (KYr) | | Outcomes | | Incidence (per KYr) | |
|---|---|---|---|---|---|---|---|---|
| | T | C | T | C | T | C | T | C |
| **PS-matching / on-treatment TAR** | | | | | | | | |
| CCAE | 46,342 | 83,132 | 40.4 | 70.6 | 215 | 429 | 5.32 (4.64 - 6.08) | 6.07 (5.51 - 6.67) |
| DAG | 13,419 | 2,361 | 14.3 | 2.4 | 0 | 0 | | |
| ODOD | 28,348 | 68,425 | 23.8 | 64.0 | 227 | 825 | 9.55 (8.35 - 10.9) | 12.9 (12.0 - 13.8) |
| MDCR | 8,611 | 17,208 | 9.0 | 18.0 | 115 | 280 | 12.8 (10.6 - 15.4) | 15.5 (13.8 - 17.4) |
| MDCD | 3,879 | 9,389 | 3.0 | 6.7 | 34 | 99 | 11.5 (7.95 - 16.0) | 14.7 (12.0 - 17.9) |
| Open Claims | 406,847 | 868,427 | 387.9 | 940.2 | 2,133 | 6,664 | 5.50 (5.27 - 5.74) | 7.09 (6.92 - 7.26) |
| OEHR | 35,898 | 94,369 | 17.7 | 52.1 | 106 | 433 | 5.98 (4.89 - 7.23) | 8.31 (7.54 - 9.13) |
| VA | 0 | 0 | 0.0 | 0.0 | 0 | 0 | | |
| SIDIAP | 26,300 | 7,143 | 76.9 | 20.2 | 180 | 42 | 2.34 (2.01 - 2.71) | 2.08 (1.50 - 2.81) |
| Meta-analysis | 556,225 | 1,148,093 | 558.6 | 1171.9 | 3,010 | 8,772 | 5.39 (5.20 - 5.58) | 7.49 (7.33 - 7.64) |
| Meta-analysis (w/o USOC) | 149,378 | 279,666 | 170.7 | 231.7 | 877 | 2,108 | 5.14 (4.80 - 5.49) | 9.10 (8.71 - 9.49) |
| **PS-stratification / on-treatment TAR** | | | | | | | | |
| CCAE | 51,682 | 84,598 | 45.2 | 71.9 | 231 | 432 | 5.11 (4.47 - 5.81) | 6.01 (5.46 - 6.61) |
| DAG | 13,447 | 2,736 | 14.3 | 2.8 | 0 | 0 | | |
| ODOD | 29,945 | 68,430 | 25.2 | 64.0 | 235 | 825 | 9.31 (8.16 - 10.6) | 12.9 (12.0 - 13.8) |
| MDCR | 9,130 | 17,670 | 9.6 | 18.6 | 120 | 288 | 12.5 (10.4 - 15.0) | 15.5 (13.8 - 17.4) |
| MDCD | 3,971 | 9,409 | 3.0 | 6.7 | 37 | 99 | 12.2 (8.56 - 16.8) | 14.7 (11.9 - 17.9) |
| Open Claims | 414,179 | 868,440 | 397.1 | 940.2 | 2,192 | 6,664 | 5.52 (5.29 - 5.76) | 7.09 (6.92 - 7.26) |
| OEHR | 37,709 | 94,398 | 18.8 | 52.1 | 116 | 434 | 6.17 (5.09 - 7.39) | 8.32 (7.56 - 9.14) |
| VA | 25,018 | 227,520 | 23.4 | 241.4 | 65 | 1,220 | 2.78 (2.14 - 3.54) | 5.05 (4.77 - 5.35) |
| SIDIAP | 26,383 | 9,182 | 76.9 | 27.3 | 181 | 52 | 2.35 (2.02 - 2.72) | 1.91 (1.42 - 2.50) |
| Meta-analysis | 572,999 | 1,152,127 | 575.9 | 1180.8 | 3,112 | 8,794 | 5.40 (5.22 - 5.60) | 7.45 (7.29 - 7.61) |
| Meta-analysis (w/o USOC) | 158,820 | 283,687 | 178.8 | 240.6 | 920 | 2,130 | 5.14 (4.82 - 5.49) | 8.85 (8.48 - 9.24) |
| **PS-matching / on-treatment TAR without escalation** | | | | | | | | |
| CCAE | 46,342 | 83,132 | 32.7 | 61.5 | 169 | 370 | 5.17 (4.42 - 6.02) | 6.02 (5.42 - 6.66) |
| DAG | 13,419 | 2,361 | 12.9 | 2.2 | 0 | 0 | | |
| ODOD | 28,348 | 68,425 | 19.5 | 57.1 | 184 | 737 | 9.45 (8.14 - 10.9) | 12.9 (12.0 - 13.9) |
| MDCR | 8,611 | 17,208 | 7.7 | 16.4 | 96 | 246 | 12.5 (10.1 - 15.3) | 15.0 (13.1 - 16.9) |
| MDCD | 3,879 | 9,389 | 2.3 | 5.6 | 28 | 82 | 12.4 (8.25 - 17.9) | 14.5 (11.5 - 18.0) |
| Open Claims | 406,847 | 868,427 | 300.8 | 796.8 | 1,643 | 5,681 | 5.46 (5.20 - 5.73) | 7.13 (6.95 - 7.32) |
| OEHR | 35,898 | 94,369 | 14.4 | 45.4 | 70 | 302 | 4.86 (3.79 - 6.14) | 6.66 (5.93 - 7.45) |
| VA | 0 | 0 | 0.0 | 0.0 | 0 | 0 | | |
| SIDIAP | 26,300 | 7,143 | 56.4 | 16.9 | 132 | 32 | 2.34 (1.96 - 2.77) | 1.89 (1.30 - 2.67) |
| Meta-analysis | 556,225 | 1,148,093 | 433.7 | 999.7 | 2,322 | 7,450 | 5.35 (5.14 - 5.58) | 7.45 (7.28 - 7.62) |
| Meta-analysis (w/o USOC) | 149,378 | 279,666 | 132.9 | 202.9 | 679 | 1,769 | 5.11 (4.73 - 5.51) | 8.72 (8.32 - 9.13) |
| **PS-stratification / on-treatment TAR without escalation** | | | | | | | | |
| CCAE | 51,682 | 84,598 | 36.6 | 62.6 | 182 | 373 | 4.97 (4.28 - 5.75) | 5.96 (5.37 - 6.59) |
| DAG | 13,447 | 2,736 | 12.9 | 2.6 | 0 | 0 | | |
| ODOD | 29,945 | 68,430 | 20.6 | 57.1 | 190 | 737 | 9.21 (7.95 - 10.6) | 12.9 (12.0 - 13.9) |
| MDCR | 9,130 | 17,670 | 8.2 | 17.0 | 100 | 252 | 12.2 (9.93 - 14.9) | 14.9 (13.1 - 16.8) |
| MDCD | 3,971 | 9,409 | 2.3 | 5.7 | 31 | 82 | 13.4 (9.12 - 19.1) | 14.5 (11.5 - 18.0) |
| Open Claims | 414,179 | 868,440 | 307.8 | 796.8 | 1,685 | 5,681 | 5.47 (5.22 - 5.74) | 7.13 (6.95 - 7.32) |
| OEHR | 37,709 | 94,398 | 15.3 | 45.4 | 78 | 303 | 5.10 (4.03 - 6.36) | 6.68 (5.95 - 7.47) |
| VA | 25,018 | 227,520 | 19.0 | 215.1 | 50 | 1,037 | 2.63 (1.95 - 3.47) | 4.82 (4.53 - 5.12) |
| SIDIAP | 26,383 | 9,182 | 56.5 | 22.6 | 133 | 40 | 2.35 (1.97 - 2.79) | 1.77 (1.27 - 2.41) |
| Meta-analysis | 572,999 | 1,152,127 | 447.3 | 1007.0 | 2,399 | 7,468 | 5.36 (5.15 - 5.58) | 7.42 (7.25 - 7.59) |
| Meta-analysis (w/o USOC) | 158,820 | 283,687 | 139.5 | 210.2 | 714 | 1,787 | 5.12 (4.75 - 5.51) | 8.50 (8.11 - 8.90) |

##### 4.6.4 Acute renal failure

**Supplementary Table 423:** Exposures, outcomes and risk estimates for Acute renal failure in DPP4I (male) (T) vs SU (male) (C) new-user cohorts under four different propensity score (PS) adjustment / time-at-risk (TAR) designs across data sources. We report total patient exposure time in 1,000 patient-year (KYr) units, uncalibrated and calibrated hazard ratios (HRs) and their 95% confidence intervals (CIs). Greyed out entries did not pass study diagnostics and are included only for completeness.

| | Patients | | Exposure (KYr) | | Outcomes | | Incidence (per KYr) | |
|---|---|---|---|---|---|---|---|---|
| | T | C | T | C | T | C | T | C |
| **PS-matching / on-treatment TAR** | | | | | | | | |
| CCAE | 46,613 | 83,347 | 40.7 | 71.0 | 345 | 734 | 8.47 (7.60 - 9.41) | 10.3 (9.61 - 11.1) |
| DAG | 13,419 | 2,361 | 14.3 | 2.4 | 0 | 0 | | |
| ODOD | 27,580 | 66,579 | 23.0 | 61.5 | 630 | 2,226 | 27.4 (25.3 - 29.6) | 36.2 (34.7 - 37.7) |
| MDCR | 8,428 | 16,789 | 8.7 | 17.5 | 323 | 727 | 37.3 (33.3 - 41.6) | 41.6 (38.6 - 44.7) |
| MDCD | 3,730 | 8,975 | 2.8 | 6.4 | 101 | 277 | 35.9 (29.3 - 43.7) | 43.3 (38.3 - 48.7) |
| Open Claims | 404,418 | 862,829 | 384.6 | 930.8 | 5,819 | 16,972 | 15.1 (14.7 - 15.5) | 18.2 (18.0 - 18.5) |
| OEHR | 35,818 | 94,028 | 17.6 | 51.4 | 329 | 1,132 | 18.7 (16.8 - 20.9) | 22.0 (20.7 - 23.3) |
| VA | 0 | 0 | 0.0 | 0.0 | 0 | 0 | | |
| SIDIAP | 26,766 | 7,212 | 78.4 | 20.5 | 34 | <5 | 0.43 (0.30 - 0.61) | < 0.24 (0.08 - 0.57) |
| Meta-analysis | 553,353 | 1,139,759 | 555.7 | 1159.2 | 7,581 | <22,073 | 13.6 (13.3 - 14.0) | < 19.0 (18.8 - 19.3) |
| Meta-analysis (w/o USOC) | 148,935 | 276,930 | 171.1 | 228.3 | 1,762 | <5,101 | 10.3 (9.82 - 10.8) | < 22.3 (21.7 - 23.0) |
| **PS-stratification / on-treatment TAR** | | | | | | | | |
| CCAE | 51,990 | 84,940 | 45.6 | 72.3 | 388 | 744 | 8.51 (7.69 - 9.40) | 10.3 (9.56 - 11.1) |
| DAG | 13,447 | 2,736 | 14.3 | 2.8 | 0 | 0 | | |
| ODOD | 29,183 | 66,582 | 24.5 | 61.5 | 659 | 2,227 | 26.9 (24.9 - 29.1) | 36.2 (34.7 - 37.7) |
| MDCR | 8,907 | 17,252 | 9.2 | 18.0 | 339 | 741 | 36.8 (33.0 - 40.9) | 41.1 (38.2 - 44.2) |
| MDCD | 3,816 | 8,991 | 2.9 | 6.4 | 103 | 277 | 35.7 (29.1 - 43.2) | 43.2 (38.3 - 48.6) |
| Open Claims | 411,928 | 862,839 | 394.0 | 930.8 | 5,966 | 16,973 | 15.1 (14.8 - 15.5) | 18.2 (18.0 - 18.5) |
| OEHR | 37,636 | 94,058 | 18.6 | 51.5 | 341 | 1,133 | 18.3 (16.4 - 20.3) | 22.0 (20.8 - 23.3) |
| VA | 24,628 | 226,342 | 22.9 | 238.2 | 295 | 3,666 | 12.9 (11.4 - 14.4) | 15.4 (14.9 - 15.9) |
| SIDIAP | 26,859 | 9,265 | 78.5 | 27.6 | 34 | 8 | 0.43 (0.30 - 0.61) | 0.29 (0.12 - 0.57) |
| Meta-analysis | 570,319 | 1,143,927 | 573.3 | 1168.2 | 7,830 | 22,103 | 13.7 (13.4 - 14.0) | 18.9 (18.7 - 19.2) |
| Meta-analysis (w/o USOC) | 158,391 | 281,088 | 179.3 | 237.4 | 1,864 | 5,130 | 10.4 (9.93 - 10.9) | 21.6 (21.0 - 22.2) |
| **PS-matching / on-treatment TAR without escalation** | | | | | | | | |
| CCAE | 46,613 | 83,347 | 32.9 | 61.7 | 274 | 625 | 8.33 (7.37 - 9.37) | 10.1 (9.34 - 10.9) |
| DAG | 13,419 | 2,361 | 12.9 | 2.2 | 0 | 0 | | |
| ODOD | 27,580 | 66,579 | 18.9 | 55.0 | 506 | 1,985 | 26.8 (24.5 - 29.3) | 36.1 (34.5 - 37.7) |
| MDCR | 8,428 | 16,789 | 7.4 | 16.0 | 269 | 648 | 36.3 (32.1 - 40.9) | 40.6 (37.5 - 43.8) |
| MDCD | 3,730 | 8,975 | 2.1 | 5.4 | 78 | 226 | 36.5 (28.8 - 45.5) | 41.9 (36.6 - 47.7) |
| Open Claims | 404,418 | 862,829 | 298.6 | 789.9 | 4,531 | 14,223 | 15.2 (14.7 - 15.6) | 18.0 (17.7 - 18.3) |
| OEHR | 35,818 | 94,028 | 14.3 | 45.0 | 224 | 833 | 15.6 (13.6 - 17.8) | 18.5 (17.3 - 19.8) |
| VA | 0 | 0 | 0.0 | 0.0 | 0 | 0 | | |
| SIDIAP | 26,766 | 7,212 | 57.4 | 17.1 | 22 | <5 | 0.38 (0.24 - 0.58) | < 0.29 (0.09 - 0.68) |
| Meta-analysis | 553,353 | 1,139,759 | 431.7 | 990.1 | 5,904 | <18,545 | 13.7 (13.3 - 14.0) | < 18.7 (18.5 - 19.0) |
| Meta-analysis (w/o USOC) | 148,935 | 276,930 | 133.1 | 200.2 | 1,373 | <4,322 | 10.3 (9.78 - 10.9) | < 21.6 (20.9 - 22.2) |
| **PS-stratification / on-treatment TAR without escalation** | | | | | | | | |
| CCAE | 51,990 | 84,940 | 36.9 | 63.0 | 307 | 635 | 8.32 (7.42 - 9.31) | 10.1 (9.31 - 10.9) |
| DAG | 13,447 | 2,736 | 12.9 | 2.6 | 0 | 0 | | |
| ODOD | 29,183 | 66,582 | 20.0 | 55.0 | 531 | 1,986 | 26.5 (24.3 - 28.9) | 36.1 (34.6 - 37.8) |
| MDCR | 8,907 | 17,252 | 7.9 | 16.5 | 283 | 661 | 35.9 (31.8 - 40.3) | 40.1 (37.1 - 43.3) |
| MDCD | 3,816 | 8,991 | 2.2 | 5.4 | 78 | 226 | 35.6 (28.1 - 44.4) | 41.8 (36.6 - 47.7) |
| Open Claims | 411,928 | 862,839 | 305.7 | 789.9 | 4,632 | 14,223 | 15.2 (14.7 - 15.6) | 18.0 (17.7 - 18.3) |
| OEHR | 37,636 | 94,058 | 15.2 | 45.0 | 234 | 834 | 15.4 (13.4 - 17.4) | 18.5 (17.3 - 19.8) |
| VA | 24,628 | 226,342 | 18.6 | 212.7 | 212 | 2,979 | 11.4 (9.90 - 13.0) | 14.0 (13.5 - 14.5) |
| SIDIAP | 26,859 | 9,265 | 57.5 | 22.8 | 22 | 7 | 0.38 (0.24 - 0.58) | 0.31 (0.12 - 0.63) |
| Meta-analysis | 570,319 | 1,143,927 | 445.4 | 997.6 | 6,087 | 18,572 | 13.7 (13.3 - 14.0) | 18.6 (18.3 - 18.9) |
| Meta-analysis (w/o USOC) | 158,391 | 281,088 | 139.7 | 207.7 | 1,455 | 4,349 | 10.4 (9.88 - 11.0) | 20.9 (20.3 - 21.6) |

##### 4.6.5 Glycemic control

**Supplementary Table 424:** Exposures, outcomes and risk estimates for Glycemic control in DPP4I (male) (T) vs SU (male) (C) new-user cohorts under four different propensity score (PS) adjustment / time-at-risk (TAR) designs across data sources. We report total patient exposure time in 1,000 patient-year (KYr) units, uncalibrated and calibrated hazard ratios (HRs) and their 95% confidence intervals (CIs). Greyed out entries did not pass study diagnostics and are included only for completeness.

| | Patients | | Exposure (KYr) | | Outcomes | | Incidence (per KYr) | |
|---|---|---|---|---|---|---|---|---|
| | T | C | T | C | T | C | T | C |
| **PS-matching / on-treatment TAR** | | | | | | | | |
| CCAE | 47,014 | 84,418 | 41.1 | 71.9 | 113 | 318 | 2.75 (2.26 - 3.30) | 4.43 (3.95 - 4.94) |
| DAG | 5,039 | 1,014 | 4.5 | 0.9 | 675 | 131 | 151 (139 - 162) | 154 (129 - 183) |
| ODOD | 20,061 | 52,049 | 15.1 | 43.2 | 1,578 | 4,237 | 104 (99.3 - 110) | 98.2 (95.2 - 101) |
| MDCR | 9,004 | 18,079 | 9.4 | 19.0 | 11 | 80 | 1.17 (0.58 - 2.09) | 4.21 (3.34 - 5.24) |
| MDCD | 4,087 | 9,851 | 3.1 | 7.1 | 0 | 0 | | |
| Open Claims | 422,431 | 903,346 | 403.8 | 982.0 | 0 | 0 | | |
| OEHR | 36,445 | 95,879 | 18.1 | 53.4 | 0 | 0 | | |
| VA | 0 | 0 | 0.0 | 0.0 | 0 | 0 | | |
| SIDIAP | 4,064 | 1,774 | 6.9 | 2.9 | 1,719 | 644 | 249 (237 - 261) | 226 (209 - 244) |
| Meta-analysis | 85,182 | 157,334 | 77.0 | 137.7 | 4,096 | 5,410 | 53.2 (51.6 - 54.8) | 39.3 (38.2 - 40.3) |
| Meta-analysis (w/o USOC) | 85,182 | 157,334 | 77.0 | 137.7 | 4,096 | 5,410 | 53.2 (51.6 - 54.8) | 39.3 (38.2 - 40.3) |
| **PS-stratification / on-treatment TAR** | | | | | | | | |
| CCAE | 52,351 | 85,573 | 46.0 | 72.7 | 131 | 490 | 2.85 (2.38 - 3.38) | 6.74 (6.16 - 7.36) |
| DAG | 5,066 | 1,195 | 4.5 | 1.0 | 683 | 161 | 152 (140 - 163) | 162 (138 - 190) |
| ODOD | 20,910 | 52,056 | 15.7 | 43.2 | 1,678 | 4,237 | 107 (102 - 112) | 98.2 (95.2 - 101) |
| MDCR | 9,515 | 18,352 | 10.0 | 19.3 | 12 | 174 | 1.20 (0.62 - 2.09) | 9.04 (7.74 - 10.5) |
| MDCD | 4,183 | 9,871 | 3.2 | 7.1 | 0 | 0 | | |
| Open Claims | 429,825 | 903,360 | 413.2 | 982.0 | 0 | 0 | | |
| OEHR | 38,289 | 95,909 | 19.2 | 53.4 | 0 | 0 | | |
| VA | 24,844 | 221,635 | 23.3 | 233.0 | 21 | 499 | 0.90 (0.56 - 1.38) | 2.14 (1.96 - 2.34) |
| SIDIAP | 4,096 | 2,569 | 6.9 | 4.3 | 1,729 | 944 | 249 (238 - 261) | 218 (204 - 232) |
| Meta-analysis | 91,938 | 159,745 | 83.1 | 140.5 | 4,233 | 6,006 | 50.9 (49.4 - 52.5) | 42.8 (41.7 - 43.9) |
| Meta-analysis (w/o USOC) | 91,938 | 159,745 | 83.1 | 140.5 | 4,233 | 6,006 | 50.9 (49.4 - 52.5) | 42.8 (41.7 - 43.9) |
| **PS-matching / on-treatment TAR without escalation** | | | | | | | | |
| CCAE | 47,014 | 84,418 | 33.2 | 62.4 | 90 | 290 | 2.71 (2.18 - 3.33) | 4.65 (4.13 - 5.22) |
| DAG | 5,039 | 1,014 | 4.1 | 0.8 | 618 | 123 | 152 (140 - 164) | 160 (133 - 191) |
| ODOD | 20,061 | 52,049 | 12.4 | 38.6 | 1,280 | 3,854 | 104 (97.9 - 109) | 99.8 (96.7 - 103) |
| MDCR | 9,004 | 18,079 | 8.0 | 17.3 | 10 | 74 | 1.25 (0.60 - 2.29) | 4.27 (3.35 - 5.36) |
| MDCD | 4,087 | 9,851 | 2.4 | 5.9 | 0 | 0 | | |
| Open Claims | 422,431 | 903,346 | 313.0 | 831.5 | 0 | 0 | | |
| OEHR | 36,445 | 95,879 | 14.7 | 46.3 | 0 | 0 | | |
| VA | 0 | 0 | 0.0 | 0.0 | 0 | 0 | | |
| SIDIAP | 4,064 | 1,774 | 4.9 | 2.4 | 1,302 | 569 | 266 (251 - 280) | 238 (218 - 258) |
| Meta-analysis | 85,182 | 157,334 | 62.6 | 121.5 | 3,300 | 4,910 | 52.7 (51.0 - 54.6) | 40.4 (39.3 - 41.6) |
| Meta-analysis (w/o USOC) | 85,182 | 157,334 | 62.6 | 121.5 | 3,300 | 4,910 | 52.7 (51.0 - 54.6) | 40.4 (39.3 - 41.6) |
| **PS-stratification / on-treatment TAR without escalation** | | | | | | | | |
| CCAE | 52,351 | 85,573 | 37.1 | 63.2 | 108 | 451 | 2.91 (2.39 - 3.51) | 7.14 (6.50 - 7.83) |
| DAG | 5,066 | 1,195 | 4.1 | 0.9 | 625 | 152 | 153 (141 - 165) | 168 (142 - 197) |
| ODOD | 20,910 | 52,056 | 12.9 | 38.6 | 1,361 | 3,854 | 106 (100 - 112) | 99.8 (96.7 - 103) |
| MDCR | 9,515 | 18,352 | 8.5 | 17.6 | 11 | 164 | 1.29 (0.64 - 2.31) | 9.34 (7.97 - 10.9) |
| MDCD | 4,183 | 9,871 | 2.4 | 6.0 | 0 | 0 | | |
| Open Claims | 429,825 | 903,360 | 320.1 | 831.5 | 0 | 0 | | |
| OEHR | 38,289 | 95,909 | 15.6 | 46.3 | 0 | 0 | | |
| VA | 24,844 | 221,635 | 18.9 | 207.4 | 18 | 467 | 0.95 (0.56 - 1.50) | 2.25 (2.05 - 2.47) |
| SIDIAP | 4,096 | 2,569 | 4.9 | 3.6 | 1,312 | 828 | 266 (252 - 281) | 230 (214 - 246) |
| Meta-analysis | 91,938 | 159,745 | 67.5 | 123.8 | 3,417 | 5,449 | 50.6 (48.9 - 52.3) | 44.0 (42.8 - 45.2) |
| Meta-analysis (w/o USOC) | 91,938 | 159,745 | 67.5 | 123.8 | 3,417 | 5,449 | 50.6 (48.9 - 52.3) | 44.0 (42.8 - 45.2) |

##### 4.6.6 Hospitalization with heart failure

**Supplementary Table 425:** Exposures, outcomes and risk estimates for Hospitalization with heart failure in DPP4I (male) (T) vs SU (male) (C) new-user cohorts under four different propensity score (PS) adjustment / time-at-risk (TAR) designs across data sources. We report total patient exposure time in 1,000 patient-year (KYr) units, uncalibrated and calibrated hazard ratios (HRs) and their 95% confidence intervals (CIs). Greyed out entries did not pass study diagnostics and are included only for completeness.

| | Patients | | Exposure (KYr) | | Outcomes | | Incidence (per KYr) | |
|---|---|---|---|---|---|---|---|---|
| | T | C | T | C | T | C | T | C |
| **PS-matching / on-treatment TAR** | | | | | | | | |
| CCAE | 46,539 | 83,340 | 40.7 | 71.0 | 238 | 531 | 5.85 (5.13 - 6.64) | 7.48 (6.86 - 8.14) |
| DAG | 13,419 | 2,361 | 14.3 | 2.4 | 0 | 0 | | |
| ODOD | 27,628 | 66,531 | 23.1 | 61.6 | 428 | 1,697 | 18.5 (16.8 - 20.3) | 27.5 (26.2 - 28.9) |
| MDCR | 8,253 | 16,385 | 8.5 | 17.1 | 272 | 657 | 31.9 (28.2 - 35.9) | 38.4 (35.5 - 41.5) |
| MDCD | 3,699 | 8,827 | 2.8 | 6.3 | 62 | 187 | 22.1 (17.0 - 28.4) | 29.7 (25.6 - 34.3) |
| Open Claims | 403,516 | 860,385 | 384.8 | 930.1 | 4,057 | 12,803 | 10.5 (10.2 - 10.9) | 13.8 (13.5 - 14.0) |
| OEHR | 35,630 | 93,613 | 17.4 | 51.1 | 225 | 920 | 12.9 (11.3 - 14.7) | 18.0 (16.8 - 19.2) |
| VA | 0 | 0 | 0.0 | 0.0 | 0 | 0 | | |
| SIDIAP | 26,675 | 7,201 | 78.1 | 20.5 | 108 | 25 | 1.38 (1.13 - 1.67) | 1.22 (0.79 - 1.80) |
| Meta-analysis | 551,940 | 1,136,282 | 555.5 | 1157.7 | 5,390 | 16,820 | 9.70 (9.45 - 9.97) | 14.5 (14.3 - 14.8) |
| Meta-analysis (w/o USOC) | 148,424 | 275,897 | 170.8 | 227.6 | 1,333 | 4,017 | 7.81 (7.39 - 8.24) | 17.6 (17.1 - 18.2) |
| **PS-stratification / on-treatment TAR** | | | | | | | | |
| CCAE | 51,915 | 84,822 | 45.6 | 72.2 | 256 | 536 | 5.62 (4.95 - 6.35) | 7.42 (6.81 - 8.08) |
| DAG | 13,447 | 2,736 | 14.3 | 2.8 | 0 | 0 | | |
| ODOD | 29,198 | 66,535 | 24.6 | 61.6 | 448 | 1,697 | 18.2 (16.6 - 20.0) | 27.5 (26.2 - 28.9) |
| MDCR | 8,741 | 16,748 | 9.1 | 17.5 | 284 | 671 | 31.1 (27.6 - 35.0) | 38.2 (35.4 - 41.3) |
| MDCD | 3,781 | 8,844 | 2.9 | 6.3 | 62 | 187 | 21.7 (16.6 - 27.8) | 29.7 (25.6 - 34.2) |
| Open Claims | 411,051 | 860,397 | 394.2 | 930.1 | 4,180 | 12,803 | 10.6 (10.3 - 10.9) | 13.8 (13.5 - 14.0) |
| OEHR | 37,434 | 93,642 | 18.5 | 51.2 | 239 | 920 | 12.9 (11.3 - 14.6) | 18.0 (16.8 - 19.2) |
| VA | 24,760 | 225,345 | 23.1 | 237.6 | 201 | 2,899 | 8.71 (7.55 - 10.00) | 12.2 (11.8 - 12.7) |
| SIDIAP | 26,764 | 9,253 | 78.2 | 27.6 | 108 | 33 | 1.38 (1.13 - 1.67) | 1.20 (0.82 - 1.68) |
| Meta-analysis | 568,884 | 1,140,241 | 573.1 | 1166.5 | 5,577 | 16,847 | 9.73 (9.48 - 9.99) | 14.4 (14.2 - 14.7) |
| Meta-analysis (w/o USOC) | 157,833 | 279,844 | 178.9 | 236.5 | 1,397 | 4,044 | 7.81 (7.41 - 8.23) | 17.1 (16.6 - 17.6) |
| **PS-matching / on-treatment TAR without escalation** | | | | | | | | |
| CCAE | 46,539 | 83,340 | 32.9 | 61.8 | 194 | 454 | 5.90 (5.10 - 6.79) | 7.35 (6.69 - 8.06) |
| DAG | 13,419 | 2,361 | 12.9 | 2.2 | 0 | 0 | | |
| ODOD | 27,628 | 66,531 | 19.0 | 55.0 | 356 | 1,497 | 18.8 (16.9 - 20.8) | 27.2 (25.8 - 28.6) |
| MDCR | 8,253 | 16,385 | 7.3 | 15.6 | 235 | 601 | 32.2 (28.2 - 36.6) | 38.5 (35.5 - 41.7) |
| MDCD | 3,699 | 8,827 | 2.1 | 5.3 | 51 | 156 | 23.9 (17.8 - 31.4) | 29.6 (25.1 - 34.6) |
| Open Claims | 403,516 | 860,385 | 298.5 | 788.6 | 3,166 | 10,900 | 10.6 (10.2 - 11.0) | 13.8 (13.6 - 14.1) |
| OEHR | 35,630 | 93,613 | 14.2 | 44.8 | 164 | 704 | 11.5 (9.83 - 13.4) | 15.7 (14.6 - 16.9) |
| VA | 0 | 0 | 0.0 | 0.0 | 0 | 0 | | |
| SIDIAP | 26,675 | 7,201 | 57.2 | 17.1 | 79 | 22 | 1.38 (1.09 - 1.72) | 1.29 (0.81 - 1.95) |
| Meta-analysis | 551,940 | 1,136,282 | 431.2 | 988.1 | 4,245 | 14,334 | 9.85 (9.55 - 10.1) | 14.5 (14.3 - 14.7) |
| Meta-analysis (w/o USOC) | 148,424 | 275,897 | 132.7 | 199.5 | 1,079 | 3,434 | 8.13 (7.65 - 8.63) | 17.2 (16.6 - 17.8) |
| **PS-stratification / on-treatment TAR without escalation** | | | | | | | | |
| CCAE | 51,915 | 84,822 | 36.8 | 62.9 | 209 | 459 | 5.67 (4.93 - 6.50) | 7.30 (6.64 - 8.00) |
| DAG | 13,447 | 2,736 | 12.9 | 2.6 | 0 | 0 | | |
| ODOD | 29,198 | 66,535 | 20.1 | 55.0 | 374 | 1,497 | 18.6 (16.8 - 20.6) | 27.2 (25.8 - 28.6) |
| MDCR | 8,741 | 16,748 | 7.8 | 16.1 | 246 | 614 | 31.6 (27.7 - 35.8) | 38.3 (35.3 - 41.4) |
| MDCD | 3,781 | 8,844 | 2.2 | 5.3 | 51 | 156 | 23.5 (17.5 - 30.8) | 29.5 (25.1 - 34.5) |
| Open Claims | 411,051 | 860,397 | 305.6 | 788.6 | 3,260 | 10,900 | 10.7 (10.3 - 11.0) | 13.8 (13.6 - 14.1) |
| OEHR | 37,434 | 93,642 | 15.1 | 44.8 | 176 | 704 | 11.6 (9.99 - 13.5) | 15.7 (14.6 - 16.9) |
| VA | 24,760 | 225,345 | 18.8 | 212.0 | 158 | 2,488 | 8.42 (7.16 - 9.84) | 11.7 (11.3 - 12.2) |
| SIDIAP | 26,764 | 9,253 | 57.3 | 22.8 | 79 | 27 | 1.38 (1.09 - 1.72) | 1.18 (0.78 - 1.72) |
| Meta-analysis | 568,884 | 1,140,241 | 444.9 | 995.5 | 4,395 | 14,357 | 9.88 (9.59 - 10.2) | 14.4 (14.2 - 14.7) |
| Meta-analysis (w/o USOC) | 157,833 | 279,844 | 139.3 | 206.8 | 1,135 | 3,457 | 8.15 (7.68 - 8.63) | 16.7 (16.2 - 17.3) |

##### 4.6.7 Measured renal dysfunction

**Supplementary Table 426:** Exposures, outcomes and risk estimates for Measured renal dysfunction in DPP4I (male) (T) vs SU (male) (C) new-user cohorts under four different propensity score (PS) adjustment / time-at-risk (TAR) designs across data sources. We report total patient exposure time in 1,000 patient-year (KYr) units, uncalibrated and calibrated hazard ratios (HRs) and their 95% confidence intervals (CIs). Greyed out entries did not pass study diagnostics and are included only for completeness.

| | Patients | | Exposure (KYr) | | Outcomes | | Incidence (per KYr) | |
|---|---|---|---|---|---|---|---|---|
| | T | C | T | C | T | C | T | C |
| **PS-matching / on-treatment TAR** | | | | | | | | |
| CCAE | 47,371 | 85,067 | 41.5 | 72.6 | 14 | 30 | 0.34 (0.18 - 0.57) | 0.41 (0.28 - 0.59) |
| DAG | 13,419 | 2,361 | 14.3 | 2.4 | 0 | 0 | | |
| ODOD | 29,183 | 70,640 | 24.5 | 66.4 | 51 | 146 | 2.08 (1.55 - 2.74) | 2.20 (1.86 - 2.59) |
| MDCR | 9,055 | 18,278 | 9.4 | 19.3 | <5 | 8 | < 0.53 (0.17 - 1.24) | 0.42 (0.18 - 0.82) |
| MDCD | 4,087 | 9,851 | 3.1 | 7.1 | 0 | 0 | | |
| Open Claims | 422,431 | 903,346 | 403.8 | 982.0 | 0 | 0 | | |
| OEHR | 36,445 | 95,879 | 18.1 | 53.4 | 0 | 0 | | |
| VA | 0 | 0 | 0.0 | 0.0 | 0 | 0 | | |
| SIDIAP | 26,580 | 7,201 | 77.4 | 20.4 | 421 | 28 | 5.44 (4.93 - 5.98) | 1.37 (0.91 - 1.98) |
| Meta-analysis | 103,134 | 162,908 | 143.4 | 159.4 | 486 | 204 | 3.39 (3.09 - 3.70) | 1.28 (1.11 - 1.47) |
| Meta-analysis (w/o USOC) | 103,134 | 162,908 | 143.4 | 159.4 | 486 | 204 | 3.39 (3.09 - 3.70) | 1.28 (1.11 - 1.47) |
| **PS-stratification / on-treatment TAR** | | | | | | | | |
| CCAE | 52,816 | 86,589 | 46.4 | 73.9 | 17 | 30 | 0.37 (0.21 - 0.59) | 0.41 (0.27 - 0.58) |
| DAG | 13,447 | 2,736 | 14.3 | 2.8 | 0 | 0 | | |
| ODOD | 30,796 | 70,646 | 26.0 | 66.4 | 54 | 146 | 2.08 (1.56 - 2.71) | 2.20 (1.86 - 2.59) |
| MDCR | 9,568 | 18,672 | 10.0 | 19.7 | <5 | 8 | < 0.50 (0.16 - 1.16) | 0.41 (0.17 - 0.80) |
| MDCD | 4,183 | 9,871 | 3.2 | 7.1 | 0 | 0 | | |
| Open Claims | 429,825 | 903,360 | 413.2 | 982.0 | 0 | 0 | | |
| OEHR | 38,289 | 95,909 | 19.2 | 53.4 | 0 | 0 | | |
| VA | 25,292 | 229,407 | 23.6 | 243.1 | 98 | 1,265 | 4.15 (3.37 - 5.05) | 5.20 (4.92 - 5.50) |
| SIDIAP | 26,665 | 9,253 | 77.5 | 27.6 | 424 | 40 | 5.47 (4.96 - 6.02) | 1.45 (1.04 - 1.98) |
| Meta-analysis | 110,277 | 166,488 | 149.9 | 167.8 | 495 | 216 | 3.30 (3.02 - 3.61) | 1.29 (1.12 - 1.47) |
| Meta-analysis (w/o USOC) | 110,277 | 166,488 | 149.9 | 167.8 | 495 | 216 | 3.30 (3.02 - 3.61) | 1.29 (1.12 - 1.47) |
| **PS-matching / on-treatment TAR without escalation** | | | | | | | | |
| CCAE | 47,371 | 85,067 | 33.5 | 63.1 | 11 | 26 | 0.33 (0.16 - 0.59) | 0.41 (0.27 - 0.60) |
| DAG | 13,419 | 2,361 | 12.9 | 2.2 | 0 | 0 | | |
| ODOD | 29,183 | 70,640 | 20.0 | 59.1 | 45 | 126 | 2.24 (1.64 - 3.00) | 2.13 (1.78 - 2.54) |
| MDCR | 9,055 | 18,278 | 8.1 | 17.6 | 0 | 7 | | 0.40 (0.16 - 0.82) |
| MDCD | 4,087 | 9,851 | 2.4 | 5.9 | 0 | 0 | | |
| Open Claims | 422,431 | 903,346 | 313.0 | 831.5 | 0 | 0 | | |
| OEHR | 36,445 | 95,879 | 14.7 | 46.3 | 0 | 0 | | |
| VA | 0 | 0 | 0.0 | 0.0 | 0 | 0 | | |
| SIDIAP | 26,580 | 7,201 | 56.7 | 17.1 | 320 | 19 | 5.65 (5.04 - 6.30) | 1.11 (0.67 - 1.74) |
| Meta-analysis | 103,134 | 162,908 | 110.2 | 139.3 | 376 | 171 | 3.41 (3.07 - 3.77) | 1.23 (1.05 - 1.43) |
| Meta-analysis (w/o USOC) | 103,134 | 162,908 | 110.2 | 139.3 | 376 | 171 | 3.41 (3.07 - 3.77) | 1.23 (1.05 - 1.43) |
| **PS-stratification / on-treatment TAR without escalation** | | | | | | | | |
| CCAE | 52,816 | 86,589 | 37.5 | 64.3 | 13 | 26 | 0.35 (0.18 - 0.59) | 0.40 (0.26 - 0.59) |
| DAG | 13,447 | 2,736 | 12.9 | 2.6 | 0 | 0 | | |
| ODOD | 30,796 | 70,646 | 21.2 | 59.1 | 48 | 126 | 2.26 (1.67 - 3.00) | 2.13 (1.77 - 2.54) |
| MDCR | 9,568 | 18,672 | 8.6 | 18.0 | 0 | 7 | | 0.39 (0.16 - 0.80) |
| MDCD | 4,183 | 9,871 | 2.4 | 6.0 | 0 | 0 | | |
| Open Claims | 429,825 | 903,360 | 320.1 | 831.5 | 0 | 0 | | |
| OEHR | 38,289 | 95,909 | 15.6 | 46.3 | 0 | 0 | | |
| VA | 25,292 | 229,407 | 19.2 | 216.6 | 77 | 1,037 | 4.01 (3.17 - 5.02) | 4.79 (4.50 - 5.09) |
| SIDIAP | 26,665 | 9,253 | 56.8 | 22.8 | 323 | 26 | 5.69 (5.09 - 6.35) | 1.14 (0.75 - 1.67) |
| Meta-analysis | 110,277 | 166,488 | 115.5 | 146.2 | 384 | 178 | 3.32 (3.00 - 3.67) | 1.22 (1.05 - 1.41) |
| Meta-analysis (w/o USOC) | 110,277 | 166,488 | 115.5 | 146.2 | 384 | 178 | 3.32 (3.00 - 3.67) | 1.22 (1.05 - 1.41) |

##### 4.6.8 Stroke

**Supplementary Table 427:** Exposures, outcomes and risk estimates for Stroke in DPP4I (male) (T) vs SU (male) (C) new-user cohorts under four different propensity score (PS) adjustment / time-at-risk (TAR) designs across data sources. We report total patient exposure time in 1,000 patient-year (KYr) units, uncalibrated and calibrated hazard ratios (HRs) and their 95% confidence intervals (CIs). Greyed out entries did not pass study diagnostics and are included only for completeness.

| | Patients | | Exposure (KYr) | | Outcomes | | Incidence (per KYr) | |
|---|---|---|---|---|---|---|---|---|
| | T | C | T | C | T | C | T | C |
| **PS-matching / on-treatment TAR** | | | | | | | | |
| CCAE | 46,880 | 83,957 | 41.1 | 71.6 | 138 | 265 | 3.36 (2.82 - 3.97) | 3.70 (3.27 - 4.17) |
| DAG | 13,419 | 2,361 | 14.3 | 2.4 | 0 | 0 | | |
| ODOD | 28,456 | 68,809 | 23.9 | 64.3 | 212 | 769 | 8.87 (7.72 - 10.1) | 12.0 (11.1 - 12.8) |
| MDCR | 8,667 | 17,383 | 9.0 | 18.2 | 133 | 318 | 14.8 (12.4 - 17.5) | 17.5 (15.6 - 19.5) |
| MDCD | 3,910 | 9,416 | 3.0 | 6.7 | 33 | 100 | 11.1 (7.63 - 15.6) | 14.8 (12.1 - 18.1) |
| Open Claims | 410,222 | 876,424 | 392.0 | 951.0 | 2,009 | 6,194 | 5.13 (4.90 - 5.35) | 6.51 (6.35 - 6.68) |
| OEHR | 36,122 | 94,871 | 17.9 | 52.5 | 82 | 296 | 4.58 (3.65 - 5.69) | 5.64 (5.01 - 6.32) |
| VA | 0 | 0 | 0.0 | 0.0 | 0 | 0 | | |
| SIDIAP | 26,445 | 7,160 | 77.2 | 20.3 | 222 | 33 | 2.87 (2.51 - 3.28) | 1.63 (1.12 - 2.28) |
| Meta-analysis | 560,702 | 1,158,020 | 564.0 | 1184.8 | 2,829 | 7,975 | 5.02 (4.83 - 5.20) | 6.73 (6.58 - 6.88) |
| Meta-analysis (w/o USOC) | 150,480 | 281,596 | 172.1 | 233.7 | 820 | 1,781 | 4.77 (4.45 - 5.10) | 7.62 (7.27 - 7.98) |
| **PS-stratification / on-treatment TAR** | | | | | | | | |
| CCAE | 52,301 | 85,457 | 46.0 | 72.9 | 152 | 268 | 3.31 (2.80 - 3.88) | 3.68 (3.25 - 4.14) |
| DAG | 13,447 | 2,736 | 14.3 | 2.8 | 0 | 0 | | |
| ODOD | 30,047 | 68,815 | 25.4 | 64.3 | 223 | 770 | 8.79 (7.68 - 10.0) | 12.0 (11.1 - 12.8) |
| MDCR | 9,165 | 17,767 | 9.6 | 18.7 | 141 | 324 | 14.7 (12.4 - 17.4) | 17.3 (15.5 - 19.3) |
| MDCD | 3,997 | 9,434 | 3.1 | 6.8 | 33 | 100 | 10.8 (7.44 - 15.2) | 14.8 (12.1 - 18.0) |
| Open Claims | 417,616 | 876,438 | 401.3 | 951.0 | 2,067 | 6,194 | 5.15 (4.93 - 5.38) | 6.51 (6.35 - 6.68) |
| OEHR | 37,946 | 94,901 | 19.0 | 52.5 | 87 | 296 | 4.58 (3.67 - 5.65) | 5.64 (5.01 - 6.32) |
| VA | 25,218 | 230,647 | 23.6 | 245.2 | 64 | 859 | 2.71 (2.09 - 3.46) | 3.50 (3.27 - 3.75) |
| SIDIAP | 26,532 | 9,197 | 77.3 | 27.4 | 222 | 47 | 2.87 (2.51 - 3.27) | 1.72 (1.26 - 2.28) |
| Meta-analysis | 577,604 | 1,162,009 | 581.6 | 1193.6 | 2,925 | 7,999 | 5.03 (4.85 - 5.22) | 6.70 (6.56 - 6.85) |
| Meta-analysis (w/o USOC) | 159,988 | 285,571 | 180.3 | 242.6 | 858 | 1,805 | 4.76 (4.45 - 5.09) | 7.44 (7.10 - 7.79) |
| **PS-matching / on-treatment TAR without escalation** | | | | | | | | |
| CCAE | 46,880 | 83,957 | 33.1 | 62.3 | 108 | 239 | 3.26 (2.67 - 3.93) | 3.84 (3.37 - 4.36) |
| DAG | 13,419 | 2,361 | 12.9 | 2.2 | 0 | 0 | | |
| ODOD | 28,456 | 68,809 | 19.5 | 57.3 | 186 | 686 | 9.52 (8.20 - 11.0) | 12.0 (11.1 - 12.9) |
| MDCR | 8,667 | 17,383 | 7.7 | 16.6 | 113 | 290 | 14.7 (12.1 - 17.7) | 17.5 (15.5 - 19.6) |
| MDCD | 3,910 | 9,416 | 2.3 | 5.6 | 23 | 81 | 10.2 (6.46 - 15.3) | 14.4 (11.4 - 17.8) |
| Open Claims | 410,222 | 876,424 | 303.9 | 805.4 | 1,601 | 5,349 | 5.27 (5.01 - 5.53) | 6.64 (6.46 - 6.82) |
| OEHR | 36,122 | 94,871 | 14.5 | 45.6 | 60 | 225 | 4.14 (3.16 - 5.32) | 4.93 (4.31 - 5.62) |
| VA | 0 | 0 | 0.0 | 0.0 | 0 | 0 | | |
| SIDIAP | 26,445 | 7,160 | 56.6 | 17.0 | 148 | 31 | 2.61 (2.21 - 3.07) | 1.83 (1.24 - 2.60) |
| Meta-analysis | 560,702 | 1,158,020 | 437.7 | 1009.8 | 2,239 | 6,901 | 5.12 (4.91 - 5.33) | 6.83 (6.67 - 7.00) |
| Meta-analysis (w/o USOC) | 150,480 | 281,596 | 133.7 | 204.4 | 638 | 1,552 | 4.77 (4.41 - 5.16) | 7.59 (7.22 - 7.98) |
| **PS-stratification / on-treatment TAR without escalation** | | | | | | | | |
| CCAE | 52,301 | 85,457 | 37.1 | 63.4 | 121 | 242 | 3.26 (2.70 - 3.89) | 3.82 (3.35 - 4.33) |
| DAG | 13,447 | 2,736 | 12.9 | 2.6 | 0 | 0 | | |
| ODOD | 30,047 | 68,815 | 20.7 | 57.3 | 194 | 687 | 9.37 (8.10 - 10.8) | 12.0 (11.1 - 12.9) |
| MDCR | 9,165 | 17,767 | 8.2 | 17.0 | 118 | 296 | 14.4 (11.9 - 17.3) | 17.4 (15.4 - 19.5) |
| MDCD | 3,997 | 9,434 | 2.3 | 5.7 | 23 | 81 | 9.95 (6.31 - 14.9) | 14.3 (11.4 - 17.8) |
| Open Claims | 417,616 | 876,438 | 311.0 | 805.4 | 1,640 | 5,349 | 5.27 (5.02 - 5.53) | 6.64 (6.46 - 6.82) |
| OEHR | 37,946 | 94,901 | 15.4 | 45.6 | 64 | 225 | 4.15 (3.20 - 5.30) | 4.93 (4.31 - 5.62) |
| VA | 25,218 | 230,647 | 19.2 | 218.4 | 41 | 740 | 2.14 (1.53 - 2.90) | 3.39 (3.15 - 3.64) |
| SIDIAP | 26,532 | 9,197 | 56.7 | 22.7 | 148 | 40 | 2.61 (2.21 - 3.07) | 1.77 (1.26 - 2.40) |
| Meta-analysis | 577,604 | 1,162,009 | 451.4 | 1017.1 | 2,308 | 6,920 | 5.11 (4.91 - 5.33) | 6.80 (6.64 - 6.97) |
| Meta-analysis (w/o USOC) | 159,988 | 285,571 | 140.4 | 211.7 | 668 | 1,571 | 4.76 (4.40 - 5.13) | 7.42 (7.06 - 7.80) |

##### 4.6.9 Sudden cardiac death

**Supplementary Table 428:** Exposures, outcomes and risk estimates for Sudden cardiac death in DPP4I (male) (T) vs SU (male) (C) new-user cohorts under four different propensity score (PS) adjustment / time-at-risk (TAR) designs across data sources. We report total patient exposure time in 1,000 patient-year (KYr) units, uncalibrated and calibrated hazard ratios (HRs) and their 95% confidence intervals (CIs). Greyed out entries did not pass study diagnostics and are included only for completeness.

| | Patients | | Exposure (KYr) | | Outcomes | | Incidence (per KYr) | |
|---|---|---|---|---|---|---|---|---|
| | T | C | T | C | T | C | T | C |
| **PS-matching / on-treatment TAR** | | | | | | | | |
| CCAE | 47,385 | 85,092 | 41.5 | 72.7 | 35 | 111 | 0.84 (0.59 - 1.17) | 1.53 (1.26 - 1.84) |
| DAG | 13,419 | 2,361 | 14.3 | 2.4 | 0 | 0 | | |
| ODOD | 29,198 | 70,628 | 24.5 | 66.4 | 83 | 253 | 3.38 (2.69 - 4.19) | 3.81 (3.35 - 4.31) |
| MDCR | 9,027 | 18,209 | 9.4 | 19.2 | 40 | 69 | 4.25 (3.03 - 5.78) | 3.60 (2.80 - 4.55) |
| MDCD | 4,070 | 9,811 | 3.1 | 7.1 | 12 | 32 | 3.86 (1.99 - 6.74) | 4.53 (3.10 - 6.39) |
| Open Claims | 421,389 | 901,146 | 402.7 | 979.2 | 635 | 1,870 | 1.58 (1.46 - 1.70) | 1.91 (1.82 - 2.00) |
| OEHR | 36,417 | 95,802 | 18.1 | 53.3 | 29 | 104 | 1.60 (1.07 - 2.30) | 1.95 (1.59 - 2.36) |
| VA | 0 | 0 | 0.0 | 0.0 | 0 | 0 | | |
| SIDIAP | 26,792 | 7,215 | 78.5 | 20.5 | 5 | <5 | 0.06 (0.02 - 0.15) | < 0.24 (0.08 - 0.57) |
| Meta-analysis | 547,486 | 1,180,688 | 499.3 | 1197.8 | 834 | 2,439 | 1.67 (1.56 - 1.79) | 2.04 (1.96 - 2.12) |
| Meta-analysis (w/o USOC) | 126,097 | 279,542 | 96.7 | 218.6 | 199 | 569 | 2.06 (1.78 - 2.37) | 2.60 (2.39 - 2.83) |
| **PS-stratification / on-treatment TAR** | | | | | | | | |
| CCAE | 52,834 | 86,613 | 46.5 | 73.9 | 36 | 112 | 0.77 (0.54 - 1.07) | 1.52 (1.25 - 1.82) |
| DAG | 13,447 | 2,736 | 14.3 | 2.8 | 0 | 0 | | |
| ODOD | 30,801 | 70,635 | 26.0 | 66.4 | 87 | 253 | 3.34 (2.68 - 4.12) | 3.81 (3.35 - 4.31) |
| MDCR | 9,537 | 18,603 | 10.0 | 19.7 | 40 | 71 | 3.99 (2.85 - 5.44) | 3.61 (2.82 - 4.55) |
| MDCD | 4,163 | 9,831 | 3.2 | 7.1 | 12 | 32 | 3.76 (1.94 - 6.56) | 4.52 (3.09 - 6.38) |
| Open Claims | 428,796 | 901,160 | 412.0 | 979.2 | 650 | 1,870 | 1.58 (1.46 - 1.70) | 1.91 (1.82 - 2.00) |
| OEHR | 38,259 | 95,832 | 19.2 | 53.3 | 30 | 104 | 1.56 (1.05 - 2.23) | 1.95 (1.59 - 2.36) |
| VA | 25,610 | 233,587 | 24.0 | 248.8 | <10 | 132 | < 0.42 (0.20 - 0.77) | 0.53 (0.44 - 0.63) |
| SIDIAP | 26,884 | 9,267 | 78.6 | 27.7 | 5 | <5 | 0.06 (0.02 - 0.15) | < 0.18 (0.06 - 0.42) |
| Meta-analysis | 564,390 | 1,182,674 | 516.9 | 1199.6 | 855 | 2,442 | 1.65 (1.55 - 1.77) | 2.04 (1.96 - 2.12) |
| Meta-analysis (w/o USOC) | 135,594 | 281,514 | 104.9 | 220.4 | 205 | 572 | 1.95 (1.70 - 2.24) | 2.60 (2.39 - 2.82) |
| **PS-matching / on-treatment TAR without escalation** | | | | | | | | |
| CCAE | 47,385 | 85,092 | 33.5 | 63.1 | 29 | 96 | 0.87 (0.58 - 1.24) | 1.52 (1.23 - 1.86) |
| DAG | 13,419 | 2,361 | 12.9 | 2.2 | 0 | 0 | | |
| ODOD | 29,198 | 70,628 | 20.1 | 59.2 | 74 | 223 | 3.69 (2.89 - 4.63) | 3.77 (3.29 - 4.30) |
| MDCR | 9,027 | 18,209 | 8.0 | 17.5 | 27 | 58 | 3.36 (2.22 - 4.89) | 3.31 (2.52 - 4.28) |
| MDCD | 4,070 | 9,811 | 2.4 | 5.9 | 9 | 27 | 3.80 (1.74 - 7.21) | 4.56 (3.01 - 6.64) |
| Open Claims | 421,389 | 901,146 | 312.2 | 829.2 | 490 | 1,586 | 1.57 (1.43 - 1.72) | 1.91 (1.82 - 2.01) |
| OEHR | 36,417 | 95,802 | 14.6 | 46.2 | 19 | 73 | 1.30 (0.78 - 2.03) | 1.58 (1.24 - 1.98) |
| VA | 0 | 0 | 0.0 | 0.0 | 0 | 0 | | |
| SIDIAP | 26,792 | 7,215 | 57.5 | 17.1 | <5 | <5 | < 0.09 (0.03 - 0.20) | < 0.29 (0.09 - 0.68) |
| Meta-analysis | 547,486 | 1,180,688 | 390.8 | 1021.1 | 648 | 2,063 | 1.66 (1.53 - 1.79) | 2.02 (1.93 - 2.11) |
| Meta-analysis (w/o USOC) | 126,097 | 279,542 | 78.6 | 192.0 | 158 | 477 | 2.01 (1.71 - 2.35) | 2.48 (2.27 - 2.72) |
| **PS-stratification / on-treatment TAR without escalation** | | | | | | | | |
| CCAE | 52,834 | 86,613 | 37.6 | 64.3 | 30 | 97 | 0.80 (0.54 - 1.14) | 1.51 (1.22 - 1.84) |
| DAG | 13,447 | 2,736 | 12.9 | 2.6 | 0 | 0 | | |
| ODOD | 30,801 | 70,635 | 21.3 | 59.2 | 78 | 223 | 3.67 (2.90 - 4.58) | 3.77 (3.29 - 4.30) |
| MDCR | 9,537 | 18,603 | 8.5 | 18.0 | 27 | 60 | 3.16 (2.08 - 4.60) | 3.34 (2.55 - 4.30) |
| MDCD | 4,163 | 9,831 | 2.4 | 5.9 | 9 | 27 | 3.71 (1.70 - 7.04) | 4.55 (3.00 - 6.63) |
| Open Claims | 428,796 | 901,160 | 319.2 | 829.2 | 501 | 1,586 | 1.57 (1.44 - 1.71) | 1.91 (1.82 - 2.01) |
| OEHR | 38,259 | 95,832 | 15.6 | 46.3 | 20 | 73 | 1.28 (0.78 - 1.98) | 1.58 (1.24 - 1.98) |
| VA | 25,610 | 233,587 | 19.5 | 221.5 | <10 | 100 | < 0.51 (0.25 - 0.94) | 0.45 (0.37 - 0.55) |
| SIDIAP | 26,884 | 9,267 | 57.6 | 22.8 | <5 | <5 | < 0.09 (0.03 - 0.20) | < 0.22 (0.07 - 0.51) |
| Meta-analysis | 564,390 | 1,182,674 | 404.5 | 1022.8 | 665 | 2,066 | 1.64 (1.52 - 1.77) | 2.02 (1.93 - 2.11) |
| Meta-analysis (w/o USOC) | 135,594 | 281,514 | 85.3 | 193.6 | 164 | 480 | 1.92 (1.64 - 2.24) | 2.48 (2.26 - 2.71) |

##### 4.6.10 Abnormal weight gain

**Supplementary Table 429:** Exposures, outcomes and risk estimates for Abnormal weight gain in DPP4I (male) (T) vs SU (male) (C) new-user cohorts under four different propensity score (PS) adjustment / time-at-risk (TAR) designs across data sources. We report total patient exposure time in 1,000 patient-year (KYr) units, uncalibrated and calibrated hazard ratios (HRs) and their 95% confidence intervals (CIs). Greyed out entries did not pass study diagnostics and are included only for completeness.

| | Patients | | Exposure (KYr) | | Outcomes | | Incidence (per KYr) | |
|---|---|---|---|---|---|---|---|---|
| | T | C | T | C | T | C | T | C |
| **PS-matching / on-treatment TAR** | | | | | | | | |
| CCAE | 46,233 | 83,342 | 40.5 | 71.0 | 132 | 292 | 3.26 (2.73 - 3.87) | 4.12 (3.66 - 4.62) |
| DAG | 13,346 | 2,345 | 14.2 | 2.4 | 16 | <5 | 1.13 (0.64 - 1.83) | < 2.07 (0.67 - 4.82) |
| ODOD | 28,438 | 69,041 | 23.8 | 64.6 | 143 | 374 | 6.02 (5.07 - 7.09) | 5.79 (5.21 - 6.40) |
| MDCR | 8,956 | 18,124 | 9.4 | 19.0 | 19 | 68 | 2.03 (1.22 - 3.17) | 3.57 (2.78 - 4.53) |
| MDCD | 3,898 | 9,467 | 3.0 | 6.7 | 20 | 67 | 6.77 (4.13 - 10.5) | 9.93 (7.70 - 12.6) |
| Open Claims | 411,272 | 882,968 | 392.5 | 958.3 | 687 | 1,841 | 1.75 (1.62 - 1.89) | 1.92 (1.83 - 2.01) |
| OEHR | 35,591 | 93,969 | 17.6 | 52.0 | 78 | 286 | 4.44 (3.51 - 5.54) | 5.50 (4.88 - 6.18) |
| VA | 0 | 0 | 0.0 | 0.0 | 0 | 0 | | |
| SIDIAP | 26,515 | 7,179 | 77.6 | 20.3 | 105 | 37 | 1.35 (1.11 - 1.64) | 1.82 (1.28 - 2.51) |
| Meta-analysis | 560,903 | 1,164,090 | 564.2 | 1191.9 | 1,184 | 2,965 | 2.10 (1.98 - 2.22) | 2.49 (2.40 - 2.58) |
| Meta-analysis (w/o USOC) | 149,631 | 281,122 | 171.8 | 233.7 | 497 | 1,124 | 2.89 (2.64 - 3.16) | 4.81 (4.53 - 5.10) |
| **PS-stratification / on-treatment TAR** | | | | | | | | |
| CCAE | 51,500 | 84,920 | 45.3 | 72.3 | 152 | 292 | 3.36 (2.85 - 3.94) | 4.04 (3.59 - 4.53) |
| DAG | 13,373 | 2,715 | 14.2 | 2.8 | 16 | <5 | 1.12 (0.64 - 1.82) | < 1.77 (0.58 - 4.14) |
| ODOD | 29,960 | 69,047 | 25.1 | 64.6 | 157 | 374 | 6.24 (5.30 - 7.30) | 5.79 (5.21 - 6.40) |
| MDCR | 9,468 | 18,518 | 10.0 | 19.5 | 22 | 69 | 2.21 (1.38 - 3.35) | 3.54 (2.75 - 4.48) |
| MDCD | 3,988 | 9,487 | 3.0 | 6.8 | 22 | 67 | 7.25 (4.54 - 11.0) | 9.91 (7.68 - 12.6) |
| Open Claims | 418,304 | 882,982 | 401.3 | 958.3 | 710 | 1,841 | 1.77 (1.64 - 1.90) | 1.92 (1.83 - 2.01) |
| OEHR | 37,353 | 93,998 | 18.6 | 52.0 | 84 | 286 | 4.50 (3.59 - 5.58) | 5.50 (4.88 - 6.18) |
| VA | 25,119 | 230,220 | 23.5 | 244.6 | 11 | 407 | 0.47 (0.23 - 0.84) | 1.66 (1.51 - 1.83) |
| SIDIAP | 26,606 | 9,227 | 77.7 | 27.4 | 105 | 45 | 1.35 (1.10 - 1.64) | 1.64 (1.20 - 2.20) |
| Meta-analysis | 577,179 | 1,168,179 | 581.1 | 1200.9 | 1,252 | 2,974 | 2.15 (2.04 - 2.28) | 2.48 (2.39 - 2.57) |
| Meta-analysis (w/o USOC) | 158,875 | 285,197 | 179.8 | 242.6 | 542 | 1,133 | 3.02 (2.77 - 3.28) | 4.67 (4.40 - 4.95) |
| **PS-matching / on-treatment TAR without escalation** | | | | | | | | |
| CCAE | 46,233 | 83,342 | 32.7 | 61.7 | 105 | 251 | 3.21 (2.63 - 3.89) | 4.07 (3.58 - 4.60) |
| DAG | 13,346 | 2,345 | 12.8 | 2.2 | 14 | <5 | 1.09 (0.60 - 1.83) | < 2.26 (0.73 - 5.28) |
| ODOD | 28,438 | 69,041 | 19.5 | 57.7 | 110 | 326 | 5.64 (4.64 - 6.80) | 5.65 (5.05 - 6.30) |
| MDCR | 8,956 | 18,124 | 8.0 | 17.3 | 13 | 62 | 1.63 (0.87 - 2.78) | 3.57 (2.74 - 4.58) |
| MDCD | 3,898 | 9,467 | 2.3 | 5.7 | 13 | 49 | 5.77 (3.07 - 9.87) | 8.65 (6.40 - 11.4) |
| Open Claims | 411,272 | 882,968 | 304.8 | 812.4 | 517 | 1,524 | 1.70 (1.55 - 1.85) | 1.88 (1.78 - 1.97) |
| OEHR | 35,591 | 93,969 | 14.3 | 45.2 | 60 | 237 | 4.20 (3.21 - 5.41) | 5.24 (4.60 - 5.96) |
| VA | 0 | 0 | 0.0 | 0.0 | 0 | 0 | | |
| SIDIAP | 26,515 | 7,179 | 56.8 | 17.0 | 81 | 31 | 1.42 (1.13 - 1.77) | 1.83 (1.24 - 2.59) |
| Meta-analysis | 560,903 | 1,164,090 | 438.4 | 1017.0 | 899 | 2,480 | 2.05 (1.92 - 2.19) | 2.44 (2.34 - 2.54) |
| Meta-analysis (w/o USOC) | 149,631 | 281,122 | 133.6 | 204.6 | 382 | 956 | 2.86 (2.58 - 3.16) | 4.67 (4.38 - 4.98) |
| **PS-stratification / on-treatment TAR without escalation** | | | | | | | | |
| CCAE | 51,500 | 84,920 | 36.6 | 62.9 | 121 | 251 | 3.31 (2.74 - 3.95) | 3.99 (3.51 - 4.51) |
| DAG | 13,373 | 2,715 | 12.8 | 2.6 | 14 | <5 | 1.09 (0.60 - 1.83) | < 1.93 (0.63 - 4.52) |
| ODOD | 29,960 | 69,047 | 20.6 | 57.7 | 122 | 326 | 5.92 (4.92 - 7.07) | 5.65 (5.05 - 6.30) |
| MDCR | 9,468 | 18,518 | 8.5 | 17.8 | 16 | 63 | 1.88 (1.08 - 3.06) | 3.54 (2.72 - 4.53) |
| MDCD | 3,988 | 9,487 | 2.3 | 5.7 | 14 | 49 | 6.07 (3.32 - 10.2) | 8.63 (6.38 - 11.4) |
| Open Claims | 418,304 | 882,982 | 311.5 | 812.4 | 534 | 1,524 | 1.71 (1.57 - 1.87) | 1.88 (1.78 - 1.97) |
| OEHR | 37,353 | 93,998 | 15.2 | 45.2 | 66 | 237 | 4.35 (3.37 - 5.54) | 5.24 (4.60 - 5.95) |
| VA | 25,119 | 230,220 | 19.1 | 217.9 | <10 | 332 | < 0.52 (0.25 - 0.96) | 1.52 (1.36 - 1.70) |
| SIDIAP | 26,606 | 9,227 | 56.9 | 22.7 | 81 | 39 | 1.42 (1.13 - 1.77) | 1.72 (1.22 - 2.35) |
| Meta-analysis | 577,179 | 1,168,179 | 451.6 | 1024.4 | 954 | 2,489 | 2.11 (1.98 - 2.25) | 2.43 (2.34 - 2.53) |
| Meta-analysis (w/o USOC) | 158,875 | 285,197 | 140.1 | 212.0 | 420 | 965 | 3.00 (2.72 - 3.30) | 4.55 (4.27 - 4.85) |

##### 4.6.11 Abnormal weight loss

**Supplementary Table 430:** Exposures, outcomes and risk estimates for Abnormal weight loss in DPP4I (male) (T) vs SU (male) (C) new-user cohorts under four different propensity score (PS) adjustment / time-at-risk (TAR) designs across data sources. We report total patient exposure time in 1,000 patient-year (KYr) units, uncalibrated and calibrated hazard ratios (HRs) and their 95% confidence intervals (CIs). Greyed out entries did not pass study diagnostics and are included only for completeness.

| | Patients | | Exposure (KYr) | | Outcomes | | Incidence (per KYr) | |
|---|---|---|---|---|---|---|---|---|
| | T | C | T | C | T | C | T | C |
| **PS-matching / on-treatment TAR** | | | | | | | | |
| CCAE | 46,342 | 83,167 | 40.5 | 71.0 | 261 | 346 | 6.44 (5.68 - 7.27) | 4.87 (4.37 - 5.42) |
| DAG | 13,155 | 2,317 | 14.0 | 2.4 | 89 | 13 | 6.38 (5.12 - 7.85) | 5.48 (2.92 - 9.37) |
| ODOD | 27,995 | 67,645 | 23.3 | 63.4 | 353 | 834 | 15.1 (13.6 - 16.8) | 13.2 (12.3 - 14.1) |
| MDCR | 8,637 | 17,435 | 9.0 | 18.4 | 128 | 243 | 14.2 (11.8 - 16.9) | 13.2 (11.6 - 15.0) |
| MDCD | 3,890 | 9,392 | 3.0 | 6.7 | 45 | 81 | 15.2 (11.1 - 20.3) | 12.1 (9.58 - 15.0) |
| Open Claims | 407,051 | 871,924 | 388.0 | 946.2 | 2,452 | 4,960 | 6.32 (6.07 - 6.57) | 5.24 (5.10 - 5.39) |
| OEHR | 35,345 | 92,794 | 17.4 | 51.2 | 203 | 551 | 11.7 (10.1 - 13.4) | 10.8 (9.87 - 11.7) |
| VA | 0 | 0 | 0.0 | 0.0 | 0 | 0 | | |
| SIDIAP | 26,520 | 7,185 | 77.5 | 20.4 | 259 | 39 | 3.34 (2.95 - 3.77) | 1.91 (1.36 - 2.62) |
| Meta-analysis | 568,935 | 1,151,859 | 572.7 | 1179.7 | 3,790 | 7,067 | 6.62 (6.41 - 6.83) | 5.99 (5.85 - 6.13) |
| Meta-analysis (w/o USOC) | 161,884 | 279,935 | 184.7 | 233.5 | 1,338 | 2,107 | 7.24 (6.86 - 7.64) | 9.02 (8.64 - 9.42) |
| **PS-stratification / on-treatment TAR** | | | | | | | | |
| CCAE | 51,674 | 84,669 | 45.3 | 72.2 | 298 | 348 | 6.58 (5.85 - 7.37) | 4.82 (4.32 - 5.35) |
| DAG | 13,183 | 2,676 | 14.0 | 2.8 | 89 | 15 | 6.36 (5.11 - 7.83) | 5.43 (3.04 - 8.96) |
| ODOD | 29,544 | 67,651 | 24.8 | 63.4 | 379 | 834 | 15.3 (13.8 - 16.9) | 13.2 (12.3 - 14.1) |
| MDCR | 9,144 | 17,821 | 9.6 | 18.9 | 137 | 244 | 14.3 (12.0 - 16.9) | 12.9 (11.4 - 14.7) |
| MDCD | 3,979 | 9,412 | 3.0 | 6.7 | 46 | 81 | 15.2 (11.1 - 20.2) | 12.0 (9.56 - 15.0) |
| Open Claims | 414,330 | 871,935 | 397.1 | 946.3 | 2,536 | 4,960 | 6.39 (6.14 - 6.64) | 5.24 (5.10 - 5.39) |
| OEHR | 37,139 | 92,820 | 18.5 | 51.3 | 221 | 552 | 12.0 (10.4 - 13.7) | 10.8 (9.89 - 11.7) |
| VA | 24,656 | 226,229 | 23.0 | 240.4 | 212 | 1,476 | 9.21 (8.01 - 10.5) | 6.14 (5.83 - 6.46) |
| SIDIAP | 26,609 | 9,240 | 77.6 | 27.5 | 259 | 43 | 3.34 (2.94 - 3.77) | 1.56 (1.13 - 2.10) |
| Meta-analysis | 585,602 | 1,156,224 | 589.9 | 1189.0 | 3,965 | 7,077 | 6.72 (6.51 - 6.93) | 5.95 (5.81 - 6.09) |
| Meta-analysis (w/o USOC) | 171,272 | 284,289 | 192.8 | 242.8 | 1,429 | 2,117 | 7.41 (7.03 - 7.81) | 8.72 (8.35 - 9.10) |
| **PS-matching / on-treatment TAR without escalation** | | | | | | | | |
| CCAE | 46,342 | 83,167 | 32.7 | 61.7 | 202 | 298 | 6.18 (5.35 - 7.09) | 4.83 (4.30 - 5.41) |
| DAG | 13,155 | 2,317 | 12.6 | 2.2 | 78 | 11 | 6.20 (4.90 - 7.74) | 5.07 (2.53 - 9.08) |
| ODOD | 27,995 | 67,645 | 19.1 | 56.5 | 298 | 733 | 15.6 (13.9 - 17.5) | 13.0 (12.1 - 13.9) |
| MDCR | 8,637 | 17,435 | 7.7 | 16.8 | 112 | 215 | 14.6 (12.0 - 17.5) | 12.8 (11.2 - 14.6) |
| MDCD | 3,890 | 9,392 | 2.3 | 5.6 | 29 | 62 | 12.9 (8.62 - 18.5) | 11.0 (8.45 - 14.1) |
| Open Claims | 407,051 | 871,924 | 300.9 | 801.7 | 1,928 | 4,225 | 6.41 (6.12 - 6.70) | 5.27 (5.11 - 5.43) |
| OEHR | 35,345 | 92,794 | 14.2 | 44.6 | 148 | 447 | 10.5 (8.84 - 12.3) | 10.0 (9.12 - 11.0) |
| VA | 0 | 0 | 0.0 | 0.0 | 0 | 0 | | |
| SIDIAP | 26,520 | 7,185 | 56.8 | 17.0 | 194 | 29 | 3.42 (2.95 - 3.93) | 1.70 (1.14 - 2.45) |
| Meta-analysis | 568,935 | 1,151,859 | 446.2 | 1006.0 | 2,989 | 6,020 | 6.70 (6.46 - 6.94) | 5.98 (5.83 - 6.14) |
| Meta-analysis (w/o USOC) | 161,884 | 279,935 | 145.3 | 204.4 | 1,061 | 1,795 | 7.30 (6.87 - 7.75) | 8.78 (8.38 - 9.20) |
| **PS-stratification / on-treatment TAR without escalation** | | | | | | | | |
| CCAE | 51,674 | 84,669 | 36.6 | 62.9 | 232 | 300 | 6.33 (5.54 - 7.20) | 4.77 (4.25 - 5.34) |
| DAG | 13,183 | 2,676 | 12.6 | 2.5 | 78 | 12 | 6.19 (4.89 - 7.72) | 4.73 (2.45 - 8.27) |
| ODOD | 29,544 | 67,651 | 20.3 | 56.5 | 318 | 733 | 15.7 (14.0 - 17.5) | 13.0 (12.1 - 13.9) |
| MDCR | 9,144 | 17,821 | 8.2 | 17.2 | 121 | 216 | 14.8 (12.3 - 17.7) | 12.5 (10.9 - 14.3) |
| MDCD | 3,979 | 9,412 | 2.3 | 5.6 | 30 | 62 | 13.0 (8.78 - 18.6) | 11.0 (8.43 - 14.1) |
| Open Claims | 414,330 | 871,935 | 307.7 | 801.7 | 1,999 | 4,225 | 6.50 (6.21 - 6.79) | 5.27 (5.11 - 5.43) |
| OEHR | 37,139 | 92,820 | 15.0 | 44.6 | 164 | 448 | 10.9 (9.30 - 12.7) | 10.1 (9.14 - 11.0) |
| VA | 24,656 | 226,229 | 18.7 | 214.1 | 174 | 1,280 | 9.30 (7.97 - 10.8) | 5.98 (5.66 - 6.32) |
| SIDIAP | 26,609 | 9,240 | 56.9 | 22.8 | 194 | 32 | 3.41 (2.95 - 3.93) | 1.41 (0.96 - 1.98) |
| Meta-analysis | 585,602 | 1,156,224 | 459.7 | 1013.8 | 3,136 | 6,028 | 6.82 (6.59 - 7.07) | 5.95 (5.80 - 6.10) |
| Meta-analysis (w/o USOC) | 171,272 | 284,289 | 151.9 | 212.1 | 1,137 | 1,803 | 7.48 (7.06 - 7.93) | 8.50 (8.11 - 8.90) |

##### 4.6.12 Acute pancreatitis

**Supplementary Table 431:** Exposures, outcomes and risk estimates for Acute pancreatitis in DPP4I (male) (T) vs SU (male) (C) new-user cohorts under four different propensity score (PS) adjustment / time-at-risk (TAR) designs across data sources. We report total patient exposure time in 1,000 patient-year (KYr) units, uncalibrated and calibrated hazard ratios (HRs) and their 95% confidence intervals (CIs). Greyed out entries did not pass study diagnostics and are included only for completeness.

| | Patients | | Exposure (KYr) | | Outcomes | | Incidence (per KYr) | |
|---|---|---|---|---|---|---|---|---|
| | T | C | T | C | T | C | T | C |
| **PS-matching / on-treatment TAR** | | | | | | | | |
| CCAE | 47,045 | 84,234 | 41.2 | 71.9 | 104 | 199 | 2.52 (2.06 - 3.06) | 2.77 (2.40 - 3.18) |
| DAG | 13,419 | 2,361 | 14.3 | 2.4 | 0 | 0 | | |
| ODOD | 28,998 | 69,988 | 24.4 | 65.7 | 62 | 238 | 2.54 (1.95 - 3.26) | 3.62 (3.18 - 4.11) |
| MDCR | 8,967 | 18,042 | 9.3 | 19.0 | 38 | 70 | 4.07 (2.88 - 5.59) | 3.69 (2.87 - 4.66) |
| MDCD | 4,011 | 9,564 | 3.1 | 6.9 | 16 | 56 | 5.21 (2.98 - 8.47) | 8.12 (6.13 - 10.5) |
| Open Claims | 417,329 | 889,404 | 398.9 | 966.5 | 793 | 2,182 | 1.99 (1.85 - 2.13) | 2.26 (2.16 - 2.35) |
| OEHR | 36,266 | 95,267 | 18.0 | 52.9 | 43 | 164 | 2.39 (1.73 - 3.22) | 3.10 (2.64 - 3.61) |
| VA | 0 | 0 | 0.0 | 0.0 | 0 | 0 | | |
| SIDIAP | 26,706 | 7,204 | 78.2 | 20.5 | 30 | 8 | 0.38 (0.26 - 0.55) | 0.39 (0.17 - 0.77) |
| Meta-analysis | 569,322 | 1,173,703 | 573.1 | 1203.3 | 1,086 | 2,917 | 1.89 (1.78 - 2.01) | 2.42 (2.34 - 2.51) |
| Meta-analysis (w/o USOC) | 151,993 | 284,299 | 174.2 | 236.9 | 293 | 735 | 1.68 (1.49 - 1.89) | 3.10 (2.88 - 3.34) |
| **PS-stratification / on-treatment TAR** | | | | | | | | |
| CCAE | 52,487 | 85,745 | 46.2 | 73.2 | 112 | 201 | 2.43 (2.00 - 2.92) | 2.75 (2.38 - 3.15) |
| DAG | 13,447 | 2,736 | 14.3 | 2.8 | 0 | 0 | | |
| ODOD | 30,607 | 69,996 | 25.9 | 65.7 | 67 | 238 | 2.59 (2.01 - 3.29) | 3.62 (3.18 - 4.11) |
| MDCR | 9,484 | 18,431 | 9.9 | 19.5 | 38 | 72 | 3.82 (2.70 - 5.24) | 3.70 (2.90 - 4.66) |
| MDCD | 4,102 | 9,580 | 3.1 | 6.9 | 18 | 56 | 5.72 (3.39 - 9.04) | 8.11 (6.13 - 10.5) |
| Open Claims | 424,808 | 889,411 | 408.4 | 966.5 | 813 | 2,182 | 1.99 (1.86 - 2.13) | 2.26 (2.16 - 2.35) |
| OEHR | 38,129 | 95,297 | 19.1 | 52.9 | 44 | 164 | 2.30 (1.67 - 3.09) | 3.10 (2.64 - 3.61) |
| VA | 25,396 | 231,330 | 23.8 | 246.1 | 42 | 510 | 1.76 (1.27 - 2.38) | 2.07 (1.90 - 2.26) |
| SIDIAP | 26,821 | 9,254 | 78.3 | 27.6 | 30 | 12 | 0.38 (0.26 - 0.55) | 0.43 (0.22 - 0.76) |
| Meta-analysis | 586,438 | 1,177,714 | 591.0 | 1212.2 | 1,122 | 2,925 | 1.90 (1.79 - 2.01) | 2.41 (2.33 - 2.50) |
| Meta-analysis (w/o USOC) | 161,630 | 288,303 | 182.6 | 245.8 | 309 | 743 | 1.69 (1.51 - 1.89) | 3.02 (2.81 - 3.25) |
| **PS-matching / on-treatment TAR without escalation** | | | | | | | | |
| CCAE | 47,045 | 84,234 | 33.3 | 62.5 | 75 | 168 | 2.25 (1.77 - 2.82) | 2.69 (2.30 - 3.13) |
| DAG | 13,419 | 2,361 | 12.9 | 2.2 | 0 | 0 | | |
| ODOD | 28,998 | 69,988 | 20.0 | 58.6 | 48 | 208 | 2.41 (1.77 - 3.19) | 3.55 (3.08 - 4.07) |
| MDCR | 8,967 | 18,042 | 8.0 | 17.3 | 36 | 61 | 4.52 (3.17 - 6.26) | 3.52 (2.70 - 4.53) |
| MDCD | 4,011 | 9,564 | 2.3 | 5.8 | 12 | 50 | 5.14 (2.66 - 8.98) | 8.66 (6.43 - 11.4) |
| Open Claims | 417,329 | 889,404 | 309.4 | 819.0 | 582 | 1,807 | 1.88 (1.73 - 2.04) | 2.21 (2.11 - 2.31) |
| OEHR | 36,266 | 95,267 | 14.6 | 46.0 | 31 | 120 | 2.13 (1.44 - 3.02) | 2.61 (2.16 - 3.12) |
| VA | 0 | 0 | 0.0 | 0.0 | 0 | 0 | | |
| SIDIAP | 26,706 | 7,204 | 57.3 | 17.1 | 26 | 7 | 0.45 (0.30 - 0.66) | 0.41 (0.16 - 0.84) |
| Meta-analysis | 569,322 | 1,173,703 | 444.9 | 1026.3 | 810 | 2,421 | 1.82 (1.70 - 1.95) | 2.36 (2.27 - 2.45) |
| Meta-analysis (w/o USOC) | 151,993 | 284,299 | 135.5 | 207.3 | 228 | 614 | 1.68 (1.47 - 1.92) | 2.96 (2.73 - 3.21) |
| **PS-stratification / on-treatment TAR without escalation** | | | | | | | | |
| CCAE | 52,487 | 85,745 | 37.3 | 63.7 | 81 | 170 | 2.17 (1.72 - 2.70) | 2.67 (2.28 - 3.10) |
| DAG | 13,447 | 2,736 | 12.9 | 2.6 | 0 | 0 | | |
| ODOD | 30,607 | 69,996 | 21.1 | 58.6 | 52 | 208 | 2.46 (1.84 - 3.23) | 3.55 (3.08 - 4.07) |
| MDCR | 9,484 | 18,431 | 8.5 | 17.8 | 36 | 63 | 4.24 (2.97 - 5.87) | 3.55 (2.72 - 4.54) |
| MDCD | 4,102 | 9,580 | 2.4 | 5.8 | 13 | 50 | 5.44 (2.90 - 9.31) | 8.65 (6.42 - 11.4) |
| Open Claims | 424,808 | 889,411 | 316.6 | 819.0 | 596 | 1,807 | 1.88 (1.73 - 2.04) | 2.21 (2.11 - 2.31) |
| OEHR | 38,129 | 95,297 | 15.5 | 46.0 | 32 | 120 | 2.06 (1.41 - 2.91) | 2.61 (2.16 - 3.12) |
| VA | 25,396 | 231,330 | 19.3 | 219.3 | 31 | 424 | 1.60 (1.09 - 2.28) | 1.93 (1.75 - 2.13) |
| SIDIAP | 26,821 | 9,254 | 57.4 | 22.8 | 26 | 11 | 0.45 (0.30 - 0.66) | 0.48 (0.24 - 0.86) |
| Meta-analysis | 586,438 | 1,177,714 | 458.9 | 1033.7 | 836 | 2,429 | 1.82 (1.70 - 1.95) | 2.35 (2.26 - 2.45) |
| Meta-analysis (w/o USOC) | 161,630 | 288,303 | 142.3 | 214.6 | 240 | 622 | 1.69 (1.48 - 1.91) | 2.90 (2.67 - 3.13) |

##### 4.6.13 Bone fracture

**Supplementary Table 432:** Exposures, outcomes and risk estimates for Bone fracture in DPP4I (male) (T) vs SU (male) (C) new-user cohorts under four different propensity score (PS) adjustment / time-at-risk (TAR) designs across data sources. We report total patient exposure time in 1,000 patient-year (KYr) units, uncalibrated and calibrated hazard ratios (HRs) and their 95% confidence intervals (CIs). Greyed out entries did not pass study diagnostics and are included only for completeness.

| | Patients | | Exposure (KYr) | | Outcomes | | Incidence (per KYr) | |
|---|---|---|---|---|---|---|---|---|
| | T | C | T | C | T | C | T | C |
| **PS-matching / on-treatment TAR** | | | | | | | | |
| CCAE | 44,226 | 79,432 | 38.1 | 66.8 | 555 | 1,030 | 14.6 (13.4 - 15.8) | 15.4 (14.5 - 16.4) |
| DAG | 11,999 | 2,144 | 12.6 | 2.2 | 208 | 41 | 16.5 (14.3 - 18.9) | 18.7 (13.4 - 25.4) |
| ODOD | 27,120 | 66,034 | 22.4 | 61.0 | 438 | 1,355 | 19.6 (17.8 - 21.5) | 22.2 (21.0 - 23.4) |
| MDCR | 8,218 | 16,511 | 8.3 | 17.0 | 228 | 465 | 27.4 (23.9 - 31.1) | 27.3 (24.9 - 29.9) |
| MDCD | 3,584 | 8,620 | 2.7 | 6.1 | 84 | 204 | 31.1 (24.8 - 38.5) | 33.4 (29.0 - 38.3) |
| Open Claims | 378,952 | 811,712 | 358.5 | 873.7 | 4,866 | 12,692 | 13.6 (13.2 - 14.0) | 14.5 (14.3 - 14.8) |
| OEHR | 34,463 | 90,094 | 16.9 | 49.2 | 271 | 893 | 16.1 (14.2 - 18.1) | 18.2 (17.0 - 19.4) |
| VA | 0 | 0 | 0.0 | 0.0 | 0 | 0 | | |
| SIDIAP | 24,737 | 6,841 | 72.1 | 19.2 | 682 | 202 | 9.46 (8.76 - 10.2) | 10.5 (9.14 - 12.1) |
| Meta-analysis | 533,299 | 1,081,388 | 531.6 | 1095.1 | 7,332 | 16,882 | 13.8 (13.5 - 14.1) | 15.4 (15.2 - 15.7) |
| Meta-analysis (w/o USOC) | 154,347 | 269,676 | 173.1 | 221.4 | 2,466 | 4,190 | 14.2 (13.7 - 14.8) | 18.9 (18.4 - 19.5) |
| **PS-stratification / on-treatment TAR** | | | | | | | | |
| CCAE | 49,297 | 80,914 | 42.6 | 68.1 | 633 | 1,044 | 14.9 (13.7 - 16.1) | 15.3 (14.4 - 16.3) |
| DAG | 12,024 | 2,488 | 12.6 | 2.6 | 209 | 44 | 16.6 (14.4 - 19.0) | 17.2 (12.5 - 23.0) |
| ODOD | 28,643 | 66,040 | 23.8 | 61.0 | 462 | 1,355 | 19.4 (17.7 - 21.3) | 22.2 (21.0 - 23.4) |
| MDCR | 8,670 | 16,876 | 8.8 | 17.5 | 242 | 473 | 27.4 (24.0 - 31.0) | 27.1 (24.7 - 29.7) |
| MDCD | 3,661 | 8,636 | 2.8 | 6.1 | 85 | 204 | 30.7 (24.5 - 38.0) | 33.3 (28.9 - 38.2) |
| Open Claims | 385,636 | 811,724 | 366.8 | 873.7 | 4,994 | 12,692 | 13.6 (13.2 - 14.0) | 14.5 (14.3 - 14.8) |
| OEHR | 36,215 | 90,122 | 17.9 | 49.2 | 284 | 894 | 15.8 (14.1 - 17.8) | 18.2 (17.0 - 19.4) |
| VA | 23,057 | 213,933 | 21.6 | 226.0 | 216 | 2,435 | 9.99 (8.70 - 11.4) | 10.8 (10.3 - 11.2) |
| SIDIAP | 24,839 | 8,816 | 72.2 | 25.8 | 682 | 266 | 9.45 (8.75 - 10.2) | 10.3 (9.10 - 11.6) |
| Meta-analysis | 548,985 | 1,085,616 | 547.6 | 1103.9 | 7,591 | 16,972 | 13.9 (13.6 - 14.2) | 15.4 (15.1 - 15.6) |
| Meta-analysis (w/o USOC) | 163,349 | 273,892 | 180.7 | 230.2 | 2,597 | 4,280 | 14.4 (13.8 - 14.9) | 18.6 (18.0 - 19.2) |
| **PS-matching / on-treatment TAR without escalation** | | | | | | | | |
| CCAE | 44,226 | 79,432 | 30.9 | 58.3 | 449 | 895 | 14.5 (13.2 - 15.9) | 15.4 (14.4 - 16.4) |
| DAG | 11,999 | 2,144 | 11.4 | 2.0 | 194 | 41 | 17.0 (14.7 - 19.6) | 20.6 (14.8 - 27.9) |
| ODOD | 27,120 | 66,034 | 18.4 | 54.5 | 352 | 1,222 | 19.1 (17.1 - 21.2) | 22.4 (21.2 - 23.7) |
| MDCR | 8,218 | 16,511 | 7.1 | 15.5 | 199 | 430 | 27.8 (24.1 - 32.0) | 27.7 (25.1 - 30.4) |
| MDCD | 3,584 | 8,620 | 2.1 | 5.2 | 62 | 171 | 30.2 (23.1 - 38.7) | 33.1 (28.4 - 38.5) |
| Open Claims | 378,952 | 811,712 | 278.7 | 741.7 | 3,739 | 10,693 | 13.4 (13.0 - 13.9) | 14.4 (14.1 - 14.7) |
| OEHR | 34,463 | 90,094 | 13.7 | 42.9 | 198 | 745 | 14.4 (12.5 - 16.6) | 17.4 (16.1 - 18.7) |
| VA | 0 | 0 | 0.0 | 0.0 | 0 | 0 | | |
| SIDIAP | 24,737 | 6,841 | 53.0 | 16.0 | 502 | 171 | 9.47 (8.66 - 10.3) | 10.7 (9.13 - 12.4) |
| Meta-analysis | 533,299 | 1,081,388 | 415.3 | 936.0 | 5,695 | 14,368 | 13.7 (13.4 - 14.1) | 15.4 (15.1 - 15.6) |
| Meta-analysis (w/o USOC) | 154,347 | 269,676 | 136.7 | 194.3 | 1,956 | 3,675 | 14.3 (13.7 - 15.0) | 18.9 (18.3 - 19.5) |
| **PS-stratification / on-treatment TAR without escalation** | | | | | | | | |
| CCAE | 49,297 | 80,914 | 34.6 | 59.4 | 511 | 909 | 14.8 (13.5 - 16.1) | 15.3 (14.3 - 16.3) |
| DAG | 12,024 | 2,488 | 11.4 | 2.3 | 195 | 43 | 17.1 (14.8 - 19.7) | 18.4 (13.3 - 24.8) |
| ODOD | 28,643 | 66,040 | 19.6 | 54.5 | 372 | 1,222 | 19.0 (17.1 - 21.1) | 22.4 (21.2 - 23.7) |
| MDCR | 8,670 | 16,876 | 7.6 | 16.0 | 211 | 437 | 27.8 (24.2 - 31.8) | 27.4 (24.8 - 30.0) |
| MDCD | 3,661 | 8,636 | 2.1 | 5.2 | 63 | 171 | 30.0 (23.1 - 38.4) | 33.1 (28.3 - 38.4) |
| Open Claims | 385,636 | 811,724 | 285.0 | 741.7 | 3,843 | 10,693 | 13.5 (13.1 - 13.9) | 14.4 (14.1 - 14.7) |
| OEHR | 36,215 | 90,122 | 14.6 | 42.9 | 209 | 745 | 14.3 (12.4 - 16.4) | 17.4 (16.1 - 18.7) |
| VA | 23,057 | 213,933 | 17.6 | 201.8 | 166 | 2,146 | 9.44 (8.06 - 11.0) | 10.6 (10.2 - 11.1) |
| SIDIAP | 24,839 | 8,816 | 53.1 | 21.4 | 502 | 225 | 9.46 (8.65 - 10.3) | 10.5 (9.18 - 12.0) |
| Meta-analysis | 548,985 | 1,085,616 | 427.9 | 943.3 | 5,906 | 14,445 | 13.8 (13.5 - 14.2) | 15.3 (15.1 - 15.6) |
| Meta-analysis (w/o USOC) | 163,349 | 273,892 | 142.9 | 201.7 | 2,063 | 3,752 | 14.4 (13.8 - 15.1) | 18.6 (18.0 - 19.2) |

##### 4.6.14 Diabetic ketoacidosis

**Supplementary Table 433:** Exposures, outcomes and risk estimates for Diabetic ketoacidosis in DPP4I (male) (T) vs SU (male) (C) new-user cohorts under four different propensity score (PS) adjustment / time-at-risk (TAR) designs across data sources. We report total patient exposure time in 1,000 patient-year (KYr) units, uncalibrated and calibrated hazard ratios (HRs) and their 95% confidence intervals (CIs). Greyed out entries did not pass study diagnostics and are included only for completeness.

| | Patients | | Exposure (KYr) | | Outcomes | | Incidence (per KYr) | |
|---|---|---|---|---|---|---|---|---|
| | T | C | T | C | T | C | T | C |
| **PS-matching / on-treatment TAR** | | | | | | | | |
| CCAE | 47,391 | 85,013 | 41.6 | 72.6 | 34 | 50 | 0.82 (0.57 - 1.14) | 0.69 (0.51 - 0.91) |
| DAG | 13,419 | 2,361 | 14.3 | 2.4 | 0 | 0 | | |
| ODOD | 29,199 | 70,642 | 24.5 | 66.5 | 37 | 87 | 1.51 (1.06 - 2.08) | 1.31 (1.05 - 1.61) |
| MDCR | 9,041 | 18,266 | 9.4 | 19.2 | 7 | 23 | 0.74 (0.30 - 1.53) | 1.20 (0.76 - 1.79) |
| MDCD | 4,072 | 9,788 | 3.1 | 7.0 | 5 | 39 | 1.61 (0.52 - 3.76) | 5.54 (3.94 - 7.57) |
| Open Claims | 421,605 | 900,893 | 403.0 | 979.8 | 299 | 788 | 0.74 (0.66 - 0.83) | 0.80 (0.75 - 0.86) |
| OEHR | 36,419 | 95,809 | 18.1 | 53.3 | 24 | 62 | 1.33 (0.85 - 1.98) | 1.16 (0.89 - 1.49) |
| VA | 0 | 0 | 0.0 | 0.0 | 0 | 0 | | |
| SIDIAP | 26,798 | 7,217 | 78.5 | 20.5 | 0 | <5 | | < 0.24 (0.08 - 0.57) |
| Meta-analysis | 547,727 | 1,180,411 | 499.7 | 1198.5 | 406 | 1,049 | 0.81 (0.74 - 0.90) | 0.88 (0.82 - 0.93) |
| Meta-analysis (w/o USOC) | 126,122 | 279,518 | 96.7 | 218.7 | 107 | 261 | 1.11 (0.91 - 1.34) | 1.19 (1.05 - 1.35) |
| **PS-stratification / on-treatment TAR** | | | | | | | | |
| CCAE | 52,836 | 86,530 | 46.5 | 73.9 | 36 | 52 | 0.77 (0.54 - 1.07) | 0.70 (0.53 - 0.92) |
| DAG | 13,447 | 2,736 | 14.3 | 2.8 | 0 | 0 | | |
| ODOD | 30,816 | 70,649 | 26.1 | 66.5 | 38 | 87 | 1.46 (1.03 - 2.00) | 1.31 (1.05 - 1.61) |
| MDCR | 9,553 | 18,658 | 10.0 | 19.7 | 7 | 23 | 0.70 (0.28 - 1.44) | 1.17 (0.74 - 1.75) |
| MDCD | 4,168 | 9,808 | 3.2 | 7.1 | 5 | 39 | 1.57 (0.51 - 3.66) | 5.53 (3.93 - 7.56) |
| Open Claims | 429,005 | 900,907 | 412.3 | 979.8 | 304 | 788 | 0.74 (0.66 - 0.83) | 0.80 (0.75 - 0.86) |
| OEHR | 38,263 | 95,839 | 19.2 | 53.3 | 24 | 62 | 1.25 (0.80 - 1.86) | 1.16 (0.89 - 1.49) |
| VA | 25,614 | 233,668 | 24.0 | 248.9 | 11 | 126 | 0.46 (0.23 - 0.82) | 0.51 (0.42 - 0.60) |
| SIDIAP | 26,891 | 9,270 | 78.6 | 27.7 | 0 | <5 | | < 0.18 (0.06 - 0.42) |
| Meta-analysis | 564,641 | 1,182,391 | 517.3 | 1200.3 | 414 | 1,051 | 0.80 (0.73 - 0.88) | 0.88 (0.82 - 0.93) |
| Meta-analysis (w/o USOC) | 135,636 | 281,484 | 105.0 | 220.5 | 110 | 263 | 1.05 (0.86 - 1.26) | 1.19 (1.05 - 1.35) |
| **PS-matching / on-treatment TAR without escalation** | | | | | | | | |
| CCAE | 47,391 | 85,013 | 33.5 | 63.1 | 27 | 38 | 0.80 (0.53 - 1.17) | 0.60 (0.43 - 0.83) |
| DAG | 13,419 | 2,361 | 12.9 | 2.2 | 0 | 0 | | |
| ODOD | 29,199 | 70,642 | 20.1 | 59.2 | 26 | 65 | 1.29 (0.85 - 1.90) | 1.10 (0.85 - 1.40) |
| MDCR | 9,041 | 18,266 | 8.0 | 17.5 | 6 | 19 | 0.75 (0.27 - 1.62) | 1.08 (0.65 - 1.69) |
| MDCD | 4,072 | 9,788 | 2.4 | 5.9 | 5 | 31 | 2.12 (0.69 - 4.94) | 5.25 (3.57 - 7.46) |
| Open Claims | 421,605 | 900,893 | 312.5 | 830.0 | 212 | 606 | 0.68 (0.59 - 0.78) | 0.73 (0.67 - 0.79) |
| OEHR | 36,419 | 95,809 | 14.7 | 46.3 | 17 | 35 | 1.16 (0.68 - 1.86) | 0.76 (0.53 - 1.05) |
| VA | 0 | 0 | 0.0 | 0.0 | 0 | 0 | | |
| SIDIAP | 26,798 | 7,217 | 57.5 | 17.1 | 0 | <5 | | < 0.29 (0.09 - 0.68) |
| Meta-analysis | 547,727 | 1,180,411 | 391.2 | 1022.0 | 293 | 794 | 0.75 (0.67 - 0.84) | 0.78 (0.72 - 0.83) |
| Meta-analysis (w/o USOC) | 126,122 | 279,518 | 78.7 | 192.1 | 81 | 188 | 1.03 (0.82 - 1.28) | 0.98 (0.84 - 1.13) |
| **PS-stratification / on-treatment TAR without escalation** | | | | | | | | |
| CCAE | 52,836 | 86,530 | 37.6 | 64.3 | 28 | 40 | 0.75 (0.50 - 1.08) | 0.62 (0.44 - 0.85) |
| DAG | 13,447 | 2,736 | 12.9 | 2.6 | 0 | 0 | | |
| ODOD | 30,816 | 70,649 | 21.3 | 59.3 | 26 | 65 | 1.22 (0.80 - 1.79) | 1.10 (0.85 - 1.40) |
| MDCR | 9,553 | 18,658 | 8.6 | 18.0 | 6 | 19 | 0.70 (0.26 - 1.53) | 1.06 (0.64 - 1.65) |
| MDCD | 4,168 | 9,808 | 2.4 | 5.9 | 5 | 31 | 2.06 (0.67 - 4.82) | 5.24 (3.56 - 7.44) |
| Open Claims | 429,005 | 900,907 | 319.5 | 830.0 | 217 | 606 | 0.68 (0.59 - 0.78) | 0.73 (0.67 - 0.79) |
| OEHR | 38,263 | 95,839 | 15.6 | 46.3 | 17 | 35 | 1.09 (0.64 - 1.75) | 0.76 (0.53 - 1.05) |
| VA | 25,614 | 233,668 | 19.5 | 221.6 | <10 | 91 | < 0.51 (0.25 - 0.94) | 0.41 (0.33 - 0.50) |
| SIDIAP | 26,891 | 9,270 | 57.6 | 22.9 | 0 | <5 | | < 0.22 (0.07 - 0.51) |
| Meta-analysis | 564,641 | 1,182,391 | 404.9 | 1023.7 | 299 | 796 | 0.74 (0.66 - 0.83) | 0.78 (0.72 - 0.83) |
| Meta-analysis (w/o USOC) | 135,636 | 281,484 | 85.4 | 193.8 | 82 | 190 | 0.96 (0.76 - 1.19) | 0.98 (0.85 - 1.13) |

##### 4.6.15 Diarrhea

**Supplementary Table 434:** Exposures, outcomes and risk estimates for Diarrhea in DPP4I (male) (T) vs SU (male) (C) new-user cohorts under four different propensity score (PS) adjustment / time-at-risk (TAR) designs across data sources. We report total patient exposure time in 1,000 patient-year (KYr) units, uncalibrated and calibrated hazard ratios (HRs) and their 95% confidence intervals (CIs). Greyed out entries did not pass study diagnostics and are included only for completeness.

| | Patients | | Exposure (KYr) | | Outcomes | | Incidence (per KYr) | |
|---|---|---|---|---|---|---|---|---|
| | T | C | T | C | T | C | T | C |
| **PS-matching / on-treatment TAR** | | | | | | | | |
| CCAE | 43,507 | 78,291 | 37.5 | 65.8 | 767 | 1,393 | 20.5 (19.0 - 22.0) | 21.2 (20.1 - 22.3) |
| DAG | 13,237 | 2,325 | 14.1 | 2.4 | 28 | 8 | 1.99 (1.32 - 2.88) | 3.34 (1.44 - 6.58) |
| ODOD | 26,135 | 63,538 | 21.3 | 58.0 | 630 | 1,910 | 29.5 (27.3 - 31.9) | 32.9 (31.5 - 34.4) |
| MDCR | 8,110 | 16,353 | 8.3 | 16.7 | 251 | 494 | 30.3 (26.7 - 34.3) | 29.5 (27.0 - 32.2) |
| MDCD | 3,468 | 8,433 | 2.6 | 5.9 | 111 | 269 | 42.4 (34.9 - 51.1) | 45.8 (40.5 - 51.6) |
| Open Claims | 380,477 | 819,460 | 358.9 | 879.4 | 5,372 | 13,028 | 15.0 (14.6 - 15.4) | 14.8 (14.6 - 15.1) |
| OEHR | 33,533 | 88,165 | 16.2 | 47.1 | 447 | 1,556 | 27.5 (25.1 - 30.2) | 33.0 (31.4 - 34.7) |
| VA | 0 | 0 | 0.0 | 0.0 | 0 | 0 | | |
| SIDIAP | 25,832 | 7,060 | 75.5 | 20.0 | 573 | 91 | 7.59 (6.98 - 8.24) | 4.56 (3.67 - 5.60) |
| Meta-analysis | 534,299 | 1,083,625 | 534.3 | 1095.2 | 8,179 | 18,749 | 15.3 (15.0 - 15.6) | 17.1 (16.9 - 17.4) |
| Meta-analysis (w/o USOC) | 153,822 | 264,165 | 175.4 | 215.9 | 2,807 | 5,721 | 16.0 (15.4 - 16.6) | 26.5 (25.8 - 27.2) |
| **PS-stratification / on-treatment TAR** | | | | | | | | |
| CCAE | 48,527 | 79,796 | 41.9 | 67.0 | 874 | 1,409 | 20.9 (19.5 - 22.3) | 21.0 (19.9 - 22.1) |
| DAG | 13,265 | 2,696 | 14.1 | 2.8 | 28 | 8 | 1.99 (1.32 - 2.87) | 2.86 (1.24 - 5.64) |
| ODOD | 27,571 | 63,544 | 22.6 | 58.0 | 676 | 1,910 | 29.9 (27.7 - 32.2) | 32.9 (31.5 - 34.4) |
| MDCR | 8,555 | 16,728 | 8.8 | 17.2 | 264 | 507 | 30.0 (26.5 - 33.8) | 29.5 (27.0 - 32.2) |
| MDCD | 3,545 | 8,450 | 2.7 | 5.9 | 111 | 269 | 41.4 (34.1 - 49.8) | 45.7 (40.4 - 51.5) |
| Open Claims | 387,075 | 819,471 | 367.0 | 879.4 | 5,531 | 13,028 | 15.1 (14.7 - 15.5) | 14.8 (14.6 - 15.1) |
| OEHR | 35,251 | 88,189 | 17.2 | 47.1 | 470 | 1,557 | 27.3 (24.9 - 29.9) | 33.0 (31.4 - 34.7) |
| VA | 22,232 | 208,766 | 20.7 | 218.0 | 408 | 3,934 | 19.7 (17.9 - 21.7) | 18.0 (17.5 - 18.6) |
| SIDIAP | 25,996 | 9,089 | 75.6 | 26.9 | 575 | 112 | 7.60 (6.99 - 8.25) | 4.16 (3.43 - 5.01) |
| Meta-analysis | 549,785 | 1,087,963 | 549.8 | 1104.3 | 8,529 | 18,800 | 15.5 (15.2 - 15.8) | 17.0 (16.8 - 17.3) |
| Meta-analysis (w/o USOC) | 162,710 | 268,492 | 182.9 | 225.0 | 2,998 | 5,772 | 16.4 (15.8 - 17.0) | 25.7 (25.0 - 26.3) |
| **PS-matching / on-treatment TAR without escalation** | | | | | | | | |
| CCAE | 43,507 | 78,291 | 30.4 | 57.5 | 628 | 1,216 | 20.6 (19.1 - 22.3) | 21.2 (20.0 - 22.4) |
| DAG | 13,237 | 2,325 | 12.7 | 2.2 | 25 | 7 | 1.97 (1.28 - 2.91) | 3.20 (1.29 - 6.59) |
| ODOD | 26,135 | 63,538 | 17.6 | 52.0 | 525 | 1,712 | 29.9 (27.4 - 32.5) | 33.0 (31.4 - 34.6) |
| MDCR | 8,110 | 16,353 | 7.1 | 15.3 | 216 | 457 | 30.4 (26.5 - 34.8) | 29.8 (27.1 - 32.7) |
| MDCD | 3,468 | 8,433 | 2.0 | 5.0 | 81 | 230 | 40.4 (32.1 - 50.3) | 45.9 (40.2 - 52.3) |
| Open Claims | 380,477 | 819,460 | 279.7 | 748.2 | 4,159 | 10,955 | 14.9 (14.4 - 15.3) | 14.6 (14.4 - 14.9) |
| OEHR | 33,533 | 88,165 | 13.3 | 41.4 | 313 | 1,242 | 23.5 (21.0 - 26.3) | 30.0 (28.4 - 31.7) |
| VA | 0 | 0 | 0.0 | 0.0 | 0 | 0 | | |
| SIDIAP | 25,832 | 7,060 | 55.4 | 16.7 | 436 | 75 | 7.88 (7.15 - 8.65) | 4.49 (3.53 - 5.63) |
| Meta-analysis | 521,062 | 1,081,300 | 405.5 | 936.0 | 6,358 | 15,887 | 15.7 (15.3 - 16.1) | 17.0 (16.7 - 17.2) |
| Meta-analysis (w/o USOC) | 140,585 | 261,840 | 125.8 | 187.8 | 2,199 | 4,932 | 17.5 (16.8 - 18.2) | 26.3 (25.5 - 27.0) |
| **PS-stratification / on-treatment TAR without escalation** | | | | | | | | |
| CCAE | 48,527 | 79,796 | 34.1 | 58.6 | 711 | 1,229 | 20.9 (19.4 - 22.5) | 21.0 (19.8 - 22.2) |
| DAG | 13,265 | 2,696 | 12.7 | 2.6 | 25 | 7 | 1.97 (1.27 - 2.90) | 2.73 (1.10 - 5.63) |
| ODOD | 27,571 | 63,544 | 18.6 | 52.0 | 561 | 1,712 | 30.2 (27.7 - 32.8) | 32.9 (31.4 - 34.5) |
| MDCR | 8,555 | 16,728 | 7.6 | 15.8 | 227 | 470 | 30.1 (26.3 - 34.2) | 29.8 (27.2 - 32.6) |
| MDCD | 3,545 | 8,450 | 2.1 | 5.0 | 81 | 230 | 39.5 (31.4 - 49.1) | 45.8 (40.1 - 52.1) |
| Open Claims | 387,075 | 819,471 | 285.9 | 748.2 | 4,278 | 10,955 | 15.0 (14.5 - 15.4) | 14.6 (14.4 - 14.9) |
| OEHR | 35,251 | 88,189 | 14.1 | 41.4 | 332 | 1,243 | 23.5 (21.1 - 26.2) | 30.0 (28.4 - 31.8) |
| VA | 22,232 | 208,766 | 16.9 | 195.3 | 329 | 3,449 | 19.5 (17.4 - 21.7) | 17.7 (17.1 - 18.3) |
| SIDIAP | 25,996 | 9,089 | 55.5 | 22.3 | 438 | 91 | 7.89 (7.17 - 8.67) | 4.09 (3.29 - 5.02) |
| Meta-analysis | 549,785 | 1,087,963 | 430.5 | 945.7 | 6,653 | 15,937 | 15.5 (15.1 - 15.8) | 16.9 (16.6 - 17.1) |
| Meta-analysis (w/o USOC) | 162,710 | 268,492 | 144.6 | 197.6 | 2,375 | 4,982 | 16.4 (15.8 - 17.1) | 25.2 (24.5 - 25.9) |

##### 4.6.16 Genitourinary infection

**Supplementary Table 435:** Exposures, outcomes and risk estimates for Genitourinary infection in DPP4I (male) (T) vs SU (male) (C) new-user cohorts under four different propensity score (PS) adjustment / time-at-risk (TAR) designs across data sources. We report total patient exposure time in 1,000 patient-year (KYr) units, uncalibrated and calibrated hazard ratios (HRs) and their 95% confidence intervals (CIs). Greyed out entries did not pass study diagnostics and are included only for completeness.

| | Patients | | Exposure (KYr) | | Outcomes | | Incidence (per KYr) | |
|---|---|---|---|---|---|---|---|---|
| | T | C | T | C | T | C | T | C |
| **PS-matching / on-treatment TAR** | | | | | | | | |
| CCAE | 43,222 | 78,021 | 37.1 | 65.5 | 762 | 1,325 | 20.6 (19.1 - 22.1) | 20.2 (19.2 - 21.4) |
| DAG | 11,640 | 2,076 | 12.0 | 2.1 | 282 | 45 | 23.5 (20.8 - 26.4) | 21.4 (15.6 - 28.7) |
| ODOD | 25,325 | 61,682 | 20.7 | 55.7 | 756 | 2,375 | 36.5 (33.9 - 39.2) | 42.7 (41.0 - 44.4) |
| MDCR | 7,685 | 15,540 | 7.8 | 15.9 | 346 | 718 | 44.1 (39.6 - 49.0) | 45.0 (41.8 - 48.4) |
| MDCD | 3,601 | 8,687 | 2.7 | 6.1 | 82 | 192 | 30.7 (24.4 - 38.1) | 31.7 (27.4 - 36.5) |
| Open Claims | 371,458 | 802,493 | 350.2 | 858.0 | 6,759 | 16,701 | 19.3 (18.8 - 19.8) | 19.5 (19.2 - 19.8) |
| OEHR | 34,221 | 90,073 | 16.7 | 48.8 | 379 | 1,241 | 22.7 (20.5 - 25.1) | 25.4 (24.0 - 26.9) |
| VA | 0 | 0 | 0.0 | 0.0 | 0 | 0 | | |
| SIDIAP | 23,919 | 6,791 | 68.7 | 18.6 | 1,582 | 340 | 23.0 (21.9 - 24.2) | 18.3 (16.4 - 20.3) |
| Meta-analysis | 521,071 | 1,065,363 | 515.9 | 1070.5 | 10,948 | 22,937 | 21.2 (20.8 - 21.6) | 21.4 (21.1 - 21.7) |
| Meta-analysis (w/o USOC) | 149,613 | 262,870 | 165.6 | 212.6 | 4,189 | 6,236 | 25.3 (24.5 - 26.1) | 29.3 (28.6 - 30.1) |
| **PS-stratification / on-treatment TAR** | | | | | | | | |
| CCAE | 48,125 | 79,426 | 41.4 | 66.6 | 868 | 1,336 | 20.9 (19.6 - 22.4) | 20.1 (19.0 - 21.2) |
| DAG | 11,719 | 2,418 | 12.1 | 2.4 | 285 | 51 | 23.6 (20.9 - 26.5) | 20.9 (15.6 - 27.5) |
| ODOD | 26,717 | 61,687 | 21.9 | 55.7 | 803 | 2,375 | 36.6 (34.1 - 39.2) | 42.7 (41.0 - 44.4) |
| MDCR | 8,073 | 15,881 | 8.3 | 16.4 | 365 | 729 | 44.1 (39.7 - 48.9) | 44.6 (41.4 - 47.9) |
| MDCD | 3,670 | 8,703 | 2.7 | 6.1 | 83 | 192 | 30.4 (24.2 - 37.7) | 31.6 (27.3 - 36.4) |
| Open Claims | 377,661 | 802,507 | 357.8 | 858.0 | 6,968 | 16,701 | 19.5 (19.0 - 19.9) | 19.5 (19.2 - 19.8) |
| OEHR | 35,941 | 90,101 | 17.7 | 48.8 | 402 | 1,242 | 22.7 (20.6 - 25.1) | 25.5 (24.1 - 26.9) |
| VA | 22,854 | 211,170 | 21.3 | 220.7 | 298 | 3,737 | 14.0 (12.5 - 15.7) | 16.9 (16.4 - 17.5) |
| SIDIAP | 24,002 | 8,800 | 68.7 | 25.3 | 1,585 | 443 | 23.1 (21.9 - 24.2) | 17.5 (15.9 - 19.2) |
| Meta-analysis | 535,908 | 1,069,523 | 530.7 | 1079.2 | 11,359 | 23,069 | 21.4 (21.0 - 21.8) | 21.4 (21.1 - 21.7) |
| Meta-analysis (w/o USOC) | 158,247 | 267,016 | 172.9 | 221.2 | 4,391 | 6,368 | 25.4 (24.7 - 26.2) | 28.8 (28.1 - 29.5) |
| **PS-matching / on-treatment TAR without escalation** | | | | | | | | |
| CCAE | 43,222 | 78,021 | 30.1 | 57.1 | 621 | 1,159 | 20.6 (19.0 - 22.3) | 20.3 (19.2 - 21.5) |
| DAG | 11,640 | 2,076 | 10.9 | 1.9 | 248 | 43 | 22.8 (20.0 - 25.8) | 22.5 (16.3 - 30.4) |
| ODOD | 25,325 | 61,682 | 17.0 | 49.7 | 634 | 2,157 | 37.3 (34.4 - 40.3) | 43.4 (41.6 - 45.3) |
| MDCR | 7,685 | 15,540 | 6.7 | 14.6 | 293 | 650 | 43.6 (38.8 - 48.9) | 44.6 (41.2 - 48.2) |
| MDCD | 3,601 | 8,687 | 2.0 | 5.1 | 64 | 162 | 31.4 (24.2 - 40.1) | 31.7 (27.0 - 37.0) |
| Open Claims | 371,458 | 802,493 | 271.8 | 727.9 | 5,351 | 14,313 | 19.7 (19.2 - 20.2) | 19.7 (19.3 - 20.0) |
| OEHR | 34,221 | 90,073 | 13.6 | 42.6 | 292 | 980 | 21.5 (19.1 - 24.1) | 23.0 (21.6 - 24.5) |
| VA | 0 | 0 | 0.0 | 0.0 | 0 | 0 | | |
| SIDIAP | 23,919 | 6,791 | 50.5 | 15.6 | 1,151 | 278 | 22.8 (21.5 - 24.1) | 17.8 (15.8 - 20.0) |
| Meta-analysis | 521,071 | 1,065,363 | 402.6 | 914.4 | 8,654 | 19,742 | 21.5 (21.0 - 22.0) | 21.6 (21.3 - 21.9) |
| Meta-analysis (w/o USOC) | 149,613 | 262,870 | 130.9 | 186.6 | 3,303 | 5,429 | 25.2 (24.4 - 26.1) | 29.1 (28.3 - 29.9) |
| **PS-stratification / on-treatment TAR without escalation** | | | | | | | | |
| CCAE | 48,125 | 79,426 | 33.7 | 58.2 | 701 | 1,169 | 20.8 (19.3 - 22.4) | 20.1 (19.0 - 21.3) |
| DAG | 11,719 | 2,418 | 11.0 | 2.2 | 251 | 49 | 22.9 (20.2 - 25.9) | 22.0 (16.3 - 29.1) |
| ODOD | 26,717 | 61,687 | 18.0 | 49.7 | 670 | 2,157 | 37.3 (34.5 - 40.2) | 43.4 (41.6 - 45.3) |
| MDCR | 8,073 | 15,881 | 7.1 | 15.0 | 309 | 660 | 43.5 (38.8 - 48.7) | 44.1 (40.8 - 47.6) |
| MDCD | 3,670 | 8,703 | 2.1 | 5.1 | 65 | 162 | 31.2 (24.1 - 39.8) | 31.7 (27.0 - 36.9) |
| Open Claims | 377,661 | 802,507 | 277.4 | 727.9 | 5,531 | 14,313 | 19.9 (19.4 - 20.5) | 19.7 (19.3 - 20.0) |
| OEHR | 35,941 | 90,101 | 14.4 | 42.6 | 311 | 981 | 21.5 (19.2 - 24.1) | 23.0 (21.6 - 24.5) |
| VA | 22,854 | 211,170 | 17.3 | 197.1 | 243 | 3,218 | 14.1 (12.4 - 16.0) | 16.3 (15.8 - 16.9) |
| SIDIAP | 24,002 | 8,800 | 50.6 | 21.0 | 1,154 | 355 | 22.8 (21.5 - 24.2) | 16.9 (15.2 - 18.8) |
| Meta-analysis | 535,908 | 1,069,523 | 414.2 | 921.7 | 8,992 | 19,846 | 21.7 (21.3 - 22.2) | 21.5 (21.2 - 21.8) |
| Meta-analysis (w/o USOC) | 158,247 | 267,016 | 136.8 | 193.8 | 3,461 | 5,533 | 25.3 (24.5 - 26.2) | 28.5 (27.8 - 29.3) |

##### 4.6.17 Hyperkalemia

**Supplementary Table 436:** Exposures, outcomes and risk estimates for Hyperkalemia in DPP4I (male) (T) vs SU (male) (C) new-user cohorts under four different propensity score (PS) adjustment / time-at-risk (TAR) designs across data sources. We report total patient exposure time in 1,000 patient-year (KYr) units, uncalibrated and calibrated hazard ratios (HRs) and their 95% confidence intervals (CIs). Greyed out entries did not pass study diagnostics and are included only for completeness.

| | Patients | | Exposure (KYr) | | Outcomes | | Incidence (per KYr) | |
|---|---|---|---|---|---|---|---|---|
| | T | C | T | C | T | C | T | C |
| **PS-matching / on-treatment TAR** | | | | | | | | |
| CCAE | 46,699 | 83,866 | 40.6 | 71.2 | 296 | 570 | 7.29 (6.48 - 8.17) | 8.01 (7.37 - 8.70) |
| DAG | 12,541 | 2,235 | 13.1 | 2.3 | 207 | 34 | 15.8 (13.7 - 18.1) | 14.8 (10.3 - 20.7) |
| ODOD | 27,972 | 67,536 | 23.1 | 62.0 | 485 | 1,534 | 21.0 (19.1 - 22.9) | 24.7 (23.5 - 26.0) |
| MDCR | 8,781 | 17,691 | 9.1 | 18.4 | 150 | 353 | 16.5 (14.0 - 19.4) | 19.2 (17.2 - 21.3) |
| MDCD | 3,914 | 9,426 | 2.9 | 6.7 | 51 | 158 | 17.4 (12.9 - 22.8) | 23.6 (20.1 - 27.6) |
| Open Claims | 412,547 | 882,505 | 391.3 | 950.3 | 3,221 | 8,249 | 8.23 (7.95 - 8.52) | 8.68 (8.49 - 8.87) |
| OEHR | 35,645 | 93,824 | 17.4 | 51.2 | 279 | 839 | 16.0 (14.2 - 18.0) | 16.4 (15.3 - 17.5) |
| VA | 0 | 0 | 0.0 | 0.0 | 0 | 0 | | |
| SIDIAP | 25,263 | 6,967 | 72.5 | 19.5 | 934 | 207 | 12.9 (12.1 - 13.7) | 10.6 (9.21 - 12.2) |
| Meta-analysis | 573,362 | 1,164,050 | 570.1 | 1181.6 | 5,623 | 11,944 | 9.86 (9.61 - 10.1) | 10.1 (9.93 - 10.3) |
| Meta-analysis (w/o USOC) | 160,815 | 281,545 | 178.8 | 231.3 | 2,402 | 3,695 | 13.4 (12.9 - 14.0) | 16.0 (15.5 - 16.5) |
| **PS-stratification / on-treatment TAR** | | | | | | | | |
| CCAE | 52,064 | 85,370 | 45.4 | 72.4 | 323 | 573 | 7.12 (6.36 - 7.94) | 7.91 (7.28 - 8.59) |
| DAG | 12,561 | 2,595 | 13.1 | 2.7 | 208 | 38 | 15.9 (13.8 - 18.2) | 14.2 (10.1 - 19.5) |
| ODOD | 29,538 | 67,542 | 24.6 | 62.0 | 515 | 1,534 | 21.0 (19.2 - 22.9) | 24.7 (23.5 - 26.0) |
| MDCR | 9,303 | 18,074 | 9.7 | 18.9 | 160 | 357 | 16.5 (14.1 - 19.3) | 18.9 (17.0 - 21.0) |
| MDCD | 4,006 | 9,470 | 3.0 | 6.7 | 52 | 158 | 17.2 (12.9 - 22.6) | 23.5 (20.0 - 27.4) |
| Open Claims | 419,848 | 882,516 | 400.5 | 950.3 | 3,291 | 8,250 | 8.22 (7.94 - 8.50) | 8.68 (8.49 - 8.87) |
| OEHR | 37,432 | 93,852 | 18.5 | 51.2 | 294 | 841 | 15.9 (14.1 - 17.8) | 16.4 (15.3 - 17.6) |
| VA | 24,038 | 219,997 | 22.1 | 228.7 | 315 | 3,958 | 14.2 (12.7 - 15.9) | 17.3 (16.8 - 17.9) |
| SIDIAP | 25,338 | 8,957 | 72.6 | 26.3 | 935 | 288 | 12.9 (12.1 - 13.7) | 10.9 (9.72 - 12.3) |
| Meta-analysis | 590,090 | 1,168,376 | 587.3 | 1190.5 | 5,778 | 12,039 | 9.84 (9.59 - 10.1) | 10.1 (9.93 - 10.3) |
| Meta-analysis (w/o USOC) | 170,242 | 285,860 | 186.8 | 240.2 | 2,487 | 3,789 | 13.3 (12.8 - 13.8) | 15.8 (15.3 - 16.3) |
| **PS-matching / on-treatment TAR without escalation** | | | | | | | | |
| CCAE | 46,699 | 83,866 | 32.8 | 61.9 | 236 | 496 | 7.19 (6.30 - 8.17) | 8.01 (7.32 - 8.75) |
| DAG | 12,541 | 2,235 | 11.8 | 2.1 | 182 | 32 | 15.4 (13.2 - 17.8) | 15.4 (10.5 - 21.7) |
| ODOD | 27,972 | 67,536 | 19.0 | 55.4 | 395 | 1,356 | 20.8 (18.8 - 23.0) | 24.5 (23.2 - 25.8) |
| MDCR | 8,781 | 17,691 | 7.8 | 16.8 | 126 | 321 | 16.2 (13.5 - 19.3) | 19.1 (17.1 - 21.3) |
| MDCD | 3,914 | 9,426 | 2.2 | 5.6 | 37 | 129 | 16.5 (11.6 - 22.7) | 23.0 (19.2 - 27.3) |
| Open Claims | 412,547 | 882,505 | 303.8 | 805.8 | 2,531 | 6,892 | 8.33 (8.01 - 8.66) | 8.55 (8.35 - 8.76) |
| OEHR | 35,645 | 93,824 | 14.2 | 44.7 | 212 | 625 | 14.9 (13.0 - 17.1) | 14.0 (12.9 - 15.1) |
| VA | 0 | 0 | 0.0 | 0.0 | 0 | 0 | | |
| SIDIAP | 25,263 | 6,967 | 53.3 | 16.3 | 674 | 167 | 12.6 (11.7 - 13.6) | 10.2 (8.73 - 11.9) |
| Meta-analysis | 573,362 | 1,164,050 | 445.0 | 1008.6 | 4,393 | 10,018 | 9.87 (9.58 - 10.2) | 9.93 (9.74 - 10.1) |
| Meta-analysis (w/o USOC) | 160,815 | 281,545 | 141.2 | 202.8 | 1,862 | 3,126 | 13.2 (12.6 - 13.8) | 15.4 (14.9 - 16.0) |
| **PS-stratification / on-treatment TAR without escalation** | | | | | | | | |
| CCAE | 52,064 | 85,370 | 36.7 | 63.1 | 258 | 499 | 7.02 (6.19 - 7.93) | 7.91 (7.23 - 8.64) |
| DAG | 12,561 | 2,595 | 11.9 | 2.4 | 183 | 36 | 15.4 (13.3 - 17.8) | 14.8 (10.3 - 20.4) |
| ODOD | 29,538 | 67,542 | 20.1 | 55.4 | 420 | 1,356 | 20.9 (18.9 - 23.0) | 24.5 (23.2 - 25.8) |
| MDCR | 9,303 | 18,074 | 8.3 | 17.2 | 134 | 325 | 16.2 (13.6 - 19.2) | 18.8 (16.8 - 21.0) |
| MDCD | 4,006 | 9,470 | 2.3 | 5.7 | 37 | 129 | 16.1 (11.3 - 22.2) | 22.8 (19.0 - 27.1) |
| Open Claims | 419,848 | 882,516 | 310.7 | 805.8 | 2,584 | 6,893 | 8.32 (8.00 - 8.64) | 8.55 (8.35 - 8.76) |
| OEHR | 37,432 | 93,852 | 15.1 | 44.7 | 223 | 626 | 14.8 (12.9 - 16.9) | 14.0 (12.9 - 15.1) |
| VA | 24,038 | 219,997 | 18.0 | 204.4 | 243 | 3,343 | 13.5 (11.9 - 15.3) | 16.4 (15.8 - 16.9) |
| SIDIAP | 25,338 | 8,957 | 53.4 | 21.8 | 675 | 233 | 12.6 (11.7 - 13.6) | 10.7 (9.36 - 12.2) |
| Meta-analysis | 590,090 | 1,168,376 | 458.4 | 1016.1 | 4,514 | 10,097 | 9.85 (9.56 - 10.1) | 9.94 (9.74 - 10.1) |
| Meta-analysis (w/o USOC) | 170,242 | 285,860 | 147.7 | 210.3 | 1,930 | 3,204 | 13.1 (12.5 - 13.7) | 15.2 (14.7 - 15.8) |

##### 4.6.18 Hypoglycemia

**Supplementary Table 437:** Exposures, outcomes and risk estimates for Hypoglycemia in DPP4I (male) (T) vs SU (male) (C) new-user cohorts under four different propensity score (PS) adjustment / time-at-risk (TAR) designs across data sources. We report total patient exposure time in 1,000 patient-year (KYr) units, uncalibrated and calibrated hazard ratios (HRs) and their 95% confidence intervals (CIs). Greyed out entries did not pass study diagnostics and are included only for completeness.

| | Patients | | Exposure (KYr) | | Outcomes | | Incidence (per KYr) | |
|---|---|---|---|---|---|---|---|---|
| | T | C | T | C | T | C | T | C |
| **PS-matching / on-treatment TAR** | | | | | | | | |
| CCAE | 46,846 | 84,213 | 40.9 | 71.6 | 112 | 544 | 2.74 (2.25 - 3.29) | 7.60 (6.97 - 8.26) |
| DAG | 13,335 | 2,343 | 14.2 | 2.4 | 23 | 12 | 1.62 (1.03 - 2.43) | 4.96 (2.57 - 8.67) |
| ODOD | 28,883 | 69,988 | 24.2 | 65.3 | 121 | 1,203 | 5.00 (4.15 - 5.98) | 18.4 (17.4 - 19.5) |
| MDCR | 8,973 | 18,118 | 9.4 | 19.0 | 33 | 322 | 3.53 (2.43 - 4.95) | 17.0 (15.2 - 18.9) |
| MDCD | 4,004 | 9,629 | 3.0 | 6.9 | 25 | 176 | 8.23 (5.33 - 12.2) | 25.7 (22.0 - 29.7) |
| Open Claims | 416,792 | 892,347 | 397.9 | 966.4 | 949 | 7,417 | 2.38 (2.24 - 2.54) | 7.67 (7.50 - 7.85) |
| OEHR | 36,212 | 95,196 | 17.9 | 52.5 | 100 | 761 | 5.59 (4.55 - 6.80) | 14.5 (13.5 - 15.6) |
| VA | 0 | 0 | 0.0 | 0.0 | 0 | 0 | | |
| SIDIAP | 26,767 | 7,208 | 78.3 | 20.4 | 51 | 64 | 0.65 (0.48 - 0.86) | 3.14 (2.42 - 4.01) |
| Meta-analysis | 581,812 | 1,179,042 | 585.9 | 1204.5 | 1,414 | 10,499 | 2.41 (2.29 - 2.54) | 8.72 (8.55 - 8.88) |
| Meta-analysis (w/o USOC) | 165,020 | 286,695 | 187.9 | 238.1 | 465 | 3,082 | 2.47 (2.25 - 2.71) | 12.9 (12.5 - 13.4) |
| **PS-stratification / on-treatment TAR** | | | | | | | | |
| CCAE | 52,171 | 85,721 | 45.8 | 72.9 | 125 | 549 | 2.73 (2.27 - 3.25) | 7.53 (6.92 - 8.19) |
| DAG | 13,363 | 2,719 | 14.2 | 2.8 | 23 | 18 | 1.62 (1.02 - 2.42) | 6.38 (3.78 - 10.1) |
| ODOD | 30,452 | 69,994 | 25.6 | 65.3 | 127 | 1,203 | 4.95 (4.13 - 5.89) | 18.4 (17.4 - 19.5) |
| MDCR | 9,475 | 18,512 | 9.9 | 19.4 | 34 | 326 | 3.42 (2.37 - 4.78) | 16.8 (15.0 - 18.7) |
| MDCD | 4,097 | 9,647 | 3.1 | 6.9 | 25 | 176 | 8.02 (5.19 - 11.8) | 25.6 (22.0 - 29.7) |
| Open Claims | 424,020 | 892,361 | 407.1 | 966.4 | 972 | 7,418 | 2.39 (2.24 - 2.54) | 7.68 (7.50 - 7.85) |
| OEHR | 38,025 | 95,226 | 19.0 | 52.5 | 104 | 763 | 5.48 (4.48 - 6.64) | 14.5 (13.5 - 15.6) |
| VA | 25,385 | 231,973 | 23.8 | 245.8 | 78 | 2,126 | 3.28 (2.60 - 4.10) | 8.65 (8.28 - 9.02) |
| SIDIAP | 26,860 | 9,258 | 78.4 | 27.5 | 51 | 77 | 0.65 (0.48 - 0.86) | 2.80 (2.21 - 3.50) |
| Meta-analysis | 598,463 | 1,183,438 | 603.3 | 1213.8 | 1,461 | 10,530 | 2.42 (2.30 - 2.55) | 8.68 (8.51 - 8.84) |
| Meta-analysis (w/o USOC) | 174,443 | 291,077 | 196.1 | 247.4 | 489 | 3,112 | 2.49 (2.28 - 2.72) | 12.6 (12.1 - 13.0) |
| **PS-matching / on-treatment TAR without escalation** | | | | | | | | |
| CCAE | 46,846 | 84,213 | 33.1 | 62.3 | 75 | 454 | 2.27 (1.78 - 2.84) | 7.29 (6.63 - 7.99) |
| DAG | 13,335 | 2,343 | 12.8 | 2.2 | 16 | 11 | 1.25 (0.71 - 2.03) | 4.99 (2.49 - 8.92) |
| ODOD | 28,883 | 69,988 | 19.9 | 58.3 | 63 | 1,046 | 3.17 (2.44 - 4.06) | 17.9 (16.9 - 19.1) |
| MDCR | 8,973 | 18,118 | 8.0 | 17.3 | 19 | 294 | 2.38 (1.43 - 3.71) | 17.0 (15.1 - 19.1) |
| MDCD | 4,004 | 9,629 | 2.3 | 5.8 | 10 | 142 | 4.32 (2.07 - 7.94) | 24.6 (20.7 - 29.0) |
| Open Claims | 416,792 | 892,347 | 308.8 | 819.3 | 524 | 6,105 | 1.70 (1.55 - 1.85) | 7.45 (7.27 - 7.64) |
| OEHR | 36,212 | 95,196 | 14.5 | 45.7 | 49 | 567 | 3.37 (2.49 - 4.45) | 12.4 (11.4 - 13.5) |
| VA | 0 | 0 | 0.0 | 0.0 | 0 | 0 | | |
| SIDIAP | 26,767 | 7,208 | 57.4 | 17.0 | 16 | 50 | 0.28 (0.16 - 0.45) | 2.93 (2.18 - 3.87) |
| Meta-analysis | 568,477 | 1,176,699 | 444.0 | 1025.7 | 756 | 8,658 | 1.70 (1.58 - 1.83) | 8.44 (8.26 - 8.62) |
| Meta-analysis (w/o USOC) | 151,685 | 284,352 | 135.2 | 206.4 | 232 | 2,553 | 1.72 (1.50 - 1.95) | 12.4 (11.9 - 12.9) |
| **PS-stratification / on-treatment TAR without escalation** | | | | | | | | |
| CCAE | 52,171 | 85,721 | 37.1 | 63.5 | 83 | 459 | 2.24 (1.78 - 2.78) | 7.23 (6.59 - 7.93) |
| DAG | 13,363 | 2,719 | 12.8 | 2.6 | 16 | 17 | 1.25 (0.71 - 2.02) | 6.58 (3.83 - 10.5) |
| ODOD | 30,452 | 69,994 | 21.0 | 58.3 | 67 | 1,046 | 3.19 (2.47 - 4.05) | 17.9 (16.9 - 19.1) |
| MDCR | 9,475 | 18,512 | 8.5 | 17.8 | 20 | 297 | 2.36 (1.44 - 3.64) | 16.7 (14.9 - 18.7) |
| MDCD | 4,097 | 9,647 | 2.4 | 5.8 | 10 | 142 | 4.21 (2.02 - 7.74) | 24.6 (20.7 - 29.0) |
| Open Claims | 424,020 | 892,361 | 315.7 | 819.3 | 537 | 6,106 | 1.70 (1.56 - 1.85) | 7.45 (7.27 - 7.64) |
| OEHR | 38,025 | 95,226 | 15.4 | 45.7 | 51 | 569 | 3.30 (2.46 - 4.34) | 12.4 (11.4 - 13.5) |
| VA | 25,385 | 231,973 | 19.3 | 219.1 | 44 | 1,738 | 2.28 (1.66 - 3.06) | 7.93 (7.56 - 8.31) |
| SIDIAP | 26,860 | 9,258 | 57.5 | 22.8 | 16 | 60 | 0.28 (0.16 - 0.45) | 2.64 (2.01 - 3.39) |
| Meta-analysis | 598,463 | 1,183,438 | 470.5 | 1035.6 | 800 | 8,696 | 1.70 (1.58 - 1.82) | 8.40 (8.22 - 8.58) |
| Meta-analysis (w/o USOC) | 174,443 | 291,077 | 154.7 | 216.4 | 263 | 2,590 | 1.70 (1.50 - 1.92) | 12.0 (11.5 - 12.4) |

##### 4.6.19 Hypotension

**Supplementary Table 438:** Exposures, outcomes and risk estimates for Hypotension in DPP4I (male) (T) vs SU (male) (C) new-user cohorts under four different propensity score (PS) adjustment / time-at-risk (TAR) designs across data sources. We report total patient exposure time in 1,000 patient-year (KYr) units, uncalibrated and calibrated hazard ratios (HRs) and their 95% confidence intervals (CIs). Greyed out entries did not pass study diagnostics and are included only for completeness.

| | Patients | | Exposure (KYr) | | Outcomes | | Incidence (per KYr) | |
|---|---|---|---|---|---|---|---|---|
| | T | C | T | C | T | C | T | C |
| **PS-matching / on-treatment TAR** | | | | | | | | |
| CCAE | 46,654 | 83,703 | 40.6 | 71.3 | 317 | 525 | 7.80 (6.97 - 8.71) | 7.36 (6.75 - 8.02) |
| DAG | 13,110 | 2,308 | 13.9 | 2.4 | 69 | 14 | 4.95 (3.85 - 6.27) | 5.91 (3.23 - 9.91) |
| ODOD | 27,836 | 67,242 | 23.1 | 62.2 | 491 | 1,548 | 21.2 (19.4 - 23.2) | 24.9 (23.7 - 26.2) |
| MDCR | 8,487 | 17,145 | 8.7 | 17.9 | 242 | 474 | 27.8 (24.4 - 31.5) | 26.5 (24.2 - 29.0) |
| MDCD | 3,796 | 9,127 | 2.9 | 6.4 | 94 | 183 | 32.9 (26.6 - 40.3) | 28.4 (24.5 - 32.9) |
| Open Claims | 407,233 | 871,628 | 387.0 | 942.2 | 3,663 | 8,953 | 9.46 (9.16 - 9.78) | 9.50 (9.31 - 9.70) |
| OEHR | 35,607 | 93,678 | 17.4 | 51.2 | 325 | 954 | 18.7 (16.7 - 20.8) | 18.6 (17.5 - 19.8) |
| VA | 0 | 0 | 0.0 | 0.0 | 0 | 0 | | |
| SIDIAP | 26,630 | 7,194 | 77.8 | 20.4 | 147 | 20 | 1.89 (1.60 - 2.22) | 0.98 (0.60 - 1.51) |
| Meta-analysis | 569,353 | 1,152,025 | 571.5 | 1174.1 | 5,348 | 12,671 | 9.36 (9.11 - 9.61) | 10.8 (10.6 - 11.0) |
| Meta-analysis (w/o USOC) | 162,120 | 280,397 | 184.5 | 231.9 | 1,685 | 3,718 | 9.13 (8.70 - 9.58) | 16.0 (15.5 - 16.6) |
| **PS-stratification / on-treatment TAR** | | | | | | | | |
| CCAE | 52,015 | 85,196 | 45.5 | 72.5 | 348 | 530 | 7.66 (6.87 - 8.50) | 7.31 (6.70 - 7.95) |
| DAG | 13,137 | 2,665 | 14.0 | 2.8 | 69 | 16 | 4.94 (3.85 - 6.26) | 5.79 (3.31 - 9.41) |
| ODOD | 29,403 | 67,247 | 24.6 | 62.2 | 520 | 1,548 | 21.2 (19.4 - 23.1) | 24.9 (23.7 - 26.2) |
| MDCR | 8,981 | 17,519 | 9.3 | 18.3 | 263 | 486 | 28.3 (25.0 - 32.0) | 26.5 (24.2 - 29.0) |
| MDCD | 3,870 | 9,169 | 2.9 | 6.5 | 95 | 186 | 32.6 (26.3 - 39.8) | 28.7 (24.7 - 33.1) |
| Open Claims | 414,563 | 871,639 | 396.3 | 942.2 | 3,770 | 8,954 | 9.51 (9.21 - 9.82) | 9.50 (9.31 - 9.70) |
| OEHR | 37,425 | 93,706 | 18.5 | 51.2 | 345 | 957 | 18.7 (16.7 - 20.7) | 18.7 (17.5 - 19.9) |
| VA | 24,071 | 221,458 | 22.3 | 232.2 | 285 | 3,230 | 12.8 (11.4 - 14.4) | 13.9 (13.4 - 14.4) |
| SIDIAP | 26,719 | 9,245 | 77.9 | 27.6 | 147 | 24 | 1.89 (1.59 - 2.22) | 0.87 (0.56 - 1.30) |
| Meta-analysis | 586,113 | 1,156,386 | 588.9 | 1183.3 | 5,557 | 12,701 | 9.44 (9.19 - 9.69) | 10.7 (10.5 - 10.9) |
| Meta-analysis (w/o USOC) | 171,550 | 284,747 | 192.6 | 241.1 | 1,787 | 3,747 | 9.28 (8.85 - 9.72) | 15.5 (15.0 - 16.0) |
| **PS-matching / on-treatment TAR without escalation** | | | | | | | | |
| CCAE | 46,654 | 83,703 | 32.8 | 62.0 | 256 | 432 | 7.80 (6.87 - 8.81) | 6.97 (6.32 - 7.65) |
| DAG | 13,110 | 2,308 | 12.6 | 2.2 | 61 | 13 | 4.86 (3.72 - 6.24) | 6.02 (3.20 - 10.3) |
| ODOD | 27,836 | 67,242 | 19.0 | 55.5 | 383 | 1,397 | 20.2 (18.2 - 22.3) | 25.2 (23.9 - 26.5) |
| MDCR | 8,487 | 17,145 | 7.4 | 16.3 | 208 | 415 | 28.0 (24.3 - 32.1) | 25.4 (23.0 - 28.0) |
| MDCD | 3,796 | 9,127 | 2.2 | 5.4 | 74 | 153 | 34.0 (26.7 - 42.7) | 28.3 (24.0 - 33.1) |
| Open Claims | 407,233 | 871,628 | 300.2 | 798.5 | 2,840 | 7,508 | 9.46 (9.12 - 9.81) | 9.40 (9.19 - 9.62) |
| OEHR | 35,607 | 93,678 | 14.2 | 44.7 | 222 | 666 | 15.6 (13.6 - 17.8) | 14.9 (13.8 - 16.1) |
| VA | 0 | 0 | 0.0 | 0.0 | 0 | 0 | | |
| SIDIAP | 26,630 | 7,194 | 57.0 | 17.1 | 116 | 18 | 2.04 (1.68 - 2.44) | 1.06 (0.63 - 1.67) |
| Meta-analysis | 569,353 | 1,152,025 | 445.4 | 1001.8 | 4,160 | 10,602 | 9.34 (9.06 - 9.63) | 10.6 (10.4 - 10.8) |
| Meta-analysis (w/o USOC) | 162,120 | 280,397 | 145.2 | 203.2 | 1,320 | 3,094 | 9.09 (8.61 - 9.60) | 15.2 (14.7 - 15.8) |
| **PS-stratification / on-treatment TAR without escalation** | | | | | | | | |
| CCAE | 52,015 | 85,196 | 36.8 | 63.2 | 278 | 437 | 7.56 (6.70 - 8.50) | 6.92 (6.28 - 7.60) |
| DAG | 13,137 | 2,665 | 12.6 | 2.5 | 61 | 15 | 4.85 (3.71 - 6.23) | 5.94 (3.32 - 9.79) |
| ODOD | 29,403 | 67,247 | 20.1 | 55.5 | 407 | 1,397 | 20.3 (18.3 - 22.3) | 25.2 (23.9 - 26.5) |
| MDCR | 8,981 | 17,519 | 7.9 | 16.8 | 226 | 426 | 28.5 (24.9 - 32.5) | 25.4 (23.1 - 28.0) |
| MDCD | 3,870 | 9,169 | 2.2 | 5.5 | 75 | 156 | 33.7 (26.5 - 42.3) | 28.6 (24.3 - 33.5) |
| Open Claims | 414,563 | 871,639 | 307.2 | 798.5 | 2,921 | 7,509 | 9.51 (9.17 - 9.86) | 9.40 (9.19 - 9.62) |
| OEHR | 37,425 | 93,706 | 15.1 | 44.7 | 240 | 667 | 15.9 (13.9 - 18.0) | 14.9 (13.8 - 16.1) |
| VA | 24,071 | 221,458 | 18.1 | 207.3 | 210 | 2,664 | 11.6 (10.1 - 13.3) | 12.8 (12.4 - 13.3) |
| SIDIAP | 26,719 | 9,245 | 57.1 | 22.8 | 116 | 21 | 2.03 (1.68 - 2.44) | 0.92 (0.57 - 1.41) |
| Meta-analysis | 586,113 | 1,156,386 | 459.0 | 1009.5 | 4,324 | 10,628 | 9.42 (9.14 - 9.71) | 10.5 (10.3 - 10.7) |
| Meta-analysis (w/o USOC) | 171,550 | 284,747 | 151.8 | 210.9 | 1,403 | 3,119 | 9.24 (8.76 - 9.74) | 14.8 (14.3 - 15.3) |

##### 4.6.20 Joint pain

**Supplementary Table 439:** Exposures, outcomes and risk estimates for Joint pain in DPP4I (male) (T) vs SU (male) (C) new-user cohorts under four different propensity score (PS) adjustment / time-at-risk (TAR) designs across data sources. We report total patient exposure time in 1,000 patient-year (KYr) units, uncalibrated and calibrated hazard ratios (HRs) and their 95% confidence intervals (CIs). Greyed out entries did not pass study diagnostics and are included only for completeness.

| | Patients | | Exposure (KYr) | | Outcomes | | Incidence (per KYr) | |
|---|---|---|---|---|---|---|---|---|
| | T | C | T | C | T | C | T | C |
| **PS-matching / on-treatment TAR** | | | | | | | | |
| CCAE | 45,710 | 82,050 | 39.8 | 69.5 | 329 | 581 | 8.27 (7.40 - 9.21) | 8.36 (7.70 - 9.07) |
| DAG | 11,582 | 2,086 | 12.0 | 2.1 | 264 | 33 | 21.9 (19.4 - 24.8) | 15.7 (10.8 - 22.0) |
| ODOD | 27,948 | 67,874 | 23.3 | 63.3 | 250 | 646 | 10.7 (9.44 - 12.1) | 10.2 (9.43 - 11.0) |
| MDCR | 8,776 | 17,714 | 9.1 | 18.6 | 82 | 162 | 9.00 (7.16 - 11.2) | 8.72 (7.43 - 10.2) |
| MDCD | 3,800 | 9,203 | 2.9 | 6.5 | 49 | 85 | 17.1 (12.7 - 22.6) | 13.0 (10.4 - 16.1) |
| Open Claims | 403,632 | 867,294 | 384.4 | 940.1 | 2,011 | 4,493 | 5.23 (5.00 - 5.46) | 4.78 (4.64 - 4.92) |
| OEHR | 35,054 | 92,497 | 17.2 | 50.9 | 139 | 431 | 8.06 (6.78 - 9.52) | 8.47 (7.69 - 9.31) |
| VA | 0 | 0 | 0.0 | 0.0 | 0 | 0 | | |
| SIDIAP | 22,584 | 6,504 | 65.1 | 17.6 | 1,213 | 378 | 18.6 (17.6 - 19.7) | 21.5 (19.4 - 23.8) |
| Meta-analysis | 559,086 | 1,145,222 | 553.8 | 1168.5 | 4,337 | 6,809 | 7.83 (7.60 - 8.07) | 5.83 (5.69 - 5.97) |
| Meta-analysis (w/o USOC) | 155,454 | 277,928 | 169.4 | 228.5 | 2,326 | 2,316 | 13.7 (13.2 - 14.3) | 10.1 (9.73 - 10.6) |
| **PS-stratification / on-treatment TAR** | | | | | | | | |
| CCAE | 50,908 | 83,525 | 44.5 | 70.7 | 376 | 582 | 8.46 (7.63 - 9.36) | 8.23 (7.58 - 8.93) |
| DAG | 11,607 | 2,420 | 12.1 | 2.5 | 264 | 40 | 21.9 (19.3 - 24.7) | 16.3 (11.6 - 22.2) |
| ODOD | 29,498 | 67,881 | 24.7 | 63.3 | 265 | 646 | 10.7 (9.47 - 12.1) | 10.2 (9.43 - 11.0) |
| MDCR | 9,268 | 18,102 | 9.7 | 19.1 | 90 | 162 | 9.29 (7.47 - 11.4) | 8.50 (7.24 - 9.92) |
| MDCD | 3,898 | 9,221 | 2.9 | 6.5 | 51 | 87 | 17.3 (12.9 - 22.8) | 13.3 (10.7 - 16.4) |
| Open Claims | 410,700 | 867,316 | 393.3 | 940.1 | 2,082 | 4,493 | 5.29 (5.07 - 5.53) | 4.78 (4.64 - 4.92) |
| OEHR | 36,839 | 92,525 | 18.3 | 50.9 | 152 | 431 | 8.31 (7.04 - 9.74) | 8.47 (7.69 - 9.31) |
| VA | 22,503 | 206,997 | 21.2 | 218.8 | 108 | 1,714 | 5.09 (4.18 - 6.15) | 7.84 (7.47 - 8.22) |
| SIDIAP | 22,781 | 8,424 | 65.3 | 23.9 | 1,214 | 511 | 18.6 (17.6 - 19.7) | 21.4 (19.6 - 23.4) |
| Meta-analysis | 575,499 | 1,149,414 | 570.7 | 1176.9 | 4,494 | 6,952 | 7.87 (7.65 - 8.11) | 5.91 (5.77 - 6.05) |
| Meta-analysis (w/o USOC) | 164,799 | 282,098 | 177.4 | 236.8 | 2,412 | 2,459 | 13.6 (13.1 - 14.2) | 10.4 (9.98 - 10.8) |
| **PS-matching / on-treatment TAR without escalation** | | | | | | | | |
| CCAE | 45,710 | 82,050 | 32.2 | 60.4 | 267 | 507 | 8.29 (7.33 - 9.35) | 8.39 (7.67 - 9.15) |
| DAG | 11,582 | 2,086 | 10.9 | 1.9 | 249 | 32 | 22.9 (20.1 - 25.9) | 16.7 (11.4 - 23.5) |
| ODOD | 27,948 | 67,874 | 19.1 | 56.5 | 213 | 576 | 11.1 (9.70 - 12.7) | 10.2 (9.38 - 11.1) |
| MDCR | 8,776 | 17,714 | 7.8 | 16.9 | 68 | 147 | 8.75 (6.80 - 11.1) | 8.67 (7.33 - 10.2) |
| MDCD | 3,800 | 9,203 | 2.2 | 5.5 | 35 | 70 | 15.8 (11.0 - 22.0) | 12.8 (9.96 - 16.1) |
| Open Claims | 403,632 | 867,294 | 298.3 | 796.8 | 1,525 | 3,799 | 5.11 (4.86 - 5.38) | 4.77 (4.62 - 4.92) |
| OEHR | 35,054 | 92,497 | 14.0 | 44.3 | 109 | 354 | 7.77 (6.38 - 9.38) | 8.00 (7.18 - 8.87) |
| VA | 0 | 0 | 0.0 | 0.0 | 0 | 0 | | |
| SIDIAP | 22,584 | 6,504 | 47.9 | 14.8 | 875 | 320 | 18.3 (17.1 - 19.5) | 21.6 (19.3 - 24.1) |
| Meta-analysis | 559,086 | 1,145,222 | 432.4 | 997.2 | 3,341 | 5,805 | 7.73 (7.47 - 7.99) | 5.82 (5.67 - 5.97) |
| Meta-analysis (w/o USOC) | 155,454 | 277,928 | 134.1 | 200.4 | 1,816 | 2,006 | 13.5 (12.9 - 14.2) | 10.0 (9.58 - 10.5) |
| **PS-stratification / on-treatment TAR without escalation** | | | | | | | | |
| CCAE | 50,908 | 83,525 | 36.0 | 61.6 | 310 | 508 | 8.61 (7.68 - 9.63) | 8.25 (7.55 - 9.00) |
| DAG | 11,607 | 2,420 | 10.9 | 2.2 | 249 | 39 | 22.8 (20.1 - 25.9) | 17.4 (12.4 - 23.8) |
| ODOD | 29,498 | 67,881 | 20.3 | 56.5 | 226 | 576 | 11.2 (9.75 - 12.7) | 10.2 (9.38 - 11.1) |
| MDCR | 9,268 | 18,102 | 8.3 | 17.4 | 76 | 147 | 9.20 (7.25 - 11.5) | 8.45 (7.14 - 9.93) |
| MDCD | 3,898 | 9,221 | 2.3 | 5.5 | 36 | 72 | 15.9 (11.1 - 22.0) | 13.1 (10.3 - 16.5) |
| Open Claims | 410,700 | 867,316 | 305.1 | 796.8 | 1,578 | 3,799 | 5.17 (4.92 - 5.43) | 4.77 (4.62 - 4.92) |
| OEHR | 36,839 | 92,525 | 14.9 | 44.3 | 120 | 354 | 8.06 (6.68 - 9.64) | 7.99 (7.18 - 8.87) |
| VA | 22,503 | 206,997 | 17.2 | 195.4 | 85 | 1,548 | 4.93 (3.94 - 6.10) | 7.92 (7.53 - 8.33) |
| SIDIAP | 22,781 | 8,424 | 48.1 | 19.9 | 875 | 424 | 18.2 (17.0 - 19.4) | 21.3 (19.3 - 23.4) |
| Meta-analysis | 575,499 | 1,149,414 | 445.7 | 1004.3 | 3,470 | 5,919 | 7.79 (7.53 - 8.05) | 5.89 (5.74 - 6.05) |
| Meta-analysis (w/o USOC) | 164,799 | 282,098 | 140.7 | 207.4 | 1,892 | 2,120 | 13.5 (12.9 - 14.1) | 10.2 (9.79 - 10.7) |

##### 4.6.21 Lower extremity amputation

**Supplementary Table 440:** Exposures, outcomes and risk estimates for Lower extremity amputation in DPP4I (male) (T) vs SU (male) (C) new-user cohorts under four different propensity score (PS) adjustment / time-at-risk (TAR) designs across data sources. We report total patient exposure time in 1,000 patient-year (KYr) units, uncalibrated and calibrated hazard ratios (HRs) and their 95% confidence intervals (CIs). Greyed out entries did not pass study diagnostics and are included only for completeness.

| | Patients | | Exposure (KYr) | | Outcomes | | Incidence (per KYr) | |
|---|---|---|---|---|---|---|---|---|
| | T | C | T | C | T | C | T | C |
| **PS-matching / on-treatment TAR** | | | | | | | | |
| CCAE | 47,380 | 85,055 | 41.5 | 72.6 | 33 | 75 | 0.79 (0.55 - 1.12) | 1.03 (0.81 - 1.29) |
| DAG | 13,419 | 2,361 | 14.3 | 2.4 | 0 | 0 | | |
| ODOD | 29,180 | 70,617 | 24.5 | 66.4 | 27 | 102 | 1.10 (0.73 - 1.60) | 1.54 (1.25 - 1.86) |
| MDCR | 9,031 | 18,228 | 9.4 | 19.2 | 10 | 28 | 1.06 (0.51 - 1.95) | 1.46 (0.97 - 2.11) |
| MDCD | 4,059 | 9,787 | 3.1 | 7.1 | 9 | 13 | 2.92 (1.33 - 5.53) | 1.84 (0.98 - 3.15) |
| Open Claims | 421,580 | 901,057 | 402.8 | 979.0 | 240 | 832 | 0.60 (0.52 - 0.68) | 0.85 (0.79 - 0.91) |
| OEHR | 36,410 | 95,792 | 18.1 | 53.3 | 15 | 83 | 0.83 (0.46 - 1.37) | 1.56 (1.24 - 1.93) |
| VA | 0 | 0 | 0.0 | 0.0 | 0 | 0 | | |
| SIDIAP | 26,751 | 7,203 | 78.3 | 20.4 | 32 | 11 | 0.41 (0.28 - 0.58) | 0.54 (0.27 - 0.96) |
| Meta-analysis | 574,391 | 1,187,739 | 577.7 | 1218.0 | 366 | 1,144 | 0.63 (0.57 - 0.70) | 0.94 (0.89 - 1.00) |
| Meta-analysis (w/o USOC) | 152,811 | 286,682 | 174.9 | 239.0 | 126 | 312 | 0.72 (0.60 - 0.86) | 1.31 (1.16 - 1.46) |
| **PS-stratification / on-treatment TAR** | | | | | | | | |
| CCAE | 52,823 | 86,574 | 46.5 | 73.9 | 33 | 76 | 0.71 (0.49 - 1.00) | 1.03 (0.81 - 1.29) |
| DAG | 13,447 | 2,736 | 14.3 | 2.8 | 0 | 0 | | |
| ODOD | 30,790 | 70,624 | 26.0 | 66.4 | 28 | 102 | 1.08 (0.71 - 1.55) | 1.54 (1.25 - 1.86) |
| MDCR | 9,542 | 18,619 | 10.0 | 19.7 | 10 | 28 | 1.00 (0.48 - 1.84) | 1.42 (0.95 - 2.06) |
| MDCD | 4,154 | 9,807 | 3.2 | 7.1 | 9 | 13 | 2.83 (1.30 - 5.38) | 1.84 (0.98 - 3.15) |
| Open Claims | 428,977 | 901,071 | 412.2 | 979.1 | 244 | 832 | 0.59 (0.52 - 0.67) | 0.85 (0.79 - 0.91) |
| OEHR | 38,251 | 95,822 | 19.2 | 53.3 | 16 | 83 | 0.83 (0.48 - 1.35) | 1.56 (1.24 - 1.93) |
| VA | 25,577 | 233,270 | 24.0 | 248.3 | 13 | 201 | 0.54 (0.29 - 0.93) | 0.81 (0.70 - 0.93) |
| SIDIAP | 26,844 | 9,248 | 78.4 | 27.6 | 32 | 20 | 0.41 (0.28 - 0.58) | 0.73 (0.44 - 1.12) |
| Meta-analysis | 591,381 | 1,191,765 | 595.4 | 1226.9 | 372 | 1,154 | 0.62 (0.56 - 0.69) | 0.94 (0.89 - 1.00) |
| Meta-analysis (w/o USOC) | 162,404 | 290,694 | 183.2 | 247.9 | 128 | 322 | 0.70 (0.58 - 0.83) | 1.30 (1.16 - 1.45) |
| **PS-matching / on-treatment TAR without escalation** | | | | | | | | |
| CCAE | 47,380 | 85,055 | 33.5 | 63.1 | 25 | 59 | 0.75 (0.48 - 1.10) | 0.94 (0.71 - 1.21) |
| DAG | 13,419 | 2,361 | 12.9 | 2.2 | 0 | 0 | | |
| ODOD | 29,180 | 70,617 | 20.1 | 59.2 | 20 | 86 | 1.00 (0.61 - 1.54) | 1.45 (1.16 - 1.79) |
| MDCR | 9,031 | 18,228 | 8.0 | 17.5 | 10 | 25 | 1.25 (0.60 - 2.29) | 1.43 (0.92 - 2.11) |
| MDCD | 4,059 | 9,787 | 2.4 | 5.9 | 7 | 10 | 2.98 (1.20 - 6.13) | 1.70 (0.81 - 3.12) |
| Open Claims | 421,580 | 901,057 | 312.3 | 829.1 | 181 | 670 | 0.58 (0.50 - 0.67) | 0.81 (0.75 - 0.87) |
| OEHR | 36,410 | 95,792 | 14.6 | 46.2 | 10 | 44 | 0.68 (0.33 - 1.26) | 0.95 (0.69 - 1.28) |
| VA | 0 | 0 | 0.0 | 0.0 | 0 | 0 | | |
| SIDIAP | 26,751 | 7,203 | 57.3 | 17.1 | 21 | 7 | 0.37 (0.23 - 0.56) | 0.41 (0.16 - 0.84) |
| Meta-analysis | 570,332 | 1,177,952 | 446.0 | 1032.2 | 267 | 891 | 0.60 (0.53 - 0.67) | 0.86 (0.81 - 0.92) |
| Meta-analysis (w/o USOC) | 148,752 | 276,895 | 133.6 | 203.1 | 86 | 221 | 0.64 (0.51 - 0.79) | 1.09 (0.95 - 1.24) |
| **PS-stratification / on-treatment TAR without escalation** | | | | | | | | |
| CCAE | 52,823 | 86,574 | 37.6 | 64.3 | 25 | 60 | 0.67 (0.43 - 0.98) | 0.93 (0.71 - 1.20) |
| DAG | 13,447 | 2,736 | 12.9 | 2.6 | 0 | 0 | | |
| ODOD | 30,790 | 70,624 | 21.3 | 59.2 | 21 | 86 | 0.99 (0.61 - 1.51) | 1.45 (1.16 - 1.79) |
| MDCR | 9,542 | 18,619 | 8.5 | 17.9 | 10 | 25 | 1.17 (0.56 - 2.15) | 1.39 (0.90 - 2.06) |
| MDCD | 4,154 | 9,807 | 2.4 | 5.9 | 7 | 10 | 2.90 (1.17 - 5.98) | 1.69 (0.81 - 3.11) |
| Open Claims | 428,977 | 901,071 | 319.4 | 829.1 | 185 | 670 | 0.58 (0.50 - 0.67) | 0.81 (0.75 - 0.87) |
| OEHR | 38,251 | 95,822 | 15.6 | 46.3 | 11 | 44 | 0.71 (0.35 - 1.26) | 0.95 (0.69 - 1.28) |
| VA | 25,577 | 233,270 | 19.5 | 221.1 | 11 | 150 | 0.57 (0.28 - 1.01) | 0.68 (0.57 - 0.80) |
| SIDIAP | 26,844 | 9,248 | 57.4 | 22.8 | 21 | 12 | 0.37 (0.23 - 0.56) | 0.53 (0.27 - 0.92) |
| Meta-analysis | 587,227 | 1,181,958 | 459.8 | 1039.6 | 273 | 897 | 0.59 (0.53 - 0.67) | 0.86 (0.81 - 0.92) |
| Meta-analysis (w/o USOC) | 158,250 | 280,887 | 140.4 | 210.5 | 88 | 227 | 0.63 (0.50 - 0.77) | 1.08 (0.94 - 1.23) |

##### 4.6.22 Nausea

**Supplementary Table 441:** Exposures, outcomes and risk estimates for Nausea in DPP4I (male) (T) vs SU (male) (C) new-user cohorts under four different propensity score (PS) adjustment / time-at-risk (TAR) designs across data sources. We report total patient exposure time in 1,000 patient-year (KYr) units, uncalibrated and calibrated hazard ratios (HRs) and their 95% confidence intervals (CIs). Greyed out entries did not pass study diagnostics and are included only for completeness.

| | Patients | | Exposure (KYr) | | Outcomes | | Incidence (per KYr) | |
|---|---|---|---|---|---|---|---|---|
| | T | C | T | C | T | C | T | C |
| **PS-matching / on-treatment TAR** | | | | | | | | |
| CCAE | 43,894 | 78,606 | 38.0 | 66.6 | 863 | 1,393 | 22.7 (21.2 - 24.3) | 20.9 (19.8 - 22.0) |
| DAG | 12,654 | 2,255 | 13.4 | 2.3 | 115 | 25 | 8.57 (7.07 - 10.3) | 11.0 (7.11 - 16.2) |
| ODOD | 26,343 | 63,660 | 21.7 | 58.8 | 677 | 1,934 | 31.2 (28.9 - 33.6) | 32.9 (31.4 - 34.4) |
| MDCR | 8,327 | 16,640 | 8.6 | 17.4 | 229 | 533 | 26.7 (23.4 - 30.4) | 30.7 (28.1 - 33.4) |
| MDCD | 3,262 | 7,837 | 2.4 | 5.5 | 134 | 341 | 55.4 (46.4 - 65.7) | 61.5 (55.2 - 68.4) |
| Open Claims | 383,482 | 820,475 | 364.1 | 889.5 | 5,625 | 13,804 | 15.4 (15.0 - 15.9) | 15.5 (15.3 - 15.8) |
| OEHR | 34,141 | 89,528 | 16.6 | 48.5 | 443 | 1,327 | 26.8 (24.3 - 29.4) | 27.4 (25.9 - 28.9) |
| VA | 0 | 0 | 0.0 | 0.0 | 0 | 0 | | |
| SIDIAP | 26,189 | 7,117 | 76.4 | 20.1 | 345 | 73 | 4.51 (4.05 - 5.02) | 3.63 (2.85 - 4.57) |
| Meta-analysis | 538,292 | 1,086,118 | 541.3 | 1108.7 | 8,431 | 19,430 | 15.6 (15.2 - 15.9) | 17.5 (17.3 - 17.8) |
| Meta-analysis (w/o USOC) | 154,810 | 265,643 | 177.1 | 219.2 | 2,806 | 5,626 | 15.8 (15.3 - 16.4) | 25.7 (25.0 - 26.3) |
| **PS-stratification / on-treatment TAR** | | | | | | | | |
| CCAE | 49,023 | 80,057 | 42.5 | 67.8 | 977 | 1,411 | 23.0 (21.6 - 24.5) | 20.8 (19.7 - 21.9) |
| DAG | 12,680 | 2,609 | 13.4 | 2.7 | 115 | 31 | 8.55 (7.06 - 10.3) | 11.6 (7.92 - 16.5) |
| ODOD | 27,811 | 63,666 | 23.0 | 58.8 | 724 | 1,934 | 31.4 (29.2 - 33.8) | 32.9 (31.4 - 34.4) |
| MDCR | 8,802 | 17,104 | 9.1 | 17.9 | 238 | 541 | 26.1 (22.9 - 29.6) | 30.2 (27.7 - 32.9) |
| MDCD | 3,340 | 7,870 | 2.5 | 5.6 | 138 | 343 | 55.6 (46.7 - 65.7) | 61.5 (55.2 - 68.4) |
| Open Claims | 390,539 | 820,488 | 373.0 | 889.5 | 5,784 | 13,805 | 15.5 (15.1 - 15.9) | 15.5 (15.3 - 15.8) |
| OEHR | 35,896 | 89,556 | 17.6 | 48.5 | 462 | 1,328 | 26.3 (23.9 - 28.8) | 27.4 (25.9 - 28.9) |
| VA | 23,930 | 221,000 | 22.4 | 233.9 | 192 | 2,238 | 8.56 (7.39 - 9.85) | 9.57 (9.18 - 9.97) |
| SIDIAP | 26,278 | 9,165 | 76.5 | 27.1 | 345 | 101 | 4.51 (4.04 - 5.01) | 3.72 (3.03 - 4.52) |
| Meta-analysis | 554,369 | 1,090,515 | 557.7 | 1118.0 | 8,783 | 19,494 | 15.7 (15.4 - 16.1) | 17.4 (17.2 - 17.7) |
| Meta-analysis (w/o USOC) | 163,830 | 270,027 | 184.7 | 228.5 | 2,999 | 5,689 | 16.2 (15.7 - 16.8) | 24.9 (24.3 - 25.6) |
| **PS-matching / on-treatment TAR without escalation** | | | | | | | | |
| CCAE | 43,894 | 78,606 | 30.8 | 58.2 | 711 | 1,201 | 23.1 (21.4 - 24.8) | 20.6 (19.5 - 21.8) |
| DAG | 12,654 | 2,255 | 12.1 | 2.1 | 103 | 24 | 8.50 (6.94 - 10.3) | 11.5 (7.40 - 17.2) |
| ODOD | 26,343 | 63,660 | 17.9 | 52.6 | 558 | 1,706 | 31.2 (28.7 - 33.9) | 32.4 (30.9 - 34.0) |
| MDCR | 8,327 | 16,640 | 7.3 | 15.9 | 200 | 476 | 27.3 (23.7 - 31.4) | 30.0 (27.4 - 32.8) |
| MDCD | 3,262 | 7,837 | 1.9 | 4.7 | 108 | 296 | 58.1 (47.7 - 70.2) | 62.7 (55.8 - 70.3) |
| Open Claims | 383,482 | 820,475 | 283.6 | 756.3 | 4,294 | 11,432 | 15.1 (14.7 - 15.6) | 15.1 (14.8 - 15.4) |
| OEHR | 34,141 | 89,528 | 13.6 | 42.5 | 327 | 1,021 | 24.1 (21.6 - 26.9) | 24.0 (22.6 - 25.6) |
| VA | 0 | 0 | 0.0 | 0.0 | 0 | 0 | | |
| SIDIAP | 26,189 | 7,117 | 56.0 | 16.8 | 260 | 60 | 4.64 (4.09 - 5.24) | 3.57 (2.73 - 4.60) |
| Meta-analysis | 538,292 | 1,086,118 | 423.2 | 949.1 | 6,561 | 16,216 | 15.5 (15.1 - 15.9) | 17.1 (16.8 - 17.4) |
| Meta-analysis (w/o USOC) | 154,810 | 265,643 | 139.6 | 192.8 | 2,267 | 4,784 | 16.2 (15.6 - 16.9) | 24.8 (24.1 - 25.5) |
| **PS-stratification / on-treatment TAR without escalation** | | | | | | | | |
| CCAE | 49,023 | 80,057 | 34.5 | 59.3 | 801 | 1,218 | 23.2 (21.6 - 24.9) | 20.5 (19.4 - 21.7) |
| DAG | 12,680 | 2,609 | 12.1 | 2.4 | 103 | 30 | 8.48 (6.92 - 10.3) | 12.3 (8.30 - 17.6) |
| ODOD | 27,811 | 63,666 | 18.9 | 52.6 | 596 | 1,706 | 31.5 (29.0 - 34.1) | 32.4 (30.9 - 34.0) |
| MDCR | 8,802 | 17,104 | 7.8 | 16.4 | 207 | 484 | 26.6 (23.1 - 30.5) | 29.5 (27.0 - 32.3) |
| MDCD | 3,340 | 7,870 | 1.9 | 4.7 | 111 | 298 | 58.2 (47.9 - 70.1) | 62.8 (55.8 - 70.3) |
| Open Claims | 390,539 | 820,488 | 290.3 | 756.3 | 4,408 | 11,433 | 15.2 (14.7 - 15.6) | 15.1 (14.8 - 15.4) |
| OEHR | 35,896 | 89,556 | 14.4 | 42.5 | 340 | 1,021 | 23.6 (21.2 - 26.2) | 24.0 (22.6 - 25.5) |
| VA | 23,930 | 221,000 | 18.3 | 208.8 | 158 | 1,880 | 8.64 (7.35 - 10.1) | 9.00 (8.60 - 9.42) |
| SIDIAP | 26,278 | 9,165 | 56.1 | 22.5 | 260 | 78 | 4.63 (4.09 - 5.23) | 3.47 (2.74 - 4.33) |
| Meta-analysis | 554,369 | 1,090,515 | 436.1 | 956.8 | 6,826 | 16,268 | 15.7 (15.3 - 16.0) | 17.0 (16.7 - 17.3) |
| Meta-analysis (w/o USOC) | 163,830 | 270,027 | 145.8 | 200.5 | 2,418 | 4,835 | 16.6 (15.9 - 17.3) | 24.1 (23.4 - 24.8) |

##### 4.6.23 Peripheral edema

**Supplementary Table 442:** Exposures, outcomes and risk estimates for Peripheral edema in DPP4I (male) (T) vs SU (male) (C) new-user cohorts under four different propensity score (PS) adjustment / time-at-risk (TAR) designs across data sources. We report total patient exposure time in 1,000 patient-year (KYr) units, uncalibrated and calibrated hazard ratios (HRs) and their 95% confidence intervals (CIs). Greyed out entries did not pass study diagnostics and are included only for completeness.

| | Patients | | Exposure (KYr) | | Outcomes | | Incidence (per KYr) | |
|---|---|---|---|---|---|---|---|---|
| | T | C | T | C | T | C | T | C |
| **PS-matching / on-treatment TAR** | | | | | | | | |
| CCAE | 41,136 | 73,891 | 34.9 | 60.8 | 1,297 | 2,567 | 37.2 (35.2 - 39.3) | 42.2 (40.6 - 43.9) |
| DAG | 11,877 | 2,119 | 12.4 | 2.1 | 281 | 58 | 22.6 (20.0 - 25.4) | 27.4 (20.8 - 35.4) |
| ODOD | 23,587 | 57,021 | 19.0 | 49.9 | 1,128 | 3,813 | 59.5 (56.1 - 63.1) | 76.4 (74.0 - 78.8) |
| MDCR | 6,975 | 14,141 | 6.9 | 13.9 | 563 | 1,213 | 82.0 (75.4 - 89.1) | 87.5 (82.6 - 92.5) |
| MDCD | 2,950 | 7,106 | 2.1 | 4.7 | 165 | 426 | 79.8 (68.0 - 92.9) | 90.0 (81.6 - 99.0) |
| Open Claims | 344,503 | 741,086 | 321.0 | 779.2 | 9,680 | 26,509 | 30.2 (29.6 - 30.8) | 34.0 (33.6 - 34.4) |
| OEHR | 31,993 | 83,626 | 15.2 | 43.5 | 600 | 2,420 | 39.4 (36.3 - 42.7) | 55.6 (53.4 - 57.9) |
| VA | 0 | 0 | 0.0 | 0.0 | 0 | 0 | | |
| SIDIAP | 24,872 | 6,927 | 71.8 | 19.3 | 1,087 | 241 | 15.1 (14.3 - 16.1) | 12.5 (11.0 - 14.2) |
| Meta-analysis | 487,893 | 985,917 | 483.3 | 973.5 | 14,801 | 37,247 | 30.6 (30.1 - 31.1) | 38.3 (37.9 - 38.7) |
| Meta-analysis (w/o USOC) | 143,390 | 244,831 | 162.3 | 194.3 | 5,121 | 10,738 | 31.6 (30.7 - 32.4) | 55.3 (54.2 - 56.3) |
| **PS-stratification / on-treatment TAR** | | | | | | | | |
| CCAE | 45,839 | 75,314 | 39.0 | 62.0 | 1,443 | 2,594 | 37.0 (35.1 - 38.9) | 41.9 (40.3 - 43.5) |
| DAG | 11,913 | 2,451 | 12.5 | 2.4 | 281 | 72 | 22.5 (20.0 - 25.3) | 29.4 (23.0 - 37.1) |
| ODOD | 24,999 | 57,024 | 20.2 | 49.9 | 1,204 | 3,814 | 59.6 (56.3 - 63.1) | 76.4 (74.0 - 78.8) |
| MDCR | 7,379 | 14,485 | 7.3 | 14.3 | 597 | 1,238 | 81.7 (75.3 - 88.6) | 86.8 (82.0 - 91.8) |
| MDCD | 3,021 | 7,122 | 2.1 | 4.7 | 171 | 426 | 80.4 (68.8 - 93.4) | 89.9 (81.5 - 98.8) |
| Open Claims | 350,809 | 741,094 | 328.7 | 779.2 | 9,964 | 26,510 | 30.3 (29.7 - 30.9) | 34.0 (33.6 - 34.4) |
| OEHR | 33,646 | 83,671 | 16.2 | 43.6 | 637 | 2,420 | 39.4 (36.4 - 42.5) | 55.6 (53.4 - 57.8) |
| VA | 20,139 | 190,948 | 18.4 | 193.3 | 620 | 7,649 | 33.7 (31.1 - 36.5) | 39.6 (38.7 - 40.5) |
| SIDIAP | 24,956 | 8,914 | 71.9 | 26.1 | 1,089 | 305 | 15.1 (14.3 - 16.1) | 11.7 (10.4 - 13.1) |
| Meta-analysis | 502,562 | 990,075 | 497.9 | 982.3 | 15,386 | 37,379 | 30.9 (30.4 - 31.4) | 38.1 (37.7 - 38.4) |
| Meta-analysis (w/o USOC) | 151,753 | 248,981 | 169.2 | 203.0 | 5,422 | 10,869 | 32.0 (31.2 - 32.9) | 53.5 (52.5 - 54.6) |
| **PS-matching / on-treatment TAR without escalation** | | | | | | | | |
| CCAE | 41,136 | 73,891 | 28.4 | 53.3 | 1,044 | 2,254 | 36.7 (34.5 - 39.0) | 42.3 (40.6 - 44.1) |
| DAG | 11,877 | 2,119 | 11.2 | 1.9 | 265 | 57 | 23.6 (20.8 - 26.6) | 29.3 (22.2 - 38.0) |
| ODOD | 23,587 | 57,021 | 15.7 | 44.9 | 931 | 3,391 | 59.2 (55.5 - 63.1) | 75.5 (73.0 - 78.1) |
| MDCR | 6,975 | 14,141 | 5.9 | 12.7 | 473 | 1,109 | 79.8 (72.8 - 87.4) | 87.0 (82.0 - 92.3) |
| MDCD | 2,950 | 7,106 | 1.6 | 4.0 | 138 | 361 | 86.5 (72.6 - 102) | 89.5 (80.5 - 99.3) |
| Open Claims | 344,503 | 741,086 | 250.2 | 664.6 | 7,527 | 22,576 | 30.1 (29.4 - 30.8) | 34.0 (33.5 - 34.4) |
| OEHR | 31,993 | 83,626 | 12.5 | 38.4 | 442 | 1,940 | 35.3 (32.1 - 38.7) | 50.5 (48.3 - 52.8) |
| VA | 0 | 0 | 0.0 | 0.0 | 0 | 0 | | |
| SIDIAP | 24,872 | 6,927 | 52.8 | 16.2 | 792 | 192 | 15.0 (14.0 - 16.1) | 11.9 (10.2 - 13.7) |
| Meta-analysis | 487,893 | 985,917 | 378.3 | 836.1 | 11,612 | 31,880 | 30.7 (30.1 - 31.3) | 38.1 (37.7 - 38.6) |
| Meta-analysis (w/o USOC) | 143,390 | 244,831 | 128.2 | 171.5 | 4,085 | 9,304 | 31.9 (30.9 - 32.9) | 54.3 (53.2 - 55.4) |
| **PS-stratification / on-treatment TAR without escalation** | | | | | | | | |
| CCAE | 45,839 | 75,314 | 31.8 | 54.4 | 1,159 | 2,279 | 36.4 (34.3 - 38.6) | 41.9 (40.2 - 43.7) |
| DAG | 11,913 | 2,451 | 11.3 | 2.3 | 265 | 70 | 23.5 (20.8 - 26.5) | 31.0 (24.2 - 39.2) |
| ODOD | 24,999 | 57,024 | 16.7 | 44.9 | 994 | 3,392 | 59.5 (55.9 - 63.3) | 75.5 (73.0 - 78.1) |
| MDCR | 7,379 | 14,485 | 6.3 | 13.1 | 502 | 1,132 | 79.6 (72.8 - 86.9) | 86.2 (81.3 - 91.4) |
| MDCD | 3,021 | 7,122 | 1.6 | 4.0 | 143 | 361 | 87.5 (73.8 - 103) | 89.4 (80.4 - 99.1) |
| Open Claims | 350,809 | 741,094 | 256.0 | 664.6 | 7,757 | 22,577 | 30.3 (29.6 - 31.0) | 34.0 (33.5 - 34.4) |
| OEHR | 33,646 | 83,671 | 13.3 | 38.4 | 467 | 1,940 | 35.0 (31.9 - 38.4) | 50.5 (48.3 - 52.8) |
| VA | 20,139 | 190,948 | 15.0 | 173.7 | 491 | 6,668 | 32.8 (29.9 - 35.8) | 38.4 (37.5 - 39.3) |
| SIDIAP | 24,956 | 8,914 | 52.8 | 21.6 | 794 | 246 | 15.0 (14.0 - 16.1) | 11.4 (9.99 - 12.9) |
| Meta-analysis | 502,562 | 990,075 | 389.9 | 843.4 | 12,081 | 31,997 | 31.0 (30.4 - 31.5) | 37.9 (37.5 - 38.4) |
| Meta-analysis (w/o USOC) | 151,753 | 248,981 | 133.9 | 178.8 | 4,324 | 9,420 | 32.3 (31.3 - 33.3) | 52.7 (51.6 - 53.8) |

##### 4.6.24 Photosensitivity

**Supplementary Table 443:** Exposures, outcomes and risk estimates for Photosensitivity in DPP4I (male) (T) vs SU (male) (C) new-user cohorts under four different propensity score (PS) adjustment / time-at-risk (TAR) designs across data sources. We report total patient exposure time in 1,000 patient-year (KYr) units, uncalibrated and calibrated hazard ratios (HRs) and their 95% confidence intervals (CIs). Greyed out entries did not pass study diagnostics and are included only for completeness.

| | Patients | | Exposure (KYr) | | Outcomes | | Incidence (per KYr) | |
|---|---|---|---|---|---|---|---|---|
| | T | C | T | C | T | C | T | C |
| **PS-matching / on-treatment TAR** | | | | | | | | |
| CCAE | 47,032 | 84,575 | 41.1 | 72.1 | 53 | 95 | 1.29 (0.97 - 1.69) | 1.32 (1.07 - 1.61) |
| DAG | 13,381 | 2,353 | 14.3 | 2.4 | 5 | <5 | 0.35 (0.11 - 0.82) | < 2.06 (0.67 - 4.80) |
| ODOD | 28,904 | 70,090 | 24.2 | 65.7 | 44 | 120 | 1.82 (1.32 - 2.44) | 1.83 (1.51 - 2.18) |
| MDCR | 8,842 | 17,915 | 9.2 | 18.8 | 43 | 75 | 4.70 (3.40 - 6.33) | 3.99 (3.14 - 5.01) |
| MDCD | 4,062 | 9,815 | 3.1 | 7.1 | <5 | <5 | < 1.62 (0.53 - 3.78) | < 0.71 (0.23 - 1.65) |
| Open Claims | 418,047 | 895,307 | 399.0 | 970.9 | 213 | 484 | 0.53 (0.46 - 0.61) | 0.50 (0.46 - 0.54) |
| OEHR | 36,240 | 95,230 | 18.0 | 52.9 | 23 | 110 | 1.28 (0.81 - 1.92) | 2.08 (1.71 - 2.51) |
| VA | 0 | 0 | 0.0 | 0.0 | 0 | 0 | | |
| SIDIAP | 26,751 | 7,209 | 78.4 | 20.5 | 7 | 9 | 0.09 (0.04 - 0.18) | 0.44 (0.20 - 0.83) |
| Meta-analysis | 539,065 | 1,163,117 | 491.4 | 1180.4 | 376 | 884 | 0.77 (0.69 - 0.85) | 0.75 (0.70 - 0.80) |
| Meta-analysis (w/o USOC) | 121,018 | 267,810 | 92.5 | 209.5 | 163 | 400 | 1.76 (1.50 - 2.06) | 1.91 (1.73 - 2.11) |
| **PS-stratification / on-treatment TAR** | | | | | | | | |
| CCAE | 52,413 | 86,087 | 46.0 | 73.4 | 65 | 96 | 1.41 (1.09 - 1.80) | 1.31 (1.06 - 1.60) |
| DAG | 13,408 | 2,730 | 14.3 | 2.8 | 5 | <5 | 0.35 (0.11 - 0.82) | < 1.76 (0.57 - 4.12) |
| ODOD | 30,504 | 70,097 | 25.7 | 65.7 | 51 | 120 | 1.98 (1.48 - 2.61) | 1.83 (1.51 - 2.18) |
| MDCR | 9,340 | 18,307 | 9.7 | 19.3 | 47 | 77 | 4.83 (3.55 - 6.42) | 4.00 (3.16 - 5.00) |
| MDCD | 4,156 | 9,835 | 3.2 | 7.1 | <5 | <5 | < 1.57 (0.51 - 3.67) | < 0.71 (0.23 - 1.65) |
| Open Claims | 425,304 | 895,320 | 408.1 | 970.9 | 221 | 484 | 0.54 (0.47 - 0.62) | 0.50 (0.46 - 0.54) |
| OEHR | 38,071 | 95,260 | 19.1 | 52.9 | 23 | 110 | 1.21 (0.76 - 1.81) | 2.08 (1.71 - 2.51) |
| VA | 25,264 | 231,142 | 23.7 | 245.5 | <10 | 367 | < 0.42 (0.20 - 0.78) | 1.49 (1.35 - 1.66) |
| SIDIAP | 26,844 | 9,260 | 78.5 | 27.6 | 7 | 11 | 0.09 (0.04 - 0.18) | 0.40 (0.20 - 0.71) |
| Meta-analysis | 555,632 | 1,165,071 | 508.6 | 1182.2 | 407 | 887 | 0.80 (0.72 - 0.88) | 0.75 (0.70 - 0.80) |
| Meta-analysis (w/o USOC) | 130,328 | 269,751 | 100.5 | 211.2 | 186 | 403 | 1.85 (1.59 - 2.14) | 1.91 (1.73 - 2.10) |
| **PS-matching / on-treatment TAR without escalation** | | | | | | | | |
| CCAE | 47,032 | 84,575 | 33.2 | 62.7 | 43 | 88 | 1.30 (0.94 - 1.75) | 1.40 (1.13 - 1.73) |
| DAG | 13,381 | 2,353 | 12.9 | 2.2 | 5 | <5 | 0.39 (0.13 - 0.91) | < 2.26 (0.73 - 5.26) |
| ODOD | 28,904 | 70,090 | 19.8 | 58.5 | 37 | 112 | 1.87 (1.31 - 2.57) | 1.91 (1.58 - 2.30) |
| MDCR | 8,842 | 17,915 | 7.8 | 17.1 | 36 | 70 | 4.61 (3.23 - 6.39) | 4.08 (3.18 - 5.16) |
| MDCD | 4,062 | 9,815 | 2.4 | 5.9 | <5 | <5 | < 2.13 (0.69 - 4.96) | < 0.84 (0.27 - 1.97) |
| Open Claims | 418,047 | 895,307 | 309.3 | 822.1 | 180 | 443 | 0.58 (0.50 - 0.67) | 0.54 (0.49 - 0.59) |
| OEHR | 36,240 | 95,230 | 14.6 | 45.9 | 20 | 98 | 1.37 (0.84 - 2.12) | 2.14 (1.73 - 2.60) |
| VA | 0 | 0 | 0.0 | 0.0 | 0 | 0 | | |
| SIDIAP | 26,751 | 7,209 | 57.4 | 17.1 | 6 | 7 | 0.10 (0.04 - 0.23) | 0.41 (0.16 - 0.84) |
| Meta-analysis | 539,065 | 1,163,117 | 384.7 | 1006.3 | 316 | 811 | 0.82 (0.73 - 0.92) | 0.81 (0.75 - 0.86) |
| Meta-analysis (w/o USOC) | 121,018 | 267,810 | 75.4 | 184.2 | 136 | 368 | 1.80 (1.51 - 2.13) | 2.00 (1.80 - 2.21) |
| **PS-stratification / on-treatment TAR without escalation** | | | | | | | | |
| CCAE | 52,413 | 86,087 | 37.2 | 63.8 | 54 | 89 | 1.45 (1.09 - 1.90) | 1.39 (1.12 - 1.72) |
| DAG | 13,408 | 2,730 | 12.9 | 2.6 | 5 | <5 | 0.39 (0.13 - 0.91) | < 1.93 (0.63 - 4.50) |
| ODOD | 30,504 | 70,097 | 21.0 | 58.5 | 43 | 112 | 2.05 (1.48 - 2.76) | 1.91 (1.58 - 2.30) |
| MDCR | 9,340 | 18,307 | 8.3 | 17.6 | 40 | 72 | 4.81 (3.44 - 6.56) | 4.09 (3.20 - 5.15) |
| MDCD | 4,156 | 9,835 | 2.4 | 5.9 | <5 | <5 | < 2.07 (0.67 - 4.84) | < 0.84 (0.27 - 1.97) |
| Open Claims | 425,304 | 895,320 | 316.2 | 822.1 | 187 | 443 | 0.59 (0.51 - 0.68) | 0.54 (0.49 - 0.59) |
| OEHR | 38,071 | 95,260 | 15.5 | 45.9 | 20 | 98 | 1.29 (0.79 - 2.00) | 2.13 (1.73 - 2.60) |
| VA | 25,264 | 231,142 | 19.2 | 218.6 | <10 | 336 | < 0.52 (0.25 - 0.96) | 1.54 (1.38 - 1.71) |
| SIDIAP | 26,844 | 9,260 | 57.5 | 22.8 | 6 | 8 | 0.10 (0.04 - 0.23) | 0.35 (0.15 - 0.69) |
| Meta-analysis | 555,632 | 1,165,071 | 398.1 | 1008.0 | 344 | 814 | 0.86 (0.78 - 0.96) | 0.81 (0.75 - 0.87) |
| Meta-analysis (w/o USOC) | 130,328 | 269,751 | 82.0 | 185.9 | 157 | 371 | 1.92 (1.63 - 2.24) | 2.00 (1.80 - 2.21) |

##### 4.6.25 Venous thromboembolism

**Supplementary Table 444:** Exposures, outcomes and risk estimates for Venous thromboembolism in DPP4I (male) (T) vs SU (male) (C) new-user cohorts under four different propensity score (PS) adjustment / time-at-risk (TAR) designs across data sources. We report total patient exposure time in 1,000 patient-year (KYr) units, uncalibrated and calibrated hazard ratios (HRs) and their 95% confidence intervals (CIs). Greyed out entries did not pass study diagnostics and are included only for completeness.

| | Patients | | Exposure (KYr) | | Outcomes | | Incidence (per KYr) | |
|---|---|---|---|---|---|---|---|---|
| | T | C | T | C | T | C | T | C |
| **PS-matching / on-treatment TAR** | | | | | | | | |
| CCAE | 46,500 | 83,393 | 40.6 | 70.9 | 184 | 427 | 4.53 (3.90 - 5.24) | 6.02 (5.46 - 6.62) |
| DAG | 12,703 | 2,266 | 13.4 | 2.3 | 90 | 24 | 6.69 (5.38 - 8.23) | 10.3 (6.61 - 15.3) |
| ODOD | 28,387 | 68,587 | 23.8 | 63.9 | 214 | 787 | 9.01 (7.84 - 10.3) | 12.3 (11.5 - 13.2) |
| MDCR | 8,678 | 17,475 | 9.0 | 18.3 | 125 | 293 | 13.9 (11.6 - 16.6) | 16.0 (14.2 - 18.0) |
| MDCD | 3,893 | 9,404 | 3.0 | 6.8 | 44 | 71 | 14.9 (10.8 - 19.9) | 10.5 (8.20 - 13.3) |
| Open Claims | 408,514 | 873,694 | 389.4 | 945.3 | 1,917 | 5,766 | 4.92 (4.71 - 5.15) | 6.10 (5.94 - 6.26) |
| OEHR | 35,569 | 93,391 | 17.5 | 51.3 | 120 | 499 | 6.85 (5.68 - 8.19) | 9.72 (8.89 - 10.6) |
| VA | 0 | 0 | 0.0 | 0.0 | 0 | 0 | | |
| SIDIAP | 26,225 | 7,125 | 76.6 | 20.2 | 275 | 61 | 3.59 (3.18 - 4.04) | 3.02 (2.31 - 3.88) |
| Meta-analysis | 570,469 | 1,155,335 | 573.2 | 1178.9 | 2,969 | 7,928 | 5.18 (4.99 - 5.37) | 6.72 (6.58 - 6.87) |
| Meta-analysis (w/o USOC) | 161,955 | 281,641 | 183.9 | 233.7 | 1,052 | 2,162 | 5.72 (5.38 - 6.08) | 9.25 (8.87 - 9.65) |
| **PS-stratification / on-treatment TAR** | | | | | | | | |
| CCAE | 51,836 | 84,899 | 45.4 | 72.2 | 204 | 430 | 4.49 (3.89 - 5.15) | 5.96 (5.41 - 6.55) |
| DAG | 12,728 | 2,625 | 13.5 | 2.7 | 91 | 28 | 6.76 (5.44 - 8.29) | 10.3 (6.86 - 14.9) |
| ODOD | 29,968 | 68,594 | 25.2 | 63.9 | 223 | 787 | 8.83 (7.71 - 10.1) | 12.3 (11.5 - 13.2) |
| MDCR | 9,176 | 17,858 | 9.6 | 18.7 | 128 | 298 | 13.4 (11.2 - 15.9) | 15.9 (14.1 - 17.8) |
| MDCD | 3,976 | 9,424 | 3.0 | 6.8 | 45 | 71 | 14.8 (10.8 - 19.8) | 10.5 (8.19 - 13.2) |
| Open Claims | 415,805 | 873,708 | 398.6 | 945.3 | 1,959 | 5,766 | 4.92 (4.70 - 5.14) | 6.10 (5.94 - 6.26) |
| OEHR | 37,345 | 93,445 | 18.6 | 51.4 | 124 | 501 | 6.67 (5.55 - 7.96) | 9.75 (8.92 - 10.6) |
| VA | 24,743 | 227,637 | 23.1 | 241.0 | 103 | 1,296 | 4.46 (3.64 - 5.40) | 5.38 (5.09 - 5.68) |
| SIDIAP | 26,327 | 9,159 | 76.7 | 27.2 | 276 | 77 | 3.60 (3.19 - 4.05) | 2.83 (2.23 - 3.53) |
| Meta-analysis | 587,161 | 1,159,712 | 590.6 | 1188.1 | 3,050 | 7,958 | 5.16 (4.98 - 5.35) | 6.70 (6.55 - 6.85) |
| Meta-analysis (w/o USOC) | 171,356 | 286,004 | 192.1 | 242.9 | 1,091 | 2,192 | 5.68 (5.35 - 6.03) | 9.03 (8.65 - 9.41) |
| **PS-matching / on-treatment TAR without escalation** | | | | | | | | |
| CCAE | 46,500 | 83,393 | 32.8 | 61.7 | 154 | 358 | 4.70 (3.99 - 5.50) | 5.80 (5.22 - 6.43) |
| DAG | 12,703 | 2,266 | 12.1 | 2.1 | 85 | 23 | 7.01 (5.60 - 8.67) | 10.8 (6.87 - 16.3) |
| ODOD | 28,387 | 68,587 | 19.4 | 57.0 | 177 | 691 | 9.10 (7.81 - 10.5) | 12.1 (11.2 - 13.1) |
| MDCR | 8,678 | 17,475 | 7.7 | 16.7 | 103 | 264 | 13.4 (11.0 - 16.3) | 15.8 (14.0 - 17.9) |
| MDCD | 3,893 | 9,404 | 2.3 | 5.7 | 36 | 57 | 15.9 (11.2 - 22.1) | 10.1 (7.63 - 13.1) |
| Open Claims | 408,514 | 873,694 | 302.2 | 801.1 | 1,485 | 4,927 | 4.91 (4.67 - 5.17) | 6.15 (5.98 - 6.32) |
| OEHR | 35,569 | 93,391 | 14.2 | 44.7 | 92 | 369 | 6.46 (5.21 - 7.93) | 8.26 (7.43 - 9.14) |
| VA | 0 | 0 | 0.0 | 0.0 | 0 | 0 | | |
| SIDIAP | 26,225 | 7,125 | 56.2 | 16.9 | 180 | 49 | 3.20 (2.75 - 3.71) | 2.90 (2.15 - 3.84) |
| Meta-analysis | 570,469 | 1,155,335 | 446.8 | 1005.8 | 2,312 | 6,738 | 5.17 (4.97 - 5.39) | 6.70 (6.54 - 6.86) |
| Meta-analysis (w/o USOC) | 161,955 | 281,641 | 144.7 | 204.7 | 827 | 1,811 | 5.72 (5.33 - 6.12) | 8.85 (8.44 - 9.26) |
| **PS-stratification / on-treatment TAR without escalation** | | | | | | | | |
| CCAE | 51,836 | 84,899 | 36.7 | 62.9 | 168 | 361 | 4.57 (3.91 - 5.32) | 5.74 (5.16 - 6.37) |
| DAG | 12,728 | 2,625 | 12.1 | 2.5 | 86 | 27 | 7.08 (5.66 - 8.74) | 10.9 (7.18 - 15.8) |
| ODOD | 29,968 | 68,594 | 20.6 | 57.0 | 183 | 691 | 8.88 (7.64 - 10.3) | 12.1 (11.2 - 13.1) |
| MDCR | 9,176 | 17,858 | 8.2 | 17.1 | 105 | 269 | 12.9 (10.5 - 15.6) | 15.7 (13.9 - 17.7) |
| MDCD | 3,976 | 9,424 | 2.3 | 5.7 | 37 | 57 | 16.0 (11.3 - 22.1) | 10.1 (7.61 - 13.0) |
| Open Claims | 415,805 | 873,708 | 309.1 | 801.1 | 1,522 | 4,927 | 4.92 (4.68 - 5.18) | 6.15 (5.98 - 6.32) |
| OEHR | 37,345 | 93,445 | 15.1 | 44.7 | 96 | 370 | 6.36 (5.15 - 7.76) | 8.27 (7.45 - 9.16) |
| VA | 24,743 | 227,637 | 18.8 | 214.8 | 74 | 1,065 | 3.94 (3.09 - 4.94) | 4.96 (4.66 - 5.26) |
| SIDIAP | 26,327 | 9,159 | 56.3 | 22.5 | 180 | 59 | 3.20 (2.75 - 3.70) | 2.62 (1.99 - 3.38) |
| Meta-analysis | 587,161 | 1,159,712 | 460.5 | 1013.5 | 2,377 | 6,761 | 5.16 (4.96 - 5.37) | 6.67 (6.51 - 6.83) |
| Meta-analysis (w/o USOC) | 171,356 | 286,004 | 151.4 | 212.4 | 855 | 1,834 | 5.65 (5.28 - 6.04) | 8.64 (8.24 - 9.04) |

##### 4.6.26 Vomiting

**Supplementary Table 445:** Exposures, outcomes and risk estimates for Vomiting in DPP4I (male) (T) vs SU (male) (C) new-user cohorts under four different propensity score (PS) adjustment / time-at-risk (TAR) designs across data sources. We report total patient exposure time in 1,000 patient-year (KYr) units, uncalibrated and calibrated hazard ratios (HRs) and their 95% confidence intervals (CIs). Greyed out entries did not pass study diagnostics and are included only for completeness.

| | Patients | | Exposure (KYr) | | Outcomes | | Incidence (per KYr) | |
|---|---|---|---|---|---|---|---|---|
| | T | C | T | C | T | C | T | C |
| **PS-matching / on-treatment TAR** | | | | | | | | |
| CCAE | 44,822 | 80,290 | 39.0 | 68.2 | 634 | 1,015 | 16.2 (15.0 - 17.6) | 14.9 (14.0 - 15.8) |
| DAG | 12,643 | 2,255 | 13.4 | 2.3 | 116 | 25 | 8.65 (7.15 - 10.4) | 11.0 (7.11 - 16.2) |
| ODOD | 27,096 | 65,489 | 22.4 | 60.9 | 519 | 1,409 | 23.1 (21.2 - 25.2) | 23.1 (21.9 - 24.4) |
| MDCR | 8,485 | 16,955 | 8.7 | 17.7 | 190 | 460 | 21.7 (18.8 - 25.1) | 26.0 (23.7 - 28.5) |
| MDCD | 3,406 | 8,130 | 2.6 | 5.8 | 111 | 279 | 43.4 (35.7 - 52.2) | 48.2 (42.7 - 54.2) |
| Open Claims | 391,579 | 836,022 | 372.6 | 909.3 | 4,488 | 10,879 | 12.0 (11.7 - 12.4) | 12.0 (11.7 - 12.2) |
| OEHR | 34,905 | 91,536 | 17.1 | 50.0 | 282 | 954 | 16.5 (14.7 - 18.6) | 19.1 (17.9 - 20.3) |
| VA | 0 | 0 | 0.0 | 0.0 | 0 | 0 | | |
| SIDIAP | 26,125 | 7,109 | 76.3 | 20.1 | 368 | 83 | 4.82 (4.34 - 5.34) | 4.13 (3.29 - 5.13) |
| Meta-analysis | 549,061 | 1,107,786 | 552.2 | 1134.3 | 6,708 | 15,104 | 12.1 (11.9 - 12.4) | 13.3 (13.1 - 13.5) |
| Meta-analysis (w/o USOC) | 157,482 | 271,764 | 179.5 | 225.0 | 2,220 | 4,225 | 12.4 (11.9 - 12.9) | 18.8 (18.2 - 19.4) |
| **PS-stratification / on-treatment TAR** | | | | | | | | |
| CCAE | 50,059 | 81,748 | 43.6 | 69.5 | 717 | 1,031 | 16.4 (15.2 - 17.7) | 14.8 (14.0 - 15.8) |
| DAG | 12,669 | 2,608 | 13.4 | 2.7 | 116 | 31 | 8.63 (7.13 - 10.4) | 11.7 (7.92 - 16.5) |
| ODOD | 28,618 | 65,496 | 23.8 | 60.9 | 555 | 1,409 | 23.3 (21.4 - 25.3) | 23.1 (21.9 - 24.4) |
| MDCR | 8,966 | 17,423 | 9.3 | 18.3 | 202 | 467 | 21.7 (18.8 - 24.9) | 25.6 (23.3 - 28.0) |
| MDCD | 3,484 | 8,165 | 2.6 | 5.8 | 117 | 280 | 44.7 (37.0 - 53.6) | 48.1 (42.6 - 54.1) |
| Open Claims | 398,740 | 836,034 | 381.7 | 909.3 | 4,622 | 10,880 | 12.1 (11.8 - 12.5) | 12.0 (11.7 - 12.2) |
| OEHR | 36,697 | 91,564 | 18.1 | 50.0 | 295 | 954 | 16.3 (14.5 - 18.2) | 19.1 (17.9 - 20.3) |
| VA | 24,357 | 224,001 | 22.9 | 237.7 | 156 | 1,701 | 6.82 (5.79 - 7.98) | 7.16 (6.82 - 7.50) |
| SIDIAP | 26,215 | 9,149 | 76.4 | 27.1 | 368 | 114 | 4.82 (4.34 - 5.34) | 4.21 (3.47 - 5.05) |
| Meta-analysis | 565,448 | 1,112,187 | 569.0 | 1143.5 | 6,992 | 15,166 | 12.3 (12.0 - 12.6) | 13.3 (13.1 - 13.5) |
| Meta-analysis (w/o USOC) | 166,708 | 276,153 | 187.3 | 234.2 | 2,370 | 4,286 | 12.7 (12.1 - 13.2) | 18.3 (17.8 - 18.9) |
| **PS-matching / on-treatment TAR without escalation** | | | | | | | | |
| CCAE | 44,822 | 80,290 | 31.6 | 59.5 | 527 | 891 | 16.7 (15.3 - 18.2) | 15.0 (14.0 - 16.0) |
| DAG | 12,643 | 2,255 | 12.1 | 2.1 | 105 | 24 | 8.67 (7.09 - 10.5) | 11.5 (7.40 - 17.2) |
| ODOD | 27,096 | 65,489 | 18.5 | 54.4 | 435 | 1,234 | 23.6 (21.4 - 25.9) | 22.7 (21.4 - 24.0) |
| MDCR | 8,485 | 16,955 | 7.5 | 16.2 | 164 | 404 | 22.0 (18.8 - 25.6) | 25.0 (22.6 - 27.5) |
| MDCD | 3,406 | 8,130 | 2.0 | 4.9 | 94 | 239 | 48.0 (38.8 - 58.8) | 48.8 (42.8 - 55.4) |
| Open Claims | 391,579 | 836,022 | 289.9 | 772.3 | 3,390 | 8,989 | 11.7 (11.3 - 12.1) | 11.6 (11.4 - 11.9) |
| OEHR | 34,905 | 91,536 | 13.9 | 43.7 | 189 | 718 | 13.6 (11.7 - 15.7) | 16.4 (15.2 - 17.7) |
| VA | 0 | 0 | 0.0 | 0.0 | 0 | 0 | | |
| SIDIAP | 26,125 | 7,109 | 55.9 | 16.8 | 280 | 67 | 5.01 (4.44 - 5.63) | 4.00 (3.10 - 5.08) |
| Meta-analysis | 549,061 | 1,107,786 | 431.3 | 969.8 | 5,184 | 12,566 | 12.0 (11.7 - 12.4) | 13.0 (12.7 - 13.2) |
| Meta-analysis (w/o USOC) | 157,482 | 271,764 | 141.4 | 197.5 | 1,794 | 3,577 | 12.7 (12.1 - 13.3) | 18.1 (17.5 - 18.7) |
| **PS-stratification / on-treatment TAR without escalation** | | | | | | | | |
| CCAE | 50,059 | 81,748 | 35.4 | 60.6 | 593 | 907 | 16.8 (15.4 - 18.2) | 15.0 (14.0 - 16.0) |
| DAG | 12,669 | 2,608 | 12.1 | 2.4 | 105 | 30 | 8.65 (7.07 - 10.5) | 12.3 (8.30 - 17.6) |
| ODOD | 28,618 | 65,496 | 19.6 | 54.4 | 466 | 1,234 | 23.8 (21.7 - 26.1) | 22.7 (21.4 - 24.0) |
| MDCR | 8,966 | 17,423 | 7.9 | 16.7 | 172 | 411 | 21.7 (18.5 - 25.1) | 24.6 (22.3 - 27.1) |
| MDCD | 3,484 | 8,165 | 2.0 | 4.9 | 98 | 240 | 48.9 (39.7 - 59.6) | 48.6 (42.7 - 55.2) |
| Open Claims | 398,740 | 836,034 | 296.7 | 772.3 | 3,494 | 8,990 | 11.8 (11.4 - 12.2) | 11.6 (11.4 - 11.9) |
| OEHR | 36,697 | 91,564 | 14.8 | 43.7 | 198 | 718 | 13.4 (11.6 - 15.4) | 16.4 (15.2 - 17.7) |
| VA | 24,357 | 224,001 | 18.6 | 212.1 | 128 | 1,427 | 6.89 (5.74 - 8.19) | 6.73 (6.38 - 7.09) |
| SIDIAP | 26,215 | 9,149 | 56.0 | 22.4 | 280 | 87 | 5.00 (4.43 - 5.62) | 3.88 (3.11 - 4.78) |
| Meta-analysis | 565,448 | 1,112,187 | 444.6 | 977.6 | 5,406 | 12,617 | 12.2 (11.8 - 12.5) | 12.9 (12.7 - 13.1) |
| Meta-analysis (w/o USOC) | 166,708 | 276,153 | 147.8 | 205.3 | 1,912 | 3,627 | 12.9 (12.4 - 13.5) | 17.7 (17.1 - 18.3) |
